# Characteristics that modify the effect of small-quantity lipid-based nutrient supplementation on child anemia and micronutrient status: an individual participant data meta-analysis of randomized controlled trials

**DOI:** 10.1101/2021.02.08.21251234

**Authors:** K. Ryan Wessells, Charles D. Arnold, Christine P. Stewart, Elizabeth L. Prado, Souheila Abbeddou, Seth Adu-Afarwuah, Benjamin F. Arnold, Per Ashorn, Ulla Ashorn, Elodie Becquey, Kenneth H. Brown, Kendra A. Byrd, Rebecca K. Campbell, Parul Christian, Lia C.H. Fernald, Yue-Mei Fan, Emanuela Galasso, Sonja Y. Hess, Lieven Huybregts, Josh M. Jorgensen, Marion Kiprotich, Emma Kortekangas, Anna Lartey, Agnes Le Port, Jef L. Leroy, Audrie Lin, Kenneth Maleta, Susana L. Matias, Mduduzi NN Mbuya, Malay K. Mridha, Kuda Mutasa, Abu Mohd. Naser, Rina R. Paul, Harriet Okronipa, Jean-Bosco Ouédraogo, Amy J. Pickering, Mahbubur Rahman, Kerry Schulze, Laura E. Smith, Ann M. Weber, Amanda Zongrone, Kathryn G. Dewey

**Author notes:** **Corresponding Author:** K. Ryan Wessells, Department of Nutrition, University of California, One Shields Ave., Davis, CA 95616. Registry and registry number for systematic reviews or meta-analyses: Registered at www.crd.york.ac.uk/PROSPERO as CRD42020156663. Data described in the manuscript, code book, and analytic code will not be made available because they are compiled from 13 different trials, and access is under the control of the investigators of each of those trials. Parul Christian is a member of the Journal’s Editorial Board.

## Abstract

**Background:** Small-quantity lipid-based nutrient supplements (SQ-LNS) have been shown to reduce the prevalence of anemia and iron deficiency among infants and young children, but effects on other micronutrients are less well known. Identifying subgroups who may experience greater benefits from SQ-LNS, or who are more likely to respond to the intervention, may facilitate the development of public health policies and programs.

**Objective:** Our objective was to identify study-level and individual-level modifiers of the effect of SQ-LNS on child hematological and micronutrient status outcomes.

**Methods:** We conducted a two-stage meta-analysis of individual participant data from 13 randomized controlled trials of SQ-LNS provided to children 6 to 24 months of age in low- and middle-income countries (n = 15,946). Outcomes were hemoglobin (Hb), inflammation-adjusted plasma ferritin, soluble transferrin receptor, zinc, retinol and retinol binding protein (RBP), and erythrocyte zinc protoporphyrin, and respective dichotomous outcomes indicative of anemia and micronutrient deficiency. We generated study-specific estimates of SQ-LNS vs. control, including main effects and subgroup estimates for individual-level effect modifiers, and pooled the estimates using fixed-effects models. We used random effects meta-regression to examine potential study-level effect modifiers.

**Results:** Provision of SQ-LNS decreased the prevalence of anemia (Hb < 110 g/L) by 16% (relative reduction), iron deficiency (plasma ferritin < 12 µg/L) by 56% and iron deficiency anemia (IDA; Hb < 110 g/L and plasma ferritin < 12 µg/L) by 64%. We observed positive effects of SQ-LNS on hematological and iron status outcomes within all subgroups of the study-level and individual-level effect modifiers, but effects were larger in certain subgroups. For example, effects of SQ-LNS on anemia and iron status were greater in trials that provided SQ-LNS for > 12 months and provided 9 mg/d vs. < 9 mg iron/d, and among later-born (vs. first-born) children. There was no effect of SQ-LNS on plasma zinc or retinol, but there was a 7% increase in plasma RBP and a 56% reduction in vitamin A deficiency (RBP < 0.70 µmol/L), with little evidence of effect modification by individual-level characteristics.

**Conclusions:** SQ-LNS provided to infants and young children 6-24 months of age can substantially reduce the prevalence of anemia, iron deficiency, and IDA across a range of individual, population and study design characteristics. Policy-makers and program planners should consider SQ-LNS within intervention packages to prevent anemia and iron deficiency. This study was registered at www.crd.york.ac.uk/PROSPERO as CRD42020156663.

## Introduction

Micronutrient deficiencies are estimated to affect two billion people globally (1, 2) and roughly one-third of the world’s population is anemic (3). Infants and young children in low- and middle-income countries are particularly vulnerable to micronutrient undernutrition, owing in part to low micronutrient stores at birth, inadequate dietary intake of bioavailable micronutrients, and increased micronutrient requirements due to infection or malabsorption (4, 5). Deficiencies of iron, zinc, and vitamin A in these populations are associated with increased morbidity and mortality and delayed psychomotor and neurocognitive development (1).

Nutritional strategies to prevent micronutrient deficiencies include dietary diversification and modification, provision of supplements, fortification (i.e. large-scale food fortification, biofortification and targeted in-home fortification), and supplemental feeding (e.g., fortified blended foods and lipid-based nutrient supplements). Small-quantity lipid-based nutrient supplements (SQ-LNS) have been designed to prevent malnutrition and provide multiple micronutrients embedded in a food base that also provides energy (100-120 kcal/d), protein and essential fatty acids. This combination of macro- and micro-nutrients in SQ-LNS has the potential to address multiple nutritional deficiencies simultaneously, thus reducing undernutrition (6).

A 2019 Cochrane Systematic Review and meta-analysis showed that SQ- and medium-quantity LNS (MQ-LNS) (∼125 – 250 kcal/d) given during the period of complementary feeding (6-24 months) significantly reduced the prevalence of anemia by 21% (RR 0.79, 95% CI 0.69 to 0.90; 5 studies; anemia as defined by trialists) compared to no intervention (7). Similarly, a 2020 meta-analysis demonstrated a 16% reduction in the prevalence of anemia (RR 0.84, 95% CI 0.75, 0.93; 8 studies; anemia as defined by trialists) among children who received SQ- or MQ-LNS in comparison to a control group (8). To-date, no meta-analysis has assessed the impact of SQ-LNS provided during the period of complementary feeding on the micronutrient status of infants and young children. However, individual studies have demonstrated significant reductions in the prevalence of iron deficiency and iron deficiency anemia (9–16), although there is heterogeneity in the magnitude of effects. Some, but not all, studies have shown significant improvements in vitamin A, B12 and folate status (11-13, 15, 17); no studies have reported an effect of SQ-LNS on zinc status (9, 11, 15).

Differences in study design and context and characteristics of study participants may modify the effect of SQ-LNS on anemia and biomarkers of micronutrient status. The identification of subgroups of infants and young children who experience greater benefits from SQ-LNS, or who are more likely to respond to the intervention, is useful in informing the development of public health policies and programs (18). Thus, we conducted an individual participant data (IPD) meta-analysis of randomized controlled trials (RCTs) of SQ-LNS provided to infants and young children 6 to 24 months of age. The objectives of this analysis were to (1) generate pooled estimates of the effects of SQ-LNS provided to infants and young children 6 to 24 months of age on hemoglobin concentration and selected biomarkers of micronutrient status, and (2) identify study- and individual-level modifiers of the effect of SQ-LNS on these outcomes in the same populations. Two companion articles report results for other outcome domains, growth (19) and development (20), from the same IPD meta-analysis.

## Methods

Methods broadly followed those presented in a companion article (19). This systematic review and IPD meta-analysis was pre-registered through PROSPERO (CRD42020156663) (21). A detailed protocol and statistical analysis plan were developed prior to analyses and are available online (22), and we have reported results according to PRISMA-IPD guidelines (23). The analyses were approved by the institutional review board of the University of California, Davis (1463609–1). All individual trial protocols were approved by their relevant institutional ethics committees.

### Inclusion and exclusion criteria for this systematic review and IPD meta-analysis

Trials were eligible for inclusion if they were individual or cluster RCTs with either longitudinal follow-up or repeated cross-sectional data collection, were conducted in low- or middle-income countries (24), provided SQ-LNS (< ∼125 kcal/d) to study participants for at least three months during any part of the age range between 6-24 months of age, and reported at least one outcome of interest.

We excluded trials in which SQ-LNS was used for the treatment, not prevention, of malnutrition (i.e., only children with moderate to severe malnutrition were eligible to participate), as well as studies specifically conducted in hospitalized populations or among children with a pre-existing disease. Trials were excluded if the only available comparison group received other types of non-LNS child supplementation (e.g., multiple micronutrient powder, fortified blended food) or if SQ-LNS provision was combined with an additional supplemental food or nutrients within a single arm (e.g., SQ-LNS + food rations vs. control), and there was no appropriate comparison group that would allow isolation of the SQ-LNS effect (e.g., food rations alone).

Trials in which there were multiple relevant SQ-LNS interventions (e.g., varying dosages or formulations of SQ-LNS in different arms), or that combined provision of SQ-LNS with other non-nutritional interventions (e.g., water, sanitation and hygiene (WASH)), were eligible fo inclusion. In such trials, all arms that provided SQ-LNS were combined into one group, and all non-LNS arms were combined into a single comparison group for each trial (herein labeled “Control”), excluding intervention arms that received non-LNS child supplementation (e.g., multiple micronutrient powder, fortified-blended food). We also conducted a sensitivity analysis restricting the comparison to specified contrasts of intervention arms within multiple intervention trials (see below).

Individual children were included in the analyses if their age at baseline allowed them to receive at least three months of intervention (supplementation or control group components) between 6 and 24 months of age, and if biochemical outcome assessment occurred within 3 months of the intended, trial-defined end of supplementation.

### Search methods and identification of studies

We identified potential studies for inclusion in the IPD analysis from a recent systematic review and meta-analysis of LNS (7). We then repeated the database search strategy employed by Das *et al.*(7), using the same keyword and controlled vocabulary search terms, to capture additional studies (both completed and on-going) indexed in one of 23 international or regional electronic databases and 2 trial registers between July 1, 2018 and May 1, 2019 (*see **Supplemental Methods***). One of the authors (KRW) reviewed the titles and abstracts of all studies included in the 2019 Cochrane review, as well as the additional studies identified by the repeated search strategy, to select all potentially relevant studies for full text review. The full-text reports of all potentially relevant records were reviewed. Trials were assessed against the inclusion and exclusion criteria detailed above. In September 2019, the same author reviewed the previously identified on-going studies that met inclusion and exclusion criteria to determine if results for outcomes of interest had been subsequently published.

### Data collection and harmonization

We invited all principal investigators of eligible studies to participate in this IPD meta-analysis. Individual investigators were asked to provide de-identified individual participant data (primary data) for pre-specified variables (defined above and detailed in a data dictionary provided to investigators). The IPD analyst (CDA) collected and managed the individual participant data, and communicated with investigators to request any missing variables or other information.

### IPD integrity

We conducted a complete-case, intention to treat analysis (25). We checked data for completeness by crosschecking sample sizes with study protocols and publications. We also crosschecked the data provided against reported values for each trial to ensure consistency. Variables were assessed for outliers and low frequency categories. Relevant model assumptions were assessed (e.g. Shapiro-Wilk normality testing and Breusch-Pagan heteroscedasticity testing) and outcome variables were appropriately transformed before subsequent analyses, as necessary.

### Assessment of risk of bias in each study and quality of evidence across studies

Two independent reviewers (KRW and CDA) assessed risk of bias using the criteria outlined in the Cochrane Handbook for Systematic Reviews of Interventions Version 5.1.0 (26). We assessed each trial against the following criteria: random sequence generation and allocation concealment (selection bias), blinding of participants and personnel (performance bias), blinding of outcome assessment (detection bias), incomplete outcome data (attrition bias), selective reporting (reporting bias), and other sources of bias. Reviewers also assessed the quality of evidence for each primary and secondary outcome across all trials based on the five GRADE criteria: risk of bias, inconsistency of effect, imprecision, indirectness and publication bias (27). Any discrepancies were resolved by discussion or consultation with the core working group, as needed.

### Specification of outcomes and effect measures

We specified outcomes *a priori* in the statistical analysis plan (22). Outcomes of interest are listed in **Box 1**. For the estimation of the main effects, we pre-specified hemoglobin concentration, anemia, moderate-to-severe anemia, and biomarkers of iron status as primary outcomes in the statistical analysis plan, and present results for all outcomes. Hemoglobin concentrations were adjusted for altitude, as necessary (28). Ferritin, sTfR, ZPP, zinc, retinol and RBP concentrations were adjusted for inflammation (i.e., C-reactive protein (CRP) and/or α-1-acid glycoprotein (AGP) concentrations, as available), using a regression correction approach adapted from the Biomarkers Reflecting Inflammation and Nutritional Determinants of Anemia (BRINDA) project and described in detail elsewhere (29); results are presented for inflammation-adjusted concentrations.

#### Box 1. Specification and definitions of outcome variables^1^

| **Outcome variables** | **Definitions of dichotomous outcome variables** |
| --- | --- |
| Hemoglobin (Hb; g/L) |  |
| Anemia | Hb < 110 g/L (80) |
| Moderate to severe anemia | Hb < 100 g/L (80) |
| Plasma ferritin (µg/L) |  |
| Iron deficiency (low plasma ferritin concentration) | Plasma ferritin < 12 µg/L (81) |
| Iron deficiency anemia | Plasma ferritin < 12 µg/L Hb < 110 g/L (28) |
| Plasma soluble transferrin receptor concentration (sTfR; mg/L) |  |
| Elevated plasma sTfR concentration | Plasma sTfR > 8.3 mg/L (82) |
| Erythrocyte zinc protoporphyrin concentration (ZPP; µmol/mol heme) |  |
| Elevated ZPP | Erythrocyte ZPP > 70 µmol/mol heme (83) |
| Plasma zinc concentration (µg/dL) |  |
| Low plasma zinc concentration | Plasma zinc < 65 µg/dL (84) |
| Plasma retinol concentration (µmol/L) |  |
| Low retinol | Plasma retinol < 0.70 µmol/L (85) |
| Marginal retinol | Plasma retinol < 1.05 µmol/L (85) |
| Plasma retinol binding protein concentration (RBP; µmol/L) |  |
| Low RBP | Plasma RBP < 0.70 µmol/L (86) |
| Marginal RBP | Plasma RBP < 1.05 µmol/L (86) |

Hb, hemoglobin; RBP, retinol binding protein; sTfR, soluble transferrin receptor; ZPP, zinc protoporphyrin.
<sup>1</sup>Plasma hepcidin concentration (ng/mL), low hepcidin (< 5.5 ng/mL) (87), plasma folate concentration (nmol/L), low folate (< 10 nmol/L) (88), plasma vitamin B12 concentration (pmol/L), low vitamin B12 (< 150 pmol/L), and depleted vitamin B12 (< 221 pmol/L) (89) were also considered as outcome variables, however data were not yet available from a sufficient number of trials to be included in the present analyses.

The principal measure of effect for normally distributed continuous outcomes was the mean difference (MD) between the intervention and comparison groups at endline, defined as the principal post-intervention time point as reported by each longitudinal trial or at an age closest to the end of the supplementation period for cross-sectional samples. For natural log transformed outcomes, the principal measure of effect was the ratio of geometric means (GMR) between the two groups at endline. For binary outcomes, the principal measure of effect was the prevalence ratio (PR; relative difference in proportions between groups) at endline. We also estimated prevalence differences (PD; in absolute percentage points) but considered them as secondary assessments of binary outcomes because such estimates are less consistent than prevalence ratios (26).

The treatment comparison of interest was child SQ-LNS vs. control. In the intervention groups, child SQ-LNS may have been provided along with other nutrition-sensitive co-interventions (e.g., WASH or child morbidity monitoring and treatment). The control groups consisted of passive or active comparison arms which provided no intervention or an intervention without any type of LNS or other child supplement (e.g., WASH). Several trials (or arms within trials) have delivered SQ-LNS to children whose mothers received maternal SQ-LNS during pregnancy and postpartum. We had originally planned to include trial arms that provided both maternal and child SQ-LNS in a sensitivity analysis only (i.e., the all-trials analysis), as maternal supplementation may have an additive effect on child outcomes when SQ-LNS is provided to both mothers and to their children. However, to maximize study inclusion and participant sample size, we decided post-hoc that if the main effects did not differ between the child-LNS-only analysis and the all-trials analysis (including maternal plus child LNS arms) by more than 20% for mean differences or by more than 0.05 for ratios of geometric means or prevalence ratios, the results of the all-trials analyses would be presented as the principal findings. Two additional sensitivity analyses were also conducted, as described below.

### Synthesis methods and exploration of variation in effects

We separately investigated: 1) full sample main effects of the intervention, 2) effect modification by study-level characteristics, and 3) effect modification by individual-level characteristics. For all three sets of analyses, we used a two-stage approach, which is preferred when the analysis includes cluster-randomized trials (30). In the first stage, we generated intervention effect estimates within each individual study. For longitudinal studies, we controlled for baseline status of the outcome variable, if available, to gain efficiency. For cluster-randomized trials, we used robust standard errors with randomization clusters as the independent unit (31). In the second stage, we pooled the first stage estimates using inverse-variance weighted fixed effects. We also conducted sensitivity analyses in which we pooled estimates using inverse-variance weighted random effects (32). Pooled estimates were only generated if three or more study or sub-study SQ-LNS vs. control comparisons were available for inclusion in the pooled estimate (e.g., 3 or more comparisons were represented within a study-level effect modification category).

1) *Full sample main effects of the intervention:* We first estimated the intervention effect for each study. We then pooled the first stage estimates to generate a pooled point estimate, 95% confidence interval, and corresponding p-value.
2) *Effect modification by study level characteristics:* We identified potential study-level effect modifiers prior to receipt of data, and categorized individual studies based on the distribution of effect modifier values across all studies (**Box 2**). We used random effects meta-regression to test the association between each effect modifier and the intervention. In the first stage of analysis, we estimated the parameter corresponding to the intervention effect as described above. In the second stage, we used a bivariate random effects meta-regression to test the association between the study-specific intervention effect and study level characteristics.
3) *Effect modification by individual level characteristics*: We identified potential individual-level characteristics based on a comprehensive review of effect modifiers considered by individual trials (either listed *a priori* in statistical analysis plans or as published) or selected based on biological plausibility (Box 2). We estimated the parameter corresponding to the interaction term of the effect modifier and the intervention for each study (31), as follows. For categorical effect modifiers, we first recoded them to create binary variables if needed, and then determined the interaction between the intervention and the binary effect modifier. We transformed continuous effect modifiers into binary variables by modeling the relationship within each study using splines and then pooling the first stage estimates to generate a pooled, fitted line. We defined programmatically useful dichotomous cutoffs based on the pooled fitted spline results and relevant context. We then generated pooled intervention effect estimates within each category to determine how the intervention effect in one subgroup differed from the intervention effect in the specified reference subgroup.

#### Box 2. Potential effect modifiers

| Study-level effect modifiers | Individual-level effect modifiers |
| --- | --- |
| Geographic region (WHO region: African vs. South-East Asia Region)<br>Anemia burden (country-specific, closest in time to the study: $< 60\%$ vs. $\geq 60\%$)<sup>1</sup><br>Malaria prevalence (country-specific, closest in time to the study: $< 10\%$ vs. $\geq 10\%$)<sup>2</sup><br>Inflammation prevalence (study-specific: elevated CRP $\leq 25\%$ and/or elevated AGP $\leq 50\%$ vs. elevated CRP $> 25\%$ and/or elevated AGP $> 50\%$)<sup>3</sup><br>Water quality (study-specific: $< 75\%$ vs. $\geq 75\%$ prevalence of improved drinking water)<sup>4</sup><br>Sanitation (study specific: $< 50\%$ vs. $\geq 50\%$ prevalence of improved sanitation)<sup>5</sup><br>Duration of child supplementation (study target: $> 12$ mo vs. $\leq 12$ mo)<br>Iron dose in the SQ-LNS product (9 mg/d vs. $< 9$ mg/d)<sup>6</sup><br>Child age at baseline or endline<br>Frequency of contact for intervention delivery or outcome assessments during the study (weekly vs. monthly)<br>Compliance (average percent compliance in LNS group $\geq 80\%$ vs. $< 80\%$)<sup>7</sup> | Maternal BMI ($< 20$ kg/m<sup>2</sup> vs. $\geq 20$ kg/m<sup>2</sup>)<br>Maternal age ($< 25$ y vs. $\geq 25$ y)<br>Maternal education (no formal or incomplete primary vs. complete primary or greater)<br>Child sex (female vs. male)<br>Child birth order (first born vs. later born)<br>Child baseline acute malnutrition (WLZ $< -2$ SD or MUAC $< 125$ mm vs. WLZ $\geq -2$ SD and MUAC $\geq 125$ mm; if MUAC not measured, WLZ $< -2$ SD vs. WLZ $> -2$ SD)<br>Child baseline anemia status (Hb $\geq 110$ g/L vs. $< 110$ g/L); iron outcomes only<br>Child high-dose vitamin A supplementation (receipt within six months prior to outcome assessment vs. non-receipt)<br>Child inflammation at time of outcome assessment (CRP $\leq 5$ mg/L and AGP $\leq 1$ g/L vs. not)<br>Household socio-economic status ($<$ study median vs. ≥ study median)<sup>8</sup><br>Food security (moderate to severe food insecurity vs. mild to secure)<br>Water quality (unimproved vs. improved)<sup>5</sup><br>Sanitation (unimproved vs. improved)<sup>6</sup><br>Season at the time of outcome assessment (rainy vs. dry)<sup>9</sup> |

<sup>1</sup> Country-specific prevalence of anemia among children 6-59 months, based on national surveys (see Supplemental Table 3)
<sup>2</sup> Country-specific prevalence of malaria, based on World Malaria Report 2018 (see Supplemental Table 3) (50)
<sup>3</sup> Elevated CRP defined as $> 5$ mg/L, elevated AGP defined as $> 1$ g/L.
<sup>4</sup> study-specific prevalence (see Supplemental Table 3) (90)
<sup>5</sup> study-specific prevalence (see Supplemental Table 3) (91)
<sup>6</sup> study-specific (see Supplemental Tables 2 and 3)
<sup>7</sup> study-specific, as reported based on a study-defined indicator (see Supplemental Table 3)
<sup>8</sup> based on a study-defined, study-specific assets index
<sup>9</sup> rainy vs. dry, based on study- and child-specific average rainfall during the month of measurement and two months prior (see Supplemental Methods and Supplemental Table 4)

In all analyses, heterogeneity was assessed using I^2^ and Tau^2^ statistics, within strata when relevant (33). We used a p-value of < 0.05 for main effects and a p-diff or p-for-interaction < 0.10 for effect modification by study-level or individual-level characteristics, respectively. Because biochemical outcomes are inter-related and the effect modification analyses are inherently exploratory, we did not adjust for multiple hypothesis testing (34).

To aid in interpretation of individual-level effect modification, we evaluated the results for binary outcomes to identify what we will call the “cutoff effect”. The distribution of the continuous outcome relative to the cutoff for the corresponding binary outcome (e.g., distribution of hemoglobin around the 110 g/L anemia cutoff) in the 2 effect modifier subgroups can influence the prevalence ratio and prevalence difference. When the mean in each of the 2 subgroups falls in a different location relative to the cutoff, the proportion of children close to the cutoff may be different between subgroups. This can lead to a greater reduction in the adverse binary outcome within one subgroup than within the other even if the shift in the mean value due to SQ-LNS is the same in both subgroups. To examine this, we simulated what would happen if we shifted the distribution of the non-reference effect modification subgroup to align with the reference subgroup (see Box 2), while maintaining the observed intervention effect mean difference in the continuous outcome within each subgroup. Based on ad-hoc, pragmatic criteria, if the p-for-interaction shifted from < 0.1 to > 0.2, we concluded that the cutoff effect explained the apparent effect modification; if it shifted from < 0.1 to > 0.1 but < 0.2, we concluded that the cutoff effect partially contributed to the apparent effect modification.

### Additional sensitivity analyses

For the child SQ-LNS only and all-trials analyses, we combined all non-LNS arms into a ‘Control’ group for that trial. However, substantial variations in trial design (e.g., integration of SQ-LNS supplementation with WASH interventions or enhanced morbidity monitoring and treatment, use of passive vs. active control arms) might influence the effect size of the estimates. Therefore, we conducted sensitivity analyses based on child-LNS only analyses in which we (1) separated the comparisons within trials that contained multi-component arms, thus restricting the comparisons to pairs of arms with the same non-nutrition components (e.g. SQ-LNS+WASH vs. WASH and SQ-LNS vs. control) or (2) excluded passive control arms, i.e., restricting the comparison to SQ-LNS vs. active control arms (**Supplemental Table 1**). These analyses are considered exploratory.

## Results

### Literature search and trial characteristics

We identified 15 trials that met our inclusion criteria for the child SQ-LNS only and/or all-trials analyses, 13 of which (35–48) provided individual participant biochemical data and are included in this analysis (9-13, 15-17, 39, 46-48) (**Figure 1**). In one study biochemical data were not collected (49), and investigators for one trial were unable to participate (14). An overview of each trial included in the analysis is provided in **Table 1**; additional details are provided in Supplemental Table 1. One trial was designed *a priori* to present results separately for HIV exposed and HIV un-exposed children (47, 48); therefore we considered it herein as two comparisons in all analyses and presentation of results. The PROMIS trials in Burkina Faso and Mali each included a longitudinal cohort and repeated cross-sectional surveys (at baseline and endline); however biochemical samples were collected only in the cross-sectional surveys (39, 46).

**Figure 1:**
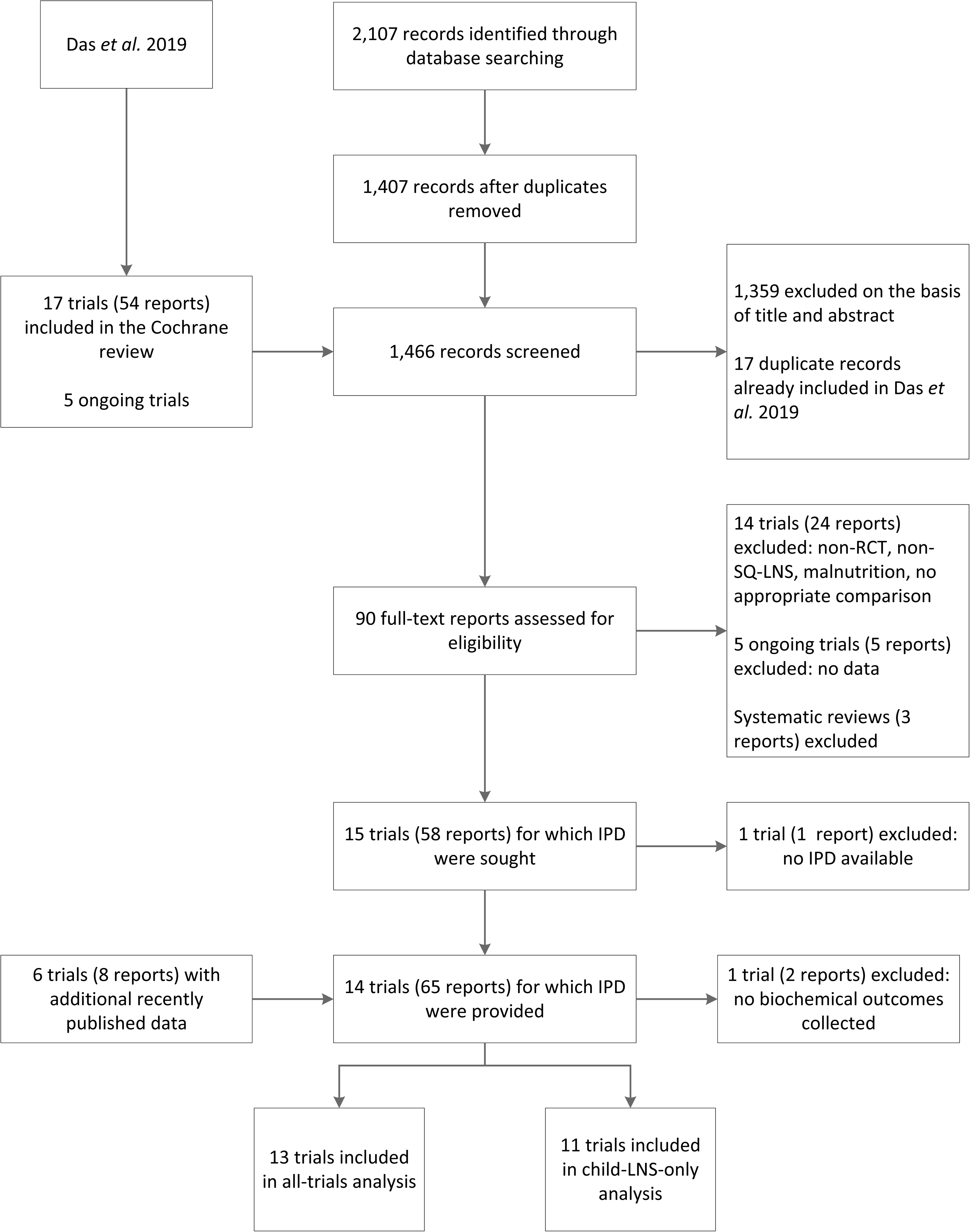
Study flow diagram

**Table 1.**
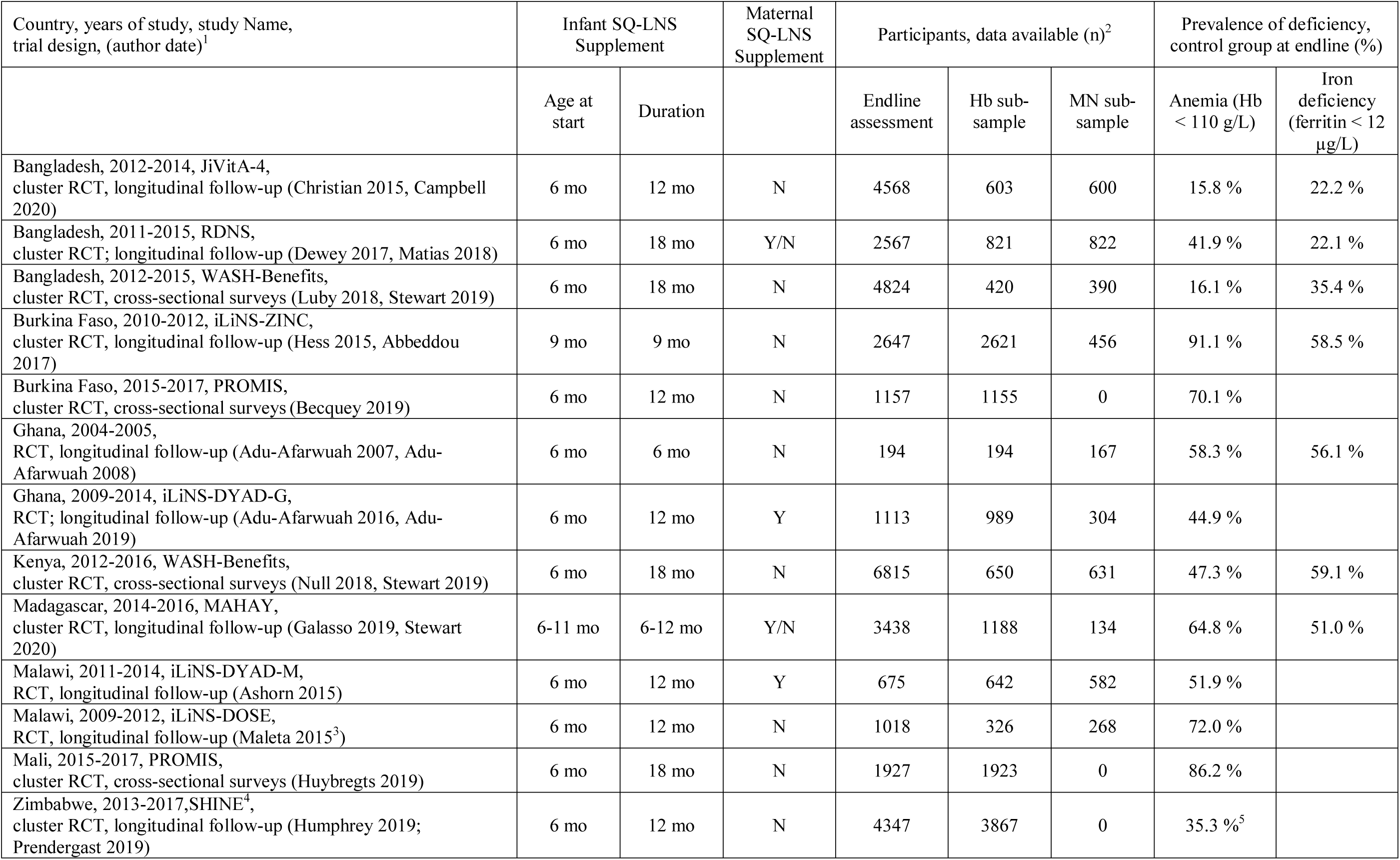
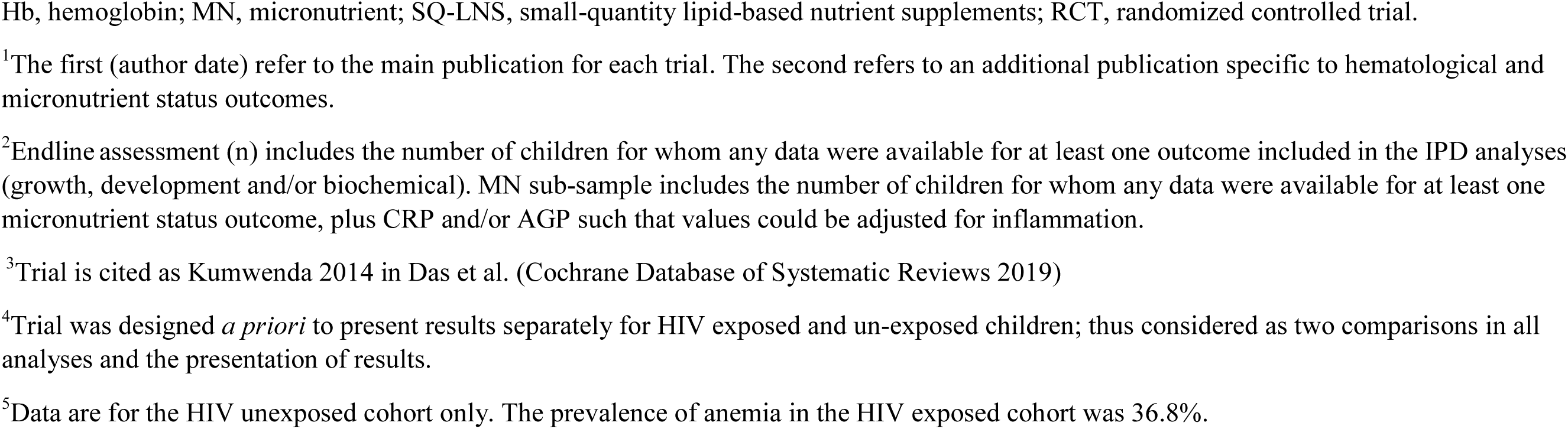
Characteristics of trials included in the individual participant data analysis

| Country, years of study, study Name, trial design, (author date)<sup>1</sup> | Infant SQ-LNS Supplement |  | Maternal SQ-LNS Supplement | Participants, data available (n)<sup>2</sup> |  |  | Prevalence of deficiency, control group at endline (%) |  |
| --- | --- | --- | --- | --- | --- | --- | --- | --- |
|  | Age at start | Duration |  | Endline assessment | Hb sub-sample | MN sub-sample | Anemia (Hb < 110 g/L) | Iron deficiency (ferritin < 12 µg/L) |
| Bangladesh, 2012-2014, JiVitA-4, cluster RCT, longitudinal follow-up (Christian 2015, Campbell 2020) | 6 mo | 12 mo | N | 4568 | 603 | 600 | 15.8 % | 22.2 % |
| Bangladesh, 2011-2015, RDNS, cluster RCT; longitudinal follow-up (Dewey 2017, Matias 2018) | 6 mo | 18 mo | Y/N | 2567 | 821 | 822 | 41.9 % | 22.1 % |
| Bangladesh, 2012-2015, WASH-Benefits, cluster RCT, cross-sectional surveys (Luby 2018, Stewart 2019) | 6 mo | 18 mo | N | 4824 | 420 | 390 | 16.1 % | 35.4 % |
| Burkina Faso, 2010-2012, iLiNS-ZINC, cluster RCT, longitudinal follow-up (Hess 2015, Abbeddou 2017) | 9 mo | 9 mo | N | 2647 | 2621 | 456 | 91.1 % | 58.5 % |
| Burkina Faso, 2015-2017, PROMIS, cluster RCT, cross-sectional surveys (Becquey 2019) | 6 mo | 12 mo | N | 1157 | 1155 | 0 | 70.1 % |  |
| Ghana, 2004-2005, RCT, longitudinal follow-up (Adu-Afarwuah 2007, Adu-Afarwuah 2008) | 6 mo | 6 mo | N | 194 | 194 | 167 | 58.3 % | 56.1 % |
| Ghana, 2009-2014, iLiNS-DYAD-G, RCT; longitudinal follow-up (Adu-Afarwuah 2016, Adu-Afarwuah 2019) | 6 mo | 12 mo | Y | 1113 | 989 | 304 | 44.9 % |  |
| Kenya, 2012-2016, WASH-Benefits, cluster RCT, cross-sectional surveys (Null 2018, Stewart 2019) | 6 mo | 18 mo | N | 6815 | 650 | 631 | 47.3 % | 59.1 % |
| Madagascar, 2014-2016, MAHAY, cluster RCT, longitudinal follow-up (Galasso 2019, Stewart 2020) | 6-11 mo | 6-12 mo | Y/N | 3438 | 1188 | 134 | 64.8 % | 51.0 % |
| Malawi, 2011-2014, iLiNS-DYAD-M, RCT, longitudinal follow-up (Ashorn 2015) | 6 mo | 12 mo | Y | 675 | 642 | 582 | 51.9 % |  |
| Malawi, 2009-2012, iLiNS-DOSE, RCT, longitudinal follow-up (Maleta 2015<sup>3</sup>) | 6 mo | 12 mo | N | 1018 | 326 | 268 | 72.0 % |  |
| Mali, 2015-2017, PROMIS, cluster RCT, cross-sectional surveys (Huybregts 2019) | 6 mo | 18 mo | N | 1927 | 1923 | 0 | 86.2 % |  |
| Zimbabwe, 2013-2017, SHINE<sup>4</sup>, cluster RCT, longitudinal follow-up (Humphrey 2019; Prendergast 2019) | 6 mo | 12 mo | N | 4347 | 3867 | 0 | 35.3 %<sup>5</sup> |  |

Hb, hemoglobin; MN, micronutrient; SQ-LNS, small-quantity lipid-based nutrient supplements; RCT, randomized controlled trial.
<sup>1</sup>The first (author date) refer to the main publication for each trial. The second refers to an additional publication specific to hematological and micronutrient status outcomes.
<sup>2</sup>Endline assessment (n) includes the number of children for whom any data were available for at least one outcome included in the IPD analyses (growth, development and/or biochemical). MN sub-sample includes the number of children for whom any data were available for at least one micronutrient status outcome, plus CRP and/or AGP such that values could be adjusted for inflammation.
<sup>3</sup>Trial is cited as Kumwenda 2014 in Das et al. (Cochrane Database of Systematic Reviews 2019)
<sup>4</sup>Trial was designed *a priori* to present results separately for HIV exposed and un-exposed children; thus considered as two comparisons in all analyses and the presentation of results.
<sup>5</sup>Data are for the HIV unexposed cohort only. The prevalence of anemia in the HIV exposed cohort was 36.8%.

The 13 trials in these analyses were conducted in 9 countries in Sub-Saharan Africa and South Asia, and included a total of 15,946 infants and young children with biochemical data. Eleven trials began child supplementation with SQ-LNS at 6 months of age; for the remaining two trials, SQ-LNS supplementation began when infants were 9 months of age in one (38), and between 6-11 months of age in the other (43). Supplementation occurred for 6 to 18 months in duration, depending on the trial. Four trials included intervention arms that also provided SQ-LNS to mothers during pregnancy and the first 6 months postpartum (36, 41, 43, 44). The majority of trials provided a peanut- and milk-based SQ-LNS providing approximately 120 kcal/d and 1 RDA of most micronutrients (**Supplemental Table 2**). However, the iron dose per day varied among trials. Four trials provided 9 mg/d iron (36, 37, 40, 42), 8 trials provided 6 mg/d iron (38, 39, 41, 43–48), and one trial provided 3.3 – 9 mg/d iron depending on study arm and child age (35).

Categorizations and descriptive information for potential study- and individual-level effect modifiers, by trial, are presented in **Supplemental Tables 3 - 4**, respectively. Characteristics ranged widely across trials. Based on data from national surveys, the prevalence of anemia among children 6-59 months ranged from 36% to 88%, and the prevalence of malaria (presumed and confirmed cases) ranged from < 1% to 59% (50). Based on data from individual trials, the study-level prevalence of elevated CRP ranged from 11% to 34% and the prevalence of elevated AGP ranged from 29% to 68%.

All trials measured hemoglobin concentration (n = 15,398), and 11 reported at least one biomarker of iron status (n = 1,542 – 3,078) (Supplemental Table 1, **Supplemental Table 5**). Fewer studies assessed zinc (3 studies; n = 1,133) or vitamin A (9 studies; n = 1,236 – 2,314) status. Assessments of hepcidin, folate and vitamin B12 concentrations were planned in 3-4 studies each, but data were not yet available from a sufficient number of trials to be included in the present analysis. Median biomarker concentrations and prevalence of dichotomous study outcomes among control groups by trial are presented in Table 1 (anemia and iron deficiency) and **Supplemental Table 6** (all outcomes). In control groups at endline, the prevalence of anemia ranged from 16% to 91%, the prevalence of iron deficiency ranged from 22% to 59%, and the prevalence of IDA ranged from 9% to 55%.

In general, trials were considered to have a low risk of bias, with the exception of lack of masking of participants owing to the nature of the intervention (**Supplemental Table 7** and **Supplemental Figure 1**).

### Main effects of SQ-LNS on anemia and micronutrient status

Results from the all-trials analyses, inclusive of maternal + child SQ-LNS trials/arms, are presented below, and in **Table 2**, because the results from the all-trials and child-LNS-only analyses were similar for all outcomes (**Supplemental Figures 2A to 2D**). Forest plots for all main effects by outcome and individual study are presented below for selected outcomes and as supplemental materials for all other outcomes (**Supplemental Figures 3A to 3AA**).

**Table 2.**
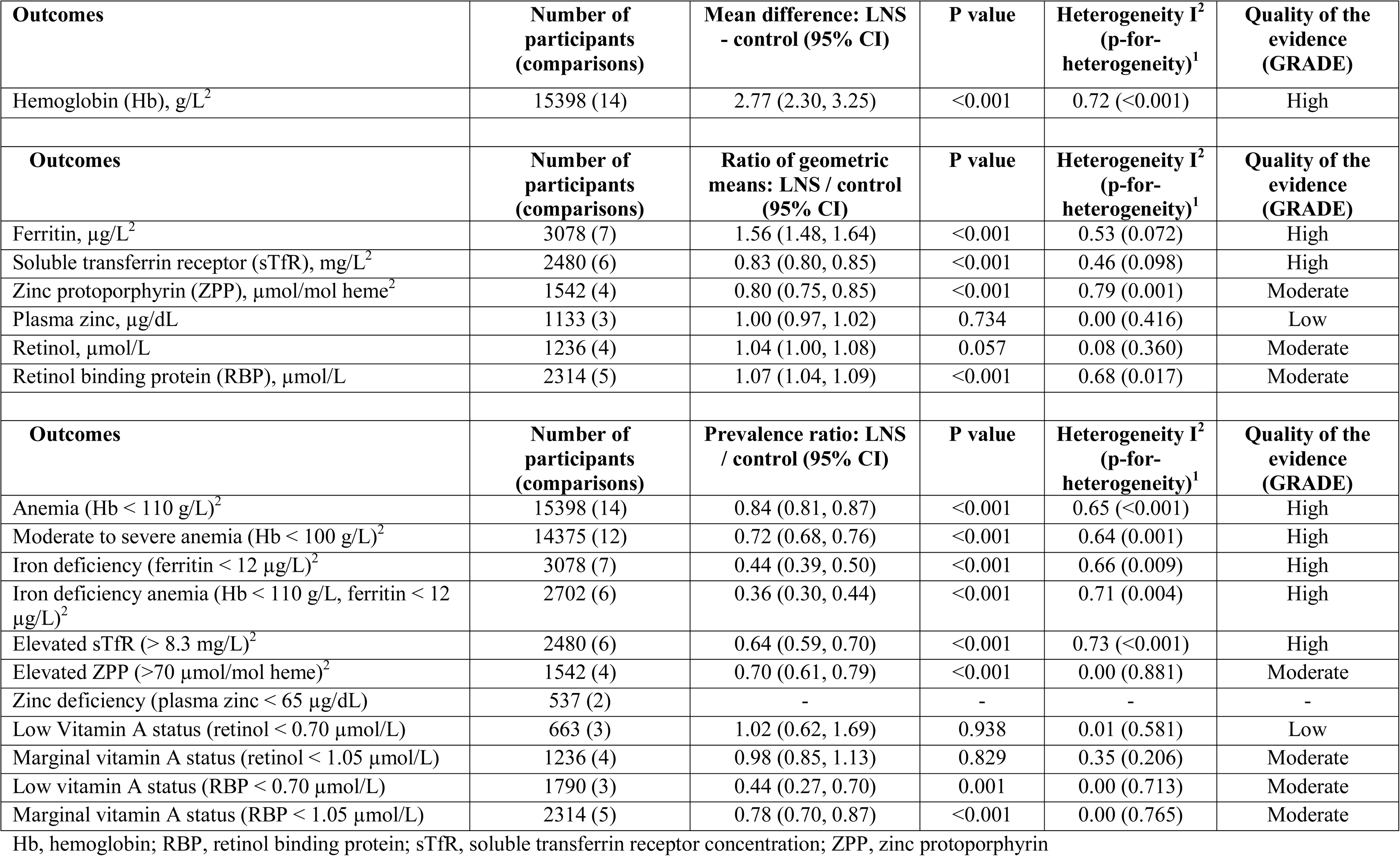
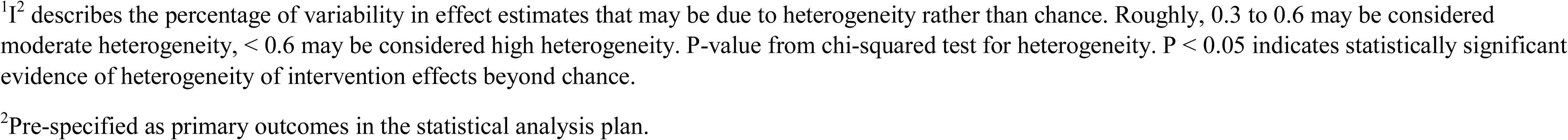
The effect of SQ-LNS on hemoglobin and micronutrient status

| **Outcomes** | **Number of participants (comparisons)** | **Mean difference: LNS - control (95% CI)** | **P value** | **Heterogeneity I<sup>2</sup> (p-for-heterogeneity)<sup>1</sup>** | **Quality of the evidence (GRADE)** |
| --- | --- | --- | --- | --- | --- |
| Hemoglobin (Hb), g/L<sup>2</sup> | 15398 (14) | 2.77 (2.30, 3.25) | <0.001 | 0.72 (<0.001) | High |
| **Outcomes** | **Number of participants (comparisons)** | **Ratio of geometric means: LNS / control (95% CI)** | **P value** | **Heterogeneity I<sup>2</sup> (p-for-heterogeneity)<sup>1</sup>** | **Quality of the evidence (GRADE)** |
| Ferritin, µg/L<sup>2</sup> | 3078 (7) | 1.56 (1.48, 1.64) | <0.001 | 0.53 (0.072) | High |
| Soluble transferrin receptor (sTfR), mg/L<sup>2</sup> | 2480 (6) | 0.83 (0.80, 0.85) | <0.001 | 0.46 (0.098) | High |
| Zinc protoporphyrin (ZPP), µmol/mol heme<sup>2</sup> | 1542 (4) | 0.80 (0.75, 0.85) | <0.001 | 0.79 (0.001) | Moderate |
| Plasma zinc, µg/dL | 1133 (3) | 1.00 (0.97, 1.02) | 0.734 | 0.00 (0.416) | Low |
| Retinol, µmol/L | 1236 (4) | 1.04 (1.00, 1.08) | 0.057 | 0.08 (0.360) | Moderate |
| Retinol binding protein (RBP), µmol/L | 2314 (5) | 1.07 (1.04, 1.09) | <0.001 | 0.68 (0.017) | Moderate |
| **Outcomes** | **Number of participants (comparisons)** | **Prevalence ratio: LNS / control (95% CI)** | **P value** | **Heterogeneity I<sup>2</sup> (p-for-heterogeneity)<sup>1</sup>** | **Quality of the evidence (GRADE)** |
| Anemia (Hb < 110 g/L)<sup>2</sup> | 15398 (14) | 0.84 (0.81, 0.87) | <0.001 | 0.65 (<0.001) | High |
| Moderate to severe anemia (Hb < 100 g/L)<sup>2</sup> | 14375 (12) | 0.72 (0.68, 0.76) | <0.001 | 0.64 (0.001) | High |
| Iron deficiency (ferritin < 12 µg/L)<sup>2</sup> | 3078 (7) | 0.44 (0.39, 0.50) | <0.001 | 0.66 (0.009) | High |
| Iron deficiency anemia (Hb < 110 g/L, ferritin < 12 µg/L)<sup>2</sup> | 2702 (6) | 0.36 (0.30, 0.44) | <0.001 | 0.71 (0.004) | High |
| Elevated sTfR (> 8.3 mg/L)<sup>2</sup> | 2480 (6) | 0.64 (0.59, 0.70) | <0.001 | 0.73 (<0.001) | High |
| Elevated ZPP (>70 µmol/mol heme)<sup>2</sup> | 1542 (4) | 0.70 (0.61, 0.79) | <0.001 | 0.00 (0.881) | Moderate |
| Zinc deficiency (plasma zinc < 65 µg/dL) | 537 (2) | - | - | - | - |
| Low Vitamin A status (retinol < 0.70 µmol/L) | 663 (3) | 1.02 (0.62, 1.69) | 0.938 | 0.01 (0.581) | Low |
| Marginal vitamin A status (retinol < 1.05 µmol/L) | 1236 (4) | 0.98 (0.85, 1.13) | 0.829 | 0.35 (0.206) | Moderate |
| Low vitamin A status (RBP < 0.70 µmol/L) | 1790 (3) | 0.44 (0.27, 0.70) | 0.001 | 0.00 (0.713) | Moderate |
| Marginal vitamin A status (RBP < 1.05 µmol/L) | 2314 (5) | 0.78 (0.70, 0.87) | <0.001 | 0.00 (0.765) | Moderate |

Hb, hemoglobin; RBP, retinol binding protein; sTfR, soluble transferrin receptor concentration; ZPP, zinc protoporphyrin
<sup>1</sup> $I^2$ describes the percentage of variability in effect estimates that may be due to heterogeneity rather than chance. Roughly, 0.3 to 0.6 may be considered moderate heterogeneity, < 0.6 may be considered high heterogeneity. P-value from chi-squared test for heterogeneity. $P < 0.05$ indicates statistically significant evidence of heterogeneity of intervention effects beyond chance.
<sup>2</sup>Pre-specified as primary outcomes in the statistical analysis plan.

SQ-LNS had a significant positive effect on all biomarkers of hematological and iron status (Table 2**, Figures 2-3**). Compared to control children, hemoglobin concentrations were 2.77 g/L higher among children who received SQ-LNS; additionally, ferritin concentrations were 56% higher, and sTfR and ZPP concentrations were 17% lower among children who received SQ-LNS vs. control. SQ-LNS reduced the prevalence of anemia by 16% (10 percentage points), iron deficiency by 56% (22 percentage points), and IDA by 64% (14 percentage points). Differences in plasma zinc and retinol concentrations between those who received SQ-LNS and those who did not were not significant. RBP concentrations were 7% higher among children who received SQ-LNS compared to the control group. SQ-LNS reduced the prevalences of low and marginal vitamin A status, as measured by RBP, by 56% (3 percentage points) and 22% (8 percentage points), respectively; there was no effect of SQ-LNS on vitamin A status as measured by retinol. We rated the quality of evidence for hemoglobin, ferritin and sTfR concentrations, as well as anemia, moderate-to-severe anemia, iron deficiency, elevated sTfR and IDA as high, based on the GRADE criteria listed above in Methods: at least 6 randomized controlled trials were available for these outcomes and the total sample size was ∼> 2,500, the risk of bias was low, all trials were directly aimed at evaluating SQ-LNS, funnel plots revealed no indication of publication bias, and the direction of the effect was consistent even though the magnitude of the effect across trials differed (i.e., moderate to high heterogeneity). For all other outcomes, we rated the quality of the evidence as low to moderate, primarily because of limited availability of data, i.e. fewer trials and participants.

**Figure 2:**
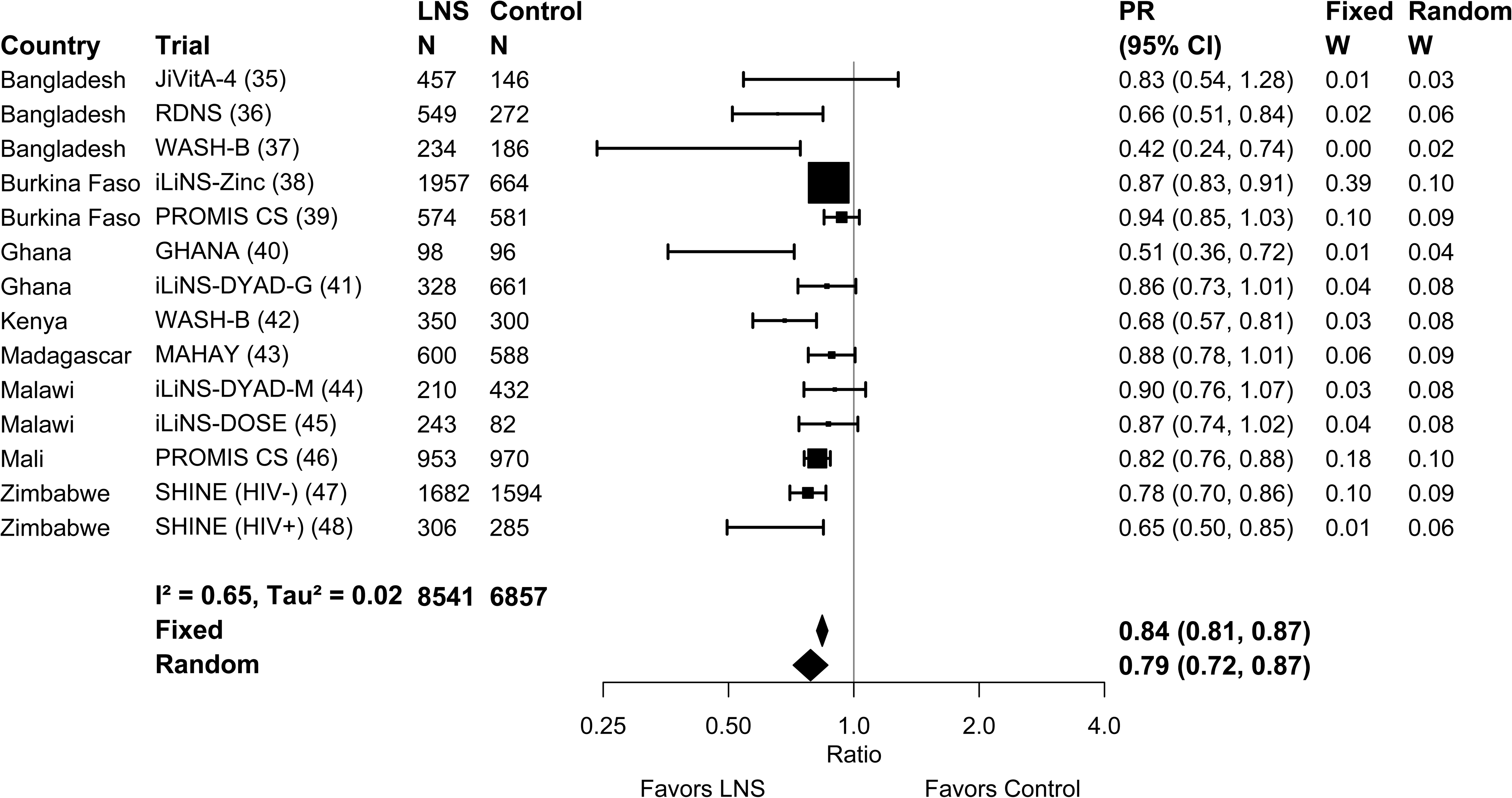
Forest plot of the effect of SQ-LNS on anemia prevalence

**Figure 3:**
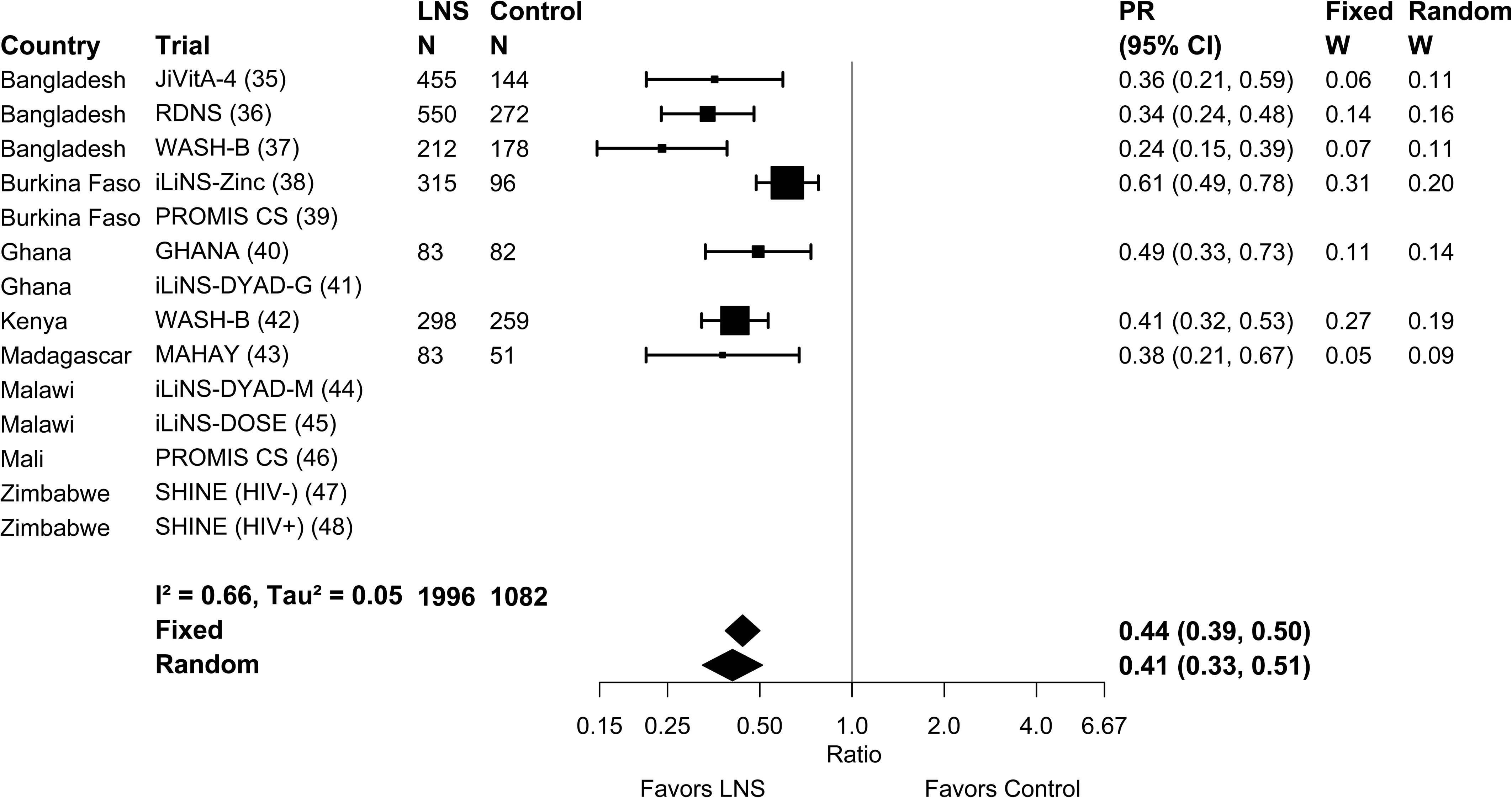
Forest plot of the effect of SQ-LNS on prevalence of iron deficiency (ferritin < 12 µg/L)

Mean differences, ratios of geometric means and prevalence ratios for all outcomes were either identical or greater in random vs. fixed effects models (Supplemental Figures 3A to 3AA). The point estimates of the main effects were similar in the sensitivity analyses in which multi-component arms (e.g. WASH + SQ-LNS) were compared to reference groups that had the same components without SQ-LNS and in which passive control trials were excluded (Supplemental Figures 2A-D). For example, the prevalence ratios for anemia ranged from 0.82-0.84 and those for iron deficiency were 0.36-0.44. There were no differences in statistical significance between models in which outcomes were adjusted vs. not adjusted for inflammation (data not shown).

### Effect modification by study-level characteristics

The effects of SQ-LNS on hemoglobin, anemia and biomarkers of iron status, stratified by study-level characteristics, are presented in **Figures 4A to 4H**, and an overview of these results is presented in **Table 3**. Forest plots for all outcomes stratified by study-level effect modifiers are presented in **Supplemental Figures 4A to 4AA.** We were unable to generate pooled estimates for effect modification by any study-level characteristics for ZPP and biomarkers of zinc and vitamin A status (i.e., retinol and RBP) due to the limited number of studies that measured these biomarkers. For iron status biomarkers (ferritin and sTfR), water quality and sanitation sub-groups captured the same trials, so these characteristics are considered as a single effect modifier at the study-level. Effect modification results were generally consistent across all sensitivity analyses (**Supplemental Figures 5A to 5AA**) and between models in which outcomes were inflammation-adjusted or not (data not shown); the results presented below refer to the all-trials analyses of inflammation-adjusted outcomes.

**Figure 4:**
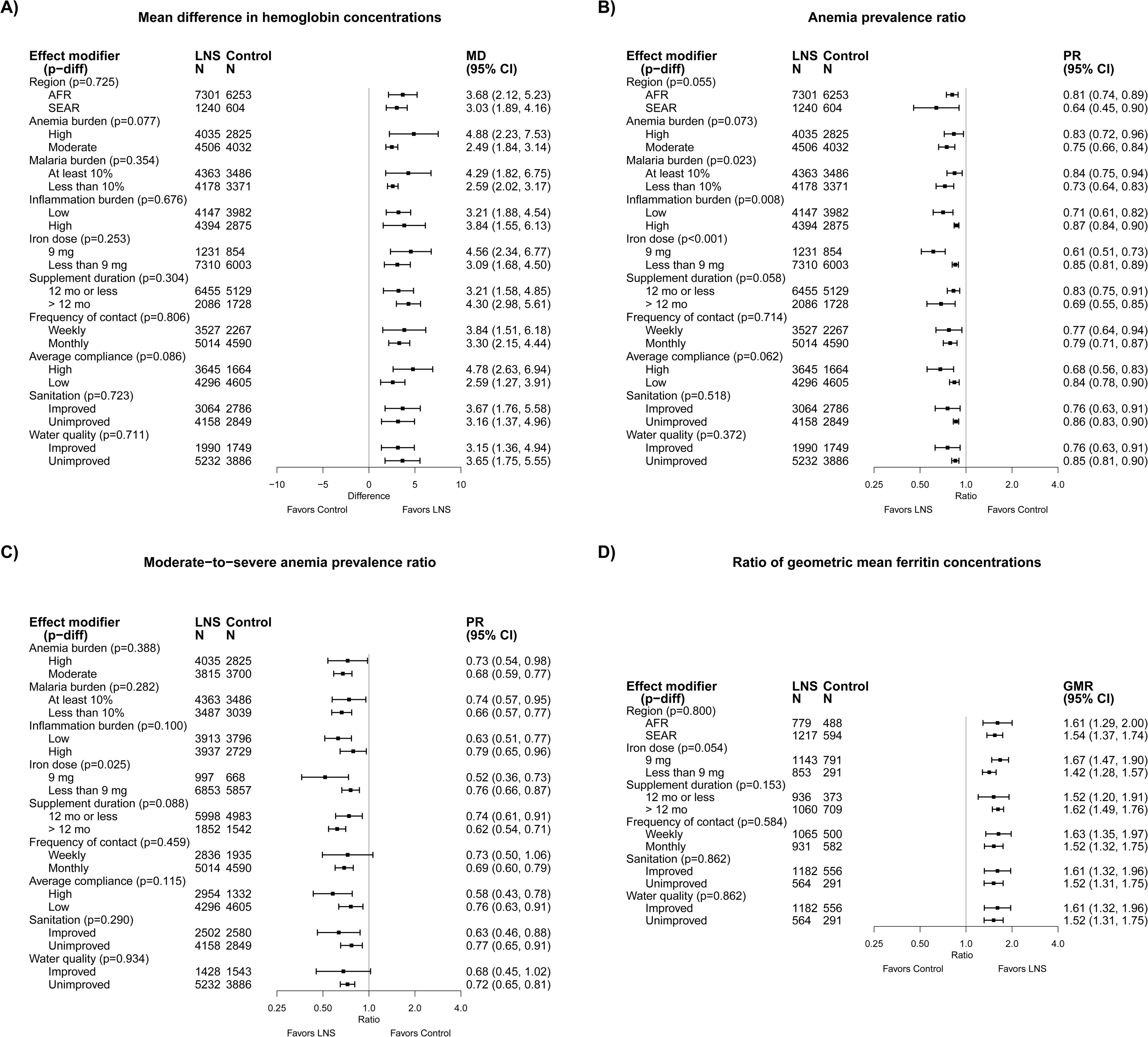
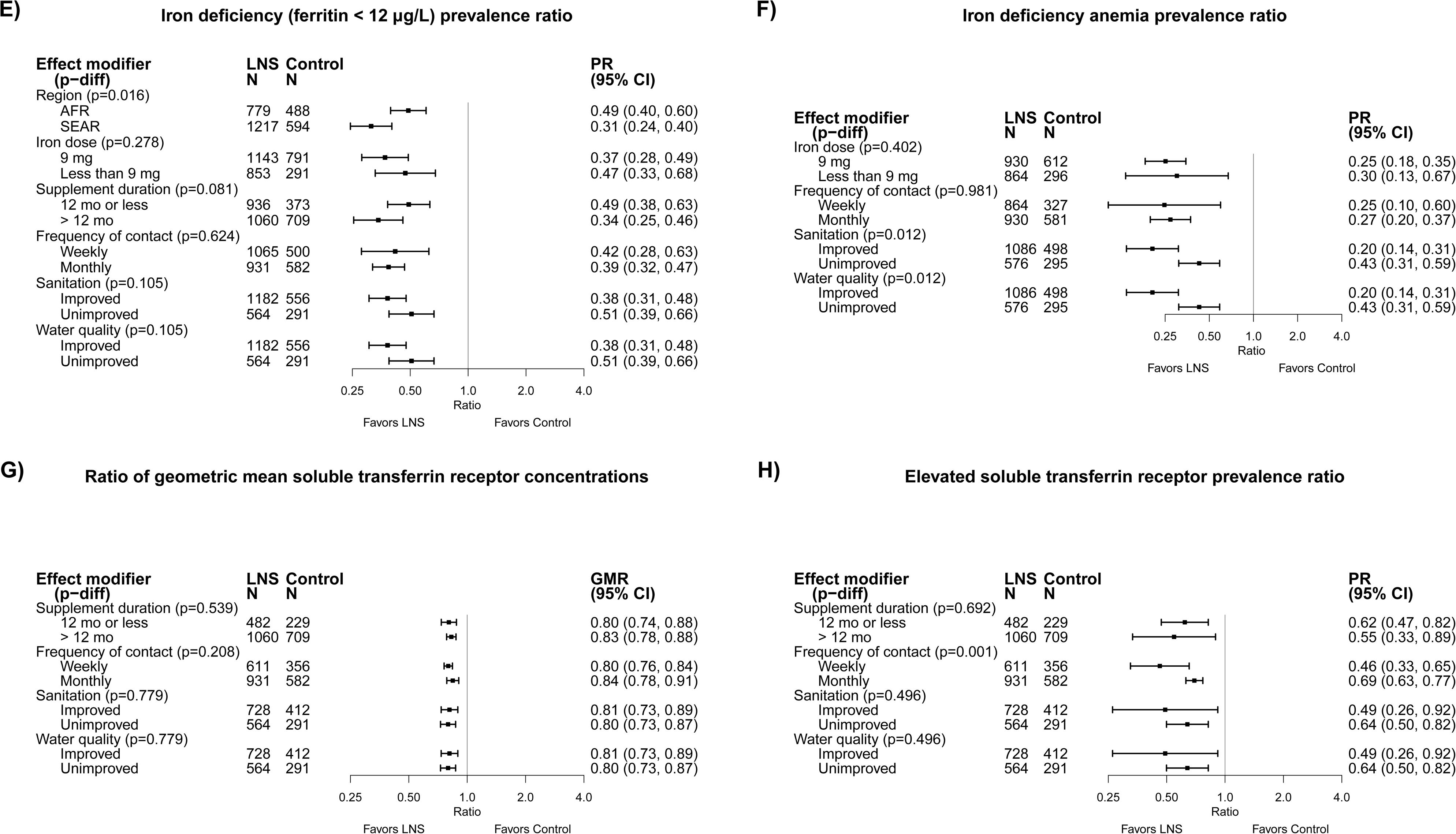
The effect of SQ-LNS provided to children 6-24 months of age compared with a control on (A) hemoglobin concentration, (B) anemia prevalence, (C) moderate-to-severe anemia prevalence, (D) ferritin concentration, (E) iron deficiency, (F) iron deficiency anemia, (G) soluble transferrin receptor concentration, and (H) elevated soluble transferrin receptor, stratified by study-level characteristics. Due to the limited number of studies, we were unable to examine study-level effect modification on all outcomes for all potential modifiers of interest. GMR, ratio of geometric means, MD, mean difference, PR, prevalence ratio, p-diff, p value for the difference in the effect of SQ-LNS between the two levels of the effect modifier.

**Table 3.**
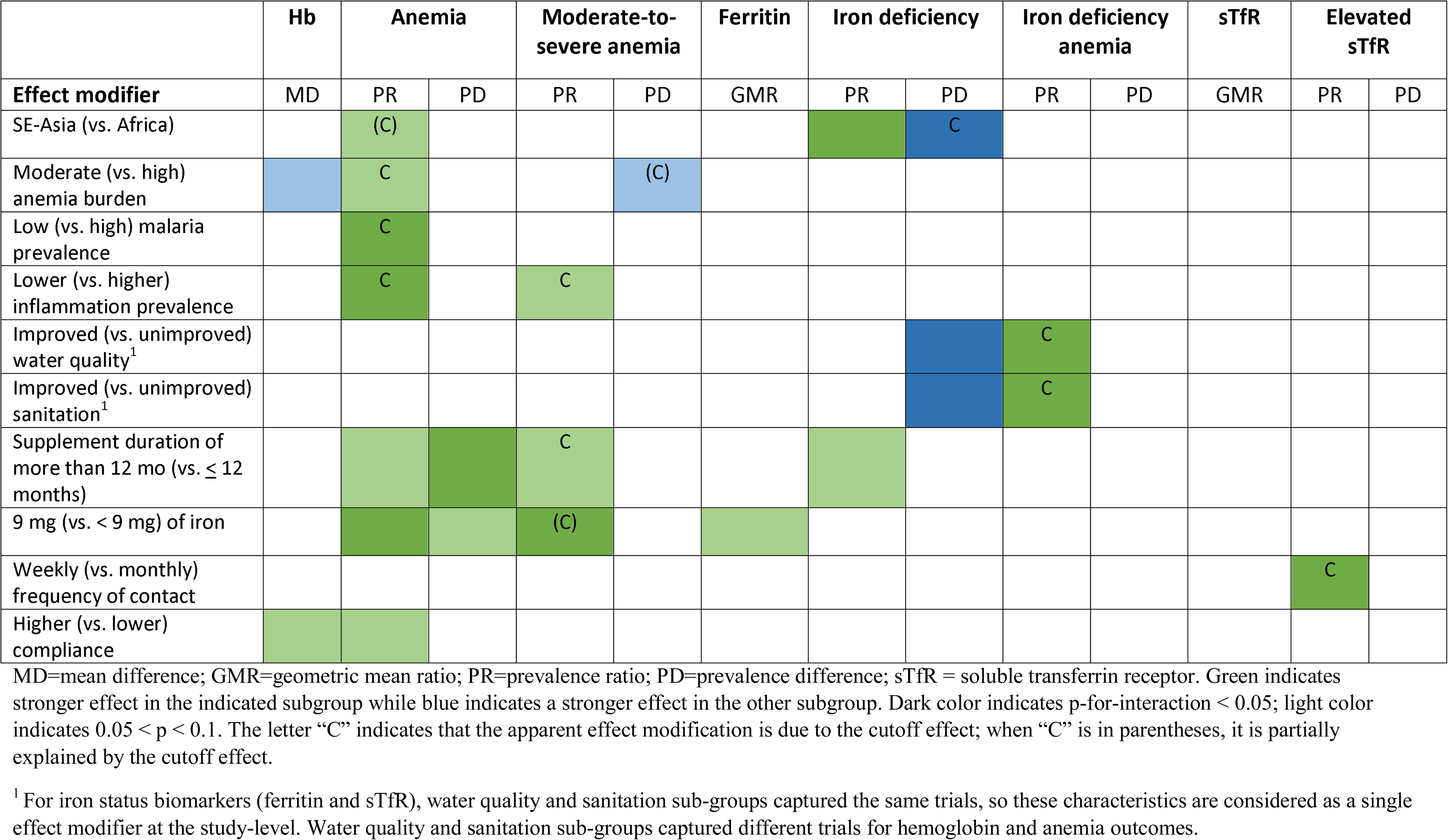
Overview of study-level effect modification

In all analyses of effect modification by study-level characteristics, the 95% confidence interval of the mean hemoglobin difference, and the ratios of geometric mean ferritin and sTfR concentrations, excluded the null for both strata, indicating that there were significant improvements in hemoglobin, ferritin and sTfR concentrations among children receiving SQ-LNS regardless of study-level characteristics. Similarly, the upper bound of the 95% confidence interval was < 1 for the prevalence ratio and < 0 for the prevalence difference for both categories in all comparisons, indicating that there were significant reductions in the prevalence of anemia, iron deficiency and IDA among children receiving SQ-LNS regardless of study-level characteristics. However, there were some significant interactions, which are described below.

#### Hemoglobin concentrations and anemia

Reductions in anemia prevalence due to SQ-LNS were greater among studies conducted in South Asia than in Africa (PR: 0.64 vs. 0.81, Figure 4B, Supplemental Figure 4B1). We observed a greater effect of SQ-LNS on hemoglobin concentrations among high anemia burden countries, compared to countries with a moderate burden of anemia (MD: 4.88 g/L vs. 2.49 g/L, Figure 4A, Supplemental Figure 4A2), although the effect of SQ-LNS on anemia prevalence was lower in high anemia burden countries than in moderate anemia burden countries (PR: 0.83 vs. 0.75, Figure 4B, Supplemental Figure 4B2). In addition, there was a greater effect of SQ-LNS on the prevalence of anemia among studies conducted in countries with a low vs. high malaria prevalence (PR: 0.73 vs. 0.84, Figure 4B, Supplemental Figure 4B3) and at sites with a low inflammation prevalence compared to those with a higher inflammation prevalence (PR: 0.71 vs. 0.87, Figure 4B, Supplemental Figure 4B4). Study-level access to improved sanitation or water quality did not significantly modify the effects of SQ-LNS on hemoglobin concentrations (Figure 4A, Supplemental Figures 4A4 and 4A5) or the prevalence of anemia (Figure 4B, Supplemental Figures 4B4 and 4B5).

Among studies that provided supplements for longer than 12 months compared to 12 months or less, there was a greater effect of SQ-LNS on the prevalence of both anemia (PR: 0.69 vs. 0.83, Figure 4B, Supplemental Figure 4B7) and moderate-to-severe anemia (PR: 0.62 vs. 0.74, Figure 4C and Supplemental Figure 4D7). Similarly, among studies that provided a higher iron dose (9 mg/d) compared to those providing a lower dose (< 9 mg/d), we observed a greater effect of SQ-LNS on the prevalence of anemia (PR: 0.61 vs. 0.85, Figure 4B, Supplemental Figure 4B8) and moderate-to-severe anemia (PR: 0.52 vs. 0.76, Supplemental Figure 4D8). There was no effect modification by frequency of contact (weekly vs. monthly), but we did observe a greater effect of SQ-LNS on hemoglobin concentrations and the prevalence of anemia among studies that reported high compliance, compared to those reporting lower compliance (MD: 4.78 g/L vs. 2.59 g/L, Figure 4A and Supplemental Figure 4A10 and PR: 0.68 vs. 0.84, Figure 4B and Supplemental Figure 4B10, respectively).

#### Biomarkers of iron status and prevalence of deficiency

The effect of SQ-LNS on the prevalence of iron deficiency (inflammation-adjusted ferritin concentration < 12 µg/L) was greater among trials conducted in South Asia than in Africa (PR: 0.31 vs. 0.49, Figure 4E and Supplemental Figure 4G1), although the percentage point reduction in iron deficiency associated with SQ-LNS was lower among sites in South Asia (vs. Africa) (18 vs. 30 percentage points; P = 0.026, Supplemental Figure 4H1). Study level sanitation and water quality did not significantly modify the effects of SQ-LNS on ferritin or sTfR concentrations or the relative reductions in the prevalences of iron deficiency and high sTfR (Figures 4D-4E, 4G-4H, Supplemental Figures 4F5-6, 4G5-6, 4K5-6, 4L5-6). We did observe greater effects of SQ-LNS on the relative reduction in the prevalence of IDA among trials conducted in sites where a greater proportion of households had access to improved sanitation and water quality, compared to sites with lower sanitation and water quality (PR: 0.20 vs. 0.43, Figure 2F, Supplemental Figure 4I5-6), consistent with the non-significant differences in the relative reductions in the prevalence of iron deficiency (PR: 0.38 vs. 0.51; P-for-diff = 0.105, Figure 4E, Supplemental Figure 4G6). However, the percentage point reduction in iron deficiency due to SQ-LNS was actually greater among trials conducted in sites where a lower (vs. higher) proportion of households had access to improved sanitation and water quality (28 vs. 18 percentage points; P-for-diff = 0.006, Supplemental Figures 4H5-6).

We observed a greater effect of SQ-LNS on the prevalence of iron deficiency among studies that provided SQ-LNS for longer than 12 months compared to studies of shorter durations (PR: 0.34 vs. 0.49, Figure 4E, Supplemental Figure 4G7). In addition, we observed a greater effect of SQ-LNS on ferritin concentrations among studies that provided 9 mg/d of iron, compared to studies that provided < 9 mg/d (GMR: 1.67 vs. 1.42, Figure 4D and Supplemental Figure 4F8). Frequency of contact did not significantly modify the effects of SQ-LNS on ferritin or sTfR concentrations, nor on prevalences of iron deficiency or IDA. However, the effect of SQ-LNS on elevated sTfR (indicative of impaired functional iron status) was greater among studies in which children received weekly vs. monthly study visits (PR: 0.46 vs. 0.69, Figure 4H and Supplemental Figure 4L9).

#### Overview of study-level effect modification

Table 3 shows that numerous characteristics of study context and design (e.g., anemia burden, iron dose, duration of supplementation) modified the effect of SQ-LNS on hematological and iron status biomarkers. When there is no significant effect modification for a continuous outcome (e.g., hemoglobin concentration) but there is for the prevalence ratio or prevalence difference for the corresponding binary outcome (e.g. anemia), the results could be due to the “cutoff effect”, as described in Methods. Our simulations identified several cutoff effects (Table 3). For example, we observed greater effects of SQ-LNS on the prevalence of anemia in sites with a moderate (vs. high) burden of anemia, a low (vs. high) malaria burden, and a low (vs. high) inflammation burden, three study-level characteristics that tended to cluster together. However, the effects of SQ-LNS on hemoglobin concentrations were greater in studies with a higher burden of anemia and mean hemoglobin differences due to SQ-LNS did not differ significantly by malaria or inflammation burden. Simulations indicated that the observed effect modification by study-level anemia, malaria and inflammation burden with respect to the prevalence ratio for anemia appears to be due to the cut-off effect (i.e., differences in the population distribution of hemoglobin between sub-groups). Among control group children, mean hemoglobin concentrations were higher in sites with lower burdens of anemia, malaria and inflammation than in sites with higher prevalences of these factors ( ≈ 114 g/L vs. ≈100 g/L). As a result, a greater proportion of children who received SQ-LNS shifted across the cut-off from anemic to non-anemic in the former sites than in the latter. The cutoff effect also appeared to contribute, at least partially, to the apparent effect modification by a) region, for the prevalence of anemia and the prevalence difference for iron deficiency, b) improved sanitation and water quality, for the prevalence of IDA, c) supplementation duration and iron dose, for the prevalence of moderate-to-severe anemia, and d) frequency of contact, for the prevalence of elevated sTfR.

### Effect modification by individual-level characteristics

The effects of SQ-LNS on hemoglobin (anemia) and biomarkers of iron status, stratified by individual-level (i.e., child, maternal and household) characteristics, are presented in **Figures 5A-5H** and an overview of individual-level effect modification is provided in **Table 4**. Forest plots of all outcomes by potential individual-level effect modifiers are presented in **Supplemental Figures 6A to 6AA and 7A to 7AA.** For some biomarkers of micronutrient status, we were unable to generate pooled estimates for effect modification by certain potential individual-level effect modifiers because fewer than three trials (comparisons) assessed both the outcome and the effect modifier of interest and/or there were too few individuals with the outcome categorized into each stratum (e.g., ferritin concentration by baseline anemia status or household sanitation). Effect modification results were generally consistent across all sensitivity analyses and between the fixed and random effects models (**Supplemental Figures 8A to 8AA**). Because all-trials analyses maximized the sample size, some potential effect modifiers were significant in the all-trials, but not in the sensitivity analyses. However, the directionality of effect modification was consistent across all analyses. In addition, results were generally consistent between models in which outcomes were inflammation-adjusted or not (data not shown). Thus, the results presented below refer to the fixed effects models for all-trials analyses of inflammation-adjusted outcomes.

**Figure 5:**
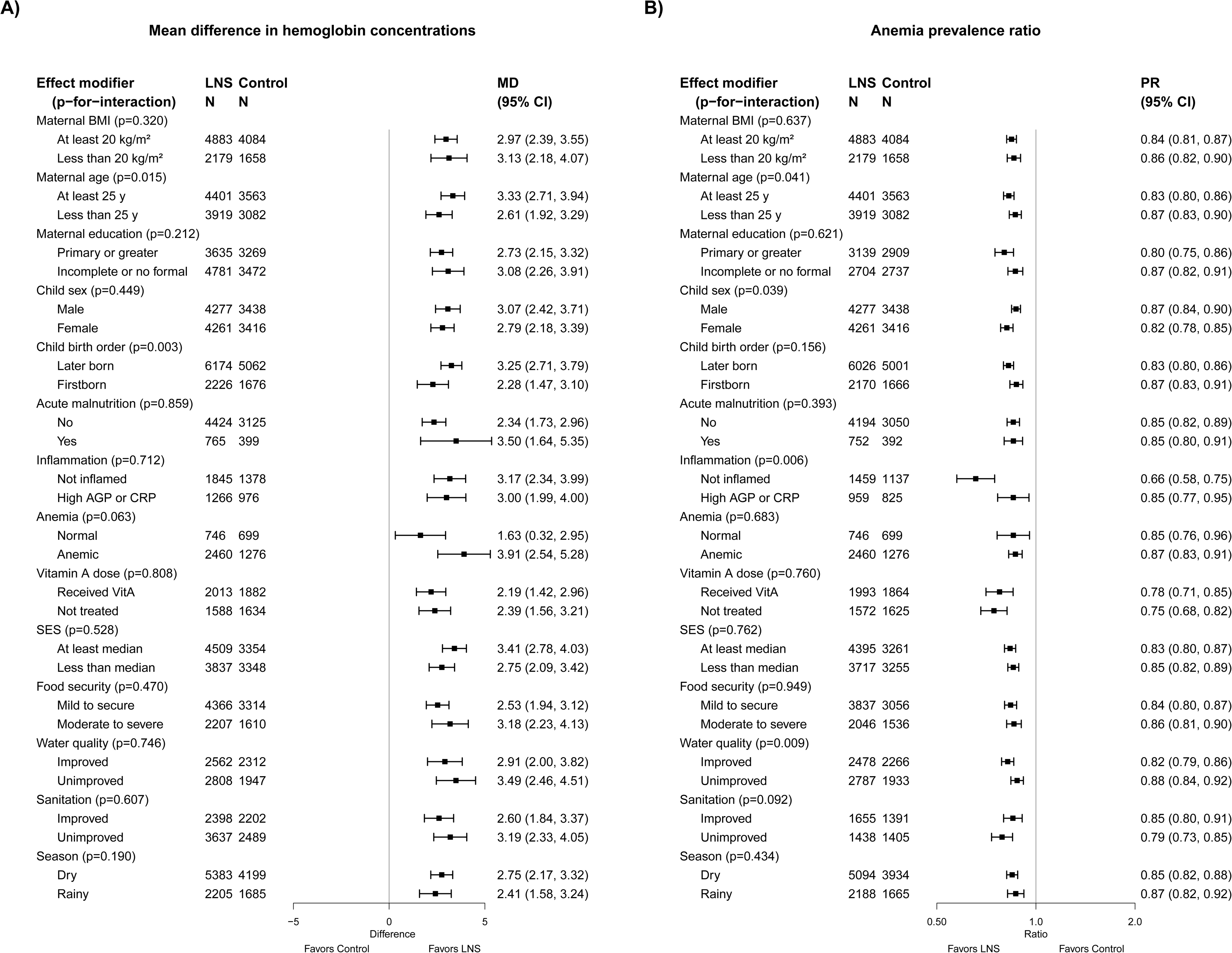
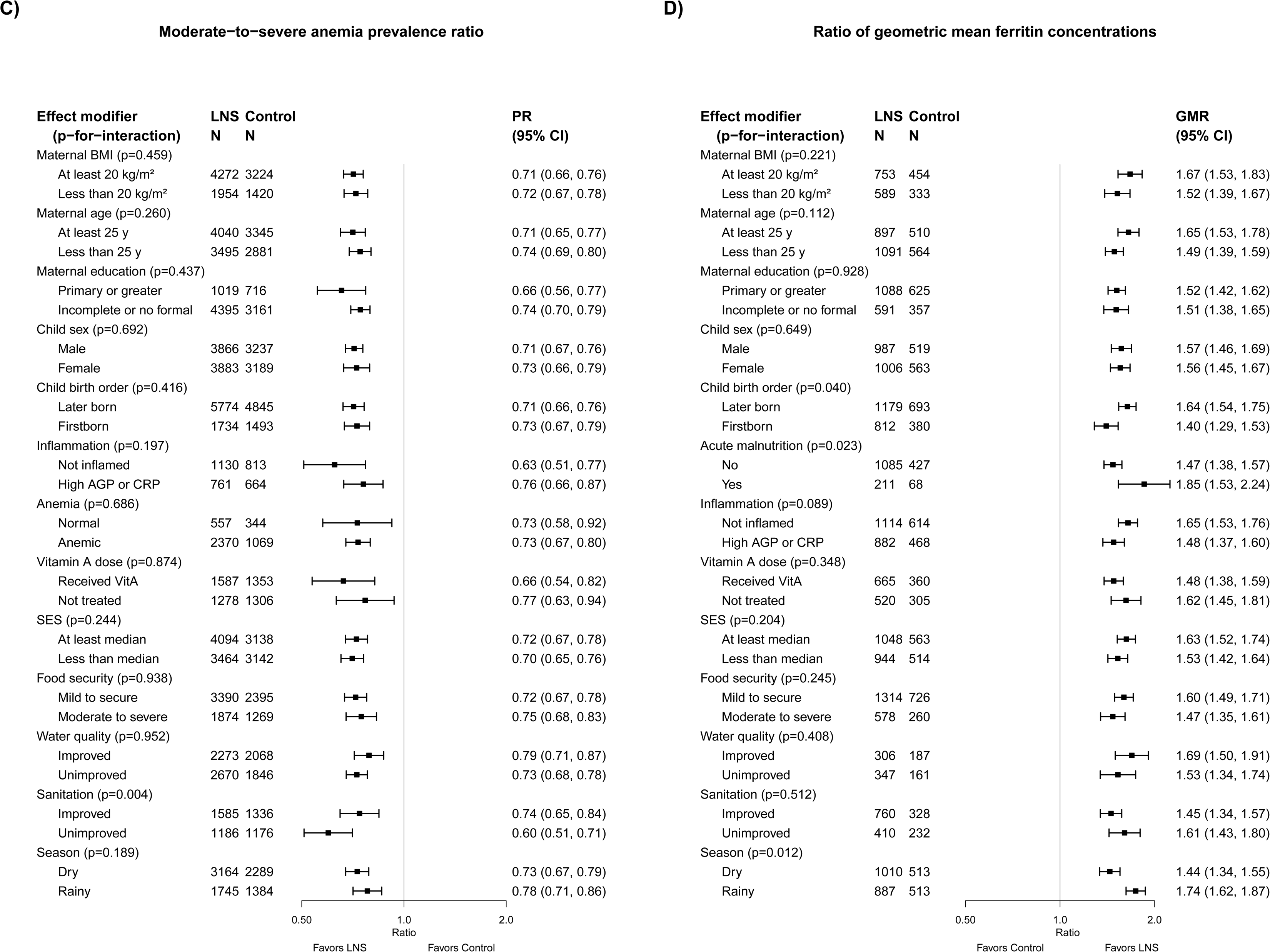
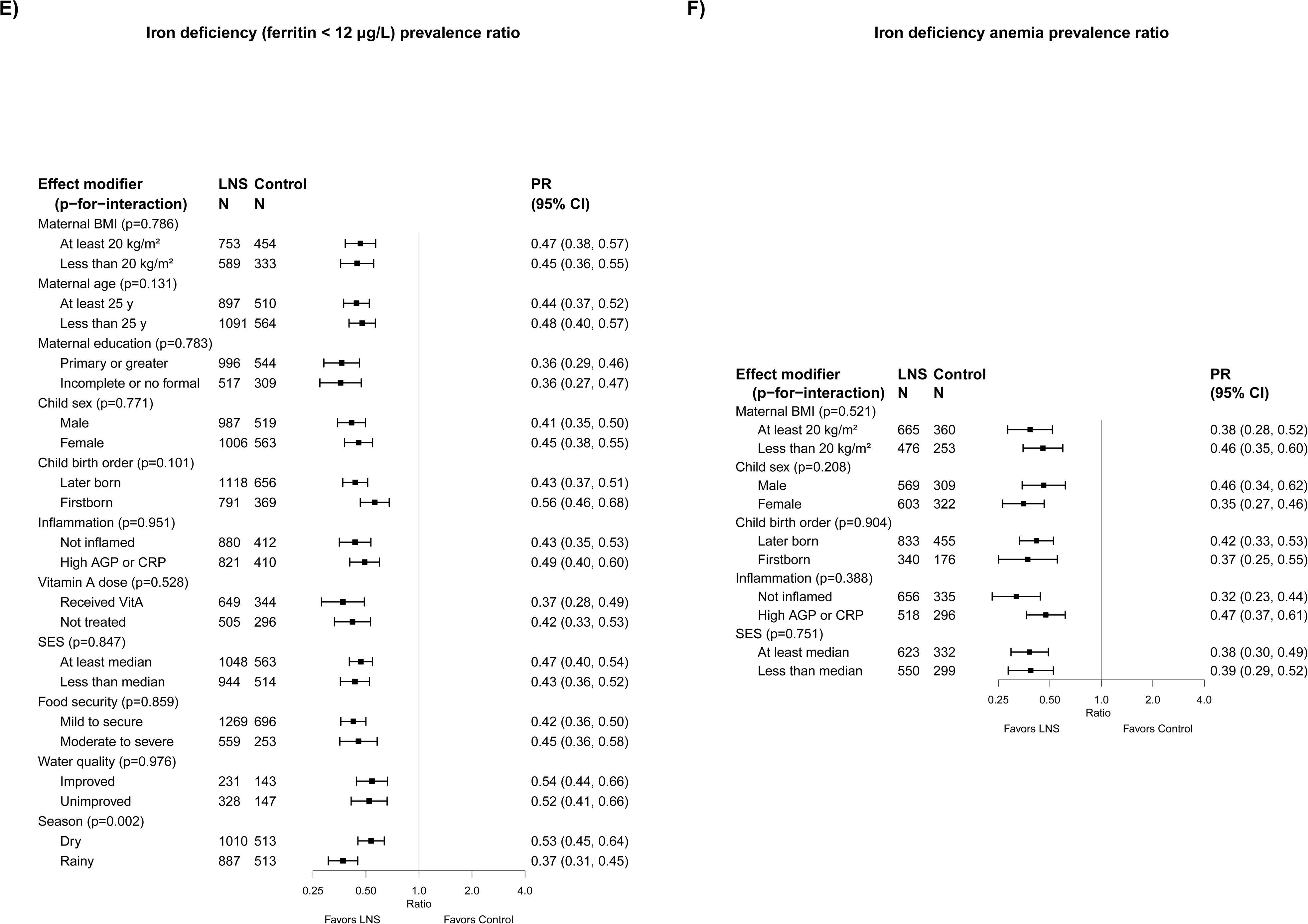
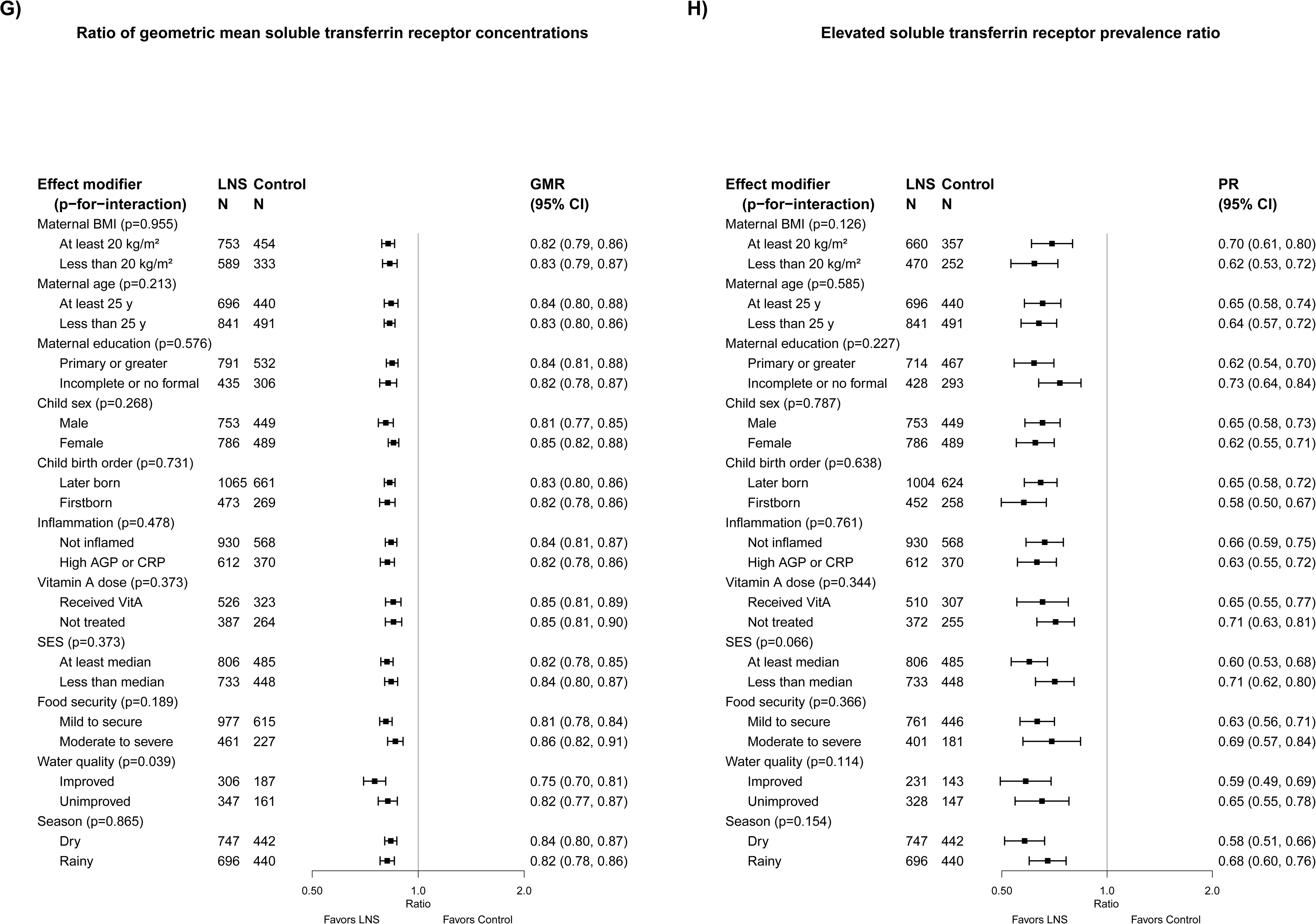
The effect of SQ-LNS provided to children 6-24 months of age compared with a control on (A) hemoglobin concentration, (B) anemia prevalence, (C) moderate-to-severe anemia prevalence, (D) ferritin concentration, (E) iron deficiency, (F) iron deficiency anemia, (G) soluble transferrin receptor concentration, and (H) elevated soluble transferrin receptor, stratified by individual-level maternal, child and household characteristics. For some biomarkers of micronutrient status, we were unable to generate pooled estimates for effect modification by certain potential individual-level effect modifiers due to an insufficient number of comparisons. GMR, ratio of geometric means, MD, mean difference, PR, prevalence ratio, p-for-interaction, p value for the interaction indicating the difference in effects of SQ-LNS between the two levels of the effect modifier.

**Table 4.**
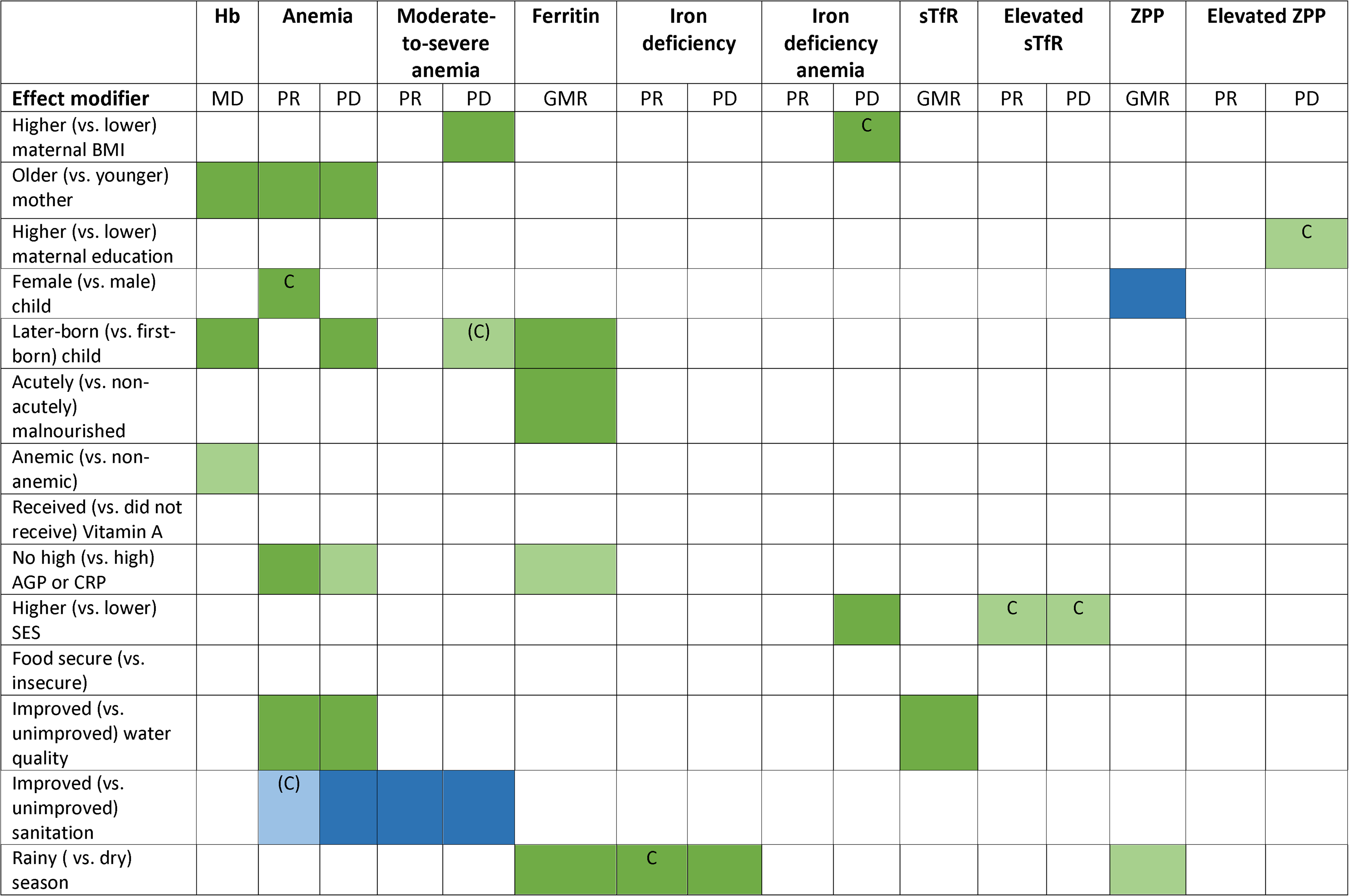
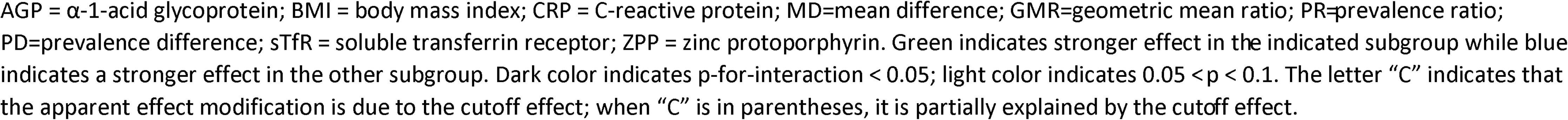
Overview of individual-level effect modification

#### Hemoglobin concentration and prevalence of anemia

In general, maternal characteristics did not significantly modify the effect of SQ-LNS on hemoglobin concentration or anemia, with two exceptions: there was a greater absolute reduction in the prevalence of moderate-to-severe anemia (8 vs. 6 percentage points, P-for-interaction = 0.047; Supplemental Figure 6E1) among children of mothers with a higher (vs. lower) BMI, and there was a greater effect of SQ-LNS on hemoglobin concentration and anemia prevalence among children of older compared to younger mothers (MD: 3.33 g/L vs. 2.61 g/L and PR: 0.83 vs. 0.87, respectively; Figures 5A-B, Supplemental Figures 6A2 and 6B2).

Regarding child characteristics, SQ-LNS had a greater effect among later-born vs. first-born children on hemoglobin concentrations (MD: 3.25 g/L vs. 2.28 g/L; Figure 5A, Supplemental Figure 6A5) and the percentage point reduction in the prevalence of anemia (12 vs. 7 percentage points; P-for-interaction = 0.024, Supplemental Figure 6C5), although not on the relative reduction in anemia prevalence (Figure 5B, Supplemental Figure 6B5). The effect of SQ-LNS on hemoglobin concentration did not differ by child sex (Figure 5A, Supplemental Figure 6A4) but we observed a greater effect of SQ-LNS on the prevalence of anemia among female vs. male children (PR: 0.82 vs. 0.87; Figure 5B, Supplemental Figure 6B4). SQ-LNS had a greater effect on hemoglobin concentrations, but not the prevalence of anemia, among children who were anemic at baseline (vs. those who were not) (MD: 3.91 g/L vs. 1.63 g/L; Figures 5A and 5B, Supplemental Figures 6A7 and 6B7). The effect of SQ-LNS on hemoglobin concentration did not differ by inflammation status at endline (Figure 5A, Supplemental Figure 6A9), however, SQ-LNS had a greater effect on the prevalence of anemia among children without (vs. with) inflammation at endline (PR: 0.66 vs. 0.85; Figure 5B, Supplemental Figure 6B9). Child acute malnutrition at baseline and receipt of high-dose vitamin A supplements did not significantly modify the effects of SQ-LNS on hemoglobin concentrations or the prevalence of anemia.

Household water quality and sanitation did not significantly modify the effects of SQ-LNS on hemoglobin concentrations (Figure 5A, Supplemental Figure 7A3-4). However, there was a greater effect of SQ-LNS on anemia prevalence among children in households with improved water quality than among children in households with unimproved water quality (PR: 0.82 vs. 0.88; Figure 5B, Supplemental Figure 7B3). Conversely, the effects of SQ-LNS on prevalences of anemia and moderate to severe anemia were greater among children in households with unimproved vs. improved sanitation (Figure 5B-C, Supplemental Figures 7B4 and 7D4). Socio-economic status (SES), food security, and season of outcome measurement did not significantly modify the effects of SQ-LNS on hemoglobin concentrations or the prevalence of anemia.

#### Biomarkers of iron status and prevalence of iron deficiency and iron deficiency anemia

In general, maternal characteristics (i.e., BMI, age and education) did not significantly modify the effect of SQ-LNS on iron status biomarkers (i.e., ferritin, sTfR or ZPP), with the exception of a greater absolute reduction in the prevalence of IDA (21 vs. 13 percentage points, P-for-interaction = 0.023; Supplemental Figure 6J1) among children of mothers with a higher (vs. lower) BMI, and greater absolute reductions in the prevalence of elevated ZPP among children of mothers with more (vs. less) formal education (14 vs. 9 percentage points, P-for-interaction = 0.080; Supplemental Figure 6P3).

We observed a greater effect of SQ-LNS on ferritin concentrations among later-born children, compared to first-born children (GMR: 1.64 vs. 1.40; Figure 5D and Supplemental Figure 6F5), among children who were acutely malnourished (vs. non-malnourished) at baseline (GMR: 1.85 vs. 1.47; Figure 5D and Supplemental Figure 6F6), and among children without (vs. with) inflammation at endline (GMR: 1.65 vs. 1.48; Figure 5D and Supplemental Figure 6F9). In addition, we observed a greater effect of SQ-LNS on ZPP concentrations in male vs. female children (GMR: 0.76 vs. 0.82; Supplemental Figure 6N4). No individual-level child characteristics modified the effect of SQ-LNS on the prevalence of iron deficiency, elevated sTfR or ZPP, or IDA.

Household characteristics (i.e., SES, food security, water quality and sanitation) did not modify the effect of SQ-LNS on ferritin or ZPP concentrations or the prevalence of iron deficiency, elevated ZPP or IDA, with the exception of greater absolute reductions in the prevalence of IDA among children of higher (vs. lower) SES (20 vs. 12 percentage points, P-for-interaction = 0.039; Supplemental Figure 7J1). We observed a greater effect of SQ-LNS on the prevalence of elevated sTfR among children in households with SES above (vs. below) the study-specific median (PR: 0.60 vs. 0.71; Figure 5H, Supplemental Figure 7L1). In addition, SQ-LNS had a greater effect on sTfR concentrations among children in households with improved (vs. unimproved) water quality (GMR: 0.75 vs. 0.82; Figure 5G, Supplemental Figure 7K3).

There was a greater effect of SQ-LNS on ferritin concentrations, and the prevalence of iron deficiency, in the rainy season than in the dry season (GMR: 1.74 vs. 1.44; PR: 0.37 vs. 0.53; Figure 5D-E, Supplemental Figures 7F6, 7G6). In addition, there was a greater effect of SQ-LNS on ZPP concentrations in the rainy season vs. the dry season, (GMR: 0.76 vs. 0.84; Supplemental Figure 7N6).

#### Biomarkers of zinc and vitamin A status

The effect of SQ-LNS on plasma zinc concentrations was greater among children of older vs. younger mothers (GMR: 1.03 vs. 0.96; Supplemental Figure 6Q2), and in the rainy season as compared to the dry season (GMR: 1.02 vs. 0.99; Supplemental Figure 7Q6). However, there was no main effect of SQ-LNS on plasma zinc concentrations, and in both aforementioned cases of effect modification, the confidence intervals around the estimates in both sub-groups included one; therefore, it is likely that there was no true effect on plasma zinc concentration in either sub-group. No other maternal, child or household characteristics significantly modified the effect of SQ-LNS on plasma zinc concentrations.

Maternal characteristics did not modify the effect of SQ-LNS on vitamin A status. There was a greater effect of SQ-LNS on RBP concentrations among children with (vs. without) inflammation at endline (GMR: 1.10 vs. 1.04; Supplemental Figure 6W9). There was also a greater effect of SQ-LNS on the prevalence of marginal vitamin A status (RBP < 1.05 µmol/L) among children in households with higher (vs. lower) SES (PR: 0.70 vs. 0.87; Supplemental Figure 7Z1). In addition, we observed a greater effect of SQ-LNS on retinol concentrations among children who were acutely malnourished (vs. non-malnourished) at baseline (GMR: 1.14 vs. 1.02; Supplemental Figure 6R6) and among children in mildly insecure or food secure (vs. moderately to severely food insecure) households (GMR: 1.06 vs. 0.99; Supplemental Figure 7R2), although there was no main effect of SQ-LNS on plasma retinol concentrations.

#### Overview of individual-level effect modification

Table 4 and **Supplemental Table 8** show that some characteristics (e.g., birth order, household water quality and sanitation, season and child inflammation) modified the effect of SQ-LNS on a number of hematological and iron status biomarkers while others (e.g., maternal education, child sex, child anemia at baseline, high-dose vitamin A supplementation, and household food security) exhibited effect modification for only a few outcomes.

The results of the simulations to identify cutoff effects are indicated in Table 4 and Supplemental Table 8. For example, among children who received SQ-LNS vs. control, the prevalence of anemia was reduced by 18% among females and by 13% among males. However, there was no significant effect modification for the continuous outcome (i.e., hemoglobin concentration). Mean hemoglobin concentrations in the control group at endline were greater among females than males (107 g/L vs. 105 g/L) and effect modification by sex became non-significant for anemia prevalence in the simulation models; therefore, the cut-off effect is the most likely explanation for these findings. The cut-off effect appeared to explain effect modification by maternal BMI, with regard to the prevalence difference for IDA, and by maternal education with regard to the prevalence difference for elevated ZPP. For the prevalence of elevated sTfR (ratio and difference), the cutoff effect appeared to explain effect modification by SES. The cutoff effect also appeared to contribute, at least partially, to the apparent effect modification by sanitation for relative anemia prevalence (but not the prevalence difference for anemia, nor the prevalence ratio or prevalence difference for moderate-to-severe anemia).

For maternal age, however, there was significant effect modification for hemoglobin concentrations and both the prevalence ratio and prevalence difference for anemia, so this was not due to the cutoff effect. In addition, the cutoff effect did not explain effect modification for anemia by maternal BMI, birth order, inflammation or water quality, for iron deficiency by season of outcome assessment, or for IDA and vitamin A deficiency by SES.

## Discussion

This individual participant data analysis included 13 RCTs in nine different countries with a total sample size of ∼15,000 children. SQ-LNS substantially reduced the prevalence of anemia, moderate-to-severe anemia, iron deficiency, and IDA among infants and young children who received SQ-LNS for 3-18 months, relative to control children. The beneficial effects of SQ-LNS on hemoglobin appeared to be greater among studies that were conducted in countries with a high burden of anemia (>60%); greater beneficial effects of SQ-LNS on anemia and iron status were observed among studies that provided SQ-LNS with a higher dose of iron and for a longer duration. Several of the individual-level characteristics also appeared to modify the effects of SQ-LNS on anemia and iron status. However, even when the magnitude of effect differed between sub-groups, the magnitude of the modifying effects was generally small and we observed positive effects of SQ-LNS within all sub-groups, indicating the potential of SQ-LNS to provide benefits across a range of individual, population and study design characteristics. This was also the case for the growth (19) and development (20) domains of this meta-analysis.

### Main effects of SQ-LNS on anemia and micronutrient status

Children who received SQ-LNS had significantly higher hemoglobin concentrations (2.77 g/L) relative to the control, and SQ-LNS reduced the prevalence of anemia and moderate to severe anemia by 16% (10 percentage points) and 28% (7 percentage points), respectively. Although these effects are smaller than what was reported in the 2019 Cochrane systematic review and meta-analysis of LNS (SQ- or MQ-LNS) for hemoglobin (MD: 5.74 (2.27, 9.30) g/L) and anemia (RR: 0.79 (0.69, 0.90)) (7), our meta-analysis includes a larger number of trials and individual participants (N = 13 trials and 15,562 participants) as compared to the Das *et al.* meta-analysis (N = 4-5 trials and 2,332-4,518 participants), which increases the precision of the effect estimates. Furthermore, we report estimates for iron deficiency and IDA, which were not included in the previous meta-analysis of SQ- and MQ-LNS. SQ-LNS reduced the prevalence of iron deficiency (plasma ferritin < 12 µg/L) by 56% (22 percentage points) and the prevalence of IDA by 64% (14 percentage points). At endline, the prevalence of IDA across all studies among children in the control groups was 23.5%, compared with 7.9% among children who received SQ-LNS. These large relative and absolute reductions in the prevalence of iron deficiency and IDA due to SQ-LNS are important given that iron deficiency is the most common documented micronutrient deficiency globally (4) and IDA is associated with compromised mental, motor, socio-emotional and neural development (51).

The findings of the present SQ-LNS meta-analysis are similar to those reported in a recent Cochrane systematic review and meta-analysis of the effect of multiple micronutrient powders (MNP) on hemoglobin (MD: 2.74 (1.95, 3.53) g/L) and anemia (RR: 0.82 (0.76, 0.90)) (N = 16-20 trials and 9,927-10,509 participants) (52). Suchdev *et al.* also reported a 53% reduction in the prevalence of iron deficiency (as defined by trialists) by MNP (N = 7 trials and 1,634 participants); the effect of MNP on the prevalence of IDA was not reported. Another meta-analysis of MNP efficacy trials by Tam *et al.* reported relative reductions in the prevalence of iron deficiency and IDA of 50% and 55%, respectively (8). Of note, in most of the MNP trials, the dose of elemental iron was 10-12.5 mg/d (primarily as ferrous fumarate), whereas the SQ-LNS trials in our analyses provided only 6-9 mg/d (primarily as ferrous sulfate).

In the present analyses, there was considerable heterogeneity in the effect of SQ-LNS on anemia (reductions ranging from 3 to 29 percentage points), iron deficiency (reductions ranging from 14 to 35 percentage points), and IDA (reductions ranging from 8 to 29 percentage points). This heterogeneity may be due to differences in population characteristics (e.g., baseline prevalence of anemia, iron deficiency and IDA, proportion of anemia attributable to nutritional deficiencies vs. underlying burdens of infection and other causes) and study-design characteristics. The more modest relative reduction in the prevalence of anemia than in IDA may reflect the influence of non-nutritional causes of anemia in these populations (e.g., genetic hemoglobin disorders and infection and inflammation including malaria, intestinal parasites and schistosomiasis). In addition, although SQ-LNS substantially reduced the prevalence of IDA, it did not completely eliminate IDA, possibly reflecting factors such as poor iron absorption (due to anti-nutritional compounds, gastric pH or intestinal or systemic inflammation) (1) or a hemoglobin cutoff for anemia that is set inappropriately high for infants and young children, especially in African populations (53–55). However, it is noteworthy that the endline prevalence of iron deficiency among children who received SQ-LNS was approximately 16% (vs. 40% among children in the control groups), which is comparable to the prevalence of iron deficiency among children 12-36 months of age in the United States of America (13.5%) (56).

We observed significant increases in RBP concentrations among children who received SQ-LNS, although the relative increase was small (7%). The difference in plasma retinol was in the same direction, but not statistically significant. Our ability to draw conclusions was limited due to small sample sizes (n = 663-1236). However, in populations with adequate vitamin A stores, serum retinol may not respond to an intervention due to homeostatic regulation (57). At endline, the prevalence of vitamin A deficiency (retinol or RBP < 0.70 µmol/L) in the control groups was low, ranging from 1.1 to 16.2%. In the majority of studies in this analysis that assessed vitamin A status (7 of 9), more than half of study participants had received high-dose vitamin A supplementation (100,000 – 200,000 IU) within the 6 months prior to outcome assessment. Therefore, in some study contexts, the daily low-dose of vitamin A provided in SQ-LNS may not have provided an additive benefit. However, SQ-LNS did reduce the prevalence of vitamin A deficiency (RBP < 0.70 µmol/L) by 56% (corresponding to a modest 3 percentage point reduction, due to the low overall prevalence of deficiency) indicating the potential of this intervention to improve the status of those at-risk. There was no overall effect of the intervention on plasma zinc concentrations, consistent with results from zinc-fortified food and MNP trials (52, 58), potentially due to the low bioavailability of zinc in SQ-LNS when provided as part of a phytate-rich food matrix (59), or differences in post-absorptive metabolism between zinc supplements and zinc-fortified foods (60).

### Effect modification

For both study- and individual-level effect modifiers, it is important to distinguish between the potential to *benefit* and the potential to *respond* (18). The potential to benefit is more likely when a population or an individual child is more vulnerable, e.g. due to a high prevalence of anemia or micronutrient deficiencies in the study population at baseline. However, in some cases, children who are more vulnerable may actually be *less* likely to respond to a nutritional intervention because of other constraints on nutritional status such as infection and inflammation or inadequate care. Thus, we will attempt to frame the following discussion of effect modifiers in this context.

### Effect modification by study-level characteristics

Studies included in this analysis were conducted in a variety of geographic and environmental contexts (e.g., different regions, burdens of anemia and infection including malaria, community-level access to improved water, sanitation and hygiene, etc.) and used varying intervention designs (e.g. durations of supplementation, iron dose, frequency of contact between participants and study staff) and this enables us to examine whether these study context and design features modify the effects of SQ-LNS. However, it is difficult to disentangle the impact attributable to one effect modifier from the influence of other characteristics of the study and thus these results should be interpreted with caution. For example, studies that provided a higher iron dose were more likely to provide SQ-LNS supplements for a longer duration, be conducted in South Asia, and have a higher average compliance with supplementation (>80 %), thus making it difficult to disentangle the impact attributable to each specific study-level characteristic. In addition, it is important to note that due to concerns about adverse effects of iron supplementation in malaria endemic areas (61, 62), a number of the SQ-LNS studies in these analyses were purposefully designed to provide a lower iron dose in countries with a high burden of malaria (6).

The effect of SQ-LNS on hemoglobin concentrations was greater in studies with a higher burden of anemia (>60%), suggesting a greater potential to benefit from SQ-LNS in such populations. Similarly, the effect of MNP on hemoglobin concentrations has been shown to be greater in populations that were anemic at baseline vs. populations with mixed/unknown baseline anemia status (MD: 4.53 g/L vs. 3.05 g/L) (52). However, even in populations with a lower burden of anemia, both SQ-LNS and MNP significantly increased hemoglobin concentrations and reduced the prevalence of anemia. In the present analyses, increases in hemoglobin concentrations due to SQ-LNS were not significantly different between populations with and without a high burden of malaria (4.29 g/L vs. 2.59 g/L) or inflammation (3.84 g/L vs. 3.21 g/L). Thus, these results, coupled with the observation that statistically significant effects of SQ-LNS were found in both subgroups, do not support the hypothesis that populations with a high burden of malaria and inflammation have less of a potential to respond to iron interventions due to increased hepcidin concentrations and down-regulation of iron absorption (63, 64).

Effect modification by study-level water quality and sanitation did not reach statistical significance for the majority of hematological and iron status outcomes, with the exception of a greater percentage point reduction in iron deficiency due to SQ-LNS in study sites with a lower proportion of households with access to improved water quality and sanitation. This is likely due to a higher prevalence of iron deficiency among children in the study sites with less access to improved water quality and sanitation; among control group children, prevalence of iron deficiency was 57% in those study sites, compared to 28% in sites with greater access.

Greater reductions in the prevalence of anemia and iron deficiency were observed in trials that provided SQ-LNS for longer than 12 months, compared to a shorter duration (6-12 months). In addition, studies that reported higher average compliance (>80 %) showed greater increases in hemoglobin concentrations and reductions in the prevalence of anemia, but we did not observe significant effect modification by frequency of contact with programmatic or study staff. Finally, we observed greater effects of SQ-LNS on the prevalence of anemia and increase in ferritin concentrations among studies that provided 9 mg/d vs. < 9 mg/d iron. Although it may be difficult to disentangle the impact attributable to each of these aforementioned study-level characteristics, they all suggest a greater effect among studies in which children received a higher total iron dose, either through a longer duration of supplementation, by higher compliance, or a higher daily iron dose. Consistent with this, meta-analyses of the effectiveness of MNP and iron supplements in infants and young children have also shown higher iron doses to have a greater impact on iron and hematological status (8, 52).

### Effect modification by individual-level characteristics

The effects of SQ-LNS on hemoglobin concentrations and the prevalence of anemia were greater among children of older mothers compared to those of younger mothers (relative reductions in anemia of 17% vs. 13%, respectively). Similarly, SQ-LNS had a larger effect on both hemoglobin and ferritin concentrations among later-born, as compared with first-born children, and reduced the prevalence of anemia in these sub-groups by 12 vs. 7 percentage points, respectively. SQ-LNS also had a larger effect on stunting, underweight and MUAC among later-born vs. first-born children (19). These results likely indicate a greater potential to benefit among later-born children. For example, hemoglobin concentrations among control group children in this IPD analysis were lower among later-born than first-born children (105.5 g/L vs. 108.4 g/L). Later-born children, who have one or more older siblings, may compete more for caregiving and family resources than first-born children, making them more vulnerable to anemia and iron deficiency and therefore more likely to benefit from nutritional supplementation targeted specifically to young children. In addition, later-born children, who are more likely to be born to older mothers, may be more vulnerable to micronutrient deficiencies due to poor maternal physiological status owing to high fertility rates and short inter-pregnancy intervals (65, 66).

The effects of SQ-LNS on biomarkers of hematological and iron status (i.e., ferritin, sTfR, ZPP) did not differ by child sex, with the exception of greater reductions in ZPP concentrations due to SQ-LNS among males vs. females (24% vs. 18%). The former may have had a greater potential to benefit, given that median ZPP concentrations were higher among control group males than females in this analysis (62.0 µmol/mol heme vs. 53.1 µmol/mol heme), and males tend to be more vulnerable to iron deficiency in infancy (67). However, both sexes responded positively to the intervention, with a 10 percentage point reduction in the prevalence of anemia and a 20-23 percentage point reduction in the prevalence of iron deficiency. Children who were anemic at baseline had greater increases in hemoglobin concentrations due to SQ-LNS than children who were not anemic at baseline, indicating a greater potential to benefit. A recent MNP trial also showed greater increases in hemoglobin concentrations due to MNP among children who were anemic vs. non-anemic at baseline (68). In addition, these results are consistent with our study-level effect modification results, which indicated that increases in hemoglobin due to SQ-LNS were greater in populations with a high vs. moderate burden of anemia. Increases in ferritin concentrations due to SQ-LNS were larger among children who were acutely malnourished at baseline, compared to those who were not. Among children in the control groups, median ferritin concentrations were lower among children with vs. without acute malnutrition (15.5 µg/L vs. 19.5 µg/L), also suggestive of a greater potential to benefit.

Although differences in the ability of populations to benefit or respond to SQ-LNS by the study-level burden of malaria or inflammation appeared to be explained by the cut-off effect, we did observe some differences at the individual level. SQ-LNS reduced the prevalence of anemia by 34% (a 12 percentage point difference) among children without concurrent inflammation vs. 15% (a 5 percentage point difference) among children with elevated CRP and/or AGP concentrations, indicating a greater potential to respond among those without inflammation. As inflammation is itself a cause of anemia (i.e., anemia of chronic disease) and can inhibit absorption of iron (63), it is likely that a higher proportion of anemia in the subgroup of non-inflamed children was amenable to nutritional supplementation. Among children with inflammation, non-nutritional causes of anemia may have been more prevalent (e.g., anemia attributable to acute or chronic infection and inflammation) (29, 69). In the present analyses, the proportion of anemia not due to iron deficiency was greater (54% vs. 35%) among children with inflammation vs. those without. In addition, it is possible that children without inflammation had a greater fractional absorption of iron relative to those with inflammation (70). Hepcidin, an important regulator of systemic iron balance, is elevated by inflammation thereby inhibiting iron absorption (63). While a few trials measured hepcidin, we had insufficient data to examine it in this analysis. However, greater increases in ferritin concentrations due to SQ-LNS were observed among children without inflammation (65% vs. 48%), although there were no differences in the effect of SQ-LNS on iron deficiency by inflammation status.

Greater effects of SQ-LNS on reductions in the prevalence of anemia, and reductions in sTfR concentrations, were seen among children in households with improved (vs. unimproved) water quality. Poor quality water or sanitation may increase the risk of anemia due to infection with soil transmitted helminths or environmental enteric dysfunction, which may increase inflammation or reduce nutrient absorption. Among children in the control group, the prevalence of anemia was higher among children without (vs. with) access to improved water quality (69% vs. 56%). Thus, this effect modification might be seen as consistent with the inflammation effect modification results described above. Children with less exposure to gastrointestinal pathogens through contaminated water supplies may have less inflammation and thus may be more able to respond to the SQ-LNS intervention. However, these findings are in contrast with the observed greater effects of SQ-LNS on anemia and moderate-to-severe anemia among children in households with unimproved (vs. improved) sanitation. This may indicate a greater potential to benefit, given that the prevalences of anemia and moderate-to-severe anemia were higher among control group children in households with unimproved sanitation (by 7 and 10 percentage points, respectively). The contradictory effect modification results for water quality and sanitation are difficult to explain. It is notable, though, that water quality and sanitation did not modify the effect of SQ-LNS on hemoglobin concentrations, and there was evidence of beneficial effects of SQ-LNS in all subgroups of children.

Greater effects of SQ-LNS on ferritin concentrations and the prevalence of iron deficiency were seen among children for whom biomarkers were assessed in the rainy season. Among children in the control groups in this analysis, median ferritin concentrations were lower (13.9 µg/L vs. 16.1 µg/L), and the prevalence of iron deficiency higher (by 6 percentage points) when outcome assessments occurred during the rainy (vs. dry) season. Thus, it is possible that this represents a greater potential to benefit, whereby the supplemental iron provided by SQ-LNS compensates for seasonal differences in dietary intake and/or constraints on iron absorption (e.g., infection and inflammation). However, iron status at a given point in time reflects the cumulative effect of intake, absorption and utilization over many months, so it is unclear whether effect modification by season is due to short-term, acute phenomena or longer-term exposure to adverse conditions that is correlated with season at outcome assessment. Other household-level characteristics, including SES and food security, did not generally modify the effect of SQ-LNS on anemia or iron biomarker outcomes.

With regard to individual-level effect modification of outcomes related to zinc and vitamin A status, observed differences between the stratum-specific point estimates were generally small even when there were statistically significant p-values. In the latter situation, results were generally consistent with effect modification for hematological and iron status biomarkers, whereby children of older mothers, children who were acutely malnourished at baseline, children in households of higher SES and with greater food security, and children assessed during the rainy season had greater increases in biomarker concentrations or reductions in the prevalence of deficiency vs. those in the respective comparison sub-groups.

### Strengths and limitations

Strengths of the present analyses include the large sample size, the substantial number of high-quality RCTs available, and the availability of individual participant data for all but one of the eligible trials. In addition, we were able to report the effects of SQ-LNS on multiple indicators of micronutrient status that had not been included in prior meta-analyses. The majority of trials used the same analytical laboratory and platform for several biomarkers (ferritin, sTfR, RBP; Vit-Min Lab) or standardized analytical methods (ZPP), lending strength to the findings. The trial sites were diverse in terms of study context (e.g., burdens of anemia, malaria and inflammation) and design (e.g., iron dose and duration of supplementation), which provided heterogeneity for exploration of study-level effect modifiers. The findings were generally consistent across sensitivity analyses, as well as between fixed- and random-effects models, adding strength to the conclusions.

Several limitations must be considered. Bangladesh was the only country represented in these analyses outside of sub-Saharan Africa. Although hemoglobin data were available for all thirteen trials, fewer studies assessed biomarkers of iron, zinc and vitamin A status (3-7 studies) and sample sizes for each of those biomarkers were smaller (1,133 – 3,078 vs. 15,398 for hemoglobin). In addition, data were not yet available from a sufficient number of studies to be able to investigate the effects of SQ-LNS on additional biomarkers, including plasma hepcidin, folate, and vitamin B12 concentrations. Furthermore, not all trials assessed both CRP and AGP in all children at endline, potentially limiting the accuracy of adjustments for inflammation and/or reducing the available sample size. It should also be noted that a single assessment of inflammation may not adequately characterize the inflammation status of individual children over time, and the effects that average inflammation status over the entire period of supplementation may have on micronutrient status.

Due to limitations in the data (e.g., number of studies that assessed biomarker outcomes or effect modifiers of interest, or a low prevalence of the binary outcome or proportion of children within one of the effect modifier subgroups), we were unable to generate effect estimates of SQ-LNS on all biomarkers by all potential effect modifiers for all trials. Overall, statistical power for study-level effect modification was constrained by the limited number of trials, so there may be meaningful differences in effect estimates between categories of trials even if the p-diff for interaction was not significant. On the other hand, the individual-level effect modification analyses involved multiple effect modifiers and numerous outcomes, so several of the significant p-for-interaction values are likely due to chance. As stated in the Methods section, we did not adjust for multiple hypothesis testing because the effect modification analyses are inherently exploratory. Finally, effect modification results should be interpreted with caution, as potential effect modifiers may be inter-related or confounded by unmeasured variables. This is particularly important for study-level characteristics because, as previously noted, it was not possible to completely disentangle the impact attributable to a specific effect modifier from the impact attributable to other characteristics of the study (e.g., study design, context, implementation, etc.).

### Programmatic Implications

The present findings suggest that policy-makers and program planners should consider SQ-LNS in the mix of interventions to prevent anemia and iron and vitamin A deficiencies. The overall effects of SQ-LNS on iron deficiency (56% reduction) and IDA (64% reduction) were substantial and may improve child neurological development and immune function (71). Although the overall effects on anemia and moderate-to-severe anemia were more modest (16% and 28% reductions, respectively), this likely reflects the presence of anemias in these populations that are not nutrition responsive (e.g., genetic hemoglobin disorders and infection and inflammation). Alternative interventions, including micronutrient supplementation, MNP and food fortification are also effective in reducing anemia and the prevalence of iron deficiency (8, 52); however, SQ-LNS provides the added benefits of reducing mortality (72), stunting and wasting (19) and improving developmental outcomes (20).

In terms of program design, the effect modification results herein suggest that a greater impact of SQ-LNS on hematological and iron status outcomes may be obtained by providing formulations with the higher dose of iron (9 mg/d) and/or providing SQ-LNS for the entire window from 6-24 months. The iron dose of fortified products has been of concern because of evidence that iron-containing supplements and MNP may increase susceptibility to malaria in endemic regions, as well as respiratory and gastrointestinal infections (61, 73–75), although two recent systematic reviews and meta-analyses did not indicate any increase in the risk of diarrhea or malaria from these interventions (52, 76). The present analysis did not examine potential adverse impacts (e.g., morbidity or mortality) of SQ-LNS by iron dose, malaria burden or other potential effect modifiers. However, most published trials have not reported differences in diarrheal or malarial morbidity between SQ-LNS and control groups (40, 42–44, 47, 48, 77–79), and some reported beneficial effects of SQ-LNS on diarrheal prevalence (37) and duration of pneumonia, diarrhea and dysentery (35). In addition, a recent meta-analysis reported an overall 27% reduction in the risk of mortality among children who received SQ-LNS vs. control (72). The majority of studies providing the higher iron dose were in lower malaria burden sites (36, 37, 42), and as such, we cannot recommend universal use of higher iron dose SQ-LNS supplements at this time. In areas with a low prevalence of malaria and inflammation, or in malaria endemic areas with a well-integrated control program with appropriate surveillance and the prevention and management of malaria, providing the higher iron dose (9 mg/d) in SQ-LNS may be appropriate and have a greater impact on iron status and hematological parameters. Provision of SQ-LNS containing 9 mg/d iron for 18 months would cumulatively provide approximately 4,925 mg of iron, compared with the 2,200 mg of iron provided by 12 months of SQ-LNS containing 6 mg/d iron, but would increase programmatic costs.

The effect modification results for biochemical outcomes generally did not provide a strong rationale for *targeting* SQ-LNS only to the most vulnerable children or populations. Even in situations that were indicative of a greater potential to benefit among one sub-group compared to another (e.g., greater increases in hemoglobin concentrations among later-born vs. first-born children and among anemic vs. non-anemic children, greater increases in ferritin concentrations among acutely malnourished vs. non-malnourished children), both sub-groups responded positively and appeared to benefit from the intervention. However, some of the results suggested that a greater impact of SQ-LNS may be obtained by *combining* supplementation with interventions that address factors related to the potential to respond. Integrating the provision of SQ-LNS with interventions to address the prevention and control of infection and inflammation (e.g., household and/or community level improvements in water, sanitation and hygiene, use of insecticide treated bed nets, and surveillance and treatment of diarrhea and malaria) may increase nutrient (iron) absorption, and thus increase the efficacy of the supplement.

There is now substantial evidence demonstrating the efficacy of SQ-LNS for prevention of anemia and iron deficiency. However, further research is needed to investigate the efficacy of SQ-LNS for prevention of other vitamin and mineral deficiencies. In the present analysis, there was insufficient evidence to assess the effect of SQ-LNS on folate and vitamin B12 status, and no trials assessed status regarding other B-vitamins, vitamin C, vitamin D and vitamin E, or minerals other than iron and zinc. Additional research is also needed on the optimal iron dose in SQ-LNS and the duration of supplementation, with regard to not only iron status and anemia but also morbidity, the costs and benefits of shorter vs. longer supplementation, and the potential to align the intervention with periods of increased vulnerability. Finally, further research would be useful to determine the optimal doses and formulations of micronutrients to include in SQ-LNS, and to investigate the effect of additional compounds, such as phytase or galacto-oligosaccharides, that may improve mineral bioavailability.

## Supporting information

PRISMA Checklist

## Data Availability

Data described in the manuscript, code book, and analytic code will not be made available because they are compiled from 13 different trials, and access is under the control of the investigators of each of those trials.

## Abbreviations

AGP: α-1-acid glycoprotein
BMI: body-mass index
CRP: C-reactive protein
GMR: geometric means ratio
Hb: hemoglobin
IPD: individual participant data
LNS: lipid-based nutrient supplement
MD: mean difference
MN: micronutrient
MNP: multiple micronutrient powder
MQ-LNS: medium-quantity lipid-based nutrient supplements
MUAC: mid-upper arm circumference
PD: prevalence difference
pF: plasma ferritin
PR: prevalence ratio
pZn: plasma zinc
RBP: retinol binding protein
RCT: randomized controlled trial
SES: socio-economic status
SQ-LNS: small-quantity lipid-based nutrient supplements
sTfR: soluble transferrin receptor
WASH: water sanitation and hygiene
WAZ: weight-for-age z-score
WLZ: weight-for-length z-score
ZPP: zinc protoporphyrin

## Acknowledgments

We thank all of the co-investigators, collaborators, study teams, participants and local communities involved in the trials included in these analyses. These trials benefitted from the contributions of many partner organizations, including: icddr,b (JiVitA-4, Rang-Din Nutrition Study and WASH Benefits trial in Bangladesh); the World Food Program (JiVitA-4 trial in Bangladesh); the Health District of Dandé and the relevant local health-care authorities (iLiNS-ZINC trial in Burkina Faso); AfricSanté and Helen Keller International (PROMIS trials in Burkina Faso and Mali); Innovations for Poverty Action and the Kenya Medical Research Institute (WASH-Benefits trial in Kenya); Unité Programme National de Nutrition Communautaire, Government of Madagascar, Institute Pasteur, and World Bank Health and Nutrition and Population Global Practice (MAHAY trial in Madagascar); the Ministry of Health and Child Care in Harare, Chirumanzu and Shurugwi districts, and Midlands Province (SHINE trial in Zimbabwe); the International Lipid-based Nutrient Supplements Project Steering Committee (iLiNS Project trials); and Nutriset (for development of SQ-LNS). We thank Emily Smith for advice on IPD analysis methods.

The authors’ responsibilities were as follows—KRW: drafted the manuscript with input from KGD, CDA, ELP, CPS and other coauthors; KRW, CDA, KGD, ELP and CPS: wrote the statistical analysis plan; BFA, PA, EB, LH: reviewed, contributed to, and approved the statistical analysis plan; KRW and CDA: compiled the data; CDA: conducted the data analysis; and all authors: read, contributed to, and approved the final manuscript.

Supported by Bill & Melinda Gates Foundation grant OPP49817 (to KGD). BA received travel support (airfare and hotel) covered by the Bill & Melinda Gates Foundation to attend meetings in Seattle during the period of this IPD analysis project; PC was an employee of the Bill & Melinda Gates Foundation when this project was conceived until December 2019. All other authors report no conflicts of interest.

## Supplemental Tables

Supplemental Table 1: Characteristics of trials included in the individual participant data analysis, outcomes assessed and the analytic contrasts in which they were included

Supplemental Table 2: Amount of SQ-LNS provided (g/day) and nutrient value (per daily ration)

Supplemental Table 3: Descriptive information on potential study-level effect modifiers, by trial

Supplemental Table 4: Descriptive information on potential individual-level effect modifiers, by trial

Supplemental Table 5: Description of biochemical assessments

Supplemental Table 6: Biomarker outcomes at endline among control groups, by trial

Supplemental Table 7: Risk of bias assessment in each trial

Supplemental Table 8: Overview of individual-level effect modification for zinc and vitamin A

## Supplemental Figures

Supplemental Figure 1: Summary risk of bias as a percentage of all included studies

Supplemental Figure 2: Sensitivity analyses of main effects of SQ-LNS on biochemical outcomes

Supplemental Figure 3: Forest plots for all main effects of SQ-LNS on biochemical outcomes

Supplemental Figure 4: Forest plots for effects of SQ-LNS on biochemical outcomes stratified by study-level effect modifiers

Supplemental Figure 5: Sensitivity analyses of effect modification of SQ-LNS on biochemical outcomes by study-level effect modifiers

Supplemental Figure 6: Forest plots for effects of SQ-LNS on biochemical outcomes stratified by individual-level maternal and child effect modifiers

Supplemental Figure 7: Forest plots effects of SQ-LNS on biochemical outcomes stratified by individual-level household effect modifiers

Supplemental Figure 8: Sensitivity analyses of effect modification of SQ-LNS on biochemical outcomes by individual-level effect modifiers

