## Supplemental Tables 1-6 for "Characteristics that modify the effect of small-quantity lipid-based nutrient supplementation on child anemia and micronutrient status: an individual participant data meta-analysis of randomized controlled trials"

**Supplemental Table 1:** Characteristics of trials included in the individual participant data analysis, outcomes assessed and the analytic contrasts in which they were included

| Country | Years of study | Study Name | Trial design | Author | Intervention Groups | Selection of biochemistry sub-sample | Endline samples collected during supplementation | Outcomes assessed | | | | | | | | | | Analysis Contrasts | | | |
|---|---|---|---|---|---|---|---|---|---|---|---|---|---|---|---|---|---|---|---|---|---|
| | | | | | | | | Hb | Ferritin | sTfR | ZPP | Hepcidin | Zinc | Retinol | RBP | Folate | B12 | All-trials analysis | Child-LNS only analysis | Separation of multi-component arms | Passive control arms excluded |
| Bangladesh | 2012-2014 | JiVitA-4 | cluster RCT; longitudinal follow-up | Christian, 2015 (35); Campbell, 2020 (15) | Plumpy'Doz[1] + IYCF counseling | Subset selected to be geographically contiguous, accessible, socioeconomically representative and balanced by number of participants per treatment arm | Y | Y | Y | ----- | ----- | ----- | Y | Y | ----- | ----- | ----- | LNS | LNS | LNS | LNS |
| | | | | | Chickpea based LNS + IYCF counseling | | | | | | | | | | | | | LNS | LNS | LNS | LNS |
| | | | | | Rice-lentil LNS + IYCF counseling | | | | | | | | | | | | | LNS | LNS | LNS | LNS |
|  |  |  |  |  | Wheat-soy blend (++) + IYCF counseling |  |  |  |  |  |  |  |  |  |  |  |  | ----- | ----- | ----- | ----- |
| | | | | | IYCF counseling only (Control) | | | | | | | | | | | | | control | control | control | control |
| Bangladesh | 2011-2015 | RDNS | cluster RCT; longitudinal follow-up | Dewey, 2017 (36); Matias, 2018 (10) | LNS-LNS: maternal SQ-LNS in pregnancy + 6 mo postpartum, child SQ-LNS 6-24 mo | During enrollment, a total of 1346 (unborn) children were randomly assigned to the child biochemical sample | Y | Y | Y | Y | ----- | ----- | ----- | ----- | Y | ----- | ----- | LNS | ----- | ----- | ----- |
| | | | | | IFA-LNS: maternal IFA in pregnancy + 3 mo postpartum, child SQ-LNS 6-24 mo | | | | | | | | | | | | | LNS | LNS | LNS | LNS |
|  |  |  |  |  | IFA-MNP: maternal IFA in pregnancy + 3 mo postpartum, child MNP 6-24 mo |  |  |  |  |  |  |  |  |  |  |  |  | ----- | ----- | ----- | ----- |
| | | | | | IFA-Control: maternal IFA in pregnancy + 3 mo postpartum, no child supplementation | | | | | | | | | | | | | control | control | control | control |
| Bangladesh | 2012-2015 | WASH-Benefits | cluster RCT; cross-sectional surveys | Luby, 2018 (37); Stewart, 2019 (12) | Nutrition: SQ-LNS + IYCF counseling | Sub-study clusters were selected based on the logistical feasibility of the preservation and collection of biological specimens, as well as their transport to the central laboratory<br>Exclusion criteria included symptoms of moderate to severe dehydration on the day of the survey | Dependent on age | Y | Y | Y | ----- | Y | ----- | ----- | Y | Y | Y | LNS | LNS | LNS | LNS |
|  |  |  |  |  | Water: family received chlorine for drinking water |  |  |  |  |  |  |  |  |  |  |  |  | control | control | ----- | control |
|  |  |  |  |  | Sanitation: family received upgraded latrine, sani-scoop and child potty |  |  |  |  |  |  |  |  |  |  |  |  | control | control | ----- | control |
|  |  |  |  |  | Handwashing: family received handwashing stations with soap |  |  |  |  |  |  |  |  |  |  |  |  | control | control | ----- | control |
| | | | | | WASH: family received all water, sanitation and hygiene interventions | | | | | | | | | | | | | control | control | control-WSH | control |
| | | | | | WASH + nutrition: all water, sanitation, hygiene and nutrition interventions | | | | | | | | | | | | | LNS | LNS | LNS-WSH | LNS |
|  |  |  |  |  | Passive control (no intervention) |  |  |  |  |  |  |  |  |  |  |  |  | control | control | control | ----- |
| Burkina Faso | 2010-2012 | iLiNS-ZINC | cluster RCT; longitudinal follow-up | Hess, 2015 (38); Abbeddou 2017 (11) | LNS-Zn0: SQ-LNS containing 0 mg/d Zn and placebo tablet[2] | Subset of 1065 children were randomly selected; one child included per concession<br>Excluded children with reported fever or diarrhea symptoms on the day of enrollment or on the scheduled blood collection day. | Y | Y | Y | Y | Y | ----- | Y | ----- | Y | ----- | ----- | LNS | LNS | LNS | ----- |
|  |  |  |  |  | LNS-Zn5: SQ-LNS containing 5 mg/d Zn and placebo tablet |  |  |  |  |  |  |  |  |  |  |  |  | LNS | LNS | LNS | ----- |
|  |  |  |  |  | LNS-Zn10: SQ-LNS containing 10 mg/d Zn and placebo tablet |  |  |  |  |  |  |  |  |  |  |  |  | LNS | LNS | LNS | ----- |
|  |  |  |  |  | LNS-TabZn5: SQ-LNS containing 0 mg/d Zn and Zn tablet containing 5 mg/d Zn |  |  |  |  |  |  |  |  |  |  |  |  | LNS | LNS | LNS | ----- |
|  |  |  |  |  | Passive control (no intervention) |  |  |  |  |  |  |  |  |  |  |  |  | control | control | control | ----- |
| Burkina Faso | 2015-2017 | PROMIS | cluster RCT; cross-sectional surveys | Becquey, 2019 (39) | SQ-LNS + IYCF counseling | N/A (all children in cross-sectional survey ≥ 3 months of age) | Y | Y | ----- | ----- | ----- | ----- | ----- | ----- | ----- | ----- | ----- | LNS | LNS | LNS | LNS |
| | | | | | Active control (standard of care) | | | | | | | | | | | | | control | control | control | control |
| Ghana | 2004-2005 |  | RCT; longitudinal follow-up | Adu-Afarwuah, 2007 (40); Adu-Afarwuah, 2008 (9) | SQ-LNS | N/A | Y | Y | Y | Y | ----- | ----- | Y | Y | ----- | ----- | ----- | LNS | LNS | LNS | ----- |
|  |  |  |  |  | MNP |  |  |  |  |  |  |  |  |  |  |  |  | ----- | ----- | ----- | ----- |
|  |  |  |  |  | Nutritabs (MMN) |  |  |  |  |  |  |  |  |  |  |  |  | ----- | ----- | ----- | ----- |
|  |  |  |  |  | Passive control (no intervention) |  |  |  |  |  |  |  |  |  |  |  |  | control | control | control | ----- |
| Ghana | 2009-2014 | iLiNS-DYAD-G | RCT; longitudinal follow-up | Adu-Afarwuah, 2016 (41); Adu-Afarwuah, 2019 (16) | LNS: maternal SQ-LNS in pregnancy + 6 mo postpartum, child SQ-LNS 6-18 mo | ZPP: measured in all; CRP/AGP: The subsample for the CRP and AGP analyses was selected from the children whose mothers were not pregnant during the period when the temporary mislabeling occurred | Y | Y | ----- | ----- | Y | ----- | ----- | Y | ----- | Y[3] | Y[3] | LNS | ----- | ----- | ----- |
|  |  |  |  |  | MMN: maternal MMN in pregnancy + 6 mo postpartum, no child supplementation |  |  |  |  |  |  |  |  |  |  |  |  | control | ----- | ----- | ----- |
|  |  |  |  |  | IFA: maternal IFA in pregnancy and placebo for 6 mo postpartum, no child supplementation |  |  |  |  |  |  |  |  |  |  |  |  | control | ----- | ----- | ----- |
| Kenya | 2012-2016 | WASH-Benefits | cluster RCT; cross-sectional surveys | Null, 2018 (42); Stewart, 2019 (12) | Nutrition: SQ-LNS + IYCF counseling | Sub-study clusters were selected based on the logistical feasibility of the preservation and collection of biological specimens, as well as their transport to the central laboratory<br>Exclusion criteria included symptoms of moderate to severe dehydration on the day of the survey. | Dependent on age | Y | Y | Y | ----- | Y | ----- | ----- | Y | Y | Y | LNS | LNS | LNS | LNS |
|  |  |  |  |  | Water: family received chlorine for drinking water |  |  |  |  |  |  |  |  |  |  |  |  | control | control | ----- | control |
|  |  |  |  |  | Sanitation: family received upgraded latrine, sani-scoop and child potty |  |  |  |  |  |  |  |  |  |  |  |  | control | control | ----- | control |
|  |  |  |  |  | Handwashing: family received handwashing stations with soap |  |  |  |  |  |  |  |  |  |  |  |  | control | control | ----- | control |
| | | | | | WASH: family received all water, sanitation and hygiene interventions | | | | | | | | | | | | | control | control | control-WSH | control |
| | | | | | WASH + nutrition: all water, sanitation, hygiene and nutrition interventions | | | | | | | | | | | | | LNS | LNS | LNS-WSH | LNS |
|  |  |  |  |  | Passive control (no intervention) |  |  |  |  |  |  |  |  |  |  |  |  | control | control | control | ----- |
| | | | | | Active control (visits to measure MUAC) | | | | | | | | | | | | | control | control | control | control |
| Madagascar | 2014-2016 | MAHAY | cluster RCT; longitudinal follow-up | Galasso, 2019 (43); Stewart, 2020 (13) | T4: early child stimulation + IYCF counseling | Subset of sites randomly samples among sites accessible via paved road; random sample of children from each site in the youngest age cohort (18-24 mo at endline) | Dependent on age | Y | Y | Y | ----- | ----- | ----- | ----- | Y | ----- | ----- | ----- | ----- | ----- | ----- |
|  |  |  |  |  | T3: maternal SQ-LNS in pregnancy + 6 mo postpartum, child SQ-LNS 6-18 mo + IYCF counseling |  |  |  |  |  |  |  |  |  |  |  |  | LNS | ----- | ----- | ----- |
| | | | | | T2: child SQ-LNS 6-18 mo + IYCF counseling | | | | | | | | | | | | | LNS | LNS | LNS | LNS |
| | | | | | T1: IYCF counseling | | | | | | | | | | | | | control | control | control | control |
| | | | | | T0: Control (standard of care) | | | | | | | | | | | | | control | control | control | control |
| Malawi | 2011-2014 | iLiNS-DYAD-M | RCT; longitudinal follow-up | Ashorn, 2015 (44) | LNS: maternal SQ-LNS in pregnancy + 6 mo postpartum, child SQ-LNS 6-18 mo | N/A | Y | Y | ----- | ----- | Y | ----- | ----- | Y | ----- | Y[3] | Y[3] | LNS | ----- | ----- | ----- |
|  |  |  |  |  | MMN: maternal MMN in pregnancy + 6 mo postpartum, no child supplementation |  |  |  |  |  |  |  |  |  |  |  |  | control | ----- | ----- | ----- |
|  |  |  |  |  | IFA: maternal IFA in pregnancy and placebo for 6 mo postpartum, no child supplementation |  |  |  |  |  |  |  |  |  |  |  |  | control | ----- | ----- | ----- |
| Malawi | 2009-2012 | iLiNS-DOSE | RCT; longitudinal follow-up | Maleta, 2015 (45)[4] | SQ-LNS containing milk (10 g/d) | Random sub-sample of 100 children/group was selected | Y | Y | ----- | ----- | Y | ----- | ----- | ----- | ----- | ----- | ----- | LNS | LNS | LNS | LNS |
| | | | | | SQ-LNS containing milk (20 g/d) | | | | | | | | | | | | | LNS | LNS | LNS | LNS |
| | | | | | SQ-LNS without milk (20 g/d) | | | | | | | | | | | | | LNS | LNS | LNS | LNS |
|  |  |  |  |  | MQ-LNS containing milk (40 g/d) |  |  |  |  |  |  |  |  |  |  |  |  | ----- | ----- | ----- | ----- |
|  |  |  |  |  | MQ-LNS without milk (40 g/d) |  |  |  |  |  |  |  |  |  |  |  |  | ----- | ----- | ----- | ----- |
| | | | | | Active control | | | | | | | | | | | | | control | control | control | control |
| Mali | 2015-2017 | PROMIS | cluster RCT; cross-sectional surveys | Huybregts, 2019 (46) | SQ-LNS + IYCF counseling | N/A (all children in cross-sectional survey) | Y | Y | ----- | ----- | ----- | ----- | ----- | ----- | ----- | ----- | ----- | LNS | LNS | LNS | LNS |

| | | | | | | | | | | | | | | | | | | | | | |
|---|---|---|---|---|---|---|---|---|---|---|---|---|---|---|---|---|---|---|---|---|---|
| | | | | | Active control (standard of care) + IYCF counseling | | | | | | | | | | | | | control | control | control | control |
| Zimbabwe | 2013-2017 | SHINE | cluster RCT; longitudinal follow-up | Humphrey, 2019 (47); Prendergast, 2019 (48)[5] | IYCF: child SQ-LNS + IYCF counseling | Sub-study included infants of women reaching 32 weeks' gestation visit from 1 May 2014 through the end of the trial; all HIV exposed infants included | Dependent on age | Y | Y[3] | Y[3] | ----- | Y[3] | ----- | ----- | ----- | ----- | ----- | LNS | LNS | LNS | LNS |
| | | | | | WASH : family received ventilated improved pit latrine, handwashing stations, soap, chlorine, child play space | | | | | | | | | | | | | control | control | control-WSH | control |
| | | | | | WASH and IYCF: child SQ-LNS + IYCF counseling, family received ventilated improved pit latrine, handwashing stations, soap, chlorine, child play space | | | | | | | | | | | | | LNS | LNS | LNS-WSH | LNS |
| | | | | | Active control (standard of care) | | | | | | | | | | | | | control | control | control | control |

IFA, iron-folic acid; IYCF, infant and young child feeding; LNS, lipid-based nutrient supplement; MMN, multiple micronutrients; MNP, multiple micronutrient powder; MQ-LNS, medium-quantity lipid-based nutrient supplements; ORS, oral rehydration solution; SQ-LNS, small-quantity lipid-based nutrient supplements; WASH, water sanitation and hygiene; WSB, wheat soy blend; RCT, randomized controlled trial.

[1] All supplements were isocaloric, children 6-12 mo received 125 kcal/d, children 12-18 mo received 250 kcal/d
[2] All children in the four intervention groups received ORS for diarrhea and treatment for malaria
[3] Data not yet available
[4] Trial is cited as Kumwenda 2014 in Das et al. (Cochrane Database of Systematic Reviews 2019)
[5] Trial was designed *a priori* to present results separately for HIV exposed and un-exposed children; thus considered as two comparisons in all analyses and the presentation of results

**Supplemental Table 1. Amount of LNS provided (g/day) and nutrient value (per daily ration)**

| | Nutributter[1] | iLiNS formulation[2,3] | Revised iLiNS formulation[4] | Plumpy'Doz (6-11 mo)[5] | Plumpy'Doz (12-18 mo)[5] | Rice-lentil LNS (6-11 mo)[5] | Rice-lentil LNS (12-18 mo)[5] | Chickpea LNS (6-11 mo)[5] | Chickpea LNS (12-18 mo)[5] |
|---|---|---|---|---|---|---|---|---|---|
| Ration (g/day) | 20 | 20 | 20 | 23.2 | 46.4 | 25.7 | 51.4 | 23.6 | 47.2 |
| Total energy (kcal) | 108 | 118 | 118 | 123.4 | 246.8 | 133.9 | 267.8 | 128.6 | 257.2 |
| Protein (g) | 2.56 | 2.6 | 2.6 | 2.9 | 5.9 | 2.8 | 5.7 | 3.5 | 7.1 |
| Fat (g) | 7.08 | 9.6 | 9.6 | 7.9 | 15.8 | 6.9 | 13.9 | 6.6 | 13.2 |
| Linoleic acid (g) | 1.29 | 4.46 | 4.46 | | | | | | |
| α-Linolenic acid (g) | 0.29 | 0.58 | 0.58 | | | | | | |
| Vitamin A (µg RE) | 400 | 400 | 400 | 200 | 400 | 117.7 | 235.4 | 118.5 | 236.9 |
| Vitamin C (mg) | 30 | 30 | 30 | | | | | | |
| Vitamin B1 (mg) | 0.3 | 0.3 | 0.5 | 0.3 | 0.5 | 0.3 | 0.6 | 0.3 | 0.6 |
| Vitamin B2 (mg) | 0.4 | s | 0.5 | 0.3 | 0.5 | 0.2 | 0.5 | 0.2 | 0.5 |
| Niacin (mg) | 4 | 4 | 6 | 2.8 | 5.6 | 2.6 | 5.1 | 2.4 | 4.7 |
| Folic acid (µg) | 80 | 80 | 150 | 80 | 160.1 | 100 | 199.9 | 118.2 | 236.5 |
| Pantothenic acid (mg) | 1.8 | 1.8 | 2 | 1 | 2 | 1.1 | 2.2 | 1 | 2 |
| Vitamin B6 (mg) | 0.3 | 0.3 | 0.5 | 0.3 | 0.5 | 0.3 | 0.6 | 0.3 | 0.6 |
| Vitamin B12 (µg) | 0.5 | 0.5 | 0.9 | 0.4 | 0.8 | 0.5 | 0.9 | 0.5 | 0.9 |
| Vitamin D (IU) 1 IU = 0.025 ug | 0 | 200 | 200 | | | 226.2 | 452.3 | 226.6 | 453.1 |
| Vitamin E (mg) | 0 | 6 | 6 | 3 | 6 | 4.9 | 9.8 | 4.7 | 9.4 |
| Vitamin K (µg) | 0 | 30 | 30 | | | 11.3 | 22.6 | 11.3 | 22.7 |
| Iron (mg) | 9 | 6 | 9 | 4.5 | 9 | 3.3 | 6.7 | 3.5 | 7.1 |
| Zinc (mg) | 4 | 8 | 8 | 2 | 4 | 2.4 | 4.8 | 2.5 | 5.1 |
| Copper (mg) | 0.2 | 0.34 | 0.34 | 0.2 | 0.3 | 0.2 | 0.4 | 0.3 | 0.5 |
| Calcium (mg) | 100 | 280 | 280 | 193.5 | 387 | 208.2 | 416.3 | 219.5 | 439 |
| Phosphorus (mg) | 82 | 190 | 190 | 137.6 | 275.2 | 61.7 | 123.4 | 72.9 | 145.8 |
| Potassium (mg) | 152 | 200 | 200 | 155 | 310 | 206.6 | 413.3 | 220.7 | 441.3 |
| Magnesium (mg) | 16 | 40 | 40 | 29.9 | 59.9 | 41.6 | 83.3 | 47.7 | 95.3 |
| Selenium (µg) | 10 | 20 | 20 | 8.6 | 17.2 | 7.5 | 14.9 | 7.6 | 15.1 |
| Iodine (µg) | 90 | 90 | 90 | 27.6 | 55.2 | 33.7 | 67.3 | 33.7 | 67.5 |
| Manganese (mg) | 0.08 | 1.2 | 1.2 | 0.1 | 0.1 | 0.5 | 0.9 | 0.4 | 0.9 |

[1]Provided by Adu-Afarwuah 2007 (40)

[2]Provided by Hess 2015 (38), Becquey 2019 (39), Adu-Afarwuah 2016 (41), Galasso 2019 (43), Ashorn 2015 (44), Maleta 2015 (45), Huybregts 2019 (46), Humphrey 2019 (47), Prendergast 2019 (48).

[3]Hess 2015 (38) provided 0-10 mg zinc/d in the SQ-LNS product, plus a 5 mg/d zinc supplement in one intervention arm. Maleta 2015 (45) provided 10-40 g/d of LNS, with and without milk powder, varying by intervention arm (the 40 g arms were not included in this IPD analysis); the micronutrient composition of the LNS was identical across intervention arms, but there were differences in total kcal, protein, fat, linoleic acid and α-linolenic acid.

[4]Provided by Dewey 2017 (36), Luby 2018 (37), Null 2018 (42)

[5]Provided by Christian 2015 (35). Product and quantities differed by intervention arm and age.

**Supplemental Table 3: Descriptive information on potential study-level effect modifiers, by trial**

| Country | Author | Geographic Region | Malaria Prevalence (%) | Anemia Prevalence (%) | Inflammation (CRP > 5 mg/L) | Inflammation AGP > 1 g/L) | Water quality (% improved) | Sanitation (% improved) | Duration of supplementation | Iron dose (mg/d) | Frequency of contact | Average SQ-LNS compliance (%) | Compliance definition (in the SQ-LNS group) | Source (Anemia prevlance) |
|---|---|---|---|---|---|---|---|---|---|---|---|---|---|---|
| Bangladesh | Christian, 2015 (35); Campbell, 2020 (15) | SEAR | 0.2[a] | 51.3%[a] | 21%[b] | 59%[b] | 100.0[a] | 77.0[a] | ≤ 12 mo | 3.3-9 mg/d[1] | Weekly | 93.0[a] | % of total intended SQ-LNS consumed (quantity * day) | National Institute of Population Research and Training (NIPORT), Mitra and Associates, and ICF International. 2013. Bangladesh Demographic and Health Survey 2011. Dhaka, Bangladesh and Calverton, Maryland, USA: NIPORT, Mitra and |
| Bangladesh | Dewey, 2017 (36); Matias, 2018 (10) | SEAR | 0.2[a] | 51.3%[a] | 14%[a] | 32%[a] | 100.0[a] | 71.1[a] | > 12 mo | 9 mg/d | Monthly | 97.4[a] | % reporting "high adherence (≥ 4 days/week) | National Institute of Population Research and Training (NIPORT), Mitra and Associates, and ICF International. 2013. Bangladesh Demographic and Health Survey 2011. Dhaka, Bangladesh and Calverton, Maryland, USA: NIPORT, Mitra and |
| Bangladesh | Luby, 2018 (37); Stewart, 2019 (12) | SEAR | 0.1[a] | 51.3%[a] | 11%[a] | 29%[a] | 88.5[a] | 94.7[a] | > 12 mo | 9 mg/d | Weekly | 93.0[a] | Number of sachets consumed in 14 days prior to annual survey/14 | National Institute of Population Research and Training (NIPORT), Mitra and Associates, and ICF International. 2013. Bangladesh Demographic and Health Survey 2011. Dhaka, Bangladesh and Calverton, Maryland, USA: NIPORT, Mitra and |
| Burkina Faso | Hess, 2015 (38); Abbeddou 2017 (11) | AFR | 59.1[b] | 87.8%[b] | 32%[b] | 59%[b] | 26.7[b] | 2.3[b] | ≤ 12 mo | 6 mg/d | Weekly | 96.8[a] | % of days SQ-LNS reported consumed | Institut National de la Statistique et de la Démographie (INSD) et ICF International, 2012. Enquête Démographique et de Santé et à Indicateurs Multiples du Burkina Faso 2010. Calverton, Maryland, USA : INSD et ICF International. |
| Burkina Faso | Becquey, 2019 (39)[2] | AFR | 42.9[b] | 87.8%[b] | --- | --- | 50.6[b] | 53.4[a] | ≤ 12 mo | 6 mg/d | Monthly | 37.0[b] | caregiver reported receiving SQ-LNS in the previous month | Institut National de la Statistique et de la Démographie (INSD) et ICF International, 2012. Enquête Démographique et de Santé et à Indicateurs Multiples du Burkina Faso 2010. Calverton, Maryland, USA : INSD et ICF International. |
| Ghana | Adu-Afarwuah, 2007 (40); Adu-Afarwuah, 2008 (9) | AFR | 37.4[b] | 76.1%[b] | 18%[a] | --- | 91.9[a] | 91.5[a] | ≤ 12 mo | 9 mg/d | Weekly | 88.2[a] | % of days SQ-LNS reported consumed | Ghana Statistical Service (GSS), Noguchi Memorial Institute for Medical Research (NMIMR), and ORC Macro. 2004. Ghana Demographic and Health Survey 2003. Calverton, Maryland: GSS, NMIMR, and ORC Macro. |
| Ghana | Adu-Afarwuah, 2016 (41); Adu-Afarwuah, 2019 (16) | AFR | 36.8[b] | 57.0%[a] | 18%[a] | 44%[a] | 98.4[a] | 97.3[a] | ≤ 12 mo | 6 mg/d | Weekly | 73.5[b] | % of days SQ-LNS reported consumed | Ghana Statistical Service, 2011. Ghana Multiple Indicator Cluster Survey with an Enhanced Malaria Module and Biomarker, 2011, Final Report. Accra, Ghana. |
| Kenya | Null, 2018 (42); Stewart, 2019 (12) | AFR | 8.5[a] | 36.3%[a] | 21%[a] | 47%[a] | 68.0[b] | 15.8[b] | > 12 mo | 9 mg/d | Monthly | 115.0[a] | Number of sachets consumed in 14 days prior to annual survey/14 | National Malaria Control Programme (NMCP), Kenya National Bureau of Statistics (KNBS), and ICF International. 2016. Kenya Malaria Indicator Survey 2015. Nairobi, Kenya, and Rockville, Maryland, USA: NMCP, KNBS, and ICF International. |
| Madagascar | Galasso, 2019 (43); Stewart, 2020 (13) | AFR | 5.5[a] | 51.0%[a] | 19%[a] | 41%[a] | 26.9[b] | 0.0[b] | ≤ 12 mo | 6 mg/d | Monthly | - | Data unavailable. | Institut National de la Statistique (INSTAT), Programme National de lutte contre le Paludisme (PNLP), Institut Pasteur de Madagascar (IPM) et ICF International. 2013. Enquête sur les Indicateurs du Paludisme (EIPM) 2013. Calverton, MD, USA : |
| Malawi | Ashorn, 2015 (44) | AFR | 26.8[b] | 62.5%[b] | 30%[b] | 68%[b] | 91.7[a] | 9.2[b] | ≤ 12 mo | 6 mg/d | Weekly | 77.1[b] | % of days SQ-LNS reported consumed | National Statistical Office (NSO) and ICF Macro. 2011. Malawi Demographic and Health Survey 2010. Zomba, Malawi, and Calverton, Maryland, USA: NSO and ICF Macro. |
| Malawi | Maleta, 2015 (45) | AFR | 30.3[b] | 62.5%[b] | 34%[b] | 66%[b] | 91.6[a] | 2.9[b] | ≤ 12 mo | 6 mg/d | Weekly | 71.6[b] | % of days SQ-LNS reported consumed (considering missed delivery visits) | National Statistical Office (NSO) and ICF Macro. 2011. Malawi Demographic and Health Survey 2010. Zomba, Malawi, and Calverton, Maryland, USA: NSO and ICF Macro. |
| Mali | Huybregts, 2019 (46)[2] | AFR | 39.1[b] | 81.8%[b] | --- | --- | 50.8[b] | 75.3[a] | > 12 mo | 6 mg/d | Monthly | 47.0[b] | caregiver reported receiving SQ-LNS in the previous month | Institut National de la Statistique (INSTAT), Cellule de Planification et de Statistique Secteur Santé-Développement Social et Promotion de la Famille (CPS/SS-DS-PF), et ICF. 2019. Sixième Enquête Démographique et de Santé au Mali 2018. |
| Zimbabwe | Humphrey, 2019 (47); Prendergast 2019 (48)[3] | AFR | 9.0[a] | 36.8%[a] | 21%[a] | --- | 63.6[b] | 34.0[b] | < 12 mo | 6 mg/d | Monthly | 73.5[b] | received > 11 (80% of expected) deliveries * consumed SQ-LNS in past 24 h (at 12 month visit) | Zimbabwe National Statistics Agency and ICF International. 2016. Zimbabwe Demographic and Health Survey 2015: Final Report. Rockville, Maryland, USA: Zimbabwe National Statistics Agency (ZIMSTAT) and ICF International. |

AFR, Africa Region; AGP, alpha-1-acid glycoprotein; CRP, C-reactive protein; SEAR, South-East Asia Region; SQ-LNS, small quantity lipid-based nutrient supplements

[1]Iron dose varied by age and SQ-LNS intervention group (see Supplemental Table 2)

[2]Study-level effect modifier categorization based on the longitudinal cohort, for consistency with the growth and development domains of this individual participant data analysis

[3]Study-level effect modifier categorization was the same for both HIV exposed and HIV un-exposed children

Superscripts a and b designate the two categories, as described in Notes below.

**Notes:**

Geograhic region: based on WHO regions

Malaria prevalence: Data extracted from Annex, Data table F: Population at risk and reestimated malaria cases and deaths, 2010-2017 (wmr2018-annex-table-f.xls); Point estimate (presumed and confirmed malaria cases), divided by population at risk, per 100 persons. Trials were categorized as (a) low burden when malaria was < 10% and (b) high burden when malaria was ≥10%.

Prevalence of anemia: Data extracted from DHS/MICS, survey closest in time to the start of child supplementation; prevaelence of Hb < 110 g/L among children 6-59 months (sources cited in table). Trials were categorized as (a) moderate burden when anemia was < 60% and (b) high burden when anemia was >60%.

Inflammation was defined as CRP > 5 mg/L and AGP > 1 g/L. Prevalence of inflammation was based on study-specific data at outcome assessment. Inflammation was defined as (a) low burden when high CRP prevalence < 25% and high AGP < 50%, and (b) high burden was defined as high CRP prevalence > 25% or high AGP > 50%.

Water quality: Data based on sample prevalences of improved source water quality. Study-level water quality was considered as (a) improved if the main source of drinking water for ≥ 75% of participants was improved; study-level water quality was considered as (b) unimproved if the main source of drinking water for < 75% of participants was improved. "Improved water sources are those that have the potential to deliver safe water by nature of their design and construction, and include: piped water, boreholes or tubewells, protected dug wells, protected springs, rainwater and packaged or delivered water". Unimproved water sources include: water from an unprotected dug well or unprotected spring, or surface water (e.g., river, dam, lake, pond, stream, canal or irrigation canal). (washdata.org/monitoring/drinking-water).

Sanitation: Data based on sample prevalences of improved sanitation. Study-level sanitation was considered as (a) improved if sanitation services for ≥ 50% of participants were improved; study-level sanitation was considered as (b) unimproved if sanitation services for < 50% of participants were improved. "Improved sanitation facilities are those designed to hygienically separate excreta from human contact, and include: flush/pour flush to piped sewer system, septic tanks or pit latrines; ventilated improved pit latrines, composting toilets or pit latrines with slabs". Unimproved sanitation services include: use of pit latrines without a slab or platform, hanging latrines or bucket latrines, or open defecation. (https://washdata.org/monitoring/sanitation)

Compliance: Data extracted from publication; study-specific definitions of compliance are noted in the table. Trials were categorized as (a) high compliance when complinace was ≥ 80% or (b) low compliance when compliance was < 80% compliance.

**Supplemental Table 4: Descriptive information on potential individual-level effect modifiers, by trial**

| Country | Author | Maternal BMI < 20 kg/m$^2$ (%) | Maternal age < 25 y (%) | Maternal education, completed primary (%) | Child sex, male (%) | Child birth order, first born (%) | Prevalence of baseline anemia (%) | Prevalence of baseline acute malnutrition (%) | Inflammation at endline (%) | Receipt of high-dose vitamin A (%) | Baseline food security, moderate/severe insecurity (%) | SES index, below median (%) | Improved source water quality (%) | Improved sanitation access (%) | Season at endline, dry (%) |
|---|---|---|---|---|---|---|---|---|---|---|---|---|---|---|---|
| Bangladesh | Christian, 2015 (35); Campbell, 2020 (15) | | 54.2 | 65.2 | 50.8 | 75.4* | | 20.3 | 61.3 | 50.6 | 24.9[a] | 46.3 | 100.0 | 79.3 | 55.6 |
| Bangladesh | Dewey, 2017 (36); Matias, 2018 (10) | 58.2 | 71.7 | 73.1 | 48.5 | 37.8 | 60.8 | 6.0 | 34.5 | 81.2 | 38.2[b] | 50.2 | 100.0 | 69.3 | 61.8 |
| Bangladesh | Luby, 2018 (37); Stewart, 2019 (12) | 51.4 | 58.9 | 73.3 | 48.8 | 37.5 | | | 22.8 | 60.3 | 20.2[b] | 50.7 | 78.8 | 92.5 | 48.6 |
| Burkina Faso | Hess, 2015 (38); Abbeddou 2017 (11) | 38.8 | 41.3 | 3.7 | 50.6 | 22.1 | 90.9 | 25.7 | 61.4 | | 48.9[b] | 41.4 | 26.7 | 2.3 | 71.5 |
| Burkina Faso | Becquey, 2019 (39) | 40.5 | 53.8 | 8.1 | 53.0 | 15.4 | | | | | | 49.6 | 63.8 | 41.1 | 45.7 |
| Ghana | Adu-Afarwuah, 2007 (40); Adu-Afarwuah, 2008 (9) | 9.2 | 29.0 | 88.3 | 52.1 | 40.0 | | | 18.1 | 59.3 | | 46.3 | 91.9 | 91.5 | 57.7 |
| Ghana | Adu-Afarwuah, 2016 (41); Adu-Afarwuah, 2019 (16) | 15.3 | 39.0 | 78.8 | 48.2 | 33.7 | 35.3 | 8.2 | 45.4 | 63.3 | 30.3[b] | 50.1 | 98.4 | 97.2 | 59.2 |
| Kenya | Null, 2018 (42); Stewart, 2019 (12) | 21.6 | 40.9 | 46.3 | 47.8 | 18.9 | | | 49.2 | 11.6 | 11.7[b] | 43.9 | 70.1 | 13.4 | 27.3 |
| Madagascar | Galasso, 2019 (43); Stewart, 2020 (13) | | 47.0 | 22.8 | 47.6 | 23.0 | | | 43.3 | | 28.4[b] | 49.0 | 26.4 | 0.0 | 88.7 |
| Malawi | Ashorn, 2015 (44) | 40.0 | 50.2 | 15.7 | 47.4 | 20.8 | 65.7 | 8.4 | 68.9 | 33.2 | 71.6[b] | 46.7 | 91.5 | 9.4 | 70.8 |
| Malawi | Maleta, 2015 (45) | 25.8 | 45.2 | 24.4 | 50.8 | 23.7 | 62.7 | 5.7 | 67.9 | | 73.9[b] | 49.2 | 91.8 | 2.9 | 59.4 |
| Mali | Huybregts, 2019 (46) | 27.5 | 47.5 | 10.6 | 52.3 | 14.4 | | | | | | 49.7 | 59.7 | 74.9 | 100.0 |
| Zimbabwe | Humphrey, 2019 (HIV-unexposed) (47) | 15.4 | 47.6 | 96.4 | 50.2 | 26.7 | | 5.6 | 21.0 | 57.0 | 18.7[c] | 49.3 | 62.6 | 33.3 | 92.6 |
| Zimbabwe | Prendergast, 2019 (HIV-exposed) (48) | 18.1 | 22.9 | 94.3 | 49.9 | 19.7 | | 8.4 | 15.5 | 54.3 | 25.1[c] | 49.3 | 61.8 | 30.6 | 93.7 |

**Abbreviations**: BMI, body mass index; SES, socio-economic status

**Notes:**

Anemia: Hemoglobin < 110 g/L

Baseline hemoglobin and anthropometric status (acute malnutrition) were measured at enrollment into the study or start of supplementation if supplementation did not begin at enrollment. These data are not available for studies with a repeated cross-sectional survey design, as the same individual children were not included in both baseline and endline surveys.

Acute malnutrition: weight-for-length Z-score < -2 SD or mid-upper arm circumfernece < 125 mm

High-dose vitamin A: received in 6 months prior to outcome assessment (yes/no)

Food security scales used: a) Food Access Survey Tool; b) Household Food Insecurity Access Scale (HFAIS); c) Coping Strategy Index

Water and sanitation references - as for study-level effect modifiers

Season is defined at time of outcome assessment as a dichotomous "Rainy" vs "Dry" category based on child-specific average rainfall during the month of measurement and 2 months prior. In Mali (Huybregts 2019), there was little variation in rainfall during the endline cross-sectional survey, as it was conducted in the dry season (February-March).

*Data on birth order were not available for all children. Consequently, first-born vs later-born status was estimated based on the number of children under 5 years old in the household.

**Supplemental Table 5:** Description of biochemical assessments

| Country | Author | Hemoglobin (Hb) | | | | | Micronutrient status biomarkers | | | | | Inflammation biomarkers | |
|---|---|---|---|---|---|---|---|---|---|---|---|---|---|
| | | Hb sample | Hb method | MN sample | Ferritin | sTfR | ZPP | Zinc | Retinol | RBP | CRP | AGP | Malaria |
| Bangladesh | Christian, 2015 (35); Campbell, 2020 (15) | Venous | Hemocue Hb 301 | Venous, serum | Automated chemiluminescent immunoassay (Immulite 2000) | ----- | ----- | Graphite furnace AAS | Reverse-phase HPLC | ----- | Automated chemiluminescent asasy (Immulite 2000) | Radial immunodiffusion (Kent Laboratories) | ----- |
| Bangladesh | Dewey, 2017 (36); Matias, 2018 (10) | Capillary | Hemocue Hb 301 | Capillary, serum | Combined sandwich ELISA | Combined sandwich ELISA | ----- | ----- | ----- | Combined sandwich ELISA | Combined sandwich ELISA | Combined sandwich ELISA | ----- |
| Bangladesh | Luby, 2018 (37); Stewart, 2019 (12) | Venous | Hemocue Hb 301 | Venous, serum | Combined sandwich ELISA | Combined sandwich ELISA | ----- | ----- | ----- | Combined sandwich ELISA | Combined sandwich ELISA | Combined sandwich ELISA | Y |
| Burkina Faso | Hess, 2015 (38); Abbeddou 2017 (11) | Capillary | Hemocue Hb 201+ | Venous, plasma | Combined sandwich ELISA | Combined sandwich ELISA | Hematoflurometer (Aviv Biomedical); washed RBC | ICP-AES | ----- | Combined sandwich ELISA | Combined sandwich ELISA | Combined sandwich ELISA | RDT (Bioline Malaria Ag P.f/Pan) |
| Burkina Faso | Becquey, 2019 (39) | Capillary | Hemocue Hb 201+ | ----- | ----- | ----- | ----- | ----- | ----- | ----- | ----- | ----- | ----- |
| Ghana | Adu-Afarwuah, 2007 (40); Adu-Afarwuah, 2008 (9) | Venous | Hemocue B-Hemoglobin | Venous, plasma | Coat-A-Count Immunoradiometric Assay Kit (Diagnostic Products) | ELISA (Ramco Laboratories) | ----- | ICP-AES | Reverse-phase HPLC | ----- | Radial immunodiffusion (Binding Site) | ----- | Blood smear (Giemsa stain) |
| Ghana | Adu-Afarwuah, 2016 (41); Adu-Afarwuah, 2019 (16) | Venous | Hemocue Hb 301 | Venous, plasma | ----- | ----- | Hematoflurometer (Aviv Biomedical); washed RBC | ----- | Reverse-phase HPLC | ----- | Cobas Integra 400 plus Automatic Analyzer (Roche); particle-enhanced turbidimetric assay | Cobas Integra 400 plus Automatic Analyzer (Roche); particle-enhanced turbidimetric assay | RDT (Clearview Malarial Combo, Vision Biotech; HRP2 and P.aldolase) |
| Kenya | Null, 2018 (42); Stewart, 2019 (12) | Venous | Hemocue Hb 301 | Venous, serum | Combined sandwich ELISA | Combined sandwich ELISA | ----- | ----- | ----- | Combined sandwich ELISA | Combined sandwich ELISA | Combined sandwich ELISA | RDT (Alere; SD Bioline Malaria Ag P.f/P.v) |
| Madagascar | Galasso, 2019 (43); Stewart, 2020 (13) | Capillary | Hemocue Hb 301 | Capillary, serum | Combined sandwich ELISA | Combined sandwich ELISA | ----- | ----- | ----- | Combined sandwich ELISA | Combined sandwich ELISA | Combined sandwich ELISA | ----- |
| Malawi | Ashorn, 2015 (44) | Capillary | Hemocue Hb 201+ | Venous, plasma | ----- | ----- | Hematoflurometer (Aviv Biomedical); washed RBC | ----- | Reverse-phase HPLC | ----- | Cobas Integra 400 plus Automatic Analyzer (Roche); particle-enhanced turbidimetric assay | Cobas Integra 400 plus Automatic Analyzer (Roche); particle-enhanced turbidimetric assay | RDT |
| Malawi | Maleta, 2015 (45) | Capillary | Hemocue Hb 201+ | ----- |  | ----- | Hematoflurometer (Aviv Biomedical); washed RBC | ----- | ----- | ----- | Cobas Integra 400 plus Automatic Analyzer (Roche); particle-enhanced turbidimetric assay | Cobas Integra 400 plus Automatic Analyzer (Roche); particle-enhanced turbidimetric assay | RDT |
| Mali | Huybregts, 2019 (46) | Capillary | Hemocue Hb 201+ | ----- | ----- | ----- | ----- | ----- | ----- | ----- | ----- | ----- | ----- |
| Zimbabwe | Humphrey, 2019 (47); Prendergast 2019 (48) | Venous | Hemocue Hb 301[1] | Venous, plasma | ----- | ----- | ----- | ----- | ----- | ----- | ----- | ----- | ----- |

AAS, atomic absorption spectroscopy; AGP, alpha-1-acid glycoprotein; CRP, C-reactive protein; Hb, hemoglobin; HPLC, high performance liquid chromatography; ICP-AES, inductively coupled plasma atomic emission spectroscopy; MN, micronutrient; RBC, red blood cells; RBP, retinol binding protein; RDT, rapid diagnostic test; sTfR, soluble transferrin receptor; ZPP, zinc protoporphyrin

[1]Hemoglobin data were altitude adjusted (27)

**Supplemental Table 6: Biomarker outcomes at endline among control groups, by trial**

| Country | Author | Hemoglobin, g/L | Anemia, Hb < 110 g/L | Moderate to severe anemia, Hb < 100 g/L | Ferritin, µg/L | Iron deficiency, ferritin < 12 µg/L | Iron deficiency anemia | sTfR, mg/L | sTfR > 8.3 mg/L | ZPP, µmol/mol heme | ZPP > 70 µmol/mol heme | Plasma zinc, µg/dL | Zinc < 65 µg/dL | Retinol, µmol/L | Retinol < 0.70 µmol | Retinol < 1.05 µmol | RBP, µmol | RBP < 0.70 µmol | RBP < 1.05 µmol |
|---|---|---|---|---|---|---|---|---|---|---|---|---|---|---|---|---|---|---|---|
| Bangladesh | Christian, 2015 (35); Campbell, 2020 (15) | 118 ± 9 | 15.8 | 1.4 | 23.4 (13.2, 37.2) | 22.2 | 9.0 | | | | | 88.3 (80.2, 99.8) | 2.1 | 1.35 (1.10, 1.70) | 2.9 | 19.4 | | | |
| Bangladesh | Dewey, 2017 (36); Matias, 2018 (10) | 112 ± 14 | 41.9 | 16.5 | 21.1 (12.8, 32.0) | 22.1 | 14.7 | 8.3 (7.0, 11.3) | 50.0 | | | | | | | | 1.19 (1.01, 1.44) | 3.3 | 29.0 |
| Bangladesh | Luby, 2018 (37); Stewart, 2019 (12) | 118 ± 9 | 16.1 | 3.2 | 15.9 (8.9, 25.6) | 35.4 | 10.1 | 7.5 (6.2, 8.6) | 31.5 | | | | | | | | 1.14 (0.96, 1.35) | 1.1 | 35.4 |
| Burkina Faso | Hess, 2015 (38); Abbeddou 2017 (11) | 89 ± 16 | 91.1 | 75.0 | 9.7 (6.2, 18.6) | 58.5 | 54.5 | 9.9 (6.6, 12.6) | 61.3 | 122.2 (70.7, 181.1) | 75.2 | 68.7 (62.7, 77.4) | 36.2 | | | | 1.06 (0.89, 1.20) | 8.5 | 49.1 |
| Burkina Faso | Becquey, 2019 (39) | 103 ± 13 | 70.1 | 36.7 | | | | | | | | | | | | | | | |
| Ghana | Adu-Afarwuah, 2007 (40); Adu-Afarwuah, 2008 (9) | 106 ± 15 | 58.3 | 32.3 | 9.3 (3.2, 29.1) | 56.1 | 35.4 | 9.6 (7.4, 12.8) | 62.2 | | | 60.0 (53.9, 68.4) | 68.1 | 1.08 (0.80, 1.33) | 16.2 | 48.6 | | | |
| Ghana | Adu-Afarwuah, 2016 (41); Adu-Afarwuah, 2019 (16) | 112 ± 11 | 44.9 | 5.4 | | | | | | 48.1 (33.8, 68.5) | 24.3 | | | 1.01 (0.87, 1.20) | 5.6 | 56.6 | | | |
| Kenya | Null, 2018 (42); Stewart, 2019 (12) | 110 ± 13 | 47.3 | 21.3 | 10.1 (6.3, 16.7) | 59.1 | 33.3 | 10.9 (8.2, 15.5) | 73.0 | | | | | | | | 0.97 (0.80, 1.16) | 9.3 | 59.5 |
| Madagascar | Galasso, 2019 (43); Stewart, 2020 (13) | 104 ± 16 | 64.8 | 32.8 | 11.6 (9.3, 18.5) | 51.0 | 27.5 | 9.0 (7.2, 13.5) | 58.8 | | | | | | | | 1.09 (0.95, 1.28) | 2.0 | 39.2 |
| Malawi | Ashorn, 2015 (44) | 108 ± 15 | 51.9 | 23.6 | | | | | | 53.6 (36.1, 85.1) | 33.6 | | | 1.05 (0.87, 1.20) | 9.2 | 48.5 | | | |
| Malawi | Maleta, 2015 (45) | 101 ± 14 | 72.0 | 46.3 | | | | | | 66.4 (40.1, 110.4) | 44.8 | | | | | | | | |
| Mali | Huybregts, 2019 (46) | 96 ± 13 | 86.2 | 60.2 | | | | | | | | | | | | | | | |
| Zimbabwe | Humphrey, 2019 (HIV-unexposed) (47) | 114 ± 12 | 35.3 | 10.6 | NA | | | NA | | | | | | | | | | | |
| Zimbabwe | Prendergast, 2019 (HIV-exposed) (48) | 115 ± 13 | 36.8 | 10.2 | NA | | | NA | | | | | | | | | | | |

Hb, hemoglobin; NA, data not yet available; RBP, retinol binding protein; sTfR, soluble transferrin receptor; ZPP, zinc protoporphyrin
Values are mean ± SD, median (Q1, Q3) or prevalences
All continuous biomarkers, with the exception of hemoglobin, were natural log transformed prior to analysis
Ferritin, sTfR, ZPP, zinc, retinol and RBP concentrations were adjusted for inflammation (i.e., C-reactive protein (CRP) and/or α-1-acid glycoprotein (AGP) concentrations, as available), using a regression correction approach adapted from the Biomarkers Reflecting Inflammation and Nutritional Determinants of Anemia (BRINDA) project (28)
