## Supplemental Table 7 for "Characteristics that modify the effect of small-quantity lipid-based nutrient supplementation on child anemia and micronutrient status: an individual participant data meta-analysis of randomized controlled trials"

### Supplemental Table 7: Risk of bias assessment in each trial

| Country | Author | Random sequence generation | Allocation concealment | Blinding participants | Outcome assessment[1] | Incomplete outcome | Selective reporting | Other |
|---|---|---|---|---|---|---|---|---|
| Bangladesh | Christian, 2015 (35); Campbell, 2020 (15) | low | low | high | low | low | low | low |
| Bangladesh | Dewey, 2017 (36); Matias, 2018 (10) | low | low | high | low | low | low | low |
| Bangladesh | Luby, 2018 (37); Stewart, 2019 (12) | low | low | high | high | low | low | low |
| Burkina Faso | Hess, 2015 (38); Abbeddou 2017 (11) | low | low | high | low | low | low | low |
| Burkina Faso | Becquey, 2019 (39) | low | low | high | low | low | low | low |
| Ghana | Adu-Afarwuah, 2007 (40); Adu-Afarwuah, 2008 (9) | low | low | high | low | low | low | low |
| Ghana | Adu-Afarwuah, 2016 (41); Adu-Afarwuah, 2019 (16) | low | low | high | low | low | low | low |
| Kenya | Null, 2018 (42); Stewart, 2019 (12) | low | low | high | low | high | low | low |
| Madagascar | Galasso, 2019 (43); Stewart, 2020 (13) | low | low | high | low | low | low | low |
| Malawi | Ashorn, 2015 (44) | low | low | high | low | low | low | low |
| Malawi | Maleta, 2015 (45) | low | low | high | low | low | low | low |
| Mali | Huybregts, 2019 (46) | low | low | high | low | low | low | low |
| Zimbabwe | Humphrey, 2019 (47); Prendergast, 2019 (48) | low | low | high | low | low | low | low |

[1]Due to the nature of the intervention, blinding of participants was not possible. We considered biochemical outcome assessment to be at low risk of bias in general, owing to the automated nature of hemoglobin assessments and the likelihood that laboratory technicians who performed the biochemical assessments were not aware of group allocation

| Adu-Afarwuah, 2007 (40); Adu-Afarwuah, 2008 (9) | | |
|---|---|---|
| **Bias** | **Authors' judgement** | **Support for judgement** |
| Random sequence generation (selection bias) | Low risk | **Quote:** "we randomly selected ~75% of the total number of eligible infants to enter the intervention trial. This was done on a weekly basis, when infants were 5 mo of age, by entering the identification numbers of the eligible infants in a dataset, and using an SAS data step (ranuni [1] le 0.75) to select those for the intervention…the NI infants were randomly selected from the pool of initially eligible infants"<br>**Comment:** adequately done |
| Allocation concealment (selection bias) | Low risk | **Quote:** "the infants were randomly assigned (with the use of opaque envelopes with group designation) to receive SP, NT, or NB until 12 mo of age"<br>**Comment:** adequately done |
| Blinding of participants and personnel (performance bias) | High risk | **Quote**: "NT was provided to the mothers in plastic bags, and the NB (20 g/d) was provided in foil packs with screw caps"<br>**Comment:** not adequate |
| Blinding of outcome assessment (detection bias) | Low risk | **Personal communication with investigator (SAA):** "At both 6 and 12 months of age, all children were brought to the laboratory of the Koforidua Central Hospital, where laboratory technologist of the Hospital collected the venous blood and determined Hb and malaria RDT and later sTfR and CRP. Frozen plasma samples were sent on ice to the Radiotherapy Department of the Korle Bu Teaching Hospital in Accra, where technicians completed the ferritin analysis. So, as in this case of the anthropometric measurements, it was not possible for the laboratory analysts to know the group assignments".<br>**Comment:** adequately done |
| Incomplete outcome data (attrition bias) | Low risk | **Attrition**: SP group = 98/105; NT group = 102/105; NB group = 98/103; Control group = 96/97<br>**Comment:** Minimal attrition and reasons given for loss to follow-up; Hb data available for ~95-99% of enrolled children |
| Selective reporting (reporting bias) | Low risk | **Comment**: The trial was registered at clinicaltrials.gov (NCT00379158); outcomes described in the methods section reported in the results section |
| Other bias | Low risk | **Comment**: no other potential sources of bias reported |

**Funding**
Supported by the Nestlé Foundation with additional support from USAID's MGL Research Program through ILSI.

| **Adu-Afarwuah, 2016 (41); Adu-Afarwuah, 2019 (16)** | | |
|---|---|---|
| **Bias** | **Authors' judgement** | **Support for judgement** |
| Random sequence generation (selection bias) | Low risk | **Quote**: "The study statistician at University of California, Davis developed group allocations with the use of a computer generated (SAS version 9.3; SAS Institute) randomization scheme in blocks of 9"<br>**Comment:** adequately done; |
| Allocation concealment (selection bias) | Low risk | **Quote**: "At each enrollment, the study nurse offered sealed, opaque envelopes bearing group allocations, 9 envelopes at a time, and the woman picked one to reveal the allocation"<br>**Comment:** adequately done |
| Blinding of participants and personnel (performance bias) | High risk | **Quote:** "It was not possible to blind study workers and participants to the capsules (IFA and MMN supplements) compared with the LNS supplements because of their different appearances" |
| Blinding of outcome assessment (detection bias) | Low risk | **Quote**: "It was not possible to blind study workers and participants to the capsules (IFA and MMN supplements) compared with the LNS supplements because of their apparent differences, but laboratory staff, anthropometrists, and data analysts had no knowledge of group assignment until all preliminary analyses had been completed"<br>**Comment:** adequately done |
| Incomplete outcome data (attrition bias) | Low risk | **Attrition:** IFA group = 393/408; MMN group = 401/411; LNS group = 391/409<br>**Comment:** Minimal attrition and reasons given for loss to follow-up; Hb data available for ~83% of enrolled children; similar across study arms |
| Selective reporting (reporting bias) | Low risk | **Comment**: The trial was registered at clinicaltrials.gov (NCT00970866); SAP available online; outcomes described in the methods section reported in the results section |
| Other bias | Low risk | **Comment**: no other potential sources of bias reported |
| **Funding**<br>Funded by a grant to the University of California, Davis, from the Bill & Melinda Gates Foundation. | | |

| **Ashorn, 2015 (44)** | | |
|---|---|---|
| **Bias** | **Authors' judgement** | **Support for judgement** |
| Random sequence generation (selection bias) | Low risk | **Quote:** "Researcher not involved with the trial created individual randomisation slips (in blocks of 9)"<br>**Comment**: Additional details on randomization provided in Ashorn *et al.* AJCN 2015. |
| Allocation concealment (selection bias) | Low risk | **Quote**: "…packed them in sealed, numbered, opaque randomization envelopes that were stored in numerical order…Eligible pregnant women were requested to choose 1 of the top 6 envelopes in the stack, and the contents of the envelope indicated her participant number and group allocation""<br>**Comment:** adequately done |
| Blinding of participants and personnel (performance bias) | High risk | **Quote:** "The IFA and MMN interventions were provided with double-masked procedures…For the LNS group, we used single-masked procedures; that is field workers who delivered the supplements knew which mothers were receiving LNS, and the participants were advised not to disclose information about their supplements to anyone other than an iLiNS team member"<br>**Comment:** not done |
| Blinding of outcome assessment (detection bias) | Low risk | **Quote:** "The data collectors who performed the anthropometric measurements or assessed other outcomes were not aware of group allocation. Researchers responsible for the data cleaning remained blind to the trial code, until the database was considered fully cleaned"<br>**Comment**: adequately done |
| Incomplete outcome data (attrition bias) | Low risk | **Attrition:** IFA group = 220/223; MMN group = 222/233; LNS group = 214/222<br>**Comment:** Minimal attrition and reasons given for loss to follow-up; Hb data available for ~95% of enrolled children |
| Selective reporting (reporting bias) | Low risk | **Comment**: The trial was registered at clinicaltrials.gov ( NCT01239693 ); SAP available online; outcomes described in the methods section reported in the results section |
| Other bias | Low risk | **Comment:** no other potential sources of bias reported |
| **Funding**<br>Supported in part by a grant to the University of California, Davis, from the Bill & Melinda Gates Foundation, with additional funding from the Office of Health, Infectious Diseases, and Nutrition, Bureau for Global Health, US Agency for International Development (USAID) under terms of cooperative agreement AID-OAAA-1200005, through the Food and Nutrition Technical Assistance III Project (FANTA), managed by FHI 360. For data management and statistical analysis, the team received additional support from the Academy of Finland grant 252075 and the Medical Research Fund of Tampere University Hospital grant 9M004. YBC was supported by the Singapore Ministry of Health's National Medical Council under its Clinician Scientist Award. | | |

| **Becquey, 2019 (39)** | | |
|---|---|---|
| **Bias** | **Authors' judgement** | **Support for judgement** |
| Random sequence generation (selection bias) | Low risk | **Quote:** "simple(i.e. non-stratified) random allocation was used"; "randomization took place at a community event…with local health authorities"; "32 identical pieces of paper with either 'control' or 'intervention' written on them were mixed in a bag for randomization"<br>"In each health center catchment area, a census was conducted 1 month prior to the cross-sectional baseline and endline surveys to identify all pregnant women and eligible children. A random sample of children was drawn from the census list."<br>**Comment**: adequately done |
| Allocation concealment (selection bias) | Low risk | **Quote:** "32 identical pieces of paper with either 'control' or 'intervention' written on them were mixed in a bag for randomization"<br>**Comment**: adequately done |
| Blinding of participants and personnel (performance bias) | High risk | **Quote:** "non-masked, community-based, trial"<br>**Comment**: blinding of participants who received no intervention was not possible |
| Blinding of outcome assessment (detection bias) | Low risk | **Quote:**<br>**Comment:** Non-blinded trial; assumed to be low-risk due to automated nature of hemoglobin assessment |
| Incomplete outcome data (attrition bias) | Low risk | **Attrition:** Hemoglobin data are available from the cross-sectional survey component of this trial.<br>**Comment:** Hb assessed/included in present analyses in ~53-55% of enrolled children (not all enrolled children were eligible to be included in the present analyses, due to enrollment of children 0-18 months of age), similar across trial arms |
| Selective reporting (reporting bias) | Low risk | **Comment**: trial registered as NCT02245152 at ClinicalTrials.gov, published protocol (Huybregts BMC Public Health 2017); outcomes described in the methods section reported in the results section; data made available to IPD investigators. |
| Other bias | Low risk | **Comment**: Hb assessed in all children; no other potential sources of bias reported |

**Funding**
The PROMIS studies were funded by Global Affairs Canada (GAC) (https://www.international.gc.ca/); grant 52308/5252/0200 and CGIAR Agriculture for Nutrition and Health (A4NH) program (http://a4nh.cgiar.org/) led by the International Food Policy Research Institute (IFPRI), with no role by either funder in study design, data collection and analysis, decision to publish, or preparation of the manuscript.

| **Christian, 2015 (35); Campbell, 2020 (15)** | | |
|---|---|---|
| **Bias** | **Authors' judgement** | **Support for judgement** |
| Random sequence generation (selection bias) | Low risk | **Quote:** "A random-number seed was selected by a statistician not involved in the study, using a random number generator, and a random number between 0 and 1 drawn from a uniform distribution was assigned to each sector"<br>**Comment:** adequately done |
| Allocation concealment (selection bias) | Low risk | **Quote: "**Cluster-randomization of the 596 predefined communities in JiVitA, called 'sectors', was done by blocks of 19 (total 32 blocks, last block had 7 sectors). A random-number seed was selected by a statistician not involved in the study, using a random number generator, and a random number between 0 and 1 drawn from a uniform distribution was assigned to each sector. Additionally, a block number was assigned to each sector in groups of 19. For blocks 1–31, the first five sectors by sort order were assigned to treatment group 1, the next five to treatment group 2, and so on. For block 32, the two larger controls were assigned two sectors and the intervention groups 1 sector each.<br>**Comment**: central randomization of a cluster-randomized trial |
| Blinding of participants and personnel (performance bias) | High risk | **Quote:** "Our trial was unblinded"<br>**Comment:** not done |
| Blinding of outcome assessment (detection bias) | Low risk | **Quote:**<br>**Comment**: not-blinded trial; assumed to be low risk due to automated nature of hemoglobin assessment and analysis of biomarkers of MN status at a non-field based laboratory. |
| Incomplete outcome data (attrition bias) | Low risk | **Attrition:** control group = 1312/1591; Plumpy'Doz group = 1395/1599; rice lentil group = 785/901; chickpea group = 786/920; WSB++ group = 789/928; "Most missing samples were due to parental refusal (n = 34) or the child being unavailable within the eligibility period (n = 30); the remaining (n = 12) were due to other reasons, such as moving."<br>**Comments**: reasons given for loss to follow-up; missing outcome data balanced in numbers across intervention groups; Hb and MN biomarker data available for ~10-14% of enrolled children, similar across trial arms (selected sub-study) |
| Selective reporting (reporting bias) | Low risk | **Comment:** trial registered as NCT01562379 at ClinicalTrials.gov, outcomes described in the methodology section reported in the results section |
| Other bias | Low risk | **Quote:** "In a 93-sector area (the "sub-study")—selected to yield a target sample size of 750 participants (150 per arm), to be contiguous and accessible by road, socioeconomically representative, and balanced by intervention arm—additional assessments were conducted for secondary outcomes, including blood collection at 18 mo of age to assess biochemical markers of nutritional status."<br>**Comment:** sub-sample may not be representative of the entire population; no other potential sources of bias reported |

**Funding**
The JiVitA-4 study was funded by the US Department of Agriculture, NIFA under the FANEP [Award no. 2010-38418-21732]. In kind support in the form of micronutrient premix for the local food supplements was provided by DSM, Basel, Switzerland and Plumpy'doz was provided by Nutriset (Malaunay, France).

| Dewey, 2017 (36); Matias, 2018 (10) | | |
|---|---|---|
| **Bias** | **Authors' judgement** | **Support for judgement** |
| Random sequence generation (selection bias) | Low risk | **Quote:** "For the randomization, the study statistician at UCD first stratified all 64 clusters in the 11 unions by subdistrict and union and then randomly assigned each cluster to 1 of the 4 arms (each containing 16 clusters)"<br>**Comment:** adequately done |
| Allocation concealment (selection bias) | Low risk | **Quote:** "For the randomization, the study statistician at UCD first stratified all 64 clusters in the 11 unions by subdistrict and union and then randomly assigned each cluster to 1 of the 4 arms (each containing 16 clusters)"<br>**Comment**: central randomization of a cluster-randomized trial |
| Blinding of participants and personnel (performance bias) | High risk | **Comment**: participant blinding not possible due to the nature of the intervention (LNS, MNP, Control) |
| Blinding of outcome assessment (detection bias) | Low risk | **Quote:** "The trial was a researcher-blind, longitudinal, cluster randomized effectiveness trial"<br>**Comment:** adequately done |
| Incomplete outcome data (attrition bias) | Low risk | **Attrition**: LNS-LNS = 884/1047; IFA-LNS = 785/930; IFA-MNP = 895/1052; IFA-Control = 816/982<br>**Comment:** reasons given for loss to follow-up; missing outcome data balanced in numbers across intervention groups; Hb and MN biomarker data available for ~30% of enrolled children, similar across trial arms (randomly assigned to child biochemical sub-sample at enrollment) |
| Selective reporting (reporting bias) | Low risk | **Comment:** The trial was registered at ClinicalTrials.gov ( NCT01715038 ); outcomes described in the methods section reported in the results section |
| Other bias | Low risk | **Quote:** "During enrollment, a total of 1346 (unborn) children were randomly assigned to the child biochemical subsample"<br>**Comment:** no other potential sources of bias reported |
| **Funding**<br>Supported by the Office of Health, Infectious Diseases, and Nutrition, Bureau for Global Health, US Agency for International Development (USAID) under the terms of cooperative agreement AID-OAA-A-12-00005, through the Food and Nutrition Technical Assistance III Project (FANTA), managed by FHI 360. Our research intervention was incorporated into the community heath and development program of LAMB, which was supported by Plan-Bangladesh in 6 of the 11 study unions. Nutriset S.A.S. prepared the lipid-based nutrient supplements for this trial, and Hudson Pharmaceuticals Ltd. prepared the iron and folic acid tablets. | | |

| Hess, 2015 (38); Abbeddou 2017 (11) | | |
|---|---|---|
| **Bias** | **Authors' judgement** | **Support for judgement** |
| Random sequence generation (selection bias) | Low risk | **Quote:** "computer-generated an assignment within strata to participate in the intervention cohort ... The same statistician, who was blinded to the intervention, generated a random allocation sequence at the level of the concession for the enrollment of eligible infants in the intervention cohort"<br>**Comment:** adequately done; Hb assessed in all enrolled children; MN biomarkers in a random sub-sample |
| Allocation concealment (selection bias) | Low risk | **Quote: "**The same statistician, who was blinded to the intervention, generated a random allocation sequence at the level of the concession for the enrollment of eligible infants in the intervention cohort"<br>**Comment:** central randomization of a cluster-randomized trial |
| Blinding of participants and personnel (performance bias) | High risk | **Quote:** "The trial was partially masked, as all participants, field staff and researchers remained blinded to the four intervention groups until data analyses were completed, but were aware which communities were assigned to intervention cohort and non-intervention cohort"<br>**Comment:** intervention and non-intervention cohorts non-blinded |
| Blinding of outcome assessment (detection bias) | Low risk | **Quote:** "The trial was partially masked, as all participants, field staff and researchers remained blinded to the four intervention groups until data analyses were completed, but were aware which communities were assigned to intervention cohort and non-intervention cohort."<br>**Comment:** IC and NIC non-blinded; assumed to be low risk due to automated nature of hemoglobin and ZPP assessment and analysis of biomarkers of MN status at a non-field based laboratory. |
| Incomplete outcome data (attrition bias) | Low risk | **Attrition:** LNS-Zn0 group = 489/602; LNS-Zn5 group = 499/613; LNS-Zn10 group = 491/603; LNS-TabZn5 group = 481/617; NIC group = 666/785<br>**Comment:** reasons for loss to follow-up mentioned; Hb data available for ~80-85% of enrolled children, similar across trial arms |
| Selective reporting (reporting bias) | Low risk | **Comment**: Protocol attached as a supplement in the study paper; registered at ClinicalTrials.gov as NCT 00944281; outcomes described in the methods section reported in the results section |
| Other bias | Low risk | **Quote:** "A subset of 1065 children from the IC and NIC were randomly selected for the biochemistry subgroup"<br>**Comment:** no other potential sources of bias reported |
| **Funding**<br>The project was funded by a grant from the Bill & Melinda Gates Foundation to the University of California, Davis. The funder had no role in study design, data collection and analysis, decision to publish, or preparation of the manuscript. | | |

| **Humphrey, 2019 (47); Prendergast, 2019 (48)** | | |
|---|---|---|
| **Bias** | **Authors' judgement** | **Support for judgement** |
| Random sequence generation (selection bias) | Low risk | **Quote:** "clusters were allocated (1:1:1:1) to one of four treatment groups"; "the study's statistician used a constrained randomization technique to identify 500 allocation schemes…From these, 10 allocations were randomly selected. The final allocation was selected at a public randomization event attended by elected representatives"<br>**Comments:** Additional details available in Supplementary Materials (Appendix) and at https://osf.io/w93hy and in (SHINE Trial Team, Clin Infect Dis, 2015) |
| Allocation concealment (selection bias) | Low risk | **Quote:** "the study's statistician used a constrained randomization technique to identify 500 allocation schemes…From these, 10 allocations were randomly selected. The final allocation was selected at a public randomization event attended by elected representatives"<br>**Comments:** Additional details available at https://osf.io/w93hy and in (SHINE Trial Team, Clin Infect Dis, 2015). |
| Blinding of participants and personnel (performance bias) | High risk | **Quote**: "masking of participants and fieldworkers was not possible because of the obvious visual differences between interventions"<br>**Comment:** not done |
| Blinding of outcome assessment (detection bias) | Low risk | **Quote**: "masking of participants and fieldworkers was not possible because of the obvious visual differences between interventions, but investigators were blinded to treatment groups until the final analysis of each pre-specified outcome."<br>**Comment:** not done; assumed to be low risk due to automated nature of hemoglobin assessment |
| Incomplete outcome data (attrition bias) | Low risk | **Comment:** attrition similar across all seven arms with reasons given; Hb data available for 77-83% of enrolled children, similar across trial arms |
| Selective reporting (reporting bias) | Low risk | **Comment:** trial registered as NCT01824940 at ClinicalTrials.gov, published protocol (SHINE Trial Team, Clin Infect Dis 2015), research and statistical analysis plan available at https://osf.io/w93hy ; outcomes described in the methods section reported in the results section |
| Other bias | Low risk | **Comment**: Hb assessed in all participants; no other potential sources of bias reported |
| **Funding**<br>The SHINE trial is funded by the Bill & Melinda Gates Foundation (OPP1021542 to Johns Hopkins Bloomberg School of Public Health and OPP1143707 to Zvitambo Institute for Maternal and Child Health Research), the UK Department for International Development, the Wellcome Trust (093768/Z/10/Z and 108065/Z/15/Z), the Swiss Agency for Development and Cooperation (8106727), UNICEF (PCA-2017-0002), and the US National Institutes of Health (R01 HD060338/HD/NICHD). | | |

| **Huybregts, 2019 (46)** | | |
|---|---|---|
| **Bias** | **Authors' judgement** | **Support for judgement** |
| Random sequence generation (selection bias) | Low risk | **Quote**: "we applied stratified random allocation of the HC catchment areas to control and intervention study groups"; "we first stratified the [health centers] by hierarchical clustering"; "random allocation to control or intervention groups was conducted within each stratum during a community ceremony…forty-eight identical pieces of paper with either 'control' or 'intervention' were mixed in a bag…each [health center] director drew one piece of paper"<br>**Comment:** adequately done |
| Allocation concealment (selection bias) | Low risk | "random allocation to control or intervention groups was conducted within each stratum during a community ceremony…forty-eight identical pieces of paper with either 'control' or 'intervention' were mixed in a bag…each [health center] director drew one piece of paper"<br>**Comment**: adequately done |
| Blinding of participants and personnel (performance bias) | High risk | **Quote**: "non-masked, community-based, trial"<br>**Comment**: blinding of participants who received no intervention was not possible |
| Blinding of outcome assessment (detection bias) | Low risk | **Quote:** "We used a two-arm, cluster-randomized, non-blinded effectiveness trial"<br>**Comment**: not done, cluster randomized trial at level of HC; assumed to be low risk due to automated nature of hemoglobin assessment |
| Incomplete outcome data (attrition bias) | Low risk | **Attrition:** Hemoglobin data are available from the cross-sectional survey component of this trial.<br>**Comment**: Hb assessed in ~82-84% of enrolled children (not all enrolled children were eligible to be included in the present analyses, due to enrollment of children 6-23 months of age), similar across trial arms |
| Selective reporting (reporting bias) | Low risk | **Comment**: trial registered as NCT02323815 at ClinicalTrials.gov, published protocol (Huybregts BMC Public Health 2017); outcomes described in the methods section reported in the results section; data made available to IPD investigators. |
| Other bias | Low risk | **Comment**: Hb assessed in all children; no other potential sources of bias reported |
| **Funding**<br>The PROMIS studies were funded by: Global Affairs Canada (GAC) (https://www.international.gc.ca/); grant 52308/5252/0200, and CGIAR Agriculture for Nutrition and Health (A4NH) program (http://a4nh.cgiar.org/) led by the International Food Policy Research Institute (IFPRI), with no role by either funder in study design, data collection and analysis, decision to publish, or preparation of the manuscript. | | |

| **Luby, 2018 (37); Stewart, 2019 (12)** | | |
|---|---|---|
| **Bias** | **Authors' judgement** | **Support for judgement** |
| Random sequence generation (selection bias) | Low risk | **Quote**: Clusters were randomly allocated to treatment using a random number generator by a coinvestigator at University of California, Berkeley (BFA). Each of the eight geographically adjacent clusters was block randomized to the double-sized control arm or one of the six interventions…Geographical matching ensured that arms were balanced across locations and time of measurement."<br>**Comment**: adequately done |
| Allocation concealment (selection bias) | Low risk | **Quote** "Clusters were randomly allocated to treatment using a random number generator by a coinvestigator at University of California, Berkeley (BFA)."<br>**Comment**: central randomization of a cluster-randomized trial |
| Blinding of participants and personnel (performance bias) | High risk | **Comment:** Not done due to the nature of the intervention |
| Blinding of outcome assessment (detection bias) | High risk | **Quote: "**Interventions included distinct visible components so neither participants nor data collectors were masked to intervention assignment, although the data collection and intervention teams were different individuals"' "masked technicians completed the laboratory analysis"; "There were delays in the timing of sample collection in the Bangladesh study due to security concerns arising from civil unrest … the interpretation is complicated by the delayed timing of sample collection relative to when LNS supplementation ceased and by the seasonal imbalance in sample collection between groups";<br>**Comment**: timing of endline assessments differed by intervention arm due to civil unrest; passive control arm; blinding not possible due to the nature of the intervention; assessment and the masked analysis of biomarkers of MN status at a non-field based laboratory |
| Incomplete outcome data (attrition bias) | Low risk | **Quote:**<br>**Comment:** attrition similar across all seven arms with reasons given; Hb assessed in 18-23% of enrolled children (not all enrolled children were eligible to be included in the present analyses, due to assessment > 3 months after study-defined end of supplementation) |
| Selective reporting (reporting bias) | Low risk | **Comment** The trial was registered at clinicaltrials.gov (NCT 01590095); SAP and trial protocol available, and published (Arnold BMJ Open 2013); outcomes described in the methods section reported in the results section |
| Other bias | Low risk | **Quote:** "Sub-study clusters were selected from the N, WSH, WSH+N, and control arm. Clusters were selected based on the logistical feasibility of the preservation and collection of biological specimens, as well as their transport to the central laboratory. Index households with live-born infants residing in selected clusters were invited to participate in the sub-study activities" |

| | | |
|---|---|---|
| | | **Comment:** no other potential sources of bias reported |

**Funding**
Supported by a global development grant (OPPGD759) from the Bill & Melinda Gates Foundation to the University of California, Berkeley, CA, USA.

| **Maleta, 2015 (45)** | | |
|---|---|---|
| **Bias** | **Authors' judgement** | **Support for judgement** |
| Random sequence generation (selection bias) | Low risk | **Quote:** "We used block randomization and a set of opaque envelopes to assign participants to the intervention groups. The randomization list and envelopes were prepared by a study statistician not involved in trial implementation"<br>**Comment:** adequately done |
| Allocation concealment (selection bias) | Low risk | **Quote:** "We used block randomization and a set of opaque envelopes to assign participants to the intervention groups."<br>**Comment:** adequately done |
| Blinding of participants and personnel (performance bias) | High risk | **Comment**: participant blinding not possible due to the nature of the intervention (LNS, Control) |
| Blinding of outcome assessment (detection bias) | Low risk | **Quote:** "…and the code was not disclosed to the researchers or to those assessing the outcomes until all data had been entered and verified in a database." "For the LNS group, we used single-masked procedures (i.e., fieldworkers who delivered the supplements knew which children were receiving LNSs, but those who performed the anthropometric measurements or assessed other outcomes were not aware of group allocation)."<br>**Comment:** adequately done; assume to be low risk due to automated nature of Hb and ZPP measurement. |
| Incomplete outcome data (attrition bias) | Low risk | **Attrition:** 40g/day milk-free LNS group =239/324; 40g/day milk LNS group = 242/322; 20g/day milk-free LNS group = 247/323; 20g/day milk LNS group = 236/322;10g/day milk LNS group = 221/321; control group = 242/320<br>**Comment**: similar levels of attrition across groups, reasons for dropout provided; Hb available for 25-26% of enrolled children |
| Selective reporting (reporting bias) | Low risk | **Comment**: The trial was registered at clinicaltrials.gov ( NCT00945698); SAP and trial protocol available online; outcomes described in the methods section reported in the results section |
| Other bias | Low risk | **Comment:** The biochemistry sub-sample was randomly selected (100 infants/group); no other potential sources of bias reported |
| **Funding**<br>Funded by a grant to the University of California, Davis, from the Bill & Melinda Gates Foundation. | | |

| **Null, 2018 (42); Stewart, 2019 (12)** | | |
|---|---|---|
| **Bias** | **Authors' judgement** | **Support for judgement** |
| Random sequence generation (selection bias) | Low risk | **Quote:** "Clusters were randomly allocated to treatment at the University of California, Berkeley using a random number generator with reproducible seed"<br>**Comment:** adequately done |
| Allocation concealment (selection bias) | Low risk | **Quote:** "Clusters were randomly allocated to treatment at the University of California, Berkeley using a random number generator with reproducible seed"<br>**Comment**: central randomization of a cluster-randomized trial |
| Blinding of participants and personnel (performance bias) | High risk | **Quote:** "Masking participants was not possible"<br>**Comment:** blinding of participants was not possible due to the nature of the intervention |
| Blinding of outcome assessment (detection bias) | Low risk | **Quote**: "The health promoters and staff who delivered the interventions were not involved in data collection, but the data collection team could have inferred treatment status if they saw intervention materials in study communities."<br>**Comment:** blinding not possible due to the nature of the intervention; assumed to be low risk due to automated nature of hemoglobin assessment and the masked analysis of biomarkers of MN status at a non-field based laboratory. |
| Incomplete outcome data (attrition bias) | High risk | **Quote:** "Among those who attended the study visit, an additional 608 refused to provide a blood sample and 142 had missing data due to insufficient blood volume collected or other reasons"; **"**In Kenya, there were high rates of refusal for the blood draw, effectively halving the sample size of the baseline cohort when combined with other reasons for losses to follow-up. This high rate of attrition could have led to some selection bias in the study sample"<br>**Comment:** attrition similar across all seven arms with reasons given; Hb concentrations were assessed in 24-32% of enrolled children |
| Selective reporting (reporting bias) | Low risk | **Comment:** Trial registered as NCT01704105 at ClincialTrials.gov. SAP and trial protocol available online, and published (Arnold BMJ Open 2013); outcomes described in the methods section reported in the results section |
| Other bias | Low risk | **Quote:** "Clusters were selected based on the logistical feasibility of the preservation and collection of biological specimens, as well as their transport to the central laboratory"<br>**Comment:** no additional potential sources of bias reported |
| **Funding**<br>Supported in part by Global Development grant OPPGD759 from the Bill & Melinda Gates Foundation to the University of California, Berkeley, CA, USA, and grant AID-OAA-F-13-00040 from United States Agency for International Development (USAID) to Innovations for Poverty Action. This manuscript was made possible by the generous support of the American people through the USAID. The contents are the responsibility of the authors and do not necessarily reflect the views of USAID or the US Government. | | |

| **Galasso, 2019 (43); Stewart, 2020 (13)** | | |
|---|---|---|
| **Bias** | **Authors' judgement** | **Support for judgement** |
| Random sequence generation (selection bias) | Low risk | **Quote: "**a random generator was used to block-randomise five sites per intervention group per region"… "An up-to-date registry of government-programme eligible women and children… was used as a sampling frame to select households eligible for enrolment in the trial. 30 households were randomly sampled per site, stratified by children's age at baseline"; "A subset of 64 sites was randomly sampled among the list of sites that were accessible via a paved road (82 out of the 125).<br>**Comment:** adequately done |
| Allocation concealment (selection bias) | Low risk | **Comment:** central randomization of a cluster-randomized trial |
| Blinding of participants and personnel (performance bias) | Low risk | **Quote:** "Due to the nature of the interventions, masking of participants and community health workers was not possible."<br>**Comment:** not done; assumed to be low risk due to automated nature of hemoglobin assessment and the masked analysis of biomarkers of MN status at a non-field based laboratory. |
| Blinding of outcome assessment (detection bias) | Low risk | **Quote:** "Due to the nature of the interventions, masking of participants and community health workers was not possible. Data analysts were not blinded to intervention group assignment due to differences in survey information"<br>**Comment:** not done; assumed to be low risk due to automated nature of hemoglobin assessment and the masked analysis of biomarkers of MN status at a non-field based laboratory |
| Incomplete outcome data (attrition bias) | Low risk | **Quote:** "Mothers or children who died before the final assessment were not replaced. Children who had permanently moved outside the programme site catchment area before final assessment were replaced with a randomly drawn child from the site within the same age range. Children and their households who returned to the site between the baseline and final assessment were re-interviewed."<br>**Comment**: similar levels of attrition across groups, reasons for dropout provided; anemia was assessed in ~39-40% of enrolled children, similar across trial arms (not all enrolled children were eligible to be included in the present analyses, due to enrollment of children 6-11 months of age, and assessment > 3 months after study-defined end of supplementation) |
| Selective reporting (reporting bias) | Low risk | **Quote:** "This trial has been registered with the ISRCTN registry, number ISRCTN14393738."<br>**Comment:** published protocol (Fernald BMC Public Health 2016); outcomes described in the methods section reported in the results section |
| Other bias | Low risk | **Quote:** "A random draw of 16 sites each from T0, T2, T3, and T4 was selected for the collection of blood samples to estimate the impact of the interventions on micronutrient status (T1 was excluded owing to cost constraints).Within each of the selected sites, a |

| | | |
|---|---|---|
| | | random sample of 6 children in the youngest age cohort (18–24 mo at endline) was selected for the assessment of micronutrient biomarkers. The roster of children alive and measured at midline was used as a sampling frame for this selection."<br>**Comment:** no other potential sources of bias reported |

**Funding**
Funded by the Eunice Kennedy Shriver National Institutes of Child Health and Human Development, Strategic Impact Evaluation Fund, Early Learning Partnership Program, World Bank Innovation Grant, Grand Challenges Canada, World Bank Research Committee, Japan Nutrition Trust Fund, Power of Nutrition Trust Fund.
