## Supplemental Table 8 for "Characteristics that modify the effect of small-quantity lipid-based nutrient supplementation on child anemia and micronutrient status: an individual participant data meta-analysis of randomized controlled trials"

**Table 8. Supplemental overview of individual-level effect modification**

| | Plasma zinc | Retinol | Marginal retinol | | RBP | Marginal RBP | |
|---|---|---|---|---|---|---|---|
| **Effect modifier** | GMR | GMR | PR | PD | GMR | PR | PD |
| Higher (vs. lower) maternal BMI | | | | | | | |
| Older (vs. younger) mother | | | | | | | |
| Higher (vs. lower) maternal education | | | | | | | |
| Female (vs. male) child | | | | | | | |
| Later-born (vs. first-born) child | | | | | | | |
| Acutely (vs. non-acutely) malnourished | | | | | | | |
| Anemic (vs. non-anemic) | | | | | | | |
| Received (vs. did not receive) Vitamin A | | | | | | | |
| No high (vs. high) AGP or CRP | | | | | | | (C) |
| Higher (vs. lower) SES | | | | | | | |
| Food secure (vs. insecure) | | | | | | | |
| Improved (vs. unimproved) water quality | | | | | | | |
| Improved (vs. unimproved) sanitation | | | | | | | |
| Rainy (vs. dry) season | | | | | | | |

MD=mean difference; GMR=geometric mean ratio; PR=prevalence ratio; PD=prevalence difference. Green indicates stronger effect in the indicated subgroup while blue indicates a stronger effect in the other subgroup. Dark color indicates p-for-interaction $< 0.05$; light color indicates $0.05 < p < 0.1$. The letter "C" indicates that the apparent effect modification is due to the cutoff effect; when "C" is in parentheses, it is partially explained by the cutoff effect.
