## Supplemental Figures 1-8 for "Characteristics that modify the effect of small-quantity lipid-based nutrient supplementation on child anemia and micronutrient status: an individual participant data meta-analysis of randomized controlled trials"

Supplemental Figure 1: Summary risk of bias as a percentage of all included studies for the effects of SQ-LNS on biochemical outcomes

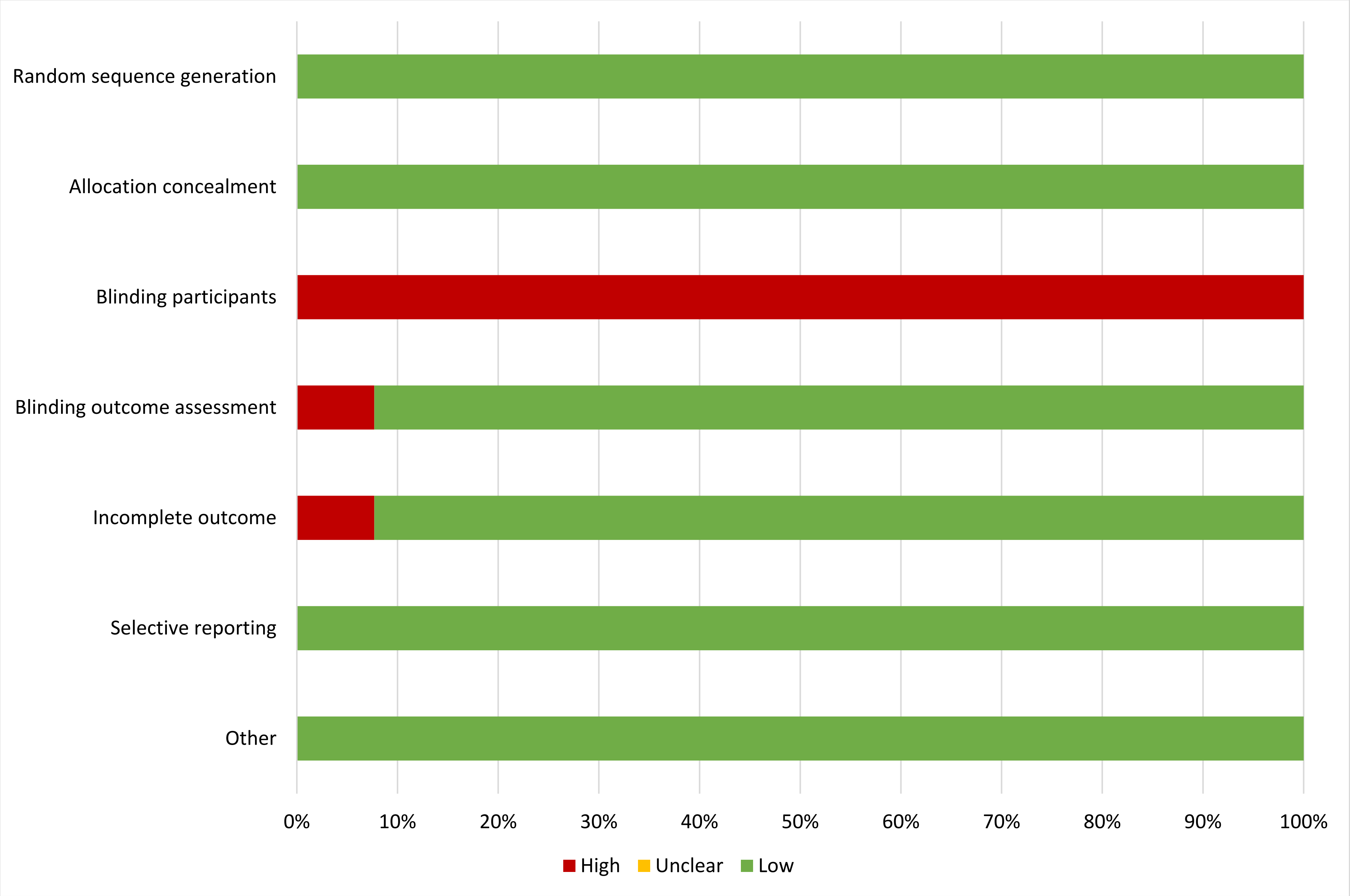

### Supplemental figure 2: Sensitivity analyses of main effects of SQ-LNS on biochemical outcomes

These figures show the pooled estimates of intervention effects by different pooling methods and different sensitivity analyses. For continuous outcomes, the intervention effect is measured by the difference in mean of the LNS group minus control. For log transformed continuous outcomes, the intervention effect is measured by the ratio of geometric means, the effect estimate is the geometric mean in the LNS group divided by the geometric mean in the control group. For dichotomous outcomes analyzed via prevalence ratios, the effect estimate is the prevalence in the LNS group divided by the prevalence in the control group. For dichotomous outcomes analyzed via prevalence differences, the effect estimate is the prevalence in the LNS group minus the prevalence in the control group.
The labels on the left y-axis indicate which outcome is assessed. The different columns correspond to sensitivity analyses in which intervention group categorization differs. All-trial analysis includes all trials; Child-LNS-only excludes trial arms that provided both maternal and child LNS; Multi-component analysis separates comparisons within trials that included multi-component interventions, so that the SQ-LNS vs. no SQ-LNS comparisons were conducted separately between pairs of arms that included the same non-nutrition components (e.g. SQ-LNS+WASH vs. WASH; SQ-LNS vs. Control); Passive arms excluded analysis excludes passive control arms. Depending on the sensitivity analysis, there may not have been enough comparisons available to generate a pooled estimate.
sTfR, soluble transferrin receptor; ZPP, zinc protoporphyrin; RBP, retinol binding protein.

#### Supplemental figure 2A: Mean differences in hemoglobin concentration

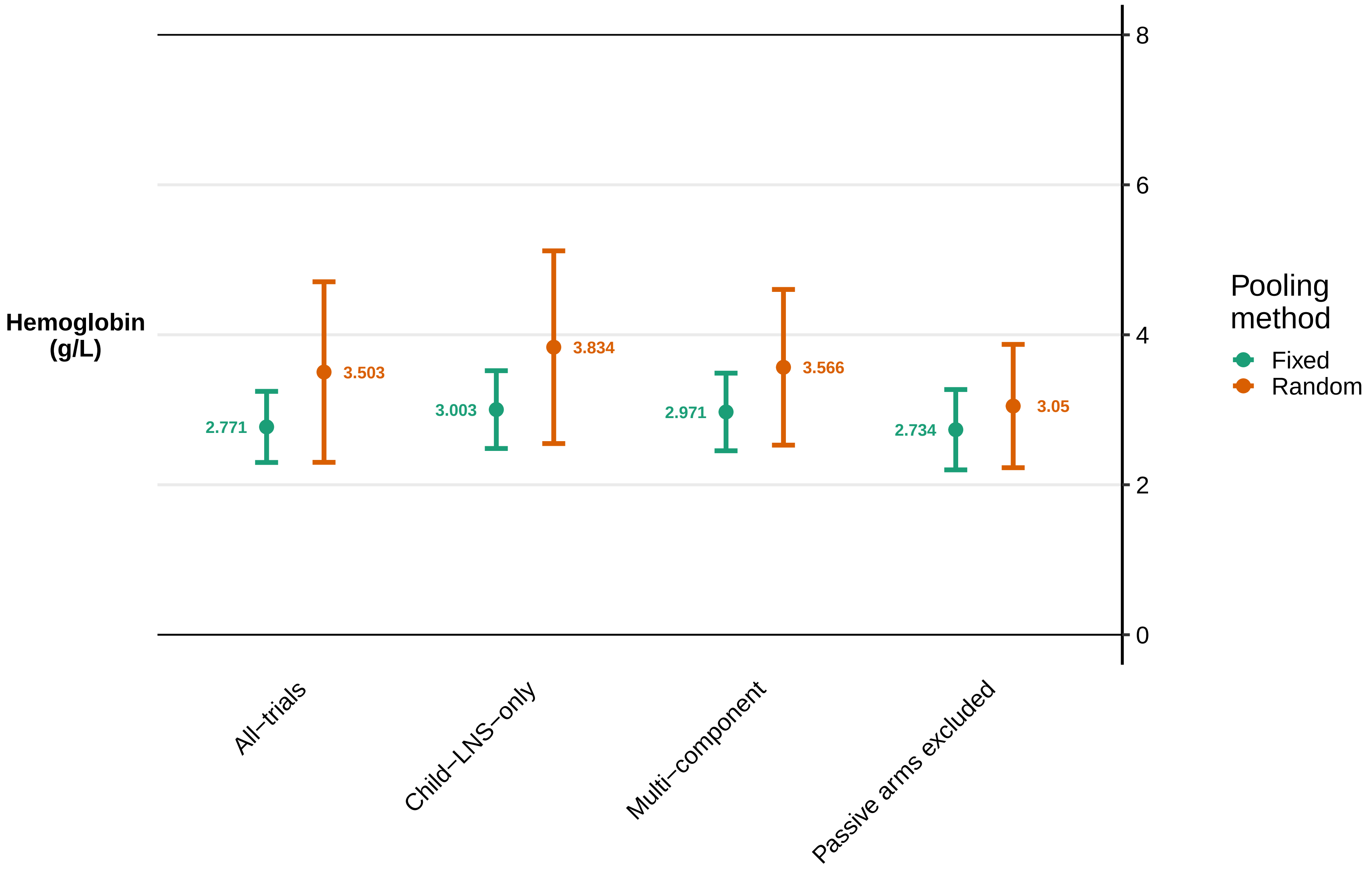

#### Supplemental figure 2B:Geometric mean ratios for log transformed continuous outcomes

##### Supplemental figure 2B1: Geometric mean ratios for ferritin, sTfR, and ZPP

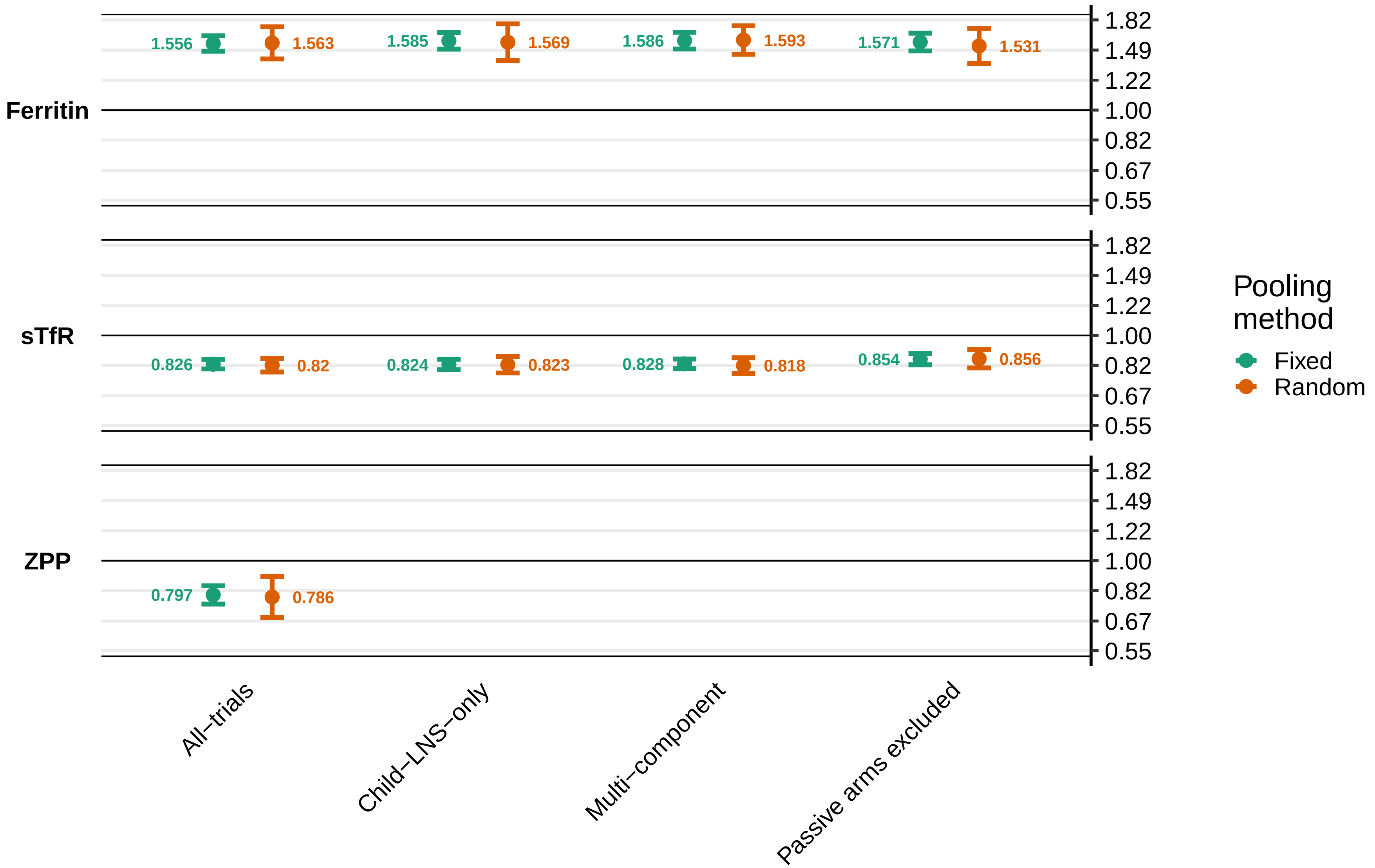

#### Supplemental figure 2B2: Geometric mean ratios for plasma zinc, retinol, and RBP

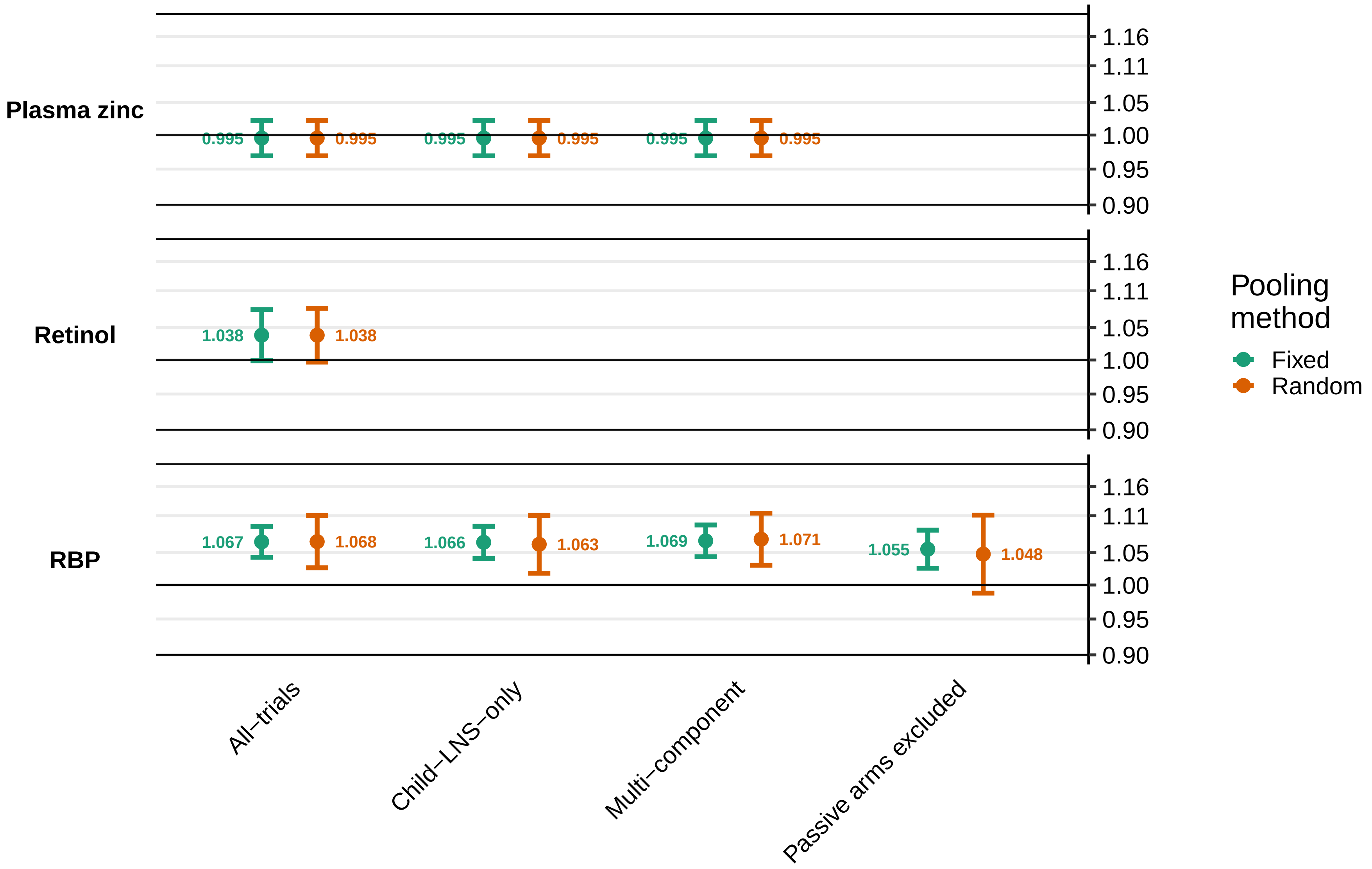

#### Supplemental figure 2C: Prevalence ratios for dichotomous outcomes

##### Supplemental figure 2C1: Prevalence ratios for anemia, moderate-to-severe anemia, iron deficiency, iron deficiency anemia, elevated sTfR, and elevated ZPP

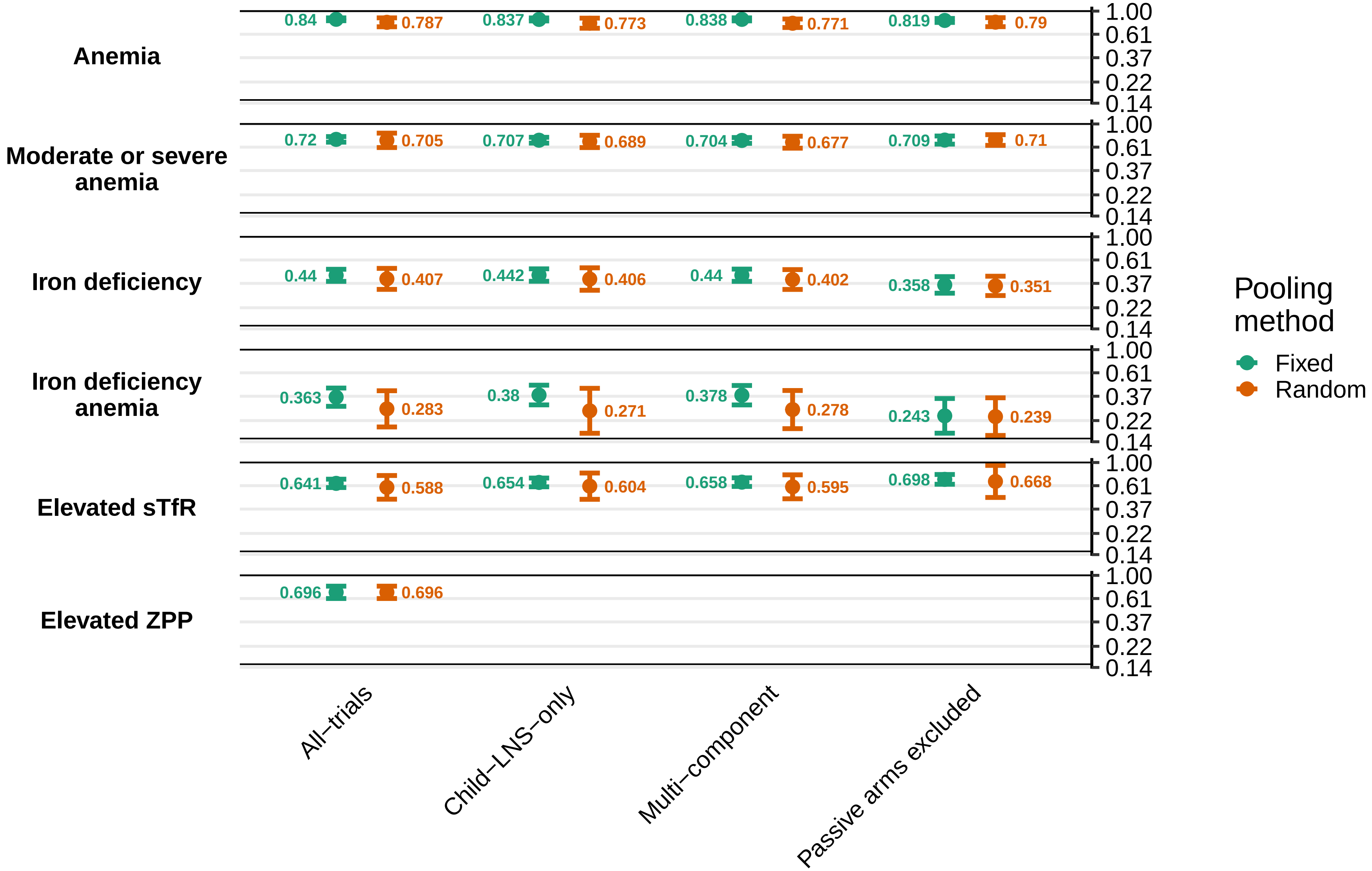

#### Supplemental figure 2C2: Prevalence ratios for low and marginal vitamin A status

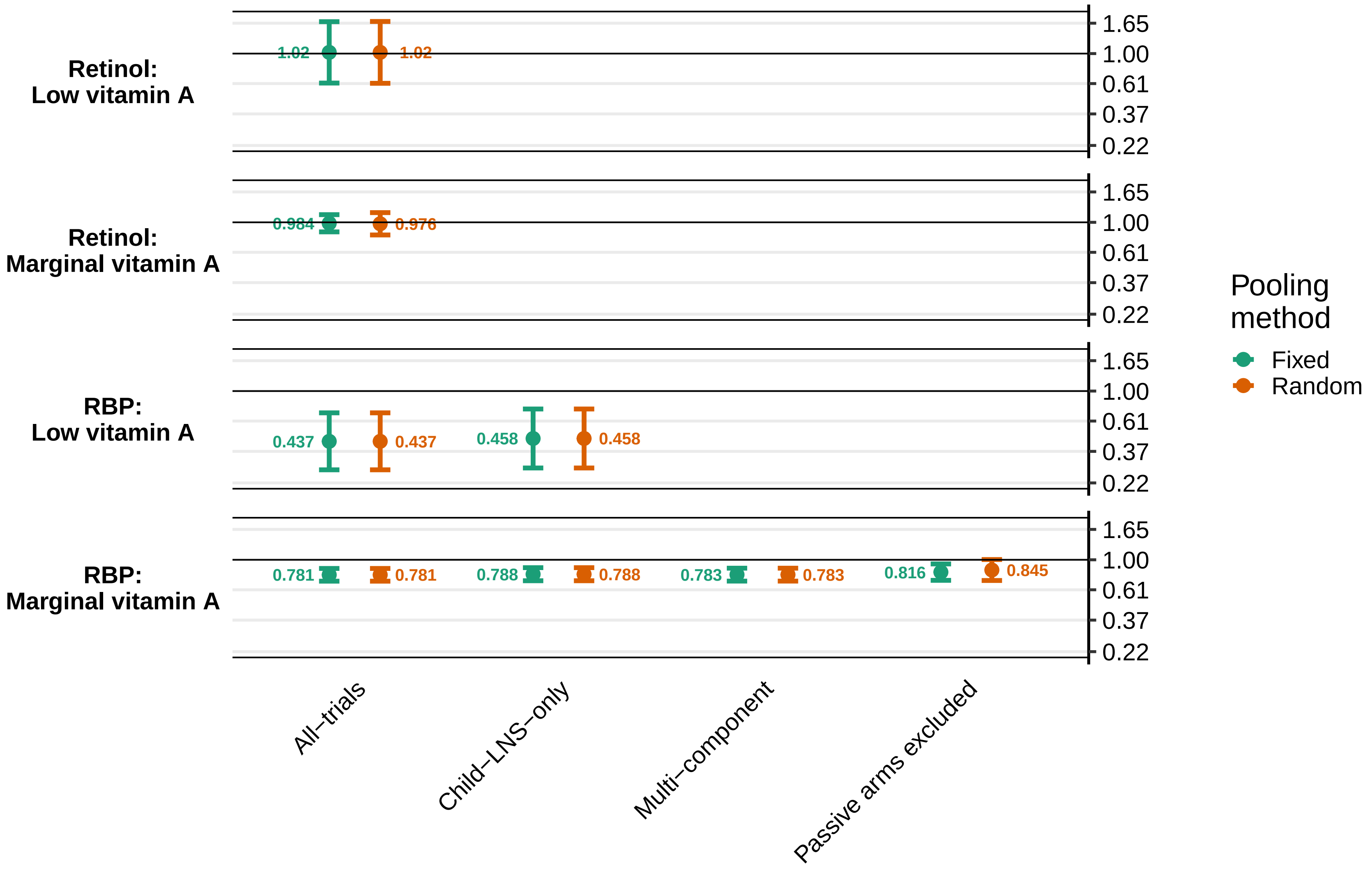

#### Supplemental figure 2D: Prevalence differences for dichotomous outcomes

##### Supplemental figure 2D1: Prevalence differences for anemia, moderate-to-severe anemia, iron deficiency, iron deficiency anemia, elevated sTfR, and elevated ZPP

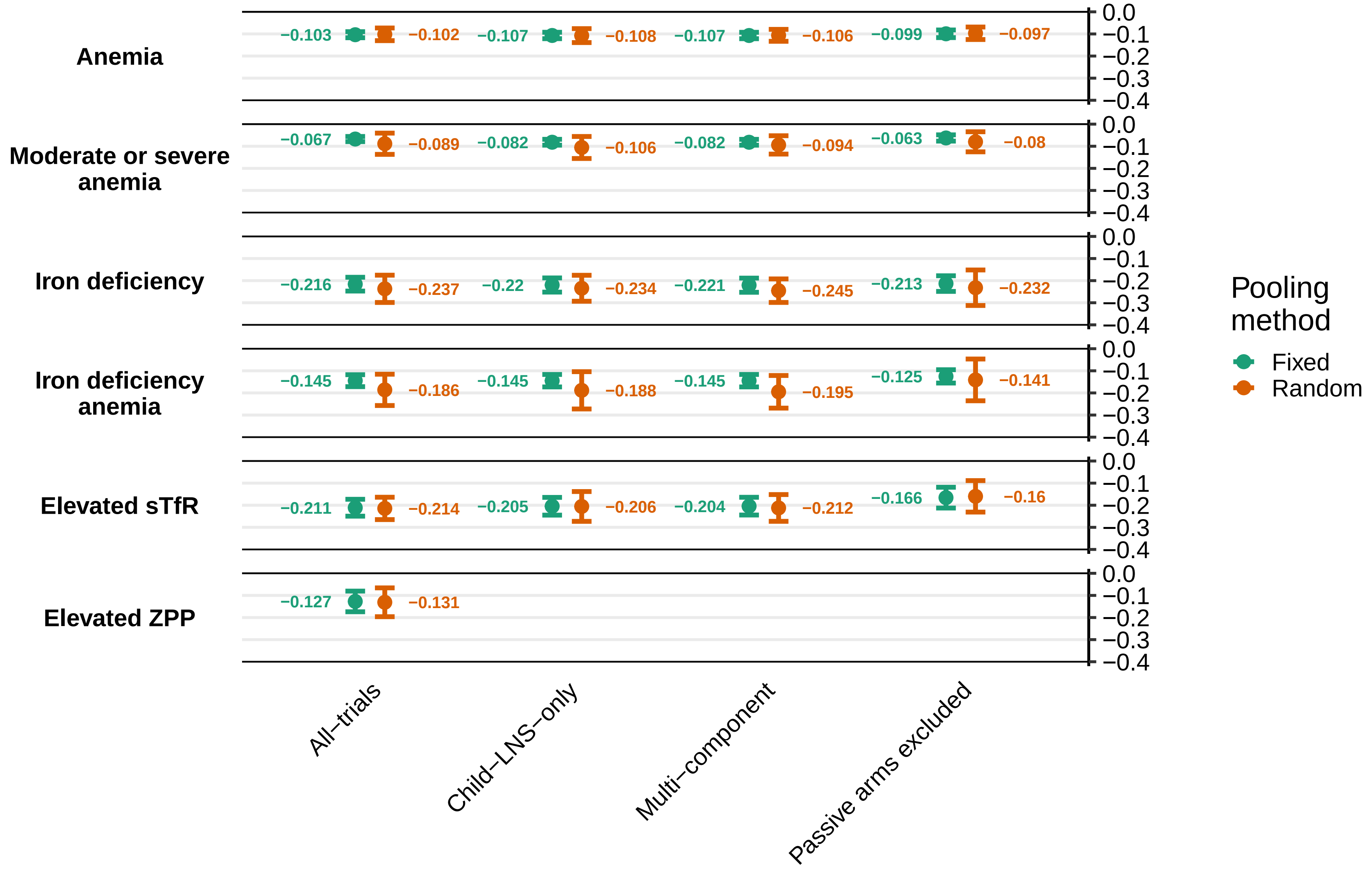

#### Supplemental figure 2D2: Prevalence ratios for low and marginal vitamin A status

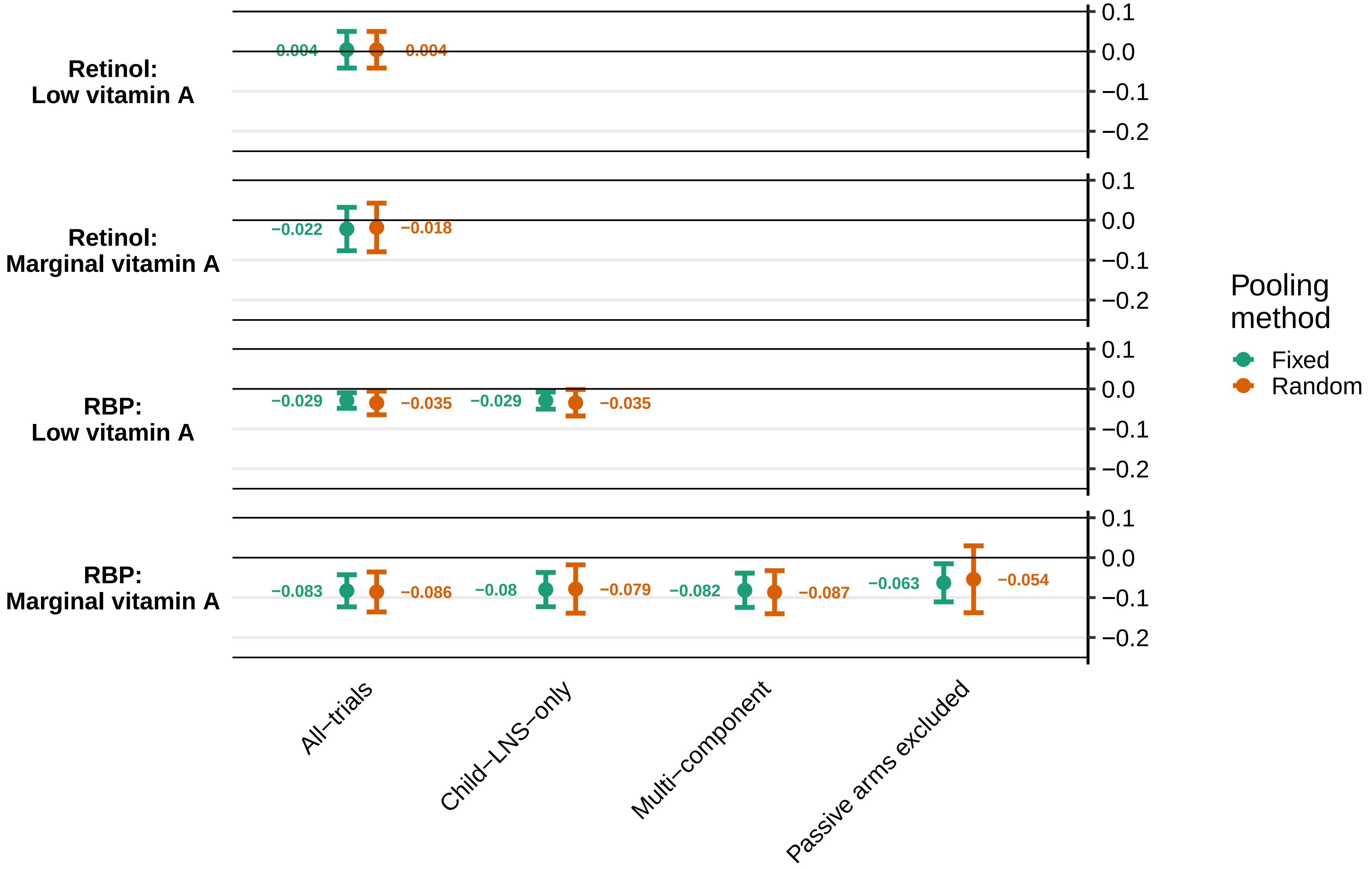

### Supplemental figure 3: Forest plots for all main effects of SQ-LNS on biochemical outcomes

#### Contents

**Supplemental figure 3A: Mean difference in hemoglobin concentration** 3

**Supplemental figure 3B: Anemia prevalence ratio** 4

**Supplemental figure 3C: Anemia prevalence difference** 5

**Supplemental figure 3D: Moderate-to-severe anemia prevalence ratio** 6

**Supplemental figure 3E: Moderate-to-severe anemia prevalence difference** 7

**Supplemental figure 3F: Geometric mean ratio of ferritin concentration** 8

**Supplemental figure 3G: Iron deficiency (ferritin < 12 µg/L) prevalence ratio** 9

**Supplemental figure 3H: Iron deficiency (ferritin < 12 µg/L) prevalence difference** 10

**Supplemental figure 3I: Iron deficiency anemia prevalence ratio** 11

**Supplemental figure 3J: Iron deficiency anemia prevalence difference** 12

**Supplemental figure 3K: Geometric mean ratio of soluble transferrin receptor concentration** 13

**Supplemental figure 3L: Elevated soluble transferrin receptor prevalence ratio** 14

**Supplemental figure 3M: Elevated soluble transferrin receptor prevalence difference** 15

**Supplemental figure 3N: Geometric mean ratio of zinc protoporphyrin concentration** 16

**Supplemental figure 3O: Elevated zinc protoporphyrin prevalence ratio** 17

**Supplemental figure 3P: Elevated zinc protoporphyrin prevalence difference** 18

**Supplemental figure 3Q: Geometric mean ratio of plasma zinc concentration** 19

**Supplemental figure 3R: Geometric mean ratio of retinol concentration** 20

**Supplemental figure 3S: Low vitamin A (retinol < 0.70 µmol/L) prevalence ratio** 21

**Supplemental figure 3T: Low vitamin A (retinol < 0.70 µmol/L) prevalence difference** 22

**Supplemental figure 3U: Marginal vitamin A (retinol < 1.05 µmol/L) prevalence ratio** 23

**Supplemental figure 3V: Marginal vitamin A (retinol < 1.05 µmol/L) prevalence difference** 24

**Supplemental figure 3W: Geometric mean ratio of retinol binding protein concentration** 25

**Supplemental figure 3X: Low vitamin A status (RBP < 0.70 µmol/L) prevalence ratio** 26

**Supplemental figure 3Y: Low vitamin A status (RBP < 0.70 µmol/L) prevalence difference** 27

**Supplemental figure 3Z: Marginal vitamin A status (RBP < 1.05 µmol/L) prevalence ratio** 28

**Supplemental figure 3AA: Marginal vitamin A status (RBP < 1.05 μmol/L) prevalence difference** **29**

These figures are forest plots showing the study-level estimates of intervention effect with the pooled estimate in the bottom summary rows. For continuous outcomes the intervention effect is measured by the difference in mean of the LNS group minus control. For log transformed continuous outcomes, the intervention effect is measured by the ratio of geometric means, the effect estimate is the geometric mean in the LNS group divided by the geometric mean in the control group. For dichotomous outcomes analyzed via prevalence ratios the effect estimate is the prevalence in the LNS group divided by the prevalence in the control group. For dichotomous outcomes analyzed via prevalence differences the effect estimate is the prevalence in the LNS group minus the prevalence in the control group. The labels on the left y-axis correspond to trial level information. The values on the right indicate the study level effect estimate, confidence interval, and weighting for deriving the pooled estimate.
RBP, retinol binding protein.

### Supplemental figure 3A: Mean difference in hemoglobin concentration

| Country | Trial | LNS N | Control N | MD (95% CI) | Fixed W | Random W |
|---|---|---|---|---|---|---|
| Bangladesh | JiVitA−4 (35) | 457 | 146 | 2.21 (0.34, 4.07) | 0.06 | 0.08 |
| Bangladesh | RDNS (36) | 549 | 272 | 4.10 (1.86, 6.34) | 0.04 | 0.07 |
| Bangladesh | WASH−B (37) | 234 | 186 | 3.09 (1.22, 4.97) | 0.06 | 0.08 |
| Burkina Faso | iLiNS−Zinc (38) | 1957 | 664 | 8.47 (5.47, 11.46) | 0.03 | 0.06 |
| Burkina Faso | PROMIS CS (39) | 574 | 581 | 1.77 (−0.11, 3.64) | 0.06 | 0.08 |
| Ghana | GHANA (40) | 98 | 96 | 8.84 (4.87, 12.81) | 0.01 | 0.05 |
| Ghana | iLiNS−DYADG (41) | 328 | 661 | 1.24 (−0.14, 2.61) | 0.12 | 0.09 |
| Kenya | WASH−B (42) | 350 | 300 | 3.94 (2.05, 5.84) | 0.06 | 0.08 |
| Madagascar | MAHAY (43) | 600 | 588 | 2.77 (−0.12, 5.67) | 0.03 | 0.06 |
| Malawi | iLiNS−DYADM (44) | 210 | 432 | 1.12 (−1.45, 3.69) | 0.03 | 0.07 |
| Malawi | iLiNS−DOSE (45) | 243 | 82 | 3.76 (0.02, 7.51) | 0.02 | 0.05 |
| Mali | PROMIS CS (46) | 953 | 970 | 6.25 (4.16, 8.34) | 0.05 | 0.08 |
| Zimbabwe | SHINE (HIV−) (47) | 1682 | 1594 | 2.13 (1.36, 2.91) | 0.37 | 0.09 |
| Zimbabwe | SHINE (HIV+) (48) | 306 | 285 | 2.78 (0.38, 5.19) | 0.04 | 0.07 |
| | **I² = 0.72, Tau² = 3.88** | **8541** | **6857** | | | |
| | **Fixed** | | | **2.77 (2.30, 3.25)** | | |
| | **Random** | | | **3.50 (2.30, 4.71)** | | |

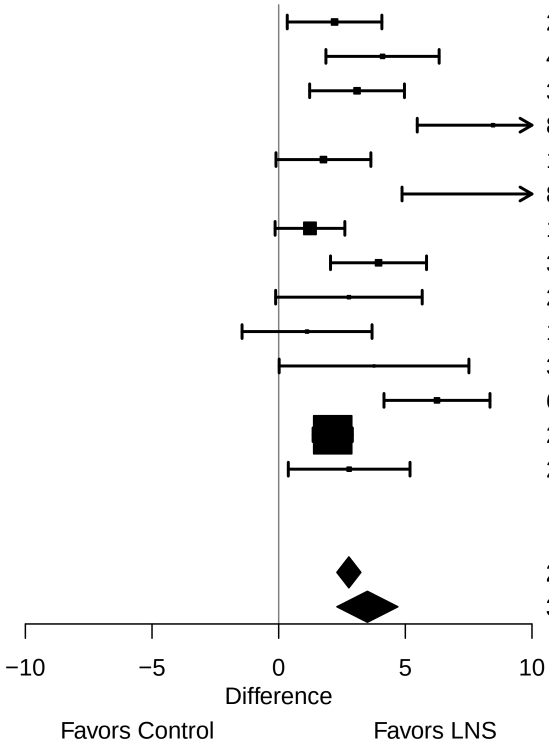

### Supplemental figure 3B: Anemia prevalence ratio

| Country | Trial | LNS N | Control N | PR (95% CI) | Fixed W | Random W |
|---|---|---|---|---|---|---|
| Bangladesh | JiVitA−4 (35) | 457 | 146 | 0.83 (0.54, 1.28) | 0.01 | 0.03 |
| Bangladesh | RDNS (36) | 549 | 272 | 0.66 (0.51, 0.84) | 0.02 | 0.06 |
| Bangladesh | WASH−B (37) | 234 | 186 | 0.42 (0.24, 0.74) | 0.00 | 0.02 |
| Burkina Faso | iLiNS−Zinc (38) | 1957 | 664 | 0.87 (0.83, 0.91) | 0.39 | 0.10 |
| Burkina Faso | PROMIS CS (39) | 574 | 581 | 0.94 (0.85, 1.03) | 0.10 | 0.09 |
| Ghana | GHANA (40) | 98 | 96 | 0.51 (0.36, 0.72) | 0.01 | 0.04 |
| Ghana | iLiNS−DYADG (41) | 328 | 661 | 0.86 (0.73, 1.01) | 0.04 | 0.08 |
| Kenya | WASH−B (42) | 350 | 300 | 0.68 (0.57, 0.81) | 0.03 | 0.08 |
| Madagascar | MAHAY (43) | 600 | 588 | 0.88 (0.78, 1.01) | 0.06 | 0.09 |
| Malawi | iLiNS−DYADM (44) | 210 | 432 | 0.90 (0.76, 1.07) | 0.03 | 0.08 |
| Malawi | iLiNS−DOSE (45) | 243 | 82 | 0.87 (0.74, 1.02) | 0.04 | 0.08 |
| Mali | PROMIS CS (46) | 953 | 970 | 0.82 (0.76, 0.88) | 0.18 | 0.10 |
| Zimbabwe | SHINE (HIV−) (47) | 1682 | 1594 | 0.78 (0.70, 0.86) | 0.10 | 0.09 |
| Zimbabwe | SHINE (HIV+) (48) | 306 | 285 | 0.65 (0.50, 0.85) | 0.01 | 0.06 |
| | **$I^2$ = 0.65, $Tau^2$ = 0.02** | **8541** | **6857** | | | |
| | **Fixed** | | | **0.84 (0.81, 0.87)** | | |
| | **Random** | | | **0.79 (0.72, 0.87)** | | |

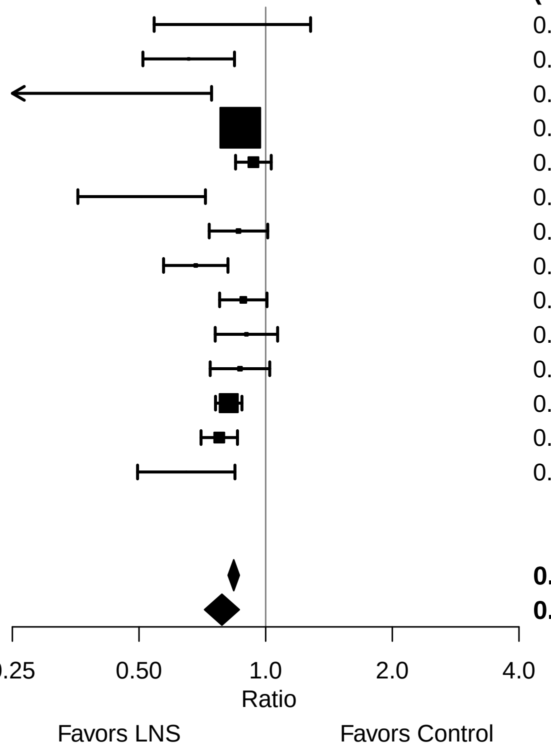

### Supplemental figure 3C: Anemia prevalence difference

| Country | Trial | LNS N | Control N | PD (95% CI) | Fixed W | Random W |
|---|---|---|---|---|---|---|
| Bangladesh | JiVitA−4 (35) | 457 | 146 | −0.03 (−0.09, 0.04) | 0.04 | 0.07 |
| Bangladesh | RDNS (36) | 549 | 272 | −0.14 (−0.21, −0.07) | 0.04 | 0.07 |
| Bangladesh | WASH−B (37) | 234 | 186 | −0.09 (−0.16, −0.03) | 0.05 | 0.07 |
| Burkina Faso | iLiNS−Zinc (38) | 1957 | 664 | −0.12 (−0.15, −0.09) | 0.24 | 0.10 |
| Burkina Faso | PROMIS CS (39) | 574 | 581 | −0.05 (−0.10, 0.01) | 0.07 | 0.08 |
| Ghana | GHANA (40) | 98 | 96 | −0.29 (−0.42, −0.15) | 0.01 | 0.03 |
| Ghana | iLiNS−DYADG (41) | 328 | 661 | −0.06 (−0.13, 0.00) | 0.04 | 0.07 |
| Kenya | WASH−B (42) | 350 | 300 | −0.15 (−0.23, −0.08) | 0.03 | 0.06 |
| Madagascar | MAHAY (43) | 600 | 588 | −0.07 (−0.13, −0.02) | 0.06 | 0.08 |
| Malawi | iLiNS−DYADM (44) | 210 | 432 | −0.05 (−0.13, 0.03) | 0.03 | 0.06 |
| Malawi | iLiNS−DOSE (45) | 243 | 82 | −0.10 (−0.21, 0.02) | 0.01 | 0.04 |
| Mali | PROMIS CS (46) | 953 | 970 | −0.16 (−0.19, −0.12) | 0.14 | 0.10 |
| Zimbabwe | SHINE (HIV−) (47) | 1682 | 1594 | −0.08 (−0.11, −0.05) | 0.19 | 0.10 |
| Zimbabwe | SHINE (HIV+) (48) | 306 | 285 | −0.13 (−0.20, −0.06) | 0.03 | 0.06 |
| | **I² = 0.64, Tau² = 0.00** | **8541** | **6857** | | | |
| | **Fixed** | | | **−0.10 (−0.12, −0.09)** | | |
| | **Random** | | | **−0.10 (−0.13, −0.07)** | | |

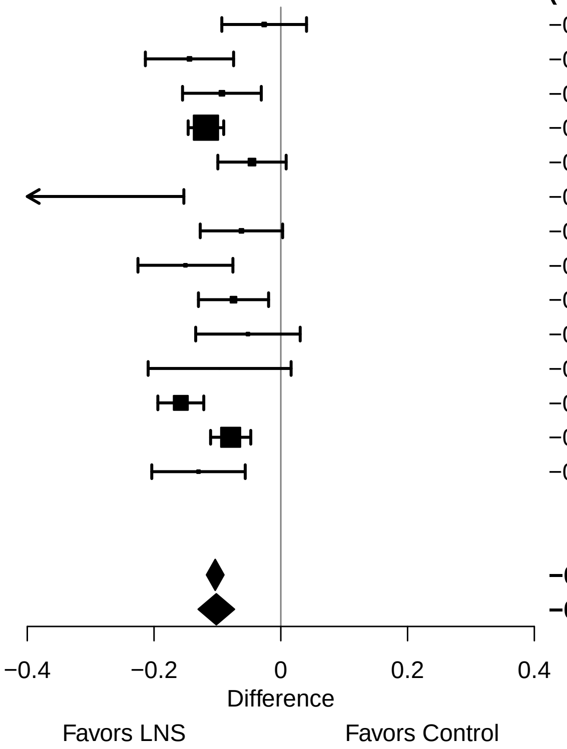

### Supplemental figure 3D: Moderate-to-severe anemia prevalence ratio

| Country | Trial | LNS N | Control N | PR (95% CI) | Fixed W | Random W |
|---|---|---|---|---|---|---|
| Bangladesh | JiVitA−4 (35) | | | | | |
| Bangladesh | RDNS (36) | 549 | 272 | 0.54 (0.38, 0.76) | 0.03 | 0.08 |
| Bangladesh | WASH−B (37) | | | | | |
| Burkina Faso | iLiNS−Zinc (38) | 1957 | 664 | 0.72 (0.65, 0.78) | 0.44 | 0.12 |
| Burkina Faso | PROMIS CS (39) | 574 | 581 | 0.88 (0.73, 1.07) | 0.10 | 0.11 |
| Ghana | GHANA (40) | 98 | 96 | 0.32 (0.16, 0.61) | 0.01 | 0.04 |
| Ghana | iLiNS−DYADG (41) | 328 | 661 | 0.84 (0.47, 1.51) | 0.01 | 0.04 |
| Kenya | WASH−B (42) | 350 | 300 | 0.63 (0.45, 0.88) | 0.03 | 0.08 |
| Madagascar | MAHAY (43) | 600 | 588 | 0.81 (0.64, 1.03) | 0.06 | 0.10 |
| Malawi | iLiNS−DYADM (44) | 210 | 432 | 1.15 (0.87, 1.52) | 0.05 | 0.09 |
| Malawi | iLiNS−DOSE (45) | 243 | 82 | 0.72 (0.54, 0.96) | 0.04 | 0.09 |
| Mali | PROMIS CS (46) | 953 | 970 | 0.63 (0.54, 0.74) | 0.15 | 0.11 |
| Zimbabwe | SHINE (HIV−) (47) | 1682 | 1594 | 0.67 (0.53, 0.84) | 0.07 | 0.10 |
| Zimbabwe | SHINE (HIV+) (48) | 306 | 285 | 0.51 (0.30, 0.89) | 0.01 | 0.05 |
| | **$I^2$ = 0.64, $Tau^2$ = 0.05** | **7850** | **6525** | | | |
| | **Fixed** | | | **0.72 (0.68, 0.76)** | | |
| | **Random** | | | **0.70 (0.60, 0.82)** | | |

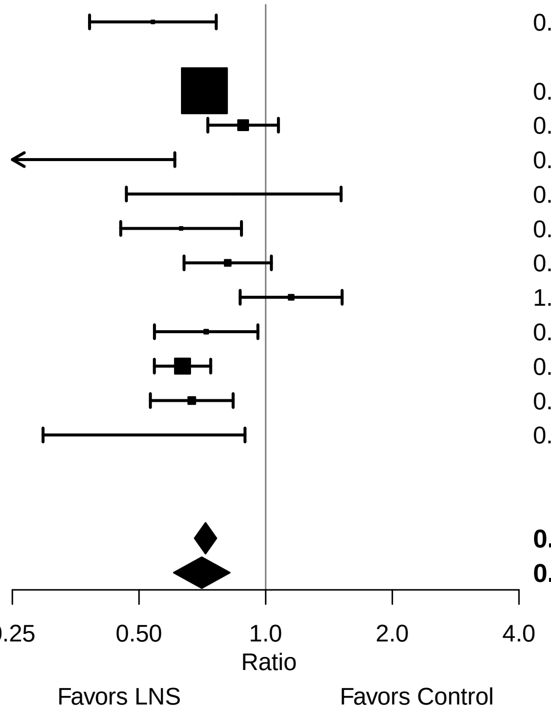

#### Supplemental figure 3E: Moderate-to-severe anemia prevalence difference

| Country | Trial | LNS N | Control N | PD (95% CI) | Fixed W | Random W |
|---|---|---|---|---|---|---|
| Bangladesh | JiVitA−4 (35) | | | | | |
| Bangladesh | RDNS (36) | 549 | 272 | −0.08 (−0.13, −0.03) | 0.05 | 0.09 |
| Bangladesh | WASH−B (37) | | | | | |
| Burkina Faso | iLiNS−Zinc (38) | 1957 | 664 | −0.21 (−0.25, −0.17) | 0.09 | 0.09 |
| Burkina Faso | PROMIS CS (39) | 574 | 581 | −0.04 (−0.10, 0.01) | 0.05 | 0.09 |
| Ghana | GHANA (40) | 98 | 96 | −0.22 (−0.33, −0.11) | 0.01 | 0.06 |
| Ghana | iLiNS−DYADG (41) | 328 | 661 | −0.01 (−0.04, 0.02) | 0.17 | 0.09 |
| Kenya | WASH−B (42) | 350 | 300 | −0.08 (−0.14, −0.02) | 0.04 | 0.08 |
| Madagascar | MAHAY (43) | 600 | 588 | −0.06 (−0.11, −0.01) | 0.05 | 0.09 |
| Malawi | iLiNS−DYADM (44) | 210 | 432 | 0.04 (−0.04, 0.11) | 0.03 | 0.08 |
| Malawi | iLiNS−DOSE (45) | 243 | 82 | −0.13 (−0.25, −0.01) | 0.01 | 0.06 |
| Mali | PROMIS CS (46) | 953 | 970 | −0.22 (−0.26, −0.18) | 0.07 | 0.09 |
| Zimbabwe | SHINE (HIV−) (47) | 1682 | 1594 | −0.04 (−0.05, −0.02) | 0.36 | 0.09 |
| Zimbabwe | SHINE (HIV+) (48) | 306 | 285 | −0.05 (−0.09, −0.01) | 0.07 | 0.09 |
| | **I² = 0.92, Tau² = 0.01** | **7850** | **6525** | | | |
| | **Fixed** | | | **−0.07 (−0.08, −0.06)** | | |
| | **Random** | | | **−0.09 (−0.14, −0.04)** | | |

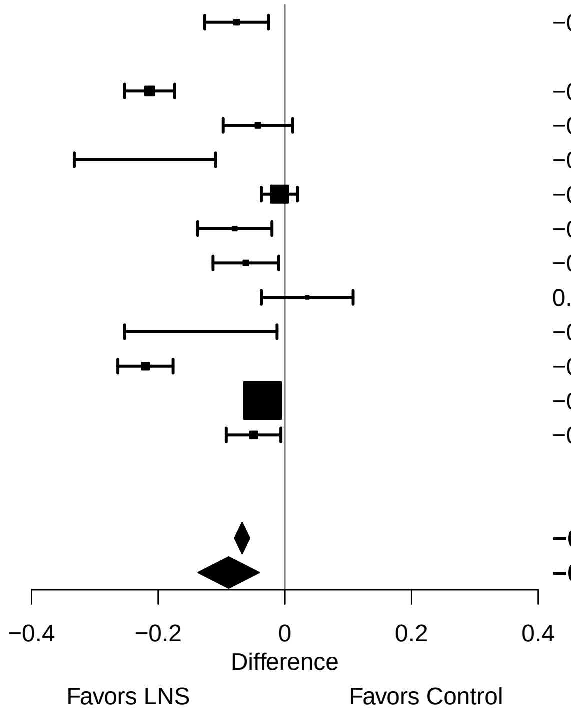

#### Supplemental figure 3F: Geometric mean ratio of ferritin concentration

| Country | Trial | LNS N | Control N | GMR (95% CI) | Fixed W | Random W |
|---|---|---|---|---|---|---|
| Bangladesh | JiVitA−4 (35) | 455 | 144 | 1.39 (1.17, 1.64) | 0.09 | 0.15 |
| Bangladesh | RDNS (36) | 550 | 272 | 1.52 (1.40, 1.64) | 0.44 | 0.21 |
| Bangladesh | WASH−B (37) | 212 | 178 | 1.74 (1.50, 2.01) | 0.12 | 0.16 |
| Burkina Faso | iLiNS−Zinc (38) | 315 | 96 | 1.55 (1.31, 1.84) | 0.09 | 0.15 |
| Burkina Faso | PROMIS CS (39) | | | | | |
| Ghana | GHANA (40) | 83 | 82 | 2.52 (1.49, 4.25) | 0.01 | 0.04 |
| Ghana | iLiNS−DYADG (41) | | | | | |
| Kenya | WASH−B (42) | 298 | 259 | 1.69 (1.50, 1.91) | 0.19 | 0.18 |
| Madagascar | MAHAY (43) | 83 | 51 | 1.29 (1.05, 1.59) | 0.06 | 0.13 |
| Malawi | iLiNS−DYADM (44) | | | | | |
| Malawi | iLiNS−DOSE (45) | | | | | |
| Mali | PROMIS CS (46) | | | | | |
| Zimbabwe | SHINE (HIV−) (47) | | | | | |
| Zimbabwe | SHINE (HIV+) (48) | | | | | |
| | **$I^2$ = 0.53, $Tau^2$ = 0.01** | **1996** | **1082** | | | |
| | **Fixed** | | | **1.56 (1.48, 1.64)** | | |
| | **Random** | | | **1.56 (1.40, 1.74)** | | |

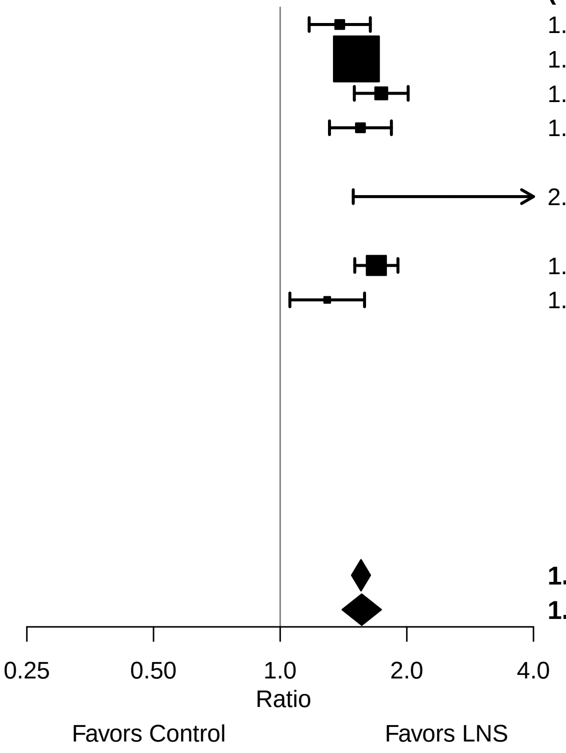

### Supplemental figure 3G: Iron deficiency (ferritin < 12 µg/L) prevalence ratio

| Country | Trial | LNS N | Control N | PR (95% CI) | Fixed W | Random W |
|---|---|---|---|---|---|---|
| Bangladesh | JiVitA−4 (35) | 455 | 144 | 0.36 (0.21, 0.59) | 0.06 | 0.11 |
| Bangladesh | RDNS (36) | 550 | 272 | 0.34 (0.24, 0.48) | 0.14 | 0.16 |
| Bangladesh | WASH−B (37) | 212 | 178 | 0.24 (0.15, 0.39) | 0.07 | 0.11 |
| Burkina Faso | iLiNS−Zinc (38) | 315 | 96 | 0.61 (0.49, 0.78) | 0.31 | 0.20 |
| Burkina Faso | PROMIS CS (39) | | | | | |
| Ghana | GHANA (40) | 83 | 82 | 0.49 (0.33, 0.73) | 0.11 | 0.14 |
| Ghana | iLiNS−DYADG (41) | | | | | |
| Kenya | WASH−B (42) | 298 | 259 | 0.41 (0.32, 0.53) | 0.27 | 0.19 |
| Madagascar | MAHAY (43) | 83 | 51 | 0.38 (0.21, 0.67) | 0.05 | 0.09 |
| Malawi | iLiNS−DYADM (44) | | | | | |
| Malawi | iLiNS−DOSE (45) | | | | | |
| Mali | PROMIS CS (46) | | | | | |
| Zimbabwe | SHINE (HIV−) (47) | | | | | |
| Zimbabwe | SHINE (HIV+) (48) | | | | | |
| | **I² = 0.66, Tau² = 0.05** | **1996** | **1082** | | | |
| | **Fixed** | | | **0.44 (0.39, 0.50)** | | |
| | **Random** | | | **0.41 (0.33, 0.51)** | | |

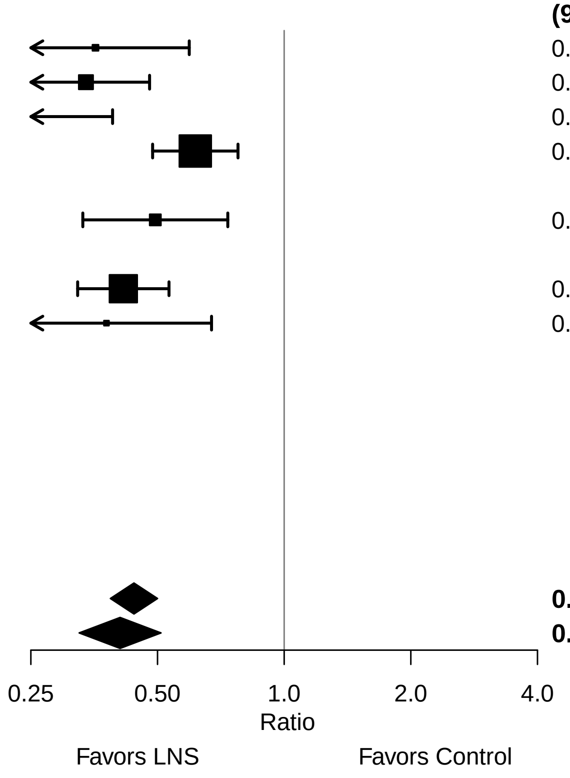

### Supplemental figure 3H: Iron deficiency (ferritin < 12 µg/L) prevalence difference

| Country | Trial | LNS N | Control N | PD (95% CI) | Fixed W | Random W |
|---|---|---|---|---|---|---|
| Bangladesh | JiVitA−4 (35) | 455 | 144 | −0.14 (−0.22, −0.07) | 0.19 | 0.17 |
| Bangladesh | RDNS (36) | 550 | 272 | −0.15 (−0.20, −0.09) | 0.33 | 0.19 |
| Bangladesh | WASH−B (37) | 212 | 178 | −0.27 (−0.35, −0.19) | 0.15 | 0.16 |
| Burkina Faso | iLiNS−Zinc (38) | 315 | 96 | −0.23 (−0.33, −0.12) | 0.08 | 0.13 |
| Burkina Faso | PROMIS CS (39) | | | | | |
| Ghana | GHANA (40) | 83 | 82 | −0.28 (−0.43, −0.14) | 0.05 | 0.10 |
| Ghana | iLiNS−DYADG (41) | | | | | |
| Kenya | WASH−B (42) | 298 | 259 | −0.35 (−0.42, −0.27) | 0.16 | 0.16 |
| Madagascar | MAHAY (43) | 83 | 51 | −0.32 (−0.48, −0.15) | 0.04 | 0.09 |
| Malawi | iLiNS−DYADM (44) | | | | | |
| Malawi | iLiNS−DOSE (45) | | | | | |
| Mali | PROMIS CS (46) | | | | | |
| Zimbabwe | SHINE (HIV−) (47) | | | | | |
| Zimbabwe | SHINE (HIV+) (48) | | | | | |
| | **I² = 0.76, Tau² = 0.00** | **1996** | **1082** | | | |
| | **Fixed** | | | **−0.22 (−0.25, −0.18)** | | |
| | **Random** | | | **−0.24 (−0.30, −0.18)** | | |

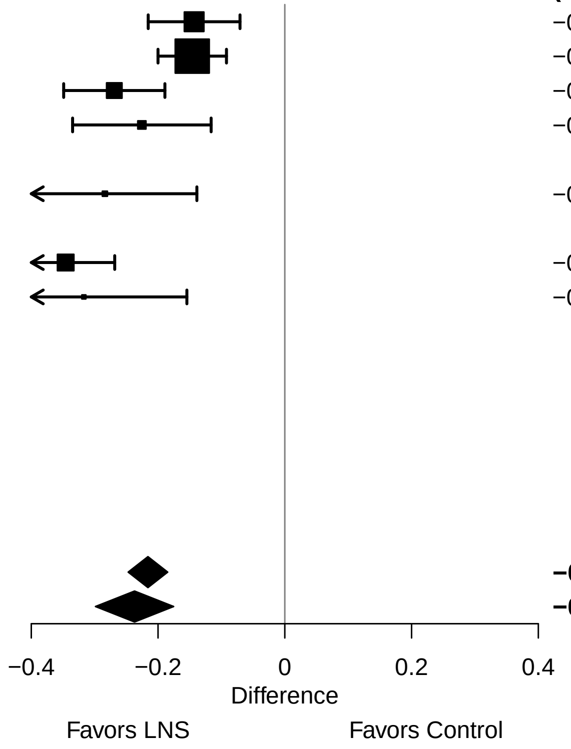

#### Supplemental figure 3I: Iron deficiency anemia prevalence ratio

| Country | Trial | LNS N | Control N | PR (95% CI) | Fixed W | Random W |
|---|---|---|---|---|---|---|
| Bangladesh | JiVitA−4 (35) | 454 | 144 | 0.12 (0.04, 0.34) | 0.04 | 0.10 |
| Bangladesh | RDNS (36) | 549 | 272 | 0.25 (0.15, 0.41) | 0.15 | 0.20 |
| Bangladesh | WASH−B (37) | | | | | |
| Burkina Faso | iLiNS−Zinc (38) | 327 | 101 | 0.51 (0.39, 0.67) | 0.54 | 0.26 |
| Burkina Faso | PROMIS CS (39) | | | | | |
| Ghana | GHANA (40) | 83 | 82 | 0.17 (0.07, 0.42) | 0.05 | 0.11 |
| Ghana | iLiNS−DYADG (41) | | | | | |
| Kenya | WASH−B (42) | 298 | 258 | 0.28 (0.18, 0.45) | 0.18 | 0.21 |
| Madagascar | MAHAY (43) | 83 | 51 | 0.31 (0.13, 0.71) | 0.05 | 0.12 |
| Malawi | iLiNS−DYADM (44) | | | | | |
| Malawi | iLiNS−DOSE (45) | | | | | |
| Mali | PROMIS CS (46) | | | | | |
| Zimbabwe | SHINE (HIV−) (47) | | | | | |
| Zimbabwe | SHINE (HIV+) (48) | | | | | |
| | **$I^2$ = 0.71, $Tau^2$ = 0.13** | **1794** | **908** | | | |
| | **Fixed** | | | **0.36 (0.30, 0.44)** | | |
| | **Random** | | | **0.28 (0.19, 0.42)** | | |

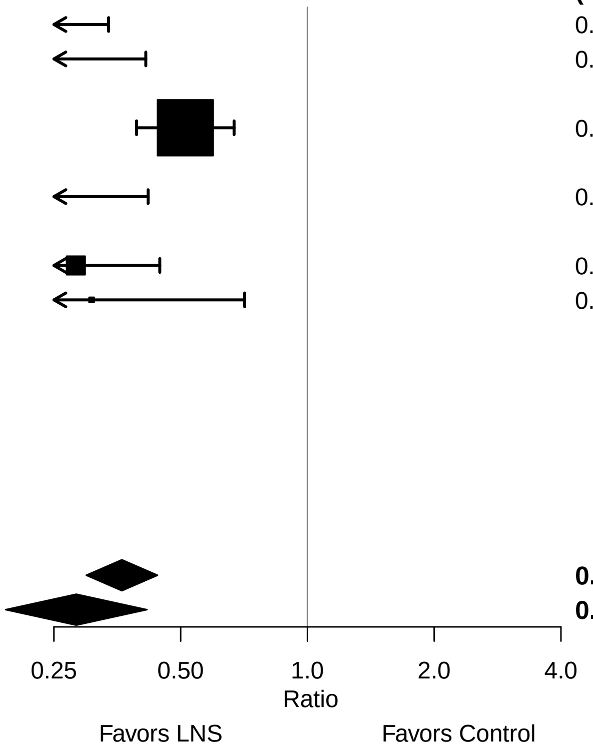

#### Supplemental figure 3J: Iron deficiency anemia prevalence difference

| Country | Trial | LNS N | Control N | PD (95% CI) | Fixed W | Random W |
|---|---|---|---|---|---|---|
| Bangladesh | JiVitA−4 (35) | 454 | 144 | −0.08 (−0.13, −0.03) | 0.32 | 0.20 |
| Bangladesh | RDNS (36) | 549 | 272 | −0.11 (−0.16, −0.07) | 0.36 | 0.20 |
| Bangladesh | WASH−B (37) | | | | | |
| Burkina Faso | iLiNS−Zinc (38) | 327 | 101 | −0.27 (−0.37, −0.16) | 0.06 | 0.15 |
| Burkina Faso | PROMIS CS (39) | | | | | |
| Ghana | GHANA (40) | 83 | 82 | −0.29 (−0.41, −0.18) | 0.05 | 0.14 |
| Ghana | iLiNS−DYADG (41) | | | | | |
| Kenya | WASH−B (42) | 298 | 258 | −0.24 (−0.31, −0.17) | 0.17 | 0.19 |
| Madagascar | MAHAY (43) | 83 | 51 | −0.19 (−0.33, −0.05) | 0.04 | 0.12 |
| Malawi | iLiNS−DYADM (44) | | | | | |
| Malawi | iLiNS−DOSE (45) | | | | | |
| Mali | PROMIS CS (46) | | | | | |
| Zimbabwe | SHINE (HIV−) (47) | | | | | |
| Zimbabwe | SHINE (HIV+) (48) | | | | | |
| | **$I^2$ = 0.83, $Tau^2$ = 0.01** | **1794** | **908** | | | |
| | **Fixed** | | | **−0.14 (−0.17, −0.12)** | | |
| | **Random** | | | **−0.19 (−0.26, −0.12)** | | |

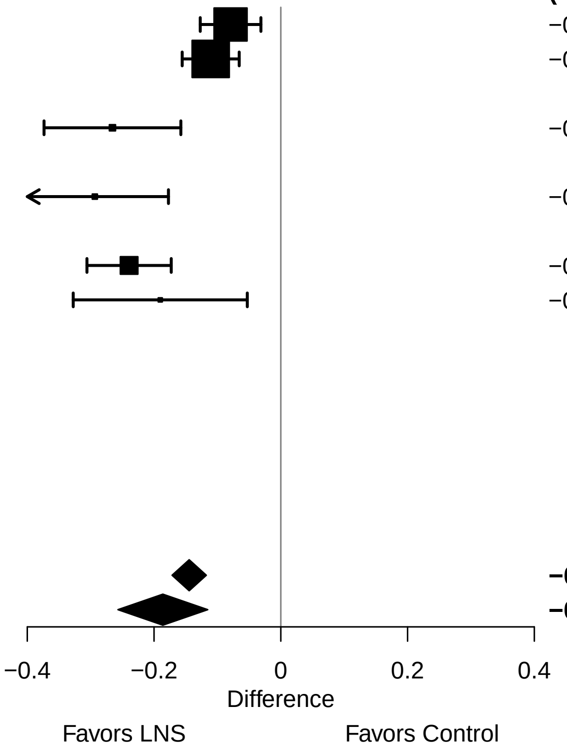

### Supplemental figure 3K: Geometric mean ratio of soluble transferrin receptor concentration

| Country | Trial | LNS N | Control N | GMR (95% CI) | Fixed W | Random W |
|---|---|---|---|---|---|---|
| Bangladesh | JiVitA−4 (35) | | | | | |
| Bangladesh | RDNS (36) | 550 | 272 | 0.87 (0.83, 0.92) | 0.35 | 0.26 |
| Bangladesh | WASH−B (37) | 212 | 178 | 0.82 (0.76, 0.87) | 0.23 | 0.22 |
| Burkina Faso | iLiNS−Zinc (38) | 315 | 96 | 0.76 (0.68, 0.84) | 0.09 | 0.13 |
| Burkina Faso | PROMIS CS (39) | | | | | |
| Ghana | GHANA (40) | 84 | 82 | 0.80 (0.72, 0.89) | 0.09 | 0.12 |
| Ghana | iLiNS−DYADG (41) | | | | | |
| Kenya | WASH−B (42) | 298 | 259 | 0.79 (0.74, 0.85) | 0.20 | 0.21 |
| Madagascar | MAHAY (43) | 83 | 51 | 0.90 (0.77, 1.05) | 0.04 | 0.07 |
| Malawi | iLiNS−DYADM (44) | | | | | |
| Malawi | iLiNS−DOSE (45) | | | | | |
| Mali | PROMIS CS (46) | | | | | |
| Zimbabwe | SHINE (HIV−) (47) | | | | | |
| Zimbabwe | SHINE (HIV+) (48) | | | | | |
| | **I² = 0.46, Tau² = 0.00** | **1542** | **938** | | | |
| | **Fixed** | | | **0.83 (0.80, 0.85)** | | |
| | **Random** | | | **0.82 (0.78, 0.86)** | | |

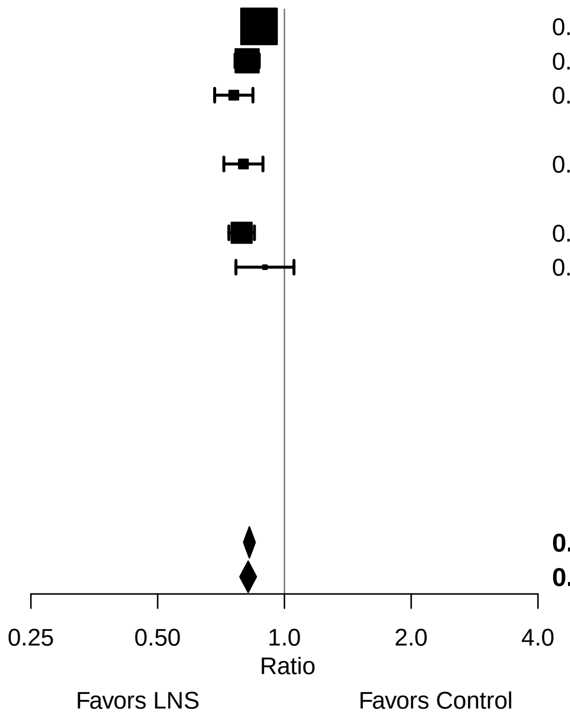

#### Supplemental figure 3L: Elevated soluble transferrin receptor prevalence ratio

| Country | Trial | LNS N | Control N | PR (95% CI) | Fixed W | Random W |
|---|---|---|---|---|---|---|
| Bangladesh | JiVitA−4 (35) | | | | | |
| Bangladesh | RDNS (36) | 550 | 272 | 0.70 (0.59, 0.84) | 0.24 | 0.19 |
| Bangladesh | WASH−B (37) | 212 | 178 | 0.31 (0.21, 0.47) | 0.05 | 0.14 |
| Burkina Faso | iLiNS−Zinc (38) | 315 | 96 | 0.51 (0.38, 0.68) | 0.09 | 0.16 |
| Burkina Faso | PROMIS CS (39) | | | | | |
| Ghana | GHANA (40) | 84 | 82 | 0.57 (0.41, 0.80) | 0.07 | 0.15 |
| Ghana | iLiNS−DYADG (41) | | | | | |
| Kenya | WASH−B (42) | 298 | 259 | 0.67 (0.59, 0.77) | 0.46 | 0.20 |
| Madagascar | MAHAY (43) | 83 | 51 | 0.82 (0.60, 1.12) | 0.08 | 0.16 |
| Malawi | iLiNS−DYADM (44) | | | | | |
| Malawi | iLiNS−DOSE (45) | | | | | |
| Mali | PROMIS CS (46) | | | | | |
| Zimbabwe | SHINE (HIV−) (47) | | | | | |
| Zimbabwe | SHINE (HIV+) (48) | | | | | |
| | **I² = 0.73, Tau² = 0.08** | **1542** | **938** | | | |
| | **Fixed** | | | **0.64 (0.59, 0.70)** | | |
| | **Random** | | | **0.59 (0.46, 0.76)** | | |

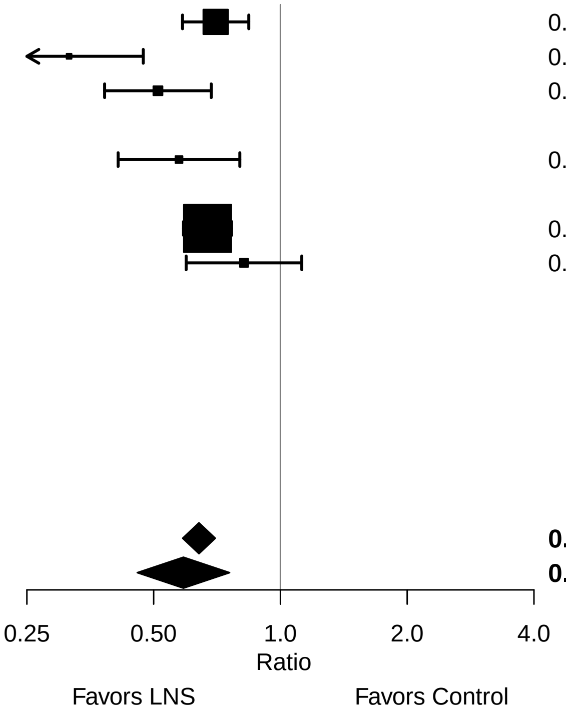

#### Supplemental figure 3M: Elevated soluble transferrin receptor prevalence difference

| Country | Trial | LNS N | Control N | PD (95% CI) | Fixed W | Random W |
|---|---|---|---|---|---|---|
| Bangladesh | JiVitA−4 (35) | | | | | |
| Bangladesh | RDNS (36) | 550 | 272 | −0.15 (−0.22, −0.08) | 0.28 | 0.24 |
| Bangladesh | WASH−B (37) | 212 | 178 | −0.22 (−0.29, −0.14) | 0.23 | 0.22 |
| Burkina Faso | iLiNS−Zinc (38) | 315 | 96 | −0.30 (−0.40, −0.19) | 0.13 | 0.16 |
| Burkina Faso | PROMIS CS (39) | | | | | |
| Ghana | GHANA (40) | 84 | 82 | −0.26 (−0.41, −0.12) | 0.07 | 0.09 |
| Ghana | iLiNS−DYADG (41) | | | | | |
| Kenya | WASH−B (42) | 298 | 259 | −0.24 (−0.32, −0.16) | 0.24 | 0.22 |
| Madagascar | MAHAY (43) | 83 | 51 | −0.11 (−0.28, 0.07) | 0.05 | 0.07 |
| Malawi | iLiNS−DYADM (44) | | | | | |
| Malawi | iLiNS−DOSE (45) | | | | | |
| Mali | PROMIS CS (46) | | | | | |
| Zimbabwe | SHINE (HIV−) (47) | | | | | |
| Zimbabwe | SHINE (HIV+) (48) | | | | | |
| | **I² = 0.37, Tau² = 0.00** | **1542** | **938** | | | |
| | **Fixed** | | | **−0.21 (−0.25, −0.17)** | | |
| | **Random** | | | **−0.21 (−0.26, −0.16)** | | |

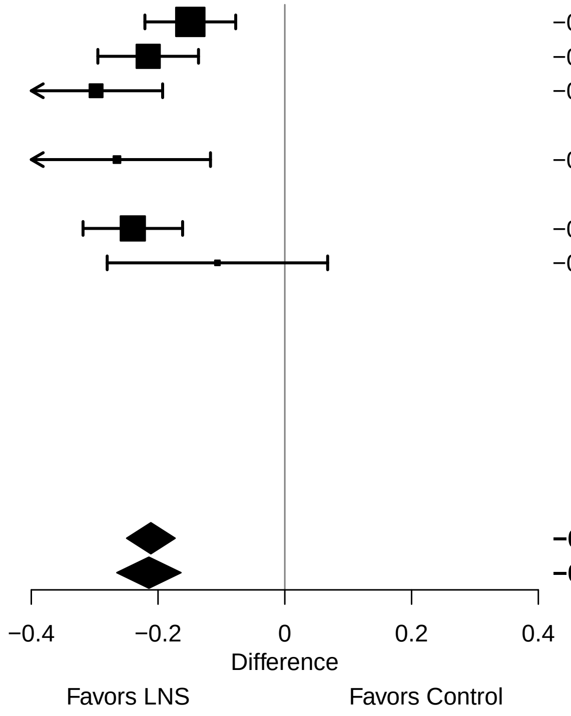

### Supplemental figure 3N: Geometric mean ratio of zinc protoporphyrin concentration

| Country | Trial | LNS N | Control N | GMR (95% CI) | Fixed W | Random W |
|---|---|---|---|---|---|---|
| Bangladesh | JiVitA−4 (35) | | | | | |
| Bangladesh | RDNS (36) | | | | | |
| Bangladesh | WASH−B (37) | | | | | |
| Burkina Faso | iLiNS−Zinc (38) | 312 | 95 | 0.64 (0.57, 0.74) | 0.21 | 0.24 |
| Burkina Faso | PROMIS CS (39) | | | | | |
| Ghana | GHANA (40) | | | | | |
| Ghana | iLiNS−DYADG (41) | 100 | 202 | 0.86 (0.76, 0.98) | 0.23 | 0.25 |
| Kenya | WASH−B (42) | | | | | |
| Madagascar | MAHAY (43) | | | | | |
| Malawi | iLiNS−DYADM (44) | 190 | 375 | 0.87 (0.78, 0.96) | 0.37 | 0.27 |
| Malawi | iLiNS−DOSE (45) | 181 | 87 | 0.78 (0.68, 0.89) | 0.19 | 0.24 |
| Mali | PROMIS CS (46) | | | | | |
| Zimbabwe | SHINE (HIV−) (47) | | | | | |
| Zimbabwe | SHINE (HIV+) (48) | | | | | |
| | **$I^2$ = 0.79, $Tau^2$ = 0.02** | **783** | **759** | | | |
| | **Fixed** | | | **0.80 (0.75, 0.85)** | | |
| | **Random** | | | **0.79 (0.69, 0.90)** | | |

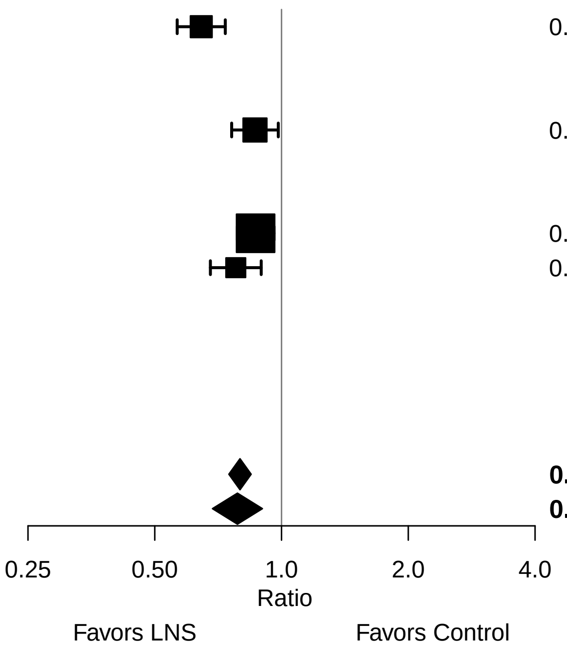

### Supplemental figure 3O: Elevated zinc protoporphyrin prevalence ratio

| Country | Trial | LNS N | Control N | PR (95% CI) | Fixed W | Random W |
|---|---|---|---|---|---|---|
| Bangladesh | JiVitA−4 (35) | | | | | |
| Bangladesh | RDNS (36) | | | | | |
| Bangladesh | WASH−B (37) | | | | | |
| Burkina Faso | iLiNS−Zinc (38) | 312 | 95 | 0.71 (0.60, 0.85) | 0.53 | 0.53 |
| Burkina Faso | PROMIS CS (39) | | | | | |
| Ghana | GHANA (40) | | | | | |
| Ghana | iLiNS−DYADG (41) | 100 | 202 | 0.74 (0.46, 1.20) | 0.07 | 0.07 |
| Kenya | WASH−B (42) | | | | | |
| Madagascar | MAHAY (43) | | | | | |
| Malawi | iLiNS−DYADM (44) | 190 | 375 | 0.70 (0.53, 0.94) | 0.20 | 0.20 |
| Malawi | iLiNS−DOSE (45) | 181 | 87 | 0.62 (0.46, 0.85) | 0.19 | 0.19 |
| Mali | PROMIS CS (46) | | | | | |
| Zimbabwe | SHINE (HIV−) (47) | | | | | |
| Zimbabwe | SHINE (HIV+) (48) | | | | | |
| | **$I^2$ = 0.00, $Tau^2$ = 0.00** | **783** | **759** | | | |
| | **Fixed** | | | **0.70 (0.61, 0.79)** | | |
| | **Random** | | | **0.70 (0.61, 0.79)** | | |

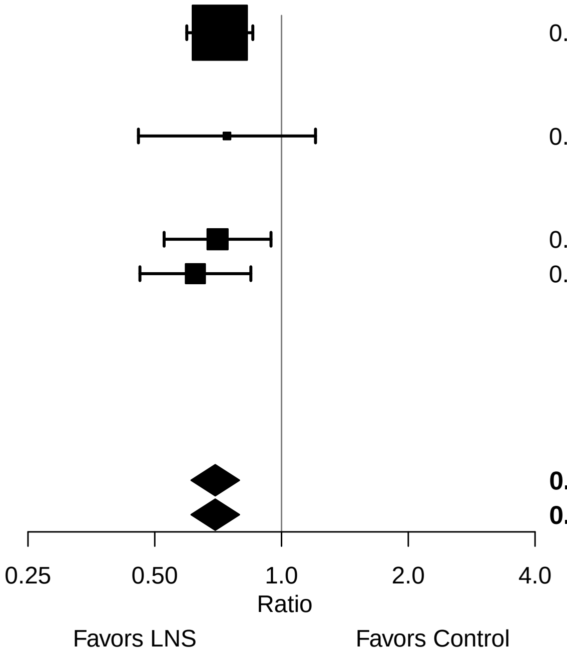

### Supplemental figure 3P: Elevated zinc protoporphyrin prevalence difference

| Country | Trial | LNS N | Control N | PD (95% CI) | Fixed W | Random W |
|---|---|---|---|---|---|---|
| Bangladesh | JiVitA−4 (35) | | | | | |
| Bangladesh | RDNS (36) | | | | | |
| Bangladesh | WASH−B (37) | | | | | |
| Burkina Faso | iLiNS−Zinc (38) | 312 | 95 | −0.21 (−0.31, −0.11) | 0.22 | 0.24 |
| Burkina Faso | PROMIS CS (39) | | | | | |
| Ghana | GHANA (40) | | | | | |
| Ghana | iLiNS−DYADG (41) | 100 | 202 | −0.06 (−0.16, 0.03) | 0.24 | 0.25 |
| Kenya | WASH−B (42) | | | | | |
| Madagascar | MAHAY (43) | | | | | |
| Malawi | iLiNS−DYADM (44) | 190 | 375 | −0.10 (−0.18, −0.02) | 0.37 | 0.31 |
| Malawi | iLiNS−DOSE (45) | 181 | 87 | −0.17 (−0.28, −0.05) | 0.18 | 0.21 |
| Mali | PROMIS CS (46) | | | | | |
| Zimbabwe | SHINE (HIV−) (47) | | | | | |
| Zimbabwe | SHINE (HIV+) (48) | | | | | |
| | **I² = 0.46, Tau² = 0.00** | **783** | **759** | | | |
| | **Fixed** | | | **−0.13 (−0.17, −0.08)** | | |
| | **Random** | | | **−0.13 (−0.20, −0.07)** | | |

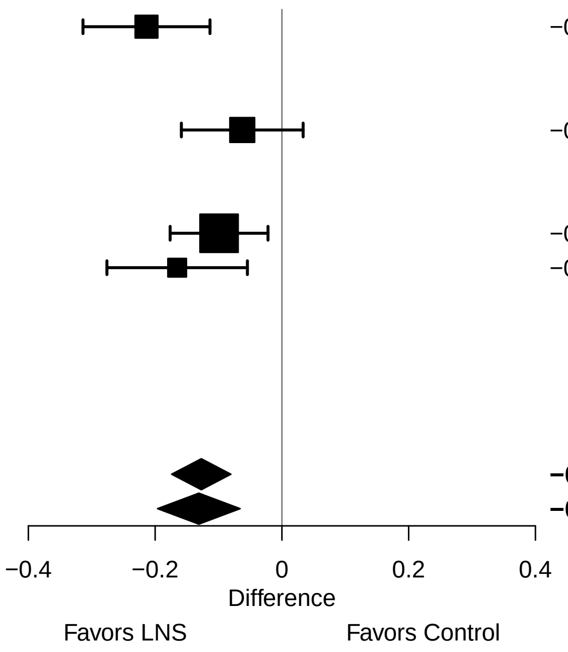

#### Supplemental figure 3Q: Geometric mean ratio of plasma zinc concentration

| Country | Trial | LNS N | Control N | GMR (95% CI) | Fixed W | Random W |
|---|---|---|---|---|---|---|
| Bangladesh | JiVitA−4 (35) | 452 | 144 | 0.96 (0.91, 1.02) | 0.23 | 0.23 |
| Bangladesh | RDNS (36) | | | | | |
| Bangladesh | WASH−B (37) | | | | | |
| Burkina Faso | iLiNS−Zinc (38) | 314 | 96 | 1.01 (0.97, 1.04) | 0.62 | 0.62 |
| Burkina Faso | PROMIS CS (39) | | | | | |
| Ghana | GHANA (40) | 58 | 69 | 1.00 (0.94, 1.08) | 0.15 | 0.15 |
| Ghana | iLiNS−DYADG (41) | | | | | |
| Kenya | WASH−B (42) | | | | | |
| Madagascar | MAHAY (43) | | | | | |
| Malawi | iLiNS−DYADM (44) | | | | | |
| Malawi | iLiNS−DOSE (45) | | | | | |
| Mali | PROMIS CS (46) | | | | | |
| Zimbabwe | SHINE (HIV−) (47) | | | | | |
| Zimbabwe | SHINE (HIV+) (48) | | | | | |
| | **$I^2$ = 0.00, $Tau^2$ = 0.00** | **824** | **309** | | | |
| | **Fixed** | | | **1.00 (0.97, 1.02)** | | |
| | **Random** | | | **1.00 (0.97, 1.02)** | | |

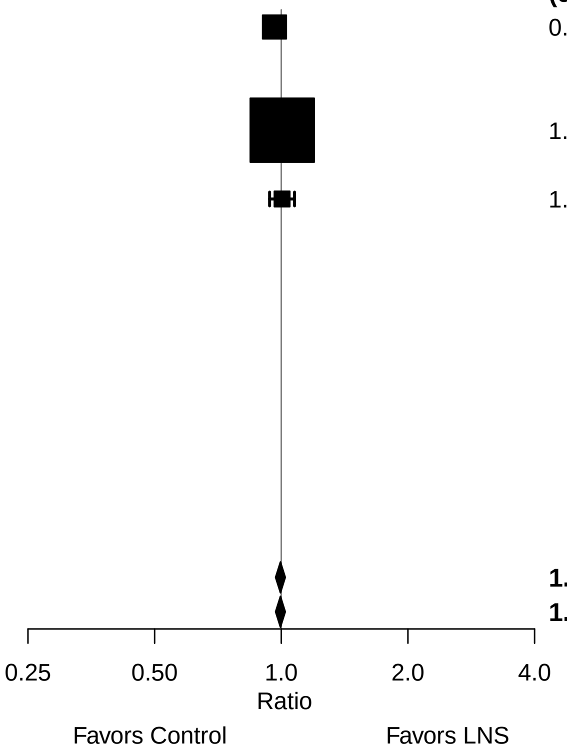

#### Supplemental figure 3R: Geometric mean ratio of retinol concentration

| Country | Trial | LNS N | Control N | GMR (95% CI) | Fixed W | Random W |
|---|---|---|---|---|---|---|
| Bangladesh | JiVitA−4 (35) | 434 | 139 | 1.08 (1.02, 1.16) | 0.35 | 0.35 |
| Bangladesh | RDNS (36) | | | | | |
| Bangladesh | WASH−B (37) | | | | | |
| Burkina Faso | iLiNS−Zinc (38) | | | | | |
| Burkina Faso | PROMIS CS (39) | | | | | |
| Ghana | GHANA (40) | 63 | 74 | 1.07 (0.93, 1.23) | 0.08 | 0.08 |
| Ghana | iLiNS−DYADG (41) | 96 | 196 | 1.01 (0.95, 1.07) | 0.36 | 0.35 |
| Kenya | WASH−B (42) | | | | | |
| Madagascar | MAHAY (43) | | | | | |
| Malawi | iLiNS−DYADM (44) | 71 | 163 | 1.00 (0.92, 1.09) | 0.21 | 0.21 |
| Malawi | iLiNS−DOSE (45) | | | | | |
| Mali | PROMIS CS (46) | | | | | |
| Zimbabwe | SHINE (HIV−) (47) | | | | | |
| Zimbabwe | SHINE (HIV+) (48) | | | | | |
| | **$I^2$ = 0.08, $Tau^2$ = 0.00** | **664** | **572** | | | |
| | **Fixed** | | | **1.04 (1.00, 1.08)** | | |
| | **Random** | | | **1.04 (1.00, 1.08)** | | |

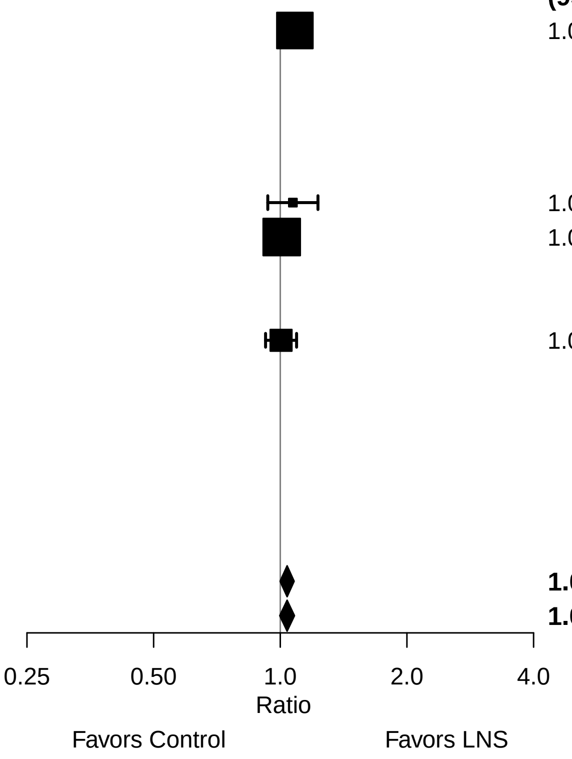

#### Supplemental figure 3S: Low vitamin A (retinol < 0.70 µmol/L) prevalence ratio

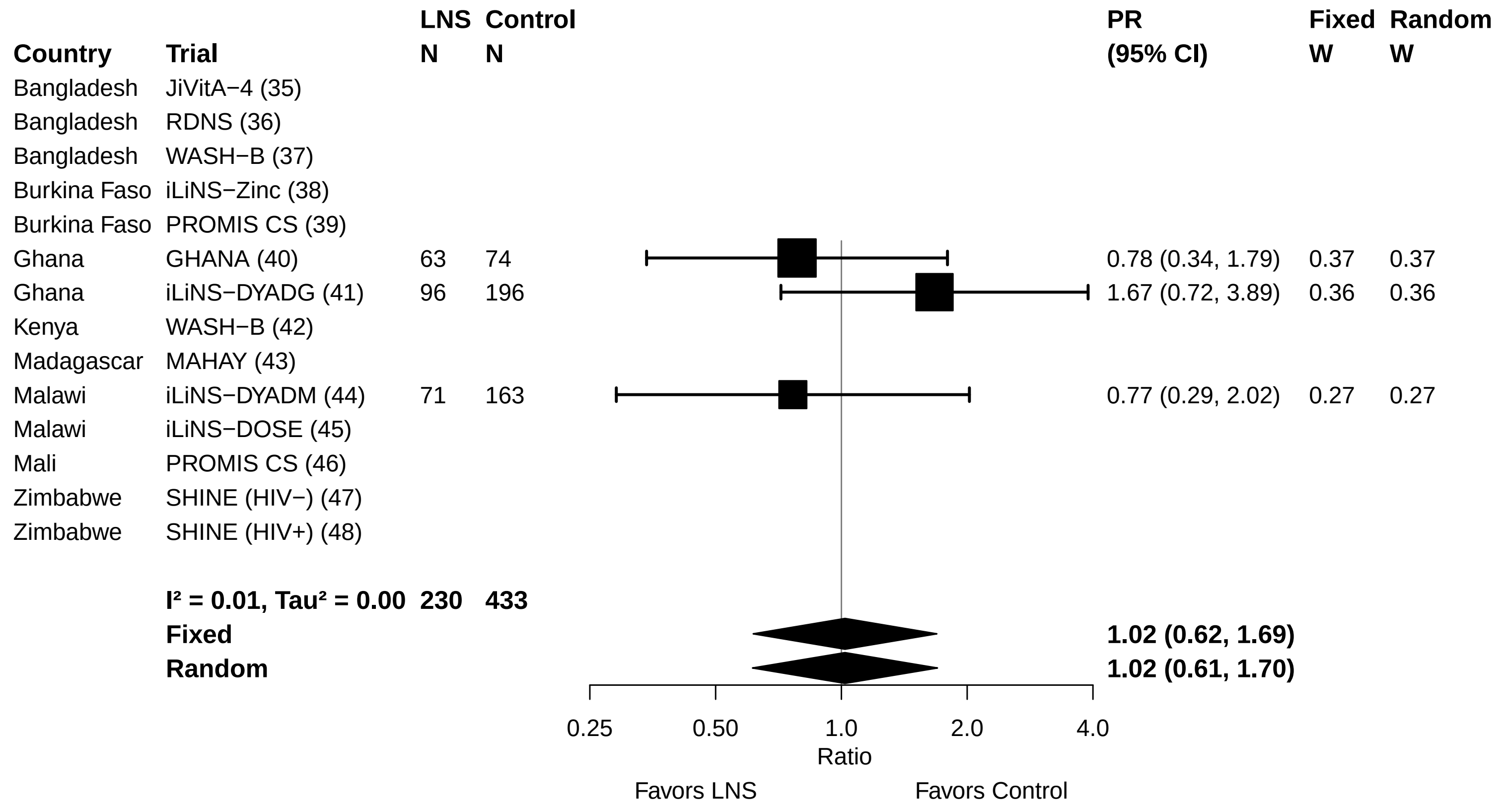

### Supplemental figure 3T: Low vitamin A (retinol < 0.70 µmol/L) prevalence difference

| Country | Trial | LNS N | Control N | PD (95% CI) | Fixed W | Random W |
|---|---|---|---|---|---|---|
| Bangladesh | JiVitA−4 (35) | | | | | |
| Bangladesh | RDNS (36) | | | | | |
| Bangladesh | WASH−B (37) | | | | | |
| Burkina Faso | iLiNS−Zinc (38) | | | | | |
| Burkina Faso | PROMIS CS (39) | | | | | |
| Ghana | GHANA (40) | 63 | 74 | −0.04 (−0.15, 0.08) | 0.15 | 0.15 |
| Ghana | iLiNS−DYADG (41) | 96 | 196 | 0.04 (−0.03, 0.10) | 0.47 | 0.47 |
| Kenya | WASH−B (42) | | | | | |
| Madagascar | MAHAY (43) | | | | | |
| Malawi | iLiNS−DYADM (44) | 71 | 163 | −0.02 (−0.10, 0.05) | 0.38 | 0.38 |
| Malawi | iLiNS−DOSE (45) | | | | | |
| Mali | PROMIS CS (46) | | | | | |
| Zimbabwe | SHINE (HIV−) (47) | | | | | |
| Zimbabwe | SHINE (HIV+) (48) | | | | | |
| | **I² = 0.00, Tau² = 0.00** | **230** | **433** | | | |
| | **Fixed** | | | **0.00 (−0.04, 0.05)** | | |
| | **Random** | | | **0.00 (−0.04, 0.05)** | | |

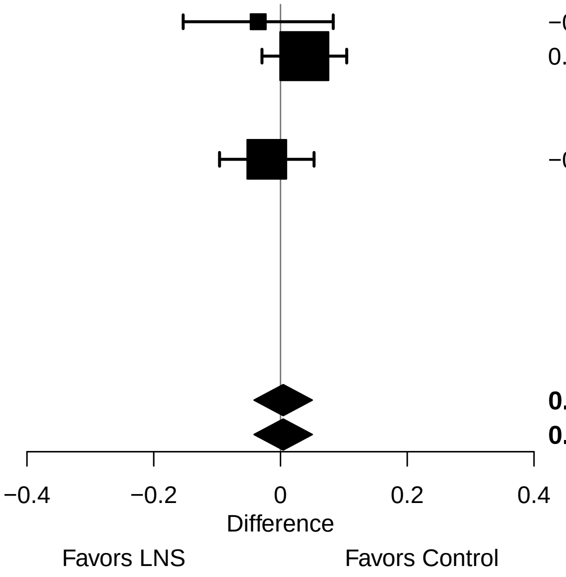

#### Supplemental figure 3U: Marginal vitamin A (retinol < 1.05 µmol/L) prevalence ratio

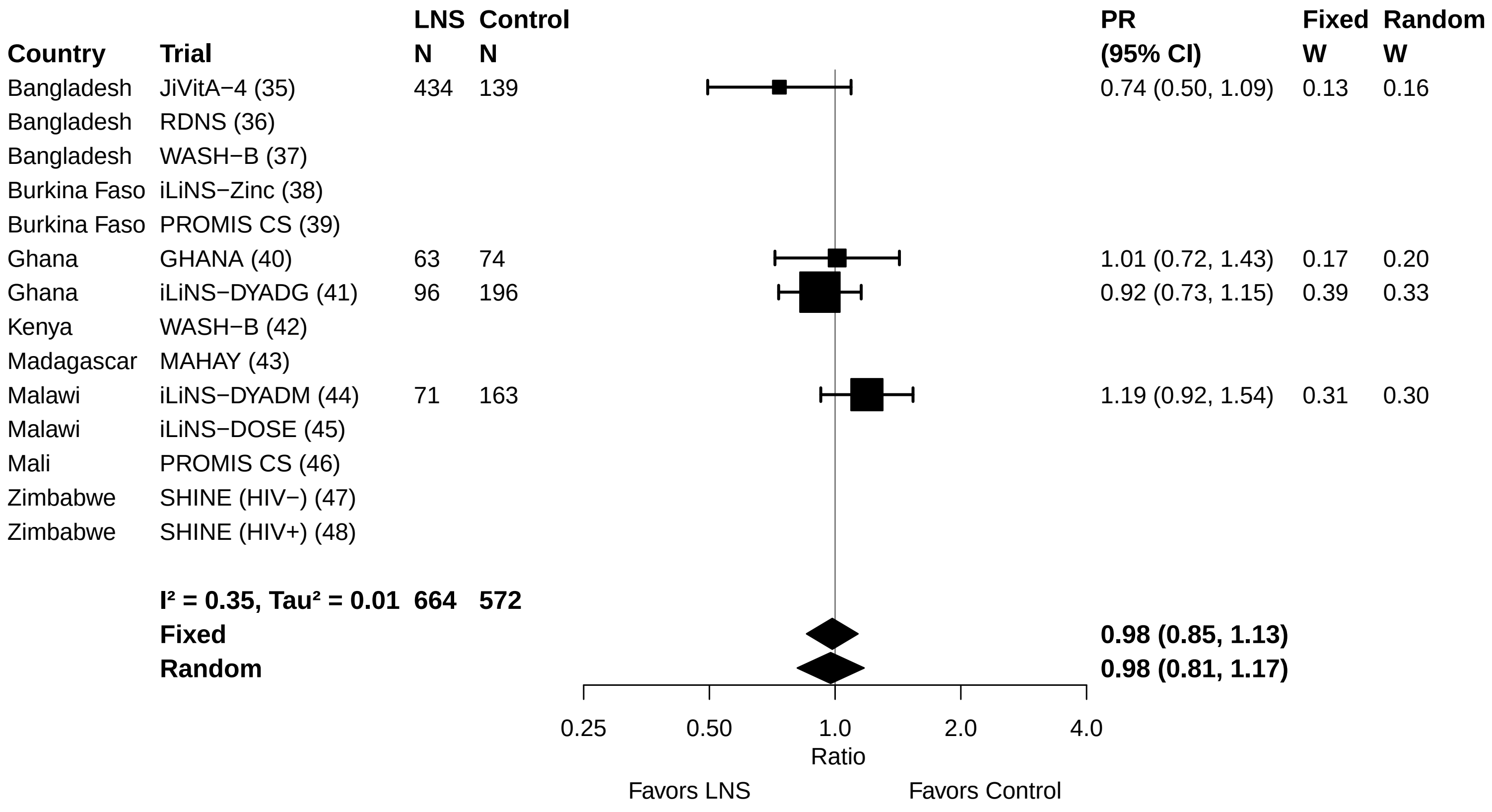

#### Supplemental figure 3V: Marginal vitamin A (retinol < 1.05 µmol/L) prevalence difference

| Country | Trial | LNS N | Control N | PD (95% CI) | Fixed W | Random W |
|---|---|---|---|---|---|---|
| Bangladesh | JiVitA−4 (35) | 434 | 139 | −0.05 (−0.13, 0.02) | 0.54 | 0.49 |
| Bangladesh | RDNS (36) | | | | | |
| Bangladesh | WASH−B (37) | | | | | |
| Burkina Faso | iLiNS−Zinc (38) | | | | | |
| Burkina Faso | PROMIS CS (39) | | | | | |
| Ghana | GHANA (40) | 63 | 74 | 0.01 (−0.16, 0.17) | 0.10 | 0.12 |
| Ghana | iLiNS−DYADG (41) | 96 | 196 | −0.05 (−0.17, 0.08) | 0.20 | 0.22 |
| Kenya | WASH−B (42) | | | | | |
| Madagascar | MAHAY (43) | | | | | |
| Malawi | iLiNS−DYADM (44) | 71 | 163 | 0.09 (−0.05, 0.23) | 0.15 | 0.17 |
| Malawi | iLiNS−DOSE (45) | | | | | |
| Mali | PROMIS CS (46) | | | | | |
| Zimbabwe | SHINE (HIV−) (47) | | | | | |
| Zimbabwe | SHINE (HIV+) (48) | | | | | |
| | **I² = 0.14, Tau² = 0.00** | **664** | **572** | | | |
| | **Fixed** | | | **−0.02 (−0.08, 0.03)** | | |
| | **Random** | | | **−0.02 (−0.08, 0.04)** | | |

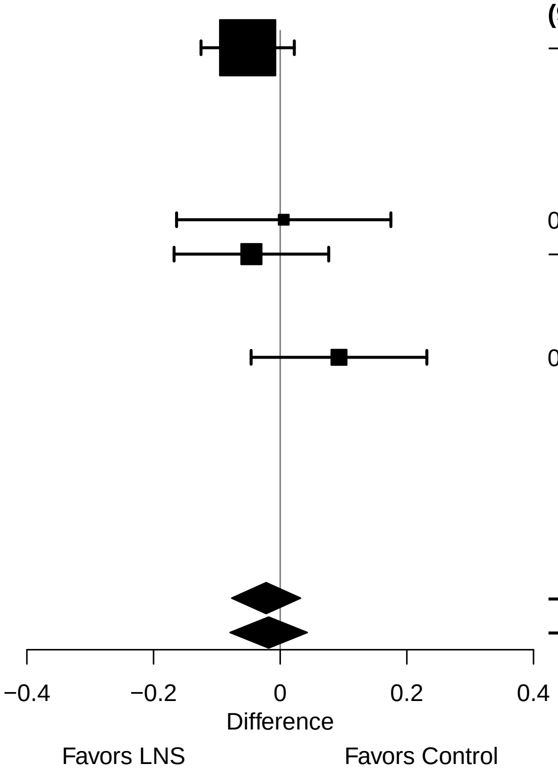

#### Supplemental figure 3W: Geometric mean ratio of retinol binding protein concentration

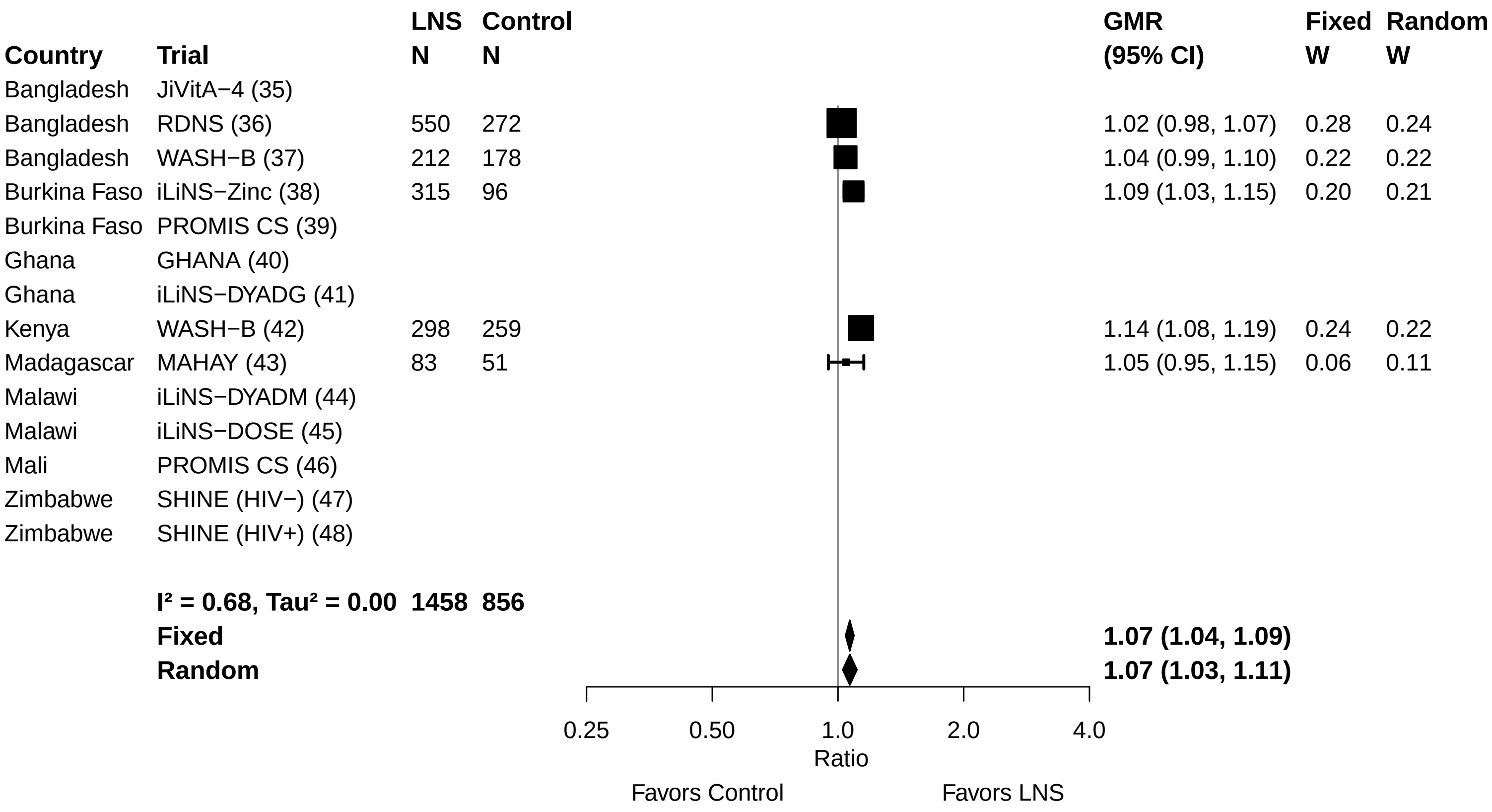

### Supplemental figure 3X: Low vitamin A status (RBP < 0.70 µmol/L) prevalence ratio

| Country | Trial | LNS N | Control N | PR (95% CI) | Fixed W | Random W |
|---|---|---|---|---|---|---|
| Bangladesh | JiVitA−4 (35) | | | | | |
| Bangladesh | RDNS (36) | 550 | 272 | 0.55 (0.23, 1.29) | 0.30 | 0.30 |
| Bangladesh | WASH−B (37) | | | | | |
| Burkina Faso | iLiNS−Zinc (38) | 315 | 96 | 0.47 (0.22, 1.04) | 0.36 | 0.36 |
| Burkina Faso | PROMIS CS (39) | | | | | |
| Ghana | GHANA (40) | | | | | |
| Ghana | iLiNS−DYADG (41) | | | | | |
| Kenya | WASH−B (42) | 298 | 259 | 0.33 (0.15, 0.73) | 0.34 | 0.34 |
| Madagascar | MAHAY (43) | | | | | |
| Malawi | iLiNS−DYADM (44) | | | | | |
| Malawi | iLiNS−DOSE (45) | | | | | |
| Mali | PROMIS CS (46) | | | | | |
| Zimbabwe | SHINE (HIV−) (47) | | | | | |
| Zimbabwe | SHINE (HIV+) (48) | | | | | |
| | **I² = 0.00, Tau² = 0.00** | **1163** | **627** | | | |
| | **Fixed** | | | **0.44 (0.27, 0.70)** | | |
| | **Random** | | | **0.44 (0.27, 0.70)** | | |

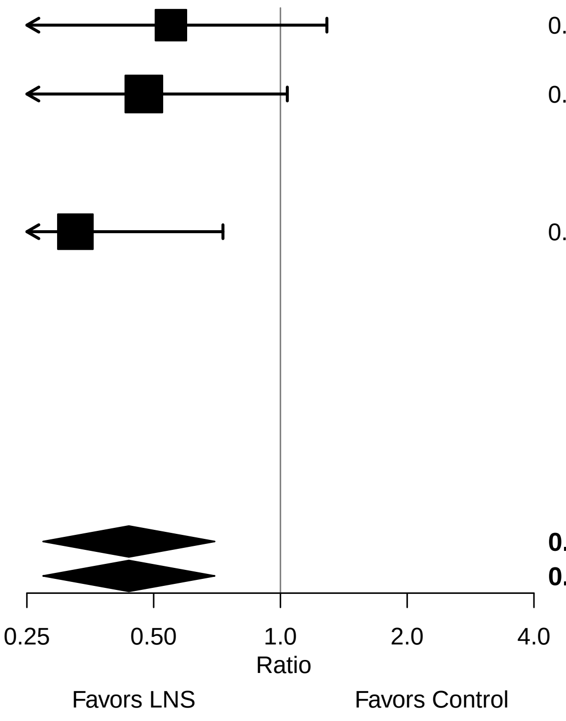

### Supplemental figure 3Y: Low vitamin A status (RBP < 0.70 μmol/L) prevalence difference

| Country | Trial | LNS N | Control N | PD (95% CI) | Fixed W | Random W |
|---|---|---|---|---|---|---|
| Bangladesh | JiVitA−4 (35) | | | | | |
| Bangladesh | RDNS (36) | 550 | 272 | −0.01 (−0.04, 0.01) | 0.65 | 0.50 |
| Bangladesh | WASH−B (37) | | | | | |
| Burkina Faso | iLiNS−Zinc (38) | 315 | 96 | −0.04 (−0.10, 0.02) | 0.11 | 0.19 |
| Burkina Faso | PROMIS CS (39) | | | | | |
| Ghana | GHANA (40) | | | | | |
| Ghana | iLiNS−DYADG (41) | | | | | |
| Kenya | WASH−B (42) | 298 | 259 | −0.06 (−0.10, −0.02) | 0.23 | 0.31 |
| Madagascar | MAHAY (43) | | | | | |
| Malawi | iLiNS−DYADM (44) | | | | | |
| Malawi | iLiNS−DOSE (45) | | | | | |
| Mali | PROMIS CS (46) | | | | | |
| Zimbabwe | SHINE (HIV−) (47) | | | | | |
| Zimbabwe | SHINE (HIV+) (48) | | | | | |
| | **I² = 0.52, Tau² = 0.00** | **1163** | **627** | | | |
| | **Fixed** | | | **−0.03 (−0.05, −0.01)** | | |
| | **Random** | | | **−0.04 (−0.06, −0.01)** | | |

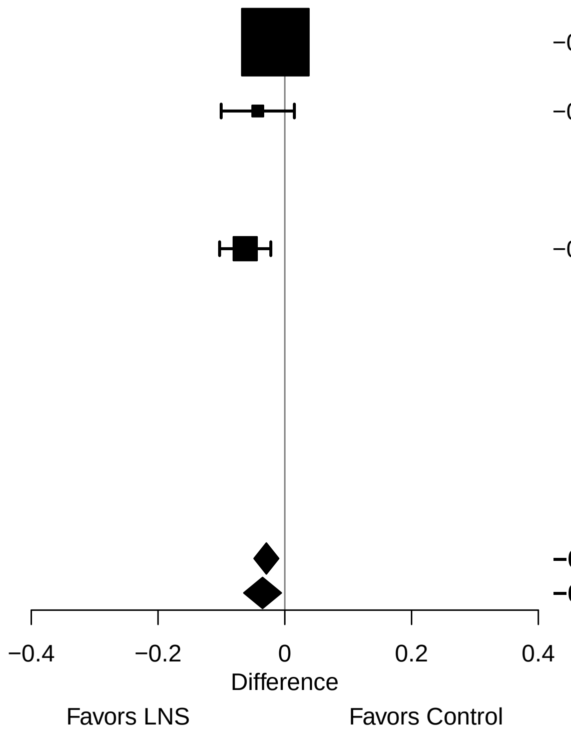

#### Supplemental figure 3Z: Marginal vitamin A status (RBP < 1.05 µmol/L) prevalence ratio

| Country | Trial | LNS N | Control N | PR (95% CI) | Fixed W | Random W |
|---|---|---|---|---|---|---|
| Bangladesh | JiVitA−4 (35) | | | | | |
| Bangladesh | RDNS (36) | 550 | 272 | 0.87 (0.68, 1.12) | 0.17 | 0.17 |
| Bangladesh | WASH−B (37) | 212 | 178 | 0.85 (0.65, 1.12) | 0.14 | 0.14 |
| Burkina Faso | iLiNS−Zinc (38) | 315 | 96 | 0.76 (0.63, 0.93) | 0.29 | 0.29 |
| Burkina Faso | PROMIS CS (39) | | | | | |
| Ghana | GHANA (40) | | | | | |
| Ghana | iLiNS−DYADG (41) | | | | | |
| Kenya | WASH−B (42) | 298 | 259 | 0.72 (0.61, 0.86) | 0.35 | 0.35 |
| Madagascar | MAHAY (43) | 83 | 51 | 0.83 (0.48, 1.42) | 0.04 | 0.04 |
| Malawi | iLiNS−DYADM (44) | | | | | |
| Malawi | iLiNS−DOSE (45) | | | | | |
| Mali | PROMIS CS (46) | | | | | |
| Zimbabwe | SHINE (HIV−) (47) | | | | | |
| Zimbabwe | SHINE (HIV+) (48) | | | | | |
| | **$I^2$ = 0.00, $Tau^2$ = 0.00** | **1458** | **856** | | | |
| | **Fixed** | | | **0.78 (0.70, 0.87)** | | |
| | **Random** | | | **0.78 (0.70, 0.87)** | | |

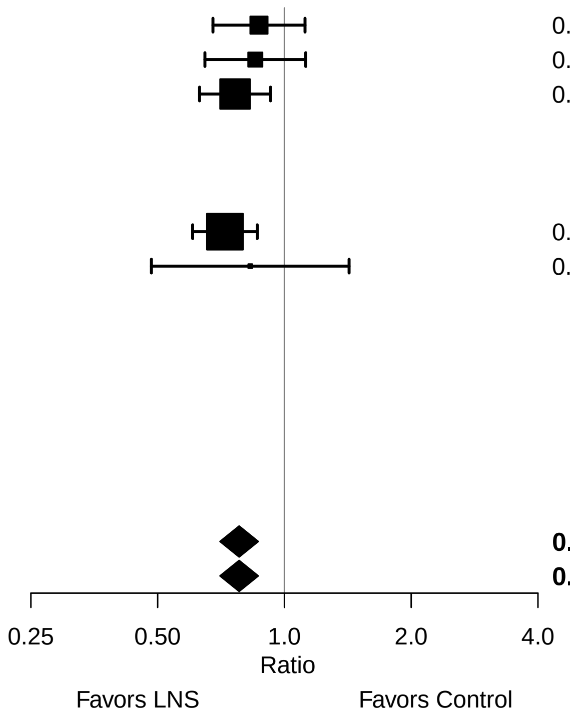

### Supplemental figure 3AA: Marginal vitamin A status (RBP < 1.05 µmol/L) prevalence difference

| Country | Trial | LNS N | Control N | PD (95% CI) | Fixed W | Random W |
|---|---|---|---|---|---|---|
| Bangladesh | JiVitA−4 (35) | | | | | |
| Bangladesh | RDNS (36) | 550 | 272 | −0.04 (−0.10, 0.03) | 0.38 | 0.32 |
| Bangladesh | WASH−B (37) | 212 | 178 | −0.05 (−0.15, 0.04) | 0.18 | 0.20 |
| Burkina Faso | iLiNS−Zinc (38) | 315 | 96 | −0.11 (−0.22, −0.01) | 0.14 | 0.16 |
| Burkina Faso | PROMIS CS (39) | | | | | |
| Ghana | GHANA (40) | | | | | |
| Ghana | iLiNS−DYADG (41) | | | | | |
| Kenya | WASH−B (42) | 298 | 259 | −0.17 (−0.25, −0.08) | 0.24 | 0.24 |
| Madagascar | MAHAY (43) | 83 | 51 | −0.07 (−0.24, 0.10) | 0.06 | 0.08 |
| Malawi | iLiNS−DYADM (44) | | | | | |
| Malawi | iLiNS−DOSE (45) | | | | | |
| Mali | PROMIS CS (46) | | | | | |
| Zimbabwe | SHINE (HIV−) (47) | | | | | |
| Zimbabwe | SHINE (HIV+) (48) | | | | | |
| | **I² = 0.38, Tau² = 0.00** | **1458** | **856** | | | |
| | **Fixed** | | | **−0.08 (−0.12, −0.04)** | | |
| | **Random** | | | **−0.09 (−0.14, −0.04)** | | |

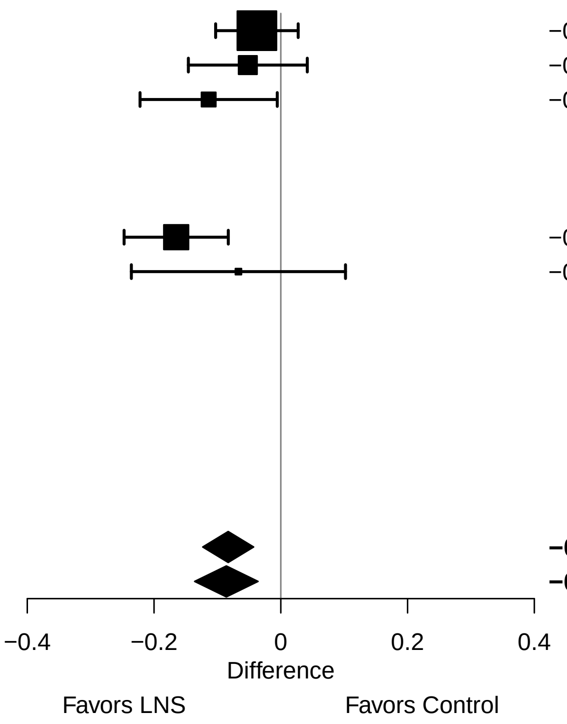

### Supplemental figure 4: Forest plots for effects of SQ-LNS on biochemical outcomes stratified by study-level effect modifiers

These figures are forest plots showing the study-level effect modification of intervention effects. Each figure shows the study-level estimates along with the corresponding pooled estimate grouped by study-level effect modifier category. For continuous outcomes the intervention effect is measured by the difference in mean of the LNS group minus control. For log transformed continuous outcomes, the intervention effect is measured by the ratio of geometric means, the effect estimate is the geometric mean in the LNS group divided by the geometric mean in the control group. For dichotomous outcomes analyzed via prevalence ratios, the effect estimate is the prevalence in the LNS group divided by the prevalence in the control group. For dichotomous outcomes analyzed via prevalence differences, the effect estimate is the prevalence in the LNS group minus the prevalence in the control group. The labels on the left y-axis correspond to trial level information. The values on the right indicate the study level effect estimate, confidence interval, and weighting for deriving the pooled estimates.

Due to the limited number of studies, we were able to examine only a few, if any, study-level effect modifiers for sTfR, ZPP, plasma zinc, retinol and RBP, and could not examine differences in the effect of SQ-LNS on ferritin by study-level anemia, malaria or inflammation prevalence.

RBP, retinol binding protein.

### Supplemental figure 4A: Mean difference in hemoglobin concentration

#### 4A1: Stratified by Geographic region

**Geographic region**
**(p−diff = 0.725)**

| Country | Trial | N | N | MD (95% CI) | W |
|---|---|---|---|---|---|
| **Geographic region − SEAR** | | | | | |
| Bangladesh | JiVitA−4 (35) | 457 | 146 | 2.21 (0.34, 4.07) | 0.34 |
| Bangladesh | RDNS (36) | 549 | 272 | 4.10 (1.86, 6.34) | 0.32 |
| Bangladesh | WASH−B (37) | 234 | 186 | 3.09 (1.22, 4.97) | 0.34 |
| **I² = 0.00, Tau² = 0.00** | | **1240** | **604** | **3.03 (1.89, 4.16)** | |
| **Geographic region − AFR** | | | | | |
| Burkina Faso | iLiNS−Zinc (38) | 1957 | 664 | 8.47 (5.47, 11.46) | 0.08 |
| Burkina Faso | PROMIS CS (39) | 574 | 581 | 1.77 (−0.11, 3.64) | 0.10 |
| Ghana | GHANA (40) | 98 | 96 | 8.84 (4.87, 12.81) | 0.06 |
| Ghana | iLiNS−DYADG (41) | 328 | 661 | 1.24 (−0.14, 2.61) | 0.11 |
| Kenya | WASH−B (42) | 350 | 300 | 3.94 (2.05, 5.84) | 0.10 |
| Madagascar | MAHAY (43) | 600 | 588 | 2.77 (−0.12, 5.67) | 0.08 |
| Malawi | iLiNS−DYADM (44) | 210 | 432 | 1.12 (−1.45, 3.69) | 0.09 |
| Malawi | iLiNS−DOSE (45) | 243 | 82 | 3.76 (0.02, 7.51) | 0.07 |
| Mali | PROMIS CS (46) | 953 | 970 | 6.25 (4.16, 8.34) | 0.10 |
| Zimbabwe | SHINE (HIV−) (47) | 1682 | 1594 | 2.13 (1.36, 2.91) | 0.12 |
| Zimbabwe | SHINE (HIV+) (48) | 306 | 285 | 2.78 (0.38, 5.19) | 0.09 |
| **I² = 0.78, Tau² = 5.36** | | **7301** | **6253** | **3.68 (2.12, 5.23)** | |

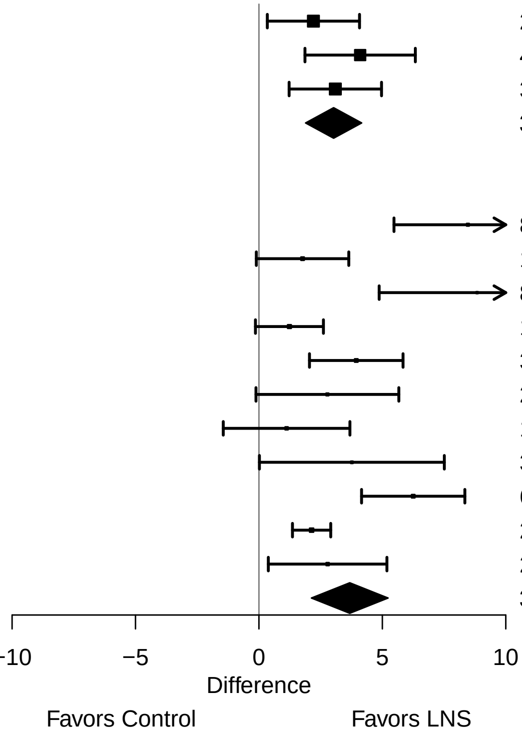

**Supplemental figure 4A: Mean difference in hemoglobin concentration**

#### 4A2: Stratified by Anemia burden

**Anemia burden**
**(p−diff = 0.077)**

| Country | Trial | N | N | MD (95% CI) | W |
|---|---|---|---|---|---|
| **Anemia burden − Moderate** | | | | | |
| Bangladesh | JiVitA−4 (35) | 457 | 146 | 2.21 (0.34, 4.07) | 0.13 |
| Bangladesh | RDNS (36) | 549 | 272 | 4.10 (1.86, 6.34) | 0.12 |
| Bangladesh | WASH−B (37) | 234 | 186 | 3.09 (1.22, 4.97) | 0.13 |
| Ghana | iLiNS−DYADG (41) | 328 | 661 | 1.24 (−0.14, 2.61) | 0.14 |
| Kenya | WASH−B (42) | 350 | 300 | 3.94 (2.05, 5.84) | 0.13 |
| Madagascar | MAHAY (43) | 600 | 588 | 2.77 (−0.12, 5.67) | 0.10 |
| Zimbabwe | SHINE (HIV−) (47) | 1682 | 1594 | 2.13 (1.36, 2.91) | 0.15 |
| Zimbabwe | SHINE (HIV+) (48) | 306 | 285 | 2.78 (0.38, 5.19) | 0.11 |
| **$I^2$ = 0.20, $Tau^2$ = 0.16** | | **4506** | **4032** | **2.49 (1.84, 3.14)** | |
| **Anemia burden − High** | | | | | |
| Burkina Faso | iLiNS−Zinc (38) | 1957 | 664 | 8.47 (5.47, 11.46) | 0.16 |
| Burkina Faso | PROMIS CS (39) | 574 | 581 | 1.77 (−0.11, 3.64) | 0.21 |
| Ghana | GHANA (40) | 98 | 96 | 8.84 (4.87, 12.81) | 0.12 |
| Malawi | iLiNS−DYADM (44) | 210 | 432 | 1.12 (−1.45, 3.69) | 0.18 |
| Malawi | iLiNS−DOSE (45) | 243 | 82 | 3.76 (0.02, 7.51) | 0.13 |
| Mali | PROMIS CS (46) | 953 | 970 | 6.25 (4.16, 8.34) | 0.20 |
| **$I^2$ = 0.83, $Tau^2$ = 8.80** | | **4035** | **2825** | **4.88 (2.23, 7.53)** | |

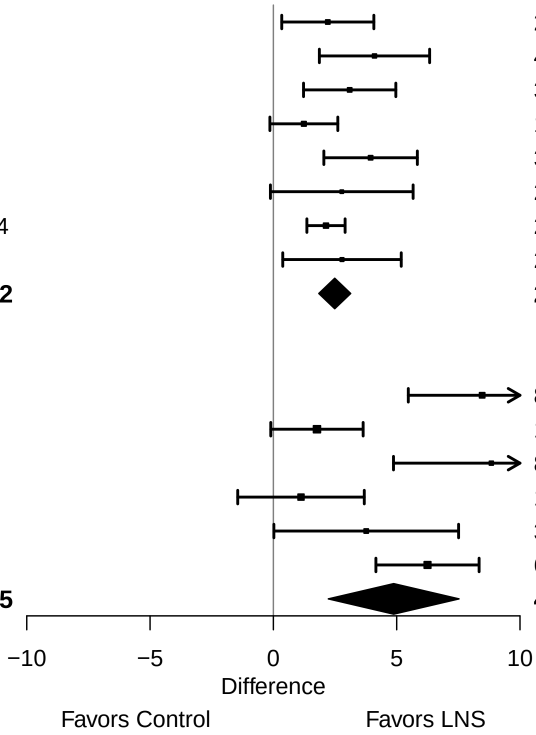

#### Supplemental figure 4A: Mean difference in hemoglobin concentration

#### 4A3: Stratified by Malaria prevalence

**Malaria prevalence**
**(p−diff = 0.354)**

| **Malaria prevalence − Less than 10%** | | | | **MD** | |
|---|---|---|---|---|---|
| **Country** | **Trial** | **N** | **N** | **(95% CI)** | **W** |
| Bangladesh | JiVitA−4 (35) | 457 | 146 | 2.21 (0.34, 4.07) | 0.15 |
| Bangladesh | RDNS (36) | 549 | 272 | 4.10 (1.86, 6.34) | 0.14 |
| Bangladesh | WASH−B (37) | 234 | 186 | 3.09 (1.22, 4.97) | 0.15 |
| Kenya | WASH−B (42) | 350 | 300 | 3.94 (2.05, 5.84) | 0.15 |
| Madagascar | MAHAY (43) | 600 | 588 | 2.77 (−0.12, 5.67) | 0.12 |
| Zimbabwe | SHINE (HIV−) (47) | 1682 | 1594 | 2.13 (1.36, 2.91) | 0.17 |
| Zimbabwe | SHINE (HIV+) (48) | 306 | 285 | 2.78 (0.38, 5.19) | 0.13 |
| **I² = 0.00, Tau² = 0.00** | | **4178** | **3371** | **2.59 (2.02, 3.17)** | |
| **Malaria prevalence − At least 10%** | | | | | |
| Burkina Faso | iLiNS−Zinc (38) | 1957 | 664 | 8.47 (5.47, 11.46) | 0.13 |
| Burkina Faso | PROMIS CS (39) | 574 | 581 | 1.77 (−0.11, 3.64) | 0.17 |
| Ghana | GHANA (40) | 98 | 96 | 8.84 (4.87, 12.81) | 0.10 |
| Ghana | iLiNS−DYADG (41) | 328 | 661 | 1.24 (−0.14, 2.61) | 0.19 |
| Malawi | iLiNS−DYADM (44) | 210 | 432 | 1.12 (−1.45, 3.69) | 0.14 |
| Malawi | iLiNS−DOSE (45) | 243 | 82 | 3.76 (0.02, 7.51) | 0.11 |
| Mali | PROMIS CS (46) | 953 | 970 | 6.25 (4.16, 8.34) | 0.16 |
| **I² = 0.85, Tau² = 9.15** | | **4363** | **3486** | **4.29 (1.82, 6.75)** | |

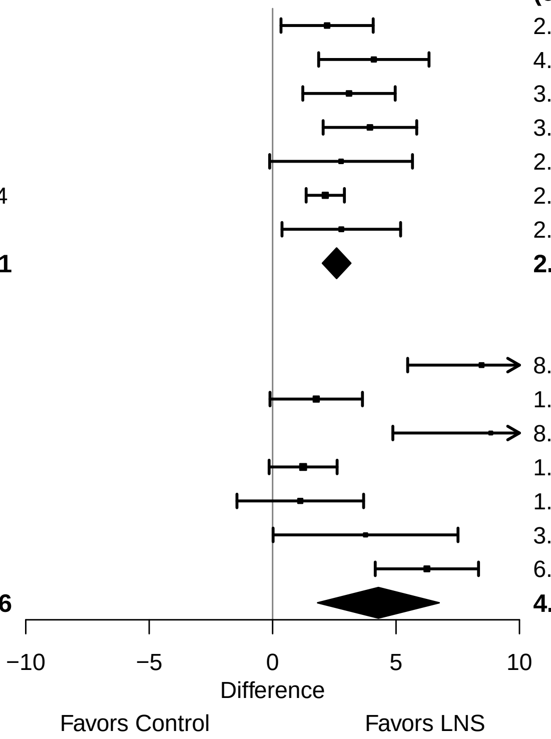

**Supplemental figure 4A: Mean difference in hemoglobin concentration**

#### 4A4: Stratified by Inflammation burden

**Inflammation burden**
**(p−diff = 0.676)**

| Country | Trial | N | N | MD (95% CI) | W |
|---|---|---|---|---|---|
| **Inflammation burden − Low** | | | | | |
| Bangladesh | RDNS (36) | 549 | 272 | 4.10 (1.86, 6.34) | 0.12 |
| Bangladesh | WASH−B (37) | 234 | 186 | 3.09 (1.22, 4.97) | 0.13 |
| Ghana | GHANA (40) | 98 | 96 | 8.84 (4.87, 12.81) | 0.08 |
| Ghana | iLiNS−DYADG (41) | 328 | 661 | 1.24 (−0.14, 2.61) | 0.15 |
| Kenya | WASH−B (42) | 350 | 300 | 3.94 (2.05, 5.84) | 0.13 |
| Madagascar | MAHAY (43) | 600 | 588 | 2.77 (−0.12, 5.67) | 0.11 |
| Zimbabwe | SHINE (HIV−) (47) | 1682 | 1594 | 2.13 (1.36, 2.91) | 0.16 |
| Zimbabwe | SHINE (HIV+) (48) | 306 | 285 | 2.78 (0.38, 5.19) | 0.12 |
| **I² = 0.62, Tau² = 2.50** | | **4147** | **3982** | **3.21 (1.88, 4.54)** | |
| **Inflammation burden − High** | | | | | |
| Bangladesh | JiVitA−4 (35) | 457 | 146 | 2.21 (0.34, 4.07) | 0.19 |
| Burkina Faso | iLiNS−Zinc (38) | 1957 | 664 | 8.47 (5.47, 11.46) | 0.15 |
| Burkina Faso | PROMIS CS (39) | 574 | 581 | 1.77 (−0.11, 3.64) | 0.19 |
| Malawi | iLiNS−DYADM (44) | 210 | 432 | 1.12 (−1.45, 3.69) | 0.16 |
| Malawi | iLiNS−DOSE (45) | 243 | 82 | 3.76 (0.02, 7.51) | 0.12 |
| Mali | PROMIS CS (46) | 953 | 970 | 6.25 (4.16, 8.34) | 0.18 |
| **I² = 0.81, Tau² = 6.54** | | **4394** | **2875** | **3.84 (1.55, 6.13)** | |

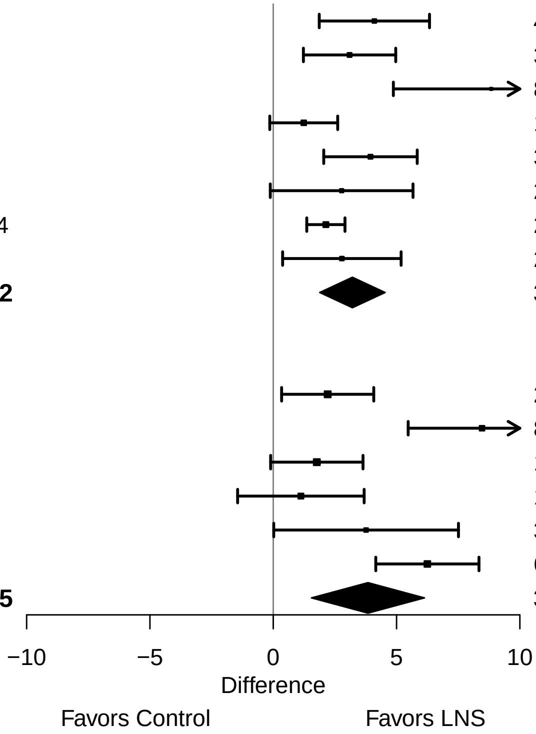

#### Supplemental figure 4A: Mean difference in hemoglobin concentration

#### 4A5: Stratified by Source water quality

**Source water quality**
**(p−diff = 0.711)**

| Country | Trial | N | N | MD (95% CI) | W |
|---|---|---|---|---|---|
| **Source water quality − Improved** | | | | | |
| Bangladesh | JiVitA−4 (35) | 457 | 146 | 2.21 (0.34, 4.07) | 0.17 |
| Bangladesh | RDNS (36) | 549 | 272 | 4.10 (1.86, 6.34) | 0.16 |
| Bangladesh | WASH−B (37) | 105 | 60 | 3.25 (−0.27, 6.78) | 0.12 |
| Ghana | GHANA (40) | 98 | 96 | 8.84 (4.87, 12.81) | 0.10 |
| Ghana | iLiNS−DYADG (41) | 328 | 661 | 1.24 (−0.14, 2.61) | 0.19 |
| Malawi | iLiNS−DYADM (44) | 210 | 432 | 1.12 (−1.45, 3.69) | 0.15 |
| Malawi | iLiNS−DOSE (45) | 243 | 82 | 3.76 (0.02, 7.51) | 0.11 |
| **I² = 0.64, Tau² = 3.93** | | **1990** | **1749** | **3.15 (1.36, 4.94)** | |
| **Source water quality − Unimproved** | | | | | |
| Burkina Faso | iLiNS−Zinc (38) | 1957 | 664 | 8.47 (5.47, 11.46) | 0.12 |
| Burkina Faso | PROMIS CS (39) | 574 | 581 | 1.77 (−0.11, 3.64) | 0.16 |
| Kenya | WASH−B (42) | 196 | 162 | 3.20 (0.41, 6.00) | 0.13 |
| Madagascar | MAHAY (43) | 600 | 588 | 2.77 (−0.12, 5.67) | 0.13 |
| Mali | PROMIS CS (46) | 953 | 970 | 6.25 (4.16, 8.34) | 0.15 |
| Zimbabwe | SHINE (HIV−) (47) | 819 | 795 | 2.12 (0.93, 3.31) | 0.18 |
| Zimbabwe | SHINE (HIV+) (48) | 133 | 126 | 1.52 (−1.46, 4.49) | 0.13 |
| **I² = 0.78, Tau² = 5.06** | | **5232** | **3886** | **3.65 (1.75, 5.55)** | |

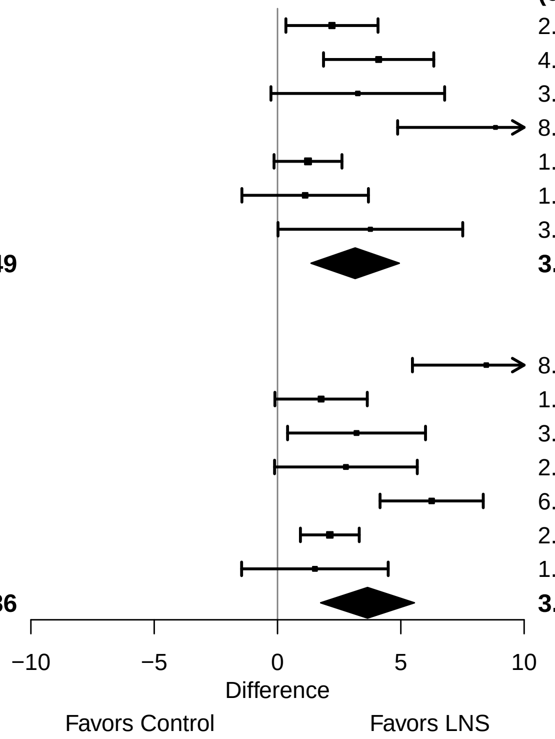

#### Supplemental figure 4A: Mean difference in hemoglobin concentration

### 4A6: Stratified by Sanitation

**Sanitation**
**(p−diff = 0.723)**

| Country | Trial | N | N | MD (95% CI) | W |
|---|---|---|---|---|---|
| **Sanitation − Improved** | | | | | |
| Bangladesh | JiVitA−4 (35) | 457 | 146 | 2.21 (0.34, 4.07) | 0.16 |
| Bangladesh | RDNS (36) | 549 | 272 | 4.10 (1.86, 6.34) | 0.15 |
| Bangladesh | WASH−B (37) | 105 | 60 | 3.25 (−0.27, 6.78) | 0.11 |
| Burkina Faso | PROMIS CS (39) | 574 | 581 | 1.77 (−0.11, 3.64) | 0.16 |
| Ghana | GHANA (40) | 98 | 96 | 8.84 (4.87, 12.81) | 0.10 |
| Ghana | iLiNS−DYADG (41) | 328 | 661 | 1.24 (−0.14, 2.61) | 0.17 |
| Mali | PROMIS CS (46) | 953 | 970 | 6.25 (4.16, 8.34) | 0.15 |
| **I² = 0.78, Tau² = 5.14** | | **3064** | **2786** | **3.67 (1.76, 5.58)** | |
| **Sanitation − Unimproved** | | | | | |
| Burkina Faso | iLiNS−Zinc (38) | 1957 | 664 | 8.47 (5.47, 11.46) | 0.13 |
| Kenya | WASH−B (42) | 196 | 162 | 3.20 (0.41, 6.00) | 0.14 |
| Madagascar | MAHAY (43) | 600 | 588 | 2.77 (−0.12, 5.67) | 0.14 |
| Malawi | iLiNS−DYADM (44) | 210 | 432 | 1.12 (−1.45, 3.69) | 0.15 |
| Malawi | iLiNS−DOSE (45) | 243 | 82 | 3.76 (0.02, 7.51) | 0.11 |
| Zimbabwe | SHINE (HIV−) (47) | 819 | 795 | 2.12 (0.93, 3.31) | 0.19 |
| Zimbabwe | SHINE (HIV+) (48) | 133 | 126 | 1.52 (−1.46, 4.49) | 0.13 |
| **I² = 0.66, Tau² = 3.92** | | **4158** | **2849** | **3.16 (1.37, 4.96)** | |

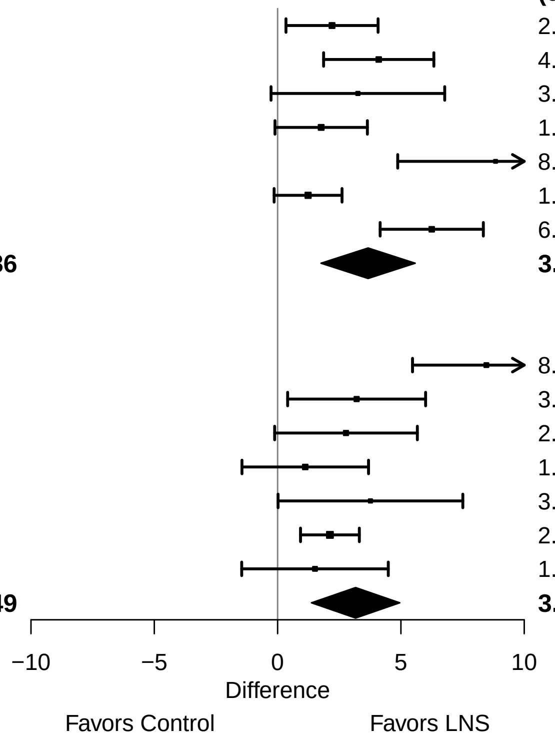

**Supplemental figure 4A: Mean difference in hemoglobin concentration**

#### 4A7: Stratified by Supplement duration

**Supplement duration**
**(p−diff = 0.304)**

| Country | Trial | N | N | MD (95% CI) | W |
|---|---|---|---|---|---|
| **Supplement duration − 12m or less** | | | | | |
| Bangladesh | JiVitA−4 (35) | 457 | 146 | 2.21 (0.34, 4.07) | 0.11 |
| Burkina Faso | iLiNS−Zinc (38) | 1957 | 664 | 8.47 (5.47, 11.46) | 0.09 |
| Burkina Faso | PROMIS CS (39) | 574 | 581 | 1.77 (−0.11, 3.64) | 0.11 |
| Ghana | GHANA (40) | 98 | 96 | 8.84 (4.87, 12.81) | 0.07 |
| Ghana | iLiNS−DYADG (41) | 328 | 661 | 1.24 (−0.14, 2.61) | 0.12 |
| Madagascar | MAHAY (43) | 600 | 588 | 2.77 (−0.12, 5.67) | 0.09 |
| Malawi | iLiNS−DYADM (44) | 210 | 432 | 1.12 (−1.45, 3.69) | 0.10 |
| Malawi | iLiNS−DOSE (45) | 243 | 82 | 3.76 (0.02, 7.51) | 0.07 |
| Zimbabwe | SHINE (HIV−) (47) | 1682 | 1594 | 2.13 (1.36, 2.91) | 0.13 |
| Zimbabwe | SHINE (HIV+) (48) | 306 | 285 | 2.78 (0.38, 5.19) | 0.10 |
| **I² = 0.71, Tau² = 5.37** | | **6455** | **5129** | **3.21 (1.58, 4.85)** | |
| **Supplement duration − > 12m** | | | | | |
| Bangladesh | RDNS (36) | 549 | 272 | 4.10 (1.86, 6.34) | 0.24 |
| Bangladesh | WASH−B (37) | 234 | 186 | 3.09 (1.22, 4.97) | 0.26 |
| Kenya | WASH−B (42) | 350 | 300 | 3.94 (2.05, 5.84) | 0.26 |
| Mali | PROMIS CS (46) | 953 | 970 | 6.25 (4.16, 8.34) | 0.25 |
| **I² = 0.41, Tau² = 0.74** | | **2086** | **1728** | **4.30 (2.98, 5.61)** | |

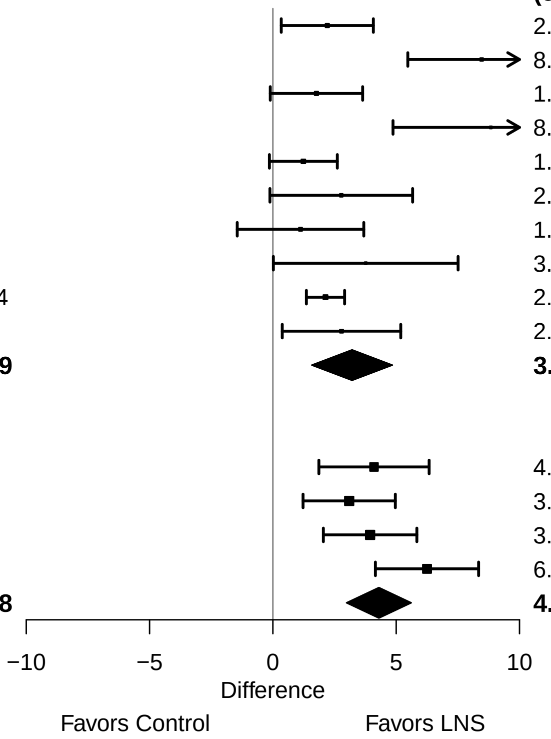

#### Supplemental figure 4A: Mean difference in hemoglobin concentration

### 4A8: Stratified by Iron dose

**Iron dose**
**(p−diff = 0.253)**

| Country | Trial | N | N | MD (95% CI) | W |
|---|---|---|---|---|---|
| **Iron dose − Less than 9 mg** | | | | | |
| Bangladesh | JiVitA−4 (35) | 457 | 146 | 2.21 (0.34, 4.07) | 0.11 |
| Burkina Faso | iLiNS−Zinc (38) | 1957 | 664 | 8.47 (5.47, 11.46) | 0.08 |
| Burkina Faso | PROMIS CS (39) | 574 | 581 | 1.77 (−0.11, 3.64) | 0.11 |
| Ghana | iLiNS−DYADG (41) | 328 | 661 | 1.24 (−0.14, 2.61) | 0.12 |
| Madagascar | MAHAY (43) | 600 | 588 | 2.77 (−0.12, 5.67) | 0.09 |
| Malawi | iLiNS−DYADM (44) | 210 | 432 | 1.12 (−1.45, 3.69) | 0.09 |
| Malawi | iLiNS−DOSE (45) | 243 | 82 | 3.76 (0.02, 7.51) | 0.07 |
| Mali | PROMIS CS (46) | 953 | 970 | 6.25 (4.16, 8.34) | 0.10 |
| Zimbabwe | SHINE (HIV−) (47) | 1682 | 1594 | 2.13 (1.36, 2.91) | 0.13 |
| Zimbabwe | SHINE (HIV+) (48) | 306 | 285 | 2.78 (0.38, 5.19) | 0.10 |
| **$I^2$ = 0.74, $Tau^2$ = 3.86** | | **7310** | **6003** | **3.09 (1.68, 4.50)** | |
| **Iron dose − 9 mg** | | | | | |
| Bangladesh | RDNS (36) | 549 | 272 | 4.10 (1.86, 6.34) | 0.26 |
| Bangladesh | WASH−B (37) | 234 | 186 | 3.09 (1.22, 4.97) | 0.28 |
| Ghana | GHANA (40) | 98 | 96 | 8.84 (4.87, 12.81) | 0.17 |
| Kenya | WASH−B (42) | 350 | 300 | 3.94 (2.05, 5.84) | 0.28 |
| **$I^2$ = 0.55, $Tau^2$ = 3.56** | | **1231** | **854** | **4.56 (2.34, 6.77)** | |

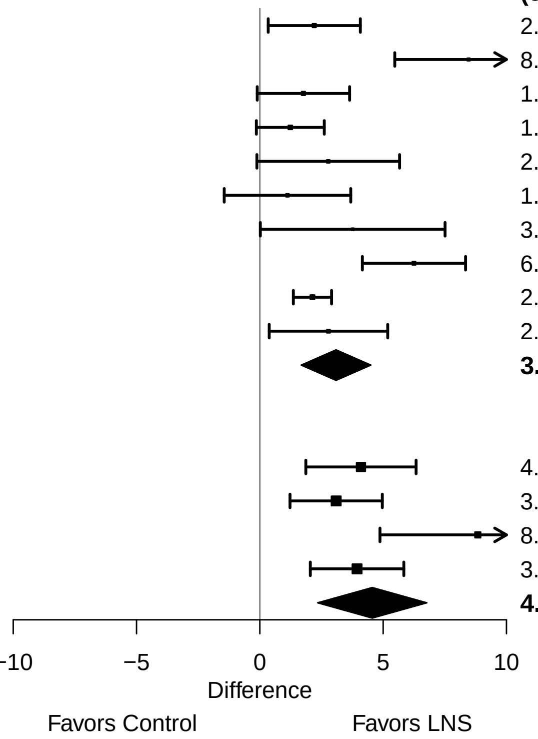

**Supplemental figure 4A: Mean difference in hemoglobin concentration**

#### 4A9: Stratified by Frequency of contact

**Frequency of contact**
**(p−diff = 0.806)**

| Country | Trial | N | N | MD (95% CI) | W |
|---|---|---|---|---|---|
| **Frequency of contact − Monthly** | | | | | |
| Bangladesh | RDNS (36) | 549 | 272 | 4.10 (1.86, 6.34) | 0.14 |
| Burkina Faso | PROMIS CS (39) | 574 | 581 | 1.77 (−0.11, 3.64) | 0.15 |
| Kenya | WASH−B (42) | 350 | 300 | 3.94 (2.05, 5.84) | 0.15 |
| Madagascar | MAHAY (43) | 600 | 588 | 2.77 (−0.12, 5.67) | 0.12 |
| Mali | PROMIS CS (46) | 953 | 970 | 6.25 (4.16, 8.34) | 0.14 |
| Zimbabwe | SHINE (HIV−) (47) | 1682 | 1594 | 2.13 (1.36, 2.91) | 0.18 |
| Zimbabwe | SHINE (HIV+) (48) | 306 | 285 | 2.78 (0.38, 5.19) | 0.13 |
| **I² = 0.65, Tau² = 1.36** | | **5014** | **4590** | **3.30 (2.15, 4.44)** | |
| **Frequency of contact − Weekly** | | | | | |
| Bangladesh | JiVitA−4 (35) | 457 | 146 | 2.21 (0.34, 4.07) | 0.17 |
| Bangladesh | WASH−B (37) | 234 | 186 | 3.09 (1.22, 4.97) | 0.17 |
| Burkina Faso | iLiNS−Zinc (38) | 1957 | 664 | 8.47 (5.47, 11.46) | 0.13 |
| Ghana | GHANA (40) | 98 | 96 | 8.84 (4.87, 12.81) | 0.10 |
| Ghana | iLiNS−DYADG (41) | 328 | 661 | 1.24 (−0.14, 2.61) | 0.18 |
| Malawi | iLiNS−DYADM (44) | 210 | 432 | 1.12 (−1.45, 3.69) | 0.14 |
| Malawi | iLiNS−DOSE (45) | 243 | 82 | 3.76 (0.02, 7.51) | 0.11 |
| **I² = 0.80, Tau² = 8.10** | | **3527** | **2267** | **3.84 (1.51, 6.18)** | |

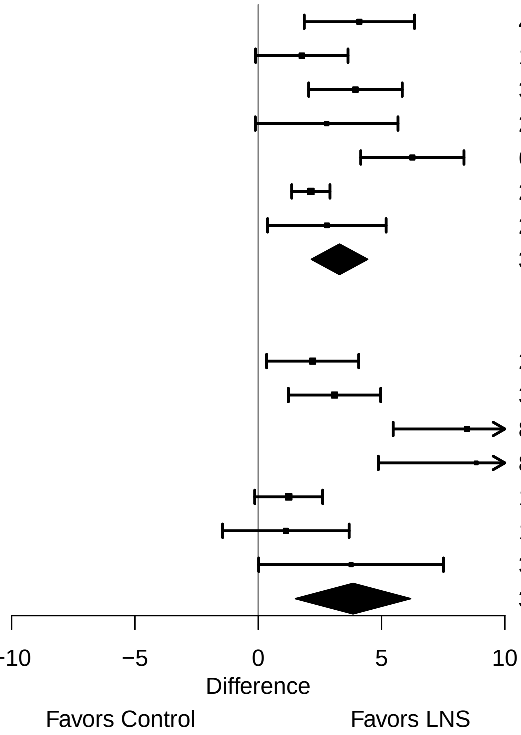

#### Supplemental figure 4A: Mean difference in hemoglobin concentration

### 4A10: Stratified by Average SQ-LNS compliance

**Average SQ−LNS compliance**
**(p−diff = 0.086)**

| Country | Trial | N | N | MD (95% CI) | W |
|---|---|---|---|---|---|
| **Average SQ−LNS compliance − Low** | | | | | |
| Burkina Faso | PROMIS CS (39) | 574 | 581 | 1.77 (−0.11, 3.64) | 0.15 |
| Ghana | iLiNS−DYADG (41) | 328 | 661 | 1.24 (−0.14, 2.61) | 0.16 |
| Malawi | iLiNS−DYADM (44) | 210 | 432 | 1.12 (−1.45, 3.69) | 0.13 |
| Malawi | iLiNS−DOSE (45) | 243 | 82 | 3.76 (0.02, 7.51) | 0.10 |
| Mali | PROMIS CS (46) | 953 | 970 | 6.25 (4.16, 8.34) | 0.14 |
| Zimbabwe | SHINE (HIV−) (47) | 1682 | 1594 | 2.13 (1.36, 2.91) | 0.18 |
| Zimbabwe | SHINE (HIV+) (48) | 306 | 285 | 2.78 (0.38, 5.19) | 0.14 |
| **$I^2$ = 0.67, $Tau^2$ = 2.09** | | **4296** | **4605** | **2.59 (1.27, 3.91)** | |
| **Average SQ−LNS compliance − High** | | | | | |
| Bangladesh | JiVitA−4 (35) | 457 | 146 | 2.21 (0.34, 4.07) | 0.19 |
| Bangladesh | RDNS (36) | 549 | 272 | 4.10 (1.86, 6.34) | 0.17 |
| Bangladesh | WASH−B (37) | 234 | 186 | 3.09 (1.22, 4.97) | 0.19 |
| Burkina Faso | iLiNS−Zinc (38) | 1957 | 664 | 8.47 (5.47, 11.46) | 0.15 |
| Ghana | GHANA (40) | 98 | 96 | 8.84 (4.87, 12.81) | 0.12 |
| Kenya | WASH−B (42) | 350 | 300 | 3.94 (2.05, 5.84) | 0.19 |
| **$I^2$ = 0.73, $Tau^2$ = 5.66** | | **3645** | **1664** | **4.78 (2.63, 6.94)** | |

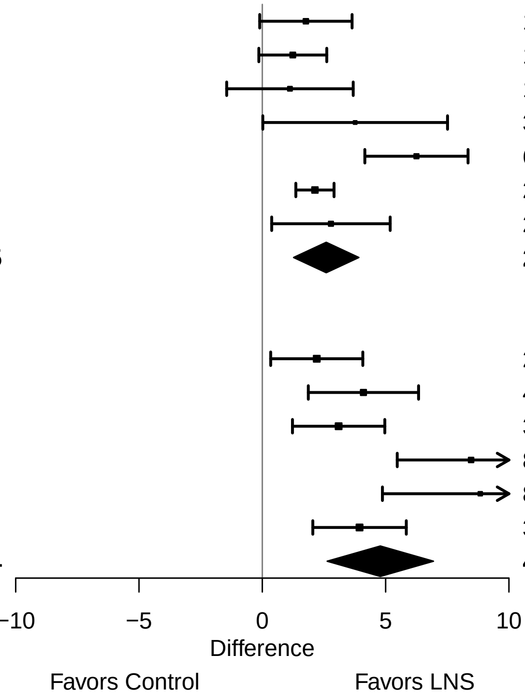

### Supplemental figure 4B: Anemia prevalence ratio

#### 4B1: Stratified by Geographic region

**Geographic region**
**(p−diff = 0.055)**

| **Geographic region − SEAR** | | | | **PR** | |
|---|---|---|---|---|---|
| **Country** | **Trial** | **N** | **N** | **(95% CI)** | **W** |
| Bangladesh | JiVitA−4 (35) | 457 | 146 | 0.83 (0.54, 1.28) | 0.29 |
| Bangladesh | RDNS (36) | 549 | 272 | 0.66 (0.51, 0.84) | 0.52 |
| Bangladesh | WASH−B (37) | 234 | 186 | 0.42 (0.24, 0.74) | 0.19 |
| **I² = 0.43, Tau² = 0.05** | | **1240** | **604** | **0.64 (0.45, 0.90)** | |
| **Geographic region − AFR** | | | | | |
| Burkina Faso | iLiNS−Zinc (38) | 1957 | 664 | 0.87 (0.83, 0.91) | 0.11 |
| Burkina Faso | PROMIS CS (39) | 574 | 581 | 0.94 (0.85, 1.03) | 0.11 |
| Ghana | GHANA (40) | 98 | 96 | 0.51 (0.36, 0.72) | 0.05 |
| Ghana | iLiNS−DYADG (41) | 328 | 661 | 0.86 (0.73, 1.01) | 0.09 |
| Kenya | WASH−B (42) | 350 | 300 | 0.68 (0.57, 0.81) | 0.09 |
| Madagascar | MAHAY (43) | 600 | 588 | 0.88 (0.78, 1.01) | 0.10 |
| Malawi | iLiNS−DYADM (44) | 210 | 432 | 0.90 (0.76, 1.07) | 0.09 |
| Malawi | iLiNS−DOSE (45) | 243 | 82 | 0.87 (0.74, 1.02) | 0.09 |
| Mali | PROMIS CS (46) | 953 | 970 | 0.82 (0.76, 0.88) | 0.11 |
| Zimbabwe | SHINE (HIV−) (47) | 1682 | 1594 | 0.78 (0.70, 0.86) | 0.10 |
| Zimbabwe | SHINE (HIV+) (48) | 306 | 285 | 0.65 (0.50, 0.85) | 0.06 |
| **I² = 0.64, Tau² = 0.02** | | **7301** | **6253** | **0.81 (0.74, 0.89)** | |

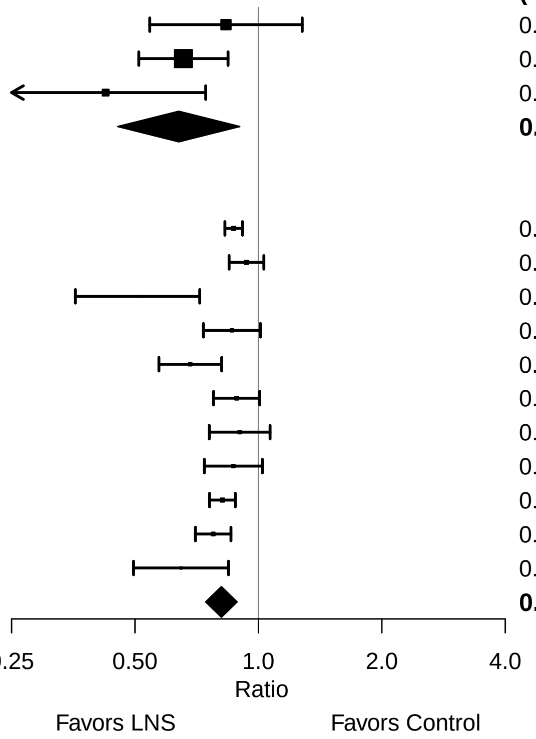

#### Supplemental figure 4B: Anemia prevalence ratio

#### 4B2: Stratified by Anemia burden

**Anemia burden**
**(p−diff = 0.073)**

| Country | Trial | N | N | PR (95% CI) | W |
|---|---|---|---|---|---|
| **Anemia burden − Moderate** | | | | | |
| Bangladesh | JiVitA−4 (35) | 457 | 146 | 0.83 (0.54, 1.28) | 0.07 |
| Bangladesh | RDNS (36) | 549 | 272 | 0.66 (0.51, 0.84) | 0.12 |
| Bangladesh | WASH−B (37) | 234 | 186 | 0.42 (0.24, 0.74) | 0.04 |
| Ghana | iLiNS−DYADG (41) | 328 | 661 | 0.86 (0.73, 1.01) | 0.16 |
| Kenya | WASH−B (42) | 350 | 300 | 0.68 (0.57, 0.81) | 0.15 |
| Madagascar | MAHAY (43) | 600 | 588 | 0.88 (0.78, 1.01) | 0.17 |
| Zimbabwe | SHINE (HIV−) (47) | 1682 | 1594 | 0.78 (0.70, 0.86) | 0.18 |
| Zimbabwe | SHINE (HIV+) (48) | 306 | 285 | 0.65 (0.50, 0.85) | 0.11 |
| **$I^2$ = 0.55, $Tau^2$ = 0.02** | | **4506** | **4032** | **0.75 (0.66, 0.84)** | |
| **Anemia burden − High** | | | | | |
| Burkina Faso | iLiNS−Zinc (38) | 1957 | 664 | 0.87 (0.83, 0.91) | 0.20 |
| Burkina Faso | PROMIS CS (39) | 574 | 581 | 0.94 (0.85, 1.03) | 0.19 |
| Ghana | GHANA (40) | 98 | 96 | 0.51 (0.36, 0.72) | 0.09 |
| Malawi | iLiNS−DYADM (44) | 210 | 432 | 0.90 (0.76, 1.07) | 0.16 |
| Malawi | iLiNS−DOSE (45) | 243 | 82 | 0.87 (0.74, 1.02) | 0.16 |
| Mali | PROMIS CS (46) | 953 | 970 | 0.82 (0.76, 0.88) | 0.20 |
| **$I^2$ = 0.64, $Tau^2$ = 0.03** | | **4035** | **2825** | **0.83 (0.72, 0.96)** | |

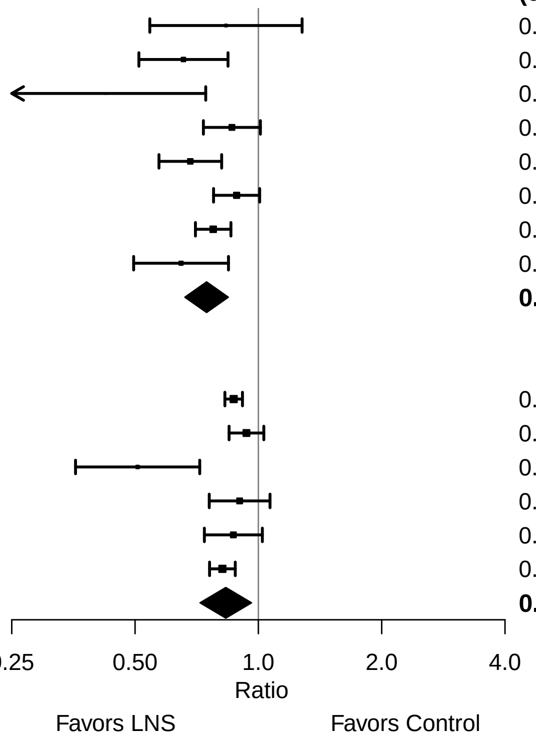

#### Supplemental figure 4B: Anemia prevalence ratio

#### 4B3: Stratified by Malaria prevalence

**Malaria prevalence**
**(p−diff = 0.023)**

| Country | Trial | N | N | PR (95% CI) | W |
|---|---|---|---|---|---|
| **Malaria prevalence − Less than 10%** | | | | | |
| Bangladesh | JiVitA−4 (35) | 457 | 146 | 0.83 (0.54, 1.28) | 0.08 |
| Bangladesh | RDNS (36) | 549 | 272 | 0.66 (0.51, 0.84) | 0.14 |
| Bangladesh | WASH−B (37) | 234 | 186 | 0.42 (0.24, 0.74) | 0.05 |
| Kenya | WASH−B (42) | 350 | 300 | 0.68 (0.57, 0.81) | 0.18 |
| Madagascar | MAHAY (43) | 600 | 588 | 0.88 (0.78, 1.01) | 0.20 |
| Zimbabwe | SHINE (HIV−) (47) | 1682 | 1594 | 0.78 (0.70, 0.86) | 0.22 |
| Zimbabwe | SHINE (HIV+) (48) | 306 | 285 | 0.65 (0.50, 0.85) | 0.13 |
| **I² = 0.57, Tau² = 0.02** | | **4178** | **3371** | **0.73 (0.64, 0.83)** | |
| **Malaria prevalence − At least 10%** | | | | | |
| Burkina Faso | iLiNS−Zinc (38) | 1957 | 664 | 0.87 (0.83, 0.91) | 0.18 |
| Burkina Faso | PROMIS CS (39) | 574 | 581 | 0.94 (0.85, 1.03) | 0.16 |
| Ghana | GHANA (40) | 98 | 96 | 0.51 (0.36, 0.72) | 0.08 |
| Ghana | iLiNS−DYADG (41) | 328 | 661 | 0.86 (0.73, 1.01) | 0.14 |
| Malawi | iLiNS−DYADM (44) | 210 | 432 | 0.90 (0.76, 1.07) | 0.14 |
| Malawi | iLiNS−DOSE (45) | 243 | 82 | 0.87 (0.74, 1.02) | 0.14 |
| Mali | PROMIS CS (46) | 953 | 970 | 0.82 (0.76, 0.88) | 0.17 |
| **I² = 0.57, Tau² = 0.02** | | **4363** | **3486** | **0.84 (0.75, 0.94)** | |

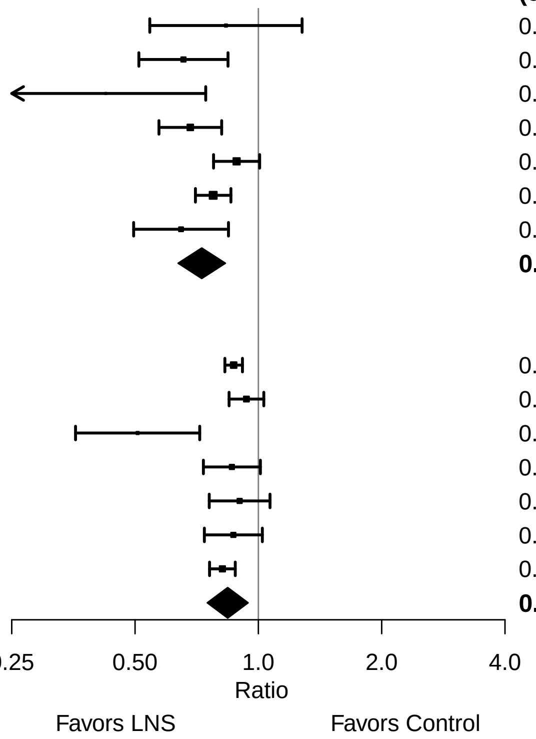

##### Supplemental figure 4B: Anemia prevalence ratio

#### 4B4: Stratified by Inflammation burden

**Inflammation burden**
**(p−diff = 0.008)**

| Country | Trial | N | N | PR (95% CI) | W |
|---|---|---|---|---|---|
| **Inflammation burden − Low** | | | | | |
| Bangladesh | RDNS (36) | 549 | 272 | 0.66 (0.51, 0.84) | 0.12 |
| Bangladesh | WASH−B (37) | 234 | 186 | 0.42 (0.24, 0.74) | 0.04 |
| Ghana | GHANA (40) | 98 | 96 | 0.51 (0.36, 0.72) | 0.08 |
| Ghana | iLiNS−DYADG (41) | 328 | 661 | 0.86 (0.73, 1.01) | 0.15 |
| Kenya | WASH−B (42) | 350 | 300 | 0.68 (0.57, 0.81) | 0.15 |
| Madagascar | MAHAY (43) | 600 | 588 | 0.88 (0.78, 1.01) | 0.17 |
| Zimbabwe | SHINE (HIV−) (47) | 1682 | 1594 | 0.78 (0.70, 0.86) | 0.18 |
| Zimbabwe | SHINE (HIV+) (48) | 306 | 285 | 0.65 (0.50, 0.85) | 0.11 |
| **$I^2$ = 0.67, $Tau^2$ = 0.03** | | **4147** | **3982** | **0.71 (0.61, 0.82)** | |
| **Inflammation burden − High** | | | | | |
| Bangladesh | JiVitA−4 (35) | 457 | 146 | 0.83 (0.54, 1.28) | 0.07 |
| Burkina Faso | iLiNS−Zinc (38) | 1957 | 664 | 0.87 (0.83, 0.91) | 0.21 |
| Burkina Faso | PROMIS CS (39) | 574 | 581 | 0.94 (0.85, 1.03) | 0.19 |
| Malawi | iLiNS−DYADM (44) | 210 | 432 | 0.90 (0.76, 1.07) | 0.16 |
| Malawi | iLiNS−DOSE (45) | 243 | 82 | 0.87 (0.74, 1.02) | 0.16 |
| Mali | PROMIS CS (46) | 953 | 970 | 0.82 (0.76, 0.88) | 0.20 |
| **$I^2$ = 0.03, $Tau^2$ = 0.00** | | **4394** | **2875** | **0.87 (0.84, 0.90)** | |

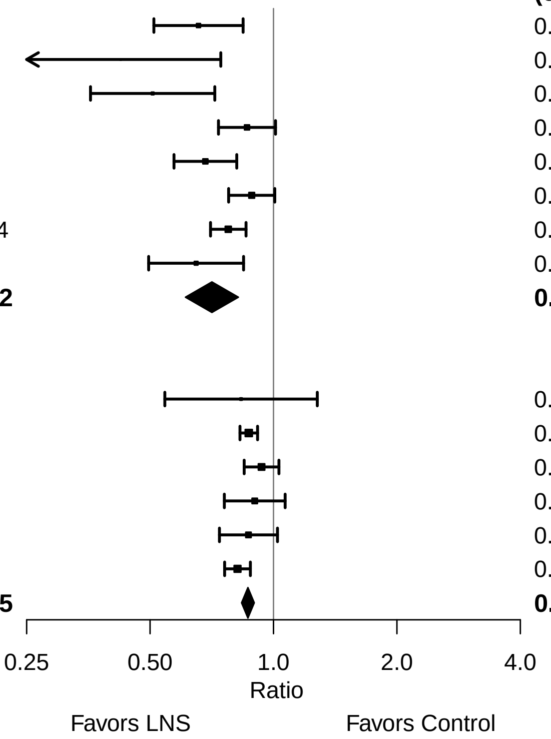

#### Supplemental figure 4B: Anemia prevalence ratio

##### 4B5: Stratified by Source water quality

**Source water quality**
**(p−diff = 0.372)**

| Country | Trial | N | N | PR (95% CI) | W |
|---|---|---|---|---|---|
| **Source water quality − Improved** | | | | | |
| Bangladesh | JiVitA−4 (35) | 457 | 146 | 0.83 (0.54, 1.28) | 0.07 |
| Bangladesh | RDNS (36) | 549 | 272 | 0.66 (0.51, 0.84) | 0.15 |
| Bangladesh | WASH−B (37) | 105 | 60 | 0.31 (0.10, 1.02) | 0.01 |
| Ghana | GHANA (40) | 98 | 96 | 0.51 (0.36, 0.72) | 0.10 |
| Ghana | iLiNS−DYADG (41) | 328 | 661 | 0.86 (0.73, 1.01) | 0.23 |
| Malawi | iLiNS−DYADM (44) | 210 | 432 | 0.90 (0.76, 1.07) | 0.22 |
| Malawi | iLiNS−DOSE (45) | 243 | 82 | 0.87 (0.74, 1.02) | 0.23 |
| **I² = 0.60, Tau² = 0.04** | | **1990** | **1749** | **0.76 (0.63, 0.91)** | |
| **Source water quality − Unimproved** | | | | | |
| Burkina Faso | iLiNS−Zinc (38) | 1957 | 664 | 0.87 (0.83, 0.91) | 0.20 |
| Burkina Faso | PROMIS CS (39) | 574 | 581 | 0.94 (0.85, 1.03) | 0.18 |
| Kenya | WASH−B (42) | 196 | 162 | 0.74 (0.58, 0.94) | 0.09 |
| Madagascar | MAHAY (43) | 600 | 588 | 0.88 (0.78, 1.01) | 0.15 |
| Mali | PROMIS CS (46) | 953 | 970 | 0.82 (0.76, 0.88) | 0.19 |
| Zimbabwe | SHINE (HIV−) (47) | 819 | 795 | 0.76 (0.65, 0.88) | 0.14 |
| Zimbabwe | SHINE (HIV+) (48) | 133 | 126 | 0.79 (0.53, 1.18) | 0.05 |
| **I² = 0.37, Tau² = 0.00** | | **5232** | **3886** | **0.85 (0.81, 0.90)** | |

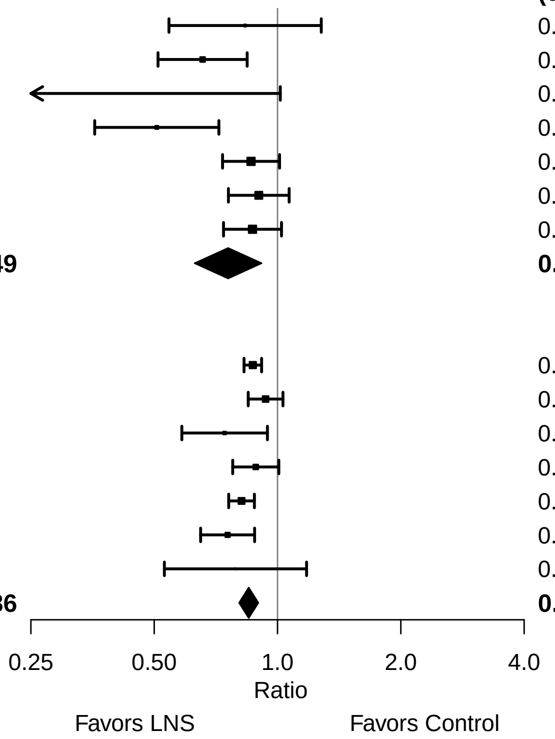

#### Supplemental figure 4B: Anemia prevalence ratio

##### 4B6: Stratified by Sanitation

**Sanitation**
**(p−diff = 0.518)**

| Country | Trial | N | N | PR (95% CI) | W |
|---|---|---|---|---|---|
| **Sanitation − Improved** | | | | | |
| Bangladesh | JiVitA−4 (35) | 457 | 146 | 0.83 (0.54, 1.28) | 0.06 |
| Bangladesh | RDNS (36) | 549 | 272 | 0.66 (0.51, 0.84) | 0.13 |
| Bangladesh | WASH−B (37) | 105 | 60 | 0.31 (0.10, 1.02) | 0.01 |
| Burkina Faso | PROMIS CS (39) | 574 | 581 | 0.94 (0.85, 1.03) | 0.25 |
| Ghana | GHANA (40) | 98 | 96 | 0.51 (0.36, 0.72) | 0.08 |
| Ghana | iLiNS−DYADG (41) | 328 | 661 | 0.86 (0.73, 1.01) | 0.19 |
| Mali | PROMIS CS (46) | 953 | 970 | 0.82 (0.76, 0.88) | 0.27 |
| **I² = 0.70, Tau² = 0.04** | | **3064** | **2786** | **0.76 (0.63, 0.91)** | |
| **Sanitation − Unimproved** | | | | | |
| Burkina Faso | iLiNS−Zinc (38) | 1957 | 664 | 0.87 (0.83, 0.91) | 0.22 |
| Kenya | WASH−B (42) | 196 | 162 | 0.74 (0.58, 0.94) | 0.10 |
| Madagascar | MAHAY (43) | 600 | 588 | 0.88 (0.78, 1.01) | 0.17 |
| Malawi | iLiNS−DYADM (44) | 210 | 432 | 0.90 (0.76, 1.07) | 0.14 |
| Malawi | iLiNS−DOSE (45) | 243 | 82 | 0.87 (0.74, 1.02) | 0.15 |
| Zimbabwe | SHINE (HIV−) (47) | 819 | 795 | 0.76 (0.65, 0.88) | 0.16 |
| Zimbabwe | SHINE (HIV+) (48) | 133 | 126 | 0.79 (0.53, 1.18) | 0.05 |
| **I² = 0.00, Tau² = 0.00** | | **4158** | **2849** | **0.86 (0.83, 0.90)** | |

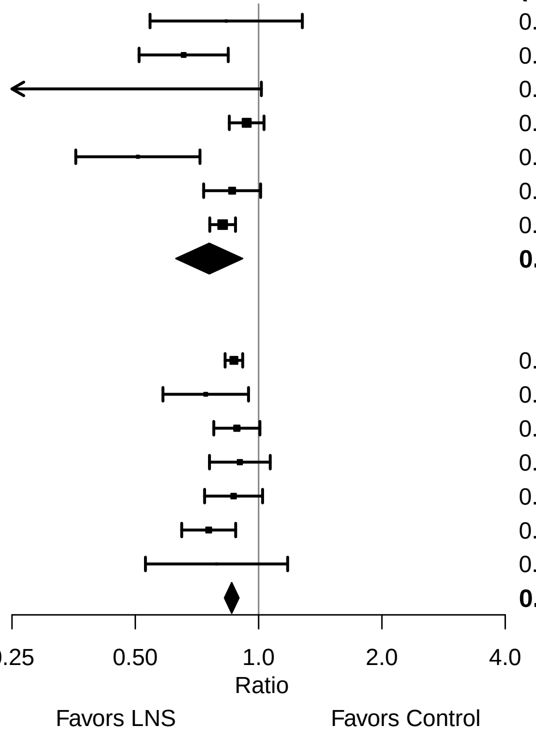

#### Supplemental figure 4B: Anemia prevalence ratio

### 4B7: Stratified by Supplement duration

**Supplement duration**
**(p−diff = 0.058)**

**Supplement duration − 12m or less**

| Country | Trial | N | N | PR (95% CI) | W |
|---|---|---|---|---|---|
| Bangladesh | JiVitA−4 (35) | 457 | 146 | 0.83 (0.54, 1.28) | 0.05 |
| Burkina Faso | iLiNS−Zinc (38) | 1957 | 664 | 0.87 (0.83, 0.91) | 0.14 |
| Burkina Faso | PROMIS CS (39) | 574 | 581 | 0.94 (0.85, 1.03) | 0.13 |
| Ghana | GHANA (40) | 98 | 96 | 0.51 (0.36, 0.72) | 0.06 |
| Ghana | iLiNS−DYADG (41) | 328 | 661 | 0.86 (0.73, 1.01) | 0.11 |
| Madagascar | MAHAY (43) | 600 | 588 | 0.88 (0.78, 1.01) | 0.12 |
| Malawi | iLiNS−DYADM (44) | 210 | 432 | 0.90 (0.76, 1.07) | 0.10 |
| Malawi | iLiNS−DOSE (45) | 243 | 82 | 0.87 (0.74, 1.02) | 0.11 |
| Zimbabwe | SHINE (HIV−) (47) | 1682 | 1594 | 0.78 (0.70, 0.86) | 0.12 |
| Zimbabwe | SHINE (HIV+) (48) | 306 | 285 | 0.65 (0.50, 0.85) | 0.08 |
| **$I^2$ = 0.57, $Tau^2$ = 0.01** | | **6455** | **5129** | **0.83 (0.75, 0.91)** | |

**Supplement duration − > 12m**

| Country | Trial | N | N | PR (95% CI) | W |
|---|---|---|---|---|---|
| Bangladesh | RDNS (36) | 549 | 272 | 0.66 (0.51, 0.84) | 0.23 |
| Bangladesh | WASH−B (37) | 234 | 186 | 0.42 (0.24, 0.74) | 0.09 |
| Kenya | WASH−B (42) | 350 | 300 | 0.68 (0.57, 0.81) | 0.30 |
| Mali | PROMIS CS (46) | 953 | 970 | 0.82 (0.76, 0.88) | 0.38 |
| **$I^2$ = 0.70, $Tau^2$ = 0.03** | | **2086** | **1728** | **0.69 (0.55, 0.85)** | |

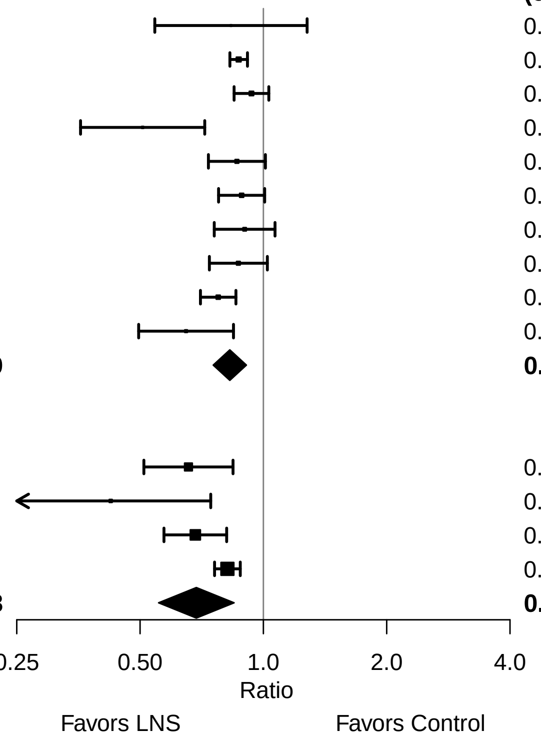

#### Supplemental figure 4B: Anemia prevalence ratio

### 4B8: Stratified by Iron dose

**Iron dose**
**(p−diff = 0.000)**

**Iron dose − Less than 9 mg**

| **Country** | **Trial** | **N** | **N** | **PR (95% CI)** | **W** |
|---|---|---|---|---|---|
| Bangladesh | JiVitA−4 (35) | 457 | 146 | 0.83 (0.54, 1.28) | 0.04 |
| Burkina Faso | iLiNS−Zinc (38) | 1957 | 664 | 0.87 (0.83, 0.91) | 0.13 |
| Burkina Faso | PROMIS CS (39) | 574 | 581 | 0.94 (0.85, 1.03) | 0.12 |
| Ghana | iLiNS−DYADG (41) | 328 | 661 | 0.86 (0.73, 1.01) | 0.10 |
| Madagascar | MAHAY (43) | 600 | 588 | 0.88 (0.78, 1.01) | 0.11 |
| Malawi | iLiNS−DYADM (44) | 210 | 432 | 0.90 (0.76, 1.07) | 0.10 |
| Malawi | iLiNS−DOSE (45) | 243 | 82 | 0.87 (0.74, 1.02) | 0.10 |
| Mali | PROMIS CS (46) | 953 | 970 | 0.82 (0.76, 0.88) | 0.12 |
| Zimbabwe | SHINE (HIV−) (47) | 1682 | 1594 | 0.78 (0.70, 0.86) | 0.12 |
| Zimbabwe | SHINE (HIV+) (48) | 306 | 285 | 0.65 (0.50, 0.85) | 0.07 |
| **I² = 0.35, Tau² = 0.00** | | **7310** | **6003** | **0.85 (0.81, 0.89)** | |

**Iron dose − 9 mg**

| **Country** | **Trial** | **N** | **N** | **PR (95% CI)** | **W** |
|---|---|---|---|---|---|
| Bangladesh | RDNS (36) | 549 | 272 | 0.66 (0.51, 0.84) | 0.30 |
| Bangladesh | WASH−B (37) | 234 | 186 | 0.42 (0.24, 0.74) | 0.11 |
| Ghana | GHANA (40) | 98 | 96 | 0.51 (0.36, 0.72) | 0.21 |
| Kenya | WASH−B (42) | 350 | 300 | 0.68 (0.57, 0.81) | 0.38 |
| **I² = 0.30, Tau² = 0.01** | | **1231** | **854** | **0.61 (0.51, 0.73)** | |

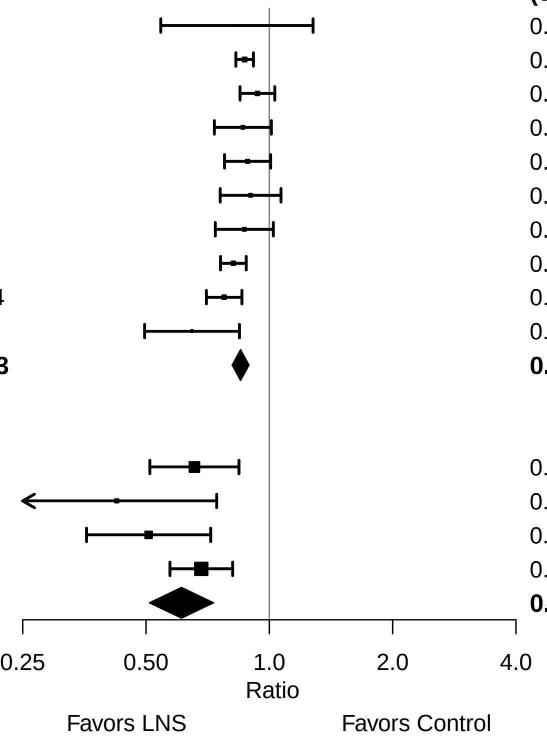

#### Supplemental figure 4B: Anemia prevalence ratio

##### 4B9: Stratified by Frequency of contact

**Frequency of contact**
**(p−diff = 0.714)**

| Country | Trial | N | N | PR (95% CI) | W |
|---|---|---|---|---|---|
| **Frequency of contact − Monthly** | | | | | |
| Bangladesh | RDNS (36) | 549 | 272 | 0.66 (0.51, 0.84) | 0.11 |
| Burkina Faso | PROMIS CS (39) | 574 | 581 | 0.94 (0.85, 1.03) | 0.17 |
| Kenya | WASH−B (42) | 350 | 300 | 0.68 (0.57, 0.81) | 0.14 |
| Madagascar | MAHAY (43) | 600 | 588 | 0.88 (0.78, 1.01) | 0.15 |
| Mali | PROMIS CS (46) | 953 | 970 | 0.82 (0.76, 0.88) | 0.17 |
| Zimbabwe | SHINE (HIV−) (47) | 1682 | 1594 | 0.78 (0.70, 0.86) | 0.16 |
| Zimbabwe | SHINE (HIV+) (48) | 306 | 285 | 0.65 (0.50, 0.85) | 0.10 |
| **I² = 0.70, Tau² = 0.01** | | **5014** | **4590** | **0.79 (0.71, 0.87)** | |
| **Frequency of contact − Weekly** | | | | | |
| Bangladesh | JiVitA−4 (35) | 457 | 146 | 0.83 (0.54, 1.28) | 0.08 |
| Bangladesh | WASH−B (37) | 234 | 186 | 0.42 (0.24, 0.74) | 0.05 |
| Burkina Faso | iLiNS−Zinc (38) | 1957 | 664 | 0.87 (0.83, 0.91) | 0.23 |
| Ghana | GHANA (40) | 98 | 96 | 0.51 (0.36, 0.72) | 0.10 |
| Ghana | iLiNS−DYADG (41) | 328 | 661 | 0.86 (0.73, 1.01) | 0.18 |
| Malawi | iLiNS−DYADM (44) | 210 | 432 | 0.90 (0.76, 1.07) | 0.18 |
| Malawi | iLiNS−DOSE (45) | 243 | 82 | 0.87 (0.74, 1.02) | 0.18 |
| **I² = 0.61, Tau² = 0.05** | | **3527** | **2267** | **0.77 (0.64, 0.94)** | |

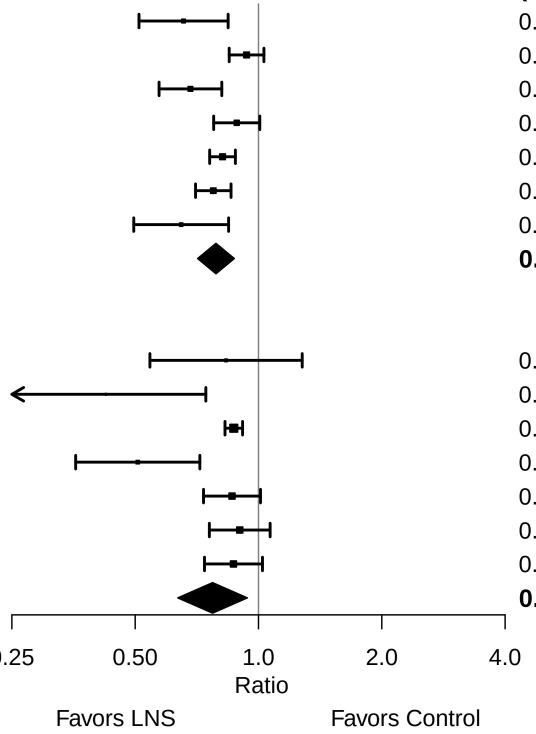

**Supplemental figure 4B: Anemia prevalence ratio**

#### 4B10: Stratified by Average SQ-LNS compliance

**Average SQ−LNS compliance**
**(p−diff = 0.062)**

| Country | Trial | N | N | PR (95% CI) | W |
|---|---|---|---|---|---|
| **Average SQ−LNS compliance − Low** | | | | | |
| Burkina Faso | PROMIS CS (39) | 574 | 581 | 0.94 (0.85, 1.03) | 0.16 |
| Ghana | iLiNS−DYADG (41) | 328 | 661 | 0.86 (0.73, 1.01) | 0.14 |
| Malawi | iLiNS−DYADM (44) | 210 | 432 | 0.90 (0.76, 1.07) | 0.13 |
| Malawi | iLiNS−DOSE (45) | 243 | 82 | 0.87 (0.74, 1.02) | 0.14 |
| Mali | PROMIS CS (46) | 953 | 970 | 0.82 (0.76, 0.88) | 0.17 |
| Zimbabwe | SHINE (HIV−) (47) | 1682 | 1594 | 0.78 (0.70, 0.86) | 0.16 |
| Zimbabwe | SHINE (HIV+) (48) | 306 | 285 | 0.65 (0.50, 0.85) | 0.10 |
| **$I^2$ = 0.51, $Tau^2$ = 0.00** | | **4296** | **4605** | **0.84 (0.78, 0.90)** | |
| **Average SQ−LNS compliance − High** | | | | | |
| Bangladesh | JiVitA−4 (35) | 457 | 146 | 0.83 (0.54, 1.28) | 0.10 |
| Bangladesh | RDNS (36) | 549 | 272 | 0.66 (0.51, 0.84) | 0.18 |
| Bangladesh | WASH−B (37) | 234 | 186 | 0.42 (0.24, 0.74) | 0.07 |
| Burkina Faso | iLiNS−Zinc (38) | 1957 | 664 | 0.87 (0.83, 0.91) | 0.29 |
| Ghana | GHANA (40) | 98 | 96 | 0.51 (0.36, 0.72) | 0.13 |
| Kenya | WASH−B (42) | 350 | 300 | 0.68 (0.57, 0.81) | 0.23 |
| **$I^2$ = 0.80, $Tau^2$ = 0.04** | | **3645** | **1664** | **0.68 (0.56, 0.83)** | |

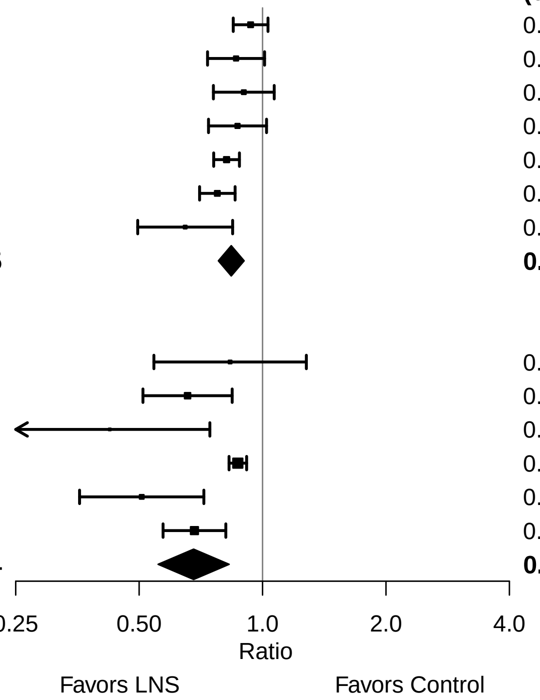

### Supplemental figure 4C: Anemia prevalence difference

#### 4C1: Stratified by Geographic region

**Geographic region**
**(p−diff = 0.590)**

| **Geographic region − SEAR** | | | | **PD** | |
|---|---|---|---|---|---|
| **Country** | **Trial** | **N** | **N** | **(95% CI)** | **W** |
| Bangladesh | JiVitA−4 (35) | 457 | 146 | −0.03 (−0.09, 0.04) | 0.33 |
| Bangladesh | RDNS (36) | 549 | 272 | −0.14 (−0.21, −0.07) | 0.32 |
| Bangladesh | WASH−B (37) | 234 | 186 | −0.09 (−0.16, −0.03) | 0.35 |
| **I² = 0.65, Tau² = 0.00** | | **1240** | **604** | **−0.09 (−0.15, −0.02)** | |
| **Geographic region − AFR** | | | | | |
| Burkina Faso | iLiNS−Zinc (38) | 1957 | 664 | −0.12 (−0.15, −0.09) | 0.13 |
| Burkina Faso | PROMIS CS (39) | 574 | 581 | −0.05 (−0.10, 0.01) | 0.10 |
| Ghana | GHANA (40) | 98 | 96 | −0.29 (−0.42, −0.15) | 0.04 |
| Ghana | iLiNS−DYADG (41) | 328 | 661 | −0.06 (−0.13, 0.00) | 0.09 |
| Kenya | WASH−B (42) | 350 | 300 | −0.15 (−0.23, −0.08) | 0.08 |
| Madagascar | MAHAY (43) | 600 | 588 | −0.07 (−0.13, −0.02) | 0.10 |
| Malawi | iLiNS−DYADM (44) | 210 | 432 | −0.05 (−0.13, 0.03) | 0.07 |
| Malawi | iLiNS−DOSE (45) | 243 | 82 | −0.10 (−0.21, 0.02) | 0.05 |
| Mali | PROMIS CS (46) | 953 | 970 | −0.16 (−0.19, −0.12) | 0.12 |
| Zimbabwe | SHINE (HIV−) (47) | 1682 | 1594 | −0.08 (−0.11, −0.05) | 0.13 |
| Zimbabwe | SHINE (HIV+) (48) | 306 | 285 | −0.13 (−0.20, −0.06) | 0.08 |
| **I² = 0.66, Tau² = 0.00** | | **7301** | **6253** | **−0.11 (−0.14, −0.07)** | |

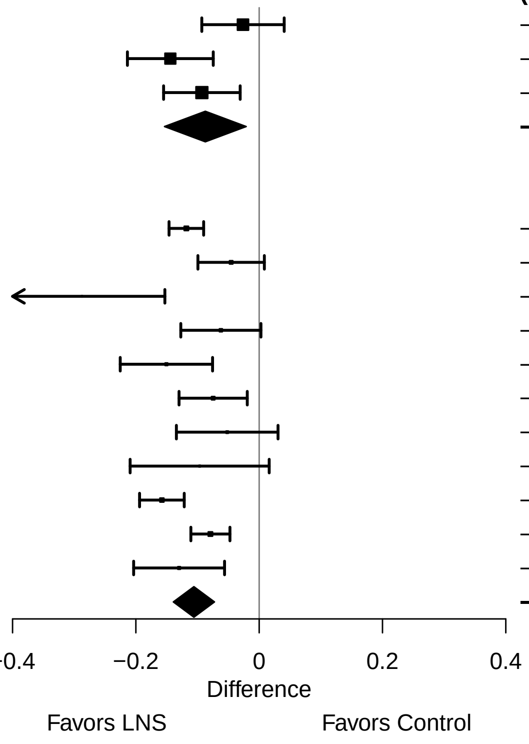

**Supplemental figure 4C: Anemia prevalence difference**

#### 4C2: Stratified by Anemia burden

**Anemia burden**
**(p−diff = 0.407)**

| Country | Trial | N | N | PD (95% CI) | W |
|---|---|---|---|---|---|
| **Anemia burden − Moderate** | | | | | |
| Bangladesh | JiVitA−4 (35) | 457 | 146 | −0.03 (−0.09, 0.04) | 0.12 |
| Bangladesh | RDNS (36) | 549 | 272 | −0.14 (−0.21, −0.07) | 0.11 |
| Bangladesh | WASH−B (37) | 234 | 186 | −0.09 (−0.16, −0.03) | 0.13 |
| Ghana | iLiNS−DYADG (41) | 328 | 661 | −0.06 (−0.13, 0.00) | 0.12 |
| Kenya | WASH−B (42) | 350 | 300 | −0.15 (−0.23, −0.08) | 0.11 |
| Madagascar | MAHAY (43) | 600 | 588 | −0.07 (−0.13, −0.02) | 0.13 |
| Zimbabwe | SHINE (HIV−) (47) | 1682 | 1594 | −0.08 (−0.11, −0.05) | 0.17 |
| Zimbabwe | SHINE (HIV+) (48) | 306 | 285 | −0.13 (−0.20, −0.06) | 0.11 |
| **I² = 0.35, Tau² = 0.00** | | **4506** | **4032** | **−0.09 (−0.12, −0.06)** | |
| **Anemia burden − High** | | | | | |
| Burkina Faso | iLiNS−Zinc (38) | 1957 | 664 | −0.12 (−0.15, −0.09) | 0.25 |
| Burkina Faso | PROMIS CS (39) | 574 | 581 | −0.05 (−0.10, 0.01) | 0.20 |
| Ghana | GHANA (40) | 98 | 96 | −0.29 (−0.42, −0.15) | 0.08 |
| Malawi | iLiNS−DYADM (44) | 210 | 432 | −0.05 (−0.13, 0.03) | 0.14 |
| Malawi | iLiNS−DOSE (45) | 243 | 82 | −0.10 (−0.21, 0.02) | 0.10 |
| Mali | PROMIS CS (46) | 953 | 970 | −0.16 (−0.19, −0.12) | 0.23 |
| **I² = 0.75, Tau² = 0.00** | | **4035** | **2825** | **−0.12 (−0.18, −0.05)** | |

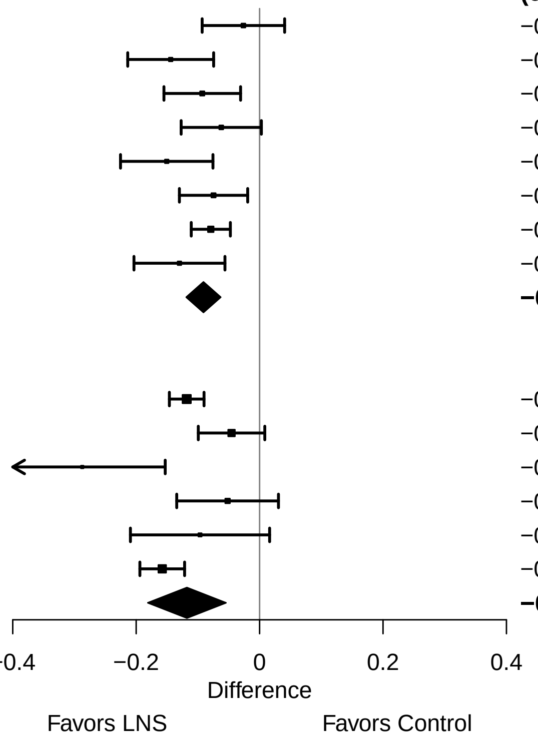

#### Supplemental figure 4C: Anemia prevalence difference

### 4C3: Stratified by Malaria prevalence

**Malaria prevalence**
**(p−diff = 0.702)**

| Country | Trial | N | N | PD (95% CI) | W |
|---|---|---|---|---|---|
| **Malaria prevalence − Less than 10%** | | | | | |
| Bangladesh | JiVitA−4 (35) | 457 | 146 | −0.03 (−0.09, 0.04) | 0.13 |
| Bangladesh | RDNS (36) | 549 | 272 | −0.14 (−0.21, −0.07) | 0.13 |
| Bangladesh | WASH−B (37) | 234 | 186 | −0.09 (−0.16, −0.03) | 0.14 |
| Kenya | WASH−B (42) | 350 | 300 | −0.15 (−0.23, −0.08) | 0.12 |
| Madagascar | MAHAY (43) | 600 | 588 | −0.07 (−0.13, −0.02) | 0.15 |
| Zimbabwe | SHINE (HIV−) (47) | 1682 | 1594 | −0.08 (−0.11, −0.05) | 0.19 |
| Zimbabwe | SHINE (HIV+) (48) | 306 | 285 | −0.13 (−0.20, −0.06) | 0.12 |
| **$I^2$ = 0.41, $Tau^2$ = 0.00** | | **4178** | **3371** | **−0.09 (−0.13, −0.06)** | |
| **Malaria prevalence − At least 10%** | | | | | |
| Burkina Faso | iLiNS−Zinc (38) | 1957 | 664 | −0.12 (−0.15, −0.09) | 0.21 |
| Burkina Faso | PROMIS CS (39) | 574 | 581 | −0.05 (−0.10, 0.01) | 0.17 |
| Ghana | GHANA (40) | 98 | 96 | −0.29 (−0.42, −0.15) | 0.07 |
| Ghana | iLiNS−DYADG (41) | 328 | 661 | −0.06 (−0.13, 0.00) | 0.15 |
| Malawi | iLiNS−DYADM (44) | 210 | 432 | −0.05 (−0.13, 0.03) | 0.12 |
| Malawi | iLiNS−DOSE (45) | 243 | 82 | −0.10 (−0.21, 0.02) | 0.09 |
| Mali | PROMIS CS (46) | 953 | 970 | −0.16 (−0.19, −0.12) | 0.20 |
| **$I^2$ = 0.74, $Tau^2$ = 0.00** | | **4363** | **3486** | **−0.11 (−0.16, −0.05)** | |

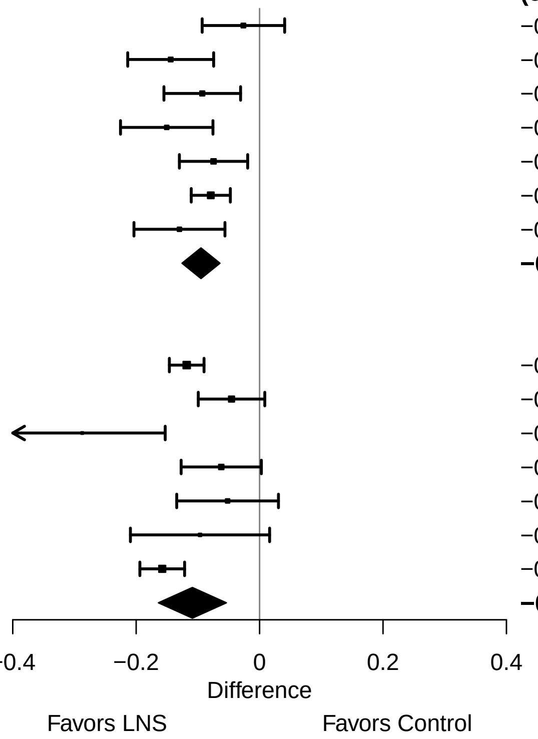

#### Supplemental figure 4C: Anemia prevalence difference

### 4C4: Stratified by Inflammation burden

**Inflammation burden**
**(p−diff = 0.423)**

| Country | Trial | N | N | PD (95% CI) | W |
|---|---|---|---|---|---|
| **Inflammation burden − Low** | | | | | |
| Bangladesh | RDNS (36) | 549 | 272 | −0.14 (−0.21, −0.07) | 0.12 |
| Bangladesh | WASH−B (37) | 234 | 186 | −0.09 (−0.16, −0.03) | 0.13 |
| Ghana | GHANA (40) | 98 | 96 | −0.29 (−0.42, −0.15) | 0.06 |
| Ghana | iLiNS−DYADG (41) | 328 | 661 | −0.06 (−0.13, 0.00) | 0.13 |
| Kenya | WASH−B (42) | 350 | 300 | −0.15 (−0.23, −0.08) | 0.12 |
| Madagascar | MAHAY (43) | 600 | 588 | −0.07 (−0.13, −0.02) | 0.14 |
| Zimbabwe | SHINE (HIV−) (47) | 1682 | 1594 | −0.08 (−0.11, −0.05) | 0.18 |
| Zimbabwe | SHINE (HIV+) (48) | 306 | 285 | −0.13 (−0.20, −0.06) | 0.12 |
| **$I^2$ = 0.54, $Tau^2$ = 0.00** | | **4147** | **3982** | **−0.11 (−0.15, −0.07)** | |
| **Inflammation burden − High** | | | | | |
| Bangladesh | JiVitA−4 (35) | 457 | 146 | −0.03 (−0.09, 0.04) | 0.16 |
| Burkina Faso | iLiNS−Zinc (38) | 1957 | 664 | −0.12 (−0.15, −0.09) | 0.23 |
| Burkina Faso | PROMIS CS (39) | 574 | 581 | −0.05 (−0.10, 0.01) | 0.18 |
| Malawi | iLiNS−DYADM (44) | 210 | 432 | −0.05 (−0.13, 0.03) | 0.13 |
| Malawi | iLiNS−DOSE (45) | 243 | 82 | −0.10 (−0.21, 0.02) | 0.09 |
| Mali | PROMIS CS (46) | 953 | 970 | −0.16 (−0.19, −0.12) | 0.21 |
| **$I^2$ = 0.76, $Tau^2$ = 0.00** | | **4394** | **2875** | **−0.09 (−0.13, −0.05)** | |

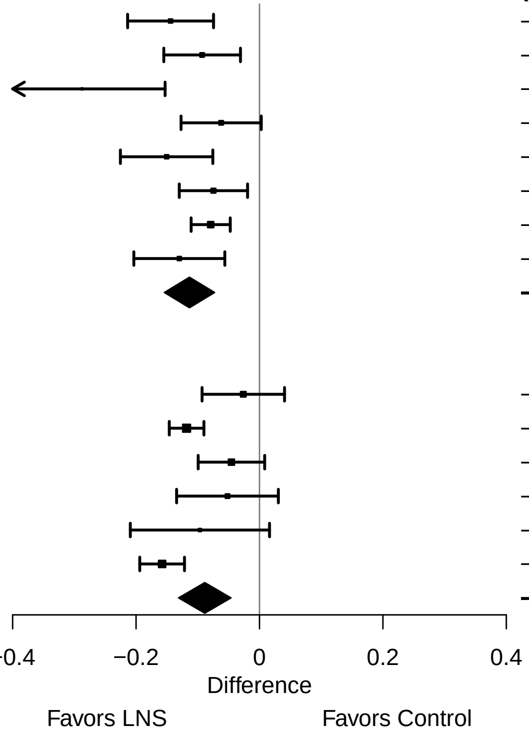

#### Supplemental figure 4C: Anemia prevalence difference

### 4C5: Stratified by Source water quality

**Source water quality**
**(p−diff = 0.943)**

**Source water quality − Improved**

| Country | Trial | N | N | PD (95% CI) | W |
|---|---|---|---|---|---|
| Bangladesh | JiVitA−4 (35) | 457 | 146 | −0.03 (−0.09, 0.04) | 0.18 |
| Bangladesh | RDNS (36) | 549 | 272 | −0.14 (−0.21, −0.07) | 0.18 |
| Bangladesh | WASH−B (37) | 105 | 60 | −0.13 (−0.23, −0.02) | 0.11 |
| Ghana | GHANA (40) | 98 | 96 | −0.29 (−0.42, −0.15) | 0.08 |
| Ghana | iLiNS−DYADG (41) | 328 | 661 | −0.06 (−0.13, 0.00) | 0.19 |
| Malawi | iLiNS−DYADM (44) | 210 | 432 | −0.05 (−0.13, 0.03) | 0.15 |
| Malawi | iLiNS−DOSE (45) | 243 | 82 | −0.10 (−0.21, 0.02) | 0.11 |
| **I² = 0.63, Tau² = 0.00** | | **1990** | **1749** | **−0.10 (−0.16, −0.04)** | |

**Source water quality − Unimproved**

| Country | Trial | N | N | PD (95% CI) | W |
|---|---|---|---|---|---|
| Burkina Faso | iLiNS−Zinc (38) | 1957 | 664 | −0.12 (−0.15, −0.09) | 0.19 |
| Burkina Faso | PROMIS CS (39) | 574 | 581 | −0.05 (−0.10, 0.01) | 0.15 |
| Kenya | WASH−B (42) | 196 | 162 | −0.13 (−0.23, −0.02) | 0.08 |
| Madagascar | MAHAY (43) | 600 | 588 | −0.07 (−0.13, −0.02) | 0.15 |
| Mali | PROMIS CS (46) | 953 | 970 | −0.16 (−0.19, −0.12) | 0.18 |
| Zimbabwe | SHINE (HIV−) (47) | 819 | 795 | −0.08 (−0.13, −0.04) | 0.17 |
| Zimbabwe | SHINE (HIV+) (48) | 133 | 126 | −0.07 (−0.18, 0.04) | 0.08 |
| **I² = 0.62, Tau² = 0.00** | | **5232** | **3886** | **−0.10 (−0.13, −0.07)** | |

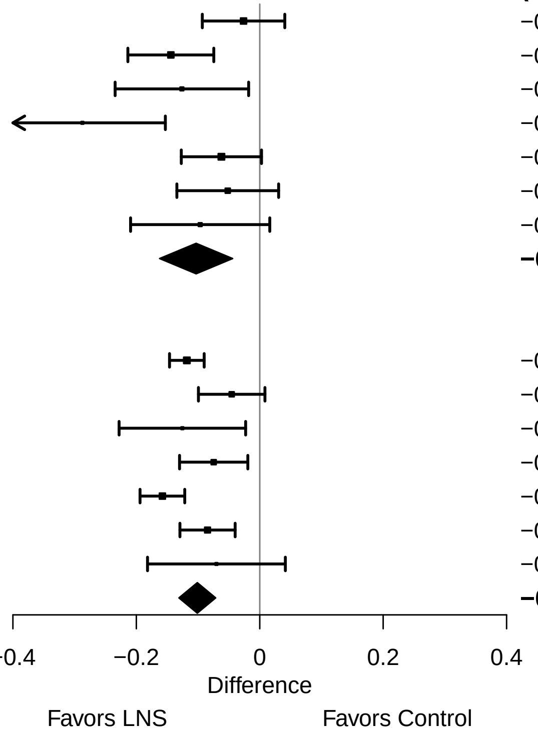

#### Supplemental figure 4C: Anemia prevalence difference

### 4C6: Stratified by Sanitation

**Sanitation**
**(p−diff = 0.565)**

| Country | Trial | N | N | PD (95% CI) | W |
|---|---|---|---|---|---|
| **Sanitation − Improved** | | | | | |
| Bangladesh | JiVitA−4 (35) | 457 | 146 | −0.03 (−0.09, 0.04) | 0.15 |
| Bangladesh | RDNS (36) | 549 | 272 | −0.14 (−0.21, −0.07) | 0.15 |
| Bangladesh | WASH−B (37) | 105 | 60 | −0.13 (−0.23, −0.02) | 0.09 |
| Burkina Faso | PROMIS CS (39) | 574 | 581 | −0.05 (−0.10, 0.01) | 0.18 |
| Ghana | GHANA (40) | 98 | 96 | −0.29 (−0.42, −0.15) | 0.07 |
| Ghana | iLiNS−DYADG (41) | 328 | 661 | −0.06 (−0.13, 0.00) | 0.15 |
| Mali | PROMIS CS (46) | 953 | 970 | −0.16 (−0.19, −0.12) | 0.21 |
| **I² = 0.79, Tau² = 0.01** | | **3064** | **2786** | **−0.11 (−0.17, −0.05)** | |
| **Sanitation − Unimproved** | | | | | |
| Burkina Faso | iLiNS−Zinc (38) | 1957 | 664 | −0.12 (−0.15, −0.09) | 0.23 |
| Kenya | WASH−B (42) | 196 | 162 | −0.13 (−0.23, −0.02) | 0.10 |
| Madagascar | MAHAY (43) | 600 | 588 | −0.07 (−0.13, −0.02) | 0.17 |
| Malawi | iLiNS−DYADM (44) | 210 | 432 | −0.05 (−0.13, 0.03) | 0.13 |
| Malawi | iLiNS−DOSE (45) | 243 | 82 | −0.10 (−0.21, 0.02) | 0.09 |
| Zimbabwe | SHINE (HIV−) (47) | 819 | 795 | −0.08 (−0.13, −0.04) | 0.20 |
| Zimbabwe | SHINE (HIV+) (48) | 133 | 126 | −0.07 (−0.18, 0.04) | 0.09 |
| **I² = 0.00, Tau² = 0.00** | | **4158** | **2849** | **−0.10 (−0.12, −0.08)** | |

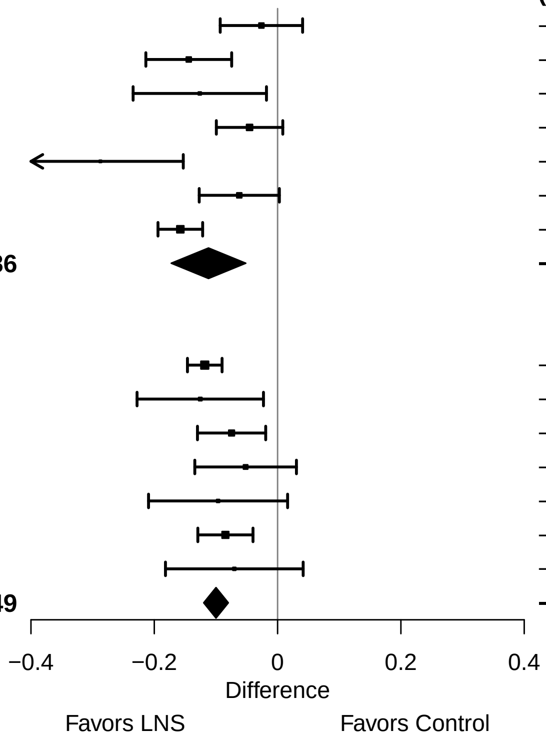

#### Supplemental figure 4C: Anemia prevalence difference

### 4C7: Stratified by Supplement duration

**Supplement duration**
**(p−diff = 0.018)**

| Country | Trial | N | N | PD (95% CI) | W |
|---|---|---|---|---|---|
| **Supplement duration − 12m or less** | | | | | |
| Bangladesh | JiVitA−4 (35) | 457 | 146 | −0.03 (−0.09, 0.04) | 0.10 |
| Burkina Faso | iLiNS−Zinc (38) | 1957 | 664 | −0.12 (−0.15, −0.09) | 0.15 |
| Burkina Faso | PROMIS CS (39) | 574 | 581 | −0.05 (−0.10, 0.01) | 0.12 |
| Ghana | GHANA (40) | 98 | 96 | −0.29 (−0.42, −0.15) | 0.05 |
| Ghana | iLiNS−DYADG (41) | 328 | 661 | −0.06 (−0.13, 0.00) | 0.10 |
| Madagascar | MAHAY (43) | 600 | 588 | −0.07 (−0.13, −0.02) | 0.11 |
| Malawi | iLiNS−DYADM (44) | 210 | 432 | −0.05 (−0.13, 0.03) | 0.08 |
| Malawi | iLiNS−DOSE (45) | 243 | 82 | −0.10 (−0.21, 0.02) | 0.06 |
| Zimbabwe | SHINE (HIV−) (47) | 1682 | 1594 | −0.08 (−0.11, −0.05) | 0.14 |
| Zimbabwe | SHINE (HIV+) (48) | 306 | 285 | −0.13 (−0.20, −0.06) | 0.09 |
| **I² = 0.58, Tau² = 0.00** | | **6455** | **5129** | **−0.09 (−0.12, −0.05)** | |
| **Supplement duration − > 12m** | | | | | |
| Bangladesh | RDNS (36) | 549 | 272 | −0.14 (−0.21, −0.07) | 0.23 |
| Bangladesh | WASH−B (37) | 234 | 186 | −0.09 (−0.16, −0.03) | 0.25 |
| Kenya | WASH−B (42) | 350 | 300 | −0.15 (−0.23, −0.08) | 0.21 |
| Mali | PROMIS CS (46) | 953 | 970 | −0.16 (−0.19, −0.12) | 0.32 |
| **I² = 0.06, Tau² = 0.00** | | **2086** | **1728** | **−0.14 (−0.17, −0.12)** | |

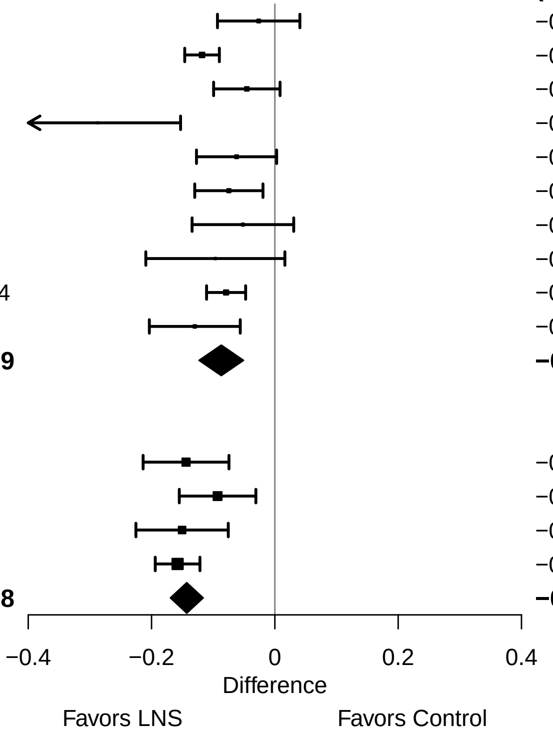

#### Supplemental figure 4C: Anemia prevalence difference

### 4C8: Stratified by Iron dose

**Iron dose**
**(p−diff = 0.055)**

| Country | Trial | N | N | PD (95% CI) | W |
|---|---|---|---|---|---|
| **Iron dose − Less than 9 mg** | | | | | |
| Bangladesh | JiVitA−4 (35) | 457 | 146 | −0.03 (−0.09, 0.04) | 0.09 |
| Burkina Faso | iLiNS−Zinc (38) | 1957 | 664 | −0.12 (−0.15, −0.09) | 0.13 |
| Burkina Faso | PROMIS CS (39) | 574 | 581 | −0.05 (−0.10, 0.01) | 0.11 |
| Ghana | iLiNS−DYADG (41) | 328 | 661 | −0.06 (−0.13, 0.00) | 0.09 |
| Madagascar | MAHAY (43) | 600 | 588 | −0.07 (−0.13, −0.02) | 0.10 |
| Malawi | iLiNS−DYADM (44) | 210 | 432 | −0.05 (−0.13, 0.03) | 0.08 |
| Malawi | iLiNS−DOSE (45) | 243 | 82 | −0.10 (−0.21, 0.02) | 0.05 |
| Mali | PROMIS CS (46) | 953 | 970 | −0.16 (−0.19, −0.12) | 0.13 |
| Zimbabwe | SHINE (HIV−) (47) | 1682 | 1594 | −0.08 (−0.11, −0.05) | 0.13 |
| Zimbabwe | SHINE (HIV+) (48) | 306 | 285 | −0.13 (−0.20, −0.06) | 0.08 |
| **I² = 0.65, Tau² = 0.00** | | **7310** | **6003** | **−0.09 (−0.12, −0.06)** | |
| **Iron dose − 9 mg** | | | | | |
| Bangladesh | RDNS (36) | 549 | 272 | −0.14 (−0.21, −0.07) | 0.29 |
| Bangladesh | WASH−B (37) | 234 | 186 | −0.09 (−0.16, −0.03) | 0.31 |
| Ghana | GHANA (40) | 98 | 96 | −0.29 (−0.42, −0.15) | 0.14 |
| Kenya | WASH−B (42) | 350 | 300 | −0.15 (−0.23, −0.08) | 0.27 |
| **I² = 0.57, Tau² = 0.00** | | **1231** | **854** | **−0.15 (−0.23, −0.08)** | |

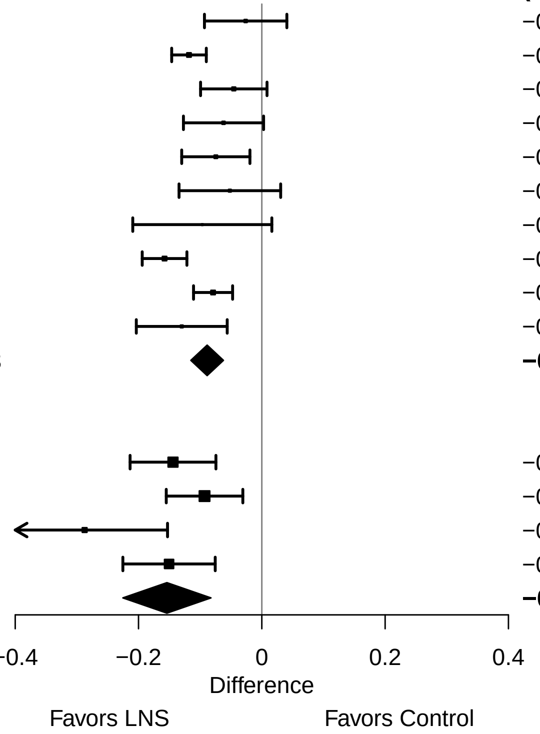

#### Supplemental figure 4C: Anemia prevalence difference

### 4C9: Stratified by Frequency of contact

**Frequency of contact**
**(p−diff = 0.524)**

**Frequency of contact − Monthly**

| Country | Trial | N | N | PD (95% CI) | W |
|---|---|---|---|---|---|
| Bangladesh | RDNS (36) | 549 | 272 | −0.14 (−0.21, −0.07) | 0.12 |
| Burkina Faso | PROMIS CS (39) | 574 | 581 | −0.05 (−0.10, 0.01) | 0.15 |
| Kenya | WASH−B (42) | 350 | 300 | −0.15 (−0.23, −0.08) | 0.12 |
| Madagascar | MAHAY (43) | 600 | 588 | −0.07 (−0.13, −0.02) | 0.14 |
| Mali | PROMIS CS (46) | 953 | 970 | −0.16 (−0.19, −0.12) | 0.17 |
| Zimbabwe | SHINE (HIV−) (47) | 1682 | 1594 | −0.08 (−0.11, −0.05) | 0.18 |
| Zimbabwe | SHINE (HIV+) (48) | 306 | 285 | −0.13 (−0.20, −0.06) | 0.12 |
| **I² = 0.69, Tau² = 0.00** | | **5014** | **4590** | **−0.11 (−0.14, −0.08)** | |

**Frequency of contact − Weekly**

| Country | Trial | N | N | PD (95% CI) | W |
|---|---|---|---|---|---|
| Bangladesh | JiVitA−4 (35) | 457 | 146 | −0.03 (−0.09, 0.04) | 0.16 |
| Bangladesh | WASH−B (37) | 234 | 186 | −0.09 (−0.16, −0.03) | 0.16 |
| Burkina Faso | iLiNS−Zinc (38) | 1957 | 664 | −0.12 (−0.15, −0.09) | 0.23 |
| Ghana | GHANA (40) | 98 | 96 | −0.29 (−0.42, −0.15) | 0.07 |
| Ghana | iLiNS−DYADG (41) | 328 | 661 | −0.06 (−0.13, 0.00) | 0.16 |
| Malawi | iLiNS−DYADM (44) | 210 | 432 | −0.05 (−0.13, 0.03) | 0.13 |
| Malawi | iLiNS−DOSE (45) | 243 | 82 | −0.10 (−0.21, 0.02) | 0.09 |
| **I² = 0.63, Tau² = 0.00** | | **3527** | **2267** | **−0.09 (−0.15, −0.04)** | |

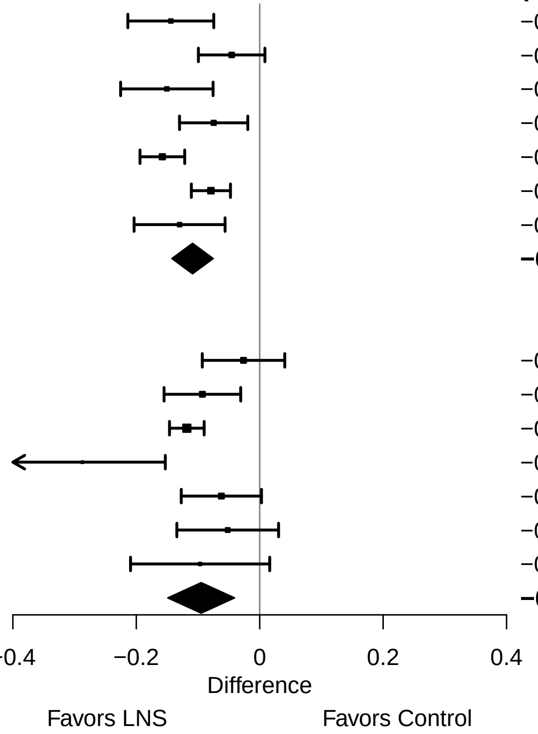

#### Supplemental figure 4C: Anemia prevalence difference

#### 4C10: Stratified by Average SQ-LNS compliance

**Average SQ−LNS compliance**
**(p−diff = 0.327)**

| Country | Trial | N | N | PD (95% CI) | W |
|---|---|---|---|---|---|
| **Average SQ−LNS compliance − Low** | | | | | |
| Burkina Faso | PROMIS CS (39) | 574 | 581 | −0.05 (−0.10, 0.01) | 0.16 |
| Ghana | iLiNS−DYADG (41) | 328 | 661 | −0.06 (−0.13, 0.00) | 0.14 |
| Malawi | iLiNS−DYADM (44) | 210 | 432 | −0.05 (−0.13, 0.03) | 0.12 |
| Malawi | iLiNS−DOSE (45) | 243 | 82 | −0.10 (−0.21, 0.02) | 0.08 |
| Mali | PROMIS CS (46) | 953 | 970 | −0.16 (−0.19, −0.12) | 0.18 |
| Zimbabwe | SHINE (HIV−) (47) | 1682 | 1594 | −0.08 (−0.11, −0.05) | 0.19 |
| Zimbabwe | SHINE (HIV+) (48) | 306 | 285 | −0.13 (−0.20, −0.06) | 0.13 |
| **I² = 0.68, Tau² = 0.00** | | **4296** | **4605** | **−0.09 (−0.13, −0.06)** | |
| **Average SQ−LNS compliance − High** | | | | | |
| Bangladesh | JiVitA−4 (35) | 457 | 146 | −0.03 (−0.09, 0.04) | 0.17 |
| Bangladesh | RDNS (36) | 549 | 272 | −0.14 (−0.21, −0.07) | 0.17 |
| Bangladesh | WASH−B (37) | 234 | 186 | −0.09 (−0.16, −0.03) | 0.18 |
| Burkina Faso | iLiNS−Zinc (38) | 1957 | 664 | −0.12 (−0.15, −0.09) | 0.24 |
| Ghana | GHANA (40) | 98 | 96 | −0.29 (−0.42, −0.15) | 0.08 |
| Kenya | WASH−B (42) | 350 | 300 | −0.15 (−0.23, −0.08) | 0.16 |
| **I² = 0.67, Tau² = 0.00** | | **3645** | **1664** | **−0.13 (−0.19, −0.06)** | |

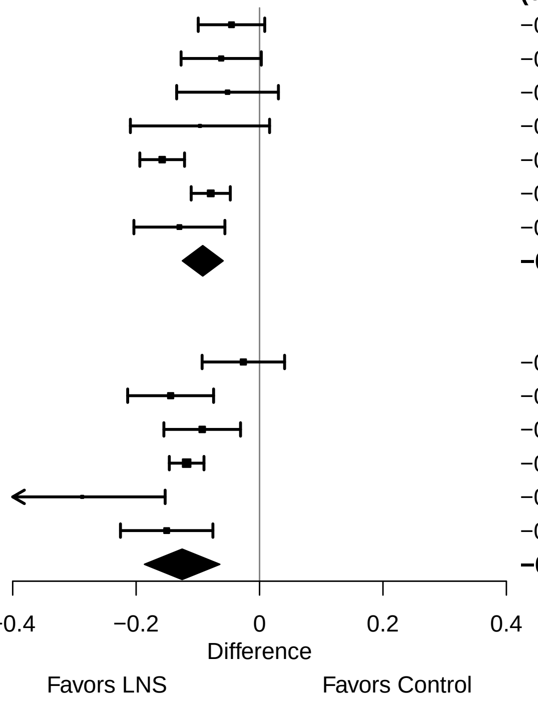

### Supplemental figure 4D: Moderate-to-severe anemia prevalence ratio

#### 4D1: Stratified by Geographic region (insufficient comparisons)

#### Supplemental figure 4D: Moderate-to-severe anemia prevalence ratio

### 4D2: Stratified by Anemia burden

**Anemia burden**
**(p−diff = 0.388)**

| Country | Trial | N | N | PR (95% CI) | W |
|---|---|---|---|---|---|
| **Anemia burden − Moderate** | | | | | |
| Bangladesh | JiVitA−4 (35) | | | | |
| Bangladesh | RDNS (36) | 549 | 272 | 0.54 (0.38, 0.76) | 0.17 |
| Bangladesh | WASH−B (37) | | | | |
| Ghana | iLiNS−DYADG (41) | 328 | 661 | 0.84 (0.47, 1.51) | 0.10 |
| Kenya | WASH−B (42) | 350 | 300 | 0.63 (0.45, 0.88) | 0.18 |
| Madagascar | MAHAY (43) | 600 | 588 | 0.81 (0.64, 1.03) | 0.22 |
| Zimbabwe | SHINE (HIV−) (47) | 1682 | 1594 | 0.67 (0.53, 0.84) | 0.22 |
| Zimbabwe | SHINE (HIV+) (48) | 306 | 285 | 0.51 (0.30, 0.89) | 0.11 |
| **$I^2$ = 0.10, $Tau^2$ = 0.00** | | **3815** | **3700** | **0.68 (0.59, 0.77)** | |
| **Anemia burden − High** | | | | | |
| Burkina Faso | iLiNS−Zinc (38) | 1957 | 664 | 0.72 (0.65, 0.78) | 0.22 |
| Burkina Faso | PROMIS CS (39) | 574 | 581 | 0.88 (0.73, 1.07) | 0.19 |
| Ghana | GHANA (40) | 98 | 96 | 0.32 (0.16, 0.61) | 0.07 |
| Malawi | iLiNS−DYADM (44) | 210 | 432 | 1.15 (0.87, 1.52) | 0.16 |
| Malawi | iLiNS−DOSE (45) | 243 | 82 | 0.72 (0.54, 0.96) | 0.16 |
| Mali | PROMIS CS (46) | 953 | 970 | 0.63 (0.54, 0.74) | 0.20 |
| **$I^2$ = 0.79, $Tau^2$ = 0.12** | | **4035** | **2825** | **0.73 (0.54, 0.98)** | |

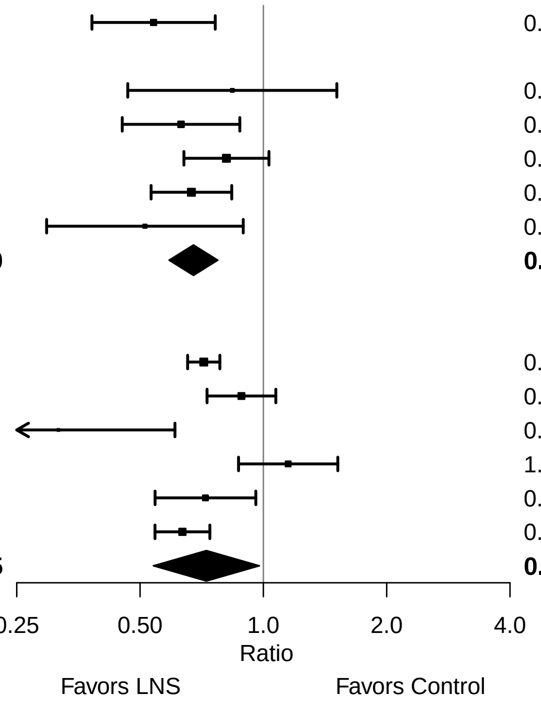

#### Supplemental figure 4D: Moderate-to-severe anemia prevalence ratio

### 4D3: Stratified by Malaria prevalence

**Malaria prevalence**
**(p−diff = 0.282)**

**Malaria prevalence − Less than 10%**

| Country | Trial | N | N | PR (95% CI) | W |
|---|---|---|---|---|---|
| Bangladesh | JiVitA−4 (35) | | | | |
| Bangladesh | RDNS (36) | 549 | 272 | 0.54 (0.38, 0.76) | 0.19 |
| Bangladesh | WASH−B (37) | | | | |
| Kenya | WASH−B (42) | 350 | 300 | 0.63 (0.45, 0.88) | 0.20 |
| Madagascar | MAHAY (43) | 600 | 588 | 0.81 (0.64, 1.03) | 0.24 |
| Zimbabwe | SHINE (HIV−) (47) | 1682 | 1594 | 0.67 (0.53, 0.84) | 0.25 |
| Zimbabwe | SHINE (HIV+) (48) | 306 | 285 | 0.51 (0.30, 0.89) | 0.12 |
| **I² = 0.20, Tau² = 0.01** | | **3487** | **3039** | **0.66 (0.57, 0.77)** | |

**Malaria prevalence − At least 10%**

| Country | Trial | N | N | PR (95% CI) | W |
|---|---|---|---|---|---|
| Burkina Faso | iLiNS−Zinc (38) | 1957 | 664 | 0.72 (0.65, 0.78) | 0.20 |
| Burkina Faso | PROMIS CS (39) | 574 | 581 | 0.88 (0.73, 1.07) | 0.18 |
| Ghana | GHANA (40) | 98 | 96 | 0.32 (0.16, 0.61) | 0.06 |
| Ghana | iLiNS−DYADG (41) | 328 | 661 | 0.84 (0.47, 1.51) | 0.07 |
| Malawi | iLiNS−DYADM (44) | 210 | 432 | 1.15 (0.87, 1.52) | 0.15 |
| Malawi | iLiNS−DOSE (45) | 243 | 82 | 0.72 (0.54, 0.96) | 0.15 |
| Mali | PROMIS CS (46) | 953 | 970 | 0.63 (0.54, 0.74) | 0.19 |
| **I² = 0.75, Tau² = 0.09** | | **4363** | **3486** | **0.74 (0.57, 0.95)** | |

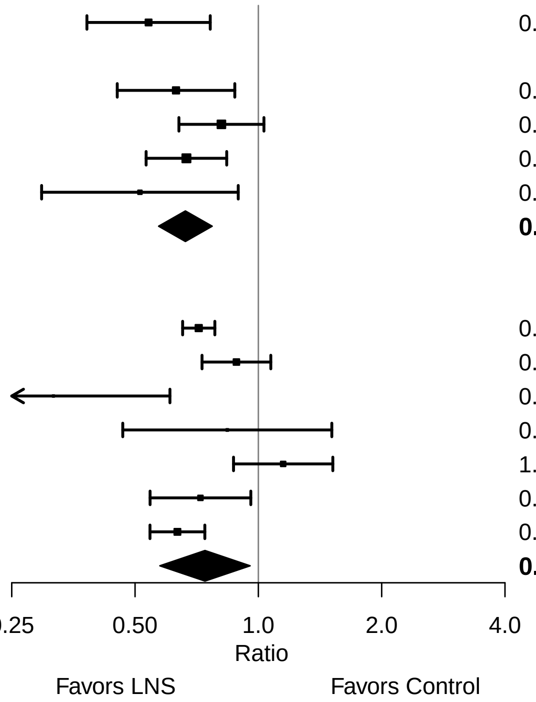

#### Supplemental figure 4D: Moderate-to-severe anemia prevalence ratio

### 4D4: Stratified by Inflammation burden

**Inflammation burden**
**(p−diff = 0.100)**

| Country | Trial | N | N | PR (95% CI) | W |
|---|---|---|---|---|---|
| **Inflammation burden − Low** | | | | | |
| Bangladesh | RDNS (36) | 549 | 272 | 0.54 (0.38, 0.76) | 0.16 |
| Bangladesh | WASH−B (37) | | | | |
| Ghana | GHANA (40) | 98 | 96 | 0.32 (0.16, 0.61) | 0.08 |
| Ghana | iLiNS−DYADG (41) | 328 | 661 | 0.84 (0.47, 1.51) | 0.09 |
| Kenya | WASH−B (42) | 350 | 300 | 0.63 (0.45, 0.88) | 0.17 |
| Madagascar | MAHAY (43) | 600 | 588 | 0.81 (0.64, 1.03) | 0.20 |
| Zimbabwe | SHINE (HIV−) (47) | 1682 | 1594 | 0.67 (0.53, 0.84) | 0.21 |
| Zimbabwe | SHINE (HIV+) (48) | 306 | 285 | 0.51 (0.30, 0.89) | 0.10 |
| **I² = 0.43, Tau² = 0.03** | | **3913** | **3796** | **0.63 (0.51, 0.77)** | |
| **Inflammation burden − High** | | | | | |
| Bangladesh | JiVitA−4 (35) | | | | |
| Burkina Faso | iLiNS−Zinc (38) | 1957 | 664 | 0.72 (0.65, 0.78) | 0.23 |
| Burkina Faso | PROMIS CS (39) | 574 | 581 | 0.88 (0.73, 1.07) | 0.20 |
| Malawi | iLiNS−DYADM (44) | 210 | 432 | 1.15 (0.87, 1.52) | 0.17 |
| Malawi | iLiNS−DOSE (45) | 243 | 82 | 0.72 (0.54, 0.96) | 0.17 |
| Mali | PROMIS CS (46) | 953 | 970 | 0.63 (0.54, 0.74) | 0.22 |
| **I² = 0.77, Tau² = 0.04** | | **3937** | **2729** | **0.79 (0.65, 0.96)** | |

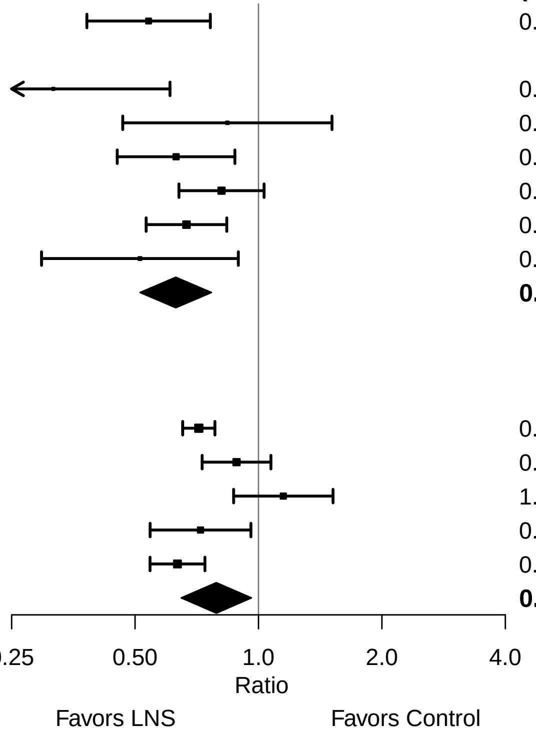

#### Supplemental figure 4D: Moderate-to-severe anemia prevalence ratio

### 4D5: Stratified by Source water quality

**Source water quality**
**(p−diff = 0.934)**

| Country | Trial | N | N | PR (95% CI) | W |
|---|---|---|---|---|---|
| **Source water quality − Improved** | | | | | |
| Bangladesh | JiVitA−4 (35) | | | | |
| Bangladesh | RDNS (36) | 549 | 272 | 0.54 (0.38, 0.76) | 0.23 |
| Bangladesh | WASH−B (37) | | | | |
| Ghana | GHANA (40) | 98 | 96 | 0.32 (0.16, 0.61) | 0.11 |
| Ghana | iLiNS−DYADG (41) | 328 | 661 | 0.84 (0.47, 1.51) | 0.13 |
| Malawi | iLiNS−DYADM (44) | 210 | 432 | 1.15 (0.87, 1.52) | 0.26 |
| Malawi | iLiNS−DOSE (45) | 243 | 82 | 0.72 (0.54, 0.96) | 0.26 |
| **I² = 0.79, Tau² = 0.17** | | **1428** | **1543** | **0.68 (0.45, 1.02)** | |
| **Source water quality − Unimproved** | | | | | |
| Burkina Faso | iLiNS−Zinc (38) | 1957 | 664 | 0.72 (0.65, 0.78) | 0.20 |
| Burkina Faso | PROMIS CS (39) | 574 | 581 | 0.88 (0.73, 1.07) | 0.18 |
| Kenya | WASH−B (42) | 196 | 162 | 0.80 (0.50, 1.27) | 0.10 |
| Madagascar | MAHAY (43) | 600 | 588 | 0.81 (0.64, 1.03) | 0.16 |
| Mali | PROMIS CS (46) | 953 | 970 | 0.63 (0.54, 0.74) | 0.19 |
| Zimbabwe | SHINE (HIV−) (47) | 819 | 795 | 0.59 (0.43, 0.81) | 0.14 |
| Zimbabwe | SHINE (HIV+) (48) | 133 | 126 | 0.52 (0.20, 1.30) | 0.04 |
| **I² = 0.41, Tau² = 0.01** | | **5232** | **3886** | **0.72 (0.65, 0.81)** | |

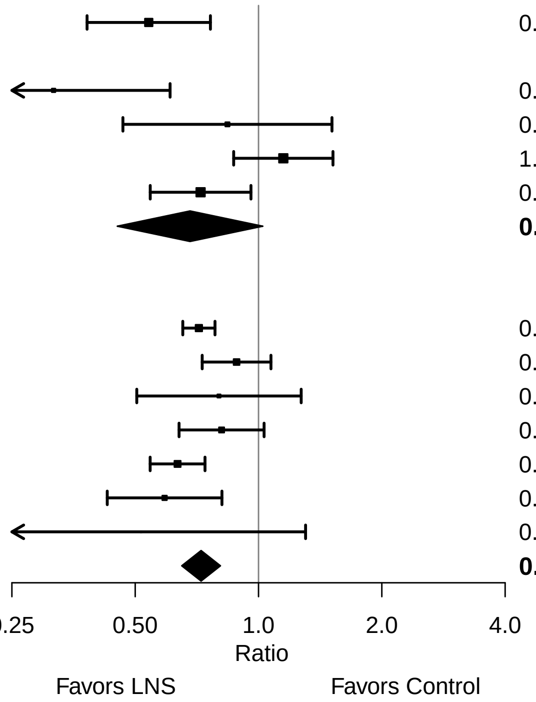

#### Supplemental figure 4D: Moderate-to-severe anemia prevalence ratio

### 4D6: Stratified by Sanitation

**Sanitation**
**(p−diff = 0.290)**

| Country | Trial | N | N | PR (95% CI) | W |
|---|---|---|---|---|---|
| **Sanitation − Improved** | | | | | |
| Bangladesh | JiVitA−4 (35) | | | | |
| Bangladesh | RDNS (36) | 549 | 272 | 0.54 (0.38, 0.76) | 0.20 |
| Bangladesh | WASH−B (37) | | | | |
| Burkina Faso | PROMIS CS (39) | 574 | 581 | 0.88 (0.73, 1.07) | 0.28 |
| Ghana | GHANA (40) | 98 | 96 | 0.32 (0.16, 0.61) | 0.10 |
| Ghana | iLiNS−DYADG (41) | 328 | 661 | 0.84 (0.47, 1.51) | 0.12 |
| Mali | PROMIS CS (46) | 953 | 970 | 0.63 (0.54, 0.74) | 0.30 |
| **I² = 0.74, Tau² = 0.10** | | **2502** | **2580** | **0.63 (0.46, 0.88)** | |
| **Sanitation − Unimproved** | | | | | |
| Burkina Faso | iLiNS−Zinc (38) | 1957 | 664 | 0.72 (0.65, 0.78) | 0.22 |
| Kenya | WASH−B (42) | 196 | 162 | 0.80 (0.50, 1.27) | 0.11 |
| Madagascar | MAHAY (43) | 600 | 588 | 0.81 (0.64, 1.03) | 0.17 |
| Malawi | iLiNS−DYADM (44) | 210 | 432 | 1.15 (0.87, 1.52) | 0.16 |
| Malawi | iLiNS−DOSE (45) | 243 | 82 | 0.72 (0.54, 0.96) | 0.16 |
| Zimbabwe | SHINE (HIV−) (47) | 819 | 795 | 0.59 (0.43, 0.81) | 0.15 |
| Zimbabwe | SHINE (HIV+) (48) | 133 | 126 | 0.52 (0.20, 1.30) | 0.04 |
| **I² = 0.55, Tau² = 0.02** | | **4158** | **2849** | **0.77 (0.65, 0.91)** | |

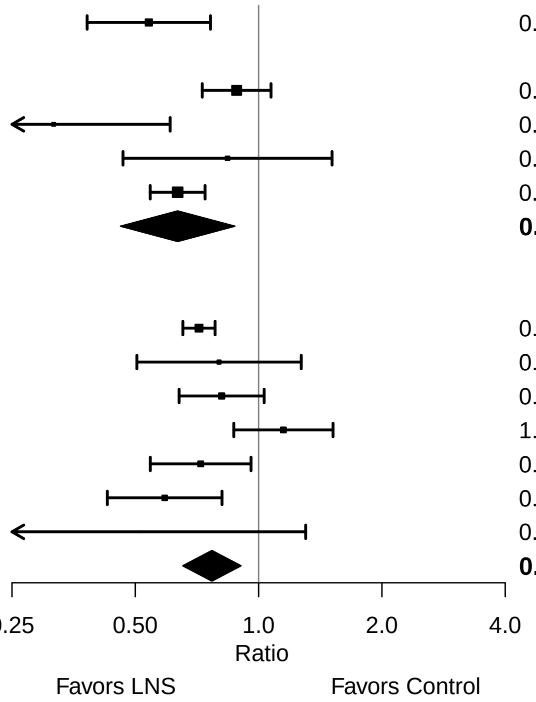

**Supplemental figure 4D: Moderate-to-severe anemia prevalence ratio**

#### 4D7: Stratified by Supplement duration

**Supplement duration**
**(p−diff = 0.088)**

| Country | Trial | N | N | PR (95% CI) | W |
|---|---|---|---|---|---|
| **Supplement duration − 12m or less** | | | | | |
| Bangladesh | JiVitA−4 (35) | | | | |
| Burkina Faso | iLiNS−Zinc (38) | 1957 | 664 | 0.72 (0.65, 0.78) | 0.17 |
| Burkina Faso | PROMIS CS (39) | 574 | 581 | 0.88 (0.73, 1.07) | 0.14 |
| Ghana | GHANA (40) | 98 | 96 | 0.32 (0.16, 0.61) | 0.05 |
| Ghana | iLiNS−DYADG (41) | 328 | 661 | 0.84 (0.47, 1.51) | 0.06 |
| Madagascar | MAHAY (43) | 600 | 588 | 0.81 (0.64, 1.03) | 0.13 |
| Malawi | iLiNS−DYADM (44) | 210 | 432 | 1.15 (0.87, 1.52) | 0.12 |
| Malawi | iLiNS−DOSE (45) | 243 | 82 | 0.72 (0.54, 0.96) | 0.12 |
| Zimbabwe | SHINE (HIV−) (47) | 1682 | 1594 | 0.67 (0.53, 0.84) | 0.14 |
| Zimbabwe | SHINE (HIV+) (48) | 306 | 285 | 0.51 (0.30, 0.89) | 0.07 |
| **I² = 0.65, Tau² = 0.07** | | **5998** | **4983** | **0.74 (0.61, 0.91)** | |
| **Supplement duration − > 12m** | | | | | |
| Bangladesh | RDNS (36) | 549 | 272 | 0.54 (0.38, 0.76) | 0.29 |
| Bangladesh | WASH−B (37) | | | | |
| Kenya | WASH−B (42) | 350 | 300 | 0.63 (0.45, 0.88) | 0.30 |
| Mali | PROMIS CS (46) | 953 | 970 | 0.63 (0.54, 0.74) | 0.42 |
| **I² = 0.00, Tau² = 0.00** | | **1852** | **1542** | **0.62 (0.54, 0.71)** | |

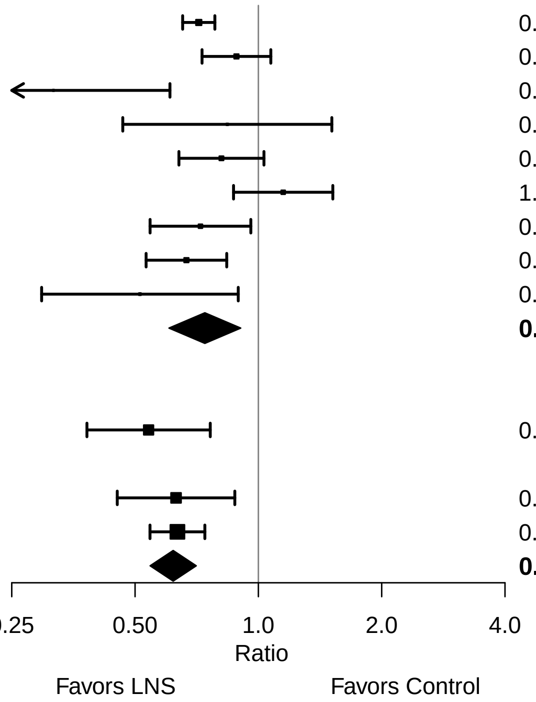

##### Supplemental figure 4D: Moderate-to-severe anemia prevalence ratio

#### 4D8: Stratified by Iron dose

**Iron dose**
**(p−diff = 0.025)**

| Country | Trial | N | N | PR (95% CI) | W |
|---|---|---|---|---|---|
| **Iron dose − Less than 9 mg** | | | | | |
| Bangladesh | JiVitA−4 (35) | | | | |
| Burkina Faso | iLiNS−Zinc (38) | 1957 | 664 | 0.72 (0.65, 0.78) | 0.15 |
| Burkina Faso | PROMIS CS (39) | 574 | 581 | 0.88 (0.73, 1.07) | 0.13 |
| Ghana | iLiNS−DYADG (41) | 328 | 661 | 0.84 (0.47, 1.51) | 0.06 |
| Madagascar | MAHAY (43) | 600 | 588 | 0.81 (0.64, 1.03) | 0.12 |
| Malawi | iLiNS−DYADM (44) | 210 | 432 | 1.15 (0.87, 1.52) | 0.11 |
| Malawi | iLiNS−DOSE (45) | 243 | 82 | 0.72 (0.54, 0.96) | 0.11 |
| Mali | PROMIS CS (46) | 953 | 970 | 0.63 (0.54, 0.74) | 0.14 |
| Zimbabwe | SHINE (HIV−) (47) | 1682 | 1594 | 0.67 (0.53, 0.84) | 0.12 |
| Zimbabwe | SHINE (HIV+) (48) | 306 | 285 | 0.51 (0.30, 0.89) | 0.06 |
| **I² = 0.61, Tau² = 0.02** | | **6853** | **5857** | **0.76 (0.66, 0.87)** | |
| **Iron dose − 9 mg** | | | | | |
| Bangladesh | RDNS (36) | 549 | 272 | 0.54 (0.38, 0.76) | 0.39 |
| Bangladesh | WASH−B (37) | | | | |
| Ghana | GHANA (40) | 98 | 96 | 0.32 (0.16, 0.61) | 0.20 |
| Kenya | WASH−B (42) | 350 | 300 | 0.63 (0.45, 0.88) | 0.41 |
| **I² = 0.41, Tau² = 0.05** | | **997** | **668** | **0.52 (0.36, 0.73)** | |

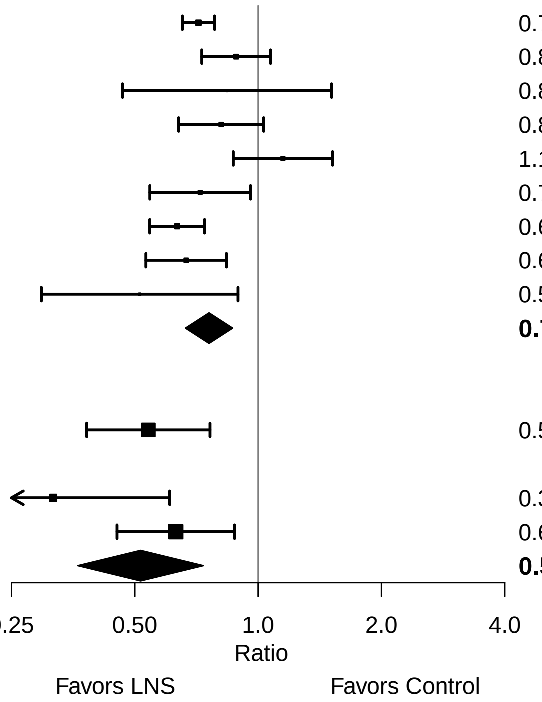

#### Supplemental figure 4D: Moderate-to-severe anemia prevalence ratio

### 4D9: Stratified by Frequency of contact

**Frequency of contact**
**(p−diff = 0.459)**

| Country | Trial | N | N | PR (95% CI) | W |
|---|---|---|---|---|---|
| **Frequency of contact − Monthly** | | | | | |
| Bangladesh | RDNS (36) | 549 | 272 | 0.54 (0.38, 0.76) | 0.12 |
| Burkina Faso | PROMIS CS (39) | 574 | 581 | 0.88 (0.73, 1.07) | 0.17 |
| Kenya | WASH−B (42) | 350 | 300 | 0.63 (0.45, 0.88) | 0.13 |
| Madagascar | MAHAY (43) | 600 | 588 | 0.81 (0.64, 1.03) | 0.16 |
| Mali | PROMIS CS (46) | 953 | 970 | 0.63 (0.54, 0.74) | 0.18 |
| Zimbabwe | SHINE (HIV−) (47) | 1682 | 1594 | 0.67 (0.53, 0.84) | 0.16 |
| Zimbabwe | SHINE (HIV+) (48) | 306 | 285 | 0.51 (0.30, 0.89) | 0.08 |
| **I² = 0.52, Tau² = 0.02** | | **5014** | **4590** | **0.69 (0.60, 0.79)** | |
| **Frequency of contact − Weekly** | | | | | |
| Bangladesh | JiVitA−4 (35) | | | | |
| Bangladesh | WASH−B (37) | | | | |
| Burkina Faso | iLiNS−Zinc (38) | 1957 | 664 | 0.72 (0.65, 0.78) | 0.32 |
| Ghana | GHANA (40) | 98 | 96 | 0.32 (0.16, 0.61) | 0.10 |
| Ghana | iLiNS−DYADG (41) | 328 | 661 | 0.84 (0.47, 1.51) | 0.12 |
| Malawi | iLiNS−DYADM (44) | 210 | 432 | 1.15 (0.87, 1.52) | 0.23 |
| Malawi | iLiNS−DOSE (45) | 243 | 82 | 0.72 (0.54, 0.96) | 0.23 |
| **I² = 0.76, Tau² = 0.15** | | **2836** | **1935** | **0.73 (0.50, 1.06)** | |

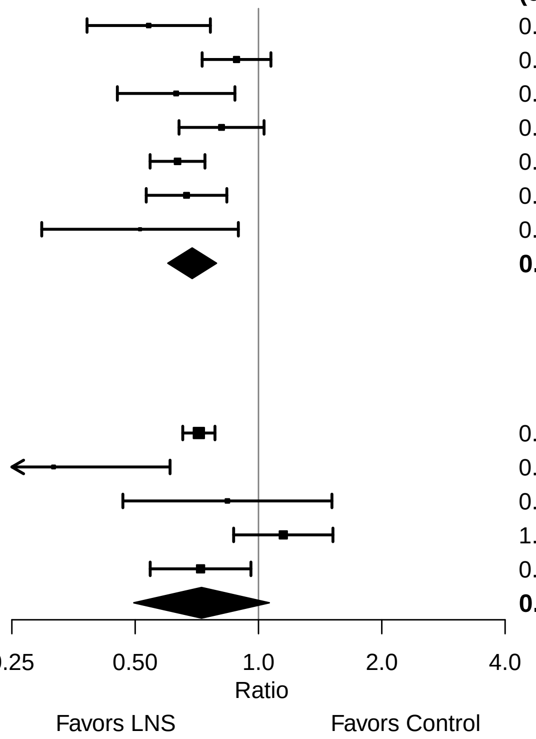

#### Supplemental figure 4D: Moderate-to-severe anemia prevalence ratio

### 4D10: Stratified by Average SQ-LNS compliance

**Average SQ−LNS compliance**
**(p−diff = 0.115)**

| Country | Trial | N | N | PR (95% CI) | W |
|---|---|---|---|---|---|
| **Average SQ−LNS compliance − Low** | | | | | |
| Burkina Faso | PROMIS CS (39) | 574 | 581 | 0.88 (0.73, 1.07) | 0.18 |
| Ghana | iLiNS−DYADG (41) | 328 | 661 | 0.84 (0.47, 1.51) | 0.08 |
| Malawi | iLiNS−DYADM (44) | 210 | 432 | 1.15 (0.87, 1.52) | 0.15 |
| Malawi | iLiNS−DOSE (45) | 243 | 82 | 0.72 (0.54, 0.96) | 0.15 |
| Mali | PROMIS CS (46) | 953 | 970 | 0.63 (0.54, 0.74) | 0.19 |
| Zimbabwe | SHINE (HIV−) (47) | 1682 | 1594 | 0.67 (0.53, 0.84) | 0.17 |
| Zimbabwe | SHINE (HIV+) (48) | 306 | 285 | 0.51 (0.30, 0.89) | 0.09 |
| **I² = 0.69, Tau² = 0.04** | | **4296** | **4605** | **0.76 (0.63, 0.91)** | |
| **Average SQ−LNS compliance − High** | | | | | |
| Bangladesh | JiVitA−4 (35) | | | | |
| Bangladesh | RDNS (36) | 549 | 272 | 0.54 (0.38, 0.76) | 0.25 |
| Bangladesh | WASH−B (37) | | | | |
| Burkina Faso | iLiNS−Zinc (38) | 1957 | 664 | 0.72 (0.65, 0.78) | 0.37 |
| Ghana | GHANA (40) | 98 | 96 | 0.32 (0.16, 0.61) | 0.13 |
| Kenya | WASH−B (42) | 350 | 300 | 0.63 (0.45, 0.88) | 0.25 |
| **I² = 0.64, Tau² = 0.06** | | **2954** | **1332** | **0.58 (0.43, 0.78)** | |

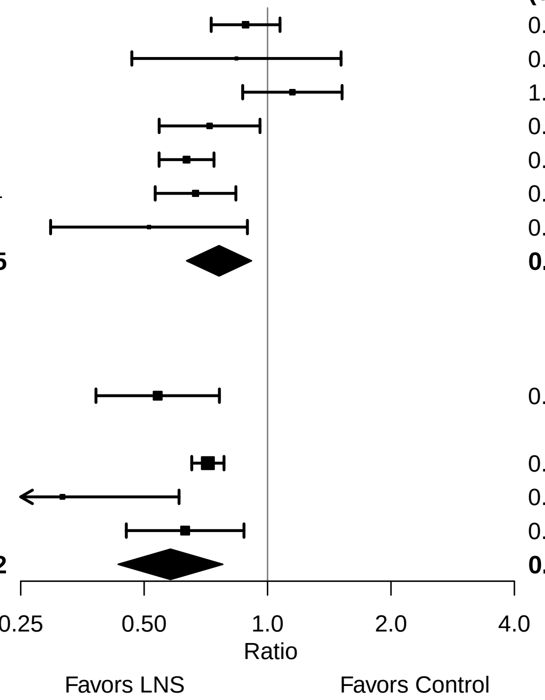

### Supplemental figure 4E: Moderate-to-severe anemia prevalence difference

#### 4E1: Stratified by Geographic region (insufficient comparisons)

#### Supplemental figure 4E: Moderate-to-severe anemia prevalence difference

#### 4E2: Stratified by Anemia burden

**Anemia burden**
**(p−diff = 0.065)**

| Country | Trial | N | N | PD (95% CI) | W |
|---|---|---|---|---|---|
| **Anemia burden − Moderate** | | | | | |
| Bangladesh | JiVitA−4 (35) | | | | |
| Bangladesh | RDNS (36) | 549 | 272 | −0.08 (−0.13, −0.03) | 0.16 |
| Bangladesh | WASH−B (37) | | | | |
| Ghana | iLiNS−DYADG (41) | 328 | 661 | −0.01 (−0.04, 0.02) | 0.17 |
| Kenya | WASH−B (42) | 350 | 300 | −0.08 (−0.14, −0.02) | 0.16 |
| Madagascar | MAHAY (43) | 600 | 588 | −0.06 (−0.11, −0.01) | 0.16 |
| Zimbabwe | SHINE (HIV−) (47) | 1682 | 1594 | −0.04 (−0.05, −0.02) | 0.18 |
| Zimbabwe | SHINE (HIV+) (48) | 306 | 285 | −0.05 (−0.09, −0.01) | 0.17 |
| **I² = 0.46, Tau² = 0.00** | | **3815** | **3700** | **−0.04 (−0.07, −0.02)** | |
| **Anemia burden − High** | | | | | |
| Burkina Faso | iLiNS−Zinc (38) | 1957 | 664 | −0.21 (−0.25, −0.17) | 0.19 |
| Burkina Faso | PROMIS CS (39) | 574 | 581 | −0.04 (−0.10, 0.01) | 0.18 |
| Ghana | GHANA (40) | 98 | 96 | −0.22 (−0.33, −0.11) | 0.14 |
| Malawi | iLiNS−DYADM (44) | 210 | 432 | 0.04 (−0.04, 0.11) | 0.17 |
| Malawi | iLiNS−DOSE (45) | 243 | 82 | −0.13 (−0.25, −0.01) | 0.13 |
| Mali | PROMIS CS (46) | 953 | 970 | −0.22 (−0.26, −0.18) | 0.19 |
| **I² = 0.92, Tau² = 0.01** | | **4035** | **2825** | **−0.13 (−0.22, −0.04)** | |

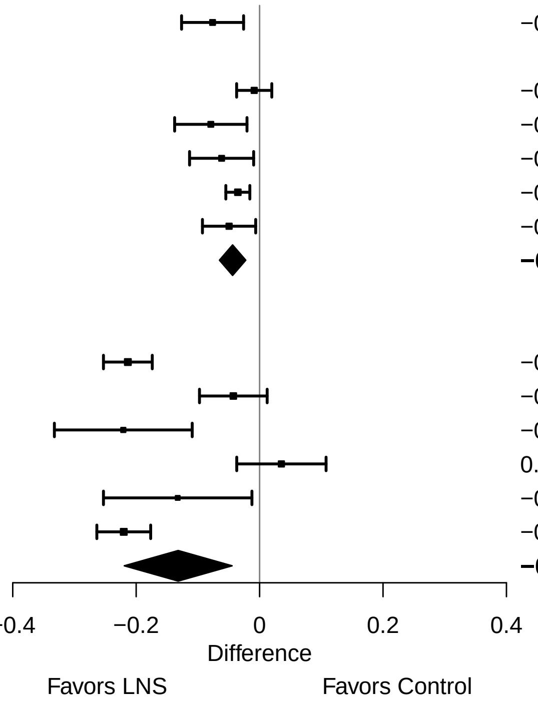

#### Supplemental figure 4E: Moderate-to-severe anemia prevalence difference

### 4E3: Stratified by Malaria prevalence

**Malaria prevalence**
**(p−diff = 0.293)**

| Country | Trial | N | N | PD (95% CI) | W |
|---|---|---|---|---|---|
| **Malaria prevalence − Less than 10%** | | | | | |
| Bangladesh | JiVitA−4 (35) | | | | |
| Bangladesh | RDNS (36) | 549 | 272 | −0.08 (−0.13, −0.03) | 0.20 |
| Bangladesh | WASH−B (37) | | | | |
| Kenya | WASH−B (42) | 350 | 300 | −0.08 (−0.14, −0.02) | 0.19 |
| Madagascar | MAHAY (43) | 600 | 588 | −0.06 (−0.11, −0.01) | 0.20 |
| Zimbabwe | SHINE (HIV−) (47) | 1682 | 1594 | −0.04 (−0.05, −0.02) | 0.21 |
| Zimbabwe | SHINE (HIV+) (48) | 306 | 285 | −0.05 (−0.09, −0.01) | 0.20 |
| **I² = 0.03, Tau² = 0.00** | | **3487** | **3039** | **−0.05 (−0.06, −0.03)** | |
| **Malaria prevalence − At least 10%** | | | | | |
| Burkina Faso | iLiNS−Zinc (38) | 1957 | 664 | −0.21 (−0.25, −0.17) | 0.16 |
| Burkina Faso | PROMIS CS (39) | 574 | 581 | −0.04 (−0.10, 0.01) | 0.15 |
| Ghana | GHANA (40) | 98 | 96 | −0.22 (−0.33, −0.11) | 0.11 |
| Ghana | iLiNS−DYADG (41) | 328 | 661 | −0.01 (−0.04, 0.02) | 0.17 |
| Malawi | iLiNS−DYADM (44) | 210 | 432 | 0.04 (−0.04, 0.11) | 0.14 |
| Malawi | iLiNS−DOSE (45) | 243 | 82 | −0.13 (−0.25, −0.01) | 0.11 |
| Mali | PROMIS CS (46) | 953 | 970 | −0.22 (−0.26, −0.18) | 0.16 |
| **I² = 0.95, Tau² = 0.01** | | **4363** | **3486** | **−0.11 (−0.19, −0.03)** | |

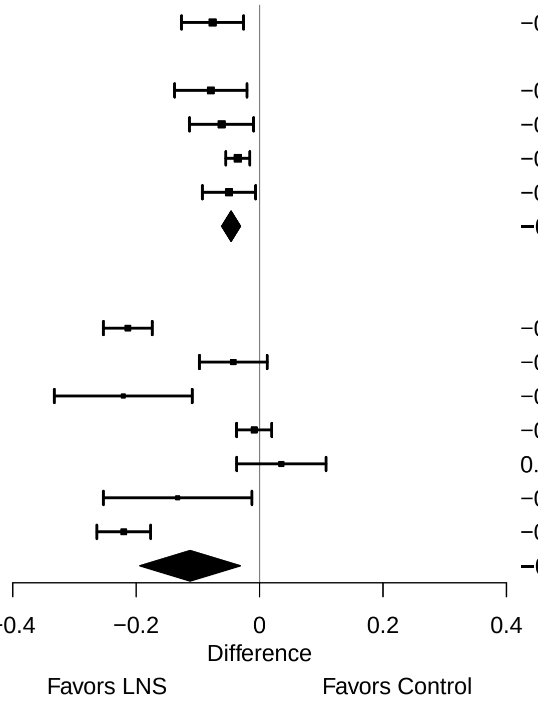

#### Supplemental figure 4E: Moderate-to-severe anemia prevalence difference

### 4E4: Stratified by Inflammation burden

**Inflammation burden**
**(p−diff = 0.312)**

| Country | Trial | N | N | PD (95% CI) | W |
|---|---|---|---|---|---|
| **Inflammation burden − Low** | | | | | |
| Bangladesh | RDNS (36) | 549 | 272 | −0.08 (−0.13, −0.03) | 0.15 |
| Bangladesh | WASH−B (37) | | | | |
| Ghana | GHANA (40) | 98 | 96 | −0.22 (−0.33, −0.11) | 0.11 |
| Ghana | iLiNS−DYADG (41) | 328 | 661 | −0.01 (−0.04, 0.02) | 0.16 |
| Kenya | WASH−B (42) | 350 | 300 | −0.08 (−0.14, −0.02) | 0.14 |
| Madagascar | MAHAY (43) | 600 | 588 | −0.06 (−0.11, −0.01) | 0.14 |
| Zimbabwe | SHINE (HIV−) (47) | 1682 | 1594 | −0.04 (−0.05, −0.02) | 0.16 |
| Zimbabwe | SHINE (HIV+) (48) | 306 | 285 | −0.05 (−0.09, −0.01) | 0.15 |
| **$I^2$ = 0.69, $Tau^2$ = 0.00** | | **3913** | **3796** | **−0.06 (−0.10, −0.02)** | |
| **Inflammation burden − High** | | | | | |
| Bangladesh | JiVitA−4 (35) | | | | |
| Burkina Faso | iLiNS−Zinc (38) | 1957 | 664 | −0.21 (−0.25, −0.17) | 0.22 |
| Burkina Faso | PROMIS CS (39) | 574 | 581 | −0.04 (−0.10, 0.01) | 0.21 |
| Malawi | iLiNS−DYADM (44) | 210 | 432 | 0.04 (−0.04, 0.11) | 0.20 |
| Malawi | iLiNS−DOSE (45) | 243 | 82 | −0.13 (−0.25, −0.01) | 0.15 |
| Mali | PROMIS CS (46) | 953 | 970 | −0.22 (−0.26, −0.18) | 0.22 |
| **$I^2$ = 0.93, $Tau^2$ = 0.01** | | **3937** | **2729** | **−0.12 (−0.21, −0.02)** | |

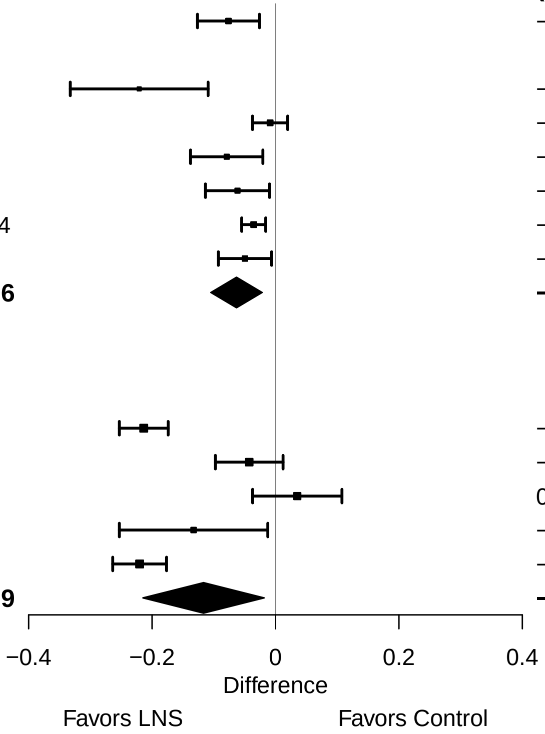

#### Supplemental figure 4E: Moderate-to-severe anemia prevalence difference

#### 4E5: Stratified by Source water quality

**Source water quality**
**(p−diff = 0.626)**

| Country | Trial | N | N | PD (95% CI) | W |
|---|---|---|---|---|---|
| **Source water quality − Improved** | | | | | |
| Bangladesh | JiVitA−4 (35) | | | | |
| Bangladesh | RDNS (36) | 549 | 272 | −0.08 (−0.13, −0.03) | 0.23 |
| Bangladesh | WASH−B (37) | | | | |
| Ghana | GHANA (40) | 98 | 96 | −0.22 (−0.33, −0.11) | 0.17 |
| Ghana | iLiNS−DYADG (41) | 328 | 661 | −0.01 (−0.04, 0.02) | 0.24 |
| Malawi | iLiNS−DYADM (44) | 210 | 432 | 0.04 (−0.04, 0.11) | 0.21 |
| Malawi | iLiNS−DOSE (45) | 243 | 82 | −0.13 (−0.25, −0.01) | 0.16 |
| **$I^2$ = 0.82, $Tau^2$ = 0.01** | | **1428** | **1543** | **−0.07 (−0.16, 0.01)** | |
| **Source water quality − Unimproved** | | | | | |
| Burkina Faso | iLiNS−Zinc (38) | 1957 | 664 | −0.21 (−0.25, −0.17) | 0.15 |
| Burkina Faso | PROMIS CS (39) | 574 | 581 | −0.04 (−0.10, 0.01) | 0.14 |
| Kenya | WASH−B (42) | 196 | 162 | −0.04 (−0.12, 0.04) | 0.13 |
| Madagascar | MAHAY (43) | 600 | 588 | −0.06 (−0.11, −0.01) | 0.14 |
| Mali | PROMIS CS (46) | 953 | 970 | −0.22 (−0.26, −0.18) | 0.15 |
| Zimbabwe | SHINE (HIV−) (47) | 819 | 795 | −0.04 (−0.07, −0.01) | 0.15 |
| Zimbabwe | SHINE (HIV+) (48) | 133 | 126 | −0.04 (−0.10, 0.02) | 0.14 |
| **$I^2$ = 0.93, $Tau^2$ = 0.01** | | **5232** | **3886** | **−0.10 (−0.16, −0.03)** | |

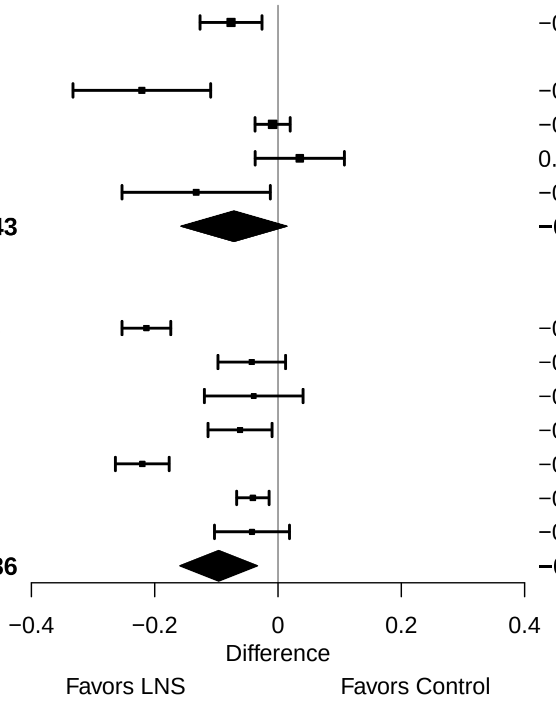

#### Supplemental figure 4E: Moderate-to-severe anemia prevalence difference

### 4E6: Stratified by Sanitation

**Sanitation**
**(p−diff = 0.481)**

| Country | Trial | N | N | PD (95% CI) | W |
|---|---|---|---|---|---|
| **Sanitation − Improved** | | | | | |
| Bangladesh | JiVitA−4 (35) | | | | |
| Bangladesh | RDNS (36) | 549 | 272 | −0.08 (−0.13, −0.03) | 0.21 |
| Bangladesh | WASH−B (37) | | | | |
| Burkina Faso | PROMIS CS (39) | 574 | 581 | −0.04 (−0.10, 0.01) | 0.20 |
| Ghana | GHANA (40) | 98 | 96 | −0.22 (−0.33, −0.11) | 0.15 |
| Ghana | iLiNS−DYADG (41) | 328 | 661 | −0.01 (−0.04, 0.02) | 0.22 |
| Mali | PROMIS CS (46) | 953 | 970 | −0.22 (−0.26, −0.18) | 0.21 |
| **I² = 0.94, Tau² = 0.01** | | **2502** | **2580** | **−0.11 (−0.20, −0.02)** | |
| **Sanitation − Unimproved** | | | | | |
| Burkina Faso | iLiNS−Zinc (38) | 1957 | 664 | −0.21 (−0.25, −0.17) | 0.16 |
| Kenya | WASH−B (42) | 196 | 162 | −0.04 (−0.12, 0.04) | 0.13 |
| Madagascar | MAHAY (43) | 600 | 588 | −0.06 (−0.11, −0.01) | 0.15 |
| Malawi | iLiNS−DYADM (44) | 210 | 432 | 0.04 (−0.04, 0.11) | 0.14 |
| Malawi | iLiNS−DOSE (45) | 243 | 82 | −0.13 (−0.25, −0.01) | 0.11 |
| Zimbabwe | SHINE (HIV−) (47) | 819 | 795 | −0.04 (−0.07, −0.01) | 0.16 |
| Zimbabwe | SHINE (HIV+) (48) | 133 | 126 | −0.04 (−0.10, 0.02) | 0.15 |
| **I² = 0.91, Tau² = 0.01** | | **4158** | **2849** | **−0.07 (−0.13, −0.01)** | |

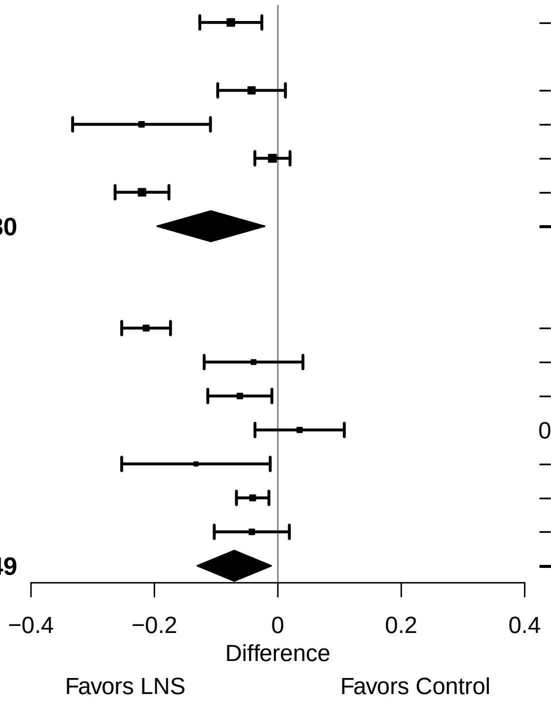

#### Supplemental figure 4E: Moderate-to-severe anemia prevalence difference

### 4E7: Stratified by Supplement duration

**Supplement duration**
**(p−diff = 0.363)**

**Supplement duration − 12m or less**

| Country | Trial | N | N | PD (95% CI) | W |
|---|---|---|---|---|---|
| Bangladesh | JiVitA−4 (35) | | | | |
| Burkina Faso | iLiNS−Zinc (38) | 1957 | 664 | −0.21 (−0.25, −0.17) | 0.12 |
| Burkina Faso | PROMIS CS (39) | 574 | 581 | −0.04 (−0.10, 0.01) | 0.12 |
| Ghana | GHANA (40) | 98 | 96 | −0.22 (−0.33, −0.11) | 0.09 |
| Ghana | iLiNS−DYADG (41) | 328 | 661 | −0.01 (−0.04, 0.02) | 0.13 |
| Madagascar | MAHAY (43) | 600 | 588 | −0.06 (−0.11, −0.01) | 0.12 |
| Malawi | iLiNS−DYADM (44) | 210 | 432 | 0.04 (−0.04, 0.11) | 0.11 |
| Malawi | iLiNS−DOSE (45) | 243 | 82 | −0.13 (−0.25, −0.01) | 0.08 |
| Zimbabwe | SHINE (HIV−) (47) | 1682 | 1594 | −0.04 (−0.05, −0.02) | 0.13 |
| Zimbabwe | SHINE (HIV+) (48) | 306 | 285 | −0.05 (−0.09, −0.01) | 0.12 |
| **I² = 0.91, Tau² = 0.01** | | **5998** | **4983** | **−0.08 (−0.13, −0.02)** | |

**Supplement duration − > 12m**

| Country | Trial | N | N | PD (95% CI) | W |
|---|---|---|---|---|---|
| Bangladesh | RDNS (36) | 549 | 272 | −0.08 (−0.13, −0.03) | 0.33 |
| Bangladesh | WASH−B (37) | | | | |
| Kenya | WASH−B (42) | 350 | 300 | −0.08 (−0.14, −0.02) | 0.32 |
| Mali | PROMIS CS (46) | 953 | 970 | −0.22 (−0.26, −0.18) | 0.34 |
| **I² = 0.91, Tau² = 0.01** | | **1852** | **1542** | **−0.13 (−0.22, −0.03)** | |

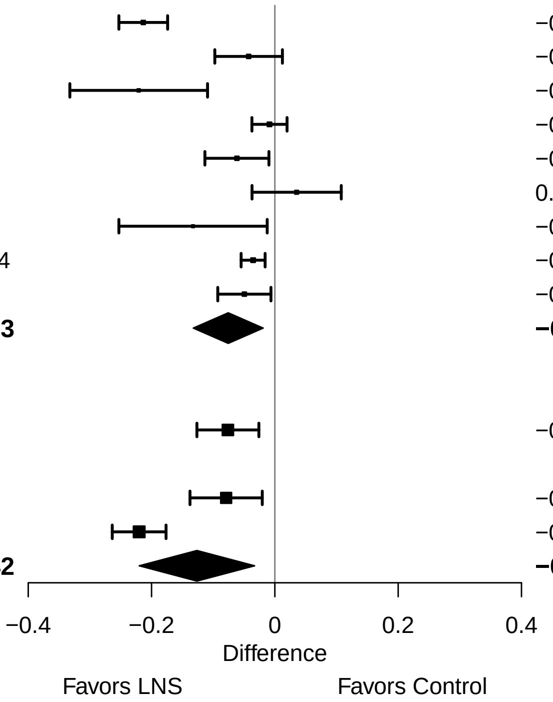

#### Supplemental figure 4E: Moderate-to-severe anemia prevalence difference

### 4E8: Stratified by Iron dose

**Iron dose**
**(p−diff = 0.542)**

| Country | Trial | N | N | PD (95% CI) | W |
|---|---|---|---|---|---|
| **Iron dose − Less than 9 mg** | | | | | |
| Bangladesh | JiVitA−4 (35) | | | | |
| Burkina Faso | iLiNS−Zinc (38) | 1957 | 664 | −0.21 (−0.25, −0.17) | 0.12 |
| Burkina Faso | PROMIS CS (39) | 574 | 581 | −0.04 (−0.10, 0.01) | 0.11 |
| Ghana | iLiNS−DYADG (41) | 328 | 661 | −0.01 (−0.04, 0.02) | 0.12 |
| Madagascar | MAHAY (43) | 600 | 588 | −0.06 (−0.11, −0.01) | 0.11 |
| Malawi | iLiNS−DYADM (44) | 210 | 432 | 0.04 (−0.04, 0.11) | 0.10 |
| Malawi | iLiNS−DOSE (45) | 243 | 82 | −0.13 (−0.25, −0.01) | 0.08 |
| Mali | PROMIS CS (46) | 953 | 970 | −0.22 (−0.26, −0.18) | 0.12 |
| Zimbabwe | SHINE (HIV−) (47) | 1682 | 1594 | −0.04 (−0.05, −0.02) | 0.12 |
| Zimbabwe | SHINE (HIV+) (48) | 306 | 285 | −0.05 (−0.09, −0.01) | 0.12 |
| **I² = 0.94, Tau² = 0.01** | | **6853** | **5857** | **−0.08 (−0.14, −0.02)** | |
| **Iron dose − 9 mg** | | | | | |
| Bangladesh | RDNS (36) | 549 | 272 | −0.08 (−0.13, −0.03) | 0.37 |
| Bangladesh | WASH−B (37) | | | | |
| Ghana | GHANA (40) | 98 | 96 | −0.22 (−0.33, −0.11) | 0.27 |
| Kenya | WASH−B (42) | 350 | 300 | −0.08 (−0.14, −0.02) | 0.36 |
| **I² = 0.65, Tau² = 0.00** | | **997** | **668** | **−0.11 (−0.20, −0.03)** | |

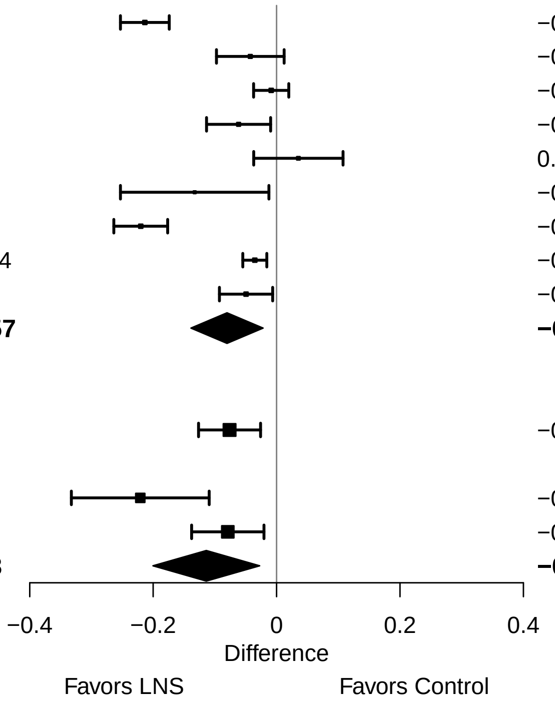

#### Supplemental figure 4E: Moderate-to-severe anemia prevalence difference

### 4E9: Stratified by Frequency of contact

**Frequency of contact**
**(p−diff = 0.680)**

| Country | Trial | N | N | PD (95% CI) | W |
|---|---|---|---|---|---|
| **Frequency of contact − Monthly** | | | | | |
| Bangladesh | RDNS (36) | 549 | 272 | −0.08 (−0.13, −0.03) | 0.14 |
| Burkina Faso | PROMIS CS (39) | 574 | 581 | −0.04 (−0.10, 0.01) | 0.14 |
| Kenya | WASH−B (42) | 350 | 300 | −0.08 (−0.14, −0.02) | 0.14 |
| Madagascar | MAHAY (43) | 600 | 588 | −0.06 (−0.11, −0.01) | 0.14 |
| Mali | PROMIS CS (46) | 953 | 970 | −0.22 (−0.26, −0.18) | 0.14 |
| Zimbabwe | SHINE (HIV−) (47) | 1682 | 1594 | −0.04 (−0.05, −0.02) | 0.15 |
| Zimbabwe | SHINE (HIV+) (48) | 306 | 285 | −0.05 (−0.09, −0.01) | 0.14 |
| **I² = 0.90, Tau² = 0.00** | | **5014** | **4590** | **−0.08 (−0.13, −0.03)** | |
| **Frequency of contact − Weekly** | | | | | |
| Bangladesh | JiVitA−4 (35) | | | | |
| Bangladesh | WASH−B (37) | | | | |
| Burkina Faso | iLiNS−Zinc (38) | 1957 | 664 | −0.21 (−0.25, −0.17) | 0.23 |
| Ghana | GHANA (40) | 98 | 96 | −0.22 (−0.33, −0.11) | 0.16 |
| Ghana | iLiNS−DYADG (41) | 328 | 661 | −0.01 (−0.04, 0.02) | 0.24 |
| Malawi | iLiNS−DYADM (44) | 210 | 432 | 0.04 (−0.04, 0.11) | 0.20 |
| Malawi | iLiNS−DOSE (45) | 243 | 82 | −0.13 (−0.25, −0.01) | 0.16 |
| **I² = 0.95, Tau² = 0.01** | | **2836** | **1935** | **−0.10 (−0.21, 0.00)** | |

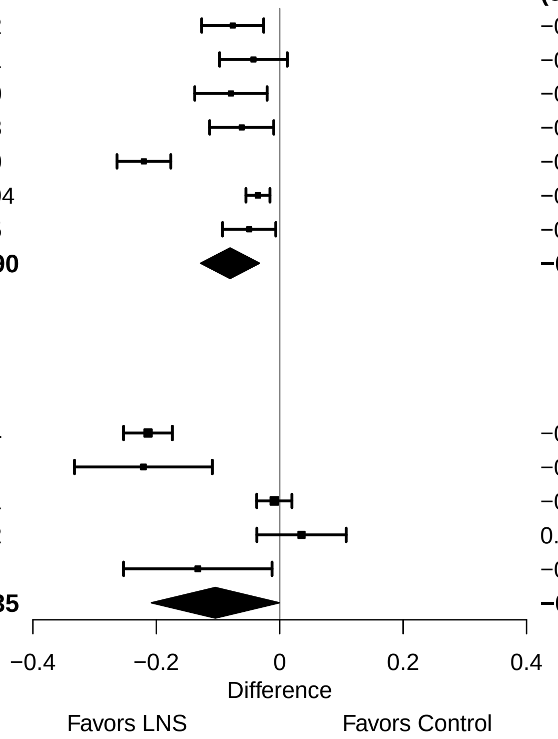

#### Supplemental figure 4E: Moderate-to-severe anemia prevalence difference

### 4E10: Stratified by Average SQ-LNS compliance

**Average SQ−LNS compliance**
**(p−diff = 0.128)**

| **Average SQ−LNS compliance − Low** | | | | **PD** | |
|---|---|---|---|---|---|
| **Country** | **Trial** | **N** | **N** | **(95% CI)** | **W** |
| Burkina Faso | PROMIS CS (39) | 574 | 581 | −0.04 (−0.10, 0.01) | 0.14 |
| Ghana | iLiNS−DYADG (41) | 328 | 661 | −0.01 (−0.04, 0.02) | 0.16 |
| Malawi | iLiNS−DYADM (44) | 210 | 432 | 0.04 (−0.04, 0.11) | 0.13 |
| Malawi | iLiNS−DOSE (45) | 243 | 82 | −0.13 (−0.25, −0.01) | 0.10 |
| Mali | PROMIS CS (46) | 953 | 970 | −0.22 (−0.26, −0.18) | 0.15 |
| Zimbabwe | SHINE (HIV−) (47) | 1682 | 1594 | −0.04 (−0.05, −0.02) | 0.16 |
| Zimbabwe | SHINE (HIV+) (48) | 306 | 285 | −0.05 (−0.09, −0.01) | 0.15 |
| **I² = 0.92, Tau² = 0.01** | | **4296** | **4605** | **−0.06 (−0.13, 0.00)** | |
| **Average SQ−LNS compliance − High** | | | | | |
| Bangladesh | JiVitA−4 (35) | | | | |
| Bangladesh | RDNS (36) | 549 | 272 | −0.08 (−0.13, −0.03) | 0.27 |
| Bangladesh | WASH−B (37) | | | | |
| Burkina Faso | iLiNS−Zinc (38) | 1957 | 664 | −0.21 (−0.25, −0.17) | 0.28 |
| Ghana | GHANA (40) | 98 | 96 | −0.22 (−0.33, −0.11) | 0.20 |
| Kenya | WASH−B (42) | 350 | 300 | −0.08 (−0.14, −0.02) | 0.26 |
| **I² = 0.88, Tau² = 0.01** | | **2954** | **1332** | **−0.14 (−0.22, −0.06)** | |

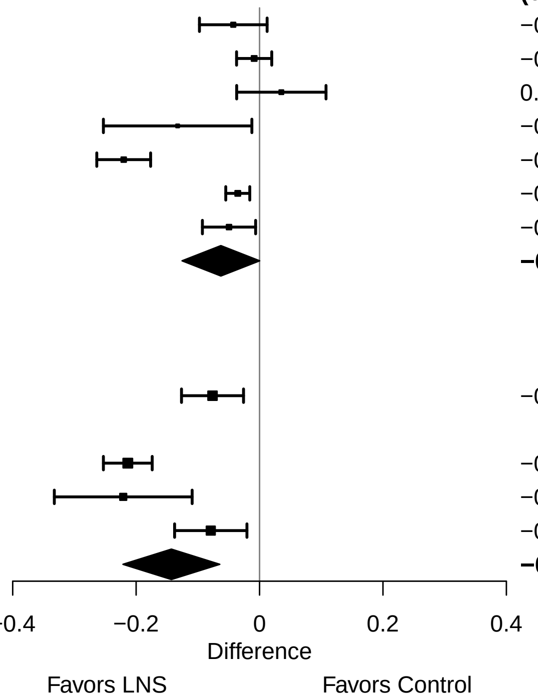

### Supplemental figure 4F: Geometric mean ratio of ferritin concentration

#### 4F1: Stratified by Geographic region

**Geographic region**
**(p−diff = 0.800)**

| Country | Trial | N | N | GMR (95% CI) | W |
|---|---|---|---|---|---|
| **Geographic region − SEAR** | | | | | |
| Bangladesh | JiVitA−4 (35) | 455 | 144 | 1.39 (1.17, 1.64) | 0.29 |
| Bangladesh | RDNS (36) | 550 | 272 | 1.52 (1.40, 1.64) | 0.40 |
| Bangladesh | WASH−B (37) | 212 | 178 | 1.74 (1.50, 2.01) | 0.31 |
| **I² = 0.53, Tau² = 0.01** | | **1217** | **594** | **1.54 (1.37, 1.74)** | |
| **Geographic region − AFR** | | | | | |
| Burkina Faso | iLiNS−Zinc (38) | 315 | 96 | 1.55 (1.31, 1.84) | 0.30 |
| Burkina Faso | PROMIS CS (39) | | | | |
| Ghana | GHANA (40) | 83 | 82 | 2.52 (1.49, 4.25) | 0.07 |
| Ghana | iLiNS−DYADG (41) | | | | |
| Kenya | WASH−B (42) | 298 | 259 | 1.69 (1.50, 1.91) | 0.37 |
| Madagascar | MAHAY (43) | 83 | 51 | 1.29 (1.05, 1.59) | 0.26 |
| Malawi | iLiNS−DYADM (44) | | | | |
| Malawi | iLiNS−DOSE (45) | | | | |
| Mali | PROMIS CS (46) | | | | |
| Zimbabwe | SHINE (HIV−) (47) | | | | |
| Zimbabwe | SHINE (HIV+) (48) | | | | |
| **I² = 0.63, Tau² = 0.04** | | **779** | **488** | **1.61 (1.29, 2.00)** | |

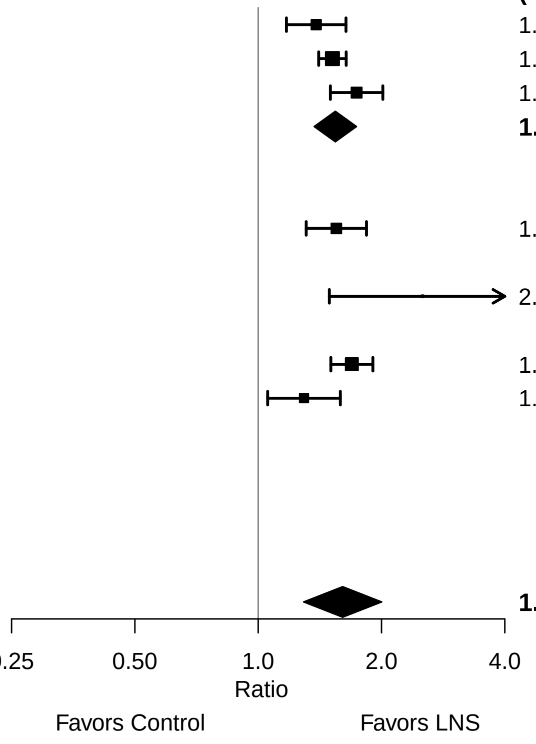

**Supplemental figure 4F: Geometric mean ratio of ferritin concentration**

#### 4F2: Stratified by Anemia burden (insufficient comparisons)

**Supplemental figure 4F: Geometric mean ratio of ferritin concentration**

#### 4F3: Stratified by Malaria prevalence (insufficient comparisons)

**Supplemental figure 4F: Geometric mean ratio of ferritin concentration**

#### 4F4: Stratified by Inflammation burden (insufficient comparisons)

#### Supplemental figure 4F: Geometric mean ratio of ferritin concentration

### 4F5: Stratified by Source water quality

**Source water quality**
**(p−diff = 0.862)**

| Country | Trial | N | N | GMR (95% CI) | W |
|---|---|---|---|---|---|
| **Source water quality − Improved** | | | | | |
| Bangladesh | JiVitA−4 (35) | 455 | 144 | 1.39 (1.17, 1.64) | 0.30 |
| Bangladesh | RDNS (36) | 550 | 272 | 1.52 (1.40, 1.64) | 0.45 |
| Bangladesh | WASH−B (37) | 94 | 58 | 1.75 (1.33, 2.31) | 0.18 |
| Ghana | GHANA (40) | 83 | 82 | 2.52 (1.49, 4.25) | 0.07 |
| Ghana | iLiNS−DYADG (41) | | | | |
| Malawi | iLiNS−DYADM (44) | | | | |
| Malawi | iLiNS−DOSE (45) | | | | |
| **I² = 0.48, Tau² = 0.03** | | **1182** | **556** | **1.61 (1.32, 1.96)** | |
| **Source water quality − Unimproved** | | | | | |
| Burkina Faso | iLiNS−Zinc (38) | 315 | 96 | 1.55 (1.31, 1.84) | 0.34 |
| Burkina Faso | PROMIS CS (39) | | | | |
| Kenya | WASH−B (42) | 166 | 144 | 1.67 (1.44, 1.94) | 0.38 |
| Madagascar | MAHAY (43) | 83 | 51 | 1.29 (1.05, 1.59) | 0.28 |
| Mali | PROMIS CS (46) | | | | |
| Zimbabwe | SHINE (HIV−) (47) | | | | |
| Zimbabwe | SHINE (HIV+) (48) | | | | |
| **I² = 0.49, Tau² = 0.01** | | **564** | **291** | **1.52 (1.31, 1.75)** | |

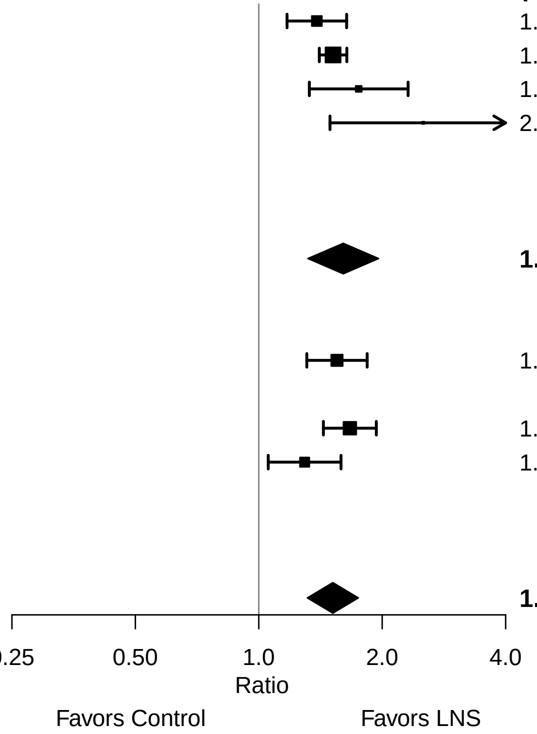

#### Supplemental figure 4F: Geometric mean ratio of ferritin concentration

### 4F6: Stratified by Sanitation

**Sanitation**
**(p−diff = 0.862)**

| Country | Trial | N | N | GMR (95% CI) | W |
|---|---|---|---|---|---|
| **Sanitation − Improved** | | | | | |
| Bangladesh | JiVitA−4 (35) | 455 | 144 | 1.39 (1.17, 1.64) | 0.30 |
| Bangladesh | RDNS (36) | 550 | 272 | 1.52 (1.40, 1.64) | 0.45 |
| Bangladesh | WASH−B (37) | 94 | 58 | 1.75 (1.33, 2.31) | 0.18 |
| Burkina Faso | PROMIS CS (39) | | | | |
| Ghana | GHANA (40) | 83 | 82 | 2.52 (1.49, 4.25) | 0.07 |
| Ghana | iLiNS−DYADG (41) | | | | |
| Mali | PROMIS CS (46) | | | | |
| **I² = 0.48, Tau² = 0.03** | | **1182** | **556** | **1.61 (1.32, 1.96)** | |
| **Sanitation − Unimproved** | | | | | |
| Burkina Faso | iLiNS−Zinc (38) | 315 | 96 | 1.55 (1.31, 1.84) | 0.34 |
| Kenya | WASH−B (42) | 166 | 144 | 1.67 (1.44, 1.94) | 0.38 |
| Madagascar | MAHAY (43) | 83 | 51 | 1.29 (1.05, 1.59) | 0.28 |
| Malawi | iLiNS−DYADM (44) | | | | |
| Malawi | iLiNS−DOSE (45) | | | | |
| Zimbabwe | SHINE (HIV−) (47) | | | | |
| Zimbabwe | SHINE (HIV+) (48) | | | | |
| **I² = 0.49, Tau² = 0.01** | | **564** | **291** | **1.52 (1.31, 1.75)** | |

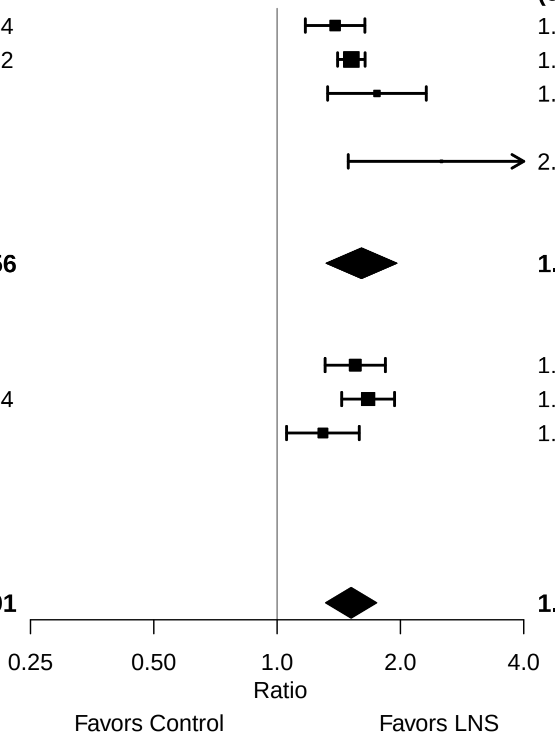

#### Supplemental figure 4F: Geometric mean ratio of ferritin concentration

### 4F7: Stratified by Supplement duration

**Supplement duration**
**(p−diff = 0.153)**

| Country | Trial | N | N | GMR (95% CI) | W |
|---|---|---|---|---|---|
| **Supplement duration − 12m or less** | | | | | |
| Bangladesh | JiVitA−4 (35) | 455 | 144 | 1.39 (1.17, 1.64) | 0.32 |
| Burkina Faso | iLiNS−Zinc (38) | 315 | 96 | 1.55 (1.31, 1.84) | 0.32 |
| Burkina Faso | PROMIS CS (39) | | | | |
| Ghana | GHANA (40) | 83 | 82 | 2.52 (1.49, 4.25) | 0.08 |
| Ghana | iLiNS−DYADG (41) | | | | |
| Madagascar | MAHAY (43) | 83 | 51 | 1.29 (1.05, 1.59) | 0.28 |
| Malawi | iLiNS−DYADM (44) | | | | |
| Malawi | iLiNS−DOSE (45) | | | | |
| Zimbabwe | SHINE (HIV−) (47) | | | | |
| Zimbabwe | SHINE (HIV+) (48) | | | | |
| **I² = 0.53, Tau² = 0.04** | | **936** | **373** | **1.52 (1.20, 1.91)** | |
| **Supplement duration − > 12m** | | | | | |
| Bangladesh | RDNS (36) | 550 | 272 | 1.52 (1.40, 1.64) | 0.38 |
| Bangladesh | WASH−B (37) | 212 | 178 | 1.74 (1.50, 2.01) | 0.29 |
| Kenya | WASH−B (42) | 298 | 259 | 1.69 (1.50, 1.91) | 0.33 |
| Mali | PROMIS CS (46) | | | | |
| **I² = 0.48, Tau² = 0.00** | | **1060** | **709** | **1.62 (1.49, 1.76)** | |

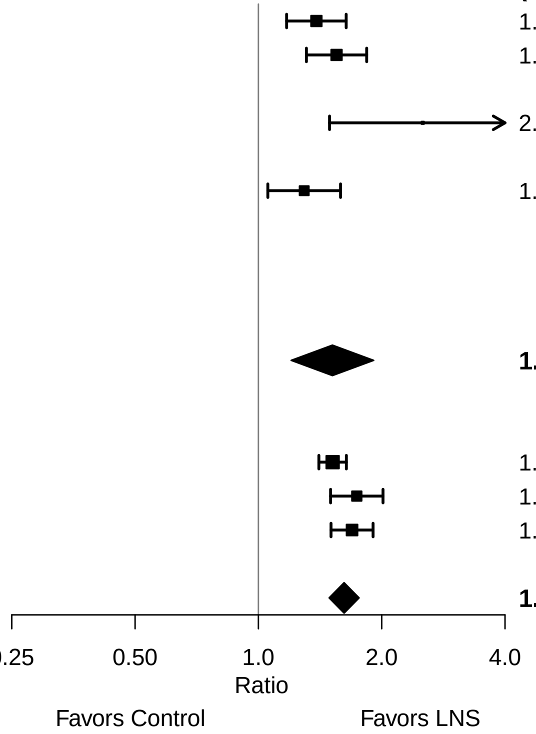

**Supplemental figure 4F: Geometric mean ratio of ferritin concentration**

#### 4F8: Stratified by Iron dose

**Iron dose**
**(p−diff = 0.054)**

| Country | Trial | N | N | GMR (95% CI) | W |
|---|---|---|---|---|---|
| **Iron dose − Less than 9 mg** | | | | | |
| Bangladesh | JiVitA−4 (35) | 455 | 144 | 1.39 (1.17, 1.64) | 0.35 |
| Burkina Faso | iLiNS−Zinc (38) | 315 | 96 | 1.55 (1.31, 1.84) | 0.35 |
| Burkina Faso | PROMIS CS (39) | | | | |
| Ghana | iLiNS−DYADG (41) | | | | |
| Madagascar | MAHAY (43) | 83 | 51 | 1.29 (1.05, 1.59) | 0.30 |
| Malawi | iLiNS−DYADM (44) | | | | |
| Malawi | iLiNS−DOSE (45) | | | | |
| Mali | PROMIS CS (46) | | | | |
| Zimbabwe | SHINE (HIV−) (47) | | | | |
| Zimbabwe | SHINE (HIV+) (48) | | | | |
| **I² = 0.00, Tau² = 0.00** | | **853** | **291** | **1.42 (1.28, 1.57)** | |
| **Iron dose − 9 mg** | | | | | |
| Bangladesh | RDNS (36) | 550 | 272 | 1.52 (1.40, 1.64) | 0.35 |
| Bangladesh | WASH−B (37) | 212 | 178 | 1.74 (1.50, 2.01) | 0.28 |
| Ghana | GHANA (40) | 83 | 82 | 2.52 (1.49, 4.25) | 0.06 |
| Kenya | WASH−B (42) | 298 | 259 | 1.69 (1.50, 1.91) | 0.31 |
| **I² = 0.56, Tau² = 0.01** | | **1143** | **791** | **1.67 (1.47, 1.90)** | |

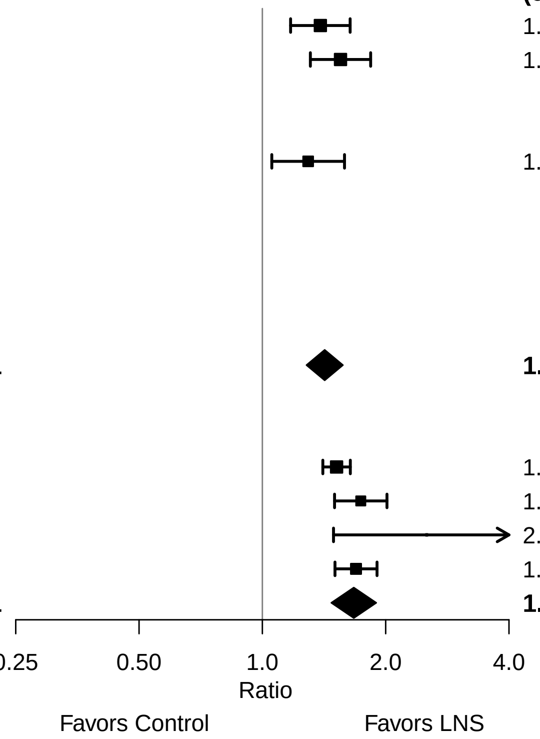

#### Supplemental figure 4F: Geometric mean ratio of ferritin concentration

#### 4F9: Stratified by Frequency of contact

**Frequency of contact**
**(p−diff = 0.584)**

| Country | Trial | N | N | GMR (95% CI) | W |
|---|---|---|---|---|---|
| **Frequency of contact − Monthly** | | | | | |
| Bangladesh | RDNS (36) | 550 | 272 | 1.52 (1.40, 1.64) | 0.40 |
| Burkina Faso | PROMIS CS (39) | | | | |
| Kenya | WASH−B (42) | 298 | 259 | 1.69 (1.50, 1.91) | 0.35 |
| Madagascar | MAHAY (43) | 83 | 51 | 1.29 (1.05, 1.59) | 0.24 |
| Mali | PROMIS CS (46) | | | | |
| Zimbabwe | SHINE (HIV−) (47) | | | | |
| Zimbabwe | SHINE (HIV+) (48) | | | | |
| **I² = 0.63, Tau² = 0.01** | | **931** | **582** | **1.52 (1.32, 1.75)** | |
| **Frequency of contact − Weekly** | | | | | |
| Bangladesh | JiVitA−4 (35) | 455 | 144 | 1.39 (1.17, 1.64) | 0.30 |
| Bangladesh | WASH−B (37) | 212 | 178 | 1.74 (1.50, 2.01) | 0.33 |
| Burkina Faso | iLiNS−Zinc (38) | 315 | 96 | 1.55 (1.31, 1.84) | 0.30 |
| Ghana | GHANA (40) | 83 | 82 | 2.52 (1.49, 4.25) | 0.07 |
| Ghana | iLiNS−DYADG (41) | | | | |
| Malawi | iLiNS−DYADM (44) | | | | |
| Malawi | iLiNS−DOSE (45) | | | | |
| **I² = 0.57, Tau² = 0.02** | | **1065** | **500** | **1.63 (1.35, 1.97)** | |

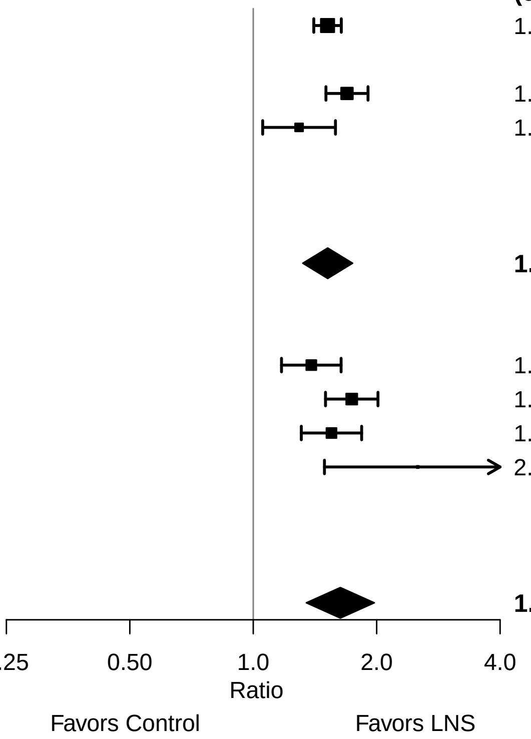

**Supplemental figure 4F: Geometric mean ratio of ferritin concentration**

#### 4F10: Stratified by Average SQ-LNS compliance (insufficient comparisons)

### Supplemental figure 4G: Iron deficiency (ferritin < 12 µg/L) prevalence ratio

#### 4G1: Stratified by Geographic region

**Geographic region**
**(p−diff = 0.016)**

| Country | Trial | N | N | PR (95% CI) | W |
|---|---|---|---|---|---|
| **Geographic region − SEAR** | | | | | |
| Bangladesh | JiVitA−4 (35) | 455 | 144 | 0.36 (0.21, 0.59) | 0.29 |
| Bangladesh | RDNS (36) | 550 | 272 | 0.34 (0.24, 0.48) | 0.41 |
| Bangladesh | WASH−B (37) | 212 | 178 | 0.24 (0.15, 0.39) | 0.30 |
| **I² = 0.00, Tau² = 0.00** | | **1217** | **594** | **0.31 (0.24, 0.40)** | |
| **Geographic region − AFR** | | | | | |
| Burkina Faso | iLiNS−Zinc (38) | 315 | 96 | 0.61 (0.49, 0.78) | 0.32 |
| Burkina Faso | PROMIS CS (39) | | | | |
| Ghana | GHANA (40) | 83 | 82 | 0.49 (0.33, 0.73) | 0.23 |
| Ghana | iLiNS−DYADG (41) | | | | |
| Kenya | WASH−B (42) | 298 | 259 | 0.41 (0.32, 0.53) | 0.31 |
| Madagascar | MAHAY (43) | 83 | 51 | 0.38 (0.21, 0.67) | 0.15 |
| Malawi | iLiNS−DYADM (44) | | | | |
| Malawi | iLiNS−DOSE (45) | | | | |
| Mali | PROMIS CS (46) | | | | |
| Zimbabwe | SHINE (HIV−) (47) | | | | |
| Zimbabwe | SHINE (HIV+) (48) | | | | |
| **I² = 0.51, Tau² = 0.02** | | **779** | **488** | **0.49 (0.40, 0.60)** | |

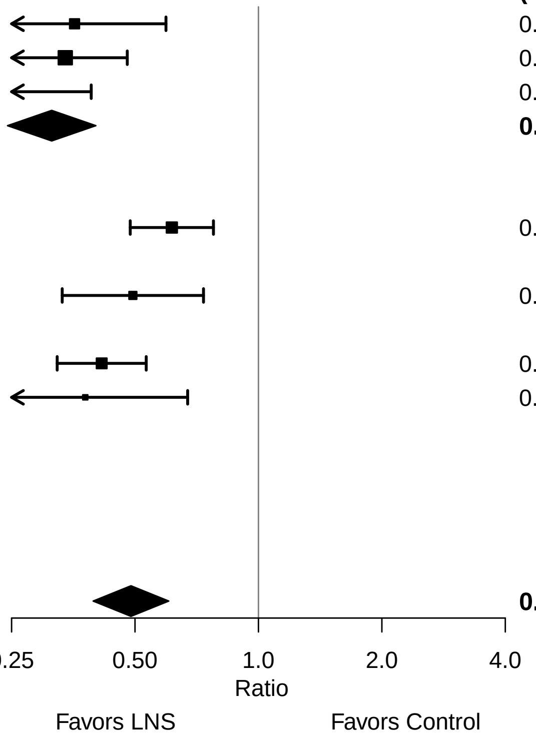

**Supplemental figure 4G: Iron deficiency (ferritin < 12 µg/L) prevalence ratio**

#### 4G2: Stratified by Anemia burden (insufficient comparisons)

**Supplemental figure 4G: Iron deficiency (ferritin < 12 µg/L) prevalence ratio**

#### 4G3: Stratified by Malaria prevalence (insufficient comparisons)

**Supplemental figure 4G: Iron deficiency (ferritin < 12 µg/L) prevalence ratio**

#### 4G4: Stratified by Inflammation burden (insufficient comparisons)

#### Supplemental figure 4G: Iron deficiency (ferritin < 12 µg/L) prevalence ratio

#### 4G5: Stratified by Source water quality

**Source water quality**
**(p−diff = 0.105)**

| Country | Trial | N | N | PR (95% CI) | W |
|---|---|---|---|---|---|
| **Source water quality − Improved** | | | | | |
| Bangladesh | JiVitA−4 (35) | 455 | 144 | 0.36 (0.21, 0.59) | 0.21 |
| Bangladesh | RDNS (36) | 550 | 272 | 0.34 (0.24, 0.48) | 0.36 |
| Bangladesh | WASH−B (37) | 94 | 58 | 0.31 (0.15, 0.64) | 0.12 |
| Ghana | GHANA (40) | 83 | 82 | 0.49 (0.33, 0.73) | 0.31 |
| Ghana | iLiNS−DYADG (41) | | | | |
| Malawi | iLiNS−DYADM (44) | | | | |
| Malawi | iLiNS−DOSE (45) | | | | |
| **I² = 0.00, Tau² = 0.00** | | **1182** | **556** | **0.38 (0.31, 0.48)** | |
| **Source water quality − Unimproved** | | | | | |
| Burkina Faso | iLiNS−Zinc (38) | 315 | 96 | 0.61 (0.49, 0.78) | 0.49 |
| Burkina Faso | PROMIS CS (39) | | | | |
| Kenya | WASH−B (42) | 166 | 144 | 0.45 (0.32, 0.63) | 0.34 |
| Madagascar | MAHAY (43) | 83 | 51 | 0.38 (0.21, 0.67) | 0.17 |
| Mali | PROMIS CS (46) | | | | |
| Zimbabwe | SHINE (HIV−) (47) | | | | |
| Zimbabwe | SHINE (HIV+) (48) | | | | |
| **I² = 0.47, Tau² = 0.02** | | **564** | **291** | **0.51 (0.39, 0.66)** | |

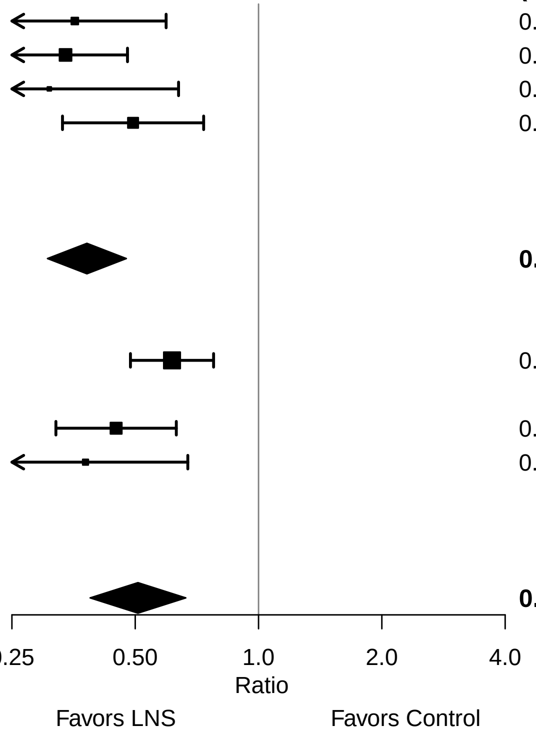

#### Supplemental figure 4G: Iron deficiency (ferritin < 12 µg/L) prevalence ratio

### 4G6: Stratified by Sanitation

**Sanitation**
**(p−diff = 0.105)**

| Country | Trial | N | N | PR (95% CI) | W |
|---|---|---|---|---|---|
| **Sanitation − Improved** | | | | | |
| Bangladesh | JiVitA−4 (35) | 455 | 144 | 0.36 (0.21, 0.59) | 0.21 |
| Bangladesh | RDNS (36) | 550 | 272 | 0.34 (0.24, 0.48) | 0.36 |
| Bangladesh | WASH−B (37) | 94 | 58 | 0.31 (0.15, 0.64) | 0.12 |
| Burkina Faso | PROMIS CS (39) | | | | |
| Ghana | GHANA (40) | 83 | 82 | 0.49 (0.33, 0.73) | 0.31 |
| Ghana | iLiNS−DYADG (41) | | | | |
| Mali | PROMIS CS (46) | | | | |
| **I² = 0.00, Tau² = 0.00** | | **1182** | **556** | **0.38 (0.31, 0.48)** | |
| **Sanitation − Unimproved** | | | | | |
| Burkina Faso | iLiNS−Zinc (38) | 315 | 96 | 0.61 (0.49, 0.78) | 0.49 |
| Kenya | WASH−B (42) | 166 | 144 | 0.45 (0.32, 0.63) | 0.34 |
| Madagascar | MAHAY (43) | 83 | 51 | 0.38 (0.21, 0.67) | 0.17 |
| Malawi | iLiNS−DYADM (44) | | | | |
| Malawi | iLiNS−DOSE (45) | | | | |
| Zimbabwe | SHINE (HIV−) (47) | | | | |
| Zimbabwe | SHINE (HIV+) (48) | | | | |
| **I² = 0.47, Tau² = 0.02** | | **564** | **291** | **0.51 (0.39, 0.66)** | |

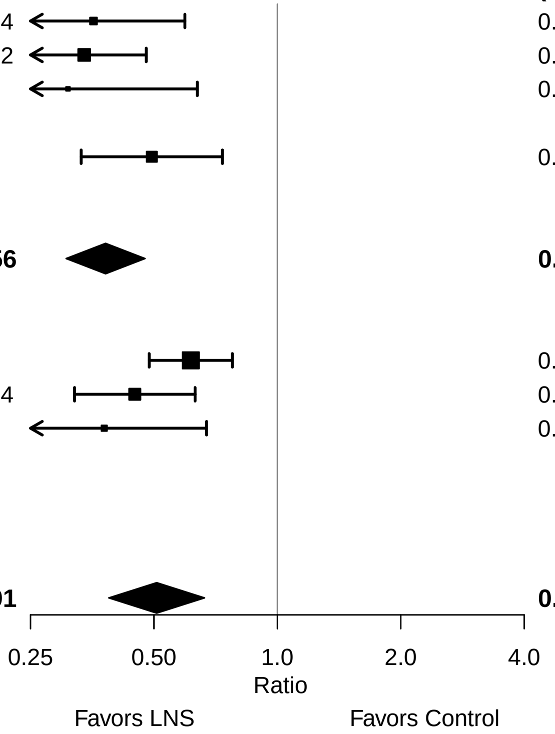

#### Supplemental figure 4G: Iron deficiency (ferritin < 12 µg/L) prevalence ratio

##### 4G7: Stratified by Supplement duration

**Supplement duration**
**(p−diff = 0.081)**

| Country | Trial | N | N | PR (95% CI) | W |
|---|---|---|---|---|---|
| **Supplement duration − 12m or less** | | | | | |
| Bangladesh | JiVitA−4 (35) | 455 | 144 | 0.36 (0.21, 0.59) | 0.20 |
| Burkina Faso | iLiNS−Zinc (38) | 315 | 96 | 0.61 (0.49, 0.78) | 0.36 |
| Burkina Faso | PROMIS CS (39) | | | | |
| Ghana | GHANA (40) | 83 | 82 | 0.49 (0.33, 0.73) | 0.26 |
| Ghana | iLiNS−DYADG (41) | | | | |
| Madagascar | MAHAY (43) | 83 | 51 | 0.38 (0.21, 0.67) | 0.18 |
| Malawi | iLiNS−DYADM (44) | | | | |
| Malawi | iLiNS−DOSE (45) | | | | |
| Zimbabwe | SHINE (HIV−) (47) | | | | |
| Zimbabwe | SHINE (HIV+) (48) | | | | |
| **I² = 0.43, Tau² = 0.03** | | **936** | **373** | **0.49 (0.38, 0.63)** | |
| **Supplement duration − > 12m** | | | | | |
| Bangladesh | RDNS (36) | 550 | 272 | 0.34 (0.24, 0.48) | 0.34 |
| Bangladesh | WASH−B (37) | 212 | 178 | 0.24 (0.15, 0.39) | 0.25 |
| Kenya | WASH−B (42) | 298 | 259 | 0.41 (0.32, 0.53) | 0.41 |
| Mali | PROMIS CS (46) | | | | |
| **I² = 0.50, Tau² = 0.04** | | **1060** | **709** | **0.34 (0.25, 0.46)** | |

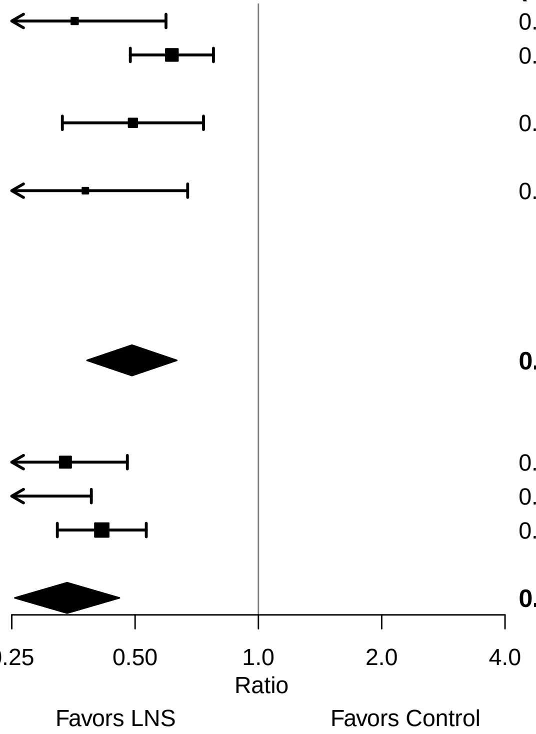

**Supplemental figure 4G: Iron deficiency (ferritin < 12 µg/L) prevalence ratio**

#### 4G8: Stratified by Iron dose

**Iron dose**
**(p−diff = 0.278)**

| Country | Trial | N | N | PR (95% CI) | W |
|---|---|---|---|---|---|
| **Iron dose − Less than 9 mg** | | | | | |
| Bangladesh | JiVitA−4 (35) | 455 | 144 | 0.36 (0.21, 0.59) | 0.27 |
| Burkina Faso | iLiNS−Zinc (38) | 315 | 96 | 0.61 (0.49, 0.78) | 0.49 |
| Burkina Faso | PROMIS CS (39) | | | | |
| Ghana | iLiNS−DYADG (41) | | | | |
| Madagascar | MAHAY (43) | 83 | 51 | 0.38 (0.21, 0.67) | 0.24 |
| Malawi | iLiNS−DYADM (44) | | | | |
| Malawi | iLiNS−DOSE (45) | | | | |
| Mali | PROMIS CS (46) | | | | |
| Zimbabwe | SHINE (HIV−) (47) | | | | |
| Zimbabwe | SHINE (HIV+) (48) | | | | |
| **I² = 0.61, Tau² = 0.06** | | **853** | **291** | **0.47 (0.33, 0.68)** | |
| **Iron dose − 9 mg** | | | | | |
| Bangladesh | RDNS (36) | 550 | 272 | 0.34 (0.24, 0.48) | 0.26 |
| Bangladesh | WASH−B (37) | 212 | 178 | 0.24 (0.15, 0.39) | 0.19 |
| Ghana | GHANA (40) | 83 | 82 | 0.49 (0.33, 0.73) | 0.23 |
| Kenya | WASH−B (42) | 298 | 259 | 0.41 (0.32, 0.53) | 0.32 |
| **I² = 0.50, Tau² = 0.05** | | **1143** | **791** | **0.37 (0.28, 0.49)** | |

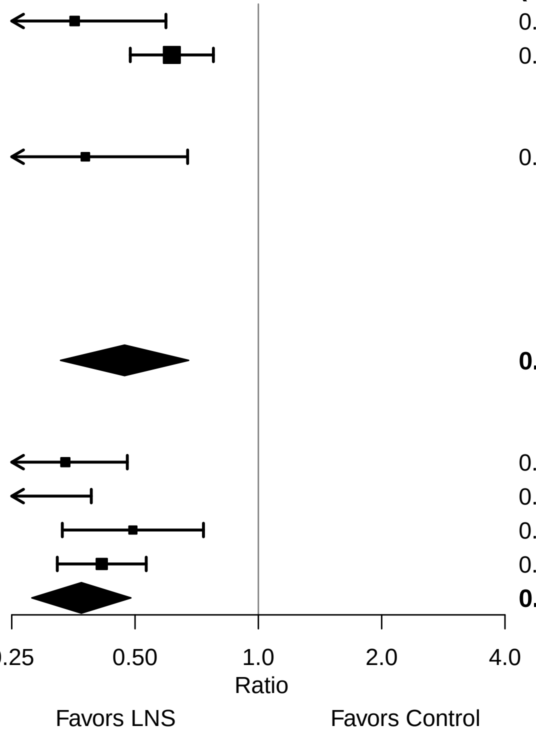

**Supplemental figure 4G: Iron deficiency (ferritin < 12 µg/L) prevalence ratio**

#### 4G9: Stratified by Frequency of contact

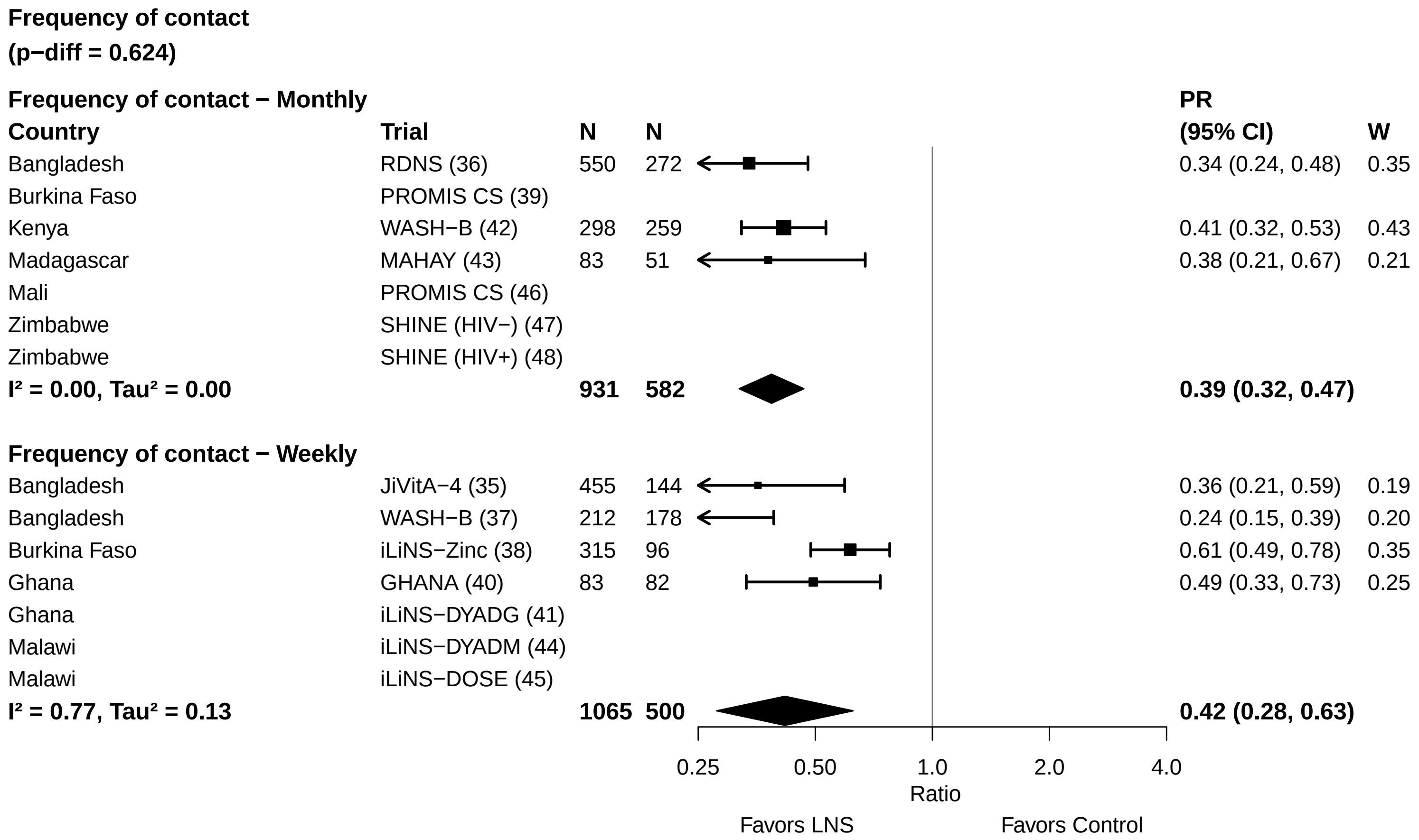

**Supplemental figure 4G: Iron deficiency (ferritin < 12 µg/L) prevalence ratio**

#### 4G10: Stratified by Average SQ-LNS compliance (insufficient comparisons)

### Supplemental figure 4H: Iron deficiency (ferritin < 12 μg/L) prevalence difference

#### 4H1: Stratified by Geographic region

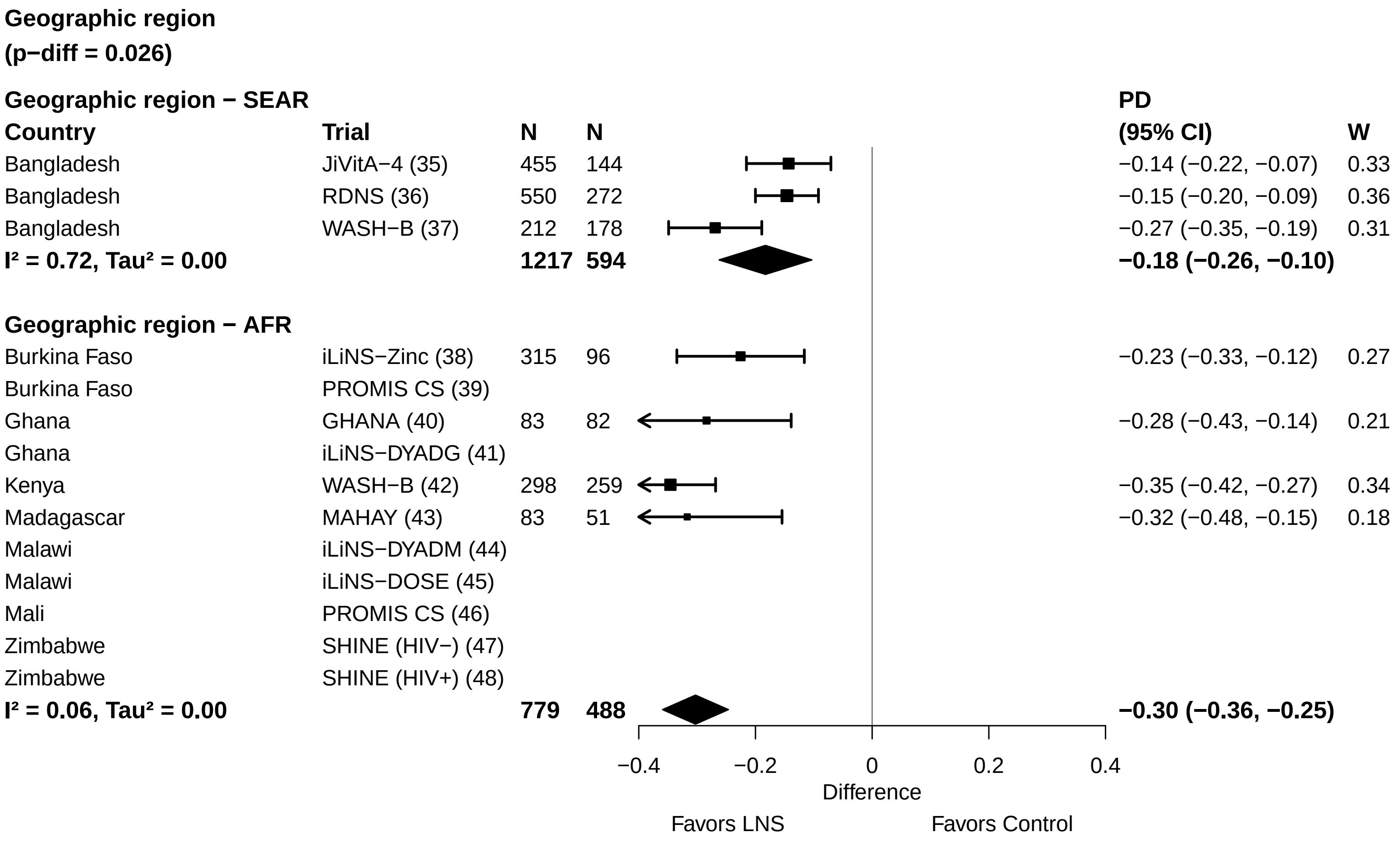

**Supplemental figure 4H: Iron deficiency (ferritin < 12 µg/L) prevalence difference**

#### 4H2: Stratified by Anemia burden (insufficient comparisons)

**Supplemental figure 4H: Iron deficiency (ferritin < 12 µg/L) prevalence difference**

#### 4H3: Stratified by Malaria prevalence (insufficient comparisons)

**Supplemental figure 4H: Iron deficiency (ferritin < 12 µg/L) prevalence difference**

#### 4H4: Stratified by Inflammation burden (insufficient comparisons)

**Supplemental figure 4H: Iron deficiency (ferritin < 12 µg/L) prevalence difference**

#### 4H5: Stratified by Source water quality

**Source water quality**
**(p−diff = 0.006)**

**Source water quality − Improved**

| Country | Trial | N | N | PD (95% CI) | W |
|---|---|---|---|---|---|
| Bangladesh | JiVitA−4 (35) | 455 | 144 | −0.14 (−0.22, −0.07) | 0.31 |
| Bangladesh | RDNS (36) | 550 | 272 | −0.15 (−0.20, −0.09) | 0.36 |
| Bangladesh | WASH−B (37) | 94 | 58 | −0.24 (−0.38, −0.10) | 0.17 |
| Ghana | GHANA (40) | 83 | 82 | −0.28 (−0.43, −0.14) | 0.16 |
| Ghana | iLiNS−DYADG (41) | | | | |
| Malawi | iLiNS−DYADM (44) | | | | |
| Malawi | iLiNS−DOSE (45) | | | | |
| **I² = 0.33, Tau² = 0.00** | | **1182** | **556** | **−0.18 (−0.24, −0.12)** | |

**Source water quality − Unimproved**

| Country | Trial | N | N | PD (95% CI) | W |
|---|---|---|---|---|---|
| Burkina Faso | iLiNS−Zinc (38) | 315 | 96 | −0.23 (−0.33, −0.12) | 0.38 |
| Burkina Faso | PROMIS CS (39) | | | | |
| Kenya | WASH−B (42) | 166 | 144 | −0.33 (−0.43, −0.22) | 0.39 |
| Madagascar | MAHAY (43) | 83 | 51 | −0.32 (−0.48, −0.15) | 0.23 |
| Mali | PROMIS CS (46) | | | | |
| Zimbabwe | SHINE (HIV−) (47) | | | | |
| Zimbabwe | SHINE (HIV+) (48) | | | | |
| **I² = 0.00, Tau² = 0.00** | | **564** | **291** | **−0.28 (−0.35, −0.22)** | |

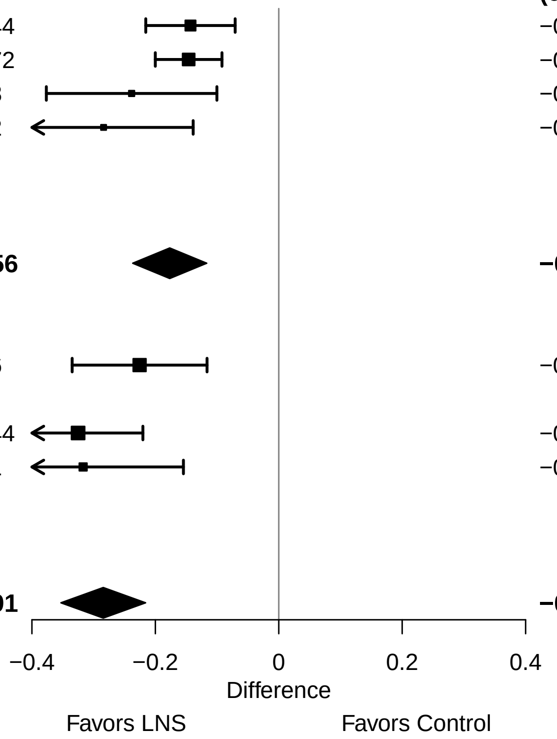

#### Supplemental figure 4H: Iron deficiency (ferritin < 12 µg/L) prevalence difference

#### 4H6: Stratified by Sanitation

**Sanitation**
**(p−diff = 0.006)**

| Country | Trial | N | N | PD (95% CI) | W |
|---|---|---|---|---|---|
| **Sanitation − Improved** | | | | | |
| Bangladesh | JiVitA−4 (35) | 455 | 144 | −0.14 (−0.22, −0.07) | 0.31 |
| Bangladesh | RDNS (36) | 550 | 272 | −0.15 (−0.20, −0.09) | 0.36 |
| Bangladesh | WASH−B (37) | 94 | 58 | −0.24 (−0.38, −0.10) | 0.17 |
| Burkina Faso | PROMIS CS (39) | | | | |
| Ghana | GHANA (40) | 83 | 82 | −0.28 (−0.43, −0.14) | 0.16 |
| Ghana | iLiNS−DYADG (41) | | | | |
| Mali | PROMIS CS (46) | | | | |
| **I² = 0.33, Tau² = 0.00** | | **1182** | **556** | **−0.18 (−0.24, −0.12)** | |
| **Sanitation − Unimproved** | | | | | |
| Burkina Faso | iLiNS−Zinc (38) | 315 | 96 | −0.23 (−0.33, −0.12) | 0.38 |
| Kenya | WASH−B (42) | 166 | 144 | −0.33 (−0.43, −0.22) | 0.39 |
| Madagascar | MAHAY (43) | 83 | 51 | −0.32 (−0.48, −0.15) | 0.23 |
| Malawi | iLiNS−DYADM (44) | | | | |
| Malawi | iLiNS−DOSE (45) | | | | |
| Zimbabwe | SHINE (HIV−) (47) | | | | |
| Zimbabwe | SHINE (HIV+) (48) | | | | |
| **I² = 0.00, Tau² = 0.00** | | **564** | **291** | **−0.28 (−0.35, −0.22)** | |

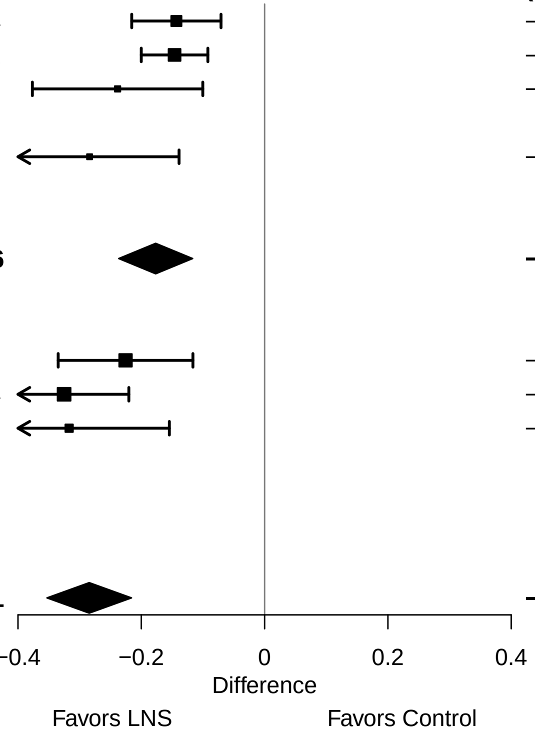

#### Supplemental figure 4H: Iron deficiency (ferritin < 12 μg/L) prevalence difference

##### 4H7: Stratified by Supplement duration

**Supplement duration**
**(p−diff = 0.767)**

| Country | Trial | N | N | PD (95% CI) | W |
|---|---|---|---|---|---|
| **Supplement duration − 12m or less** | | | | | |
| Bangladesh | JiVitA−4 (35) | 455 | 144 | −0.14 (−0.22, −0.07) | 0.35 |
| Burkina Faso | iLiNS−Zinc (38) | 315 | 96 | −0.23 (−0.33, −0.12) | 0.27 |
| Burkina Faso | PROMIS CS (39) | | | | |
| Ghana | GHANA (40) | 83 | 82 | −0.28 (−0.43, −0.14) | 0.20 |
| Ghana | iLiNS−DYADG (41) | | | | |
| Madagascar | MAHAY (43) | 83 | 51 | −0.32 (−0.48, −0.15) | 0.18 |
| Malawi | iLiNS−DYADM (44) | | | | |
| Malawi | iLiNS−DOSE (45) | | | | |
| Zimbabwe | SHINE (HIV−) (47) | | | | |
| Zimbabwe | SHINE (HIV+) (48) | | | | |
| **I² = 0.49, Tau² = 0.00** | | **936** | **373** | **−0.22 (−0.30, −0.14)** | |
| **Supplement duration − > 12m** | | | | | |
| Bangladesh | RDNS (36) | 550 | 272 | −0.15 (−0.20, −0.09) | 0.37 |
| Bangladesh | WASH−B (37) | 212 | 178 | −0.27 (−0.35, −0.19) | 0.31 |
| Kenya | WASH−B (42) | 298 | 259 | −0.35 (−0.42, −0.27) | 0.32 |
| Mali | PROMIS CS (46) | | | | |
| **I² = 0.89, Tau² = 0.01** | | **1060** | **709** | **−0.25 (−0.37, −0.14)** | |

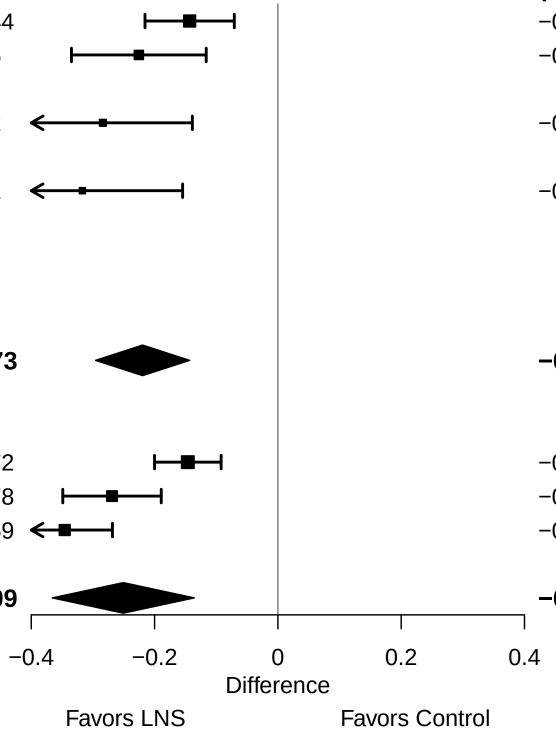

#### Supplemental figure 4H: Iron deficiency (ferritin < 12 µg/L) prevalence difference

#### 4H8: Stratified by Iron dose

**Iron dose**
**(p−diff = 0.548)**

| Country | Trial | N | N | PD (95% CI) | W |
|---|---|---|---|---|---|
| **Iron dose − Less than 9 mg** | | | | | |
| Bangladesh | JiVitA−4 (35) | 455 | 144 | −0.14 (−0.22, −0.07) | 0.44 |
| Burkina Faso | iLiNS−Zinc (38) | 315 | 96 | −0.23 (−0.33, −0.12) | 0.34 |
| Burkina Faso | PROMIS CS (39) | | | | |
| Ghana | iLiNS−DYADG (41) | | | | |
| Madagascar | MAHAY (43) | 83 | 51 | −0.32 (−0.48, −0.15) | 0.23 |
| Malawi | iLiNS−DYADM (44) | | | | |
| Malawi | iLiNS−DOSE (45) | | | | |
| Mali | PROMIS CS (46) | | | | |
| Zimbabwe | SHINE (HIV−) (47) | | | | |
| Zimbabwe | SHINE (HIV+) (48) | | | | |
| **I² = 0.54, Tau² = 0.00** | | **853** | **291** | **−0.21 (−0.30, −0.11)** | |
| **Iron dose − 9 mg** | | | | | |
| Bangladesh | RDNS (36) | 550 | 272 | −0.15 (−0.20, −0.09) | 0.31 |
| Bangladesh | WASH−B (37) | 212 | 178 | −0.27 (−0.35, −0.19) | 0.26 |
| Ghana | GHANA (40) | 83 | 82 | −0.28 (−0.43, −0.14) | 0.16 |
| Kenya | WASH−B (42) | 298 | 259 | −0.35 (−0.42, −0.27) | 0.27 |
| **I² = 0.84, Tau² = 0.01** | | **1143** | **791** | **−0.26 (−0.34, −0.17)** | |

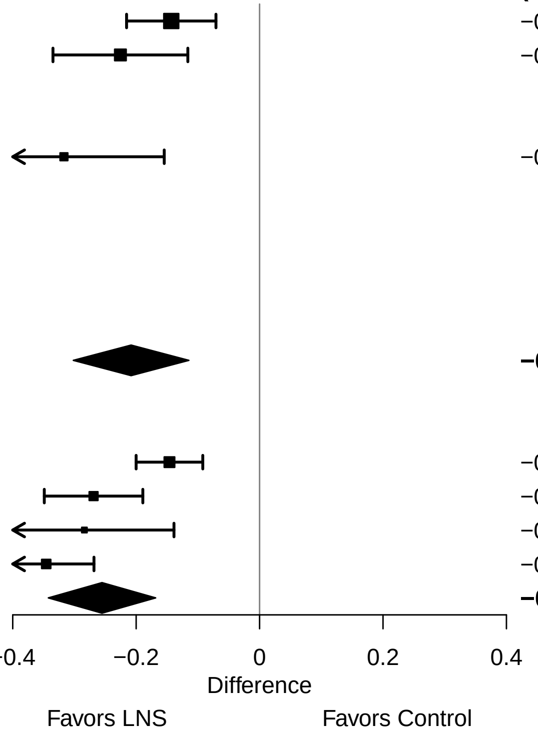

#### Supplemental figure 4H: Iron deficiency (ferritin < 12 µg/L) prevalence difference

### 4H9: Stratified by Frequency of contact

**Frequency of contact**
**(p−diff = 0.650)**

| | | | | | |
|---|---|---|---|---|---|
| **Frequency of contact − Monthly** | | | | | |
| **Country** | **Trial** | **N** | **N** | **PD (95% CI)** | **W** |
| Bangladesh | RDNS (36) | 550 | 272 | −0.15 (−0.20, −0.09) | 0.43 |
| Burkina Faso | PROMIS CS (39) | | | | |
| Kenya | WASH−B (42) | 298 | 259 | −0.35 (−0.42, −0.27) | 0.37 |
| Madagascar | MAHAY (43) | 83 | 51 | −0.32 (−0.48, −0.15) | 0.20 |
| Mali | PROMIS CS (46) | | | | |
| Zimbabwe | SHINE (HIV−) (47) | | | | |
| Zimbabwe | SHINE (HIV+) (48) | | | | |
| **I² = 0.89, Tau² = 0.01** | | **931** | **582** | **−0.26 (−0.39, −0.13)** | |
| **Frequency of contact − Weekly** | | | | | |
| Bangladesh | JiVitA−4 (35) | 455 | 144 | −0.14 (−0.22, −0.07) | 0.30 |
| Bangladesh | WASH−B (37) | 212 | 178 | −0.27 (−0.35, −0.19) | 0.29 |
| Burkina Faso | iLiNS−Zinc (38) | 315 | 96 | −0.23 (−0.33, −0.12) | 0.23 |
| Ghana | GHANA (40) | 83 | 82 | −0.28 (−0.43, −0.14) | 0.18 |
| Ghana | iLiNS−DYADG (41) | | | | |
| Malawi | iLiNS−DYADM (44) | | | | |
| Malawi | iLiNS−DOSE (45) | | | | |
| **I² = 0.53, Tau² = 0.00** | | **1065** | **500** | **−0.22 (−0.29, −0.16)** | |

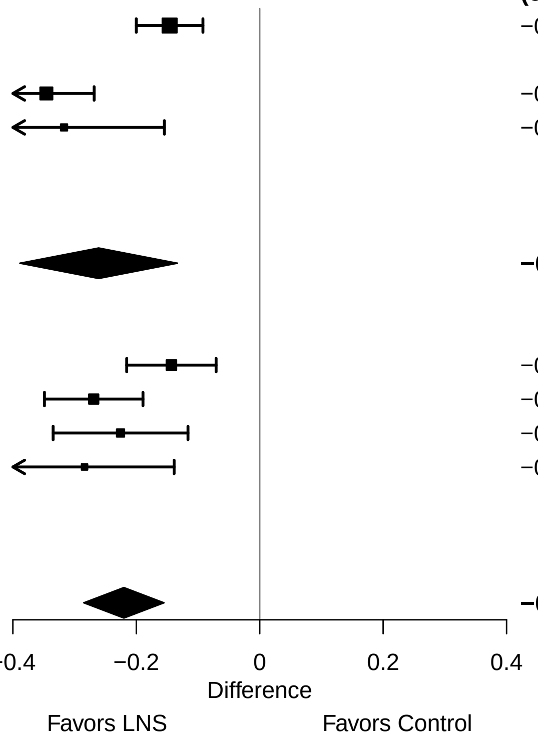

**Supplemental figure 4H: Iron deficiency (ferritin < 12 µg/L) prevalence difference**

#### 4H10: Stratified by Average SQ-LNS compliance (insufficient comparisons)

### Supplemental figure 4I: Iron deficiency anemia prevalence ratio

#### 4I1: Stratified by Geographic region (insufficient comparisons)

**Supplemental figure 4I: Iron deficiency anemia prevalence ratio**

#### 4I2: Stratified by Anemia burden (insufficient comparisons)

**Supplemental figure 4I: Iron deficiency anemia prevalence ratio**

#### 4I3: Stratified by Malaria prevalence (insufficient comparisons)

**Supplemental figure 4I: Iron deficiency anemia prevalence ratio**

#### 4I4: Stratified by Inflammation burden (insufficient comparisons)

#### Supplemental figure 4I: Iron deficiency anemia prevalence ratio

### 4I5: Stratified by Source water quality

**Source water quality**
**(p−diff = 0.012)**

| Country | Trial | N | N | PR (95% CI) | W |
|---|---|---|---|---|---|
| **Source water quality − Improved** | | | | | |
| Bangladesh | JiVitA−4 (35) | 454 | 144 | 0.12 (0.04, 0.34) | 0.24 |
| Bangladesh | RDNS (36) | 549 | 272 | 0.25 (0.15, 0.41) | 0.48 |
| Bangladesh | WASH−B (37) | | | | |
| Ghana | GHANA (40) | 83 | 82 | 0.17 (0.07, 0.42) | 0.28 |
| Ghana | iLiNS−DYADG (41) | | | | |
| Malawi | iLiNS−DYADM (44) | | | | |
| Malawi | iLiNS−DOSE (45) | | | | |
| **I² = 0.00, Tau² = 0.00** | | **1086** | **498** | **0.20 (0.14, 0.31)** | |
| **Source water quality − Unimproved** | | | | | |
| Burkina Faso | iLiNS−Zinc (38) | 327 | 101 | 0.51 (0.39, 0.67) | 0.45 |
| Burkina Faso | PROMIS CS (39) | | | | |
| Kenya | WASH−B (42) | 166 | 143 | 0.33 (0.19, 0.56) | 0.33 |
| Madagascar | MAHAY (43) | 83 | 51 | 0.31 (0.13, 0.71) | 0.22 |
| Mali | PROMIS CS (46) | | | | |
| Zimbabwe | SHINE (HIV−) (47) | | | | |
| Zimbabwe | SHINE (HIV+) (48) | | | | |
| **I² = 0.35, Tau² = 0.03** | | **576** | **295** | **0.43 (0.31, 0.59)** | |

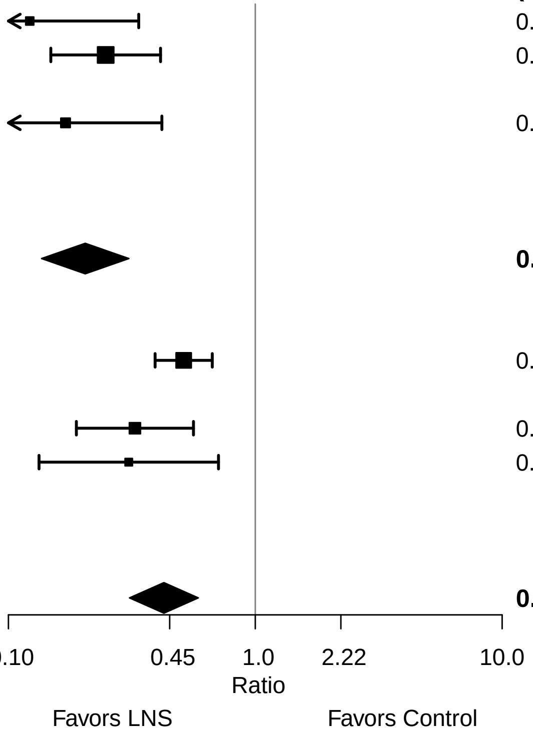

**Supplemental figure 4I: Iron deficiency anemia prevalence ratio**

#### 4I6: Stratified by Sanitation

**Sanitation**
**(p−diff = 0.012)**

| Country | Trial | N | N | PR (95% CI) | W |
|---|---|---|---|---|---|
| **Sanitation − Improved** | | | | | |
| Bangladesh | JiVitA−4 (35) | 454 | 144 | 0.12 (0.04, 0.34) | 0.24 |
| Bangladesh | RDNS (36) | 549 | 272 | 0.25 (0.15, 0.41) | 0.48 |
| Bangladesh | WASH−B (37) | | | | |
| Burkina Faso | PROMIS CS (39) | | | | |
| Ghana | GHANA (40) | 83 | 82 | 0.17 (0.07, 0.42) | 0.28 |
| Ghana | iLiNS−DYADG (41) | | | | |
| Mali | PROMIS CS (46) | | | | |
| **I² = 0.00, Tau² = 0.00** | | **1086** | **498** | **0.20 (0.14, 0.31)** | |
| **Sanitation − Unimproved** | | | | | |
| Burkina Faso | iLiNS−Zinc (38) | 327 | 101 | 0.51 (0.39, 0.67) | 0.45 |
| Kenya | WASH−B (42) | 166 | 143 | 0.33 (0.19, 0.56) | 0.33 |
| Madagascar | MAHAY (43) | 83 | 51 | 0.31 (0.13, 0.71) | 0.22 |
| Malawi | iLiNS−DYADM (44) | | | | |
| Malawi | iLiNS−DOSE (45) | | | | |
| Zimbabwe | SHINE (HIV−) (47) | | | | |
| Zimbabwe | SHINE (HIV+) (48) | | | | |
| **I² = 0.35, Tau² = 0.03** | | **576** | **295** | **0.43 (0.31, 0.59)** | |

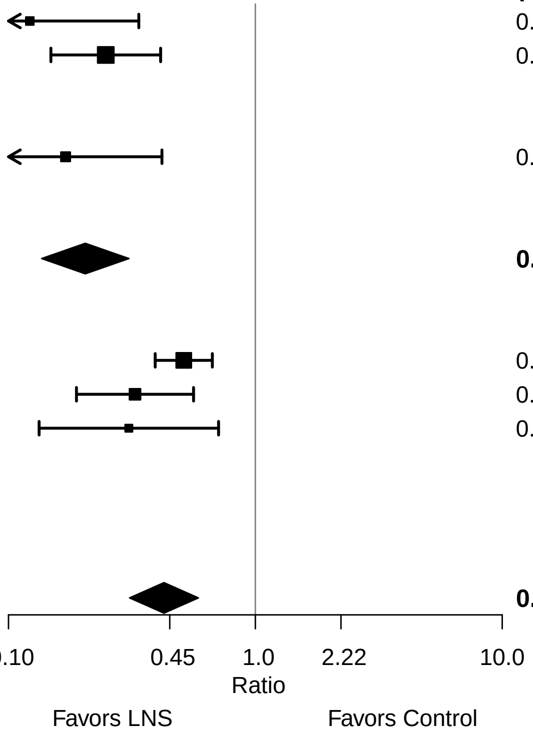

**Supplemental figure 4I: Iron deficiency anemia prevalence ratio**

#### 4I7: Stratified by Supplement duration (insufficient comparisons)

**Supplemental figure 4I: Iron deficiency anemia prevalence ratio**

#### 4I8: Stratified by Iron dose

**Iron dose**
**(p−diff = 0.402)**

| Country | Trial | N | N | PR (95% CI) | W |
|---|---|---|---|---|---|
| **Iron dose − Less than 9 mg** | | | | | |
| Bangladesh | JiVitA−4 (35) | 454 | 144 | 0.12 (0.04, 0.34) | 0.20 |
| Burkina Faso | iLiNS−Zinc (38) | 327 | 101 | 0.51 (0.39, 0.67) | 0.54 |
| Burkina Faso | PROMIS CS (39) | | | | |
| Ghana | iLiNS−DYADG (41) | | | | |
| Madagascar | MAHAY (43) | 83 | 51 | 0.31 (0.13, 0.71) | 0.26 |
| Malawi | iLiNS−DYADM (44) | | | | |
| Malawi | iLiNS−DOSE (45) | | | | |
| Mali | PROMIS CS (46) | | | | |
| Zimbabwe | SHINE (HIV−) (47) | | | | |
| Zimbabwe | SHINE (HIV+) (48) | | | | |
| **I² = 0.75, Tau² = 0.37** | | **864** | **296** | **0.30 (0.13, 0.67)** | |
| **Iron dose − 9 mg** | | | | | |
| Bangladesh | RDNS (36) | 549 | 272 | 0.25 (0.15, 0.41) | 0.38 |
| Bangladesh | WASH−B (37) | | | | |
| Ghana | GHANA (40) | 83 | 82 | 0.17 (0.07, 0.42) | 0.22 |
| Kenya | WASH−B (42) | 298 | 258 | 0.28 (0.18, 0.45) | 0.40 |
| **I² = 0.00, Tau² = 0.00** | | **930** | **612** | **0.25 (0.18, 0.35)** | |

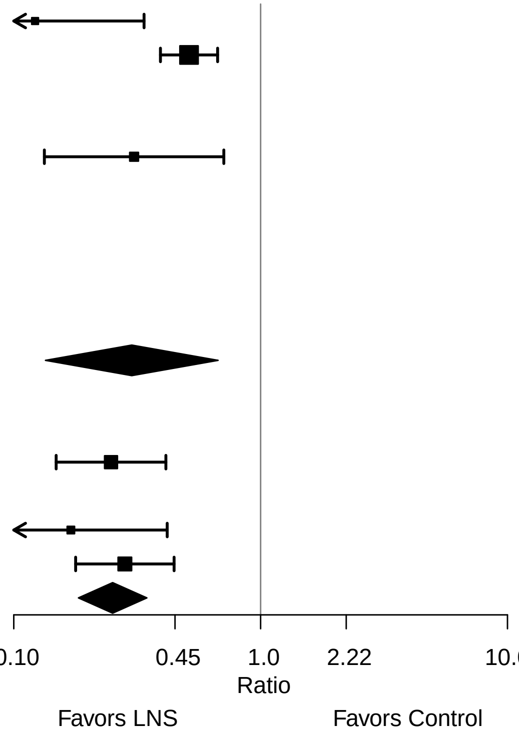

**Supplemental figure 4I: Iron deficiency anemia prevalence ratio**

#### 4I9: Stratified by Frequency of contact

**Frequency of contact**
**(p−diff = 0.981)**

| Country | Trial | N | N | PR (95% CI) | W |
|---|---|---|---|---|---|
| **Frequency of contact − Monthly** | | | | | |
| Bangladesh | RDNS (36) | 549 | 272 | 0.25 (0.15, 0.41) | 0.37 |
| Burkina Faso | PROMIS CS (39) | | | | |
| Kenya | WASH−B (42) | 298 | 258 | 0.28 (0.18, 0.45) | 0.40 |
| Madagascar | MAHAY (43) | 83 | 51 | 0.31 (0.13, 0.71) | 0.23 |
| Mali | PROMIS CS (46) | | | | |
| Zimbabwe | SHINE (HIV−) (47) | | | | |
| Zimbabwe | SHINE (HIV+) (48) | | | | |
| **I² = 0.00, Tau² = 0.00** | | **930** | **581** | **0.27 (0.20, 0.37)** | |
| **Frequency of contact − Weekly** | | | | | |
| Bangladesh | JiVitA−4 (35) | 454 | 144 | 0.12 (0.04, 0.34) | 0.21 |
| Bangladesh | WASH−B (37) | | | | |
| Burkina Faso | iLiNS−Zinc (38) | 327 | 101 | 0.51 (0.39, 0.67) | 0.55 |
| Ghana | GHANA (40) | 83 | 82 | 0.17 (0.07, 0.42) | 0.24 |
| Ghana | iLiNS−DYADG (41) | | | | |
| Malawi | iLiNS−DYADM (44) | | | | |
| Malawi | iLiNS−DOSE (45) | | | | |
| **I² = 0.83, Tau² = 0.46** | | **864** | **327** | **0.25 (0.10, 0.60)** | |

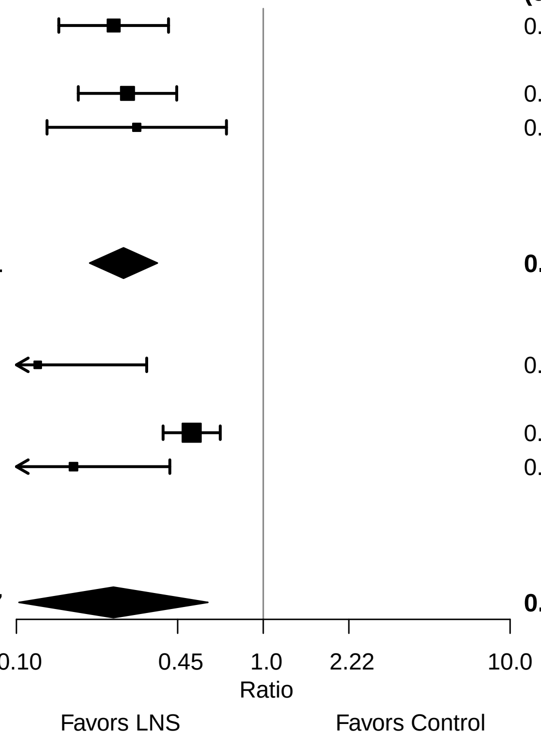

**Supplemental figure 4I: Iron deficiency anemia prevalence ratio**

#### 4I10: Stratified by Average SQ-LNS compliance (insufficient comparisons)

### Supplemental figure 4J: Iron deficiency anemia prevalence difference

#### 4J1: Stratified by Geographic region (insufficient comparisons)

**Supplemental figure 4J: Iron deficiency anemia prevalence difference**

#### 4J2: Stratified by Anemia burden (insufficient comparisons)

**Supplemental figure 4J: Iron deficiency anemia prevalence difference**

#### 4J3: Stratified by Malaria prevalence (insufficient comparisons)

**Supplemental figure 4J: Iron deficiency anemia prevalence difference**

#### 4J4: Stratified by Inflammation burden (insufficient comparisons)

#### Supplemental figure 4J: Iron deficiency anemia prevalence difference

### 4J5: Stratified by Source water quality

**Source water quality**
**(p−diff = 0.222)**

| Country | Trial | N | N | PD (95% CI) | W |
|---|---|---|---|---|---|
| **Source water quality − Improved** | | | | | |
| Bangladesh | JiVitA−4 (35) | 454 | 144 | −0.08 (−0.13, −0.03) | 0.37 |
| Bangladesh | RDNS (36) | 549 | 272 | −0.11 (−0.16, −0.07) | 0.38 |
| Bangladesh | WASH−B (37) | | | | |
| Ghana | GHANA (40) | 83 | 82 | −0.29 (−0.41, −0.18) | 0.25 |
| Ghana | iLiNS−DYADG (41) | | | | |
| Malawi | iLiNS−DYADM (44) | | | | |
| Malawi | iLiNS−DOSE (45) | | | | |
| **I² = 0.82, Tau² = 0.01** | | **1086** | **498** | **−0.15 (−0.28, −0.03)** | |
| **Source water quality − Unimproved** | | | | | |
| Burkina Faso | iLiNS−Zinc (38) | 327 | 101 | −0.27 (−0.37, −0.16) | 0.34 |
| Burkina Faso | PROMIS CS (39) | | | | |
| Kenya | WASH−B (42) | 166 | 143 | −0.21 (−0.30, −0.12) | 0.38 |
| Madagascar | MAHAY (43) | 83 | 51 | −0.19 (−0.33, −0.05) | 0.28 |
| Mali | PROMIS CS (46) | | | | |
| Zimbabwe | SHINE (HIV−) (47) | | | | |
| Zimbabwe | SHINE (HIV+) (48) | | | | |
| **I² = 0.00, Tau² = 0.00** | | **576** | **295** | **−0.23 (−0.29, −0.16)** | |

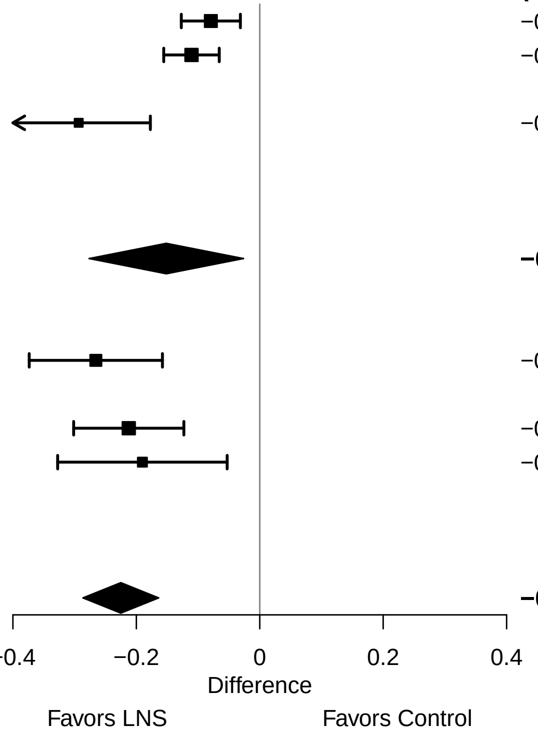

#### Supplemental figure 4J: Iron deficiency anemia prevalence difference

### 4J6: Stratified by Sanitation

**Sanitation**
**(p−diff = 0.222)**

| Country | Trial | N | N | PD (95% CI) | W |
|---|---|---|---|---|---|
| **Sanitation − Improved** | | | | | |
| Bangladesh | JiVitA−4 (35) | 454 | 144 | −0.08 (−0.13, −0.03) | 0.37 |
| Bangladesh | RDNS (36) | 549 | 272 | −0.11 (−0.16, −0.07) | 0.38 |
| Bangladesh | WASH−B (37) | | | | |
| Burkina Faso | PROMIS CS (39) | | | | |
| Ghana | GHANA (40) | 83 | 82 | −0.29 (−0.41, −0.18) | 0.25 |
| Ghana | iLiNS−DYADG (41) | | | | |
| Mali | PROMIS CS (46) | | | | |
| **I² = 0.82, Tau² = 0.01** | | **1086** | **498** | **−0.15 (−0.28, −0.03)** | |
| **Sanitation − Unimproved** | | | | | |
| Burkina Faso | iLiNS−Zinc (38) | 327 | 101 | −0.27 (−0.37, −0.16) | 0.34 |
| Kenya | WASH−B (42) | 166 | 143 | −0.21 (−0.30, −0.12) | 0.38 |
| Madagascar | MAHAY (43) | 83 | 51 | −0.19 (−0.33, −0.05) | 0.28 |
| Malawi | iLiNS−DYADM (44) | | | | |
| Malawi | iLiNS−DOSE (45) | | | | |
| Zimbabwe | SHINE (HIV−) (47) | | | | |
| Zimbabwe | SHINE (HIV+) (48) | | | | |
| **I² = 0.00, Tau² = 0.00** | | **576** | **295** | **−0.23 (−0.29, −0.16)** | |

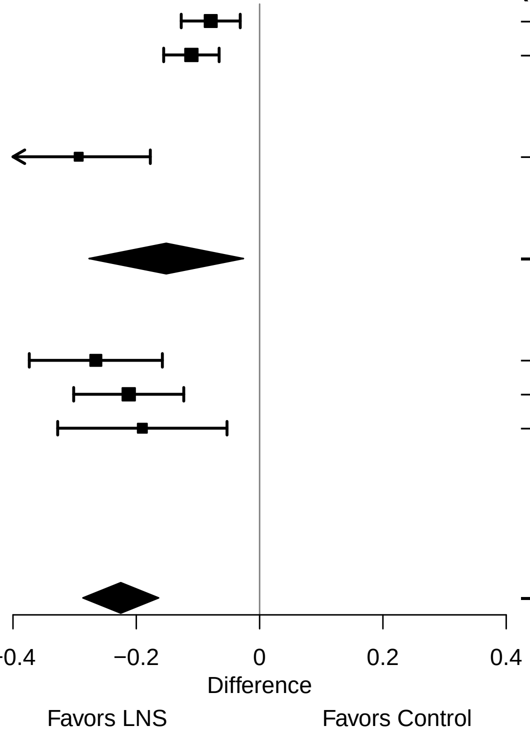

**Supplemental figure 4J: Iron deficiency anemia prevalence difference**

#### 4J7: Stratified by Supplement duration (insufficient comparisons)

**Supplemental figure 4J: Iron deficiency anemia prevalence difference**

#### 4J8: Stratified by Iron dose

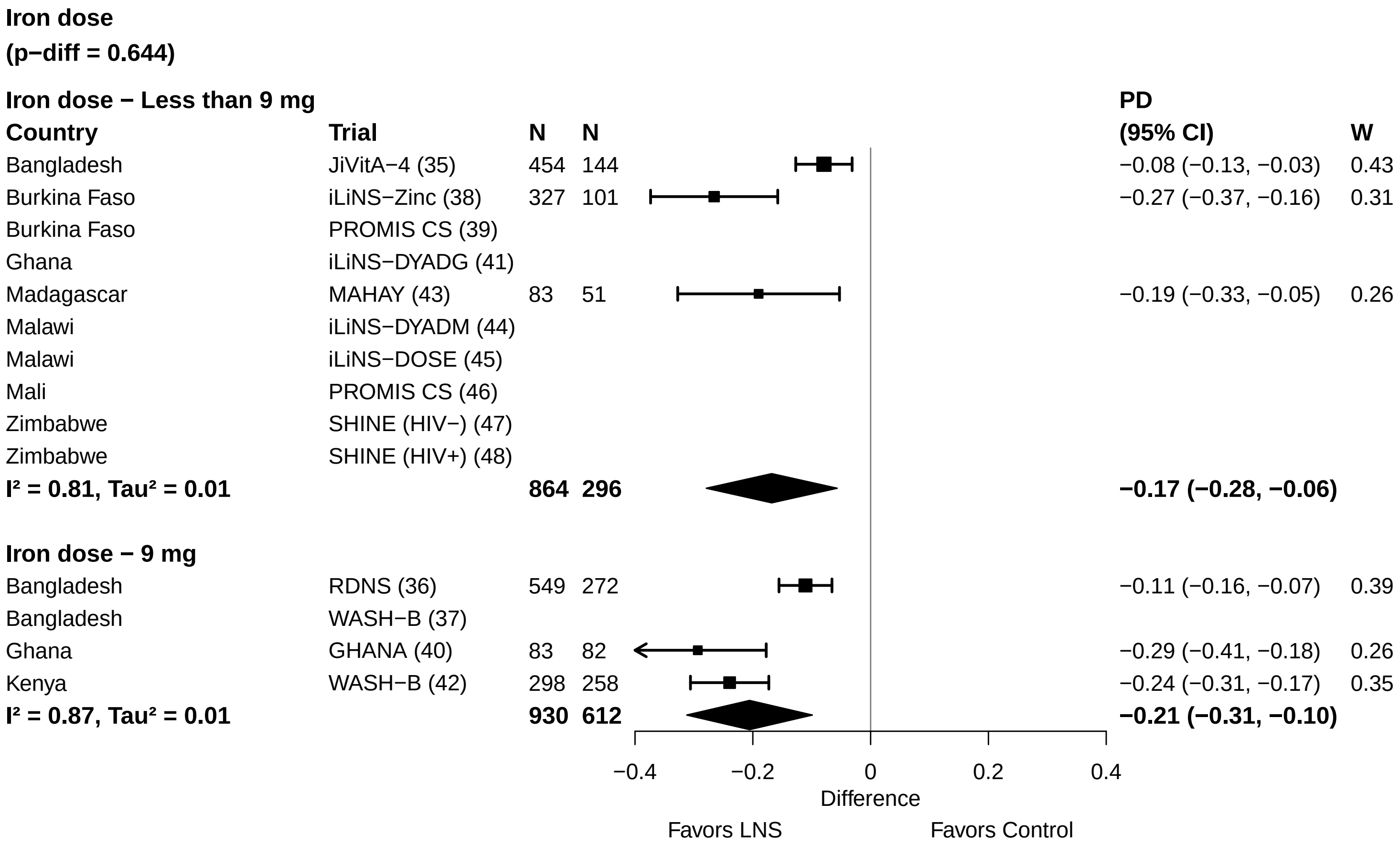

#### Supplemental figure 4J: Iron deficiency anemia prevalence difference

#### 4J9: Stratified by Frequency of contact

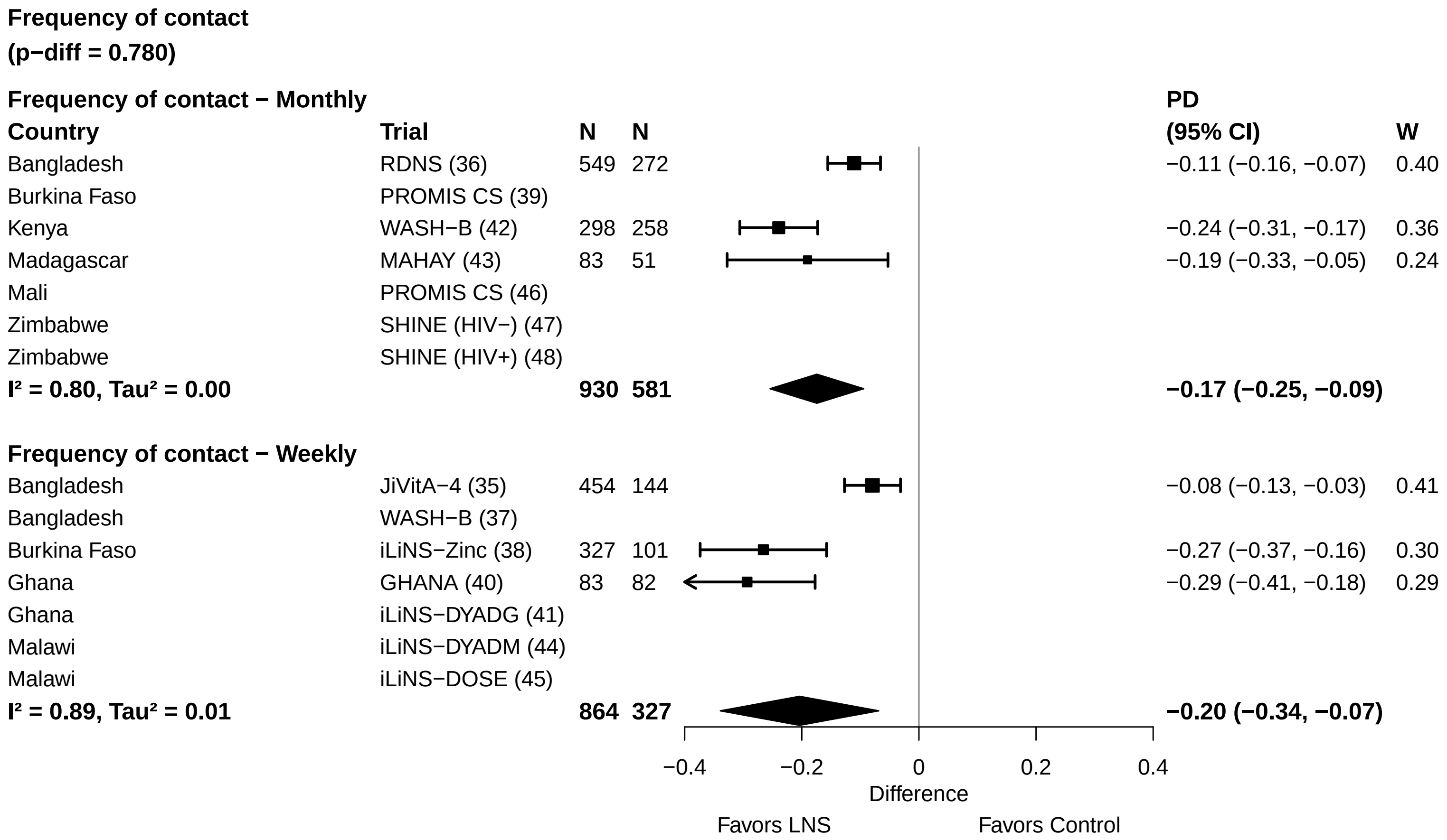

**Supplemental figure 4J: Iron deficiency anemia prevalence difference**

#### 4J10: Stratified by Average SQ-LNS compliance (insufficient comparisons)

### Supplemental figure 4K: Geometric mean ratio of soluble transferrin receptor concentration

#### 4K1: Stratified by Geographic region (insufficient comparisons)

**Supplemental figure 4K: Geometric mean ratio of soluble transferrin receptor concentration**

#### 4K2: Stratified by Anemia burden (insufficient comparisons)

**Supplemental figure 4K: Geometric mean ratio of soluble transferrin receptor concentration**

#### 4K3: Stratified by Malaria prevalence (insufficient comparisons)

**Supplemental figure 4K: Geometric mean ratio of soluble transferrin receptor concentration**

#### 4K4: Stratified by Inflammation burden (insufficient comparisons)

#### Supplemental figure 4K: Geometric mean ratio of soluble transferrin receptor concentration

### 4K5: Stratified by Source water quality

**Source water quality**
**(p−diff = 0.779)**

| Country | Trial | N | N | GMR (95% CI) | W |
|---|---|---|---|---|---|
| **Source water quality − Improved** | | | | | |
| Bangladesh | JiVitA−4 (35) | | | | |
| Bangladesh | RDNS (36) | 550 | 272 | 0.87 (0.83, 0.92) | 0.48 |
| Bangladesh | WASH−B (37) | 94 | 58 | 0.73 (0.63, 0.84) | 0.22 |
| Ghana | GHANA (40) | 84 | 82 | 0.80 (0.72, 0.89) | 0.30 |
| Ghana | iLiNS−DYADG (41) | | | | |
| Malawi | iLiNS−DYADM (44) | | | | |
| Malawi | iLiNS−DOSE (45) | | | | |
| **I² = 0.69, Tau² = 0.01** | | **728** | **412** | **0.81 (0.73, 0.89)** | |
| **Source water quality − Unimproved** | | | | | |
| Burkina Faso | iLiNS−Zinc (38) | 315 | 96 | 0.76 (0.68, 0.84) | 0.35 |
| Burkina Faso | PROMIS CS (39) | | | | |
| Kenya | WASH−B (42) | 166 | 144 | 0.78 (0.72, 0.85) | 0.43 |
| Madagascar | MAHAY (43) | 83 | 51 | 0.90 (0.77, 1.05) | 0.21 |
| Mali | PROMIS CS (46) | | | | |
| Zimbabwe | SHINE (HIV−) (47) | | | | |
| Zimbabwe | SHINE (HIV+) (48) | | | | |
| **I² = 0.37, Tau² = 0.00** | | **564** | **291** | **0.80 (0.73, 0.87)** | |

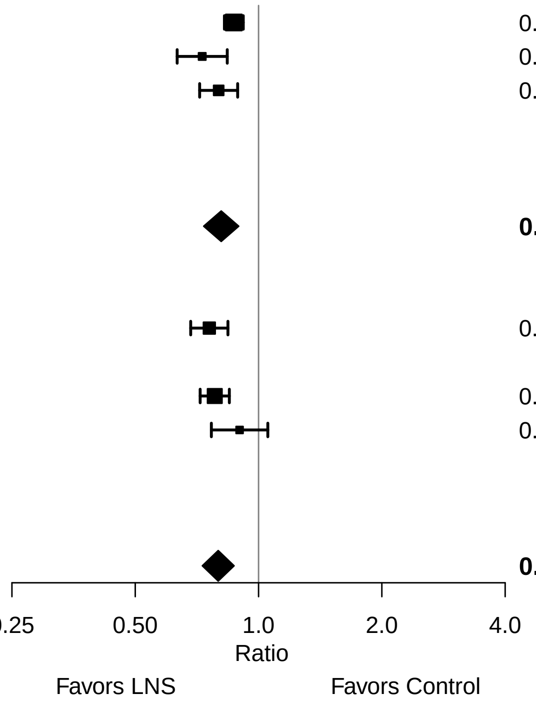

**Supplemental figure 4K: Geometric mean ratio of soluble transferrin receptor concentration**

#### 4K6: Stratified by Sanitation

**Sanitation**
**(p−diff = 0.779)**

| Country | Trial | N | N | GMR (95% CI) | W |
|---|---|---|---|---|---|
| **Sanitation − Improved** | | | | | |
| Bangladesh | JiVitA−4 (35) | | | | |
| Bangladesh | RDNS (36) | 550 | 272 | 0.87 (0.83, 0.92) | 0.48 |
| Bangladesh | WASH−B (37) | 94 | 58 | 0.73 (0.63, 0.84) | 0.22 |
| Burkina Faso | PROMIS CS (39) | | | | |
| Ghana | GHANA (40) | 84 | 82 | 0.80 (0.72, 0.89) | 0.30 |
| Ghana | iLiNS−DYADG (41) | | | | |
| Mali | PROMIS CS (46) | | | | |
| **I² = 0.69, Tau² = 0.01** | | **728** | **412** | **0.81 (0.73, 0.89)** | |
| **Sanitation − Unimproved** | | | | | |
| Burkina Faso | iLiNS−Zinc (38) | 315 | 96 | 0.76 (0.68, 0.84) | 0.35 |
| Kenya | WASH−B (42) | 166 | 144 | 0.78 (0.72, 0.85) | 0.43 |
| Madagascar | MAHAY (43) | 83 | 51 | 0.90 (0.77, 1.05) | 0.21 |
| Malawi | iLiNS−DYADM (44) | | | | |
| Malawi | iLiNS−DOSE (45) | | | | |
| Zimbabwe | SHINE (HIV−) (47) | | | | |
| Zimbabwe | SHINE (HIV+) (48) | | | | |
| **I² = 0.37, Tau² = 0.00** | | **564** | **291** | **0.80 (0.73, 0.87)** | |

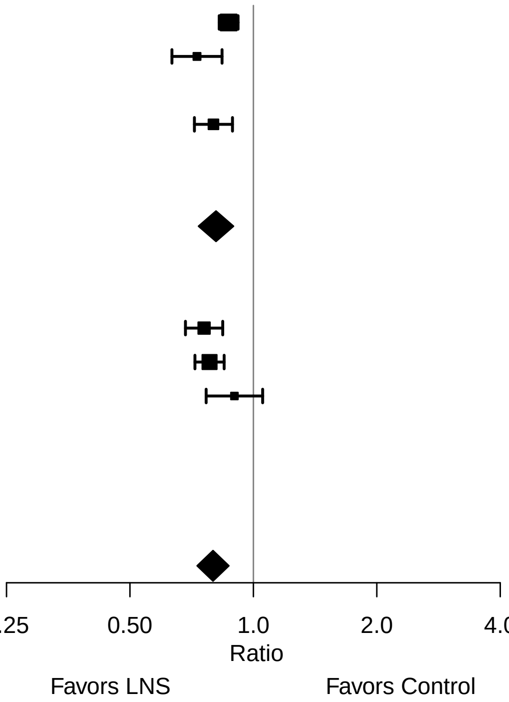

#### Supplemental figure 4K: Geometric mean ratio of soluble transferrin receptor concentration

#### 4K7: Stratified by Supplement duration

**Supplement duration**
**(p−diff = 0.539)**

| Country | Trial | N | N | GMR (95% CI) | W |
|---|---|---|---|---|---|
| **Supplement duration − 12m or less** | | | | | |
| Bangladesh | JiVitA−4 (35) | | | | |
| Burkina Faso | iLiNS−Zinc (38) | 315 | 96 | 0.76 (0.68, 0.84) | 0.40 |
| Burkina Faso | PROMIS CS (39) | | | | |
| Ghana | GHANA (40) | 84 | 82 | 0.80 (0.72, 0.89) | 0.39 |
| Ghana | iLiNS−DYADG (41) | | | | |
| Madagascar | MAHAY (43) | 83 | 51 | 0.90 (0.77, 1.05) | 0.21 |
| Malawi | iLiNS−DYADM (44) | | | | |
| Malawi | iLiNS−DOSE (45) | | | | |
| Zimbabwe | SHINE (HIV−) (47) | | | | |
| Zimbabwe | SHINE (HIV+) (48) | | | | |
| **I² = 0.35, Tau² = 0.00** | | **482** | **229** | **0.80 (0.74, 0.88)** | |
| **Supplement duration − > 12m** | | | | | |
| Bangladesh | RDNS (36) | 550 | 272 | 0.87 (0.83, 0.92) | 0.38 |
| Bangladesh | WASH−B (37) | 212 | 178 | 0.82 (0.76, 0.87) | 0.32 |
| Kenya | WASH−B (42) | 298 | 259 | 0.79 (0.74, 0.85) | 0.30 |
| Mali | PROMIS CS (46) | | | | |
| **I² = 0.61, Tau² = 0.00** | | **1060** | **709** | **0.83 (0.78, 0.88)** | |

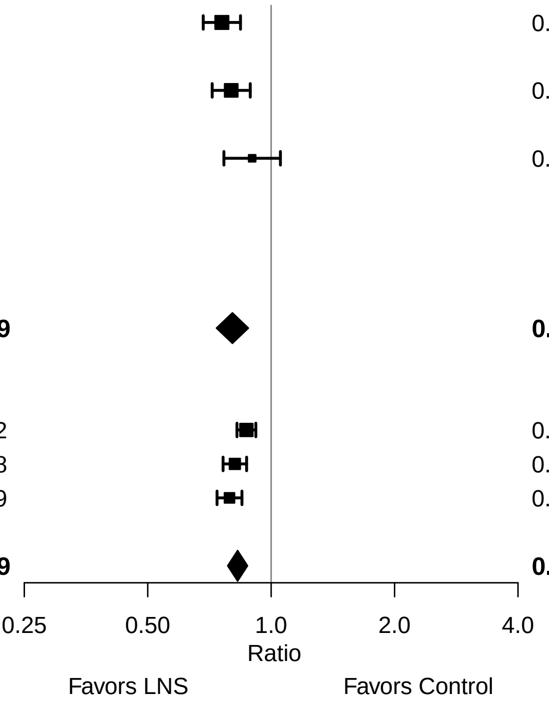

**Supplemental figure 4K: Geometric mean ratio of soluble transferrin receptor concentration**

#### 4K8: Stratified by Iron dose (insufficient comparisons)

#### Supplemental figure 4K: Geometric mean ratio of soluble transferrin receptor concentration

### 4K9: Stratified by Frequency of contact

**Frequency of contact**
**(p−diff = 0.208)**

| Country | Trial | N | N | GMR (95% CI) | W |
|---|---|---|---|---|---|
| **Frequency of contact − Monthly** | | | | | |
| Bangladesh | RDNS (36) | 550 | 272 | 0.87 (0.83, 0.92) | 0.49 |
| Burkina Faso | PROMIS CS (39) | | | | |
| Kenya | WASH−B (42) | 298 | 259 | 0.79 (0.74, 0.85) | 0.39 |
| Madagascar | MAHAY (43) | 83 | 51 | 0.90 (0.77, 1.05) | 0.13 |
| Mali | PROMIS CS (46) | | | | |
| Zimbabwe | SHINE (HIV−) (47) | | | | |
| Zimbabwe | SHINE (HIV+) (48) | | | | |
| **I² = 0.61, Tau² = 0.00** | | **931** | **582** | **0.84 (0.78, 0.91)** | |
| **Frequency of contact − Weekly** | | | | | |
| Bangladesh | JiVitA−4 (35) | | | | |
| Bangladesh | WASH−B (37) | 212 | 178 | 0.82 (0.76, 0.87) | 0.46 |
| Burkina Faso | iLiNS−Zinc (38) | 315 | 96 | 0.76 (0.68, 0.84) | 0.27 |
| Ghana | GHANA (40) | 84 | 82 | 0.80 (0.72, 0.89) | 0.26 |
| Ghana | iLiNS−DYADG (41) | | | | |
| Malawi | iLiNS−DYADM (44) | | | | |
| Malawi | iLiNS−DOSE (45) | | | | |
| **I² = 0.00, Tau² = 0.00** | | **611** | **356** | **0.80 (0.76, 0.84)** | |

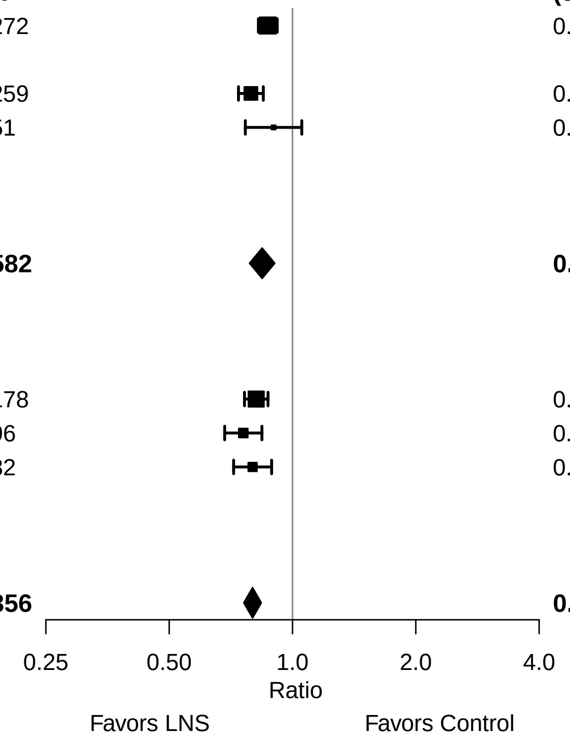

**Supplemental figure 4K: Geometric mean ratio of soluble transferrin receptor concentration**

#### 4K10: Stratified by Average SQ-LNS compliance (insufficient comparisons)

### Supplemental figure 4L: Elevated soluble transferrin receptor prevalence ratio

#### 4L1: Stratified by Geographic region (insufficient comparisons)

**Supplemental figure 4L: Elevated soluble transferrin receptor prevalence ratio**

#### 4L2: Stratified by Anemia burden (insufficient comparisons)

**Supplemental figure 4L: Elevated soluble transferrin receptor prevalence ratio**

#### 4L3: Stratified by Malaria prevalence (insufficient comparisons)

**Supplemental figure 4L: Elevated soluble transferrin receptor prevalence ratio**

#### 4L4: Stratified by Inflammation burden (insufficient comparisons)

##### Supplemental figure 4L: Elevated soluble transferrin receptor prevalence ratio

#### 4L5: Stratified by Source water quality

**Source water quality**
**(p−diff = 0.496)**

| Country | Trial | N | N | PR (95% CI) | W |
|---|---|---|---|---|---|
| **Source water quality − Improved** | | | | | |
| Bangladesh | JiVitA−4 (35) | | | | |
| Bangladesh | RDNS (36) | 550 | 272 | 0.70 (0.59, 0.84) | 0.44 |
| Bangladesh | WASH−B (37) | 94 | 58 | 0.23 (0.11, 0.48) | 0.20 |
| Ghana | GHANA (40) | 84 | 82 | 0.57 (0.41, 0.80) | 0.37 |
| Ghana | iLiNS−DYADG (41) | | | | |
| Malawi | iLiNS−DYADM (44) | | | | |
| Malawi | iLiNS−DOSE (45) | | | | |
| **I² = 0.77, Tau² = 0.25** | | **728** | **412** | **0.49 (0.26, 0.92)** | |
| **Source water quality − Unimproved** | | | | | |
| Burkina Faso | iLiNS−Zinc (38) | 315 | 96 | 0.51 (0.38, 0.68) | 0.32 |
| Burkina Faso | PROMIS CS (39) | | | | |
| Kenya | WASH−B (42) | 166 | 144 | 0.64 (0.54, 0.75) | 0.37 |
| Madagascar | MAHAY (43) | 83 | 51 | 0.82 (0.60, 1.12) | 0.31 |
| Mali | PROMIS CS (46) | | | | |
| Zimbabwe | SHINE (HIV−) (47) | | | | |
| Zimbabwe | SHINE (HIV+) (48) | | | | |
| **I² = 0.57, Tau² = 0.03** | | **564** | **291** | **0.64 (0.50, 0.82)** | |

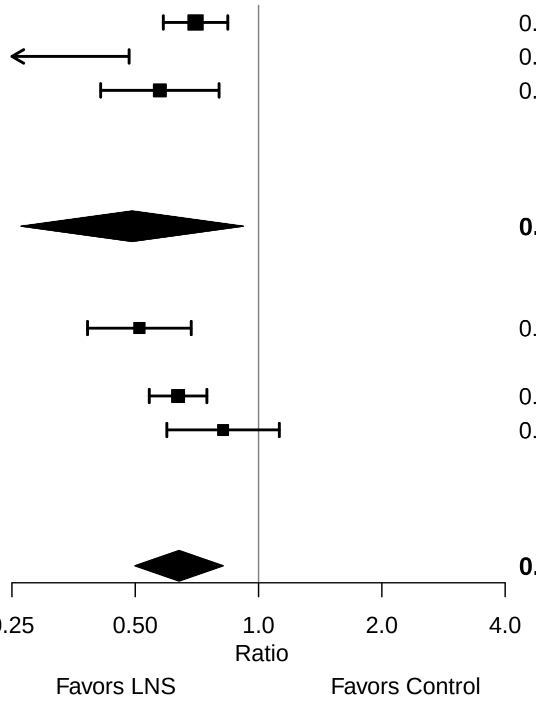

**Supplemental figure 4L: Elevated soluble transferrin receptor prevalence ratio**

#### 4L6: Stratified by Sanitation

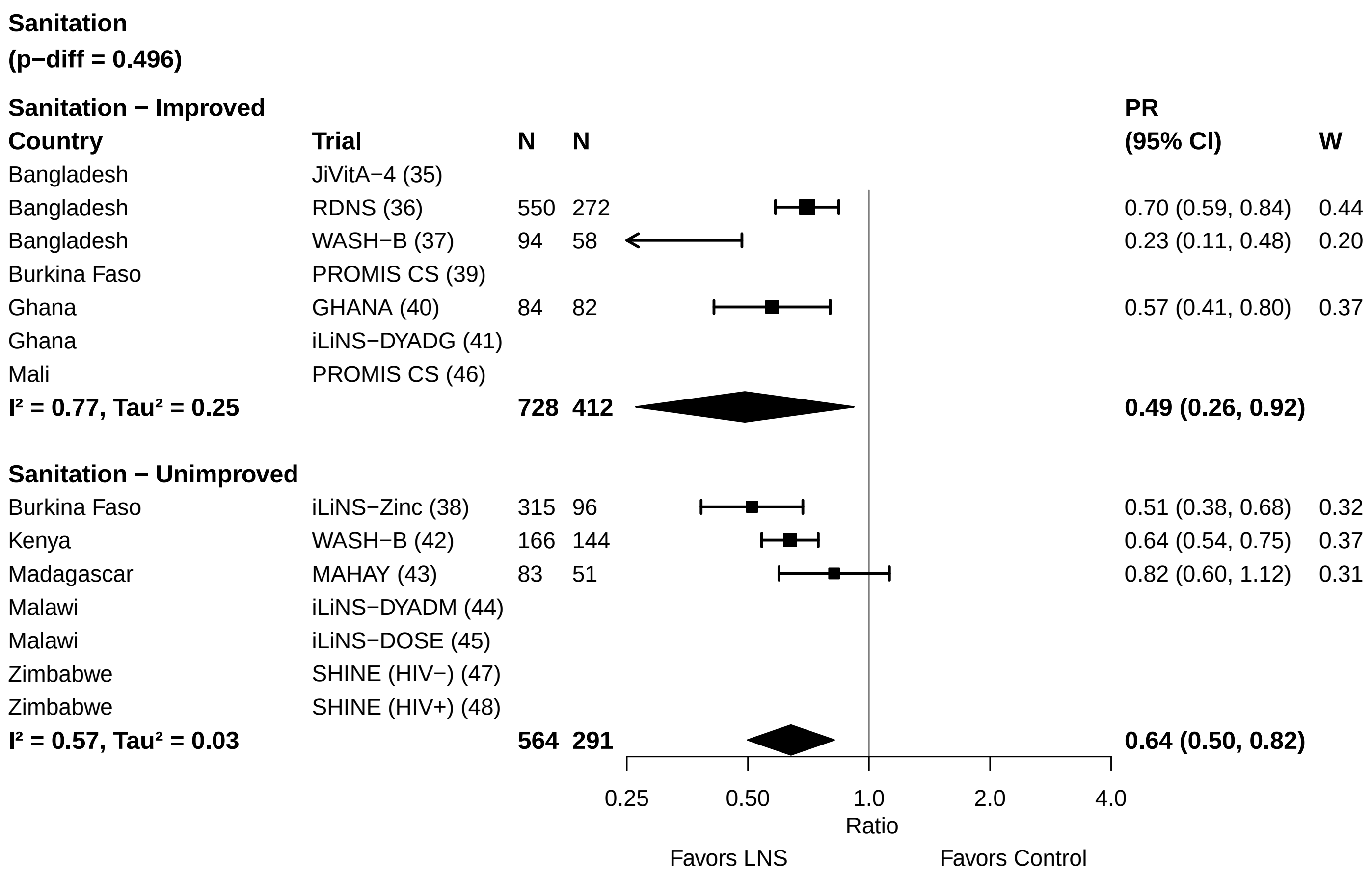

#### Supplemental figure 4L: Elevated soluble transferrin receptor prevalence ratio

### 4L7: Stratified by Supplement duration

**Supplement duration**
**(p−diff = 0.692)**

| Country | Trial | N | N | PR (95% CI) | W |
|---|---|---|---|---|---|
| **Supplement duration − 12m or less** | | | | | |
| Bangladesh | JiVitA−4 (35) | | | | |
| Burkina Faso | iLiNS−Zinc (38) | 315 | 96 | 0.51 (0.38, 0.68) | 0.34 |
| Burkina Faso | PROMIS CS (39) | | | | |
| Ghana | GHANA (40) | 84 | 82 | 0.57 (0.41, 0.80) | 0.32 |
| Ghana | iLiNS−DYADG (41) | | | | |
| Madagascar | MAHAY (43) | 83 | 51 | 0.82 (0.60, 1.12) | 0.33 |
| Malawi | iLiNS−DYADM (44) | | | | |
| Malawi | iLiNS−DOSE (45) | | | | |
| Zimbabwe | SHINE (HIV−) (47) | | | | |
| Zimbabwe | SHINE (HIV+) (48) | | | | |
| **$I^2$ = 0.59, $Tau^2$ = 0.04** | | **482** | **229** | **0.62 (0.47, 0.82)** | |
| **Supplement duration − > 12m** | | | | | |
| Bangladesh | RDNS (36) | 550 | 272 | 0.70 (0.59, 0.84) | 0.36 |
| Bangladesh | WASH−B (37) | 212 | 178 | 0.31 (0.21, 0.47) | 0.26 |
| Kenya | WASH−B (42) | 298 | 259 | 0.67 (0.59, 0.77) | 0.38 |
| Mali | PROMIS CS (46) | | | | |
| **$I^2$ = 0.85, $Tau^2$ = 0.17** | | **1060** | **709** | **0.55 (0.33, 0.89)** | |

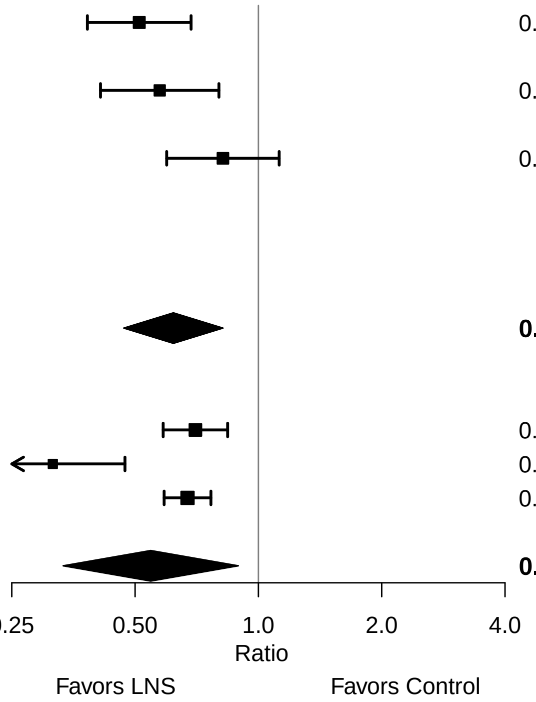

**Supplemental figure 4L: Elevated soluble transferrin receptor prevalence ratio**

#### 4L8: Stratified by Iron dose (insufficient comparisons)

**Supplemental figure 4L: Elevated soluble transferrin receptor prevalence ratio**

#### 4L9: Stratified by Frequency of contact

**Frequency of contact**
**(p−diff = 0.001)**

| Country | Trial | N | N | PR (95% CI) | W |
|---|---|---|---|---|---|
| **Frequency of contact − Monthly** | | | | | |
| Bangladesh | RDNS (36) | 550 | 272 | 0.70 (0.59, 0.84) | 0.35 |
| Burkina Faso | PROMIS CS (39) | | | | |
| Kenya | WASH−B (42) | 298 | 259 | 0.67 (0.59, 0.77) | 0.36 |
| Madagascar | MAHAY (43) | 83 | 51 | 0.82 (0.60, 1.12) | 0.29 |
| Mali | PROMIS CS (46) | | | | |
| Zimbabwe | SHINE (HIV−) (47) | | | | |
| Zimbabwe | SHINE (HIV+) (48) | | | | |
| **I² = 0.00, Tau² = 0.00** | | **931** | **582** | **0.69 (0.63, 0.77)** | |
| **Frequency of contact − Weekly** | | | | | |
| Bangladesh | JiVitA−4 (35) | | | | |
| Bangladesh | WASH−B (37) | 212 | 178 | 0.31 (0.21, 0.47) | 0.30 |
| Burkina Faso | iLiNS−Zinc (38) | 315 | 96 | 0.51 (0.38, 0.68) | 0.36 |
| Ghana | GHANA (40) | 84 | 82 | 0.57 (0.41, 0.80) | 0.34 |
| Ghana | iLiNS−DYADG (41) | | | | |
| Malawi | iLiNS−DYADM (44) | | | | |
| Malawi | iLiNS−DOSE (45) | | | | |
| **I² = 0.63, Tau² = 0.06** | | **611** | **356** | **0.46 (0.33, 0.65)** | |

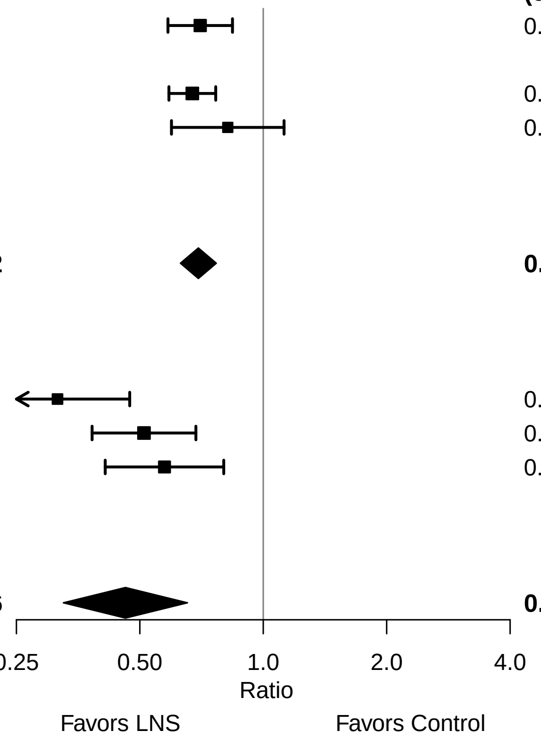

##### Supplemental figure 4L: Elevated soluble transferrin receptor prevalence ratio

#### 4L10: Stratified by Average SQ-LNS compliance (insufficient comparisons)

#### Supplemental figure 4M: Elevated soluble transferrin receptor prevalence difference

##### 4M1: Stratified by Geographic region (insufficient comparisons)

**Supplemental figure 4M: Elevated soluble transferrin receptor prevalence difference**

#### 4M2: Stratified by Anemia burden (insufficient comparisons)

**Supplemental figure 4M: Elevated soluble transferrin receptor prevalence difference**

#### 4M3: Stratified by Malaria prevalence (insufficient comparisons)

**Supplemental figure 4M: Elevated soluble transferrin receptor prevalence difference**

#### 4M4: Stratified by Inflammation burden (insufficient comparisons)

**Supplemental figure 4M: Elevated soluble transferrin receptor prevalence difference**

#### 4M5: Stratified by Source water quality

**Source water quality**
**(p−diff = 0.841)**

| Country | Trial | N | N | PD (95% CI) | W |
|---|---|---|---|---|---|
| **Source water quality − Improved** | | | | | |
| Bangladesh | JiVitA−4 (35) | | | | |
| Bangladesh | RDNS (36) | 550 | 272 | −0.15 (−0.22, −0.08) | 0.49 |
| Bangladesh | WASH−B (37) | 94 | 58 | −0.32 (−0.46, −0.18) | 0.26 |
| Ghana | GHANA (40) | 84 | 82 | −0.26 (−0.41, −0.12) | 0.25 |
| Ghana | iLiNS−DYADG (41) | | | | |
| Malawi | iLiNS−DYADM (44) | | | | |
| Malawi | iLiNS−DOSE (45) | | | | |
| **I² = 0.63, Tau² = 0.00** | | **728** | **412** | **−0.23 (−0.33, −0.12)** | |
| **Source water quality − Unimproved** | | | | | |
| Burkina Faso | iLiNS−Zinc (38) | 315 | 96 | −0.30 (−0.40, −0.19) | 0.39 |
| Burkina Faso | PROMIS CS (39) | | | | |
| Kenya | WASH−B (42) | 166 | 144 | −0.27 (−0.37, −0.16) | 0.39 |
| Madagascar | MAHAY (43) | 83 | 51 | −0.11 (−0.28, 0.07) | 0.21 |
| Mali | PROMIS CS (46) | | | | |
| Zimbabwe | SHINE (HIV−) (47) | | | | |
| Zimbabwe | SHINE (HIV+) (48) | | | | |
| **I² = 0.43, Tau² = 0.00** | | **564** | **291** | **−0.24 (−0.35, −0.14)** | |

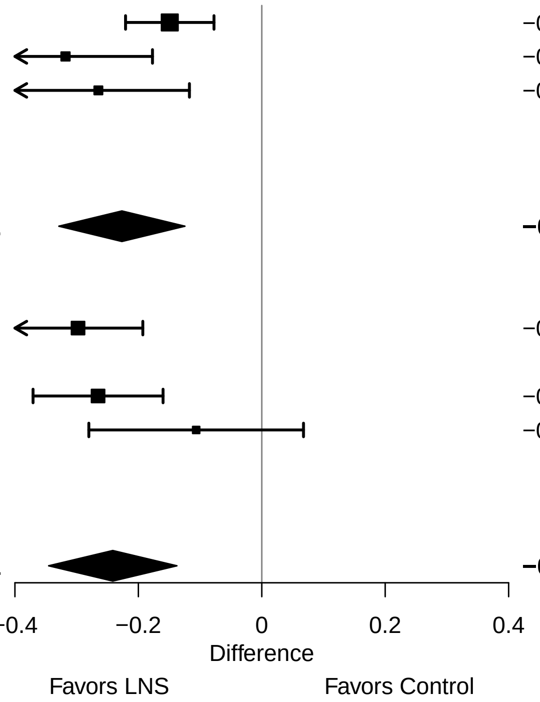

#### Supplemental figure 4M: Elevated soluble transferrin receptor prevalence difference

### 4M6: Stratified by Sanitation

**Sanitation**
**(p−diff = 0.841)**

| Country | Trial | N | N | PD (95% CI) | W |
|---|---|---|---|---|---|
| **Sanitation − Improved** | | | | | |
| Bangladesh | JiVitA−4 (35) | | | | |
| Bangladesh | RDNS (36) | 550 | 272 | −0.15 (−0.22, −0.08) | 0.49 |
| Bangladesh | WASH−B (37) | 94 | 58 | −0.32 (−0.46, −0.18) | 0.26 |
| Burkina Faso | PROMIS CS (39) | | | | |
| Ghana | GHANA (40) | 84 | 82 | −0.26 (−0.41, −0.12) | 0.25 |
| Ghana | iLiNS−DYADG (41) | | | | |
| Mali | PROMIS CS (46) | | | | |
| **I² = 0.63, Tau² = 0.00** | | **728** | **412** | **−0.23 (−0.33, −0.12)** | |
| **Sanitation − Unimproved** | | | | | |
| Burkina Faso | iLiNS−Zinc (38) | 315 | 96 | −0.30 (−0.40, −0.19) | 0.39 |
| Kenya | WASH−B (42) | 166 | 144 | −0.27 (−0.37, −0.16) | 0.39 |
| Madagascar | MAHAY (43) | 83 | 51 | −0.11 (−0.28, 0.07) | 0.21 |
| Malawi | iLiNS−DYADM (44) | | | | |
| Malawi | iLiNS−DOSE (45) | | | | |
| Zimbabwe | SHINE (HIV−) (47) | | | | |
| Zimbabwe | SHINE (HIV+) (48) | | | | |
| **I² = 0.43, Tau² = 0.00** | | **564** | **291** | **−0.24 (−0.35, −0.14)** | |

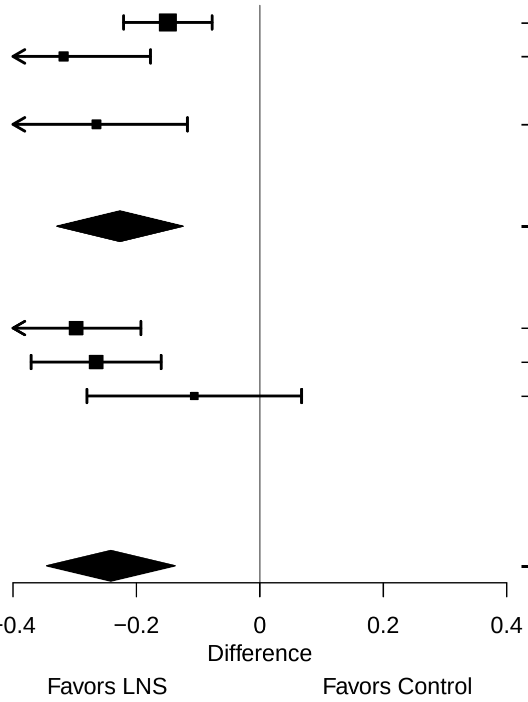

**Supplemental figure 4M: Elevated soluble transferrin receptor prevalence difference**

#### 4M7: Stratified by Supplement duration

**Supplement duration**
**(p−diff = 0.385)**

| Country | Trial | N | N | PD (95% CI) | W |
|---|---|---|---|---|---|
| **Supplement duration − 12m or less** | | | | | |
| Bangladesh | JiVitA−4 (35) | | | | |
| Burkina Faso | iLiNS−Zinc (38) | 315 | 96 | −0.30 (−0.40, −0.19) | 0.48 |
| Burkina Faso | PROMIS CS (39) | | | | |
| Ghana | GHANA (40) | 84 | 82 | −0.26 (−0.41, −0.12) | 0.29 |
| Ghana | iLiNS−DYADG (41) | | | | |
| Madagascar | MAHAY (43) | 83 | 51 | −0.11 (−0.28, 0.07) | 0.22 |
| Malawi | iLiNS−DYADM (44) | | | | |
| Malawi | iLiNS−DOSE (45) | | | | |
| Zimbabwe | SHINE (HIV−) (47) | | | | |
| Zimbabwe | SHINE (HIV+) (48) | | | | |
| **I² = 0.42, Tau² = 0.00** | | **482** | **229** | **−0.24 (−0.35, −0.13)** | |
| **Supplement duration − > 12m** | | | | | |
| Bangladesh | RDNS (36) | 550 | 272 | −0.15 (−0.22, −0.08) | 0.36 |
| Bangladesh | WASH−B (37) | 212 | 178 | −0.22 (−0.29, −0.14) | 0.32 |
| Kenya | WASH−B (42) | 298 | 259 | −0.24 (−0.32, −0.16) | 0.32 |
| Mali | PROMIS CS (46) | | | | |
| **I² = 0.35, Tau² = 0.00** | | **1060** | **709** | **−0.20 (−0.25, −0.15)** | |

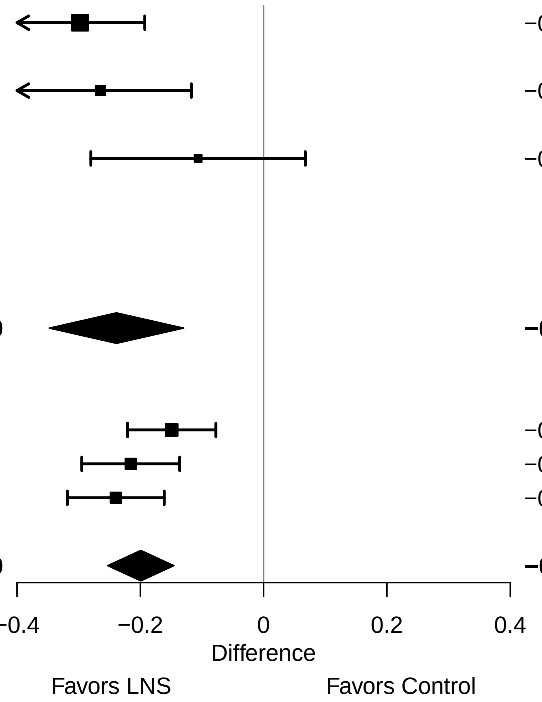

**Supplemental figure 4M: Elevated soluble transferrin receptor prevalence difference**

#### 4M8: Stratified by Iron dose (insufficient comparisons)

**Supplemental figure 4M: Elevated soluble transferrin receptor prevalence difference**

#### 4M9: Stratified by Frequency of contact

**Frequency of contact**
**(p−diff = 0.152)**

| Country | Trial | N | N | PD (95% CI) | W |
|---|---|---|---|---|---|
| **Frequency of contact − Monthly** | | | | | |
| Bangladesh | RDNS (36) | 550 | 272 | −0.15 (−0.22, −0.08) | 0.45 |
| Burkina Faso | PROMIS CS (39) | | | | |
| Kenya | WASH−B (42) | 298 | 259 | −0.24 (−0.32, −0.16) | 0.41 |
| Madagascar | MAHAY (43) | 83 | 51 | −0.11 (−0.28, 0.07) | 0.13 |
| Mali | PROMIS CS (46) | | | | |
| Zimbabwe | SHINE (HIV−) (47) | | | | |
| Zimbabwe | SHINE (HIV+) (48) | | | | |
| **I² = 0.45, Tau² = 0.00** | | **931** | **582** | **−0.18 (−0.25, −0.11)** | |
| **Frequency of contact − Weekly** | | | | | |
| Bangladesh | JiVitA−4 (35) | | | | |
| Bangladesh | WASH−B (37) | 212 | 178 | −0.22 (−0.29, −0.14) | 0.47 |
| Burkina Faso | iLiNS−Zinc (38) | 315 | 96 | −0.30 (−0.40, −0.19) | 0.33 |
| Ghana | GHANA (40) | 84 | 82 | −0.26 (−0.41, −0.12) | 0.20 |
| Ghana | iLiNS−DYADG (41) | | | | |
| Malawi | iLiNS−DYADM (44) | | | | |
| Malawi | iLiNS−DOSE (45) | | | | |
| **I² = 0.00, Tau² = 0.00** | | **611** | **356** | **−0.25 (−0.31, −0.19)** | |

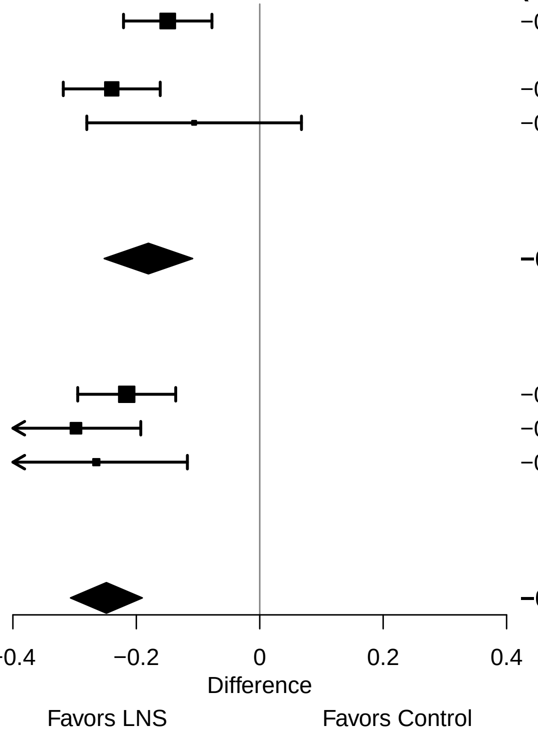

**Supplemental figure 4M: Elevated soluble transferrin receptor prevalence difference**

#### 4M10: Stratified by Average SQ-LNS compliance (insufficient comparisons)

### Supplemental figure 4N: Geometric mean ratio of zinc protoporphyrin concentration

#### 4N1: Stratified by Geographic region (insufficient comparisons)

**Supplemental figure 4N: Geometric mean ratio of zinc protoporphyrin concentration**

#### 4N2: Stratified by Anemia burden (insufficient comparisons)

**Supplemental figure 4N: Geometric mean ratio of zinc protoporphyrin concentration**

#### 4N3: Stratified by Malaria prevalence (insufficient comparisons)

**Supplemental figure 4N: Geometric mean ratio of zinc protoporphyrin concentration**

#### 4N4: Stratified by Inflammation burden (insufficient comparisons)

**Supplemental figure 4N: Geometric mean ratio of zinc protoporphyrin concentration**

#### 4N5: Stratified by Source water quality (insufficient comparisons)

**Supplemental figure 4N: Geometric mean ratio of zinc protoporphyrin concentration**

#### 4N6: Stratified by Sanitation (insufficient comparisons)

**Supplemental figure 4N: Geometric mean ratio of zinc protoporphyrin concentration**

#### 4N7: Stratified by Supplement duration (insufficient comparisons)

**Supplemental figure 4N: Geometric mean ratio of zinc protoporphyrin concentration**

#### 4N8: Stratified by Iron dose (insufficient comparisons)

**Supplemental figure 4N: Geometric mean ratio of zinc protoporphyrin concentration**

#### 4N9: Stratified by Frequency of contact (insufficient comparisons)

**Supplemental figure 4N: Geometric mean ratio of zinc protoporphyrin concentration**

#### 4N10: Stratified by Average SQ-LNS compliance (insufficient comparisons)

### Supplemental figure 4O: Elevated zinc protoporphyrin prevalence ratio

#### 4O1: Stratified by Geographic region (insufficient comparisons)

**Supplemental figure 4O: Elevated zinc protoporphyrin prevalence ratio**

#### 4O2: Stratified by Anemia burden (insufficient comparisons)

##### Supplemental figure 4O: Elevated zinc protoporphyrin prevalence ratio

#### 4O3: Stratified by Malaria prevalence (insufficient comparisons)

**Supplemental figure 4O: Elevated zinc protoporphyrin prevalence ratio**

#### 4O4: Stratified by Inflammation burden (insufficient comparisons)

#### Supplemental figure 4O: Elevated zinc protoporphyrin prevalence ratio

### 4O5: Stratified by Source water quality (insufficient comparisons)

**Supplemental figure 4O: Elevated zinc protoporphyrin prevalence ratio**

#### 4O6: Stratified by Sanitation (insufficient comparisons)

**Supplemental figure 4O: Elevated zinc protoporphyrin prevalence ratio**

#### 4O7: Stratified by Supplement duration (insufficient comparisons)

**Supplemental figure 4O: Elevated zinc protoporphyrin prevalence ratio**

#### 4O8: Stratified by Iron dose (insufficient comparisons)

**Supplemental figure 4O: Elevated zinc protoporphyrin prevalence ratio**

#### 4O9: Stratified by Frequency of contact (insufficient comparisons)

**Supplemental figure 4O: Elevated zinc protoporphyrin prevalence ratio**

#### 4O10: Stratified by Average SQ-LNS compliance (insufficient comparisons)

### Supplemental figure 4P: Elevated zinc protoporphyrin prevalence difference

#### 4P1: Stratified by Geographic region (insufficient comparisons)

**Supplemental figure 4P: Elevated zinc protoporphyrin prevalence difference**

#### 4P2: Stratified by Anemia burden (insufficient comparisons)

**Supplemental figure 4P: Elevated zinc protoporphyrin prevalence difference**

#### 4P3: Stratified by Malaria prevalence (insufficient comparisons)

**Supplemental figure 4P: Elevated zinc protoporphyrin prevalence difference**

#### 4P4: Stratified by Inflammation burden (insufficient comparisons)

**Supplemental figure 4P: Elevated zinc protoporphyrin prevalence difference**

#### 4P5: Stratified by Source water quality (insufficient comparisons)

**Supplemental figure 4P: Elevated zinc protoporphyrin prevalence difference**

#### 4P6: Stratified by Sanitation (insufficient comparisons)

**Supplemental figure 4P: Elevated zinc protoporphyrin prevalence difference**

#### 4P7: Stratified by Supplement duration (insufficient comparisons)

**Supplemental figure 4P: Elevated zinc protoporphyrin prevalence difference**

#### 4P8: Stratified by Iron dose (insufficient comparisons)

**Supplemental figure 4P: Elevated zinc protoporphyrin prevalence difference**

#### 4P9: Stratified by Frequency of contact (insufficient comparisons)

**Supplemental figure 4P: Elevated zinc protoporphyrin prevalence difference**

#### 4P10: Stratified by Average SQ-LNS compliance (insufficient comparisons)

### Supplemental figure 4Q: Geometric mean ratio of plasma zinc concentration

#### 4Q1: Stratified by Geographic region (insufficient comparisons)

**Supplemental figure 4Q: Geometric mean ratio of plasma zinc concentration**

#### 4Q2: Stratified by Anemia burden (insufficient comparisons)

**Supplemental figure 4Q: Geometric mean ratio of plasma zinc concentration**

#### 4Q3: Stratified by Malaria prevalence (insufficient comparisons)

**Supplemental figure 4Q: Geometric mean ratio of plasma zinc concentration**

#### 4Q4: Stratified by Inflammation burden (insufficient comparisons)

**Supplemental figure 4Q: Geometric mean ratio of plasma zinc concentration**

#### 4Q5: Stratified by Source water quality (insufficient comparisons)

**Supplemental figure 4Q: Geometric mean ratio of plasma zinc concentration**

#### 4Q6: Stratified by Sanitation (insufficient comparisons)

**Supplemental figure 4Q: Geometric mean ratio of plasma zinc concentration**

#### 4Q7: Stratified by Supplement duration (insufficient comparisons)

**Supplemental figure 4Q: Geometric mean ratio of plasma zinc concentration**

#### 4Q8: Stratified by Iron dose (insufficient comparisons)

**Supplemental figure 4Q: Geometric mean ratio of plasma zinc concentration**

#### 4Q9: Stratified by Frequency of contact (insufficient comparisons)

**Supplemental figure 4Q: Geometric mean ratio of plasma zinc concentration**

#### 4Q10: Stratified by Average SQ-LNS compliance (insufficient comparisons)

### Supplemental figure 4R: Geometric mean ratio of retinol concentration

#### 4R1: Stratified by Geographic region (insufficient comparisons)

**Supplemental figure 4R: Geometric mean ratio of retinol concentration**

#### 4R2: Stratified by Anemia burden (insufficient comparisons)

**Supplemental figure 4R: Geometric mean ratio of retinol concentration**

#### 4R3: Stratified by Malaria prevalence (insufficient comparisons)

**Supplemental figure 4R: Geometric mean ratio of retinol concentration**

#### 4R4: Stratified by Inflammation burden (insufficient comparisons)

**Supplemental figure 4R: Geometric mean ratio of retinol concentration**

#### 4R5: Stratified by Source water quality (insufficient comparisons)

**Supplemental figure 4R: Geometric mean ratio of retinol concentration**

#### 4R6: Stratified by Sanitation (insufficient comparisons)

**Supplemental figure 4R: Geometric mean ratio of retinol concentration**

#### 4R7: Stratified by Supplement duration (insufficient comparisons)

**Supplemental figure 4R: Geometric mean ratio of retinol concentration**

#### 4R8: Stratified by Iron dose (insufficient comparisons)

**Supplemental figure 4R: Geometric mean ratio of retinol concentration**

#### 4R9: Stratified by Frequency of contact (insufficient comparisons)

**Supplemental figure 4R: Geometric mean ratio of retinol concentration**

#### 4R10: Stratified by Average SQ-LNS compliance (insufficient comparisons)

### Supplemental figure 4S: Low vitamin A (retinol < 0.70 µmol/L) prevalence ratio

#### 4S1: Stratified by Geographic region (insufficient comparisons)

**Supplemental figure 4S: Low vitamin A (retinol < 0.70 µmol/L) prevalence ratio**

#### 4S2: Stratified by Anemia burden (insufficient comparisons)

**Supplemental figure 4S: Low vitamin A (retinol < 0.70 µmol/L) prevalence ratio**

#### 4S3: Stratified by Malaria prevalence (insufficient comparisons)

**Supplemental figure 4S: Low vitamin A (retinol < 0.70 µmol/L) prevalence ratio**

#### 4S4: Stratified by Inflammation burden (insufficient comparisons)

**Supplemental figure 4S: Low vitamin A (retinol < 0.70 µmol/L) prevalence ratio**

#### 4S5: Stratified by Source water quality (insufficient comparisons)

**Supplemental figure 4S: Low vitamin A (retinol < 0.70 µmol/L) prevalence ratio**

#### 4S6: Stratified by Sanitation (insufficient comparisons)

**Supplemental figure 4S: Low vitamin A (retinol < 0.70 µmol/L) prevalence ratio**

#### 4S7: Stratified by Supplement duration (insufficient comparisons)

**Supplemental figure 4S: Low vitamin A (retinol < 0.70 µmol/L) prevalence ratio**

#### 4S8: Stratified by Iron dose (insufficient comparisons)

**Supplemental figure 4S: Low vitamin A (retinol < 0.70 µmol/L) prevalence ratio**

#### 4S9: Stratified by Frequency of contact (insufficient comparisons)

**Supplemental figure 4S: Low vitamin A (retinol < 0.70 µmol/L) prevalence ratio**

#### 4S10: Stratified by Average SQ-LNS compliance (insufficient comparisons)

### Supplemental figure 4T: Low vitamin A (retinol < 0.70 µmol/L) prevalence difference

#### 4T1: Stratified by Geographic region (insufficient comparisons)

**Supplemental figure 4T: Low vitamin A (retinol < 0.70 µmol/L) prevalence difference**

#### 4T2: Stratified by Anemia burden (insufficient comparisons)

**Supplemental figure 4T: Low vitamin A (retinol < 0.70 µmol/L) prevalence difference**

#### 4T3: Stratified by Malaria prevalence (insufficient comparisons)

**Supplemental figure 4T: Low vitamin A (retinol < 0.70 µmol/L) prevalence difference**

#### 4T4: Stratified by Inflammation burden (insufficient comparisons)

**Supplemental figure 4T: Low vitamin A (retinol < 0.70 µmol/L) prevalence difference**

#### 4T5: Stratified by Source water quality (insufficient comparisons)

**Supplemental figure 4T: Low vitamin A (retinol < 0.70 µmol/L) prevalence difference**

#### 4T6: Stratified by Sanitation (insufficient comparisons)

**Supplemental figure 4T: Low vitamin A (retinol < 0.70 µmol/L) prevalence difference**

#### 4T7: Stratified by Supplement duration (insufficient comparisons)

**Supplemental figure 4T: Low vitamin A (retinol < 0.70 µmol/L) prevalence difference**

#### 4T8: Stratified by Iron dose (insufficient comparisons)

#### Supplemental figure 4T: Low vitamin A (retinol < 0.70 µmol/L) prevalence difference

### 4T9: Stratified by Frequency of contact (insufficient comparisons)

#### Supplemental figure 4T: Low vitamin A (retinol < 0.70 µmol/L) prevalence difference

### 4T10: Stratified by Average SQ-LNS compliance (insufficient comparisons)

### Supplemental figure 4U: Marginal vitamin A (retinol < 1.05 µmol/L) prevalence ratio

#### 4U1: Stratified by Geographic region (insufficient comparisons)

**Supplemental figure 4U: Marginal vitamin A (retinol < 1.05 µmol/L) prevalence ratio**

#### 4U2: Stratified by Anemia burden (insufficient comparisons)

**Supplemental figure 4U: Marginal vitamin A (retinol < 1.05 µmol/L) prevalence ratio**

#### 4U3: Stratified by Malaria prevalence (insufficient comparisons)

**Supplemental figure 4U: Marginal vitamin A (retinol < 1.05 µmol/L) prevalence ratio**

#### 4U4: Stratified by Inflammation burden (insufficient comparisons)

**Supplemental figure 4U: Marginal vitamin A (retinol < 1.05 µmol/L) prevalence ratio**

#### 4U5: Stratified by Source water quality (insufficient comparisons)

**Supplemental figure 4U: Marginal vitamin A (retinol < 1.05 µmol/L) prevalence ratio**

#### 4U6: Stratified by Sanitation (insufficient comparisons)

**Supplemental figure 4U: Marginal vitamin A (retinol < 1.05 µmol/L) prevalence ratio**

#### 4U7: Stratified by Supplement duration (insufficient comparisons)

**Supplemental figure 4U: Marginal vitamin A (retinol < 1.05 µmol/L) prevalence ratio**

#### 4U8: Stratified by Iron dose (insufficient comparisons)

**Supplemental figure 4U: Marginal vitamin A (retinol < 1.05 µmol/L) prevalence ratio**

#### 4U9: Stratified by Frequency of contact (insufficient comparisons)

**Supplemental figure 4U: Marginal vitamin A (retinol < 1.05 µmol/L) prevalence ratio**

#### 4U10: Stratified by Average SQ-LNS compliance (insufficient comparisons)

### Supplemental figure 4V: Marginal vitamin A (retinol < 1.05 µmol/L) prevalence difference

#### 4V1: Stratified by Geographic region (insufficient comparisons)

**Supplemental figure 4V: Marginal vitamin A (retinol < 1.05 μmol/L) prevalence difference**

#### 4V2: Stratified by Anemia burden (insufficient comparisons)

**Supplemental figure 4V: Marginal vitamin A (retinol < 1.05 µmol/L) prevalence difference**

#### 4V3: Stratified by Malaria prevalence (insufficient comparisons)

##### Supplemental figure 4V: Marginal vitamin A (retinol < 1.05 µmol/L) prevalence difference

#### 4V4: Stratified by Inflammation burden (insufficient comparisons)

**Supplemental figure 4V: Marginal vitamin A (retinol < 1.05 µmol/L) prevalence difference**

#### 4V5: Stratified by Source water quality (insufficient comparisons)

**Supplemental figure 4V: Marginal vitamin A (retinol < 1.05 µmol/L) prevalence difference**

#### 4V6: Stratified by Sanitation (insufficient comparisons)

**Supplemental figure 4V: Marginal vitamin A (retinol < 1.05 µmol/L) prevalence difference**

#### 4V7: Stratified by Supplement duration (insufficient comparisons)

**Supplemental figure 4V: Marginal vitamin A (retinol $<$ 1.05 µmol/L) prevalence difference**

#### 4V8: Stratified by Iron dose (insufficient comparisons)

**Supplemental figure 4V: Marginal vitamin A (retinol < 1.05 µmol/L) prevalence difference**

#### 4V9: Stratified by Frequency of contact (insufficient comparisons)

**Supplemental figure 4V: Marginal vitamin A (retinol $< 1.05$ µmol/L) prevalence difference**

#### 4V10: Stratified by Average SQ-LNS compliance (insufficient comparisons)

### Supplemental figure 4W: Geometric mean ratio of retinol binding protein concentration

#### 4W1: Stratified by Geographic region (insufficient comparisons)

**Supplemental figure 4W: Geometric mean ratio of retinol binding protein concentration**

#### 4W2: Stratified by Anemia burden (insufficient comparisons)

**Supplemental figure 4W: Geometric mean ratio of retinol binding protein concentration**

#### 4W3: Stratified by Malaria prevalence (insufficient comparisons)

**Supplemental figure 4W: Geometric mean ratio of retinol binding protein concentration**

#### 4W4: Stratified by Inflammation burden (insufficient comparisons)

**Supplemental figure 4W: Geometric mean ratio of retinol binding protein concentration**

#### 4W5: Stratified by Source water quality (insufficient comparisons)

**Supplemental figure 4W: Geometric mean ratio of retinol binding protein concentration**

#### 4W6: Stratified by Sanitation (insufficient comparisons)

**Supplemental figure 4W: Geometric mean ratio of retinol binding protein concentration**

#### 4W7: Stratified by Supplement duration (insufficient comparisons)

**Supplemental figure 4W: Geometric mean ratio of retinol binding protein concentration**

#### 4W8: Stratified by Iron dose (insufficient comparisons)

**Supplemental figure 4W: Geometric mean ratio of retinol binding protein concentration**

#### 4W9: Stratified by Frequency of contact (insufficient comparisons)

**Supplemental figure 4W: Geometric mean ratio of retinol binding protein concentration**

#### 4W10: Stratified by Average SQ-LNS compliance (insufficient comparisons)

### Supplemental figure 4X: Low vitamin A status (RBP < 0.70 µmol/L) prevalence ratio

#### 4X1: Stratified by Geographic region (insufficient comparisons)

**Supplemental figure 4X: Low vitamin A status (RBP < 0.70 µmol/L) prevalence ratio**

#### 4X2: Stratified by Anemia burden (insufficient comparisons)

**Supplemental figure 4X: Low vitamin A status (RBP < 0.70 µmol/L) prevalence ratio**

#### 4X3: Stratified by Malaria prevalence (insufficient comparisons)

**Supplemental figure 4X: Low vitamin A status (RBP < 0.70 µmol/L) prevalence ratio**

#### 4X4: Stratified by Inflammation burden (insufficient comparisons)

#### Supplemental figure 4X: Low vitamin A status (RBP $< 0.70$ µmol/L) prevalence ratio

### 4X5: Stratified by Source water quality (insufficient comparisons)

**Supplemental figure 4X: Low vitamin A status (RBP < 0.70 µmol/L) prevalence ratio**

#### 4X6: Stratified by Sanitation (insufficient comparisons)

**Supplemental figure 4X: Low vitamin A status (RBP < 0.70 µmol/L) prevalence ratio**

#### 4X7: Stratified by Supplement duration (insufficient comparisons)

**Supplemental figure 4X: Low vitamin A status (RBP < 0.70 µmol/L) prevalence ratio**

#### 4X8: Stratified by Iron dose (insufficient comparisons)

**Supplemental figure 4X: Low vitamin A status (RBP < 0.70 µmol/L) prevalence ratio**

#### 4X9: Stratified by Frequency of contact (insufficient comparisons)

##### Supplemental figure 4X: Low vitamin A status (RBP < 0.70 µmol/L) prevalence ratio

#### 4X10: Stratified by Average SQ-LNS compliance (insufficient comparisons)

### Supplemental figure 4Y: Low vitamin A status (RBP < 0.70 µmol/L) prevalence difference

#### 4Y1: Stratified by Geographic region (insufficient comparisons)

**Supplemental figure 4Y: Low vitamin A status (RBP < 0.70 µmol/L) prevalence difference**

#### 4Y2: Stratified by Anemia burden (insufficient comparisons)

#### Supplemental figure 4Y: Low vitamin A status (RBP $< 0.70$ µmol/L) prevalence difference

### 4Y3: Stratified by Malaria prevalence (insufficient comparisons)

**Supplemental figure 4Y: Low vitamin A status (RBP $<$ 0.70 µmol/L) prevalence difference**

#### 4Y4: Stratified by Inflammation burden (insufficient comparisons)

**Supplemental figure 4Y: Low vitamin A status (RBP < 0.70 µmol/L) prevalence difference**

#### 4Y5: Stratified by Source water quality (insufficient comparisons)

**Supplemental figure 4Y: Low vitamin A status (RBP < 0.70 µmol/L) prevalence difference**

#### 4Y6: Stratified by Sanitation (insufficient comparisons)

**Supplemental figure 4Y: Low vitamin A status (RBP < 0.70 µmol/L) prevalence difference**

#### 4Y7: Stratified by Supplement duration (insufficient comparisons)

**Supplemental figure 4Y: Low vitamin A status (RBP < 0.70 µmol/L) prevalence difference**

#### 4Y8: Stratified by Iron dose (insufficient comparisons)

**Supplemental figure 4Y: Low vitamin A status (RBP < 0.70 µmol/L) prevalence difference**

#### 4Y9: Stratified by Frequency of contact (insufficient comparisons)

**Supplemental figure 4Y: Low vitamin A status (RBP < 0.70 µmol/L) prevalence difference**

#### 4Y10: Stratified by Average SQ-LNS compliance (insufficient comparisons)

### Supplemental figure 4Z: Marginal vitamin A status (RBP < 1.05 µmol/L) prevalence ratio

#### 4Z1: Stratified by Geographic region (insufficient comparisons)

**Supplemental figure 4Z: Marginal vitamin A status (RBP < 1.05 µmol/L) prevalence ratio**

#### 4Z2: Stratified by Anemia burden (insufficient comparisons)

**Supplemental figure 4Z: Marginal vitamin A status (RBP < 1.05 µmol/L) prevalence ratio**

#### 4Z3: Stratified by Malaria prevalence (insufficient comparisons)

**Supplemental figure 4Z: Marginal vitamin A status (RBP < 1.05 µmol/L) prevalence ratio**

#### 4Z4: Stratified by Inflammation burden (insufficient comparisons)

**Supplemental figure 4Z: Marginal vitamin A status (RBP < 1.05 µmol/L) prevalence ratio**

#### 4Z5: Stratified by Source water quality (insufficient comparisons)

**Supplemental figure 4Z: Marginal vitamin A status (RBP < 1.05 µmol/L) prevalence ratio**

#### 4Z6: Stratified by Sanitation (insufficient comparisons)

**Supplemental figure 4Z: Marginal vitamin A status (RBP < 1.05 µmol/L) prevalence ratio**

#### 4Z7: Stratified by Supplement duration (insufficient comparisons)

**Supplemental figure 4Z: Marginal vitamin A status (RBP < 1.05 µmol/L) prevalence ratio**

#### 4Z8: Stratified by Iron dose (insufficient comparisons)

**Supplemental figure 4Z: Marginal vitamin A status (RBP $<$ 1.05 µmol/L) prevalence ratio**

#### 4Z9: Stratified by Frequency of contact (insufficient comparisons)

**Supplemental figure 4Z: Marginal vitamin A status (RBP < 1.05 µmol/L) prevalence ratio**

#### 4Z10: Stratified by Average SQ-LNS compliance (insufficient comparisons)

### Supplemental figure 4AA: Marginal vitamin A status (RBP < 1.05 µmol/L) prevalence difference

#### 4AA1: Stratified by Geographic region (insufficient comparisons)

##### Supplemental figure 4AA: Marginal vitamin A status (RBP < 1.05 µmol/L) prevalence difference

#### 4AA2: Stratified by Anemia burden (insufficient comparisons)

**Supplemental figure 4AA: Marginal vitamin A status (RBP < 1.05 µmol/L) prevalence difference**

#### 4AA3: Stratified by Malaria prevalence (insufficient comparisons)

**Supplemental figure 4AA: Marginal vitamin A status (RBP < 1.05 µmol/L) prevalence difference**

#### 4AA4: Stratified by Inflammation burden (insufficient comparisons)

**Supplemental figure 4AA: Marginal vitamin A status (RBP < 1.05 µmol/L) prevalence difference**

#### 4AA5: Stratified by Source water quality (insufficient comparisons)

**Supplemental figure 4AA: Marginal vitamin A status (RBP < 1.05 µmol/L) prevalence difference**

#### 4AA6: Stratified by Sanitation (insufficient comparisons)

**Supplemental figure 4AA: Marginal vitamin A status (RBP < 1.05 µmol/L) prevalence difference**

#### 4AA7: Stratified by Supplement duration (insufficient comparisons)

**Supplemental figure 4AA: Marginal vitamin A status (RBP < 1.05 µmol/L) prevalence difference**

#### 4AA8: Stratified by Iron dose (insufficient comparisons)

**Supplemental figure 4AA: Marginal vitamin A status (RBP < 1.05 µmol/L) prevalence difference**

#### 4AA9: Stratified by Frequency of contact (insufficient comparisons)

**Supplemental figure 4AA: Marginal vitamin A status (RBP < 1.05 μmol/L) prevalence difference**

#### 4AA10: Stratified by Average SQ-LNS compliance (insufficient comparisons)

### Supplemental figure 5: Sensitivity analyses of effect modification of SQ-LNS on biochemical outcomes by study-level effect modifiers

These figures show the pooled estimates of effect modification by different sensitivity analyses. For continuous outcomes, the intervention effect is measured by the difference in mean of the LNS group minus control. For log transformed continuous outcomes, the intervention effect is measured by the ratio of geometric means, the effect estimate is the geometric mean in the LNS group divided by the geometric mean in the control group. For dichotomous outcomes analyzed via prevalence ratios, the effect estimate is the prevalence in the LNS group divided by the prevalence in the control group. For dichotomous outcomes analyzed via prevalence differences, the effect estimate is the prevalence in the LNS group minus the prevalence in the control group.
The labels on the left y-axis indicate which outcome is assessed. The different columns correspond to sensitivity analyses in which intervention group categorization differs. All-trial analysis includes all trials; Child-LNS-only excludes trial arms that provided both maternal and child LNS; Multi-component analysis separates comparisons within trials that included multi-component interventions, so that the SQ-LNS vs. no SQ-LNS comparisons were conducted separately between pairs of arms that included the same non-nutrition components (e.g. SQ-LNS+WASH vs. WASH; SQ-LNS vs. Control); Passive arms excluded analysis excludes passive control arms. Depending on the sensitivity analysis, there may not have been enough comparisons available to generate a pooled estimate.
sTfR, soluble transferrin receptor; ZPP, zinc protoporphyrin; RBP, retinol binding protein.

##### Supplemental figure 5A: Difference in mean differences in hemoglobin concentration

###### 5A1: By study context effect modifiers

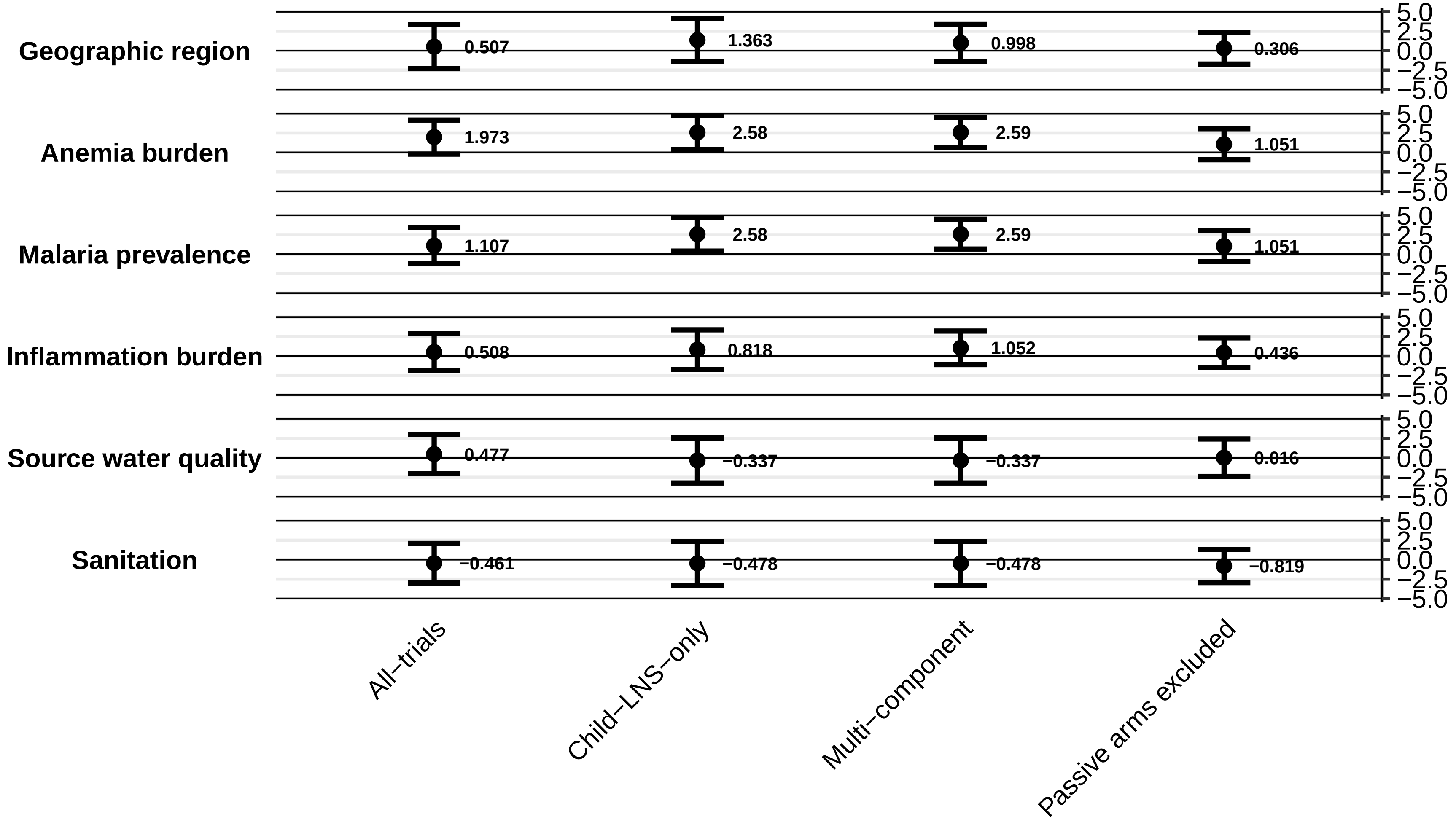

**Supplemental figure 5A: Difference in mean differences in hemoglobin concentration**

#### 5A2: By study design effect modifiers

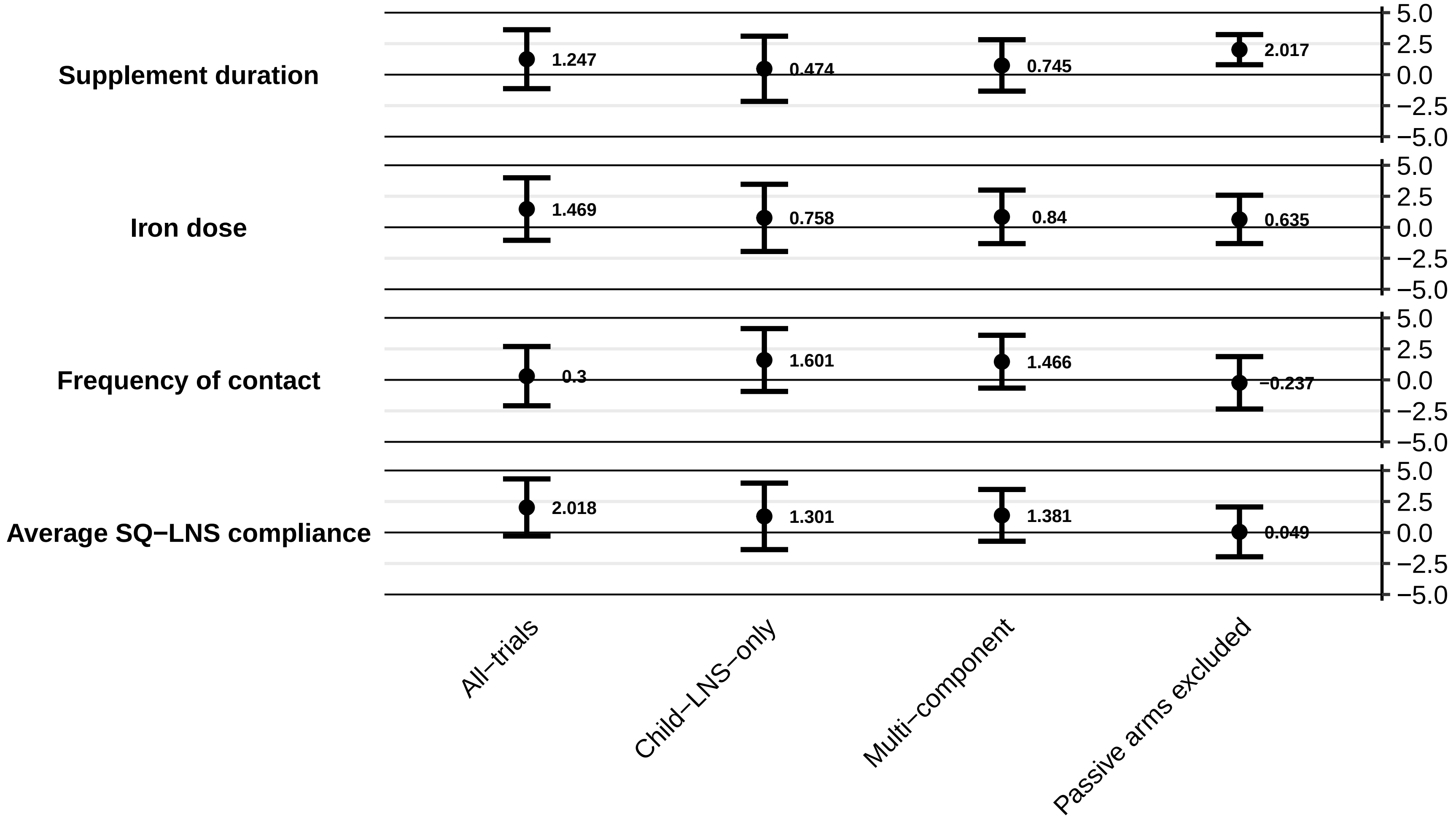

#### Supplemental figure 5B: Ratio of anemia prevalence ratios

##### 5B1: By study context effect modifiers

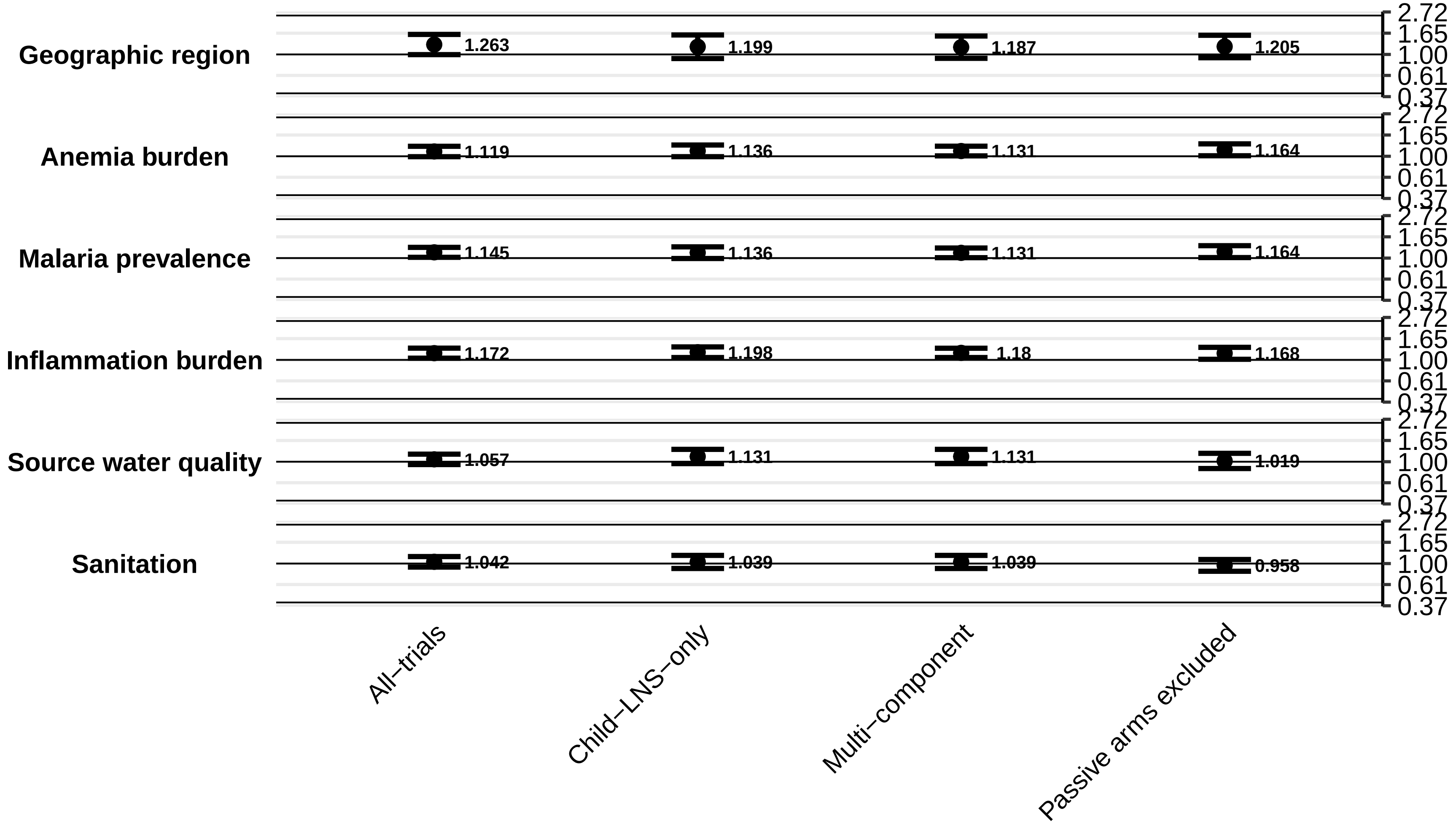

Supplemental figure 5B: Ratio of anemia prevalence ratios

#### 5B2: By study design effect modifiers

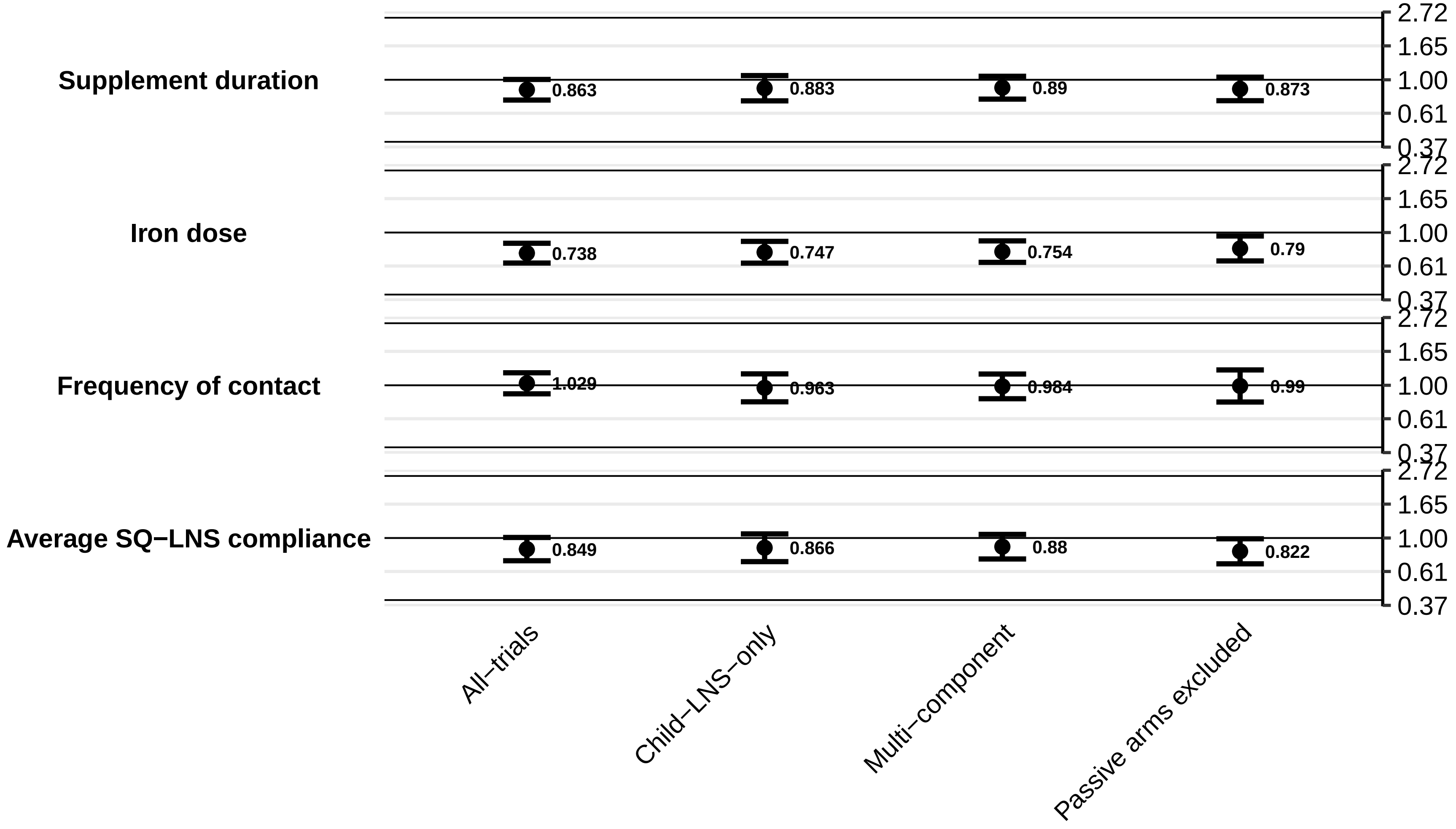

### Supplemental figure 5C: Difference in anemia prevalence differences

#### 5C1: By study context effect modifiers

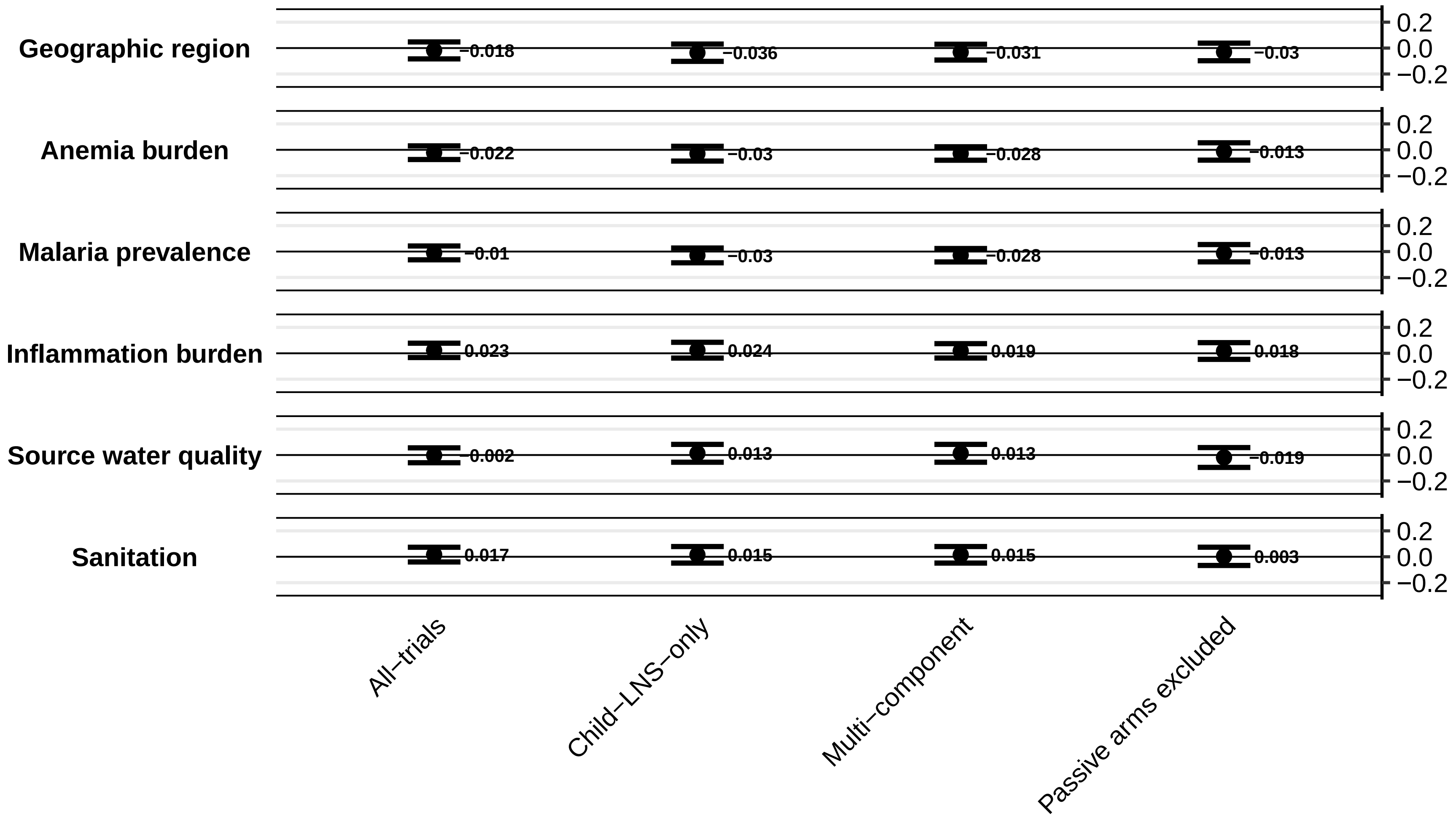

Supplemental figure 5C: Difference in anemia prevalence differences

#### 5C2: By study design effect modifiers

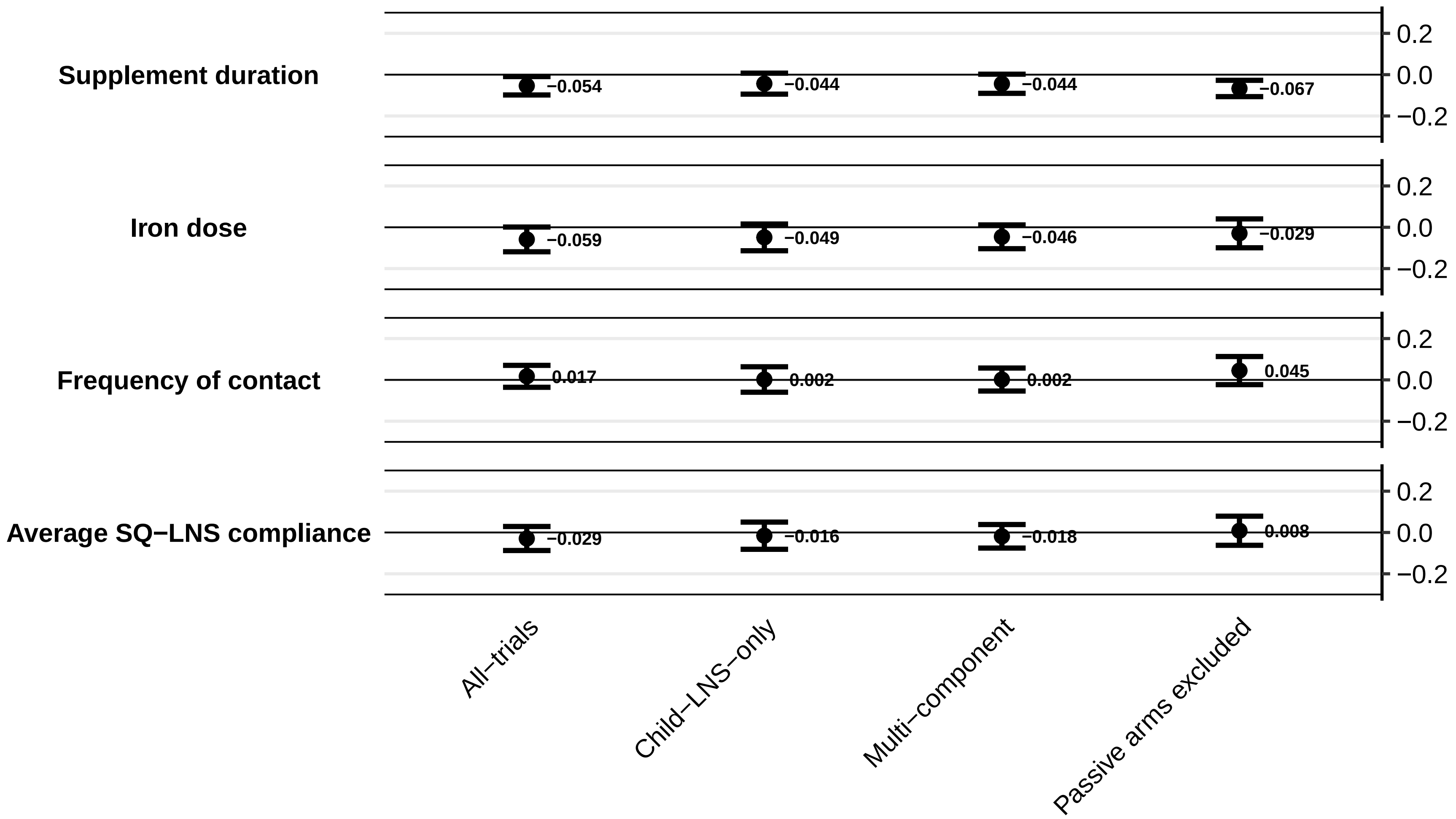

#### Supplemental figure 5D: Ratio of moderate-to-severe anemia prevalence ratios

##### 5D1: By study context effect modifiers

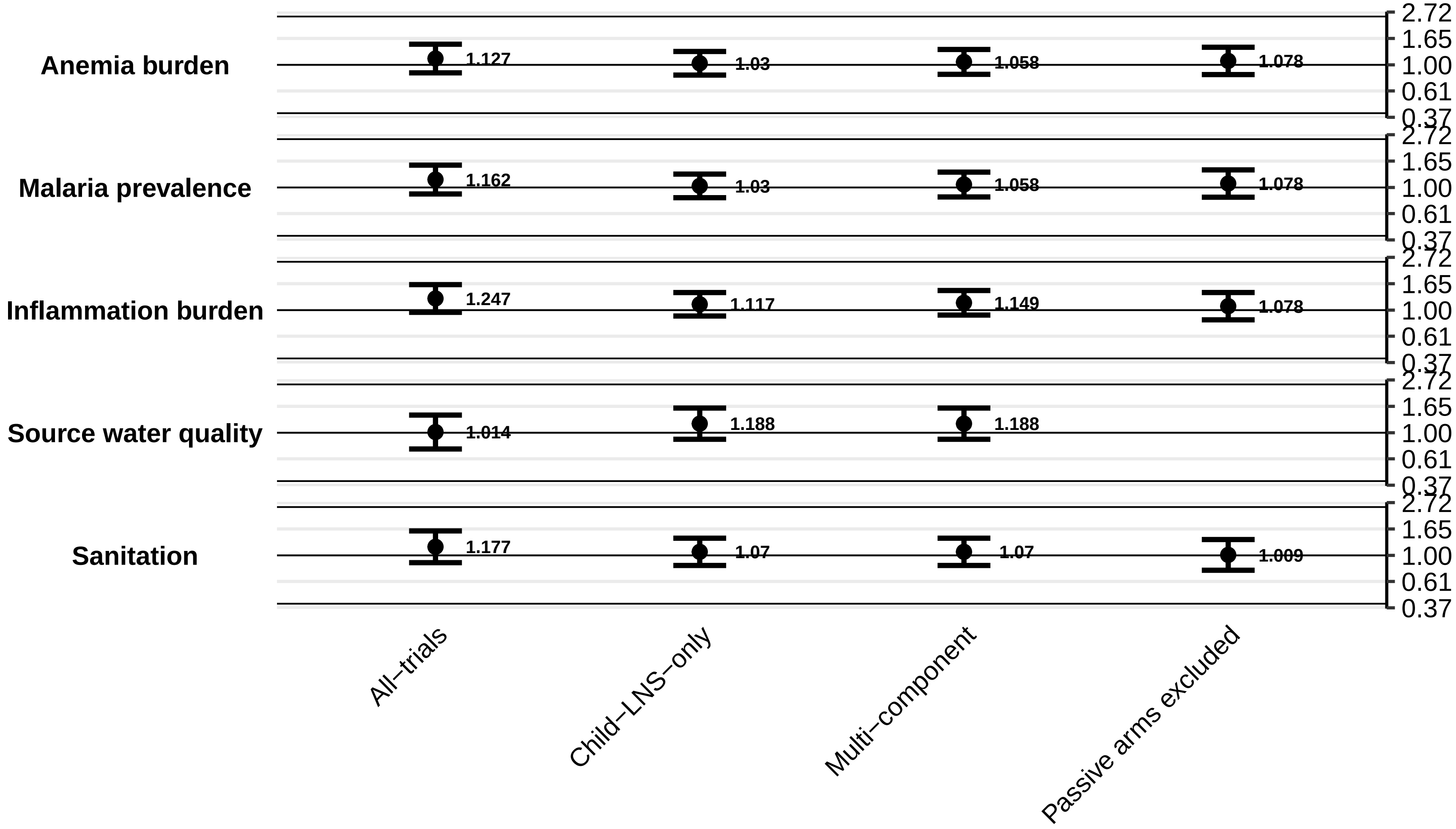

**Supplemental figure 5D: Ratio of moderate-to-severe anemia prevalence ratios**

#### 5D2: By study design effect modifiers

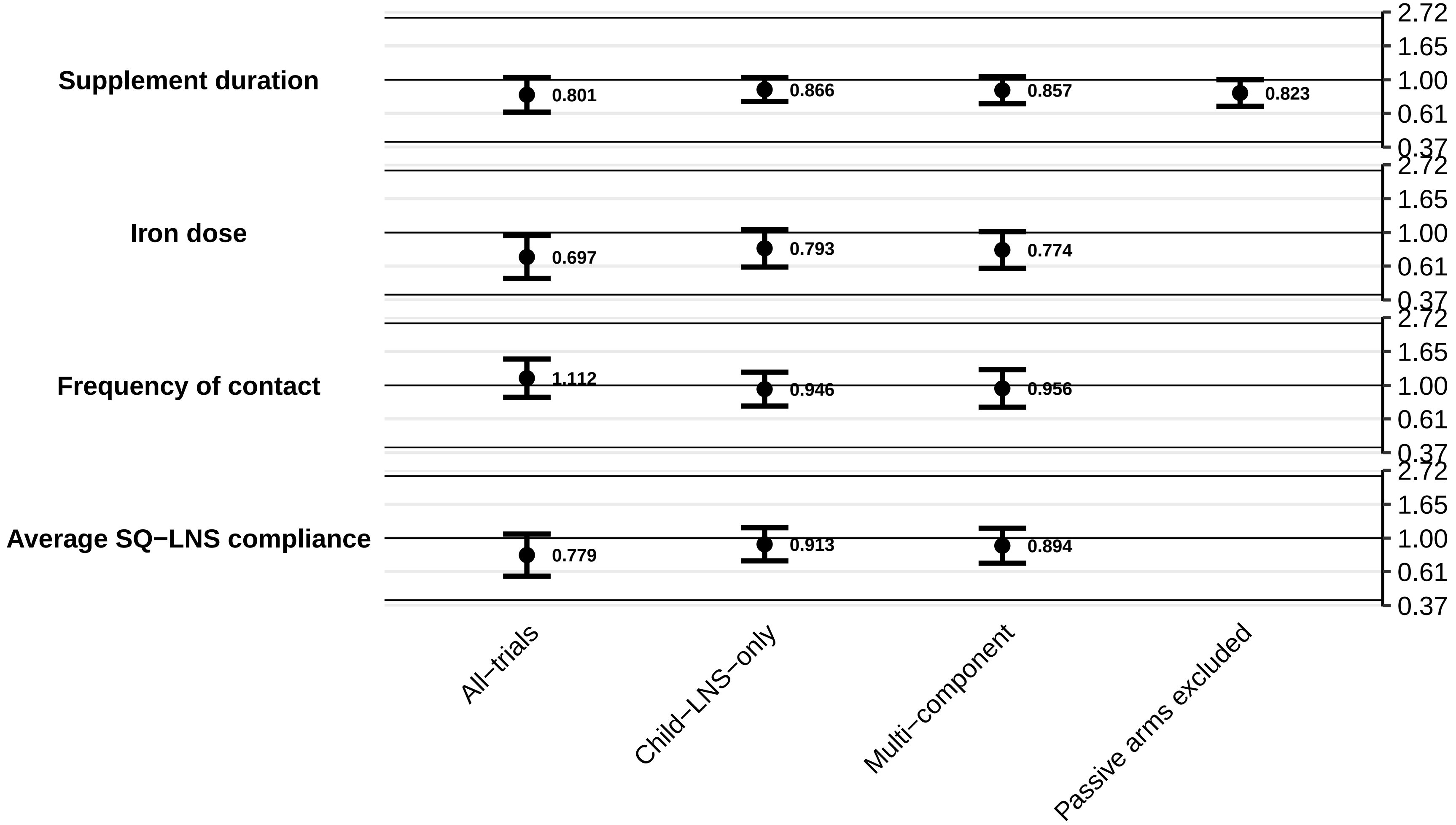

#### Supplemental figure 5E: Difference in moderate-to-severe anemia prevalence differences

##### 5E1: By study context effect modifiers

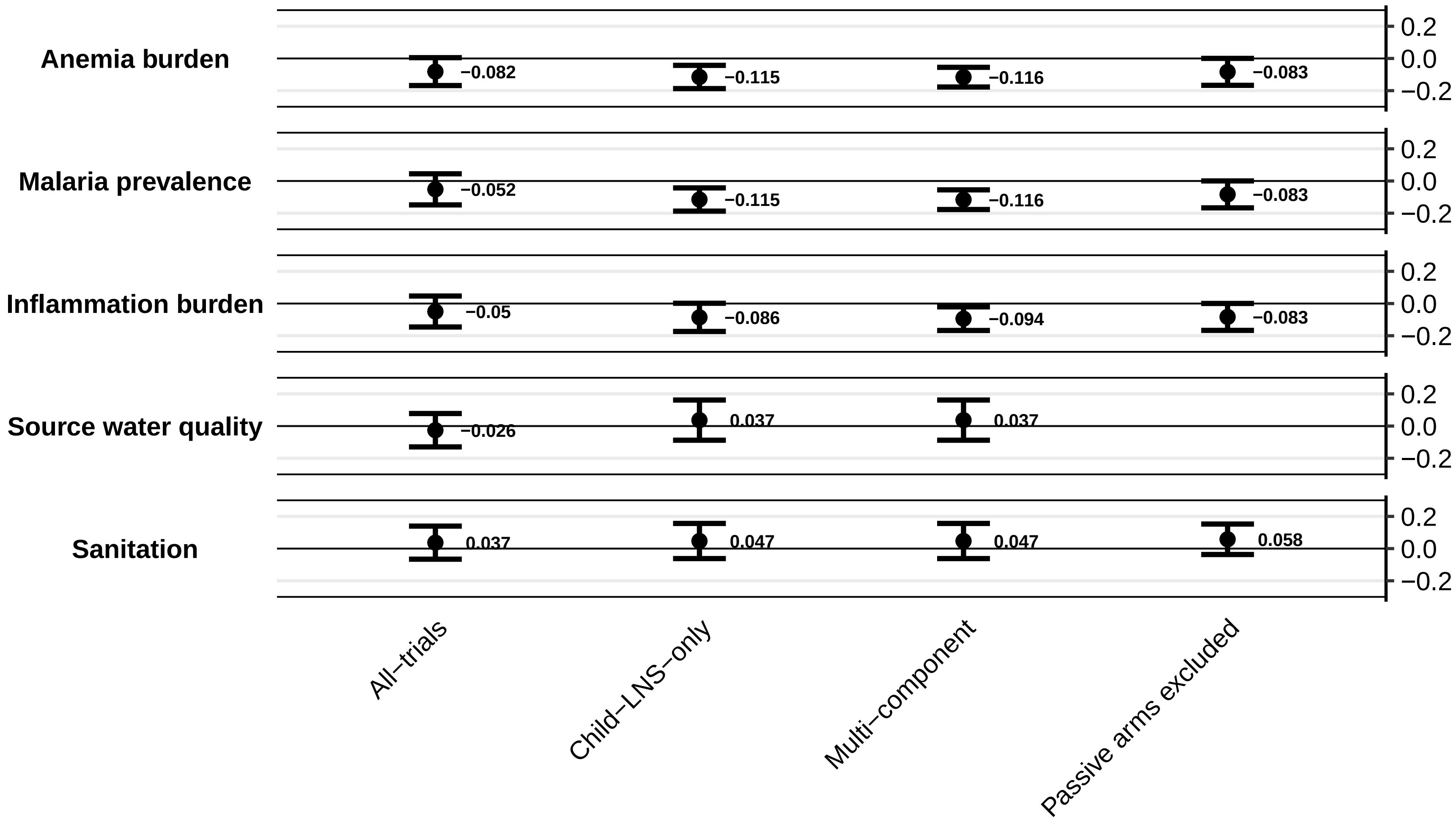

**Supplemental figure 5E: Difference in moderate-to-severe anemia prevalence differences**

#### 5E2: By study design effect modifiers

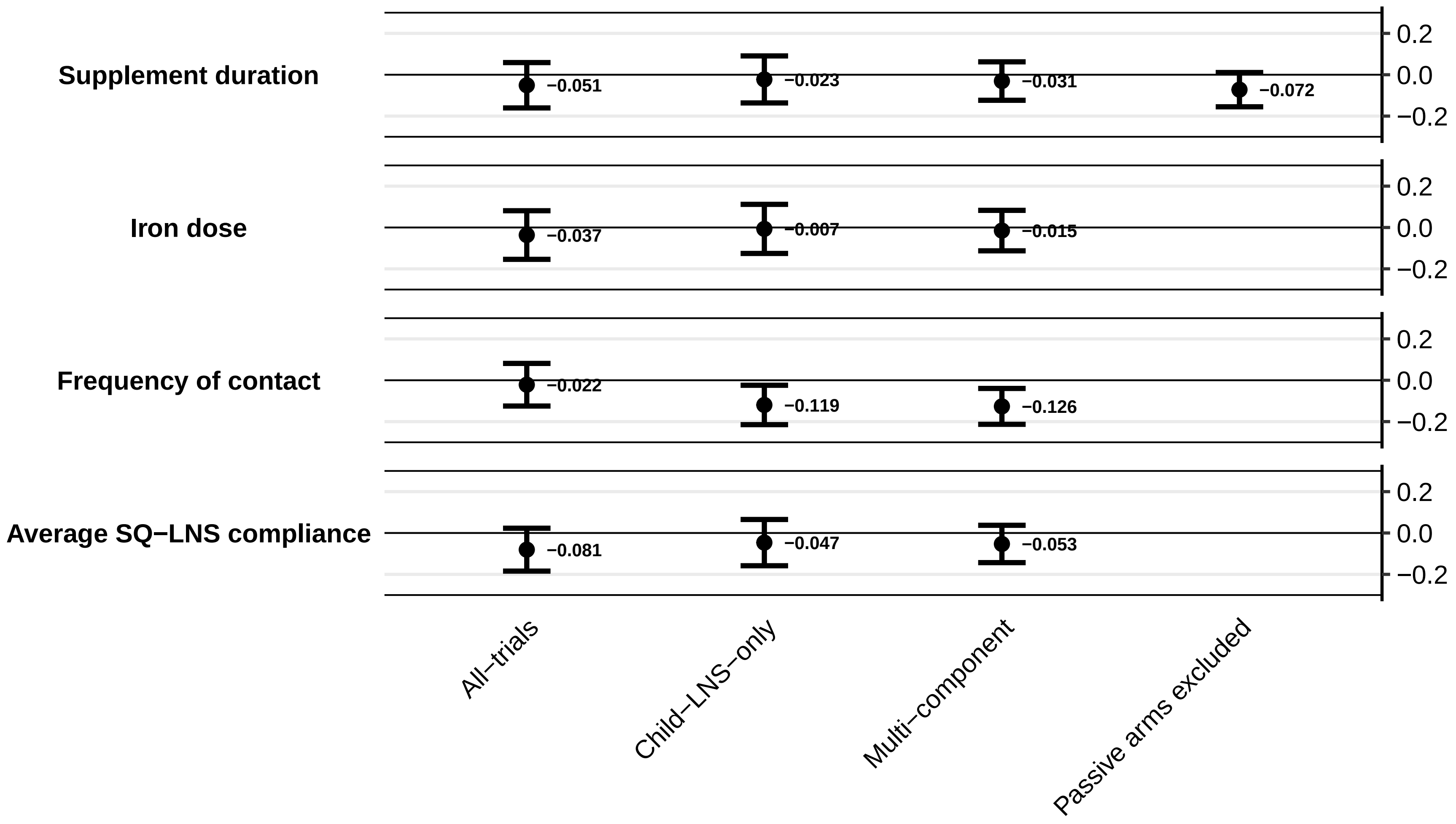

#### Supplemental figure 5F: Ratio of geometric mean ratios of ferritin concentration

##### 5F1: By study context effect modifiers

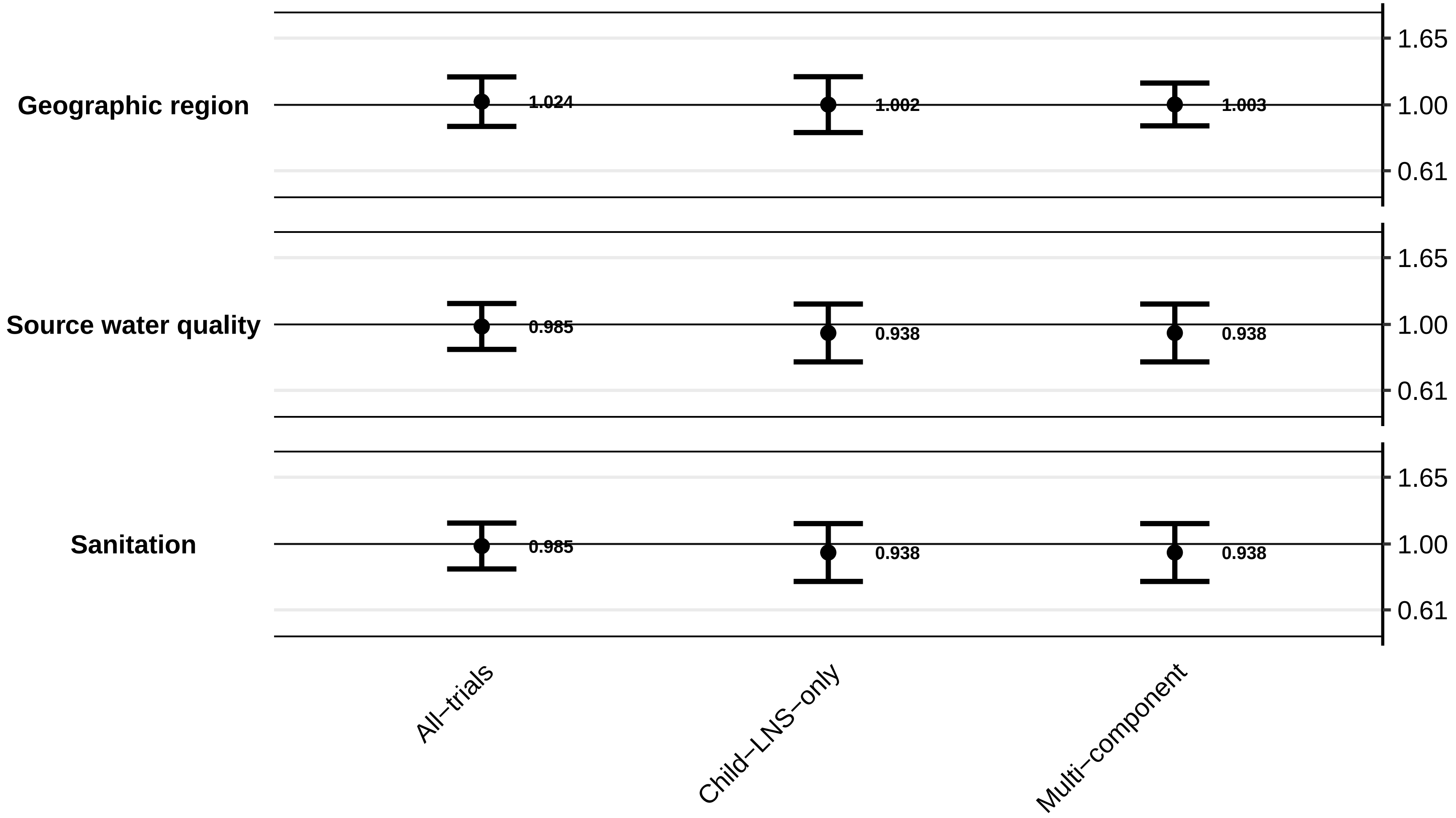

**Supplemental figure 5F: Ratio of geometric mean ratios of ferritin concentration**

#### 5F2: By study design effect modifiers

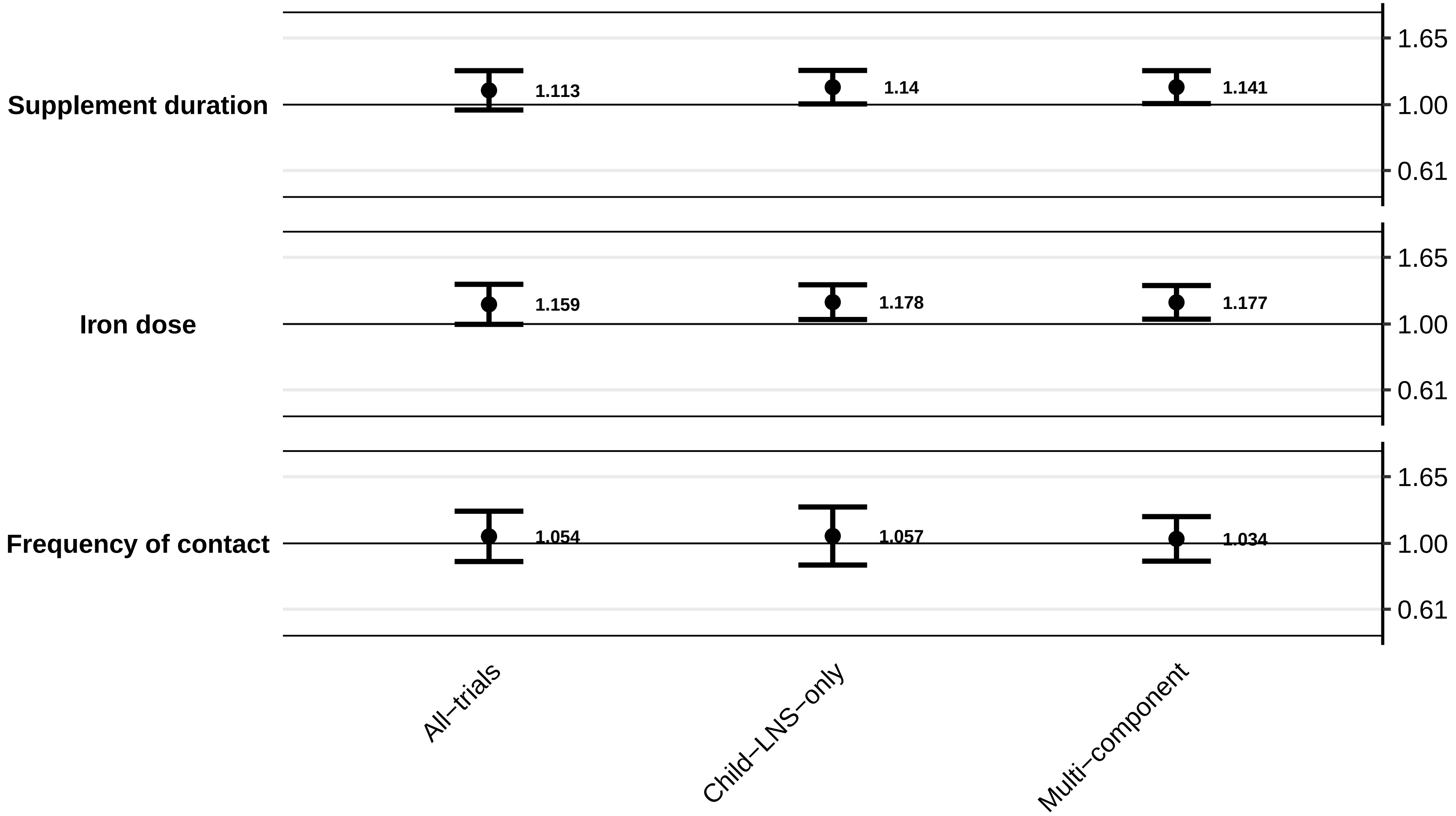

#### Supplemental figure 5G: Ratio of iron deficiency (ferritin < 12 µg/L) prevalence ratios

##### 5G1: By study context effect modifiers

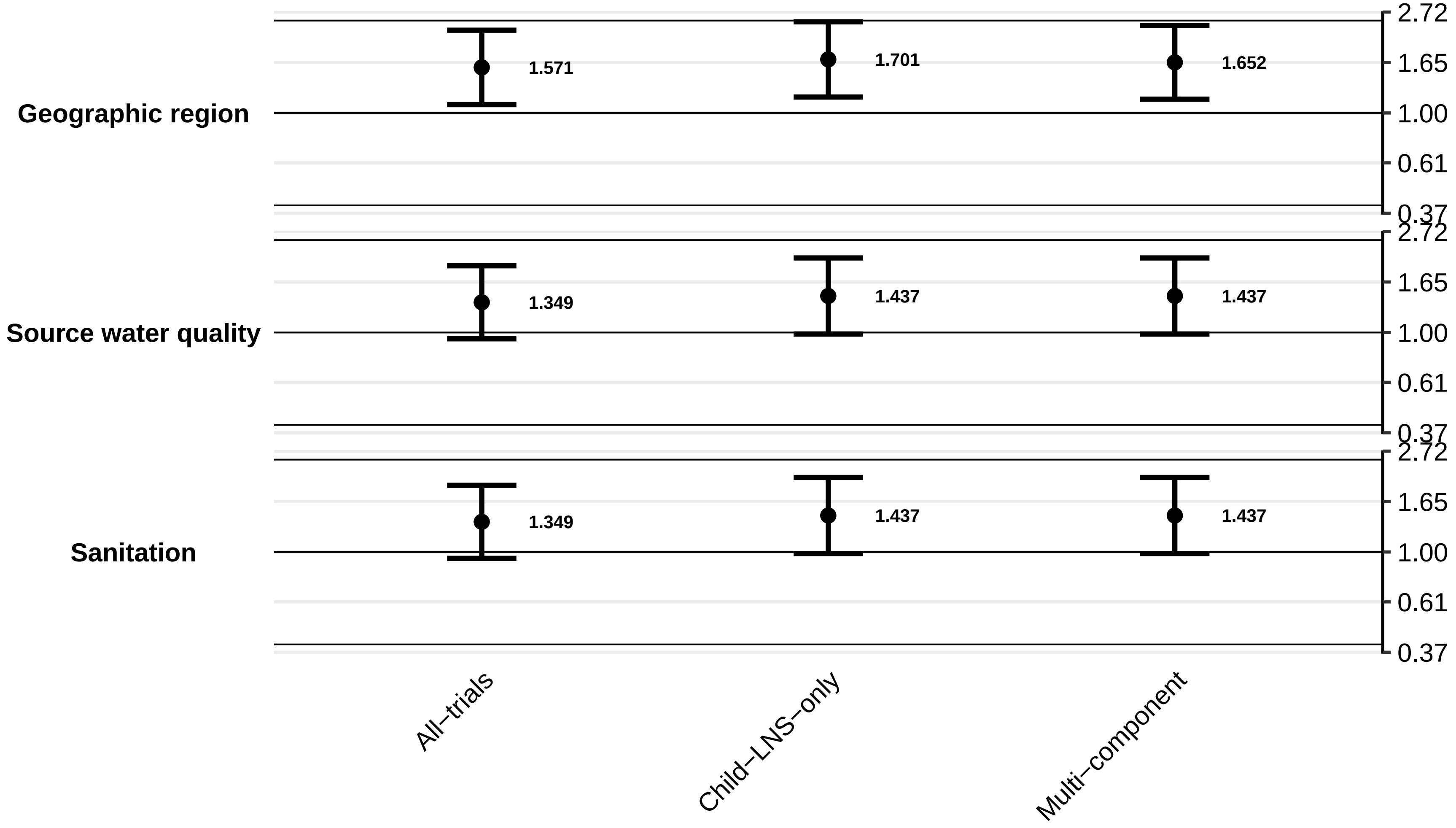

**Supplemental figure 5G: Ratio of iron deficiency (ferritin < 12 µg/L) prevalence ratios**

#### 5G2: By study design effect modifiers

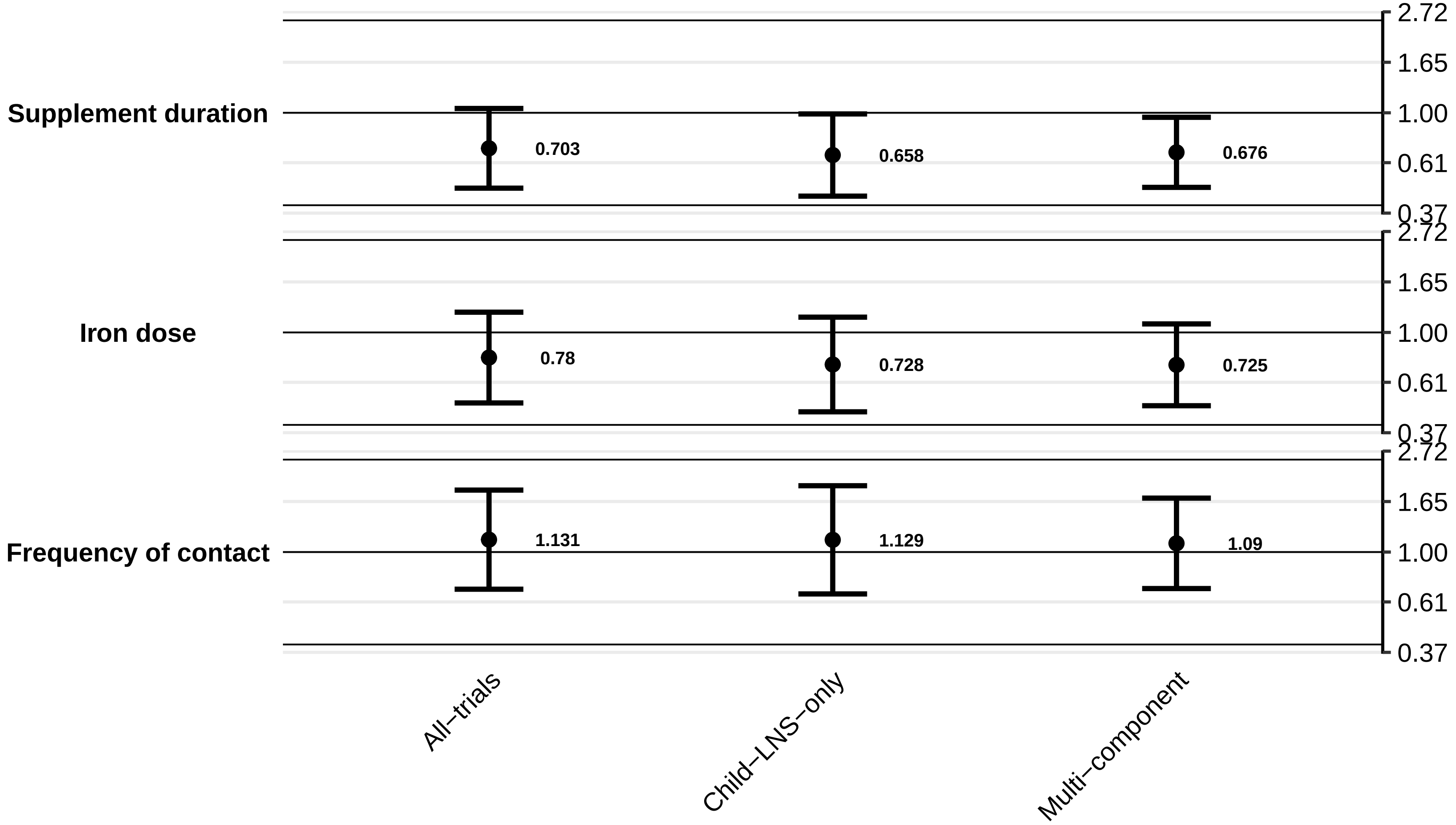

#### Supplemental figure 5H: Difference in iron deficiency (ferritin < 12 µg/L) prevalence differences

##### 5H1: By study context effect modifiers

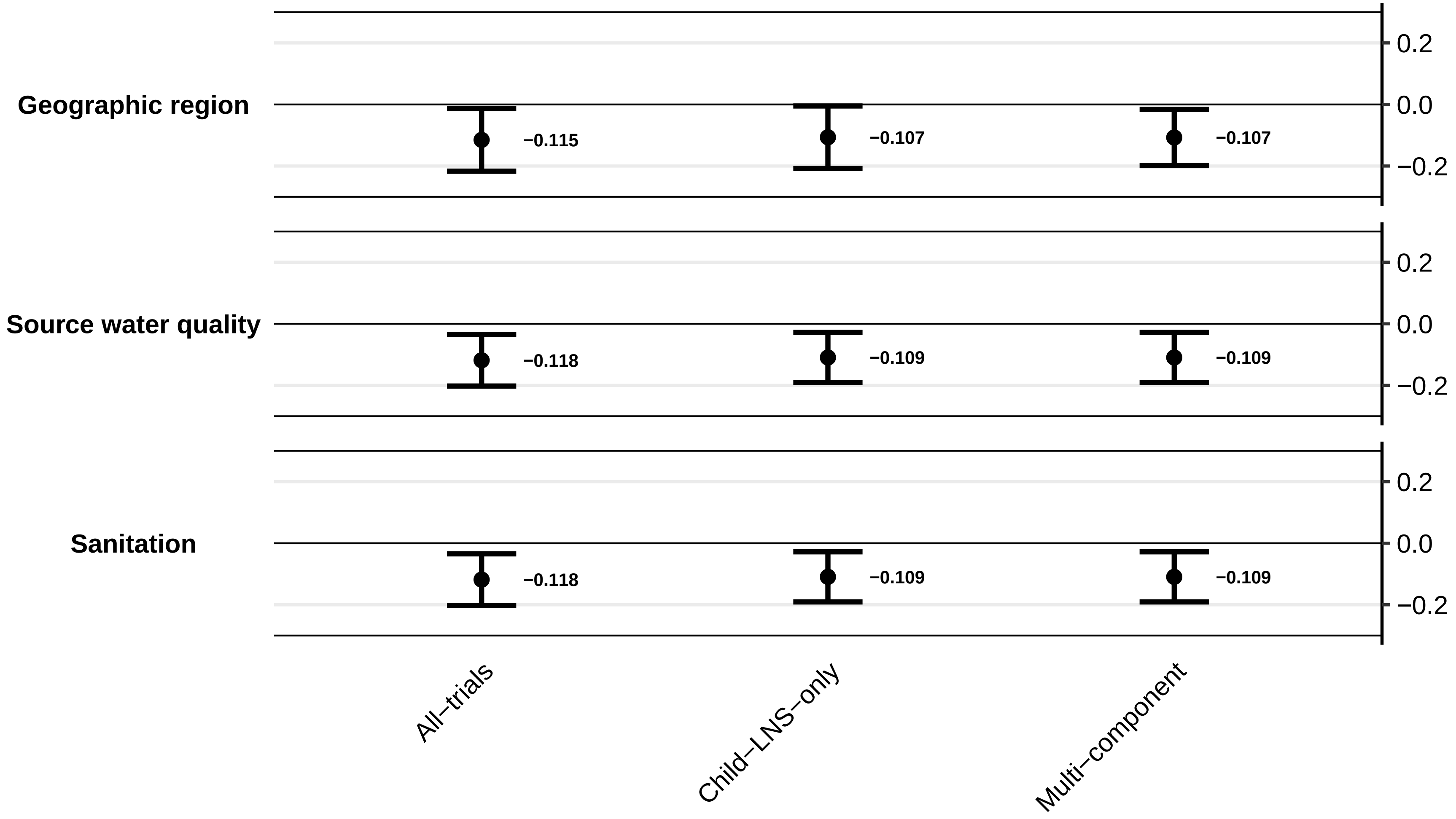

**Supplemental figure 5H: Difference in iron deficiency (ferritin < 12 μg/L) prevalence differences**

#### 5H2: By study design effect modifiers

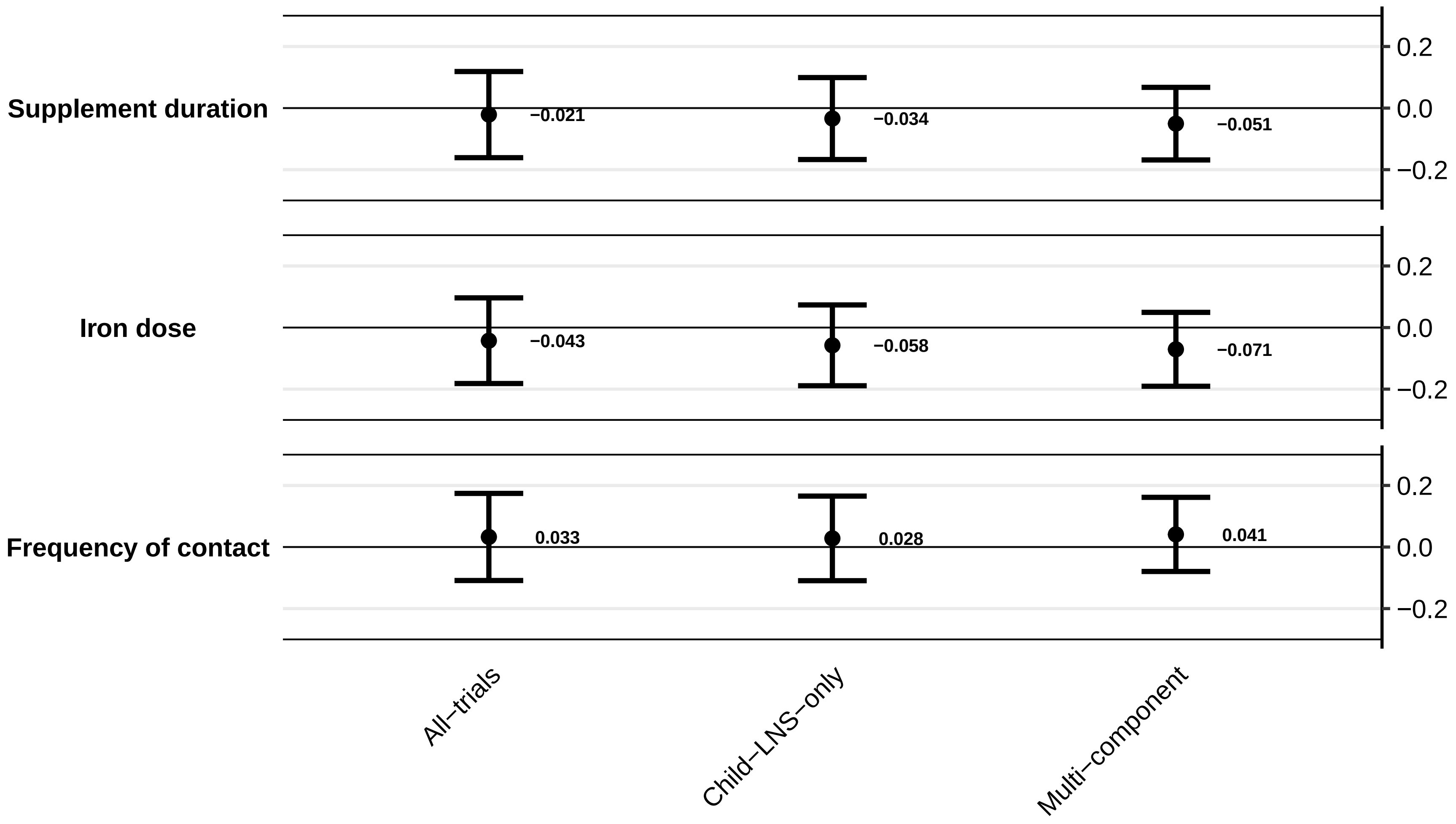

#### Supplemental figure 5I: Ratio of iron deficiency anemia prevalence ratios

##### 5I1: By study context effect modifiers

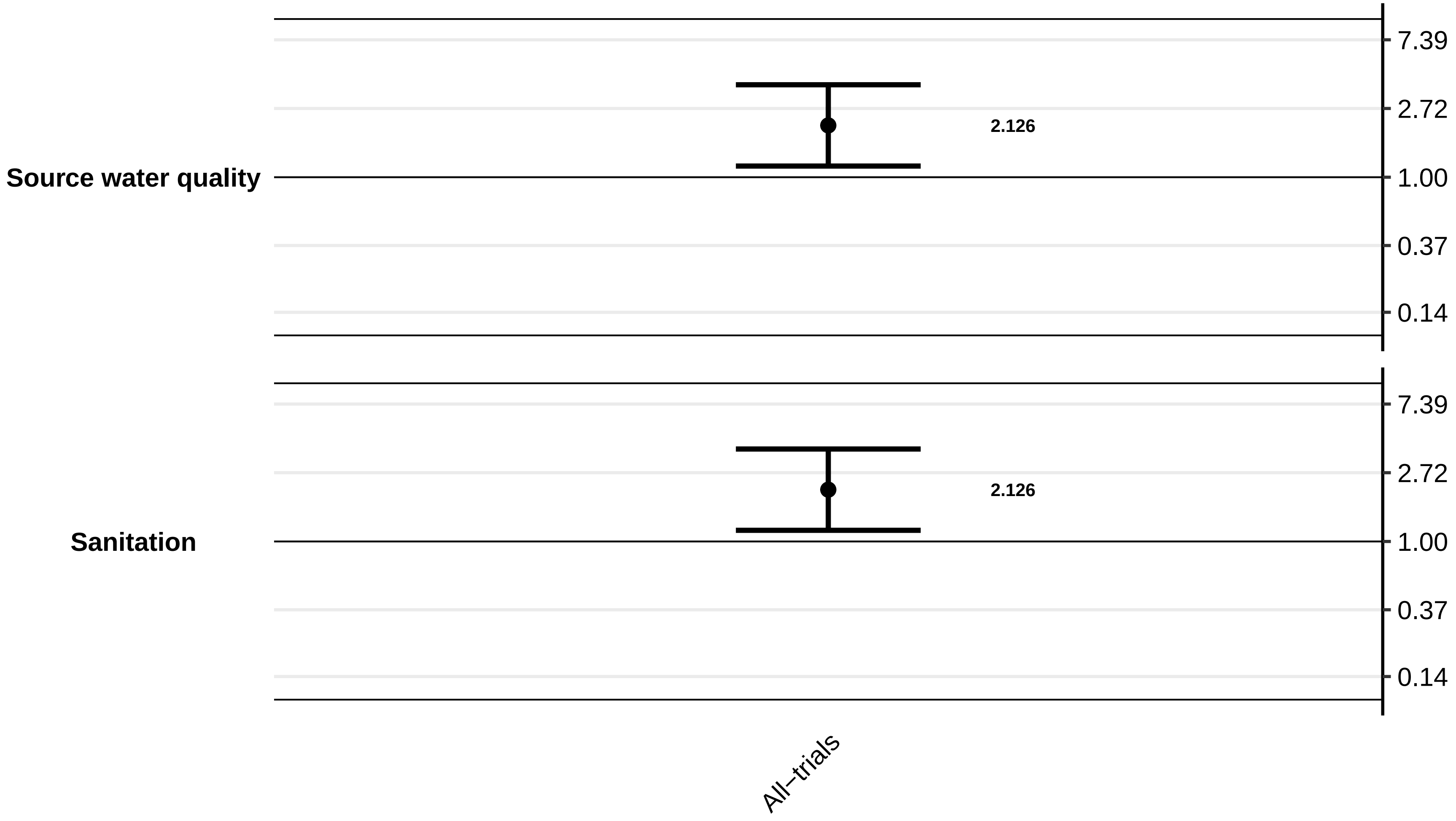

**Supplemental figure 5I: Ratio of iron deficiency anemia prevalence ratios**

#### 5I2: By study design effect modifiers

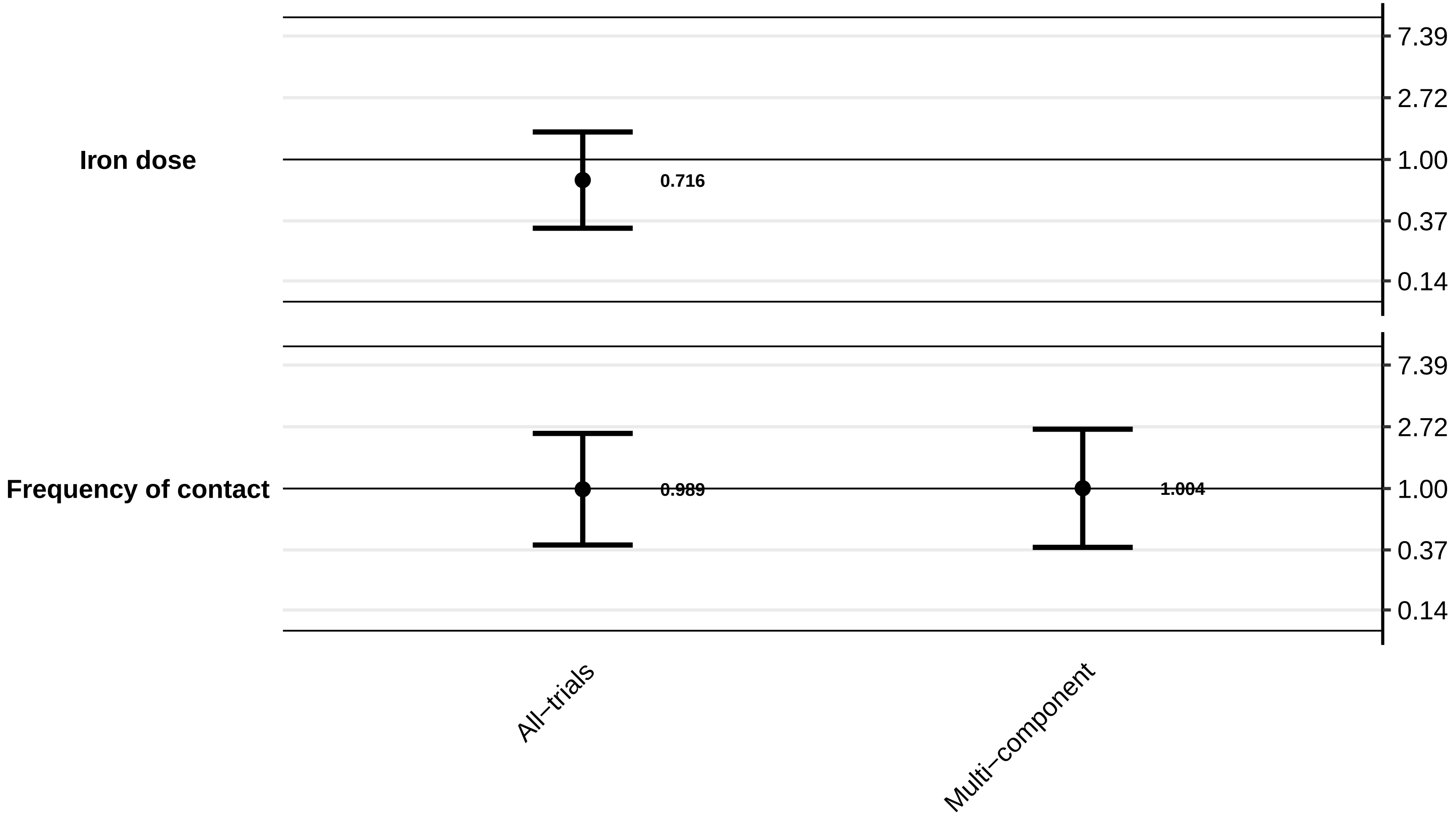

#### Supplemental figure 5J: Difference in iron deficiency anemia prevalence differences

##### 5J1: By study context effect modifiers

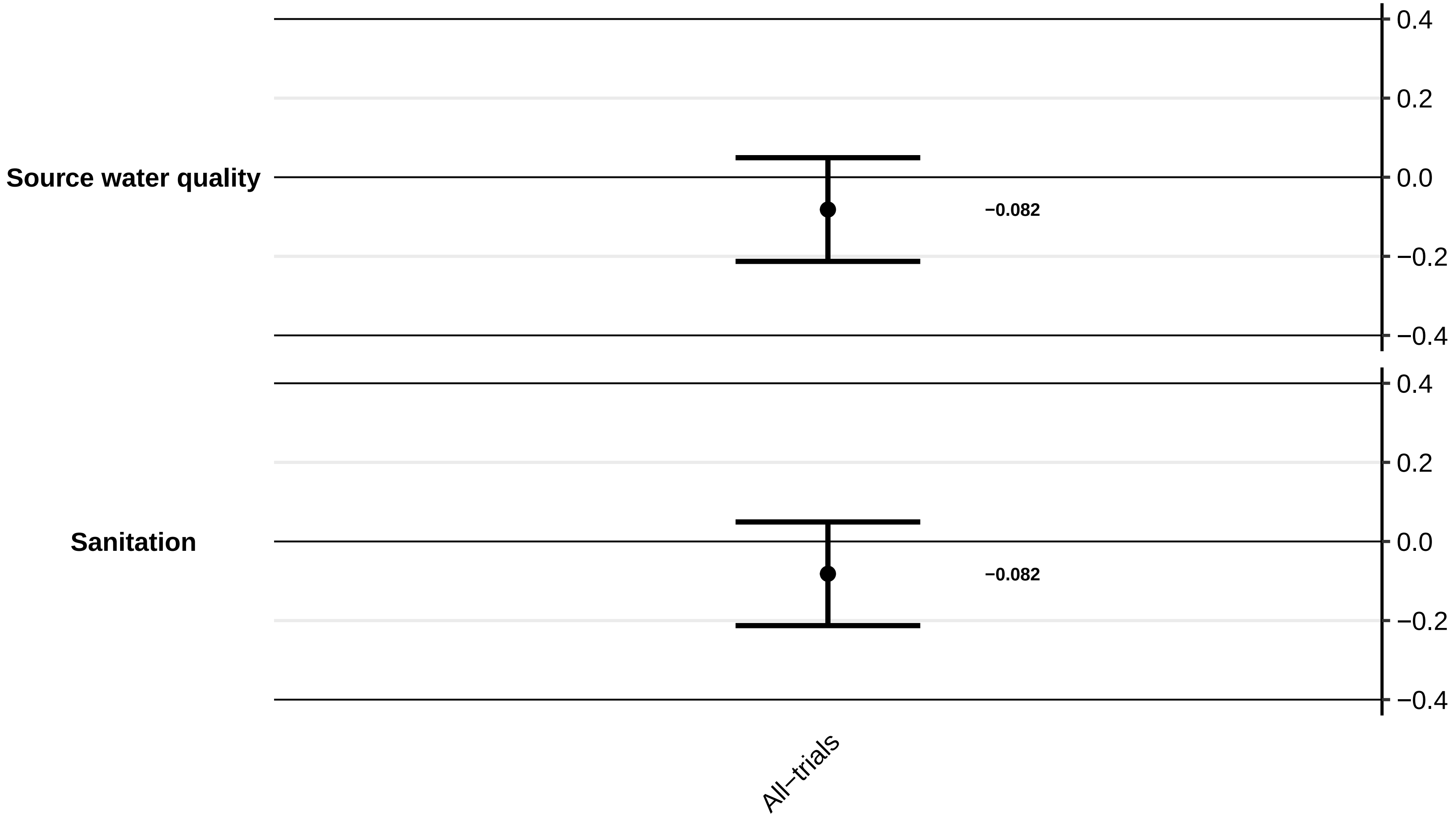

**Supplemental figure 5J: Difference in iron deficiency anemia prevalence differences**

#### 5J2: By study design effect modifiers

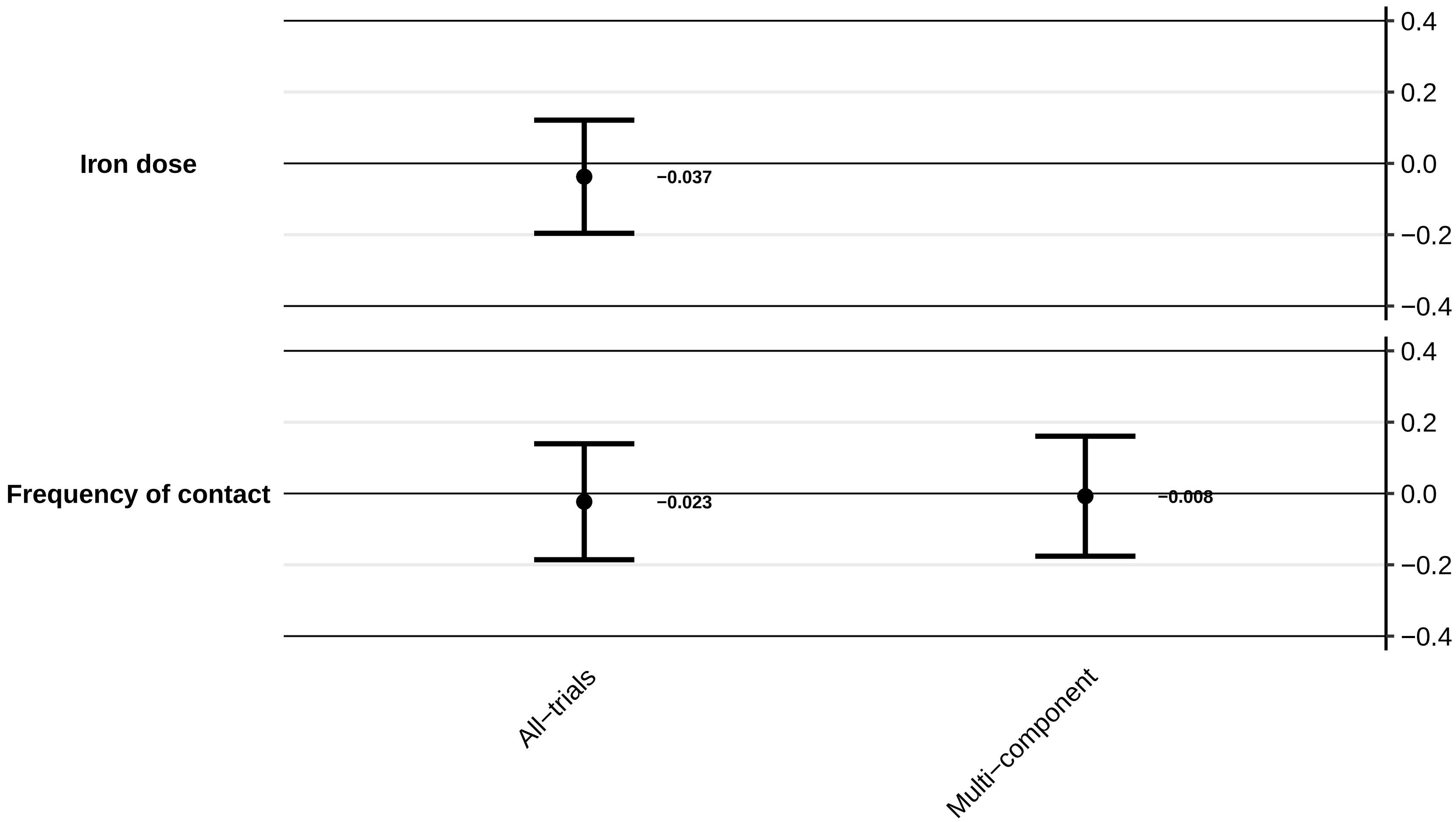

##### Supplemental figure 5K: Ratio of geometric mean ratios of soluble transferrin receptor concentration

###### 5K1: By study context effect modifiers

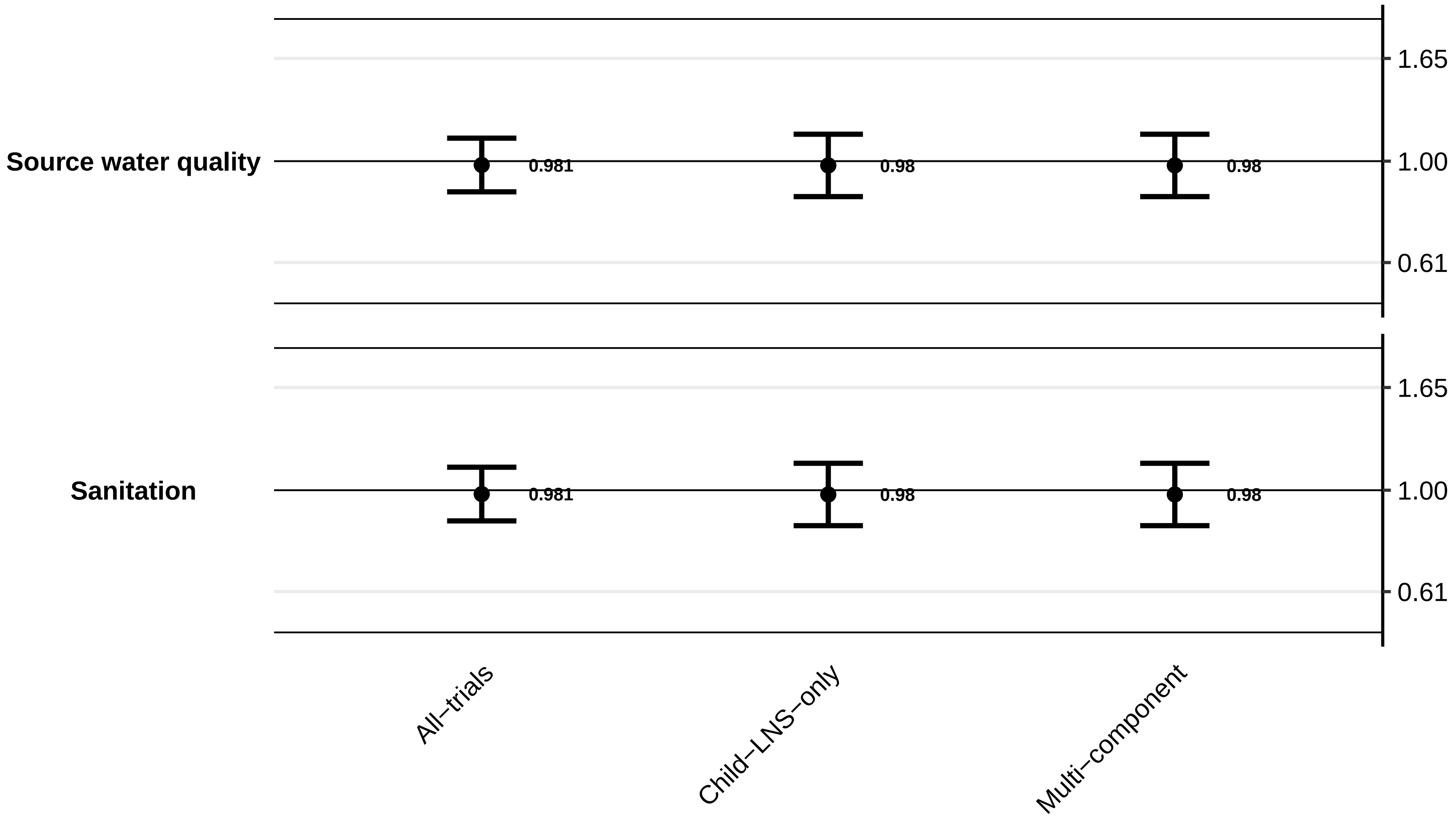

**Supplemental figure 5K: Ratio of geometric mean ratios of soluble transferrin receptor concentration**

#### 5K2: By study design effect modifiers

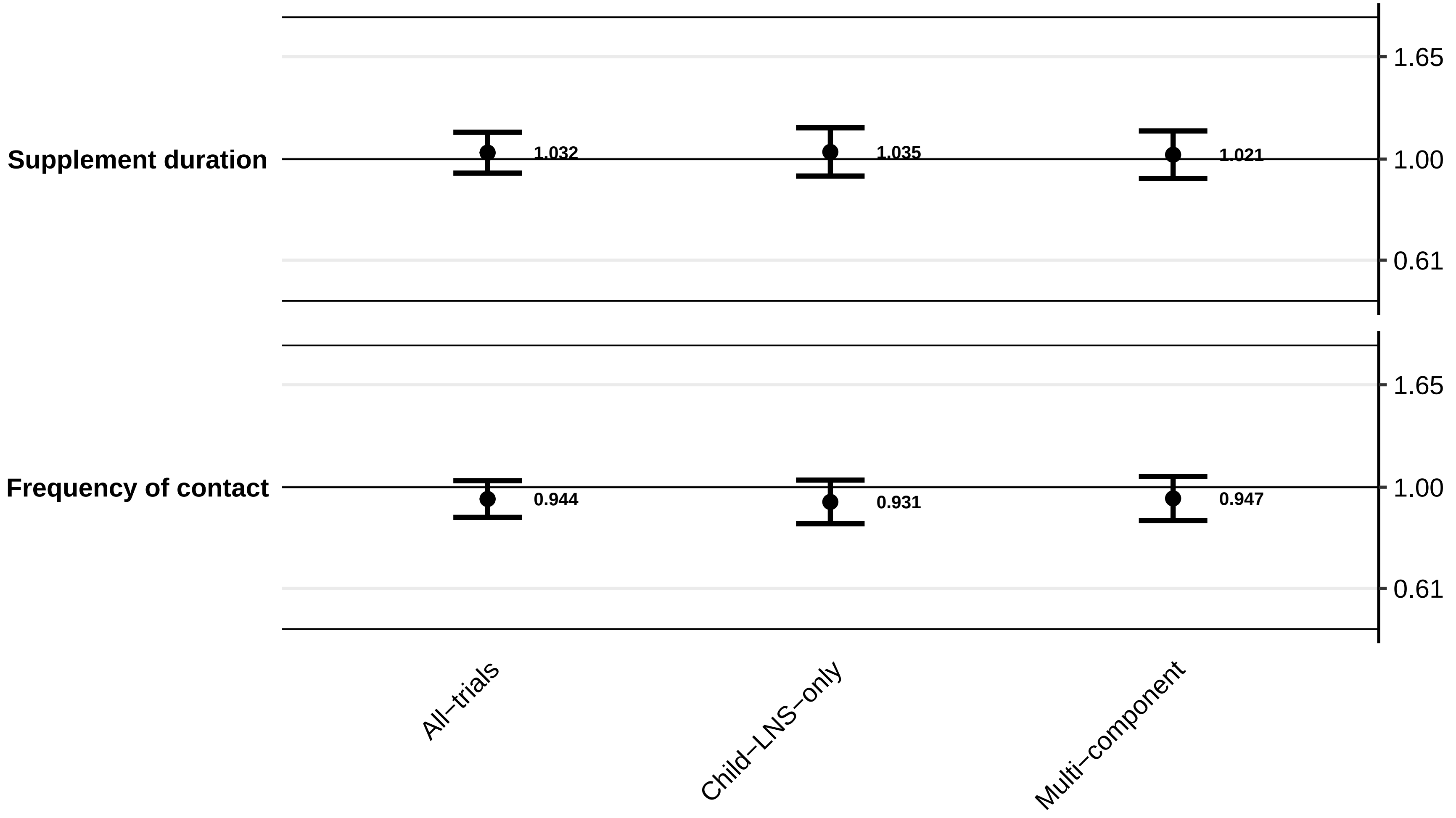

#### Supplemental figure 5L: Ratio of elevated soluble transferrin receptor prevalence ratios

##### 5L1: By study context effect modifiers

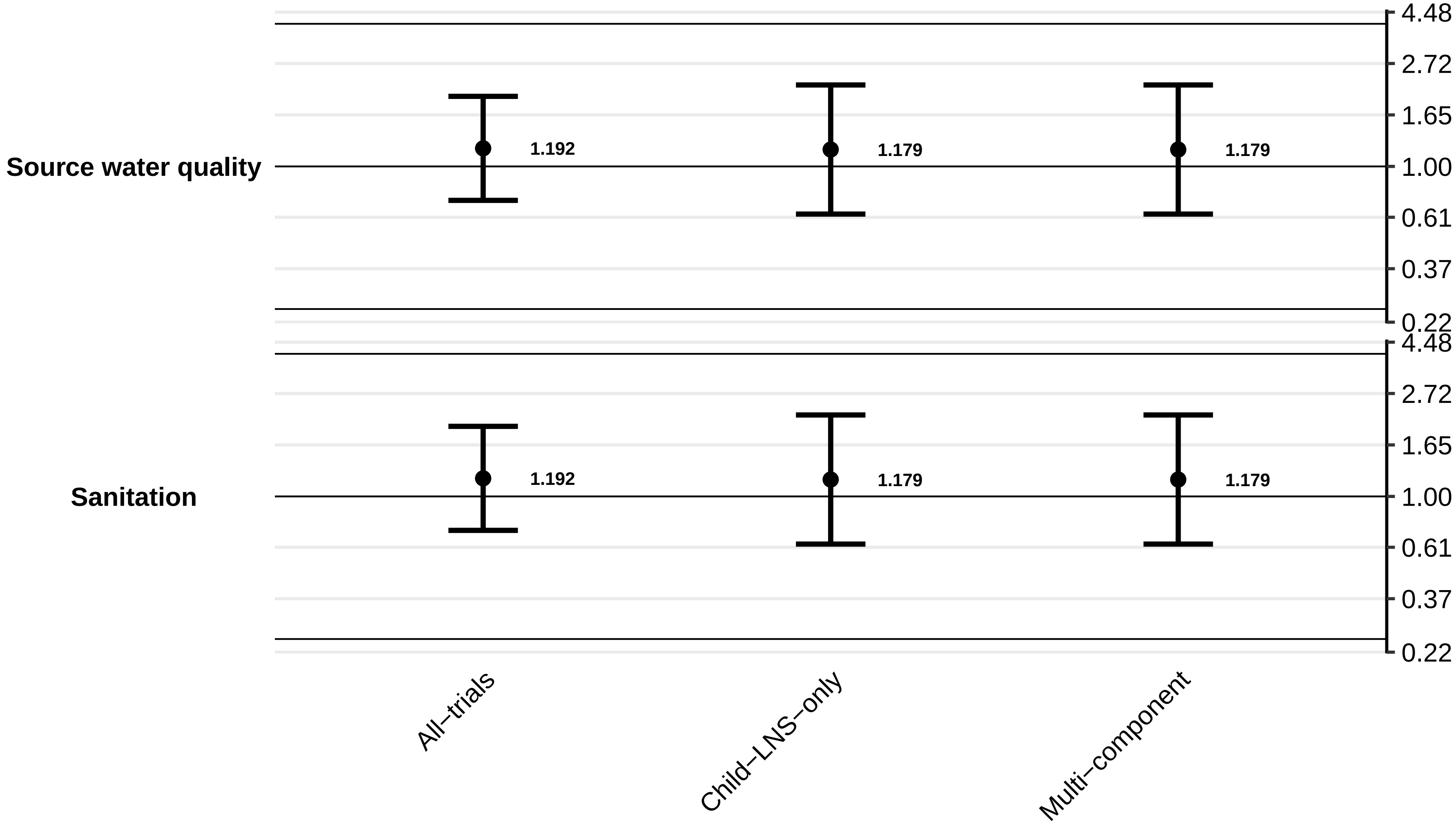

**Supplemental figure 5L: Ratio of elevated soluble transferrin receptor prevalence ratios**

#### 5L2: By study design effect modifiers

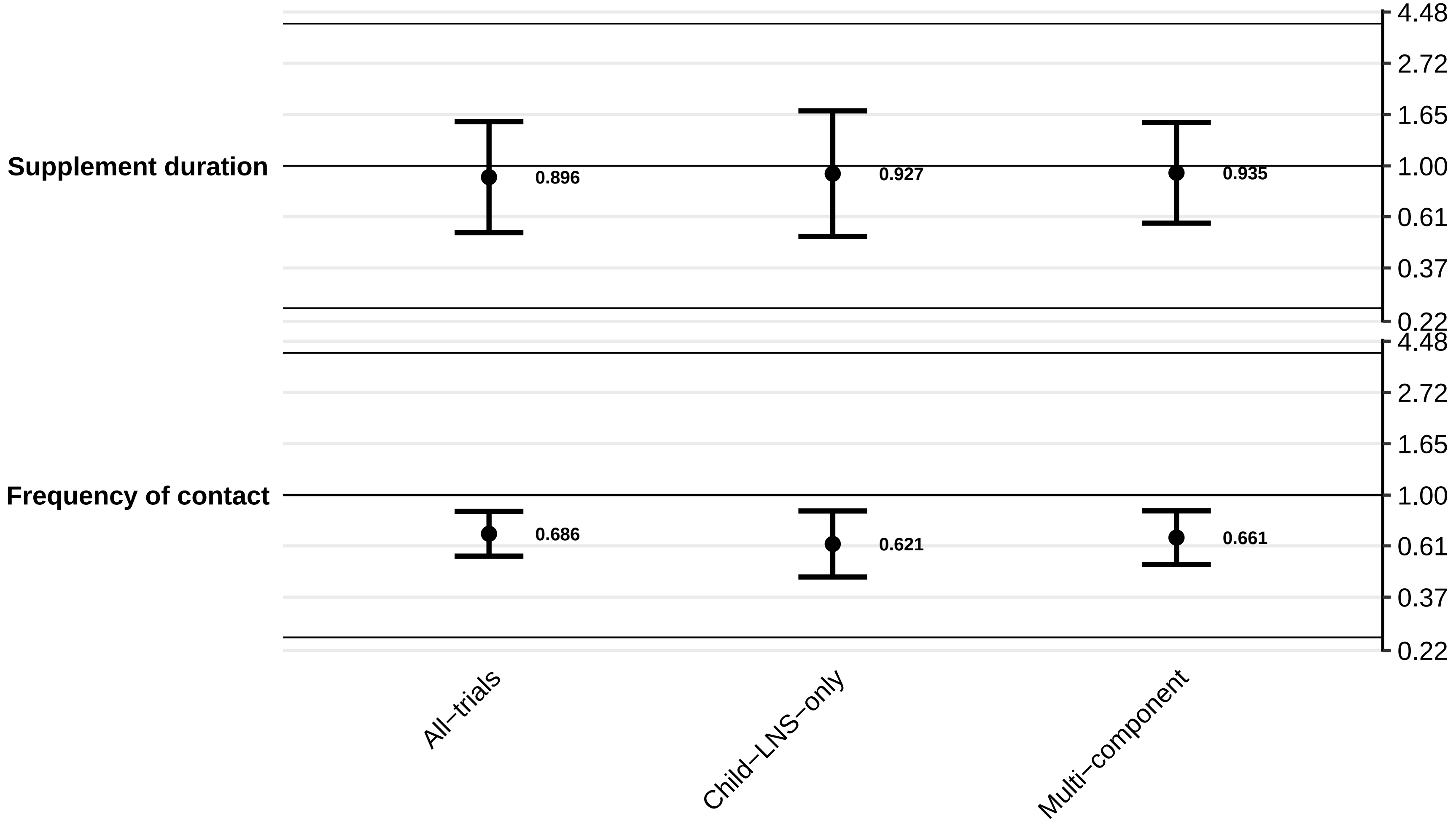

#### Supplemental figure 5M: Difference in elevated soluble transferrin receptor prevalence differences

##### 5M1: By study context effect modifiers

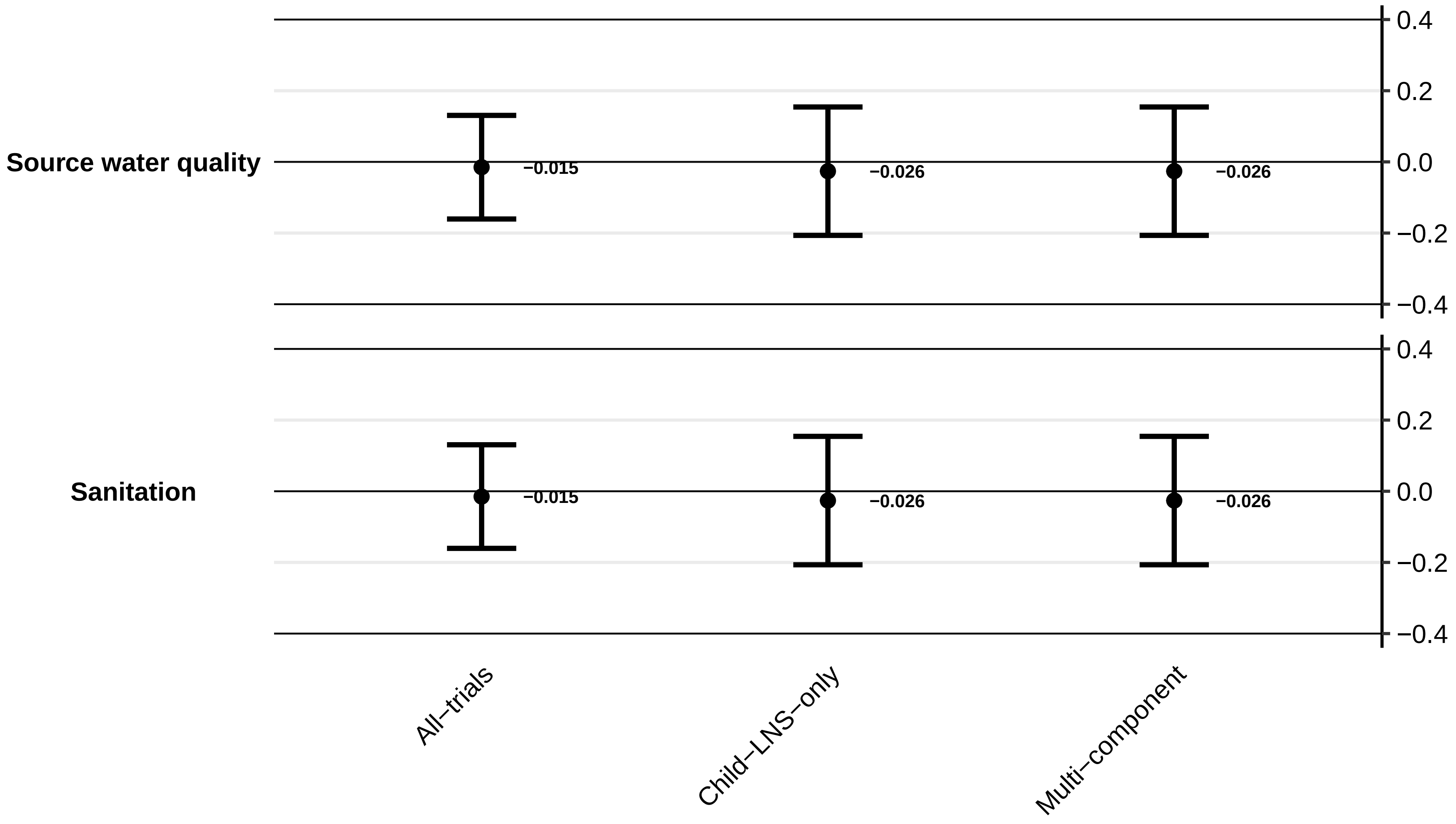

**Supplemental figure 5M: Difference in elevated soluble transferrin receptor prevalence differences**

#### 5M2: By study design effect modifiers

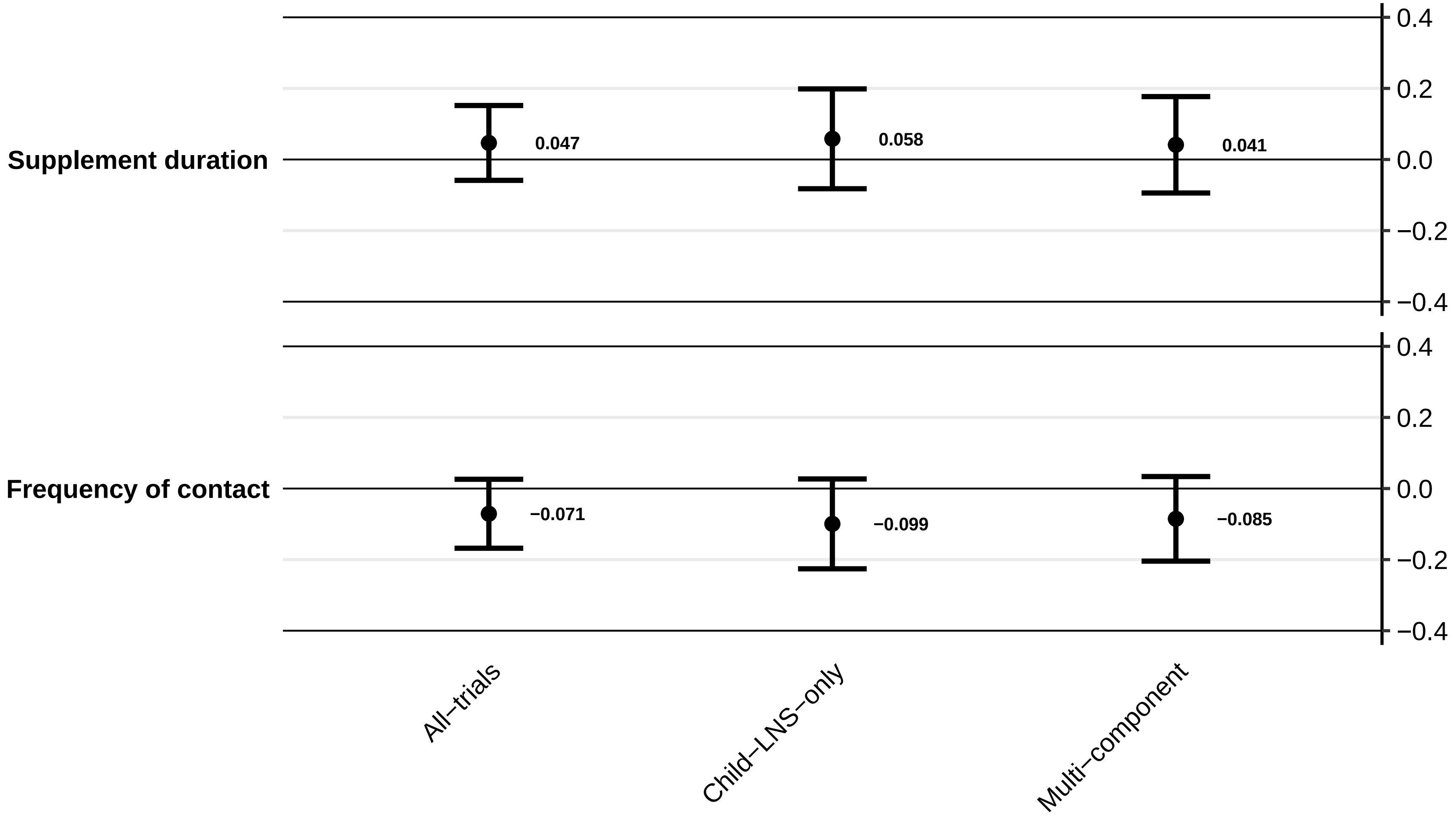

#### Supplemental figure 5N: Ratio of geometric mean ratios of zinc protoporphyrin concentration

##### 5N1: By study context effect modifiers (insufficient comparisons)

**Supplemental figure 5N: Ratio of geometric mean ratios of zinc protoporphyrin concentration**

#### 5N2: By study design effect modifiers (insufficient comparisons)

#### Supplemental figure 5O: Ratio of elevated zinc protoporphyrin prevalence ratios

##### 5O1: By study context effect modifiers (insufficient comparisons)

**Supplemental figure 5O: Ratio of elevated zinc protoporphyrin prevalence ratios**

#### 5O2: By study design effect modifiers (insufficient comparisons)

#### Supplemental figure 5P: Difference in elevated zinc protoporphyrin prevalence differences

##### 5P1: By study context effect modifiers (insufficient comparisons)

**Supplemental figure 5P: Difference in elevated zinc protoporphyrin prevalence differences**

#### 5P2: By study design effect modifiers (insufficient comparisons)

#### Supplemental figure 5Q: Ratio of geometric mean ratios of plasma zinc concentration

##### 5Q1: By study context effect modifiers (insufficient comparisons)

**Supplemental figure 5Q: Ratio of geometric mean ratios of plasma zinc concentration**

#### 5Q2: By study design effect modifiers (insufficient comparisons)

#### Supplemental figure 5R: Ratio of geometric mean ratios of retinol concentration

##### 5R1: By study context effect modifiers (insufficient comparisons)

**Supplemental figure 5R: Ratio of geometric mean ratios of retinol concentration**

#### 5R2: By study design effect modifiers (insufficient comparisons)

#### Supplemental figure 5S: Ratio of low vitamin A (retinol < 0.70 µmol/L) prevalence ratios

##### 5S1: By study context effect modifiers (insufficient comparisons)

**Supplemental figure 5S: Ratio of low vitamin A (retinol < 0.70 µmol/L) prevalence ratios**

#### 5S2: By study design effect modifiers (insufficient comparisons)

#### Supplemental figure 5T: Difference in low vitamin A (retinol < 0.70 µmol/L) prevalence differences

##### 5T1: By study context effect modifiers (insufficient comparisons)

**Supplemental figure 5T: Difference in low vitamin A (retinol < 0.70 µmol/L) prevalence differences**

#### 5T2: By study design effect modifiers (insufficient comparisons)

#### Supplemental figure 5U: Ratio of marginal vitamin A (retinol < 1.05 µmol/L) prevalence ratios

##### 5U1: By study context effect modifiers (insufficient comparisons)

**Supplemental figure 5U: Ratio of marginal vitamin A (retinol < 1.05 µmol/L) prevalence ratios**

#### 5U2: By study design effect modifiers (insufficient comparisons)

#### Supplemental figure 5V: Difference in marginal vitamin A (retinol < 1.05 µmol/L) prevalence differences

##### 5V1: By study context effect modifiers (insufficient comparisons)

**Supplemental figure 5V: Difference in marginal vitamin A (retinol < 1.05 µmol/L) prevalence differences**

#### 5V2: By study design effect modifiers (insufficient comparisons)

#### Supplemental figure 5W: Ratio of geometric mean ratio of retinol binding protein concentrations

##### 5W1: By study context effect modifiers (insufficient comparisons)

**Supplemental figure 5W: Ratio of geometric mean ratio of retinol binding protein concentrations**

#### 5W2: By study design effect modifiers (insufficient comparisons)

#### Supplemental figure 5X: Ratio of Low vitamin A status (RBP < 0.70 µmol/L) prevalence ratios

##### 5X1: By study context effect modifiers (insufficient comparisons)

**Supplemental figure 5X: Ratio of Low vitamin A status (RBP < 0.70 µmol/L) prevalence ratios**

#### 5X2: By study design effect modifiers (insufficient comparisons)

#### Supplemental figure 5Y: Difference in low vitamin A status (RBP < 0.70 µmol/L) prevalence differences

##### 5Y1: By study context effect modifiers (insufficient comparisons)

**Supplemental figure 5Y: Difference in low vitamin A status (RBP $< 0.70$ µmol/L) prevalence differences**

#### 5Y2: By study design effect modifiers (insufficient comparisons)

#### Supplemental figure 5Z: Ratio of marginal vitamin A status (RBP < 1.05 µmol/L) prevalence ratios

##### 5Z1: By study context effect modifiers (insufficient comparisons)

**Supplemental figure 5Z: Ratio of marginal vitamin A status (RBP < 1.05 µmol/L) prevalence ratios**

#### 5Z2: By study design effect modifiers (insufficient comparisons)

#### Supplemental figure 5AA: Difference in marginal vitamin A status (RBP < 1.05 µmol/L) prevalence differences

##### 5AA1: By study context effect modifiers (insufficient comparisons)

**Supplemental figure 5AA: Difference in marginal vitamin A status (RBP < 1.05 µmol/L) prevalence differences**

#### 5AA2: By study design effect modifiers (insufficient comparisons)

### Supplemental figure 6: Forest plots for effects of SQ-LNS on biochemical outcomes stratified by individual-level maternal and child effect modifiers

Supplemental figure 6: Forest plots for effects of SQ-LNS on biochemical outcomes stratified by individual-level maternal and child effect modifiers

These figures are forest plots showing the individual-level effect modification of intervention effects. Each figure has the estimates of intervention effect stratified within study by individual-level effect modifier category. For continuous outcomes the intervention effect is measured by the difference in mean of the LNS group minus control. For log transformed continuous outcomes, the intervention effect is measured by the ratio of geometric means, the effect estimate is the geometric mean in the LNS group divided by the geometric mean in the control group. For dichotomous outcomes analyzed via prevalence ratios, the effect estimate is the prevalence in the LNS group divided by the prevalence in the control group. For dichotomous outcomes analyzed via prevalence differences, the effect estimate is the prevalence in the LNS group minus the prevalence in the control group. The labels on the far left correspond to trial level information. In the middle left and on the right the values indicate the study level effect estimate, confidence interval, and weighting for deriving the pooled estimates is shown by subgroup.
Not all trials were included in all individual-level effect modification analyses, either because they did not measure the biomarker outcome or the effect modifier of interest (e.g., baseline anemia or acute malnutrition, receipt of high-dose vitamin A supplement), or because the prevalence of the binary outcome or proportion of children within one of the effect modifier subgroups was too low to allow us to generate effect estimates.
RBP, retinol binding protein.

### Supplemental figure 6A: Mean difference in hemoglobin concentration

#### 6A1: Stratified by Maternal BMI

**P−for−interaction = 0.320**
**Difference in MDs = −0.49 (−1.46, 0.48)**

| | | At least 20 kg/m² | | | | | | Less than 20 kg/m² | | | | | |
|---|---|---|---|---|---|---|---|---|---|---|---|---|---|
| Country | Trial | LNS N | Control N | Control Mean | MD (95% CI) | Fixed W | Random W | LNS N | Control N | Control Mean | MD (95% CI) | Fixed W | Random W |
| Bangladesh | JiVitA−4 (35) | | | | | | | | | | | | |
| Bangladesh | RDNS (36) | 227 | 102 | 111.1 | 5.00 (1.85, 8.15) | 0.03 | 0.08 | 300 | 160 | 113.1 | 3.69 (0.90, 6.49) | 0.11 | 0.11 |
| Bangladesh | WASH−B (37) | 102 | 102 | 118.0 | 4.54 (2.27, 6.81) | 0.07 | 0.10 | 132 | 84 | 118.3 | 1.91 (−0.70, 4.51) | 0.13 | 0.11 |
| Burkina Faso | iLiNS−Zinc (38) | 1217 | 390 | 88.5 | 8.29 (5.51, 11.07) | 0.04 | 0.09 | 738 | 274 | 88.7 | 8.73 (5.13, 12.33) | 0.07 | 0.09 |
| Burkina Faso | PROMIS CS (39) | 335 | 349 | 102.4 | 1.13 (−0.92, 3.19) | 0.08 | 0.10 | 239 | 227 | 102.7 | 2.79 (−0.12, 5.69) | 0.11 | 0.10 |
| Ghana | GHANA (40) | | | | | | | | | | | | |
| Ghana | iLiNS−DYADG (41) | 288 | 536 | 112.4 | 0.99 (−0.49, 2.48) | 0.15 | 0.11 | 35 | 114 | 110.2 | 2.33 (−1.69, 6.35) | 0.05 | 0.08 |
| Kenya | WASH−B (42) | 266 | 227 | 109.3 | 4.70 (2.53, 6.88) | 0.07 | 0.10 | 72 | 60 | 113.0 | 0.17 (−3.87, 4.21) | 0.05 | 0.08 |
| Madagascar | MAHAY (43) | | | | | | | | | | | | |
| Malawi | iLiNS−DYADM (44) | 124 | 258 | 107.8 | 1.99 (−1.35, 5.33) | 0.03 | 0.07 | 85 | 171 | 108.0 | 0.42 (−3.59, 4.42) | 0.06 | 0.08 |
| Malawi | iLiNS−DOSE (45) | 179 | 52 | 100.3 | 6.04 (1.31, 10.78) | 0.02 | 0.05 | 61 | 29 | 100.6 | −0.01 (−6.19, 6.16) | 0.02 | 0.05 |
| Mali | PROMIS CS (46) | 702 | 685 | 95.9 | 6.19 (3.84, 8.54) | 0.06 | 0.09 | 246 | 279 | 95.4 | 6.43 (4.14, 8.73) | 0.17 | 0.12 |
| Zimbabwe | SHINE (HIV−) (47) | 1222 | 1161 | 114.3 | 2.26 (1.34, 3.18) | 0.40 | 0.12 | 213 | 220 | 114.8 | 1.10 (−1.04, 3.23) | 0.20 | 0.12 |
| Zimbabwe | SHINE (HIV+) (48) | 221 | 222 | 114.7 | 2.78 (−0.19, 5.76) | 0.04 | 0.08 | 58 | 40 | 114.4 | 3.48 (−1.98, 8.94) | 0.03 | 0.06 |
| | | **4883** | **4084** | | **$I^2$ = 0.76, $Tau^2$ = 3.84** | | | **2179** | **1658** | | **$I^2$ = 0.63, $Tau^2$ = 4.11** | | |
| **Fixed** | | | | | **2.97 (2.39, 3.55)** | | | | | | **3.13 (2.18, 4.07)** | | |
| **Random** | | | | | **3.79 (2.40, 5.17)** | | | | | | **3.01 (1.42, 4.61)** | | |

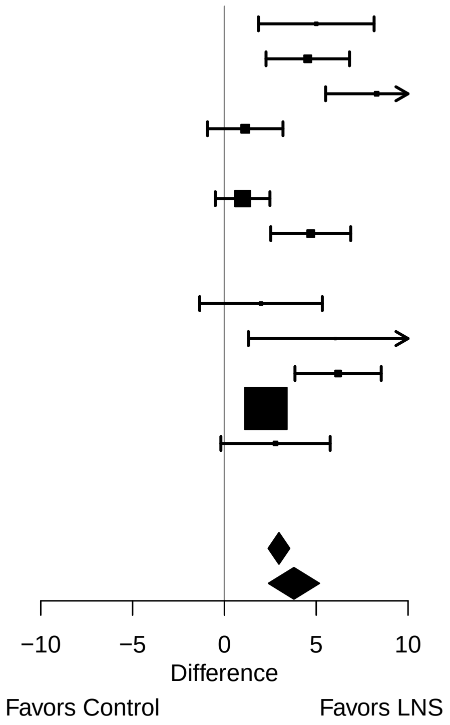

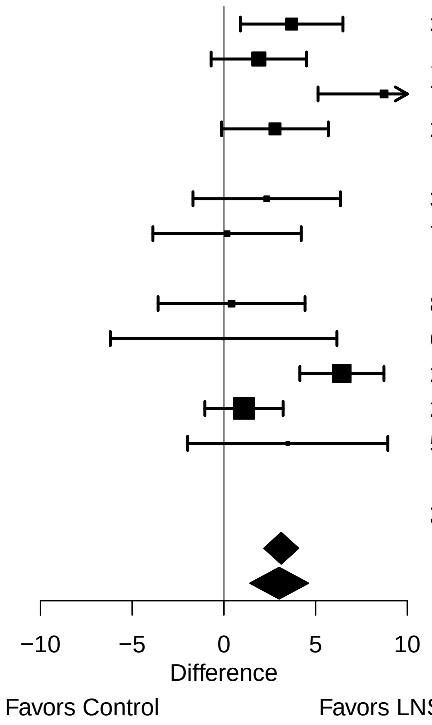

#### Supplemental figure 6A: Mean difference in hemoglobin concentration

#### 6A2: Stratified by Maternal age

**P−for−interaction = 0.015**
**Difference in MDs = −1.03 (−1.86, −0.20)**

| Country | Trial | At least 25 y: LNS N | Control N | Control Mean | MD (95% CI) | Fixed W | Random W | Less than 25 y: LNS N | Control N | Control Mean | MD (95% CI) | Fixed W | Random W |
|---|---|---|---|---|---|---|---|---|---|---|---|---|---|
| Bangladesh | JiVitA−4 (35) | 203 | 72 | 117.4 | 2.75 (0.13, 5.37) | 0.06 | 0.08 | 251 | 73 | 118.5 | 1.59 (−1.10, 4.28) | 0.07 | 0.08 |
| Bangladesh | RDNS (36) | 152 | 81 | 113.2 | 4.42 (0.28, 8.55) | 0.02 | 0.05 | 397 | 191 | 112.1 | 4.01 (1.98, 6.05) | 0.11 | 0.10 |
| Bangladesh | WASH−B (37) | 89 | 83 | 117.9 | 3.48 (1.50, 5.46) | 0.10 | 0.09 | 144 | 103 | 118.3 | 2.90 (0.33, 5.46) | 0.07 | 0.08 |
| Burkina Faso | iLiNS−Zinc (38) | 1132 | 399 | 88.4 | 9.30 (5.99, 12.60) | 0.03 | 0.07 | 812 | 262 | 88.9 | 7.22 (3.66, 10.78) | 0.04 | 0.06 |
| Burkina Faso | PROMIS CS (39) | 271 | 263 | 103.5 | 2.24 (−0.19, 4.67) | 0.06 | 0.08 | 303 | 318 | 101.9 | 1.28 (−1.17, 3.74) | 0.08 | 0.09 |
| Ghana | GHANA (40) | 69 | 63 | 106.8 | 9.03 (4.63, 13.44) | 0.02 | 0.05 | 29 | 25 | 101.0 | 10.43 (2.08, 18.78) | 0.01 | 0.01 |
| Ghana | iLiNS−DYADG (41) | 206 | 397 | 112.7 | 1.46 (−0.23, 3.16) | 0.13 | 0.09 | 122 | 264 | 110.8 | 0.71 (−1.61, 3.03) | 0.09 | 0.09 |
| Kenya | WASH−B (42) | 213 | 168 | 110.7 | 3.54 (1.20, 5.87) | 0.07 | 0.08 | 133 | 131 | 108.7 | 4.21 (1.02, 7.39) | 0.05 | 0.07 |
| Madagascar | MAHAY (43) | 301 | 329 | 104.6 | 2.38 (−1.14, 5.90) | 0.03 | 0.06 | 299 | 259 | 103.2 | 3.34 (0.16, 6.52) | 0.05 | 0.07 |
| Malawi | iLiNS−DYADM (44) | 99 | 220 | 109.5 | 0.07 (−3.51, 3.64) | 0.03 | 0.06 | 111 | 212 | 106.6 | 2.27 (−1.40, 5.94) | 0.04 | 0.06 |
| Malawi | iLiNS−DOSE (45) | 135 | 50 | 100.8 | 3.58 (−1.36, 8.52) | 0.02 | 0.04 | 104 | 28 | 100.1 | 4.62 (−1.55, 10.78) | 0.01 | 0.02 |
| Mali | PROMIS CS (46) | 502 | 508 | 95.0 | 7.55 (5.33, 9.78) | 0.08 | 0.08 | 451 | 462 | 96.5 | 4.81 (2.39, 7.22) | 0.08 | 0.09 |
| Zimbabwe | SHINE (HIV−) (47) | 808 | 726 | 114.8 | 2.60 (1.50, 3.70) | 0.31 | 0.10 | 697 | 694 | 113.7 | 1.45 (0.18, 2.73) | 0.29 | 0.13 |
| Zimbabwe | SHINE (HIV+) (48) | 221 | 204 | 115.0 | 3.33 (0.25, 6.41) | 0.04 | 0.07 | 66 | 60 | 114.2 | 1.29 (−3.02, 5.60) | 0.03 | 0.04 |
| | | **4401** | **3563** | | **I² = 0.70, Tau² = 4.76** | | | **3919** | **3082** | | **I² = 0.46, Tau² = 1.75** | | |
| **Fixed** | | | | | **3.33 (2.71, 3.94)** | | | | | | **2.61 (1.92, 3.29)** | | |
| **Random** | | | | | **3.83 (2.44, 5.21)** | | | | | | **2.95 (1.89, 4.01)** | | |

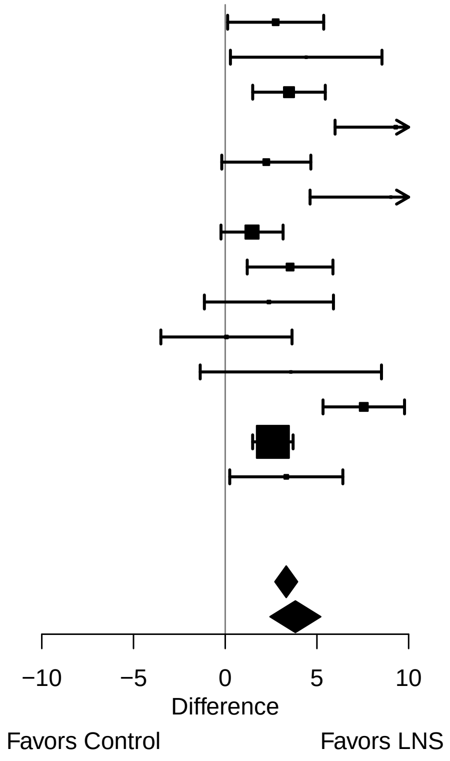

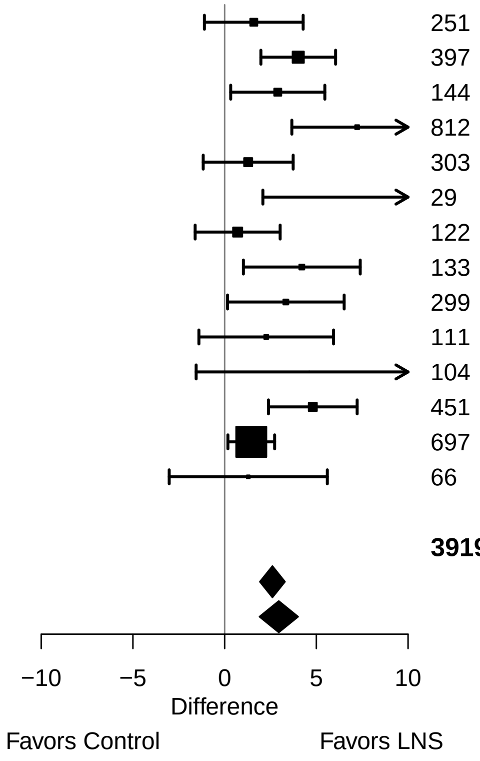

#### Supplemental figure 6A: Mean difference in hemoglobin concentration

##### 6A3: Stratified by Maternal education

**P−for−interaction = 0.212**
**Difference in MDs = −0.80 (−2.05, 0.45)**

| | | Primary or greater | | | | | | Incomplete or no formal | | | | | |
|---|---|---|---|---|---|---|---|---|---|---|---|---|---|
| Country | Trial | LNS N | Control N | Control Mean | MD (95% CI) | Fixed W | Random W | LNS N | Control N | Control Mean | MD (95% CI) | Fixed W | Random W |
| Bangladesh | JiVitA−4 (35) | 299 | 93 | 117.4 | 3.22 (0.81, 5.62) | 0.06 | 0.09 | 157 | 53 | 118.9 | 0.43 (−2.39, 3.24) | 0.09 | 0.09 |
| Bangladesh | RDNS (36) | 405 | 195 | 112.8 | 4.17 (1.54, 6.81) | 0.05 | 0.09 | 144 | 77 | 111.5 | 3.79 (−0.76, 8.34) | 0.03 | 0.06 |
| Bangladesh | WASH−B (37) | 163 | 145 | 118.5 | 2.92 (0.80, 5.03) | 0.08 | 0.10 | 71 | 41 | 116.7 | 4.02 (1.01, 7.03) | 0.07 | 0.08 |
| Burkina Faso | iLiNS−Zinc (38) | 82 | 16 | 86.0 | 11.27 (3.53, 19.01) | 0.01 | 0.03 | 1862 | 645 | 88.6 | 8.41 (5.40, 11.43) | 0.07 | 0.08 |
| Burkina Faso | PROMIS CS (39) | 46 | 47 | 103.1 | 0.39 (−4.37, 5.16) | 0.02 | 0.05 | 526 | 532 | 102.6 | 1.84 (−0.12, 3.80) | 0.18 | 0.10 |
| Ghana | GHANA (40) | 91 | 75 | 105.8 | 8.93 (4.80, 13.06) | 0.02 | 0.06 | 7 | 15 | 102.0 | 9.57 (−6.01, 25.15) | 0.00 | 0.01 |
| Ghana | iLiNS−DYADG (41) | 253 | 526 | 111.7 | 1.38 (−0.16, 2.92) | 0.14 | 0.11 | 75 | 135 | 112.8 | 0.65 (−2.42, 3.72) | 0.07 | 0.08 |
| Kenya | WASH−B (42) | 168 | 136 | 108.4 | 7.46 (4.92, 10.01) | 0.05 | 0.09 | 181 | 164 | 110.9 | 0.79 (−1.73, 3.30) | 0.11 | 0.09 |
| Madagascar | MAHAY (43) | 128 | 143 | 103.5 | 3.17 (−1.42, 7.75) | 0.02 | 0.06 | 472 | 445 | 104.2 | 2.64 (−0.58, 5.87) | 0.07 | 0.08 |
| Malawi | iLiNS−DYADM (44) | 34 | 66 | 109.6 | 4.51 (−1.73, 10.75) | 0.01 | 0.04 | 175 | 363 | 107.6 | 0.65 (−2.16, 3.47) | 0.09 | 0.09 |
| Malawi | iLiNS−DOSE (45) | 49 | 17 | 97.4 | 11.72 (2.32, 21.13) | 0.00 | 0.02 | 190 | 61 | 101.4 | 1.78 (−2.35, 5.92) | 0.04 | 0.06 |
| Mali | PROMIS CS (46) | 107 | 96 | 97.5 | 4.27 (0.60, 7.94) | 0.03 | 0.07 | 845 | 874 | 95.6 | 6.46 (4.25, 8.66) | 0.14 | 0.10 |
| Zimbabwe | SHINE (HIV−) (47) | 1536 | 1462 | 114.4 | 1.88 (1.04, 2.73) | 0.48 | 0.11 | 58 | 53 | 112.3 | 7.17 (2.94, 11.39) | 0.04 | 0.06 |
| Zimbabwe | SHINE (HIV+) (48) | 274 | 252 | 114.8 | 2.81 (0.13, 5.49) | 0.05 | 0.09 | 18 | 14 | 112.6 | 6.65 (−4.28, 17.58) | 0.01 | 0.02 |
| | | **3635** | **3269** | | **I² = 0.68, Tau² = 4.95** | | | **4781** | **3472** | | **I² = 0.68, Tau² = 4.63** | | |
| **Fixed** | | | | | **2.73 (2.15, 3.32)** | | | | | | **3.08 (2.26, 3.91)** | | |
| **Random** | | | | | **4.00 (2.50, 5.50)** | | | | | | **3.25 (1.76, 4.74)** | | |

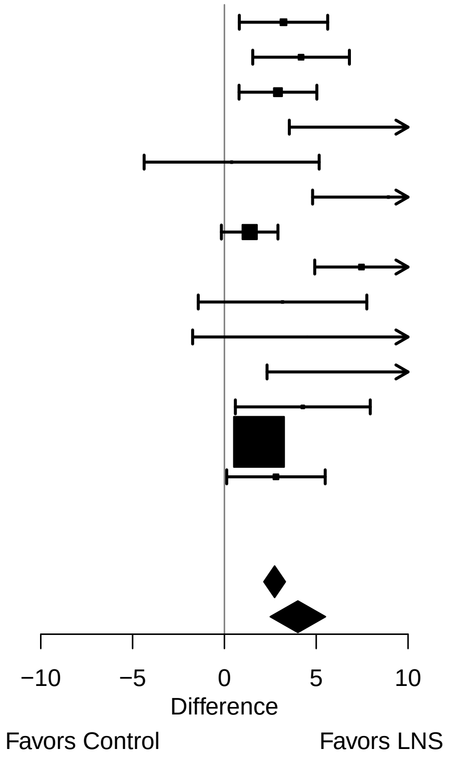

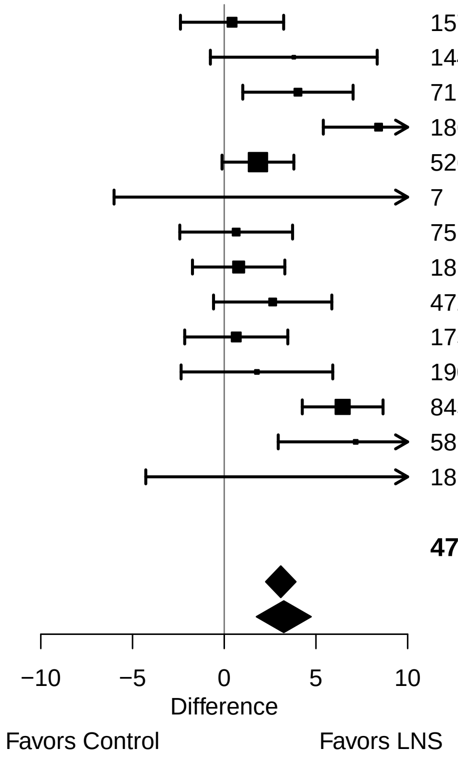

#### Supplemental figure 6A: Mean difference in hemoglobin concentration

##### 6A4: Stratified by Child sex

**P−for−interaction = 0.449**
**Difference in MDs = −0.30 (−1.08, 0.48)**

| | | Male | | | | | | Female | | | | | |
|---|---|---|---|---|---|---|---|---|---|---|---|---|---|
| | | **LNS** | **Control** | **Control** | **MD** | **Fixed** | **Random** | **LNS** | **Control** | **Control** | **MD** | **Fixed** | **Random** |
| Country | Trial | N | N | Mean | (95% CI) | W | W | N | N | Mean | (95% CI) | W | W |
| Bangladesh | JiVitA−4 (35) | 235 | 71 | 116.9 | 4.05 (1.55, 6.55) | 0.07 | 0.08 | 222 | 75 | 119.0 | 0.37 (−1.81, 2.55) | 0.08 | 0.09 |
| Bangladesh | RDNS (36) | 266 | 131 | 111.0 | 5.50 (2.06, 8.93) | 0.04 | 0.07 | 281 | 141 | 113.7 | 2.90 (0.13, 5.66) | 0.05 | 0.07 |
| Bangladesh | WASH−B (37) | 116 | 89 | 118.2 | 3.70 (1.12, 6.27) | 0.06 | 0.08 | 118 | 97 | 118.1 | 2.50 (0.10, 4.90) | 0.06 | 0.08 |
| Burkina Faso | iLiNS−Zinc (38) | 993 | 334 | 87.3 | 9.05 (6.04, 12.05) | 0.05 | 0.07 | 964 | 330 | 89.8 | 7.89 (4.36, 11.41) | 0.03 | 0.05 |
| Burkina Faso | PROMIS CS (39) | 317 | 295 | 101.1 | 1.63 (−0.56, 3.82) | 0.09 | 0.08 | 257 | 286 | 104.2 | 2.24 (0.10, 4.39) | 0.08 | 0.09 |
| Ghana | GHANA (40) | 60 | 41 | 104.5 | 11.05 (5.41, 16.68) | 0.01 | 0.04 | 38 | 55 | 106.6 | 6.36 (0.56, 12.17) | 0.01 | 0.03 |
| Ghana | iLiNS−DYADG (41) | 164 | 310 | 111.7 | 0.33 (−1.67, 2.32) | 0.10 | 0.09 | 163 | 348 | 112.2 | 2.18 (0.29, 4.08) | 0.10 | 0.10 |
| Kenya | WASH−B (42) | 168 | 145 | 110.3 | 3.13 (0.34, 5.92) | 0.05 | 0.07 | 182 | 155 | 109.4 | 4.70 (2.69, 6.70) | 0.09 | 0.09 |
| Madagascar | MAHAY (43) | 277 | 288 | 102.4 | 3.37 (−0.23, 6.96) | 0.03 | 0.06 | 323 | 300 | 105.5 | 2.10 (−1.06, 5.27) | 0.04 | 0.06 |
| Malawi | iLiNS−DYADM (44) | 103 | 202 | 106.6 | 0.87 (−2.84, 4.59) | 0.03 | 0.06 | 107 | 230 | 109.3 | 1.48 (−2.05, 5.01) | 0.03 | 0.05 |
| Malawi | iLiNS−DOSE (45) | 123 | 43 | 98.0 | 4.62 (−0.50, 9.74) | 0.02 | 0.05 | 120 | 39 | 103.6 | 2.79 (−2.65, 8.23) | 0.01 | 0.03 |
| Mali | PROMIS CS (46) | 479 | 526 | 94.6 | 6.32 (4.06, 8.58) | 0.08 | 0.08 | 474 | 444 | 97.1 | 5.98 (3.33, 8.63) | 0.05 | 0.07 |
| Zimbabwe | SHINE (HIV−) (47) | 830 | 814 | 113.9 | 1.77 (0.65, 2.89) | 0.33 | 0.10 | 852 | 780 | 114.6 | 2.46 (1.39, 3.53) | 0.32 | 0.12 |
| Zimbabwe | SHINE (HIV+) (48) | 146 | 149 | 112.8 | 3.83 (0.70, 6.97) | 0.04 | 0.07 | 160 | 136 | 116.9 | 1.46 (−1.38, 4.29) | 0.05 | 0.07 |
| | | **4277** | **3438** | | **I² = 0.74, Tau² = 5.44** | | | **4261** | **3416** | | **I² = 0.50, Tau² = 1.94** | | |
| **Fixed** | | | | | **3.07 (2.42, 3.71)** | | | | | | **2.79 (2.18, 3.39)** | | |
| **Random** | | | | | **3.91 (2.44, 5.37)** | | | | | | **2.99 (1.96, 4.02)** | | |

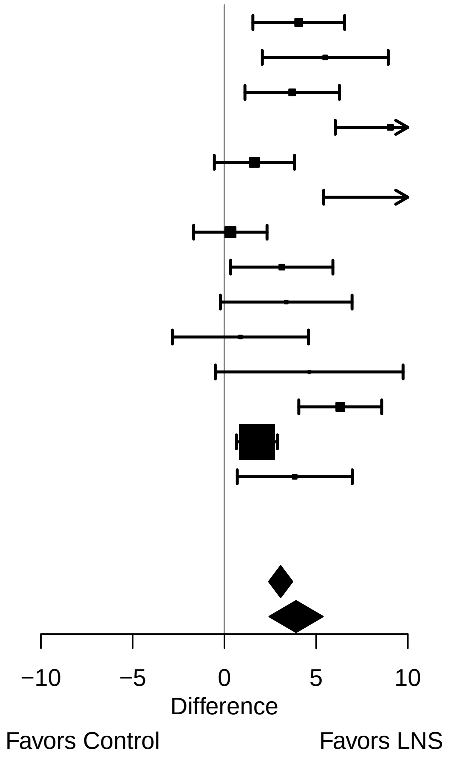

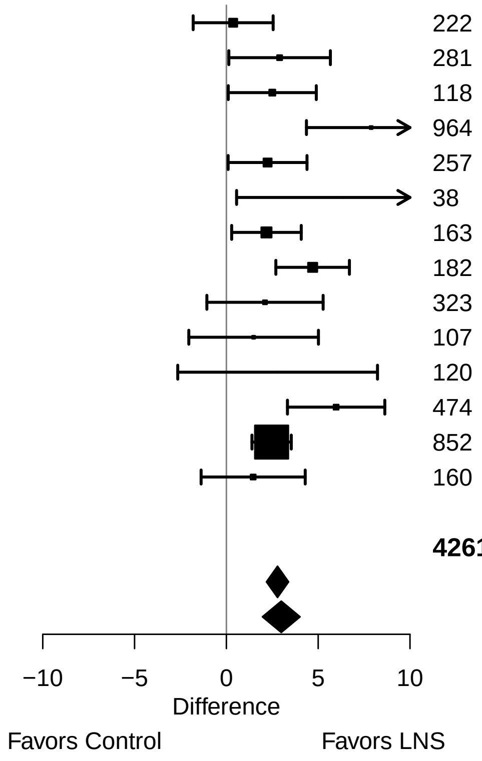

#### Supplemental figure 6A: Mean difference in hemoglobin concentration

#### 6A5: Stratified by Child birth order

**P−for−interaction = 0.003**
**Difference in MDs = −1.46 (−2.42, −0.50)**

| | | Later born | | | | | | Firstborn | | | | | |
|---|---|---|---|---|---|---|---|---|---|---|---|---|---|
| Country | Trial | LNS N | Control N | Control Mean | MD (95% CI) | Fixed W | Random W | LNS N | Control N | Control Mean | MD (95% CI) | Fixed W | Random W |
| Bangladesh | JiVitA−4 (35) | 115 | 33 | 115.1 | 4.41 (0.47, 8.34) | 0.02 | 0.05 | 341 | 112 | 118.8 | 1.62 (−0.39, 3.63) | 0.16 | 0.10 |
| Bangladesh | RDNS (36) | 339 | 171 | 112.6 | 3.88 (1.15, 6.62) | 0.04 | 0.07 | 210 | 101 | 112.1 | 4.46 (1.64, 7.29) | 0.08 | 0.09 |
| Bangladesh | WASH−B (37) | 137 | 123 | 117.7 | 3.01 (1.07, 4.94) | 0.08 | 0.09 | 95 | 61 | 118.7 | 3.11 (−0.54, 6.77) | 0.05 | 0.08 |
| Burkina Faso | iLiNS−Zinc (38) | 1522 | 520 | 88.7 | 8.30 (4.85, 11.75) | 0.02 | 0.05 | 435 | 144 | 88.0 | 9.03 (6.18, 11.87) | 0.08 | 0.09 |
| Burkina Faso | PROMIS CS (39) | 498 | 477 | 102.4 | 2.26 (0.28, 4.25) | 0.07 | 0.09 | 75 | 103 | 103.6 | −1.21 (−6.02, 3.59) | 0.03 | 0.06 |
| Ghana | GHANA (40) | 60 | 54 | 106.3 | 9.73 (4.70, 14.77) | 0.01 | 0.03 | 38 | 38 | 104.4 | 7.72 (1.26, 14.19) | 0.02 | 0.04 |
| Ghana | iLiNS−DYADG (41) | 220 | 435 | 111.9 | 2.35 (0.69, 4.01) | 0.11 | 0.10 | 108 | 226 | 112.1 | −1.03 (−3.47, 1.42) | 0.11 | 0.09 |
| Kenya | WASH−B (42) | 282 | 244 | 110.1 | 3.73 (1.59, 5.87) | 0.06 | 0.08 | 67 | 56 | 108.4 | 4.82 (−0.22, 9.86) | 0.03 | 0.06 |
| Madagascar | MAHAY (43) | 458 | 454 | 103.7 | 2.84 (−0.35, 6.02) | 0.03 | 0.06 | 141 | 131 | 105.0 | 2.43 (−1.24, 6.10) | 0.05 | 0.08 |
| Malawi | iLiNS−DYADM (44) | 165 | 342 | 108.5 | 1.61 (−1.21, 4.44) | 0.04 | 0.07 | 45 | 88 | 105.9 | −0.25 (−6.22, 5.71) | 0.02 | 0.05 |
| Malawi | iLiNS−DOSE (45) | 148 | 61 | 99.7 | 3.54 (−0.95, 8.02) | 0.01 | 0.04 | 56 | 10 | 108.2 | 0.99 (−10.07, 12.04) | 0.01 | 0.02 |
| Mali | PROMIS CS (46) | 790 | 832 | 95.7 | 6.39 (4.23, 8.56) | 0.06 | 0.08 | 146 | 126 | 95.7 | 5.42 (2.11, 8.72) | 0.06 | 0.08 |
| Zimbabwe | SHINE (HIV−) (47) | 1188 | 1100 | 114.2 | 2.70 (1.82, 3.57) | 0.39 | 0.12 | 418 | 416 | 114.4 | 0.67 (−0.89, 2.22) | 0.27 | 0.11 |
| Zimbabwe | SHINE (HIV+) (48) | 252 | 216 | 114.7 | 3.15 (0.63, 5.67) | 0.05 | 0.07 | 51 | 64 | 115.1 | 0.56 (−4.11, 5.24) | 0.03 | 0.06 |
| | | **6174** | **5062** | | **I² = 0.54, Tau² = 2.36** | | | **2226** | **1676** | | **I² = 0.72, Tau² = 5.98** | | |
| **Fixed** | | | | | **3.25 (2.71, 3.79)** | | | | | | **2.28 (1.47, 3.10)** | | |
| **Random** | | | | | **3.75 (2.68, 4.82)** | | | | | | **2.76 (1.10, 4.42)** | | |

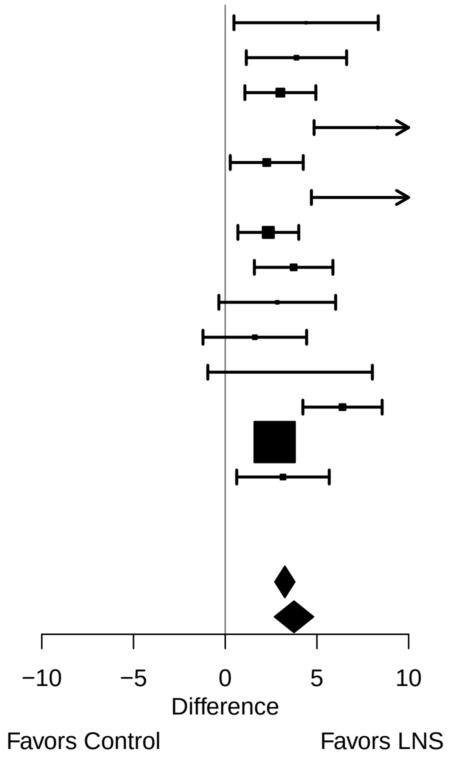

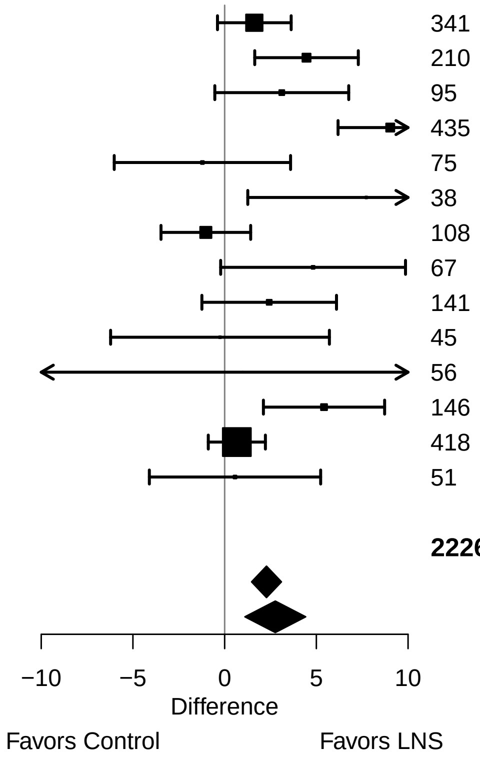

**Supplemental figure 6A: Mean difference in hemoglobin concentration**

#### 6A6: Stratified by Child baseline acute malnutrition

**P−for−interaction = 0.859**

**Difference in MDs = 0.15 (−1.54, 1.84)**

| | | No | | | | | | Yes | | | | | |
|---|---|---|---|---|---|---|---|---|---|---|---|---|---|
| Country | Trial | LNS N | Control N | Control Mean | MD (95% CI) | Fixed W | Random W | LNS N | Control N | Control Mean | MD (95% CI) | Fixed W | Random W |
| Bangladesh | JiVitA−4 (35) | 365 | 114 | 117.5 | 2.77 (0.93, 4.60) | 0.11 | 0.14 | 90 | 32 | 119.5 | 0.10 (−3.25, 3.46) | 0.31 | 0.18 |
| Bangladesh | RDNS (36) | 493 | 243 | 113.0 | 3.77 (1.44, 6.11) | 0.07 | 0.12 | 35 | 12 | 108.8 | 6.00 (−1.95, 13.94) | 0.05 | 0.10 |
| Bangladesh | WASH−B (37) | | | | | | | | | | | | |
| Burkina Faso | iLiNS−Zinc (38) | 1457 | 487 | 89.2 | 8.10 (5.54, 10.65) | 0.06 | 0.12 | 500 | 177 | 86.7 | 9.42 (4.35, 14.49) | 0.13 | 0.15 |
| Burkina Faso | PROMIS CS (39) | | | | | | | | | | | | |
| Ghana | GHANA (40) | | | | | | | | | | | | |
| Ghana | iLiNS−DYADG (41) | 285 | 556 | 111.5 | 1.51 (0.05, 2.98) | 0.18 | 0.15 | 21 | 54 | 115.4 | −1.97 (−7.31, 3.36) | 0.12 | 0.14 |
| Kenya | WASH−B (42) | | | | | | | | | | | | |
| Madagascar | MAHAY (43) | | | | | | | | | | | | |
| Malawi | iLiNS−DYADM (44) | 183 | 379 | 108.2 | 0.85 (−1.91, 3.61) | 0.05 | 0.11 | 14 | 37 | 106.4 | 2.31 (−6.74, 11.36) | 0.04 | 0.08 |
| Malawi | iLiNS−DOSE (45) | 230 | 75 | 100.5 | 4.29 (0.45, 8.14) | 0.03 | 0.08 | 13 | 7 | 101.9 | −3.34 (−20.72, 14.03) | 0.01 | 0.03 |
| Mali | PROMIS CS (46) | | | | | | | | | | | | |
| Zimbabwe | SHINE (HIV−) (47) | 1185 | 1069 | 114.9 | 1.60 (0.70, 2.51) | 0.46 | 0.16 | 66 | 67 | 114.7 | 3.54 (−0.63, 7.71) | 0.20 | 0.17 |
| Zimbabwe | SHINE (HIV+) (48) | 226 | 202 | 115.3 | 3.01 (0.05, 5.96) | 0.04 | 0.11 | 26 | 13 | 108.5 | 10.08 (5.03, 15.12) | 0.14 | 0.15 |
| | | **4424** | **3125** | | **I² = 0.74, Tau² = 3.69** | | | **765** | **399** | | **I² = 0.66, Tau² = 12.68** | | |
| **Fixed** | | | | | **2.34 (1.73, 2.96)** | | | | | | **3.50 (1.64, 5.35)** | | |
| **Random** | | | | | **3.08 (1.52, 4.65)** | | | | | | **3.88 (0.57, 7.20)** | | |

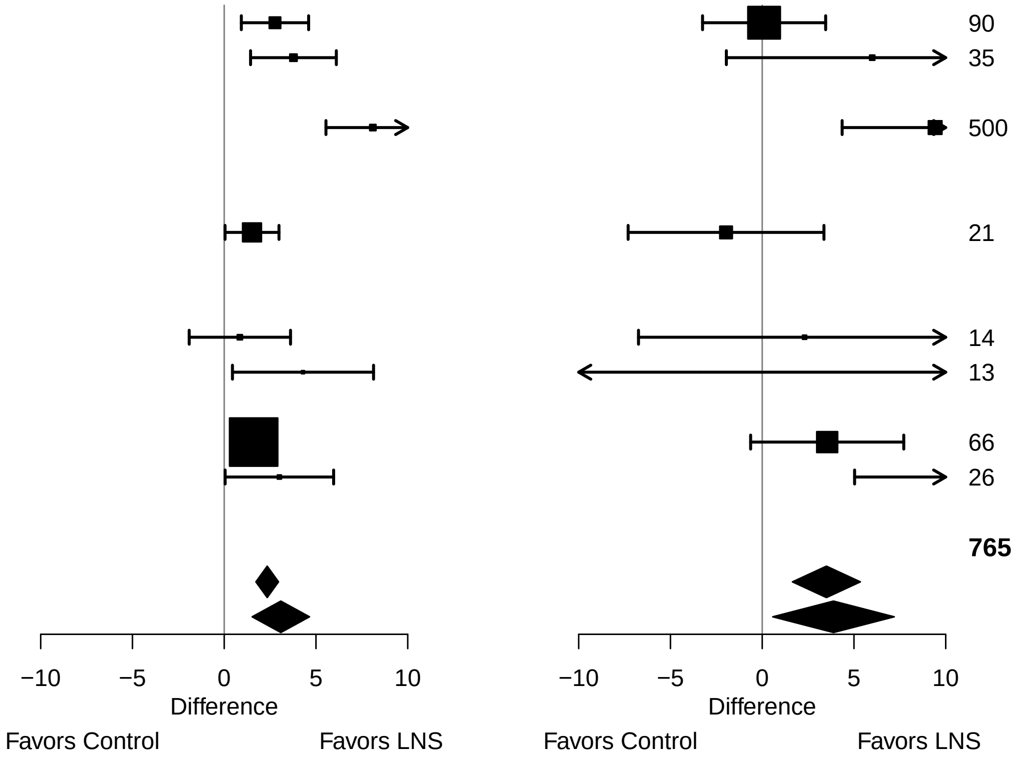

**Supplemental figure 6A: Mean difference in hemoglobin concentration**

#### 6A7: Stratified by Child baseline anemia

**P−for−interaction = 0.063**
**Difference in MDs = 1.84 (−0.10, 3.79)**

| Country | Trial | Not anemic: LNS N | Not anemic: Control N | Not anemic: Control Mean | Not anemic: MD (95% CI) | Not anemic: Fixed W | Not anemic: Random W | Anemic: LNS N | Anemic: Control N | Anemic: Control Mean | Anemic: MD (95% CI) | Anemic: Fixed W | Anemic: Random W |
|---|---|---|---|---|---|---|---|---|---|---|---|---|---|
| Bangladesh | JiVitA−4 (35) | | | | | | | | | | | | |
| Bangladesh | RDNS (36) | 213 | 93 | 118.7 | 2.31 (−1.04, 5.66) | 0.15 | 0.21 | 314 | 160 | 109.4 | 4.25 (1.53, 6.97) | 0.25 | 0.23 |
| Bangladesh | WASH−B (37) | | | | | | | | | | | | |
| Burkina Faso | iLiNS−Zinc (38) | 177 | 61 | 93.9 | 8.12 (4.37, 11.86) | 0.12 | 0.19 | 1780 | 603 | 88.0 | 8.53 (5.27, 11.79) | 0.18 | 0.20 |
| Burkina Faso | PROMIS CS (39) | | | | | | | | | | | | |
| Ghana | GHANA (40) | | | | | | | | | | | | |
| Ghana | iLiNS−DYADG (41) | 189 | 355 | 114.7 | 0.01 (−1.76, 1.78) | 0.55 | 0.26 | 90 | 207 | 107.1 | 2.85 (0.45, 5.25) | 0.33 | 0.24 |
| Kenya | WASH−B (42) | | | | | | | | | | | | |
| Madagascar | MAHAY (43) | | | | | | | | | | | | |
| Malawi | iLiNS−DYADM (44) | 66 | 145 | 112.9 | 0.22 (−4.11, 4.55) | 0.09 | 0.17 | 134 | 269 | 106.0 | 0.95 (−2.20, 4.10) | 0.19 | 0.21 |
| Malawi | iLiNS−DOSE (45) | 101 | 45 | 103.7 | 3.17 (−1.42, 7.76) | 0.08 | 0.17 | 142 | 37 | 96.9 | 3.98 (−1.99, 9.94) | 0.05 | 0.11 |
| Mali | PROMIS CS (46) | | | | | | | | | | | | |
| Zimbabwe | SHINE (HIV−) (47) | | | | | | | | | | | | |
| Zimbabwe | SHINE (HIV+) (48) | | | | | | | | | | | | |
| | | **746** | **699** | | **I² = 0.75, Tau² = 7.57** | | | **2460** | **1276** | | **I² = 0.66, Tau² = 5.13** | | |
| **Fixed** | | | | | **1.63 (0.32, 2.95)** | | | | | | **3.91 (2.54, 5.28)** | | |
| **Random** | | | | | **2.62 (−0.27, 5.50)** | | | | | | **4.06 (1.57, 6.56)** | | |

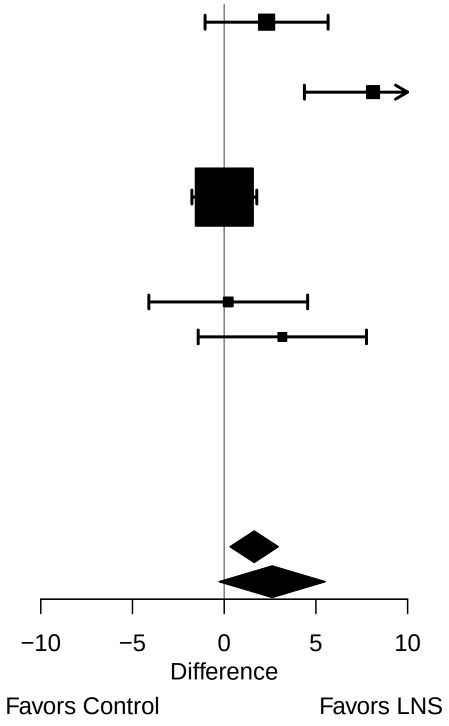

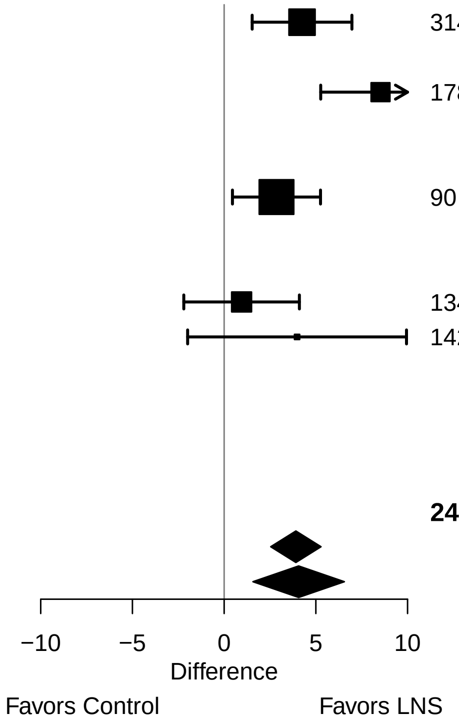

**Supplemental figure 6A: Mean difference in hemoglobin concentration**

#### 6A8: Stratified by Child high-dose vitamin A supplementation

**P−for−interaction = 0.808**
**Difference in MDs = 0.15 (−1.03, 1.33)**

| | | Received VitA | | | | | | Not treated | | | | | |
|---|---|---|---|---|---|---|---|---|---|---|---|---|---|
| | | **LNS** | **Control** | **Control** | **MD** | **Fixed** | **Random** | **LNS** | **Control** | **Control** | **MD** | **Fixed** | **Random** |
| Country | Trial | N | N | Mean | (95% CI) | W | W | N | N | Mean | (95% CI) | W | W |
| Bangladesh | JiVitA−4 (35) | 141 | 37 | 118.5 | 1.63 (−2.54, 5.81) | 0.03 | 0.09 | 133 | 41 | 118.8 | 1.17 (−1.56, 3.91) | 0.09 | 0.13 |
| Bangladesh | RDNS (36) | 444 | 223 | 112.8 | 3.91 (1.28, 6.54) | 0.09 | 0.14 | 105 | 49 | 110.5 | 5.04 (1.41, 8.68) | 0.05 | 0.10 |
| Bangladesh | WASH−B (37) | 20 | 18 | 117.3 | 3.57 (−1.97, 9.11) | 0.02 | 0.06 | 16 | 9 | 119.6 | −2.43 (−9.10, 4.23) | 0.02 | 0.04 |
| Burkina Faso | iLiNS−Zinc (38) | | | | | | | | | | | | |
| Burkina Faso | PROMIS CS (39) | | | | | | | | | | | | |
| Ghana | GHANA (40) | 50 | 62 | 105.6 | 10.67 (5.60, 15.74) | 0.02 | 0.07 | 48 | 29 | 104.7 | 8.05 (1.40, 14.70) | 0.02 | 0.04 |
| Ghana | iLiNS−DYADG (41) | 215 | 412 | 111.7 | 1.71 (−0.02, 3.44) | 0.20 | 0.19 | 113 | 249 | 112.4 | 0.40 (−1.88, 2.67) | 0.13 | 0.14 |
| Kenya | WASH−B (42) | 29 | 34 | 109.9 | 1.75 (−5.04, 8.54) | 0.01 | 0.04 | 263 | 212 | 109.7 | 4.24 (1.90, 6.58) | 0.12 | 0.14 |
| Madagascar | MAHAY (43) | | | | | | | | | | | | |
| Malawi | iLiNS−DYADM (44) | 75 | 136 | 106.6 | 2.65 (−1.96, 7.27) | 0.03 | 0.08 | 133 | 289 | 108.6 | 0.43 (−2.70, 3.56) | 0.07 | 0.11 |
| Malawi | iLiNS−DOSE (45) | | | | | | | | | | | | |
| Mali | PROMIS CS (46) | | | | | | | | | | | | |
| Zimbabwe | SHINE (HIV−) (47) | 874 | 833 | 114.7 | 1.71 (0.67, 2.74) | 0.55 | 0.22 | 665 | 622 | 114.0 | 2.57 (1.31, 3.83) | 0.43 | 0.19 |
| Zimbabwe | SHINE (HIV+) (48) | 165 | 127 | 115.8 | 2.35 (−1.09, 5.78) | 0.05 | 0.11 | 112 | 134 | 113.9 | 3.09 (−0.12, 6.29) | 0.07 | 0.11 |
| | | **2013** | **1882** | | **I² = 0.43, Tau² = 2.66** | | | **1588** | **1634** | | **I² = 0.46, Tau² = 2.70** | | |
| **Fixed** | | | | | **2.19 (1.42, 2.96)** | | | | | | **2.39 (1.56, 3.21)** | | |
| **Random** | | | | | **2.89 (1.32, 4.46)** | | | | | | **2.39 (0.89, 3.89)** | | |

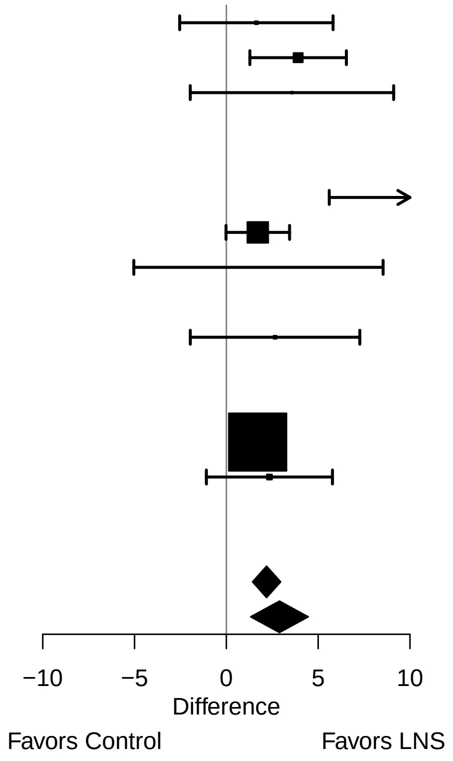

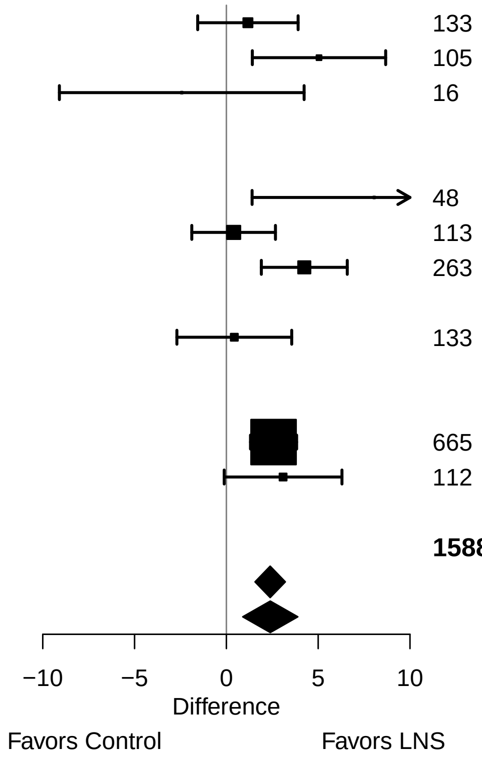

#### Supplemental figure 6A: Mean difference in hemoglobin concentration

##### 6A9: Stratified by Child inflammation

**P−for−interaction = 0.712**

**Difference in MDs = −0.25 (−1.59, 1.09)**

| Country | Trial | **Not inflamed** LNS N | Control N | Control Mean | MD (95% CI) | Fixed W | Random W | **High AGP or CRP** LNS N | Control N | Control Mean | MD (95% CI) | Fixed W | Random W |
|---|---|---|---|---|---|---|---|---|---|---|---|---|---|
| Bangladesh | JiVitA−4 (35) | 185 | 47 | 118.2 | 2.82 (0.23, 5.41) | 0.10 | 0.11 | 269 | 98 | 118.0 | 1.66 (−0.68, 4.01) | 0.18 | 0.18 |
| Bangladesh | RDNS (36) | 369 | 168 | 112.6 | 4.59 (1.81, 7.37) | 0.09 | 0.10 | 180 | 104 | 112.0 | 2.99 (0.20, 5.79) | 0.13 | 0.13 |
| Bangladesh | WASH−B (37) | 165 | 136 | 118.7 | 2.74 (0.53, 4.95) | 0.14 | 0.12 | 47 | 42 | 117.0 | 4.15 (1.57, 6.74) | 0.15 | 0.15 |
| Burkina Faso | iLiNS−Zinc (38) | 131 | 33 | 91.7 | 8.95 (1.27, 16.62) | 0.01 | 0.03 | 197 | 69 | 87.3 | 8.74 (4.22, 13.27) | 0.05 | 0.05 |
| Burkina Faso | PROMIS CS (39) | | | | | | | | | | | | |
| Ghana | GHANA (40) | 70 | 66 | 106.6 | 9.72 (5.38, 14.07) | 0.04 | 0.06 | 14 | 16 | 94.1 | 9.91 (−1.17, 20.99) | 0.01 | 0.01 |
| Ghana | iLiNS−DYADG (41) | 45 | 120 | 111.0 | 1.78 (−1.79, 5.36) | 0.05 | 0.08 | 55 | 82 | 109.4 | 2.13 (−1.29, 5.54) | 0.09 | 0.09 |
| Kenya | WASH−B (42) | 157 | 135 | 112.2 | 4.72 (2.12, 7.31) | 0.10 | 0.11 | 141 | 123 | 106.8 | 3.13 (−0.36, 6.63) | 0.08 | 0.08 |
| Madagascar | MAHAY (43) | 44 | 32 | 111.2 | 6.73 (1.94, 11.51) | 0.03 | 0.06 | 39 | 19 | 110.4 | 2.20 (−5.62, 10.01) | 0.02 | 0.02 |
| Malawi | iLiNS−DYADM (44) | 59 | 123 | 112.6 | 2.45 (−1.78, 6.68) | 0.04 | 0.07 | 138 | 269 | 105.8 | 1.87 (−1.27, 5.00) | 0.10 | 0.10 |
| Malawi | iLiNS−DOSE (45) | 20 | 6 | 105.2 | 7.34 (−2.36, 17.05) | 0.01 | 0.02 | 49 | 24 | 100.7 | 3.04 (−5.13, 11.21) | 0.02 | 0.02 |
| Mali | PROMIS CS (46) | | | | | | | | | | | | |
| Zimbabwe | SHINE (HIV−) (47) | 414 | 354 | 116.1 | 0.98 (−0.61, 2.56) | 0.27 | 0.14 | 105 | 99 | 112.0 | 2.50 (−0.09, 5.09) | 0.15 | 0.15 |
| Zimbabwe | SHINE (HIV+) (48) | 186 | 158 | 114.8 | 3.76 (1.39, 6.13) | 0.12 | 0.11 | 32 | 31 | 114.2 | 2.90 (−4.90, 10.70) | 0.02 | 0.02 |
| | | **1845** | **1378** | | **I² = 0.55, Tau² = 3.07** | | | **1266** | **976** | | **I² = 0.00, Tau² = 0.00** | | |
| **Fixed** | | | | | **3.17 (2.34, 3.99)** | | | | | | **3.00 (1.99, 4.00)** | | |
| **Random** | | | | | **3.86 (2.45, 5.27)** | | | | | | **3.00 (1.99, 4.00)** | | |

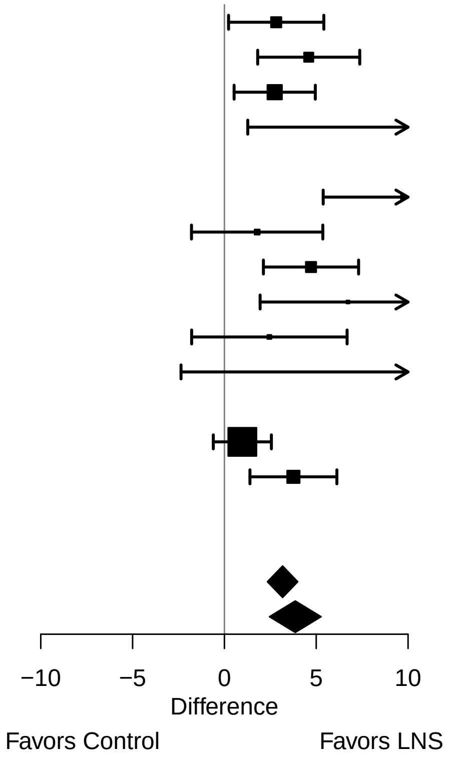

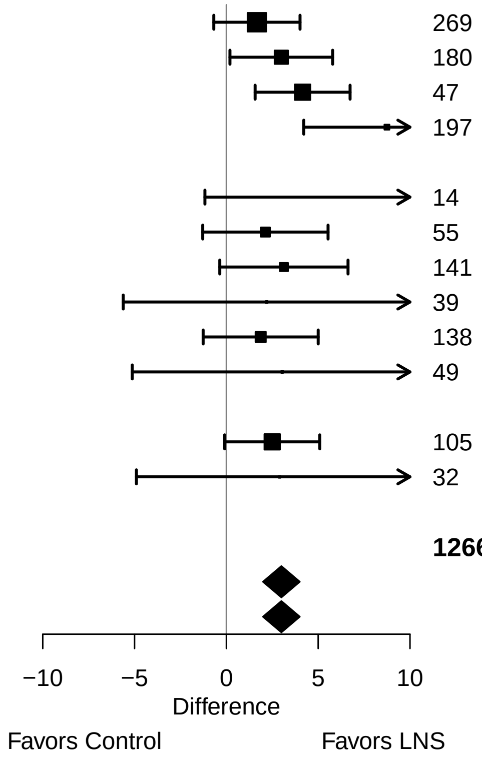

### Supplemental figure 6B: Anemia prevalence ratio

#### 6B1: Stratified by Maternal BMI

**P−for−interaction = 0.637**
**Ratio of PRs = 1.01 (0.97, 1.06)**

| | | At least 20 kg/m² | | | | | | Less than 20 kg/m² | | | | | |
|---|---|---|---|---|---|---|---|---|---|---|---|---|---|
| Country | Trial | LNS N | Control N | Control Prevalence | PR (95% CI) | Fixed W | Random W | LNS N | Control N | Control Prevalence | PR (95% CI) | Fixed W | Random W |
| Bangladesh | JiVitA−4 (35) | | | | | | | | | | | | |
| Bangladesh | RDNS (36) | 227 | 102 | 42.2 | 0.70 (0.50, 0.98) | 0.01 | 0.05 | 300 | 160 | 41.9 | 0.62 (0.46, 0.84) | 0.02 | 0.08 |
| Bangladesh | WASH−B (37) | 102 | 102 | 17.6 | 0.33 (0.14, 0.80) | 0.00 | 0.01 | 132 | 84 | 14.3 | 0.53 (0.26, 1.07) | 0.00 | 0.02 |
| Burkina Faso | iLiNS−Zinc (38) | 1217 | 390 | 91.5 | 0.87 (0.83, 0.91) | 0.49 | 0.15 | 738 | 274 | 90.5 | 0.87 (0.82, 0.93) | 0.50 | 0.18 |
| Burkina Faso | PROMIS CS (39) | 335 | 349 | 70.2 | 0.97 (0.87, 1.08) | 0.10 | 0.13 | 239 | 227 | 70.0 | 0.88 (0.75, 1.04) | 0.09 | 0.14 |
| Ghana | GHANA (40) | | | | | | | | | | | | |
| Ghana | iLiNS−DYADG (41) | 288 | 536 | 42.9 | 0.86 (0.72, 1.03) | 0.04 | 0.10 | 35 | 114 | 51.8 | 0.94 (0.64, 1.38) | 0.01 | 0.06 |
| Kenya | WASH−B (42) | 266 | 227 | 48.9 | 0.65 (0.52, 0.80) | 0.03 | 0.09 | 72 | 60 | 36.7 | 0.91 (0.62, 1.34) | 0.01 | 0.06 |
| Madagascar | MAHAY (43) | | | | | | | | | | | | |
| Malawi | iLiNS−DYADM (44) | 124 | 258 | 51.2 | 0.87 (0.69, 1.09) | 0.02 | 0.08 | 85 | 171 | 53.8 | 0.92 (0.71, 1.19) | 0.03 | 0.09 |
| Malawi | iLiNS−DOSE (45) | 179 | 52 | 75.0 | 0.77 (0.63, 0.93) | 0.03 | 0.09 | 61 | 29 | 69.0 | 1.06 (0.80, 1.40) | 0.03 | 0.08 |
| Mali | PROMIS CS (46) | 702 | 685 | 86.6 | 0.82 (0.76, 0.88) | 0.20 | 0.14 | 246 | 279 | 85.7 | 0.82 (0.74, 0.90) | 0.25 | 0.17 |
| Zimbabwe | SHINE (HIV−) (47) | 1222 | 1161 | 34.7 | 0.76 (0.67, 0.86) | 0.07 | 0.12 | 213 | 220 | 33.2 | 0.93 (0.72, 1.22) | 0.03 | 0.09 |
| Zimbabwe | SHINE (HIV+) (48) | 221 | 222 | 36.9 | 0.67 (0.48, 0.95) | 0.01 | 0.05 | 58 | 40 | 40.0 | 0.43 (0.22, 0.83) | 0.01 | 0.02 |
| | | **4883** | **4084** | | **I² = 0.61, Tau² = 0.01** | | | **2179** | **1658** | | **I² = 0.34, Tau² = 0.02** | | |
| **Fixed** | | | | | **0.84 (0.81, 0.87)** | | | | | | **0.86 (0.82, 0.90)** | | |
| **Random** | | | | | **0.80 (0.73, 0.88)** | | | | | | **0.85 (0.76, 0.94)** | | |

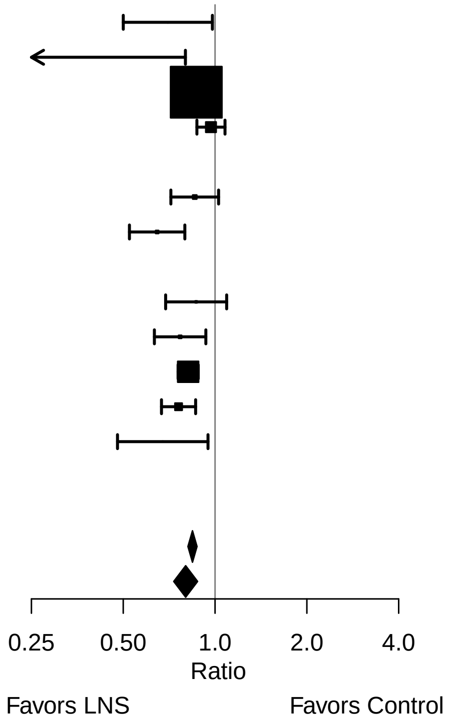

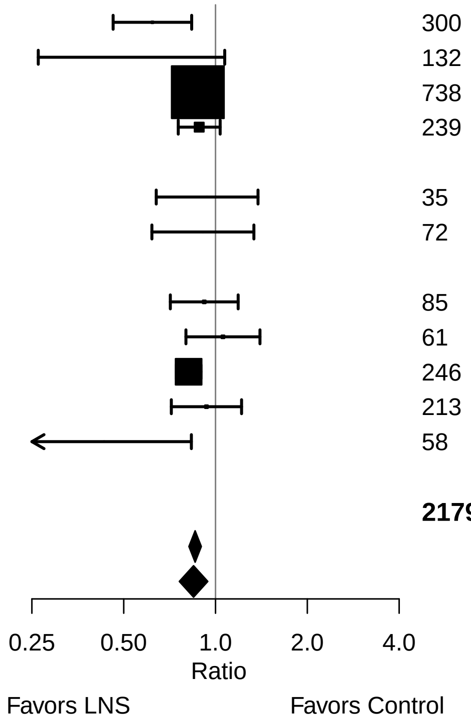

##### Supplemental figure 6B: Anemia prevalence ratio

#### 6B2: Stratified by Maternal age

**P−for−interaction = 0.041**
**Ratio of PRs = 1.05 (1.00, 1.10)**

**At least 25 y**

| Country | Trial | LNS N | Control N | Control Prevalence | PR (95% CI) | Fixed W | Random W |
|---|---|---|---|---|---|---|---|
| Bangladesh | JiVitA−4 (35) | 203 | 72 | 19.4 | 0.63 (0.35, 1.15) | 0.00 | 0.03 |
| Bangladesh | RDNS (36) | 152 | 81 | 40.7 | 0.60 (0.39, 0.92) | 0.01 | 0.04 |
| Bangladesh | WASH−B (37) | 89 | 83 | 14.5 | 0.39 (0.14, 1.06) | 0.00 | 0.01 |
| Burkina Faso | iLiNS−Zinc (38) | 1132 | 399 | 91.7 | 0.86 (0.82, 0.90) | 0.52 | 0.12 |
| Burkina Faso | PROMIS CS (39) | 271 | 263 | 67.3 | 0.89 (0.78, 1.03) | 0.07 | 0.10 |
| Ghana | GHANA (40) | 69 | 63 | 57.1 | 0.43 (0.27, 0.69) | 0.01 | 0.04 |
| Ghana | iLiNS−DYADG (41) | 206 | 397 | 42.3 | 0.80 (0.64, 1.00) | 0.03 | 0.08 |
| Kenya | WASH−B (42) | 213 | 168 | 45.2 | 0.71 (0.57, 0.87) | 0.03 | 0.08 |
| Madagascar | MAHAY (43) | 301 | 329 | 62.9 | 0.92 (0.78, 1.08) | 0.05 | 0.09 |
| Malawi | iLiNS−DYADM (44) | 99 | 220 | 47.3 | 0.96 (0.74, 1.24) | 0.02 | 0.07 |
| Malawi | iLiNS−DOSE (45) | 135 | 50 | 72.0 | 0.84 (0.69, 1.04) | 0.03 | 0.08 |
| Mali | PROMIS CS (46) | 502 | 508 | 87.8 | 0.79 (0.72, 0.86) | 0.18 | 0.11 |
| Zimbabwe | SHINE (HIV−) (47) | 808 | 726 | 33.7 | 0.71 (0.61, 0.83) | 0.05 | 0.10 |
| Zimbabwe | SHINE (HIV+) (48) | 221 | 204 | 37.7 | 0.56 (0.39, 0.80) | 0.01 | 0.05 |
| | | **4401** | **3563** | | **I² = 0.57, Tau² = 0.02** | | |
| **Fixed** | | | | | **0.83 (0.80, 0.86)** | | |
| **Random** | | | | | **0.77 (0.69, 0.86)** | | |

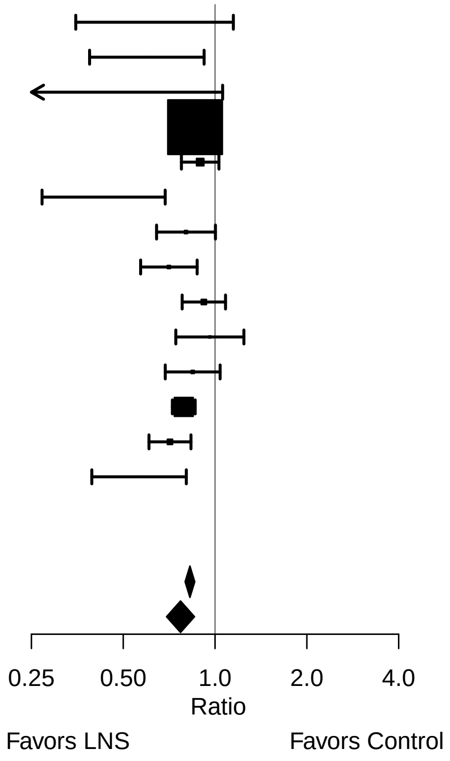

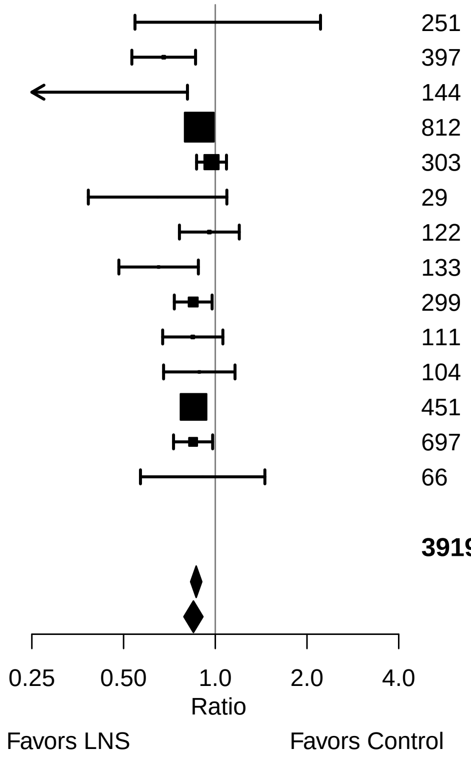

**Less than 25 y**

| Country | Trial | LNS N | Control N | Control Prevalence | PR (95% CI) | Fixed W | Random W |
|---|---|---|---|---|---|---|---|
| Bangladesh | JiVitA−4 (35) | 251 | 73 | 12.3 | 1.10 (0.54, 2.22) | 0.00 | 0.01 |
| Bangladesh | RDNS (36) | 397 | 191 | 42.4 | 0.68 (0.53, 0.86) | 0.03 | 0.06 |
| Bangladesh | WASH−B (37) | 144 | 103 | 17.5 | 0.44 (0.24, 0.81) | 0.00 | 0.01 |
| Burkina Faso | iLiNS−Zinc (38) | 812 | 262 | 90.1 | 0.89 (0.82, 0.96) | 0.28 | 0.15 |
| Burkina Faso | PROMIS CS (39) | 303 | 318 | 72.3 | 0.97 (0.87, 1.09) | 0.14 | 0.13 |
| Ghana | GHANA (40) | 29 | 25 | 64.0 | 0.65 (0.38, 1.09) | 0.01 | 0.02 |
| Ghana | iLiNS−DYADG (41) | 122 | 264 | 48.9 | 0.96 (0.76, 1.20) | 0.03 | 0.07 |
| Kenya | WASH−B (42) | 133 | 131 | 49.6 | 0.65 (0.48, 0.88) | 0.02 | 0.04 |
| Madagascar | MAHAY (43) | 299 | 259 | 67.2 | 0.85 (0.73, 0.98) | 0.09 | 0.11 |
| Malawi | iLiNS−DYADM (44) | 111 | 212 | 56.6 | 0.84 (0.67, 1.06) | 0.03 | 0.07 |
| Malawi | iLiNS−DOSE (45) | 104 | 28 | 71.4 | 0.89 (0.68, 1.16) | 0.02 | 0.05 |
| Mali | PROMIS CS (46) | 451 | 462 | 84.4 | 0.85 (0.78, 0.92) | 0.25 | 0.15 |
| Zimbabwe | SHINE (HIV−) (47) | 697 | 694 | 36.3 | 0.85 (0.73, 0.98) | 0.08 | 0.11 |
| Zimbabwe | SHINE (HIV+) (48) | 66 | 60 | 33.3 | 0.91 (0.57, 1.45) | 0.01 | 0.02 |
| | | **3919** | **3082** | | **I² = 0.33, Tau² = 0.01** | | |
| **Fixed** | | | | | **0.87 (0.83, 0.90)** | | |
| **Random** | | | | | **0.85 (0.79, 0.91)** | | |

#### Supplemental figure 6B: Anemia prevalence ratio

##### 6B3: Stratified by Maternal education

**P−for−interaction = 0.621**
**Ratio of PRs = 1.03 (0.93, 1.14)**

| | | Primary or greater | | | | | | Incomplete or no formal | | | | | |
|---|---|---|---|---|---|---|---|---|---|---|---|---|---|
| Country | Trial | LNS N | Control N | Control Prevalence | PR (95% CI) | Fixed W | Random W | LNS N | Control N | Control Prevalence | PR (95% CI) | Fixed W | Random W |
| Bangladesh | JiVitA−4 (35) | 299 | 93 | 17.2 | 0.64 (0.35, 1.19) | 0.01 | 0.05 | 157 | 53 | 13.2 | 1.30 (0.69, 2.47) | 0.01 | 0.03 |
| Bangladesh | RDNS (36) | 405 | 195 | 41.0 | 0.62 (0.47, 0.82) | 0.05 | 0.11 | 144 | 77 | 44.2 | 0.75 (0.50, 1.13) | 0.02 | 0.06 |
| Bangladesh | WASH−B (37) | 163 | 145 | 15.9 | 0.39 (0.20, 0.75) | 0.01 | 0.05 | 71 | 41 | 17.1 | 0.49 (0.18, 1.36) | 0.00 | 0.01 |
| Burkina Faso | iLiNS−Zinc (38) | | | | | | | | | | | | |
| Burkina Faso | PROMIS CS (39) | 46 | 47 | 70.2 | 1.05 (0.83, 1.34) | 0.07 | 0.11 | 526 | 532 | 69.9 | 0.93 (0.84, 1.03) | 0.26 | 0.19 |
| Ghana | GHANA (40) | | | | | | | | | | | | |
| Ghana | iLiNS−DYADG (41) | 253 | 526 | 45.4 | 0.83 (0.69, 0.99) | 0.12 | 0.13 | 75 | 135 | 43.0 | 0.99 (0.72, 1.38) | 0.03 | 0.08 |
| Kenya | WASH−B (42) | 168 | 136 | 50.0 | 0.46 (0.34, 0.63) | 0.04 | 0.10 | 181 | 164 | 45.1 | 0.91 (0.72, 1.14) | 0.05 | 0.12 |
| Madagascar | MAHAY (43) | 128 | 143 | 64.3 | 0.79 (0.62, 1.00) | 0.07 | 0.12 | 472 | 445 | 64.9 | 0.91 (0.79, 1.05) | 0.13 | 0.16 |
| Malawi | iLiNS−DYADM (44) | 34 | 66 | 48.5 | 0.79 (0.48, 1.29) | 0.02 | 0.07 | 175 | 363 | 52.9 | 0.91 (0.76, 1.09) | 0.08 | 0.14 |
| Malawi | iLiNS−DOSE (45) | | | | | | | | | | | | |
| Mali | PROMIS CS (46) | 107 | 96 | 84.4 | 0.90 (0.78, 1.03) | 0.22 | 0.14 | 845 | 874 | 86.4 | 0.81 (0.75, 0.88) | 0.42 | 0.20 |
| Zimbabwe | SHINE (HIV−) (47) | 1536 | 1462 | 34.9 | 0.80 (0.72, 0.89) | 0.37 | 0.14 | 58 | 53 | 39.6 | 0.35 (0.16, 0.75) | 0.00 | 0.02 |
| Zimbabwe | SHINE (HIV+) (48) | | | | | | | | | | | | |
| | | **3139** | **2909** | | **I² = 0.68, Tau² = 0.05** | | | **2704** | **2737** | | **I² = 0.39, Tau² = 0.02** | | |
| **Fixed** | | | | | **0.80 (0.75, 0.86)** | | | | | | **0.87 (0.82, 0.91)** | | |
| **Random** | | | | | **0.74 (0.62, 0.88)** | | | | | | **0.87 (0.78, 0.98)** | | |

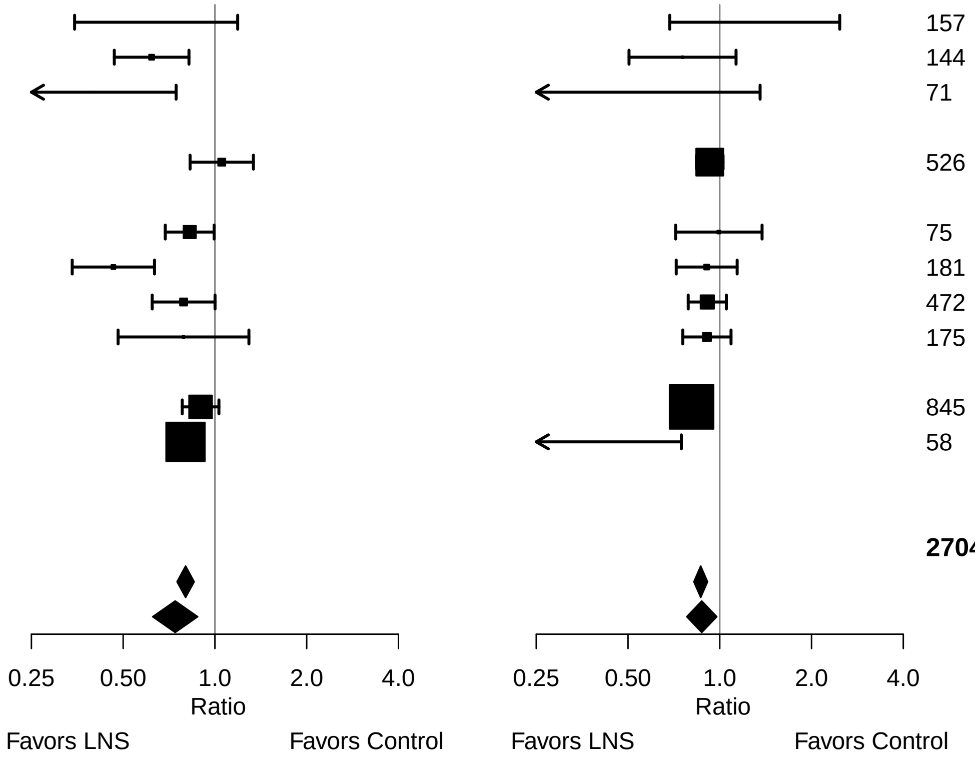

#### Supplemental figure 6B: Anemia prevalence ratio

##### 6B4: Stratified by Child sex

**P−for−interaction = 0.039**
**Ratio of PRs = 0.96 (0.92, 1.00)**

**Male**

| Country | Trial | LNS N | Control N | Control Prevalence | PR (95% CI) | Fixed W | Random W |
|---|---|---|---|---|---|---|---|
| Bangladesh | JiVitA−4 (35) | 235 | 71 | 22.5 | 0.47 (0.28, 0.78) | 0.00 | 0.04 |
| Bangladesh | RDNS (36) | 266 | 131 | 41.2 | 0.62 (0.42, 0.91) | 0.01 | 0.05 |
| Bangladesh | WASH−B (37) | 116 | 89 | 16.9 | 0.31 (0.12, 0.82) | 0.00 | 0.01 |
| Burkina Faso | iLiNS−Zinc (38) | 993 | 334 | 91.6 | 0.88 (0.85, 0.92) | 0.56 | 0.10 |
| Burkina Faso | PROMIS CS (39) | 317 | 295 | 77.3 | 0.93 (0.83, 1.05) | 0.07 | 0.09 |
| Ghana | GHANA (40) | 60 | 41 | 61.0 | 0.46 (0.29, 0.74) | 0.00 | 0.04 |
| Ghana | iLiNS−DYADG (41) | 164 | 310 | 44.8 | 0.97 (0.78, 1.20) | 0.02 | 0.08 |
| Kenya | WASH−B (42) | 168 | 145 | 44.8 | 0.81 (0.63, 1.04) | 0.02 | 0.07 |
| Madagascar | MAHAY (43) | 277 | 288 | 65.6 | 0.95 (0.83, 1.09) | 0.05 | 0.09 |
| Malawi | iLiNS−DYADM (44) | 103 | 202 | 56.4 | 0.95 (0.76, 1.18) | 0.02 | 0.08 |
| Malawi | iLiNS−DOSE (45) | 123 | 43 | 76.7 | 0.84 (0.68, 1.04) | 0.02 | 0.08 |
| Mali | PROMIS CS (46) | 479 | 526 | 89.0 | 0.84 (0.78, 0.91) | 0.17 | 0.10 |
| Zimbabwe | SHINE (HIV−) (47) | 830 | 814 | 36.2 | 0.81 (0.71, 0.94) | 0.05 | 0.09 |
| Zimbabwe | SHINE (HIV+) (48) | 146 | 149 | 40.9 | 0.62 (0.46, 0.83) | 0.01 | 0.07 |
| | | **4277** | **3438** | | **I² = 0.59, Tau² = 0.04** | | |
| **Fixed** | | | | | **0.87 (0.84, 0.90)** | | |
| **Random** | | | | | **0.79 (0.70, 0.90)** | | |

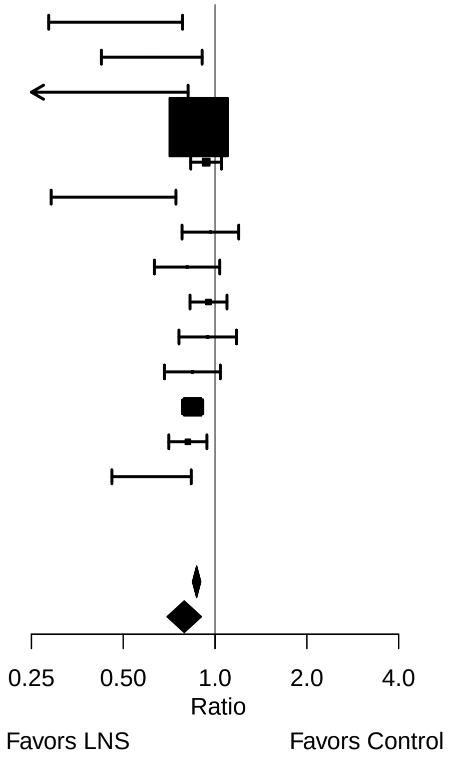

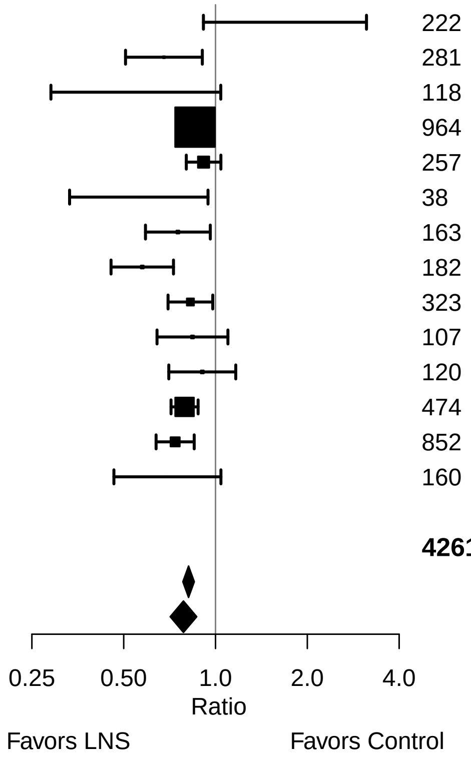

**Female**

| LNS N | Control N | Control Prevalence | PR (95% CI) | Fixed W | Random W |
|---|---|---|---|---|---|
| 222 | 75 | 9.3 | 1.69 (0.91, 3.13) | 0.00 | 0.02 |
| 281 | 141 | 42.6 | 0.68 (0.51, 0.91) | 0.02 | 0.06 |
| 118 | 97 | 15.5 | 0.55 (0.29, 1.04) | 0.00 | 0.02 |
| 964 | 330 | 90.6 | 0.86 (0.80, 0.92) | 0.38 | 0.12 |
| 257 | 286 | 62.6 | 0.91 (0.80, 1.04) | 0.11 | 0.11 |
| 38 | 55 | 56.4 | 0.56 (0.33, 0.94) | 0.01 | 0.03 |
| 163 | 348 | 44.8 | 0.75 (0.59, 0.96) | 0.03 | 0.07 |
| 182 | 155 | 49.7 | 0.58 (0.45, 0.73) | 0.03 | 0.08 |
| 323 | 300 | 64.0 | 0.83 (0.70, 0.98) | 0.07 | 0.09 |
| 107 | 230 | 47.8 | 0.84 (0.64, 1.10) | 0.03 | 0.07 |
| 120 | 39 | 66.7 | 0.90 (0.70, 1.17) | 0.03 | 0.07 |
| 474 | 444 | 82.9 | 0.79 (0.71, 0.88) | 0.18 | 0.11 |
| 852 | 780 | 34.2 | 0.74 (0.64, 0.85) | 0.09 | 0.10 |
| 160 | 136 | 32.4 | 0.70 (0.46, 1.04) | 0.01 | 0.04 |
| **4261** | **3416** | | **I² = 0.53, Tau² = 0.02** | | |
| | | | **0.82 (0.78, 0.85)** | | |
| | | | **0.78 (0.71, 0.87)** | | |

#### Supplemental figure 6B: Anemia prevalence ratio

##### 6B5: Stratified by Child birth order

**P−for−interaction = 0.156**
**Ratio of PRs = 1.04 (0.99, 1.09)**

| | | Later born | | | | | | Firstborn | | | | | |
|---|---|---|---|---|---|---|---|---|---|---|---|---|---|
| Country | Trial | LNS N | Control N | Control Prevalence | PR (95% CI) | Fixed W | Random W | LNS N | Control N | Control Prevalence | PR (95% CI) | Fixed W | Random W |
| Bangladesh | JiVitA−4 (35) | 115 | 33 | 27.3 | 0.54 (0.27, 1.09) | 0.00 | 0.03 | 341 | 112 | 12.5 | 1.01 (0.63, 1.61) | 0.01 | 0.02 |
| Bangladesh | RDNS (36) | 339 | 171 | 40.9 | 0.66 (0.49, 0.88) | 0.01 | 0.07 | 210 | 101 | 43.6 | 0.66 (0.47, 0.92) | 0.02 | 0.04 |
| Bangladesh | WASH−B (37) | 137 | 123 | 16.3 | 0.31 (0.14, 0.69) | 0.00 | 0.02 | 95 | 61 | 16.4 | 0.51 (0.20, 1.31) | 0.00 | 0.01 |
| Burkina Faso | iLiNS−Zinc (38) | 1522 | 520 | 90.4 | 0.87 (0.82, 0.92) | 0.38 | 0.11 | 435 | 144 | 93.8 | 0.87 (0.82, 0.92) | 0.64 | 0.29 |
| Burkina Faso | PROMIS CS (39) | 498 | 477 | 70.0 | 0.93 (0.84, 1.03) | 0.11 | 0.10 | 75 | 103 | 69.9 | 0.99 (0.80, 1.23) | 0.04 | 0.09 |
| Ghana | GHANA (40) | 60 | 54 | 61.1 | 0.38 (0.23, 0.63) | 0.00 | 0.04 | 38 | 38 | 55.3 | 0.71 (0.44, 1.16) | 0.01 | 0.02 |
| Ghana | iLiNS−DYADG (41) | 220 | 435 | 45.3 | 0.76 (0.62, 0.94) | 0.03 | 0.09 | 108 | 226 | 44.2 | 1.07 (0.83, 1.37) | 0.03 | 0.07 |
| Kenya | WASH−B (42) | 282 | 244 | 47.1 | 0.67 (0.54, 0.83) | 0.03 | 0.08 | 67 | 56 | 48.2 | 0.74 (0.46, 1.20) | 0.01 | 0.02 |
| Madagascar | MAHAY (43) | 458 | 454 | 66.1 | 0.90 (0.78, 1.03) | 0.06 | 0.10 | 141 | 131 | 61.1 | 0.84 (0.66, 1.05) | 0.04 | 0.08 |
| Malawi | iLiNS−DYADM (44) | 165 | 342 | 50.9 | 0.82 (0.67, 1.01) | 0.03 | 0.09 | 45 | 88 | 55.7 | 1.16 (0.87, 1.54) | 0.03 | 0.06 |
| Malawi | iLiNS−DOSE (45) | | | | | | | | | | | | |
| Mali | PROMIS CS (46) | 790 | 832 | 86.1 | 0.81 (0.76, 0.88) | 0.22 | 0.10 | 146 | 126 | 88.1 | 0.82 (0.71, 0.93) | 0.11 | 0.17 |
| Zimbabwe | SHINE (HIV−) (47) | 1188 | 1100 | 35.5 | 0.71 (0.63, 0.80) | 0.10 | 0.10 | 418 | 416 | 33.2 | 0.94 (0.77, 1.15) | 0.05 | 0.10 |
| Zimbabwe | SHINE (HIV+) (48) | 252 | 216 | 36.6 | 0.63 (0.47, 0.84) | 0.02 | 0.07 | 51 | 64 | 37.5 | 0.78 (0.46, 1.35) | 0.01 | 0.02 |
| | | **6026** | **5001** | | **I² = 0.72, Tau² = 0.04** | | | **2170** | **1666** | | **I² = 0.19, Tau² = 0.00** | | |
| **Fixed** | | | | | **0.83 (0.80, 0.86)** | | | | | | **0.87 (0.83, 0.91)** | | |
| **Random** | | | | | **0.73 (0.64, 0.84)** | | | | | | **0.88 (0.82, 0.95)** | | |

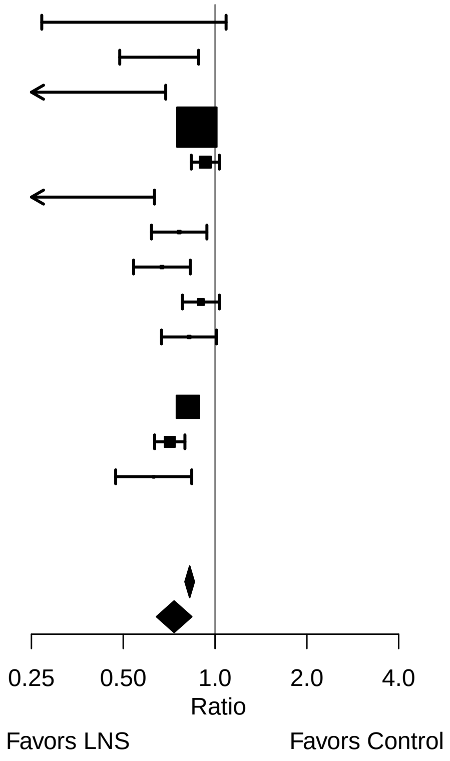

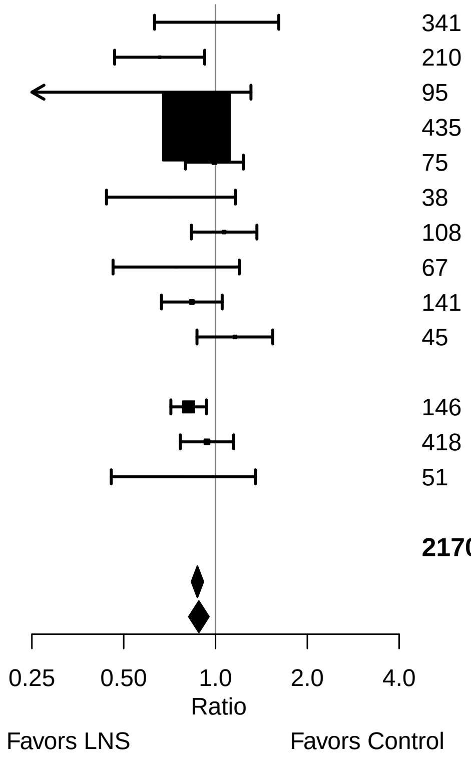

**Supplemental figure 6B: Anemia prevalence ratio**

#### 6B6: Stratified by Child baseline acute malnutrition

**P−for−interaction = 0.393**
**Ratio of PRs = 0.98 (0.93, 1.03)**

| | | No | | | | | | Yes | | | | | |
|---|---|---|---|---|---|---|---|---|---|---|---|---|---|
| Country | Trial | LNS N | Control N | Control Prevalence | PR (95% CI) | Fixed W | Random W | LNS N | Control N | Control Prevalence | PR (95% CI) | Fixed W | Random W |
| Bangladesh | JiVitA−4 (35) | 365 | 114 | 15.8 | 0.80 (0.53, 1.21) | 0.01 | 0.04 | 90 | 32 | 15.6 | 1.00 (0.40, 2.49) | 0.00 | 0.00 |
| Bangladesh | RDNS (36) | 493 | 243 | 40.3 | 0.65 (0.51, 0.85) | 0.03 | 0.09 | 35 | 12 | 41.7 | 0.82 (0.31, 2.21) | 0.00 | 0.00 |
| Bangladesh | WASH−B (37) | | | | | | | | | | | | |
| Burkina Faso | iLiNS−Zinc (38) | 1457 | 487 | 90.3 | 0.88 (0.83, 0.92) | 0.71 | 0.31 | 500 | 177 | 93.2 | 0.85 (0.80, 0.91) | 0.95 | 0.95 |
| Burkina Faso | PROMIS CS (39) | | | | | | | | | | | | |
| Ghana | GHANA (40) | | | | | | | | | | | | |
| Ghana | iLiNS−DYADG (41) | 285 | 556 | 46.4 | 0.84 (0.71, 1.00) | 0.06 | 0.15 | 21 | 54 | 31.5 | 1.06 (0.51, 2.18) | 0.01 | 0.01 |
| Kenya | WASH−B (42) | | | | | | | | | | | | |
| Madagascar | MAHAY (43) | | | | | | | | | | | | |
| Malawi | iLiNS−DYADM (44) | 183 | 379 | 51.5 | 0.90 (0.75, 1.08) | 0.06 | 0.14 | 14 | 37 | 51.4 | 1.11 (0.64, 1.93) | 0.01 | 0.01 |
| Malawi | iLiNS−DOSE (45) | | | | | | | | | | | | |
| Mali | PROMIS CS (46) | | | | | | | | | | | | |
| Zimbabwe | SHINE (HIV−) (47) | 1185 | 1069 | 32.8 | 0.80 (0.70, 0.91) | 0.11 | 0.20 | 66 | 67 | 32.8 | 0.74 (0.42, 1.31) | 0.01 | 0.01 |
| Zimbabwe | SHINE (HIV+) (48) | 226 | 202 | 34.7 | 0.63 (0.46, 0.85) | 0.02 | 0.07 | 26 | 13 | 61.5 | 0.38 (0.16, 0.86) | 0.01 | 0.01 |
| | | **4194** | **3050** | | **I² = 0.42, Tau² = 0.01** | | | **752** | **392** | | **I² = 0.00, Tau² = 0.00** | | |
| **Fixed** | | | | | **0.85 (0.82, 0.89)** | | | | | | **0.85 (0.80, 0.91)** | | |
| **Random** | | | | | **0.82 (0.75, 0.89)** | | | | | | **0.85 (0.80, 0.91)** | | |

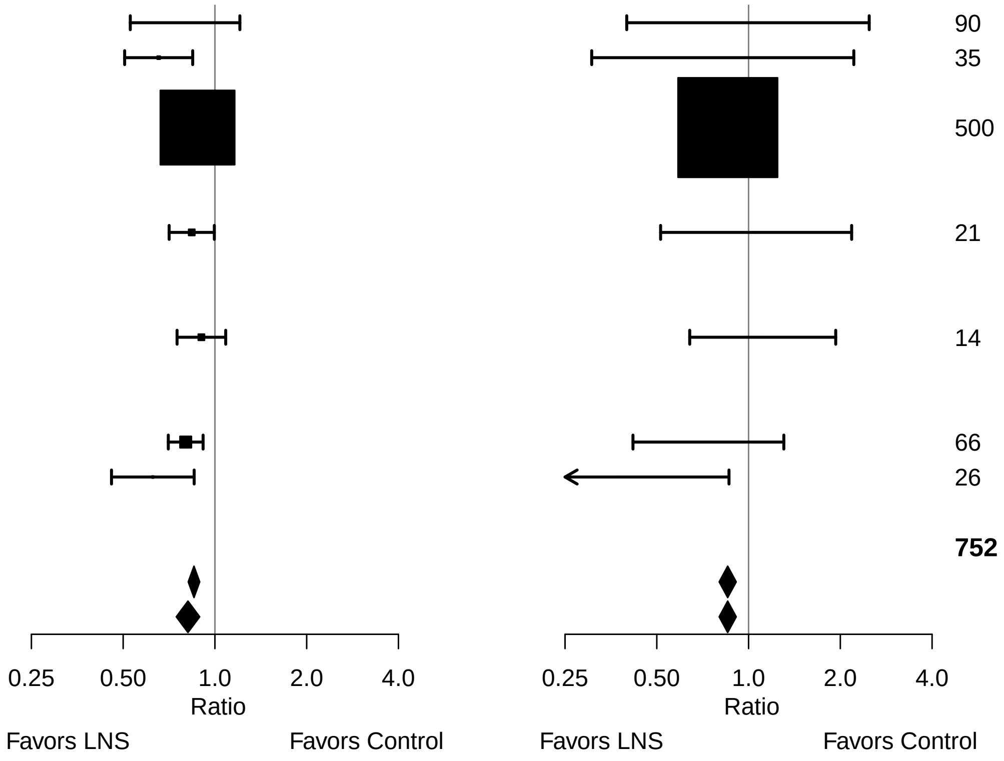

**Supplemental figure 6B: Anemia prevalence ratio**

#### 6B7: Stratified by Child baseline anemia

**P−for−interaction = 0.683**
**Ratio of PRs = 1.03 (0.91, 1.16)**

| Country | Trial | Not anemic: LNS N | Control N | Control Prevalence | PR (95% CI) | Fixed W | Random W | Anemic: LNS N | Control N | Control Prevalence | PR (95% CI) | Fixed W | Random W |
|---|---|---|---|---|---|---|---|---|---|---|---|---|---|
| Bangladesh | JiVitA−4 (35) | | | | | | | | | | | | |
| Bangladesh | RDNS (36) | 213 | 93 | 23.7 | 0.62 (0.34, 1.10) | 0.04 | 0.05 | 314 | 160 | 50.0 | 0.71 (0.55, 0.90) | 0.04 | 0.04 |
| Bangladesh | WASH−B (37) | | | | | | | | | | | | |
| Burkina Faso | iLiNS−Zinc (38) | 177 | 61 | 82.0 | 0.79 (0.68, 0.93) | 0.50 | 0.44 | 1780 | 603 | 92.0 | 0.88 (0.83, 0.92) | 0.79 | 0.79 |
| Burkina Faso | PROMIS CS (39) | | | | | | | | | | | | |
| Ghana | GHANA (40) | | | | | | | | | | | | |
| Ghana | iLiNS−DYADG (41) | 189 | 355 | 32.7 | 1.00 (0.78, 1.29) | 0.20 | 0.22 | 90 | 207 | 64.7 | 0.81 (0.65, 1.01) | 0.05 | 0.05 |
| Kenya | WASH−B (42) | | | | | | | | | | | | |
| Madagascar | MAHAY (43) | | | | | | | | | | | | |
| Malawi | iLiNS−DYADM (44) | 66 | 145 | 36.6 | 1.04 (0.71, 1.51) | 0.09 | 0.11 | 134 | 269 | 59.1 | 0.88 (0.73, 1.07) | 0.06 | 0.06 |
| Malawi | iLiNS−DOSE (45) | 101 | 45 | 64.4 | 0.85 (0.64, 1.12) | 0.17 | 0.19 | 142 | 37 | 81.1 | 0.88 (0.73, 1.06) | 0.06 | 0.06 |
| Mali | PROMIS CS (46) | | | | | | | | | | | | |
| Zimbabwe | SHINE (HIV−) (47) | | | | | | | | | | | | |
| Zimbabwe | SHINE (HIV+) (48) | | | | | | | | | | | | |
| | | **746** | **699** | | **$I^2$ = 0.13, $Tau^2$ = 0.00** | | | **2460** | **1276** | | **$I^2$ = 0.00, $Tau^2$ = 0.00** | | |
| **Fixed** | | | | | **0.85 (0.76, 0.96)** | | | | | | **0.87 (0.83, 0.91)** | | |
| **Random** | | | | | **0.86 (0.76, 0.98)** | | | | | | **0.87 (0.83, 0.91)** | | |

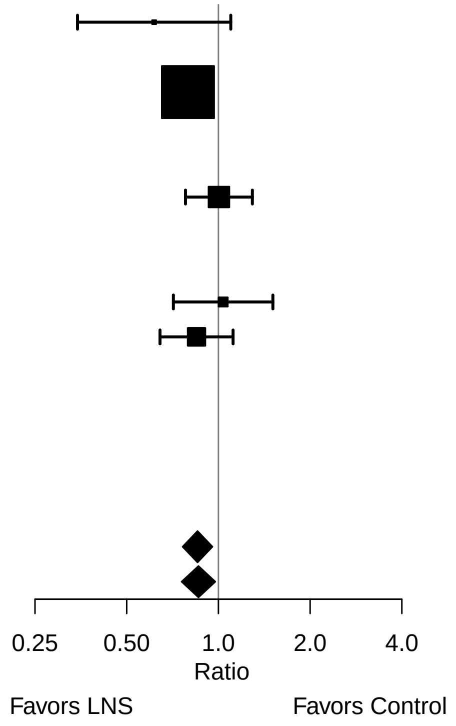

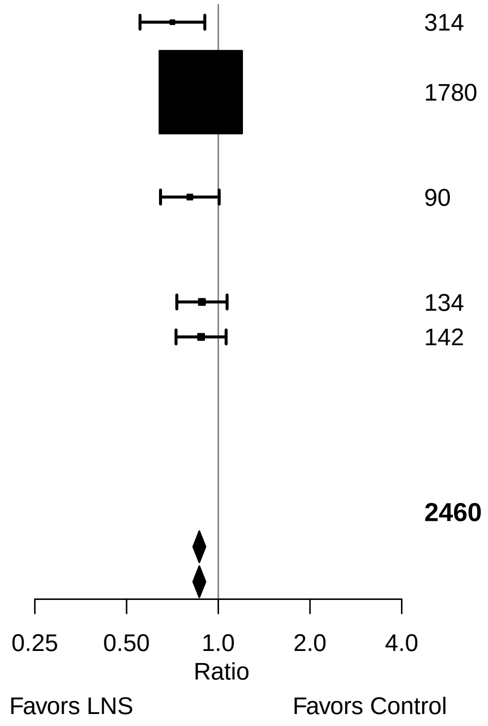

#### Supplemental figure 6B: Anemia prevalence ratio

##### 6B8: Stratified by Child high-dose vitamin A supplementation

**P−for−interaction = 0.760**
**Ratio of PRs = 0.98 (0.85, 1.12)**

| Country | Trial | Received VitA: LNS N | Control N | Control Prevalence | PR (95% CI) | Fixed W | Random W | Not treated: LNS N | Control N | Control Prevalence | PR (95% CI) | Fixed W | Random W |
|---|---|---|---|---|---|---|---|---|---|---|---|---|---|
| Bangladesh | JiVitA−4 (35) | 141 | 37 | 16.2 | 0.96 (0.38, 2.44) | 0.01 | 0.03 | 133 | 41 | 14.6 | 0.87 (0.36, 2.14) | 0.01 | 0.02 |
| Bangladesh | RDNS (36) | 444 | 223 | 40.8 | 0.67 (0.50, 0.89) | 0.11 | 0.15 | 105 | 49 | 46.9 | 0.61 (0.43, 0.87) | 0.07 | 0.09 |
| Bangladesh | WASH−B (37) | | | | | | | | | | | | |
| Burkina Faso | iLiNS−Zinc (38) | | | | | | | | | | | | |
| Burkina Faso | PROMIS CS (39) | | | | | | | | | | | | |
| Ghana | GHANA (40) | 50 | 62 | 56.5 | 0.39 (0.22, 0.69) | 0.03 | 0.06 | 48 | 29 | 65.5 | 0.57 (0.36, 0.90) | 0.04 | 0.06 |
| Ghana | iLiNS−DYADG (41) | 215 | 412 | 45.6 | 0.87 (0.71, 1.05) | 0.23 | 0.21 | 113 | 249 | 43.8 | 0.85 (0.64, 1.12) | 0.11 | 0.13 |
| Kenya | WASH−B (42) | 29 | 34 | 52.9 | 0.65 (0.36, 1.19) | 0.02 | 0.05 | 263 | 212 | 46.7 | 0.68 (0.55, 0.84) | 0.19 | 0.19 |
| Madagascar | MAHAY (43) | | | | | | | | | | | | |
| Malawi | iLiNS−DYADM (44) | 75 | 136 | 58.8 | 0.77 (0.58, 1.03) | 0.11 | 0.15 | 133 | 289 | 48.4 | 0.98 (0.79, 1.21) | 0.18 | 0.18 |
| Malawi | iLiNS−DOSE (45) | | | | | | | | | | | | |
| Mali | PROMIS CS (46) | | | | | | | | | | | | |
| Zimbabwe | SHINE (HIV−) (47) | 874 | 833 | 33.7 | 0.83 (0.72, 0.97) | 0.42 | 0.24 | 665 | 622 | 35.9 | 0.71 (0.61, 0.83) | 0.34 | 0.24 |
| Zimbabwe | SHINE (HIV+) (48) | 165 | 127 | 37.0 | 0.59 (0.41, 0.85) | 0.07 | 0.11 | 112 | 134 | 36.6 | 0.63 (0.43, 0.94) | 0.06 | 0.08 |
| | | **1993** | **1864** | | **I² = 0.39, Tau² = 0.02** | | | **1572** | **1625** | | **I² = 0.38, Tau² = 0.01** | | |
| **Fixed** | | | | | **0.78 (0.71, 0.85)** | | | | | | **0.75 (0.68, 0.82)** | | |
| **Random** | | | | | **0.73 (0.63, 0.85)** | | | | | | **0.74 (0.65, 0.84)** | | |

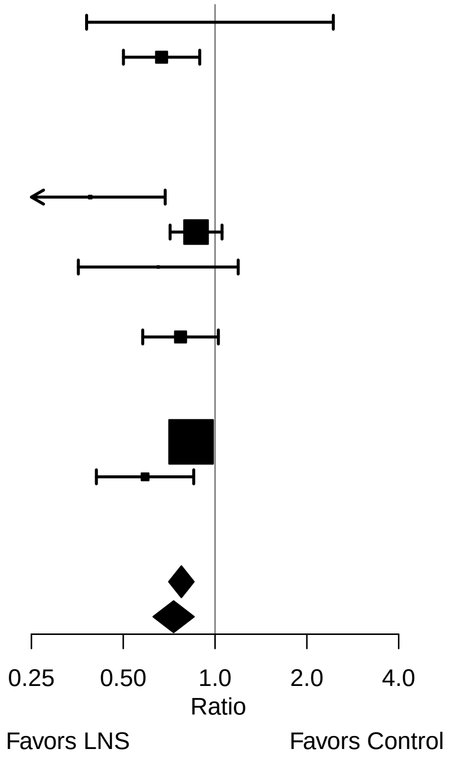

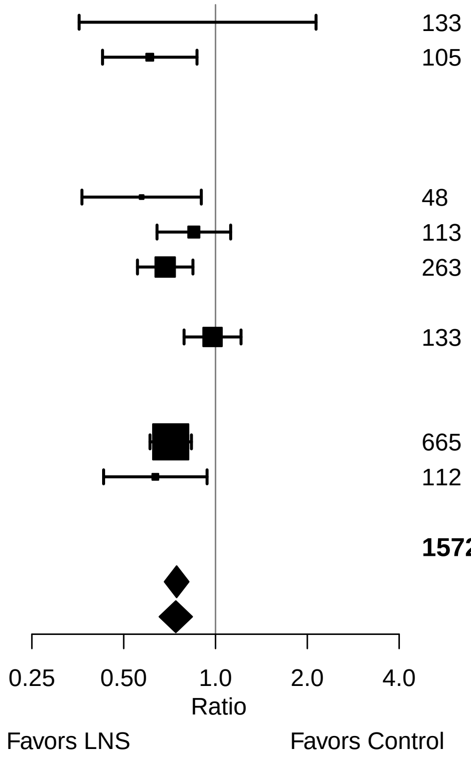

#### Supplemental figure 6B: Anemia prevalence ratio

##### 6B9: Stratified by Child inflammation

**P−for−interaction = 0.006**
**Ratio of PRs = 1.30 (1.08, 1.57)**

Not inflamed:

| Country | Trial | LNS N | Control N | Control Prevalence | PR (95% CI) | Fixed W | Random W |
|---|---|---|---|---|---|---|---|
| Bangladesh | JiVitA−4 (35) | 185 | 47 | 17.0 | 0.67 (0.27, 1.63) | 0.02 | 0.04 |
| Bangladesh | RDNS (36) | 369 | 168 | 42.3 | 0.57 (0.42, 0.77) | 0.19 | 0.18 |
| Bangladesh | WASH−B (37) | | | | | | |
| Burkina Faso | iLiNS−Zinc (38) | | | | | | |
| Burkina Faso | PROMIS CS (39) | | | | | | |
| Ghana | GHANA (40) | | | | | | |
| Ghana | iLiNS−DYADG (41) | 45 | 120 | 46.7 | 0.90 (0.61, 1.34) | 0.11 | 0.13 |
| Kenya | WASH−B (42) | 157 | 135 | 43.0 | 0.47 (0.33, 0.68) | 0.13 | 0.15 |
| Madagascar | MAHAY (43) | 44 | 32 | 43.8 | 0.42 (0.20, 0.87) | 0.03 | 0.05 |
| Malawi | iLiNS−DYADM (44) | 59 | 123 | 40.7 | 0.58 (0.35, 0.97) | 0.07 | 0.09 |
| Malawi | iLiNS−DOSE (45) | | | | | | |
| Mali | PROMIS CS (46) | | | | | | |
| Zimbabwe | SHINE (HIV−) (47) | 414 | 354 | 28.8 | 0.84 (0.66, 1.06) | 0.31 | 0.22 |
| Zimbabwe | SHINE (HIV+) (48) | 186 | 158 | 35.4 | 0.56 (0.39, 0.81) | 0.13 | 0.14 |
| | | **1459** | **1137** | | **I² = 0.46, Tau² = 0.02** | | |
| **Fixed** | | | | | **0.66 (0.58, 0.75)** | | |
| **Random** | | | | | **0.64 (0.53, 0.76)** | | |

High AGP or CRP:

| Country | Trial | LNS N | Control N | Control Prevalence | PR (95% CI) | Fixed W | Random W |
|---|---|---|---|---|---|---|---|
| Bangladesh | JiVitA−4 (35) | 269 | 98 | 14.3 | 0.99 (0.60, 1.62) | 0.05 | 0.05 |
| Bangladesh | RDNS (36) | 180 | 104 | 41.3 | 0.83 (0.61, 1.13) | 0.13 | 0.13 |
| Bangladesh | WASH−B (37) | | | | | | |
| Burkina Faso | iLiNS−Zinc (38) | | | | | | |
| Burkina Faso | PROMIS CS (39) | | | | | | |
| Ghana | GHANA (40) | | | | | | |
| Ghana | iLiNS−DYADG (41) | 55 | 82 | 53.7 | 0.81 (0.57, 1.17) | 0.09 | 0.09 |
| Kenya | WASH−B (42) | 141 | 123 | 52.8 | 0.87 (0.69, 1.10) | 0.22 | 0.22 |
| Madagascar | MAHAY (43) | 39 | 19 | 47.4 | 0.60 (0.31, 1.13) | 0.03 | 0.03 |
| Malawi | iLiNS−DYADM (44) | 138 | 269 | 57.6 | 0.93 (0.77, 1.12) | 0.35 | 0.35 |
| Malawi | iLiNS−DOSE (45) | | | | | | |
| Mali | PROMIS CS (46) | | | | | | |
| Zimbabwe | SHINE (HIV−) (47) | 105 | 99 | 43.4 | 0.72 (0.52, 1.01) | 0.11 | 0.11 |
| Zimbabwe | SHINE (HIV+) (48) | 32 | 31 | 41.9 | 0.67 (0.32, 1.39) | 0.02 | 0.02 |
| | | **959** | **825** | | **I² = 0.00, Tau² = 0.00** | | |
| **Fixed** | | | | | **0.85 (0.77, 0.95)** | | |
| **Random** | | | | | **0.85 (0.77, 0.95)** | | |

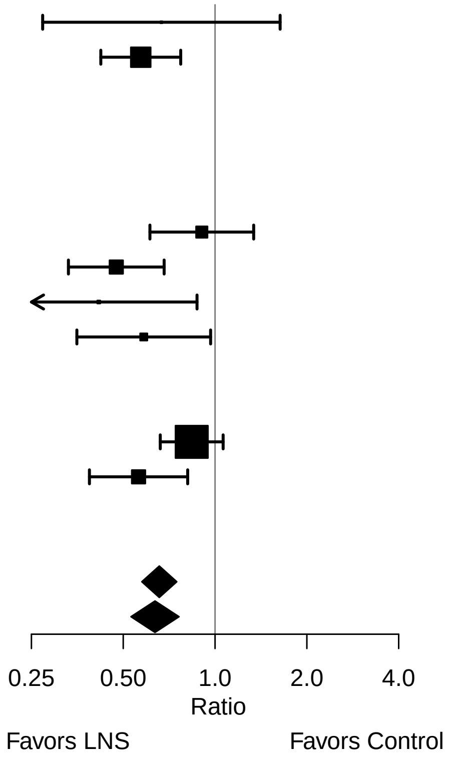

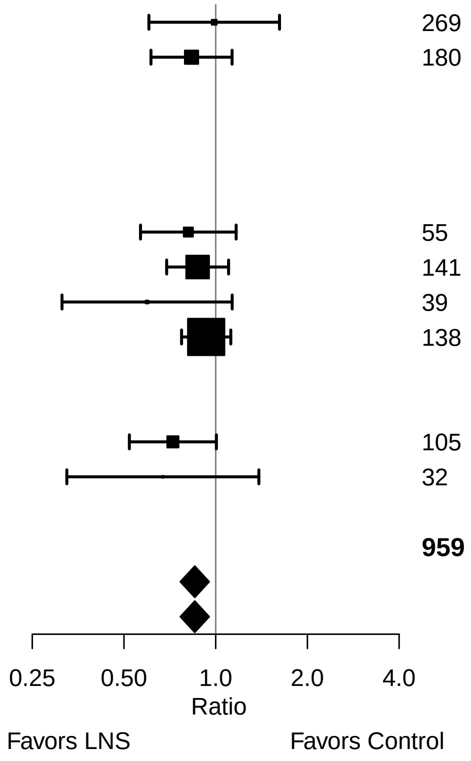

### Supplemental figure 6C: Anemia prevalence difference

#### 6C1: Stratified by Maternal BMI

**P−for−interaction = 0.397**
**Difference in PDs = 0.01 (−0.02, 0.05)**

| | | At least 20 kg/m² | | | | | | Less than 20 kg/m² | | | | | |
|---|---|---|---|---|---|---|---|---|---|---|---|---|---|
| Country | Trial | LNS N | Control N | Control Prevalence | PD (95% CI) | Fixed W | Random W | LNS N | Control N | Control Prevalence | PD (95% CI) | Fixed W | Random W |
| Bangladesh | JiVitA−4 (35) | | | | | | | | | | | | |
| Bangladesh | RDNS (36) | 227 | 102 | 42.2 | −0.13 (−0.25, 0.00) | 0.03 | 0.04 | 300 | 160 | 41.9 | −0.16 (−0.27, −0.05) | 0.07 | 0.09 |
| Bangladesh | WASH−B (37) | 102 | 102 | 17.6 | −0.12 (−0.21, −0.03) | 0.05 | 0.07 | 132 | 84 | 14.3 | −0.07 (−0.15, 0.02) | 0.12 | 0.13 |
| Burkina Faso | iLiNS−Zinc (38) | 1217 | 390 | 91.5 | −0.12 (−0.16, −0.08) | 0.23 | 0.17 | 738 | 274 | 90.5 | −0.11 (−0.17, −0.06) | 0.26 | 0.19 |
| Burkina Faso | PROMIS CS (39) | 335 | 349 | 70.2 | −0.02 (−0.09, 0.05) | 0.07 | 0.09 | 239 | 227 | 70.0 | −0.08 (−0.19, 0.02) | 0.08 | 0.09 |
| Ghana | GHANA (40) | | | | | | | | | | | | |
| Ghana | iLiNS−DYADG (41) | 288 | 536 | 42.9 | −0.06 (−0.13, 0.01) | 0.08 | 0.10 | 35 | 114 | 51.8 | −0.03 (−0.22, 0.16) | 0.02 | 0.03 |
| Kenya | WASH−B (42) | 266 | 227 | 48.9 | −0.17 (−0.26, −0.09) | 0.06 | 0.08 | 72 | 60 | 36.7 | −0.03 (−0.17, 0.10) | 0.05 | 0.06 |
| Madagascar | MAHAY (43) | | | | | | | | | | | | |
| Malawi | iLiNS−DYADM (44) | 124 | 258 | 51.2 | −0.07 (−0.18, 0.04) | 0.03 | 0.05 | 85 | 171 | 53.8 | −0.04 (−0.17, 0.09) | 0.05 | 0.06 |
| Malawi | iLiNS−DOSE (45) | 179 | 52 | 75.0 | −0.18 (−0.32, −0.03) | 0.02 | 0.03 | 61 | 29 | 69.0 | 0.04 (−0.16, 0.23) | 0.02 | 0.03 |
| Mali | PROMIS CS (46) | 702 | 685 | 86.6 | −0.16 (−0.22, −0.10) | 0.12 | 0.13 | 246 | 279 | 85.7 | −0.16 (−0.23, −0.09) | 0.18 | 0.16 |
| Zimbabwe | SHINE (HIV−) (47) | 1222 | 1161 | 34.7 | −0.08 (−0.12, −0.04) | 0.26 | 0.18 | 213 | 220 | 33.2 | −0.02 (−0.11, 0.06) | 0.12 | 0.12 |
| Zimbabwe | SHINE (HIV+) (48) | 221 | 222 | 36.9 | −0.12 (−0.22, −0.02) | 0.04 | 0.06 | 58 | 40 | 40.0 | −0.23 (−0.40, −0.05) | 0.03 | 0.04 |
| | | **4883** | **4084** | | **I² = 0.38, Tau² = 0.00** | | | **2179** | **1658** | | **I² = 0.28, Tau² = 0.00** | | |
| **Fixed** | | | | | **−0.11 (−0.13, −0.09)** | | | | | | **−0.10 (−0.13, −0.07)** | | |
| **Random** | | | | | **−0.11 (−0.13, −0.08)** | | | | | | **−0.09 (−0.13, −0.05)** | | |

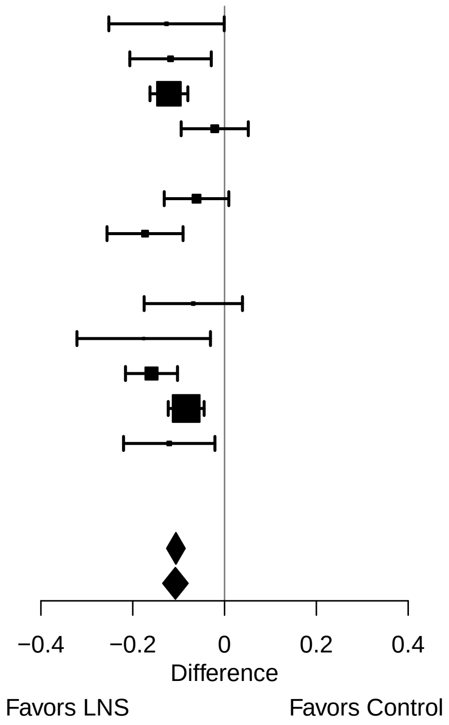

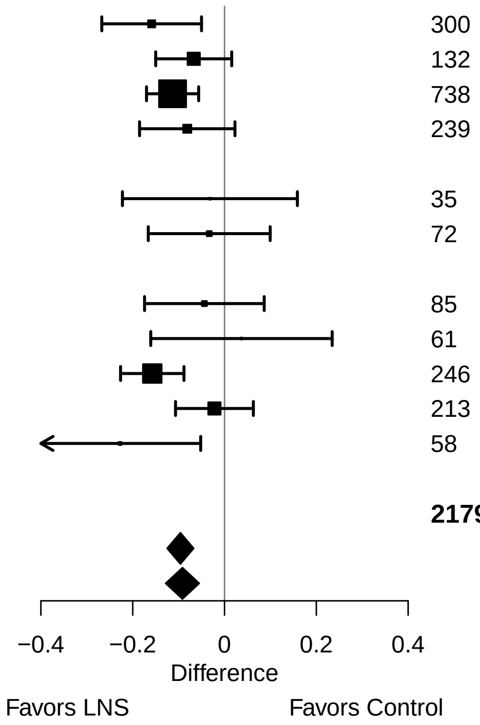

#### Supplemental figure 6C: Anemia prevalence difference

#### 6C2: Stratified by Maternal age

**P−for−interaction = 0.035**
**Difference in PDs = 0.03 (0.00, 0.06)**

| | | At least 25 y | | | | | | Less than 25 y | | | | | |
|---|---|---|---|---|---|---|---|---|---|---|---|---|---|
| Country | Trial | LNS N | Control N | Control Prevalence | PD (95% CI) | Fixed W | Random W | LNS N | Control N | Control Prevalence | PD (95% CI) | Fixed W | Random W |
| Bangladesh | JiVitA−4 (35) | 203 | 72 | 19.4 | −0.07 (−0.17, 0.03) | 0.04 | 0.06 | 251 | 73 | 12.3 | 0.01 (−0.08, 0.10) | 0.07 | 0.08 |
| Bangladesh | RDNS (36) | 152 | 81 | 40.7 | −0.16 (−0.31, −0.02) | 0.02 | 0.04 | 397 | 191 | 42.4 | −0.14 (−0.23, −0.05) | 0.07 | 0.07 |
| Bangladesh | WASH−B (37) | 89 | 83 | 14.5 | −0.09 (−0.19, 0.01) | 0.04 | 0.06 | 144 | 103 | 17.5 | −0.10 (−0.18, −0.02) | 0.09 | 0.09 |
| Burkina Faso | iLiNS−Zinc (38) | 1132 | 399 | 91.7 | −0.13 (−0.17, −0.09) | 0.24 | 0.13 | 812 | 262 | 90.1 | −0.10 (−0.17, −0.03) | 0.11 | 0.11 |
| Burkina Faso | PROMIS CS (39) | 271 | 263 | 67.3 | −0.07 (−0.16, 0.02) | 0.05 | 0.07 | 303 | 318 | 72.3 | −0.02 (−0.10, 0.06) | 0.08 | 0.09 |
| Ghana | GHANA (40) | 69 | 63 | 57.1 | −0.33 (−0.48, −0.17) | 0.02 | 0.03 | 29 | 25 | 64.0 | −0.23 (−0.49, 0.04) | 0.01 | 0.01 |
| Ghana | iLiNS−DYADG (41) | 206 | 397 | 42.3 | −0.08 (−0.17, 0.00) | 0.06 | 0.08 | 122 | 264 | 48.9 | −0.02 (−0.13, 0.09) | 0.05 | 0.05 |
| Kenya | WASH−B (42) | 213 | 168 | 45.2 | −0.13 (−0.21, −0.05) | 0.07 | 0.08 | 133 | 131 | 49.6 | −0.17 (−0.29, −0.05) | 0.04 | 0.04 |
| Madagascar | MAHAY (43) | 301 | 329 | 62.9 | −0.05 (−0.15, 0.05) | 0.04 | 0.06 | 299 | 259 | 67.2 | −0.10 (−0.19, −0.01) | 0.07 | 0.07 |
| Malawi | iLiNS−DYADM (44) | 99 | 220 | 47.3 | −0.02 (−0.14, 0.10) | 0.03 | 0.05 | 111 | 212 | 56.6 | −0.09 (−0.20, 0.03) | 0.04 | 0.05 |
| Malawi | iLiNS−DOSE (45) | 135 | 50 | 72.0 | −0.11 (−0.26, 0.04) | 0.02 | 0.04 | 104 | 28 | 71.4 | −0.08 (−0.28, 0.11) | 0.01 | 0.02 |
| Mali | PROMIS CS (46) | 502 | 508 | 87.8 | −0.18 (−0.25, −0.12) | 0.11 | 0.10 | 451 | 462 | 84.4 | −0.13 (−0.19, −0.06) | 0.13 | 0.12 |
| Zimbabwe | SHINE (HIV−) (47) | 808 | 726 | 33.7 | −0.10 (−0.14, −0.05) | 0.21 | 0.13 | 697 | 694 | 36.3 | −0.06 (−0.11, −0.01) | 0.21 | 0.16 |
| Zimbabwe | SHINE (HIV+) (48) | 221 | 204 | 37.7 | −0.16 (−0.26, −0.07) | 0.04 | 0.06 | 66 | 60 | 33.3 | −0.03 (−0.18, 0.12) | 0.02 | 0.03 |
| | | **4401** | **3563** | | **I² = 0.37, Tau² = 0.00** | | | **3919** | **3082** | | **I² = 0.23, Tau² = 0.00** | | |
| **Fixed** | | | | | **−0.12 (−0.14, −0.10)** | | | | | | **−0.08 (−0.10, −0.06)** | | |
| **Random** | | | | | **−0.12 (−0.15, −0.08)** | | | | | | **−0.08 (−0.11, −0.05)** | | |

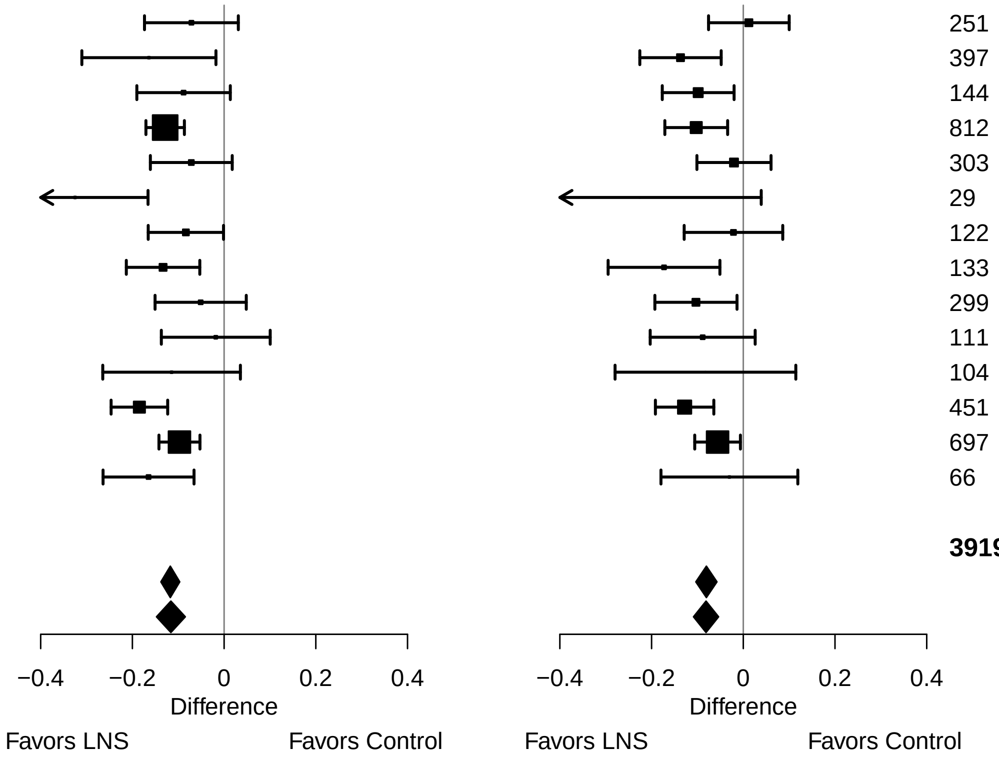

#### Supplemental figure 6C: Anemia prevalence difference

##### 6C3: Stratified by Maternal education

**P−for−interaction = 0.307**
**Difference in PDs = 0.02 (−0.02, 0.07)**

| Country | Trial | LNS N | Control N | Control Prevalence | Primary or greater PD (95% CI) | Fixed W | Random W | LNS N | Control N | Control Prevalence | Incomplete or no formal PD (95% CI) | Fixed W | Random W |
|---|---|---|---|---|---|---|---|---|---|---|---|---|---|
| Bangladesh | JiVitA−4 (35) | 299 | 93 | 17.2 | −0.06 (−0.16, 0.04) | 0.06 | 0.10 | 157 | 53 | 13.2 | 0.04 (−0.05, 0.13) | 0.10 | 0.11 |
| Bangladesh | RDNS (36) | 405 | 195 | 41.0 | −0.16 (−0.26, −0.05) | 0.05 | 0.09 | 144 | 77 | 44.2 | −0.11 (−0.27, 0.05) | 0.03 | 0.06 |
| Bangladesh | WASH−B (37) | 163 | 145 | 15.9 | −0.10 (−0.17, −0.03) | 0.12 | 0.14 | 71 | 41 | 17.1 | −0.09 (−0.21, 0.04) | 0.05 | 0.08 |
| Burkina Faso | iLiNS−Zinc (38) | | | | | | | | | | | | |
| Burkina Faso | PROMIS CS (39) | 46 | 47 | 70.2 | 0.04 (−0.14, 0.21) | 0.02 | 0.05 | 526 | 532 | 69.9 | −0.05 (−0.12, 0.02) | 0.19 | 0.14 |
| Ghana | GHANA (40) | | | | | | | | | | | | |
| Ghana | iLiNS−DYADG (41) | 253 | 526 | 45.4 | −0.08 (−0.15, 0.00) | 0.10 | 0.13 | 75 | 135 | 43.0 | 0.00 (−0.14, 0.14) | 0.04 | 0.07 |
| Kenya | WASH−B (42) | 168 | 136 | 50.0 | −0.27 (−0.37, −0.17) | 0.06 | 0.10 | 181 | 164 | 45.1 | −0.04 (−0.14, 0.06) | 0.09 | 0.10 |
| Madagascar | MAHAY (43) | 128 | 143 | 64.3 | −0.14 (−0.27, 0.00) | 0.03 | 0.07 | 472 | 445 | 64.9 | −0.06 (−0.15, 0.03) | 0.11 | 0.11 |
| Malawi | iLiNS−DYADM (44) | 34 | 66 | 48.5 | −0.10 (−0.31, 0.10) | 0.01 | 0.04 | 175 | 363 | 52.9 | −0.05 (−0.14, 0.04) | 0.11 | 0.11 |
| Malawi | iLiNS−DOSE (45) | | | | | | | | | | | | |
| Mali | PROMIS CS (46) | 107 | 96 | 84.4 | −0.09 (−0.20, 0.02) | 0.05 | 0.09 | 845 | 874 | 86.4 | −0.17 (−0.22, −0.11) | 0.26 | 0.15 |
| Zimbabwe | SHINE (HIV−) (47) | 1536 | 1462 | 34.9 | −0.07 (−0.10, −0.04) | 0.50 | 0.19 | 58 | 53 | 39.6 | −0.26 (−0.42, −0.10) | 0.03 | 0.06 |
| Zimbabwe | SHINE (HIV+) (48) | | | | | | | | | | | | |
| | | **3139** | **2909** | | **I² = 0.49, Tau² = 0.00** | | | **2704** | **2737** | | **I² = 0.60, Tau² = 0.00** | | |
| **Fixed** | | | | | **−0.09 (−0.11, −0.07)** | | | | | | **−0.08 (−0.11, −0.05)** | | |
| **Random** | | | | | **−0.10 (−0.15, −0.06)** | | | | | | **−0.07 (−0.12, −0.02)** | | |

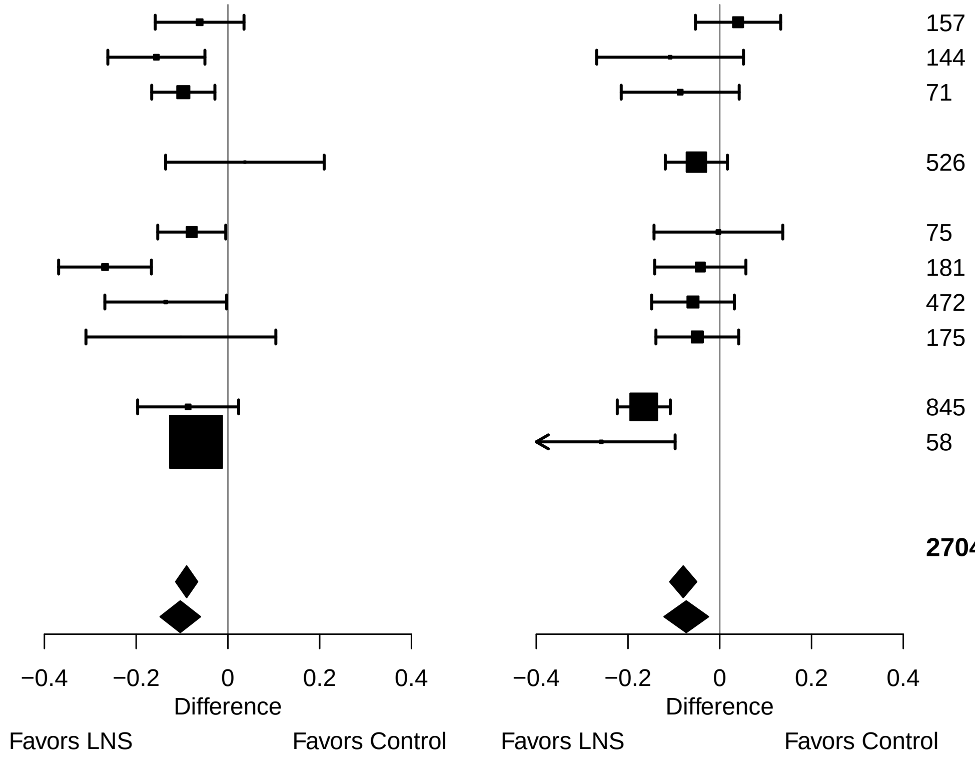

##### Supplemental figure 6C: Anemia prevalence difference

#### 6C4: Stratified by Child sex

**P−for−interaction = 0.283**
**Difference in PDs = −0.02 (−0.04, 0.01)**

**Male**

| Country | Trial | LNS N | Control N | Control Prevalence | PD (95% CI) | Fixed W | Random W |
|---|---|---|---|---|---|---|---|
| Bangladesh | JiVitA−4 (35) | 235 | 71 | 22.5 | −0.12 (−0.22, −0.02) | 0.04 | 0.06 |
| Bangladesh | RDNS (36) | 266 | 131 | 41.2 | −0.16 (−0.29, −0.02) | 0.02 | 0.04 |
| Bangladesh | WASH−B (37) | 116 | 89 | 16.9 | −0.12 (−0.22, −0.02) | 0.04 | 0.06 |
| Burkina Faso | iLiNS−Zinc (38) | 993 | 334 | 91.6 | −0.11 (−0.14, −0.07) | 0.32 | 0.15 |
| Burkina Faso | PROMIS CS (39) | 317 | 295 | 77.3 | −0.05 (−0.14, 0.04) | 0.05 | 0.08 |
| Ghana | GHANA (40) | 60 | 41 | 61.0 | −0.33 (−0.51, −0.14) | 0.01 | 0.02 |
| Ghana | iLiNS−DYADG (41) | 164 | 310 | 44.8 | −0.02 (−0.11, 0.08) | 0.05 | 0.07 |
| Kenya | WASH−B (42) | 168 | 145 | 44.8 | −0.09 (−0.19, 0.02) | 0.04 | 0.06 |
| Madagascar | MAHAY (43) | 277 | 288 | 65.6 | −0.03 (−0.12, 0.06) | 0.05 | 0.07 |
| Malawi | iLiNS−DYADM (44) | 103 | 202 | 56.4 | −0.03 (−0.15, 0.09) | 0.03 | 0.05 |
| Malawi | iLiNS−DOSE (45) | 123 | 43 | 76.7 | −0.12 (−0.28, 0.04) | 0.02 | 0.03 |
| Mali | PROMIS CS (46) | 479 | 526 | 89.0 | −0.14 (−0.20, −0.08) | 0.11 | 0.11 |
| Zimbabwe | SHINE (HIV−) (47) | 830 | 814 | 36.2 | −0.07 (−0.12, −0.02) | 0.17 | 0.13 |
| Zimbabwe | SHINE (HIV+) (48) | 146 | 149 | 40.9 | −0.16 (−0.25, −0.06) | 0.04 | 0.07 |
| | | **4277** | **3438** | | **I² = 0.32, Tau² = 0.00** | | |
| **Fixed** | | | | | **−0.10 (−0.12, −0.08)** | | |
| **Random** | | | | | **−0.10 (−0.13, −0.07)** | | |

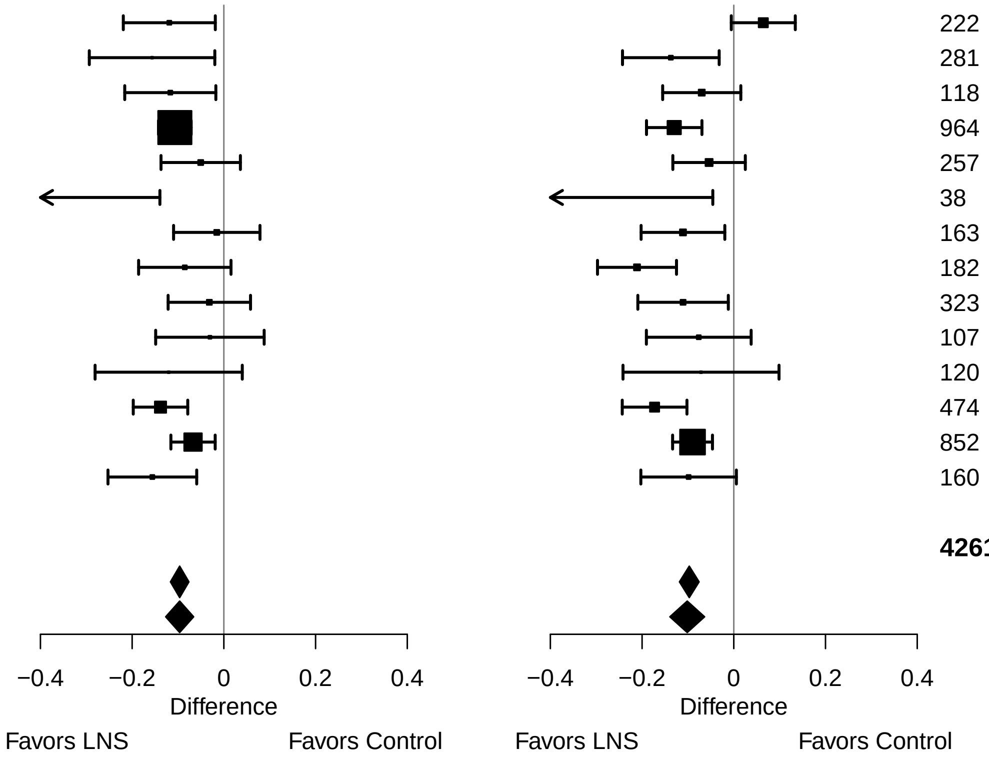

**Female**

| LNS N | Control N | Control Prevalence | PD (95% CI) | Fixed W | Random W |
|---|---|---|---|---|---|
| 222 | 75 | 9.3 | 0.06 (−0.01, 0.13) | 0.09 | 0.09 |
| 281 | 141 | 42.6 | −0.14 (−0.24, −0.03) | 0.04 | 0.06 |
| 118 | 97 | 15.5 | −0.07 (−0.16, 0.02) | 0.06 | 0.08 |
| 964 | 330 | 90.6 | −0.13 (−0.19, −0.07) | 0.13 | 0.09 |
| 257 | 286 | 62.6 | −0.05 (−0.13, 0.03) | 0.07 | 0.08 |
| 38 | 55 | 56.4 | −0.25 (−0.45, −0.05) | 0.01 | 0.03 |
| 163 | 348 | 44.8 | −0.11 (−0.20, −0.02) | 0.06 | 0.07 |
| 182 | 155 | 49.7 | −0.21 (−0.30, −0.12) | 0.06 | 0.08 |
| 323 | 300 | 64.0 | −0.11 (−0.21, −0.01) | 0.05 | 0.07 |
| 107 | 230 | 47.8 | −0.08 (−0.19, 0.04) | 0.04 | 0.06 |
| 120 | 39 | 66.7 | −0.07 (−0.24, 0.10) | 0.02 | 0.04 |
| 474 | 444 | 82.9 | −0.17 (−0.24, −0.10) | 0.09 | 0.09 |
| 852 | 780 | 34.2 | −0.09 (−0.13, −0.05) | 0.24 | 0.11 |
| 160 | 136 | 32.4 | −0.10 (−0.20, 0.01) | 0.04 | 0.06 |
| **4261** | **3416** | | **I² = 0.65, Tau² = 0.00** | | |
| | | | **−0.10 (−0.12, −0.08)** | | |
| | | | **−0.10 (−0.14, −0.06)** | | |

#### Supplemental figure 6C: Anemia prevalence difference

#### 6C5: Stratified by Child birth order

**P−for−interaction = 0.024**
**Difference in PDs = 0.04 (0.01, 0.07)**

| | | Later born | | | | | | Firstborn | | | | | |
|---|---|---|---|---|---|---|---|---|---|---|---|---|---|
| Country | Trial | LNS N | Control N | Control Prevalence | PD (95% CI) | Fixed W | Random W | LNS N | Control N | Control Prevalence | PD (95% CI) | Fixed W | Random W |
| Bangladesh | JiVitA−4 (35) | 115 | 33 | 27.3 | −0.12 (−0.29, 0.04) | 0.01 | 0.03 | 341 | 112 | 12.5 | 0.00 (−0.06, 0.06) | 0.20 | 0.14 |
| Bangladesh | RDNS (36) | 339 | 171 | 40.9 | −0.14 (−0.24, −0.04) | 0.03 | 0.06 | 210 | 101 | 43.6 | −0.15 (−0.28, −0.02) | 0.04 | 0.07 |
| Bangladesh | WASH−B (37) | 137 | 123 | 16.3 | −0.11 (−0.20, −0.03) | 0.05 | 0.07 | 95 | 61 | 16.4 | −0.08 (−0.20, 0.04) | 0.05 | 0.07 |
| Burkina Faso | iLiNS−Zinc (38) | 1522 | 520 | 90.4 | −0.12 (−0.16, −0.07) | 0.15 | 0.12 | 435 | 144 | 93.8 | −0.13 (−0.18, −0.08) | 0.29 | 0.16 |
| Burkina Faso | PROMIS CS (39) | 498 | 477 | 70.0 | −0.05 (−0.12, 0.02) | 0.07 | 0.09 | 75 | 103 | 69.9 | −0.01 (−0.16, 0.15) | 0.03 | 0.05 |
| Ghana | GHANA (40) | 60 | 54 | 61.1 | −0.38 (−0.55, −0.21) | 0.01 | 0.03 | 38 | 38 | 55.3 | −0.16 (−0.38, 0.07) | 0.01 | 0.03 |
| Ghana | iLiNS−DYADG (41) | 220 | 435 | 45.3 | −0.11 (−0.19, −0.03) | 0.06 | 0.08 | 108 | 226 | 44.2 | 0.03 (−0.08, 0.14) | 0.05 | 0.08 |
| Kenya | WASH−B (42) | 282 | 244 | 47.1 | −0.16 (−0.24, −0.08) | 0.05 | 0.08 | 67 | 56 | 48.2 | −0.12 (−0.32, 0.08) | 0.02 | 0.03 |
| Madagascar | MAHAY (43) | 458 | 454 | 66.1 | −0.07 (−0.15, 0.02) | 0.05 | 0.07 | 141 | 131 | 61.1 | −0.10 (−0.23, 0.03) | 0.04 | 0.06 |
| Malawi | iLiNS−DYADM (44) | 165 | 342 | 50.9 | −0.09 (−0.18, 0.00) | 0.04 | 0.07 | 45 | 88 | 55.7 | 0.09 (−0.09, 0.27) | 0.02 | 0.04 |
| Malawi | iLiNS−DOSE (45) | | | | | | | | | | | | |
| Mali | PROMIS CS (46) | 790 | 832 | 86.1 | −0.16 (−0.21, −0.10) | 0.12 | 0.11 | 146 | 126 | 88.1 | −0.16 (−0.26, −0.06) | 0.07 | 0.09 |
| Zimbabwe | SHINE (HIV−) (47) | 1188 | 1100 | 35.5 | −0.10 (−0.14, −0.07) | 0.31 | 0.14 | 418 | 416 | 33.2 | −0.02 (−0.09, 0.04) | 0.16 | 0.13 |
| Zimbabwe | SHINE (HIV+) (48) | 252 | 216 | 36.6 | −0.14 (−0.22, −0.05) | 0.05 | 0.07 | 51 | 64 | 37.5 | −0.08 (−0.26, 0.09) | 0.02 | 0.04 |
| | | **6026** | **5001** | | **I² = 0.35, Tau² = 0.00** | | | **2170** | **1666** | | **I² = 0.52, Tau² = 0.00** | | |
| **Fixed** | | | | | **−0.12 (−0.13, −0.10)** | | | | | | **−0.07 (−0.09, −0.04)** | | |
| **Random** | | | | | **−0.12 (−0.15, −0.09)** | | | | | | **−0.07 (−0.11, −0.03)** | | |

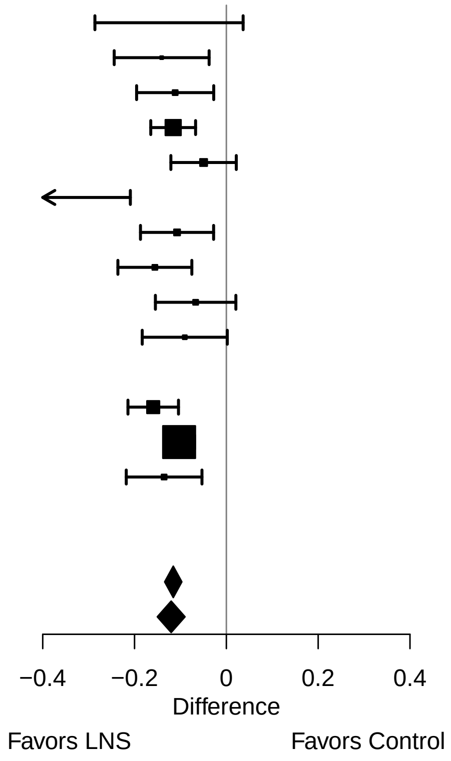

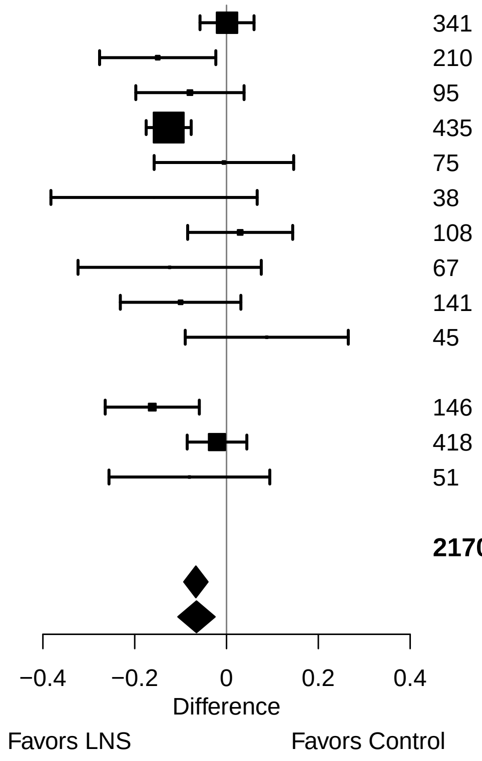

#### Supplemental figure 6C: Anemia prevalence difference

##### 6C6: Stratified by Child baseline acute malnutrition

**P−for−interaction = 0.595**
**Difference in PDs = −0.01 (−0.06, 0.04)**

| Country | Trial | No: LNS N | No: Control N | No: Control Prevalence | No: PD (95% CI) | No: Fixed W | No: Random W | Yes: LNS N | Yes: Control N | Yes: Control Prevalence | Yes: PD (95% CI) | Yes: Fixed W | Yes: Random W |
|---|---|---|---|---|---|---|---|---|---|---|---|---|---|
| Bangladesh | JiVitA−4 (35) | 365 | 114 | 15.8 | −0.03 (−0.09, 0.03) | 0.13 | 0.15 | 90 | 32 | 15.6 | 0.00 (−0.14, 0.14) | 0.11 | 0.18 |
| Bangladesh | RDNS (36) | 493 | 243 | 40.3 | −0.14 (−0.23, −0.05) | 0.06 | 0.08 | 35 | 12 | 41.7 | −0.07 (−0.45, 0.31) | 0.01 | 0.03 |
| Bangladesh | WASH−B (37) | | | | | | | | | | | | |
| Burkina Faso | iLiNS−Zinc (38) | 1457 | 487 | 90.3 | −0.11 (−0.16, −0.07) | 0.24 | 0.22 | 500 | 177 | 93.2 | −0.14 (−0.19, −0.08) | 0.72 | 0.47 |
| Burkina Faso | PROMIS CS (39) | | | | | | | | | | | | |
| Ghana | GHANA (40) | | | | | | | | | | | | |
| Ghana | iLiNS−DYADG (41) | 285 | 556 | 46.4 | −0.07 (−0.15, 0.00) | 0.10 | 0.12 | 21 | 54 | 31.5 | 0.02 (−0.22, 0.26) | 0.04 | 0.08 |
| Kenya | WASH−B (42) | | | | | | | | | | | | |
| Madagascar | MAHAY (43) | | | | | | | | | | | | |
| Malawi | iLiNS−DYADM (44) | 183 | 379 | 51.5 | −0.05 (−0.14, 0.04) | 0.06 | 0.08 | 14 | 37 | 51.4 | 0.06 (−0.25, 0.37) | 0.02 | 0.05 |
| Malawi | iLiNS−DOSE (45) | | | | | | | | | | | | |
| Mali | PROMIS CS (46) | | | | | | | | | | | | |
| Zimbabwe | SHINE (HIV−) (47) | 1185 | 1069 | 32.8 | −0.07 (−0.10, −0.03) | 0.34 | 0.26 | 66 | 67 | 32.8 | −0.09 (−0.25, 0.07) | 0.08 | 0.15 |
| Zimbabwe | SHINE (HIV+) (48) | 226 | 202 | 34.7 | −0.13 (−0.21, −0.05) | 0.07 | 0.09 | 26 | 13 | 61.5 | −0.38 (−0.73, −0.04) | 0.02 | 0.04 |
| | | **4194** | **3050** | | **I² = 0.28, Tau² = 0.00** | | | **752** | **392** | | **I² = 0.24, Tau² = 0.00** | | |
| **Fixed** | | | | | **−0.08 (−0.10, −0.06)** | | | | | | **−0.11 (−0.16, −0.06)** | | |
| **Random** | | | | | **−0.08 (−0.11, −0.05)** | | | | | | **−0.09 (−0.16, −0.02)** | | |

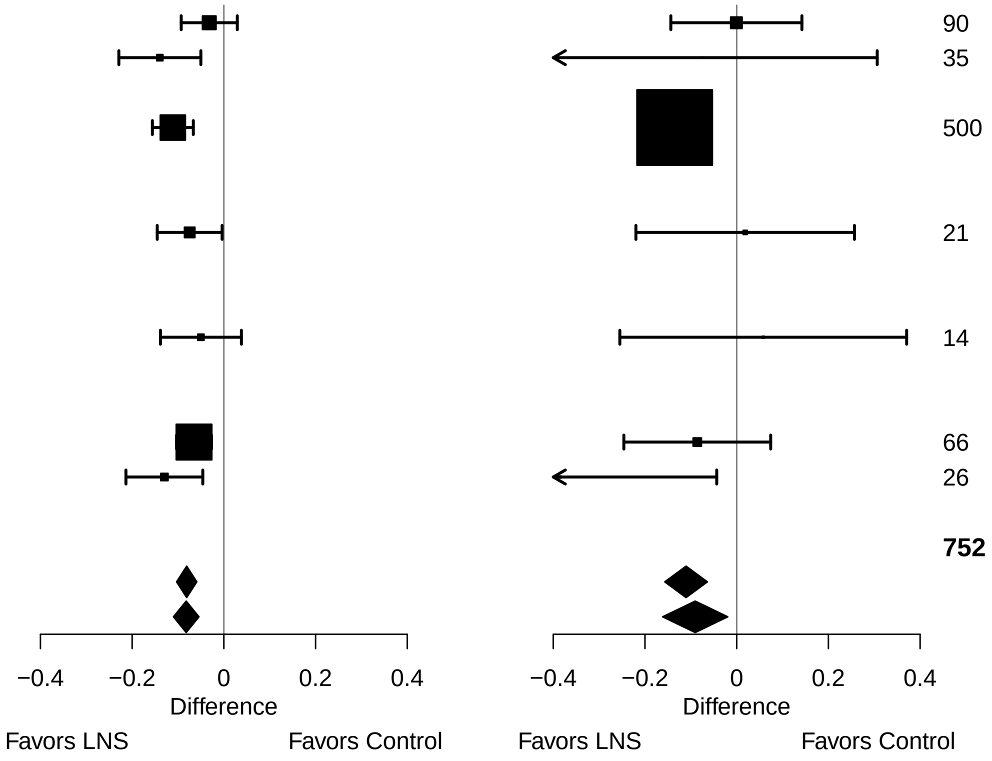

**Supplemental figure 6C: Anemia prevalence difference**

#### 6C7: Stratified by Child baseline anemia

**P−for−interaction = 0.308**
**Difference in PDs = −0.04 (−0.10, 0.03)**

| Country | Trial | LNS N | Control N | Control Prevalence | Not anemic PD (95% CI) | Fixed W | Random W | LNS N | Control N | Control Prevalence | Anemic PD (95% CI) | Fixed W | Random W |
|---|---|---|---|---|---|---|---|---|---|---|---|---|---|
| Bangladesh | JiVitA−4 (35) | | | | | | | | | | | | |
| Bangladesh | RDNS (36) | 213 | 93 | 23.7 | −0.09 (−0.21, 0.02) | 0.20 | 0.21 | 314 | 160 | 50.0 | −0.15 (−0.25, −0.04) | 0.11 | 0.11 |
| Bangladesh | WASH−B (37) | | | | | | | | | | | | |
| Burkina Faso | iLiNS−Zinc (38) | 177 | 61 | 82.0 | −0.17 (−0.28, −0.06) | 0.21 | 0.22 | 1780 | 603 | 92.0 | −0.11 (−0.16, −0.07) | 0.62 | 0.62 |
| Burkina Faso | PROMIS CS (39) | | | | | | | | | | | | |
| Ghana | GHANA (40) | | | | | | | | | | | | |
| Ghana | iLiNS−DYADG (41) | 189 | 355 | 32.7 | 0.00 (−0.08, 0.08) | 0.38 | 0.29 | 90 | 207 | 64.7 | −0.13 (−0.25, 0.00) | 0.09 | 0.09 |
| Kenya | WASH−B (42) | | | | | | | | | | | | |
| Madagascar | MAHAY (43) | | | | | | | | | | | | |
| Malawi | iLiNS−DYADM (44) | 66 | 145 | 36.6 | 0.01 (−0.13, 0.15) | 0.13 | 0.16 | 134 | 269 | 59.1 | −0.07 (−0.17, 0.03) | 0.13 | 0.13 |
| Malawi | iLiNS−DOSE (45) | 101 | 45 | 64.4 | −0.10 (−0.27, 0.07) | 0.09 | 0.12 | 142 | 37 | 81.1 | −0.10 (−0.26, 0.06) | 0.05 | 0.05 |
| Mali | PROMIS CS (46) | | | | | | | | | | | | |
| Zimbabwe | SHINE (HIV−) (47) | | | | | | | | | | | | |
| Zimbabwe | SHINE (HIV+) (48) | | | | | | | | | | | | |
| | | **746** | **699** | | **I² = 0.44, Tau² = 0.00** | | | **2460** | **1276** | | **I² = 0.00, Tau² = 0.00** | | |
| **Fixed** | | | | | **−0.06 (−0.11, −0.01)** | | | | | | **−0.11 (−0.15, −0.08)** | | |
| **Random** | | | | | **−0.06 (−0.13, 0.00)** | | | | | | **−0.11 (−0.15, −0.08)** | | |

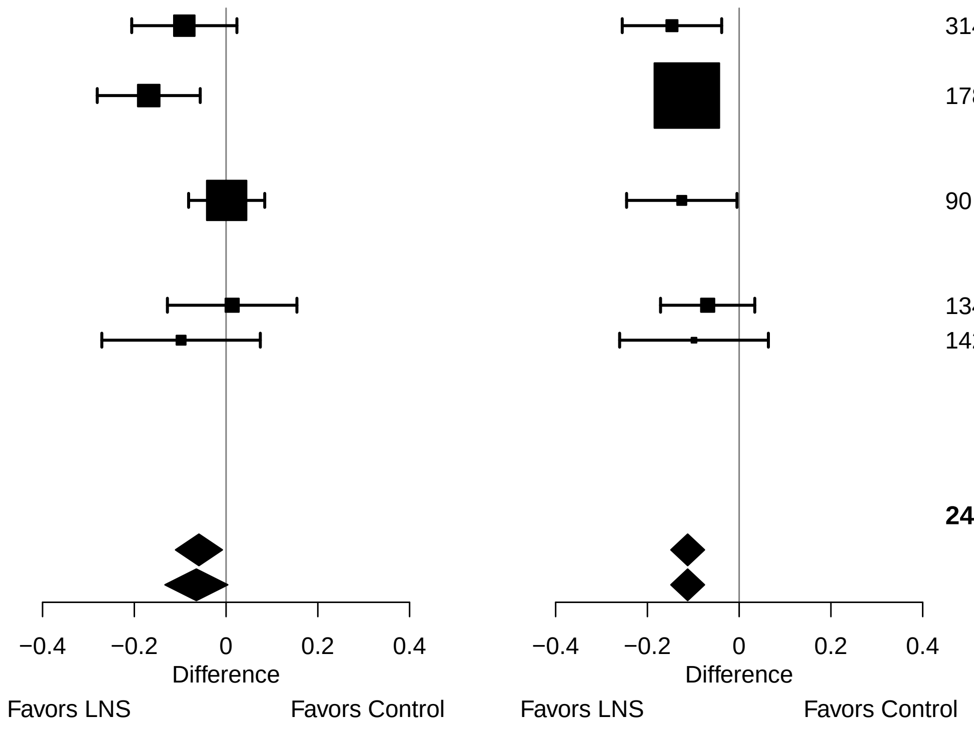

#### Supplemental figure 6C: Anemia prevalence difference

##### 6C8: Stratified by Child high-dose vitamin A supplementation

**P−for−interaction = 0.549**
**Difference in PDs = −0.01 (−0.06, 0.03)**

| | | Received VitA | | | | | | | Not treated | | | | |
|---|---|---|---|---|---|---|---|---|---|---|---|---|---|
| | | **LNS** | **Control** | **Control** | **PD** | **Fixed** | **Random** | **LNS** | **Control** | **Control** | **PD** | **Fixed** | **Random** |
| Country | Trial | N | N | Prevalence | (95% CI) | W | W | N | N | Prevalence | (95% CI) | W | W |
| Bangladesh | JiVitA−4 (35) | 141 | 37 | 16.2 | −0.01 (−0.16, 0.14) | 0.05 | 0.10 | 133 | 41 | 14.6 | −0.02 (−0.15, 0.11) | 0.06 | 0.10 |
| Bangladesh | RDNS (36) | 444 | 223 | 40.8 | −0.14 (−0.24, −0.03) | 0.10 | 0.14 | 105 | 49 | 46.9 | −0.18 (−0.33, −0.04) | 0.05 | 0.08 |
| Bangladesh | WASH−B (37) | | | | | | | | | | | | |
| Burkina Faso | iLiNS−Zinc (38) | | | | | | | | | | | | |
| Burkina Faso | PROMIS CS (39) | | | | | | | | | | | | |
| Ghana | GHANA (40) | 50 | 62 | 56.5 | −0.34 (−0.52, −0.17) | 0.03 | 0.08 | 48 | 29 | 65.5 | −0.28 (−0.50, −0.06) | 0.02 | 0.04 |
| Ghana | iLiNS−DYADG (41) | 215 | 412 | 45.6 | −0.06 (−0.14, 0.02) | 0.15 | 0.17 | 113 | 249 | 43.8 | −0.07 (−0.18, 0.04) | 0.09 | 0.12 |
| Kenya | WASH−B (42) | 29 | 34 | 52.9 | −0.18 (−0.43, 0.06) | 0.02 | 0.05 | 263 | 212 | 46.7 | −0.15 (−0.23, −0.07) | 0.15 | 0.16 |
| Madagascar | MAHAY (43) | | | | | | | | | | | | |
| Malawi | iLiNS−DYADM (44) | 75 | 136 | 58.8 | −0.13 (−0.27, 0.01) | 0.05 | 0.11 | 133 | 289 | 48.4 | −0.01 (−0.11, 0.09) | 0.10 | 0.13 |
| Malawi | iLiNS−DOSE (45) | | | | | | | | | | | | |
| Mali | PROMIS CS (46) | | | | | | | | | | | | |
| Zimbabwe | SHINE (HIV−) (47) | 874 | 833 | 33.7 | −0.06 (−0.10, −0.01) | 0.51 | 0.20 | 665 | 622 | 35.9 | −0.10 (−0.15, −0.05) | 0.45 | 0.24 |
| Zimbabwe | SHINE (HIV+) (48) | 165 | 127 | 37.0 | −0.15 (−0.26, −0.05) | 0.09 | 0.14 | 112 | 134 | 36.6 | −0.13 (−0.24, −0.02) | 0.09 | 0.12 |
| | | **1993** | **1864** | | **$I^2$ = 0.54, $Tau^2$ = 0.00** | | | **1572** | **1625** | | **$I^2$ = 0.32, $Tau^2$ = 0.00** | | |
| **Fixed** | | | | | **−0.09 (−0.12, −0.06)** | | | | | | **−0.10 (−0.13, −0.07)** | | |
| **Random** | | | | | **−0.12 (−0.18, −0.05)** | | | | | | **−0.10 (−0.15, −0.06)** | | |

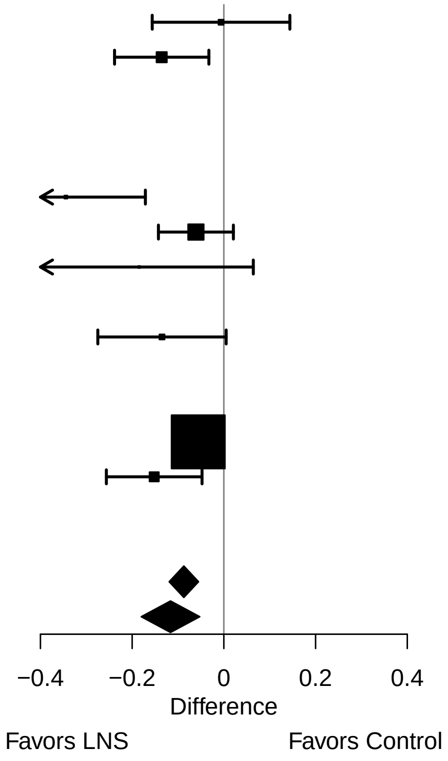

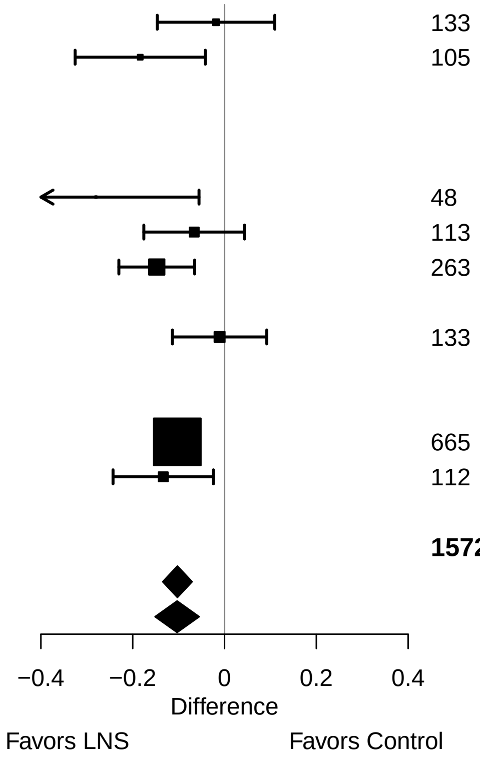

#### Supplemental figure 6C: Anemia prevalence difference

#### 6C9: Stratified by Child inflammation

**P−for−interaction = 0.068**
**Difference in PDs = 0.06 (0.00, 0.12)**

| | | Not inflamed | | | | | | High AGP or CRP | | | | | |
|---|---|---|---|---|---|---|---|---|---|---|---|---|---|
| | | **LNS** | **Control** | **Control** | **PD** | **Fixed** | **Random** | **LNS** | **Control** | **Control** | **PD** | **Fixed** | **Random** |
| Country | Trial | N | N | Prevalence | (95% CI) | W | W | N | N | Prevalence | (95% CI) | W | W |
| Bangladesh | JiVitA−4 (35) | 185 | 47 | 17.0 | −0.06 (−0.20, 0.09) | 0.07 | 0.10 | 269 | 98 | 14.3 | 0.00 (−0.07, 0.07) | 0.35 | 0.35 |
| Bangladesh | RDNS (36) | 369 | 168 | 42.3 | −0.18 (−0.29, −0.07) | 0.12 | 0.14 | 180 | 104 | 41.3 | −0.07 (−0.19, 0.05) | 0.12 | 0.12 |
| Bangladesh | WASH−B (37) | | | | | | | | | | | | |
| Burkina Faso | iLiNS−Zinc (38) | | | | | | | | | | | | |
| Burkina Faso | PROMIS CS (39) | | | | | | | | | | | | |
| Ghana | GHANA (40) | | | | | | | | | | | | |
| Ghana | iLiNS−DYADG (41) | 45 | 120 | 46.7 | −0.04 (−0.22, 0.13) | 0.05 | 0.08 | 55 | 82 | 53.7 | −0.10 (−0.27, 0.07) | 0.06 | 0.06 |
| Kenya | WASH−B (42) | 157 | 135 | 43.0 | −0.23 (−0.33, −0.12) | 0.13 | 0.15 | 141 | 123 | 52.8 | −0.07 (−0.19, 0.05) | 0.12 | 0.12 |
| Madagascar | MAHAY (43) | 44 | 32 | 43.8 | −0.26 (−0.44, −0.07) | 0.04 | 0.07 | 39 | 19 | 47.4 | −0.19 (−0.44, 0.06) | 0.03 | 0.03 |
| Malawi | iLiNS−DYADM (44) | 59 | 123 | 40.7 | −0.17 (−0.32, −0.02) | 0.07 | 0.10 | 138 | 269 | 57.6 | −0.04 (−0.14, 0.06) | 0.17 | 0.17 |
| Malawi | iLiNS−DOSE (45) | | | | | | | | | | | | |
| Mali | PROMIS CS (46) | | | | | | | | | | | | |
| Zimbabwe | SHINE (HIV−) (47) | 414 | 354 | 28.8 | −0.05 (−0.11, 0.02) | 0.35 | 0.21 | 105 | 99 | 43.4 | −0.12 (−0.24, 0.00) | 0.12 | 0.12 |
| Zimbabwe | SHINE (HIV+) (48) | 186 | 158 | 35.4 | −0.16 (−0.25, −0.06) | 0.17 | 0.16 | 32 | 31 | 41.9 | −0.14 (−0.38, 0.11) | 0.03 | 0.03 |
| | | **1459** | **1137** | | **I² = 0.53, Tau² = 0.00** | | | **959** | **825** | | **I² = 0.00, Tau² = 0.00** | | |
| **Fixed** | | | | | **−0.12 (−0.16, −0.08)** | | | | | | **−0.05 (−0.10, −0.01)** | | |
| **Random** | | | | | **−0.14 (−0.19, −0.08)** | | | | | | **−0.05 (−0.10, −0.01)** | | |

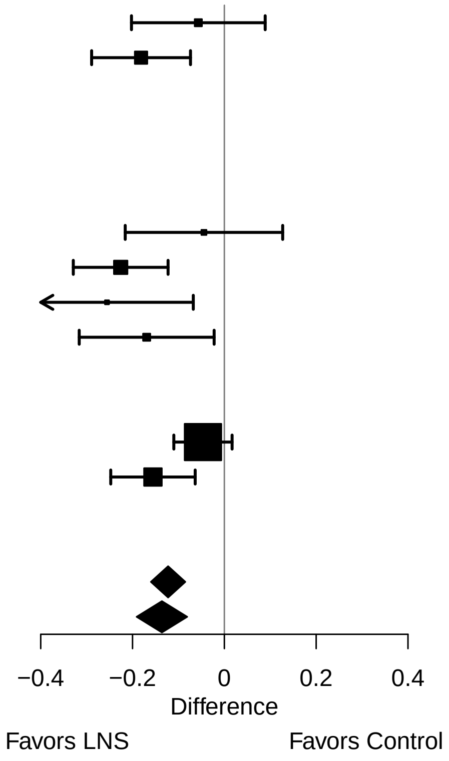

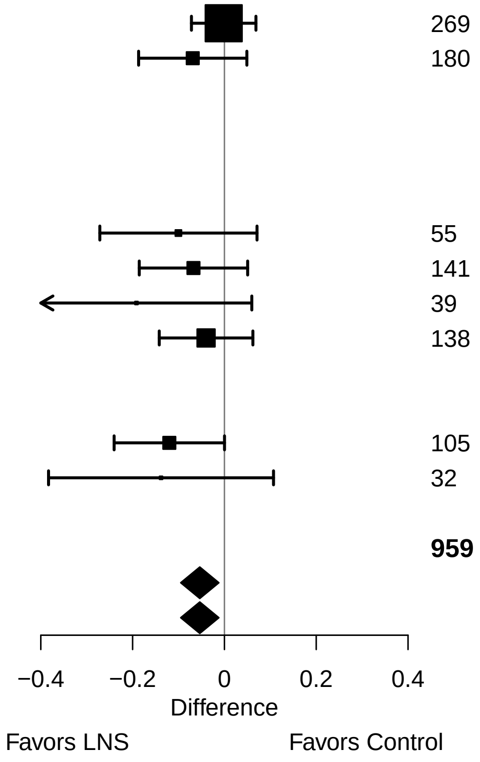

### Supplemental figure 6D: Moderate-to-severe anemia prevalence ratio

#### 6D1: Stratified by Maternal BMI

**P−for−interaction = 0.459**
**Ratio of PRs = 1.03 (0.96, 1.10)**

| | | At least 20 kg/m² | | | | | | Less than 20 kg/m² | | | | | |
|---|---|---|---|---|---|---|---|---|---|---|---|---|---|
| Country | Trial | LNS N | Control N | Control Prevalence | PR (95% CI) | Fixed W | Random W | LNS N | Control N | Control Prevalence | PR (95% CI) | Fixed W | Random W |
| Bangladesh | JiVitA−4 (35) | | | | | | | | | | | | |
| Bangladesh | RDNS (36) | 227 | 102 | 20.6 | 0.47 (0.28, 0.80) | 0.02 | 0.07 | 300 | 160 | 13.8 | 0.61 (0.37, 0.99) | 0.03 | 0.09 |
| Bangladesh | WASH−B (37) | | | | | | | | | | | | |
| Burkina Faso | iLiNS−Zinc (38) | 1217 | 390 | 76.2 | 0.71 (0.65, 0.77) | 0.59 | 0.17 | 738 | 274 | 73.4 | 0.72 (0.65, 0.81) | 0.51 | 0.22 |
| Burkina Faso | PROMIS CS (39) | 335 | 349 | 36.7 | 0.96 (0.77, 1.20) | 0.09 | 0.14 | 239 | 227 | 37.0 | 0.77 (0.57, 1.03) | 0.07 | 0.15 |
| Ghana | GHANA (40) | | | | | | | | | | | | |
| Ghana | iLiNS−DYADG (41) | | | | | | | | | | | | |
| Kenya | WASH−B (42) | 266 | 227 | 22.9 | 0.54 (0.36, 0.81) | 0.03 | 0.10 | 72 | 60 | 11.7 | 1.43 (0.67, 3.05) | 0.01 | 0.05 |
| Madagascar | MAHAY (43) | | | | | | | | | | | | |
| Malawi | iLiNS−DYADM (44) | 124 | 258 | 24.0 | 1.14 (0.80, 1.63) | 0.03 | 0.11 | 85 | 171 | 23.4 | 1.11 (0.71, 1.74) | 0.03 | 0.10 |
| Malawi | iLiNS−DOSE (45) | 179 | 52 | 46.2 | 0.60 (0.42, 0.85) | 0.04 | 0.11 | 61 | 29 | 48.3 | 0.97 (0.62, 1.52) | 0.03 | 0.10 |
| Mali | PROMIS CS (46) | 702 | 685 | 60.0 | 0.63 (0.53, 0.76) | 0.14 | 0.16 | 246 | 279 | 61.3 | 0.63 (0.54, 0.73) | 0.30 | 0.21 |
| Zimbabwe | SHINE (HIV−) (47) | 1222 | 1161 | 10.9 | 0.64 (0.50, 0.83) | 0.07 | 0.14 | 213 | 220 | 6.8 | 1.24 (0.66, 2.31) | 0.02 | 0.07 |
| Zimbabwe | SHINE (HIV+) (48) | | | | | | | | | | | | |
| | | **4272** | **3224** | | **I² = 0.66, Tau² = 0.05** | | | **1954** | **1420** | | **I² = 0.53, Tau² = 0.04** | | |
| **Fixed** | | | | | **0.71 (0.66, 0.76)** | | | | | | **0.72 (0.67, 0.78)** | | |
| **Random** | | | | | **0.70 (0.58, 0.85)** | | | | | | **0.80 (0.67, 0.97)** | | |

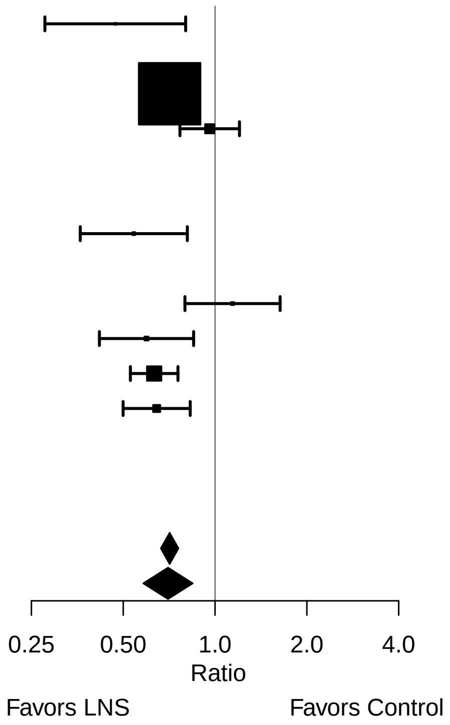

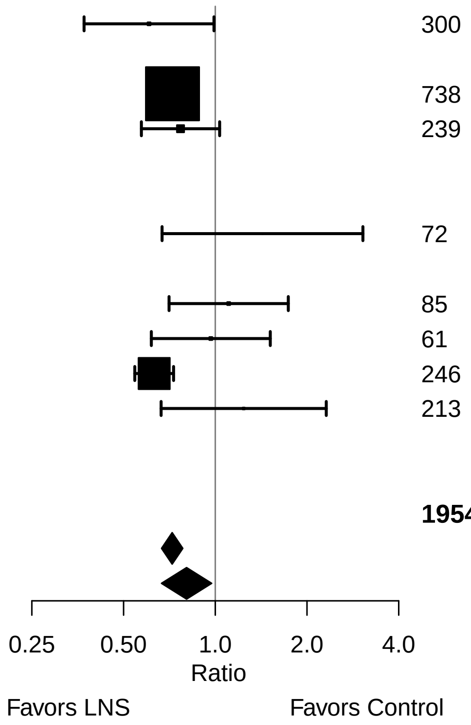

##### Supplemental figure 6D: Moderate-to-severe anemia prevalence ratio

#### 6D2: Stratified by Maternal age

**P−for−interaction = 0.260**
**Ratio of PRs = 1.06 (0.96, 1.18)**

| Country | Trial | At least 25 y: LNS N | Control N | Control Prevalence | PR (95% CI) | Fixed W | Random W | Less than 25 y: LNS N | Control N | Control Prevalence | PR (95% CI) | Fixed W | Random W |
|---|---|---|---|---|---|---|---|---|---|---|---|---|---|
| Bangladesh | JiVitA−4 (35) | | | | | | | | | | | | |
| Bangladesh | RDNS (36) | 152 | 81 | 17.3 | 0.46 (0.20, 1.04) | 0.01 | 0.03 | 397 | 191 | 16.2 | 0.57 (0.39, 0.85) | 0.04 | 0.04 |
| Bangladesh | WASH−B (37) | | | | | | | | | | | | |
| Burkina Faso | iLiNS−Zinc (38) | 1132 | 399 | 74.2 | 0.69 (0.61, 0.79) | 0.41 | 0.17 | 812 | 262 | 76.0 | 0.74 (0.67, 0.83) | 0.48 | 0.48 |
| Burkina Faso | PROMIS CS (39) | 271 | 263 | 32.7 | 0.89 (0.68, 1.17) | 0.09 | 0.12 | 303 | 318 | 39.9 | 0.88 (0.68, 1.15) | 0.08 | 0.08 |
| Ghana | GHANA (40) | | | | | | | | | | | | |
| Ghana | iLiNS−DYADG (41) | 206 | 397 | 5.0 | 0.77 (0.35, 1.72) | 0.01 | 0.03 | 122 | 264 | 6.1 | 0.95 (0.40, 2.24) | 0.01 | 0.01 |
| Kenya | WASH−B (42) | 213 | 168 | 20.8 | 0.59 (0.37, 0.93) | 0.03 | 0.07 | 133 | 131 | 22.1 | 0.68 (0.42, 1.09) | 0.02 | 0.02 |
| Madagascar | MAHAY (43) | 301 | 329 | 30.7 | 0.82 (0.59, 1.14) | 0.07 | 0.11 | 299 | 259 | 35.5 | 0.79 (0.61, 1.02) | 0.08 | 0.08 |
| Malawi | iLiNS−DYADM (44) | 99 | 220 | 21.8 | 1.34 (0.90, 1.99) | 0.05 | 0.09 | 111 | 212 | 25.5 | 0.99 (0.67, 1.47) | 0.04 | 0.04 |
| Malawi | iLiNS−DOSE (45) | 135 | 50 | 46.0 | 0.70 (0.49, 1.01) | 0.05 | 0.10 | 104 | 28 | 46.4 | 0.74 (0.46, 1.16) | 0.03 | 0.03 |
| Mali | PROMIS CS (46) | 502 | 508 | 61.4 | 0.61 (0.50, 0.73) | 0.21 | 0.15 | 451 | 462 | 58.9 | 0.67 (0.56, 0.80) | 0.17 | 0.17 |
| Zimbabwe | SHINE (HIV−) (47) | 808 | 726 | 9.4 | 0.65 (0.45, 0.92) | 0.06 | 0.10 | 697 | 694 | 12.1 | 0.72 (0.53, 0.98) | 0.06 | 0.06 |
| Zimbabwe | SHINE (HIV+) (48) | 221 | 204 | 9.8 | 0.42 (0.19, 0.93) | 0.01 | 0.03 | 66 | 60 | 10.0 | 0.91 (0.32, 2.57) | 0.01 | 0.01 |
| | | **4040** | **3345** | | **$I^2$ = 0.50, $Tau^2$ = 0.04** | | | **3495** | **2881** | | **$I^2$ = 0.00, $Tau^2$ = 0.00** | | |
| **Fixed** | | | | | **0.71 (0.65, 0.77)** | | | | | | **0.74 (0.69, 0.80)** | | |
| **Random** | | | | | **0.72 (0.62, 0.85)** | | | | | | **0.74 (0.69, 0.80)** | | |

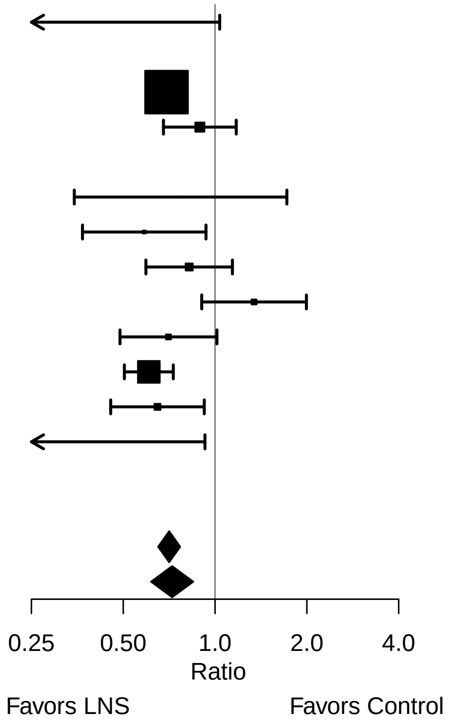

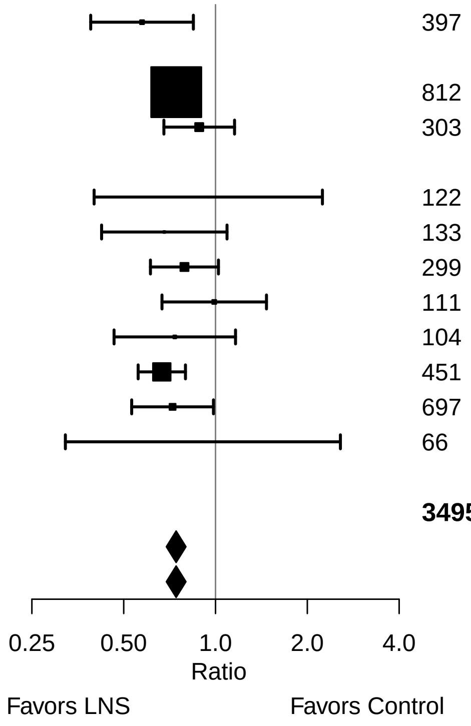

**Supplemental figure 6D: Moderate-to-severe anemia prevalence ratio**

#### 6D3: Stratified by Maternal education

**P−for−interaction = 0.437**
**Ratio of PRs = 1.07 (0.90, 1.28)**

| Country | Trial | LNS N | Control N | Control Prevalence | Primary or greater PR (95% CI) | Fixed W | Random W | LNS N | Control N | Control Prevalence | Incomplete or no formal PR (95% CI) | Fixed W | Random W |
|---|---|---|---|---|---|---|---|---|---|---|---|---|---|
| Bangladesh | JiVitA−4 (35) | | | | | | | | | | | | |
| Bangladesh | RDNS (36) | 405 | 195 | 13.3 | 0.57 (0.34, 0.98) | 0.09 | 0.12 | 144 | 77 | 24.7 | 0.51 (0.30, 0.87) | 0.01 | 0.06 |
| Bangladesh | WASH−B (37) | | | | | | | | | | | | |
| Burkina Faso | iLiNS−Zinc (38) | 82 | 16 | 68.8 | 0.79 (0.51, 1.23) | 0.14 | 0.14 | 1862 | 645 | 75.0 | 0.71 (0.65, 0.78) | 0.55 | 0.18 |
| Burkina Faso | PROMIS CS (39) | 46 | 47 | 36.2 | 1.02 (0.57, 1.85) | 0.08 | 0.11 | 526 | 532 | 36.5 | 0.88 (0.72, 1.08) | 0.10 | 0.15 |
| Ghana | GHANA (40) | | | | | | | | | | | | |
| Ghana | iLiNS−DYADG (41) | | | | | | | | | | | | |
| Kenya | WASH−B (42) | 168 | 136 | 23.5 | 0.33 (0.21, 0.52) | 0.13 | 0.14 | 181 | 164 | 19.5 | 0.96 (0.64, 1.46) | 0.02 | 0.09 |
| Madagascar | MAHAY (43) | 128 | 143 | 33.6 | 0.84 (0.57, 1.23) | 0.18 | 0.16 | 472 | 445 | 32.6 | 0.81 (0.62, 1.05) | 0.06 | 0.13 |
| Malawi | iLiNS−DYADM (44) | 34 | 66 | 22.7 | 0.78 (0.33, 1.82) | 0.04 | 0.07 | 175 | 363 | 24.0 | 1.19 (0.89, 1.61) | 0.05 | 0.12 |
| Malawi | iLiNS−DOSE (45) | 49 | 17 | 52.9 | 0.42 (0.23, 0.78) | 0.07 | 0.10 | 190 | 61 | 44.3 | 0.81 (0.59, 1.12) | 0.04 | 0.11 |
| Mali | PROMIS CS (46) | 107 | 96 | 53.1 | 0.70 (0.52, 0.96) | 0.28 | 0.18 | 845 | 874 | 61.0 | 0.63 (0.53, 0.74) | 0.16 | 0.16 |
| Zimbabwe | SHINE (HIV−) (47) | | | | | | | | | | | | |
| Zimbabwe | SHINE (HIV+) (48) | | | | | | | | | | | | |
| | | **1019** | **716** | | **I² = 0.55, Tau² = 0.08** | | | **4395** | **3161** | | **I² = 0.67, Tau² = 0.03** | | |
| **Fixed** | | | | | **0.66 (0.56, 0.77)** | | | | | | **0.74 (0.70, 0.79)** | | |
| **Random** | | | | | **0.64 (0.50, 0.84)** | | | | | | **0.79 (0.67, 0.93)** | | |

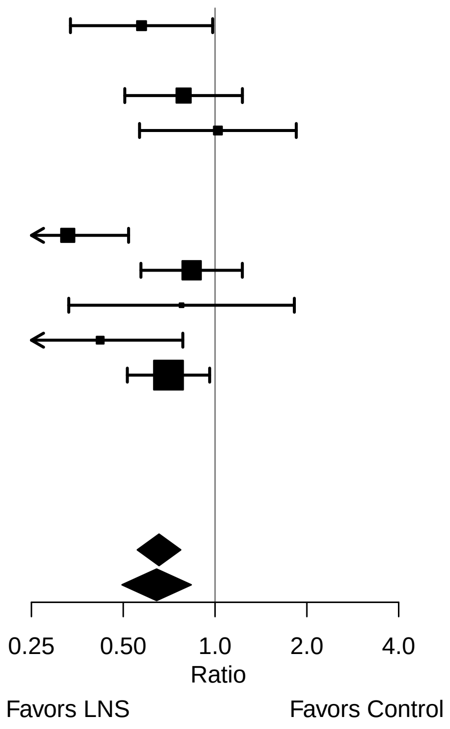

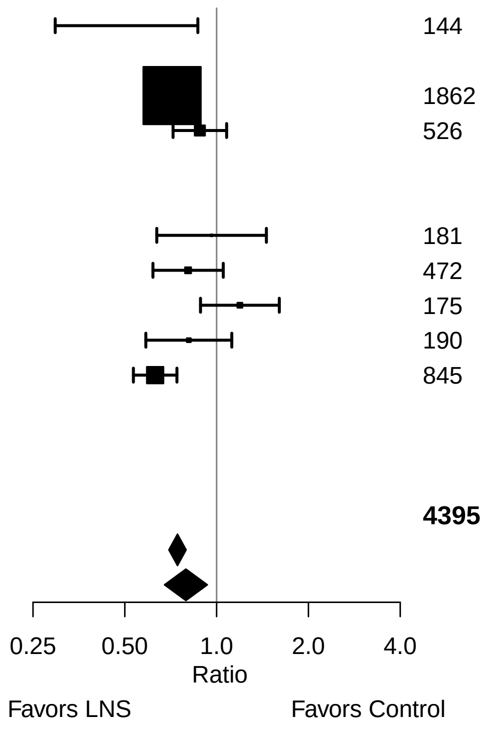

**Supplemental figure 6D: Moderate-to-severe anemia prevalence ratio**

#### 6D4: Stratified by Child sex

**P−for−interaction = 0.692**
**Ratio of PRs = 1.02 (0.93, 1.12)**

| Country | Trial | Male LNS N | Male Control N | Male Control Prevalence | Male PR (95% CI) | Male Fixed W | Male Random W | Female LNS N | Female Control N | Female Control Prevalence | Female PR (95% CI) | Female Fixed W | Female Random W |
|---|---|---|---|---|---|---|---|---|---|---|---|---|---|
| Bangladesh | JiVitA−4 (35) | | | | | | | | | | | | |
| Bangladesh | RDNS (36) | 266 | 131 | 20.6 | 0.44 (0.25, 0.78) | 0.01 | 0.06 | 281 | 141 | 12.8 | 0.67 (0.40, 1.11) | 0.03 | 0.03 |
| Bangladesh | WASH−B (37) | | | | | | | | | | | | |
| Burkina Faso | iLiNS−Zinc (38) | 993 | 334 | 79.0 | 0.70 (0.65, 0.75) | 0.65 | 0.14 | 964 | 330 | 70.9 | 0.74 (0.64, 0.85) | 0.38 | 0.38 |
| Burkina Faso | PROMIS CS (39) | 317 | 295 | 42.0 | 0.87 (0.69, 1.09) | 0.07 | 0.12 | 257 | 286 | 31.1 | 0.88 (0.67, 1.15) | 0.10 | 0.10 |
| Ghana | GHANA (40) | | | | | | | | | | | | |
| Ghana | iLiNS−DYADG (41) | 164 | 310 | 6.5 | 0.95 (0.45, 1.97) | 0.01 | 0.05 | 163 | 348 | 4.3 | 0.71 (0.26, 1.92) | 0.01 | 0.01 |
| Kenya | WASH−B (42) | 168 | 145 | 21.4 | 0.64 (0.41, 0.99) | 0.02 | 0.08 | 182 | 155 | 21.3 | 0.62 (0.40, 0.96) | 0.04 | 0.04 |
| Madagascar | MAHAY (43) | 277 | 288 | 33.0 | 0.86 (0.64, 1.16) | 0.04 | 0.10 | 323 | 300 | 32.7 | 0.77 (0.57, 1.04) | 0.08 | 0.08 |
| Malawi | iLiNS−DYADM (44) | 103 | 202 | 25.7 | 1.32 (0.92, 1.89) | 0.03 | 0.09 | 107 | 230 | 21.7 | 0.95 (0.61, 1.48) | 0.04 | 0.04 |
| Malawi | iLiNS−DOSE (45) | 123 | 43 | 51.2 | 0.74 (0.52, 1.06) | 0.03 | 0.09 | 120 | 39 | 41.0 | 0.71 (0.45, 1.13) | 0.04 | 0.04 |
| Mali | PROMIS CS (46) | 479 | 526 | 64.6 | 0.63 (0.53, 0.75) | 0.12 | 0.13 | 474 | 444 | 55.0 | 0.65 (0.53, 0.79) | 0.20 | 0.20 |
| Zimbabwe | SHINE (HIV−) (47) | 830 | 814 | 10.8 | 0.71 (0.52, 0.98) | 0.03 | 0.10 | 852 | 780 | 10.4 | 0.62 (0.45, 0.85) | 0.08 | 0.08 |
| Zimbabwe | SHINE (HIV+) (48) | 146 | 149 | 14.1 | 0.29 (0.13, 0.67) | 0.01 | 0.04 | 160 | 136 | 5.9 | 1.06 (0.45, 2.51) | 0.01 | 0.01 |
| | | **3866** | **3237** | | **I² = 0.62, Tau² = 0.07** | | | **3883** | **3189** | | **I² = 0.00, Tau² = 0.00** | | |
| **Fixed** | | | | | **0.71 (0.67, 0.76)** | | | | | | **0.73 (0.66, 0.79)** | | |
| **Random** | | | | | **0.73 (0.60, 0.89)** | | | | | | **0.73 (0.66, 0.79)** | | |

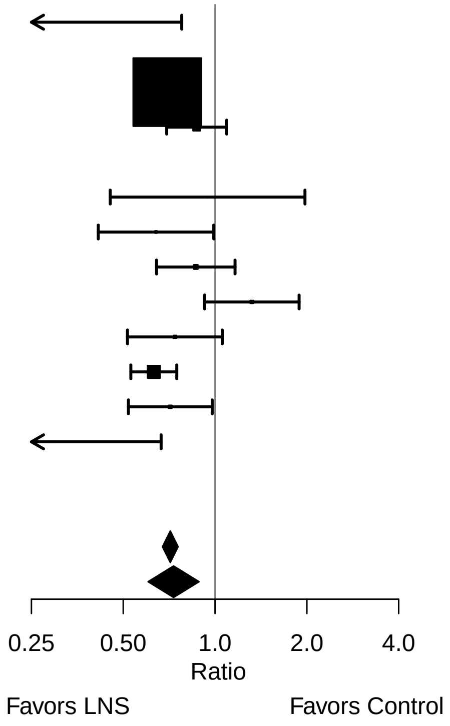

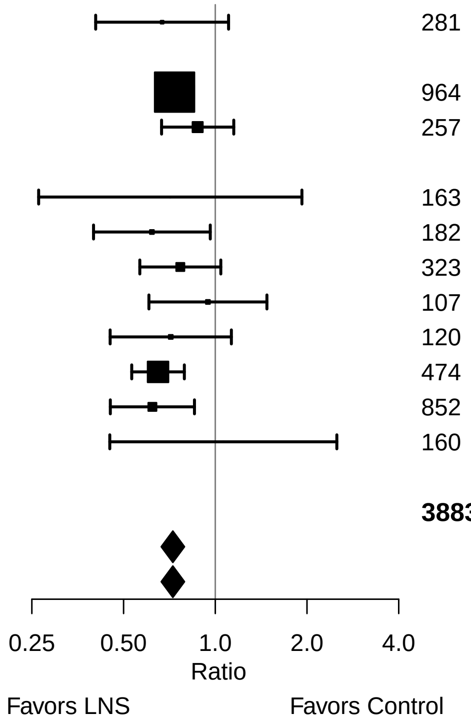

**Supplemental figure 6D: Moderate-to-severe anemia prevalence ratio**

#### 6D5: Stratified by Child birth order

**P−for−interaction = 0.416**
**Ratio of PRs = 1.05 (0.94, 1.17)**

| Country | Trial | Later born LNS N | Later born Control N | Later born Control Prevalence | Later born PR (95% CI) | Later born Fixed W | Later born Random W | Firstborn LNS N | Firstborn Control N | Firstborn Control Prevalence | Firstborn PR (95% CI) | Firstborn Fixed W | Firstborn Random W |
|---|---|---|---|---|---|---|---|---|---|---|---|---|---|
| Bangladesh | JiVitA−4 (35) | | | | | | | | | | | | |
| Bangladesh | RDNS (36) | 339 | 171 | 17.5 | 0.47 (0.29, 0.76) | 0.02 | 0.08 | 210 | 101 | 14.9 | 0.67 (0.41, 1.10) | 0.03 | 0.09 |
| Bangladesh | WASH−B (37) | | | | | | | | | | | | |
| Burkina Faso | iLiNS−Zinc (38) | 1522 | 520 | 74.0 | 0.72 (0.64, 0.81) | 0.40 | 0.13 | 435 | 144 | 78.5 | 0.70 (0.63, 0.78) | 0.70 | 0.23 |
| Burkina Faso | PROMIS CS (39) | 498 | 477 | 37.1 | 0.84 (0.68, 1.04) | 0.11 | 0.12 | 75 | 103 | 35.0 | 1.18 (0.74, 1.88) | 0.03 | 0.10 |
| Ghana | GHANA (40) | 60 | 54 | 33.3 | 0.25 (0.10, 0.63) | 0.01 | 0.03 | 38 | 38 | 31.6 | 0.42 (0.16, 1.07) | 0.01 | 0.03 |
| Ghana | iLiNS−DYADG (41) | 220 | 435 | 6.0 | 0.61 (0.28, 1.32) | 0.01 | 0.04 | 108 | 226 | 4.4 | 1.46 (0.57, 3.74) | 0.01 | 0.03 |
| Kenya | WASH−B (42) | 282 | 244 | 20.9 | 0.68 (0.46, 1.00) | 0.03 | 0.09 | 67 | 56 | 23.2 | 0.45 (0.20, 1.03) | 0.01 | 0.04 |
| Madagascar | MAHAY (43) | 458 | 454 | 33.3 | 0.81 (0.63, 1.04) | 0.08 | 0.11 | 141 | 131 | 31.3 | 0.84 (0.56, 1.27) | 0.04 | 0.11 |
| Malawi | iLiNS−DYADM (44) | 165 | 342 | 23.1 | 1.13 (0.82, 1.56) | 0.05 | 0.10 | 45 | 88 | 26.1 | 1.19 (0.68, 2.08) | 0.02 | 0.08 |
| Malawi | iLiNS−DOSE (45) | | | | | | | | | | | | |
| Mali | PROMIS CS (46) | 790 | 832 | 60.0 | 0.64 (0.55, 0.75) | 0.21 | 0.13 | 146 | 126 | 60.3 | 0.62 (0.47, 0.83) | 0.09 | 0.15 |
| Zimbabwe | SHINE (HIV−) (47) | 1188 | 1100 | 10.4 | 0.60 (0.45, 0.80) | 0.06 | 0.11 | 418 | 416 | 10.8 | 0.86 (0.58, 1.29) | 0.05 | 0.11 |
| Zimbabwe | SHINE (HIV+) (48) | 252 | 216 | 10.2 | 0.39 (0.20, 0.76) | 0.01 | 0.05 | 51 | 64 | 9.4 | 1.25 (0.45, 3.50) | 0.01 | 0.03 |
| | | **5774** | **4845** | | **I² = 0.61, Tau² = 0.07** | | | **1734** | **1493** | | **I² = 0.37, Tau² = 0.04** | | |
| **Fixed** | | | | | **0.71 (0.66, 0.76)** | | | | | | **0.73 (0.67, 0.79)** | | |
| **Random** | | | | | **0.67 (0.55, 0.81)** | | | | | | **0.79 (0.66, 0.95)** | | |

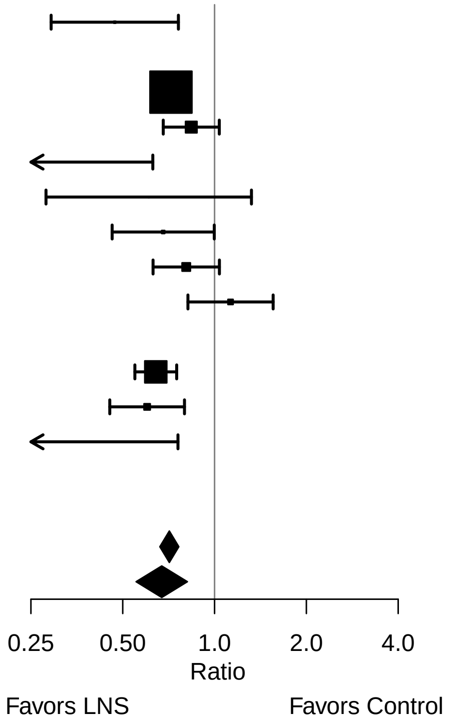

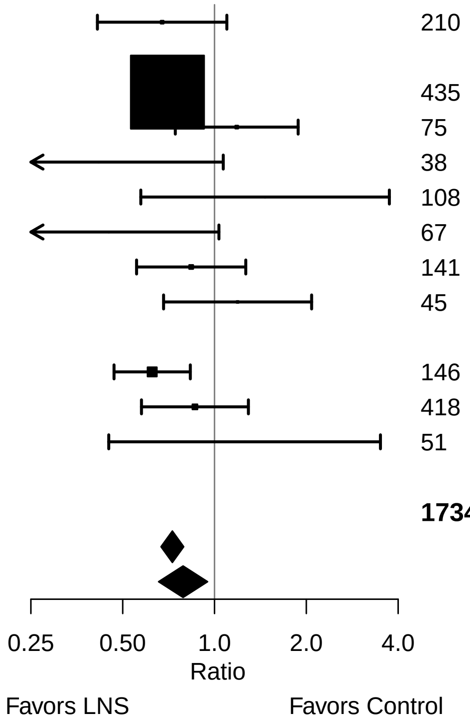

**Supplemental figure 6D: Moderate-to-severe anemia prevalence ratio**

#### 6D6: Stratified by Child baseline acute malnutrition (insufficient comparisons)

##### Supplemental figure 6D: Moderate-to-severe anemia prevalence ratio

#### 6b7: Stratified by Child baseline anemia

**P−for−interaction = 0.686**
**Ratio of PRs = 1.05 (0.82, 1.35)**

| | | Not anemic | | | | | | Anemic | | | | | |
|---|---|---|---|---|---|---|---|---|---|---|---|---|---|
| Country | Trial | LNS N | Control N | Control Prevalence | PR (95% CI) | Fixed W | Random W | LNS N | Control N | Control Prevalence | PR (95% CI) | Fixed W | Random W |
| Bangladesh | JiVitA−4 (35) | | | | | | | | | | | | |
| Bangladesh | RDNS (36) | 213 | 93 | 6.5 | 0.51 (0.15, 1.78) | 0.04 | 0.08 | 314 | 160 | 21.9 | 0.57 (0.41, 0.78) | 0.07 | 0.23 |
| Bangladesh | WASH−B (37) | | | | | | | | | | | | |
| Burkina Faso | iLiNS−Zinc (38) | 177 | 61 | 60.7 | 0.64 (0.48, 0.87) | 0.61 | 0.42 | 1780 | 603 | 76.5 | 0.72 (0.66, 0.79) | 0.80 | 0.31 |
| Burkina Faso | PROMIS CS (39) | | | | | | | | | | | | |
| Ghana | GHANA (40) | | | | | | | | | | | | |
| Ghana | iLiNS−DYADG (41) | | | | | | | | | | | | |
| Kenya | WASH−B (42) | | | | | | | | | | | | |
| Madagascar | MAHAY (43) | | | | | | | | | | | | |
| Malawi | iLiNS−DYADM (44) | 66 | 145 | 12.4 | 1.46 (0.75, 2.86) | 0.12 | 0.21 | 134 | 269 | 27.5 | 1.17 (0.85, 1.60) | 0.07 | 0.23 |
| Malawi | iLiNS−DOSE (45) | 101 | 45 | 37.8 | 0.74 (0.45, 1.20) | 0.23 | 0.29 | 142 | 37 | 56.8 | 0.72 (0.51, 1.02) | 0.06 | 0.22 |
| Mali | PROMIS CS (46) | | | | | | | | | | | | |
| Zimbabwe | SHINE (HIV−) (47) | | | | | | | | | | | | |
| Zimbabwe | SHINE (HIV+) (48) | | | | | | | | | | | | |
| | | **557** | **344** | | **I² = 0.41, Tau² = 0.07** | | | **2370** | **1069** | | **I² = 0.73, Tau² = 0.07** | | |
| **Fixed** | | | | | **0.73 (0.58, 0.92)** | | | | | | **0.73 (0.67, 0.80)** | | |
| **Random** | | | | | **0.78 (0.53, 1.14)** | | | | | | **0.76 (0.57, 1.02)** | | |

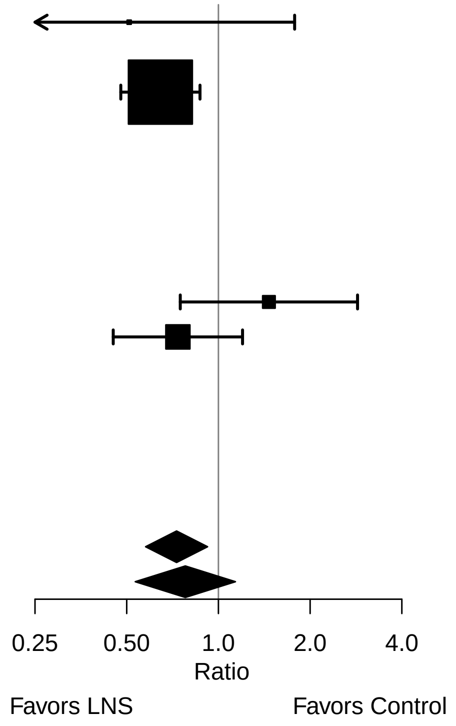

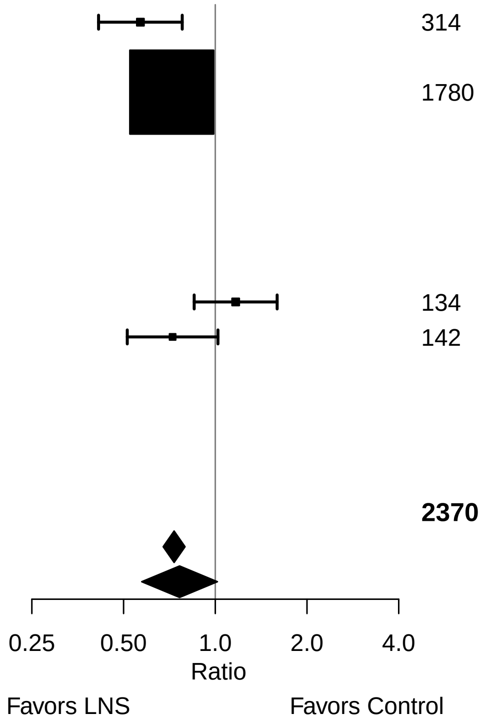

#### Supplemental figure 6D: Moderate-to-severe anemia prevalence ratio

##### 6D8: Stratified by Child high-dose vitamin A supplementation

**P−for−interaction = 0.874**
**Ratio of PRs = 1.03 (0.75, 1.40)**

| | | | | | Received VitA | | | | | | Not treated | | |
|---|---|---|---|---|---|---|---|---|---|---|---|---|---|
| | | **LNS** | **Control** | **Control** | **PR** | **Fixed** | **Random** | **LNS** | **Control** | **Control** | **PR** | **Fixed** | **Random** |
| Country | Trial | N | N | Prevalence | (95% CI) | W | W | N | N | Prevalence | (95% CI) | W | W |
| Bangladesh | JiVitA−4 (35) | | | | | | | | | | | | |
| Bangladesh | RDNS (36) | 444 | 223 | 16.6 | 0.52 (0.35, 0.75) | 0.32 | 0.30 | 105 | 49 | 16.3 | 0.64 (0.28, 1.47) | 0.06 | 0.11 |
| Bangladesh | WASH−B (37) | | | | | | | | | | | | |
| Burkina Faso | iLiNS−Zinc (38) | | | | | | | | | | | | |
| Burkina Faso | PROMIS CS (39) | | | | | | | | | | | | |
| Ghana | GHANA (40) | | | | | | | | | | | | |
| Ghana | iLiNS−DYADG (41) | | | | | | | | | | | | |
| Kenya | WASH−B (42) | 29 | 34 | 26.5 | 0.78 (0.30, 2.02) | 0.05 | 0.08 | 263 | 212 | 19.8 | 0.63 (0.42, 0.95) | 0.24 | 0.24 |
| Madagascar | MAHAY (43) | | | | | | | | | | | | |
| Malawi | iLiNS−DYADM (44) | 75 | 136 | 25.7 | 0.98 (0.61, 1.59) | 0.19 | 0.22 | 133 | 289 | 22.5 | 1.27 (0.90, 1.79) | 0.32 | 0.27 |
| Malawi | iLiNS−DOSE (45) | | | | | | | | | | | | |
| Mali | PROMIS CS (46) | | | | | | | | | | | | |
| Zimbabwe | SHINE (HIV−) (47) | 874 | 833 | 9.6 | 0.70 (0.50, 0.99) | 0.40 | 0.33 | 665 | 622 | 11.6 | 0.60 (0.42, 0.84) | 0.33 | 0.27 |
| Zimbabwe | SHINE (HIV+) (48) | 165 | 127 | 8.7 | 0.35 (0.13, 0.98) | 0.04 | 0.07 | 112 | 134 | 12.7 | 0.49 (0.21, 1.13) | 0.06 | 0.11 |
| | | **1587** | **1353** | | **I² = 0.33, Tau² = 0.03** | | | **1278** | **1306** | | **I² = 0.68, Tau² = 0.08** | | |
| **Fixed** | | | | | **0.66 (0.54, 0.82)** | | | | | | **0.77 (0.63, 0.94)** | | |
| **Random** | | | | | **0.66 (0.50, 0.88)** | | | | | | **0.73 (0.52, 1.02)** | | |

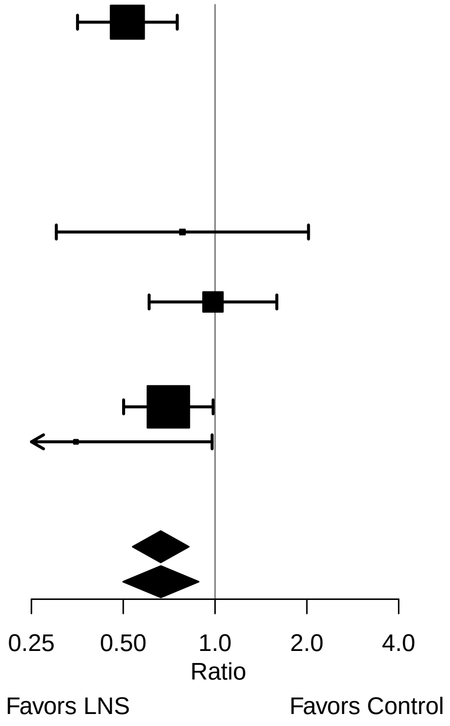

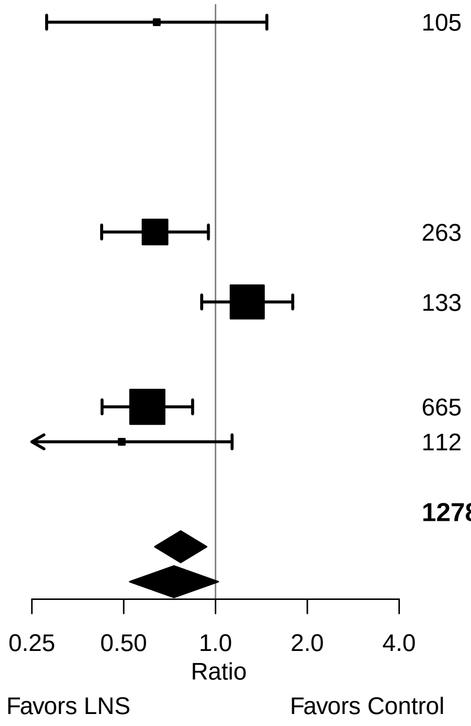

#### Supplemental figure 6D: Moderate-to-severe anemia prevalence ratio

##### 6D9: Stratified by Child inflammation

**P−for−interaction = 0.197**
**Ratio of PRs = 1.17 (0.92, 1.47)**

| Country | Trial | LNS N | Control N | Control Prevalence | Not inflamed PR (95% CI) | Fixed W | Random W | LNS N | Control N | Control Prevalence | High AGP or CRP PR (95% CI) | Fixed W | Random W |
|---|---|---|---|---|---|---|---|---|---|---|---|---|---|
| Bangladesh | JiVitA−4 (35) | | | | | | | | | | | | |
| Bangladesh | RDNS (36) | 369 | 168 | 17.3 | 0.50 (0.33, 0.76) | 0.26 | 0.26 | 180 | 104 | 15.4 | 0.61 (0.36, 1.03) | 0.07 | 0.10 |
| Bangladesh | WASH−B (37) | | | | | | | | | | | | |
| Burkina Faso | iLiNS−Zinc (38) | 131 | 33 | 69.7 | 0.61 (0.44, 0.84) | 0.43 | 0.32 | 197 | 69 | 81.2 | 0.73 (0.61, 0.86) | 0.62 | 0.49 |
| Burkina Faso | PROMIS CS (39) | | | | | | | | | | | | |
| Ghana | GHANA (40) | | | | | | | | | | | | |
| Ghana | iLiNS−DYADG (41) | | | | | | | | | | | | |
| Kenya | WASH−B (42) | 157 | 135 | 14.1 | 0.45 (0.23, 0.88) | 0.10 | 0.14 | 141 | 123 | 30.9 | 0.71 (0.47, 1.09) | 0.10 | 0.14 |
| Madagascar | MAHAY (43) | | | | | | | | | | | | |
| Malawi | iLiNS−DYADM (44) | 59 | 123 | 13.0 | 0.91 (0.40, 2.10) | 0.06 | 0.10 | 138 | 269 | 29.4 | 1.04 (0.76, 1.42) | 0.18 | 0.23 |
| Malawi | iLiNS−DOSE (45) | | | | | | | | | | | | |
| Mali | PROMIS CS (46) | | | | | | | | | | | | |
| Zimbabwe | SHINE (HIV−) (47) | 414 | 354 | 5.6 | 1.03 (0.60, 1.76) | 0.15 | 0.19 | 105 | 99 | 14.1 | 0.61 (0.29, 1.29) | 0.03 | 0.05 |
| Zimbabwe | SHINE (HIV+) (48) | | | | | | | | | | | | |
| | | **1130** | **813** | | **I² = 0.34, Tau² = 0.04** | | | **761** | **664** | | **I² = 0.22, Tau² = 0.01** | | |
| **Fixed** | | | | | **0.63 (0.51, 0.77)** | | | | | | **0.76 (0.66, 0.87)** | | |
| **Random** | | | | | **0.64 (0.48, 0.85)** | | | | | | **0.76 (0.65, 0.91)** | | |

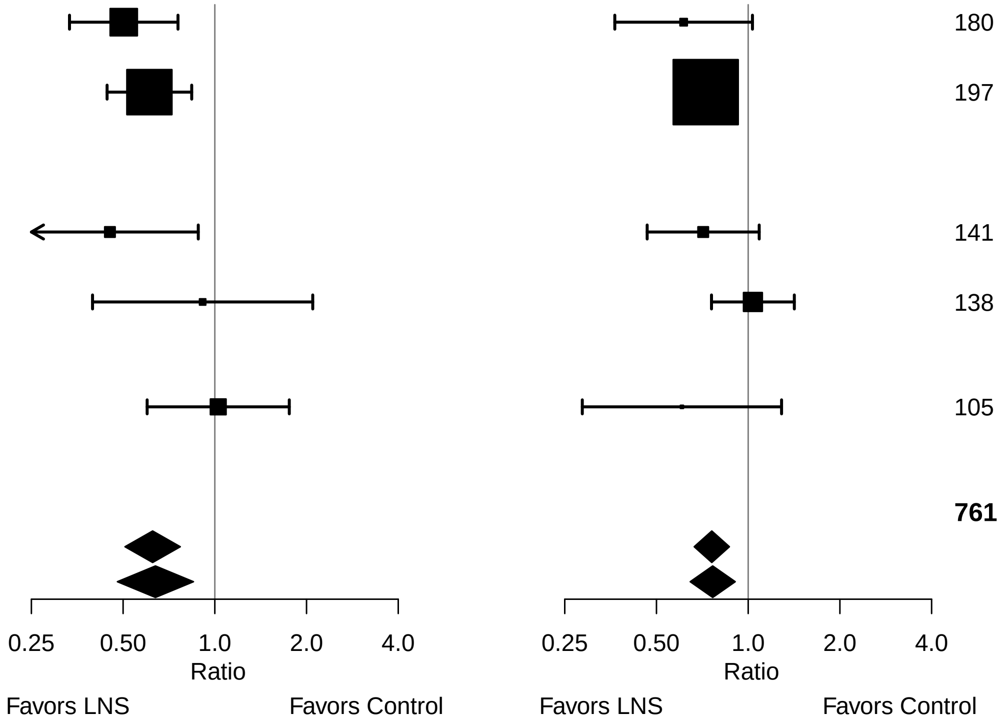

### Supplemental figure 6E: Moderate-to-severe anemia prevalence difference

#### 6E1: Stratified by Maternal BMI

**P−for−interaction = 0.047**
**Difference in PDs = 0.03 (0.00, 0.07)**

| Country | Trial | At least 20 kg/m² LNS N | Control N | Control Prevalence | PD (95% CI) | Fixed W | Random W | Less than 20 kg/m² LNS N | Control N | Control Prevalence | PD (95% CI) | Fixed W | Random W |
|---|---|---|---|---|---|---|---|---|---|---|---|---|---|
| Bangladesh | JiVitA−4 (35) | | | | | | | | | | | | |
| Bangladesh | RDNS (36) | 227 | 102 | 20.6 | −0.11 (−0.20, −0.02) | 0.04 | 0.12 | 300 | 160 | 13.8 | −0.05 (−0.11, 0.00) | 0.22 | 0.15 |
| Bangladesh | WASH−B (37) | | | | | | | | | | | | |
| Burkina Faso | iLiNS−Zinc (38) | 1217 | 390 | 76.2 | −0.22 (−0.28, −0.17) | 0.10 | 0.14 | 738 | 274 | 73.4 | −0.20 (−0.27, −0.13) | 0.14 | 0.14 |
| Burkina Faso | PROMIS CS (39) | 335 | 349 | 36.7 | −0.01 (−0.09, 0.07) | 0.05 | 0.13 | 239 | 227 | 37.0 | −0.09 (−0.19, 0.01) | 0.07 | 0.12 |
| Ghana | GHANA (40) | | | | | | | | | | | | |
| Ghana | iLiNS−DYADG (41) | | | | | | | | | | | | |
| Kenya | WASH−B (42) | 266 | 227 | 22.9 | −0.11 (−0.18, −0.03) | 0.06 | 0.13 | 72 | 60 | 11.7 | 0.05 (−0.05, 0.15) | 0.07 | 0.12 |
| Madagascar | MAHAY (43) | | | | | | | | | | | | |
| Malawi | iLiNS−DYADM (44) | 124 | 258 | 24.0 | 0.03 (−0.06, 0.13) | 0.04 | 0.12 | 85 | 171 | 23.4 | 0.02 (−0.09, 0.14) | 0.05 | 0.12 |
| Malawi | iLiNS−DOSE (45) | 179 | 52 | 46.2 | −0.19 (−0.33, −0.05) | 0.02 | 0.09 | 61 | 29 | 48.3 | −0.02 (−0.24, 0.20) | 0.01 | 0.07 |
| Mali | PROMIS CS (46) | 702 | 685 | 60.0 | −0.22 (−0.30, −0.14) | 0.05 | 0.13 | 246 | 279 | 61.3 | −0.23 (−0.30, −0.16) | 0.14 | 0.14 |
| Zimbabwe | SHINE (HIV−) (47) | 1222 | 1161 | 10.9 | −0.04 (−0.06, −0.02) | 0.65 | 0.15 | 213 | 220 | 6.8 | 0.02 (−0.03, 0.06) | 0.30 | 0.15 |
| Zimbabwe | SHINE (HIV+) (48) | | | | | | | | | | | | |
| | | **4272** | **3224** | | **I² = 0.88, Tau² = 0.01** | | | **1954** | **1420** | | **I² = 0.87, Tau² = 0.01** | | |
| **Fixed** | | | | | **−0.07 (−0.09, −0.05)** | | | | | | **−0.07 (−0.09, −0.04)** | | |
| **Random** | | | | | **−0.11 (−0.17, −0.04)** | | | | | | **−0.07 (−0.14, 0.01)** | | |

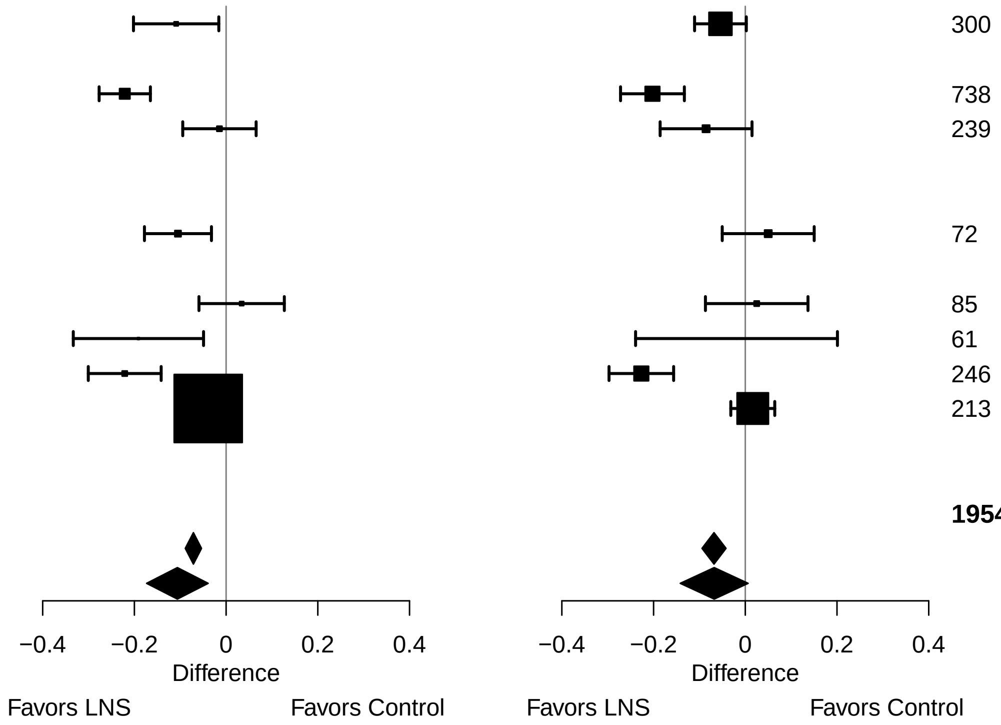

#### Supplemental figure 6E: Moderate-to-severe anemia prevalence difference

##### 6E2: Stratified by Maternal age

**P−for−interaction = 0.502**
**Difference in PDs = 0.01 (−0.02, 0.03)**

| Country | Trial | At least 25 y: LNS N | Control N | Control Prevalence | PD (95% CI) | Fixed W | Random W | Less than 25 y: LNS N | Control N | Control Prevalence | PD (95% CI) | Fixed W | Random W |
|---|---|---|---|---|---|---|---|---|---|---|---|---|---|
| Bangladesh | JiVitA−4 (35) | | | | | | | | | | | | |
| Bangladesh | RDNS (36) | 152 | 81 | 17.3 | −0.09 (−0.21, 0.02) | 0.02 | 0.07 | 397 | 191 | 16.2 | −0.07 (−0.12, −0.02) | 0.14 | 0.12 |
| Bangladesh | WASH−B (37) | | | | | | | | | | | | |
| Burkina Faso | iLiNS−Zinc (38) | 1132 | 399 | 74.2 | −0.23 (−0.31, −0.15) | 0.04 | 0.09 | 812 | 262 | 76.0 | −0.19 (−0.26, −0.13) | 0.08 | 0.10 |
| Burkina Faso | PROMIS CS (39) | 271 | 263 | 32.7 | −0.04 (−0.12, 0.05) | 0.04 | 0.09 | 303 | 318 | 39.9 | −0.05 (−0.15, 0.05) | 0.04 | 0.08 |
| Ghana | GHANA (40) | | | | | | | | | | | | |
| Ghana | iLiNS−DYADG (41) | 206 | 397 | 5.0 | −0.01 (−0.05, 0.02) | 0.23 | 0.11 | 122 | 264 | 6.1 | 0.00 (−0.05, 0.05) | 0.14 | 0.12 |
| Kenya | WASH−B (42) | 213 | 168 | 20.8 | −0.09 (−0.17, −0.01) | 0.05 | 0.09 | 133 | 131 | 22.1 | −0.07 (−0.16, 0.02) | 0.05 | 0.09 |
| Madagascar | MAHAY (43) | 301 | 329 | 30.7 | −0.05 (−0.15, 0.04) | 0.03 | 0.09 | 299 | 259 | 35.5 | −0.07 (−0.16, 0.01) | 0.05 | 0.09 |
| Malawi | iLiNS−DYADM (44) | 99 | 220 | 21.8 | 0.07 (−0.03, 0.18) | 0.03 | 0.08 | 111 | 212 | 25.5 | 0.00 (−0.10, 0.10) | 0.04 | 0.08 |
| Malawi | iLiNS−DOSE (45) | 135 | 50 | 46.0 | −0.14 (−0.29, 0.01) | 0.01 | 0.06 | 104 | 28 | 46.4 | −0.13 (−0.32, 0.07) | 0.01 | 0.03 |
| Mali | PROMIS CS (46) | 502 | 508 | 61.4 | −0.24 (−0.32, −0.16) | 0.04 | 0.09 | 451 | 462 | 58.9 | −0.20 (−0.27, −0.12) | 0.06 | 0.09 |
| Zimbabwe | SHINE (HIV−) (47) | 808 | 726 | 9.4 | −0.03 (−0.06, −0.01) | 0.41 | 0.11 | 697 | 694 | 12.1 | −0.03 (−0.06, 0.00) | 0.37 | 0.13 |
| Zimbabwe | SHINE (HIV+) (48) | 221 | 204 | 9.8 | −0.06 (−0.11, 0.00) | 0.10 | 0.10 | 66 | 60 | 10.0 | −0.01 (−0.11, 0.09) | 0.04 | 0.08 |
| | | **4040** | **3345** | | **$I^2$ = 0.81, $Tau^2$ = 0.01** | | | **3495** | **2881** | | **$I^2$ = 0.73, $Tau^2$ = 0.00** | | |
| **Fixed** | | | | | **−0.05 (−0.07, −0.03)** | | | | | | **−0.06 (−0.08, −0.04)** | | |
| **Random** | | | | | **−0.08 (−0.13, −0.03)** | | | | | | **−0.07 (−0.11, −0.03)** | | |

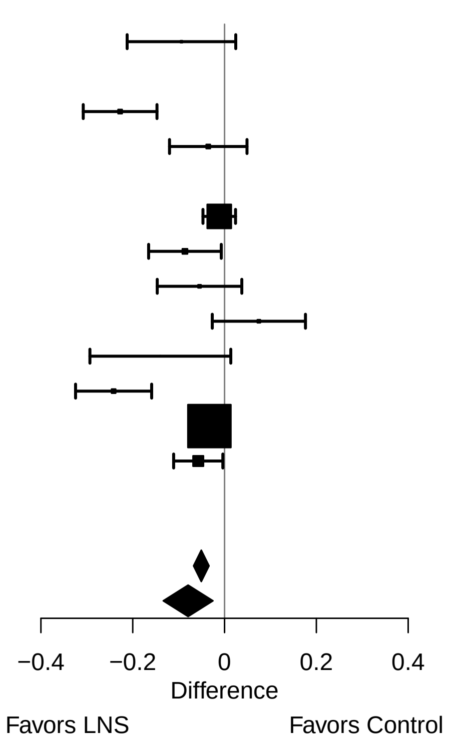

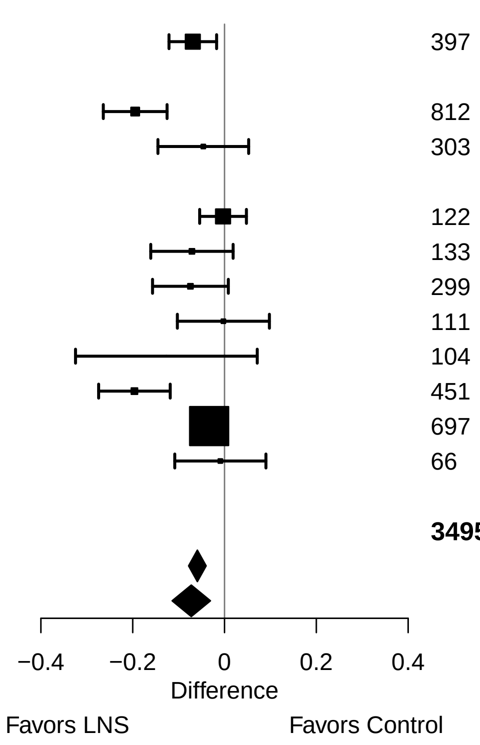

#### Supplemental figure 6E: Moderate-to-severe anemia prevalence difference

##### 6E3: Stratified by Maternal education

**P−for−interaction = 0.526**
**Difference in PDs = 0.02 (−0.04, 0.07)**

| | | Primary or greater | | | | | | Incomplete or no formal | | | | | |
|---|---|---|---|---|---|---|---|---|---|---|---|---|---|
| Country | Trial | LNS N | Control N | Control Prevalence | PD (95% CI) | Fixed W | Random W | LNS N | Control N | Control Prevalence | PD (95% CI) | Fixed W | Random W |
| Bangladesh | JiVitA−4 (35) | | | | | | | | | | | | |
| Bangladesh | RDNS (36) | 405 | 195 | 13.3 | −0.06 (−0.12, 0.01) | 0.37 | 0.29 | 144 | 77 | 24.7 | −0.12 (−0.23, −0.01) | 0.06 | 0.11 |
| Bangladesh | WASH−B (37) | | | | | | | | | | | | |
| Burkina Faso | iLiNS−Zinc (38) | 82 | 16 | 68.8 | −0.15 (−0.42, 0.13) | 0.02 | 0.03 | 1862 | 645 | 75.0 | −0.21 (−0.27, −0.16) | 0.23 | 0.14 |
| Burkina Faso | PROMIS CS (39) | 46 | 47 | 36.2 | 0.01 (−0.21, 0.22) | 0.03 | 0.05 | 526 | 532 | 36.5 | −0.04 (−0.11, 0.03) | 0.16 | 0.13 |
| Ghana | GHANA (40) | | | | | | | | | | | | |
| Ghana | iLiNS−DYADG (41) | | | | | | | | | | | | |
| Kenya | WASH−B (42) | 168 | 136 | 23.5 | −0.16 (−0.23, −0.09) | 0.31 | 0.27 | 181 | 164 | 19.5 | −0.01 (−0.09, 0.07) | 0.12 | 0.13 |
| Madagascar | MAHAY (43) | 128 | 143 | 33.6 | −0.05 (−0.17, 0.06) | 0.11 | 0.14 | 472 | 445 | 32.6 | −0.06 (−0.14, 0.02) | 0.12 | 0.13 |
| Malawi | iLiNS−DYADM (44) | 34 | 66 | 22.7 | −0.05 (−0.22, 0.12) | 0.05 | 0.08 | 175 | 363 | 24.0 | 0.05 (−0.03, 0.12) | 0.12 | 0.13 |
| Malawi | iLiNS−DOSE (45) | 49 | 17 | 52.9 | −0.32 (−0.57, −0.07) | 0.02 | 0.04 | 190 | 61 | 44.3 | −0.09 (−0.22, 0.05) | 0.04 | 0.10 |
| Mali | PROMIS CS (46) | 107 | 96 | 53.1 | −0.16 (−0.30, −0.02) | 0.08 | 0.11 | 845 | 874 | 61.0 | −0.23 (−0.30, −0.16) | 0.15 | 0.13 |
| Zimbabwe | SHINE (HIV−) (47) | | | | | | | | | | | | |
| Zimbabwe | SHINE (HIV+) (48) | | | | | | | | | | | | |
| | | **1019** | **716** | | **I² = 0.31, Tau² = 0.00** | | | **4395** | **3161** | | **I² = 0.86, Tau² = 0.01** | | |
| **Fixed** | | | | | **−0.10 (−0.14, −0.06)** | | | | | | **−0.10 (−0.13, −0.08)** | | |
| **Random** | | | | | **−0.10 (−0.15, −0.05)** | | | | | | **−0.09 (−0.16, −0.02)** | | |

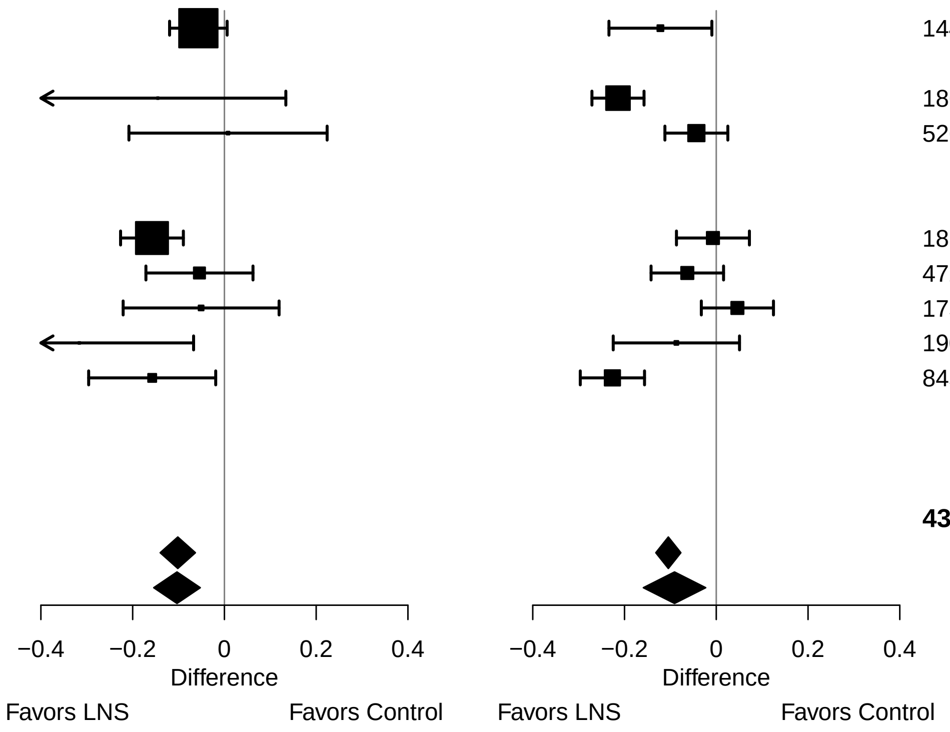

**Supplemental figure 6E: Moderate-to-severe anemia prevalence difference**

#### 6E4: Stratified by Child sex

**P−for−interaction = 0.323**
**Difference in PDs = 0.01 (−0.01, 0.04)**

| Country | Trial | Male LNS N | Male Control N | Male Control Prevalence | Male PD (95% CI) | Male Fixed W | Male Random W | Female LNS N | Female Control N | Female Control Prevalence | Female PD (95% CI) | Female Fixed W | Female Random W |
|---|---|---|---|---|---|---|---|---|---|---|---|---|---|
| Bangladesh | JiVitA−4 (35) | | | | | | | | | | | | |
| Bangladesh | RDNS (36) | 266 | 131 | 20.6 | −0.12 (−0.21, −0.02) | 0.03 | 0.08 | 281 | 141 | 12.8 | −0.04 (−0.10, 0.01) | 0.08 | 0.11 |
| Bangladesh | WASH−B (37) | | | | | | | | | | | | |
| Burkina Faso | iLiNS−Zinc (38) | 993 | 334 | 79.0 | −0.24 (−0.28, −0.19) | 0.15 | 0.11 | 964 | 330 | 70.9 | −0.19 (−0.28, −0.10) | 0.03 | 0.08 |
| Burkina Faso | PROMIS CS (39) | 317 | 295 | 42.0 | −0.05 (−0.14, 0.03) | 0.04 | 0.09 | 257 | 286 | 31.1 | −0.04 (−0.12, 0.04) | 0.04 | 0.09 |
| Ghana | GHANA (40) | | | | | | | | | | | | |
| Ghana | iLiNS−DYADG (41) | 164 | 310 | 6.5 | 0.00 (−0.05, 0.04) | 0.15 | 0.11 | 163 | 348 | 4.3 | −0.01 (−0.05, 0.02) | 0.20 | 0.12 |
| Kenya | WASH−B (42) | 168 | 145 | 21.4 | −0.08 (−0.16, 0.00) | 0.05 | 0.09 | 182 | 155 | 21.3 | −0.08 (−0.15, −0.01) | 0.05 | 0.09 |
| Madagascar | MAHAY (43) | 277 | 288 | 33.0 | −0.04 (−0.14, 0.05) | 0.04 | 0.09 | 323 | 300 | 32.7 | −0.08 (−0.16, 0.01) | 0.03 | 0.08 |
| Malawi | iLiNS−DYADM (44) | 103 | 202 | 25.7 | 0.08 (−0.02, 0.19) | 0.03 | 0.08 | 107 | 230 | 21.7 | −0.01 (−0.11, 0.08) | 0.03 | 0.08 |
| Malawi | iLiNS−DOSE (45) | 123 | 43 | 51.2 | −0.13 (−0.30, 0.04) | 0.01 | 0.06 | 120 | 39 | 41.0 | −0.13 (−0.30, 0.03) | 0.01 | 0.04 |
| Mali | PROMIS CS (46) | 479 | 526 | 64.6 | −0.24 (−0.32, −0.16) | 0.04 | 0.09 | 474 | 444 | 55.0 | −0.19 (−0.27, −0.11) | 0.04 | 0.09 |
| Zimbabwe | SHINE (HIV−) (47) | 830 | 814 | 10.8 | −0.03 (−0.06, 0.00) | 0.39 | 0.11 | 852 | 780 | 10.4 | −0.04 (−0.07, −0.01) | 0.37 | 0.13 |
| Zimbabwe | SHINE (HIV+) (48) | 146 | 149 | 14.1 | −0.10 (−0.17, −0.03) | 0.07 | 0.10 | 160 | 136 | 5.9 | 0.00 (−0.05, 0.06) | 0.10 | 0.11 |
| | | **3866** | **3237** | | **I² = 0.89, Tau² = 0.01** | | | **3883** | **3189** | | **I² = 0.68, Tau² = 0.00** | | |
| **Fixed** | | | | | **−0.08 (−0.09, −0.06)** | | | | | | **−0.04 (−0.06, −0.03)** | | |
| **Random** | | | | | **−0.09 (−0.14, −0.03)** | | | | | | **−0.06 (−0.10, −0.03)** | | |

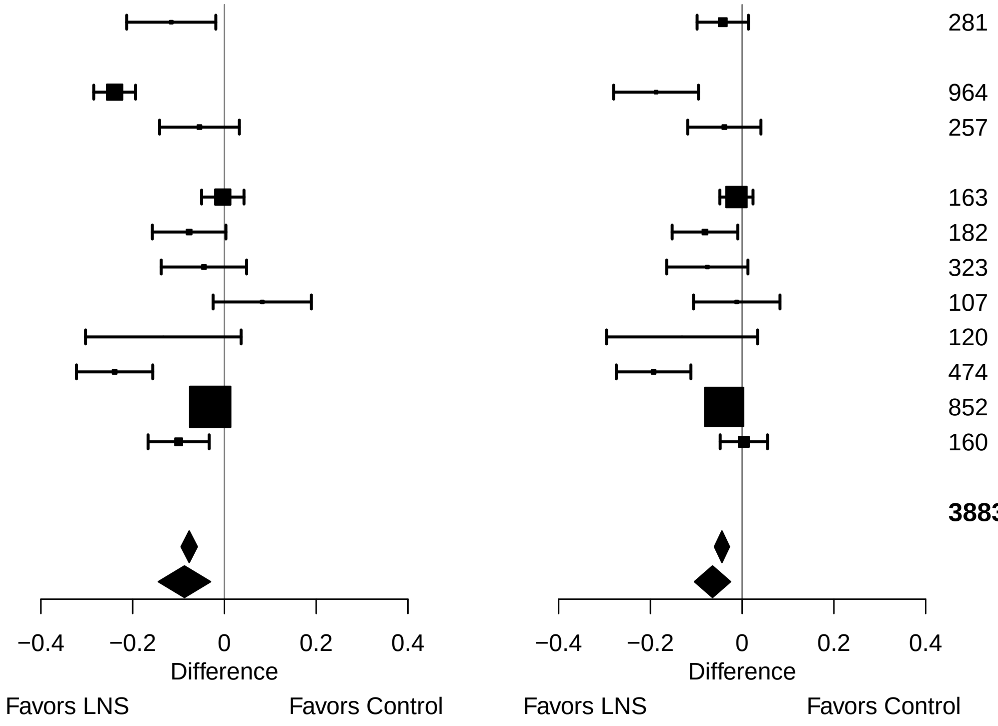

##### Supplemental figure 6E: Moderate-to-severe anemia prevalence difference

#### 6E5: Stratified by Child birth order

**P−for−interaction = 0.058**
**Difference in PDs = 0.03 (0.00, 0.06)**

| Country | Trial | Later born LNS N | Later born Control N | Later born Control Prevalence | Later born PD (95% CI) | Later born Fixed W | Later born Random W | Firstborn LNS N | Firstborn Control N | Firstborn Control Prevalence | Firstborn PD (95% CI) | Firstborn Fixed W | Firstborn Random W |
|---|---|---|---|---|---|---|---|---|---|---|---|---|---|
| Bangladesh | JiVitA−4 (35) | | | | | | | | | | | | |
| Bangladesh | RDNS (36) | 339 | 171 | 17.5 | −0.09 (−0.16, −0.02) | 0.04 | 0.09 | 210 | 101 | 14.9 | −0.05 (−0.11, 0.01) | 0.15 | 0.11 |
| Bangladesh | WASH−B (37) | | | | | | | | | | | | |
| Burkina Faso | iLiNS−Zinc (38) | 1522 | 520 | 74.0 | −0.21 (−0.28, −0.14) | 0.04 | 0.09 | 435 | 144 | 78.5 | −0.23 (−0.30, −0.17) | 0.12 | 0.11 |
| Burkina Faso | PROMIS CS (39) | 498 | 477 | 37.1 | −0.06 (−0.13, 0.01) | 0.04 | 0.09 | 75 | 103 | 35.0 | 0.06 (−0.11, 0.24) | 0.02 | 0.06 |
| Ghana | GHANA (40) | 60 | 54 | 33.3 | −0.25 (−0.39, −0.11) | 0.01 | 0.06 | 38 | 38 | 31.6 | −0.18 (−0.37, 0.00) | 0.02 | 0.06 |
| Ghana | iLiNS−DYADG (41) | 220 | 435 | 6.0 | −0.02 (−0.06, 0.01) | 0.16 | 0.10 | 108 | 226 | 4.4 | 0.02 (−0.03, 0.07) | 0.21 | 0.12 |
| Kenya | WASH−B (42) | 282 | 244 | 20.9 | −0.07 (−0.13, 0.00) | 0.05 | 0.09 | 67 | 56 | 23.2 | −0.13 (−0.27, 0.01) | 0.03 | 0.08 |
| Madagascar | MAHAY (43) | 458 | 454 | 33.3 | −0.06 (−0.14, 0.01) | 0.04 | 0.09 | 141 | 131 | 31.3 | −0.05 (−0.17, 0.07) | 0.04 | 0.09 |
| Malawi | iLiNS−DYADM (44) | 165 | 342 | 23.1 | 0.03 (−0.05, 0.11) | 0.03 | 0.09 | 45 | 88 | 26.1 | 0.05 (−0.11, 0.21) | 0.02 | 0.07 |
| Malawi | iLiNS−DOSE (45) | | | | | | | | | | | | |
| Mali | PROMIS CS (46) | 790 | 832 | 60.0 | −0.21 (−0.29, −0.14) | 0.04 | 0.09 | 146 | 126 | 60.3 | −0.23 (−0.35, −0.10) | 0.03 | 0.08 |
| Zimbabwe | SHINE (HIV−) (47) | 1188 | 1100 | 10.4 | −0.04 (−0.06, −0.02) | 0.44 | 0.11 | 418 | 416 | 10.8 | −0.01 (−0.06, 0.03) | 0.32 | 0.12 |
| Zimbabwe | SHINE (HIV+) (48) | 252 | 216 | 10.2 | −0.06 (−0.11, −0.02) | 0.10 | 0.10 | 51 | 64 | 9.4 | 0.02 (−0.08, 0.13) | 0.05 | 0.09 |
| | | **5774** | **4845** | | **I² = 0.82, Tau² = 0.01** | | | **1734** | **1493** | | **I² = 0.82, Tau² = 0.01** | | |
| **Fixed** | | | | | **−0.06 (−0.07, −0.05)** | | | | | | **−0.05 (−0.07, −0.03)** | | |
| **Random** | | | | | **−0.09 (−0.14, −0.04)** | | | | | | **−0.07 (−0.13, 0.00)** | | |

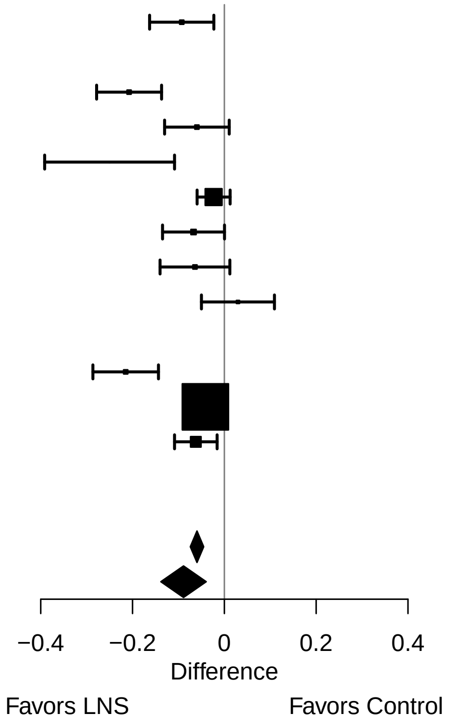

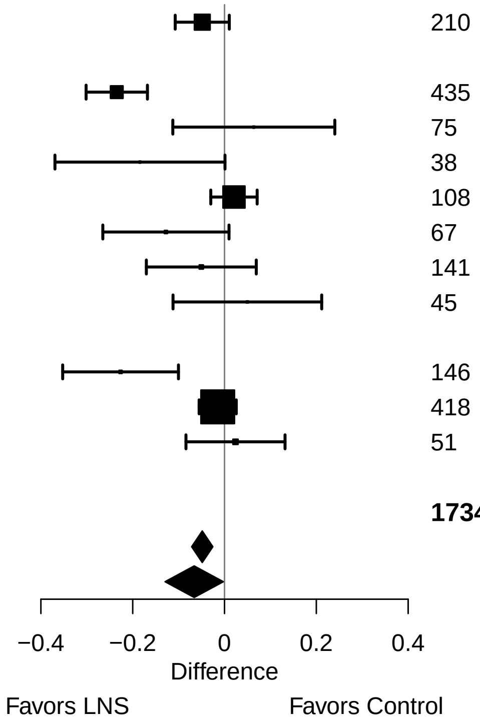

**Supplemental figure 6E: Moderate-to-severe anemia prevalence difference**

#### 6E6: Stratified by Child baseline acute malnutrition (insufficient comparisons)

#### Supplemental figure 6E: Moderate-to-severe anemia prevalence difference

##### 6E7: Stratified by Child baseline anemia

**P−for−interaction = 0.251**
**Difference in PDs = −0.04 (−0.10, 0.03)**

| Country | Trial | Not anemic LNS N | Not anemic Control N | Not anemic Control Prevalence | Not anemic PD (95% CI) | Not anemic Fixed W | Not anemic Random W | Anemic LNS N | Anemic Control N | Anemic Control Prevalence | Anemic PD (95% CI) | Anemic Fixed W | Anemic Random W |
|---|---|---|---|---|---|---|---|---|---|---|---|---|---|
| Bangladesh | JiVitA−4 (35) | | | | | | | | | | | | |
| Bangladesh | RDNS (36) | 213 | 93 | 6.5 | −0.03 (−0.10, 0.04) | 0.53 | 0.31 | 314 | 160 | 21.9 | −0.09 (−0.15, −0.04) | 0.41 | 0.29 |
| Bangladesh | WASH−B (37) | | | | | | | | | | | | |
| Burkina Faso | iLiNS−Zinc (38) | 177 | 61 | 60.7 | −0.22 (−0.37, −0.06) | 0.11 | 0.21 | 1780 | 603 | 76.5 | −0.21 (−0.28, −0.15) | 0.39 | 0.28 |
| Burkina Faso | PROMIS CS (39) | | | | | | | | | | | | |
| Ghana | GHANA (40) | | | | | | | | | | | | |
| Ghana | iLiNS−DYADG (41) | | | | | | | | | | | | |
| Kenya | WASH−B (42) | | | | | | | | | | | | |
| Madagascar | MAHAY (43) | | | | | | | | | | | | |
| Malawi | iLiNS−DYADM (44) | 66 | 145 | 12.4 | 0.06 (−0.04, 0.16) | 0.26 | 0.28 | 134 | 269 | 27.5 | 0.05 (−0.05, 0.14) | 0.16 | 0.25 |
| Malawi | iLiNS−DOSE (45) | 101 | 45 | 37.8 | −0.10 (−0.26, 0.06) | 0.10 | 0.20 | 142 | 37 | 56.8 | −0.15 (−0.33, 0.02) | 0.05 | 0.18 |
| Mali | PROMIS CS (46) | | | | | | | | | | | | |
| Zimbabwe | SHINE (HIV−) (47) | | | | | | | | | | | | |
| Zimbabwe | SHINE (HIV+) (48) | | | | | | | | | | | | |
| | | **557** | **344** | | **I² = 0.66, Tau² = 0.01** | | | **2370** | **1069** | | **I² = 0.86, Tau² = 0.01** | | |
| **Fixed** | | | | | **−0.04 (−0.09, 0.02)** | | | | | | **−0.12 (−0.16, −0.08)** | | |
| **Random** | | | | | **−0.06 (−0.17, 0.05)** | | | | | | **−0.10 (−0.21, 0.01)** | | |

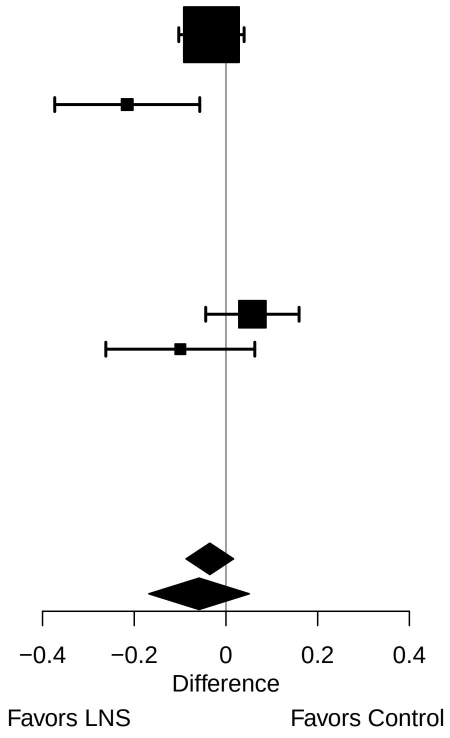

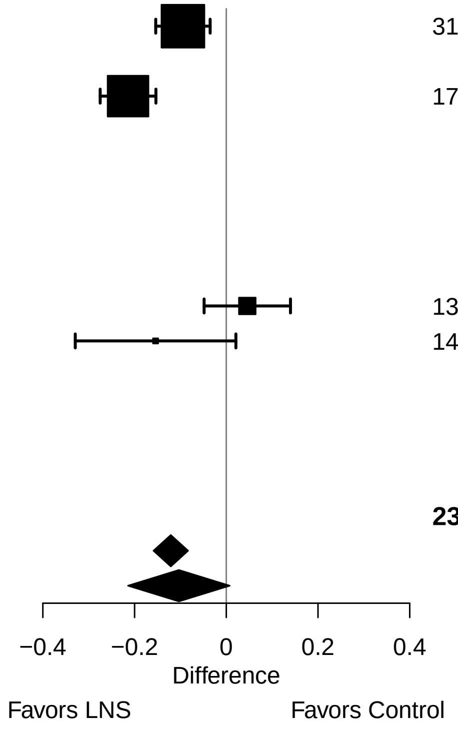

#### Supplemental figure 6E: Moderate-to-severe anemia prevalence difference

##### 6E8: Stratified by Child high-dose vitamin A supplementation

**P−for−interaction = 0.589**
**Difference in PDs = −0.01 (−0.04, 0.03)**

| Country | Trial | Received VitA: LNS N | Control N | Control Prevalence | PD (95% CI) | Fixed W | Random W | Not treated: LNS N | Control N | Control Prevalence | PD (95% CI) | Fixed W | Random W |
|---|---|---|---|---|---|---|---|---|---|---|---|---|---|
| Bangladesh | JiVitA−4 (35) | | | | | | | | | | | | |
| Bangladesh | RDNS (36) | 444 | 223 | 16.6 | −0.08 (−0.13, −0.03) | 0.17 | 0.17 | 105 | 49 | 16.3 | −0.06 (−0.18, 0.06) | 0.04 | 0.10 |
| Bangladesh | WASH−B (37) | | | | | | | | | | | | |
| Burkina Faso | iLiNS−Zinc (38) | | | | | | | | | | | | |
| Burkina Faso | PROMIS CS (39) | | | | | | | | | | | | |
| Ghana | GHANA (40) | | | | | | | | | | | | |
| Ghana | iLiNS−DYADG (41) | | | | | | | | | | | | |
| Kenya | WASH−B (42) | 29 | 34 | 26.5 | −0.06 (−0.27, 0.16) | 0.01 | 0.01 | 263 | 212 | 19.8 | −0.07 (−0.14, 0.00) | 0.13 | 0.21 |
| Madagascar | MAHAY (43) | | | | | | | | | | | | |
| Malawi | iLiNS−DYADM (44) | 75 | 136 | 25.7 | 0.00 (−0.13, 0.12) | 0.03 | 0.03 | 133 | 289 | 22.5 | 0.06 (−0.03, 0.15) | 0.08 | 0.15 |
| Malawi | iLiNS−DOSE (45) | | | | | | | | | | | | |
| Mali | PROMIS CS (46) | | | | | | | | | | | | |
| Zimbabwe | SHINE (HIV−) (47) | 874 | 833 | 9.6 | −0.03 (−0.05, 0.00) | 0.67 | 0.67 | 665 | 622 | 11.6 | −0.05 (−0.08, −0.02) | 0.65 | 0.36 |
| Zimbabwe | SHINE (HIV+) (48) | 165 | 127 | 8.7 | −0.06 (−0.12, 0.00) | 0.12 | 0.12 | 112 | 134 | 12.7 | −0.06 (−0.14, 0.01) | 0.11 | 0.19 |
| | | **1587** | **1353** | | **I² = 0.00, Tau² = 0.00** | | | **1278** | **1306** | | **I² = 0.38, Tau² = 0.00** | | |
| **Fixed** | | | | | **−0.04 (−0.06, −0.02)** | | | | | | **−0.04 (−0.07, −0.02)** | | |
| **Random** | | | | | **−0.04 (−0.06, −0.02)** | | | | | | **−0.04 (−0.08, 0.00)** | | |

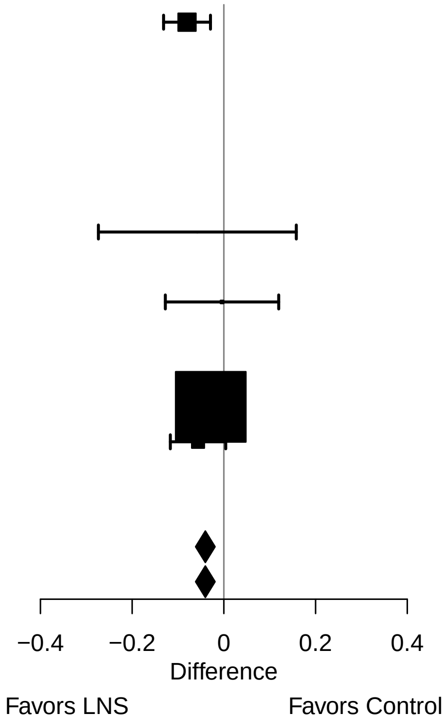

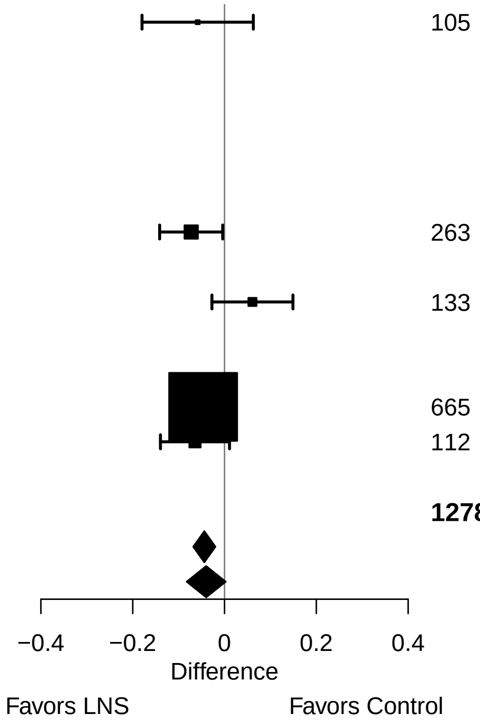

**Supplemental figure 6E: Moderate-to-severe anemia prevalence difference**

#### 6E9: Stratified by Child inflammation

**P−for−interaction = 0.777**
**Difference in PDs = −0.01 (−0.06, 0.05)**

| | | Not inflamed | | | | | | High AGP or CRP | | | | | |
|---|---|---|---|---|---|---|---|---|---|---|---|---|---|
| Country | Trial | LNS N | Control N | Control Prevalence | PD (95% CI) | Fixed W | Random W | LNS N | Control N | Control Prevalence | PD (95% CI) | Fixed W | Random W |
| Bangladesh | JiVitA−4 (35) | | | | | | | | | | | | |
| Bangladesh | RDNS (36) | 369 | 168 | 17.3 | −0.09 (−0.15, −0.02) | 0.16 | 0.23 | 180 | 104 | 15.4 | −0.06 (−0.12, 0.00) | 0.38 | 0.24 |
| Bangladesh | WASH−B (37) | | | | | | | | | | | | |
| Burkina Faso | iLiNS−Zinc (38) | 131 | 33 | 69.7 | −0.27 (−0.45, −0.09) | 0.02 | 0.11 | 197 | 69 | 81.2 | −0.22 (−0.33, −0.11) | 0.12 | 0.18 |
| Burkina Faso | PROMIS CS (39) | | | | | | | | | | | | |
| Ghana | GHANA (40) | | | | | | | | | | | | |
| Ghana | iLiNS−DYADG (41) | | | | | | | | | | | | |
| Kenya | WASH−B (42) | 157 | 135 | 14.1 | −0.08 (−0.14, −0.01) | 0.13 | 0.22 | 141 | 123 | 30.9 | −0.09 (−0.20, 0.02) | 0.12 | 0.17 |
| Madagascar | MAHAY (43) | | | | | | | | | | | | |
| Malawi | iLiNS−DYADM (44) | 59 | 123 | 13.0 | −0.01 (−0.12, 0.09) | 0.06 | 0.18 | 138 | 269 | 29.4 | 0.01 (−0.08, 0.10) | 0.17 | 0.20 |
| Malawi | iLiNS−DOSE (45) | | | | | | | | | | | | |
| Mali | PROMIS CS (46) | | | | | | | | | | | | |
| Zimbabwe | SHINE (HIV−) (47) | 414 | 354 | 5.6 | 0.00 (−0.03, 0.03) | 0.64 | 0.26 | 105 | 99 | 14.1 | −0.06 (−0.14, 0.03) | 0.21 | 0.21 |
| Zimbabwe | SHINE (HIV+) (48) | | | | | | | | | | | | |
| | | **1130** | **813** | | **I² = 0.75, Tau² = 0.01** | | | **761** | **664** | | **I² = 0.63, Tau² = 0.00** | | |
| **Fixed** | | | | | **−0.03 (−0.05, 0.00)** | | | | | | **−0.07 (−0.11, −0.03)** | | |
| **Random** | | | | | **−0.07 (−0.15, 0.01)** | | | | | | **−0.08 (−0.15, −0.01)** | | |

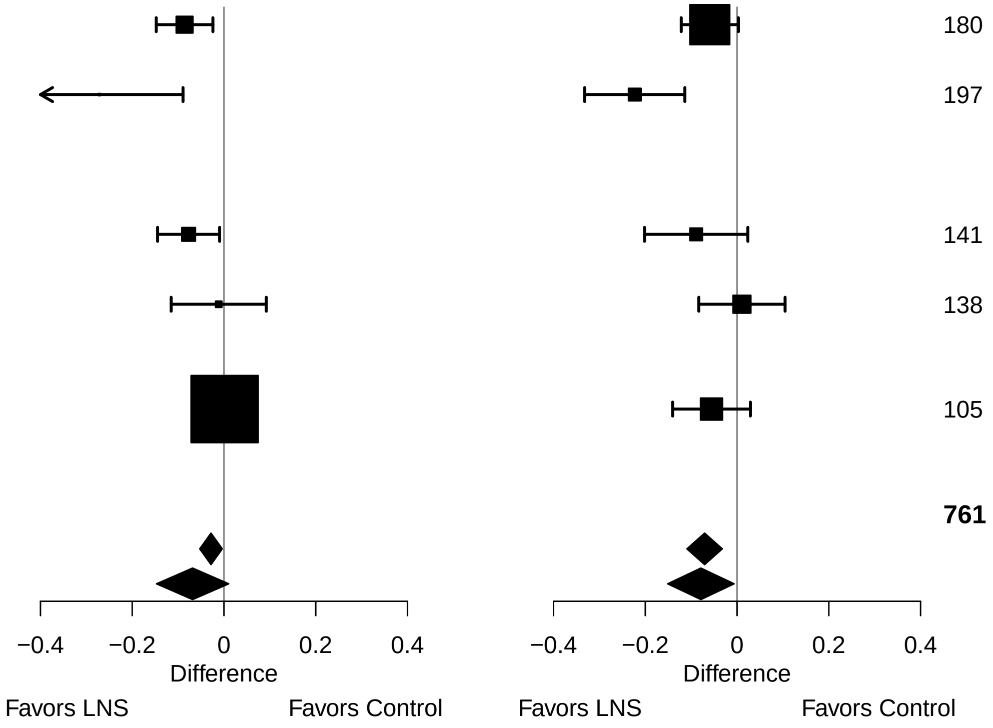

### Supplemental figure 6F: Geometric mean ratio of ferritin concentration

#### 6F1: Stratified by Maternal BMI

**P−for−interaction = 0.221**
**Ratio of GMRs = 0.91 (0.79, 1.06)**

| | | At least 20 kg/m² | | | | | | Less than 20 kg/m² | | | | | |
|---|---|---|---|---|---|---|---|---|---|---|---|---|---|
| | | **LNS** | **Control** | **Control** | **GMR** | **Fixed** | **Random** | **LNS** | **Control** | **Control** | **GMR** | **Fixed** | **Random** |
| Country | Trial | N | N | Median | (95% CI) | W | W | N | N | Median | (95% CI) | W | W |
| Bangladesh | JiVitA−4 (35) | | | | | | | | | | | | |
| Bangladesh | RDNS (36) | 228 | 102 | 19.8 | 1.63 (1.40, 1.90) | 0.33 | 0.33 | 300 | 160 | 21.6 | 1.43 (1.24, 1.64) | 0.45 | 0.44 |
| Bangladesh | WASH−B (37) | 93 | 97 | 15.1 | 1.74 (1.37, 2.20) | 0.14 | 0.14 | 119 | 81 | 16.7 | 1.74 (1.45, 2.09) | 0.25 | 0.26 |
| Burkina Faso | iLiNS−Zinc (38) | 209 | 55 | 10.6 | 1.56 (1.23, 1.96) | 0.14 | 0.14 | 106 | 41 | 8.9 | 1.56 (1.26, 1.94) | 0.18 | 0.19 |
| Burkina Faso | PROMIS CS (39) | | | | | | | | | | | | |
| Ghana | GHANA (40) | | | | | | | | | | | | |
| Ghana | iLiNS−DYADG (41) | | | | | | | | | | | | |
| Kenya | WASH−B (42) | 223 | 200 | 9.8 | 1.73 (1.50, 1.99) | 0.39 | 0.39 | 64 | 51 | 13.1 | 1.41 (1.07, 1.86) | 0.11 | 0.12 |
| Madagascar | MAHAY (43) | | | | | | | | | | | | |
| Malawi | iLiNS−DYADM (44) | | | | | | | | | | | | |
| Malawi | iLiNS−DOSE (45) | | | | | | | | | | | | |
| Mali | PROMIS CS (46) | | | | | | | | | | | | |
| Zimbabwe | SHINE (HIV−) (47) | | | | | | | | | | | | |
| Zimbabwe | SHINE (HIV+) (48) | | | | | | | | | | | | |
| | | **753** | **454** | | **I² = 0.00, Tau² = 0.00** | | | **589** | **333** | | **I² = 0.07, Tau² = 0.00** | | |
| **Fixed** | | | | | **1.67 (1.53, 1.83)** | | | | | | **1.52 (1.39, 1.67)** | | |
| **Random** | | | | | **1.67 (1.53, 1.83)** | | | | | | **1.52 (1.38, 1.68)** | | |

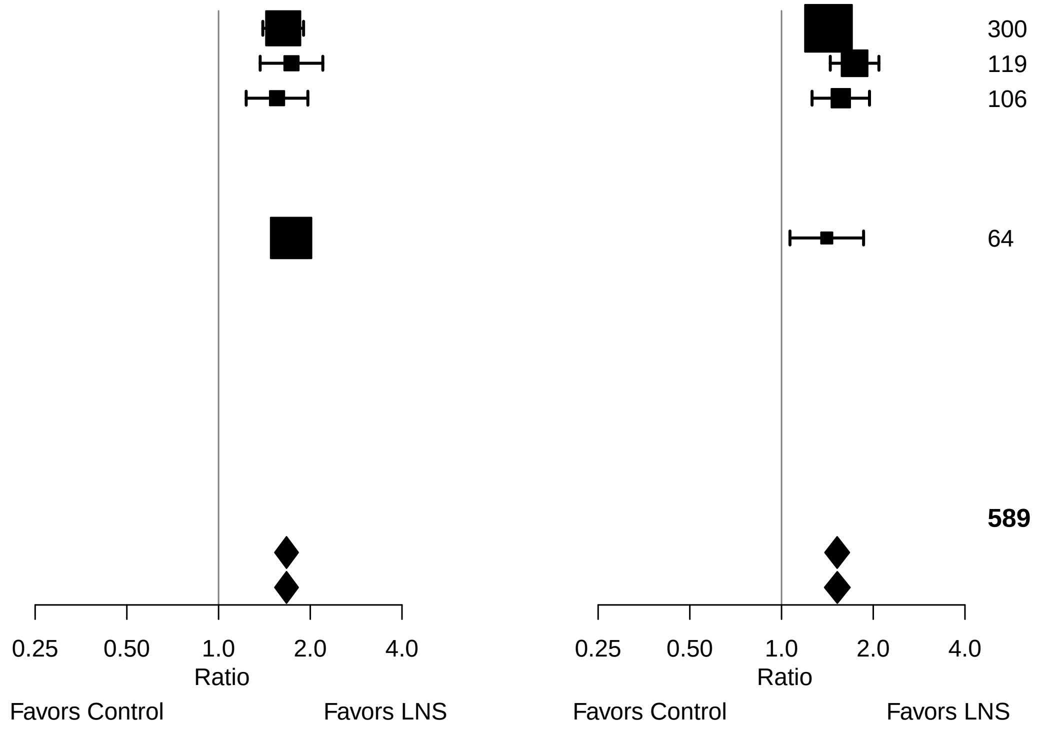

#### Supplemental figure 6F: Geometric mean ratio of ferritin concentration

#### 6F2: Stratified by Maternal age

**P−for−interaction = 0.112**
**Ratio of GMRs = 0.92 (0.83, 1.02)**

| | | At least 25 y | | | | | | Less than 25 y | | | | | |
|---|---|---|---|---|---|---|---|---|---|---|---|---|---|
| Country | Trial | LNS N | Control N | Control Median | GMR (95% CI) | Fixed W | Random W | LNS N | Control N | Control Median | GMR (95% CI) | Fixed W | Random W |
| Bangladesh | JiVitA−4 (35) | 202 | 70 | 23.2 | 1.47 (1.17, 1.85) | 0.11 | 0.11 | 250 | 73 | 25.3 | 1.33 (1.08, 1.63) | 0.10 | 0.16 |
| Bangladesh | RDNS (36) | 152 | 81 | 21.0 | 1.71 (1.47, 1.98) | 0.26 | 0.26 | 398 | 191 | 21.1 | 1.45 (1.32, 1.59) | 0.48 | 0.21 |
| Bangladesh | WASH−B (37) | 79 | 80 | 17.7 | 1.50 (1.20, 1.88) | 0.12 | 0.12 | 132 | 98 | 14.0 | 1.94 (1.64, 2.31) | 0.15 | 0.17 |
| Burkina Faso | iLiNS−Zinc (38) | 178 | 50 | 9.7 | 1.56 (1.30, 1.89) | 0.17 | 0.17 | 137 | 46 | 10.0 | 1.50 (1.19, 1.91) | 0.08 | 0.14 |
| Burkina Faso | PROMIS CS (39) | | | | | | | | | | | | |
| Ghana | GHANA (40) | 61 | 55 | 11.2 | 2.40 (1.32, 4.38) | 0.02 | 0.02 | 22 | 21 | 7.6 | 2.64 (0.81, 8.64) | 0.00 | 0.02 |
| Ghana | iLiNS−DYADG (41) | | | | | | | | | | | | |
| Kenya | WASH−B (42) | 178 | 149 | 9.5 | 1.79 (1.55, 2.08) | 0.26 | 0.26 | 116 | 109 | 10.5 | 1.53 (1.28, 1.82) | 0.14 | 0.17 |
| Madagascar | MAHAY (43) | 47 | 25 | 10.1 | 1.58 (1.17, 2.14) | 0.06 | 0.06 | 36 | 26 | 15.4 | 1.04 (0.79, 1.37) | 0.06 | 0.13 |
| Malawi | iLiNS−DYADM (44) | | | | | | | | | | | | |
| Malawi | iLiNS−DOSE (45) | | | | | | | | | | | | |
| Mali | PROMIS CS (46) | | | | | | | | | | | | |
| Zimbabwe | SHINE (HIV−) (47) | | | | | | | | | | | | |
| Zimbabwe | SHINE (HIV+) (48) | | | | | | | | | | | | |
| | | **897** | **510** | | **I² = 0.00, Tau² = 0.00** | | | **1091** | **564** | | **I² = 0.67, Tau² = 0.03** | | |
| **Fixed** | | | | | **1.65 (1.53, 1.78)** | | | | | | **1.49 (1.39, 1.59)** | | |
| **Random** | | | | | **1.65 (1.53, 1.78)** | | | | | | **1.48 (1.27, 1.72)** | | |

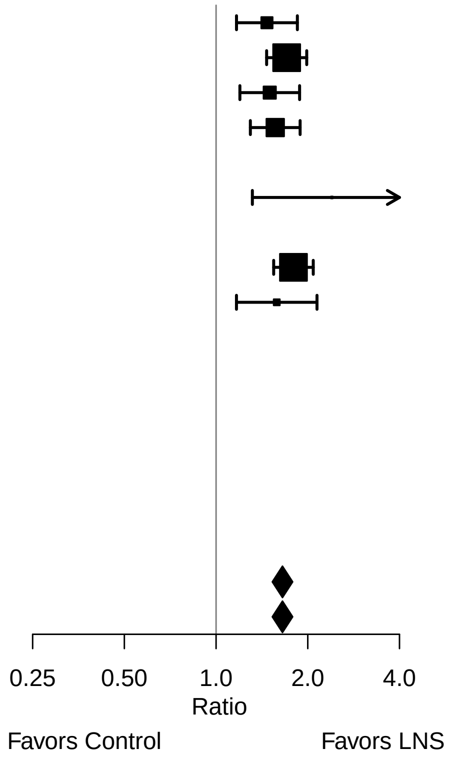

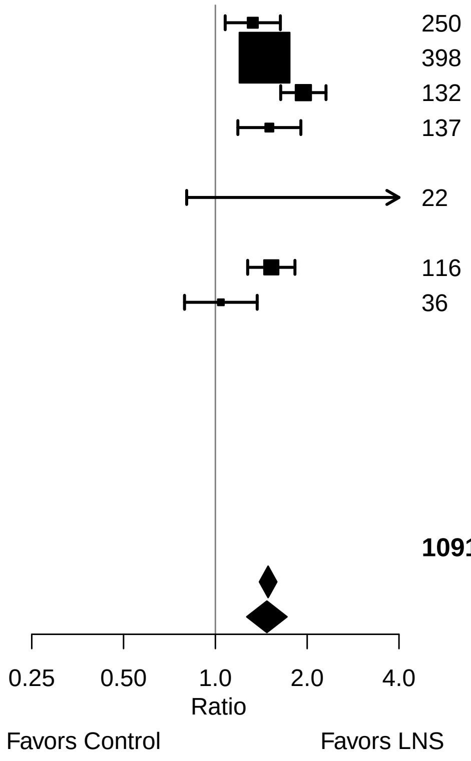

#### Supplemental figure 6F: Geometric mean ratio of ferritin concentration

#### 6F3: Stratified by Maternal education

**P−for−interaction = 0.928**
**Ratio of GMRs = 0.99 (0.89, 1.12)**

| | | Primary or greater | | | | | | Incomplete or no formal | | | | | |
|---|---|---|---|---|---|---|---|---|---|---|---|---|---|
| Country | Trial | LNS N | Control N | Control Median | GMR (95% CI) | Fixed W | Random W | LNS N | Control N | Control Median | GMR (95% CI) | Fixed W | Random W |
| Bangladesh | JiVitA−4 (35) | 298 | 93 | 21.0 | 1.57 (1.28, 1.92) | 0.10 | 0.17 | 156 | 51 | 29.8 | 1.10 (0.88, 1.38) | 0.16 | 0.19 |
| Bangladesh | RDNS (36) | 406 | 195 | 21.2 | 1.46 (1.34, 1.60) | 0.53 | 0.22 | 144 | 77 | 18.7 | 1.66 (1.42, 1.94) | 0.34 | 0.26 |
| Bangladesh | WASH−B (37) | 149 | 139 | 15.0 | 1.84 (1.56, 2.17) | 0.15 | 0.19 | 63 | 39 | 16.7 | 1.45 (1.05, 1.99) | 0.08 | 0.13 |
| Burkina Faso | iLiNS−Zinc (38) | | | | | | | | | | | | |
| Burkina Faso | PROMIS CS (39) | | | | | | | | | | | | |
| Ghana | GHANA (40) | 76 | 65 | 9.4 | 2.37 (1.35, 4.15) | 0.01 | 0.06 | 7 | 13 | 11.2 | 1.70 (0.28, 10.25) | 0.00 | 0.01 |
| Ghana | iLiNS−DYADG (41) | | | | | | | | | | | | |
| Kenya | WASH−B (42) | 143 | 117 | 10.5 | 1.71 (1.39, 2.11) | 0.09 | 0.17 | 154 | 142 | 10.0 | 1.69 (1.43, 1.99) | 0.30 | 0.25 |
| Madagascar | MAHAY (43) | 16 | 16 | 15.4 | 1.17 (0.98, 1.41) | 0.12 | 0.18 | 67 | 35 | 11.3 | 1.35 (1.04, 1.76) | 0.12 | 0.16 |
| Malawi | iLiNS−DYADM (44) | | | | | | | | | | | | |
| Malawi | iLiNS−DOSE (45) | | | | | | | | | | | | |
| Mali | PROMIS CS (46) | | | | | | | | | | | | |
| Zimbabwe | SHINE (HIV−) (47) | | | | | | | | | | | | |
| Zimbabwe | SHINE (HIV+) (48) | | | | | | | | | | | | |
| | | **1088** | **625** | | **I² = 0.71, Tau² = 0.03** | | | **591** | **357** | | **I² = 0.55, Tau² = 0.01** | | |
| **Fixed** | | | | | **1.52 (1.42, 1.62)** | | | | | | **1.51 (1.38, 1.65)** | | |
| **Random** | | | | | **1.57 (1.34, 1.84)** | | | | | | **1.46 (1.27, 1.69)** | | |

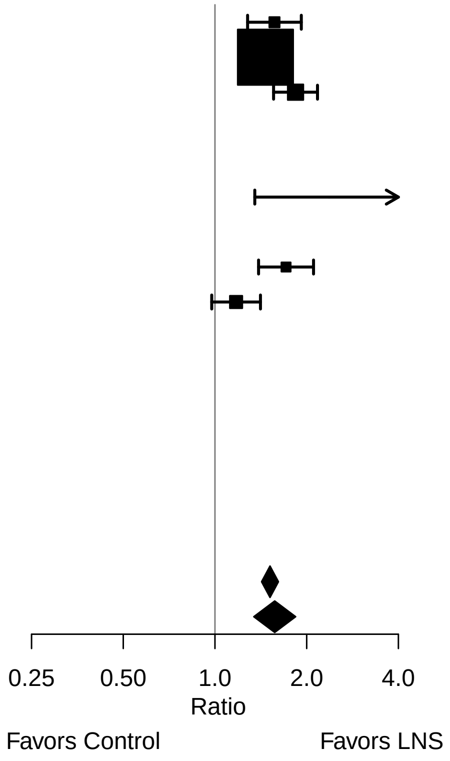

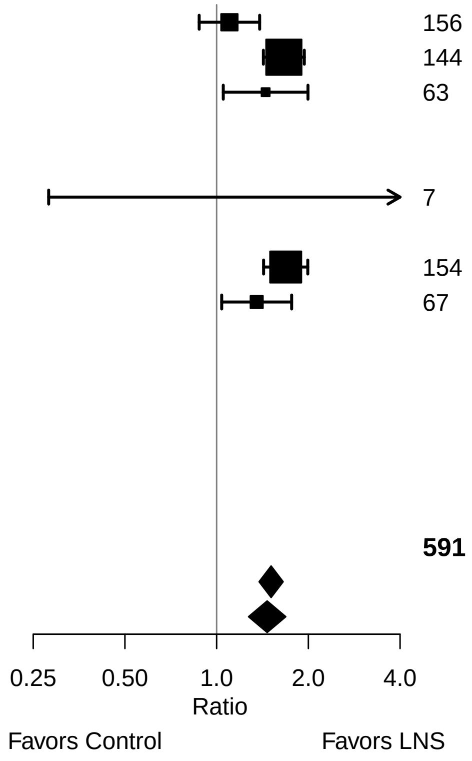

**Supplemental figure 6F: Geometric mean ratio of ferritin concentration**

#### 6F4: Stratified by Child sex

**P−for−interaction = 0.649**
**Ratio of GMRs = 0.98 (0.88, 1.08)**

| Country | Trial | Male LNS N | Male Control N | Male Control Median | Male GMR (95% CI) | Male Fixed W | Male Random W | Female LNS N | Female Control N | Female Control Median | Female GMR (95% CI) | Female Fixed W | Female Random W |
|---|---|---|---|---|---|---|---|---|---|---|---|---|---|
| Bangladesh | JiVitA−4 (35) | 235 | 70 | 21.1 | 1.45 (1.15, 1.84) | 0.10 | 0.14 | 220 | 74 | 27.7 | 1.33 (1.09, 1.64) | 0.12 | 0.15 |
| Bangladesh | RDNS (36) | 266 | 131 | 18.1 | 1.53 (1.38, 1.71) | 0.47 | 0.25 | 281 | 141 | 22.5 | 1.51 (1.33, 1.71) | 0.31 | 0.22 |
| Bangladesh | WASH−B (37) | 105 | 85 | 16.2 | 1.67 (1.32, 2.10) | 0.10 | 0.15 | 107 | 93 | 15.7 | 1.81 (1.52, 2.17) | 0.16 | 0.17 |
| Burkina Faso | iLiNS−Zinc (38) | 150 | 49 | 10.5 | 1.62 (1.23, 2.14) | 0.07 | 0.12 | 165 | 47 | 9.5 | 1.51 (1.21, 1.89) | 0.11 | 0.14 |
| Burkina Faso | PROMIS CS (39) | | | | | | | | | | | | |
| Ghana | GHANA (40) | 51 | 37 | 6.9 | 3.87 (1.83, 8.18) | 0.01 | 0.02 | 32 | 45 | 16.5 | 1.93 (0.94, 3.98) | 0.01 | 0.02 |
| Ghana | iLiNS−DYADG (41) | | | | | | | | | | | | |
| Kenya | WASH−B (42) | 146 | 125 | 10.8 | 1.65 (1.38, 1.97) | 0.17 | 0.19 | 152 | 134 | 9.9 | 1.74 (1.49, 2.03) | 0.21 | 0.19 |
| Madagascar | MAHAY (43) | 34 | 22 | 10.6 | 1.42 (1.09, 1.85) | 0.08 | 0.13 | 49 | 29 | 14.2 | 1.21 (0.93, 1.57) | 0.07 | 0.11 |
| Malawi | iLiNS−DYADM (44) | | | | | | | | | | | | |
| Malawi | iLiNS−DOSE (45) | | | | | | | | | | | | |
| Mali | PROMIS CS (46) | | | | | | | | | | | | |
| Zimbabwe | SHINE (HIV−) (47) | | | | | | | | | | | | |
| Zimbabwe | SHINE (HIV+) (48) | | | | | | | | | | | | |
| | | **987** | **519** | | **I² = 0.18, Tau² = 0.01** | | | **1006** | **563** | | **I² = 0.46, Tau² = 0.01** | | |
| **Fixed** | | | | | **1.57 (1.46, 1.69)** | | | | | | **1.56 (1.45, 1.67)** | | |
| **Random** | | | | | **1.59 (1.41, 1.79)** | | | | | | **1.54 (1.38, 1.72)** | | |

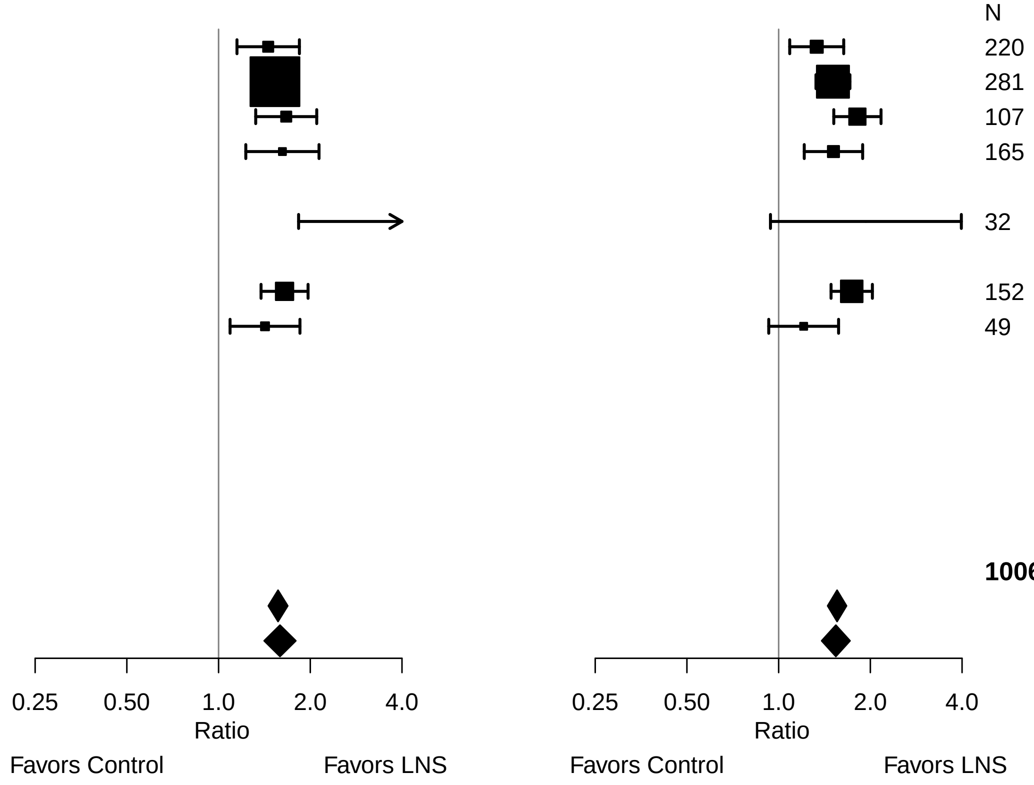

**Supplemental figure 6F: Geometric mean ratio of ferritin concentration**

#### 6F5: Stratified by Child birth order

**P−for−interaction = 0.040**
**Ratio of GMRs = 0.88 (0.78, 0.99)**

| | | Later born | | | | | | Firstborn | | | | | |
|---|---|---|---|---|---|---|---|---|---|---|---|---|---|
| Country | Trial | LNS N | Control N | Control Median | GMR (95% CI) | Fixed W | Random W | LNS N | Control N | Control Median | GMR (95% CI) | Fixed W | Random W |
| Bangladesh | JiVitA−4 (35) | 114 | 32 | 28.6 | 1.26 (0.88, 1.80) | 0.03 | 0.03 | 340 | 111 | 23.2 | 1.42 (1.20, 1.68) | 0.27 | 0.19 |
| Bangladesh | RDNS (36) | 340 | 171 | 19.8 | 1.64 (1.50, 1.80) | 0.47 | 0.47 | 210 | 101 | 22.5 | 1.33 (1.14, 1.56) | 0.31 | 0.19 |
| Bangladesh | WASH−B (37) | 124 | 119 | 16.7 | 1.75 (1.44, 2.12) | 0.11 | 0.11 | 86 | 57 | 15.1 | 1.74 (1.36, 2.22) | 0.13 | 0.16 |
| Burkina Faso | iLiNS−Zinc (38) | 249 | 71 | 9.2 | 1.59 (1.30, 1.94) | 0.10 | 0.10 | 66 | 25 | 11.4 | 1.42 (1.11, 1.83) | 0.12 | 0.16 |
| Burkina Faso | PROMIS CS (39) | | | | | | | | | | | | |
| Ghana | GHANA (40) | 54 | 50 | 11.6 | 2.27 (1.25, 4.11) | 0.01 | 0.01 | 29 | 29 | 7.6 | 2.68 (0.96, 7.46) | 0.01 | 0.03 |
| Ghana | iLiNS−DYADG (41) | | | | | | | | | | | | |
| Kenya | WASH−B (42) | 237 | 213 | 10.1 | 1.71 (1.49, 1.97) | 0.21 | 0.21 | 60 | 46 | 9.3 | 1.63 (1.23, 2.17) | 0.09 | 0.14 |
| Madagascar | MAHAY (43) | 61 | 37 | 11.3 | 1.43 (1.12, 1.82) | 0.07 | 0.07 | 21 | 11 | 16.8 | 0.91 (0.67, 1.24) | 0.08 | 0.13 |
| Malawi | iLiNS−DYADM (44) | | | | | | | | | | | | |
| Malawi | iLiNS−DOSE (45) | | | | | | | | | | | | |
| Mali | PROMIS CS (46) | | | | | | | | | | | | |
| Zimbabwe | SHINE (HIV−) (47) | | | | | | | | | | | | |
| Zimbabwe | SHINE (HIV+) (48) | | | | | | | | | | | | |
| | | **1179** | **693** | | **I² = 0.00, Tau² = 0.00** | | | **812** | **380** | | **I² = 0.55, Tau² = 0.03** | | |
| **Fixed** | | | | | **1.64 (1.54, 1.75)** | | | | | | **1.40 (1.29, 1.53)** | | |
| **Random** | | | | | **1.64 (1.54, 1.75)** | | | | | | **1.41 (1.19, 1.68)** | | |

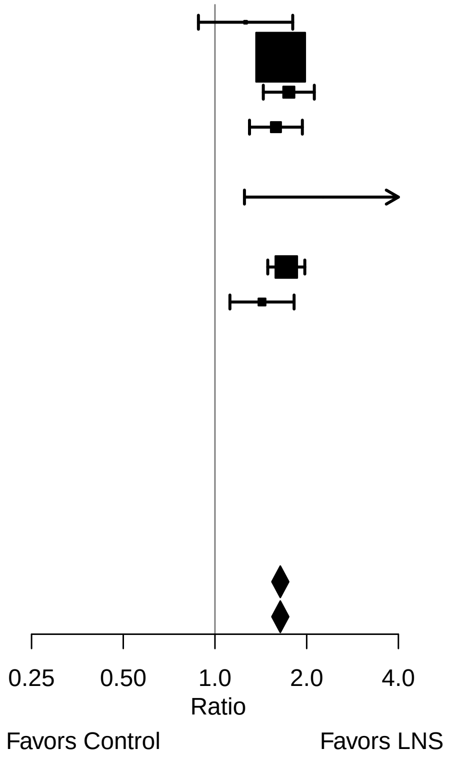

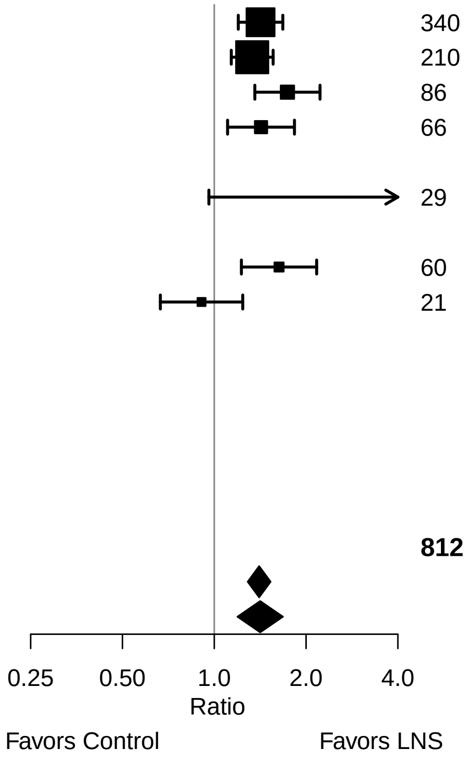

**Supplemental figure 6F: Geometric mean ratio of ferritin concentration**

#### 6F6: Stratified by Child baseline acute malnutrition

**P−for−interaction = 0.023**
**Ratio of GMRs = 1.27 (1.03, 1.56)**

| | | No | | | | | | Yes | | | | | |
|---|---|---|---|---|---|---|---|---|---|---|---|---|---|
| | | LNS | Control | Control | GMR | Fixed | Random | LNS | Control | Control | GMR | Fixed | Random |
| Country | Trial | N | N | Median | (95% CI) | W | W | N | N | Median | (95% CI) | W | W |
| Bangladesh | JiVitA−4 (35) | 363 | 112 | 23.1 | 1.37 (1.17, 1.61) | 0.17 | 0.17 | 90 | 32 | 26.4 | 1.43 (0.92, 2.20) | 0.19 | 0.22 |
| Bangladesh | RDNS (36) | 493 | 243 | 21.0 | 1.51 (1.40, 1.64) | 0.72 | 0.72 | 35 | 12 | 17.0 | 1.76 (1.28, 2.41) | 0.36 | 0.36 |
| Bangladesh | WASH−B (37) | | | | | | | | | | | | |
| Burkina Faso | iLiNS−Zinc (38) | 229 | 72 | 10.7 | 1.38 (1.12, 1.69) | 0.10 | 0.10 | 86 | 24 | 8.1 | 2.16 (1.63, 2.87) | 0.45 | 0.42 |
| Burkina Faso | PROMIS CS (39) | | | | | | | | | | | | |
| Ghana | GHANA (40) | | | | | | | | | | | | |
| Ghana | iLiNS−DYADG (41) | | | | | | | | | | | | |
| Kenya | WASH−B (42) | | | | | | | | | | | | |
| Madagascar | MAHAY (43) | | | | | | | | | | | | |
| Malawi | iLiNS−DYADM (44) | | | | | | | | | | | | |
| Malawi | iLiNS−DOSE (45) | | | | | | | | | | | | |
| Mali | PROMIS CS (46) | | | | | | | | | | | | |
| Zimbabwe | SHINE (HIV−) (47) | | | | | | | | | | | | |
| Zimbabwe | SHINE (HIV+) (48) | | | | | | | | | | | | |
| | | **1085** | **427** | | **I² = 0.00, Tau² = 0.00** | | | **211** | **68** | | **I² = 0.25, Tau² = 0.01** | | |
| **Fixed** | | | | | **1.47 (1.38, 1.57)** | | | | | | **1.85 (1.53, 2.24)** | | |
| **Random** | | | | | **1.47 (1.38, 1.57)** | | | | | | **1.83 (1.46, 2.29)** | | |

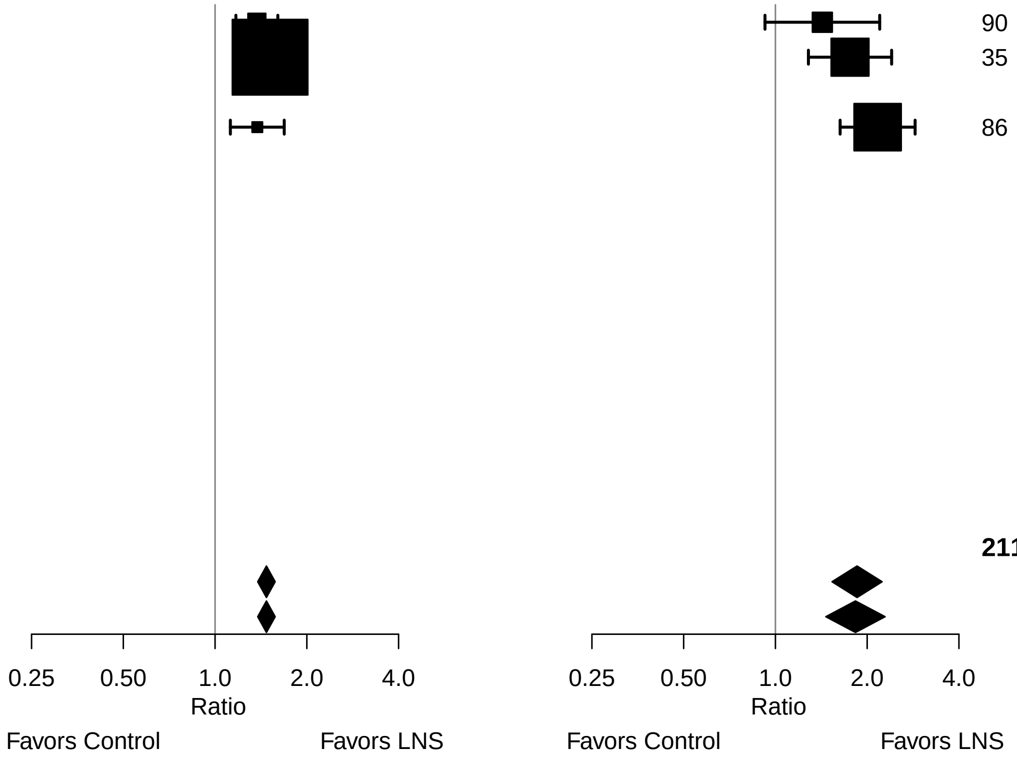

**Supplemental figure 6F: Geometric mean ratio of ferritin concentration**

#### 6F7: Stratified by Child baseline anemia (insufficient comparisons)

##### Supplemental figure 6F: Geometric mean ratio of ferritin concentration

#### 6F8: Stratified by Child high-dose vitamin A supplementation

**P−for−interaction = 0.348**
**Ratio of GMRs = 1.08 (0.92, 1.27)**

| Country | Trial | Received VitA: LNS N | Control N | Control Median | GMR (95% CI) | Fixed W | Random W | Not treated: LNS N | Control N | Control Median | GMR (95% CI) | Fixed W | Random W |
|---|---|---|---|---|---|---|---|---|---|---|---|---|---|
| Bangladesh | JiVitA−4 (35) | 140 | 37 | 24.2 | 1.42 (1.11, 1.82) | 0.08 | 0.23 | 133 | 41 | 22.6 | 1.27 (0.98, 1.65) | 0.18 | 0.21 |
| Bangladesh | RDNS (36) | 444 | 223 | 21.3 | 1.47 (1.36, 1.59) | 0.84 | 0.50 | 105 | 49 | 17.6 | 1.74 (1.40, 2.17) | 0.24 | 0.27 |
| Bangladesh | WASH−B (37) | 16 | 16 | 13.1 | 1.65 (0.99, 2.73) | 0.02 | 0.08 | 15 | 9 | 17.5 | 1.51 (0.89, 2.58) | 0.04 | 0.06 |
| Burkina Faso | iLiNS−Zinc (38) | | | | | | | | | | | | |
| Burkina Faso | PROMIS CS (39) | | | | | | | | | | | | |
| Ghana | GHANA (40) | 39 | 53 | 8.5 | 3.37 (1.53, 7.45) | 0.01 | 0.03 | 44 | 26 | 14.0 | 1.26 (0.64, 2.45) | 0.03 | 0.04 |
| Ghana | iLiNS−DYADG (41) | | | | | | | | | | | | |
| Kenya | WASH−B (42) | 26 | 31 | 12.5 | 1.49 (1.08, 2.05) | 0.05 | 0.16 | 223 | 180 | 9.5 | 1.74 (1.49, 2.03) | 0.51 | 0.43 |
| Madagascar | MAHAY (43) | | | | | | | | | | | | |
| Malawi | iLiNS−DYADM (44) | | | | | | | | | | | | |
| Malawi | iLiNS−DOSE (45) | | | | | | | | | | | | |
| Mali | PROMIS CS (46) | | | | | | | | | | | | |
| Zimbabwe | SHINE (HIV−) (47) | | | | | | | | | | | | |
| Zimbabwe | SHINE (HIV+) (48) | | | | | | | | | | | | |
| | | **665** | **360** | | **I² = 0.10, Tau² = 0.01** | | | **520** | **305** | | **I² = 0.23, Tau² = 0.00** | | |
| **Fixed** | | | | | **1.48 (1.38, 1.59)** | | | | | | **1.62 (1.45, 1.81)** | | |
| **Random** | | | | | **1.52 (1.31, 1.77)** | | | | | | **1.60 (1.40, 1.82)** | | |

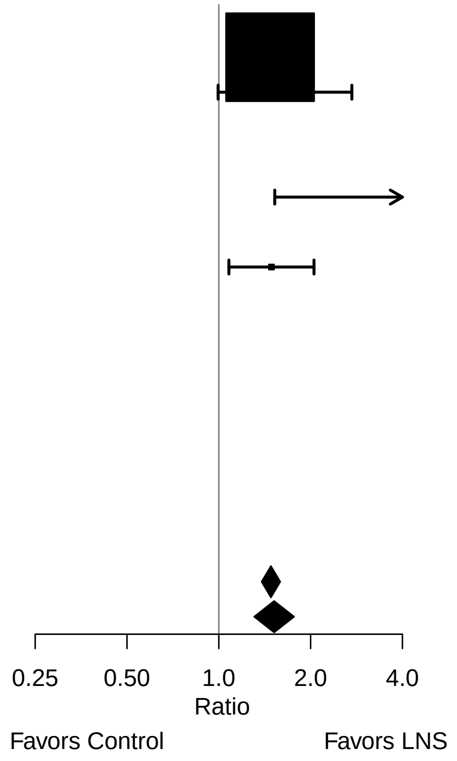

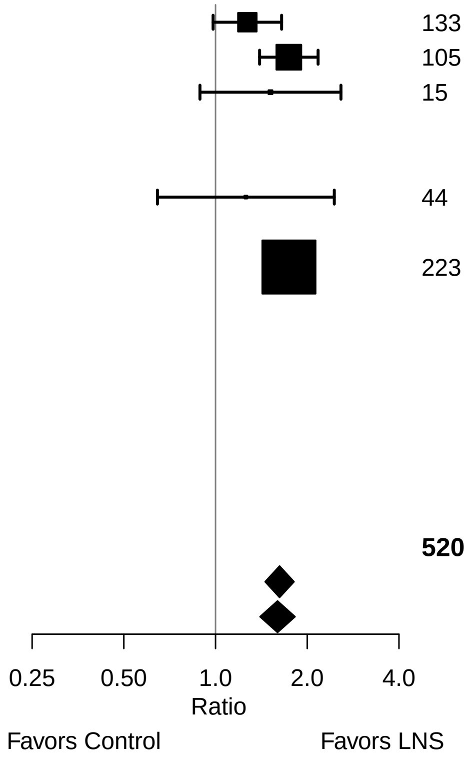

#### Supplemental figure 6F: Geometric mean ratio of ferritin concentration

#### 6F9: Stratified by Child inflammation

**P−for−interaction = 0.089**
**Ratio of GMRs = 0.90 (0.81, 1.02)**

| Country | Trial | Not inflamed LNS N | Not inflamed Control N | Not inflamed Control Median | Not inflamed GMR (95% CI) | Not inflamed Fixed W | Not inflamed Random W | High AGP or CRP LNS N | High AGP or CRP Control N | High AGP or CRP Control Median | High AGP or CRP GMR (95% CI) | High AGP or CRP Fixed W | High AGP or CRP Random W |
|---|---|---|---|---|---|---|---|---|---|---|---|---|---|
| Bangladesh | JiVitA−4 (35) | 185 | 46 | 23.4 | 1.53 (1.09, 2.14) | 0.04 | 0.12 | 270 | 98 | 24.4 | 1.31 (1.08, 1.60) | 0.16 | 0.18 |
| Bangladesh | RDNS (36) | 370 | 168 | 19.3 | 1.58 (1.42, 1.77) | 0.39 | 0.18 | 180 | 104 | 21.9 | 1.41 (1.23, 1.61) | 0.34 | 0.25 |
| Bangladesh | WASH−B (37) | 165 | 136 | 13.3 | 1.85 (1.61, 2.14) | 0.25 | 0.18 | 47 | 42 | 20.0 | 1.42 (1.04, 1.93) | 0.07 | 0.10 |
| Burkina Faso | iLiNS−Zinc (38) | 124 | 31 | 9.4 | 1.86 (1.47, 2.35) | 0.09 | 0.15 | 191 | 65 | 9.7 | 1.40 (1.12, 1.76) | 0.12 | 0.15 |
| Burkina Faso | PROMIS CS (39) | | | | | | | | | | | | |
| Ghana | GHANA (40) | 69 | 66 | 7.6 | 3.35 (1.87, 5.97) | 0.01 | 0.07 | 14 | 16 | 33.0 | 0.79 (0.28, 2.22) | 0.01 | 0.01 |
| Ghana | iLiNS−DYADG (41) | | | | | | | | | | | | |
| Kenya | WASH−B (42) | 157 | 135 | 10.1 | 1.56 (1.28, 1.89) | 0.13 | 0.16 | 141 | 124 | 10.0 | 1.86 (1.58, 2.18) | 0.24 | 0.22 |
| Madagascar | MAHAY (43) | 44 | 32 | 11.4 | 1.23 (0.97, 1.57) | 0.08 | 0.15 | 39 | 19 | 16.1 | 1.36 (0.98, 1.89) | 0.06 | 0.09 |
| Malawi | iLiNS−DYADM (44) | | | | | | | | | | | | |
| Malawi | iLiNS−DOSE (45) | | | | | | | | | | | | |
| Mali | PROMIS CS (46) | | | | | | | | | | | | |
| Zimbabwe | SHINE (HIV−) (47) | | | | | | | | | | | | |
| Zimbabwe | SHINE (HIV+) (48) | | | | | | | | | | | | |
| | | **1114** | **614** | | **$I^2$ = 0.63, $Tau^2$ = 0.04** | | | **882** | **468** | | **$I^2$ = 0.48, $Tau^2$ = 0.01** | | |
| **Fixed** | | | | | **1.65 (1.53, 1.76)** | | | | | | **1.48 (1.37, 1.60)** | | |
| **Random** | | | | | **1.68 (1.40, 2.01)** | | | | | | **1.46 (1.31, 1.64)** | | |

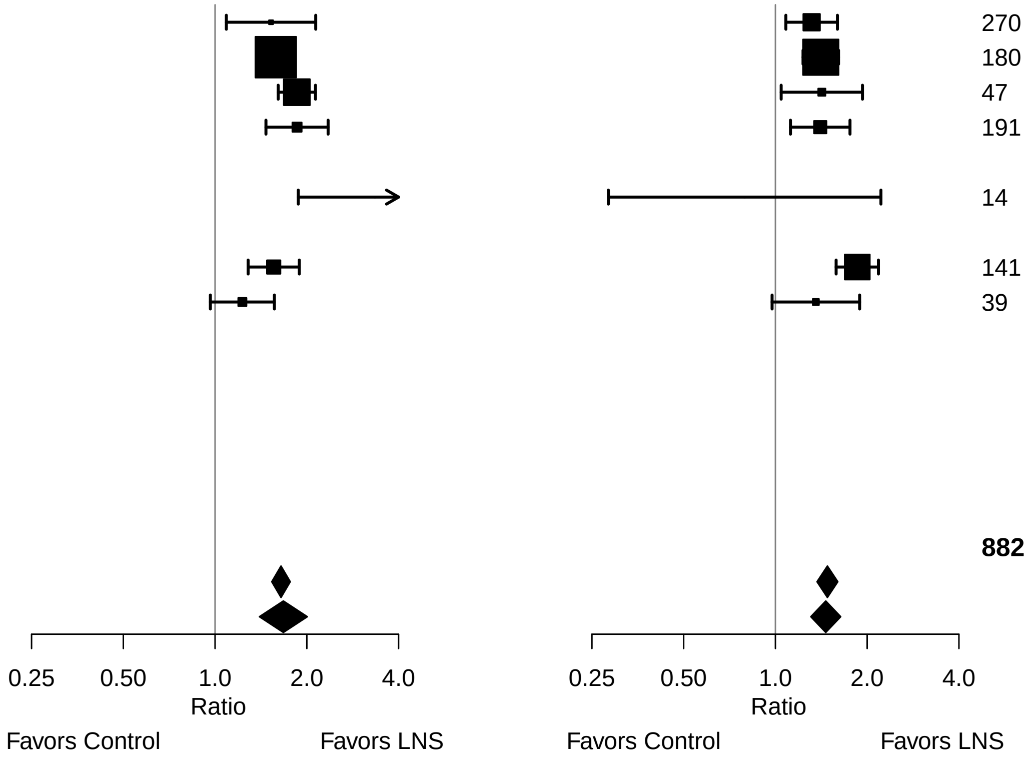

### Supplemental figure 6G: Iron deficiency (ferritin < 12 µg/L) prevalence ratio

#### 6G1: Stratified by Maternal BMI

**P−for−interaction = 0.786**
**Ratio of PRs = 0.96 (0.71, 1.30)**

**At least 20 kg/m²**

| Country | Trial | **LNS** N | **Control** N | **Control** Prevalence | **PR** (95% CI) | **Fixed** W | **Random** W |
|---|---|---|---|---|---|---|---|
| Bangladesh | JiVitA−4 (35) | | | | | | |
| Bangladesh | RDNS (36) | 228 | 102 | 22.5 | 0.23 (0.12, 0.44) | 0.10 | 0.21 |
| Bangladesh | WASH−B (37) | 93 | 97 | 36.1 | 0.30 (0.15, 0.61) | 0.08 | 0.19 |
| Burkina Faso | iLiNS−Zinc (38) | 209 | 55 | 54.1 | 0.69 (0.50, 0.94) | 0.40 | 0.30 |
| Burkina Faso | PROMIS CS (39) | | | | | | |
| Ghana | GHANA (40) | | | | | | |
| Ghana | iLiNS−DYADG (41) | | | | | | |
| Kenya | WASH−B (42) | 223 | 200 | 61.5 | 0.41 (0.30, 0.56) | 0.42 | 0.30 |
| Madagascar | MAHAY (43) | | | | | | |
| Malawi | iLiNS−DYADM (44) | | | | | | |
| Malawi | iLiNS−DOSE (45) | | | | | | |
| Mali | PROMIS CS (46) | | | | | | |
| Zimbabwe | SHINE (HIV−) (47) | | | | | | |
| Zimbabwe | SHINE (HIV+) (48) | | | | | | |
| | | **753** | **454** | | **I² = 0.77, Tau² = 0.16** | | |
| **Fixed** | | | | | **0.47 (0.38, 0.57)** | | |
| **Random** | | | | | **0.40 (0.25, 0.63)** | | |

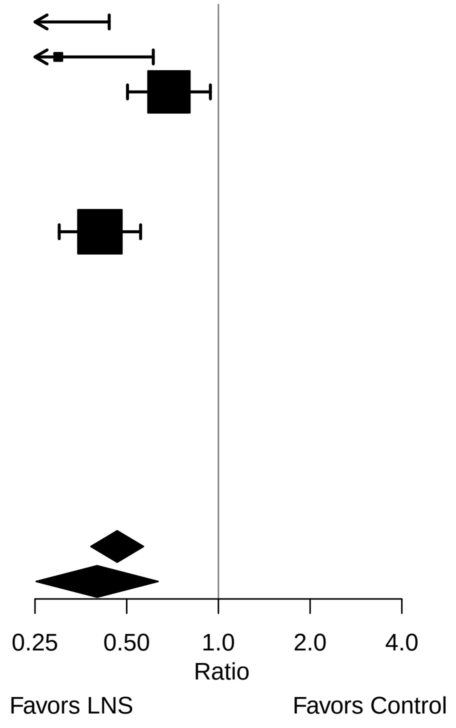

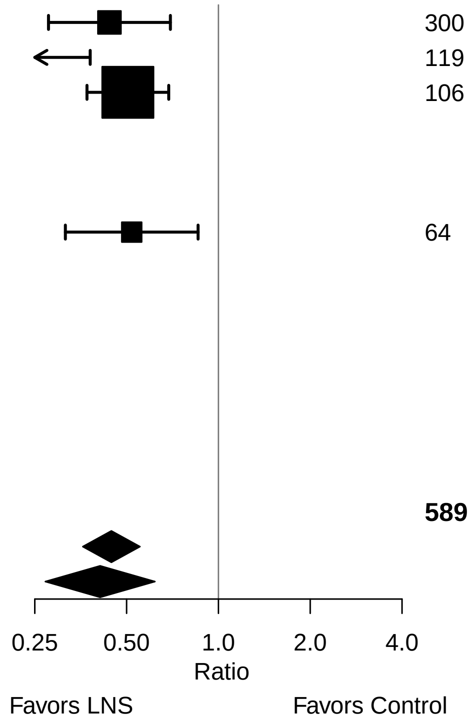

**Less than 20 kg/m²**

| Country | Trial | **LNS** N | **Control** N | **Control** Prevalence | **PR** (95% CI) | **Fixed** W | **Random** W |
|---|---|---|---|---|---|---|---|
| Bangladesh | RDNS (36) | 300 | 160 | 21.2 | 0.44 (0.28, 0.70) | 0.22 | 0.26 |
| Bangladesh | WASH−B (37) | 119 | 81 | 34.6 | 0.19 (0.10, 0.38) | 0.10 | 0.19 |
| Burkina Faso | iLiNS−Zinc (38) | 106 | 41 | 64.4 | 0.50 (0.37, 0.69) | 0.49 | 0.31 |
| Kenya | WASH−B (42) | 64 | 51 | 45.1 | 0.52 (0.31, 0.86) | 0.19 | 0.24 |
| | | **589** | **333** | | **I² = 0.56, Tau² = 0.12** | | |
| **Fixed** | | | | | **0.45 (0.36, 0.55)** | | |
| **Random** | | | | | **0.41 (0.27, 0.62)** | | |

#### Supplemental figure 6G: Iron deficiency (ferritin < 12 μg/L) prevalence ratio

##### 6G2: Stratified by Maternal age

**P−for−interaction = 0.131**
**Ratio of PRs = 1.19 (0.95, 1.48)**

| | | At least 25 y | | | | | | Less than 25 y | | | | | |
|---|---|---|---|---|---|---|---|---|---|---|---|---|---|
| Country | Trial | LNS N | Control N | Control Prevalence | PR (95% CI) | Fixed W | Random W | LNS N | Control N | Control Prevalence | PR (95% CI) | Fixed W | Random W |
| Bangladesh | JiVitA−4 (35) | 202 | 70 | 22.9 | 0.26 (0.13, 0.52) | 0.06 | 0.07 | 250 | 73 | 21.9 | 0.44 (0.23, 0.85) | 0.07 | 0.12 |
| Bangladesh | RDNS (36) | 152 | 81 | 22.2 | 0.27 (0.12, 0.60) | 0.04 | 0.05 | 398 | 191 | 22.0 | 0.37 (0.25, 0.54) | 0.20 | 0.17 |
| Bangladesh | WASH−B (37) | 79 | 80 | 27.5 | 0.41 (0.20, 0.87) | 0.05 | 0.07 | 132 | 98 | 41.8 | 0.16 (0.08, 0.31) | 0.07 | 0.12 |
| Burkina Faso | iLiNS−Zinc (38) | 178 | 50 | 59.3 | 0.55 (0.43, 0.72) | 0.41 | 0.35 | 137 | 46 | 57.7 | 0.71 (0.53, 0.95) | 0.34 | 0.19 |
| Burkina Faso | PROMIS CS (39) | | | | | | | | | | | | |
| Ghana | GHANA (40) | 61 | 55 | 52.7 | 0.47 (0.28, 0.77) | 0.11 | 0.13 | 22 | 21 | 61.9 | 0.59 (0.31, 1.12) | 0.07 | 0.12 |
| Ghana | iLiNS−DYADG (41) | | | | | | | | | | | | |
| Kenya | WASH−B (42) | 178 | 149 | 60.4 | 0.40 (0.29, 0.55) | 0.29 | 0.27 | 116 | 109 | 56.9 | 0.44 (0.30, 0.65) | 0.19 | 0.17 |
| Madagascar | MAHAY (43) | 47 | 25 | 60.0 | 0.32 (0.14, 0.71) | 0.04 | 0.06 | 36 | 26 | 42.3 | 0.46 (0.22, 0.95) | 0.06 | 0.11 |
| Malawi | iLiNS−DYADM (44) | | | | | | | | | | | | |
| Malawi | iLiNS−DOSE (45) | | | | | | | | | | | | |
| Mali | PROMIS CS (46) | | | | | | | | | | | | |
| Zimbabwe | SHINE (HIV−) (47) | | | | | | | | | | | | |
| Zimbabwe | SHINE (HIV+) (48) | | | | | | | | | | | | |
| | | **897** | **510** | | **I² = 0.21, Tau² = 0.01** | | | **1091** | **564** | | **I² = 0.69, Tau² = 0.13** | | |
| **Fixed** | | | | | **0.44 (0.37, 0.52)** | | | | | | **0.48 (0.40, 0.57)** | | |
| **Random** | | | | | **0.43 (0.35, 0.52)** | | | | | | **0.43 (0.31, 0.60)** | | |

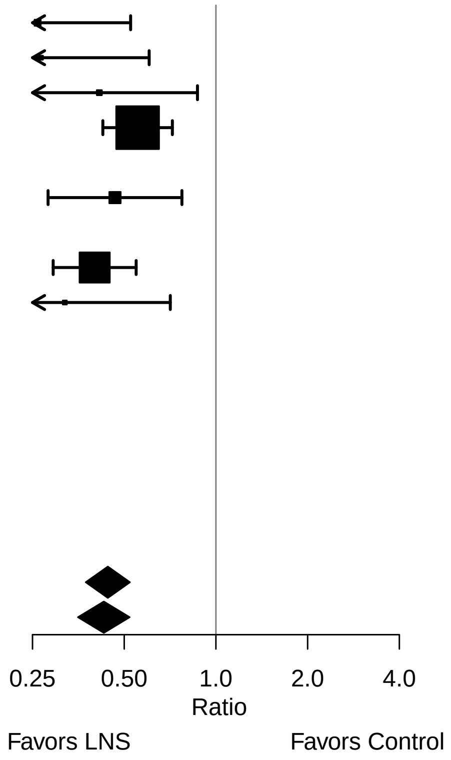

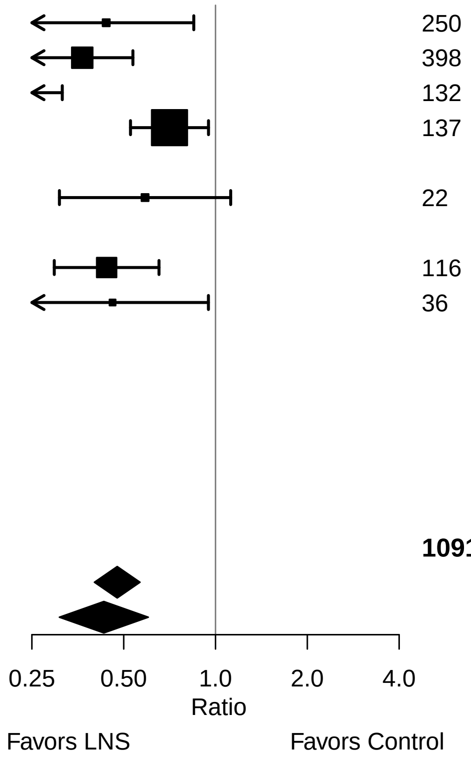

#### Supplemental figure 6G: Iron deficiency (ferritin < 12 µg/L) prevalence ratio

##### 6G3: Stratified by Maternal education

**P−for−interaction = 0.783**
**Ratio of PRs = 0.95 (0.66, 1.37)**

| | | Primary or greater | | | | | | Incomplete or no formal | | | | | |
|---|---|---|---|---|---|---|---|---|---|---|---|---|---|
| Country | Trial | LNS N | Control N | Control Prevalence | PR (95% CI) | Fixed W | Random W | LNS N | Control N | Control Prevalence | PR (95% CI) | Fixed W | Random W |
| Bangladesh | JiVitA−4 (35) | 298 | 93 | 26.9 | 0.31 (0.18, 0.55) | 0.17 | 0.18 | 156 | 51 | 13.7 | 0.51 (0.20, 1.32) | 0.08 | 0.08 |
| Bangladesh | RDNS (36) | 406 | 195 | 18.5 | 0.40 (0.26, 0.62) | 0.28 | 0.28 | 144 | 77 | 31.2 | 0.25 (0.13, 0.45) | 0.20 | 0.20 |
| Bangladesh | WASH−B (37) | 149 | 139 | 38.1 | 0.23 (0.12, 0.42) | 0.14 | 0.16 | 63 | 39 | 25.6 | 0.31 (0.14, 0.70) | 0.11 | 0.11 |
| Burkina Faso | iLiNS−Zinc (38) | | | | | | | | | | | | |
| Burkina Faso | PROMIS CS (39) | | | | | | | | | | | | |
| Ghana | GHANA (40) | | | | | | | | | | | | |
| Ghana | iLiNS−DYADG (41) | | | | | | | | | | | | |
| Kenya | WASH−B (42) | 143 | 117 | 55.6 | 0.43 (0.30, 0.62) | 0.41 | 0.38 | 154 | 142 | 62.0 | 0.40 (0.28, 0.56) | 0.61 | 0.61 |
| Madagascar | MAHAY (43) | | | | | | | | | | | | |
| Malawi | iLiNS−DYADM (44) | | | | | | | | | | | | |
| Malawi | iLiNS−DOSE (45) | | | | | | | | | | | | |
| Mali | PROMIS CS (46) | | | | | | | | | | | | |
| Zimbabwe | SHINE (HIV−) (47) | | | | | | | | | | | | |
| Zimbabwe | SHINE (HIV+) (48) | | | | | | | | | | | | |
| | | **996** | **544** | | **$I^2$ = 0.13, $Tau^2$ = 0.01** | | | **517** | **309** | | **$I^2$ = 0.00, $Tau^2$ = 0.00** | | |
| **Fixed** | | | | | **0.36 (0.29, 0.46)** | | | | | | **0.36 (0.27, 0.47)** | | |
| **Random** | | | | | **0.36 (0.28, 0.46)** | | | | | | **0.36 (0.27, 0.47)** | | |

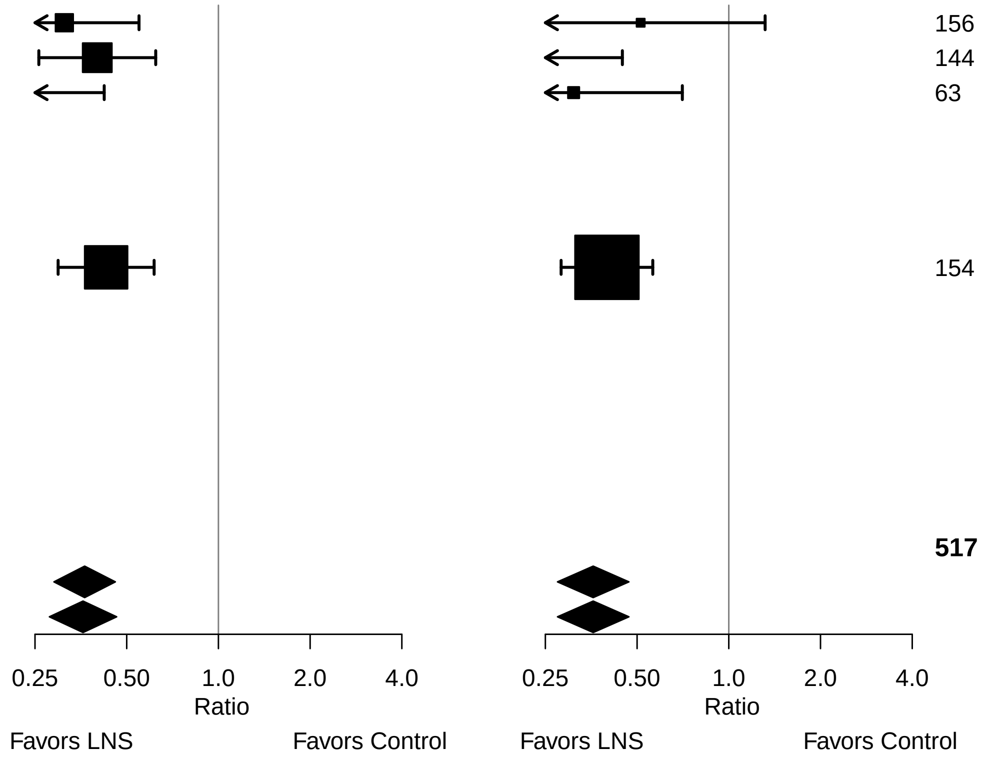

#### Supplemental figure 6G: Iron deficiency (ferritin < 12 µg/L) prevalence ratio

#### 6G4: Stratified by Child sex

**P−for−interaction = 0.771**
**Ratio of PRs = 1.04 (0.80, 1.36)**

| | | Male | | | | | | Female | | | | | |
|---|---|---|---|---|---|---|---|---|---|---|---|---|---|
| | | **LNS** | **Control** | **Control** | **PR** | **Fixed** | **Random** | **LNS** | **Control** | **Control** | **PR** | **Fixed** | **Random** |
| Country | Trial | N | N | Prevalence | (95% CI) | W | W | N | N | Prevalence | (95% CI) | W | W |
| Bangladesh | JiVitA−4 (35) | 235 | 70 | 27.1 | 0.31 (0.17, 0.56) | 0.10 | 0.11 | 220 | 74 | 17.6 | 0.41 (0.19, 0.89) | 0.06 | 0.08 |
| Bangladesh | RDNS (36) | 266 | 131 | 25.2 | 0.36 (0.23, 0.55) | 0.18 | 0.18 | 281 | 141 | 19.1 | 0.32 (0.18, 0.55) | 0.11 | 0.14 |
| Bangladesh | WASH−B (37) | 105 | 85 | 37.6 | 0.25 (0.12, 0.54) | 0.06 | 0.07 | 107 | 93 | 33.3 | 0.22 (0.11, 0.46) | 0.07 | 0.09 |
| Burkina Faso | iLiNS−Zinc (38) | 150 | 49 | 55.6 | 0.62 (0.42, 0.92) | 0.21 | 0.20 | 165 | 47 | 61.5 | 0.61 (0.45, 0.82) | 0.37 | 0.27 |
| Burkina Faso | PROMIS CS (39) | | | | | | | | | | | | |
| Ghana | GHANA (40) | 51 | 37 | 75.7 | 0.41 (0.27, 0.65) | 0.17 | 0.17 | 32 | 45 | 40.0 | 0.55 (0.26, 1.15) | 0.06 | 0.09 |
| Ghana | iLiNS−DYADG (41) | | | | | | | | | | | | |
| Kenya | WASH−B (42) | 146 | 125 | 58.4 | 0.43 (0.30, 0.62) | 0.26 | 0.23 | 152 | 134 | 59.7 | 0.40 (0.28, 0.57) | 0.25 | 0.23 |
| Madagascar | MAHAY (43) | 34 | 22 | 59.1 | 0.25 (0.09, 0.70) | 0.03 | 0.04 | 49 | 29 | 44.8 | 0.50 (0.26, 0.96) | 0.08 | 0.11 |
| Malawi | iLiNS−DYADM (44) | | | | | | | | | | | | |
| Malawi | iLiNS−DOSE (45) | | | | | | | | | | | | |
| Mali | PROMIS CS (46) | | | | | | | | | | | | |
| Zimbabwe | SHINE (HIV−) (47) | | | | | | | | | | | | |
| Zimbabwe | SHINE (HIV+) (48) | | | | | | | | | | | | |
| | | **987** | **519** | | **I² = 0.24, Tau² = 0.02** | | | **1006** | **563** | | **I² = 0.39, Tau² = 0.03** | | |
| **Fixed** | | | | | **0.41 (0.35, 0.50)** | | | | | | **0.45 (0.38, 0.55)** | | |
| **Random** | | | | | **0.41 (0.33, 0.50)** | | | | | | **0.43 (0.34, 0.55)** | | |

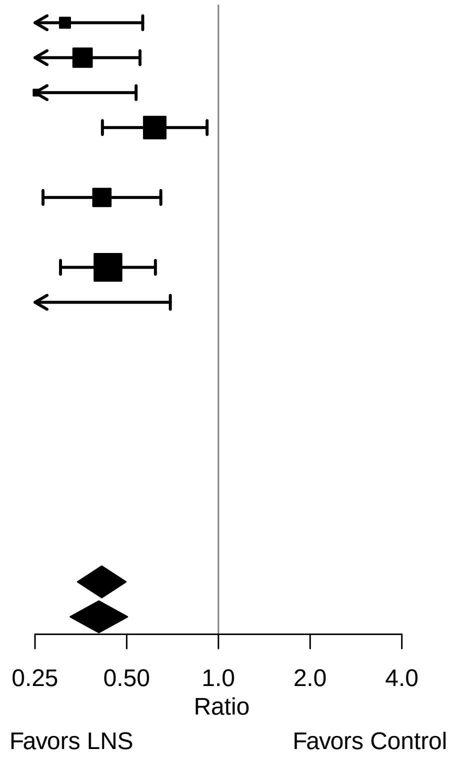

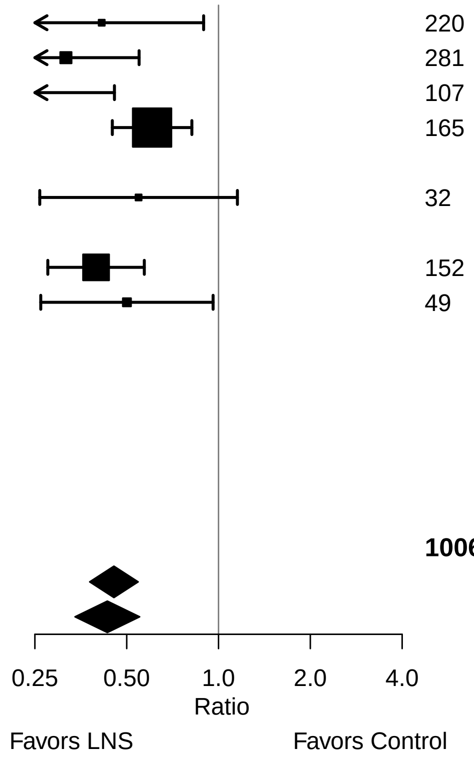

##### Supplemental figure 6G: Iron deficiency (ferritin < 12 µg/L) prevalence ratio

#### 6G5: Stratified by Child birth order

**P−for−interaction = 0.101**
**Ratio of PRs = 1.22 (0.96, 1.56)**

| | | Later born | | | | | | Firstborn | | | | | |
|---|---|---|---|---|---|---|---|---|---|---|---|---|---|
| Country | Trial | LNS N | Control N | Control Prevalence | PR (95% CI) | Fixed W | Random W | LNS N | Control N | Control Prevalence | PR (95% CI) | Fixed W | Random W |
| Bangladesh | JiVitA−4 (35) | 114 | 32 | 25.0 | 0.32 (0.11, 0.92) | 0.02 | 0.04 | 340 | 111 | 21.6 | 0.37 (0.21, 0.65) | 0.11 | 0.17 |
| Bangladesh | RDNS (36) | 340 | 171 | 22.8 | 0.32 (0.21, 0.49) | 0.15 | 0.18 | 210 | 101 | 20.8 | 0.37 (0.20, 0.66) | 0.11 | 0.16 |
| Bangladesh | WASH−B (37) | 124 | 119 | 35.3 | 0.27 (0.14, 0.53) | 0.06 | 0.09 | 86 | 57 | 36.8 | 0.16 (0.07, 0.37) | 0.05 | 0.12 |
| Burkina Faso | iLiNS−Zinc (38) | 249 | 71 | 61.3 | 0.57 (0.43, 0.74) | 0.36 | 0.30 | 66 | 25 | 51.6 | 0.82 (0.62, 1.08) | 0.48 | 0.21 |
| Burkina Faso | PROMIS CS (39) | | | | | | | | | | | | |
| Ghana | GHANA (40) | 54 | 50 | 52.0 | 0.43 (0.24, 0.75) | 0.08 | 0.11 | 29 | 29 | 58.6 | 0.65 (0.37, 1.13) | 0.11 | 0.17 |
| Ghana | iLiNS−DYADG (41) | | | | | | | | | | | | |
| Kenya | WASH−B (42) | 237 | 213 | 58.2 | 0.41 (0.31, 0.55) | 0.32 | 0.28 | 60 | 46 | 63.0 | 0.40 (0.24, 0.66) | 0.14 | 0.17 |
| Madagascar | MAHAY (43) | | | | | | | | | | | | |
| Malawi | iLiNS−DYADM (44) | | | | | | | | | | | | |
| Malawi | iLiNS−DOSE (45) | | | | | | | | | | | | |
| Mali | PROMIS CS (46) | | | | | | | | | | | | |
| Zimbabwe | SHINE (HIV−) (47) | | | | | | | | | | | | |
| Zimbabwe | SHINE (HIV+) (48) | | | | | | | | | | | | |
| | | **1118** | **656** | | **I² = 0.37, Tau² = 0.02** | | | **791** | **369** | | **I² = 0.77, Tau² = 0.21** | | |
| **Fixed** | | | | | **0.43 (0.37, 0.51)** | | | | | | **0.56 (0.46, 0.68)** | | |
| **Random** | | | | | **0.42 (0.34, 0.52)** | | | | | | **0.44 (0.28, 0.68)** | | |

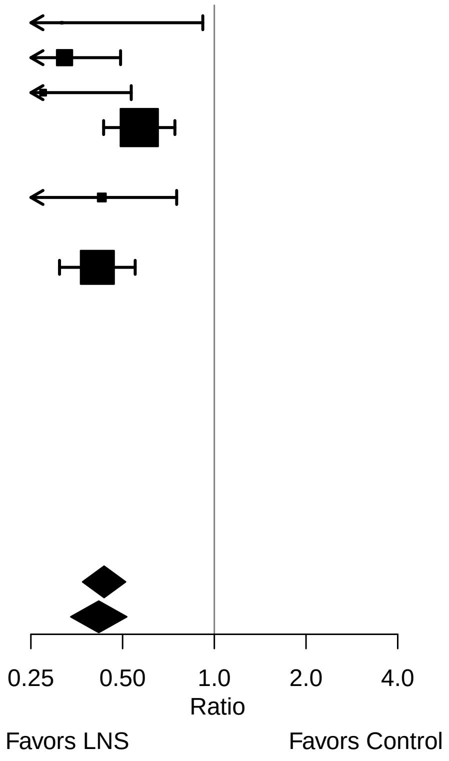

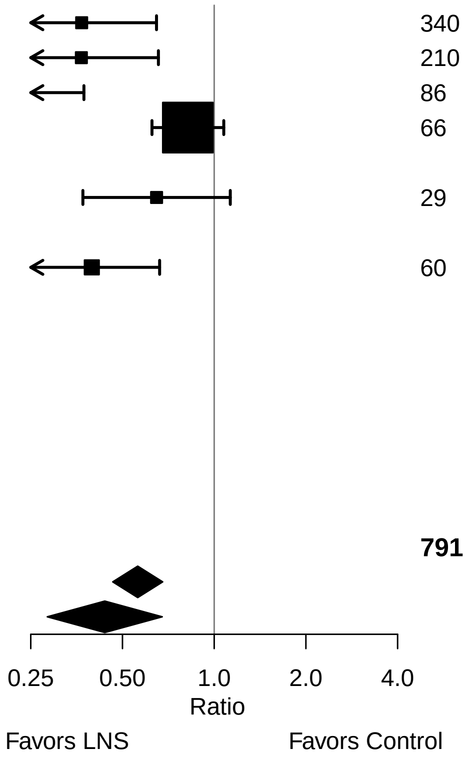

**Supplemental figure 6G: Iron deficiency (ferritin < 12 µg/L) prevalence ratio**

#### 6G6: Stratified by Child baseline acute malnutrition (insufficient comparisons)

**Supplemental figure 6G: Iron deficiency (ferritin < 12 µg/L) prevalence ratio**

#### 6G7: Stratified by Child baseline anemia (insufficient comparisons)

**Supplemental figure 6G: Iron deficiency (ferritin < 12 µg/L) prevalence ratio**

#### 6G8: Stratified by Child high-dose vitamin A supplementation

**P−for−interaction = 0.528**
**Ratio of PRs = 1.16 (0.73, 1.85)**

| | | Received VitA | | | | | | Not treated | | | | | |
|---|---|---|---|---|---|---|---|---|---|---|---|---|---|
| Country | Trial | LNS N | Control N | Control Prevalence | PR (95% CI) | Fixed W | Random W | LNS N | Control N | Control Prevalence | PR (95% CI) | Fixed W | Random W |
| Bangladesh | JiVitA−4 (35) | 140 | 37 | 29.7 | 0.17 (0.06, 0.44) | 0.08 | 0.10 | 133 | 41 | 19.5 | 0.58 (0.27, 1.25) | 0.09 | 0.16 |
| Bangladesh | RDNS (36) | 444 | 223 | 20.2 | 0.37 (0.25, 0.54) | 0.56 | 0.51 | 105 | 49 | 30.6 | 0.25 (0.11, 0.55) | 0.09 | 0.15 |
| Bangladesh | WASH−B (37) | | | | | | | | | | | | |
| Burkina Faso | iLiNS−Zinc (38) | | | | | | | | | | | | |
| Burkina Faso | PROMIS CS (39) | | | | | | | | | | | | |
| Ghana | GHANA (40) | 39 | 53 | 60.4 | 0.47 (0.27, 0.81) | 0.26 | 0.28 | 44 | 26 | 42.3 | 0.64 (0.33, 1.25) | 0.13 | 0.20 |
| Ghana | iLiNS−DYADG (41) | | | | | | | | | | | | |
| Kenya | WASH−B (42) | 26 | 31 | 48.4 | 0.40 (0.16, 0.99) | 0.09 | 0.11 | 223 | 180 | 62.2 | 0.40 (0.30, 0.53) | 0.69 | 0.48 |
| Madagascar | MAHAY (43) | | | | | | | | | | | | |
| Malawi | iLiNS−DYADM (44) | | | | | | | | | | | | |
| Malawi | iLiNS−DOSE (45) | | | | | | | | | | | | |
| Mali | PROMIS CS (46) | | | | | | | | | | | | |
| Zimbabwe | SHINE (HIV−) (47) | | | | | | | | | | | | |
| Zimbabwe | SHINE (HIV+) (48) | | | | | | | | | | | | |
| | | **649** | **344** | | **$I^2$ = 0.08, $Tau^2$ = 0.01** | | | **505** | **296** | | **$I^2$ = 0.27, $Tau^2$ = 0.05** | | |
| **Fixed** | | | | | **0.37 (0.28, 0.49)** | | | | | | **0.42 (0.33, 0.53)** | | |
| **Random** | | | | | **0.37 (0.27, 0.50)** | | | | | | **0.43 (0.30, 0.61)** | | |

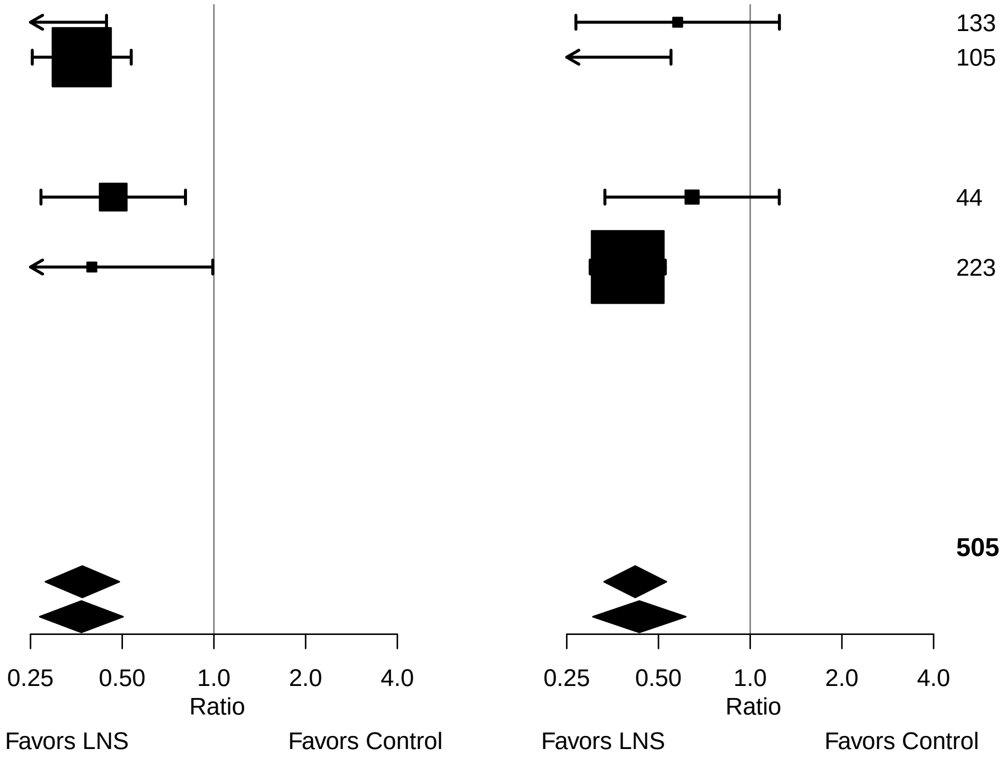

#### Supplemental figure 6G: Iron deficiency (ferritin < 12 μg/L) prevalence ratio

##### 6G9: Stratified by Child inflammation

**P−for−interaction = 0.951**
**Ratio of PRs = 0.99 (0.74, 1.33)**

| | | Not inflamed | | | | | | High AGP or CRP | | | | | |
|---|---|---|---|---|---|---|---|---|---|---|---|---|---|
| Country | Trial | LNS N | Control N | Control Prevalence | PR (95% CI) | Fixed W | Random W | LNS N | Control N | Control Prevalence | PR (95% CI) | Fixed W | Random W |
| Bangladesh | JiVitA−4 (35) | 185 | 46 | 26.1 | 0.27 (0.11, 0.66) | 0.05 | 0.08 | 270 | 98 | 20.4 | 0.42 (0.23, 0.75) | 0.11 | 0.15 |
| Bangladesh | RDNS (36) | 370 | 168 | 22.6 | 0.29 (0.18, 0.46) | 0.19 | 0.22 | 180 | 104 | 21.2 | 0.45 (0.26, 0.77) | 0.12 | 0.17 |
| Bangladesh | WASH−B (37) | | | | | | | | | | | | |
| Burkina Faso | iLiNS−Zinc (38) | 124 | 31 | 55.9 | 0.55 (0.35, 0.86) | 0.22 | 0.23 | 191 | 65 | 59.7 | 0.66 (0.50, 0.87) | 0.48 | 0.35 |
| Burkina Faso | PROMIS CS (39) | | | | | | | | | | | | |
| Ghana | GHANA (40) | | | | | | | | | | | | |
| Ghana | iLiNS−DYADG (41) | | | | | | | | | | | | |
| Kenya | WASH−B (42) | 157 | 135 | 57.8 | 0.50 (0.37, 0.67) | 0.46 | 0.35 | 141 | 124 | 60.5 | 0.33 (0.22, 0.49) | 0.24 | 0.26 |
| Madagascar | MAHAY (43) | 44 | 32 | 53.1 | 0.39 (0.19, 0.78) | 0.09 | 0.12 | 39 | 19 | 47.4 | 0.38 (0.15, 0.94) | 0.05 | 0.08 |
| Malawi | iLiNS−DYADM (44) | | | | | | | | | | | | |
| Malawi | iLiNS−DOSE (45) | | | | | | | | | | | | |
| Mali | PROMIS CS (46) | | | | | | | | | | | | |
| Zimbabwe | SHINE (HIV−) (47) | | | | | | | | | | | | |
| Zimbabwe | SHINE (HIV+) (48) | | | | | | | | | | | | |
| | | **880** | **412** | | **I² = 0.33, Tau² = 0.03** | | | **821** | **410** | | **I² = 0.55, Tau² = 0.04** | | |
| **Fixed** | | | | | **0.43 (0.35, 0.53)** | | | | | | **0.49 (0.40, 0.60)** | | |
| **Random** | | | | | **0.42 (0.32, 0.55)** | | | | | | **0.46 (0.35, 0.60)** | | |

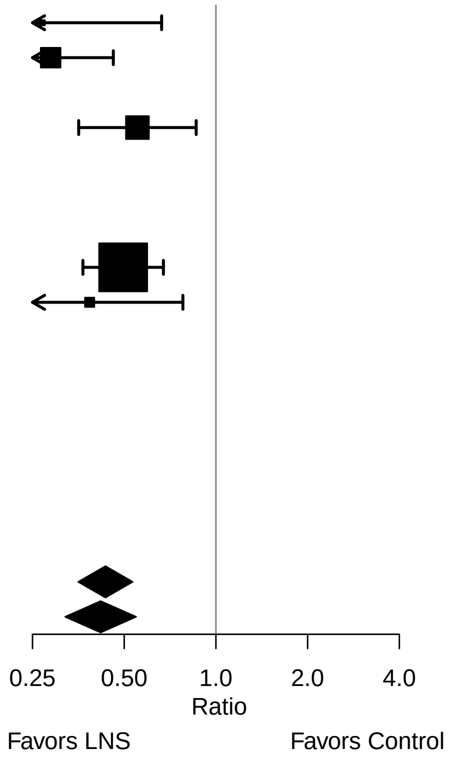

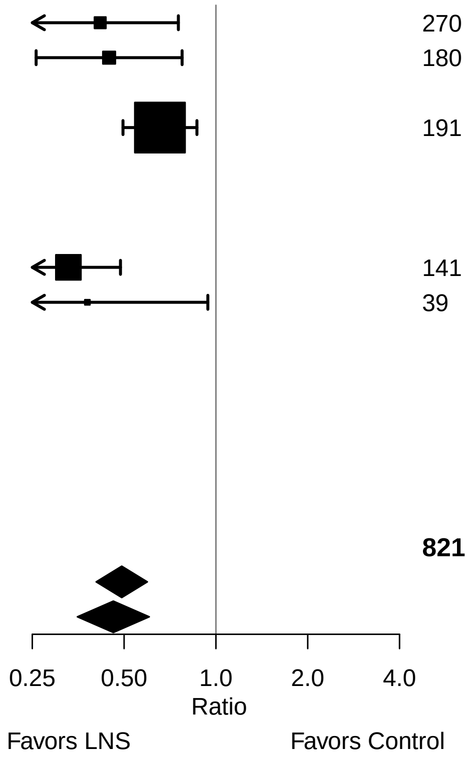

### Supplemental figure 6H: Iron deficiency (ferritin < 12 µg/L) prevalence difference

#### 6H1: Stratified by Maternal BMI

**P−for−interaction = 0.473**
**Difference in PDs = 0.03 (−0.05, 0.11)**

| | | **At least 20 kg/m²** | | | | | | **Less than 20 kg/m²** | | | | | |
|---|---|---|---|---|---|---|---|---|---|---|---|---|---|
| Country | Trial | **LNS** N | **Control** N | **Control** Prevalence | **PD** (95% CI) | **Fixed** W | **Random** W | **LNS** N | **Control** N | **Control** Prevalence | **PD** (95% CI) | **Fixed** W | **Random** W |
| Bangladesh | JiVitA−4 (35) | | | | | | | | | | | | |
| Bangladesh | RDNS (36) | 228 | 102 | 22.5 | −0.17 (−0.26, −0.09) | 0.41 | 0.30 | 300 | 160 | 21.2 | −0.12 (−0.19, −0.05) | 0.51 | 0.33 |
| Bangladesh | WASH−B (37) | 93 | 97 | 36.1 | −0.25 (−0.39, −0.12) | 0.16 | 0.22 | 119 | 81 | 34.6 | −0.28 (−0.38, −0.17) | 0.22 | 0.26 |
| Burkina Faso | iLiNS−Zinc (38) | 209 | 55 | 54.1 | −0.17 (−0.32, −0.03) | 0.14 | 0.20 | 106 | 41 | 64.4 | −0.31 (−0.43, −0.19) | 0.17 | 0.23 |
| Burkina Faso | PROMIS CS (39) | | | | | | | | | | | | |
| Ghana | GHANA (40) | | | | | | | | | | | | |
| Ghana | iLiNS−DYADG (41) | | | | | | | | | | | | |
| Kenya | WASH−B (42) | 223 | 200 | 61.5 | −0.36 (−0.46, −0.26) | 0.29 | 0.28 | 64 | 51 | 45.1 | −0.22 (−0.38, −0.05) | 0.09 | 0.17 |
| Madagascar | MAHAY (43) | | | | | | | | | | | | |
| Malawi | iLiNS−DYADM (44) | | | | | | | | | | | | |
| Malawi | iLiNS−DOSE (45) | | | | | | | | | | | | |
| Mali | PROMIS CS (46) | | | | | | | | | | | | |
| Zimbabwe | SHINE (HIV−) (47) | | | | | | | | | | | | |
| Zimbabwe | SHINE (HIV+) (48) | | | | | | | | | | | | |
| | | **753** | **454** | | **I² = 0.67, Tau² = 0.01** | | | **589** | **333** | | **I² = 0.72, Tau² = 0.01** | | |
| **Fixed** | | | | | **−0.24 (−0.30, −0.19)** | | | | | | **−0.20 (−0.25, −0.15)** | | |
| **Random** | | | | | **−0.24 (−0.33, −0.15)** | | | | | | **−0.22 (−0.31, −0.13)** | | |

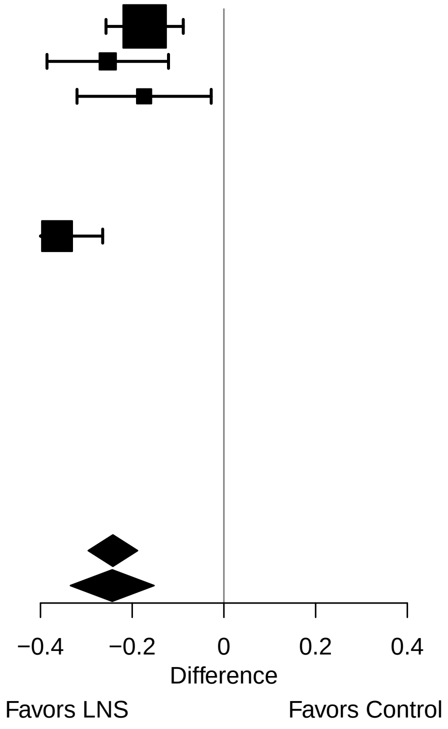

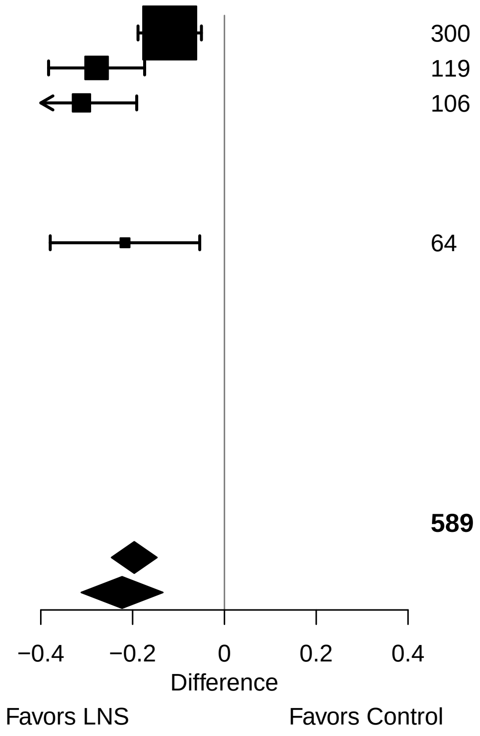

#### Supplemental figure 6H: Iron deficiency (ferritin < 12 µg/L) prevalence difference

#### 6H2: Stratified by Maternal age

**P−for−interaction = 0.779**
**Difference in PDs = 0.01 (−0.06, 0.07)**

| Country | Trial | At least 25 y: LNS N | Control N | Control Prevalence | PD (95% CI) | Fixed W | Random W | Less than 25 y: LNS N | Control N | Control Prevalence | PD (95% CI) | Fixed W | Random W |
|---|---|---|---|---|---|---|---|---|---|---|---|---|---|
| Bangladesh | JiVitA−4 (35) | 202 | 70 | 22.9 | −0.17 (−0.29, −0.05) | 0.14 | 0.15 | 250 | 73 | 21.9 | −0.12 (−0.24, 0.00) | 0.10 | 0.16 |
| Bangladesh | RDNS (36) | 152 | 81 | 22.2 | −0.16 (−0.26, −0.07) | 0.24 | 0.18 | 398 | 191 | 22.0 | −0.14 (−0.19, −0.09) | 0.60 | 0.26 |
| Bangladesh | WASH−B (37) | 79 | 80 | 27.5 | −0.16 (−0.29, −0.03) | 0.13 | 0.14 | 132 | 98 | 41.8 | −0.35 (−0.46, −0.24) | 0.12 | 0.18 |
| Burkina Faso | iLiNS−Zinc (38) | 178 | 50 | 59.3 | −0.27 (−0.38, −0.15) | 0.16 | 0.16 | 137 | 46 | 57.7 | −0.17 (−0.31, −0.02) | 0.07 | 0.13 |
| Burkina Faso | PROMIS CS (39) | | | | | | | | | | | | |
| Ghana | GHANA (40) | 61 | 55 | 52.7 | −0.28 (−0.45, −0.11) | 0.07 | 0.11 | 22 | 21 | 61.9 | −0.26 (−0.55, 0.04) | 0.02 | 0.05 |
| Ghana | iLiNS−DYADG (41) | | | | | | | | | | | | |
| Kenya | WASH−B (42) | 178 | 149 | 60.4 | −0.36 (−0.46, −0.26) | 0.21 | 0.18 | 116 | 109 | 56.9 | −0.32 (−0.46, −0.18) | 0.07 | 0.14 |
| Madagascar | MAHAY (43) | 47 | 25 | 60.0 | −0.41 (−0.62, −0.20) | 0.05 | 0.08 | 36 | 26 | 42.3 | −0.23 (−0.45, −0.01) | 0.03 | 0.08 |
| Malawi | iLiNS−DYADM (44) | | | | | | | | | | | | |
| Malawi | iLiNS−DOSE (45) | | | | | | | | | | | | |
| Mali | PROMIS CS (46) | | | | | | | | | | | | |
| Zimbabwe | SHINE (HIV−) (47) | | | | | | | | | | | | |
| Zimbabwe | SHINE (HIV+) (48) | | | | | | | | | | | | |
| | | **897** | **510** | | **I² = 0.57, Tau² = 0.00** | | | **1091** | **564** | | **I² = 0.67, Tau² = 0.00** | | |
| **Fixed** | | | | | **−0.24 (−0.29, −0.20)** | | | | | | **−0.18 (−0.22, −0.15)** | | |
| **Random** | | | | | **−0.25 (−0.32, −0.18)** | | | | | | **−0.22 (−0.29, −0.14)** | | |

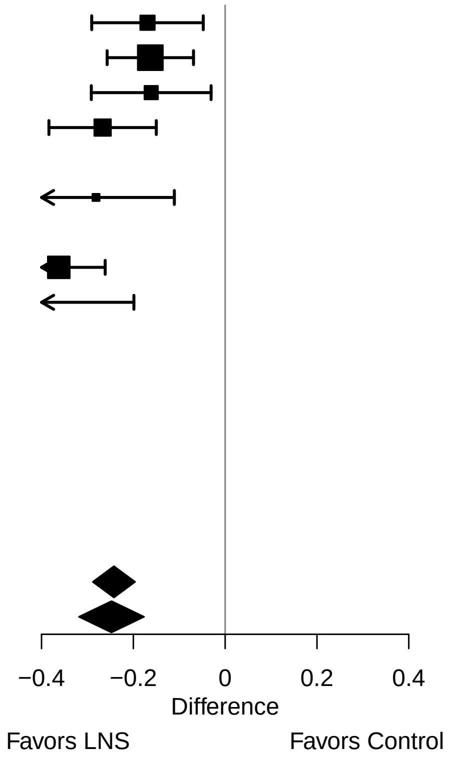

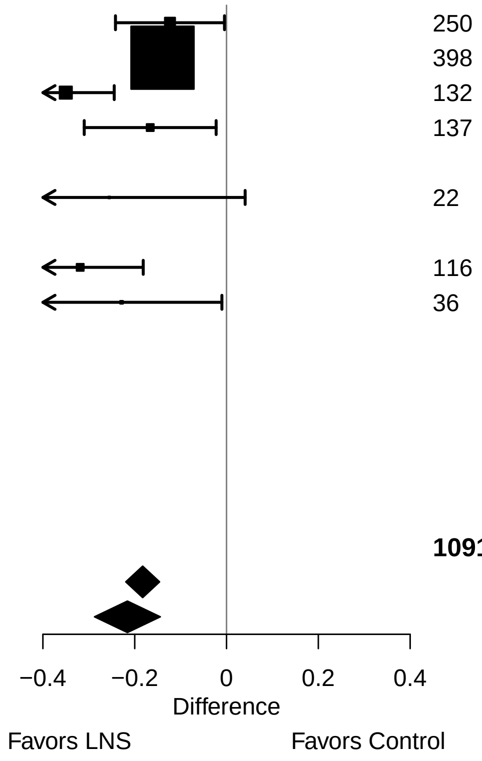

**Supplemental figure 6H: Iron deficiency (ferritin < 12 μg/L) prevalence difference**

#### 6H3: Stratified by Maternal education

**P−for−interaction = 0.963**
**Difference in PDs = 0.00 (−0.08, 0.07)**

| | | Primary or greater | | | | | | Incomplete or no formal | | | | | |
|---|---|---|---|---|---|---|---|---|---|---|---|---|---|
| | | **LNS** | **Control** | **Control** | **PD** | **Fixed** | **Random** | **LNS** | **Control** | **Control** | **PD** | **Fixed** | **Random** |
| Country | Trial | N | N | Prevalence | (95% CI) | W | W | N | N | Prevalence | (95% CI) | W | W |
| Bangladesh | JiVitA−4 (35) | 298 | 93 | 26.9 | −0.18 (−0.30, −0.07) | 0.12 | 0.23 | 156 | 51 | 13.7 | −0.07 (−0.17, 0.04) | 0.29 | 0.26 |
| Bangladesh | RDNS (36) | 406 | 195 | 18.5 | −0.11 (−0.16, −0.06) | 0.63 | 0.31 | 144 | 77 | 31.2 | −0.24 (−0.34, −0.14) | 0.32 | 0.27 |
| Bangladesh | WASH−B (37) | 149 | 139 | 38.1 | −0.29 (−0.40, −0.19) | 0.15 | 0.25 | 63 | 39 | 25.6 | −0.18 (−0.32, −0.03) | 0.15 | 0.22 |
| Burkina Faso | iLiNS−Zinc (38) | | | | | | | | | | | | |
| Burkina Faso | PROMIS CS (39) | | | | | | | | | | | | |
| Ghana | GHANA (40) | | | | | | | | | | | | |
| Ghana | iLiNS−DYADG (41) | | | | | | | | | | | | |
| Kenya | WASH−B (42) | 143 | 117 | 55.6 | −0.32 (−0.45, −0.19) | 0.10 | 0.21 | 154 | 142 | 62.0 | −0.37 (−0.49, −0.26) | 0.24 | 0.25 |
| Madagascar | MAHAY (43) | | | | | | | | | | | | |
| Malawi | iLiNS−DYADM (44) | | | | | | | | | | | | |
| Malawi | iLiNS−DOSE (45) | | | | | | | | | | | | |
| Mali | PROMIS CS (46) | | | | | | | | | | | | |
| Zimbabwe | SHINE (HIV−) (47) | | | | | | | | | | | | |
| Zimbabwe | SHINE (HIV+) (48) | | | | | | | | | | | | |
| | | **996** | **544** | | **I² = 0.81, Tau² = 0.01** | | | **517** | **309** | | **I² = 0.80, Tau² = 0.01** | | |
| **Fixed** | | | | | **−0.17 (−0.21, −0.13)** | | | | | | **−0.21 (−0.27, −0.15)** | | |
| **Random** | | | | | **−0.22 (−0.31, −0.12)** | | | | | | **−0.21 (−0.34, −0.09)** | | |

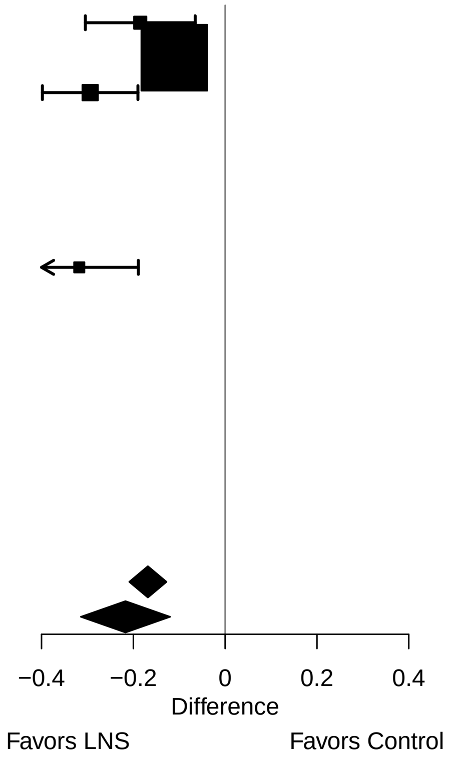

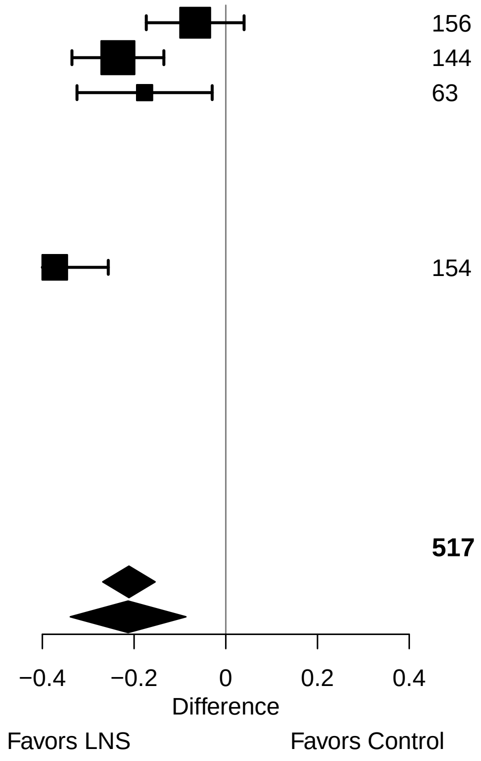

#### Supplemental figure 6H: Iron deficiency (ferritin < 12 µg/L) prevalence difference

#### 6H4: Stratified by Child sex

**P−for−interaction = 0.179**
**Difference in PDs = 0.04 (−0.02, 0.11)**

| Country | Trial | Male LNS N | Male Control N | Male Control Prevalence | Male PD (95% CI) | Male Fixed W | Male Random W | Female LNS N | Female Control N | Female Control Prevalence | Female PD (95% CI) | Female Fixed W | Female Random W |
|---|---|---|---|---|---|---|---|---|---|---|---|---|---|
| Bangladesh | JiVitA−4 (35) | 235 | 70 | 27.1 | −0.19 (−0.31, −0.06) | 0.13 | 0.16 | 220 | 74 | 17.6 | −0.10 (−0.21, 0.00) | 0.17 | 0.17 |
| Bangladesh | RDNS (36) | 266 | 131 | 25.2 | −0.16 (−0.23, −0.10) | 0.46 | 0.21 | 281 | 141 | 19.1 | −0.13 (−0.21, −0.05) | 0.33 | 0.22 |
| Bangladesh | WASH−B (37) | 105 | 85 | 37.6 | −0.28 (−0.43, −0.14) | 0.10 | 0.14 | 107 | 93 | 33.3 | −0.26 (−0.35, −0.17) | 0.23 | 0.19 |
| Burkina Faso | iLiNS−Zinc (38) | 150 | 49 | 55.6 | −0.21 (−0.39, −0.03) | 0.06 | 0.12 | 165 | 47 | 61.5 | −0.25 (−0.41, −0.09) | 0.08 | 0.11 |
| Burkina Faso | PROMIS CS (39) | | | | | | | | | | | | |
| Ghana | GHANA (40) | 51 | 37 | 75.7 | −0.44 (−0.64, −0.25) | 0.05 | 0.11 | 32 | 45 | 40.0 | −0.18 (−0.39, 0.03) | 0.04 | 0.08 |
| Ghana | iLiNS−DYADG (41) | | | | | | | | | | | | |
| Kenya | WASH−B (42) | 146 | 125 | 58.4 | −0.33 (−0.44, −0.22) | 0.15 | 0.17 | 152 | 134 | 59.7 | −0.36 (−0.49, −0.23) | 0.11 | 0.14 |
| Madagascar | MAHAY (43) | 34 | 22 | 59.1 | −0.44 (−0.65, −0.23) | 0.04 | 0.09 | 49 | 29 | 44.8 | −0.22 (−0.43, −0.02) | 0.05 | 0.08 |
| Malawi | iLiNS−DYADM (44) | | | | | | | | | | | | |
| Malawi | iLiNS−DOSE (45) | | | | | | | | | | | | |
| Mali | PROMIS CS (46) | | | | | | | | | | | | |
| Zimbabwe | SHINE (HIV−) (47) | | | | | | | | | | | | |
| Zimbabwe | SHINE (HIV+) (48) | | | | | | | | | | | | |
| | | **987** | **519** | | **$I^2$ = 0.64, $Tau^2$ = 0.01** | | | **1006** | **563** | | **$I^2$ = 0.57, $Tau^2$ = 0.00** | | |
| **Fixed** | | | | | **−0.23 (−0.28, −0.19)** | | | | | | **−0.20 (−0.24, −0.15)** | | |
| **Random** | | | | | **−0.27 (−0.36, −0.19)** | | | | | | **−0.21 (−0.28, −0.14)** | | |

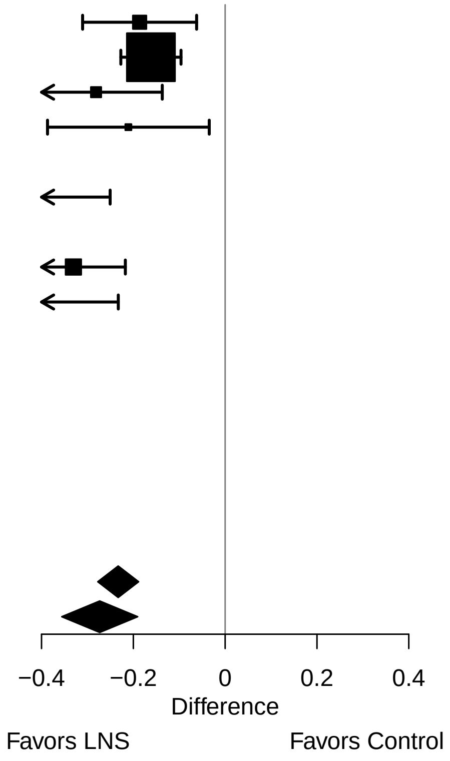

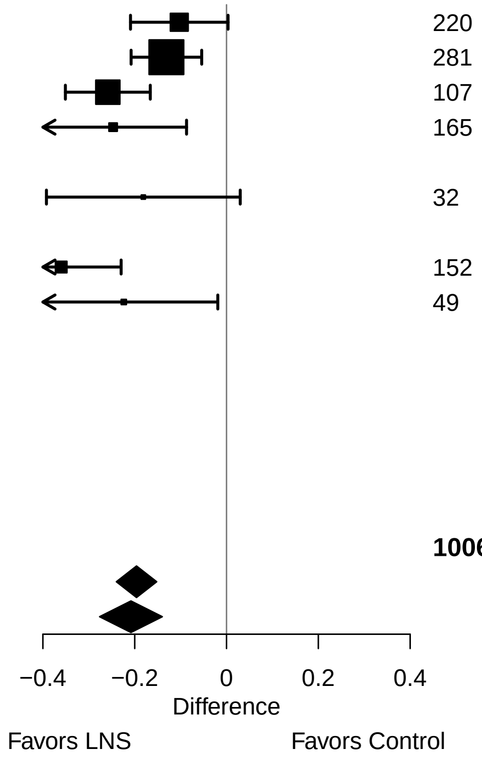

#### Supplemental figure 6H: Iron deficiency (ferritin < 12 µg/L) prevalence difference

##### 6H5: Stratified by Child birth order

**P−for−interaction = 0.638**
**Difference in PDs = 0.02 (−0.05, 0.09)**

| | | Later born | | | | | | Firstborn | | | | | |
|---|---|---|---|---|---|---|---|---|---|---|---|---|---|
| Country | Trial | LNS N | Control N | Control Prevalence | PD (95% CI) | Fixed W | Random W | LNS N | Control N | Control Prevalence | PD (95% CI) | Fixed W | Random W |
| Bangladesh | JiVitA−4 (35) | 114 | 32 | 25.0 | −0.17 (−0.39, 0.05) | 0.03 | 0.07 | 340 | 111 | 21.6 | −0.14 (−0.23, −0.04) | 0.24 | 0.21 |
| Bangladesh | RDNS (36) | 340 | 171 | 22.8 | −0.15 (−0.21, −0.10) | 0.54 | 0.31 | 210 | 101 | 20.8 | −0.13 (−0.20, −0.06) | 0.40 | 0.24 |
| Bangladesh | WASH−B (37) | 124 | 119 | 35.3 | −0.26 (−0.37, −0.14) | 0.11 | 0.17 | 86 | 57 | 36.8 | −0.31 (−0.45, −0.17) | 0.11 | 0.16 |
| Burkina Faso | iLiNS−Zinc (38) | 249 | 71 | 61.3 | −0.27 (−0.40, −0.13) | 0.08 | 0.14 | 66 | 25 | 51.6 | −0.10 (−0.21, 0.01) | 0.17 | 0.19 |
| Burkina Faso | PROMIS CS (39) | | | | | | | | | | | | |
| Ghana | GHANA (40) | 54 | 50 | 52.0 | −0.30 (−0.48, −0.12) | 0.05 | 0.10 | 29 | 29 | 58.6 | −0.21 (−0.46, 0.05) | 0.03 | 0.08 |
| Ghana | iLiNS−DYADG (41) | | | | | | | | | | | | |
| Kenya | WASH−B (42) | 237 | 213 | 58.2 | −0.34 (−0.43, −0.25) | 0.18 | 0.22 | 60 | 46 | 63.0 | −0.38 (−0.58, −0.18) | 0.05 | 0.11 |
| Madagascar | MAHAY (43) | | | | | | | | | | | | |
| Malawi | iLiNS−DYADM (44) | | | | | | | | | | | | |
| Malawi | iLiNS−DOSE (45) | | | | | | | | | | | | |
| Mali | PROMIS CS (46) | | | | | | | | | | | | |
| Zimbabwe | SHINE (HIV−) (47) | | | | | | | | | | | | |
| Zimbabwe | SHINE (HIV+) (48) | | | | | | | | | | | | |
| | | **1118** | **656** | | **I² = 0.66, Tau² = 0.00** | | | **791** | **369** | | **I² = 0.55, Tau² = 0.01** | | |
| **Fixed** | | | | | **−0.22 (−0.26, −0.18)** | | | | | | **−0.16 (−0.21, −0.12)** | | |
| **Random** | | | | | **−0.24 (−0.31, −0.18)** | | | | | | **−0.19 (−0.27, −0.10)** | | |

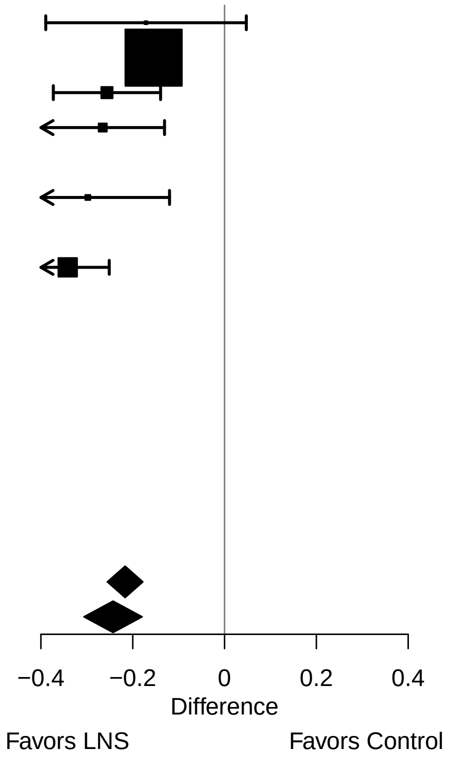

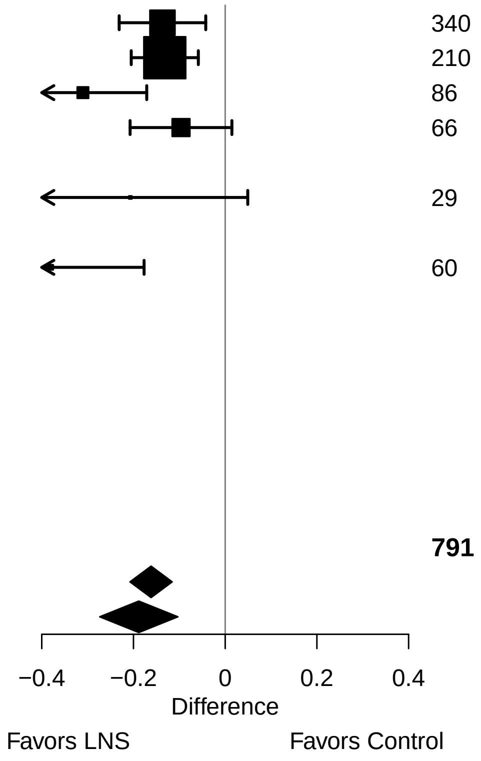

**Supplemental figure 6H: Iron deficiency (ferritin < 12 µg/L) prevalence difference**

#### 6H6: Stratified by Child baseline acute malnutrition (insufficient comparisons)

**Supplemental figure 6H: Iron deficiency (ferritin < 12 µg/L) prevalence difference**

#### 6H7: Stratified by Child baseline anemia (insufficient comparisons)

#### Supplemental figure 6H: Iron deficiency (ferritin < 12 µg/L) prevalence difference

#### 6H8: Stratified by Child high-dose vitamin A supplementation

**P−for−interaction = 0.999**
**Difference in PDs = 0.00 (−0.10, 0.10)**

| Country | Trial | Received VitA: LNS N | Control N | Control Prevalence | PD (95% CI) | Fixed W | Random W | Not treated: LNS N | Control N | Control Prevalence | PD (95% CI) | Fixed W | Random W |
|---|---|---|---|---|---|---|---|---|---|---|---|---|---|
| Bangladesh | JiVitA−4 (35) | 140 | 37 | 29.7 | −0.25 (−0.42, −0.07) | 0.05 | 0.19 | 133 | 41 | 19.5 | −0.08 (−0.22, 0.05) | 0.22 | 0.26 |
| Bangladesh | RDNS (36) | 444 | 223 | 20.2 | −0.13 (−0.17, −0.08) | 0.88 | 0.55 | 105 | 49 | 30.6 | −0.23 (−0.35, −0.11) | 0.27 | 0.27 |
| Bangladesh | WASH−B (37) | | | | | | | | | | | | |
| Burkina Faso | iLiNS−Zinc (38) | | | | | | | | | | | | |
| Burkina Faso | PROMIS CS (39) | | | | | | | | | | | | |
| Ghana | GHANA (40) | 39 | 53 | 60.4 | −0.32 (−0.52, −0.12) | 0.04 | 0.16 | 44 | 26 | 42.3 | −0.15 (−0.38, 0.08) | 0.07 | 0.17 |
| Ghana | iLiNS−DYADG (41) | | | | | | | | | | | | |
| Kenya | WASH−B (42) | 26 | 31 | 48.4 | −0.29 (−0.55, −0.03) | 0.02 | 0.10 | 223 | 180 | 62.2 | −0.38 (−0.47, −0.28) | 0.44 | 0.30 |
| Madagascar | MAHAY (43) | | | | | | | | | | | | |
| Malawi | iLiNS−DYADM (44) | | | | | | | | | | | | |
| Malawi | iLiNS−DOSE (45) | | | | | | | | | | | | |
| Mali | PROMIS CS (46) | | | | | | | | | | | | |
| Zimbabwe | SHINE (HIV−) (47) | | | | | | | | | | | | |
| Zimbabwe | SHINE (HIV+) (48) | | | | | | | | | | | | |
| | | **649** | **344** | | **I² = 0.52, Tau² = 0.00** | | | **505** | **296** | | **I² = 0.78, Tau² = 0.01** | | |
| **Fixed** | | | | | **−0.15 (−0.19, −0.11)** | | | | | | **−0.26 (−0.32, −0.19)** | | |
| **Random** | | | | | **−0.20 (−0.29, −0.11)** | | | | | | **−0.22 (−0.35, −0.09)** | | |

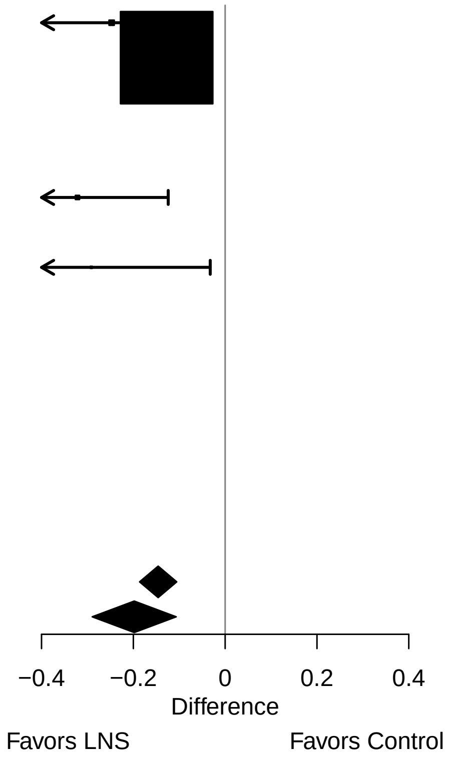

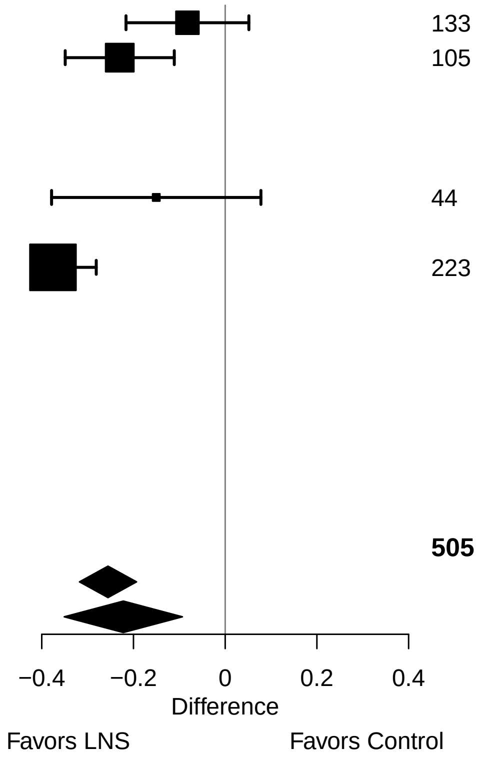

**Supplemental figure 6H: Iron deficiency (ferritin < 12 µg/L) prevalence difference**

#### 6H9: Stratified by Child inflammation

**P−for−interaction = 0.698**
**Difference in PDs = 0.01 (−0.06, 0.09)**

| Country | Trial | Not inflamed: LNS N | Control N | Control Prevalence | PD (95% CI) | Fixed W | Random W | High AGP or CRP: LNS N | Control N | Control Prevalence | PD (95% CI) | Fixed W | Random W |
|---|---|---|---|---|---|---|---|---|---|---|---|---|---|
| Bangladesh | JiVitA−4 (35) | 185 | 46 | 26.1 | −0.19 (−0.38, 0.00) | 0.05 | 0.09 | 270 | 98 | 20.4 | −0.12 (−0.22, −0.02) | 0.24 | 0.22 |
| Bangladesh | RDNS (36) | 370 | 168 | 22.6 | −0.16 (−0.21, −0.11) | 0.72 | 0.51 | 180 | 104 | 21.2 | −0.12 (−0.20, −0.04) | 0.38 | 0.24 |
| Bangladesh | WASH−B (37) | | | | | | | | | | | | |
| Burkina Faso | iLiNS−Zinc (38) | 124 | 31 | 55.9 | −0.25 (−0.43, −0.08) | 0.06 | 0.10 | 191 | 65 | 59.7 | −0.21 (−0.35, −0.06) | 0.12 | 0.19 |
| Burkina Faso | PROMIS CS (39) | | | | | | | | | | | | |
| Ghana | GHANA (40) | | | | | | | | | | | | |
| Ghana | iLiNS−DYADG (41) | | | | | | | | | | | | |
| Kenya | WASH−B (42) | 157 | 135 | 57.8 | −0.29 (−0.41, −0.18) | 0.13 | 0.21 | 141 | 124 | 60.5 | −0.41 (−0.51, −0.30) | 0.22 | 0.22 |
| Madagascar | MAHAY (43) | 44 | 32 | 53.1 | −0.33 (−0.53, −0.13) | 0.04 | 0.09 | 39 | 19 | 47.4 | −0.29 (−0.53, −0.05) | 0.04 | 0.12 |
| Malawi | iLiNS−DYADM (44) | | | | | | | | | | | | |
| Malawi | iLiNS−DOSE (45) | | | | | | | | | | | | |
| Mali | PROMIS CS (46) | | | | | | | | | | | | |
| Zimbabwe | SHINE (HIV−) (47) | | | | | | | | | | | | |
| Zimbabwe | SHINE (HIV+) (48) | | | | | | | | | | | | |
| | | **880** | **412** | | **I² = 0.39, Tau² = 0.00** | | | **821** | **410** | | **I² = 0.82, Tau² = 0.01** | | |
| **Fixed** | | | | | **−0.19 (−0.23, −0.15)** | | | | | | **−0.20 (−0.25, −0.15)** | | |
| **Random** | | | | | **−0.21 (−0.28, −0.15)** | | | | | | **−0.22 (−0.33, −0.11)** | | |

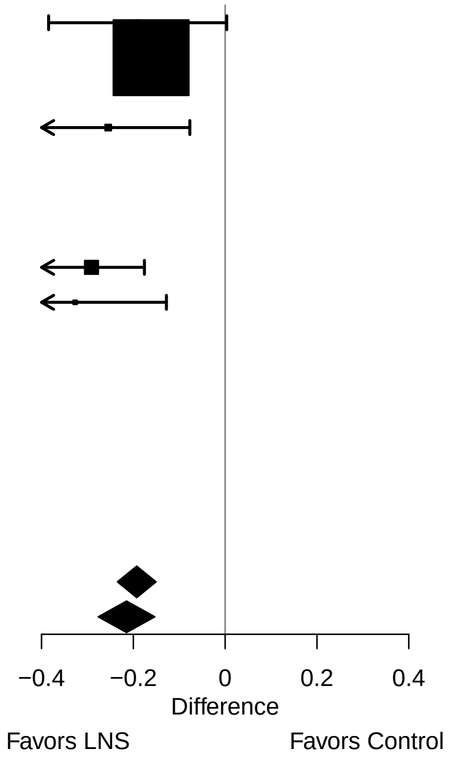

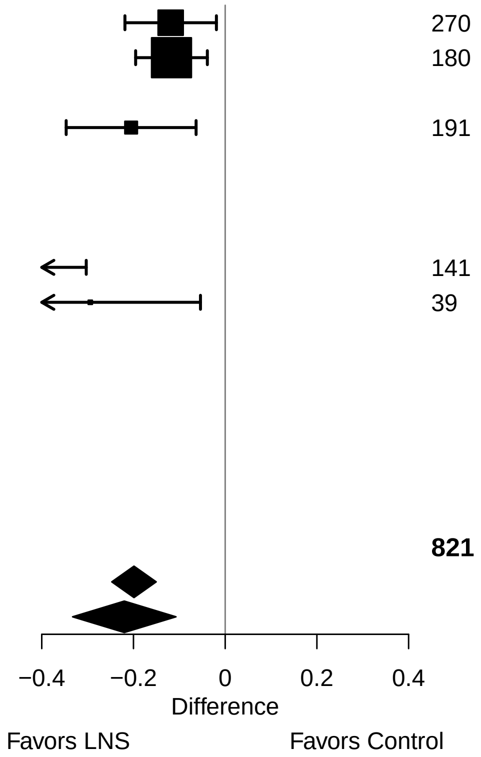

### Supplemental figure 6I: Iron deficiency anemia prevalence ratio

#### 6I1: Stratified by Maternal BMI

**P−for−interaction = 0.521**
**Ratio of PRs = 1.16 (0.74, 1.81)**

| | | At least 20 kg/m² | | | | | | Less than 20 kg/m² | | | | | |
|---|---|---|---|---|---|---|---|---|---|---|---|---|---|
| Country | Trial | LNS N | Control N | Control Prevalence | PR (95% CI) | Fixed W | Random W | LNS N | Control N | Control Prevalence | PR (95% CI) | Fixed W | Random W |
| Bangladesh | JiVitA−4 (35) | | | | | | | | | | | | |
| Bangladesh | RDNS (36) | 227 | 102 | 16.7 | 0.19 (0.08, 0.45) | 0.11 | 0.24 | 300 | 160 | 13.1 | 0.33 (0.16, 0.66) | 0.15 | 0.15 |
| Bangladesh | WASH−B (37) | | | | | | | | | | | | |
| Burkina Faso | iLiNS−Zinc (38) | 215 | 59 | 52.5 | 0.53 (0.36, 0.78) | 0.60 | 0.41 | 112 | 42 | 57.1 | 0.47 (0.35, 0.63) | 0.78 | 0.78 |
| Burkina Faso | PROMIS CS (39) | | | | | | | | | | | | |
| Ghana | GHANA (40) | | | | | | | | | | | | |
| Ghana | iLiNS−DYADG (41) | | | | | | | | | | | | |
| Kenya | WASH−B (42) | 223 | 199 | 36.7 | 0.26 (0.15, 0.45) | 0.29 | 0.35 | 64 | 51 | 13.7 | 0.68 (0.25, 1.89) | 0.07 | 0.07 |
| Madagascar | MAHAY (43) | | | | | | | | | | | | |
| Malawi | iLiNS−DYADM (44) | | | | | | | | | | | | |
| Malawi | iLiNS−DOSE (45) | | | | | | | | | | | | |
| Mali | PROMIS CS (46) | | | | | | | | | | | | |
| Zimbabwe | SHINE (HIV−) (47) | | | | | | | | | | | | |
| Zimbabwe | SHINE (HIV+) (48) | | | | | | | | | | | | |
| | | **665** | **360** | | **I² = 0.72, Tau² = 0.20** | | | **476** | **253** | | **I² = 0.00, Tau² = 0.00** | | |
| **Fixed** | | | | | **0.38 (0.28, 0.52)** | | | | | | **0.46 (0.35, 0.60)** | | |
| **Random** | | | | | **0.32 (0.17, 0.59)** | | | | | | **0.46 (0.35, 0.60)** | | |

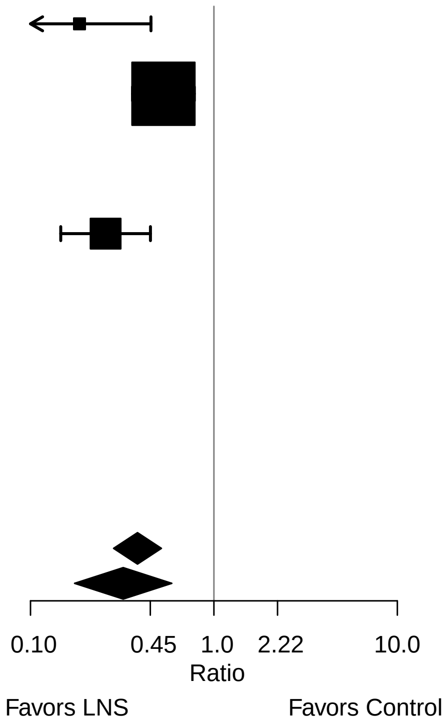

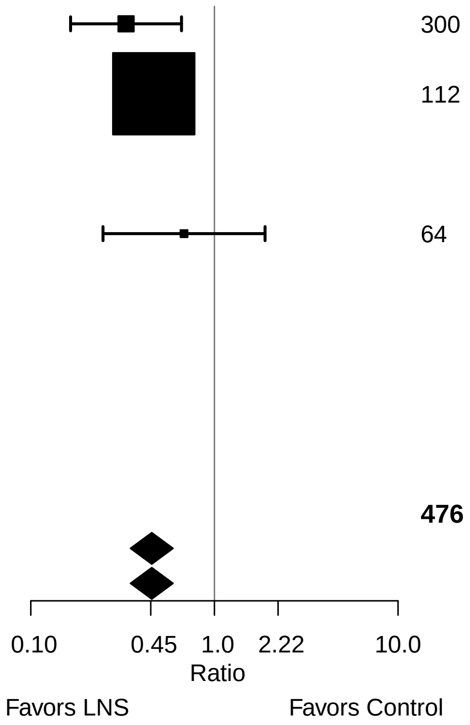

**Supplemental figure 6I: Iron deficiency anemia prevalence ratio**

#### 6I2: Stratified by Maternal age (insufficient comparisons)

**Supplemental figure 6I: Iron deficiency anemia prevalence ratio**

#### 6I3: Stratified by Maternal education (insufficient comparisons)

#### Supplemental figure 6I: Iron deficiency anemia prevalence ratio

##### 6I4: Stratified by Child sex

**P−for−interaction = 0.208**
**Ratio of PRs = 0.78 (0.53, 1.15)**

| Country | Trial | Male LNS N | Male Control N | Male Control Prevalence | Male PR (95% CI) | Male Fixed W | Male Random W | Female LNS N | Female Control N | Female Control Prevalence | Female PR (95% CI) | Female Fixed W | Female Random W |
|---|---|---|---|---|---|---|---|---|---|---|---|---|---|
| Bangladesh | JiVitA−4 (35) | | | | | | | | | | | | |
| Bangladesh | RDNS (36) | 266 | 131 | 16.0 | 0.26 (0.12, 0.57) | 0.13 | 0.23 | 281 | 141 | 13.5 | 0.24 (0.12, 0.49) | 0.15 | 0.24 |
| Bangladesh | WASH−B (37) | | | | | | | | | | | | |
| Burkina Faso | iLiNS−Zinc (38) | 157 | 53 | 50.9 | 0.57 (0.40, 0.82) | 0.65 | 0.47 | 170 | 48 | 58.3 | 0.46 (0.32, 0.66) | 0.59 | 0.44 |
| Burkina Faso | PROMIS CS (39) | | | | | | | | | | | | |
| Ghana | GHANA (40) | | | | | | | | | | | | |
| Ghana | iLiNS−DYADG (41) | | | | | | | | | | | | |
| Kenya | WASH−B (42) | 146 | 125 | 30.4 | 0.34 (0.18, 0.63) | 0.21 | 0.30 | 152 | 133 | 36.1 | 0.24 (0.14, 0.41) | 0.26 | 0.32 |
| Madagascar | MAHAY (43) | | | | | | | | | | | | |
| Malawi | iLiNS−DYADM (44) | | | | | | | | | | | | |
| Malawi | iLiNS−DOSE (45) | | | | | | | | | | | | |
| Mali | PROMIS CS (46) | | | | | | | | | | | | |
| Zimbabwe | SHINE (HIV−) (47) | | | | | | | | | | | | |
| Zimbabwe | SHINE (HIV+) (48) | | | | | | | | | | | | |
| | | **569** | **309** | | **I² = 0.55, Tau² = 0.09** | | | **603** | **322** | | **I² = 0.61, Tau² = 0.09** | | |
| **Fixed** | | | | | **0.46 (0.34, 0.62)** | | | | | | **0.35 (0.27, 0.46)** | | |
| **Random** | | | | | **0.41 (0.26, 0.65)** | | | | | | **0.32 (0.20, 0.50)** | | |

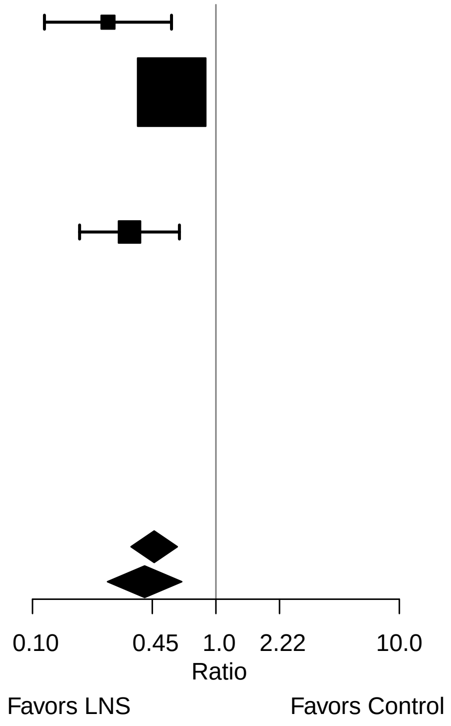

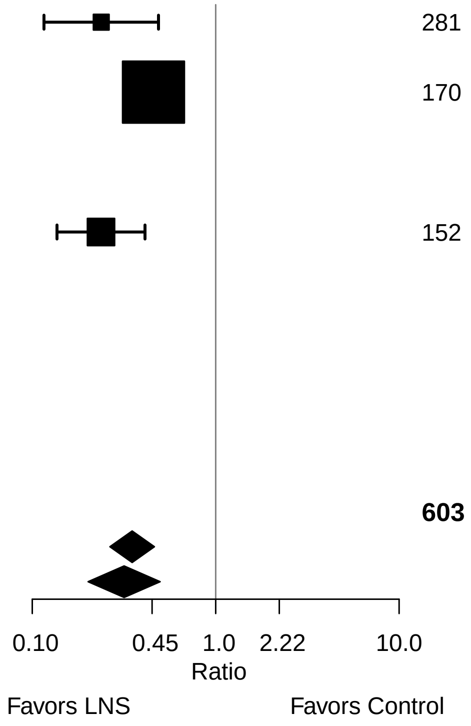

#### Supplemental figure 6I: Iron deficiency anemia prevalence ratio

#### 6I5: Stratified by Child birth order

**P−for−interaction = 0.904**
**Ratio of PRs = 0.97 (0.62, 1.53)**

| Country | Trial | LNS N | Control N | Control Prevalence | Later born PR (95% CI) | Fixed W | Random W | LNS N | Control N | Control Prevalence | Firstborn PR (95% CI) | Fixed W | Random W |
|---|---|---|---|---|---|---|---|---|---|---|---|---|---|
| Bangladesh | JiVitA−4 (35) | | | | | | | | | | | | |
| Bangladesh | RDNS (36) | 339 | 171 | 14.6 | 0.26 (0.13, 0.51) | 0.11 | 0.22 | 210 | 101 | 14.9 | 0.22 (0.10, 0.49) | 0.26 | 0.32 |
| Bangladesh | WASH−B (37) | | | | | | | | | | | | |
| Burkina Faso | iLiNS−Zinc (38) | 257 | 71 | 56.3 | 0.50 (0.38, 0.65) | 0.69 | 0.48 | 70 | 30 | 50.0 | 0.55 (0.32, 0.92) | 0.57 | 0.43 |
| Burkina Faso | PROMIS CS (39) | | | | | | | | | | | | |
| Ghana | GHANA (40) | | | | | | | | | | | | |
| Ghana | iLiNS−DYADG (41) | | | | | | | | | | | | |
| Kenya | WASH−B (42) | 237 | 213 | 32.4 | 0.30 (0.18, 0.50) | 0.20 | 0.30 | 60 | 45 | 37.8 | 0.22 (0.08, 0.58) | 0.17 | 0.25 |
| Madagascar | MAHAY (43) | | | | | | | | | | | | |
| Malawi | iLiNS−DYADM (44) | | | | | | | | | | | | |
| Malawi | iLiNS−DOSE (45) | | | | | | | | | | | | |
| Mali | PROMIS CS (46) | | | | | | | | | | | | |
| Zimbabwe | SHINE (HIV−) (47) | | | | | | | | | | | | |
| Zimbabwe | SHINE (HIV+) (48) | | | | | | | | | | | | |
| | | **833** | **455** | | **I² = 0.60, Tau² = 0.07** | | | **340** | **176** | | **I² = 0.58, Tau² = 0.16** | | |
| **Fixed** | | | | | **0.42 (0.33, 0.53)** | | | | | | **0.37 (0.25, 0.55)** | | |
| **Random** | | | | | **0.37 (0.25, 0.55)** | | | | | | **0.33 (0.18, 0.61)** | | |

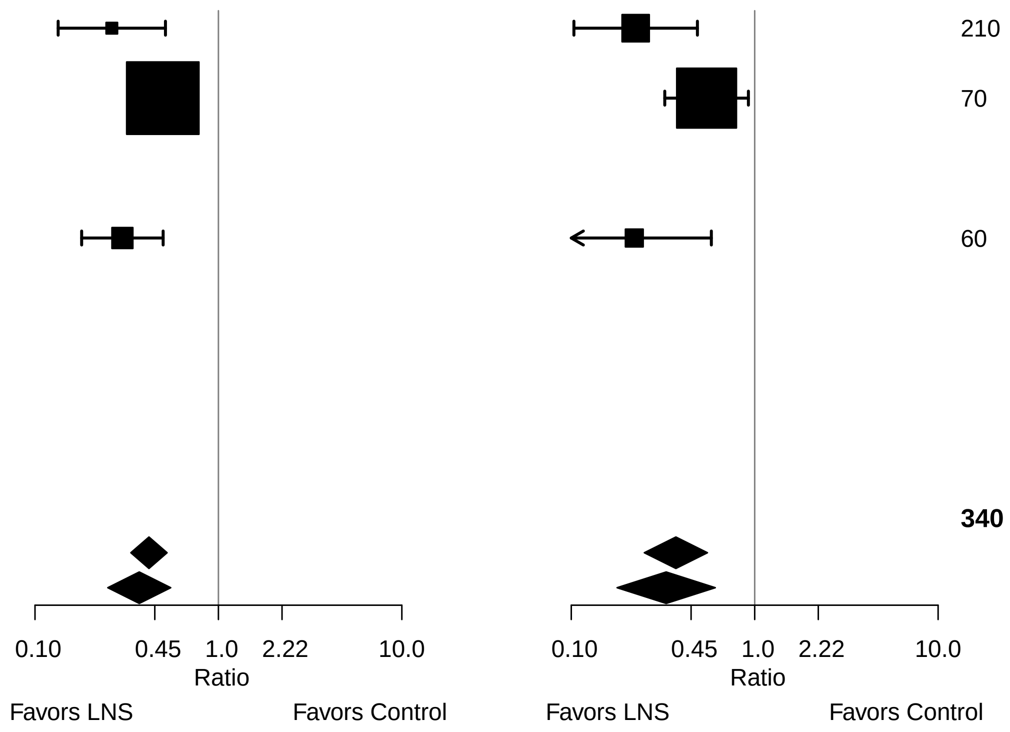

**Supplemental figure 6I: Iron deficiency anemia prevalence ratio**

#### 6I6: Stratified by Child baseline acute malnutrition (insufficient comparisons)

**Supplemental figure 6I: Iron deficiency anemia prevalence ratio**

#### 6I7: Stratified by Child baseline anemia (insufficient comparisons)

**Supplemental figure 6I: Iron deficiency anemia prevalence ratio**

#### 6I8: Stratified by Child high-dose vitamin A supplementation (insufficient comparisons)

#### Supplemental figure 6I: Iron deficiency anemia prevalence ratio

##### 6I9: Stratified by Child inflammation

**P−for−interaction = 0.388**
**Ratio of PRs = 1.21 (0.79, 1.86)**

| | | **Not inflamed** | | | | | | **High AGP or CRP** | | | | | |
|---|---|---|---|---|---|---|---|---|---|---|---|---|---|
| Country | Trial | **LNS** N | **Control** N | **Control** Prevalence | **PR** (95% CI) | **Fixed** W | **Random** W | **LNS** N | **Control** N | **Control** Prevalence | **PR** (95% CI) | **Fixed** W | **Random** W |
| Bangladesh | JiVitA−4 (35) | | | | | | | | | | | | |
| Bangladesh | RDNS (36) | 369 | 168 | 15.5 | 0.19 (0.10, 0.37) | 0.24 | 0.28 | 180 | 104 | 13.5 | 0.37 (0.18, 0.78) | 0.12 | 0.27 |
| Bangladesh | WASH−B (37) | | | | | | | | | | | | |
| Burkina Faso | iLiNS−Zinc (38) | 130 | 32 | 53.1 | 0.40 (0.24, 0.66) | 0.42 | 0.38 | 197 | 69 | 55.1 | 0.58 (0.43, 0.79) | 0.73 | 0.43 |
| Burkina Faso | PROMIS CS (39) | | | | | | | | | | | | |
| Ghana | GHANA (40) | | | | | | | | | | | | |
| Ghana | iLiNS−DYADG (41) | | | | | | | | | | | | |
| Kenya | WASH−B (42) | 157 | 135 | 33.3 | 0.34 (0.20, 0.60) | 0.34 | 0.34 | 141 | 123 | 33.3 | 0.21 (0.11, 0.42) | 0.15 | 0.29 |
| Madagascar | MAHAY (43) | | | | | | | | | | | | |
| Malawi | iLiNS−DYADM (44) | | | | | | | | | | | | |
| Malawi | iLiNS−DOSE (45) | | | | | | | | | | | | |
| Mali | PROMIS CS (46) | | | | | | | | | | | | |
| Zimbabwe | SHINE (HIV−) (47) | | | | | | | | | | | | |
| Zimbabwe | SHINE (HIV+) (48) | | | | | | | | | | | | |
| | | **656** | **335** | | **I² = 0.36, Tau² = 0.05** | | | **518** | **296** | | **I² = 0.73, Tau² = 0.18** | | |
| **Fixed** | | | | | **0.32 (0.23, 0.44)** | | | | | | **0.47 (0.37, 0.61)** | | |
| **Random** | | | | | **0.31 (0.20, 0.47)** | | | | | | **0.38 (0.21, 0.68)** | | |

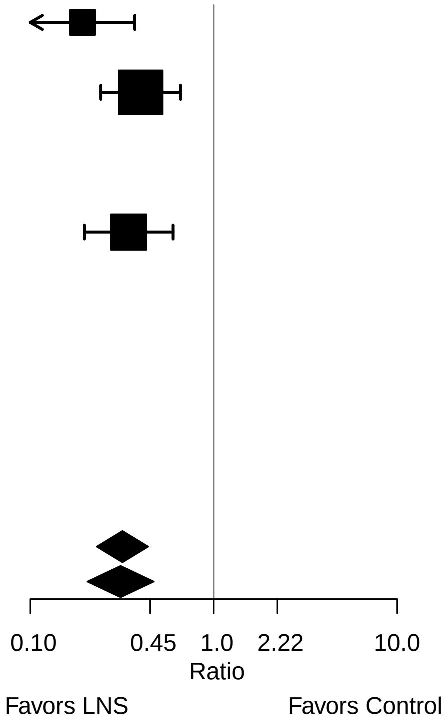

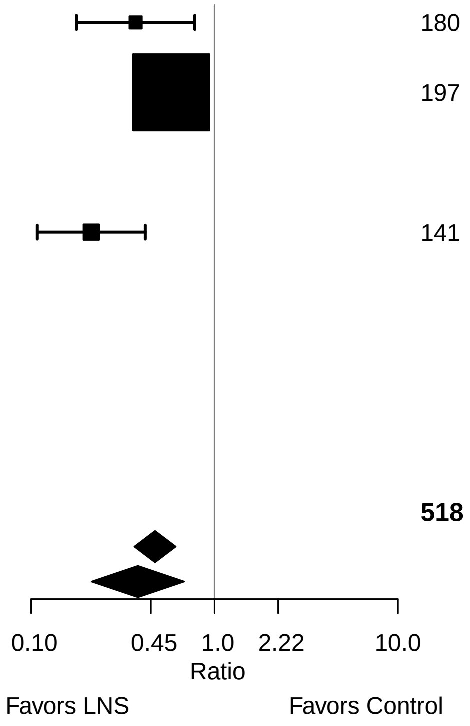

### Supplemental figure 6J: Iron deficiency anemia prevalence difference

#### 6J1: Stratified by Maternal BMI

**P−for−interaction = 0.023**
**Difference in PDs = 0.09 (0.01, 0.16)**

| | | At least 20 kg/m² | | | | | | Less than 20 kg/m² | | | | | |
|---|---|---|---|---|---|---|---|---|---|---|---|---|---|
| Country | Trial | LNS N | Control N | Control Prevalence | PD (95% CI) | Fixed W | Random W | LNS N | Control N | Control Prevalence | PD (95% CI) | Fixed W | Random W |
| Bangladesh | JiVitA−4 (35) | | | | | | | | | | | | |
| Bangladesh | RDNS (36) | 227 | 102 | 16.7 | −0.14 (−0.21, −0.06) | 0.44 | 0.40 | 300 | 160 | 13.1 | −0.09 (−0.16, −0.02) | 0.57 | 0.36 |
| Bangladesh | WASH−B (37) | | | | | | | | | | | | |
| Burkina Faso | iLiNS−Zinc (38) | 215 | 59 | 52.5 | −0.25 (−0.41, −0.09) | 0.11 | 0.20 | 112 | 42 | 57.1 | −0.30 (−0.41, −0.19) | 0.23 | 0.32 |
| Burkina Faso | PROMIS CS (39) | | | | | | | | | | | | |
| Ghana | GHANA (40) | | | | | | | | | | | | |
| Ghana | iLiNS−DYADG (41) | | | | | | | | | | | | |
| Kenya | WASH−B (42) | 223 | 199 | 36.7 | −0.27 (−0.35, −0.19) | 0.45 | 0.40 | 64 | 51 | 13.7 | −0.04 (−0.16, 0.08) | 0.20 | 0.32 |
| Madagascar | MAHAY (43) | | | | | | | | | | | | |
| Malawi | iLiNS−DYADM (44) | | | | | | | | | | | | |
| Malawi | iLiNS−DOSE (45) | | | | | | | | | | | | |
| Mali | PROMIS CS (46) | | | | | | | | | | | | |
| Zimbabwe | SHINE (HIV−) (47) | | | | | | | | | | | | |
| Zimbabwe | SHINE (HIV+) (48) | | | | | | | | | | | | |
| | | **665** | **360** | | **I² = 0.67, Tau² = 0.00** | | | **476** | **253** | | **I² = 0.84, Tau² = 0.02** | | |
| **Fixed** | | | | | **−0.21 (−0.26, −0.16)** | | | | | | **−0.13 (−0.18, −0.08)** | | |
| **Random** | | | | | **−0.21 (−0.30, −0.13)** | | | | | | **−0.14 (−0.30, 0.01)** | | |

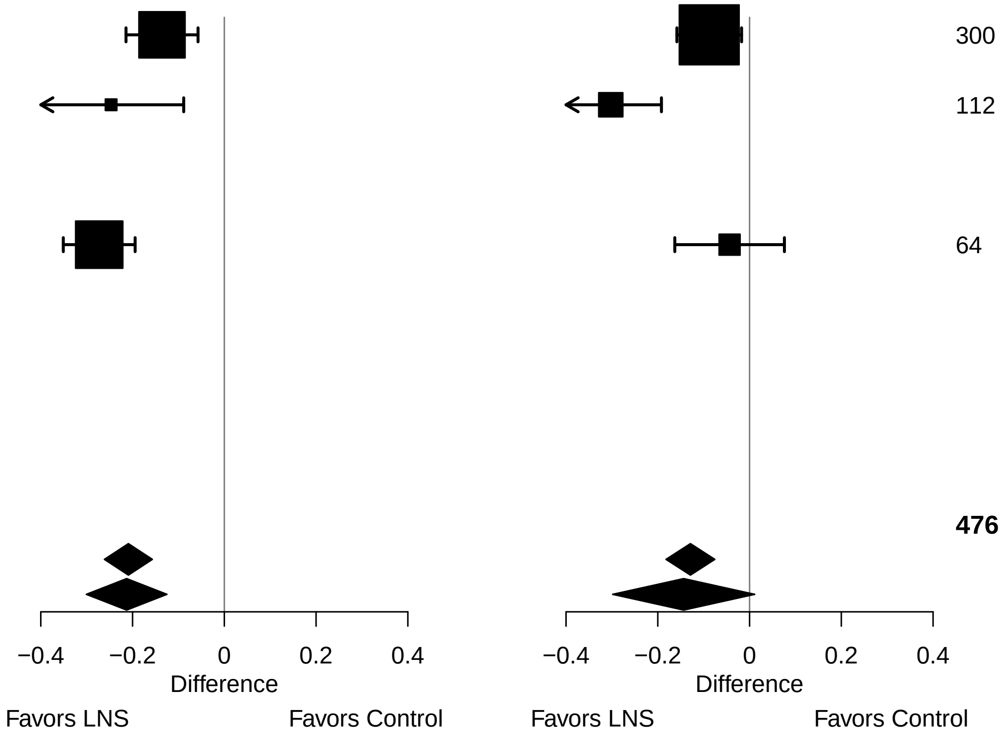

**Supplemental figure 6J: Iron deficiency anemia prevalence difference**

#### 6J2: Stratified by Maternal age (insufficient comparisons)

**Supplemental figure 6J: Iron deficiency anemia prevalence difference**

#### 6J3: Stratified by Maternal education (insufficient comparisons)

#### Supplemental figure 6J: Iron deficiency anemia prevalence difference

#### 6J4: Stratified by Child sex

**P−for−interaction = 0.555**
**Difference in PDs = −0.02 (−0.09, 0.05)**

| Country | Trial | Male LNS N | Male Control N | Male Control Prevalence | Male PD (95% CI) | Male Fixed W | Male Random W | Female LNS N | Female Control N | Female Control Prevalence | Female PD (95% CI) | Female Fixed W | Female Random W |
|---|---|---|---|---|---|---|---|---|---|---|---|---|---|
| Bangladesh | JiVitA−4 (35) | | | | | | | | | | | | |
| Bangladesh | RDNS (36) | 266 | 131 | 16.0 | −0.12 (−0.19, −0.04) | 0.56 | 0.52 | 281 | 141 | 13.5 | −0.10 (−0.16, −0.04) | 0.62 | 0.39 |
| Bangladesh | WASH−B (37) | | | | | | | | | | | | |
| Burkina Faso | iLiNS−Zinc (38) | 157 | 53 | 50.9 | −0.22 (−0.36, −0.08) | 0.15 | 0.17 | 170 | 48 | 58.3 | −0.32 (−0.47, −0.16) | 0.08 | 0.26 |
| Burkina Faso | PROMIS CS (39) | | | | | | | | | | | | |
| Ghana | GHANA (40) | | | | | | | | | | | | |
| Ghana | iLiNS−DYADG (41) | | | | | | | | | | | | |
| Kenya | WASH−B (42) | 146 | 125 | 30.4 | −0.20 (−0.31, −0.10) | 0.29 | 0.30 | 152 | 133 | 36.1 | −0.28 (−0.36, −0.19) | 0.30 | 0.36 |
| Madagascar | MAHAY (43) | | | | | | | | | | | | |
| Malawi | iLiNS−DYADM (44) | | | | | | | | | | | | |
| Malawi | iLiNS−DOSE (45) | | | | | | | | | | | | |
| Mali | PROMIS CS (46) | | | | | | | | | | | | |
| Zimbabwe | SHINE (HIV−) (47) | | | | | | | | | | | | |
| Zimbabwe | SHINE (HIV+) (48) | | | | | | | | | | | | |
| | | **569** | **309** | | **$I^2$ = 0.18, $Tau^2$ = 0.00** | | | **603** | **322** | | **$I^2$ = 0.86, $Tau^2$ = 0.01** | | |
| **Fixed** | | | | | **−0.16 (−0.21, −0.10)** | | | | | | **−0.17 (−0.22, −0.13)** | | |
| **Random** | | | | | **−0.16 (−0.22, −0.10)** | | | | | | **−0.22 (−0.35, −0.09)** | | |

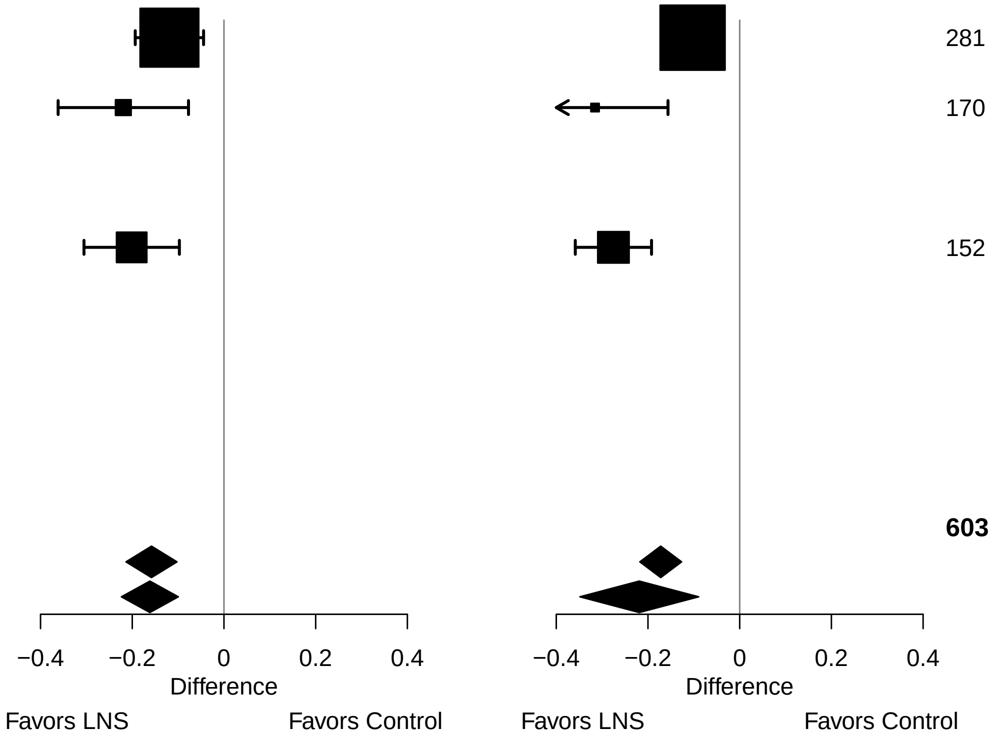

**Supplemental figure 6J: Iron deficiency anemia prevalence difference**

#### 6J5: Stratified by Child birth order

**P−for−interaction = 0.745**
**Difference in PDs = −0.01 (−0.09, 0.06)**

| Country | Trial | LNS N | Control N | Control Prevalence | Later born PD (95% CI) | Fixed W | Random W | LNS N | Control N | Control Prevalence | Firstborn PD (95% CI) | Fixed W | Random W |
|---|---|---|---|---|---|---|---|---|---|---|---|---|---|
| Bangladesh | JiVitA−4 (35) | | | | | | | | | | | | |
| Bangladesh | RDNS (36) | 339 | 171 | 14.6 | −0.11 (−0.16, −0.05) | 0.54 | 0.37 | 210 | 101 | 14.9 | −0.12 (−0.18, −0.05) | 0.82 | 0.60 |
| Bangladesh | WASH−B (37) | | | | | | | | | | | | |
| Burkina Faso | iLiNS−Zinc (38) | 257 | 71 | 56.3 | −0.28 (−0.38, −0.18) | 0.18 | 0.30 | 70 | 30 | 50.0 | −0.23 (−0.45, −0.01) | 0.08 | 0.18 |
| Burkina Faso | PROMIS CS (39) | | | | | | | | | | | | |
| Ghana | GHANA (40) | | | | | | | | | | | | |
| Ghana | iLiNS−DYADG (41) | | | | | | | | | | | | |
| Kenya | WASH−B (42) | 237 | 213 | 32.4 | −0.23 (−0.31, −0.15) | 0.28 | 0.33 | 60 | 45 | 37.8 | −0.29 (−0.49, −0.10) | 0.10 | 0.21 |
| Madagascar | MAHAY (43) | | | | | | | | | | | | |
| Malawi | iLiNS−DYADM (44) | | | | | | | | | | | | |
| Malawi | iLiNS−DOSE (45) | | | | | | | | | | | | |
| Mali | PROMIS CS (46) | | | | | | | | | | | | |
| Zimbabwe | SHINE (HIV−) (47) | | | | | | | | | | | | |
| Zimbabwe | SHINE (HIV+) (48) | | | | | | | | | | | | |
| | | **833** | **455** | | **I² = 0.83, Tau² = 0.01** | | | **340** | **176** | | **I² = 0.44, Tau² = 0.00** | | |
| **Fixed** | | | | | **−0.17 (−0.21, −0.13)** | | | | | | **−0.14 (−0.20, −0.08)** | | |
| **Random** | | | | | **−0.20 (−0.30, −0.10)** | | | | | | **−0.17 (−0.28, −0.07)** | | |

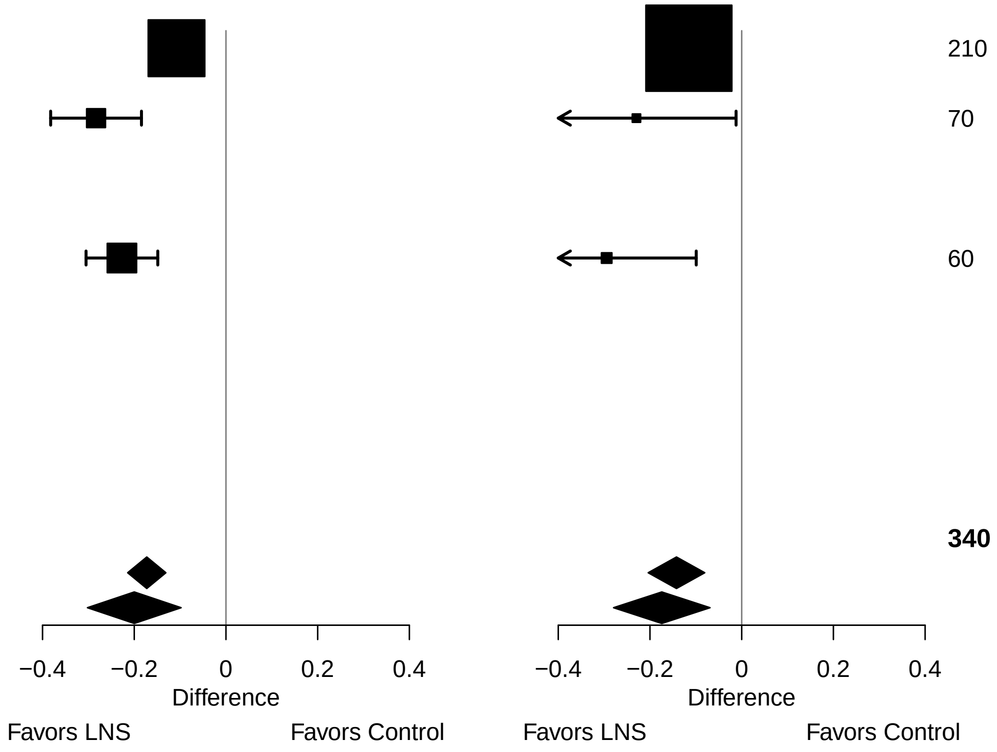

**Supplemental figure 6J: Iron deficiency anemia prevalence difference**

#### 6J6: Stratified by Child baseline acute malnutrition (insufficient comparisons)

**Supplemental figure 6J: Iron deficiency anemia prevalence difference**

#### 6J7: Stratified by Child baseline anemia (insufficient comparisons)

**Supplemental figure 6J: Iron deficiency anemia prevalence difference**

#### 6J8: Stratified by Child high-dose vitamin A supplementation (insufficient comparisons)

#### Supplemental figure 6J: Iron deficiency anemia prevalence difference

##### 6J9: Stratified by Child inflammation

**P−for−interaction = 0.590**
**Difference in PDs = 0.02 (−0.05, 0.09)**

| Country | Trial | Not inflamed: LNS N | Control N | Control Prevalence | PD (95% CI) | Fixed W | Random W | High AGP or CRP: LNS N | Control N | Control Prevalence | PD (95% CI) | Fixed W | Random W |
|---|---|---|---|---|---|---|---|---|---|---|---|---|---|
| Bangladesh | JiVitA−4 (35) | | | | | | | | | | | | |
| Bangladesh | RDNS (36) | 369 | 168 | 15.5 | −0.12 (−0.17, −0.08) | 0.76 | 0.48 | 180 | 104 | 13.5 | −0.08 (−0.15, −0.01) | 0.53 | 0.38 |
| Bangladesh | WASH−B (37) | | | | | | | | | | | | |
| Burkina Faso | iLiNS−Zinc (38) | 130 | 32 | 53.1 | −0.32 (−0.51, −0.12) | 0.05 | 0.18 | 197 | 69 | 55.1 | −0.23 (−0.36, −0.10) | 0.15 | 0.27 |
| Burkina Faso | PROMIS CS (39) | | | | | | | | | | | | |
| Ghana | GHANA (40) | | | | | | | | | | | | |
| Ghana | iLiNS−DYADG (41) | | | | | | | | | | | | |
| Kenya | WASH−B (42) | 157 | 135 | 33.3 | −0.22 (−0.32, −0.12) | 0.18 | 0.35 | 141 | 123 | 33.3 | −0.26 (−0.35, −0.17) | 0.33 | 0.35 |
| Madagascar | MAHAY (43) | | | | | | | | | | | | |
| Malawi | iLiNS−DYADM (44) | | | | | | | | | | | | |
| Malawi | iLiNS−DOSE (45) | | | | | | | | | | | | |
| Mali | PROMIS CS (46) | | | | | | | | | | | | |
| Zimbabwe | SHINE (HIV−) (47) | | | | | | | | | | | | |
| Zimbabwe | SHINE (HIV+) (48) | | | | | | | | | | | | |
| | | **656** | **335** | | **I² = 0.64, Tau² = 0.00** | | | **518** | **296** | | **I² = 0.81, Tau² = 0.01** | | |
| **Fixed** | | | | | **−0.15 (−0.20, −0.11)** | | | | | | **−0.16 (−0.22, −0.11)** | | |
| **Random** | | | | | **−0.19 (−0.29, −0.09)** | | | | | | **−0.19 (−0.30, −0.07)** | | |

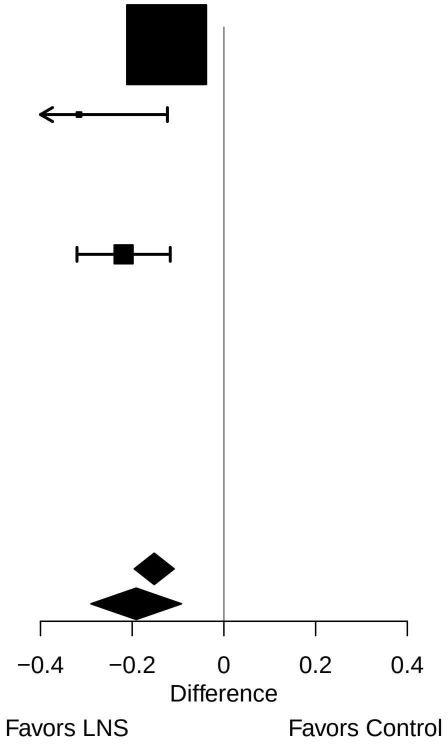

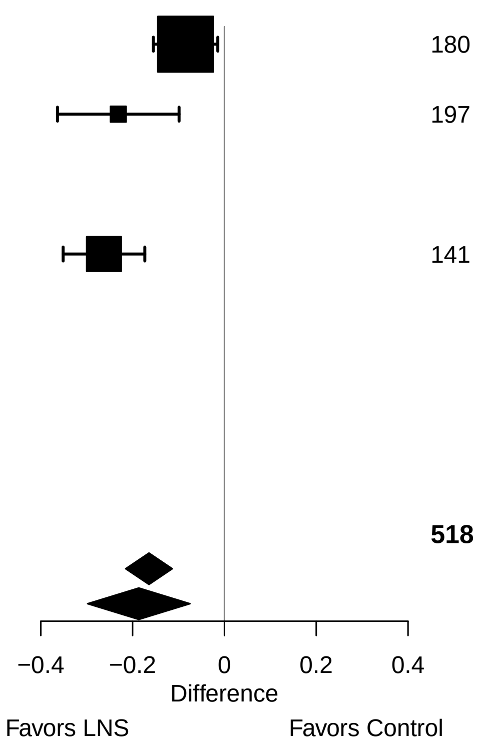

### Supplemental figure 6K: Geometric mean ratio of soluble transferrin receptor concentration

#### 6K1: Stratified by Maternal BMI

**P−for−interaction = 0.955**
**Ratio of GMRs = 1.00 (0.94, 1.06)**

| | | At least 20 kg/m² | | | | | | Less than 20 kg/m² | | | | | |
|---|---|---|---|---|---|---|---|---|---|---|---|---|---|
| | | **LNS** | **Control** | **Control** | **GMR** | **Fixed** | **Random** | **LNS** | **Control** | **Control** | **GMR** | **Fixed** | **Random** |
| Country | Trial | N | N | Median | (95% CI) | W | W | N | N | Median | (95% CI) | W | W |
| Bangladesh | JiVitA−4 (35) | | | | | | | | | | | | |
| Bangladesh | RDNS (36) | 228 | 102 | 8.1 | 0.88 (0.82, 0.94) | 0.37 | 0.29 | 300 | 160 | 8.4 | 0.87 (0.81, 0.93) | 0.45 | 0.44 |
| Bangladesh | WASH−B (37) | 93 | 97 | 7.6 | 0.81 (0.75, 0.88) | 0.28 | 0.27 | 119 | 81 | 7.5 | 0.81 (0.74, 0.89) | 0.28 | 0.28 |
| Burkina Faso | iLiNS−Zinc (38) | 209 | 55 | 10.0 | 0.74 (0.66, 0.82) | 0.15 | 0.21 | 106 | 41 | 9.9 | 0.78 (0.65, 0.93) | 0.07 | 0.07 |
| Burkina Faso | PROMIS CS (39) | | | | | | | | | | | | |
| Ghana | GHANA (40) | | | | | | | | | | | | |
| Ghana | iLiNS−DYADG (41) | | | | | | | | | | | | |
| Kenya | WASH−B (42) | 223 | 200 | 11.3 | 0.80 (0.72, 0.88) | 0.19 | 0.23 | 64 | 51 | 9.9 | 0.79 (0.71, 0.88) | 0.20 | 0.21 |
| Madagascar | MAHAY (43) | | | | | | | | | | | | |
| Malawi | iLiNS−DYADM (44) | | | | | | | | | | | | |
| Malawi | iLiNS−DOSE (45) | | | | | | | | | | | | |
| Mali | PROMIS CS (46) | | | | | | | | | | | | |
| Zimbabwe | SHINE (HIV−) (47) | | | | | | | | | | | | |
| Zimbabwe | SHINE (HIV+) (48) | | | | | | | | | | | | |
| | | **753** | **454** | | **I² = 0.65, Tau² = 0.00** | | | **589** | **333** | | **I² = 0.03, Tau² = 0.00** | | |
| **Fixed** | | | | | **0.82 (0.79, 0.86)** | | | | | | **0.83 (0.79, 0.87)** | | |
| **Random** | | | | | **0.81 (0.75, 0.87)** | | | | | | **0.83 (0.79, 0.87)** | | |

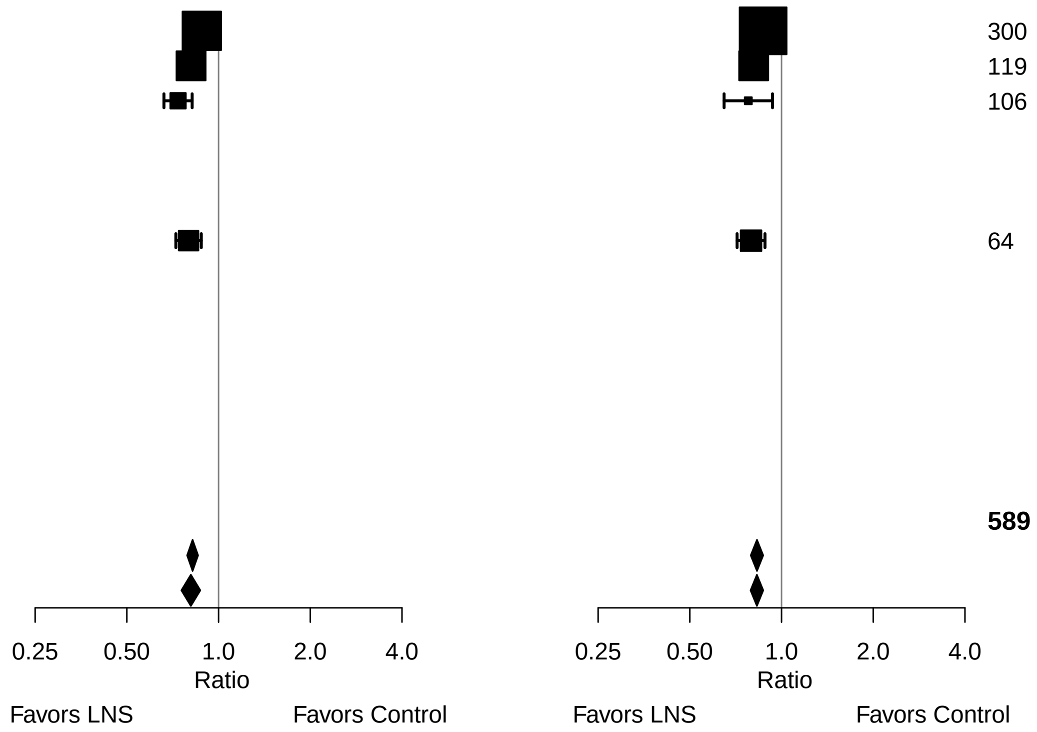

#### Supplemental figure 6K: Geometric mean ratio of soluble transferrin receptor concentration

##### 6K2: Stratified by Maternal age

**P−for−interaction = 0.213**
**Ratio of GMRs = 0.96 (0.91, 1.02)**

| | | At least 25 y | | | | | | Less than 25 y | | | | | |
|---|---|---|---|---|---|---|---|---|---|---|---|---|---|
| Country | Trial | LNS N | Control N | Control Median | GMR (95% CI) | Fixed W | Random W | LNS N | Control N | Control Median | GMR (95% CI) | Fixed W | Random W |
| Bangladesh | JiVitA−4 (35) | | | | | | | | | | | | |
| Bangladesh | RDNS (36) | 152 | 81 | 7.8 | 0.88 (0.79, 0.97) | 0.20 | 0.20 | 398 | 191 | 8.6 | 0.87 (0.83, 0.91) | 0.58 | 0.29 |
| Bangladesh | WASH−B (37) | 79 | 80 | 7.3 | 0.90 (0.82, 0.98) | 0.27 | 0.25 | 132 | 98 | 7.6 | 0.76 (0.69, 0.84) | 0.12 | 0.18 |
| Burkina Faso | iLiNS−Zinc (38) | 178 | 50 | 9.5 | 0.77 (0.67, 0.89) | 0.09 | 0.11 | 137 | 46 | 10.1 | 0.75 (0.68, 0.82) | 0.13 | 0.19 |
| Burkina Faso | PROMIS CS (39) | | | | | | | | | | | | |
| Ghana | GHANA (40) | 62 | 55 | 9.6 | 0.79 (0.70, 0.89) | 0.14 | 0.15 | 22 | 21 | 12.8 | 0.83 (0.65, 1.05) | 0.02 | 0.06 |
| Ghana | iLiNS−DYADG (41) | | | | | | | | | | | | |
| Kenya | WASH−B (42) | 178 | 149 | 10.8 | 0.80 (0.73, 0.88) | 0.26 | 0.24 | 116 | 109 | 11.3 | 0.78 (0.70, 0.87) | 0.11 | 0.18 |
| Madagascar | MAHAY (43) | 47 | 25 | 9.0 | 0.85 (0.68, 1.06) | 0.04 | 0.05 | 36 | 26 | 9.0 | 0.94 (0.78, 1.14) | 0.04 | 0.09 |
| Malawi | iLiNS−DYADM (44) | | | | | | | | | | | | |
| Malawi | iLiNS−DOSE (45) | | | | | | | | | | | | |
| Mali | PROMIS CS (46) | | | | | | | | | | | | |
| Zimbabwe | SHINE (HIV−) (47) | | | | | | | | | | | | |
| Zimbabwe | SHINE (HIV+) (48) | | | | | | | | | | | | |
| | | **696** | **440** | | **I² = 0.23, Tau² = 0.00** | | | **841** | **491** | | **I² = 0.63, Tau² = 0.00** | | |
| **Fixed** | | | | | **0.84 (0.80, 0.88)** | | | | | | **0.83 (0.80, 0.86)** | | |
| **Random** | | | | | **0.84 (0.79, 0.88)** | | | | | | **0.81 (0.76, 0.87)** | | |

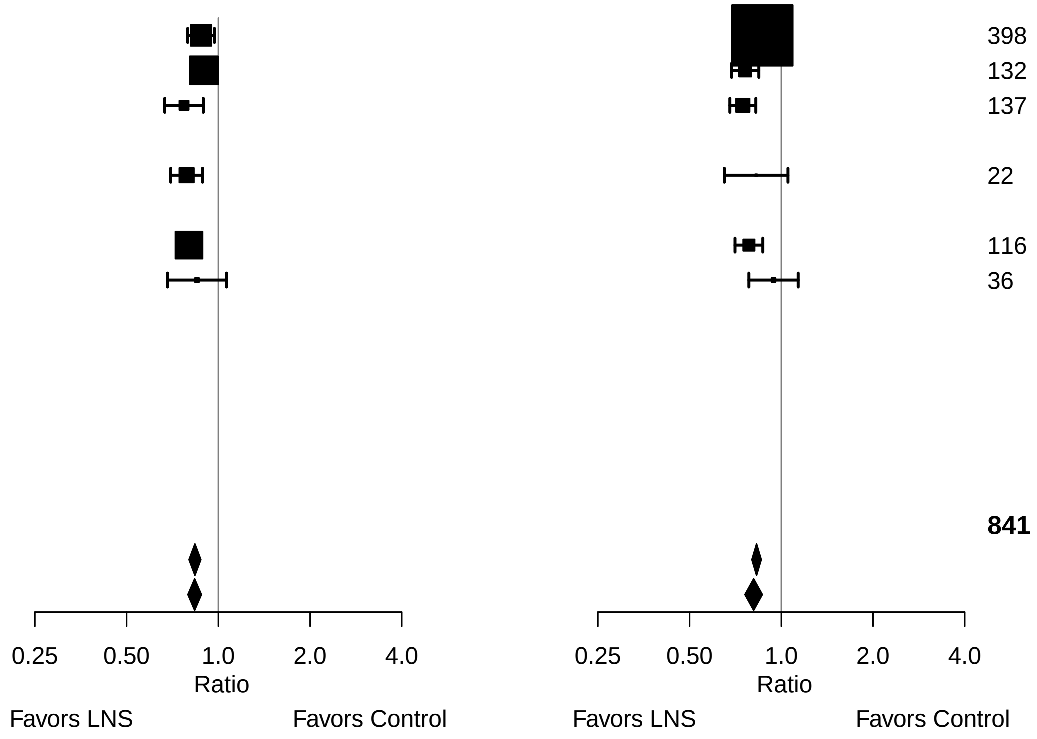

**Supplemental figure 6K: Geometric mean ratio of soluble transferrin receptor concentration**

#### 6K3: Stratified by Maternal education

**P−for−interaction = 0.576**
**Ratio of GMRs = 0.98 (0.92, 1.05)**

| | | Primary or greater | | | | | | Incomplete or no formal | | | | | |
|---|---|---|---|---|---|---|---|---|---|---|---|---|---|
| | | **LNS** | **Control** | **Control** | **GMR** | **Fixed** | **Random** | **LNS** | **Control** | **Control** | **GMR** | **Fixed** | **Random** |
| Country | Trial | N | N | Median | (95% CI) | W | W | N | N | Median | (95% CI) | W | W |
| Bangladesh | JiVitA−4 (35) | | | | | | | | | | | | |
| Bangladesh | RDNS (36) | 406 | 195 | 8.2 | 0.89 (0.84, 0.94) | 0.47 | 0.33 | 144 | 77 | 8.9 | 0.82 (0.74, 0.91) | 0.31 | 0.31 |
| Bangladesh | WASH−B (37) | 149 | 139 | 7.6 | 0.82 (0.76, 0.88) | 0.27 | 0.27 | 63 | 39 | 7.5 | 0.81 (0.71, 0.93) | 0.18 | 0.18 |
| Burkina Faso | iLiNS−Zinc (38) | | | | | | | | | | | | |
| Burkina Faso | PROMIS CS (39) | | | | | | | | | | | | |
| Ghana | GHANA (40) | 77 | 65 | 9.3 | 0.83 (0.74, 0.93) | 0.11 | 0.16 | 7 | 13 | 12.9 | 0.69 (0.52, 0.91) | 0.04 | 0.04 |
| Ghana | iLiNS−DYADG (41) | | | | | | | | | | | | |
| Kenya | WASH−B (42) | 143 | 117 | 11.3 | 0.76 (0.68, 0.84) | 0.13 | 0.19 | 154 | 142 | 10.4 | 0.82 (0.75, 0.90) | 0.38 | 0.38 |
| Madagascar | MAHAY (43) | 16 | 16 | 8.8 | 0.86 (0.67, 1.12) | 0.02 | 0.05 | 67 | 35 | 9.0 | 0.90 (0.75, 1.08) | 0.09 | 0.09 |
| Malawi | iLiNS−DYADM (44) | | | | | | | | | | | | |
| Malawi | iLiNS−DOSE (45) | | | | | | | | | | | | |
| Mali | PROMIS CS (46) | | | | | | | | | | | | |
| Zimbabwe | SHINE (HIV−) (47) | | | | | | | | | | | | |
| Zimbabwe | SHINE (HIV+) (48) | | | | | | | | | | | | |
| | | **791** | **532** | | **I² = 0.55, Tau² = 0.00** | | | **435** | **306** | | **I² = 0.00, Tau² = 0.00** | | |
| **Fixed** | | | | | **0.84 (0.81, 0.88)** | | | | | | **0.82 (0.78, 0.87)** | | |
| **Random** | | | | | **0.83 (0.79, 0.88)** | | | | | | **0.82 (0.78, 0.87)** | | |

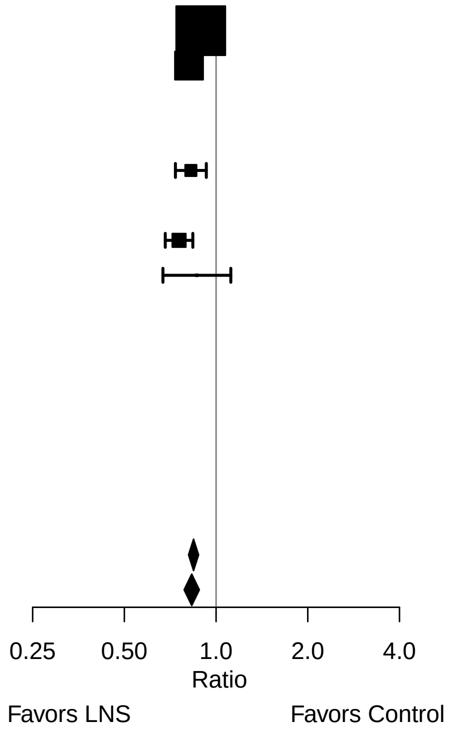

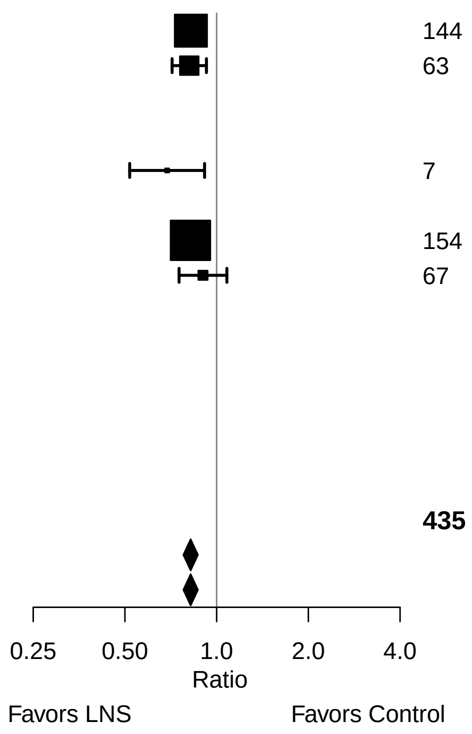

#### Supplemental figure 6K: Geometric mean ratio of soluble transferrin receptor concentration

#### 6K4: Stratified by Child sex

**P−for−interaction = 0.268**
**Ratio of GMRs = 1.03 (0.98, 1.09)**

| Country | Trial | Male LNS N | Male Control N | Male Control Median | Male GMR (95% CI) | Male Fixed W | Male Random W | Female LNS N | Female Control N | Female Control Median | Female GMR (95% CI) | Female Fixed W | Female Random W |
|---|---|---|---|---|---|---|---|---|---|---|---|---|---|
| Bangladesh | JiVitA−4 (35) | | | | | | | | | | | | |
| Bangladesh | RDNS (36) | 266 | 131 | 8.8 | 0.84 (0.77, 0.91) | 0.33 | 0.33 | 281 | 141 | 7.7 | 0.90 (0.86, 0.94) | 0.51 | 0.25 |
| Bangladesh | WASH−B (37) | 105 | 85 | 7.9 | 0.80 (0.70, 0.90) | 0.15 | 0.15 | 107 | 93 | 7.4 | 0.83 (0.76, 0.92) | 0.14 | 0.18 |
| Burkina Faso | iLiNS−Zinc (38) | 150 | 49 | 10.3 | 0.74 (0.64, 0.85) | 0.12 | 0.12 | 165 | 47 | 9.5 | 0.78 (0.70, 0.88) | 0.09 | 0.16 |
| Burkina Faso | PROMIS CS (39) | | | | | | | | | | | | |
| Ghana | GHANA (40) | 52 | 37 | 10.8 | 0.79 (0.68, 0.92) | 0.10 | 0.10 | 32 | 45 | 9.3 | 0.78 (0.67, 0.91) | 0.05 | 0.11 |
| Ghana | iLiNS−DYADG (41) | | | | | | | | | | | | |
| Kenya | WASH−B (42) | 146 | 125 | 11.1 | 0.82 (0.74, 0.90) | 0.25 | 0.25 | 152 | 134 | 10.7 | 0.77 (0.70, 0.83) | 0.17 | 0.20 |
| Madagascar | MAHAY (43) | 34 | 22 | 9.3 | 0.83 (0.67, 1.04) | 0.05 | 0.05 | 49 | 29 | 7.7 | 0.95 (0.80, 1.14) | 0.04 | 0.09 |
| Malawi | iLiNS−DYADM (44) | | | | | | | | | | | | |
| Malawi | iLiNS−DOSE (45) | | | | | | | | | | | | |
| Mali | PROMIS CS (46) | | | | | | | | | | | | |
| Zimbabwe | SHINE (HIV−) (47) | | | | | | | | | | | | |
| Zimbabwe | SHINE (HIV+) (48) | | | | | | | | | | | | |
| | | **753** | **449** | | **I² = 0.00, Tau² = 0.00** | | | **786** | **489** | | **I² = 0.70, Tau² = 0.00** | | |
| **Fixed** | | | | | **0.81 (0.77, 0.85)** | | | | | | **0.85 (0.82, 0.88)** | | |
| **Random** | | | | | **0.81 (0.77, 0.85)** | | | | | | **0.83 (0.78, 0.89)** | | |

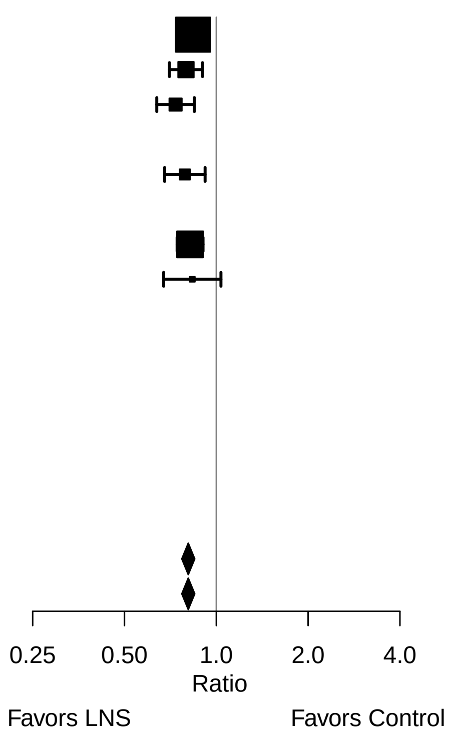

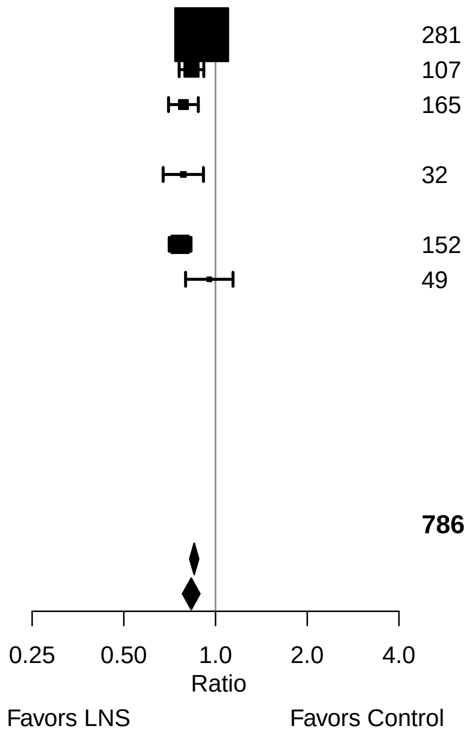

#### Supplemental figure 6K: Geometric mean ratio of soluble transferrin receptor concentration

##### 6K5: Stratified by Child birth order

**P−for−interaction = 0.731**
**Ratio of GMRs = 0.99 (0.93, 1.05)**

| | | **Later born** | | | | | | **Firstborn** | | | | | |
|---|---|---|---|---|---|---|---|---|---|---|---|---|---|
| Country | Trial | **LNS** N | **Control** N | **Control** Median | **GMR** (95% CI) | **Fixed** W | **Random** W | **LNS** N | **Control** N | **Control** Median | **GMR** (95% CI) | **Fixed** W | **Random** W |
| Bangladesh | JiVitA−4 (35) | | | | | | | | | | | | |
| Bangladesh | RDNS (36) | 340 | 171 | 8.3 | 0.87 (0.81, 0.92) | 0.29 | 0.23 | 210 | 101 | 8.1 | 0.88 (0.81, 0.94) | 0.47 | 0.28 |
| Bangladesh | WASH−B (37) | 124 | 119 | 7.6 | 0.83 (0.78, 0.89) | 0.28 | 0.23 | 86 | 57 | 7.4 | 0.79 (0.68, 0.90) | 0.13 | 0.17 |
| Burkina Faso | iLiNS−Zinc (38) | 249 | 71 | 9.0 | 0.78 (0.68, 0.90) | 0.06 | 0.11 | 66 | 25 | 10.3 | 0.70 (0.63, 0.79) | 0.20 | 0.21 |
| Burkina Faso | PROMIS CS (39) | | | | | | | | | | | | |
| Ghana | GHANA (40) | 54 | 50 | 9.8 | 0.76 (0.67, 0.86) | 0.08 | 0.12 | 30 | 29 | 9.4 | 0.88 (0.72, 1.07) | 0.07 | 0.11 |
| Ghana | iLiNS−DYADG (41) | | | | | | | | | | | | |
| Kenya | WASH−B (42) | 237 | 213 | 10.8 | 0.80 (0.74, 0.86) | 0.23 | 0.21 | 60 | 46 | 11.3 | 0.76 (0.65, 0.90) | 0.10 | 0.15 |
| Madagascar | MAHAY (43) | 61 | 37 | 8.8 | 0.95 (0.82, 1.11) | 0.05 | 0.10 | 21 | 11 | 9.4 | 0.91 (0.69, 1.18) | 0.04 | 0.07 |
| Malawi | iLiNS−DYADM (44) | | | | | | | | | | | | |
| Malawi | iLiNS−DOSE (45) | | | | | | | | | | | | |
| Mali | PROMIS CS (46) | | | | | | | | | | | | |
| Zimbabwe | SHINE (HIV−) (47) | | | | | | | | | | | | |
| Zimbabwe | SHINE (HIV+) (48) | | | | | | | | | | | | |
| | | **1065** | **661** | | **I² = 0.44, Tau² = 0.00** | | | **473** | **269** | | **I² = 0.60, Tau² = 0.00** | | |
| **Fixed** | | | | | **0.83 (0.80, 0.86)** | | | | | | **0.82 (0.78, 0.86)** | | |
| **Random** | | | | | **0.83 (0.78, 0.88)** | | | | | | **0.81 (0.74, 0.87)** | | |

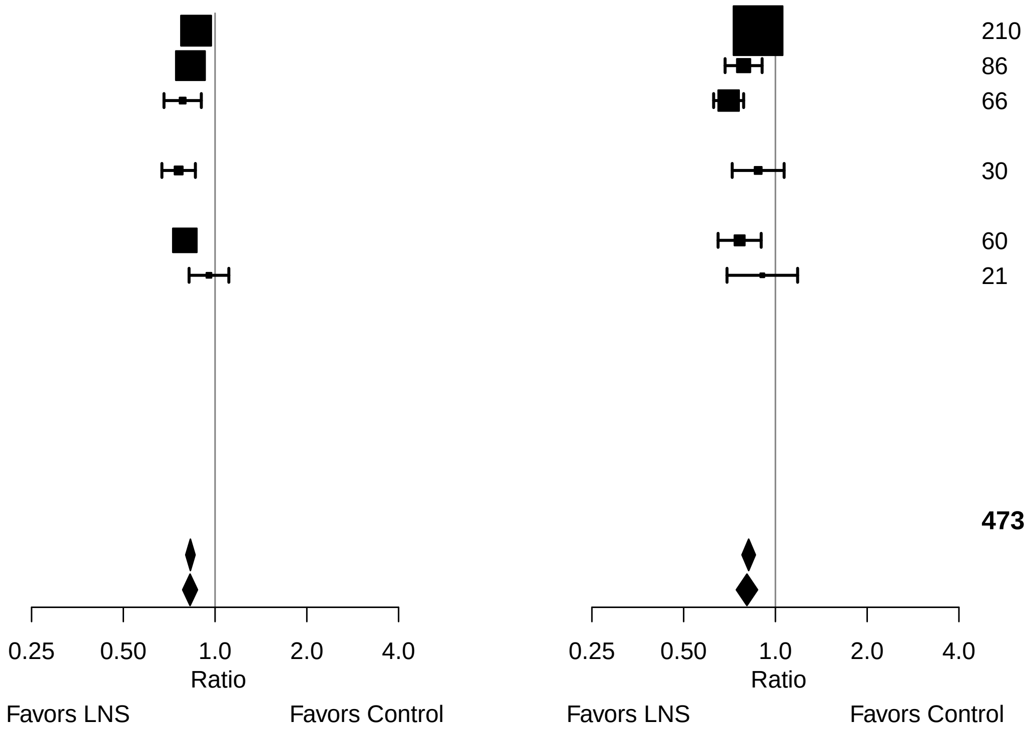

**Supplemental figure 6K: Geometric mean ratio of soluble transferrin receptor concentration**

#### 6K6: Stratified by Child baseline acute malnutrition (insufficient comparisons)

**Supplemental figure 6K: Geometric mean ratio of soluble transferrin receptor concentration**

#### 6K7: Stratified by Child baseline anemia (insufficient comparisons)

#### Supplemental figure 6K: Geometric mean ratio of soluble transferrin receptor concentration

##### 6K8: Stratified by Child high-dose vitamin A supplementation

**P−for−interaction = 0.373**
**Ratio of GMRs = 1.04 (0.96, 1.12)**

| | | Received VitA | | | | | | Not treated | | | | | |
|---|---|---|---|---|---|---|---|---|---|---|---|---|---|
| Country | Trial | LNS N | Control N | Control Median | GMR (95% CI) | Fixed W | Random W | LNS N | Control N | Control Median | GMR (95% CI) | Fixed W | Random W |
| Bangladesh | JiVitA−4 (35) | | | | | | | | | | | | |
| Bangladesh | RDNS (36) | 444 | 223 | 8.3 | 0.87 (0.82, 0.92) | 0.73 | 0.51 | 105 | 49 | 7.8 | 0.89 (0.82, 0.96) | 0.48 | 0.48 |
| Bangladesh | WASH−B (37) | 16 | 16 | 7.4 | 0.87 (0.75, 1.02) | 0.10 | 0.19 | 15 | 9 | 8.1 | 0.84 (0.62, 1.15) | 0.03 | 0.03 |
| Burkina Faso | iLiNS−Zinc (38) | | | | | | | | | | | | |
| Burkina Faso | PROMIS CS (39) | | | | | | | | | | | | |
| Ghana | GHANA (40) | 40 | 53 | 10.0 | 0.79 (0.69, 0.92) | 0.12 | 0.21 | 44 | 26 | 9.0 | 0.83 (0.70, 0.98) | 0.10 | 0.10 |
| Ghana | iLiNS−DYADG (41) | | | | | | | | | | | | |
| Kenya | WASH−B (42) | 26 | 31 | 12.0 | 0.70 (0.55, 0.88) | 0.05 | 0.10 | 223 | 180 | 11.0 | 0.81 (0.75, 0.89) | 0.39 | 0.39 |
| Madagascar | MAHAY (43) | | | | | | | | | | | | |
| Malawi | iLiNS−DYADM (44) | | | | | | | | | | | | |
| Malawi | iLiNS−DOSE (45) | | | | | | | | | | | | |
| Mali | PROMIS CS (46) | | | | | | | | | | | | |
| Zimbabwe | SHINE (HIV−) (47) | | | | | | | | | | | | |
| Zimbabwe | SHINE (HIV+) (48) | | | | | | | | | | | | |
| | | **526** | **323** | | **$I^2$ = 0.28, $Tau^2$ = 0.00** | | | **387** | **264** | | **$I^2$ = 0.00, $Tau^2$ = 0.00** | | |
| **Fixed** | | | | | **0.85 (0.81, 0.89)** | | | | | | **0.85 (0.81, 0.90)** | | |
| **Random** | | | | | **0.83 (0.77, 0.90)** | | | | | | **0.85 (0.81, 0.90)** | | |

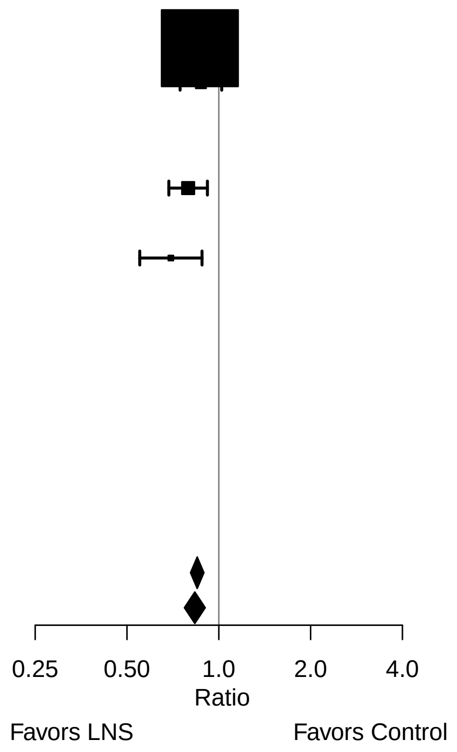

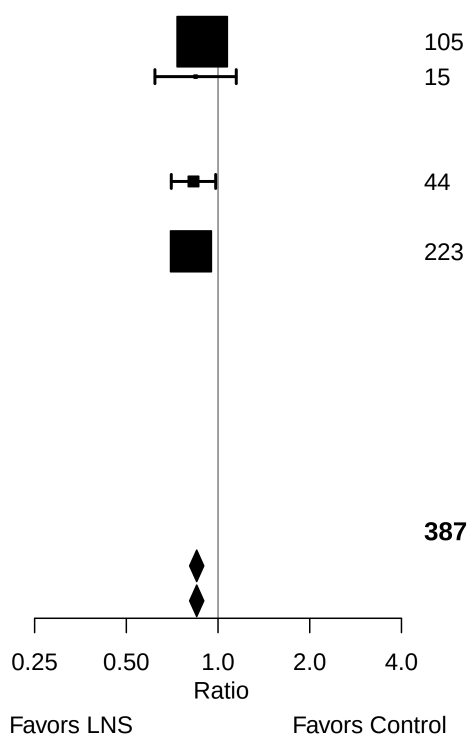

#### Supplemental figure 6K: Geometric mean ratio of soluble transferrin receptor concentration

#### 6K9: Stratified by Child inflammation

**P−for−interaction = 0.478**
**Ratio of GMRs = 0.98 (0.93, 1.03)**

| Country | Trial | Not inflamed: LNS N | Control N | Control Median | GMR (95% CI) | Fixed W | Random W | High AGP or CRP: LNS N | Control N | Control Median | GMR (95% CI) | Fixed W | Random W |
|---|---|---|---|---|---|---|---|---|---|---|---|---|---|
| Bangladesh | JiVitA−4 (35) | | | | | | | | | | | | |
| Bangladesh | RDNS (36) | 370 | 168 | 8.0 | 0.89 (0.83, 0.94) | 0.38 | 0.25 | 180 | 104 | 9.0 | 0.85 (0.79, 0.91) | 0.44 | 0.36 |
| Bangladesh | WASH−B (37) | 165 | 136 | 7.7 | 0.80 (0.74, 0.86) | 0.25 | 0.22 | 47 | 42 | 7.2 | 0.88 (0.78, 1.00) | 0.14 | 0.16 |
| Burkina Faso | iLiNS−Zinc (38) | 124 | 31 | 9.6 | 0.75 (0.66, 0.86) | 0.08 | 0.12 | 191 | 65 | 10.0 | 0.76 (0.68, 0.86) | 0.15 | 0.18 |
| Burkina Faso | PROMIS CS (39) | | | | | | | | | | | | |
| Ghana | GHANA (40) | 70 | 66 | 9.6 | 0.79 (0.70, 0.89) | 0.10 | 0.15 | 14 | 16 | 9.8 | 0.85 (0.65, 1.12) | 0.03 | 0.04 |
| Ghana | iLiNS−DYADG (41) | | | | | | | | | | | | |
| Kenya | WASH−B (42) | 157 | 135 | 10.1 | 0.83 (0.74, 0.93) | 0.12 | 0.16 | 141 | 124 | 11.7 | 0.75 (0.68, 0.83) | 0.21 | 0.23 |
| Madagascar | MAHAY (43) | 44 | 32 | 8.8 | 0.94 (0.81, 1.09) | 0.06 | 0.11 | 39 | 19 | 9.4 | 0.83 (0.62, 1.10) | 0.03 | 0.04 |
| Malawi | iLiNS−DYADM (44) | | | | | | | | | | | | |
| Malawi | iLiNS−DOSE (45) | | | | | | | | | | | | |
| Mali | PROMIS CS (46) | | | | | | | | | | | | |
| Zimbabwe | SHINE (HIV−) (47) | | | | | | | | | | | | |
| Zimbabwe | SHINE (HIV+) (48) | | | | | | | | | | | | |
| | | **930** | **568** | | **$I^2$ = 0.53, $Tau^2$ = 0.00** | | | **612** | **370** | | **$I^2$ = 0.28, $Tau^2$ = 0.00** | | |
| **Fixed** | | | | | **0.84 (0.81, 0.87)** | | | | | | **0.82 (0.78, 0.86)** | | |
| **Random** | | | | | **0.83 (0.78, 0.88)** | | | | | | **0.81 (0.77, 0.86)** | | |

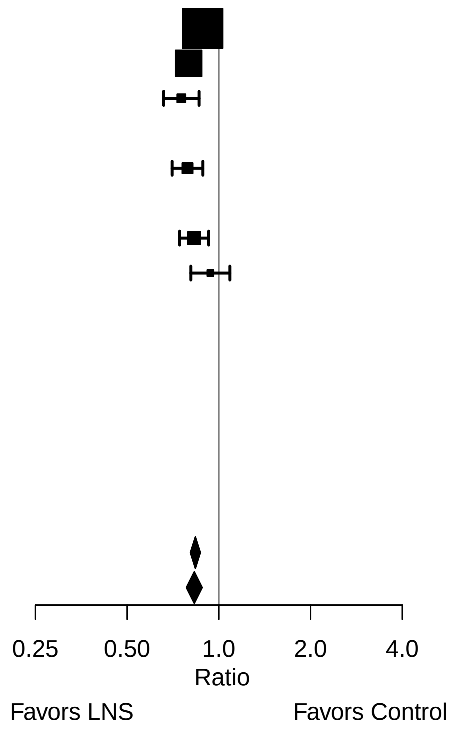

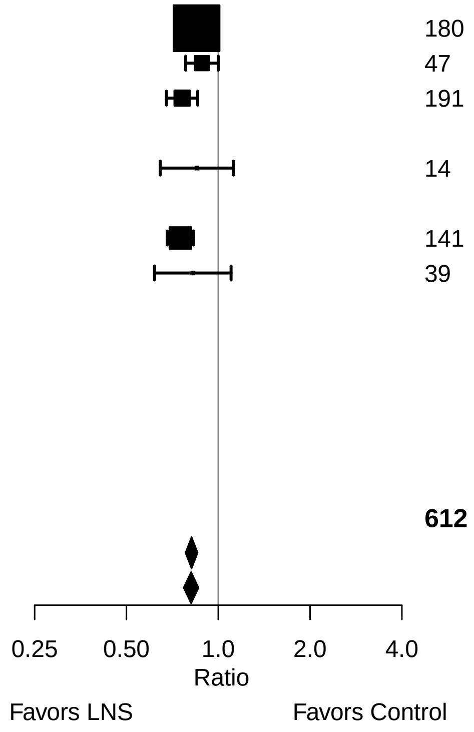

### Supplemental figure 6L: Elevated soluble transferrin receptor prevalence ratio

#### 6L1: Stratified by Maternal BMI

**P−for−interaction = 0.126**
**Ratio of PRs = 0.85 (0.69, 1.05)**

| Country | Trial | LNS N | Control N | Control Prevalence | At least 20 kg/m² PR (95% CI) | Fixed W | Random W | LNS N | Control N | Control Prevalence | Less than 20 kg/m² PR (95% CI) | Fixed W | Random W |
|---|---|---|---|---|---|---|---|---|---|---|---|---|---|
| Bangladesh | JiVitA−4 (35) | | | | | | | | | | | | |
| Bangladesh | RDNS (36) | 228 | 102 | 44.1 | 0.81 (0.61, 1.06) | 0.24 | 0.29 | 300 | 160 | 51.2 | 0.68 (0.55, 0.85) | 0.52 | 0.39 |
| Bangladesh | WASH−B (37) | | | | | | | | | | | | |
| Burkina Faso | iLiNS−Zinc (38) | 209 | 55 | 60.7 | 0.56 (0.40, 0.78) | 0.16 | 0.22 | 106 | 41 | 62.2 | 0.43 (0.30, 0.62) | 0.18 | 0.27 |
| Burkina Faso | PROMIS CS (39) | | | | | | | | | | | | |
| Ghana | GHANA (40) | | | | | | | | | | | | |
| Ghana | iLiNS−DYADG (41) | | | | | | | | | | | | |
| Kenya | WASH−B (42) | 223 | 200 | 72.0 | 0.70 (0.59, 0.83) | 0.60 | 0.49 | 64 | 51 | 76.5 | 0.65 (0.49, 0.87) | 0.30 | 0.34 |
| Madagascar | MAHAY (43) | | | | | | | | | | | | |
| Malawi | iLiNS−DYADM (44) | | | | | | | | | | | | |
| Malawi | iLiNS−DOSE (45) | | | | | | | | | | | | |
| Mali | PROMIS CS (46) | | | | | | | | | | | | |
| Zimbabwe | SHINE (HIV−) (47) | | | | | | | | | | | | |
| Zimbabwe | SHINE (HIV+) (48) | | | | | | | | | | | | |
| | | **660** | **357** | | **I² = 0.27, Tau² = 0.01** | | | **470** | **252** | | **I² = 0.59, Tau² = 0.04** | | |
| **Fixed** | | | | | **0.70 (0.61, 0.80)** | | | | | | **0.62 (0.53, 0.72)** | | |
| **Random** | | | | | **0.69 (0.58, 0.83)** | | | | | | **0.59 (0.45, 0.78)** | | |

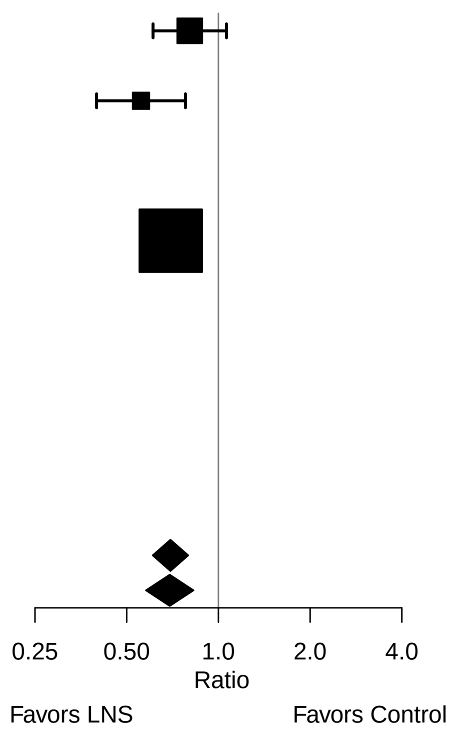

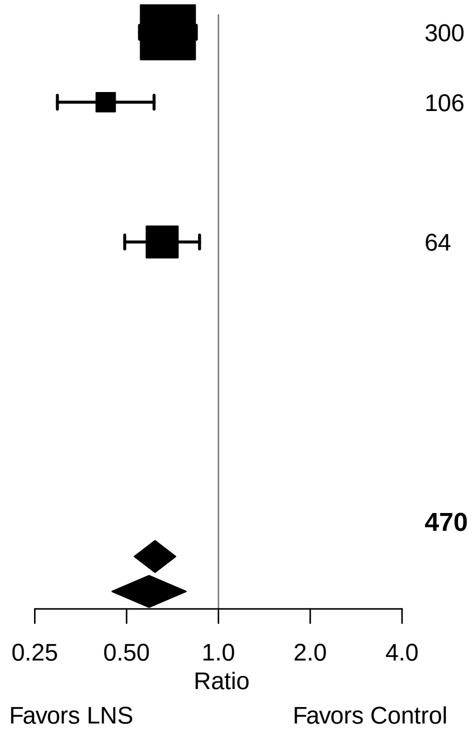

**Supplemental figure 6L: Elevated soluble transferrin receptor prevalence ratio**

#### 6L2: Stratified by Maternal age

**P−for−interaction = 0.585**
**Ratio of PRs = 0.96 (0.81, 1.13)**

| | | At least 25 y | | | | | | Less than 25 y | | | | | |
|---|---|---|---|---|---|---|---|---|---|---|---|---|---|
| Country | Trial | LNS N | Control N | Control Prevalence | PR (95% CI) | Fixed W | Random W | LNS N | Control N | Control Prevalence | PR (95% CI) | Fixed W | Random W |
| Bangladesh | JiVitA−4 (35) | | | | | | | | | | | | |
| Bangladesh | RDNS (36) | 152 | 81 | 40.7 | 0.78 (0.55, 1.09) | 0.12 | 0.13 | 398 | 191 | 53.9 | 0.68 (0.56, 0.81) | 0.41 | 0.19 |
| Bangladesh | WASH−B (37) | 79 | 80 | 26.2 | 0.58 (0.33, 1.02) | 0.05 | 0.05 | 132 | 98 | 35.7 | 0.17 (0.08, 0.34) | 0.03 | 0.13 |
| Burkina Faso | iLiNS−Zinc (38) | 178 | 50 | 61.1 | 0.49 (0.34, 0.71) | 0.10 | 0.10 | 137 | 46 | 61.5 | 0.54 (0.40, 0.75) | 0.14 | 0.18 |
| Burkina Faso | PROMIS CS (39) | | | | | | | | | | | | |
| Ghana | GHANA (40) | 62 | 55 | 63.6 | 0.53 (0.36, 0.79) | 0.09 | 0.09 | 22 | 21 | 61.9 | 0.66 (0.36, 1.21) | 0.04 | 0.14 |
| Ghana | iLiNS−DYADG (41) | | | | | | | | | | | | |
| Kenya | WASH−B (42) | 178 | 149 | 72.5 | 0.68 (0.58, 0.80) | 0.58 | 0.57 | 116 | 109 | 73.4 | 0.66 (0.53, 0.81) | 0.32 | 0.19 |
| Madagascar | MAHAY (43) | 47 | 25 | 56.0 | 0.76 (0.46, 1.26) | 0.06 | 0.06 | 36 | 26 | 61.5 | 0.90 (0.58, 1.40) | 0.07 | 0.16 |
| Malawi | iLiNS−DYADM (44) | | | | | | | | | | | | |
| Malawi | iLiNS−DOSE (45) | | | | | | | | | | | | |
| Mali | PROMIS CS (46) | | | | | | | | | | | | |
| Zimbabwe | SHINE (HIV−) (47) | | | | | | | | | | | | |
| Zimbabwe | SHINE (HIV+) (48) | | | | | | | | | | | | |
| | | **696** | **440** | | **I² = 0.01, Tau² = 0.00** | | | **841** | **491** | | **I² = 0.72, Tau² = 0.24** | | |
| **Fixed** | | | | | **0.65 (0.58, 0.74)** | | | | | | **0.64 (0.57, 0.72)** | | |
| **Random** | | | | | **0.65 (0.58, 0.74)** | | | | | | **0.56 (0.37, 0.87)** | | |

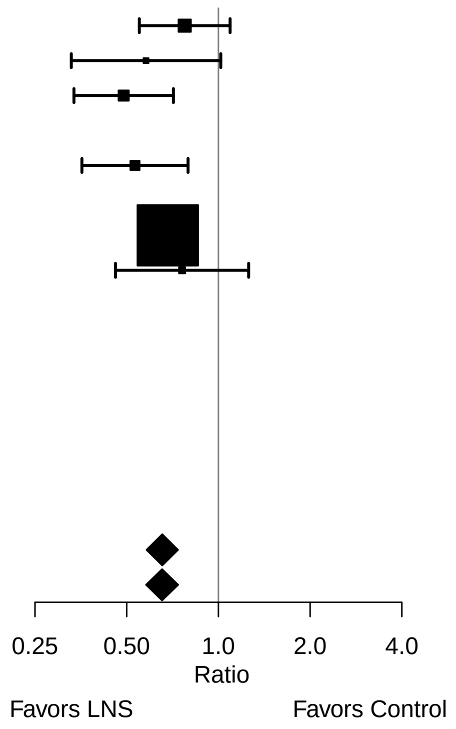

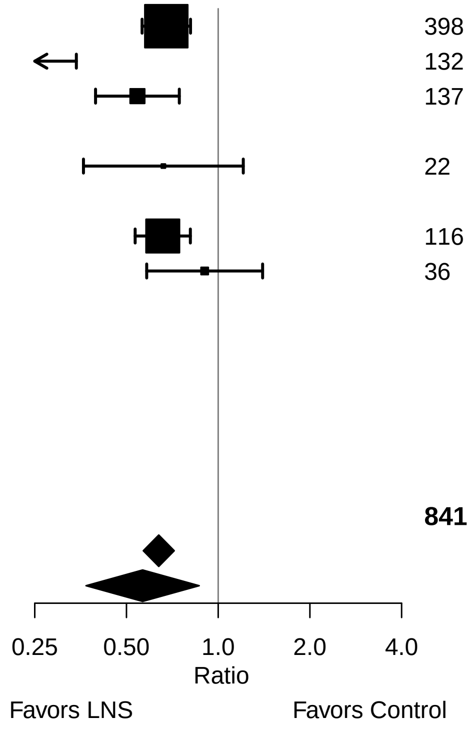

**Supplemental figure 6L: Elevated soluble transferrin receptor prevalence ratio**

#### 6L3: Stratified by Maternal education

**P−for−interaction = 0.227**
**Ratio of PRs = 1.12 (0.93, 1.35)**

| Country | Trial | LNS N | Control N | Control Prevalence | Primary or greater PR (95% CI) | Fixed W | Random W | LNS N | Control N | Control Prevalence | Incomplete or no formal PR (95% CI) | Fixed W | Random W |
|---|---|---|---|---|---|---|---|---|---|---|---|---|---|
| Bangladesh | JiVitA−4 (35) | | | | | | | | | | | | |
| Bangladesh | RDNS (36) | 406 | 195 | 47.7 | 0.71 (0.58, 0.86) | 0.45 | 0.33 | 144 | 77 | 55.8 | 0.70 (0.53, 0.91) | 0.28 | 0.28 |
| Bangladesh | WASH−B (37) | 149 | 139 | 33.1 | 0.28 (0.16, 0.50) | 0.05 | 0.20 | 63 | 39 | 25.6 | 0.43 (0.19, 1.00) | 0.03 | 0.03 |
| Burkina Faso | iLiNS−Zinc (38) | | | | | | | | | | | | |
| Burkina Faso | PROMIS CS (39) | | | | | | | | | | | | |
| Ghana | GHANA (40) | | | | | | | | | | | | |
| Ghana | iLiNS−DYADG (41) | | | | | | | | | | | | |
| Kenya | WASH−B (42) | 143 | 117 | 77.8 | 0.59 (0.49, 0.72) | 0.47 | 0.33 | 154 | 142 | 69.0 | 0.74 (0.61, 0.90) | 0.53 | 0.53 |
| Madagascar | MAHAY (43) | 16 | 16 | 50.0 | 0.63 (0.28, 1.38) | 0.03 | 0.14 | 67 | 35 | 62.9 | 0.83 (0.59, 1.17) | 0.17 | 0.17 |
| Malawi | iLiNS−DYADM (44) | | | | | | | | | | | | |
| Malawi | iLiNS−DOSE (45) | | | | | | | | | | | | |
| Mali | PROMIS CS (46) | | | | | | | | | | | | |
| Zimbabwe | SHINE (HIV−) (47) | | | | | | | | | | | | |
| Zimbabwe | SHINE (HIV+) (48) | | | | | | | | | | | | |
| | | **714** | **467** | | **I² = 0.67, Tau² = 0.10** | | | **428** | **293** | | **I² = 0.00, Tau² = 0.00** | | |
| **Fixed** | | | | | **0.62 (0.54, 0.70)** | | | | | | **0.73 (0.64, 0.84)** | | |
| **Random** | | | | | **0.55 (0.37, 0.80)** | | | | | | **0.73 (0.64, 0.84)** | | |

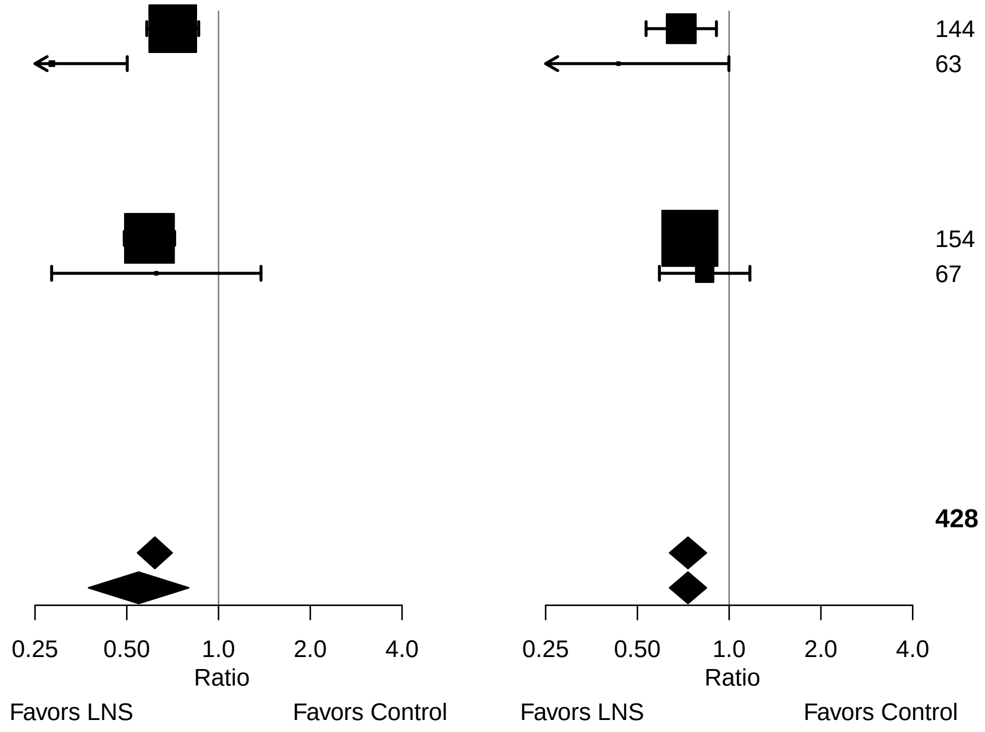

**Supplemental figure 6L: Elevated soluble transferrin receptor prevalence ratio**

#### 6L4: Stratified by Child sex

**P−for−interaction = 0.787**
**Ratio of PRs = 0.98 (0.83, 1.16)**

| Country | Trial | Male: LNS N | Male: Control N | Male: Control Prevalence | Male: PR (95% CI) | Male: Fixed W | Male: Random W | Female: LNS N | Female: Control N | Female: Control Prevalence | Female: PR (95% CI) | Female: Fixed W | Female: Random W |
|---|---|---|---|---|---|---|---|---|---|---|---|---|---|
| Bangladesh | JiVitA−4 (35) | | | | | | | | | | | | |
| Bangladesh | RDNS (36) | 266 | 131 | 58.0 | 0.70 (0.56, 0.87) | 0.28 | 0.21 | 281 | 141 | 42.6 | 0.70 (0.54, 0.91) | 0.23 | 0.22 |
| Bangladesh | WASH−B (37) | 105 | 85 | 37.6 | 0.30 (0.17, 0.54) | 0.04 | 0.10 | 107 | 93 | 25.8 | 0.33 (0.17, 0.61) | 0.04 | 0.10 |
| Burkina Faso | iLiNS−Zinc (38) | 150 | 49 | 66.7 | 0.51 (0.33, 0.78) | 0.07 | 0.14 | 165 | 47 | 55.8 | 0.53 (0.40, 0.70) | 0.19 | 0.21 |
| Burkina Faso | PROMIS CS (39) | | | | | | | | | | | | |
| Ghana | GHANA (40) | 52 | 37 | 73.0 | 0.58 (0.40, 0.84) | 0.10 | 0.16 | 32 | 45 | 53.3 | 0.47 (0.24, 0.91) | 0.04 | 0.09 |
| Ghana | iLiNS−DYADG (41) | | | | | | | | | | | | |
| Kenya | WASH−B (42) | 146 | 125 | 73.6 | 0.70 (0.58, 0.84) | 0.41 | 0.23 | 152 | 134 | 72.4 | 0.65 (0.53, 0.78) | 0.43 | 0.25 |
| Madagascar | MAHAY (43) | 34 | 22 | 72.7 | 0.77 (0.53, 1.12) | 0.09 | 0.16 | 49 | 29 | 48.3 | 0.89 (0.56, 1.41) | 0.07 | 0.14 |
| Malawi | iLiNS−DYADM (44) | | | | | | | | | | | | |
| Malawi | iLiNS−DOSE (45) | | | | | | | | | | | | |
| Mali | PROMIS CS (46) | | | | | | | | | | | | |
| Zimbabwe | SHINE (HIV−) (47) | | | | | | | | | | | | |
| Zimbabwe | SHINE (HIV+) (48) | | | | | | | | | | | | |
| | | **753** | **449** | | **$I^2$ = 0.51, $Tau^2$ = 0.05** | | | **786** | **489** | | **$I^2$ = 0.47, $Tau^2$ = 0.05** | | |
| **Fixed** | | | | | **0.65 (0.58, 0.73)** | | | | | | **0.62 (0.55, 0.71)** | | |
| **Random** | | | | | **0.60 (0.48, 0.76)** | | | | | | **0.60 (0.47, 0.76)** | | |

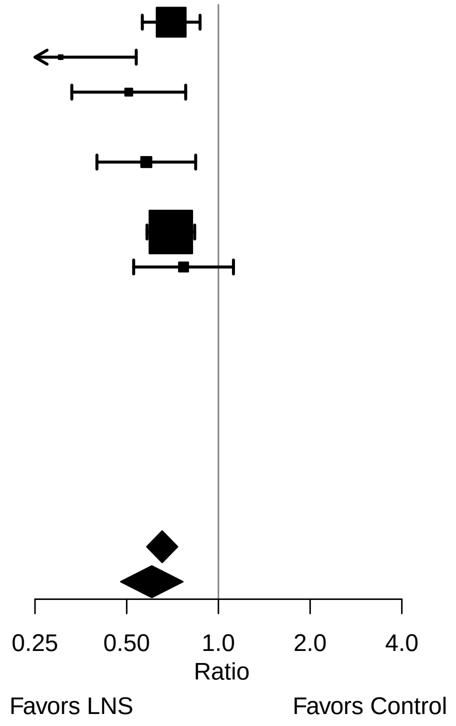

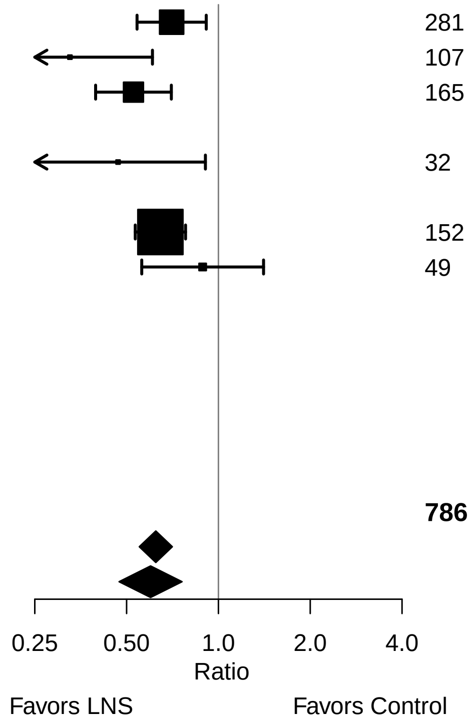

#### Supplemental figure 6L: Elevated soluble transferrin receptor prevalence ratio

##### 6L5: Stratified by Child birth order

**P−for−interaction = 0.638**
**Ratio of PRs = 0.96 (0.80, 1.15)**

| Country | Trial | Later born LNS N | Later born Control N | Later born Control Prevalence | Later born PR (95% CI) | Later born Fixed W | Later born Random W | Firstborn LNS N | Firstborn Control N | Firstborn Control Prevalence | Firstborn PR (95% CI) | Firstborn Fixed W | Firstborn Random W |
|---|---|---|---|---|---|---|---|---|---|---|---|---|---|
| Bangladesh | JiVitA−4 (35) | | | | | | | | | | | | |
| Bangladesh | RDNS (36) | 340 | 171 | 50.9 | 0.67 (0.55, 0.82) | 0.28 | 0.28 | 210 | 101 | 48.5 | 0.76 (0.59, 0.97) | 0.35 | 0.23 |
| Bangladesh | WASH−B (37) | 124 | 119 | 30.3 | 0.40 (0.25, 0.65) | 0.05 | 0.11 | 86 | 57 | 33.3 | 0.17 (0.07, 0.43) | 0.03 | 0.13 |
| Burkina Faso | iLiNS−Zinc (38) | 249 | 71 | 54.7 | 0.56 (0.38, 0.82) | 0.08 | 0.15 | 66 | 25 | 77.4 | 0.43 (0.32, 0.57) | 0.27 | 0.23 |
| Burkina Faso | PROMIS CS (39) | | | | | | | | | | | | |
| Ghana | GHANA (40) | 54 | 50 | 62.0 | 0.48 (0.30, 0.76) | 0.05 | 0.12 | 30 | 29 | 62.1 | 0.75 (0.47, 1.21) | 0.10 | 0.19 |
| Ghana | iLiNS−DYADG (41) | | | | | | | | | | | | |
| Kenya | WASH−B (42) | 237 | 213 | 72.8 | 0.70 (0.60, 0.81) | 0.55 | 0.34 | 60 | 46 | 73.9 | 0.56 (0.42, 0.75) | 0.26 | 0.22 |
| Madagascar | MAHAY (43) | | | | | | | | | | | | |
| Malawi | iLiNS−DYADM (44) | | | | | | | | | | | | |
| Malawi | iLiNS−DOSE (45) | | | | | | | | | | | | |
| Mali | PROMIS CS (46) | | | | | | | | | | | | |
| Zimbabwe | SHINE (HIV−) (47) | | | | | | | | | | | | |
| Zimbabwe | SHINE (HIV+) (48) | | | | | | | | | | | | |
| | | **1004** | **624** | | **$I^2$ = 0.43, $Tau^2$ = 0.02** | | | **452** | **258** | | **$I^2$ = 0.76, $Tau^2$ = 0.22** | | |
| **Fixed** | | | | | **0.65 (0.58, 0.72)** | | | | | | **0.58 (0.50, 0.67)** | | |
| **Random** | | | | | **0.60 (0.50, 0.72)** | | | | | | **0.52 (0.33, 0.82)** | | |

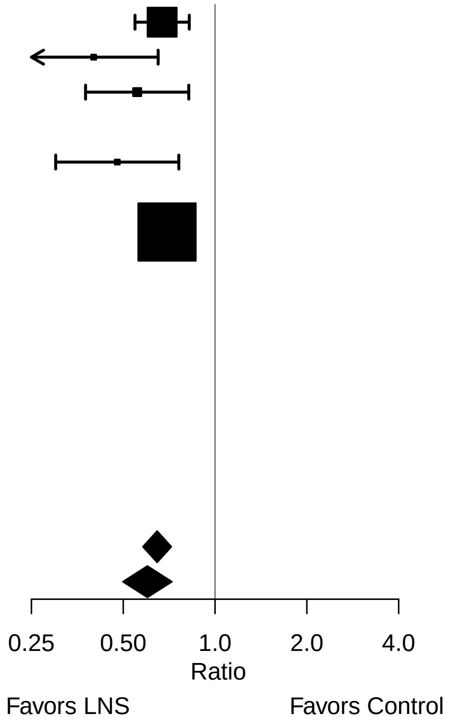

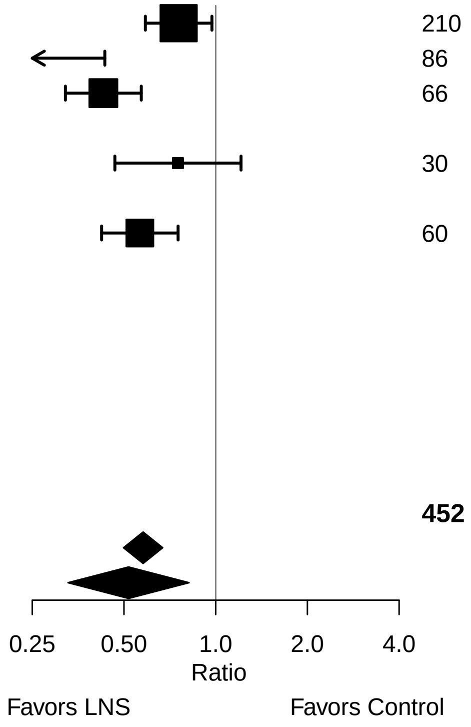

**Supplemental figure 6L: Elevated soluble transferrin receptor prevalence ratio**

#### 6L6: Stratified by Child baseline acute malnutrition (insufficient comparisons)

**Supplemental figure 6L: Elevated soluble transferrin receptor prevalence ratio**

#### 6L7: Stratified by Child baseline anemia (insufficient comparisons)

**Supplemental figure 6L: Elevated soluble transferrin receptor prevalence ratio**

#### 6L8: Stratified by Child high-dose vitamin A supplementation

**P−for−interaction = 0.344**
**Ratio of PRs = 1.14 (0.87, 1.51)**

| Country | Trial | Received VitA: LNS N | Control N | Control Prevalence | PR (95% CI) | Fixed W | Random W | Not treated: LNS N | Control N | Control Prevalence | PR (95% CI) | Fixed W | Random W |
|---|---|---|---|---|---|---|---|---|---|---|---|---|---|
| Bangladesh | JiVitA−4 (35) | | | | | | | | | | | | |
| Bangladesh | RDNS (36) | 444 | 223 | 50.7 | 0.69 (0.57, 0.84) | 0.76 | 0.69 | 105 | 49 | 46.9 | 0.73 (0.52, 1.02) | 0.14 | 0.14 |
| Bangladesh | WASH−B (37) | | | | | | | | | | | | |
| Burkina Faso | iLiNS−Zinc (38) | | | | | | | | | | | | |
| Burkina Faso | PROMIS CS (39) | | | | | | | | | | | | |
| Ghana | GHANA (40) | 40 | 53 | 66.0 | 0.61 (0.40, 0.93) | 0.16 | 0.20 | 44 | 26 | 53.8 | 0.59 (0.34, 1.03) | 0.05 | 0.05 |
| Ghana | iLiNS−DYADG (41) | | | | | | | | | | | | |
| Kenya | WASH−B (42) | 26 | 31 | 71.0 | 0.43 (0.24, 0.79) | 0.08 | 0.11 | 223 | 180 | 75.0 | 0.72 (0.63, 0.82) | 0.81 | 0.81 |
| Madagascar | MAHAY (43) | | | | | | | | | | | | |
| Malawi | iLiNS−DYADM (44) | | | | | | | | | | | | |
| Malawi | iLiNS−DOSE (45) | | | | | | | | | | | | |
| Mali | PROMIS CS (46) | | | | | | | | | | | | |
| Zimbabwe | SHINE (HIV−) (47) | | | | | | | | | | | | |
| Zimbabwe | SHINE (HIV+) (48) | | | | | | | | | | | | |
| | | **510** | **307** | | **I² = 0.12, Tau² = 0.01** | | | **372** | **255** | | **I² = 0.00, Tau² = 0.00** | | |
| **Fixed** | | | | | **0.65 (0.55, 0.77)** | | | | | | **0.71 (0.63, 0.81)** | | |
| **Random** | | | | | **0.64 (0.53, 0.79)** | | | | | | **0.71 (0.63, 0.81)** | | |

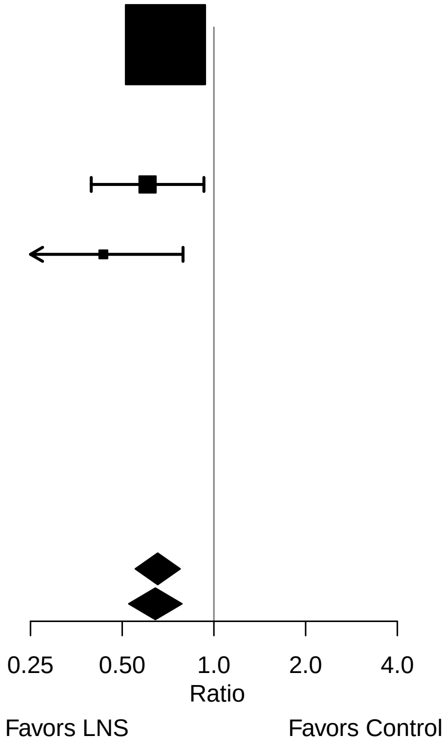

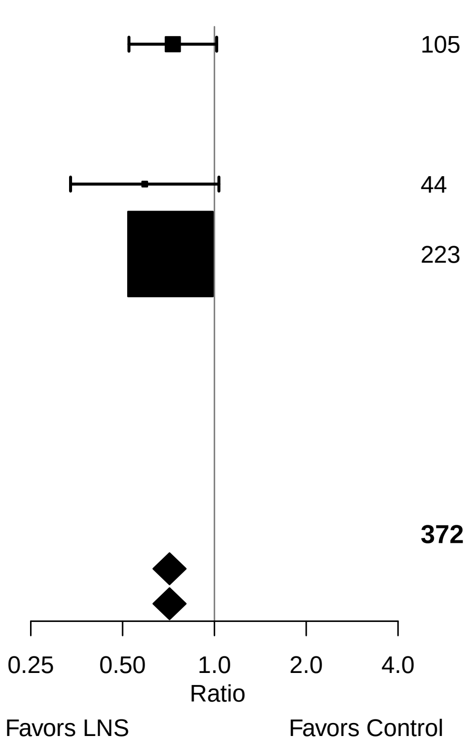

**Supplemental figure 6L: Elevated soluble transferrin receptor prevalence ratio**

#### 6L9: Stratified by Child inflammation

**P−for−interaction = 0.761**
**Ratio of PRs = 0.97 (0.82, 1.16)**

| Country | Trial | LNS N | Control N | Control Prevalence | Not inflamed PR (95% CI) | Fixed W | Random W | LNS N | Control N | Control Prevalence | High AGP or CRP PR (95% CI) | Fixed W | Random W |
|---|---|---|---|---|---|---|---|---|---|---|---|---|---|
| Bangladesh | JiVitA−4 (35) | | | | | | | | | | | | |
| Bangladesh | RDNS (36) | 370 | 168 | 43.5 | 0.77 (0.61, 0.97) | 0.28 | 0.19 | 180 | 104 | 60.6 | 0.64 (0.51, 0.82) | 0.29 | 0.29 |
| Bangladesh | WASH−B (37) | 165 | 136 | 34.6 | 0.28 (0.18, 0.45) | 0.07 | 0.15 | 47 | 42 | 21.4 | 0.50 (0.21, 1.17) | 0.02 | 0.02 |
| Burkina Faso | iLiNS−Zinc (38) | 124 | 31 | 58.8 | 0.42 (0.25, 0.68) | 0.06 | 0.14 | 191 | 65 | 62.5 | 0.58 (0.43, 0.77) | 0.19 | 0.19 |
| Burkina Faso | PROMIS CS (39) | | | | | | | | | | | | |
| Ghana | GHANA (40) | 70 | 66 | 62.1 | 0.53 (0.36, 0.78) | 0.10 | 0.16 | 14 | 16 | 62.5 | 0.80 (0.42, 1.53) | 0.04 | 0.04 |
| Ghana | iLiNS−DYADG (41) | | | | | | | | | | | | |
| Kenya | WASH−B (42) | 157 | 135 | 68.9 | 0.70 (0.57, 0.86) | 0.36 | 0.19 | 141 | 124 | 77.4 | 0.64 (0.52, 0.79) | 0.39 | 0.39 |
| Madagascar | MAHAY (43) | 44 | 32 | 53.1 | 0.98 (0.70, 1.39) | 0.13 | 0.17 | 39 | 19 | 68.4 | 0.64 (0.38, 1.06) | 0.06 | 0.06 |
| Malawi | iLiNS−DYADM (44) | | | | | | | | | | | | |
| Malawi | iLiNS−DOSE (45) | | | | | | | | | | | | |
| Mali | PROMIS CS (46) | | | | | | | | | | | | |
| Zimbabwe | SHINE (HIV−) (47) | | | | | | | | | | | | |
| Zimbabwe | SHINE (HIV+) (48) | | | | | | | | | | | | |
| | | **930** | **568** | | **I² = 0.79, Tau² = 0.16** | | | **612** | **370** | | **I² = 0.00, Tau² = 0.00** | | |
| **Fixed** | | | | | **0.66 (0.59, 0.75)** | | | | | | **0.63 (0.55, 0.72)** | | |
| **Random** | | | | | **0.58 (0.41, 0.83)** | | | | | | **0.63 (0.55, 0.72)** | | |

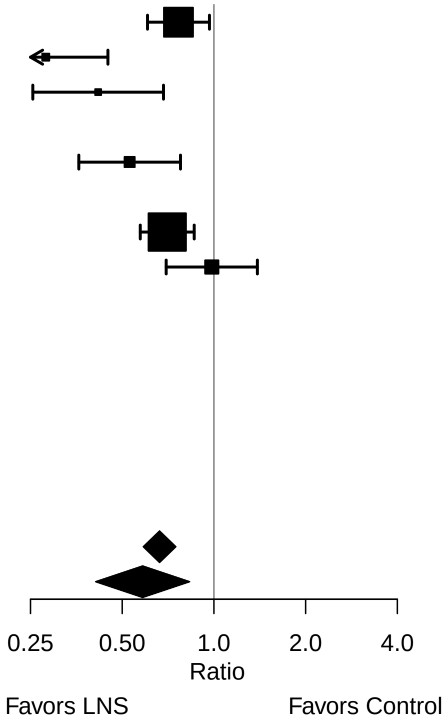

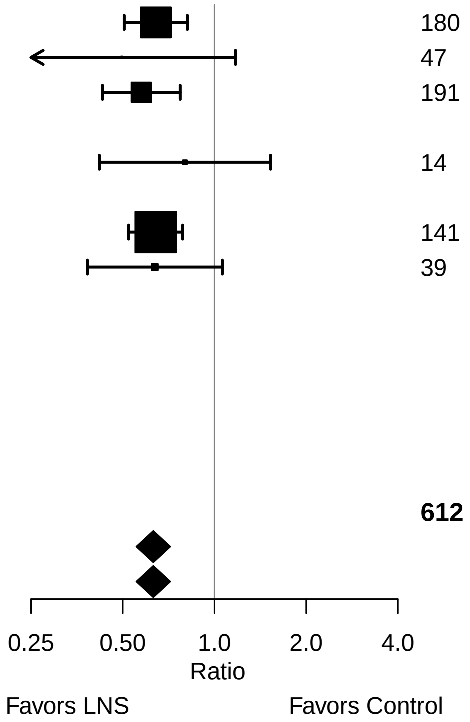

### Supplemental figure 6M: Elevated soluble transferrin receptor prevalence difference

#### 6M1: Stratified by Maternal BMI

**P−for−interaction = 0.187**
**Difference in PDs = −0.07 (−0.17, 0.03)**

| | | At least 20 kg/m² | | | | | | Less than 20 kg/m² | | | | | |
|---|---|---|---|---|---|---|---|---|---|---|---|---|---|
| Country | Trial | LNS N | Control N | Control Prevalence | PD (95% CI) | Fixed W | Random W | LNS N | Control N | Control Prevalence | PD (95% CI) | Fixed W | Random W |
| Bangladesh | JiVitA−4 (35) | | | | | | | | | | | | |
| Bangladesh | RDNS (36) | 228 | 102 | 44.1 | −0.09 (−0.20, 0.03) | 0.35 | 0.35 | 300 | 160 | 51.2 | −0.16 (−0.25, −0.07) | 0.64 | 0.50 |
| Bangladesh | WASH−B (37) | | | | | | | | | | | | |
| Burkina Faso | iLiNS−Zinc (38) | 209 | 55 | 60.7 | −0.27 (−0.42, −0.12) | 0.20 | 0.27 | 106 | 41 | 62.2 | −0.35 (−0.54, −0.16) | 0.15 | 0.23 |
| Burkina Faso | PROMIS CS (39) | | | | | | | | | | | | |
| Ghana | GHANA (40) | | | | | | | | | | | | |
| Ghana | iLiNS−DYADG (41) | | | | | | | | | | | | |
| Kenya | WASH−B (42) | 223 | 200 | 72.0 | −0.22 (−0.32, −0.12) | 0.45 | 0.38 | 64 | 51 | 76.5 | −0.26 (−0.43, −0.10) | 0.20 | 0.28 |
| Madagascar | MAHAY (43) | | | | | | | | | | | | |
| Malawi | iLiNS−DYADM (44) | | | | | | | | | | | | |
| Malawi | iLiNS−DOSE (45) | | | | | | | | | | | | |
| Mali | PROMIS CS (46) | | | | | | | | | | | | |
| Zimbabwe | SHINE (HIV−) (47) | | | | | | | | | | | | |
| Zimbabwe | SHINE (HIV+) (48) | | | | | | | | | | | | |
| | | **660** | **357** | | **I² = 0.55, Tau² = 0.00** | | | **470** | **252** | | **I² = 0.44, Tau² = 0.00** | | |
| **Fixed** | | | | | **−0.18 (−0.25, −0.11)** | | | | | | **−0.21 (−0.29, −0.14)** | | |
| **Random** | | | | | **−0.19 (−0.29, −0.08)** | | | | | | **−0.23 (−0.34, −0.13)** | | |

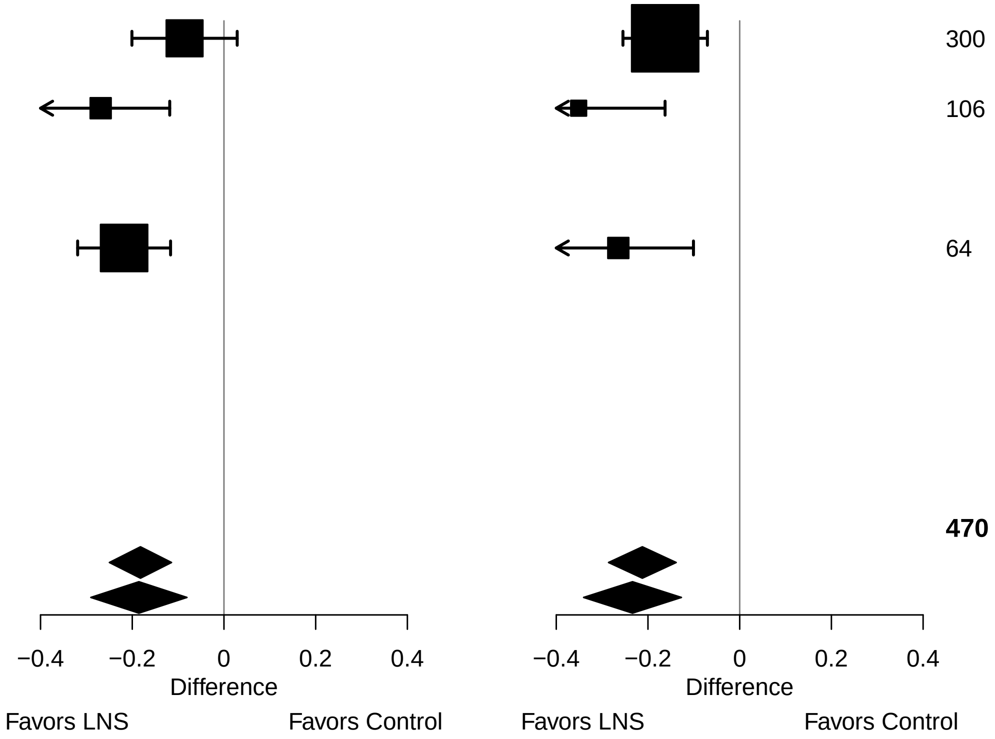

#### Supplemental figure 6M: Elevated soluble transferrin receptor prevalence difference

##### 6M2: Stratified by Maternal age

**P−for−interaction = 0.142**
**Difference in PDs = −0.06 (−0.14, 0.02)**

| | | At least 25 y | | | | | | Less than 25 y | | | | | |
|---|---|---|---|---|---|---|---|---|---|---|---|---|---|
| | | **LNS** | **Control** | **Control** | **PD** | **Fixed** | **Random** | **LNS** | **Control** | **Control** | **PD** | **Fixed** | **Random** |
| Country | Trial | N | N | Prevalence | (95% CI) | W | W | N | N | Prevalence | (95% CI) | W | W |
| Bangladesh | JiVitA−4 (35) | | | | | | | | | | | | |
| Bangladesh | RDNS (36) | 152 | 81 | 40.7 | −0.09 (−0.22, 0.04) | 0.17 | 0.18 | 398 | 191 | 53.9 | −0.17 (−0.26, −0.09) | 0.42 | 0.39 |
| Bangladesh | WASH−B (37) | 79 | 80 | 26.2 | −0.11 (−0.22, 0.00) | 0.23 | 0.21 | 132 | 98 | 35.7 | −0.30 (−0.41, −0.19) | 0.23 | 0.23 |
| Burkina Faso | iLiNS−Zinc (38) | 178 | 50 | 61.1 | −0.31 (−0.45, −0.16) | 0.14 | 0.16 | 137 | 46 | 61.5 | −0.28 (−0.46, −0.11) | 0.09 | 0.10 |
| Burkina Faso | PROMIS CS (39) | | | | | | | | | | | | |
| Ghana | GHANA (40) | 62 | 55 | 63.6 | −0.30 (−0.47, −0.12) | 0.09 | 0.13 | 22 | 21 | 61.9 | −0.21 (−0.51, 0.09) | 0.03 | 0.03 |
| Ghana | iLiNS−DYADG (41) | | | | | | | | | | | | |
| Kenya | WASH−B (42) | 178 | 149 | 72.5 | −0.23 (−0.32, −0.14) | 0.33 | 0.25 | 116 | 109 | 73.4 | −0.25 (−0.37, −0.13) | 0.19 | 0.20 |
| Madagascar | MAHAY (43) | 47 | 25 | 56.0 | −0.13 (−0.38, 0.11) | 0.05 | 0.08 | 36 | 26 | 61.5 | −0.06 (−0.32, 0.20) | 0.04 | 0.05 |
| Malawi | iLiNS−DYADM (44) | | | | | | | | | | | | |
| Malawi | iLiNS−DOSE (45) | | | | | | | | | | | | |
| Mali | PROMIS CS (46) | | | | | | | | | | | | |
| Zimbabwe | SHINE (HIV−) (47) | | | | | | | | | | | | |
| Zimbabwe | SHINE (HIV+) (48) | | | | | | | | | | | | |
| | | **696** | **440** | | **I² = 0.45, Tau² = 0.00** | | | **841** | **491** | | **I² = 0.07, Tau² = 0.00** | | |
| **Fixed** | | | | | **−0.19 (−0.25, −0.14)** | | | | | | **−0.22 (−0.28, −0.17)** | | |
| **Random** | | | | | **−0.19 (−0.27, −0.12)** | | | | | | **−0.22 (−0.28, −0.17)** | | |

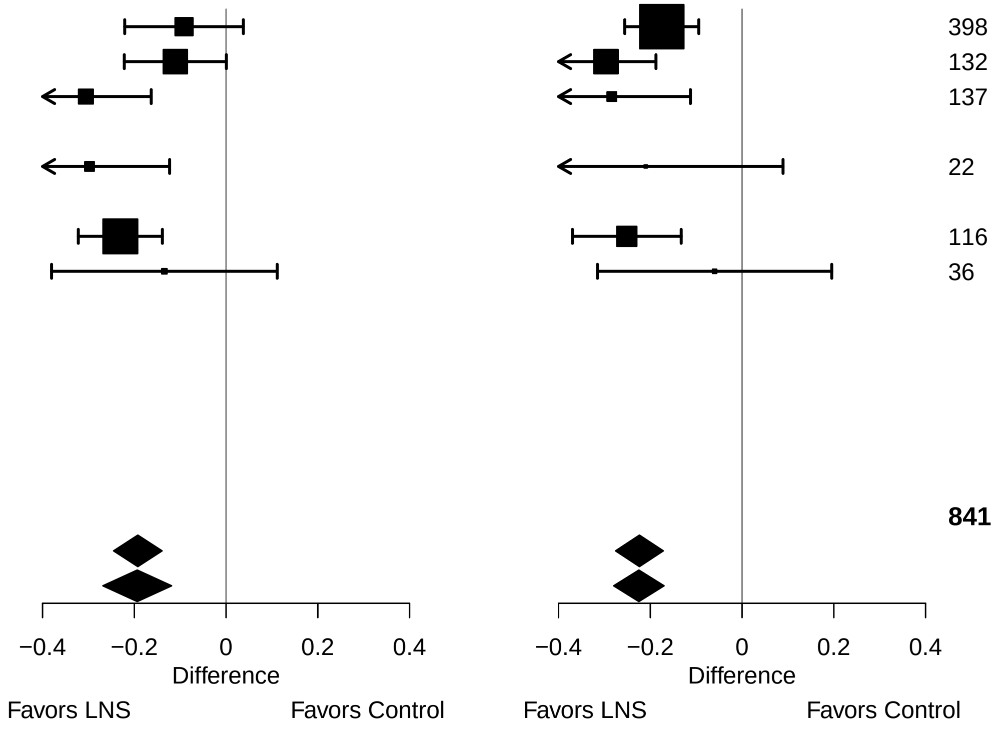

**Supplemental figure 6M: Elevated soluble transferrin receptor prevalence difference**

#### 6M3: Stratified by Maternal education

**P−for−interaction = 0.157**
**Difference in PDs = 0.07 (−0.03, 0.16)**

| | | Primary or greater | | | | | | Incomplete or no formal | | | | | |
|---|---|---|---|---|---|---|---|---|---|---|---|---|---|
| Country | Trial | LNS N | Control N | Control Prevalence | PD (95% CI) | Fixed W | Random W | LNS N | Control N | Control Prevalence | PD (95% CI) | Fixed W | Random W |
| Bangladesh | JiVitA−4 (35) | | | | | | | | | | | | |
| Bangladesh | RDNS (36) | 406 | 195 | 47.7 | −0.14 (−0.22, −0.06) | 0.43 | 0.36 | 144 | 77 | 55.8 | −0.17 (−0.29, −0.05) | 0.31 | 0.31 |
| Bangladesh | WASH−B (37) | 149 | 139 | 33.1 | −0.24 (−0.33, −0.14) | 0.32 | 0.32 | 63 | 39 | 25.6 | −0.15 (−0.30, 0.01) | 0.18 | 0.18 |
| Burkina Faso | iLiNS−Zinc (38) | | | | | | | | | | | | |
| Burkina Faso | PROMIS CS (39) | | | | | | | | | | | | |
| Ghana | GHANA (40) | | | | | | | | | | | | |
| Ghana | iLiNS−DYADG (41) | | | | | | | | | | | | |
| Kenya | WASH−B (42) | 143 | 117 | 77.8 | −0.32 (−0.43, −0.20) | 0.22 | 0.26 | 154 | 142 | 69.0 | −0.18 (−0.29, −0.07) | 0.39 | 0.39 |
| Madagascar | MAHAY (43) | 16 | 16 | 50.0 | −0.19 (−0.48, 0.11) | 0.03 | 0.06 | 67 | 35 | 62.9 | −0.11 (−0.31, 0.09) | 0.12 | 0.12 |
| Malawi | iLiNS−DYADM (44) | | | | | | | | | | | | |
| Malawi | iLiNS−DOSE (45) | | | | | | | | | | | | |
| Mali | PROMIS CS (46) | | | | | | | | | | | | |
| Zimbabwe | SHINE (HIV−) (47) | | | | | | | | | | | | |
| Zimbabwe | SHINE (HIV+) (48) | | | | | | | | | | | | |
| | | **714** | **467** | | **I² = 0.54, Tau² = 0.00** | | | **428** | **293** | | **I² = 0.00, Tau² = 0.00** | | |
| **Fixed** | | | | | **−0.21 (−0.26, −0.16)** | | | | | | **−0.16 (−0.23, −0.09)** | | |
| **Random** | | | | | **−0.22 (−0.30, −0.14)** | | | | | | **−0.16 (−0.23, −0.09)** | | |

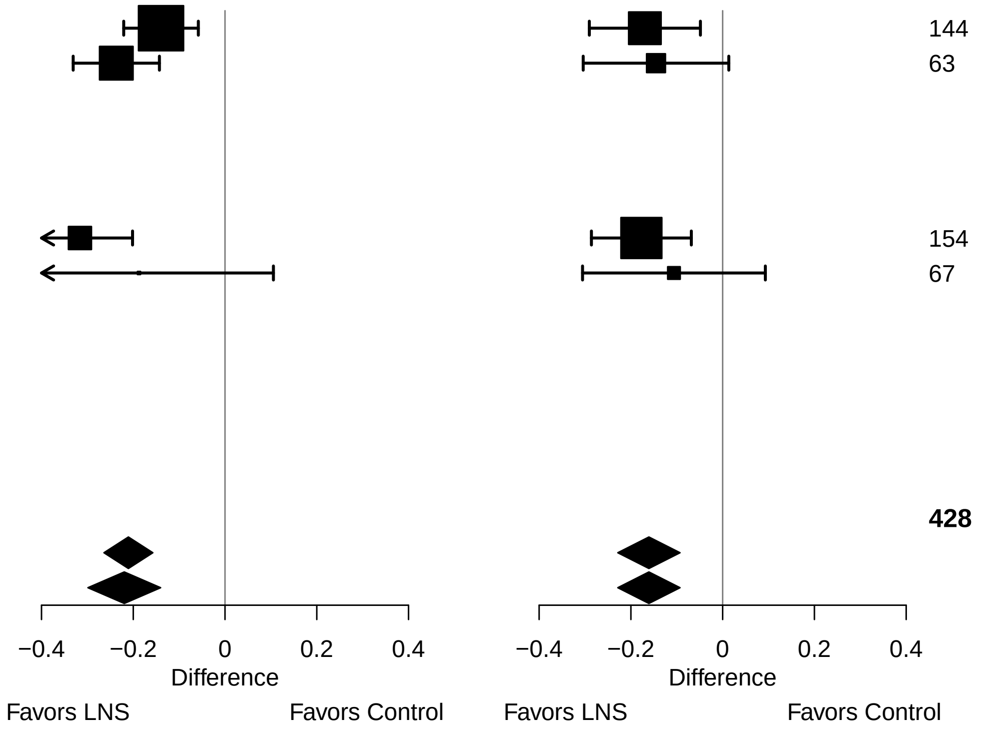

**Supplemental figure 6M: Elevated soluble transferrin receptor prevalence difference**

#### 6M4: Stratified by Child sex

**P−for−interaction = 0.322**
**Difference in PDs = 0.04 (−0.04, 0.11)**

| | | **Male** | | | | | | **Female** | | | | | |
|---|---|---|---|---|---|---|---|---|---|---|---|---|---|
| | | **LNS** | **Control** | **Control** | **PD** | **Fixed** | **Random** | **LNS** | **Control** | **Control** | **PD** | **Fixed** | **Random** |
| Country | Trial | N | N | Prevalence | (95% CI) | W | W | N | N | Prevalence | (95% CI) | W | W |
| Bangladesh | JiVitA−4 (35) | | | | | | | | | | | | |
| Bangladesh | RDNS (36) | 266 | 131 | 58.0 | −0.17 (−0.28, −0.07) | 0.29 | 0.29 | 281 | 141 | 42.6 | −0.13 (−0.22, −0.03) | 0.26 | 0.23 |
| Bangladesh | WASH−B (37) | 105 | 85 | 37.6 | −0.26 (−0.39, −0.14) | 0.22 | 0.22 | 107 | 93 | 25.8 | −0.17 (−0.28, −0.07) | 0.22 | 0.21 |
| Burkina Faso | iLiNS−Zinc (38) | 150 | 49 | 66.7 | −0.32 (−0.53, −0.10) | 0.07 | 0.07 | 165 | 47 | 55.8 | −0.28 (−0.39, −0.16) | 0.18 | 0.19 |
| Burkina Faso | PROMIS CS (39) | | | | | | | | | | | | |
| Ghana | GHANA (40) | 52 | 37 | 73.0 | −0.31 (−0.51, −0.10) | 0.08 | 0.08 | 32 | 45 | 53.3 | −0.28 (−0.50, −0.07) | 0.05 | 0.07 |
| Ghana | iLiNS−DYADG (41) | | | | | | | | | | | | |
| Kenya | WASH−B (42) | 146 | 125 | 73.6 | −0.22 (−0.33, −0.11) | 0.28 | 0.28 | 152 | 134 | 72.4 | −0.26 (−0.36, −0.16) | 0.24 | 0.22 |
| Madagascar | MAHAY (43) | 34 | 22 | 72.7 | −0.17 (−0.40, 0.06) | 0.06 | 0.06 | 49 | 29 | 48.3 | −0.05 (−0.26, 0.15) | 0.05 | 0.08 |
| Malawi | iLiNS−DYADM (44) | | | | | | | | | | | | |
| Malawi | iLiNS−DOSE (45) | | | | | | | | | | | | |
| Mali | PROMIS CS (46) | | | | | | | | | | | | |
| Zimbabwe | SHINE (HIV−) (47) | | | | | | | | | | | | |
| Zimbabwe | SHINE (HIV+) (48) | | | | | | | | | | | | |
| | | **753** | **449** | | **I² = 0.00, Tau² = 0.00** | | | **786** | **489** | | **I² = 0.37, Tau² = 0.00** | | |
| **Fixed** | | | | | **−0.23 (−0.28, −0.17)** | | | | | | **−0.20 (−0.25, −0.15)** | | |
| **Random** | | | | | **−0.23 (−0.28, −0.17)** | | | | | | **−0.20 (−0.26, −0.14)** | | |

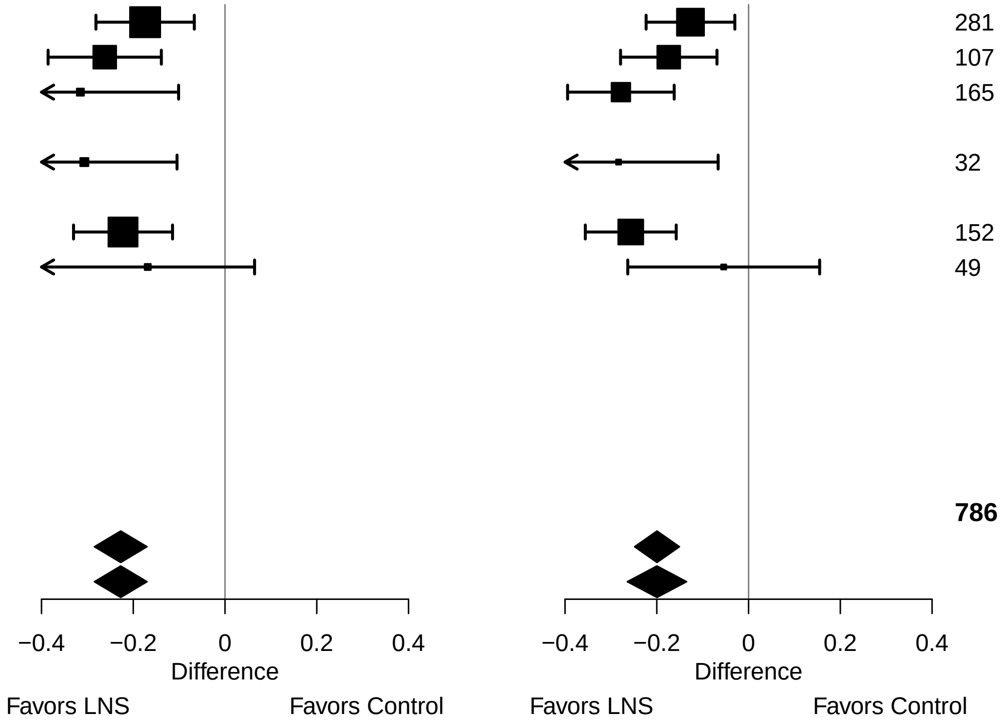

#### Supplemental figure 6M: Elevated soluble transferrin receptor prevalence difference

##### 6M5: Stratified by Child birth order

**P−for−interaction = 0.285**
**Difference in PDs = −0.05 (−0.13, 0.04)**

| Country | Trial | Later born LNS N | Later born Control N | Later born Control Prevalence | Later born PD (95% CI) | Later born Fixed W | Later born Random W | Firstborn LNS N | Firstborn Control N | Firstborn Control Prevalence | Firstborn PD (95% CI) | Firstborn Fixed W | Firstborn Random W |
|---|---|---|---|---|---|---|---|---|---|---|---|---|---|
| Bangladesh | JiVitA−4 (35) | | | | | | | | | | | | |
| Bangladesh | RDNS (36) | 340 | 171 | 50.9 | −0.17 (−0.26, −0.08) | 0.27 | 0.27 | 210 | 101 | 48.5 | −0.12 (−0.22, −0.02) | 0.36 | 0.24 |
| Bangladesh | WASH−B (37) | 124 | 119 | 30.3 | −0.18 (−0.27, −0.10) | 0.29 | 0.29 | 86 | 57 | 33.3 | −0.28 (−0.42, −0.13) | 0.18 | 0.21 |
| Burkina Faso | iLiNS−Zinc (38) | 249 | 71 | 54.7 | −0.24 (−0.41, −0.07) | 0.07 | 0.07 | 66 | 25 | 77.4 | −0.46 (−0.58, −0.34) | 0.26 | 0.23 |
| Burkina Faso | PROMIS CS (39) | | | | | | | | | | | | |
| Ghana | GHANA (40) | 54 | 50 | 62.0 | −0.32 (−0.51, −0.14) | 0.06 | 0.06 | 30 | 29 | 62.1 | −0.15 (−0.41, 0.10) | 0.06 | 0.13 |
| Ghana | iLiNS−DYADG (41) | | | | | | | | | | | | |
| Kenya | WASH−B (42) | 237 | 213 | 72.8 | −0.22 (−0.31, −0.14) | 0.30 | 0.30 | 60 | 46 | 73.9 | −0.32 (−0.48, −0.16) | 0.15 | 0.20 |
| Madagascar | MAHAY (43) | | | | | | | | | | | | |
| Malawi | iLiNS−DYADM (44) | | | | | | | | | | | | |
| Malawi | iLiNS−DOSE (45) | | | | | | | | | | | | |
| Mali | PROMIS CS (46) | | | | | | | | | | | | |
| Zimbabwe | SHINE (HIV−) (47) | | | | | | | | | | | | |
| Zimbabwe | SHINE (HIV+) (48) | | | | | | | | | | | | |
| | | **1004** | **624** | | **I² = 0.00, Tau² = 0.00** | | | **452** | **258** | | **I² = 0.79, Tau² = 0.01** | | |
| **Fixed** | | | | | **−0.20 (−0.25, −0.16)** | | | | | | **−0.27 (−0.33, −0.20)** | | |
| **Random** | | | | | **−0.20 (−0.25, −0.16)** | | | | | | **−0.27 (−0.40, −0.15)** | | |

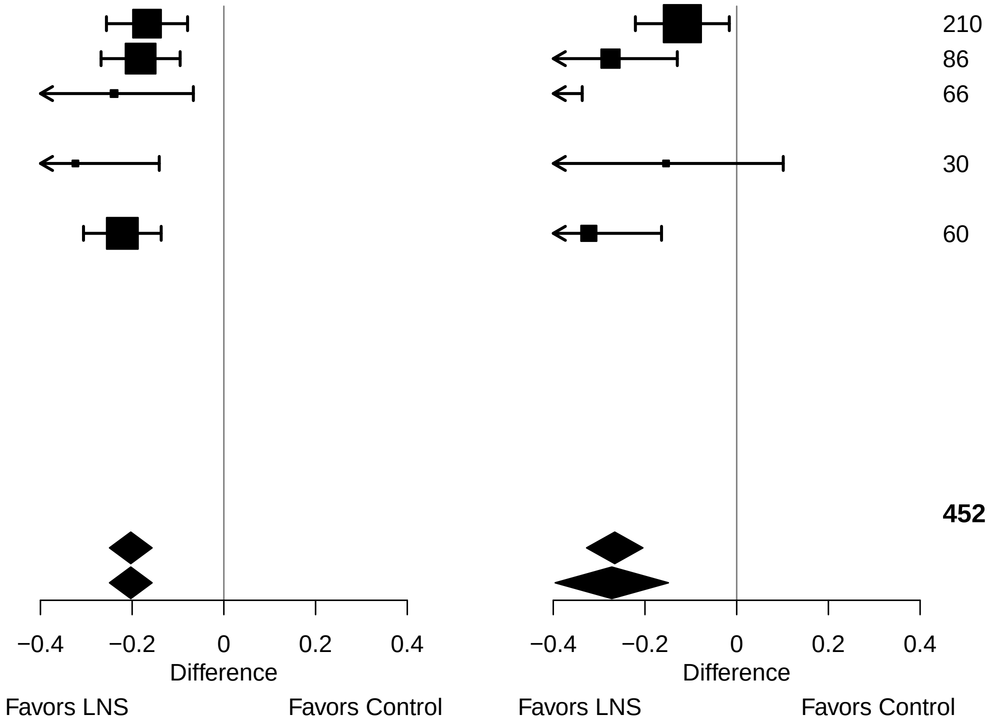

**Supplemental figure 6M: Elevated soluble transferrin receptor prevalence difference**

#### 6M6: Stratified by Child baseline acute malnutrition (insufficient comparisons)

**Supplemental figure 6M: Elevated soluble transferrin receptor prevalence difference**

#### 6M7: Stratified by Child baseline anemia (insufficient comparisons)

**Supplemental figure 6M: Elevated soluble transferrin receptor prevalence difference**

#### 6M8: Stratified by Child high-dose vitamin A supplementation

**P−for−interaction = 0.272**
**Difference in PDs = 0.08 (−0.06, 0.21)**

| | | Received VitA | | | | | | Not treated | | | | | |
|---|---|---|---|---|---|---|---|---|---|---|---|---|---|
| Country | Trial | LNS N | Control N | Control Prevalence | PD (95% CI) | Fixed W | Random W | LNS N | Control N | Control Prevalence | PD (95% CI) | Fixed W | Random W |
| Bangladesh | JiVitA−4 (35) | | | | | | | | | | | | |
| Bangladesh | RDNS (36) | 444 | 223 | 50.7 | −0.16 (−0.24, −0.07) | 0.77 | 0.52 | 105 | 49 | 46.9 | −0.13 (−0.26, 0.01) | 0.26 | 0.26 |
| Bangladesh | WASH−B (37) | | | | | | | | | | | | |
| Burkina Faso | iLiNS−Zinc (38) | | | | | | | | | | | | |
| Burkina Faso | PROMIS CS (39) | | | | | | | | | | | | |
| Ghana | GHANA (40) | 40 | 53 | 66.0 | −0.26 (−0.46, −0.06) | 0.13 | 0.27 | 44 | 26 | 53.8 | −0.22 (−0.46, 0.01) | 0.08 | 0.08 |
| Ghana | iLiNS−DYADG (41) | | | | | | | | | | | | |
| Kenya | WASH−B (42) | 26 | 31 | 71.0 | −0.40 (−0.64, −0.17) | 0.09 | 0.22 | 223 | 180 | 75.0 | −0.21 (−0.29, −0.13) | 0.66 | 0.66 |
| Madagascar | MAHAY (43) | | | | | | | | | | | | |
| Malawi | iLiNS−DYADM (44) | | | | | | | | | | | | |
| Malawi | iLiNS−DOSE (45) | | | | | | | | | | | | |
| Mali | PROMIS CS (46) | | | | | | | | | | | | |
| Zimbabwe | SHINE (HIV−) (47) | | | | | | | | | | | | |
| Zimbabwe | SHINE (HIV+) (48) | | | | | | | | | | | | |
| | | **510** | **307** | | **I² = 0.53, Tau² = 0.01** | | | **372** | **255** | | **I² = 0.00, Tau² = 0.00** | | |
| **Fixed** | | | | | **−0.19 (−0.27, −0.12)** | | | | | | **−0.19 (−0.26, −0.12)** | | |
| **Random** | | | | | **−0.24 (−0.37, −0.10)** | | | | | | **−0.19 (−0.26, −0.12)** | | |

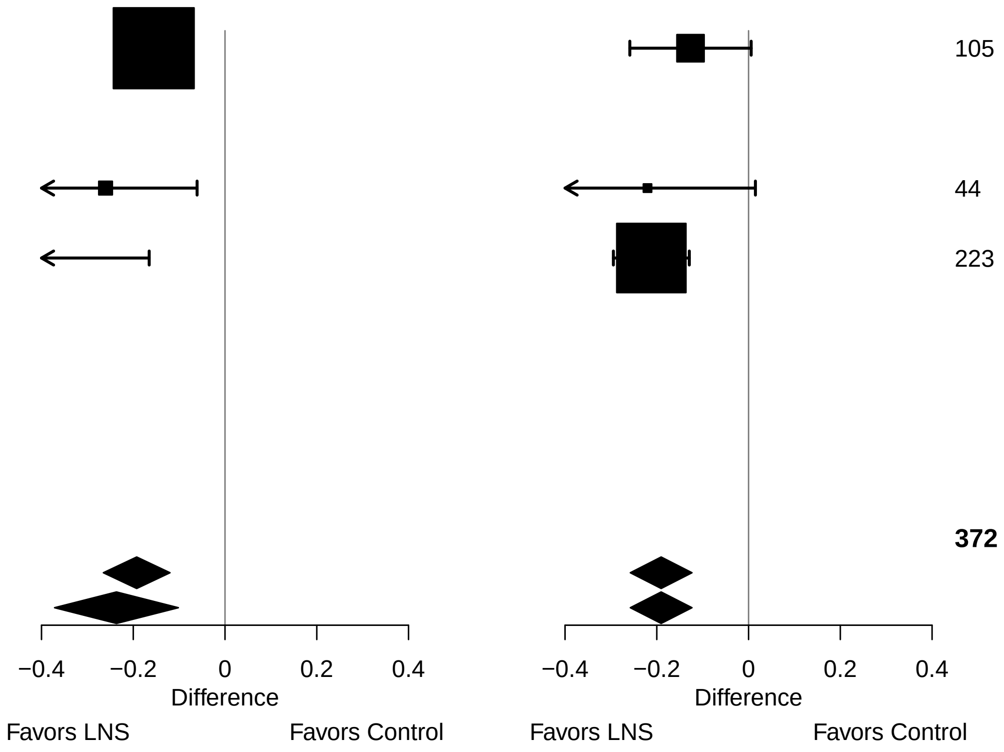

**Supplemental figure 6M: Elevated soluble transferrin receptor prevalence difference**

#### 6M9: Stratified by Child inflammation

**P−for−interaction = 0.599**
**Difference in PDs = −0.02 (−0.10, 0.06)**

| | | **Not inflamed** | | | | | | **High AGP or CRP** | | | | | |
|---|---|---|---|---|---|---|---|---|---|---|---|---|---|
| Country | Trial | **LNS** N | **Control** N | **Control** Prevalence | **PD** (95% CI) | **Fixed** W | **Random** W | **LNS** N | **Control** N | **Control** Prevalence | **PD** (95% CI) | **Fixed** W | **Random** W |
| Bangladesh | JiVitA−4 (35) | | | | | | | | | | | | |
| Bangladesh | RDNS (36) | 370 | 168 | 43.5 | −0.10 (−0.20, −0.01) | 0.28 | 0.21 | 180 | 104 | 60.6 | −0.22 (−0.32, −0.11) | 0.31 | 0.31 |
| Bangladesh | WASH−B (37) | 165 | 136 | 34.6 | −0.25 (−0.34, −0.16) | 0.31 | 0.21 | 47 | 42 | 21.4 | −0.11 (−0.23, 0.01) | 0.23 | 0.23 |
| Burkina Faso | iLiNS−Zinc (38) | 124 | 31 | 58.8 | −0.34 (−0.53, −0.14) | 0.07 | 0.12 | 191 | 65 | 62.5 | −0.27 (−0.42, −0.12) | 0.15 | 0.15 |
| Burkina Faso | PROMIS CS (39) | | | | | | | | | | | | |
| Ghana | GHANA (40) | 70 | 66 | 62.1 | −0.29 (−0.45, −0.13) | 0.09 | 0.14 | 14 | 16 | 62.5 | −0.12 (−0.49, 0.24) | 0.03 | 0.03 |
| Ghana | iLiNS−DYADG (41) | | | | | | | | | | | | |
| Kenya | WASH−B (42) | 157 | 135 | 68.9 | −0.20 (−0.32, −0.09) | 0.18 | 0.19 | 141 | 124 | 77.4 | −0.28 (−0.39, −0.16) | 0.25 | 0.25 |
| Madagascar | MAHAY (43) | 44 | 32 | 53.1 | −0.01 (−0.19, 0.17) | 0.07 | 0.13 | 39 | 19 | 68.4 | −0.25 (−0.53, 0.03) | 0.04 | 0.04 |
| Malawi | iLiNS−DYADM (44) | | | | | | | | | | | | |
| Malawi | iLiNS−DOSE (45) | | | | | | | | | | | | |
| Mali | PROMIS CS (46) | | | | | | | | | | | | |
| Zimbabwe | SHINE (HIV−) (47) | | | | | | | | | | | | |
| Zimbabwe | SHINE (HIV+) (48) | | | | | | | | | | | | |
| | | **930** | **568** | | **I² = 0.61, Tau² = 0.01** | | | **612** | **370** | | **I² = 0.00, Tau² = 0.00** | | |
| **Fixed** | | | | | **−0.19 (−0.24, −0.14)** | | | | | | **−0.21 (−0.27, −0.16)** | | |
| **Random** | | | | | **−0.20 (−0.29, −0.11)** | | | | | | **−0.21 (−0.27, −0.16)** | | |

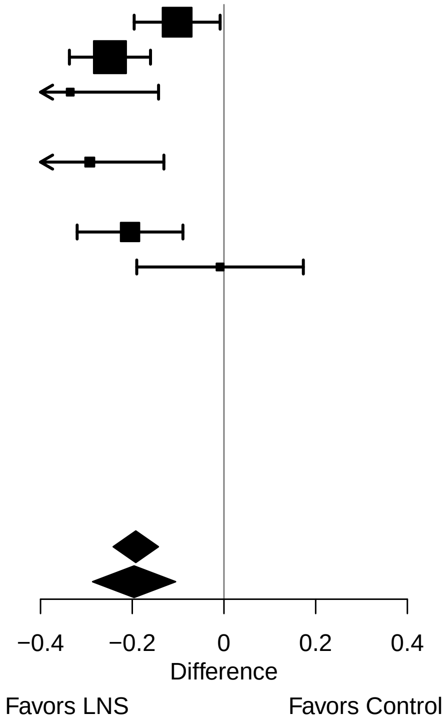

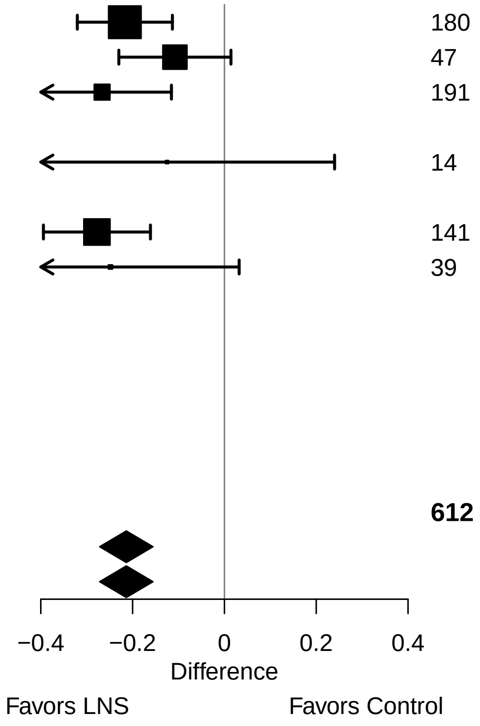

### Supplemental figure 6N: Geometric mean ratio of zinc protoporphyrin concentration

#### 6N1: Stratified by Maternal BMI

**P−for−interaction = 0.792**
**Ratio of GMRs = 0.98 (0.85, 1.13)**

| | | At least 20 $kg/m^2$ | | | | | | Less than 20 $kg/m^2$ | | | | | |
|---|---|---|---|---|---|---|---|---|---|---|---|---|---|
| Country | Trial | LNS N | Control N | Control Median | GMR (95% CI) | Fixed W | Random W | LNS N | Control N | Control Median | GMR (95% CI) | Fixed W | Random W |
| Bangladesh | JiVitA−4 (35) | | | | | | | | | | | | |
| Bangladesh | RDNS (36) | | | | | | | | | | | | |
| Bangladesh | WASH−B (37) | | | | | | | | | | | | |
| Burkina Faso | iLiNS−Zinc (38) | 208 | 55 | 123.3 | 0.63 (0.51, 0.77) | 0.14 | 0.21 | 104 | 40 | 111.5 | 0.65 (0.52, 0.82) | 0.23 | 0.26 |
| Burkina Faso | PROMIS CS (39) | | | | | | | | | | | | |
| Ghana | GHANA (40) | | | | | | | | | | | | |
| Ghana | iLiNS−DYADG (41) | 85 | 174 | 47.0 | 0.88 (0.76, 1.01) | 0.30 | 0.27 | 11 | 25 | 54.5 | 0.72 (0.50, 1.05) | 0.09 | 0.12 |
| Kenya | WASH−B (42) | | | | | | | | | | | | |
| Madagascar | MAHAY (43) | | | | | | | | | | | | |
| Malawi | iLiNS−DYADM (44) | 111 | 222 | 56.0 | 0.86 (0.75, 0.99) | 0.32 | 0.27 | 79 | 151 | 50.8 | 0.88 (0.75, 1.02) | 0.53 | 0.43 |
| Malawi | iLiNS−DOSE (45) | 127 | 65 | 67.0 | 0.78 (0.67, 0.91) | 0.24 | 0.25 | 53 | 22 | 55.8 | 0.75 (0.57, 0.99) | 0.15 | 0.19 |
| Mali | PROMIS CS (46) | | | | | | | | | | | | |
| Zimbabwe | SHINE (HIV−) (47) | | | | | | | | | | | | |
| Zimbabwe | SHINE (HIV+) (48) | | | | | | | | | | | | |
| | | **531** | **516** | | **$I^2$ = 0.64, $Tau^2$ = 0.02** | | | **247** | **238** | | **$I^2$ = 0.38, $Tau^2$ = 0.01** | | |
| **Fixed** | | | | | **0.81 (0.75, 0.87)** | | | | | | **0.79 (0.71, 0.88)** | | |
| **Random** | | | | | **0.79 (0.68, 0.91)** | | | | | | **0.77 (0.67, 0.88)** | | |

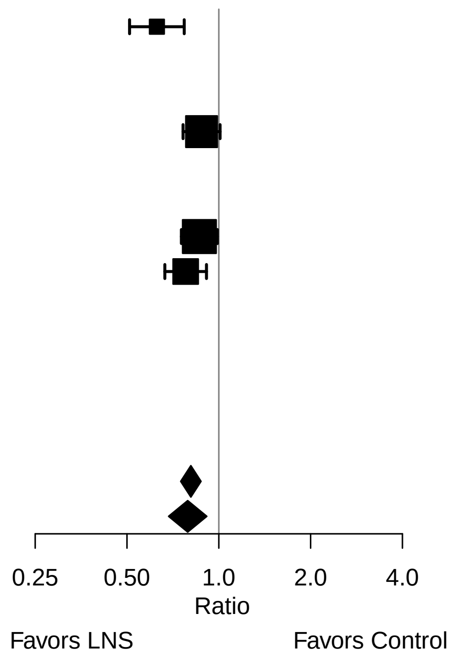

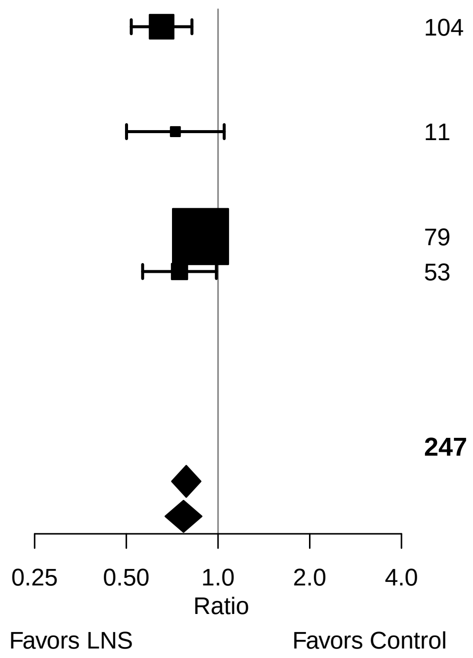

**Supplemental figure 6N: Geometric mean ratio of zinc protoporphyrin concentration**

#### 6N2: Stratified by Maternal age

**P−for−interaction = 0.998**
**Ratio of GMRs = 1.00 (0.88, 1.14)**

| Country | Trial | At least 25 y LNS N | Control N | Control Median | GMR (95% CI) | Fixed W | Random W | Less than 25 y LNS N | Control N | Control Median | GMR (95% CI) | Fixed W | Random W |
|---|---|---|---|---|---|---|---|---|---|---|---|---|---|
| Bangladesh | JiVitA−4 (35) | | | | | | | | | | | | |
| Bangladesh | RDNS (36) | | | | | | | | | | | | |
| Bangladesh | WASH−B (37) | | | | | | | | | | | | |
| Burkina Faso | iLiNS−Zinc (38) | 175 | 50 | 121.9 | 0.65 (0.53, 0.79) | 0.17 | 0.21 | 137 | 45 | 122.6 | 0.65 (0.53, 0.80) | 0.20 | 0.24 |
| Burkina Faso | PROMIS CS (39) | | | | | | | | | | | | |
| Ghana | GHANA (40) | | | | | | | | | | | | |
| Ghana | iLiNS−DYADG (41) | 65 | 128 | 45.8 | 0.88 (0.77, 1.02) | 0.32 | 0.28 | 35 | 74 | 52.1 | 0.84 (0.65, 1.07) | 0.15 | 0.20 |
| Kenya | WASH−B (42) | | | | | | | | | | | | |
| Madagascar | MAHAY (43) | | | | | | | | | | | | |
| Malawi | iLiNS−DYADM (44) | 88 | 192 | 50.4 | 0.84 (0.73, 0.97) | 0.32 | 0.28 | 102 | 183 | 57.4 | 0.89 (0.77, 1.02) | 0.45 | 0.34 |
| Malawi | iLiNS−DOSE (45) | 95 | 48 | 65.4 | 0.79 (0.66, 0.95) | 0.19 | 0.23 | 83 | 39 | 69.1 | 0.75 (0.60, 0.93) | 0.20 | 0.23 |
| Mali | PROMIS CS (46) | | | | | | | | | | | | |
| Zimbabwe | SHINE (HIV−) (47) | | | | | | | | | | | | |
| Zimbabwe | SHINE (HIV+) (48) | | | | | | | | | | | | |
| | | **423** | **418** | | **I² = 0.55, Tau² = 0.01** | | | **357** | **341** | | **I² = 0.53, Tau² = 0.01** | | |
| **Fixed** | | | | | **0.81 (0.74, 0.88)** | | | | | | **0.80 (0.73, 0.88)** | | |
| **Random** | | | | | **0.79 (0.70, 0.90)** | | | | | | **0.78 (0.68, 0.90)** | | |

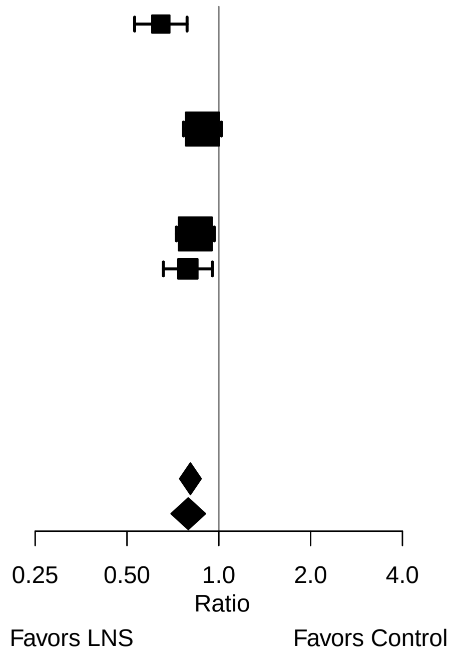

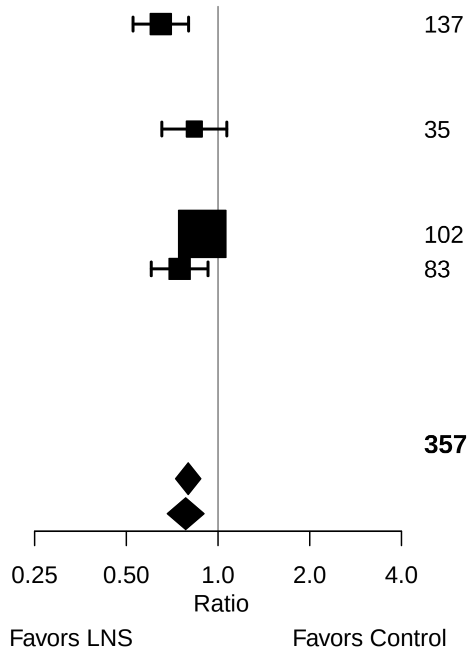

#### Supplemental figure 6N: Geometric mean ratio of zinc protoporphyrin concentration

##### 6N3: Stratified by Maternal education

**P−for−interaction = 0.117**
**Ratio of GMRs = 1.15 (0.97, 1.37)**

| Country | Trial | LNS N | Control N | Control Median | Primary or greater GMR (95% CI) | Fixed W | Random W | LNS N | Control N | Control Median | Incomplete or no formal GMR (95% CI) | Fixed W | Random W |
|---|---|---|---|---|---|---|---|---|---|---|---|---|---|
| Bangladesh | JiVitA−4 (35) | | | | | | | | | | | | |
| Bangladesh | RDNS (36) | | | | | | | | | | | | |
| Bangladesh | WASH−B (37) | | | | | | | | | | | | |
| Burkina Faso | iLiNS−Zinc (38) | | | | | | | | | | | | |
| Burkina Faso | PROMIS CS (39) | | | | | | | | | | | | |
| Ghana | GHANA (40) | | | | | | | | | | | | |
| Ghana | iLiNS−DYADG (41) | 83 | 159 | 48.4 | 0.85 (0.74, 0.98) | 0.66 | 0.56 | 17 | 43 | 43.5 | 0.92 (0.68, 1.25) | 0.08 | 0.08 |
| Kenya | WASH−B (42) | | | | | | | | | | | | |
| Madagascar | MAHAY (43) | | | | | | | | | | | | |
| Malawi | iLiNS−DYADM (44) | 31 | 53 | 61.6 | 0.79 (0.58, 1.07) | 0.14 | 0.19 | 158 | 320 | 53.1 | 0.88 (0.79, 0.98) | 0.66 | 0.66 |
| Malawi | iLiNS−DOSE (45) | 48 | 26 | 76.1 | 0.66 (0.51, 0.86) | 0.20 | 0.25 | 130 | 61 | 65.8 | 0.83 (0.70, 0.98) | 0.27 | 0.27 |
| Mali | PROMIS CS (46) | | | | | | | | | | | | |
| Zimbabwe | SHINE (HIV−) (47) | | | | | | | | | | | | |
| Zimbabwe | SHINE (HIV+) (48) | | | | | | | | | | | | |
| | | **162** | **238** | | **I² = 0.30, Tau² = 0.00** | | | **305** | **424** | | **I² = 0.00, Tau² = 0.00** | | |
| **Fixed** | | | | | **0.80 (0.71, 0.90)** | | | | | | **0.87 (0.80, 0.95)** | | |
| **Random** | | | | | **0.79 (0.68, 0.91)** | | | | | | **0.87 (0.80, 0.95)** | | |

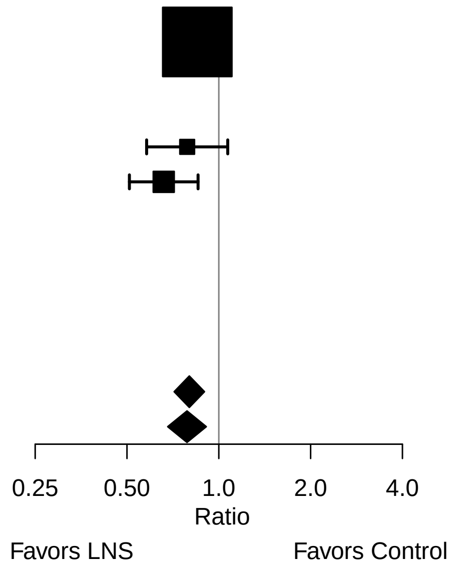

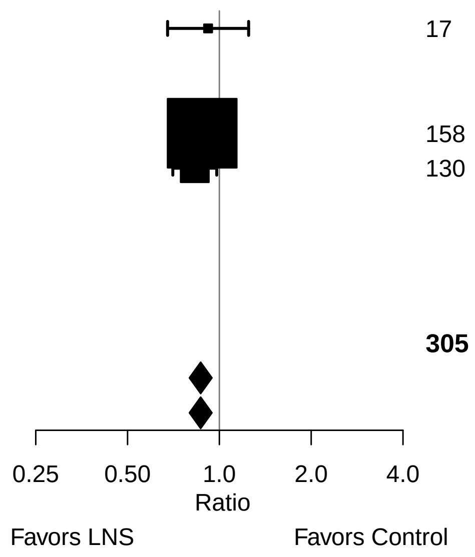

**Supplemental figure 6N: Geometric mean ratio of zinc protoporphyrin concentration**

#### 6N4: Stratified by Child sex

**P−for−interaction = 0.040**
**Ratio of GMRs = 1.13 (1.01, 1.27)**

| Country | Trial | Male LNS N | Male Control N | Male Control Median | Male GMR (95% CI) | Male Fixed W | Male Random W | Female LNS N | Female Control N | Female Control Median | Female GMR (95% CI) | Female Fixed W | Female Random W |
|---|---|---|---|---|---|---|---|---|---|---|---|---|---|
| Bangladesh | JiVitA−4 (35) | | | | | | | | | | | | |
| Bangladesh | RDNS (36) | | | | | | | | | | | | |
| Bangladesh | WASH−B (37) | | | | | | | | | | | | |
| Burkina Faso | iLiNS−Zinc (38) | 149 | 49 | 131.3 | 0.58 (0.48, 0.70) | 0.19 | 0.24 | 163 | 46 | 99.3 | 0.72 (0.63, 0.83) | 0.34 | 0.30 |
| Burkina Faso | PROMIS CS (39) | | | | | | | | | | | | |
| Ghana | GHANA (40) | | | | | | | | | | | | |
| Ghana | iLiNS−DYADG (41) | 63 | 95 | 49.1 | 0.89 (0.76, 1.04) | 0.27 | 0.26 | 36 | 106 | 46.1 | 0.85 (0.69, 1.04) | 0.16 | 0.20 |
| Kenya | WASH−B (42) | | | | | | | | | | | | |
| Madagascar | MAHAY (43) | | | | | | | | | | | | |
| Malawi | iLiNS−DYADM (44) | 95 | 183 | 57.9 | 0.81 (0.70, 0.93) | 0.35 | 0.27 | 95 | 192 | 49.0 | 0.93 (0.81, 1.07) | 0.34 | 0.30 |
| Malawi | iLiNS−DOSE (45) | 86 | 46 | 66.9 | 0.74 (0.61, 0.89) | 0.19 | 0.24 | 95 | 41 | 65.2 | 0.81 (0.66, 1.00) | 0.16 | 0.20 |
| Mali | PROMIS CS (46) | | | | | | | | | | | | |
| Zimbabwe | SHINE (HIV−) (47) | | | | | | | | | | | | |
| Zimbabwe | SHINE (HIV+) (48) | | | | | | | | | | | | |
| | | **393** | **373** | | **I² = 0.75, Tau² = 0.03** | | | **389** | **385** | | **I² = 0.52, Tau² = 0.01** | | |
| **Fixed** | | | | | **0.76 (0.70, 0.83)** | | | | | | **0.82 (0.76, 0.89)** | | |
| **Random** | | | | | **0.75 (0.63, 0.89)** | | | | | | **0.82 (0.74, 0.92)** | | |

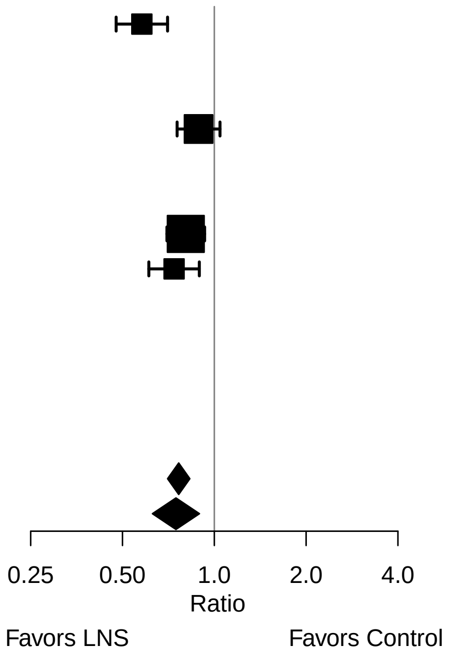

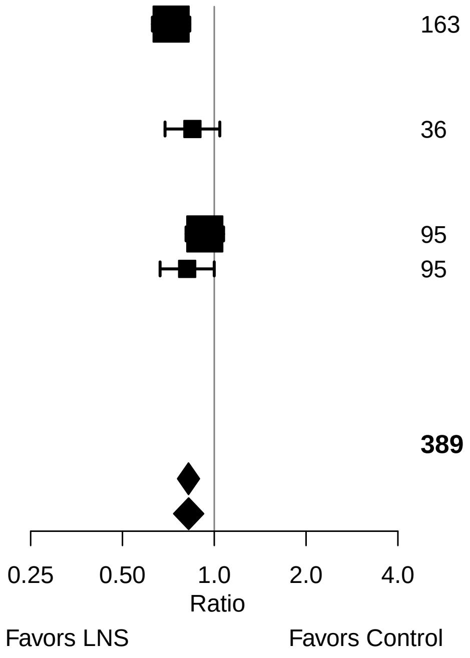

#### Supplemental figure 6N: Geometric mean ratio of zinc protoporphyrin concentration

##### 6N5: Stratified by Child birth order

**P−for−interaction = 0.311**
**Ratio of GMRs = 0.93 (0.80, 1.07)**

| | | Later born | | | | | | Firstborn | | | | | |
|---|---|---|---|---|---|---|---|---|---|---|---|---|---|
| Country | Trial | LNS N | Control N | Control Median | GMR (95% CI) | Fixed W | Random W | LNS N | Control N | Control Median | GMR (95% CI) | Fixed W | Random W |
| Bangladesh | JiVitA−4 (35) | | | | | | | | | | | | |
| Bangladesh | RDNS (36) | | | | | | | | | | | | |
| Bangladesh | WASH−B (37) | | | | | | | | | | | | |
| Burkina Faso | iLiNS−Zinc (38) | 246 | 71 | 116.3 | 0.68 (0.58, 0.79) | 0.22 | 0.24 | 66 | 24 | 157.4 | 0.56 (0.43, 0.72) | 0.24 | 0.25 |
| Burkina Faso | PROMIS CS (39) | | | | | | | | | | | | |
| Ghana | GHANA (40) | | | | | | | | | | | | |
| Ghana | iLiNS−DYADG (41) | 70 | 141 | 45.8 | 0.92 (0.79, 1.07) | 0.22 | 0.24 | 30 | 61 | 51.1 | 0.75 (0.59, 0.94) | 0.30 | 0.27 |
| Kenya | WASH−B (42) | | | | | | | | | | | | |
| Madagascar | MAHAY (43) | | | | | | | | | | | | |
| Malawi | iLiNS−DYADM (44) | 152 | 299 | 52.8 | 0.86 (0.77, 0.97) | 0.39 | 0.29 | 38 | 75 | 55.9 | 0.89 (0.72, 1.10) | 0.34 | 0.28 |
| Malawi | iLiNS−DOSE (45) | 119 | 59 | 65.5 | 0.77 (0.65, 0.92) | 0.18 | 0.23 | 33 | 19 | 58.4 | 0.90 (0.63, 1.29) | 0.12 | 0.19 |
| Mali | PROMIS CS (46) | | | | | | | | | | | | |
| Zimbabwe | SHINE (HIV−) (47) | | | | | | | | | | | | |
| Zimbabwe | SHINE (HIV+) (48) | | | | | | | | | | | | |
| | | **587** | **570** | | **I² = 0.67, Tau² = 0.01** | | | **167** | **179** | | **I² = 0.65, Tau² = 0.03** | | |
| **Fixed** | | | | | **0.82 (0.76, 0.88)** | | | | | | **0.76 (0.67, 0.86)** | | |
| **Random** | | | | | **0.81 (0.71, 0.92)** | | | | | | **0.76 (0.61, 0.94)** | | |

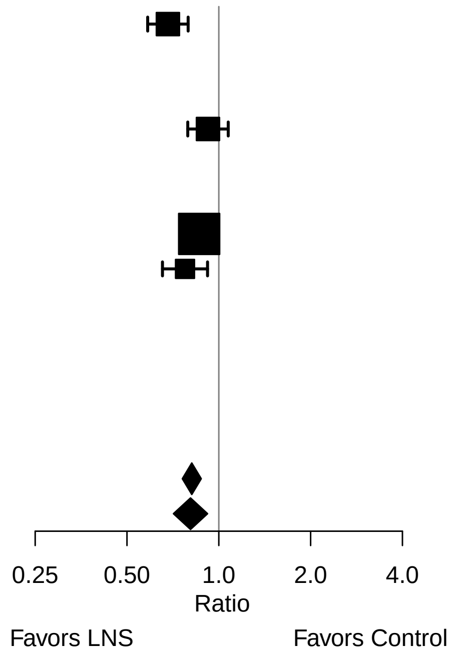

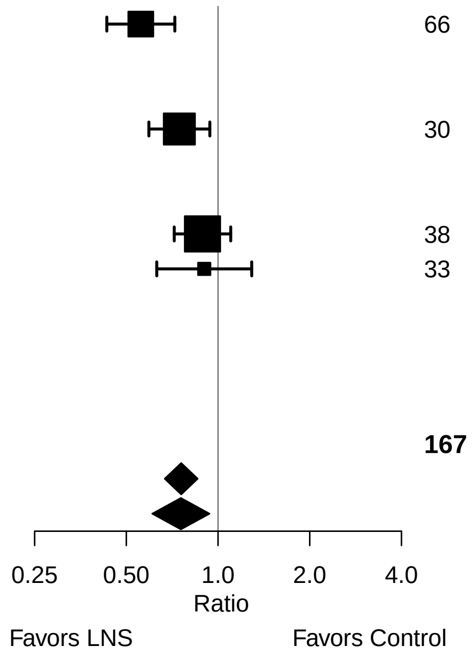

**Supplemental figure 6N: Geometric mean ratio of zinc protoporphyrin concentration**

#### 6N6: Stratified by Child baseline acute malnutrition

**P−for−interaction = 0.817**
**Ratio of GMRs = 0.98 (0.82, 1.16)**

| | | No | | | | | | Yes | | | | | |
|---|---|---|---|---|---|---|---|---|---|---|---|---|---|
| Country | Trial | LNS N | Control N | Control Median | GMR (95% CI) | Fixed W | Random W | LNS N | Control N | Control Median | GMR (95% CI) | Fixed W | Random W |
| Bangladesh | JiVitA−4 (35) | | | | | | | | | | | | |
| Bangladesh | RDNS (36) | | | | | | | | | | | | |
| Bangladesh | WASH−B (37) | | | | | | | | | | | | |
| Burkina Faso | iLiNS−Zinc (38) | 227 | 72 | 121.6 | 0.64 (0.55, 0.73) | 0.26 | 0.32 | 85 | 23 | 122.9 | 0.66 (0.54, 0.81) | 0.60 | 0.60 |
| Burkina Faso | PROMIS CS (39) | | | | | | | | | | | | |
| Ghana | GHANA (40) | | | | | | | | | | | | |
| Ghana | iLiNS−DYADG (41) | 94 | 187 | 48.1 | 0.88 (0.77, 1.01) | 0.29 | 0.33 | 6 | 15 | 45.8 | 0.62 (0.43, 0.89) | 0.20 | 0.20 |
| Kenya | WASH−B (42) | | | | | | | | | | | | |
| Madagascar | MAHAY (43) | | | | | | | | | | | | |
| Malawi | iLiNS−DYADM (44) | 165 | 330 | 53.2 | 0.87 (0.79, 0.97) | 0.45 | 0.35 | 11 | 33 | 59.2 | 0.83 (0.58, 1.20) | 0.20 | 0.20 |
| Malawi | iLiNS−DOSE (45) | | | | | | | | | | | | |
| Mali | PROMIS CS (46) | | | | | | | | | | | | |
| Zimbabwe | SHINE (HIV−) (47) | | | | | | | | | | | | |
| Zimbabwe | SHINE (HIV+) (48) | | | | | | | | | | | | |
| | | **486** | **589** | | **I² = 0.86, Tau² = 0.03** | | | **102** | **71** | | **I² = 0.00, Tau² = 0.00** | | |
| **Fixed** | | | | | **0.81 (0.75, 0.87)** | | | | | | **0.68 (0.58, 0.80)** | | |
| **Random** | | | | | **0.79 (0.64, 0.97)** | | | | | | **0.68 (0.58, 0.80)** | | |

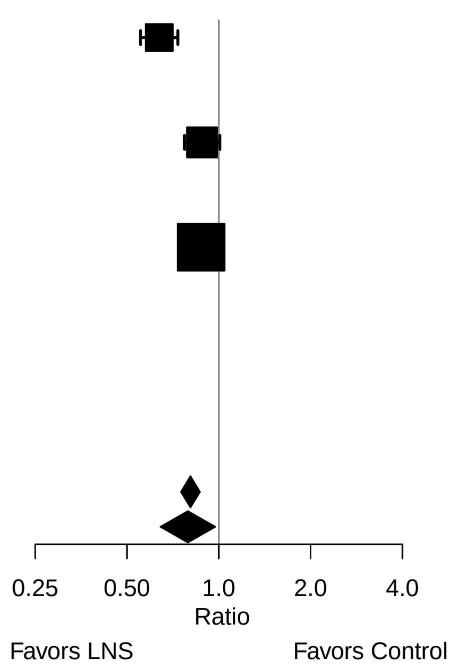

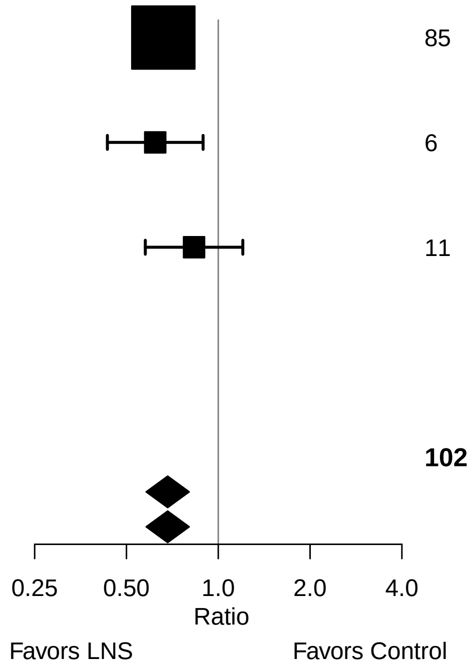

**Supplemental figure 6N: Geometric mean ratio of zinc protoporphyrin concentration**

#### 6N7: Stratified by Child baseline anemia

**P−for−interaction = 0.243**
**Ratio of GMRs = 1.09 (0.95, 1.25)**

| | | Not anemic | | | | | | Anemic | | | | | |
|---|---|---|---|---|---|---|---|---|---|---|---|---|---|
| Country | Trial | LNS N | Control N | Control Median | GMR (95% CI) | Fixed W | Random W | LNS N | Control N | Control Median | GMR (95% CI) | Fixed W | Random W |
| Bangladesh | JiVitA−4 (35) | | | | | | | | | | | | |
| Bangladesh | RDNS (36) | | | | | | | | | | | | |
| Bangladesh | WASH−B (37) | | | | | | | | | | | | |
| Burkina Faso | iLiNS−Zinc (38) | 20 | 7 | 83.9 | 0.63 (0.41, 0.98) | 0.05 | 0.06 | 292 | 88 | 122.6 | 0.65 (0.57, 0.73) | 0.35 | 0.28 |
| Burkina Faso | PROMIS CS (39) | | | | | | | | | | | | |
| Ghana | GHANA (40) | | | | | | | | | | | | |
| Ghana | iLiNS−DYADG (41) | 75 | 132 | 45.7 | 0.82 (0.71, 0.95) | 0.43 | 0.41 | 25 | 70 | 52.3 | 1.03 (0.80, 1.32) | 0.09 | 0.20 |
| Kenya | WASH−B (42) | | | | | | | | | | | | |
| Madagascar | MAHAY (43) | | | | | | | | | | | | |
| Malawi | iLiNS−DYADM (44) | 59 | 121 | 44.5 | 0.88 (0.74, 1.04) | 0.32 | 0.32 | 121 | 242 | 58.4 | 0.87 (0.77, 0.98) | 0.39 | 0.28 |
| Malawi | iLiNS−DOSE (45) | 66 | 36 | 57.3 | 0.72 (0.58, 0.89) | 0.20 | 0.22 | 115 | 50 | 70.0 | 0.82 (0.68, 0.99) | 0.17 | 0.24 |
| Mali | PROMIS CS (46) | | | | | | | | | | | | |
| Zimbabwe | SHINE (HIV−) (47) | | | | | | | | | | | | |
| Zimbabwe | SHINE (HIV+) (48) | | | | | | | | | | | | |
| | | **220** | **296** | | **I² = 0.10, Tau² = 0.00** | | | **553** | **450** | | **I² = 0.82, Tau² = 0.03** | | |
| **Fixed** | | | | | **0.81 (0.73, 0.89)** | | | | | | **0.79 (0.73, 0.85)** | | |
| **Random** | | | | | **0.81 (0.72, 0.89)** | | | | | | **0.82 (0.68, 0.99)** | | |

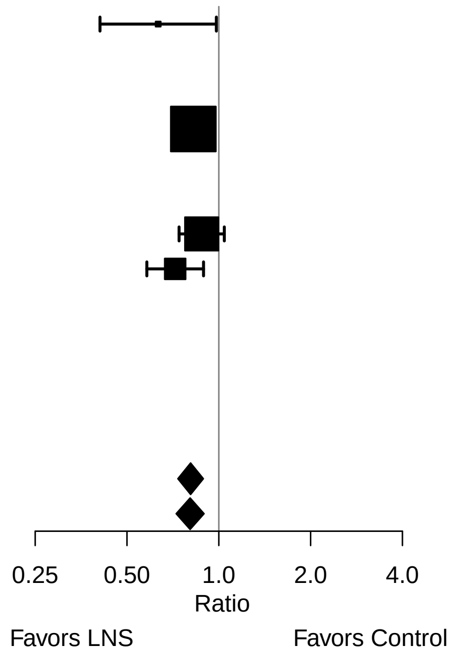

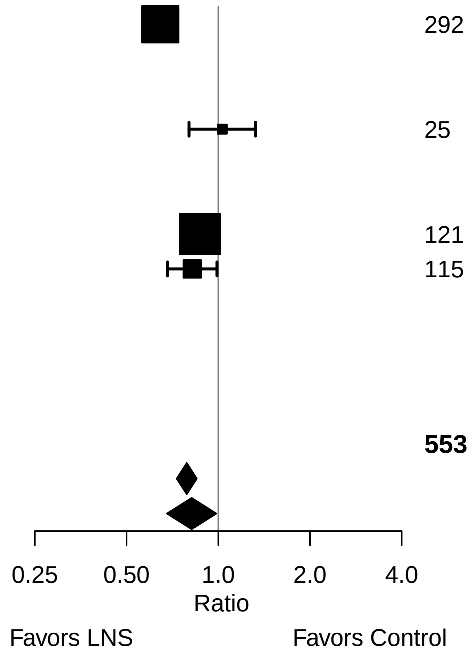

**Supplemental figure 6N: Geometric mean ratio of zinc protoporphyrin concentration**

#### 6N8: Stratified by Child high-dose vitamin A supplementation (insufficient comparisons)

**Supplemental figure 6N: Geometric mean ratio of zinc protoporphyrin concentration**

#### 6N9: Stratified by Child inflammation

**P−for−interaction = 0.777**
**Ratio of GMRs = 0.98 (0.86, 1.11)**

| Country | Trial | LNS N | Control N | Control Median | Not inflamed GMR (95% CI) | Fixed W | Random W | LNS N | Control N | Control Median | High AGP or CRP GMR (95% CI) | Fixed W | Random W |
|---|---|---|---|---|---|---|---|---|---|---|---|---|---|
| Bangladesh | JiVitA−4 (35) | | | | | | | | | | | | |
| Bangladesh | RDNS (36) | | | | | | | | | | | | |
| Bangladesh | WASH−B (37) | | | | | | | | | | | | |
| Burkina Faso | iLiNS−Zinc (38) | 123 | 30 | 126.7 | 0.61 (0.48, 0.76) | 0.20 | 0.24 | 189 | 65 | 113.4 | 0.67 (0.57, 0.78) | 0.23 | 0.24 |
| Burkina Faso | PROMIS CS (39) | | | | | | | | | | | | |
| Ghana | GHANA (40) | | | | | | | | | | | | |
| Ghana | iLiNS−DYADG (41) | 45 | 120 | 45.3 | 0.87 (0.72, 1.05) | 0.30 | 0.27 | 55 | 82 | 51.6 | 0.85 (0.72, 1.02) | 0.18 | 0.22 |
| Kenya | WASH−B (42) | | | | | | | | | | | | |
| Madagascar | MAHAY (43) | | | | | | | | | | | | |
| Malawi | iLiNS−DYADM (44) | 53 | 119 | 48.8 | 0.94 (0.78, 1.12) | 0.31 | 0.27 | 137 | 256 | 55.8 | 0.84 (0.75, 0.95) | 0.39 | 0.31 |
| Malawi | iLiNS−DOSE (45) | 62 | 26 | 60.3 | 0.79 (0.62, 1.01) | 0.18 | 0.23 | 119 | 61 | 67.0 | 0.77 (0.65, 0.91) | 0.20 | 0.23 |
| Mali | PROMIS CS (46) | | | | | | | | | | | | |
| Zimbabwe | SHINE (HIV−) (47) | | | | | | | | | | | | |
| Zimbabwe | SHINE (HIV+) (48) | | | | | | | | | | | | |
| | | **283** | **295** | | **$I^2$ = 0.67, $Tau^2$ = 0.02** | | | **500** | **464** | | **$I^2$ = 0.51, $Tau^2$ = 0.01** | | |
| **Fixed** | | | | | **0.81 (0.73, 0.90)** | | | | | | **0.79 (0.73, 0.85)** | | |
| **Random** | | | | | **0.80 (0.66, 0.96)** | | | | | | **0.78 (0.70, 0.87)** | | |

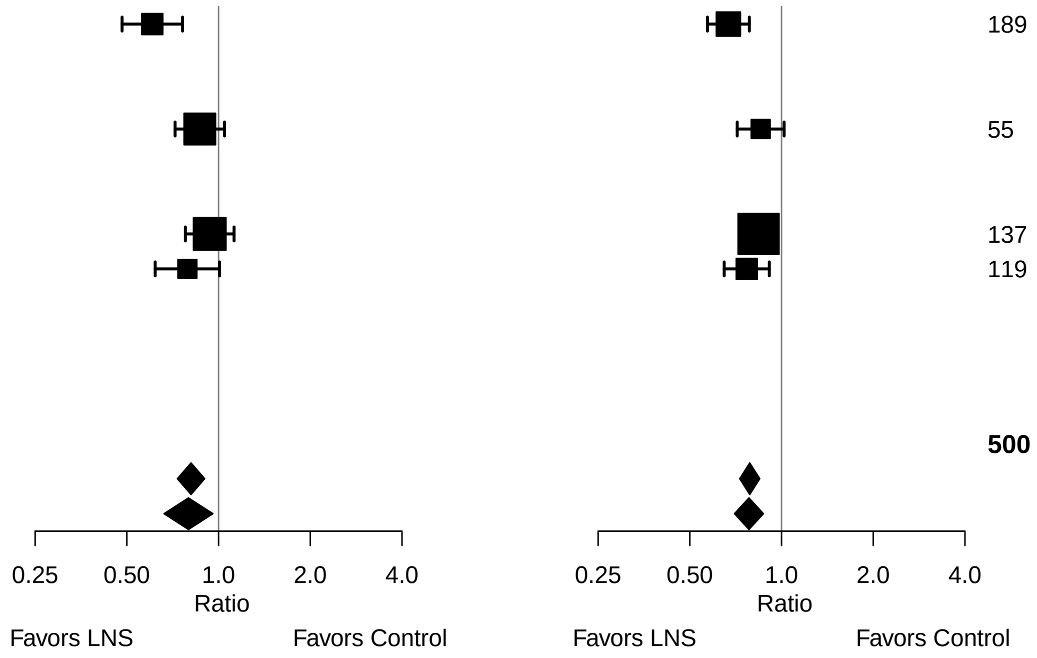

### Supplemental figure 6O: Elevated zinc protoporphyrin prevalence ratio

#### 6O1: Stratified by Maternal BMI

**P−for−interaction = 0.942**
**Ratio of PRs = 1.01 (0.78, 1.30)**

| | | At least 20 kg/m² | | | | | | Less than 20 kg/m² | | | | | |
|---|---|---|---|---|---|---|---|---|---|---|---|---|---|
| Country | Trial | LNS N | Control N | Control Prevalence | PR (95% CI) | Fixed W | Random W | LNS N | Control N | Control Prevalence | PR (95% CI) | Fixed W | Random W |
| Bangladesh | JiVitA−4 (35) | | | | | | | | | | | | |
| Bangladesh | RDNS (36) | | | | | | | | | | | | |
| Bangladesh | WASH−B (37) | | | | | | | | | | | | |
| Burkina Faso | iLiNS−Zinc (38) | 208 | 55 | 73.8 | 0.73 (0.57, 0.94) | 0.52 | 0.52 | 104 | 40 | 77.3 | 0.69 (0.56, 0.86) | 0.74 | 0.74 |
| Burkina Faso | PROMIS CS (39) | | | | | | | | | | | | |
| Ghana | GHANA (40) | | | | | | | | | | | | |
| Ghana | iLiNS−DYADG (41) | | | | | | | | | | | | |
| Kenya | WASH−B (42) | | | | | | | | | | | | |
| Madagascar | MAHAY (43) | | | | | | | | | | | | |
| Malawi | iLiNS−DYADM (44) | 111 | 222 | 36.5 | 0.64 (0.44, 0.94) | 0.23 | 0.23 | 79 | 151 | 29.1 | 0.83 (0.52, 1.31) | 0.16 | 0.16 |
| Malawi | iLiNS−DOSE (45) | 127 | 65 | 46.2 | 0.60 (0.41, 0.86) | 0.25 | 0.25 | 53 | 22 | 40.9 | 0.62 (0.35, 1.10) | 0.10 | 0.10 |
| Mali | PROMIS CS (46) | | | | | | | | | | | | |
| Zimbabwe | SHINE (HIV−) (47) | | | | | | | | | | | | |
| Zimbabwe | SHINE (HIV+) (48) | | | | | | | | | | | | |
| | | **446** | **342** | | **I² = 0.00, Tau² = 0.00** | | | **236** | **213** | | **I² = 0.00, Tau² = 0.00** | | |
| **Fixed** | | | | | **0.67 (0.56, 0.81)** | | | | | | **0.70 (0.58, 0.85)** | | |
| **Random** | | | | | **0.67 (0.56, 0.81)** | | | | | | **0.70 (0.58, 0.85)** | | |

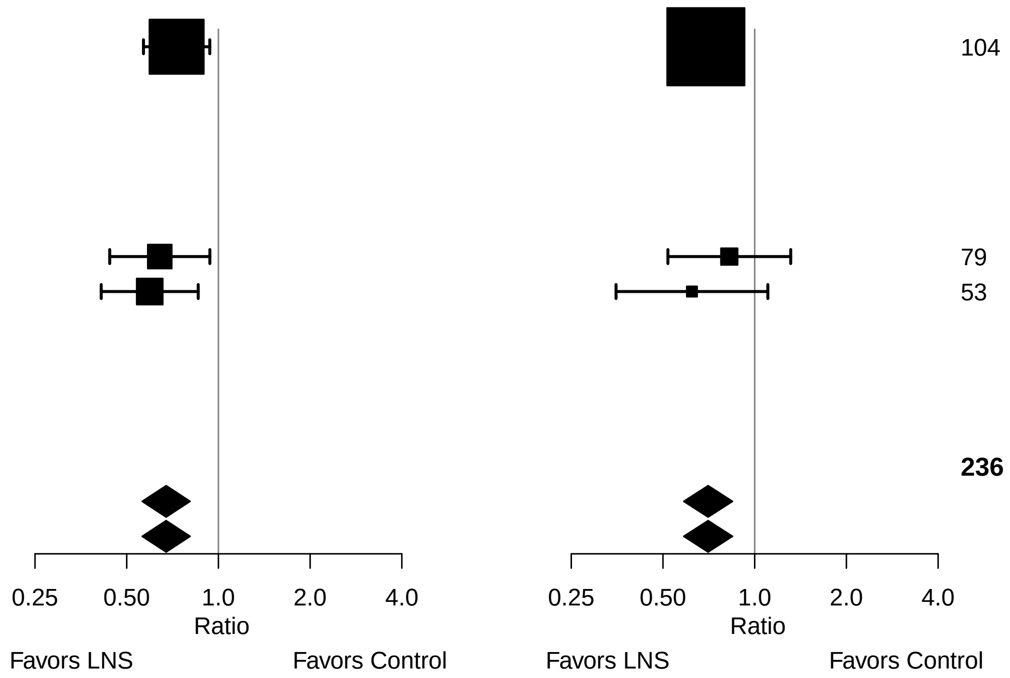

##### Supplemental figure 6O: Elevated zinc protoporphyrin prevalence ratio

#### 6O2: Stratified by Maternal age

**P−for−interaction = 0.660**
**Ratio of PRs = 1.07 (0.80, 1.43)**

| Country | Trial | At least 25 y LNS N | Control N | Control Prevalence | PR (95% CI) | Fixed W | Random W | Less than 25 y LNS N | Control N | Control Prevalence | PR (95% CI) | Fixed W | Random W |
|---|---|---|---|---|---|---|---|---|---|---|---|---|---|
| Bangladesh | JiVitA−4 (35) | | | | | | | | | | | | |
| Bangladesh | RDNS (36) | | | | | | | | | | | | |
| Bangladesh | WASH−B (37) | | | | | | | | | | | | |
| Burkina Faso | iLiNS−Zinc (38) | 175 | 50 | 74.1 | 0.66 (0.49, 0.89) | 0.49 | 0.49 | 137 | 45 | 76.5 | 0.78 (0.58, 1.03) | 0.45 | 0.45 |
| Burkina Faso | PROMIS CS (39) | | | | | | | | | | | | |
| Ghana | GHANA (40) | | | | | | | | | | | | |
| Ghana | iLiNS−DYADG (41) | 65 | 128 | 16.4 | 0.75 (0.35, 1.60) | 0.07 | 0.07 | 35 | 74 | 37.8 | 0.76 (0.41, 1.38) | 0.10 | 0.10 |
| Kenya | WASH−B (42) | | | | | | | | | | | | |
| Madagascar | MAHAY (43) | | | | | | | | | | | | |
| Malawi | iLiNS−DYADM (44) | 88 | 192 | 28.1 | 0.77 (0.49, 1.21) | 0.20 | 0.20 | 102 | 183 | 39.3 | 0.65 (0.44, 0.94) | 0.26 | 0.26 |
| Malawi | iLiNS−DOSE (45) | 95 | 48 | 41.7 | 0.56 (0.37, 0.85) | 0.24 | 0.24 | 83 | 39 | 48.7 | 0.65 (0.42, 1.01) | 0.19 | 0.19 |
| Mali | PROMIS CS (46) | | | | | | | | | | | | |
| Zimbabwe | SHINE (HIV−) (47) | | | | | | | | | | | | |
| Zimbabwe | SHINE (HIV+) (48) | | | | | | | | | | | | |
| | | **423** | **418** | | **I² = 0.00, Tau² = 0.00** | | | **357** | **341** | | **I² = 0.00, Tau² = 0.00** | | |
| **Fixed** | | | | | **0.66 (0.54, 0.81)** | | | | | | **0.72 (0.59, 0.87)** | | |
| **Random** | | | | | **0.66 (0.54, 0.81)** | | | | | | **0.72 (0.59, 0.87)** | | |

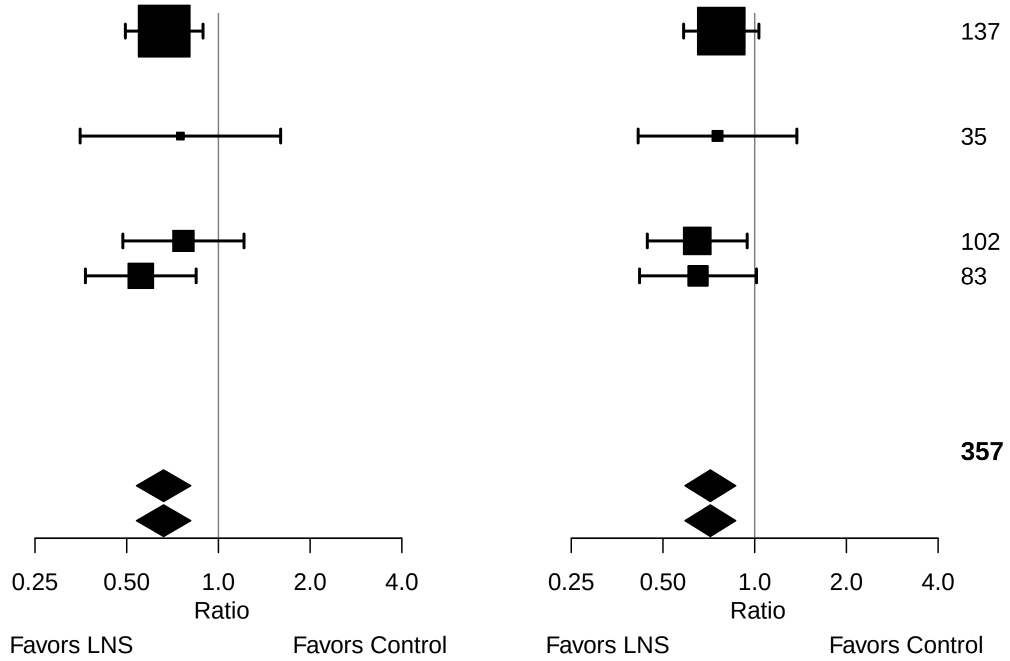

**Supplemental figure 6O: Elevated zinc protoporphyrin prevalence ratio**

#### 6O3: Stratified by Maternal education

**P−for−interaction = 0.114**
**Ratio of PRs = 1.42 (0.92, 2.20)**

| Country | Trial | Primary or greater: LNS N | Control N | Control Prevalence | PR (95% CI) | Fixed W | Random W | Incomplete or no formal: LNS N | Control N | Control Prevalence | PR (95% CI) | Fixed W | Random W |
|---|---|---|---|---|---|---|---|---|---|---|---|---|---|
| Bangladesh | JiVitA−4 (35) | | | | | | | | | | | | |
| Bangladesh | RDNS (36) | | | | | | | | | | | | |
| Bangladesh | WASH−B (37) | | | | | | | | | | | | |
| Burkina Faso | iLiNS−Zinc (38) | | | | | | | | | | | | |
| Burkina Faso | PROMIS CS (39) | | | | | | | | | | | | |
| Ghana | GHANA (40) | | | | | | | | | | | | |
| Ghana | iLiNS−DYADG (41) | 83 | 159 | 25.2 | 0.62 (0.35, 1.10) | 0.33 | 0.33 | 17 | 43 | 20.9 | 1.41 (0.55, 3.59) | 0.07 | 0.07 |
| Kenya | WASH−B (42) | | | | | | | | | | | | |
| Madagascar | MAHAY (43) | | | | | | | | | | | | |
| Malawi | iLiNS−DYADM (44) | 31 | 53 | 45.3 | 0.64 (0.34, 1.20) | 0.27 | 0.27 | 158 | 320 | 31.6 | 0.70 (0.50, 0.98) | 0.52 | 0.52 |
| Malawi | iLiNS−DOSE (45) | 48 | 26 | 53.8 | 0.47 (0.28, 0.79) | 0.40 | 0.40 | 130 | 61 | 41.0 | 0.70 (0.48, 1.02) | 0.41 | 0.41 |
| Mali | PROMIS CS (46) | | | | | | | | | | | | |
| Zimbabwe | SHINE (HIV−) (47) | | | | | | | | | | | | |
| Zimbabwe | SHINE (HIV+) (48) | | | | | | | | | | | | |
| | | **162** | **238** | | **I² = 0.00, Tau² = 0.00** | | | **305** | **424** | | **I² = 0.00, Tau² = 0.00** | | |
| **Fixed** | | | | | **0.56 (0.41, 0.78)** | | | | | | **0.74 (0.58, 0.94)** | | |
| **Random** | | | | | **0.56 (0.41, 0.78)** | | | | | | **0.74 (0.58, 0.94)** | | |

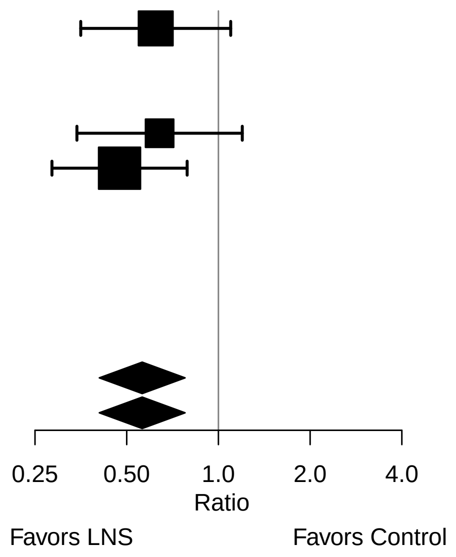

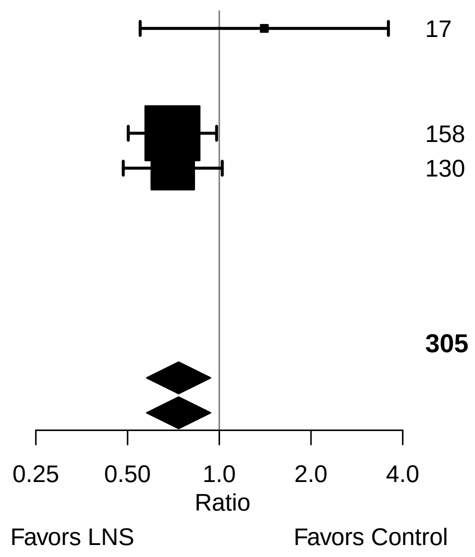

#### Supplemental figure 6O: Elevated zinc protoporphyrin prevalence ratio

##### 6O4: Stratified by Child sex

**P−for−interaction = 0.187**
**Ratio of PRs = 1.13 (0.94, 1.36)**

| Country | Trial | Male LNS N | Male Control N | Male Control Prevalence | Male PR (95% CI) | Male Fixed W | Male Random W | Female LNS N | Female Control N | Female Control Prevalence | Female PR (95% CI) | Female Fixed W | Female Random W |
|---|---|---|---|---|---|---|---|---|---|---|---|---|---|
| Bangladesh | JiVitA−4 (35) | | | | | | | | | | | | |
| Bangladesh | RDNS (36) | | | | | | | | | | | | |
| Bangladesh | WASH−B (37) | | | | | | | | | | | | |
| Burkina Faso | iLiNS−Zinc (38) | 149 | 49 | 81.5 | 0.68 (0.53, 0.86) | 0.56 | 0.56 | 163 | 46 | 68.6 | 0.76 (0.64, 0.91) | 0.73 | 0.73 |
| Burkina Faso | PROMIS CS (39) | | | | | | | | | | | | |
| Ghana | GHANA (40) | | | | | | | | | | | | |
| Ghana | iLiNS−DYADG (41) | 63 | 95 | 26.3 | 0.72 (0.39, 1.33) | 0.09 | 0.09 | 36 | 106 | 21.7 | 0.77 (0.34, 1.74) | 0.03 | 0.03 |
| Kenya | WASH−B (42) | | | | | | | | | | | | |
| Madagascar | MAHAY (43) | | | | | | | | | | | | |
| Malawi | iLiNS−DYADM (44) | 95 | 183 | 37.2 | 0.62 (0.41, 0.94) | 0.19 | 0.19 | 95 | 192 | 30.2 | 0.80 (0.53, 1.21) | 0.13 | 0.13 |
| Malawi | iLiNS−DOSE (45) | 86 | 46 | 43.5 | 0.61 (0.40, 0.95) | 0.17 | 0.17 | 95 | 41 | 46.3 | 0.67 (0.42, 1.05) | 0.11 | 0.11 |
| Mali | PROMIS CS (46) | | | | | | | | | | | | |
| Zimbabwe | SHINE (HIV−) (47) | | | | | | | | | | | | |
| Zimbabwe | SHINE (HIV+) (48) | | | | | | | | | | | | |
| | | **393** | **373** | | **I² = 0.00, Tau² = 0.00** | | | **389** | **385** | | **I² = 0.00, Tau² = 0.00** | | |
| **Fixed** | | | | | **0.66 (0.55, 0.79)** | | | | | | **0.76 (0.65, 0.88)** | | |
| **Random** | | | | | **0.66 (0.55, 0.79)** | | | | | | **0.76 (0.65, 0.88)** | | |

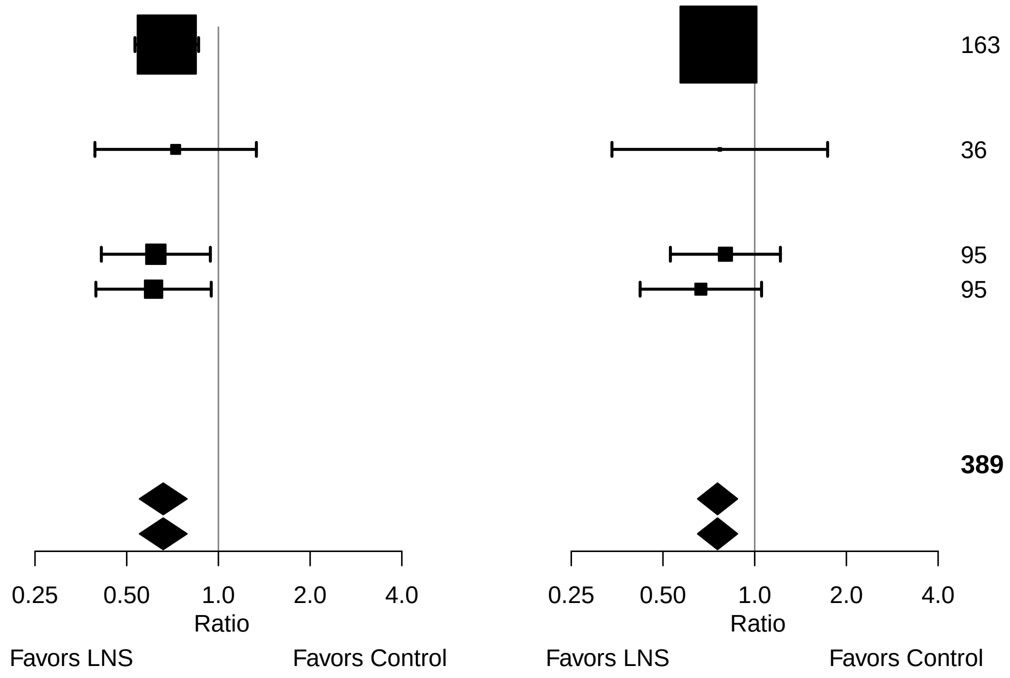

#### Supplemental figure 6O: Elevated zinc protoporphyrin prevalence ratio

#### 6O5: Stratified by Child birth order

**P−for−interaction = 0.123**
**Ratio of PRs = 1.21 (0.95, 1.53)**

| Country | Trial | LNS N | Control N | Control Prevalence | Later born PR (95% CI) | Fixed W | Random W | LNS N | Control N | Control Prevalence | Firstborn PR (95% CI) | Fixed W | Random W |
|---|---|---|---|---|---|---|---|---|---|---|---|---|---|
| Bangladesh | JiVitA−4 (35) | | | | | | | | | | | | |
| Bangladesh | RDNS (36) | | | | | | | | | | | | |
| Bangladesh | WASH−B (37) | | | | | | | | | | | | |
| Burkina Faso | iLiNS−Zinc (38) | 246 | 71 | 74.7 | 0.69 (0.56, 0.84) | 0.59 | 0.59 | 66 | 24 | 76.7 | 0.86 (0.67, 1.11) | 0.79 | 0.79 |
| Burkina Faso | PROMIS CS (39) | | | | | | | | | | | | |
| Ghana | GHANA (40) | | | | | | | | | | | | |
| Ghana | iLiNS−DYADG (41) | | | | | | | | | | | | |
| Kenya | WASH−B (42) | | | | | | | | | | | | |
| Madagascar | MAHAY (43) | | | | | | | | | | | | |
| Malawi | iLiNS−DYADM (44) | 152 | 299 | 33.4 | 0.73 (0.53, 1.00) | 0.24 | 0.24 | 38 | 75 | 34.7 | 0.61 (0.30, 1.21) | 0.11 | 0.11 |
| Malawi | iLiNS−DOSE (45) | 119 | 59 | 45.8 | 0.55 (0.37, 0.79) | 0.17 | 0.17 | 33 | 19 | 36.8 | 0.99 (0.49, 2.02) | 0.10 | 0.10 |
| Mali | PROMIS CS (46) | | | | | | | | | | | | |
| Zimbabwe | SHINE (HIV−) (47) | | | | | | | | | | | | |
| Zimbabwe | SHINE (HIV+) (48) | | | | | | | | | | | | |
| | | **517** | **429** | | **I² = 0.00, Tau² = 0.00** | | | **137** | **118** | | **I² = 0.00, Tau² = 0.00** | | |
| **Fixed** | | | | | **0.67 (0.57, 0.78)** | | | | | | **0.84 (0.67, 1.06)** | | |
| **Random** | | | | | **0.67 (0.57, 0.78)** | | | | | | **0.84 (0.67, 1.06)** | | |

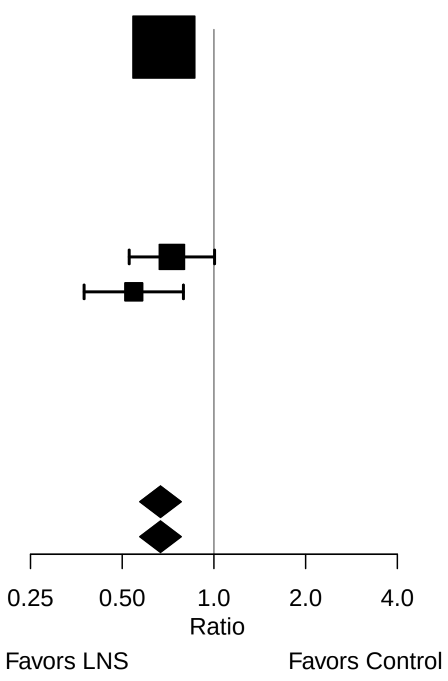

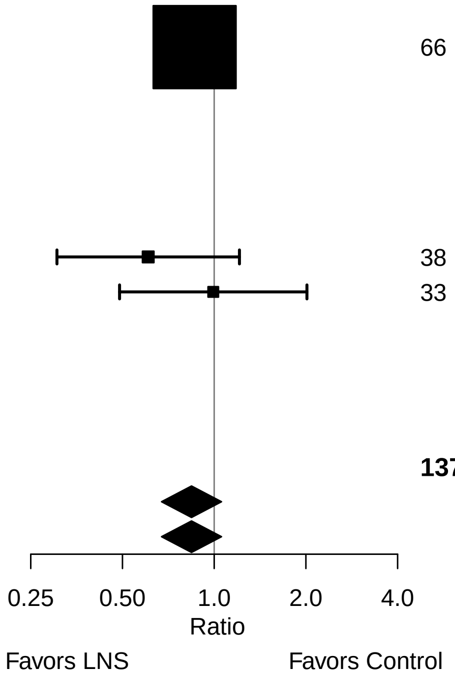

**Supplemental figure 6O: Elevated zinc protoporphyrin prevalence ratio**

#### 6O6: Stratified by Child baseline acute malnutrition (insufficient comparisons)

#### Supplemental figure 6O: Elevated zinc protoporphyrin prevalence ratio

##### 6O7: Stratified by Child baseline anemia

**P−for−interaction = 0.305**
**Ratio of PRs = 1.26 (0.81, 1.94)**

| Country | Trial | Not anemic: LNS N | Not anemic: Control N | Not anemic: Control Prevalence | Not anemic: PR (95% CI) | Not anemic: Fixed W | Not anemic: Random W | Anemic: LNS N | Anemic: Control N | Anemic: Control Prevalence | Anemic: PR (95% CI) | Anemic: Fixed W | Anemic: Random W |
|---|---|---|---|---|---|---|---|---|---|---|---|---|---|
| Bangladesh | JiVitA−4 (35) | | | | | | | | | | | | |
| Bangladesh | RDNS (36) | | | | | | | | | | | | |
| Bangladesh | WASH−B (37) | | | | | | | | | | | | |
| Burkina Faso | iLiNS−Zinc (38) | | | | | | | | | | | | |
| Burkina Faso | PROMIS CS (39) | | | | | | | | | | | | |
| Ghana | GHANA (40) | | | | | | | | | | | | |
| Ghana | iLiNS−DYADG (41) | 75 | 132 | 23.5 | 0.51 (0.26, 1.01) | 0.27 | 0.27 | 25 | 70 | 25.7 | 1.40 (0.73, 2.70) | 0.12 | 0.24 |
| Kenya | WASH−B (42) | | | | | | | | | | | | |
| Madagascar | MAHAY (43) | | | | | | | | | | | | |
| Malawi | iLiNS−DYADM (44) | 59 | 121 | 23.1 | 0.66 (0.33, 1.31) | 0.27 | 0.27 | 121 | 242 | 38.0 | 0.72 (0.51, 1.00) | 0.49 | 0.39 |
| Malawi | iLiNS−DOSE (45) | 66 | 36 | 36.1 | 0.62 (0.37, 1.05) | 0.46 | 0.46 | 115 | 50 | 50.0 | 0.62 (0.43, 0.90) | 0.39 | 0.37 |
| Mali | PROMIS CS (46) | | | | | | | | | | | | |
| Zimbabwe | SHINE (HIV−) (47) | | | | | | | | | | | | |
| Zimbabwe | SHINE (HIV+) (48) | | | | | | | | | | | | |
| | | **200** | **289** | | **I² = 0.00, Tau² = 0.00** | | | **261** | **362** | | **I² = 0.56, Tau² = 0.10** | | |
| **Fixed** | | | | | **0.60 (0.42, 0.86)** | | | | | | **0.74 (0.58, 0.93)** | | |
| **Random** | | | | | **0.60 (0.42, 0.86)** | | | | | | **0.80 (0.51, 1.25)** | | |

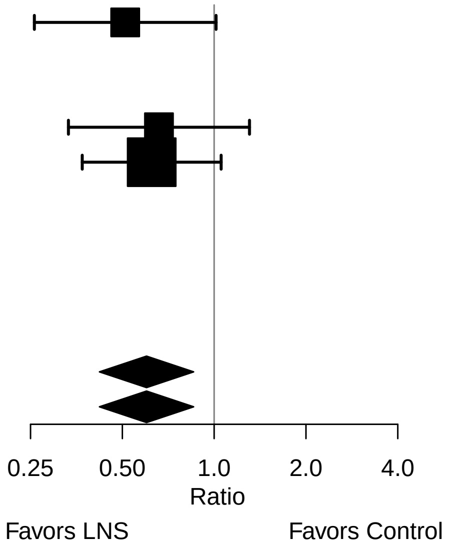

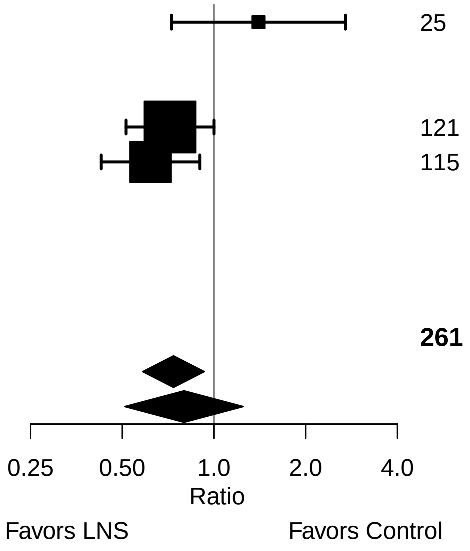

**Supplemental figure 6O: Elevated zinc protoporphyrin prevalence ratio**

#### 6O8: Stratified by Child high-dose vitamin A supplementation (insufficient comparisons)

#### Supplemental figure 6O: Elevated zinc protoporphyrin prevalence ratio

##### 6O9: Stratified by Child inflammation

**P−for−interaction = 0.980**
**Ratio of PRs = 1.00 (0.76, 1.30)**

| Country | Trial | LNS N | Control N | Control Prevalence | Not inflamed PR (95% CI) | Fixed W | Random W | LNS N | Control N | Control Prevalence | High AGP or CRP PR (95% CI) | Fixed W | Random W |
|---|---|---|---|---|---|---|---|---|---|---|---|---|---|
| Bangladesh | JiVitA−4 (35) | | | | | | | | | | | | |
| Bangladesh | RDNS (36) | | | | | | | | | | | | |
| Bangladesh | WASH−B (37) | | | | | | | | | | | | |
| Burkina Faso | iLiNS−Zinc (38) | 123 | 30 | 78.8 | 0.68 (0.51, 0.92) | 0.59 | 0.59 | 189 | 65 | 73.6 | 0.73 (0.58, 0.92) | 0.50 | 0.50 |
| Burkina Faso | PROMIS CS (39) | | | | | | | | | | | | |
| Ghana | GHANA (40) | | | | | | | | | | | | |
| Ghana | iLiNS−DYADG (41) | 45 | 120 | 25.0 | 0.62 (0.29, 1.31) | 0.09 | 0.09 | 55 | 82 | 23.2 | 0.86 (0.45, 1.67) | 0.06 | 0.06 |
| Kenya | WASH−B (42) | | | | | | | | | | | | |
| Madagascar | MAHAY (43) | | | | | | | | | | | | |
| Malawi | iLiNS−DYADM (44) | 53 | 119 | 29.4 | 0.83 (0.48, 1.44) | 0.17 | 0.17 | 137 | 256 | 35.5 | 0.66 (0.47, 0.93) | 0.22 | 0.22 |
| Malawi | iLiNS−DOSE (45) | 62 | 26 | 42.3 | 0.69 (0.38, 1.26) | 0.14 | 0.14 | 119 | 61 | 45.9 | 0.58 (0.41, 0.82) | 0.22 | 0.22 |
| Mali | PROMIS CS (46) | | | | | | | | | | | | |
| Zimbabwe | SHINE (HIV−) (47) | | | | | | | | | | | | |
| Zimbabwe | SHINE (HIV+) (48) | | | | | | | | | | | | |
| | | **283** | **295** | | **I² = 0.00, Tau² = 0.00** | | | **500** | **464** | | **I² = 0.00, Tau² = 0.00** | | |
| **Fixed** | | | | | **0.70 (0.56, 0.88)** | | | | | | **0.69 (0.58, 0.81)** | | |
| **Random** | | | | | **0.70 (0.56, 0.88)** | | | | | | **0.69 (0.58, 0.81)** | | |

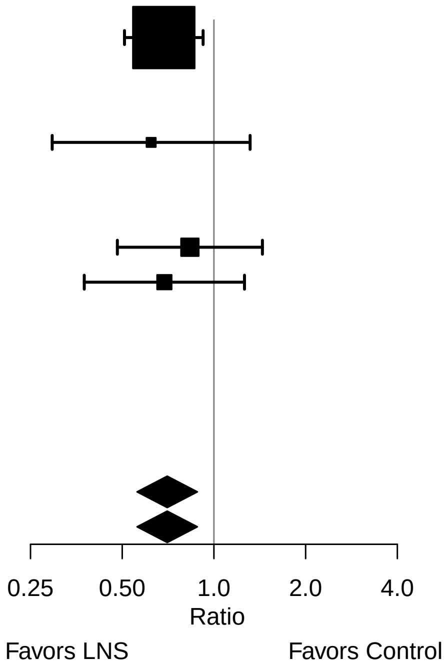

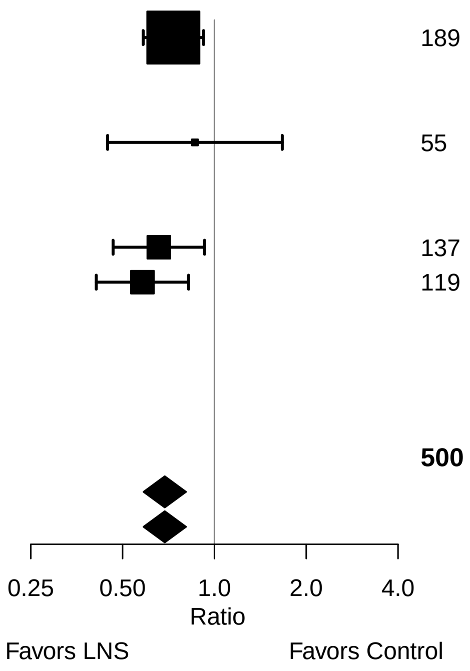

### Supplemental figure 6P: Elevated zinc protoporphyrin prevalence difference

#### 6P1: Stratified by Maternal BMI

**P−for−interaction = 0.612**
**Difference in PDs = 0.03 (−0.08, 0.14)**

| | | At least 20 kg/m² | | | | | | Less than 20 kg/m² | | | | | |
|---|---|---|---|---|---|---|---|---|---|---|---|---|---|
| Country | Trial | LNS N | Control N | Control Prevalence | PD (95% CI) | Fixed W | Random W | LNS N | Control N | Control Prevalence | PD (95% CI) | Fixed W | Random W |
| Bangladesh | JiVitA−4 (35) | | | | | | | | | | | | |
| Bangladesh | RDNS (36) | | | | | | | | | | | | |
| Bangladesh | WASH−B (37) | | | | | | | | | | | | |
| Burkina Faso | iLiNS−Zinc (38) | 208 | 55 | 73.8 | −0.20 (−0.36, −0.04) | 0.20 | 0.20 | 104 | 40 | 77.3 | −0.23 (−0.37, −0.10) | 0.39 | 0.38 |
| Burkina Faso | PROMIS CS (39) | | | | | | | | | | | | |
| Ghana | GHANA (40) | | | | | | | | | | | | |
| Ghana | iLiNS−DYADG (41) | | | | | | | | | | | | |
| Kenya | WASH−B (42) | | | | | | | | | | | | |
| Madagascar | MAHAY (43) | | | | | | | | | | | | |
| Malawi | iLiNS−DYADM (44) | 111 | 222 | 36.5 | −0.13 (−0.24, −0.02) | 0.47 | 0.47 | 79 | 151 | 29.1 | −0.05 (−0.17, 0.07) | 0.46 | 0.41 |
| Malawi | iLiNS−DOSE (45) | 127 | 65 | 46.2 | −0.17 (−0.30, −0.05) | 0.33 | 0.33 | 53 | 22 | 40.9 | −0.19 (−0.40, 0.02) | 0.15 | 0.22 |
| Mali | PROMIS CS (46) | | | | | | | | | | | | |
| Zimbabwe | SHINE (HIV−) (47) | | | | | | | | | | | | |
| Zimbabwe | SHINE (HIV+) (48) | | | | | | | | | | | | |
| | | **446** | **342** | | **I² = 0.00, Tau² = 0.00** | | | **236** | **213** | | **I² = 0.53, Tau² = 0.00** | | |
| **Fixed** | | | | | **−0.16 (−0.23, −0.09)** | | | | | | **−0.14 (−0.23, −0.06)** | | |
| **Random** | | | | | **−0.16 (−0.23, −0.09)** | | | | | | **−0.15 (−0.27, −0.03)** | | |

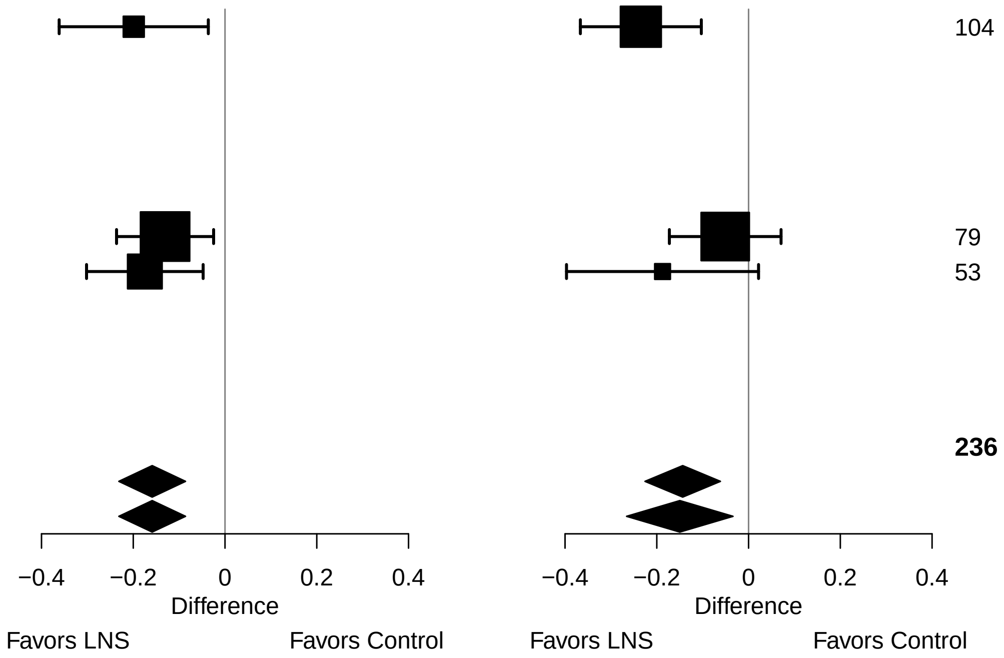

**Supplemental figure 6P: Elevated zinc protoporphyrin prevalence difference**

#### 6P2: Stratified by Maternal age

**P−for−interaction = 0.689**
**Difference in PDs = −0.02 (−0.12, 0.08)**

| Country | Trial | At least 25 y: LNS N | Control N | Control Prevalence | PD (95% CI) | Fixed W | Random W | Less than 25 y: LNS N | Control N | Control Prevalence | PD (95% CI) | Fixed W | Random W |
|---|---|---|---|---|---|---|---|---|---|---|---|---|---|
| Bangladesh | JiVitA−4 (35) | | | | | | | | | | | | |
| Bangladesh | RDNS (36) | | | | | | | | | | | | |
| Bangladesh | WASH−B (37) | | | | | | | | | | | | |
| Burkina Faso | iLiNS−Zinc (38) | 175 | 50 | 74.1 | −0.24 (−0.42, −0.07) | 0.12 | 0.17 | 137 | 45 | 76.5 | −0.17 (−0.37, 0.03) | 0.16 | 0.16 |
| Burkina Faso | PROMIS CS (39) | | | | | | | | | | | | |
| Ghana | GHANA (40) | | | | | | | | | | | | |
| Ghana | iLiNS−DYADG (41) | 65 | 128 | 16.4 | −0.04 (−0.15, 0.07) | 0.35 | 0.30 | 35 | 74 | 37.8 | −0.09 (−0.29, 0.10) | 0.17 | 0.17 |
| Kenya | WASH−B (42) | | | | | | | | | | | | |
| Madagascar | MAHAY (43) | | | | | | | | | | | | |
| Malawi | iLiNS−DYADM (44) | 88 | 192 | 28.1 | −0.07 (−0.18, 0.05) | 0.32 | 0.29 | 102 | 183 | 39.3 | −0.14 (−0.25, −0.02) | 0.47 | 0.47 |
| Malawi | iLiNS−DOSE (45) | 95 | 48 | 41.7 | −0.18 (−0.32, −0.04) | 0.21 | 0.23 | 83 | 39 | 48.7 | −0.17 (−0.34, 0.01) | 0.21 | 0.21 |
| Mali | PROMIS CS (46) | | | | | | | | | | | | |
| Zimbabwe | SHINE (HIV−) (47) | | | | | | | | | | | | |
| Zimbabwe | SHINE (HIV+) (48) | | | | | | | | | | | | |
| | | **423** | **418** | | **I² = 0.43, Tau² = 0.00** | | | **357** | **341** | | **I² = 0.00, Tau² = 0.00** | | |
| **Fixed** | | | | | **−0.10 (−0.17, −0.04)** | | | | | | **−0.14 (−0.22, −0.06)** | | |
| **Random** | | | | | **−0.11 (−0.20, −0.03)** | | | | | | **−0.14 (−0.22, −0.06)** | | |

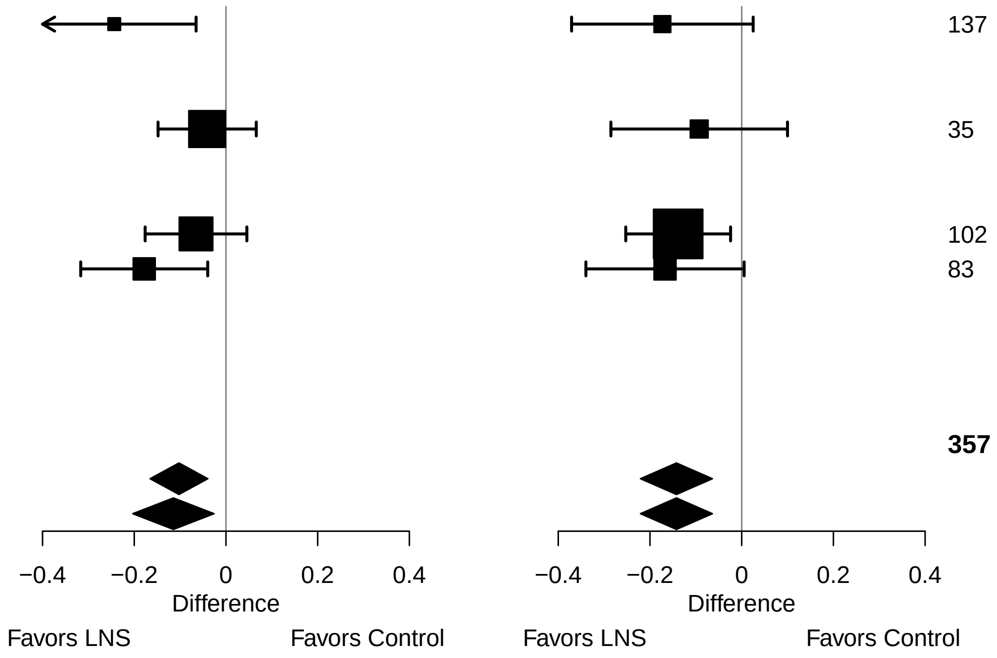

**Supplemental figure 6P: Elevated zinc protoporphyrin prevalence difference**

#### 6P3: Stratified by Maternal education

**P−for−interaction = 0.080**
**Difference in PDs = 0.13 (−0.02, 0.27)**

| Country | Trial | Primary or greater: LNS N | Control N | Control Prevalence | PD (95% CI) | Fixed W | Random W | Incomplete or no formal: LNS N | Control N | Control Prevalence | PD (95% CI) | Fixed W | Random W |
|---|---|---|---|---|---|---|---|---|---|---|---|---|---|
| Bangladesh | JiVitA−4 (35) | | | | | | | | | | | | |
| Bangladesh | RDNS (36) | | | | | | | | | | | | |
| Bangladesh | WASH−B (37) | | | | | | | | | | | | |
| Burkina Faso | iLiNS−Zinc (38) | | | | | | | | | | | | |
| Burkina Faso | PROMIS CS (39) | | | | | | | | | | | | |
| Ghana | GHANA (40) | | | | | | | | | | | | |
| Ghana | iLiNS−DYADG (41) | 83 | 159 | 25.2 | −0.09 (−0.20, 0.01) | 0.65 | 0.60 | 17 | 43 | 20.9 | 0.08 (−0.16, 0.33) | 0.08 | 0.12 |
| Kenya | WASH−B (42) | | | | | | | | | | | | |
| Madagascar | MAHAY (43) | | | | | | | | | | | | |
| Malawi | iLiNS−DYADM (44) | 31 | 53 | 45.3 | −0.16 (−0.38, 0.05) | 0.17 | 0.19 | 158 | 320 | 31.6 | −0.09 (−0.18, −0.01) | 0.63 | 0.55 |
| Malawi | iLiNS−DOSE (45) | 48 | 26 | 53.8 | −0.28 (−0.49, −0.07) | 0.18 | 0.21 | 130 | 61 | 41.0 | −0.12 (−0.25, 0.00) | 0.29 | 0.33 |
| Mali | PROMIS CS (46) | | | | | | | | | | | | |
| Zimbabwe | SHINE (HIV−) (47) | | | | | | | | | | | | |
| Zimbabwe | SHINE (HIV+) (48) | | | | | | | | | | | | |
| | | **162** | **238** | | **I² = 0.16, Tau² = 0.00** | | | **305** | **424** | | **I² = 0.13, Tau² = 0.00** | | |
| **Fixed** | | | | | **−0.14 (−0.23, −0.05)** | | | | | | **−0.09 (−0.16, −0.02)** | | |
| **Random** | | | | | **−0.15 (−0.25, −0.05)** | | | | | | **−0.08 (−0.17, 0.00)** | | |

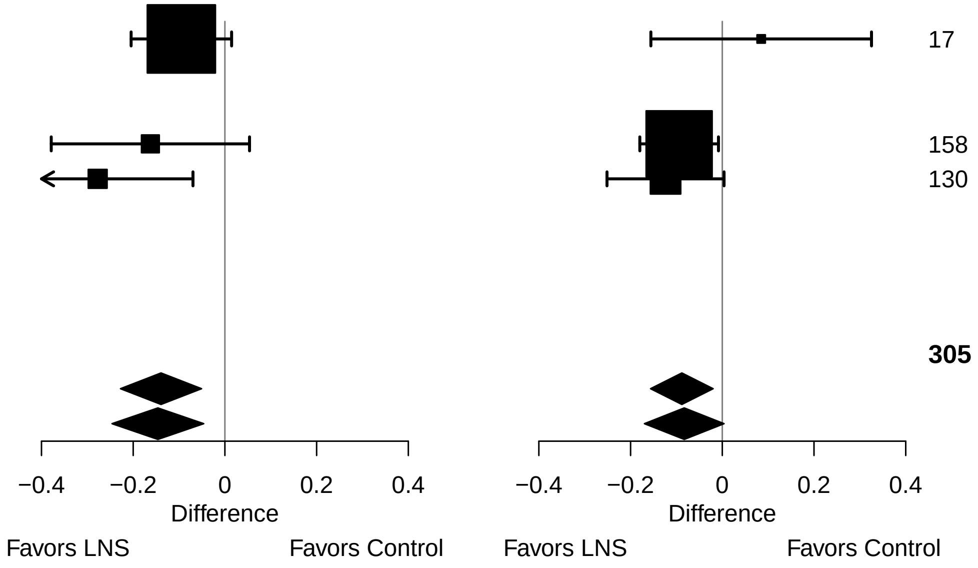

#### Supplemental figure 6P: Elevated zinc protoporphyrin prevalence difference

##### 6P4: Stratified by Child sex

**P−for−interaction = 0.276**
**Difference in PDs = 0.05 (−0.04, 0.15)**

| Country | Trial | Male LNS N | Male Control N | Male Control Prevalence | Male PD (95% CI) | Male Fixed W | Male Random W | Female LNS N | Female Control N | Female Control Prevalence | Female PD (95% CI) | Female Fixed W | Female Random W |
|---|---|---|---|---|---|---|---|---|---|---|---|---|---|
| Bangladesh | JiVitA−4 (35) | | | | | | | | | | | | |
| Bangladesh | RDNS (36) | | | | | | | | | | | | |
| Bangladesh | WASH−B (37) | | | | | | | | | | | | |
| Burkina Faso | iLiNS−Zinc (38) | 149 | 49 | 81.5 | −0.26 (−0.41, −0.10) | 0.20 | 0.20 | 163 | 46 | 68.6 | −0.17 (−0.27, −0.06) | 0.37 | 0.36 |
| Burkina Faso | PROMIS CS (39) | | | | | | | | | | | | |
| Ghana | GHANA (40) | | | | | | | | | | | | |
| Ghana | iLiNS−DYADG (41) | 63 | 95 | 26.3 | −0.07 (−0.21, 0.06) | 0.26 | 0.26 | 36 | 106 | 21.7 | −0.05 (−0.20, 0.10) | 0.16 | 0.16 |
| Kenya | WASH−B (42) | | | | | | | | | | | | |
| Madagascar | MAHAY (43) | | | | | | | | | | | | |
| Malawi | iLiNS−DYADM (44) | 95 | 183 | 37.2 | −0.14 (−0.26, −0.02) | 0.36 | 0.35 | 95 | 192 | 30.2 | −0.06 (−0.17, 0.05) | 0.31 | 0.30 |
| Malawi | iLiNS−DOSE (45) | 86 | 46 | 43.5 | −0.16 (−0.32, 0.00) | 0.19 | 0.19 | 95 | 41 | 46.3 | −0.17 (−0.32, −0.02) | 0.17 | 0.17 |
| Mali | PROMIS CS (46) | | | | | | | | | | | | |
| Zimbabwe | SHINE (HIV−) (47) | | | | | | | | | | | | |
| Zimbabwe | SHINE (HIV+) (48) | | | | | | | | | | | | |
| | | **393** | **373** | | **I² = 0.05, Tau² = 0.00** | | | **389** | **385** | | **I² = 0.04, Tau² = 0.00** | | |
| **Fixed** | | | | | **−0.15 (−0.22, −0.08)** | | | | | | **−0.12 (−0.18, −0.05)** | | |
| **Random** | | | | | **−0.15 (−0.22, −0.08)** | | | | | | **−0.12 (−0.18, −0.05)** | | |

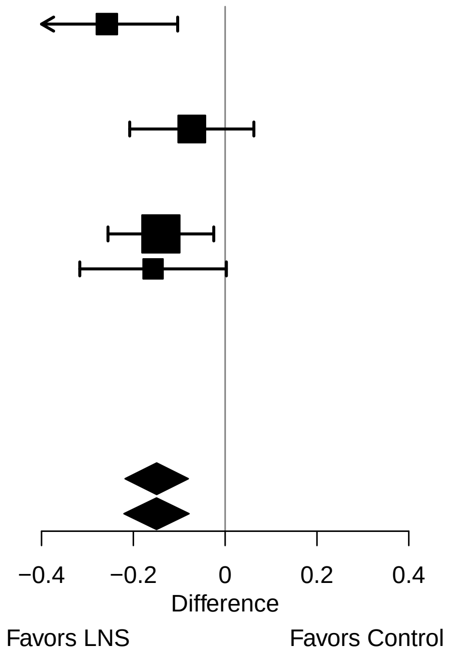

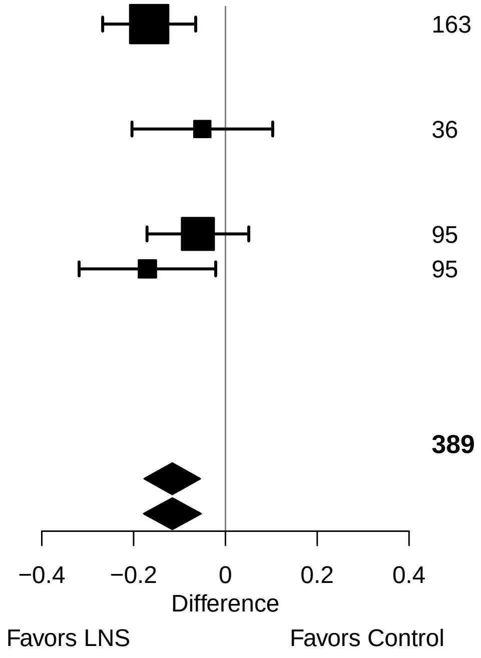

##### Supplemental figure 6P: Elevated zinc protoporphyrin prevalence difference

#### 6P5: Stratified by Child birth order

**P−for−interaction = 0.462**
**Difference in PDs = 0.05 (−0.08, 0.18)**

| Country | Trial | Later born LNS N | Later born Control N | Later born Control Prevalence | Later born PD (95% CI) | Later born Fixed W | Later born Random W | Firstborn LNS N | Firstborn Control N | Firstborn Control Prevalence | Firstborn PD (95% CI) | Firstborn Fixed W | Firstborn Random W |
|---|---|---|---|---|---|---|---|---|---|---|---|---|---|
| Bangladesh | JiVitA−4 (35) | | | | | | | | | | | | |
| Bangladesh | RDNS (36) | | | | | | | | | | | | |
| Bangladesh | WASH−B (37) | | | | | | | | | | | | |
| Burkina Faso | iLiNS−Zinc (38) | 246 | 71 | 74.7 | −0.23 (−0.35, −0.12) | 0.29 | 0.32 | 66 | 24 | 76.7 | −0.13 (−0.29, 0.04) | 0.45 | 0.45 |
| Burkina Faso | PROMIS CS (39) | | | | | | | | | | | | |
| Ghana | GHANA (40) | | | | | | | | | | | | |
| Ghana | iLiNS−DYADG (41) | | | | | | | | | | | | |
| Kenya | WASH−B (42) | | | | | | | | | | | | |
| Madagascar | MAHAY (43) | | | | | | | | | | | | |
| Malawi | iLiNS−DYADM (44) | 152 | 299 | 33.4 | −0.09 (−0.18, 0.00) | 0.47 | 0.40 | 38 | 75 | 34.7 | −0.14 (−0.31, 0.04) | 0.39 | 0.39 |
| Malawi | iLiNS−DOSE (45) | 119 | 59 | 45.8 | −0.21 (−0.34, −0.08) | 0.23 | 0.28 | 33 | 19 | 36.8 | −0.01 (−0.28, 0.27) | 0.17 | 0.17 |
| Mali | PROMIS CS (46) | | | | | | | | | | | | |
| Zimbabwe | SHINE (HIV−) (47) | | | | | | | | | | | | |
| Zimbabwe | SHINE (HIV+) (48) | | | | | | | | | | | | |
| | | **517** | **429** | | **I² = 0.55, Tau² = 0.00** | | | **137** | **118** | | **I² = 0.00, Tau² = 0.00** | | |
| **Fixed** | | | | | **−0.16 (−0.22, −0.10)** | | | | | | **−0.11 (−0.22, 0.00)** | | |
| **Random** | | | | | **−0.17 (−0.26, −0.08)** | | | | | | **−0.11 (−0.22, 0.00)** | | |

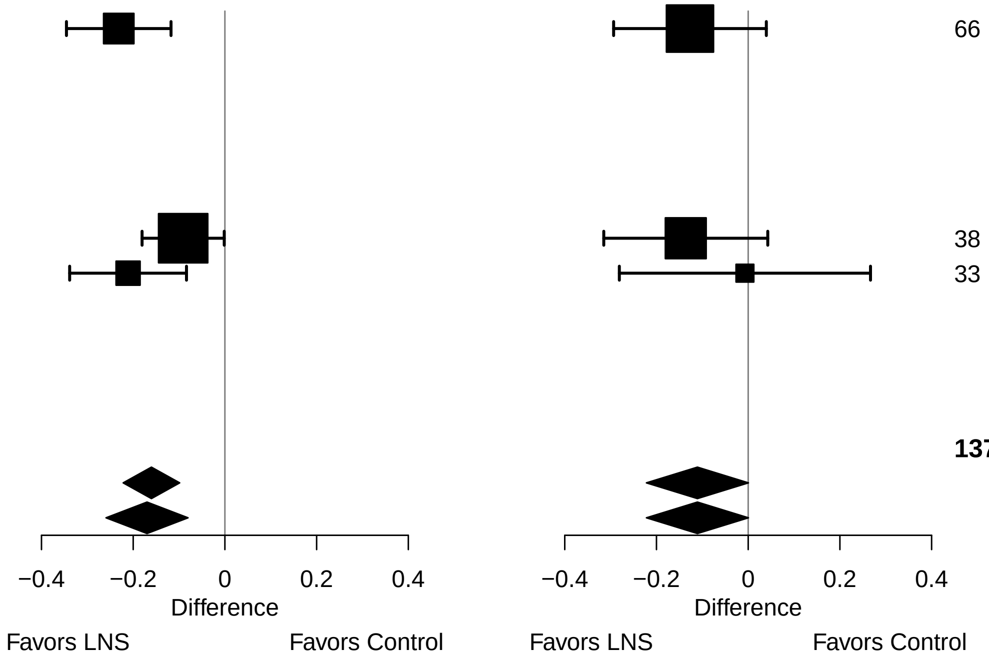

**Supplemental figure 6P: Elevated zinc protoporphyrin prevalence difference**

#### 6P6: Stratified by Child baseline acute malnutrition (insufficient comparisons)

#### Supplemental figure 6P: Elevated zinc protoporphyrin prevalence difference

### 6P7: Stratified by Child baseline anemia

**P−for−interaction = 0.663**
**Difference in PDs = 0.03 (−0.09, 0.14)**

| Country | Trial | LNS N | Control N | Control Prevalence | Not anemic PD (95% CI) | Fixed W | Random W | LNS N | Control N | Control Prevalence | Anemic PD (95% CI) | Fixed W | Random W |
|---|---|---|---|---|---|---|---|---|---|---|---|---|---|
| Bangladesh | JiVitA−4 (35) | | | | | | | | | | | | |
| Bangladesh | RDNS (36) | | | | | | | | | | | | |
| Bangladesh | WASH−B (37) | | | | | | | | | | | | |
| Burkina Faso | iLiNS−Zinc (38) | | | | | | | | | | | | |
| Burkina Faso | PROMIS CS (39) | | | | | | | | | | | | |
| Ghana | GHANA (40) | | | | | | | | | | | | |
| Ghana | iLiNS−DYADG (41) | 75 | 132 | 23.5 | −0.11 (−0.23, 0.00) | 0.45 | 0.45 | 25 | 70 | 25.7 | 0.10 (−0.10, 0.31) | 0.14 | 0.26 |
| Kenya | WASH−B (42) | | | | | | | | | | | | |
| Madagascar | MAHAY (43) | | | | | | | | | | | | |
| Malawi | iLiNS−DYADM (44) | 59 | 121 | 23.1 | −0.08 (−0.20, 0.05) | 0.35 | 0.35 | 121 | 242 | 38.0 | −0.11 (−0.21, 0.00) | 0.57 | 0.40 |
| Malawi | iLiNS−DOSE (45) | 66 | 36 | 36.1 | −0.14 (−0.31, 0.02) | 0.20 | 0.20 | 115 | 50 | 50.0 | −0.18 (−0.32, −0.03) | 0.29 | 0.34 |
| Mali | PROMIS CS (46) | | | | | | | | | | | | |
| Zimbabwe | SHINE (HIV−) (47) | | | | | | | | | | | | |
| Zimbabwe | SHINE (HIV+) (48) | | | | | | | | | | | | |
| | | **200** | **289** | | **I² = 0.00, Tau² = 0.00** | | | **261** | **362** | | **I² = 0.58, Tau² = 0.01** | | |
| **Fixed** | | | | | **−0.11 (−0.18, −0.03)** | | | | | | **−0.10 (−0.18, −0.02)** | | |
| **Random** | | | | | **−0.11 (−0.18, −0.03)** | | | | | | **−0.08 (−0.23, 0.08)** | | |

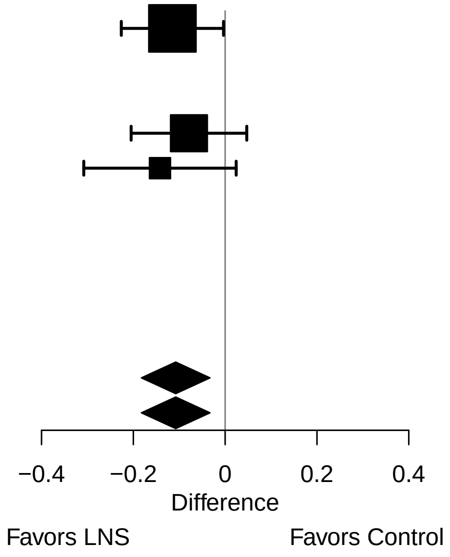

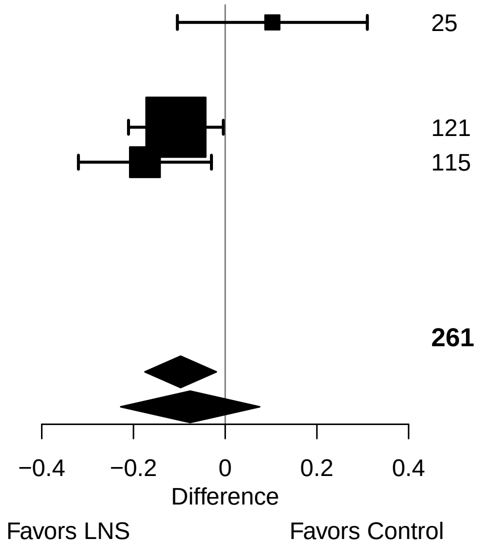

**Supplemental figure 6P: Elevated zinc protoporphyrin prevalence difference**

#### 6P8: Stratified by Child high-dose vitamin A supplementation (insufficient comparisons)

##### Supplemental figure 6P: Elevated zinc protoporphyrin prevalence difference

#### 6P9: Stratified by Child inflammation

**P−for−interaction = 0.882**
**Difference in PDs = −0.01 (−0.11, 0.09)**

| | | Not inflamed | | | | | | High AGP or CRP | | | | | |
|---|---|---|---|---|---|---|---|---|---|---|---|---|---|
| Country | Trial | LNS N | Control N | Control Prevalence | PD (95% CI) | Fixed W | Random W | LNS N | Control N | Control Prevalence | PD (95% CI) | Fixed W | Random W |
| Bangladesh | JiVitA−4 (35) | | | | | | | | | | | | |
| Bangladesh | RDNS (36) | | | | | | | | | | | | |
| Bangladesh | WASH−B (37) | | | | | | | | | | | | |
| Burkina Faso | iLiNS−Zinc (38) | 123 | 30 | 78.8 | −0.24 (−0.45, −0.04) | 0.16 | 0.16 | 189 | 65 | 73.6 | −0.20 (−0.32, −0.07) | 0.22 | 0.23 |
| Burkina Faso | PROMIS CS (39) | | | | | | | | | | | | |
| Ghana | GHANA (40) | | | | | | | | | | | | |
| Ghana | iLiNS−DYADG (41) | 45 | 120 | 25.0 | −0.09 (−0.24, 0.05) | 0.34 | 0.34 | 55 | 82 | 23.2 | −0.03 (−0.17, 0.11) | 0.17 | 0.19 |
| Kenya | WASH−B (42) | | | | | | | | | | | | |
| Madagascar | MAHAY (43) | | | | | | | | | | | | |
| Malawi | iLiNS−DYADM (44) | 53 | 119 | 29.4 | −0.05 (−0.19, 0.10) | 0.33 | 0.33 | 137 | 256 | 35.5 | −0.12 (−0.22, −0.03) | 0.39 | 0.35 |
| Malawi | iLiNS−DOSE (45) | 62 | 26 | 42.3 | −0.14 (−0.34, 0.07) | 0.17 | 0.17 | 119 | 61 | 45.9 | −0.19 (−0.31, −0.06) | 0.22 | 0.23 |
| Mali | PROMIS CS (46) | | | | | | | | | | | | |
| Zimbabwe | SHINE (HIV−) (47) | | | | | | | | | | | | |
| Zimbabwe | SHINE (HIV+) (48) | | | | | | | | | | | | |
| | | **283** | **295** | | **I² = 0.00, Tau² = 0.00** | | | **500** | **464** | | **I² = 0.16, Tau² = 0.00** | | |
| **Fixed** | | | | | **−0.11 (−0.19, −0.03)** | | | | | | **−0.14 (−0.20, −0.08)** | | |
| **Random** | | | | | **−0.11 (−0.19, −0.03)** | | | | | | **−0.14 (−0.20, −0.07)** | | |

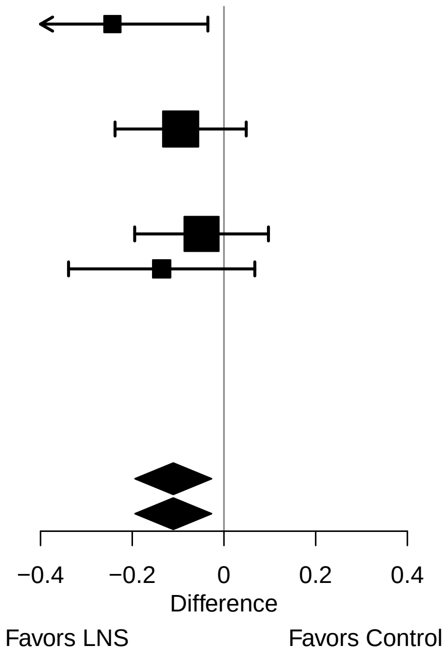

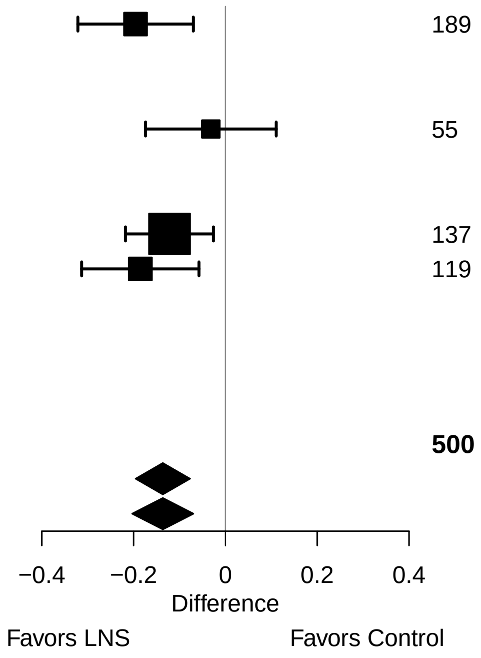

### Supplemental figure 6Q: Geometric mean ratio of plasma zinc concentration

#### 6Q1: Stratified by Maternal BMI (insufficient comparisons)

**Supplemental figure 6Q: Geometric mean ratio of plasma zinc concentration**

#### 6Q2: Stratified by Maternal age

**P−for−interaction = 0.003**
**Ratio of GMRs = 0.94 (0.90, 0.98)**

| | | At least 25 y | | | | | | Less than 25 y | | | | | |
|---|---|---|---|---|---|---|---|---|---|---|---|---|---|
| Country | Trial | LNS N | Control N | Control Median | GMR (95% CI) | Fixed W | Random W | LNS N | Control N | Control Median | GMR (95% CI) | Fixed W | Random W |
| Bangladesh | JiVitA−4 (35) | 201 | 70 | 86.1 | 1.00 (0.92, 1.08) | 0.12 | 0.12 | 248 | 73 | 91.3 | 0.93 (0.87, 1.00) | 0.31 | 0.35 |
| Bangladesh | RDNS (36) | | | | | | | | | | | | |
| Bangladesh | WASH−B (37) | | | | | | | | | | | | |
| Burkina Faso | iLiNS−Zinc (38) | 177 | 50 | 67.1 | 1.03 (1.00, 1.06) | 0.73 | 0.73 | 137 | 46 | 69.9 | 0.98 (0.93, 1.03) | 0.64 | 0.57 |
| Burkina Faso | PROMIS CS (39) | | | | | | | | | | | | |
| Ghana | GHANA (40) | 44 | 44 | 61.0 | 1.03 (0.96, 1.11) | 0.15 | 0.15 | 14 | 19 | 62.9 | 0.87 (0.74, 1.03) | 0.06 | 0.08 |
| Ghana | iLiNS−DYADG (41) | | | | | | | | | | | | |
| Kenya | WASH−B (42) | | | | | | | | | | | | |
| Madagascar | MAHAY (43) | | | | | | | | | | | | |
| Malawi | iLiNS−DYADM (44) | | | | | | | | | | | | |
| Malawi | iLiNS−DOSE (45) | | | | | | | | | | | | |
| Mali | PROMIS CS (46) | | | | | | | | | | | | |
| Zimbabwe | SHINE (HIV−) (47) | | | | | | | | | | | | |
| Zimbabwe | SHINE (HIV+) (48) | | | | | | | | | | | | |
| | | **422** | **164** | | **$I^2$ = 0.00, $Tau^2$ = 0.00** | | | **399** | **138** | | **$I^2$ = 0.21, $Tau^2$ = 0.00** | | |
| **Fixed** | | | | | **1.03 (1.00, 1.05)** | | | | | | **0.96 (0.92, 1.00)** | | |
| **Random** | | | | | **1.03 (1.00, 1.05)** | | | | | | **0.96 (0.91, 1.00)** | | |

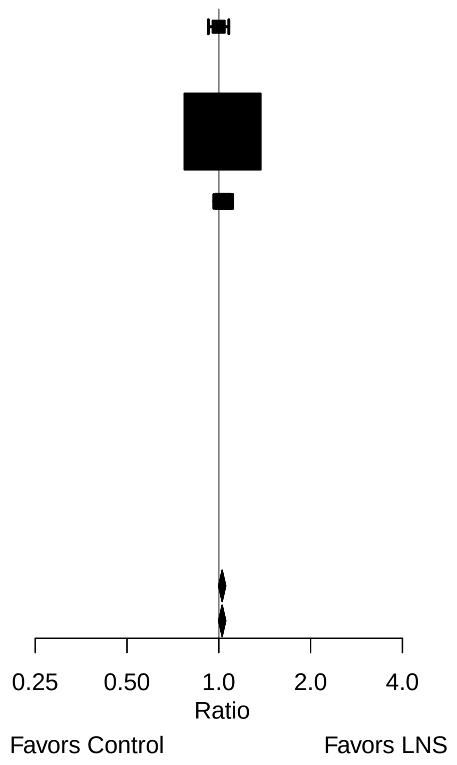

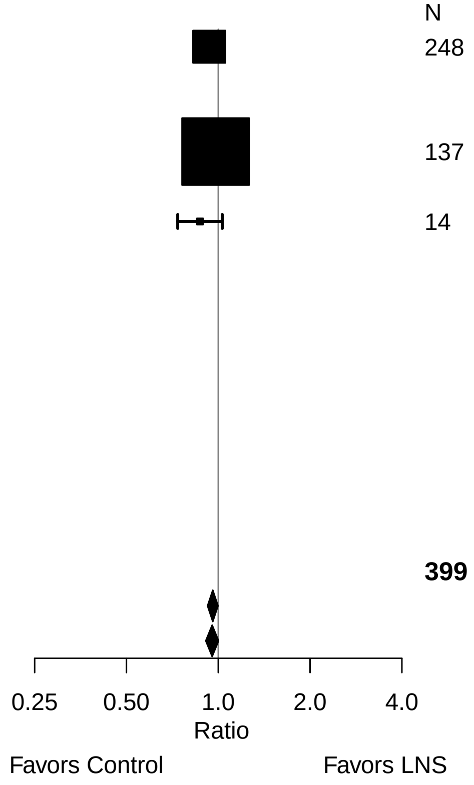

**Supplemental figure 6Q: Geometric mean ratio of plasma zinc concentration**

#### 6Q3: Stratified by Maternal education (insufficient comparisons)

**Supplemental figure 6Q: Geometric mean ratio of plasma zinc concentration**

#### 6Q4: Stratified by Child sex

**P−for−interaction = 0.728**
**Ratio of GMRs = 1.01 (0.96, 1.06)**

| Country | Trial | Male: LNS N | Male: Control N | Male: Control Median | Male: GMR (95% CI) | Male: Fixed W | Male: Random W | Female: LNS N | Female: Control N | Female: Control Median | Female: GMR (95% CI) | Female: Fixed W | Female: Random W |
|---|---|---|---|---|---|---|---|---|---|---|---|---|---|
| Bangladesh | JiVitA−4 (35) | 234 | 70 | 91.2 | 0.95 (0.88, 1.02) | 0.25 | 0.30 | 218 | 74 | 87.0 | 0.98 (0.91, 1.05) | 0.16 | 0.16 |
| Bangladesh | RDNS (36) | | | | | | | | | | | | |
| Bangladesh | WASH−B (37) | | | | | | | | | | | | |
| Burkina Faso | iLiNS−Zinc (38) | 149 | 49 | 67.8 | 1.00 (0.95, 1.05) | 0.57 | 0.47 | 165 | 47 | 69.8 | 1.01 (0.98, 1.05) | 0.76 | 0.76 |
| Burkina Faso | PROMIS CS (39) | | | | | | | | | | | | |
| Ghana | GHANA (40) | 34 | 31 | 58.6 | 1.05 (0.96, 1.15) | 0.17 | 0.23 | 24 | 38 | 61.2 | 0.96 (0.86, 1.07) | 0.08 | 0.08 |
| Ghana | iLiNS−DYADG (41) | | | | | | | | | | | | |
| Kenya | WASH−B (42) | | | | | | | | | | | | |
| Madagascar | MAHAY (43) | | | | | | | | | | | | |
| Malawi | iLiNS−DYADM (44) | | | | | | | | | | | | |
| Malawi | iLiNS−DOSE (45) | | | | | | | | | | | | |
| Mali | PROMIS CS (46) | | | | | | | | | | | | |
| Zimbabwe | SHINE (HIV−) (47) | | | | | | | | | | | | |
| Zimbabwe | SHINE (HIV+) (48) | | | | | | | | | | | | |
| | | **417** | **150** | | **I² = 0.30, Tau² = 0.00** | | | **407** | **159** | | **I² = 0.00, Tau² = 0.00** | | |
| **Fixed** | | | | | **0.99 (0.96, 1.03)** | | | | | | **1.00 (0.97, 1.03)** | | |
| **Random** | | | | | **0.99 (0.95, 1.04)** | | | | | | **1.00 (0.97, 1.03)** | | |

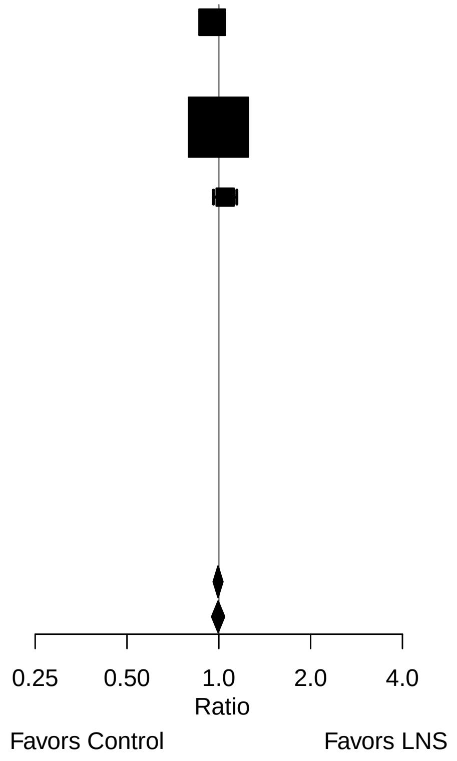

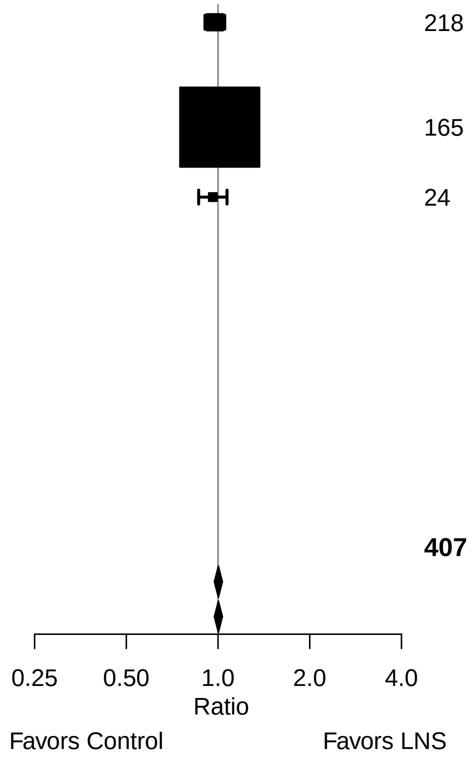

#### Supplemental figure 6Q: Geometric mean ratio of plasma zinc concentration

##### 6Q5: Stratified by Child birth order

**P−for−interaction = 0.131**
**Ratio of GMRs = 0.96 (0.91, 1.01)**

| | | **Later born** | | | | | | **Firstborn** | | | | | |
|---|---|---|---|---|---|---|---|---|---|---|---|---|---|
| | | **LNS** | **Control** | **Control** | **GMR** | **Fixed** | **Random** | **LNS** | **Control** | **Control** | **GMR** | **Fixed** | **Random** |
| Country | Trial | N | N | Median | (95% CI) | W | W | N | N | Median | (95% CI) | W | W |
| Bangladesh | JiVitA−4 (35) | 113 | 32 | 86.1 | 1.00 (0.90, 1.12) | 0.08 | 0.08 | 338 | 111 | 88.8 | 0.95 (0.90, 1.01) | 0.47 | 0.47 |
| Bangladesh | RDNS (36) | | | | | | | | | | | | |
| Bangladesh | WASH−B (37) | | | | | | | | | | | | |
| Burkina Faso | iLiNS−Zinc (38) | 248 | 71 | 66.5 | 1.02 (0.98, 1.06) | 0.77 | 0.77 | 66 | 25 | 72.6 | 0.97 (0.91, 1.03) | 0.41 | 0.41 |
| Burkina Faso | PROMIS CS (39) | | | | | | | | | | | | |
| Ghana | GHANA (40) | 38 | 43 | 61.5 | 0.99 (0.91, 1.08) | 0.15 | 0.15 | 20 | 23 | 59.8 | 1.00 (0.89, 1.12) | 0.12 | 0.12 |
| Ghana | iLiNS−DYADG (41) | | | | | | | | | | | | |
| Kenya | WASH−B (42) | | | | | | | | | | | | |
| Madagascar | MAHAY (43) | | | | | | | | | | | | |
| Malawi | iLiNS−DYADM (44) | | | | | | | | | | | | |
| Malawi | iLiNS−DOSE (45) | | | | | | | | | | | | |
| Mali | PROMIS CS (46) | | | | | | | | | | | | |
| Zimbabwe | SHINE (HIV−) (47) | | | | | | | | | | | | |
| Zimbabwe | SHINE (HIV+) (48) | | | | | | | | | | | | |
| | | **399** | **146** | | **$I^2$ = 0.00, $Tau^2$ = 0.00** | | | **424** | **159** | | **$I^2$ = 0.00, $Tau^2$ = 0.00** | | |
| **Fixed** | | | | | **1.01 (0.98, 1.05)** | | | | | | **0.97 (0.93, 1.00)** | | |
| **Random** | | | | | **1.01 (0.98, 1.05)** | | | | | | **0.97 (0.93, 1.00)** | | |

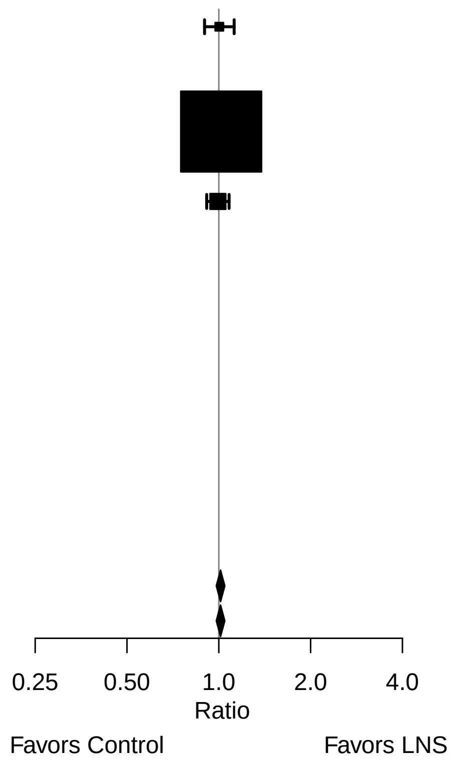

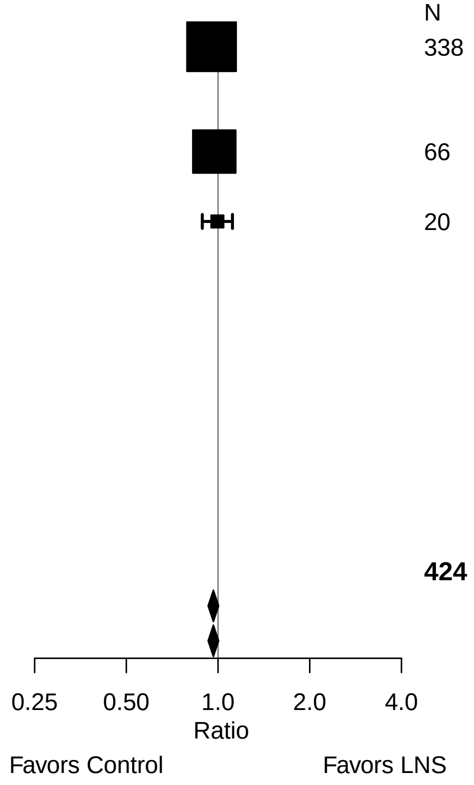

**Supplemental figure 6Q: Geometric mean ratio of plasma zinc concentration**

#### 6Q6: Stratified by Child baseline acute malnutrition (insufficient comparisons)

**Supplemental figure 6Q: Geometric mean ratio of plasma zinc concentration**

#### 6Q7: Stratified by Child baseline anemia (insufficient comparisons)

**Supplemental figure 6Q: Geometric mean ratio of plasma zinc concentration**

#### 6Q8: Stratified by Child high-dose vitamin A supplementation (insufficient comparisons)

**Supplemental figure 6Q: Geometric mean ratio of plasma zinc concentration**

#### 6Q9: Stratified by Child inflammation

**P−for−interaction = 0.425**
**Ratio of GMRs = 1.02 (0.97, 1.06)**

| | | Not inflamed | | | | | | High AGP or CRP | | | | | |
|---|---|---|---|---|---|---|---|---|---|---|---|---|---|
| Country | Trial | LNS N | Control N | Control Median | GMR (95% CI) | Fixed W | Random W | LNS N | Control N | Control Median | GMR (95% CI) | Fixed W | Random W |
| Bangladesh | JiVitA−4 (35) | 184 | 46 | 88.8 | 0.95 (0.89, 1.01) | 0.24 | 0.26 | 268 | 98 | 88.1 | 0.97 (0.91, 1.03) | 0.29 | 0.35 |
| Bangladesh | RDNS (36) | | | | | | | | | | | | |
| Bangladesh | WASH−B (37) | | | | | | | | | | | | |
| Burkina Faso | iLiNS−Zinc (38) | 124 | 31 | 68.5 | 0.99 (0.94, 1.03) | 0.58 | 0.54 | 190 | 65 | 69.0 | 1.02 (0.97, 1.06) | 0.66 | 0.56 |
| Burkina Faso | PROMIS CS (39) | | | | | | | | | | | | |
| Ghana | GHANA (40) | 48 | 54 | 59.5 | 1.03 (0.95, 1.11) | 0.18 | 0.20 | 10 | 15 | 61.5 | 0.91 (0.78, 1.06) | 0.05 | 0.09 |
| Ghana | iLiNS−DYADG (41) | | | | | | | | | | | | |
| Kenya | WASH−B (42) | | | | | | | | | | | | |
| Madagascar | MAHAY (43) | | | | | | | | | | | | |
| Malawi | iLiNS−DYADM (44) | | | | | | | | | | | | |
| Malawi | iLiNS−DOSE (45) | | | | | | | | | | | | |
| Mali | PROMIS CS (46) | | | | | | | | | | | | |
| Zimbabwe | SHINE (HIV−) (47) | | | | | | | | | | | | |
| Zimbabwe | SHINE (HIV+) (48) | | | | | | | | | | | | |
| | | **356** | **131** | | **$I^2$ = 0.12, $Tau^2$ = 0.00** | | | **468** | **178** | | **$I^2$ = 0.30, $Tau^2$ = 0.00** | | |
| **Fixed** | | | | | **0.98 (0.95, 1.02)** | | | | | | **1.00 (0.96, 1.03)** | | |
| **Random** | | | | | **0.98 (0.95, 1.02)** | | | | | | **0.99 (0.94, 1.04)** | | |

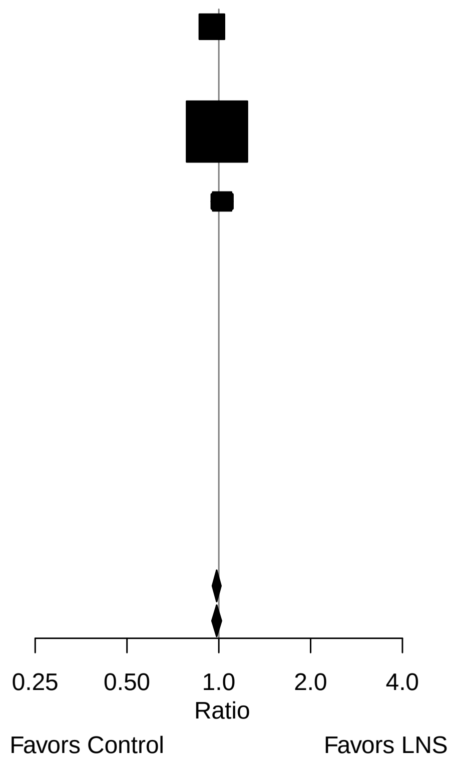

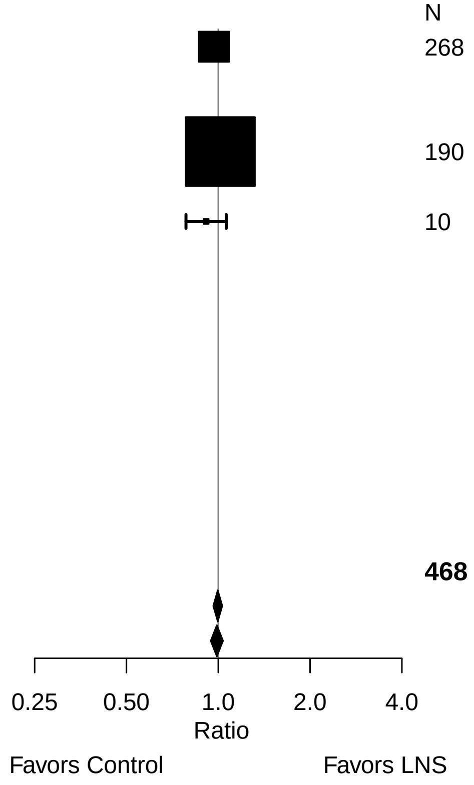

#### Supplemental figure 6R: Geometric mean ratio of retinol concentration

##### 6R1: Stratified by Maternal BMI (insufficient comparisons)

#### Supplemental figure 6R: Geometric mean ratio of retinol concentration

#### 6R2: Stratified by Maternal age

**P−for−interaction = 0.473**
**Ratio of GMRs = 0.97 (0.90, 1.05)**

| | | At least 25 y | | | | | | Less than 25 y | | | | | |
|---|---|---|---|---|---|---|---|---|---|---|---|---|---|
| | | **LNS** | **Control** | **Control** | **GMR** | **Fixed** | **Random** | **LNS** | **Control** | **Control** | **GMR** | **Fixed** | **Random** |
| Country | Trial | N | N | Median | (95% CI) | W | W | N | N | Median | (95% CI) | W | W |
| Bangladesh | JiVitA−4 (35) | 195 | 67 | 1.4 | 1.12 (1.03, 1.22) | 0.31 | 0.30 | 237 | 71 | 1.3 | 1.05 (0.97, 1.14) | 0.49 | 0.49 |
| Bangladesh | RDNS (36) | | | | | | | | | | | | |
| Bangladesh | WASH−B (37) | | | | | | | | | | | | |
| Burkina Faso | iLiNS−Zinc (38) | | | | | | | | | | | | |
| Burkina Faso | PROMIS CS (39) | | | | | | | | | | | | |
| Ghana | GHANA (40) | 47 | 47 | 1.0 | 1.13 (0.97, 1.32) | 0.10 | 0.15 | 16 | 21 | 1.2 | 0.91 (0.67, 1.24) | 0.04 | 0.04 |
| Ghana | iLiNS−DYADG (41) | 63 | 124 | 1.1 | 0.98 (0.91, 1.06) | 0.42 | 0.34 | 33 | 72 | 1.0 | 1.05 (0.93, 1.18) | 0.24 | 0.24 |
| Kenya | WASH−B (42) | | | | | | | | | | | | |
| Madagascar | MAHAY (43) | | | | | | | | | | | | |
| Malawi | iLiNS−DYADM (44) | 33 | 81 | 1.1 | 1.02 (0.91, 1.15) | 0.16 | 0.21 | 38 | 82 | 1.0 | 0.99 (0.88, 1.12) | 0.23 | 0.23 |
| Malawi | iLiNS−DOSE (45) | | | | | | | | | | | | |
| Mali | PROMIS CS (46) | | | | | | | | | | | | |
| Zimbabwe | SHINE (HIV−) (47) | | | | | | | | | | | | |
| Zimbabwe | SHINE (HIV+) (48) | | | | | | | | | | | | |
| | | **338** | **319** | | **I² = 0.52, Tau² = 0.00** | | | **324** | **246** | | **I² = 0.00, Tau² = 0.00** | | |
| **Fixed** | | | | | **1.05 (1.00, 1.10)** | | | | | | **1.03 (0.97, 1.09)** | | |
| **Random** | | | | | **1.05 (0.98, 1.13)** | | | | | | **1.03 (0.97, 1.09)** | | |

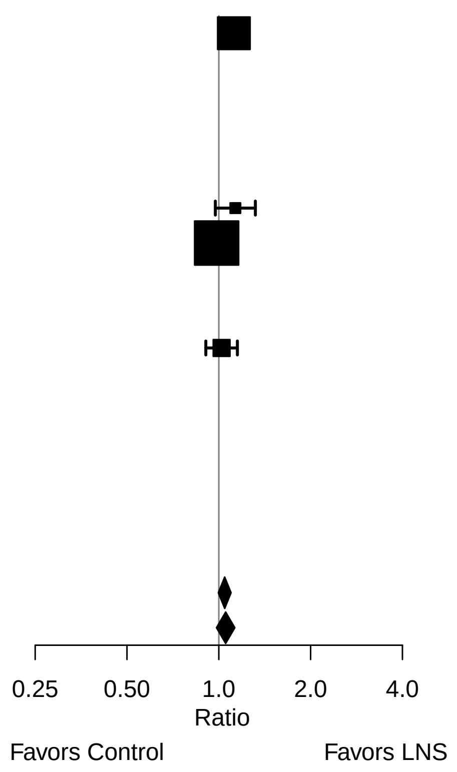

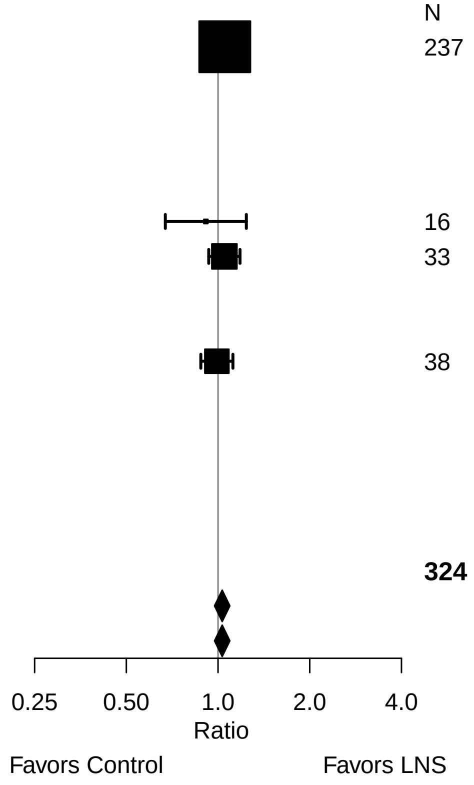

#### Supplemental figure 6R: Geometric mean ratio of retinol concentration

#### 6R3: Stratified by Maternal education

**P−for−interaction = 0.607**
**Ratio of GMRs = 0.97 (0.88, 1.08)**

| | | Primary or greater | | | | | | Incomplete or no formal | | | | | |
|---|---|---|---|---|---|---|---|---|---|---|---|---|---|
| Country | Trial | LNS N | Control N | Control Median | GMR (95% CI) | Fixed W | Random W | LNS N | Control N | Control Median | GMR (95% CI) | Fixed W | Random W |
| Bangladesh | JiVitA−4 (35) | 283 | 90 | 1.4 | 1.08 (1.00, 1.17) | 0.44 | 0.44 | 150 | 49 | 1.3 | 1.08 (0.95, 1.23) | 0.28 | 0.28 |
| Bangladesh | RDNS (36) | | | | | | | | | | | | |
| Bangladesh | WASH−B (37) | | | | | | | | | | | | |
| Burkina Faso | iLiNS−Zinc (38) | | | | | | | | | | | | |
| Burkina Faso | PROMIS CS (39) | | | | | | | | | | | | |
| Ghana | GHANA (40) | | | | | | | | | | | | |
| Ghana | iLiNS−DYADG (41) | 79 | 154 | 1.0 | 1.02 (0.95, 1.10) | 0.52 | 0.52 | 17 | 42 | 1.0 | 0.95 (0.81, 1.10) | 0.19 | 0.19 |
| Kenya | WASH−B (42) | | | | | | | | | | | | |
| Madagascar | MAHAY (43) | | | | | | | | | | | | |
| Malawi | iLiNS−DYADM (44) | 9 | 22 | 1.1 | 0.98 (0.76, 1.26) | 0.04 | 0.04 | 61 | 141 | 1.1 | 1.00 (0.91, 1.09) | 0.53 | 0.53 |
| Malawi | iLiNS−DOSE (45) | | | | | | | | | | | | |
| Mali | PROMIS CS (46) | | | | | | | | | | | | |
| Zimbabwe | SHINE (HIV−) (47) | | | | | | | | | | | | |
| Zimbabwe | SHINE (HIV+) (48) | | | | | | | | | | | | |
| | | **371** | **266** | | **I² = 0.00, Tau² = 0.00** | | | **228** | **232** | | **I² = 0.00, Tau² = 0.00** | | |
| **Fixed** | | | | | **1.05 (0.99, 1.10)** | | | | | | **1.01 (0.95, 1.08)** | | |
| **Random** | | | | | **1.05 (0.99, 1.10)** | | | | | | **1.01 (0.95, 1.08)** | | |

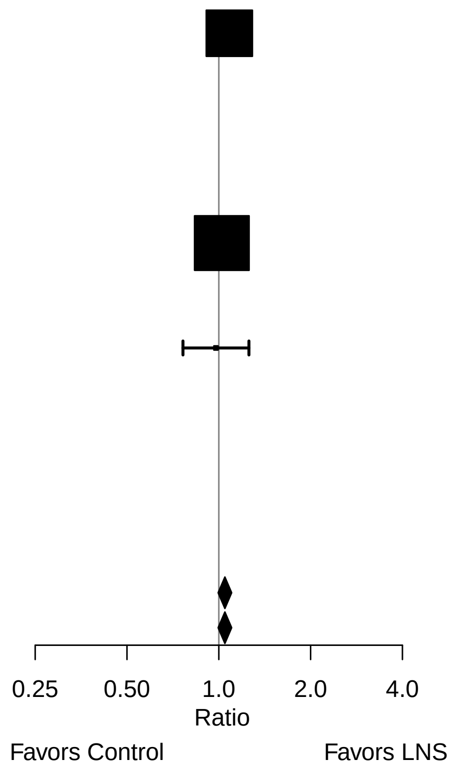

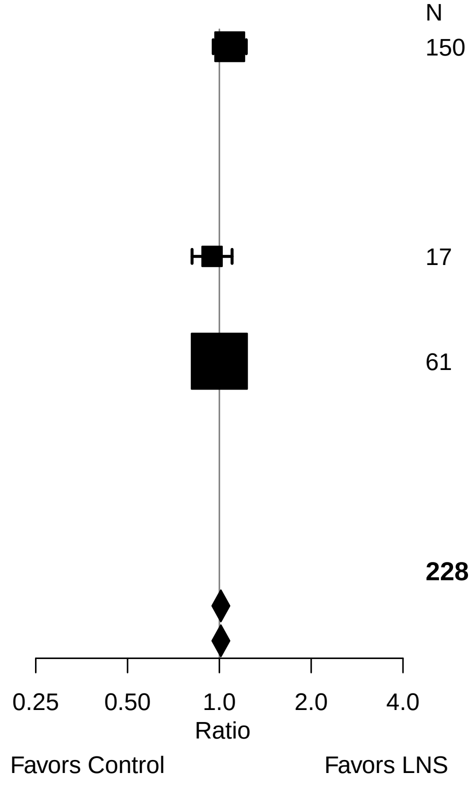

#### Supplemental figure 6R: Geometric mean ratio of retinol concentration

##### 6R4: Stratified by Child sex

**P−for−interaction = 0.315**
**Ratio of GMRs = 1.04 (0.96, 1.12)**

| Country | Trial | LNS N | Control N | Control Median | Male GMR (95% CI) | Fixed W | Random W | LNS N | Control N | Control Median | Female GMR (95% CI) | Fixed W | Random W |
|---|---|---|---|---|---|---|---|---|---|---|---|---|---|
| Bangladesh | JiVitA−4 (35) | 222 | 67 | 1.4 | 1.02 (0.93, 1.11) | 0.37 | 0.37 | 212 | 72 | 1.4 | 1.15 (1.05, 1.26) | 0.38 | 0.33 |
| Bangladesh | RDNS (36) | | | | | | | | | | | | |
| Bangladesh | WASH−B (37) | | | | | | | | | | | | |
| Burkina Faso | iLiNS−Zinc (38) | | | | | | | | | | | | |
| Burkina Faso | PROMIS CS (39) | | | | | | | | | | | | |
| Ghana | GHANA (40) | 37 | 32 | 1.1 | 1.01 (0.83, 1.23) | 0.07 | 0.07 | 26 | 42 | 1.0 | 1.13 (0.93, 1.37) | 0.08 | 0.13 |
| Ghana | iLiNS−DYADG (41) | 62 | 93 | 1.0 | 1.06 (0.97, 1.15) | 0.35 | 0.35 | 33 | 102 | 1.1 | 0.98 (0.89, 1.08) | 0.34 | 0.31 |
| Kenya | WASH−B (42) | | | | | | | | | | | | |
| Madagascar | MAHAY (43) | | | | | | | | | | | | |
| Malawi | iLiNS−DYADM (44) | 39 | 80 | 1.0 | 1.00 (0.89, 1.12) | 0.21 | 0.21 | 32 | 83 | 1.1 | 1.01 (0.89, 1.15) | 0.19 | 0.23 |
| Malawi | iLiNS−DOSE (45) | | | | | | | | | | | | |
| Mali | PROMIS CS (46) | | | | | | | | | | | | |
| Zimbabwe | SHINE (HIV−) (47) | | | | | | | | | | | | |
| Zimbabwe | SHINE (HIV+) (48) | | | | | | | | | | | | |
| | | **360** | **272** | | **I² = 0.00, Tau² = 0.00** | | | **303** | **299** | | **I² = 0.55, Tau² = 0.00** | | |
| **Fixed** | | | | | **1.03 (0.97, 1.08)** | | | | | | **1.06 (1.01, 1.13)** | | |
| **Random** | | | | | **1.03 (0.97, 1.08)** | | | | | | **1.06 (0.98, 1.15)** | | |

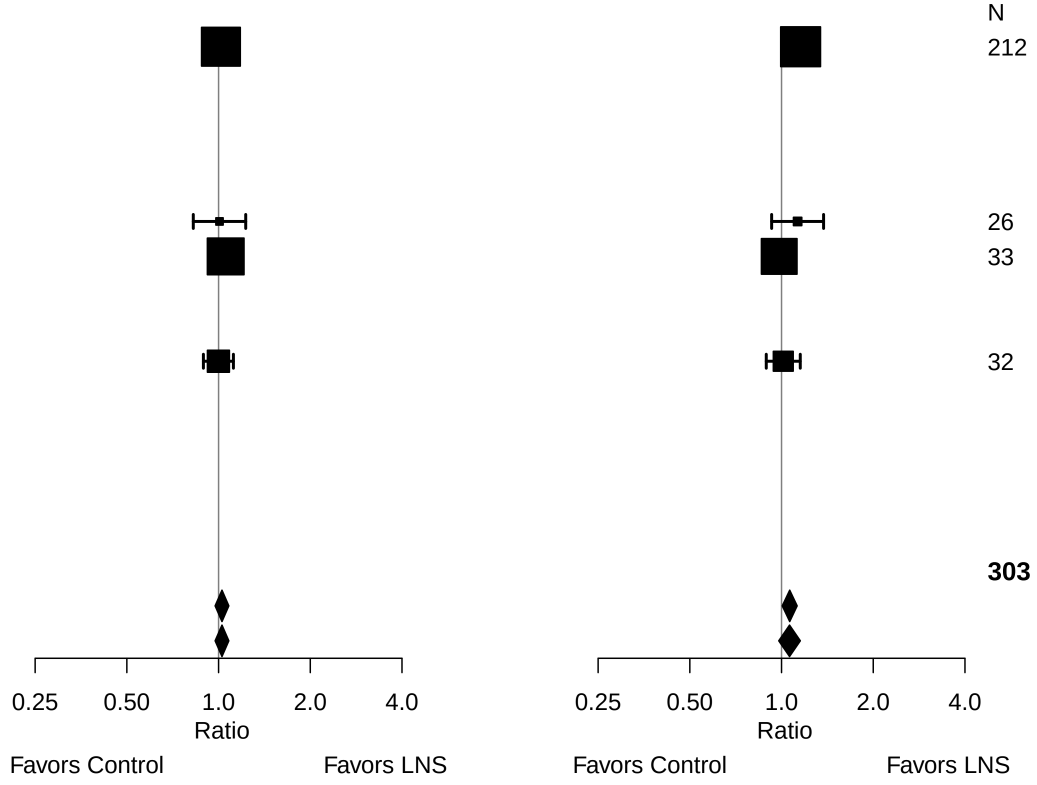

#### Supplemental figure 6R: Geometric mean ratio of retinol concentration

#### 6R5: Stratified by Child birth order

**P−for−interaction = 0.862**
**Ratio of GMRs = 0.99 (0.91, 1.09)**

| | | Later born | | | | | | Firstborn | | | | | |
|---|---|---|---|---|---|---|---|---|---|---|---|---|---|
| Country | Trial | LNS N | Control N | Control Median | GMR (95% CI) | Fixed W | Random W | LNS N | Control N | Control Median | GMR (95% CI) | Fixed W | Random W |
| Bangladesh | JiVitA−4 (35) | 106 | 31 | 1.5 | 1.04 (0.89, 1.21) | 0.11 | 0.16 | 327 | 107 | 1.3 | 1.10 (1.02, 1.17) | 0.68 | 0.57 |
| Bangladesh | RDNS (36) | | | | | | | | | | | | |
| Bangladesh | WASH−B (37) | | | | | | | | | | | | |
| Burkina Faso | iLiNS−Zinc (38) | | | | | | | | | | | | |
| Burkina Faso | PROMIS CS (39) | | | | | | | | | | | | |
| Ghana | GHANA (40) | 42 | 45 | 1.0 | 1.20 (1.02, 1.40) | 0.11 | 0.16 | 21 | 26 | 1.1 | 0.87 (0.67, 1.13) | 0.05 | 0.07 |
| Ghana | iLiNS−DYADG (41) | 68 | 137 | 1.0 | 1.01 (0.94, 1.08) | 0.47 | 0.37 | 28 | 59 | 1.0 | 1.01 (0.89, 1.14) | 0.20 | 0.26 |
| Kenya | WASH−B (42) | | | | | | | | | | | | |
| Madagascar | MAHAY (43) | | | | | | | | | | | | |
| Malawi | iLiNS−DYADM (44) | 56 | 129 | 1.1 | 1.00 (0.91, 1.09) | 0.31 | 0.31 | 15 | 34 | 1.1 | 1.04 (0.83, 1.30) | 0.06 | 0.10 |
| Malawi | iLiNS−DOSE (45) | | | | | | | | | | | | |
| Mali | PROMIS CS (46) | | | | | | | | | | | | |
| Zimbabwe | SHINE (HIV−) (47) | | | | | | | | | | | | |
| Zimbabwe | SHINE (HIV+) (48) | | | | | | | | | | | | |
| | | **272** | **342** | | **I² = 0.32, Tau² = 0.00** | | | **391** | **226** | | **I² = 0.21, Tau² = 0.00** | | |
| **Fixed** | | | | | **1.03 (0.97, 1.08)** | | | | | | **1.06 (1.00, 1.12)** | | |
| **Random** | | | | | **1.04 (0.96, 1.11)** | | | | | | **1.05 (0.97, 1.13)** | | |

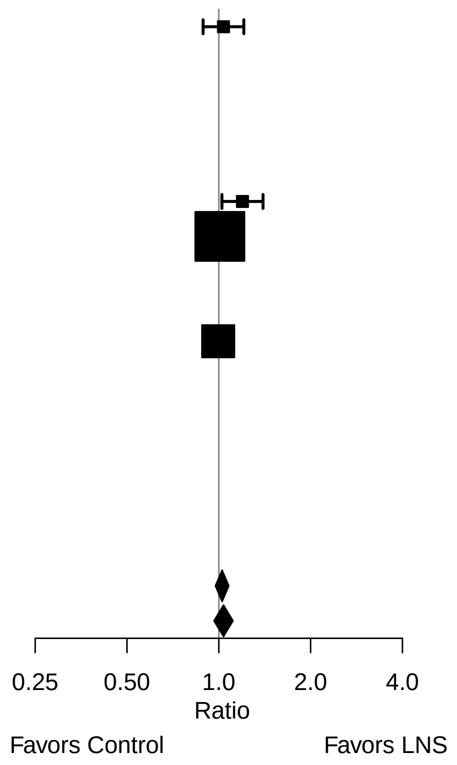

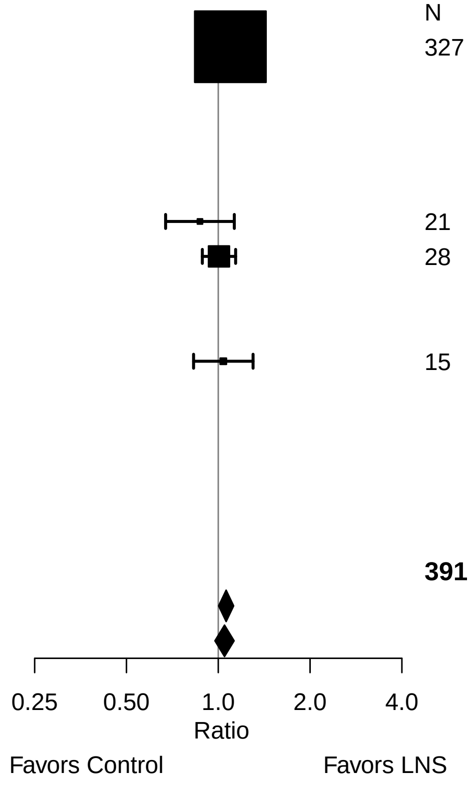

#### Supplemental figure 6R: Geometric mean ratio of retinol concentration

##### 6R6: Stratified by Child baseline acute malnutrition

**P−for−interaction = 0.075**
**Ratio of GMRs = 1.11 (0.99, 1.24)**

| | | No | | | | | | Yes | | | | | |
|---|---|---|---|---|---|---|---|---|---|---|---|---|---|
| Country | Trial | LNS N | Control N | Control Median | GMR (95% CI) | Fixed W | Random W | LNS N | Control N | Control Median | GMR (95% CI) | Fixed W | Random W |
| Bangladesh | JiVitA−4 (35) | 346 | 107 | 1.4 | 1.06 (0.99, 1.14) | 0.35 | 0.35 | 86 | 32 | 1.3 | 1.17 (1.03, 1.32) | 0.69 | 0.69 |
| Bangladesh | RDNS (36) | | | | | | | | | | | | |
| Bangladesh | WASH−B (37) | | | | | | | | | | | | |
| Burkina Faso | iLiNS−Zinc (38) | | | | | | | | | | | | |
| Burkina Faso | PROMIS CS (39) | | | | | | | | | | | | |
| Ghana | GHANA (40) | | | | | | | | | | | | |
| Ghana | iLiNS−DYADG (41) | 90 | 182 | 1.0 | 1.01 (0.94, 1.08) | 0.42 | 0.42 | 6 | 14 | 1.0 | 1.00 (0.81, 1.25) | 0.21 | 0.21 |
| Kenya | WASH−B (42) | | | | | | | | | | | | |
| Madagascar | MAHAY (43) | | | | | | | | | | | | |
| Malawi | iLiNS−DYADM (44) | 58 | 145 | 1.1 | 1.00 (0.91, 1.09) | 0.24 | 0.24 | 7 | 14 | 0.9 | 1.28 (0.92, 1.80) | 0.09 | 0.09 |
| Malawi | iLiNS−DOSE (45) | | | | | | | | | | | | |
| Mali | PROMIS CS (46) | | | | | | | | | | | | |
| Zimbabwe | SHINE (HIV−) (47) | | | | | | | | | | | | |
| Zimbabwe | SHINE (HIV+) (48) | | | | | | | | | | | | |
| | | **494** | **434** | | **I² = 0.00, Tau² = 0.00** | | | **99** | **60** | | **I² = 0.00, Tau² = 0.00** | | |
| **Fixed** | | | | | **1.02 (0.98, 1.07)** | | | | | | **1.14 (1.03, 1.26)** | | |
| **Random** | | | | | **1.02 (0.98, 1.07)** | | | | | | **1.14 (1.03, 1.26)** | | |

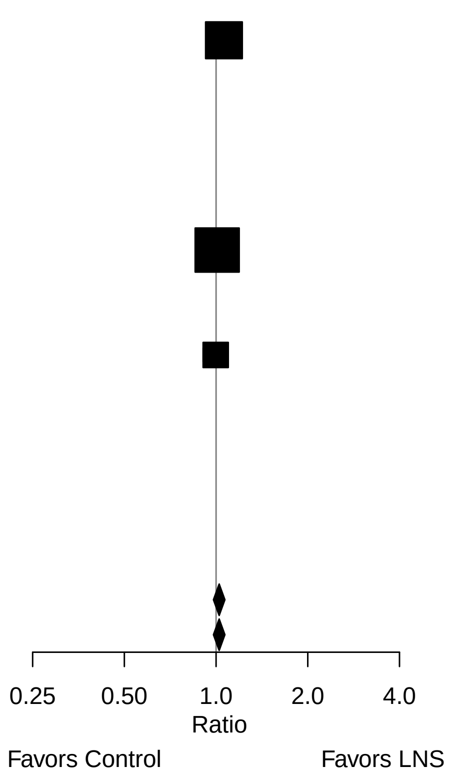

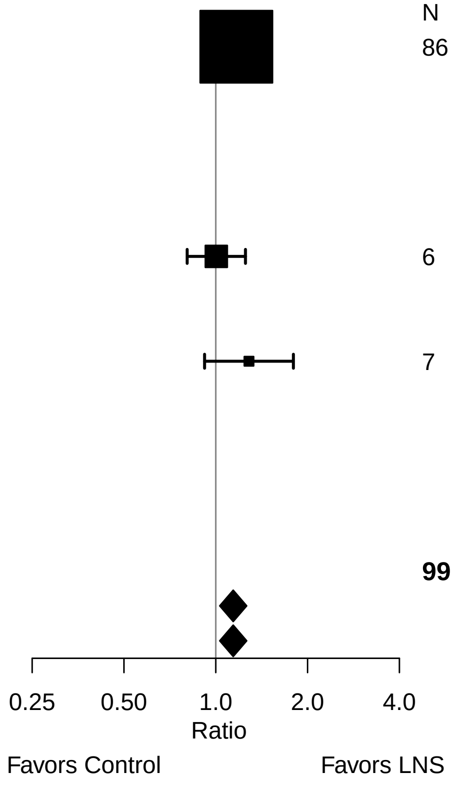

**Supplemental figure 6R: Geometric mean ratio of retinol concentration**

#### 6R7: Stratified by Child baseline anemia (insufficient comparisons)

**Supplemental figure 6R: Geometric mean ratio of retinol concentration**

#### 6R8: Stratified by Child high-dose vitamin A supplementation

**P−for−interaction = 0.577**
**Ratio of GMRs = 0.98 (0.90, 1.06)**

| | | Received VitA | | | | | | Not treated | | | | | |
|---|---|---|---|---|---|---|---|---|---|---|---|---|---|
| | | **LNS** | **Control** | **Control** | **GMR** | **Fixed** | **Random** | **LNS** | **Control** | **Control** | **GMR** | **Fixed** | **Random** |
| Country | Trial | N | N | Median | (95% CI) | W | W | N | N | Median | (95% CI) | W | W |
| Bangladesh | JiVitA−4 (35) | 136 | 36 | 1.4 | 1.10 (1.02, 1.19) | 0.41 | 0.41 | 129 | 38 | 1.4 | 1.04 (0.93, 1.17) | 0.28 | 0.28 |
| Bangladesh | RDNS (36) | | | | | | | | | | | | |
| Bangladesh | WASH−B (37) | | | | | | | | | | | | |
| Burkina Faso | iLiNS−Zinc (38) | | | | | | | | | | | | |
| Burkina Faso | PROMIS CS (39) | | | | | | | | | | | | |
| Ghana | GHANA (40) | 27 | 47 | 1.0 | 1.07 (0.90, 1.27) | 0.08 | 0.08 | 36 | 24 | 1.2 | 1.00 (0.80, 1.25) | 0.07 | 0.07 |
| Ghana | iLiNS−DYADG (41) | 71 | 143 | 1.0 | 1.00 (0.93, 1.08) | 0.43 | 0.43 | 25 | 53 | 1.1 | 1.02 (0.91, 1.15) | 0.26 | 0.26 |
| Kenya | WASH−B (42) | | | | | | | | | | | | |
| Madagascar | MAHAY (43) | | | | | | | | | | | | |
| Malawi | iLiNS−DYADM (44) | 16 | 42 | 1.0 | 1.01 (0.84, 1.20) | 0.08 | 0.08 | 54 | 119 | 1.1 | 1.00 (0.91, 1.10) | 0.39 | 0.39 |
| Malawi | iLiNS−DOSE (45) | | | | | | | | | | | | |
| Mali | PROMIS CS (46) | | | | | | | | | | | | |
| Zimbabwe | SHINE (HIV−) (47) | | | | | | | | | | | | |
| Zimbabwe | SHINE (HIV+) (48) | | | | | | | | | | | | |
| | | **250** | **268** | | **I² = 0.00, Tau² = 0.00** | | | **244** | **234** | | **I² = 0.00, Tau² = 0.00** | | |
| **Fixed** | | | | | **1.05 (1.00, 1.10)** | | | | | | **1.02 (0.96, 1.08)** | | |
| **Random** | | | | | **1.05 (1.00, 1.10)** | | | | | | **1.02 (0.96, 1.08)** | | |

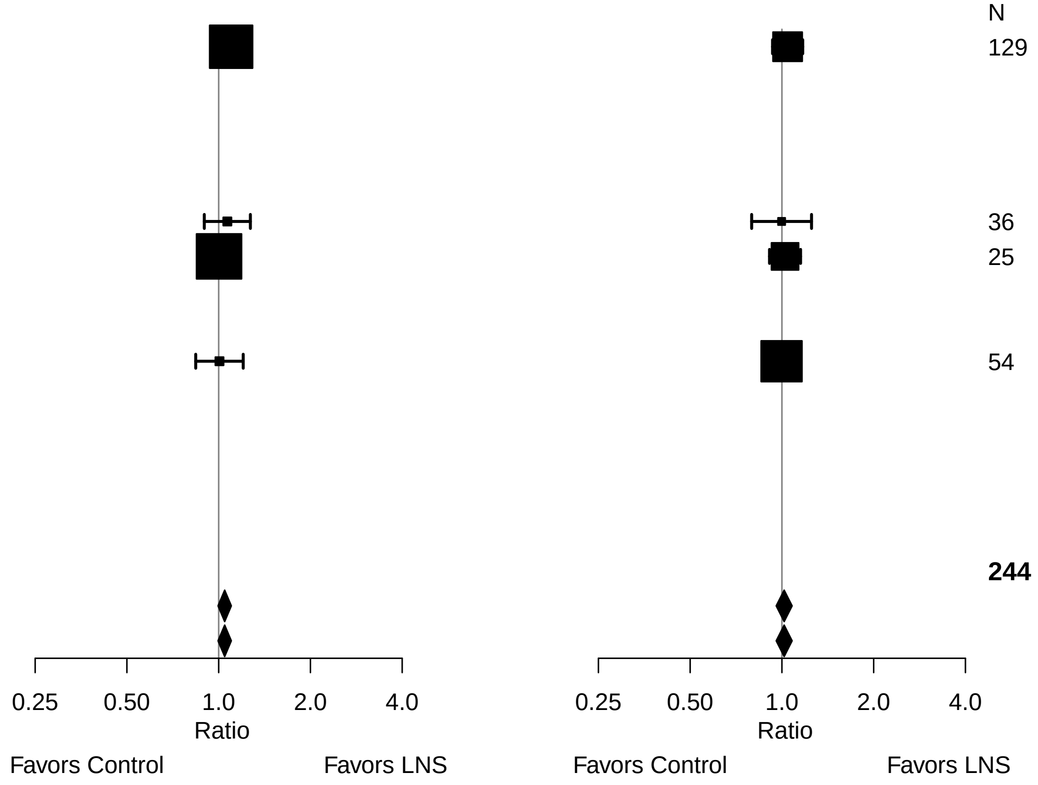

**Supplemental figure 6R: Geometric mean ratio of retinol concentration**

#### 6R9: Stratified by Child inflammation

**P−for−interaction = 0.481**
**Ratio of GMRs = 1.03 (0.95, 1.11)**

| | | Not inflamed | | | | | | High AGP or CRP | | | | | |
|---|---|---|---|---|---|---|---|---|---|---|---|---|---|
| | | **LNS** | **Control** | **Control** | **GMR** | **Fixed** | **Random** | **LNS** | **Control** | **Control** | **GMR** | **Fixed** | **Random** |
| Country | Trial | N | N | Median | (95% CI) | W | W | N | N | Median | (95% CI) | W | W |
| Bangladesh | JiVitA−4 (35) | 177 | 46 | 1.4 | 1.05 (0.97, 1.15) | 0.37 | 0.36 | 257 | 93 | 1.4 | 1.10 (1.02, 1.20) | 0.43 | 0.42 |
| Bangladesh | RDNS (36) | | | | | | | | | | | | |
| Bangladesh | WASH−B (37) | | | | | | | | | | | | |
| Burkina Faso | iLiNS−Zinc (38) | | | | | | | | | | | | |
| Burkina Faso | PROMIS CS (39) | | | | | | | | | | | | |
| Ghana | GHANA (40) | 51 | 59 | 1.1 | 1.13 (0.98, 1.30) | 0.13 | 0.14 | 12 | 15 | 1.0 | 0.86 (0.59, 1.26) | 0.02 | 0.02 |
| Ghana | iLiNS−DYADG (41) | 43 | 118 | 1.0 | 0.99 (0.91, 1.07) | 0.40 | 0.39 | 53 | 78 | 1.0 | 1.03 (0.93, 1.15) | 0.27 | 0.27 |
| Kenya | WASH−B (42) | | | | | | | | | | | | |
| Madagascar | MAHAY (43) | | | | | | | | | | | | |
| Malawi | iLiNS−DYADM (44) | 20 | 61 | 1.0 | 0.98 (0.84, 1.15) | 0.10 | 0.11 | 51 | 102 | 1.1 | 1.01 (0.91, 1.11) | 0.28 | 0.29 |
| Malawi | iLiNS−DOSE (45) | | | | | | | | | | | | |
| Mali | PROMIS CS (46) | | | | | | | | | | | | |
| Zimbabwe | SHINE (HIV−) (47) | | | | | | | | | | | | |
| Zimbabwe | SHINE (HIV+) (48) | | | | | | | | | | | | |
| | | **291** | **284** | | **I² = 0.02, Tau² = 0.00** | | | **373** | **288** | | **I² = 0.06, Tau² = 0.00** | | |
| **Fixed** | | | | | **1.03 (0.98, 1.08)** | | | | | | **1.05 (1.00, 1.11)** | | |
| **Random** | | | | | **1.03 (0.98, 1.09)** | | | | | | **1.05 (0.99, 1.11)** | | |

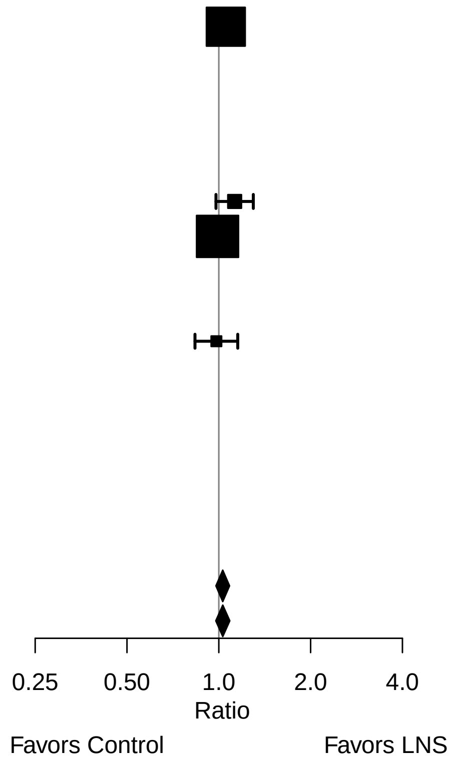

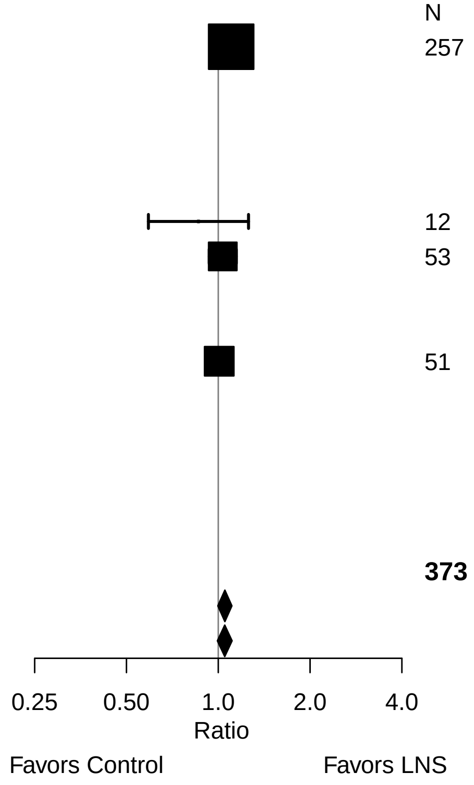

### Supplemental figure 6S: Low vitamin A (retinol < 0.70 µmol/L) prevalence ratio

#### 6S1: Stratified by Maternal BMI (insufficient comparisons)

**Supplemental figure 6S: Low vitamin A (retinol < 0.70 µmol/L) prevalence ratio**

#### 6S2: Stratified by Maternal age (insufficient comparisons)

**Supplemental figure 6S: Low vitamin A (retinol < 0.70 μmol/L) prevalence ratio**

#### 6S3: Stratified by Maternal education (insufficient comparisons)

**Supplemental figure 6S: Low vitamin A (retinol < 0.70 µmol/L) prevalence ratio**

#### 6S4: Stratified by Child sex (insufficient comparisons)

**Supplemental figure 6S: Low vitamin A (retinol < 0.70 µmol/L) prevalence ratio**

#### 6S5: Stratified by Child birth order (insufficient comparisons)

**Supplemental figure 6S: Low vitamin A (retinol < 0.70 µmol/L) prevalence ratio**

#### 6S6: Stratified by Child baseline acute malnutrition (insufficient comparisons)

**Supplemental figure 6S: Low vitamin A (retinol < 0.70 µmol/L) prevalence ratio**

#### 6S7: Stratified by Child baseline anemia (insufficient comparisons)

**Supplemental figure 6S: Low vitamin A (retinol < 0.70 µmol/L) prevalence ratio**

#### 6S8: Stratified by Child high-dose vitamin A supplementation (insufficient comparisons)

**Supplemental figure 6S: Low vitamin A (retinol < 0.70 µmol/L) prevalence ratio**

#### 6S9: Stratified by Child inflammation (insufficient comparisons)

### Supplemental figure 6T: Low vitamin A (retinol < 0.70 µmol/L) prevalence difference

#### 6T1: Stratified by Maternal BMI (insufficient comparisons)

**Supplemental figure 6T: Low vitamin A (retinol < 0.70 µmol/L) prevalence difference**

#### 6T2: Stratified by Maternal age (insufficient comparisons)

**Supplemental figure 6T: Low vitamin A (retinol $< 0.70$ µmol/L) prevalence difference**

#### 6T3: Stratified by Maternal education (insufficient comparisons)

**Supplemental figure 6T: Low vitamin A (retinol < 0.70 µmol/L) prevalence difference**

#### 6T4: Stratified by Child sex (insufficient comparisons)

**Supplemental figure 6T: Low vitamin A (retinol < 0.70 μmol/L) prevalence difference**

#### 6T5: Stratified by Child birth order (insufficient comparisons)

**Supplemental figure 6T: Low vitamin A (retinol < 0.70 µmol/L) prevalence difference**

#### 6T6: Stratified by Child baseline acute malnutrition (insufficient comparisons)

**Supplemental figure 6T: Low vitamin A (retinol < 0.70 µmol/L) prevalence difference**

#### 6T7: Stratified by Child baseline anemia (insufficient comparisons)

**Supplemental figure 6T: Low vitamin A (retinol < 0.70 µmol/L) prevalence difference**

#### 6T8: Stratified by Child high-dose vitamin A supplementation (insufficient comparisons)

**Supplemental figure 6T: Low vitamin A (retinol < 0.70 µmol/L) prevalence difference**

#### 6T9: Stratified by Child inflammation (insufficient comparisons)

#### Supplemental figure 6U: Marginal vitamin A (retinol < 1.05 µmol/L) prevalence ratio

##### 6U1: Stratified by Maternal BMI (insufficient comparisons)

#### Supplemental figure 6U: Marginal vitamin A (retinol < 1.05 µmol/L) prevalence ratio

#### 6U2: Stratified by Maternal age

**P−for−interaction = 0.912**
**Ratio of PRs = 0.98 (0.73, 1.32)**

| Country | Trial | At least 25 y: LNS N | Control N | Control Prevalence | PR (95% CI) | Fixed W | Random W | Less than 25 y: LNS N | Control N | Control Prevalence | PR (95% CI) | Fixed W | Random W |
|---|---|---|---|---|---|---|---|---|---|---|---|---|---|
| Bangladesh | JiVitA−4 (35) | 195 | 67 | 23.9 | 0.62 (0.35, 1.11) | 0.11 | 0.14 | 237 | 71 | 15.5 | 0.90 (0.51, 1.59) | 0.14 | 0.14 |
| Bangladesh | RDNS (36) | | | | | | | | | | | | |
| Bangladesh | WASH−B (37) | | | | | | | | | | | | |
| Burkina Faso | iLiNS−Zinc (38) | | | | | | | | | | | | |
| Burkina Faso | PROMIS CS (39) | | | | | | | | | | | | |
| Ghana | GHANA (40) | 47 | 47 | 53.2 | 0.96 (0.65, 1.41) | 0.24 | 0.25 | 16 | 21 | 38.1 | 1.15 (0.53, 2.50) | 0.07 | 0.07 |
| Ghana | iLiNS−DYADG (41) | 63 | 124 | 50.0 | 0.98 (0.72, 1.34) | 0.39 | 0.35 | 33 | 72 | 68.1 | 0.85 (0.61, 1.18) | 0.41 | 0.41 |
| Kenya | WASH−B (42) | | | | | | | | | | | | |
| Madagascar | MAHAY (43) | | | | | | | | | | | | |
| Malawi | iLiNS−DYADM (44) | 33 | 81 | 46.9 | 1.23 (0.84, 1.78) | 0.26 | 0.27 | 38 | 82 | 50.0 | 1.16 (0.82, 1.64) | 0.38 | 0.38 |
| Malawi | iLiNS−DOSE (45) | | | | | | | | | | | | |
| Mali | PROMIS CS (46) | | | | | | | | | | | | |
| Zimbabwe | SHINE (HIV−) (47) | | | | | | | | | | | | |
| Zimbabwe | SHINE (HIV+) (48) | | | | | | | | | | | | |
| | | **338** | **319** | | **I² = 0.21, Tau² = 0.02** | | | **324** | **246** | | **I² = 0.00, Tau² = 0.00** | | |
| **Fixed** | | | | | **0.99 (0.81, 1.19)** | | | | | | **0.98 (0.79, 1.22)** | | |
| **Random** | | | | | **0.98 (0.77, 1.23)** | | | | | | **0.98 (0.79, 1.22)** | | |

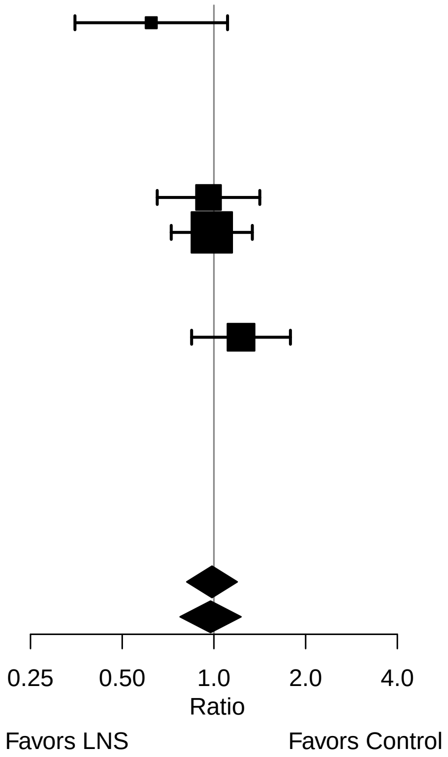

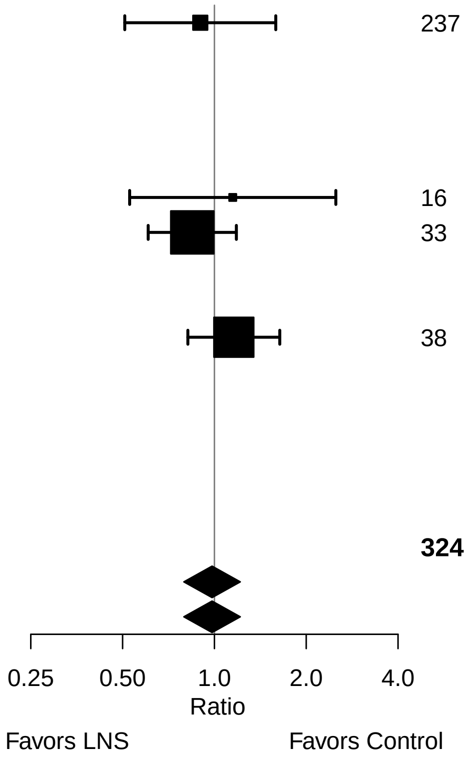

**Supplemental figure 6U: Marginal vitamin A (retinol < 1.05 µmol/L) prevalence ratio**

#### 6U3: Stratified by Maternal education (insufficient comparisons)

#### Supplemental figure 6U: Marginal vitamin A (retinol < 1.05 µmol/L) prevalence ratio

##### 6U4: Stratified by Child sex

**P−for−interaction = 0.838**
**Ratio of PRs = 1.03 (0.77, 1.38)**

| Country | Trial | Male LNS N | Male Control N | Male Control Prevalence | Male PR (95% CI) | Male Fixed W | Male Random W | Female LNS N | Female Control N | Female Control Prevalence | Female PR (95% CI) | Female Fixed W | Female Random W |
|---|---|---|---|---|---|---|---|---|---|---|---|---|---|
| Bangladesh | JiVitA−4 (35) | 222 | 67 | 17.9 | 1.03 (0.51, 2.10) | 0.07 | 0.10 | 212 | 72 | 20.8 | 0.48 (0.31, 0.73) | 0.25 | 0.25 |
| Bangladesh | RDNS (36) | | | | | | | | | | | | |
| Bangladesh | WASH−B (37) | | | | | | | | | | | | |
| Burkina Faso | iLiNS−Zinc (38) | | | | | | | | | | | | |
| Burkina Faso | PROMIS CS (39) | | | | | | | | | | | | |
| Ghana | GHANA (40) | 37 | 32 | 46.9 | 1.04 (0.63, 1.70) | 0.14 | 0.18 | 26 | 42 | 50.0 | 1.00 (0.61, 1.63) | 0.19 | 0.23 |
| Ghana | iLiNS−DYADG (41) | 62 | 93 | 66.7 | 0.75 (0.56, 1.00) | 0.41 | 0.37 | 33 | 102 | 47.1 | 1.16 (0.80, 1.68) | 0.33 | 0.27 |
| Kenya | WASH−B (42) | | | | | | | | | | | | |
| Madagascar | MAHAY (43) | | | | | | | | | | | | |
| Malawi | iLiNS−DYADM (44) | 39 | 80 | 55.0 | 1.21 (0.90, 1.63) | 0.38 | 0.36 | 32 | 83 | 42.2 | 1.11 (0.71, 1.74) | 0.23 | 0.25 |
| Malawi | iLiNS−DOSE (45) | | | | | | | | | | | | |
| Mali | PROMIS CS (46) | | | | | | | | | | | | |
| Zimbabwe | SHINE (HIV−) (47) | | | | | | | | | | | | |
| Zimbabwe | SHINE (HIV+) (48) | | | | | | | | | | | | |
| | | **360** | **272** | | **$I^2$ = 0.44, $Tau^2$ = 0.02** | | | **303** | **299** | | **$I^2$ = 0.74, $Tau^2$ = 0.13** | | |
| **Fixed** | | | | | **0.96 (0.80, 1.16)** | | | | | | **0.89 (0.72, 1.11)** | | |
| **Random** | | | | | **0.97 (0.77, 1.23)** | | | | | | **0.89 (0.59, 1.34)** | | |

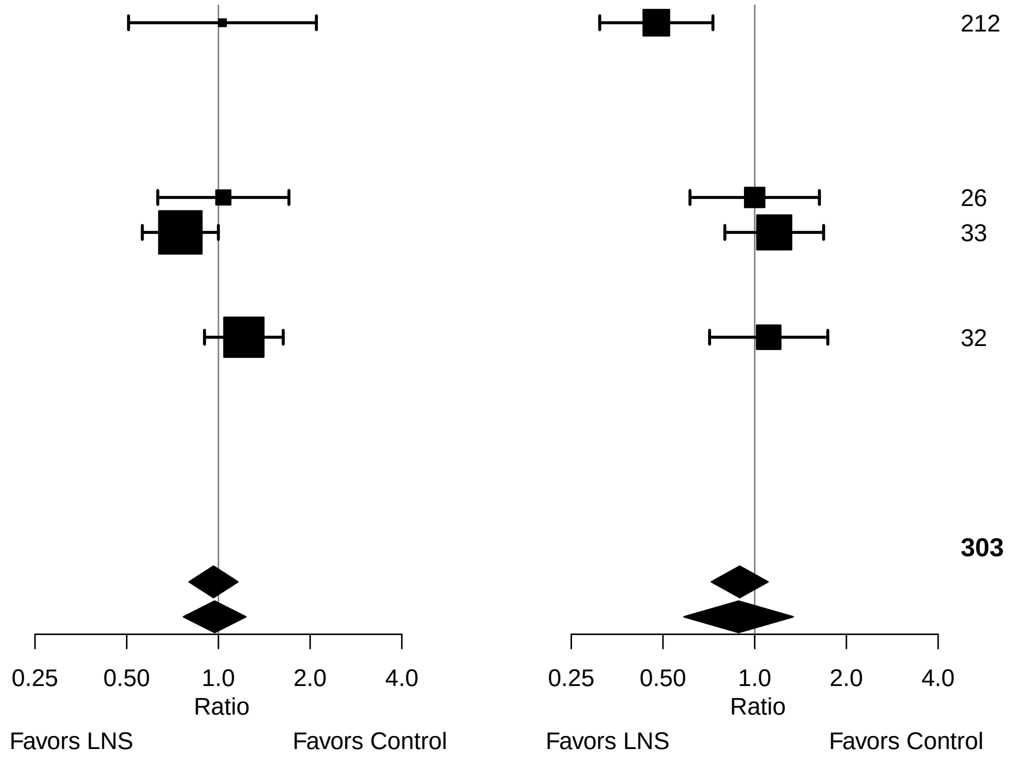

##### Supplemental figure 6U: Marginal vitamin A (retinol < 1.05 µmol/L) prevalence ratio

#### 6U5: Stratified by Child birth order

**P−for−interaction = 0.132**
**Ratio of PRs = 1.27 (0.93, 1.74)**

| Country | Trial | Later born LNS N | Later born Control N | Later born Control Prevalence | Later born PR (95% CI) | Later born Fixed W | Later born Random W | Firstborn LNS N | Firstborn Control N | Firstborn Control Prevalence | Firstborn PR (95% CI) | Firstborn Fixed W | Firstborn Random W |
|---|---|---|---|---|---|---|---|---|---|---|---|---|---|
| Bangladesh | JiVitA−4 (35) | 106 | 31 | 16.1 | 0.82 (0.29, 2.29) | 0.03 | 0.05 | 327 | 107 | 20.6 | 0.71 (0.46, 1.10) | 0.24 | 0.26 |
| Bangladesh | RDNS (36) | | | | | | | | | | | | |
| Bangladesh | WASH−B (37) | | | | | | | | | | | | |
| Burkina Faso | iLiNS−Zinc (38) | | | | | | | | | | | | |
| Burkina Faso | PROMIS CS (39) | | | | | | | | | | | | |
| Ghana | GHANA (40) | 42 | 45 | 51.1 | 0.79 (0.50, 1.26) | 0.16 | 0.20 | 21 | 26 | 46.2 | 1.44 (0.86, 2.41) | 0.17 | 0.21 |
| Ghana | iLiNS−DYADG (41) | 68 | 137 | 54.0 | 0.82 (0.60, 1.11) | 0.36 | 0.35 | 28 | 59 | 62.7 | 1.14 (0.84, 1.55) | 0.48 | 0.38 |
| Kenya | WASH−B (42) | | | | | | | | | | | | |
| Madagascar | MAHAY (43) | | | | | | | | | | | | |
| Malawi | iLiNS−DYADM (44) | 56 | 129 | 48.8 | 1.24 (0.94, 1.64) | 0.45 | 0.40 | 15 | 34 | 47.1 | 0.99 (0.52, 1.90) | 0.11 | 0.15 |
| Malawi | iLiNS−DOSE (45) | | | | | | | | | | | | |
| Mali | PROMIS CS (46) | | | | | | | | | | | | |
| Zimbabwe | SHINE (HIV−) (47) | | | | | | | | | | | | |
| Zimbabwe | SHINE (HIV+) (48) | | | | | | | | | | | | |
| | | **272** | **342** | | **I² = 0.42, Tau² = 0.02** | | | **391** | **226** | | **I² = 0.38, Tau² = 0.03** | | |
| **Fixed** | | | | | **0.98 (0.82, 1.18)** | | | | | | **1.05 (0.85, 1.29)** | | |
| **Random** | | | | | **0.96 (0.76, 1.22)** | | | | | | **1.04 (0.78, 1.38)** | | |

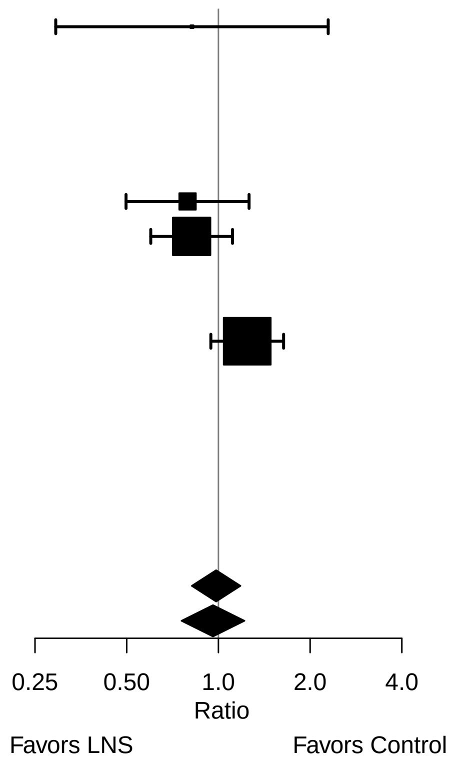

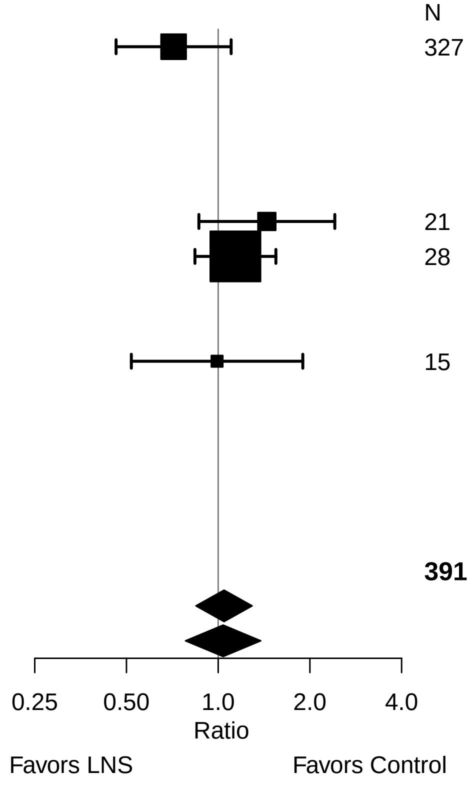

**Supplemental figure 6U: Marginal vitamin A (retinol < 1.05 µmol/L) prevalence ratio**

#### 6U6: Stratified by Child baseline acute malnutrition (insufficient comparisons)

**Supplemental figure 6U: Marginal vitamin A (retinol < 1.05 µmol/L) prevalence ratio**

#### 6U7: Stratified by Child baseline anemia (insufficient comparisons)

##### Supplemental figure 6U: Marginal vitamin A (retinol < 1.05 µmol/L) prevalence ratio

#### 6U8: Stratified by Child high-dose vitamin A supplementation

**P−for−interaction = 0.520**
**Ratio of PRs = 0.90 (0.64, 1.25)**

| | | Received VitA | | | | | | Not treated | | | | | |
|---|---|---|---|---|---|---|---|---|---|---|---|---|---|
| Country | Trial | LNS N | Control N | Control Prevalence | PR (95% CI) | Fixed W | Random W | LNS N | Control N | Control Prevalence | PR (95% CI) | Fixed W | Random W |
| Bangladesh | JiVitA−4 (35) | 136 | 36 | 13.9 | 0.74 (0.33, 1.68) | 0.05 | 0.05 | 129 | 38 | 18.4 | 0.67 (0.31, 1.44) | 0.09 | 0.09 |
| Bangladesh | RDNS (36) | | | | | | | | | | | | |
| Bangladesh | WASH−B (37) | | | | | | | | | | | | |
| Burkina Faso | iLiNS−Zinc (38) | | | | | | | | | | | | |
| Burkina Faso | PROMIS CS (39) | | | | | | | | | | | | |
| Ghana | GHANA (40) | 27 | 47 | 53.2 | 1.11 (0.74, 1.68) | 0.21 | 0.21 | 36 | 24 | 41.7 | 1.00 (0.54, 1.84) | 0.15 | 0.15 |
| Ghana | iLiNS−DYADG (41) | 71 | 143 | 59.4 | 0.92 (0.72, 1.19) | 0.56 | 0.55 | 25 | 53 | 49.1 | 0.90 (0.53, 1.51) | 0.20 | 0.20 |
| Kenya | WASH−B (42) | | | | | | | | | | | | |
| Madagascar | MAHAY (43) | | | | | | | | | | | | |
| Malawi | iLiNS−DYADM (44) | 16 | 42 | 50.0 | 1.37 (0.88, 2.15) | 0.18 | 0.18 | 54 | 119 | 47.1 | 1.14 (0.83, 1.56) | 0.56 | 0.56 |
| Malawi | iLiNS−DOSE (45) | | | | | | | | | | | | |
| Mali | PROMIS CS (46) | | | | | | | | | | | | |
| Zimbabwe | SHINE (HIV−) (47) | | | | | | | | | | | | |
| Zimbabwe | SHINE (HIV+) (48) | | | | | | | | | | | | |
| | | **250** | **268** | | **I² = 0.02, Tau² = 0.00** | | | **244** | **234** | | **I² = 0.00, Tau² = 0.00** | | |
| **Fixed** | | | | | **1.02 (0.84, 1.23)** | | | | | | **1.01 (0.80, 1.28)** | | |
| **Random** | | | | | **1.02 (0.84, 1.24)** | | | | | | **1.01 (0.80, 1.28)** | | |

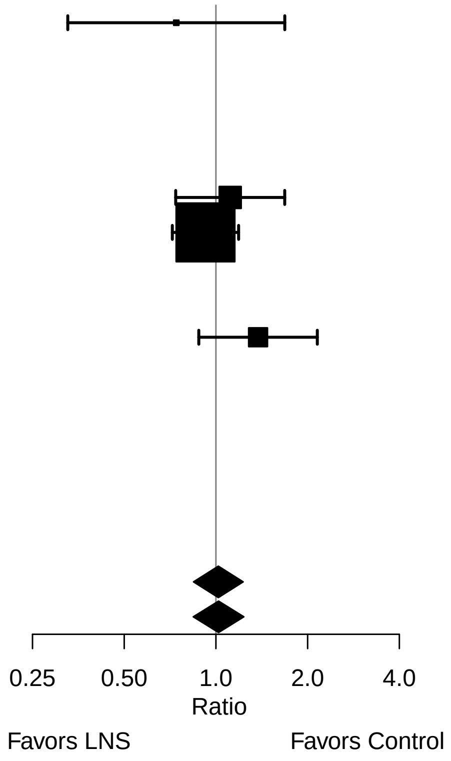

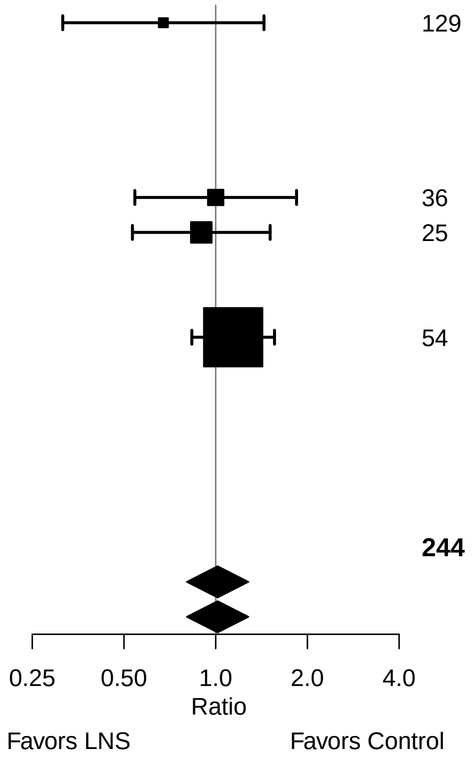

#### Supplemental figure 6U: Marginal vitamin A (retinol < 1.05 µmol/L) prevalence ratio

#### 6U9: Stratified by Child inflammation

**P−for−interaction = 0.775**
**Ratio of PRs = 0.96 (0.70, 1.30)**

| | | Not inflamed | | | | | | High AGP or CRP | | | | | |
|---|---|---|---|---|---|---|---|---|---|---|---|---|---|
| Country | Trial | LNS N | Control N | Control Prevalence | PR (95% CI) | Fixed W | Random W | LNS N | Control N | Control Prevalence | PR (95% CI) | Fixed W | Random W |
| Bangladesh | JiVitA−4 (35) | 177 | 46 | 21.7 | 0.68 (0.42, 1.08) | 0.21 | 0.30 | 257 | 93 | 18.3 | 0.77 (0.43, 1.36) | 0.14 | 0.14 |
| Bangladesh | RDNS (36) | | | | | | | | | | | | |
| Bangladesh | WASH−B (37) | | | | | | | | | | | | |
| Burkina Faso | iLiNS−Zinc (38) | | | | | | | | | | | | |
| Burkina Faso | PROMIS CS (39) | | | | | | | | | | | | |
| Ghana | GHANA (40) | | | | | | | | | | | | |
| Ghana | iLiNS−DYADG (41) | 43 | 118 | 55.9 | 0.87 (0.62, 1.23) | 0.40 | 0.35 | 53 | 78 | 57.7 | 0.95 (0.70, 1.29) | 0.47 | 0.47 |
| Kenya | WASH−B (42) | | | | | | | | | | | | |
| Madagascar | MAHAY (43) | | | | | | | | | | | | |
| Malawi | iLiNS−DYADM (44) | 20 | 61 | 52.5 | 1.43 (1.01, 2.02) | 0.39 | 0.35 | 51 | 102 | 46.1 | 1.11 (0.79, 1.56) | 0.39 | 0.39 |
| Malawi | iLiNS−DOSE (45) | | | | | | | | | | | | |
| Mali | PROMIS CS (46) | | | | | | | | | | | | |
| Zimbabwe | SHINE (HIV−) (47) | | | | | | | | | | | | |
| Zimbabwe | SHINE (HIV+) (48) | | | | | | | | | | | | |
| | | **240** | **225** | | **I² = 0.73, Tau² = 0.10** | | | **361** | **273** | | **I² = 0.00, Tau² = 0.00** | | |
| **Fixed** | | | | | **1.00 (0.81, 1.24)** | | | | | | **0.98 (0.79, 1.21)** | | |
| **Random** | | | | | **0.96 (0.63, 1.48)** | | | | | | **0.98 (0.79, 1.21)** | | |

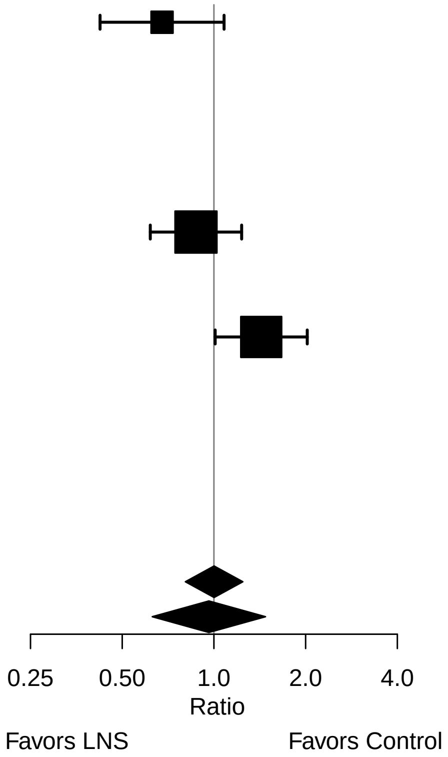

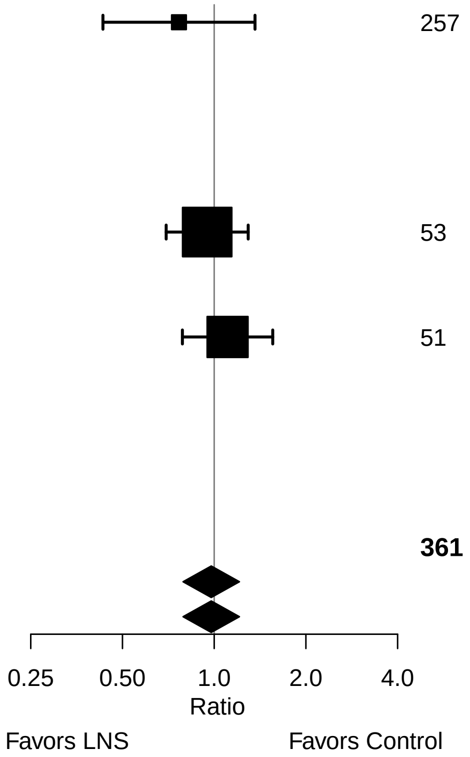

#### Supplemental figure 6V: Marginal vitamin A (retinol < 1.05 µmol/L) prevalence difference

##### 6V1: Stratified by Maternal BMI (insufficient comparisons)

#### Supplemental figure 6V: Marginal vitamin A (retinol < 1.05 μmol/L) prevalence difference

#### 6V2: Stratified by Maternal age

**P−for−interaction = 0.659**
**Difference in PDs = 0.03 (−0.09, 0.14)**

| | | At least 25 y | | | | | | Less than 25 y | | | | | |
|---|---|---|---|---|---|---|---|---|---|---|---|---|---|
| | | **LNS** | **Control** | **Control** | **PD** | **Fixed** | **Random** | **LNS** | **Control** | **Control** | **PD** | **Fixed** | **Random** |
| Country | Trial | N | N | Prevalence | (95% CI) | W | W | N | N | Prevalence | (95% CI) | W | W |
| Bangladesh | JiVitA−4 (35) | 195 | 67 | 23.9 | −0.09 (−0.21, 0.03) | 0.42 | 0.42 | 237 | 71 | 15.5 | −0.02 (−0.10, 0.07) | 0.69 | 0.69 |
| Bangladesh | RDNS (36) | | | | | | | | | | | | |
| Bangladesh | WASH−B (37) | | | | | | | | | | | | |
| Burkina Faso | iLiNS−Zinc (38) | | | | | | | | | | | | |
| Burkina Faso | PROMIS CS (39) | | | | | | | | | | | | |
| Ghana | GHANA (40) | 47 | 47 | 53.2 | −0.02 (−0.23, 0.18) | 0.15 | 0.15 | 16 | 21 | 38.1 | 0.06 (−0.27, 0.38) | 0.05 | 0.05 |
| Ghana | iLiNS−DYADG (41) | 63 | 124 | 50.0 | −0.01 (−0.16, 0.14) | 0.27 | 0.27 | 33 | 72 | 68.1 | −0.10 (−0.30, 0.09) | 0.13 | 0.13 |
| Kenya | WASH−B (42) | | | | | | | | | | | | |
| Madagascar | MAHAY (43) | | | | | | | | | | | | |
| Malawi | iLiNS−DYADM (44) | 33 | 81 | 46.9 | 0.11 (−0.10, 0.31) | 0.15 | 0.15 | 38 | 82 | 50.0 | 0.08 (−0.11, 0.27) | 0.14 | 0.14 |
| Malawi | iLiNS−DOSE (45) | | | | | | | | | | | | |
| Mali | PROMIS CS (46) | | | | | | | | | | | | |
| Zimbabwe | SHINE (HIV−) (47) | | | | | | | | | | | | |
| Zimbabwe | SHINE (HIV+) (48) | | | | | | | | | | | | |
| | | **338** | **319** | | **I² = 0.00, Tau² = 0.00** | | | **324** | **246** | | **I² = 0.00, Tau² = 0.00** | | |
| **Fixed** | | | | | **−0.03 (−0.11, 0.05)** | | | | | | **−0.01 (−0.08, 0.06)** | | |
| **Random** | | | | | **−0.03 (−0.11, 0.05)** | | | | | | **−0.01 (−0.08, 0.06)** | | |

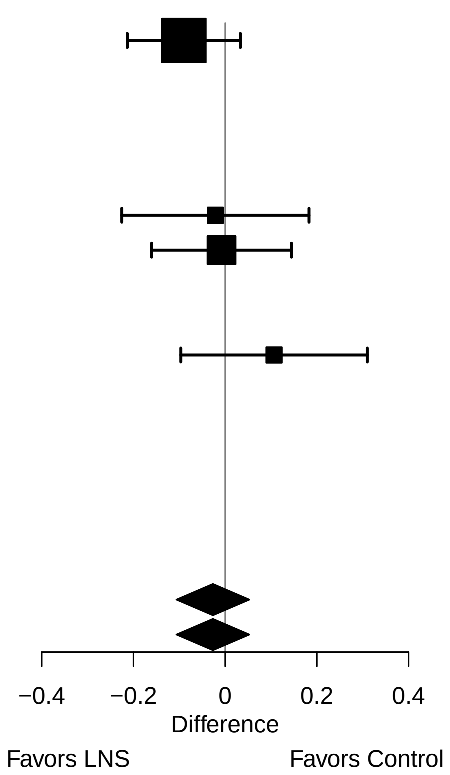

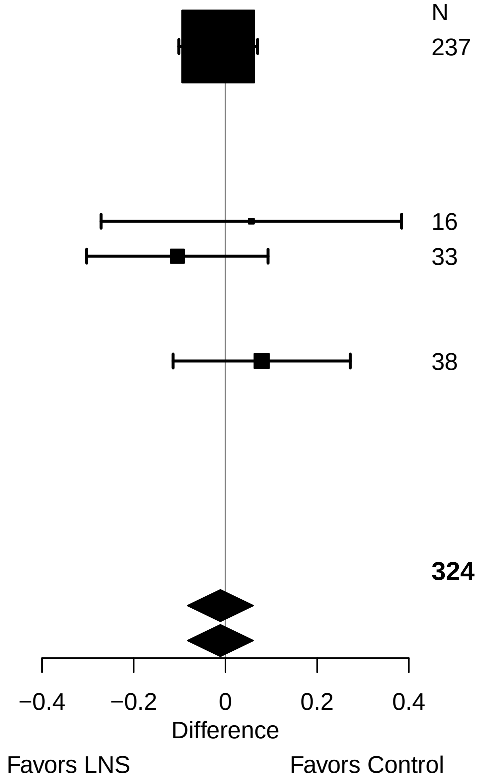

**Supplemental figure 6V: Marginal vitamin A (retinol < 1.05 µmol/L) prevalence difference**

#### 6V3: Stratified by Maternal education (insufficient comparisons)

**Supplemental figure 6V: Marginal vitamin A (retinol < 1.05 µmol/L) prevalence difference**

#### 6V4: Stratified by Child sex

**P−for−interaction = 0.578**
**Difference in PDs = −0.03 (−0.14, 0.08)**

| | | Male | | | | | | Female | | | | | |
|---|---|---|---|---|---|---|---|---|---|---|---|---|---|
| Country | Trial | LNS N | Control N | Control Prevalence | PD (95% CI) | Fixed W | Random W | LNS N | Control N | Control Prevalence | PD (95% CI) | Fixed W | Random W |
| Bangladesh | JiVitA−4 (35) | 222 | 67 | 17.9 | 0.01 (−0.12, 0.13) | 0.42 | 0.33 | 212 | 72 | 20.8 | −0.11 (−0.17, −0.05) | 0.83 | 0.58 |
| Bangladesh | RDNS (36) | | | | | | | | | | | | |
| Bangladesh | WASH−B (37) | | | | | | | | | | | | |
| Burkina Faso | iLiNS−Zinc (38) | | | | | | | | | | | | |
| Burkina Faso | PROMIS CS (39) | | | | | | | | | | | | |
| Ghana | GHANA (40) | 37 | 32 | 46.9 | 0.02 (−0.22, 0.26) | 0.12 | 0.17 | 26 | 42 | 50.0 | 0.00 (−0.25, 0.25) | 0.04 | 0.11 |
| Ghana | iLiNS−DYADG (41) | 62 | 93 | 66.7 | −0.17 (−0.32, −0.01) | 0.28 | 0.28 | 33 | 102 | 47.1 | 0.07 (−0.12, 0.27) | 0.07 | 0.16 |
| Kenya | WASH−B (42) | | | | | | | | | | | | |
| Madagascar | MAHAY (43) | | | | | | | | | | | | |
| Malawi | iLiNS−DYADM (44) | 39 | 80 | 55.0 | 0.12 (−0.07, 0.31) | 0.19 | 0.23 | 32 | 83 | 42.2 | 0.05 (−0.16, 0.25) | 0.06 | 0.15 |
| Malawi | iLiNS−DOSE (45) | | | | | | | | | | | | |
| Mali | PROMIS CS (46) | | | | | | | | | | | | |
| Zimbabwe | SHINE (HIV−) (47) | | | | | | | | | | | | |
| Zimbabwe | SHINE (HIV+) (48) | | | | | | | | | | | | |
| | | **360** | **272** | | **I² = 0.47, Tau² = 0.01** | | | **303** | **299** | | **I² = 0.43, Tau² = 0.00** | | |
| **Fixed** | | | | | **−0.02 (−0.10, 0.06)** | | | | | | **−0.08 (−0.13, −0.03)** | | |
| **Random** | | | | | **−0.02 (−0.13, 0.10)** | | | | | | **−0.04 (−0.13, 0.04)** | | |

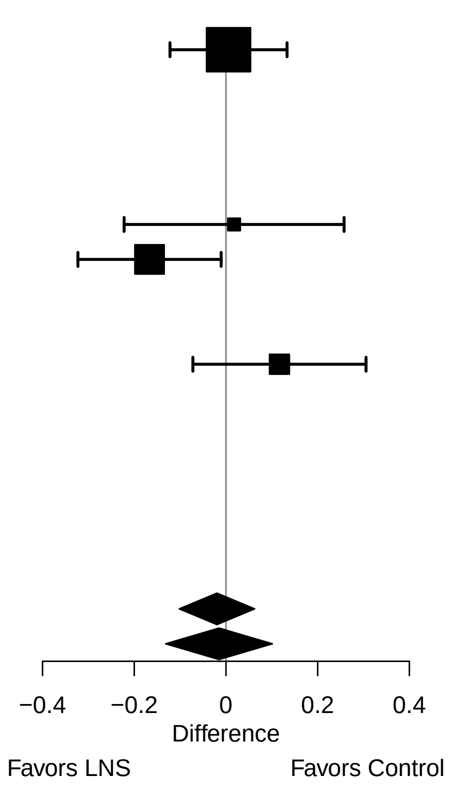

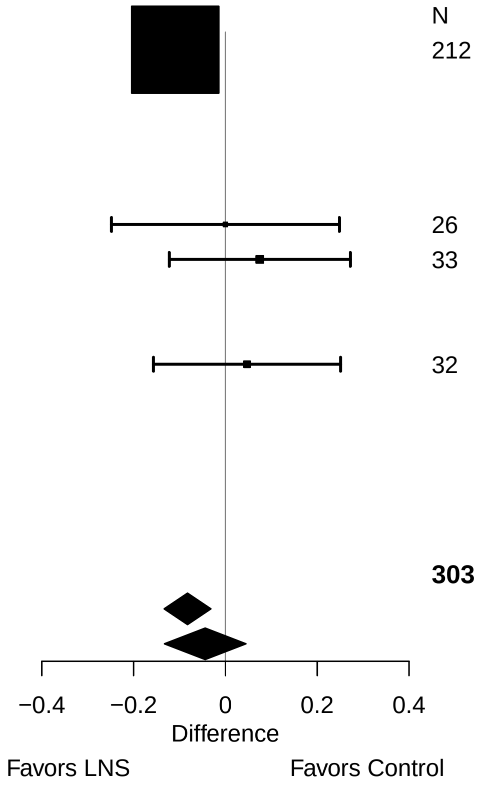

#### Supplemental figure 6V: Marginal vitamin A (retinol < 1.05 µmol/L) prevalence difference

##### 6V5: Stratified by Child birth order

**P−for−interaction = 0.432**
**Difference in PDs = 0.05 (−0.07, 0.17)**

| | | Later born | | | | | | Firstborn | | | | | |
|---|---|---|---|---|---|---|---|---|---|---|---|---|---|
| Country | Trial | LNS N | Control N | Control Prevalence | PD (95% CI) | Fixed W | Random W | LNS N | Control N | Control Prevalence | PD (95% CI) | Fixed W | Random W |
| Bangladesh | JiVitA−4 (35) | 106 | 31 | 16.1 | −0.03 (−0.19, 0.13) | 0.26 | 0.26 | 327 | 107 | 20.6 | −0.06 (−0.14, 0.02) | 0.77 | 0.60 |
| Bangladesh | RDNS (36) | | | | | | | | | | | | |
| Bangladesh | WASH−B (37) | | | | | | | | | | | | |
| Burkina Faso | iLiNS−Zinc (38) | | | | | | | | | | | | |
| Burkina Faso | PROMIS CS (39) | | | | | | | | | | | | |
| Ghana | GHANA (40) | 42 | 45 | 51.1 | −0.11 (−0.32, 0.10) | 0.15 | 0.18 | 21 | 26 | 46.2 | 0.21 (−0.08, 0.49) | 0.06 | 0.11 |
| Ghana | iLiNS−DYADG (41) | 68 | 137 | 54.0 | −0.10 (−0.24, 0.05) | 0.31 | 0.29 | 28 | 59 | 62.7 | 0.09 (−0.13, 0.30) | 0.11 | 0.19 |
| Kenya | WASH−B (42) | | | | | | | | | | | | |
| Madagascar | MAHAY (43) | | | | | | | | | | | | |
| Malawi | iLiNS−DYADM (44) | 56 | 129 | 48.8 | 0.12 (−0.04, 0.28) | 0.27 | 0.27 | 15 | 34 | 47.1 | 0.00 (−0.31, 0.31) | 0.06 | 0.10 |
| Malawi | iLiNS−DOSE (45) | | | | | | | | | | | | |
| Mali | PROMIS CS (46) | | | | | | | | | | | | |
| Zimbabwe | SHINE (HIV−) (47) | | | | | | | | | | | | |
| Zimbabwe | SHINE (HIV+) (48) | | | | | | | | | | | | |
| | | **272** | **342** | | **I² = 0.38, Tau² = 0.00** | | | **391** | **226** | | **I² = 0.28, Tau² = 0.00** | | |
| **Fixed** | | | | | **−0.02 (−0.10, 0.06)** | | | | | | **−0.02 (−0.09, 0.05)** | | |
| **Random** | | | | | **−0.02 (−0.13, 0.08)** | | | | | | **0.00 (−0.10, 0.11)** | | |

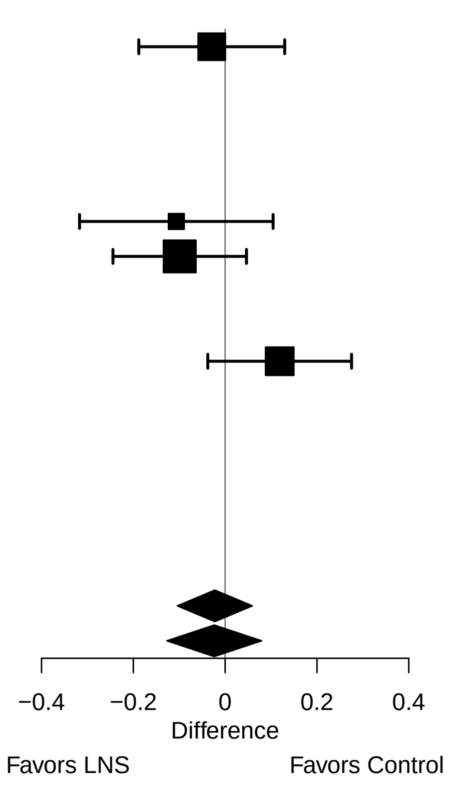

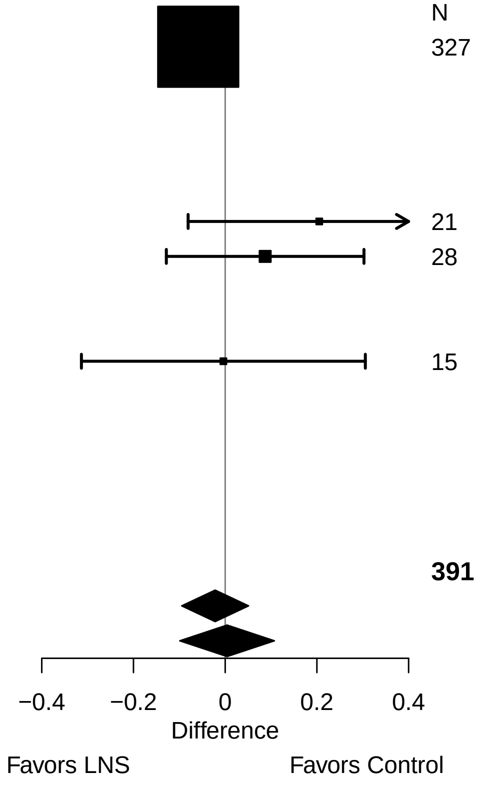

**Supplemental figure 6V: Marginal vitamin A (retinol < 1.05 µmol/L) prevalence difference**

#### 6V6: Stratified by Child baseline acute malnutrition (insufficient comparisons)

**Supplemental figure 6V: Marginal vitamin A (retinol < 1.05 µmol/L) prevalence difference**

#### 6V7: Stratified by Child baseline anemia (insufficient comparisons)

#### Supplemental figure 6V: Marginal vitamin A (retinol < 1.05 μmol/L) prevalence difference

#### 6V8: Stratified by Child high-dose vitamin A supplementation

**P−for−interaction = 0.534**
**Difference in PDs = −0.04 (−0.17, 0.09)**

| | | Received VitA | | | | | | Not treated | | | | | |
|---|---|---|---|---|---|---|---|---|---|---|---|---|---|
| | | **LNS** | **Control** | **Control** | **PD** | **Fixed** | **Random** | **LNS** | **Control** | **Control** | **PD** | **Fixed** | **Random** |
| Country | Trial | N | N | Prevalence | (95% CI) | W | W | N | N | Prevalence | (95% CI) | W | W |
| Bangladesh | JiVitA−4 (35) | 136 | 36 | 13.9 | −0.04 (−0.14, 0.07) | 0.54 | 0.54 | 129 | 38 | 18.4 | −0.06 (−0.19, 0.07) | 0.46 | 0.46 |
| Bangladesh | RDNS (36) | | | | | | | | | | | | |
| Bangladesh | WASH−B (37) | | | | | | | | | | | | |
| Burkina Faso | iLiNS−Zinc (38) | | | | | | | | | | | | |
| Burkina Faso | PROMIS CS (39) | | | | | | | | | | | | |
| Ghana | GHANA (40) | 27 | 47 | 53.2 | 0.06 (−0.18, 0.30) | 0.10 | 0.10 | 36 | 24 | 41.7 | 0.00 (−0.26, 0.26) | 0.11 | 0.11 |
| Ghana | iLiNS−DYADG (41) | 71 | 143 | 59.4 | −0.05 (−0.19, 0.10) | 0.29 | 0.29 | 25 | 53 | 49.1 | −0.05 (−0.29, 0.19) | 0.13 | 0.13 |
| Kenya | WASH−B (42) | | | | | | | | | | | | |
| Madagascar | MAHAY (43) | | | | | | | | | | | | |
| Malawi | iLiNS−DYADM (44) | 16 | 42 | 50.0 | 0.19 (−0.10, 0.47) | 0.07 | 0.07 | 54 | 119 | 47.1 | 0.07 (−0.09, 0.23) | 0.29 | 0.29 |
| Malawi | iLiNS−DOSE (45) | | | | | | | | | | | | |
| Mali | PROMIS CS (46) | | | | | | | | | | | | |
| Zimbabwe | SHINE (HIV−) (47) | | | | | | | | | | | | |
| Zimbabwe | SHINE (HIV+) (48) | | | | | | | | | | | | |
| | | **250** | **268** | | **I² = 0.00, Tau² = 0.00** | | | **244** | **234** | | **I² = 0.00, Tau² = 0.00** | | |
| **Fixed** | | | | | **−0.01 (−0.09, 0.06)** | | | | | | **−0.02 (−0.10, 0.07)** | | |
| **Random** | | | | | **−0.01 (−0.09, 0.06)** | | | | | | **−0.02 (−0.10, 0.07)** | | |

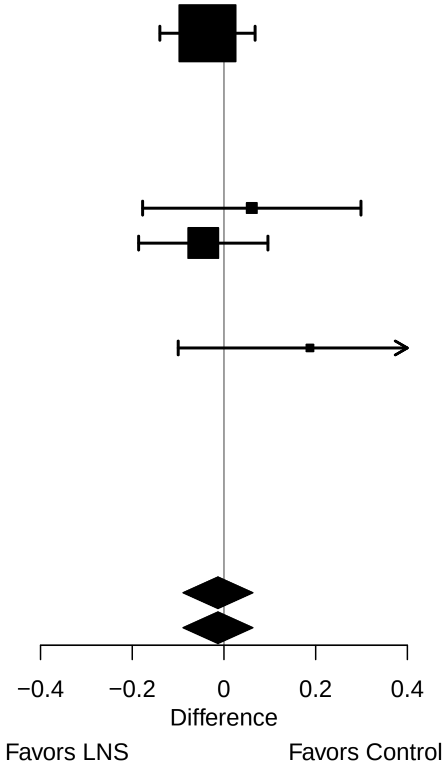

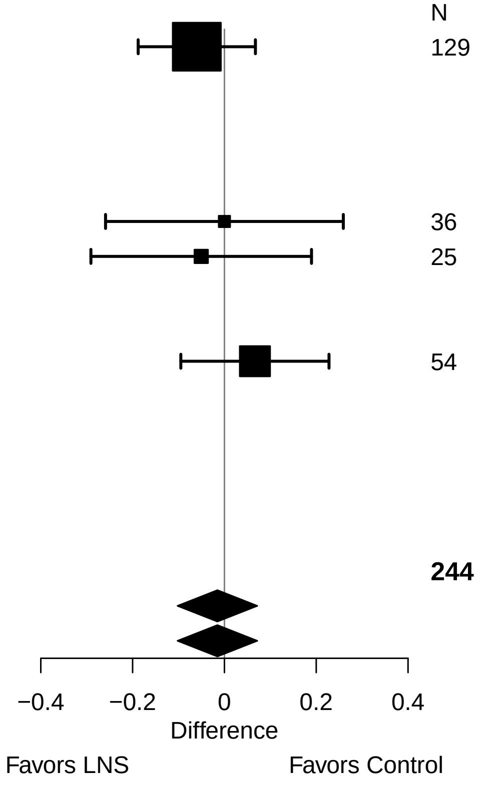

**Supplemental figure 6V: Marginal vitamin A (retinol < 1.05 µmol/L) prevalence difference**

#### 6V9: Stratified by Child inflammation

**P−for−interaction = 0.927**
**Difference in PDs = −0.01 (−0.13, 0.12)**

| | | Not inflamed | | | | | | High AGP or CRP | | | | | |
|---|---|---|---|---|---|---|---|---|---|---|---|---|---|
| | | **LNS** | **Control** | **Control** | **PD** | **Fixed** | **Random** | **LNS** | **Control** | **Control** | **PD** | **Fixed** | **Random** |
| Country | Trial | N | N | Prevalence | (95% CI) | W | W | N | N | Prevalence | (95% CI) | W | W |
| Bangladesh | JiVitA−4 (35) | 177 | 46 | 21.7 | −0.07 (−0.16, 0.02) | 0.72 | 0.43 | 257 | 93 | 18.3 | −0.04 (−0.14, 0.06) | 0.60 | 0.60 |
| Bangladesh | RDNS (36) | | | | | | | | | | | | |
| Bangladesh | WASH−B (37) | | | | | | | | | | | | |
| Burkina Faso | iLiNS−Zinc (38) | | | | | | | | | | | | |
| Burkina Faso | PROMIS CS (39) | | | | | | | | | | | | |
| Ghana | GHANA (40) | | | | | | | | | | | | |
| Ghana | iLiNS−DYADG (41) | 43 | 118 | 55.9 | −0.07 (−0.25, 0.10) | 0.19 | 0.33 | 53 | 78 | 57.7 | −0.03 (−0.20, 0.14) | 0.19 | 0.19 |
| Kenya | WASH−B (42) | | | | | | | | | | | | |
| Madagascar | MAHAY (43) | | | | | | | | | | | | |
| Malawi | iLiNS−DYADM (44) | 20 | 61 | 52.5 | 0.23 (−0.02, 0.47) | 0.09 | 0.25 | 51 | 102 | 46.1 | 0.05 (−0.12, 0.22) | 0.21 | 0.21 |
| Malawi | iLiNS−DOSE (45) | | | | | | | | | | | | |
| Mali | PROMIS CS (46) | | | | | | | | | | | | |
| Zimbabwe | SHINE (HIV−) (47) | | | | | | | | | | | | |
| Zimbabwe | SHINE (HIV+) (48) | | | | | | | | | | | | |
| | | **240** | **225** | | **I² = 0.60, Tau² = 0.02** | | | **361** | **273** | | **I² = 0.00, Tau² = 0.00** | | |
| **Fixed** | | | | | **−0.04 (−0.12, 0.03)** | | | | | | **−0.02 (−0.10, 0.06)** | | |
| **Random** | | | | | **0.00 (−0.17, 0.18)** | | | | | | **−0.02 (−0.10, 0.06)** | | |

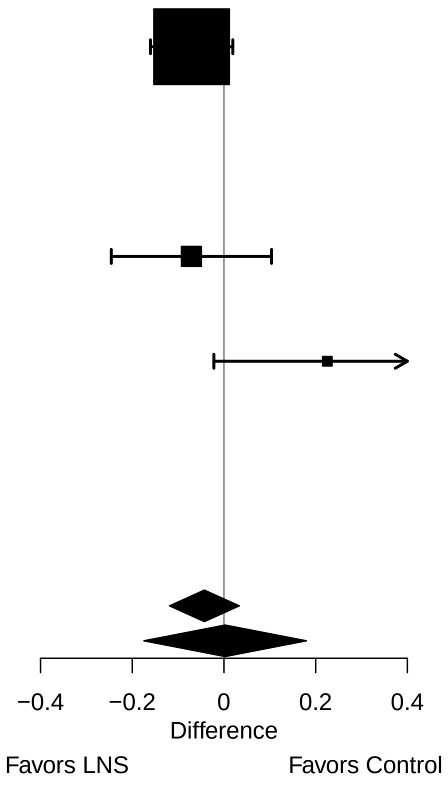

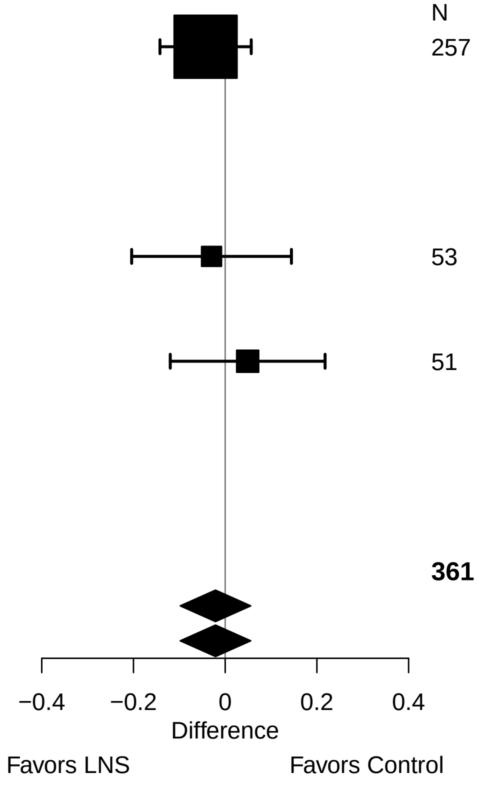

### Supplemental figure 6W: Geometric mean ratio of retinol binding protein concentration

#### 6W1: Stratified by Maternal BMI

**P−for−interaction = 0.572**
**Ratio of GMRs = 0.99 (0.95, 1.03)**

| | | At least 20 kg/m² | | | | | | Less than 20 kg/m² | | | | | |
|---|---|---|---|---|---|---|---|---|---|---|---|---|---|
| Country | Trial | LNS N | Control N | Control Median | GMR (95% CI) | Fixed W | Random W | LNS N | Control N | Control Median | GMR (95% CI) | Fixed W | Random W |
| Bangladesh | JiVitA−4 (35) | | | | | | | | | | | | |
| Bangladesh | RDNS (36) | 228 | 102 | 1.1 | 1.07 (1.00, 1.15) | 0.21 | 0.24 | 300 | 160 | 1.2 | 0.99 (0.95, 1.03) | 0.44 | 0.28 |
| Bangladesh | WASH−B (37) | 93 | 97 | 1.2 | 1.01 (0.93, 1.09) | 0.16 | 0.20 | 119 | 81 | 1.1 | 1.09 (1.03, 1.15) | 0.26 | 0.26 |
| Burkina Faso | iLiNS−Zinc (38) | 209 | 55 | 1.1 | 1.08 (1.02, 1.15) | 0.31 | 0.28 | 106 | 41 | 1.1 | 1.10 (1.03, 1.18) | 0.16 | 0.23 |
| Burkina Faso | PROMIS CS (39) | | | | | | | | | | | | |
| Ghana | GHANA (40) | | | | | | | | | | | | |
| Ghana | iLiNS−DYADG (41) | | | | | | | | | | | | |
| Kenya | WASH−B (42) | 223 | 200 | 1.0 | 1.14 (1.08, 1.21) | 0.32 | 0.28 | 64 | 51 | 1.0 | 1.14 (1.06, 1.23) | 0.14 | 0.23 |
| Madagascar | MAHAY (43) | | | | | | | | | | | | |
| Malawi | iLiNS−DYADM (44) | | | | | | | | | | | | |
| Malawi | iLiNS−DOSE (45) | | | | | | | | | | | | |
| Mali | PROMIS CS (46) | | | | | | | | | | | | |
| Zimbabwe | SHINE (HIV−) (47) | | | | | | | | | | | | |
| Zimbabwe | SHINE (HIV+) (48) | | | | | | | | | | | | |
| | | **753** | **454** | | **I² = 0.52, Tau² = 0.00** | | | **589** | **333** | | **I² = 0.83, Tau² = 0.00** | | |
| **Fixed** | | | | | **1.09 (1.05, 1.12)** | | | | | | **1.05 (1.02, 1.08)** | | |
| **Random** | | | | | **1.08 (1.03, 1.13)** | | | | | | **1.07 (1.01, 1.15)** | | |

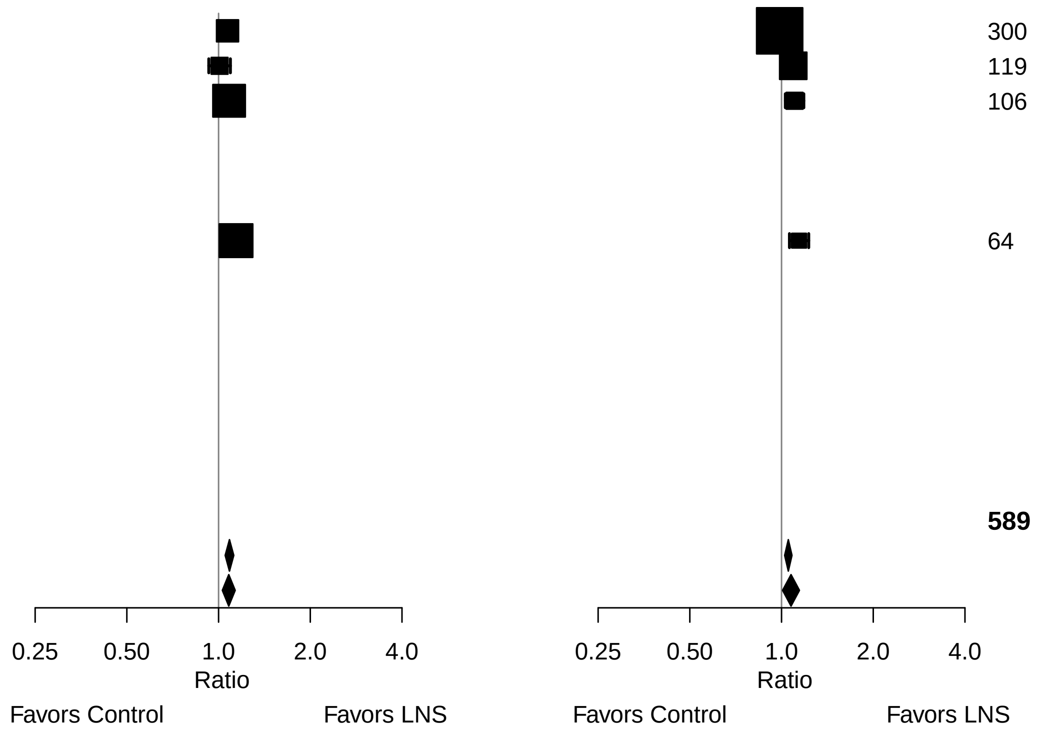

**Supplemental figure 6W: Geometric mean ratio of retinol binding protein concentration**

#### 6W2: Stratified by Maternal age

**P−for−interaction = 0.657**
**Ratio of GMRs = 0.99 (0.95, 1.03)**

| Country | Trial | At least 25 y LNS N | Control N | Control Median | GMR (95% CI) | Fixed W | Random W | Less than 25 y LNS N | Control N | Control Median | GMR (95% CI) | Fixed W | Random W |
|---|---|---|---|---|---|---|---|---|---|---|---|---|---|
| Bangladesh | JiVitA−4 (35) | | | | | | | | | | | | |
| Bangladesh | RDNS (36) | 152 | 81 | 1.2 | 1.07 (1.00, 1.15) | 0.24 | 0.23 | 398 | 191 | 1.2 | 1.00 (0.96, 1.05) | 0.43 | 0.28 |
| Bangladesh | WASH−B (37) | 79 | 80 | 1.1 | 1.02 (0.94, 1.11) | 0.18 | 0.19 | 132 | 98 | 1.2 | 1.06 (0.99, 1.13) | 0.20 | 0.22 |
| Burkina Faso | iLiNS−Zinc (38) | 178 | 50 | 1.0 | 1.08 (1.00, 1.17) | 0.17 | 0.19 | 137 | 46 | 1.1 | 1.09 (1.01, 1.18) | 0.15 | 0.19 |
| Burkina Faso | PROMIS CS (39) | | | | | | | | | | | | |
| Ghana | GHANA (40) | | | | | | | | | | | | |
| Ghana | iLiNS−DYADG (41) | | | | | | | | | | | | |
| Kenya | WASH−B (42) | 178 | 149 | 1.0 | 1.13 (1.07, 1.20) | 0.35 | 0.30 | 116 | 109 | 0.9 | 1.15 (1.06, 1.24) | 0.16 | 0.20 |
| Madagascar | MAHAY (43) | 47 | 25 | 1.1 | 0.99 (0.87, 1.13) | 0.06 | 0.09 | 36 | 26 | 1.1 | 1.11 (0.98, 1.26) | 0.06 | 0.11 |
| Malawi | iLiNS−DYADM (44) | | | | | | | | | | | | |
| Malawi | iLiNS−DOSE (45) | | | | | | | | | | | | |
| Mali | PROMIS CS (46) | | | | | | | | | | | | |
| Zimbabwe | SHINE (HIV−) (47) | | | | | | | | | | | | |
| Zimbabwe | SHINE (HIV+) (48) | | | | | | | | | | | | |
| | | **634** | **385** | | **I² = 0.34, Tau² = 0.00** | | | **819** | **470** | | **I² = 0.65, Tau² = 0.00** | | |
| **Fixed** | | | | | **1.08 (1.04, 1.12)** | | | | | | **1.05 (1.02, 1.09)** | | |
| **Random** | | | | | **1.07 (1.03, 1.12)** | | | | | | **1.07 (1.02, 1.12)** | | |

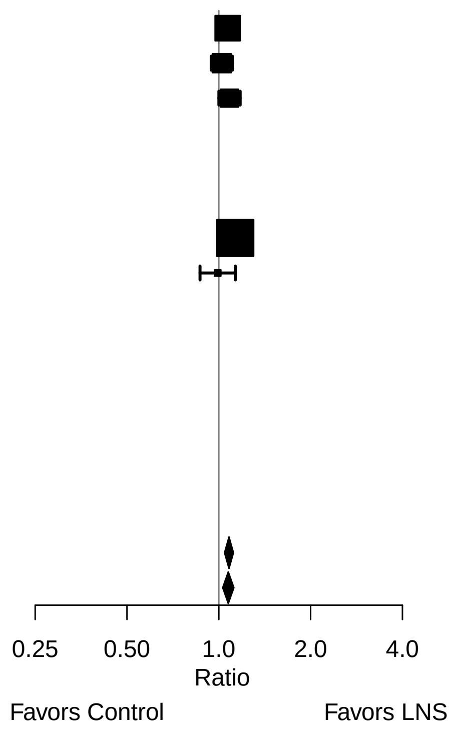

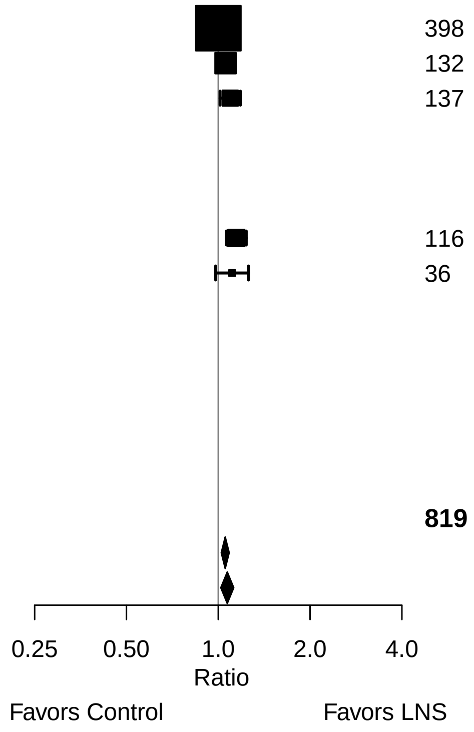

**Supplemental figure 6W: Geometric mean ratio of retinol binding protein concentration**

#### 6W3: Stratified by Maternal education

**P−for−interaction = 0.793**
**Ratio of GMRs = 1.01 (0.95, 1.07)**

| Country | Trial | Primary or greater: LNS N | Control N | Control Median | GMR (95% CI) | Fixed W | Random W | Incomplete or no formal: LNS N | Control N | Control Median | GMR (95% CI) | Fixed W | Random W |
|---|---|---|---|---|---|---|---|---|---|---|---|---|---|
| Bangladesh | JiVitA−4 (35) | | | | | | | | | | | | |
| Bangladesh | RDNS (36) | 406 | 195 | 1.2 | 1.00 (0.95, 1.06) | 0.43 | 0.33 | 144 | 77 | 1.2 | 1.06 (0.98, 1.15) | 0.26 | 0.26 |
| Bangladesh | WASH−B (37) | 149 | 139 | 1.2 | 1.05 (0.98, 1.12) | 0.28 | 0.29 | 63 | 39 | 1.1 | 1.04 (0.93, 1.15) | 0.17 | 0.19 |
| Burkina Faso | iLiNS−Zinc (38) | | | | | | | | | | | | |
| Burkina Faso | PROMIS CS (39) | | | | | | | | | | | | |
| Ghana | GHANA (40) | | | | | | | | | | | | |
| Ghana | iLiNS−DYADG (41) | | | | | | | | | | | | |
| Kenya | WASH−B (42) | 143 | 117 | 1.0 | 1.13 (1.05, 1.21) | 0.25 | 0.28 | 154 | 142 | 0.9 | 1.14 (1.07, 1.22) | 0.39 | 0.35 |
| Madagascar | MAHAY (43) | 16 | 16 | 1.1 | 1.15 (0.97, 1.37) | 0.04 | 0.09 | 67 | 35 | 1.1 | 1.02 (0.93, 1.13) | 0.18 | 0.20 |
| Malawi | iLiNS−DYADM (44) | | | | | | | | | | | | |
| Malawi | iLiNS−DOSE (45) | | | | | | | | | | | | |
| Mali | PROMIS CS (46) | | | | | | | | | | | | |
| Zimbabwe | SHINE (HIV−) (47) | | | | | | | | | | | | |
| Zimbabwe | SHINE (HIV+) (48) | | | | | | | | | | | | |
| | | **714** | **467** | | **I² = 0.64, Tau² = 0.00** | | | **428** | **293** | | **I² = 0.33, Tau² = 0.00** | | |
| **Fixed** | | | | | **1.05 (1.02, 1.09)** | | | | | | **1.08 (1.04, 1.13)** | | |
| **Random** | | | | | **1.06 (1.00, 1.13)** | | | | | | **1.08 (1.02, 1.13)** | | |

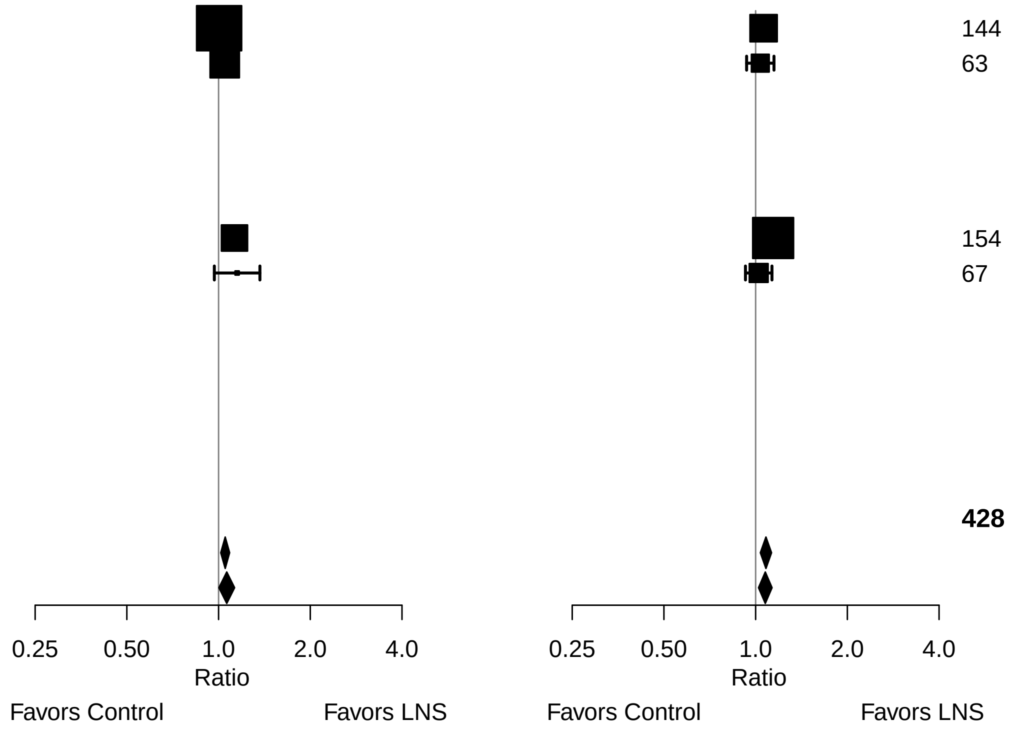

**Supplemental figure 6W: Geometric mean ratio of retinol binding protein concentration**

#### 6W4: Stratified by Child sex

**P−for−interaction = 0.100**
**Ratio of GMRs = 1.03 (0.99, 1.07)**

| Country | Trial | Male LNS N | Male Control N | Male Control Median | Male GMR (95% CI) | Male Fixed W | Male Random W | Female LNS N | Female Control N | Female Control Median | Female GMR (95% CI) | Female Fixed W | Female Random W |
|---|---|---|---|---|---|---|---|---|---|---|---|---|---|
| Bangladesh | JiVitA−4 (35) | | | | | | | | | | | | |
| Bangladesh | RDNS (36) | 266 | 131 | 1.2 | 1.00 (0.94, 1.07) | 0.28 | 0.24 | 281 | 141 | 1.2 | 1.04 (0.99, 1.08) | 0.41 | 0.33 |
| Bangladesh | WASH−B (37) | 105 | 85 | 1.2 | 1.00 (0.94, 1.07) | 0.24 | 0.23 | 107 | 93 | 1.1 | 1.08 (1.02, 1.15) | 0.18 | 0.20 |
| Burkina Faso | iLiNS−Zinc (38) | 150 | 49 | 1.0 | 1.07 (0.98, 1.16) | 0.15 | 0.18 | 165 | 47 | 1.1 | 1.11 (1.04, 1.18) | 0.19 | 0.20 |
| Burkina Faso | PROMIS CS (39) | | | | | | | | | | | | |
| Ghana | GHANA (40) | | | | | | | | | | | | |
| Ghana | iLiNS−DYADG (41) | | | | | | | | | | | | |
| Kenya | WASH−B (42) | 146 | 125 | 0.9 | 1.14 (1.07, 1.22) | 0.27 | 0.24 | 152 | 134 | 1.0 | 1.13 (1.06, 1.21) | 0.17 | 0.19 |
| Madagascar | MAHAY (43) | 34 | 22 | 1.1 | 1.03 (0.91, 1.18) | 0.07 | 0.11 | 49 | 29 | 1.1 | 1.06 (0.94, 1.19) | 0.05 | 0.07 |
| Malawi | iLiNS−DYADM (44) | | | | | | | | | | | | |
| Malawi | iLiNS−DOSE (45) | | | | | | | | | | | | |
| Mali | PROMIS CS (46) | | | | | | | | | | | | |
| Zimbabwe | SHINE (HIV−) (47) | | | | | | | | | | | | |
| Zimbabwe | SHINE (HIV+) (48) | | | | | | | | | | | | |
| | | **701** | **412** | | **I² = 0.63, Tau² = 0.00** | | | **754** | **444** | | **I² = 0.35, Tau² = 0.00** | | |
| **Fixed** | | | | | **1.05 (1.02, 1.09)** | | | | | | **1.07 (1.05, 1.10)** | | |
| **Random** | | | | | **1.05 (1.00, 1.11)** | | | | | | **1.08 (1.04, 1.11)** | | |

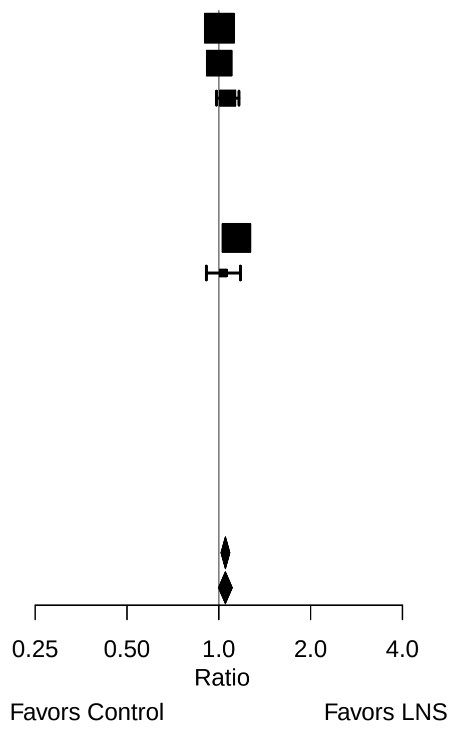

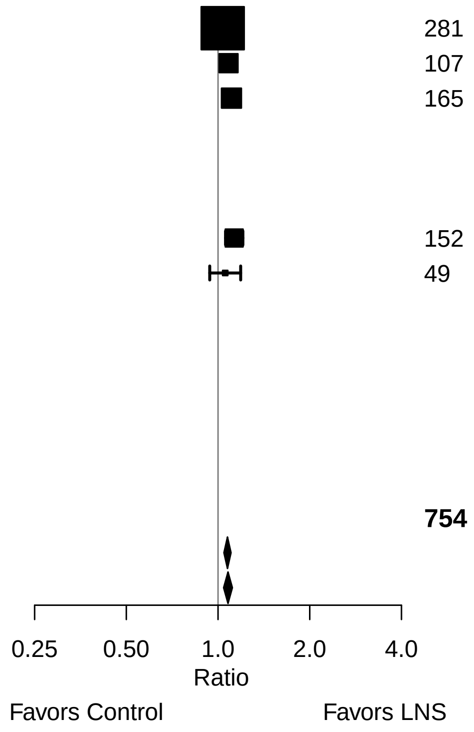

**Supplemental figure 6W: Geometric mean ratio of retinol binding protein concentration**

#### 6W5: Stratified by Child birth order

**P−for−interaction = 0.353**
**Ratio of GMRs = 0.98 (0.93, 1.03)**

| Country | Trial | Later born LNS N | Later born Control N | Later born Control Median | Later born GMR (95% CI) | Later born Fixed W | Later born Random W | Firstborn LNS N | Firstborn Control N | Firstborn Control Median | Firstborn GMR (95% CI) | Firstborn Fixed W | Firstborn Random W |
|---|---|---|---|---|---|---|---|---|---|---|---|---|---|
| Bangladesh | JiVitA−4 (35) | | | | | | | | | | | | |
| Bangladesh | RDNS (36) | 340 | 171 | 1.2 | 1.03 (0.98, 1.08) | 0.32 | 0.25 | 210 | 101 | 1.2 | 1.00 (0.93, 1.08) | 0.28 | 0.28 |
| Bangladesh | WASH−B (37) | 124 | 119 | 1.1 | 1.03 (0.96, 1.10) | 0.16 | 0.19 | 86 | 57 | 1.2 | 1.07 (0.98, 1.16) | 0.21 | 0.21 |
| Burkina Faso | iLiNS−Zinc (38) | 249 | 71 | 1.0 | 1.11 (1.04, 1.18) | 0.16 | 0.20 | 66 | 25 | 1.1 | 1.04 (0.95, 1.13) | 0.22 | 0.22 |
| Burkina Faso | PROMIS CS (39) | | | | | | | | | | | | |
| Ghana | GHANA (40) | | | | | | | | | | | | |
| Ghana | iLiNS−DYADG (41) | | | | | | | | | | | | |
| Kenya | WASH−B (42) | 237 | 213 | 1.0 | 1.15 (1.09, 1.20) | 0.30 | 0.24 | 60 | 46 | 0.9 | 1.10 (1.00, 1.21) | 0.17 | 0.17 |
| Madagascar | MAHAY (43) | 61 | 37 | 1.1 | 1.04 (0.93, 1.15) | 0.06 | 0.12 | 21 | 11 | 1.2 | 1.05 (0.93, 1.18) | 0.11 | 0.11 |
| Malawi | iLiNS−DYADM (44) | | | | | | | | | | | | |
| Malawi | iLiNS−DOSE (45) | | | | | | | | | | | | |
| Mali | PROMIS CS (46) | | | | | | | | | | | | |
| Zimbabwe | SHINE (HIV−) (47) | | | | | | | | | | | | |
| Zimbabwe | SHINE (HIV+) (48) | | | | | | | | | | | | |
| | | **1011** | **611** | | **I² = 0.68, Tau² = 0.00** | | | **443** | **240** | | **I² = 0.00, Tau² = 0.00** | | |
| **Fixed** | | | | | **1.08 (1.05, 1.11)** | | | | | | **1.05 (1.00, 1.09)** | | |
| **Random** | | | | | **1.07 (1.03, 1.12)** | | | | | | **1.05 (1.00, 1.09)** | | |

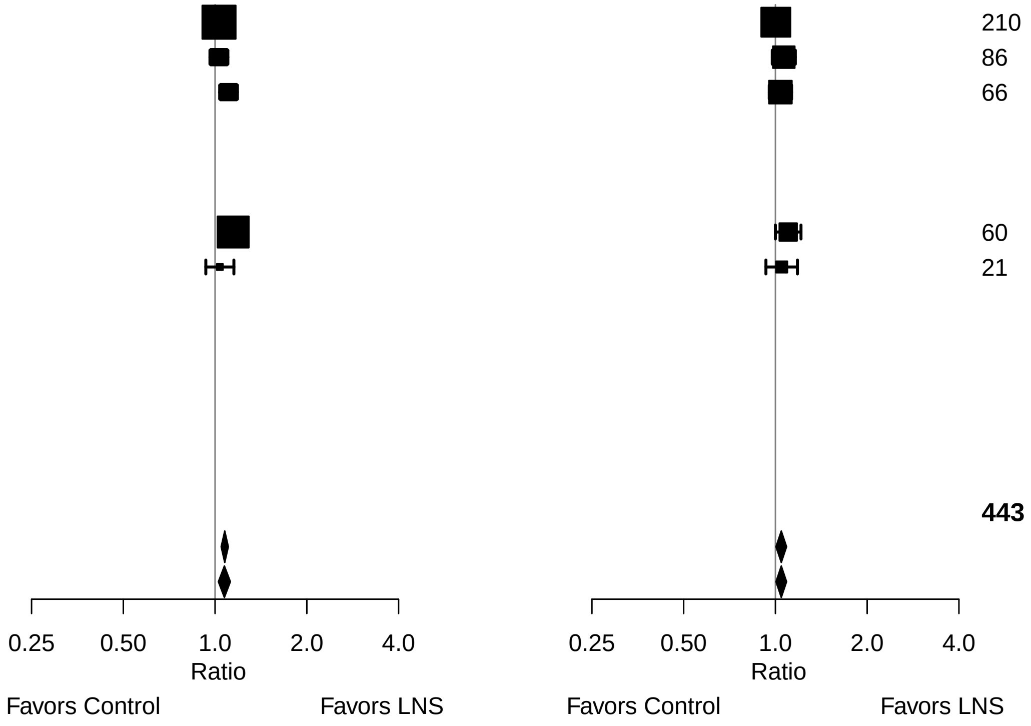

**Supplemental figure 6W: Geometric mean ratio of retinol binding protein concentration**

#### 6W6: Stratified by Child baseline acute malnutrition (insufficient comparisons)

**Supplemental figure 6W: Geometric mean ratio of retinol binding protein concentration**

#### 6W7: Stratified by Child baseline anemia (insufficient comparisons)

#### Supplemental figure 6W: Geometric mean ratio of retinol binding protein concentration

#### 6W8: Stratified by Child high-dose vitamin A supplementation

**P−for−interaction = 0.107**
**Ratio of GMRs = 0.94 (0.88, 1.01)**

| | | **Received VitA** | | | | | | **Not treated** | | | | | |
|---|---|---|---|---|---|---|---|---|---|---|---|---|---|
| Country | Trial | **LNS** N | **Control** N | **Control** Median | **GMR** (95% CI) | **Fixed** W | **Random** W | **LNS** N | **Control** N | **Control** Median | **GMR** (95% CI) | **Fixed** W | **Random** W |
| Bangladesh | JiVitA−4 (35) | | | | | | | | | | | | |
| Bangladesh | RDNS (36) | 444 | 223 | 1.2 | 1.02 (0.97, 1.06) | 0.80 | 0.40 | 105 | 49 | 1.2 | 1.04 (0.94, 1.14) | 0.22 | 0.34 |
| Bangladesh | WASH−B (37) | 16 | 16 | 1.0 | 1.21 (1.07, 1.37) | 0.11 | 0.31 | 15 | 9 | 1.2 | 0.91 (0.79, 1.06) | 0.09 | 0.24 |
| Burkina Faso | iLiNS−Zinc (38) | | | | | | | | | | | | |
| Burkina Faso | PROMIS CS (39) | | | | | | | | | | | | |
| Ghana | GHANA (40) | | | | | | | | | | | | |
| Ghana | iLiNS−DYADG (41) | | | | | | | | | | | | |
| Kenya | WASH−B (42) | 26 | 31 | 0.9 | 1.24 (1.08, 1.42) | 0.09 | 0.29 | 223 | 180 | 1.0 | 1.11 (1.05, 1.17) | 0.69 | 0.42 |
| Madagascar | MAHAY (43) | | | | | | | | | | | | |
| Malawi | iLiNS−DYADM (44) | | | | | | | | | | | | |
| Malawi | iLiNS−DOSE (45) | | | | | | | | | | | | |
| Mali | PROMIS CS (46) | | | | | | | | | | | | |
| Zimbabwe | SHINE (HIV−) (47) | | | | | | | | | | | | |
| Zimbabwe | SHINE (HIV+) (48) | | | | | | | | | | | | |
| | | **486** | **270** | | **I² = 0.85, Tau² = 0.01** | | | **343** | **238** | | **I² = 0.69, Tau² = 0.01** | | |
| **Fixed** | | | | | **1.05 (1.01, 1.10)** | | | | | | **1.07 (1.03, 1.12)** | | |
| **Random** | | | | | **1.14 (1.00, 1.29)** | | | | | | **1.03 (0.93, 1.15)** | | |

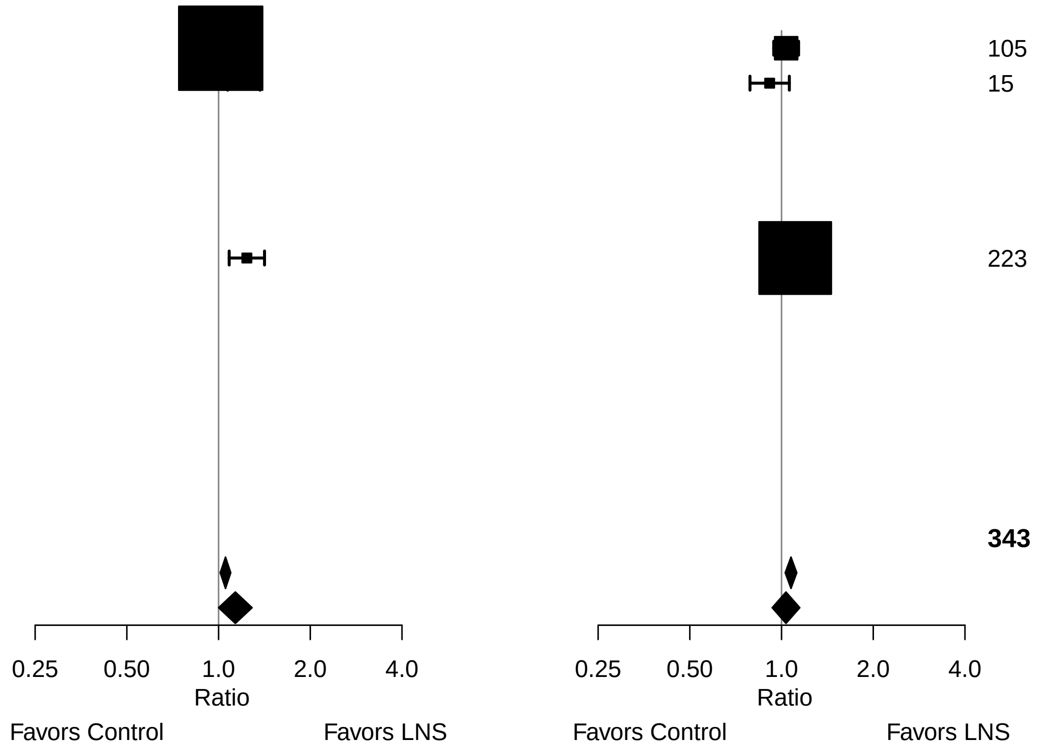

##### Supplemental figure 6W: Geometric mean ratio of retinol binding protein concentration

#### 6W9: Stratified by Child inflammation

**P−for−interaction = 0.088**
**Ratio of GMRs = 1.04 (0.99, 1.08)**

| | | Not inflamed | | | | | | High AGP or CRP | | | | | |
|---|---|---|---|---|---|---|---|---|---|---|---|---|---|
| Country | Trial | LNS N | Control N | Control Median | GMR (95% CI) | Fixed W | Random W | LNS N | Control N | Control Median | GMR (95% CI) | Fixed W | Random W |
| Bangladesh | JiVitA−4 (35) | | | | | | | | | | | | |
| Bangladesh | RDNS (36) | 370 | 168 | 1.2 | 1.00 (0.96, 1.04) | 0.40 | 0.27 | 180 | 104 | 1.1 | 1.06 (0.98, 1.14) | 0.22 | 0.22 |
| Bangladesh | WASH−B (37) | 165 | 136 | 1.2 | 1.05 (0.99, 1.11) | 0.20 | 0.22 | 47 | 42 | 1.1 | 1.02 (0.91, 1.14) | 0.11 | 0.15 |
| Burkina Faso | iLiNS−Zinc (38) | 124 | 31 | 1.1 | 1.13 (1.04, 1.22) | 0.10 | 0.16 | 191 | 65 | 1.1 | 1.07 (1.01, 1.15) | 0.33 | 0.27 |
| Burkina Faso | PROMIS CS (39) | | | | | | | | | | | | |
| Ghana | GHANA (40) | | | | | | | | | | | | |
| Ghana | iLiNS−DYADG (41) | | | | | | | | | | | | |
| Kenya | WASH−B (42) | 157 | 135 | 1.0 | 1.09 (1.04, 1.15) | 0.25 | 0.24 | 141 | 124 | 0.9 | 1.18 (1.10, 1.27) | 0.27 | 0.25 |
| Madagascar | MAHAY (43) | 44 | 32 | 1.1 | 1.00 (0.89, 1.12) | 0.05 | 0.10 | 39 | 19 | 1.1 | 1.12 (0.98, 1.29) | 0.07 | 0.11 |
| Malawi | iLiNS−DYADM (44) | | | | | | | | | | | | |
| Malawi | iLiNS−DOSE (45) | | | | | | | | | | | | |
| Mali | PROMIS CS (46) | | | | | | | | | | | | |
| Zimbabwe | SHINE (HIV−) (47) | | | | | | | | | | | | |
| Zimbabwe | SHINE (HIV+) (48) | | | | | | | | | | | | |
| | | **860** | **502** | | **I² = 0.64, Tau² = 0.00** | | | **598** | **354** | | **I² = 0.49, Tau² = 0.00** | | |
| **Fixed** | | | | | **1.04 (1.02, 1.07)** | | | | | | **1.10 (1.06, 1.14)** | | |
| **Random** | | | | | **1.05 (1.00, 1.10)** | | | | | | **1.09 (1.04, 1.15)** | | |

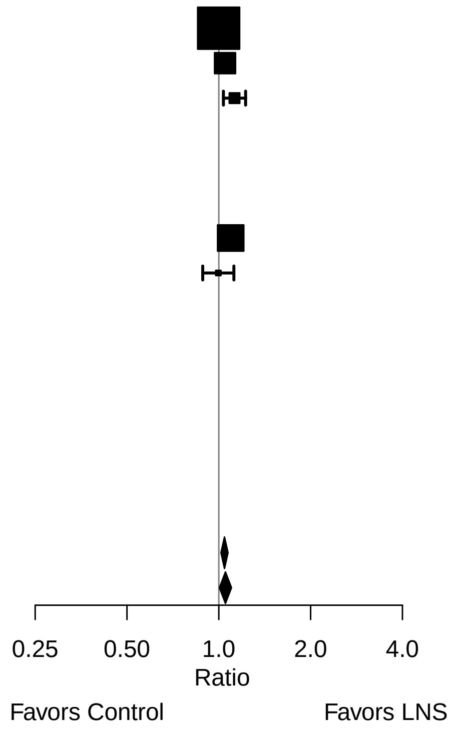

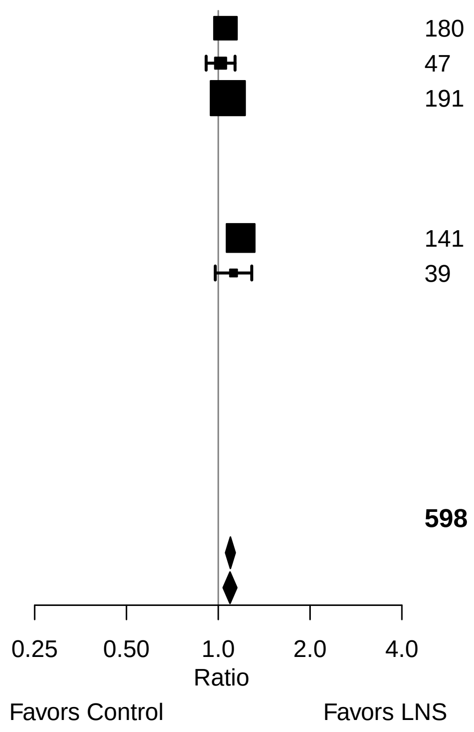

### Supplemental figure 6X: Low vitamin A status (RBP < 0.70 µmol/L) prevalence ratio

#### 6X1: Stratified by Maternal BMI (insufficient comparisons)

**Supplemental figure 6X: Low vitamin A status (RBP < 0.70 µmol/L) prevalence ratio**

#### 6X2: Stratified by Maternal age (insufficient comparisons)

**Supplemental figure 6X: Low vitamin A status (RBP < 0.70 µmol/L) prevalence ratio**

#### 6X3: Stratified by Maternal education (insufficient comparisons)

**Supplemental figure 6X: Low vitamin A status (RBP < 0.70 µmol/L) prevalence ratio**

#### 6X4: Stratified by Child sex (insufficient comparisons)

**Supplemental figure 6X: Low vitamin A status (RBP < 0.70 µmol/L) prevalence ratio**

#### 6X5: Stratified by Child birth order (insufficient comparisons)

**Supplemental figure 6X: Low vitamin A status (RBP < 0.70 µmol/L) prevalence ratio**

#### 6X6: Stratified by Child baseline acute malnutrition (insufficient comparisons)

**Supplemental figure 6X: Low vitamin A status (RBP < 0.70 µmol/L) prevalence ratio**

#### 6X7: Stratified by Child baseline anemia (insufficient comparisons)

**Supplemental figure 6X: Low vitamin A status (RBP < 0.70 µmol/L) prevalence ratio**

#### 6X8: Stratified by Child high-dose vitamin A supplementation (insufficient comparisons)

**Supplemental figure 6X: Low vitamin A status (RBP < 0.70 µmol/L) prevalence ratio**

#### 6X9: Stratified by Child inflammation (insufficient comparisons)

#### Supplemental figure 6Y: Low vitamin A status (RBP < 0.70 μmol/L) prevalence difference

##### 6Y1: Stratified by Maternal BMI (insufficient comparisons)

**Supplemental figure 6Y: Low vitamin A status (RBP < 0.70 µmol/L) prevalence difference**

#### 6Y2: Stratified by Maternal age (insufficient comparisons)

**Supplemental figure 6Y: Low vitamin A status (RBP < 0.70 µmol/L) prevalence difference**

#### 6Y3: Stratified by Maternal education (insufficient comparisons)

**Supplemental figure 6Y: Low vitamin A status (RBP $< 0.70$ µmol/L) prevalence difference**

#### 6Y4: Stratified by Child sex (insufficient comparisons)

**Supplemental figure 6Y: Low vitamin A status (RBP < 0.70 µmol/L) prevalence difference**

#### 6Y5: Stratified by Child birth order (insufficient comparisons)

**Supplemental figure 6Y: Low vitamin A status (RBP < 0.70 µmol/L) prevalence difference**

#### 6Y6: Stratified by Child baseline acute malnutrition (insufficient comparisons)

**Supplemental figure 6Y: Low vitamin A status (RBP < 0.70 µmol/L) prevalence difference**

#### 6Y7: Stratified by Child baseline anemia (insufficient comparisons)

**Supplemental figure 6Y: Low vitamin A status (RBP < 0.70 µmol/L) prevalence difference**

#### 6Y8: Stratified by Child high-dose vitamin A supplementation (insufficient comparisons)

**Supplemental figure 6Y: Low vitamin A status (RBP < 0.70 µmol/L) prevalence difference**

#### 6Y9: Stratified by Child inflammation (insufficient comparisons)

### Supplemental figure 6Z: Marginal vitamin A status (RBP < 1.05 μmol/L) prevalence ratio

#### 6Z1: Stratified by Maternal BMI

**P−for−interaction = 0.327**
**Ratio of PRs = 1.13 (0.89, 1.43)**

| Country | Trial | At least 20 kg/m²: LNS N | Control N | Control Prevalence | PR (95% CI) | Fixed W | Random W | Less than 20 kg/m²: LNS N | Control N | Control Prevalence | PR (95% CI) | Fixed W | Random W |
|---|---|---|---|---|---|---|---|---|---|---|---|---|---|
| Bangladesh | JiVitA−4 (35) | | | | | | | | | | | | |
| Bangladesh | RDNS (36) | 228 | 102 | 34.3 | 0.66 (0.47, 0.94) | 0.18 | 0.18 | 300 | 160 | 27.5 | 1.01 (0.71, 1.42) | 0.26 | 0.26 |
| Bangladesh | WASH−B (37) | 93 | 97 | 30.9 | 0.76 (0.46, 1.27) | 0.08 | 0.08 | 119 | 81 | 40.7 | 0.87 (0.61, 1.23) | 0.25 | 0.25 |
| Burkina Faso | iLiNS−Zinc (38) | 209 | 55 | 49.2 | 0.76 (0.55, 1.06) | 0.20 | 0.20 | 106 | 41 | 48.9 | 0.76 (0.54, 1.07) | 0.26 | 0.26 |
| Burkina Faso | PROMIS CS (39) | | | | | | | | | | | | |
| Ghana | GHANA (40) | | | | | | | | | | | | |
| Ghana | iLiNS−DYADG (41) | | | | | | | | | | | | |
| Kenya | WASH−B (42) | 223 | 200 | 61.5 | 0.72 (0.59, 0.88) | 0.54 | 0.54 | 64 | 51 | 56.9 | 0.69 (0.48, 0.98) | 0.23 | 0.23 |
| Madagascar | MAHAY (43) | | | | | | | | | | | | |
| Malawi | iLiNS−DYADM (44) | | | | | | | | | | | | |
| Malawi | iLiNS−DOSE (45) | | | | | | | | | | | | |
| Mali | PROMIS CS (46) | | | | | | | | | | | | |
| Zimbabwe | SHINE (HIV−) (47) | | | | | | | | | | | | |
| Zimbabwe | SHINE (HIV+) (48) | | | | | | | | | | | | |
| | | **753** | **454** | | **I² = 0.00, Tau² = 0.00** | | | **589** | **333** | | **I² = 0.00, Tau² = 0.00** | | |
| **Fixed** | | | | | **0.72 (0.62, 0.84)** | | | | | | **0.82 (0.69, 0.98)** | | |
| **Random** | | | | | **0.72 (0.62, 0.84)** | | | | | | **0.82 (0.69, 0.98)** | | |

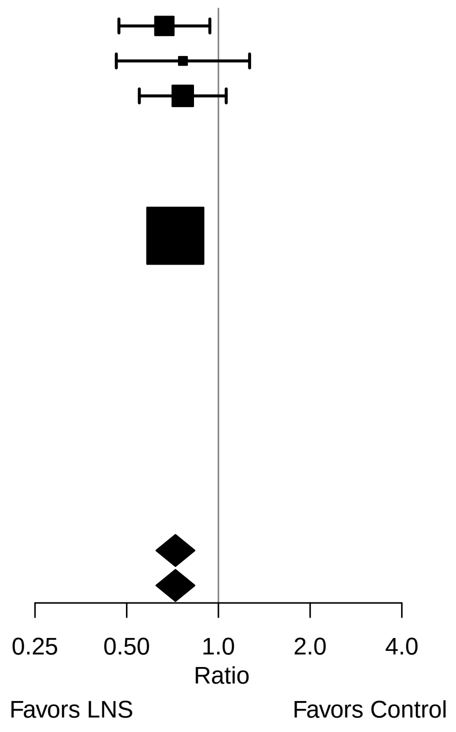

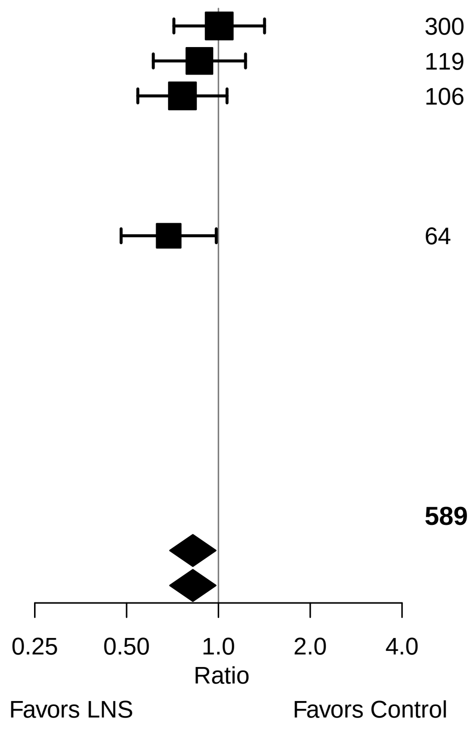

#### Supplemental figure 6Z: Marginal vitamin A status (RBP < 1.05 μmol/L) prevalence ratio

##### 6Z2: Stratified by Maternal age

**P−for−interaction = 0.255**
**Ratio of PRs = 0.89 (0.72, 1.09)**

| | | At least 25 y | | | | | | Less than 25 y | | | | | |
|---|---|---|---|---|---|---|---|---|---|---|---|---|---|
| Country | Trial | LNS N | Control N | Control Prevalence | PR (95% CI) | Fixed W | Random W | LNS N | Control N | Control Prevalence | PR (95% CI) | Fixed W | Random W |
| Bangladesh | JiVitA−4 (35) | | | | | | | | | | | | |
| Bangladesh | RDNS (36) | 152 | 81 | 29.6 | 0.87 (0.54, 1.39) | 0.10 | 0.10 | 398 | 191 | 28.8 | 0.87 (0.68, 1.11) | 0.35 | 0.34 |
| Bangladesh | WASH−B (37) | 79 | 80 | 38.8 | 1.05 (0.71, 1.54) | 0.14 | 0.14 | 132 | 98 | 32.7 | 0.72 (0.45, 1.16) | 0.09 | 0.09 |
| Burkina Faso | iLiNS−Zinc (38) | 178 | 50 | 50.0 | 0.72 (0.55, 0.93) | 0.30 | 0.30 | 137 | 46 | 48.1 | 0.83 (0.61, 1.12) | 0.22 | 0.23 |
| Burkina Faso | PROMIS CS (39) | | | | | | | | | | | | |
| Ghana | GHANA (40) | | | | | | | | | | | | |
| Ghana | iLiNS−DYADG (41) | | | | | | | | | | | | |
| Kenya | WASH−B (42) | 178 | 149 | 55.0 | 0.82 (0.65, 1.03) | 0.41 | 0.41 | 116 | 109 | 66.1 | 0.61 (0.47, 0.80) | 0.31 | 0.30 |
| Madagascar | MAHAY (43) | 47 | 25 | 40.0 | 0.90 (0.47, 1.74) | 0.05 | 0.05 | 36 | 26 | 38.5 | 0.72 (0.33, 1.58) | 0.03 | 0.04 |
| Malawi | iLiNS−DYADM (44) | | | | | | | | | | | | |
| Malawi | iLiNS−DOSE (45) | | | | | | | | | | | | |
| Mali | PROMIS CS (46) | | | | | | | | | | | | |
| Zimbabwe | SHINE (HIV−) (47) | | | | | | | | | | | | |
| Zimbabwe | SHINE (HIV+) (48) | | | | | | | | | | | | |
| | | **634** | **385** | | **I² = 0.00, Tau² = 0.00** | | | **819** | **470** | | **I² = 0.05, Tau² = 0.00** | | |
| **Fixed** | | | | | **0.82 (0.71, 0.95)** | | | | | | **0.76 (0.65, 0.87)** | | |
| **Random** | | | | | **0.82 (0.71, 0.95)** | | | | | | **0.76 (0.65, 0.88)** | | |

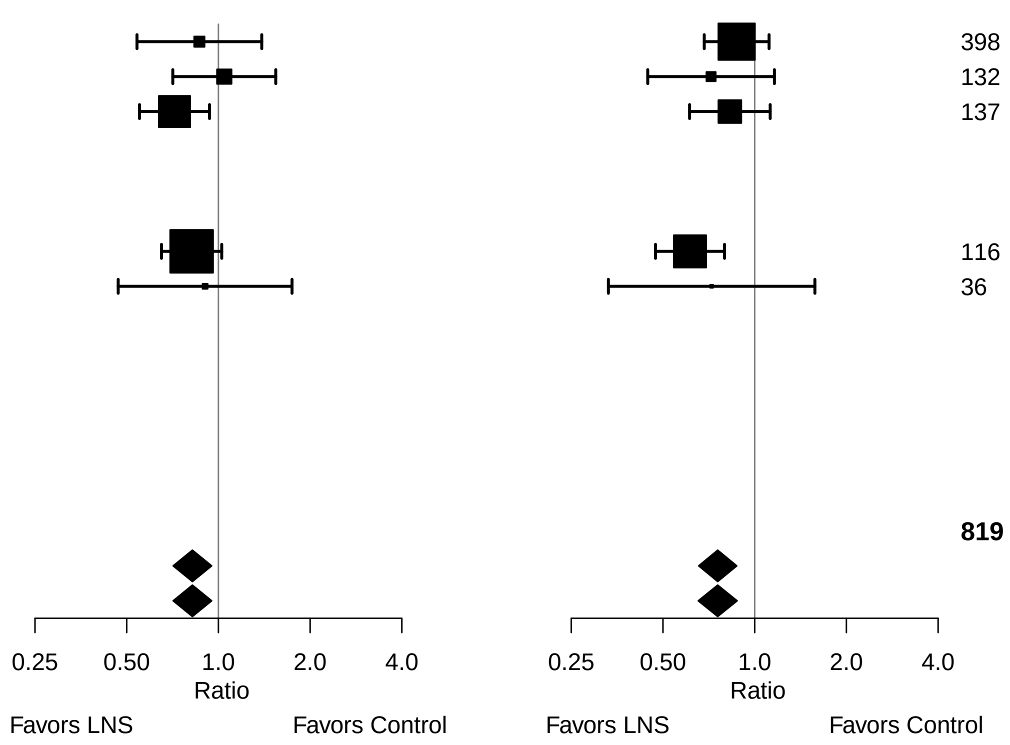

**Supplemental figure 6Z: Marginal vitamin A status (RBP < 1.05 µmol/L) prevalence ratio**

#### 6Z3: Stratified by Maternal education

**P−for−interaction = 0.772**
**Ratio of PRs = 1.04 (0.79, 1.38)**

| Country | Trial | Primary or greater: LNS N | Control N | Control Prevalence | PR (95% CI) | Fixed W | Random W | Incomplete or no formal: LNS N | Control N | Control Prevalence | PR (95% CI) | Fixed W | Random W |
|---|---|---|---|---|---|---|---|---|---|---|---|---|---|
| Bangladesh | JiVitA−4 (35) | | | | | | | | | | | | |
| Bangladesh | RDNS (36) | 406 | 195 | 27.2 | 0.87 (0.67, 1.13) | 0.42 | 0.42 | 144 | 77 | 33.8 | 0.88 (0.54, 1.45) | 0.16 | 0.16 |
| Bangladesh | WASH−B (37) | 149 | 139 | 35.3 | 0.82 (0.58, 1.15) | 0.24 | 0.24 | 63 | 39 | 35.9 | 0.93 (0.53, 1.62) | 0.12 | 0.12 |
| Burkina Faso | iLiNS−Zinc (38) | | | | | | | | | | | | |
| Burkina Faso | PROMIS CS (39) | | | | | | | | | | | | |
| Ghana | GHANA (40) | | | | | | | | | | | | |
| Ghana | iLiNS−DYADG (41) | | | | | | | | | | | | |
| Kenya | WASH−B (42) | 143 | 117 | 53.0 | 0.71 (0.53, 0.95) | 0.33 | 0.33 | 154 | 142 | 64.8 | 0.73 (0.58, 0.92) | 0.72 | 0.72 |
| Madagascar | MAHAY (43) | | | | | | | | | | | | |
| Malawi | iLiNS−DYADM (44) | | | | | | | | | | | | |
| Malawi | iLiNS−DOSE (45) | | | | | | | | | | | | |
| Mali | PROMIS CS (46) | | | | | | | | | | | | |
| Zimbabwe | SHINE (HIV−) (47) | | | | | | | | | | | | |
| Zimbabwe | SHINE (HIV+) (48) | | | | | | | | | | | | |
| | | **698** | **451** | | **I² = 0.00, Tau² = 0.00** | | | **361** | **258** | | **I² = 0.00, Tau² = 0.00** | | |
| **Fixed** | | | | | **0.80 (0.68, 0.95)** | | | | | | **0.78 (0.64, 0.94)** | | |
| **Random** | | | | | **0.80 (0.68, 0.95)** | | | | | | **0.78 (0.64, 0.94)** | | |

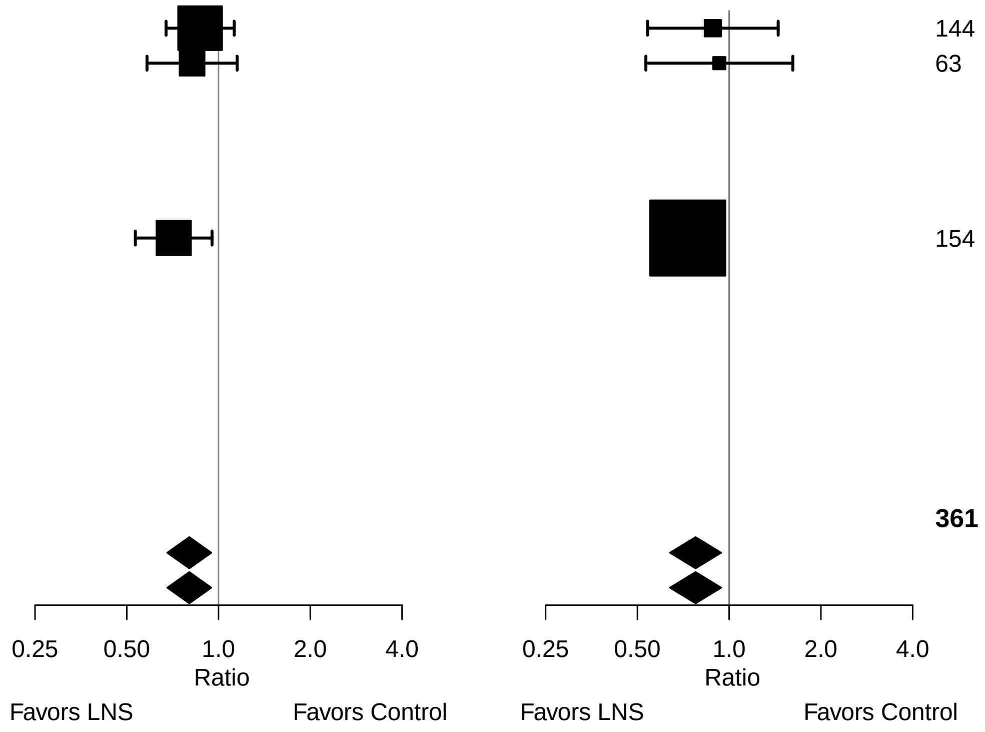

**Supplemental figure 6Z: Marginal vitamin A status (RBP < 1.05 µmol/L) prevalence ratio**

#### 6Z4: Stratified by Child sex

**P−for−interaction = 0.345**
**Ratio of PRs = 0.91 (0.74, 1.11)**

| Country | Trial | Male LNS N | Male Control N | Male Control Prevalence | Male PR (95% CI) | Male Fixed W | Male Random W | Female LNS N | Female Control N | Female Control Prevalence | Female PR (95% CI) | Female Fixed W | Female Random W |
|---|---|---|---|---|---|---|---|---|---|---|---|---|---|
| Bangladesh | JiVitA−4 (35) | | | | | | | | | | | | |
| Bangladesh | RDNS (36) | 266 | 131 | 27.5 | 0.96 (0.68, 1.36) | 0.19 | 0.19 | 281 | 141 | 30.5 | 0.78 (0.58, 1.05) | 0.25 | 0.25 |
| Bangladesh | WASH−B (37) | 105 | 85 | 32.9 | 1.04 (0.70, 1.55) | 0.15 | 0.15 | 107 | 93 | 37.6 | 0.70 (0.47, 1.03) | 0.14 | 0.14 |
| Burkina Faso | iLiNS−Zinc (38) | 150 | 49 | 57.4 | 0.74 (0.55, 0.99) | 0.27 | 0.27 | 165 | 47 | 40.4 | 0.83 (0.61, 1.12) | 0.23 | 0.23 |
| Burkina Faso | PROMIS CS (39) | | | | | | | | | | | | |
| Ghana | GHANA (40) | | | | | | | | | | | | |
| Ghana | iLiNS−DYADG (41) | | | | | | | | | | | | |
| Kenya | WASH−B (42) | 146 | 125 | 61.6 | 0.72 (0.56, 0.93) | 0.36 | 0.36 | 152 | 134 | 57.5 | 0.72 (0.56, 0.93) | 0.33 | 0.33 |
| Madagascar | MAHAY (43) | 34 | 22 | 31.8 | 0.92 (0.41, 2.08) | 0.04 | 0.04 | 49 | 29 | 44.8 | 0.77 (0.43, 1.38) | 0.06 | 0.06 |
| Malawi | iLiNS−DYADM (44) | | | | | | | | | | | | |
| Malawi | iLiNS−DOSE (45) | | | | | | | | | | | | |
| Mali | PROMIS CS (46) | | | | | | | | | | | | |
| Zimbabwe | SHINE (HIV−) (47) | | | | | | | | | | | | |
| Zimbabwe | SHINE (HIV+) (48) | | | | | | | | | | | | |
| | | **701** | **412** | | **I² = 0.00, Tau² = 0.00** | | | **754** | **444** | | **I² = 0.00, Tau² = 0.00** | | |
| **Fixed** | | | | | **0.82 (0.70, 0.95)** | | | | | | **0.76 (0.66, 0.88)** | | |
| **Random** | | | | | **0.82 (0.70, 0.95)** | | | | | | **0.76 (0.66, 0.88)** | | |

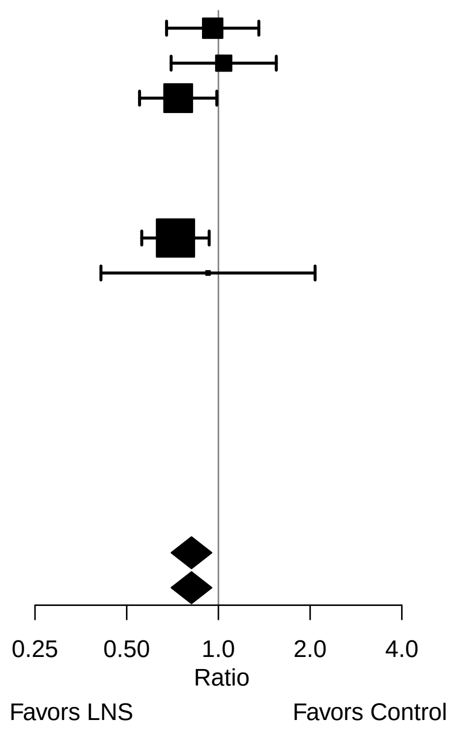

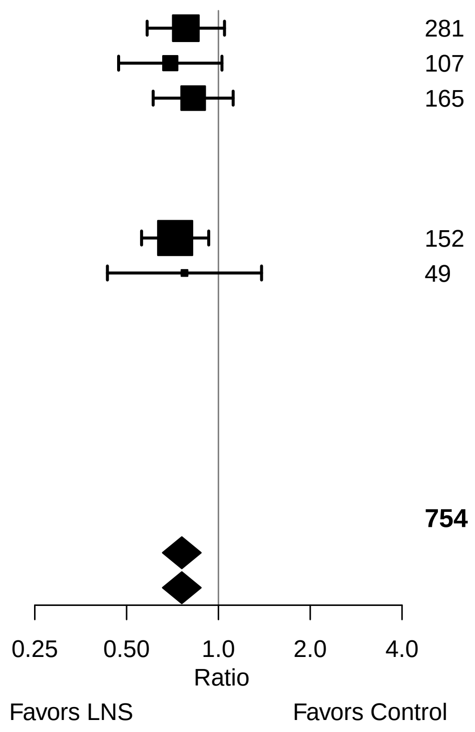

#### Supplemental figure 6Z: Marginal vitamin A status (RBP < 1.05 μmol/L) prevalence ratio

##### 6Z5: Stratified by Child birth order

**P−for−interaction = 0.310**
**Ratio of PRs = 1.13 (0.89, 1.44)**

| | | Later born | | | | | | Firstborn | | | | | |
|---|---|---|---|---|---|---|---|---|---|---|---|---|---|
| Country | Trial | LNS N | Control N | Control Prevalence | PR (95% CI) | Fixed W | Random W | LNS N | Control N | Control Prevalence | PR (95% CI) | Fixed W | Random W |
| Bangladesh | JiVitA−4 (35) | | | | | | | | | | | | |
| Bangladesh | RDNS (36) | 340 | 171 | 30.4 | 0.82 (0.59, 1.14) | 0.14 | 0.14 | 210 | 101 | 26.7 | 0.96 (0.66, 1.40) | 0.30 | 0.30 |
| Bangladesh | WASH−B (37) | 124 | 119 | 38.7 | 0.92 (0.69, 1.22) | 0.19 | 0.19 | 86 | 57 | 29.8 | 0.74 (0.39, 1.41) | 0.10 | 0.10 |
| Burkina Faso | iLiNS−Zinc (38) | 249 | 71 | 53.3 | 0.70 (0.55, 0.88) | 0.29 | 0.29 | 66 | 25 | 38.7 | 1.05 (0.65, 1.71) | 0.18 | 0.18 |
| Burkina Faso | PROMIS CS (39) | | | | | | | | | | | | |
| Ghana | GHANA (40) | | | | | | | | | | | | |
| Ghana | iLiNS−DYADG (41) | | | | | | | | | | | | |
| Kenya | WASH−B (42) | 237 | 213 | 58.2 | 0.70 (0.57, 0.86) | 0.38 | 0.38 | 60 | 46 | 65.2 | 0.77 (0.56, 1.05) | 0.42 | 0.42 |
| Madagascar | MAHAY (43) | | | | | | | | | | | | |
| Malawi | iLiNS−DYADM (44) | | | | | | | | | | | | |
| Malawi | iLiNS−DOSE (45) | | | | | | | | | | | | |
| Mali | PROMIS CS (46) | | | | | | | | | | | | |
| Zimbabwe | SHINE (HIV−) (47) | | | | | | | | | | | | |
| Zimbabwe | SHINE (HIV+) (48) | | | | | | | | | | | | |
| | | **950** | **574** | | **I² = 0.00, Tau² = 0.00** | | | **422** | **229** | | **I² = 0.00, Tau² = 0.00** | | |
| **Fixed** | | | | | **0.75 (0.67, 0.85)** | | | | | | **0.86 (0.70, 1.06)** | | |
| **Random** | | | | | **0.75 (0.67, 0.85)** | | | | | | **0.86 (0.70, 1.06)** | | |

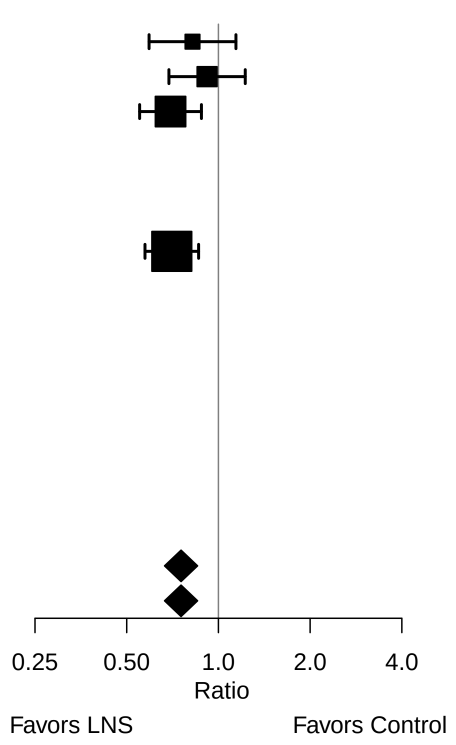

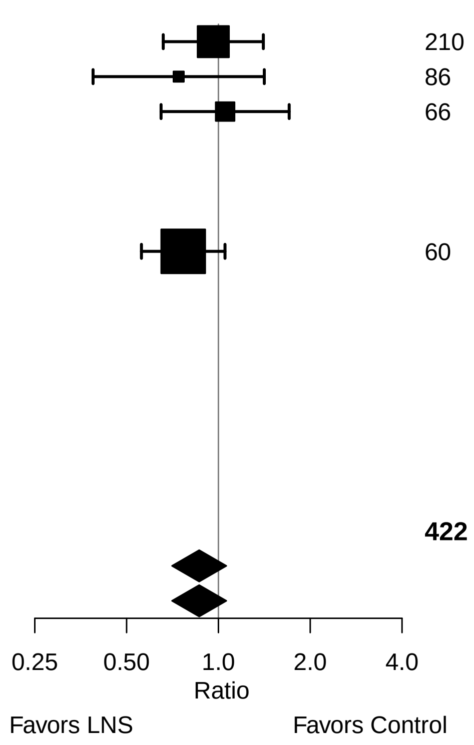

**Supplemental figure 6Z: Marginal vitamin A status (RBP < 1.05 µmol/L) prevalence ratio**

#### 6Z6: Stratified by Child baseline acute malnutrition (insufficient comparisons)

**Supplemental figure 6Z: Marginal vitamin A status (RBP < 1.05 µmol/L) prevalence ratio**

#### 6Z7: Stratified by Child baseline anemia (insufficient comparisons)

**Supplemental figure 6Z: Marginal vitamin A status (RBP < 1.05 µmol/L) prevalence ratio**

#### 6Z8: Stratified by Child high-dose vitamin A supplementation (insufficient comparisons)

**Supplemental figure 6Z: Marginal vitamin A status (RBP < 1.05 µmol/L) prevalence ratio**

#### 6Z9: Stratified by Child inflammation

**P−for−interaction = 0.223**
**Ratio of PRs = 0.87 (0.69, 1.09)**

| | | Not inflamed | | | | | | High AGP or CRP | | | | | |
|---|---|---|---|---|---|---|---|---|---|---|---|---|---|
| | | **LNS** | **Control** | **Control** | **PR** | **Fixed** | **Random** | **LNS** | **Control** | **Control** | **PR** | **Fixed** | **Random** |
| Country | Trial | N | N | Prevalence | (95% CI) | W | W | N | N | Prevalence | (95% CI) | W | W |
| Bangladesh | JiVitA−4 (35) | | | | | | | | | | | | |
| Bangladesh | RDNS (36) | 370 | 168 | 23.8 | 1.02 (0.68, 1.53) | 0.14 | 0.14 | 180 | 104 | 37.5 | 0.73 (0.54, 0.98) | 0.27 | 0.27 |
| Bangladesh | WASH−B (37) | 165 | 136 | 31.6 | 0.94 (0.63, 1.40) | 0.14 | 0.14 | 47 | 42 | 47.6 | 0.67 (0.38, 1.18) | 0.08 | 0.08 |
| Burkina Faso | iLiNS−Zinc (38) | 124 | 31 | 50.0 | 0.73 (0.53, 0.99) | 0.24 | 0.24 | 191 | 65 | 48.6 | 0.77 (0.57, 1.04) | 0.27 | 0.27 |
| Burkina Faso | PROMIS CS (39) | | | | | | | | | | | | |
| Ghana | GHANA (40) | | | | | | | | | | | | |
| Ghana | iLiNS−DYADG (41) | | | | | | | | | | | | |
| Kenya | WASH−B (42) | 157 | 135 | 56.3 | 0.77 (0.61, 0.97) | 0.43 | 0.43 | 141 | 124 | 62.9 | 0.68 (0.52, 0.88) | 0.34 | 0.34 |
| Madagascar | MAHAY (43) | 44 | 32 | 37.5 | 0.85 (0.42, 1.72) | 0.05 | 0.05 | 39 | 19 | 42.1 | 0.79 (0.37, 1.69) | 0.04 | 0.04 |
| Malawi | iLiNS−DYADM (44) | | | | | | | | | | | | |
| Malawi | iLiNS−DOSE (45) | | | | | | | | | | | | |
| Mali | PROMIS CS (46) | | | | | | | | | | | | |
| Zimbabwe | SHINE (HIV−) (47) | | | | | | | | | | | | |
| Zimbabwe | SHINE (HIV+) (48) | | | | | | | | | | | | |
| | | **860** | **502** | | **I² = 0.00, Tau² = 0.00** | | | **598** | **354** | | **I² = 0.00, Tau² = 0.00** | | |
| **Fixed** | | | | | **0.82 (0.70, 0.95)** | | | | | | **0.72 (0.61, 0.84)** | | |
| **Random** | | | | | **0.82 (0.70, 0.95)** | | | | | | **0.72 (0.61, 0.84)** | | |

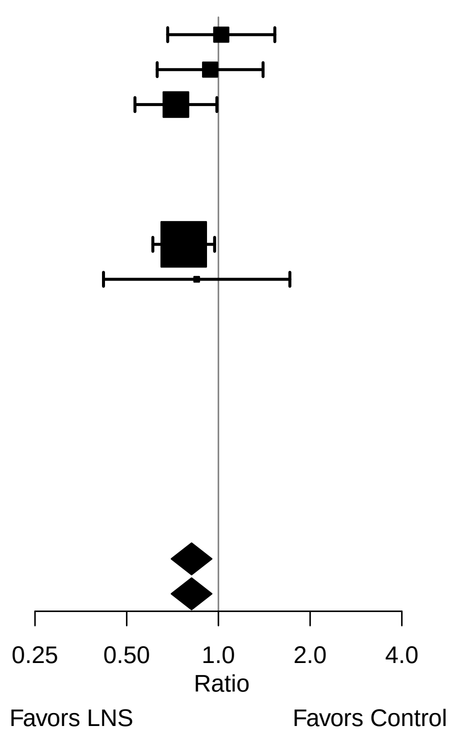

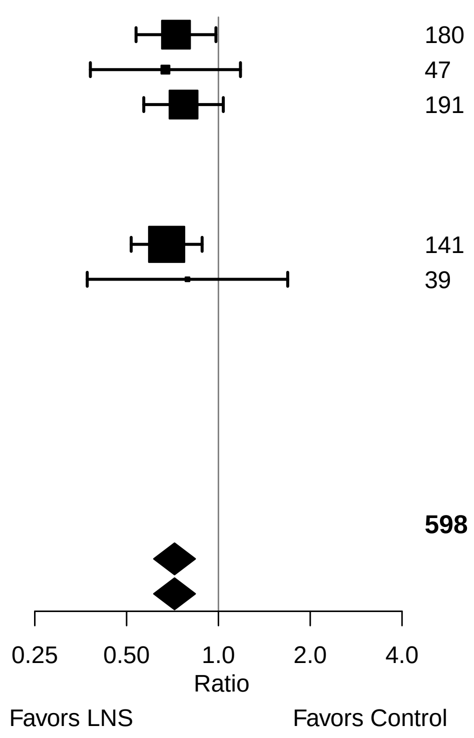

### Supplemental figure 6AA: Marginal vitamin A status (RBP < 1.05 µmol/L) prevalence difference

#### 6AA1: Stratified by Maternal BMI

**P−for−interaction = 0.234**
**Difference in PDs = 0.05 (−0.04, 0.14)**

| | | | | | **At least 20 $kg/m^2$** | | | | | | **Less than 20 $kg/m^2$** | | |
|---|---|---|---|---|---|---|---|---|---|---|---|---|---|
| | | **LNS** | **Control** | **Control** | **PD** | **Fixed** | **Random** | **LNS** | **Control** | **Control** | **PD** | **Fixed** | **Random** |
| Country | Trial | N | N | Prevalence | (95% CI) | W | W | N | N | Prevalence | (95% CI) | W | W |
| Bangladesh | JiVitA−4 (35) | | | | | | | | | | | | |
| Bangladesh | RDNS (36) | 228 | 102 | 34.3 | −0.12 (−0.21, −0.02) | 0.35 | 0.35 | 300 | 160 | 27.5 | 0.00 (−0.09, 0.10) | 0.44 | 0.38 |
| Bangladesh | WASH−B (37) | 93 | 97 | 30.9 | −0.07 (−0.21, 0.06) | 0.18 | 0.18 | 119 | 81 | 40.7 | −0.05 (−0.19, 0.08) | 0.22 | 0.24 |
| Burkina Faso | iLiNS−Zinc (38) | 209 | 55 | 49.2 | −0.12 (−0.26, 0.02) | 0.17 | 0.17 | 106 | 41 | 48.9 | −0.11 (−0.26, 0.04) | 0.17 | 0.20 |
| Burkina Faso | PROMIS CS (39) | | | | | | | | | | | | |
| Ghana | GHANA (40) | | | | | | | | | | | | |
| Ghana | iLiNS−DYADG (41) | | | | | | | | | | | | |
| Kenya | WASH−B (42) | 223 | 200 | 61.5 | −0.17 (−0.28, −0.07) | 0.30 | 0.30 | 64 | 51 | 56.9 | −0.18 (−0.34, −0.02) | 0.16 | 0.19 |
| Madagascar | MAHAY (43) | | | | | | | | | | | | |
| Malawi | iLiNS−DYADM (44) | | | | | | | | | | | | |
| Malawi | iLiNS−DOSE (45) | | | | | | | | | | | | |
| Mali | PROMIS CS (46) | | | | | | | | | | | | |
| Zimbabwe | SHINE (HIV−) (47) | | | | | | | | | | | | |
| Zimbabwe | SHINE (HIV+) (48) | | | | | | | | | | | | |
| | | **753** | **454** | | **$I^2$ = 0.00, $Tau^2$ = 0.00** | | | **589** | **333** | | **$I^2$ = 0.30, $Tau^2$ = 0.00** | | |
| **Fixed** | | | | | **−0.12 (−0.18, −0.07)** | | | | | | **−0.06 (−0.12, 0.00)** | | |
| **Random** | | | | | **−0.12 (−0.18, −0.07)** | | | | | | **−0.07 (−0.14, 0.01)** | | |

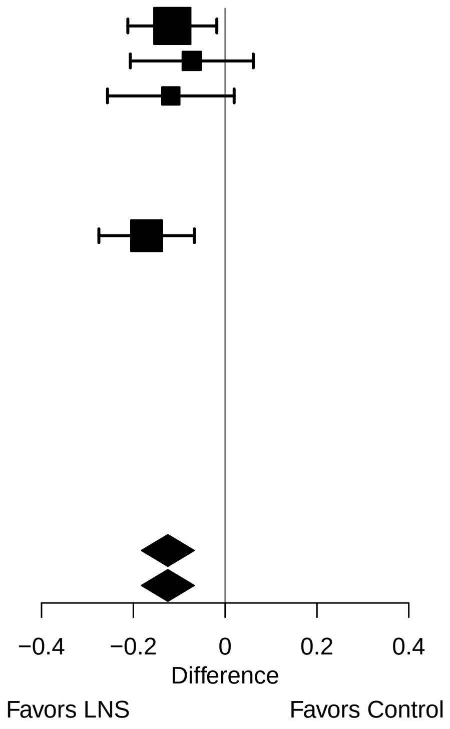

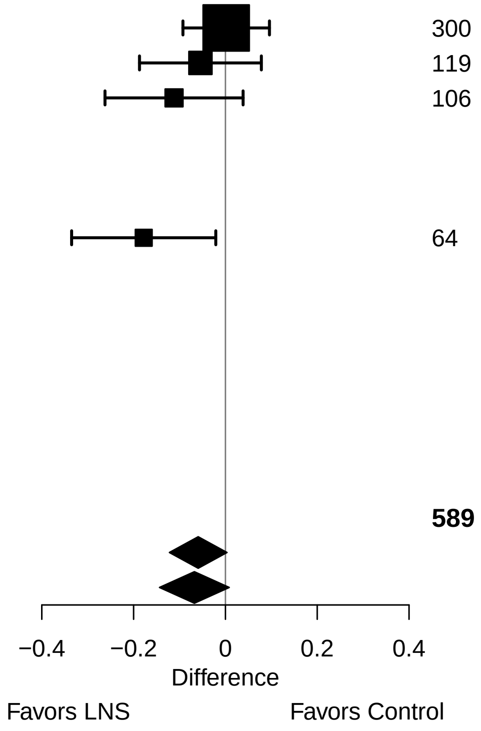

#### Supplemental figure 6AA: Marginal vitamin A status (RBP < 1.05 μmol/L) prevalence difference

#### 6AA2: Stratified by Maternal age

**P−for−interaction = 0.217**
**Difference in PDs = −0.05 (−0.14, 0.03)**

**At least 25 y**

| Country | Trial | LNS N | Control N | Control Prevalence | PD (95% CI) | Fixed W | Random W |
|---|---|---|---|---|---|---|---|
| Bangladesh | JiVitA−4 (35) | | | | | | |
| Bangladesh | RDNS (36) | 152 | 81 | 29.6 | −0.04 (−0.17, 0.09) | 0.20 | 0.20 |
| Bangladesh | WASH−B (37) | 79 | 80 | 38.8 | 0.02 (−0.14, 0.17) | 0.15 | 0.15 |
| Burkina Faso | iLiNS−Zinc (38) | 178 | 50 | 50.0 | −0.14 (−0.25, −0.04) | 0.34 | 0.34 |
| Burkina Faso | PROMIS CS (39) | | | | | | |
| Ghana | GHANA (40) | | | | | | |
| Ghana | iLiNS−DYADG (41) | | | | | | |
| Kenya | WASH−B (42) | 178 | 149 | 55.0 | −0.10 (−0.22, 0.01) | 0.26 | 0.26 |
| Madagascar | MAHAY (43) | 47 | 25 | 40.0 | −0.04 (−0.29, 0.22) | 0.05 | 0.05 |
| Malawi | iLiNS−DYADM (44) | | | | | | |
| Malawi | iLiNS−DOSE (45) | | | | | | |
| Mali | PROMIS CS (46) | | | | | | |
| Zimbabwe | SHINE (HIV−) (47) | | | | | | |
| Zimbabwe | SHINE (HIV+) (48) | | | | | | |
| | | **634** | **385** | | **I² = 0.00, Tau² = 0.00** | | |
| **Fixed** | | | | | **−0.08 (−0.14, −0.02)** | | |
| **Random** | | | | | **−0.08 (−0.14, −0.02)** | | |

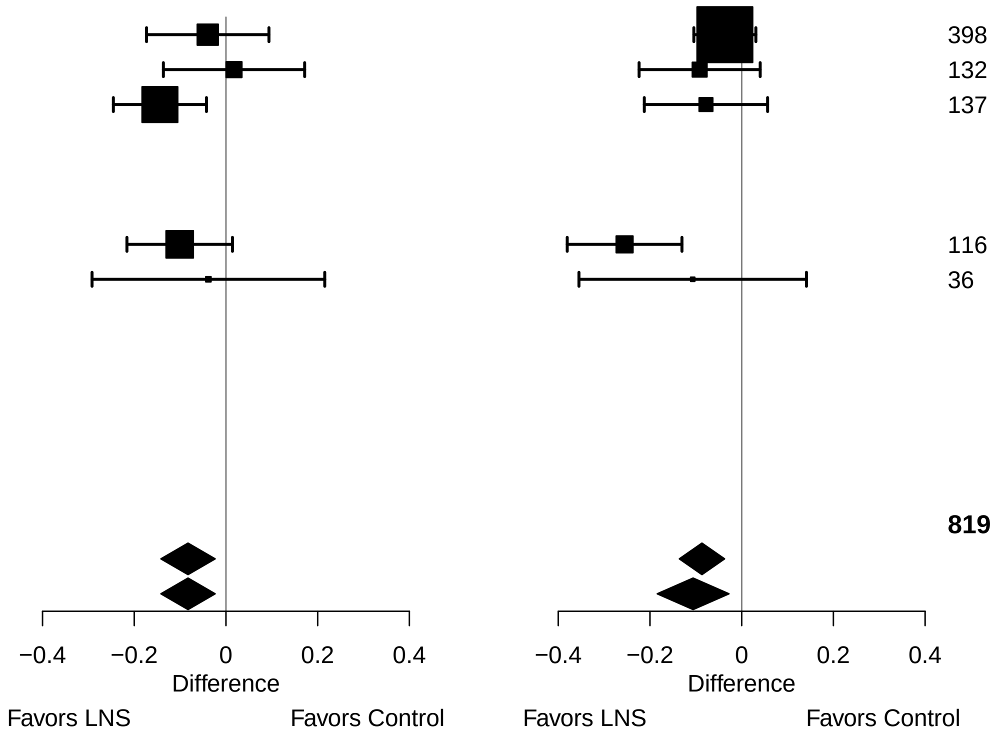

**Less than 25 y**

| Country | Trial | LNS N | Control N | Control Prevalence | PD (95% CI) | Fixed W | Random W |
|---|---|---|---|---|---|---|---|
| Bangladesh | JiVitA−4 (35) | | | | | | |
| Bangladesh | RDNS (36) | 398 | 191 | 28.8 | −0.04 (−0.10, 0.03) | 0.53 | 0.33 |
| Bangladesh | WASH−B (37) | 132 | 98 | 32.7 | −0.09 (−0.22, 0.04) | 0.14 | 0.19 |
| Burkina Faso | iLiNS−Zinc (38) | 137 | 46 | 48.1 | −0.08 (−0.21, 0.06) | 0.13 | 0.19 |
| Burkina Faso | PROMIS CS (39) | | | | | | |
| Ghana | GHANA (40) | | | | | | |
| Ghana | iLiNS−DYADG (41) | | | | | | |
| Kenya | WASH−B (42) | 116 | 109 | 66.1 | −0.26 (−0.38, −0.13) | 0.16 | 0.21 |
| Madagascar | MAHAY (43) | 36 | 26 | 38.5 | −0.11 (−0.35, 0.14) | 0.04 | 0.08 |
| Malawi | iLiNS−DYADM (44) | | | | | | |
| Malawi | iLiNS−DOSE (45) | | | | | | |
| Mali | PROMIS CS (46) | | | | | | |
| Zimbabwe | SHINE (HIV−) (47) | | | | | | |
| Zimbabwe | SHINE (HIV+) (48) | | | | | | |
| | | **819** | **470** | | **I² = 0.56, Tau² = 0.00** | | |
| **Fixed** | | | | | **−0.09 (−0.14, −0.04)** | | |
| **Random** | | | | | **−0.11 (−0.18, −0.03)** | | |

#### Supplemental figure 6AA: Marginal vitamin A status (RBP < 1.05 μmol/L) prevalence difference

##### 6AA3: Stratified by Maternal education

**P−for−interaction = 0.974**
**Difference in PDs = 0.00 (−0.10, 0.10)**

| | | Primary or greater | | | | | | Incomplete or no formal | | | | | |
|---|---|---|---|---|---|---|---|---|---|---|---|---|---|
| Country | Trial | LNS N | Control N | Control Prevalence | PD (95% CI) | Fixed W | Random W | LNS N | Control N | Control Prevalence | PD (95% CI) | Fixed W | Random W |
| Bangladesh | JiVitA−4 (35) | | | | | | | | | | | | |
| Bangladesh | RDNS (36) | 406 | 195 | 27.2 | −0.04 (−0.10, 0.03) | 0.61 | 0.55 | 144 | 77 | 33.8 | −0.04 (−0.20, 0.12) | 0.31 | 0.31 |
| Bangladesh | WASH−B (37) | 149 | 139 | 35.3 | −0.06 (−0.17, 0.04) | 0.23 | 0.27 | 63 | 39 | 35.9 | −0.03 (−0.22, 0.17) | 0.21 | 0.22 |
| Burkina Faso | iLiNS−Zinc (38) | | | | | | | | | | | | |
| Burkina Faso | PROMIS CS (39) | | | | | | | | | | | | |
| Ghana | GHANA (40) | | | | | | | | | | | | |
| Ghana | iLiNS−DYADG (41) | | | | | | | | | | | | |
| Kenya | WASH−B (42) | 143 | 117 | 53.0 | −0.15 (−0.28, −0.02) | 0.15 | 0.19 | 154 | 142 | 64.8 | −0.17 (−0.30, −0.05) | 0.49 | 0.46 |
| Madagascar | MAHAY (43) | | | | | | | | | | | | |
| Malawi | iLiNS−DYADM (44) | | | | | | | | | | | | |
| Malawi | iLiNS−DOSE (45) | | | | | | | | | | | | |
| Mali | PROMIS CS (46) | | | | | | | | | | | | |
| Zimbabwe | SHINE (HIV−) (47) | | | | | | | | | | | | |
| Zimbabwe | SHINE (HIV+) (48) | | | | | | | | | | | | |
| | | **698** | **451** | | **I² = 0.19, Tau² = 0.00** | | | **361** | **258** | | **I² = 0.17, Tau² = 0.00** | | |
| **Fixed** | | | | | **−0.06 (−0.11, −0.01)** | | | | | | **−0.10 (−0.19, −0.01)** | | |
| **Random** | | | | | **−0.06 (−0.13, 0.00)** | | | | | | **−0.10 (−0.20, 0.00)** | | |

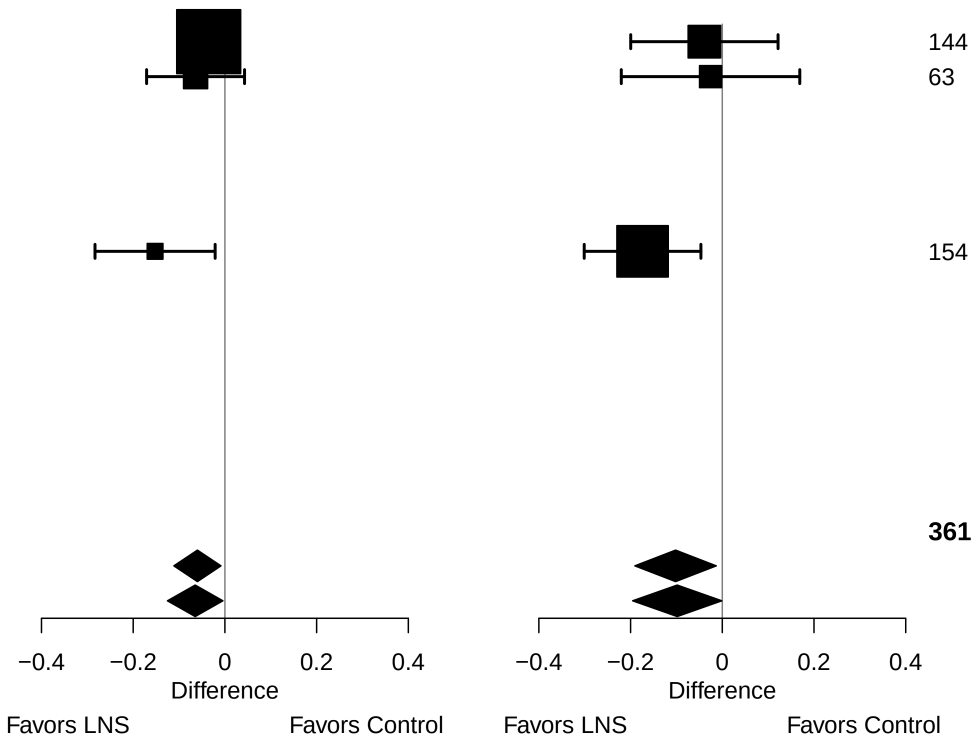

**Supplemental figure 6AA: Marginal vitamin A status (RBP < 1.05 µmol/L) prevalence difference**

#### 6AA4: Stratified by Child sex

**P−for−interaction = 0.393**
**Difference in PDs = −0.04 (−0.12, 0.05)**

| Country | Trial | Male LNS N | Male Control N | Male Control Prevalence | Male PD (95% CI) | Male Fixed W | Male Random W | Female LNS N | Female Control N | Female Control Prevalence | Female PD (95% CI) | Female Fixed W | Female Random W |
|---|---|---|---|---|---|---|---|---|---|---|---|---|---|
| Bangladesh | JiVitA−4 (35) | | | | | | | | | | | | |
| Bangladesh | RDNS (36) | 266 | 131 | 27.5 | −0.01 (−0.11, 0.08) | 0.38 | 0.31 | 281 | 141 | 30.5 | −0.07 (−0.15, 0.01) | 0.40 | 0.40 |
| Bangladesh | WASH−B (37) | 105 | 85 | 32.9 | 0.01 (−0.12, 0.15) | 0.19 | 0.21 | 107 | 93 | 37.6 | −0.11 (−0.23, 0.01) | 0.18 | 0.18 |
| Burkina Faso | iLiNS−Zinc (38) | 150 | 49 | 57.4 | −0.14 (−0.28, 0.00) | 0.18 | 0.20 | 165 | 47 | 40.4 | −0.07 (−0.18, 0.04) | 0.20 | 0.20 |
| Burkina Faso | PROMIS CS (39) | | | | | | | | | | | | |
| Ghana | GHANA (40) | | | | | | | | | | | | |
| Ghana | iLiNS−DYADG (41) | | | | | | | | | | | | |
| Kenya | WASH−B (42) | 146 | 125 | 61.6 | −0.17 (−0.30, −0.04) | 0.19 | 0.21 | 152 | 134 | 57.5 | −0.16 (−0.28, −0.04) | 0.17 | 0.17 |
| Madagascar | MAHAY (43) | 34 | 22 | 31.8 | −0.02 (−0.27, 0.23) | 0.05 | 0.08 | 49 | 29 | 44.8 | −0.10 (−0.34, 0.13) | 0.05 | 0.05 |
| Malawi | iLiNS−DYADM (44) | | | | | | | | | | | | |
| Malawi | iLiNS−DOSE (45) | | | | | | | | | | | | |
| Mali | PROMIS CS (46) | | | | | | | | | | | | |
| Zimbabwe | SHINE (HIV−) (47) | | | | | | | | | | | | |
| Zimbabwe | SHINE (HIV+) (48) | | | | | | | | | | | | |
| | | **701** | **412** | | **$I^2$ = 0.36, $Tau^2$ = 0.00** | | | **754** | **444** | | **$I^2$ = 0.00, $Tau^2$ = 0.00** | | |
| **Fixed** | | | | | **−0.06 (−0.12, 0.00)** | | | | | | **−0.09 (−0.14, −0.04)** | | |
| **Random** | | | | | **−0.07 (−0.14, 0.01)** | | | | | | **−0.09 (−0.14, −0.04)** | | |

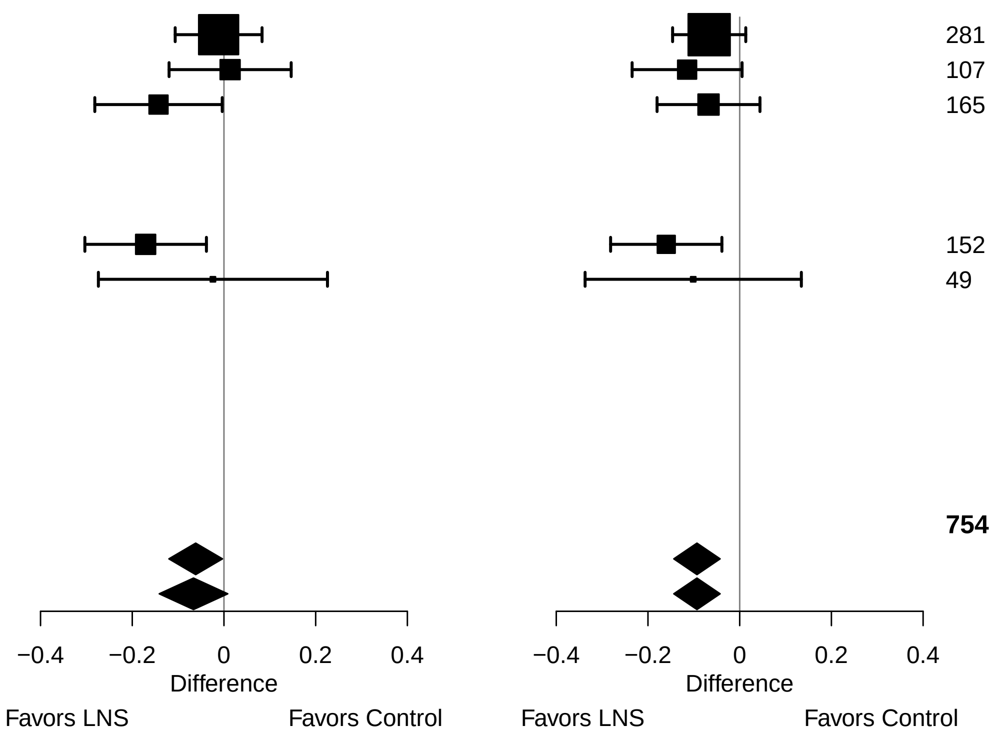

##### Supplemental figure 6AA: Marginal vitamin A status (RBP < 1.05 µmol/L) prevalence difference

#### 6AA5: Stratified by Child birth order

**P−for−interaction = 0.410**
**Difference in PDs = 0.04 (−0.05, 0.13)**

| Country | Trial | Later born LNS N | Later born Control N | Later born Control Prevalence | Later born PD (95% CI) | Later born Fixed W | Later born Random W | Firstborn LNS N | Firstborn Control N | Firstborn Control Prevalence | Firstborn PD (95% CI) | Firstborn Fixed W | Firstborn Random W |
|---|---|---|---|---|---|---|---|---|---|---|---|---|---|
| Bangladesh | JiVitA−4 (35) | | | | | | | | | | | | |
| Bangladesh | RDNS (36) | 340 | 171 | 30.4 | −0.05 (−0.15, 0.04) | 0.30 | 0.27 | 210 | 101 | 26.7 | −0.01 (−0.11, 0.09) | 0.50 | 0.50 |
| Bangladesh | WASH−B (37) | 124 | 119 | 38.7 | −0.03 (−0.14, 0.07) | 0.22 | 0.24 | 86 | 57 | 29.8 | −0.08 (−0.24, 0.09) | 0.19 | 0.19 |
| Burkina Faso | iLiNS−Zinc (38) | 249 | 71 | 53.3 | −0.16 (−0.27, −0.05) | 0.22 | 0.23 | 66 | 25 | 38.7 | 0.02 (−0.16, 0.20) | 0.16 | 0.16 |
| Burkina Faso | PROMIS CS (39) | | | | | | | | | | | | |
| Ghana | GHANA (40) | | | | | | | | | | | | |
| Ghana | iLiNS−DYADG (41) | | | | | | | | | | | | |
| Kenya | WASH−B (42) | 237 | 213 | 58.2 | −0.17 (−0.27, −0.07) | 0.26 | 0.26 | 60 | 46 | 65.2 | −0.15 (−0.33, 0.03) | 0.16 | 0.16 |
| Madagascar | MAHAY (43) | | | | | | | | | | | | |
| Malawi | iLiNS−DYADM (44) | | | | | | | | | | | | |
| Malawi | iLiNS−DOSE (45) | | | | | | | | | | | | |
| Mali | PROMIS CS (46) | | | | | | | | | | | | |
| Zimbabwe | SHINE (HIV−) (47) | | | | | | | | | | | | |
| Zimbabwe | SHINE (HIV+) (48) | | | | | | | | | | | | |
| | | **950** | **574** | | **I² = 0.48, Tau² = 0.00** | | | **422** | **229** | | **I² = 0.00, Tau² = 0.00** | | |
| **Fixed** | | | | | **−0.10 (−0.15, −0.05)** | | | | | | **−0.04 (−0.11, 0.03)** | | |
| **Random** | | | | | **−0.10 (−0.17, −0.03)** | | | | | | **−0.04 (−0.11, 0.03)** | | |

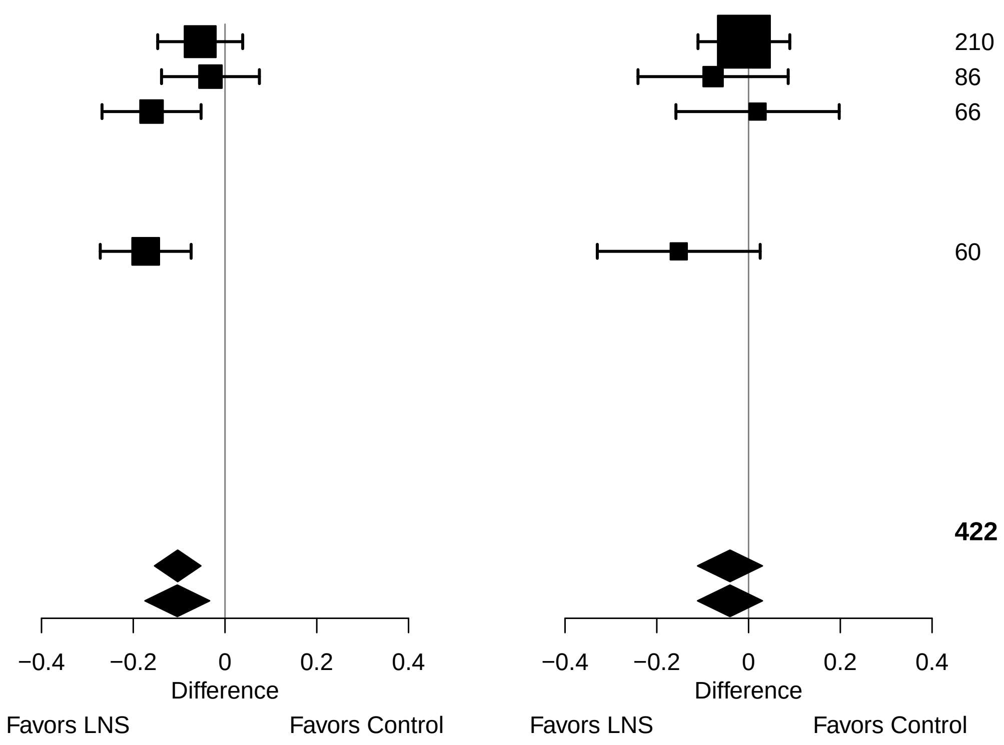

**Supplemental figure 6AA: Marginal vitamin A status (RBP < 1.05 µmol/L) prevalence difference**

#### 6AA6: Stratified by Child baseline acute malnutrition (insufficient comparisons)

**Supplemental figure 6AA: Marginal vitamin A status (RBP < 1.05 µmol/L) prevalence difference**

#### 6AA7: Stratified by Child baseline anemia (insufficient comparisons)

**Supplemental figure 6AA: Marginal vitamin A status (RBP < 1.05 µmol/L) prevalence difference**

#### 6AA8: Stratified by Child high-dose vitamin A supplementation (insufficient comparisons)

##### Supplemental figure 6AA: Marginal vitamin A status (RBP < 1.05 µmol/L) prevalence difference

#### 6AA9: Stratified by Child inflammation

**P−for−interaction = 0.080**
**Difference in PDs = −0.08 (−0.16, 0.01)**

| Country | Trial | Not inflamed: LNS N | Control N | Control Prevalence | PD (95% CI) | Fixed W | Random W | High AGP or CRP: LNS N | Control N | Control Prevalence | PD (95% CI) | Fixed W | Random W |
|---|---|---|---|---|---|---|---|---|---|---|---|---|---|
| Bangladesh | JiVitA−4 (35) | | | | | | | | | | | | |
| Bangladesh | RDNS (36) | 370 | 168 | 23.8 | 0.01 (−0.09, 0.10) | 0.35 | 0.32 | 180 | 104 | 37.5 | −0.10 (−0.20, −0.01) | 0.40 | 0.40 |
| Bangladesh | WASH−B (37) | 165 | 136 | 31.6 | −0.02 (−0.14, 0.10) | 0.22 | 0.22 | 47 | 42 | 47.6 | −0.16 (−0.37, 0.05) | 0.08 | 0.08 |
| Burkina Faso | iLiNS−Zinc (38) | 124 | 31 | 50.0 | −0.15 (−0.29, 0.00) | 0.15 | 0.16 | 191 | 65 | 48.6 | −0.11 (−0.22, 0.01) | 0.27 | 0.27 |
| Burkina Faso | PROMIS CS (39) | | | | | | | | | | | | |
| Ghana | GHANA (40) | | | | | | | | | | | | |
| Ghana | iLiNS−DYADG (41) | | | | | | | | | | | | |
| Kenya | WASH−B (42) | 157 | 135 | 56.3 | −0.13 (−0.25, −0.01) | 0.23 | 0.24 | 141 | 124 | 62.9 | −0.20 (−0.34, −0.07) | 0.21 | 0.21 |
| Madagascar | MAHAY (43) | 44 | 32 | 37.5 | −0.06 (−0.30, 0.19) | 0.05 | 0.06 | 39 | 19 | 42.1 | −0.09 (−0.38, 0.21) | 0.04 | 0.04 |
| Malawi | iLiNS−DYADM (44) | | | | | | | | | | | | |
| Malawi | iLiNS−DOSE (45) | | | | | | | | | | | | |
| Mali | PROMIS CS (46) | | | | | | | | | | | | |
| Zimbabwe | SHINE (HIV−) (47) | | | | | | | | | | | | |
| Zimbabwe | SHINE (HIV+) (48) | | | | | | | | | | | | |
| | | **860** | **502** | | **I² = 0.16, Tau² = 0.00** | | | **598** | **354** | | **I² = 0.00, Tau² = 0.00** | | |
| **Fixed** | | | | | **−0.06 (−0.11, 0.00)** | | | | | | **−0.13 (−0.19, −0.07)** | | |
| **Random** | | | | | **−0.06 (−0.12, 0.00)** | | | | | | **−0.13 (−0.19, −0.07)** | | |

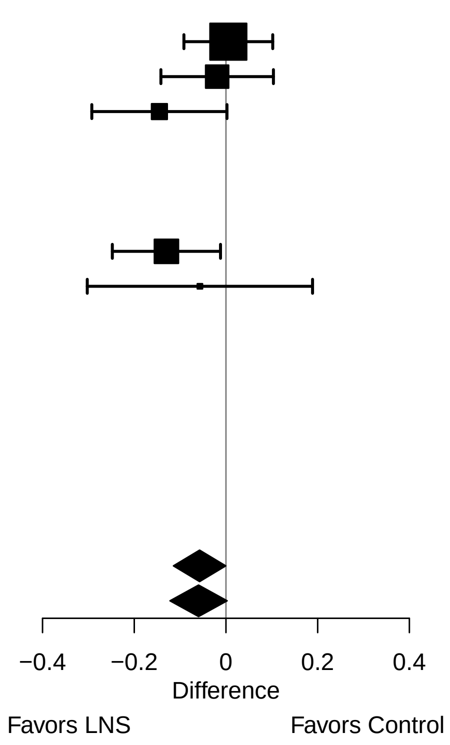

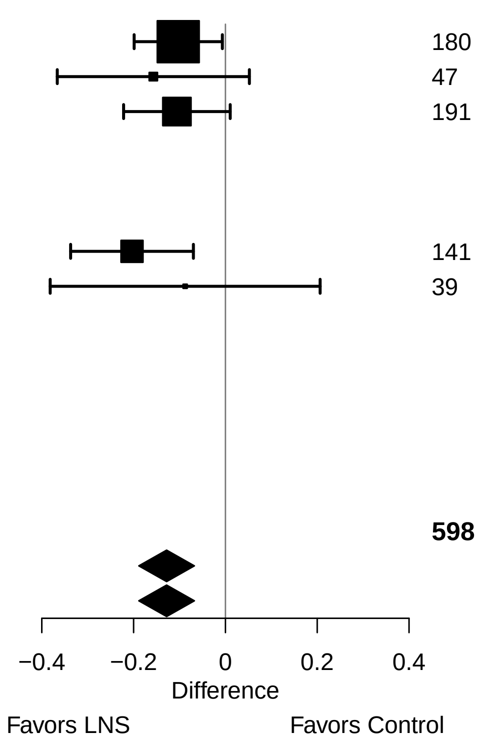

### Supplemental figure 7: Forest plots for effects of SQ-LNS on biochemical outcomes stratified by individual-level household effect modifiers

These figures are forest plots showing the individual-level effect modification of intervention effects. Each figure has the estimates of intervention effect stratified within study by individual-level effect modifier category. For continuous outcomes the intervention effect is measured by the difference in mean of the LNS group minus control. For log transformed continuous outcomes, the intervention effect is measured by the ratio of geometric means, the effect estimate is the geometric mean in the LNS group divided by the geometric mean in the control group. For dichotomous outcomes analyzed via prevalence ratios, the effect estimate is the prevalence in the LNS group divided by the prevalence in the control group. For dichotomous outcomes analyzed via prevalence differences, the effect estimate is the prevalence in the LNS group minus the prevalence in the control group. The labels on the far left correspond to trial level information. In the middle left and on the right the values indicate the study level effect estimate, confidence interval, and weighting for deriving the pooled estimates is shown by subgroup.
Not all trials were included in all individual-level effect modification analyses, either because they did not measure the biomarker outcome or the effect modifier of interest (e.g., baseline anemia or acute malnutrition, receipt of high-dose vitamin A supplement), or because the prevalence of the binary outcome or proportion of children within one of the effect modifier subgroups was too low to allow us to generate effect estimates.
RBP, retinol binding protein.

### Supplemental figure 7A: Mean difference in hemoglobin concentration

#### 7A1: Stratified by Household socio-economic status

**P−for−interaction = 0.528**
**Difference in MDs = −0.26 (−1.07, 0.55)**

**At least median**

| Country | Trial | LNS N | Control N | Control Mean | MD (95% CI) | Fixed W | Random W |
|---|---|---|---|---|---|---|---|
| Bangladesh | JiVitA−4 (35) | 244 | 79 | 117.1 | 3.51 (1.31, 5.71) | 0.08 | 0.08 |
| Bangladesh | RDNS (36) | 272 | 136 | 112.2 | 5.44 (2.82, 8.05) | 0.06 | 0.08 |
| Bangladesh | WASH−B (37) | 114 | 93 | 117.5 | 4.52 (1.84, 7.20) | 0.05 | 0.07 |
| Burkina Faso | iLiNS−Zinc (38) | 1232 | 299 | 88.9 | 8.35 (5.76, 10.95) | 0.06 | 0.08 |
| Burkina Faso | PROMIS CS (39) | 289 | 293 | 103.1 | 1.10 (−1.85, 4.06) | 0.04 | 0.07 |
| Ghana | GHANA (40) | 57 | 44 | 109.4 | 6.35 (1.27, 11.42) | 0.02 | 0.04 |
| Ghana | iLiNS−DYADG (41) | 149 | 344 | 113.3 | 0.24 (−1.66, 2.15) | 0.11 | 0.09 |
| Kenya | WASH−B (42) | 190 | 175 | 109.5 | 4.56 (2.31, 6.81) | 0.08 | 0.08 |
| Madagascar | MAHAY (43) | 278 | 311 | 104.6 | 4.08 (0.92, 7.24) | 0.04 | 0.06 |
| Malawi | iLiNS−DYADM (44) | 117 | 225 | 109.0 | 2.44 (−0.97, 5.86) | 0.03 | 0.06 |
| Malawi | iLiNS−DOSE (45) | 86 | 32 | 99.9 | 7.41 (1.69, 13.13) | 0.01 | 0.03 |
| Mali | PROMIS CS (46) | 505 | 463 | 96.8 | 5.61 (3.44, 7.78) | 0.08 | 0.08 |
| Zimbabwe | SHINE (HIV−) (47) | 821 | 722 | 114.2 | 2.19 (1.05, 3.32) | 0.30 | 0.11 |
| Zimbabwe | SHINE (HIV+) (48) | 155 | 138 | 115.0 | 3.00 (−0.27, 6.28) | 0.04 | 0.06 |
| | | **4509** | **3354** | | **I² = 0.70, Tau² = 3.18** | | |
| **Fixed** | | | | | **3.41 (2.78, 4.03)** | | |
| **Random** | | | | | **3.93 (2.73, 5.13)** | | |

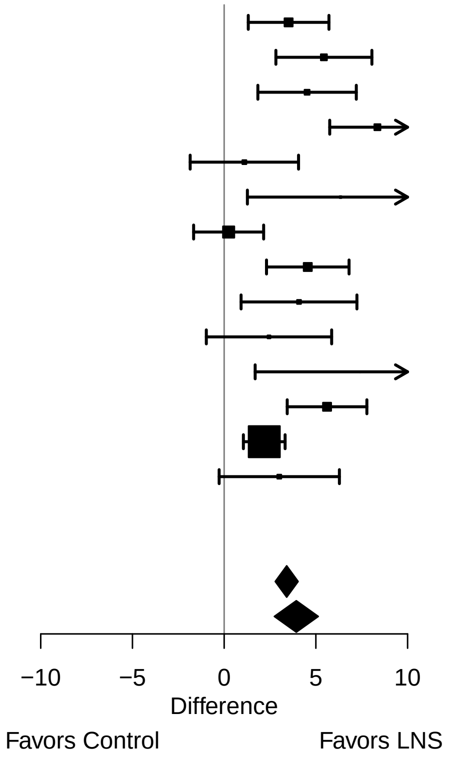

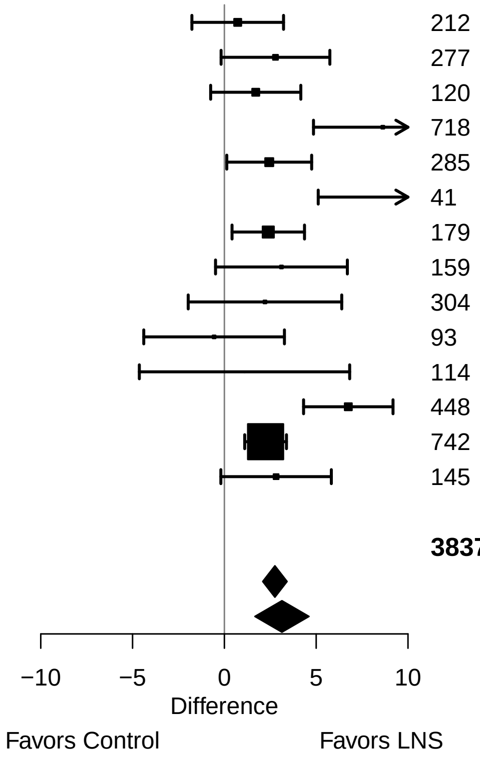

**Less than median**

| Country | Trial | LNS N | Control N | Control Mean | MD (95% CI) | Fixed W | Random W |
|---|---|---|---|---|---|---|---|
| Bangladesh | JiVitA−4 (35) | 212 | 67 | 119.0 | 0.73 (−1.77, 3.22) | 0.07 | 0.08 |
| Bangladesh | RDNS (36) | 277 | 136 | 112.6 | 2.78 (−0.18, 5.74) | 0.05 | 0.07 |
| Bangladesh | WASH−B (37) | 120 | 93 | 118.7 | 1.71 (−0.75, 4.16) | 0.07 | 0.08 |
| Burkina Faso | iLiNS−Zinc (38) | 718 | 364 | 88.3 | 8.63 (4.85, 12.40) | 0.03 | 0.06 |
| Burkina Faso | PROMIS CS (39) | 285 | 288 | 102.1 | 2.44 (0.13, 4.75) | 0.08 | 0.08 |
| Ghana | GHANA (40) | 41 | 46 | 101.6 | 11.25 (5.11, 17.40) | 0.01 | 0.04 |
| Ghana | iLiNS−DYADG (41) | 179 | 315 | 110.5 | 2.39 (0.42, 4.36) | 0.11 | 0.09 |
| Kenya | WASH−B (42) | 159 | 125 | 110.3 | 3.11 (−0.48, 6.70) | 0.03 | 0.07 |
| Madagascar | MAHAY (43) | 304 | 262 | 103.2 | 2.21 (−1.97, 6.39) | 0.03 | 0.06 |
| Malawi | iLiNS−DYADM (44) | 93 | 206 | 106.8 | −0.56 (−4.39, 3.27) | 0.03 | 0.06 |
| Malawi | iLiNS−DOSE (45) | 114 | 39 | 101.8 | 1.10 (−4.63, 6.82) | 0.01 | 0.04 |
| Mali | PROMIS CS (46) | 448 | 507 | 94.8 | 6.75 (4.31, 9.18) | 0.07 | 0.08 |
| Zimbabwe | SHINE (HIV−) (47) | 742 | 760 | 114.4 | 2.24 (1.11, 3.38) | 0.34 | 0.10 |
| Zimbabwe | SHINE (HIV+) (48) | 145 | 140 | 114.4 | 2.82 (−0.19, 5.83) | 0.05 | 0.07 |
| | | **3837** | **3348** | | **I² = 0.62, Tau² = 5.27** | | |
| **Fixed** | | | | | **2.75 (2.09, 3.42)** | | |
| **Random** | | | | | **3.14 (1.66, 4.61)** | | |

#### Supplemental figure 7A: Mean difference in hemoglobin concentration

##### 7A2: Stratified by Household food insecurity

**P−for−interaction = 0.470**
**Difference in MDs = 0.40 (−0.68, 1.48)**

| | | Mild to secure | | | | | | Moderate to severe | | | | | |
|---|---|---|---|---|---|---|---|---|---|---|---|---|---|
| Country | Trial | LNS N | Control N | Control Mean | MD (95% CI) | Fixed W | Random W | LNS N | Control N | Control Mean | MD (95% CI) | Fixed W | Random W |
| Bangladesh | JiVitA−4 (35) | 339 | 113 | 118.0 | 2.27 (−0.02, 4.55) | 0.07 | 0.10 | 117 | 33 | 117.8 | 2.15 (−0.82, 5.13) | 0.10 | 0.11 |
| Bangladesh | RDNS (36) | 342 | 166 | 113.7 | 2.94 (−0.01, 5.89) | 0.04 | 0.09 | 207 | 106 | 110.4 | 5.91 (2.94, 8.88) | 0.10 | 0.11 |
| Bangladesh | WASH−B (37) | 190 | 145 | 118.6 | 2.75 (0.55, 4.95) | 0.07 | 0.11 | 44 | 41 | 116.4 | 4.22 (0.41, 8.03) | 0.06 | 0.08 |
| Burkina Faso | iLiNS−Zinc (38) | 1016 | 320 | 87.4 | 8.87 (6.02, 11.73) | 0.04 | 0.09 | 934 | 343 | 89.6 | 8.22 (4.52, 11.92) | 0.07 | 0.09 |
| Burkina Faso | PROMIS CS (39) | | | | | | | | | | | | |
| Ghana | GHANA (40) | | | | | | | | | | | | |
| Ghana | iLiNS−DYADG (41) | 230 | 457 | 111.6 | 1.02 (−0.56, 2.61) | 0.14 | 0.12 | 97 | 201 | 112.6 | 1.91 (−0.81, 4.63) | 0.12 | 0.11 |
| Kenya | WASH−B (42) | 304 | 273 | 109.7 | 4.27 (2.23, 6.31) | 0.08 | 0.11 | 45 | 27 | 111.1 | 1.18 (−4.08, 6.44) | 0.03 | 0.06 |
| Madagascar | MAHAY (43) | 359 | 340 | 101.8 | 5.00 (1.79, 8.20) | 0.03 | 0.08 | 137 | 140 | 105.6 | −0.76 (−5.57, 4.05) | 0.04 | 0.06 |
| Malawi | iLiNS−DYADM (44) | 66 | 113 | 105.3 | 4.85 (−0.06, 9.75) | 0.01 | 0.05 | 143 | 314 | 109.0 | −0.21 (−3.23, 2.82) | 0.10 | 0.10 |
| Malawi | iLiNS−DOSE (45) | 50 | 21 | 101.6 | 2.53 (−5.96, 11.02) | 0.00 | 0.02 | 144 | 46 | 101.0 | 3.24 (−1.69, 8.18) | 0.04 | 0.06 |
| Mali | PROMIS CS (46) | | | | | | | | | | | | |
| Zimbabwe | SHINE (HIV−) (47) | 1253 | 1162 | 114.5 | 1.76 (0.88, 2.65) | 0.45 | 0.14 | 264 | 293 | 113.9 | 3.53 (1.78, 5.28) | 0.29 | 0.15 |
| Zimbabwe | SHINE (HIV+) (48) | 217 | 204 | 114.6 | 2.60 (−0.03, 5.24) | 0.05 | 0.09 | 75 | 66 | 114.9 | 3.90 (−0.54, 8.34) | 0.05 | 0.07 |
| | | **4366** | **3314** | | **I² = 0.68, Tau² = 2.85** | | | **2207** | **1610** | | **I² = 0.50, Tau² = 3.19** | | |
| **Fixed** | | | | | **2.53 (1.94, 3.12)** | | | | | | **3.18 (2.23, 4.13)** | | |
| **Random** | | | | | **3.32 (2.03, 4.62)** | | | | | | **3.13 (1.63, 4.63)** | | |

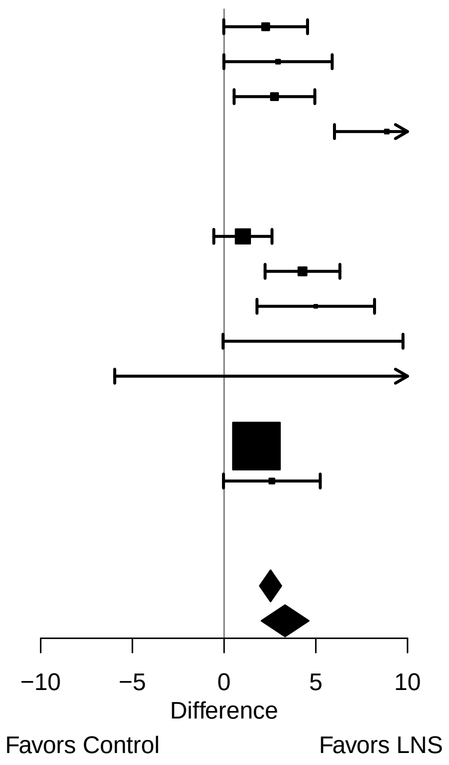

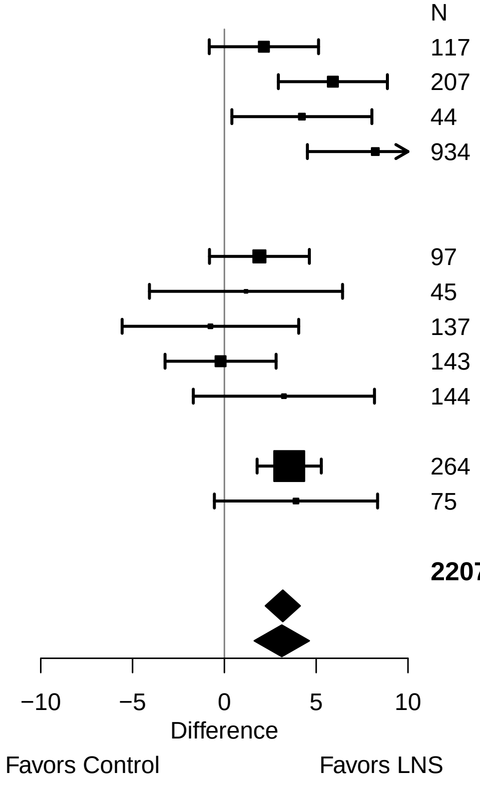

##### Supplemental figure 7A: Mean difference in hemoglobin concentration

#### 7A3: Stratified by Household source water quality

**P−for−interaction = 0.746**
**Difference in MDs = −0.21 (−1.49, 1.07)**

| | | **Improved** | | | | | | **Unimproved** | | | | | |
|---|---|---|---|---|---|---|---|---|---|---|---|---|---|
| Country | Trial | **LNS** N | **Control** N | **Control** Mean | **MD** (95% CI) | **Fixed** W | **Random** W | **LNS** N | **Control** N | **Control** Mean | **MD** (95% CI) | **Fixed** W | **Random** W |
| Bangladesh | JiVitA−4 (35) | | | | | | | | | | | | |
| Bangladesh | RDNS (36) | | | | | | | | | | | | |
| Bangladesh | WASH−B (37) | 84 | 46 | 118.5 | 3.76 (0.24, 7.28) | 0.07 | 0.11 | 21 | 14 | 118.8 | 1.26 (−4.36, 6.89) | 0.03 | 0.06 |
| Burkina Faso | iLiNS−Zinc (38) | 555 | 141 | 86.9 | 11.38 (6.98, 15.77) | 0.04 | 0.09 | 1394 | 519 | 89.0 | 7.53 (4.38, 10.68) | 0.11 | 0.13 |
| Burkina Faso | PROMIS CS (39) | 373 | 363 | 102.8 | 1.35 (−0.79, 3.48) | 0.18 | 0.13 | 201 | 217 | 102.3 | 2.49 (−0.34, 5.31) | 0.13 | 0.14 |
| Ghana | GHANA (40) | | | | | | | | | | | | |
| Ghana | iLiNS−DYADG (41) | | | | | | | | | | | | |
| Kenya | WASH−B (42) | 128 | 121 | 108.8 | 3.46 (0.26, 6.67) | 0.08 | 0.11 | 68 | 41 | 113.5 | 1.45 (−2.98, 5.89) | 0.05 | 0.08 |
| Madagascar | MAHAY (43) | 137 | 164 | 105.0 | 0.74 (−4.25, 5.74) | 0.03 | 0.08 | 433 | 408 | 103.5 | 3.80 (0.70, 6.90) | 0.11 | 0.13 |
| Malawi | iLiNS−DYADM (44) | 191 | 397 | 108.5 | 0.34 (−2.33, 3.01) | 0.12 | 0.12 | 19 | 33 | 101.5 | 10.72 (1.84, 19.59) | 0.01 | 0.03 |
| Malawi | iLiNS−DOSE (45) | | | | | | | | | | | | |
| Mali | PROMIS CS (46) | 548 | 567 | 95.3 | 6.90 (4.34, 9.45) | 0.13 | 0.12 | 364 | 388 | 96.3 | 5.43 (2.66, 8.20) | 0.14 | 0.14 |
| Zimbabwe | SHINE (HIV−) (47) | 469 | 436 | 114.6 | 2.18 (0.55, 3.81) | 0.31 | 0.14 | 259 | 281 | 114.4 | 2.25 (0.53, 3.97) | 0.35 | 0.19 |
| Zimbabwe | SHINE (HIV+) (48) | 77 | 77 | 115.0 | 1.05 (−3.12, 5.22) | 0.05 | 0.10 | 49 | 46 | 114.7 | 2.36 (−1.74, 6.47) | 0.06 | 0.09 |
| | | **2562** | **2312** | | **I² = 0.75, Tau² = 8.97** | | | **2808** | **1947** | | **I² = 0.46, Tau² = 2.60** | | |
| **Fixed** | | | | | **2.91 (2.00, 3.82)** | | | | | | **3.49 (2.46, 4.51)** | | |
| **Random** | | | | | **3.37 (1.13, 5.61)** | | | | | | **3.73 (2.14, 5.32)** | | |

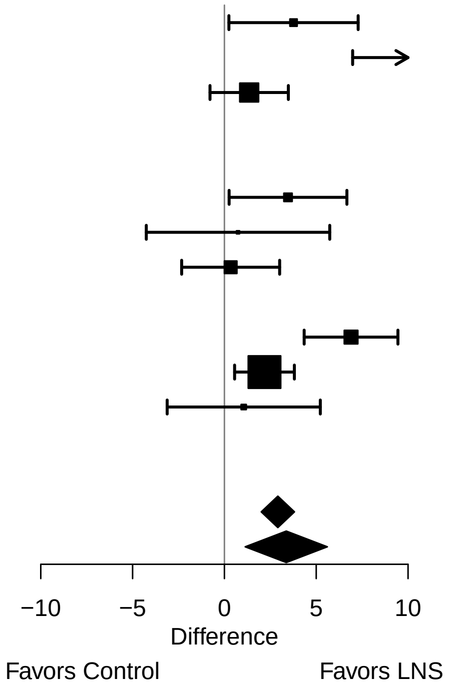

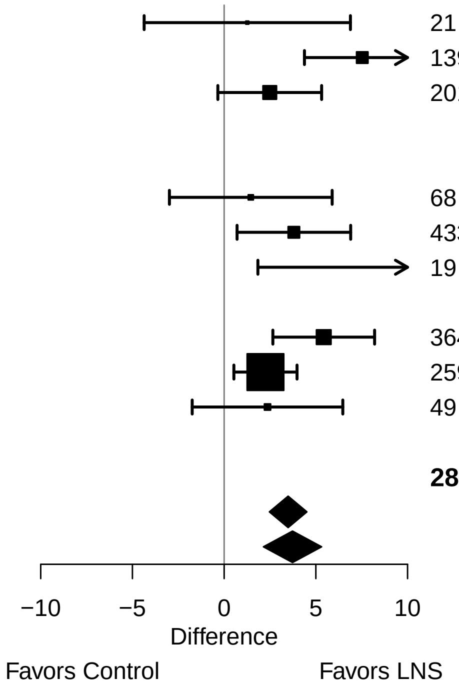

##### Supplemental figure 7A: Mean difference in hemoglobin concentration

#### 7A4: Stratified by Household sanitation

**P−for−interaction = 0.607**
**Difference in MDs = 0.32 (−0.89, 1.52)**

| Country | Trial | Improved LNS N | Improved Control N | Improved Control Mean | Improved MD (95% CI) | Improved Fixed W | Improved Random W | Unimproved LNS N | Unimproved Control N | Unimproved Control Mean | Unimproved MD (95% CI) | Unimproved Fixed W | Unimproved Random W |
|---|---|---|---|---|---|---|---|---|---|---|---|---|---|
| Bangladesh | JiVitA−4 (35) | 363 | 114 | 117.6 | 3.14 (1.07, 5.21) | 0.14 | 0.13 | 93 | 32 | 119.5 | −1.19 (−4.97, 2.59) | 0.05 | 0.09 |
| Bangladesh | RDNS (36) | 376 | 193 | 112.9 | 3.74 (1.16, 6.33) | 0.09 | 0.12 | 173 | 79 | 111.2 | 5.01 (2.00, 8.01) | 0.08 | 0.10 |
| Bangladesh | WASH−B (37) | | | | | | | | | | | | |
| Burkina Faso | iLiNS−Zinc (38) | 43 | 16 | 88.3 | 9.64 (4.27, 15.01) | 0.02 | 0.06 | 1906 | 644 | 88.6 | 8.46 (5.37, 11.55) | 0.08 | 0.10 |
| Burkina Faso | PROMIS CS (39) | 244 | 229 | 103.0 | 1.15 (−1.14, 3.44) | 0.11 | 0.13 | 329 | 350 | 102.3 | 2.15 (−0.10, 4.40) | 0.15 | 0.11 |
| Ghana | GHANA (40) | | | | | | | | | | | | |
| Ghana | iLiNS−DYADG (41) | 317 | 642 | 112.0 | 1.22 (−0.18, 2.62) | 0.30 | 0.15 | 10 | 17 | 109.8 | 2.74 (−4.84, 10.31) | 0.01 | 0.05 |
| Kenya | WASH−B (42) | 24 | 25 | 113.2 | 5.17 (−1.20, 11.55) | 0.01 | 0.04 | 172 | 137 | 109.4 | 3.07 (0.10, 6.03) | 0.08 | 0.10 |
| Madagascar | MAHAY (43) | | | | | | | | | | | | |
| Malawi | iLiNS−DYADM (44) | 20 | 39 | 107.3 | 9.44 (0.08, 18.80) | 0.01 | 0.02 | 190 | 391 | 108.1 | 0.32 (−2.34, 2.97) | 0.11 | 0.11 |
| Malawi | iLiNS−DOSE (45) | | | | | | | | | | | | |
| Mali | PROMIS CS (46) | 703 | 696 | 95.7 | 5.44 (3.17, 7.70) | 0.11 | 0.13 | 219 | 249 | 96.1 | 8.80 (5.89, 11.71) | 0.09 | 0.11 |
| Zimbabwe | SHINE (HIV−) (47) | 262 | 218 | 114.3 | 1.68 (−0.08, 3.44) | 0.19 | 0.14 | 465 | 498 | 114.5 | 2.57 (0.97, 4.18) | 0.29 | 0.12 |
| Zimbabwe | SHINE (HIV+) (48) | 46 | 30 | 114.1 | 3.56 (−1.06, 8.18) | 0.03 | 0.07 | 80 | 92 | 115.3 | 0.40 (−3.07, 3.87) | 0.06 | 0.10 |
| | | **2398** | **2202** | | **I² = 0.61, Tau² = 3.47** | | | **3637** | **2489** | | **I² = 0.78, Tau² = 8.45** | | |
| **Fixed** | | | | | **2.60 (1.84, 3.37)** | | | | | | **3.19 (2.33, 4.05)** | | |
| **Random** | | | | | **3.38 (1.84, 4.92)** | | | | | | **3.29 (1.22, 5.37)** | | |

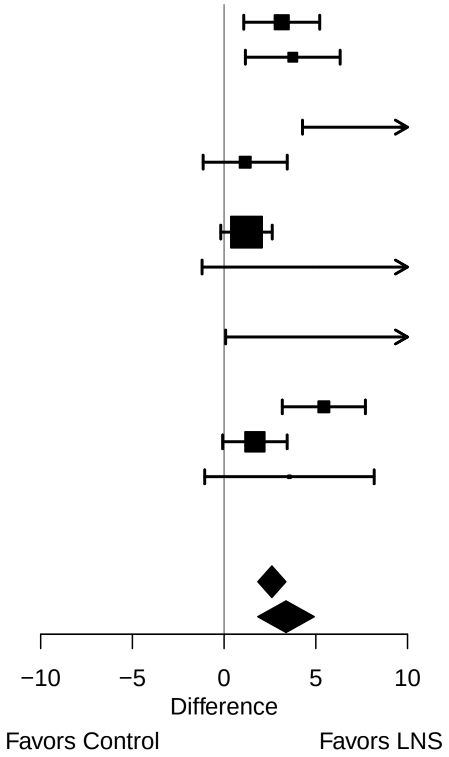

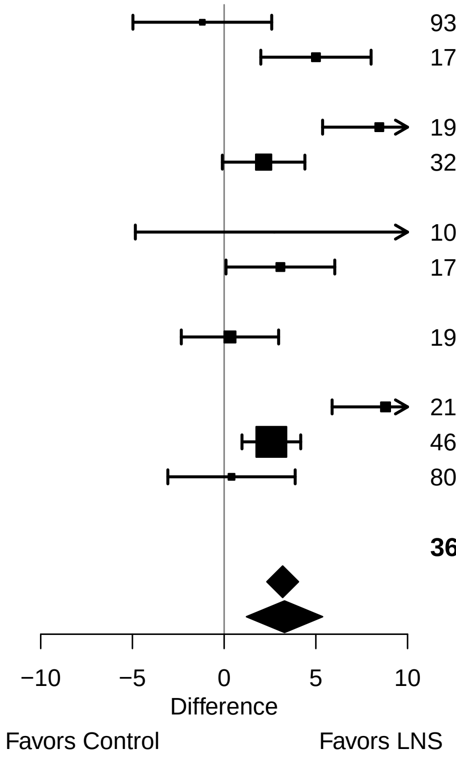

#### Supplemental figure 7A: Mean difference in hemoglobin concentration

##### 7A5: Stratified by Season at the time of assessment

**P−for−interaction = 0.190**
**Difference in MDs = −0.69 (−1.72, 0.34)**

| | | Dry | | | | | | Rainy | | | | | |
|---|---|---|---|---|---|---|---|---|---|---|---|---|---|
| | | **LNS** | **Control** | **Control** | **MD** | **Fixed** | **Random** | **LNS** | **Control** | **Control** | **MD** | **Fixed** | **Random** |
| Country | Trial | N | N | Mean | (95% CI) | W | W | N | N | Mean | (95% CI) | W | W |
| Bangladesh | JiVitA−4 (35) | 264 | 72 | 118.6 | 1.35 (−1.44, 4.14) | 0.04 | 0.08 | 193 | 74 | 117.4 | 3.14 (0.91, 5.37) | 0.14 | 0.10 |
| Bangladesh | RDNS (36) | 341 | 166 | 110.5 | 5.65 (3.05, 8.25) | 0.05 | 0.08 | 208 | 106 | 115.4 | 1.70 (−2.06, 5.46) | 0.05 | 0.07 |
| Bangladesh | WASH−B (37) | 108 | 96 | 118.5 | 2.49 (0.08, 4.90) | 0.06 | 0.09 | 126 | 90 | 117.7 | 3.70 (0.94, 6.46) | 0.09 | 0.09 |
| Burkina Faso | iLiNS−Zinc (38) | 1398 | 475 | 88.6 | 8.74 (5.82, 11.65) | 0.04 | 0.08 | 559 | 189 | 88.5 | 7.76 (3.68, 11.85) | 0.04 | 0.07 |
| Burkina Faso | PROMIS CS (39) | 283 | 245 | 102.1 | 2.36 (−0.38, 5.09) | 0.04 | 0.08 | 291 | 336 | 103.0 | 1.32 (−1.02, 3.66) | 0.13 | 0.10 |
| Ghana | GHANA (40) | 57 | 55 | 105.7 | 9.59 (4.43, 14.75) | 0.01 | 0.04 | 41 | 41 | 105.7 | 7.79 (1.53, 14.05) | 0.02 | 0.04 |
| Ghana | iLiNS−DYADG (41) | 198 | 386 | 111.8 | 1.55 (−0.21, 3.31) | 0.11 | 0.10 | 130 | 274 | 112.2 | 0.76 (−1.46, 2.97) | 0.14 | 0.11 |
| Kenya | WASH−B (42) | 100 | 88 | 111.7 | 3.74 (0.76, 6.71) | 0.04 | 0.08 | 250 | 212 | 109.0 | 4.05 (1.79, 6.32) | 0.13 | 0.10 |
| Madagascar | MAHAY (43) | 517 | 537 | 103.3 | 2.04 (−0.87, 4.95) | 0.04 | 0.08 | 83 | 51 | 110.9 | 4.51 (−0.26, 9.29) | 0.03 | 0.06 |
| Malawi | iLiNS−DYADM (44) | 147 | 305 | 109.2 | 1.95 (−1.01, 4.91) | 0.04 | 0.08 | 63 | 125 | 105.1 | −0.61 (−5.53, 4.31) | 0.03 | 0.05 |
| Malawi | iLiNS−DOSE (45) | 123 | 32 | 101.8 | −0.23 (−6.20, 5.74) | 0.01 | 0.04 | 120 | 50 | 99.8 | 6.93 (2.14, 11.72) | 0.03 | 0.06 |
| Mali | PROMIS CS (46) | | | | | | | | | | | | |
| Zimbabwe | SHINE (HIV−) (47) | 1558 | 1477 | 114.2 | 2.35 (1.53, 3.18) | 0.48 | 0.11 | 124 | 117 | 115.5 | −0.63 (−2.74, 1.47) | 0.16 | 0.11 |
| Zimbabwe | SHINE (HIV+) (48) | 289 | 265 | 115.0 | 2.69 (0.14, 5.25) | 0.05 | 0.08 | 17 | 20 | 111.2 | 3.26 (−2.80, 9.32) | 0.02 | 0.04 |
| | | **5383** | **4199** | | **I² = 0.64, Tau² = 4.40** | | | **2205** | **1685** | | **I² = 0.59, Tau² = 3.95** | | |
| **Fixed** | | | | | **2.75 (2.17, 3.32)** | | | | | | **2.41 (1.58, 3.24)** | | |
| **Random** | | | | | **3.27 (1.88, 4.66)** | | | | | | **2.92 (1.46, 4.37)** | | |

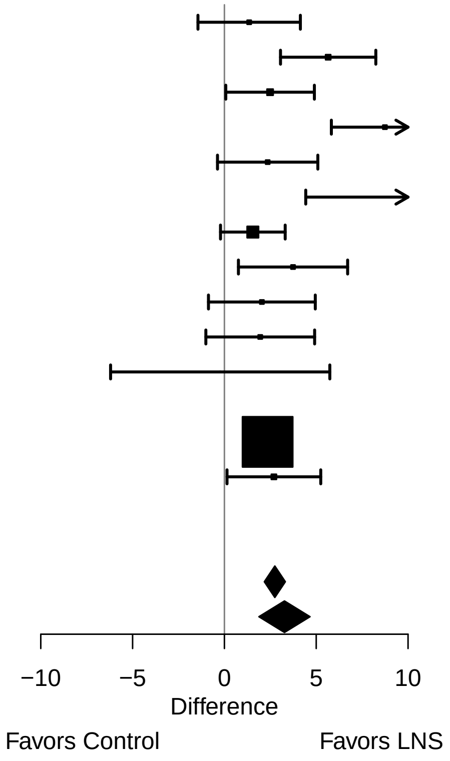

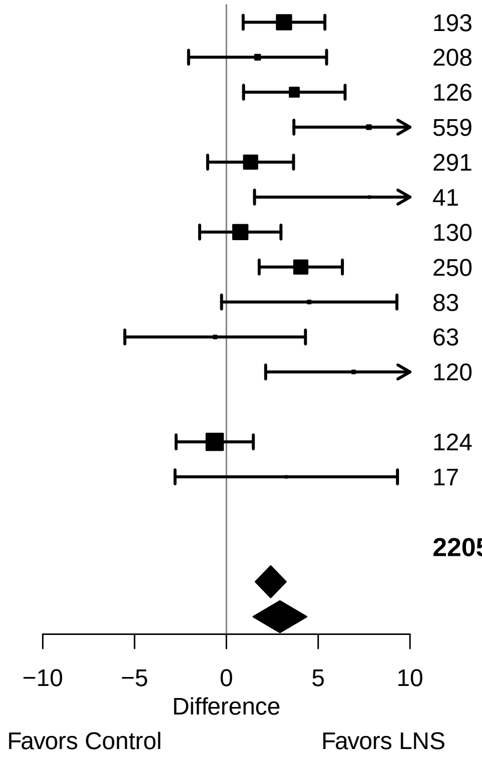

### Supplemental figure 7B: Anemia prevalence ratio

#### 7B1: Stratified by Household socio-economic status

**P−for−interaction = 0.762**
**Ratio of PRs = 0.99 (0.95, 1.04)**

| Country | Trial | At least median: LNS N | Control N | Control Prevalence | PR (95% CI) | Fixed W | Random W | Less than median: LNS N | Control N | Control Prevalence | PR (95% CI) | Fixed W | Random W |
|---|---|---|---|---|---|---|---|---|---|---|---|---|---|
| Bangladesh | JiVitA−4 (35) | 244 | 79 | 17.7 | 0.62 (0.37, 1.05) | 0.01 | 0.02 | 212 | 67 | 13.4 | 1.16 (0.64, 2.10) | 0.00 | 0.02 |
| Bangladesh | RDNS (36) | 272 | 136 | 41.9 | 0.57 (0.40, 0.81) | 0.01 | 0.04 | 277 | 136 | 41.9 | 0.74 (0.56, 0.97) | 0.02 | 0.06 |
| Bangladesh | WASH−B (37) | | | | | | | | | | | | |
| Burkina Faso | iLiNS−Zinc (38) | 1232 | 299 | 90.0 | 0.89 (0.84, 0.95) | 0.44 | 0.17 | 718 | 364 | 92.0 | 0.84 (0.79, 0.89) | 0.40 | 0.13 |
| Burkina Faso | PROMIS CS (39) | 289 | 293 | 70.0 | 0.93 (0.80, 1.08) | 0.07 | 0.11 | 285 | 288 | 70.1 | 0.94 (0.85, 1.04) | 0.16 | 0.12 |
| Ghana | GHANA (40) | 57 | 44 | 45.5 | 0.58 (0.34, 1.00) | 0.01 | 0.02 | 41 | 46 | 71.7 | 0.48 (0.30, 0.76) | 0.01 | 0.03 |
| Ghana | iLiNS−DYADG (41) | 149 | 344 | 41.6 | 0.89 (0.70, 1.13) | 0.03 | 0.07 | 179 | 315 | 48.6 | 0.83 (0.67, 1.02) | 0.03 | 0.08 |
| Kenya | WASH−B (42) | 190 | 175 | 48.0 | 0.65 (0.49, 0.85) | 0.02 | 0.06 | 159 | 125 | 46.4 | 0.73 (0.53, 1.00) | 0.01 | 0.05 |
| Madagascar | MAHAY (43) | 278 | 311 | 64.0 | 0.79 (0.65, 0.95) | 0.04 | 0.09 | 304 | 262 | 65.6 | 0.96 (0.81, 1.13) | 0.06 | 0.10 |
| Malawi | iLiNS−DYADM (44) | 117 | 225 | 48.4 | 0.83 (0.64, 1.07) | 0.02 | 0.06 | 93 | 206 | 55.8 | 0.98 (0.79, 1.23) | 0.03 | 0.08 |
| Malawi | iLiNS−DOSE (45) | 86 | 32 | 78.1 | 0.71 (0.55, 0.93) | 0.02 | 0.06 | 114 | 39 | 61.5 | 1.08 (0.84, 1.40) | 0.02 | 0.07 |
| Mali | PROMIS CS (46) | 505 | 463 | 85.7 | 0.82 (0.75, 0.89) | 0.23 | 0.15 | 448 | 507 | 86.6 | 0.82 (0.75, 0.89) | 0.18 | 0.12 |
| Zimbabwe | SHINE (HIV−) (47) | 821 | 722 | 35.3 | 0.73 (0.64, 0.84) | 0.08 | 0.12 | 742 | 760 | 34.9 | 0.78 (0.68, 0.91) | 0.07 | 0.10 |
| Zimbabwe | SHINE (HIV+) (48) | 155 | 138 | 34.1 | 0.61 (0.41, 0.90) | 0.01 | 0.03 | 145 | 140 | 40.0 | 0.66 (0.46, 0.93) | 0.01 | 0.05 |
| | | **4395** | **3261** | | **I² = 0.54, Tau² = 0.01** | | | **3717** | **3255** | | **I² = 0.51, Tau² = 0.02** | | |
| **Fixed** | | | | | **0.83 (0.80, 0.87)** | | | | | | **0.85 (0.82, 0.89)** | | |
| **Random** | | | | | **0.79 (0.73, 0.85)** | | | | | | **0.85 (0.77, 0.93)** | | |

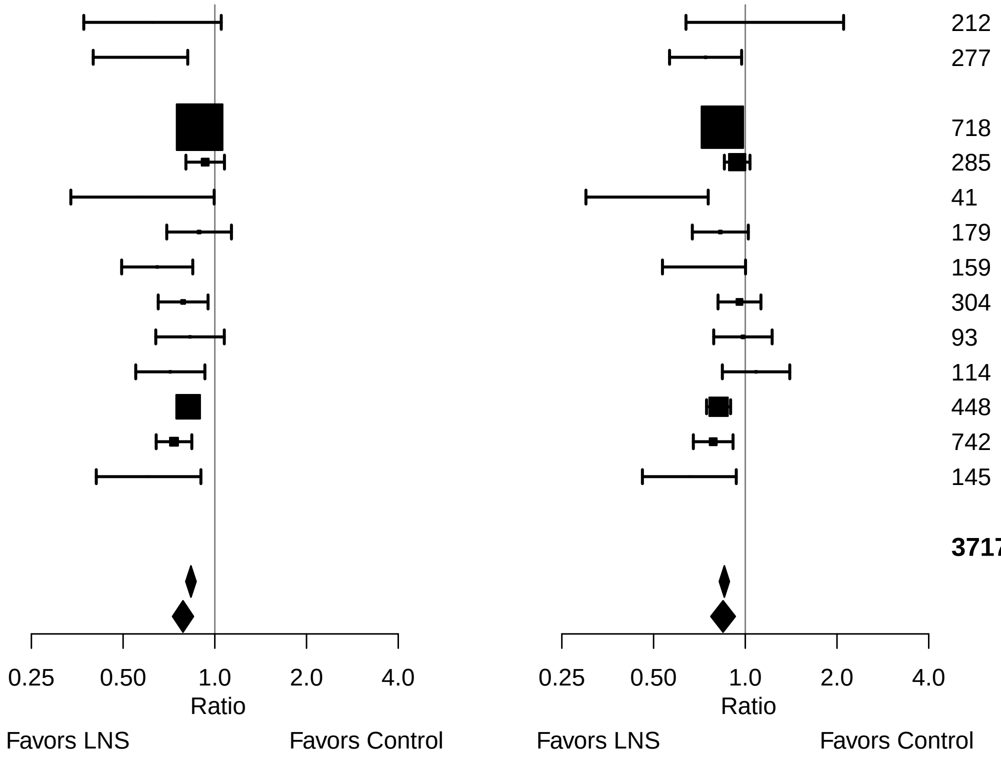

**Supplemental figure 7B: Anemia prevalence ratio**

#### 7B2: Stratified by Household food insecurity

**P−for−interaction = 0.949**
**Ratio of PRs = 1.00 (0.95, 1.05)**

| | | Mild to secure | | | | | | Moderate to severe | | | | | |
|---|---|---|---|---|---|---|---|---|---|---|---|---|---|
| Country | Trial | LNS N | Control N | Control Prevalence | PR (95% CI) | Fixed W | Random W | LNS N | Control N | Control Prevalence | PR (95% CI) | Fixed W | Random W |
| Bangladesh | JiVitA−4 (35) | | | | | | | | | | | | |
| Bangladesh | RDNS (36) | 342 | 166 | 40.4 | 0.65 (0.46, 0.92) | 0.01 | 0.06 | 207 | 106 | 44.3 | 0.66 (0.51, 0.87) | 0.04 | 0.11 |
| Bangladesh | WASH−B (37) | | | | | | | | | | | | |
| Burkina Faso | iLiNS−Zinc (38) | 1016 | 320 | 91.9 | 0.87 (0.83, 0.92) | 0.61 | 0.20 | 934 | 343 | 90.4 | 0.87 (0.82, 0.92) | 0.71 | 0.20 |
| Burkina Faso | PROMIS CS (39) | | | | | | | | | | | | |
| Ghana | GHANA (40) | | | | | | | | | | | | |
| Ghana | iLiNS−DYADG (41) | 230 | 457 | 47.0 | 0.88 (0.73, 1.05) | 0.05 | 0.12 | 97 | 201 | 40.8 | 0.81 (0.58, 1.12) | 0.02 | 0.09 |
| Kenya | WASH−B (42) | 304 | 273 | 48.7 | 0.65 (0.54, 0.78) | 0.05 | 0.12 | 45 | 27 | 33.3 | 1.13 (0.65, 1.96) | 0.01 | 0.04 |
| Madagascar | MAHAY (43) | 359 | 340 | 69.4 | 0.83 (0.72, 0.95) | 0.09 | 0.15 | 137 | 140 | 63.6 | 1.01 (0.82, 1.24) | 0.06 | 0.14 |
| Malawi | iLiNS−DYADM (44) | 66 | 113 | 58.4 | 0.78 (0.57, 1.06) | 0.02 | 0.07 | 143 | 314 | 50.0 | 0.94 (0.76, 1.15) | 0.06 | 0.14 |
| Malawi | iLiNS−DOSE (45) | 50 | 21 | 61.9 | 1.12 (0.78, 1.61) | 0.01 | 0.05 | 144 | 46 | 69.6 | 0.87 (0.69, 1.09) | 0.05 | 0.13 |
| Mali | PROMIS CS (46) | | | | | | | | | | | | |
| Zimbabwe | SHINE (HIV−) (47) | 1253 | 1162 | 34.5 | 0.80 (0.71, 0.90) | 0.13 | 0.16 | 264 | 293 | 36.9 | 0.61 (0.47, 0.79) | 0.04 | 0.11 |
| Zimbabwe | SHINE (HIV+) (48) | 217 | 204 | 35.3 | 0.64 (0.47, 0.87) | 0.02 | 0.07 | 75 | 66 | 42.4 | 0.60 (0.36, 1.00) | 0.01 | 0.05 |
| | | **3837** | **3056** | | **I² = 0.55, Tau² = 0.01** | | | **2046** | **1536** | | **I² = 0.51, Tau² = 0.02** | | |
| **Fixed** | | | | | **0.84 (0.80, 0.87)** | | | | | | **0.86 (0.81, 0.90)** | | |
| **Random** | | | | | **0.80 (0.72, 0.88)** | | | | | | **0.83 (0.73, 0.94)** | | |

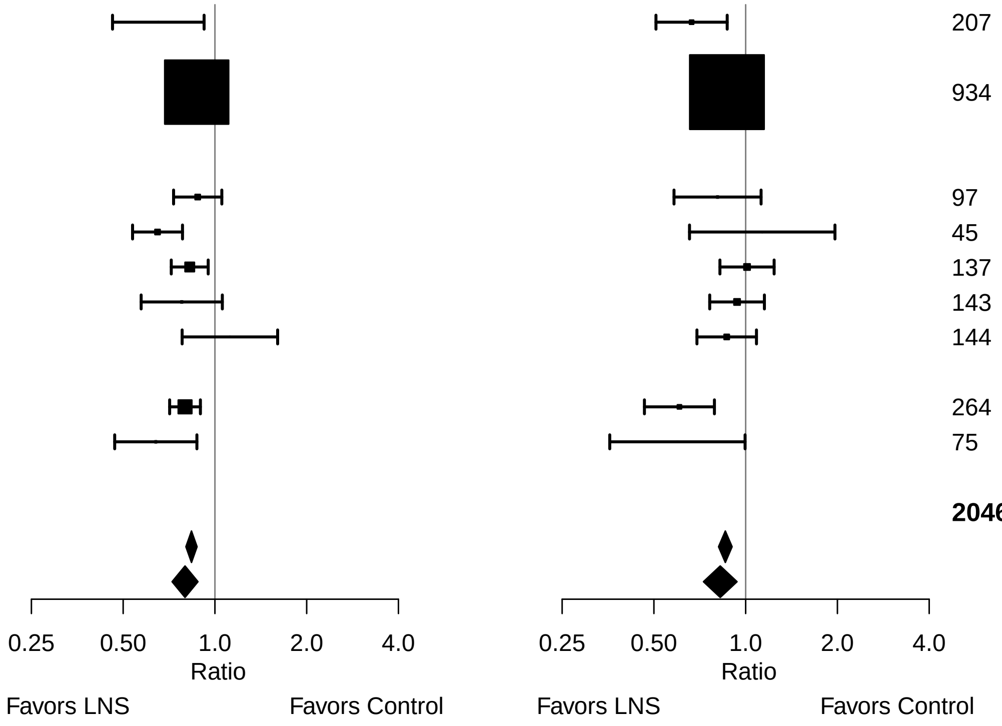

#### Supplemental figure 7B: Anemia prevalence ratio

##### 7B3: Stratified by Household source water quality

**P−for−interaction = 0.009**
**Ratio of PRs = 1.08 (1.02, 1.14)**

| Country | Trial | Improved LNS N | Improved Control N | Improved Control Prevalence | Improved PR (95% CI) | Improved Fixed W | Improved Random W | Unimproved LNS N | Unimproved Control N | Unimproved Control Prevalence | Unimproved PR (95% CI) | Unimproved Fixed W | Unimproved Random W |
|---|---|---|---|---|---|---|---|---|---|---|---|---|---|
| Bangladesh | JiVitA−4 (35) | | | | | | | | | | | | |
| Bangladesh | RDNS (36) | | | | | | | | | | | | |
| Bangladesh | WASH−B (37) | | | | | | | | | | | | |
| Burkina Faso | iLiNS−Zinc (38) | 555 | 141 | 96.5 | 0.80 (0.76, 0.85) | 0.51 | 0.30 | 1394 | 519 | 89.6 | 0.89 (0.84, 0.95) | 0.57 | 0.57 |
| Burkina Faso | PROMIS CS (39) | 373 | 363 | 71.1 | 0.92 (0.82, 1.04) | 0.13 | 0.17 | 201 | 217 | 68.2 | 0.96 (0.81, 1.12) | 0.07 | 0.07 |
| Ghana | GHANA (40) | | | | | | | | | | | | |
| Ghana | iLiNS−DYADG (41) | | | | | | | | | | | | |
| Kenya | WASH−B (42) | 128 | 121 | 52.9 | 0.72 (0.55, 0.94) | 0.03 | 0.05 | 68 | 41 | 36.6 | 0.88 (0.51, 1.52) | 0.01 | 0.01 |
| Madagascar | MAHAY (43) | 137 | 164 | 66.5 | 0.90 (0.71, 1.14) | 0.03 | 0.07 | 433 | 408 | 64.2 | 0.87 (0.75, 1.01) | 0.08 | 0.08 |
| Malawi | iLiNS−DYADM (44) | 191 | 397 | 51.1 | 0.91 (0.76, 1.09) | 0.06 | 0.10 | 19 | 33 | 60.6 | 0.78 (0.45, 1.35) | 0.01 | 0.01 |
| Malawi | iLiNS−DOSE (45) | | | | | | | | | | | | |
| Mali | PROMIS CS (46) | 548 | 567 | 86.8 | 0.81 (0.74, 0.89) | 0.20 | 0.21 | 364 | 388 | 85.6 | 0.82 (0.75, 0.90) | 0.21 | 0.21 |
| Zimbabwe | SHINE (HIV−) (47) | 469 | 436 | 33.9 | 0.69 (0.56, 0.86) | 0.04 | 0.07 | 259 | 281 | 33.8 | 0.82 (0.66, 1.03) | 0.04 | 0.04 |
| Zimbabwe | SHINE (HIV+) (48) | 77 | 77 | 36.4 | 0.68 (0.42, 1.10) | 0.01 | 0.02 | 49 | 46 | 28.3 | 1.01 (0.48, 2.12) | 0.00 | 0.00 |
| | | **2478** | **2266** | | **$I^2$ = 0.31, $Tau^2$ = 0.00** | | | **2787** | **1933** | | **$I^2$ = 0.00, $Tau^2$ = 0.00** | | |
| **Fixed** | | | | | **0.82 (0.79, 0.86)** | | | | | | **0.88 (0.84, 0.92)** | | |
| **Random** | | | | | **0.83 (0.77, 0.88)** | | | | | | **0.88 (0.84, 0.92)** | | |

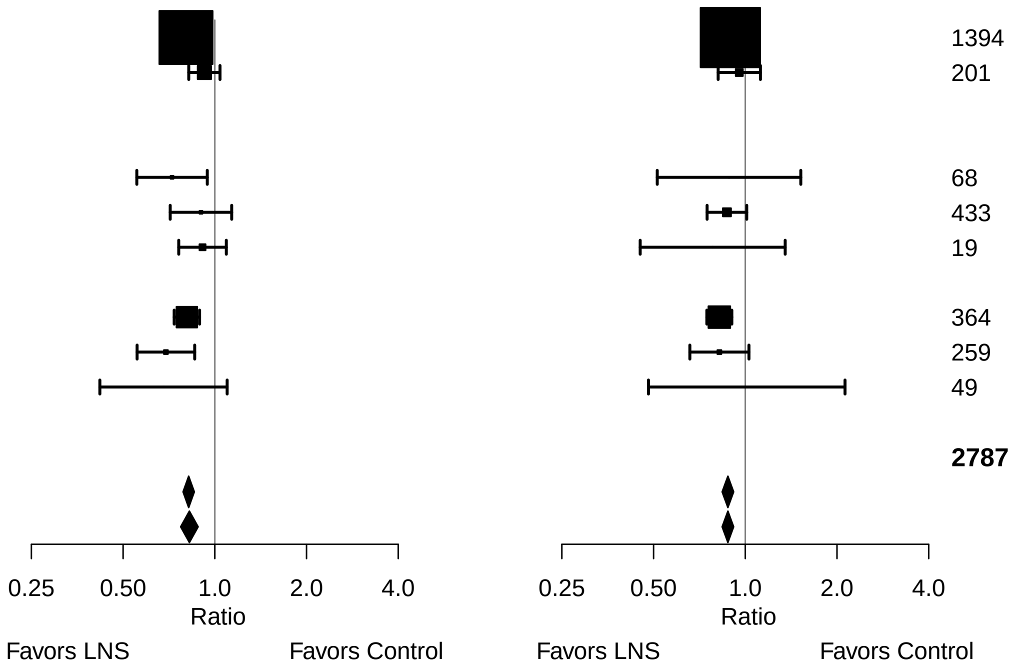

**Supplemental figure 7B: Anemia prevalence ratio**

#### 7B4: Stratified by Household sanitation

**P−for−interaction = 0.092**
**Ratio of PRs = 0.92 (0.83, 1.01)**

| Country | Trial | LNS N | Control N | Control Prevalence | Improved PR (95% CI) | Fixed W | Random W | LNS N | Control N | Control Prevalence | Unimproved PR (95% CI) | Fixed W | Random W |
|---|---|---|---|---|---|---|---|---|---|---|---|---|---|
| Bangladesh | JiVitA−4 (35) | | | | | | | | | | | | |
| Bangladesh | RDNS (36) | 376 | 193 | 38.9 | 0.72 (0.52, 0.98) | 0.04 | 0.04 | 173 | 79 | 49.4 | 0.54 (0.40, 0.72) | 0.07 | 0.14 |
| Bangladesh | WASH−B (37) | | | | | | | | | | | | |
| Burkina Faso | iLiNS−Zinc (38) | | | | | | | | | | | | |
| Burkina Faso | PROMIS CS (39) | 244 | 229 | 69.9 | 0.92 (0.81, 1.05) | 0.24 | 0.24 | 329 | 350 | 70.0 | 0.95 (0.84, 1.07) | 0.37 | 0.23 |
| Ghana | GHANA (40) | | | | | | | | | | | | |
| Ghana | iLiNS−DYADG (41) | | | | | | | | | | | | |
| Kenya | WASH−B (42) | 24 | 25 | 44.0 | 0.47 (0.20, 1.10) | 0.01 | 0.01 | 172 | 137 | 49.6 | 0.77 (0.61, 0.98) | 0.10 | 0.16 |
| Madagascar | MAHAY (43) | | | | | | | | | | | | |
| Malawi | iLiNS−DYADM (44) | | | | | | | | | | | | |
| Malawi | iLiNS−DOSE (45) | | | | | | | | | | | | |
| Mali | PROMIS CS (46) | 703 | 696 | 86.1 | 0.84 (0.77, 0.91) | 0.62 | 0.62 | 219 | 249 | 86.7 | 0.73 (0.64, 0.83) | 0.34 | 0.22 |
| Zimbabwe | SHINE (HIV−) (47) | 262 | 218 | 31.7 | 0.88 (0.70, 1.10) | 0.09 | 0.09 | 465 | 498 | 34.7 | 0.67 (0.54, 0.84) | 0.11 | 0.17 |
| Zimbabwe | SHINE (HIV+) (48) | 46 | 30 | 30.0 | 0.72 (0.34, 1.52) | 0.01 | 0.01 | 80 | 92 | 33.7 | 0.85 (0.53, 1.36) | 0.02 | 0.08 |
| | | **1655** | **1391** | | **$I^2$ = 0.00, $Tau^2$ = 0.00** | | | **1438** | **1405** | | **$I^2$ = 0.74, $Tau^2$ = 0.02** | | |
| **Fixed** | | | | | **0.85 (0.80, 0.91)** | | | | | | **0.79 (0.73, 0.85)** | | |
| **Random** | | | | | **0.85 (0.80, 0.91)** | | | | | | **0.75 (0.64, 0.88)** | | |

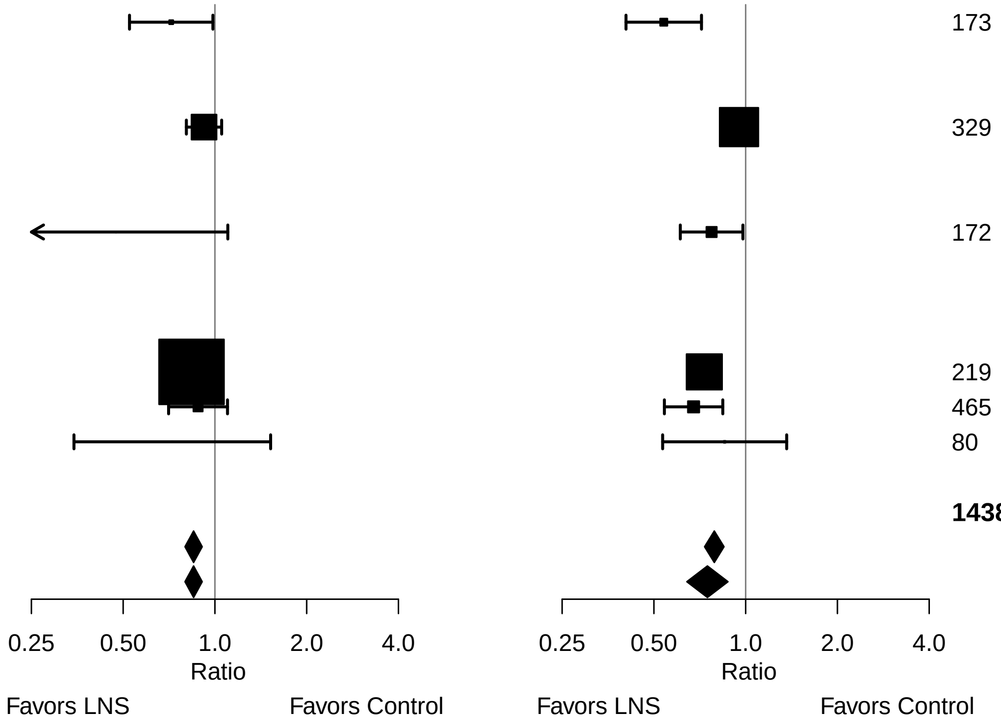

#### Supplemental figure 7B: Anemia prevalence ratio

##### 7B5: Stratified by Season at the time of assessment

**P−for−interaction = 0.434**
**Ratio of PRs = 1.03 (0.96, 1.10)**

| Country | Trial | Dry LNS N | Dry Control N | Dry Control Prevalence | Dry PR (95% CI) | Dry Fixed W | Dry Random W | Rainy LNS N | Rainy Control N | Rainy Control Prevalence | Rainy PR (95% CI) | Rainy Fixed W | Rainy Random W |
|---|---|---|---|---|---|---|---|---|---|---|---|---|---|
| Bangladesh | JiVitA−4 (35) | 264 | 72 | 13.9 | 1.09 (0.58, 2.05) | 0.00 | 0.02 | 193 | 74 | 17.6 | 0.59 (0.31, 1.12) | 0.01 | 0.04 |
| Bangladesh | RDNS (36) | 341 | 166 | 45.8 | 0.62 (0.47, 0.83) | 0.02 | 0.07 | 208 | 106 | 35.8 | 0.72 (0.48, 1.10) | 0.02 | 0.07 |
| Bangladesh | WASH−B (37) | 108 | 96 | 17.7 | 0.47 (0.23, 0.97) | 0.00 | 0.02 | 126 | 90 | 14.4 | 0.38 (0.17, 0.87) | 0.00 | 0.03 |
| Burkina Faso | iLiNS−Zinc (38) | 1398 | 475 | 92.6 | 0.86 (0.82, 0.90) | 0.58 | 0.14 | 559 | 189 | 87.3 | 0.90 (0.83, 0.98) | 0.43 | 0.14 |
| Burkina Faso | PROMIS CS (39) | 283 | 245 | 72.7 | 0.89 (0.77, 1.04) | 0.07 | 0.12 | 291 | 336 | 68.2 | 0.97 (0.86, 1.09) | 0.24 | 0.13 |
| Ghana | GHANA (40) | 57 | 55 | 60.0 | 0.50 (0.32, 0.78) | 0.01 | 0.04 | 41 | 41 | 56.1 | 0.52 (0.30, 0.90) | 0.01 | 0.05 |
| Ghana | iLiNS−DYADG (41) | 198 | 386 | 45.6 | 0.80 (0.64, 0.99) | 0.03 | 0.09 | 130 | 274 | 43.8 | 0.97 (0.76, 1.23) | 0.06 | 0.10 |
| Kenya | WASH−B (42) | 100 | 88 | 35.2 | 0.74 (0.53, 1.02) | 0.01 | 0.06 | 250 | 212 | 52.4 | 0.66 (0.55, 0.81) | 0.08 | 0.11 |
| Madagascar | MAHAY (43) | 517 | 537 | 66.7 | 0.94 (0.83, 1.07) | 0.10 | 0.12 | 83 | 51 | 45.1 | 0.51 (0.32, 0.81) | 0.02 | 0.06 |
| Malawi | iLiNS−DYADM (44) | 147 | 305 | 46.6 | 0.91 (0.72, 1.13) | 0.03 | 0.09 | 63 | 125 | 64.8 | 0.88 (0.69, 1.13) | 0.05 | 0.10 |
| Malawi | iLiNS−DOSE (45) | 123 | 32 | 68.8 | 1.01 (0.78, 1.30) | 0.02 | 0.08 | 120 | 50 | 74.0 | 0.78 (0.62, 0.97) | 0.07 | 0.11 |
| Mali | PROMIS CS (46) | | | | | | | | | | | | |
| Zimbabwe | SHINE (HIV−) (47) | 1558 | 1477 | 36.0 | 0.76 (0.68, 0.84) | 0.13 | 0.13 | 124 | 117 | 25.6 | 1.10 (0.74, 1.64) | 0.02 | 0.07 |
| Zimbabwe | SHINE (HIV+) (48) | | | | | | | | | | | | |
| | | **5094** | **3934** | | **I² = 0.54, Tau² = 0.02** | | | **2188** | **1665** | | **I² = 0.62, Tau² = 0.04** | | |
| **Fixed** | | | | | **0.85 (0.82, 0.88)** | | | | | | **0.87 (0.82, 0.92)** | | |
| **Random** | | | | | **0.82 (0.73, 0.91)** | | | | | | **0.79 (0.68, 0.91)** | | |

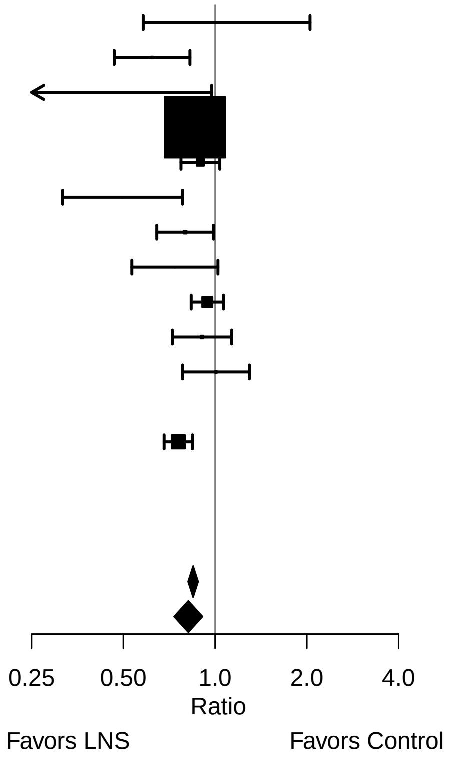

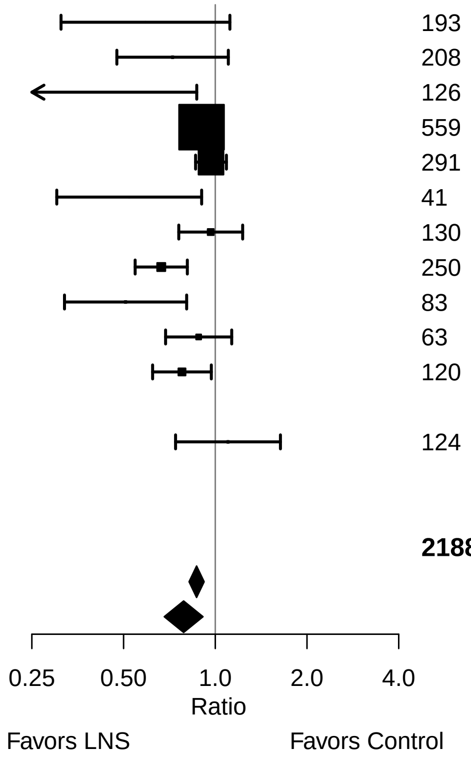

### Supplemental figure 7C: Anemia prevalence difference

#### 7C1: Stratified by Household socio-economic status

**P−for−interaction = 0.499**
**Difference in PDs = 0.01 (−0.02, 0.04)**

| | | At least median | | | | | | Less than median | | | | | |
|---|---|---|---|---|---|---|---|---|---|---|---|---|---|
| Country | Trial | LNS N | Control N | Control Prevalence | PD (95% CI) | Fixed W | Random W | LNS N | Control N | Control Prevalence | PD (95% CI) | Fixed W | Random W |
| Bangladesh | JiVitA−4 (35) | 244 | 79 | 17.7 | −0.07 (−0.14, 0.01) | 0.08 | 0.08 | 212 | 67 | 13.4 | 0.02 (−0.06, 0.10) | 0.07 | 0.09 |
| Bangladesh | RDNS (36) | 272 | 136 | 41.9 | −0.18 (−0.30, −0.06) | 0.03 | 0.04 | 277 | 136 | 41.9 | −0.11 (−0.21, −0.01) | 0.05 | 0.08 |
| Bangladesh | WASH−B (37) | | | | | | | | | | | | |
| Burkina Faso | iLiNS−Zinc (38) | 1232 | 299 | 90.0 | −0.10 (−0.15, −0.05) | 0.18 | 0.17 | 718 | 364 | 92.0 | −0.15 (−0.20, −0.10) | 0.19 | 0.10 |
| Burkina Faso | PROMIS CS (39) | 289 | 293 | 70.0 | −0.05 (−0.15, 0.05) | 0.05 | 0.05 | 285 | 288 | 70.1 | −0.04 (−0.11, 0.02) | 0.12 | 0.10 |
| Ghana | GHANA (40) | 57 | 44 | 45.5 | −0.19 (−0.38, −0.01) | 0.01 | 0.02 | 41 | 46 | 71.7 | −0.38 (−0.57, −0.18) | 0.01 | 0.04 |
| Ghana | iLiNS−DYADG (41) | 149 | 344 | 41.6 | −0.05 (−0.14, 0.05) | 0.05 | 0.06 | 179 | 315 | 48.6 | −0.08 (−0.17, 0.01) | 0.06 | 0.08 |
| Kenya | WASH−B (42) | 190 | 175 | 48.0 | −0.17 (−0.26, −0.08) | 0.05 | 0.06 | 159 | 125 | 46.4 | −0.12 (−0.25, 0.01) | 0.03 | 0.06 |
| Madagascar | MAHAY (43) | 278 | 311 | 64.0 | −0.14 (−0.24, −0.03) | 0.04 | 0.05 | 304 | 262 | 65.6 | −0.03 (−0.13, 0.08) | 0.04 | 0.07 |
| Malawi | iLiNS−DYADM (44) | 117 | 225 | 48.4 | −0.08 (−0.19, 0.03) | 0.04 | 0.04 | 93 | 206 | 55.8 | −0.01 (−0.13, 0.11) | 0.03 | 0.07 |
| Malawi | iLiNS−DOSE (45) | 86 | 32 | 78.1 | −0.22 (−0.42, −0.03) | 0.01 | 0.01 | 114 | 39 | 61.5 | 0.05 (−0.12, 0.22) | 0.02 | 0.05 |
| Mali | PROMIS CS (46) | 505 | 463 | 85.7 | −0.16 (−0.22, −0.10) | 0.13 | 0.13 | 448 | 507 | 86.6 | −0.16 (−0.23, −0.09) | 0.11 | 0.09 |
| Zimbabwe | SHINE (HIV−) (47) | 821 | 722 | 35.3 | −0.09 (−0.14, −0.05) | 0.27 | 0.24 | 742 | 760 | 34.9 | −0.08 (−0.12, −0.03) | 0.23 | 0.10 |
| Zimbabwe | SHINE (HIV+) (48) | 155 | 138 | 34.1 | −0.13 (−0.24, −0.03) | 0.04 | 0.05 | 145 | 140 | 40.0 | −0.14 (−0.25, −0.03) | 0.04 | 0.07 |
| | | **4395** | **3261** | | **$I^2$ = 0.08, $Tau^2$ = 0.00** | | | **3717** | **3255** | | **$I^2$ = 0.64, $Tau^2$ = 0.01** | | |
| **Fixed** | | | | | **−0.11 (−0.13, −0.09)** | | | | | | **−0.09 (−0.11, −0.07)** | | |
| **Random** | | | | | **−0.11 (−0.13, −0.09)** | | | | | | **−0.09 (−0.14, −0.04)** | | |

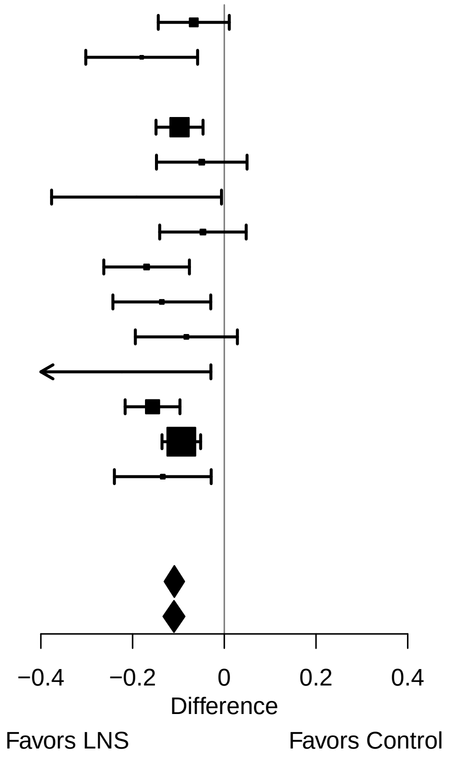

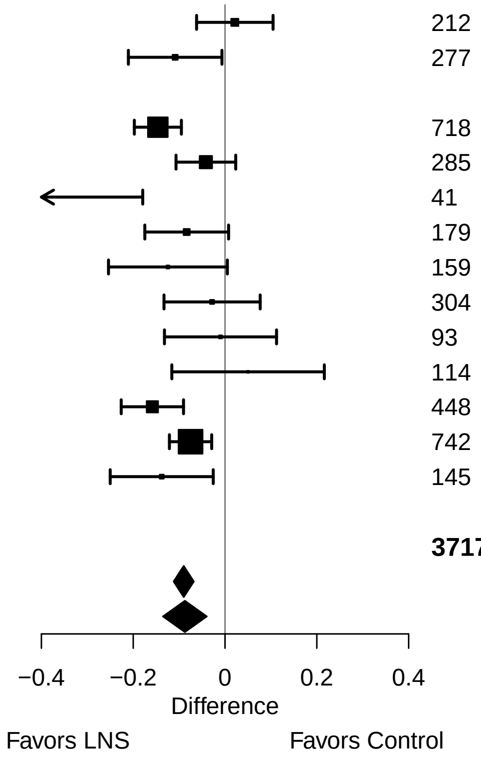

#### Supplemental figure 7C: Anemia prevalence difference

#### 7C2: Stratified by Household food insecurity

**P−for−interaction = 0.857**
**Difference in PDs = 0.00 (−0.04, 0.03)**

| | | Mild to secure | | | | | | Moderate to severe | | | | | |
|---|---|---|---|---|---|---|---|---|---|---|---|---|---|
| Country | Trial | LNS N | Control N | Control Prevalence | PD (95% CI) | Fixed W | Random W | LNS N | Control N | Control Prevalence | PD (95% CI) | Fixed W | Random W |
| Bangladesh | JiVitA−4 (35) | | | | | | | | | | | | |
| Bangladesh | RDNS (36) | 342 | 166 | 40.4 | −0.14 (−0.27, −0.01) | 0.03 | 0.05 | 207 | 106 | 44.3 | −0.15 (−0.24, −0.05) | 0.11 | 0.12 |
| Bangladesh | WASH−B (37) | | | | | | | | | | | | |
| Burkina Faso | iLiNS−Zinc (38) | 1016 | 320 | 91.9 | −0.12 (−0.17, −0.07) | 0.23 | 0.22 | 934 | 343 | 90.4 | −0.12 (−0.17, −0.07) | 0.38 | 0.26 |
| Burkina Faso | PROMIS CS (39) | | | | | | | | | | | | |
| Ghana | GHANA (40) | | | | | | | | | | | | |
| Ghana | iLiNS−DYADG (41) | 230 | 457 | 47.0 | −0.06 (−0.14, 0.02) | 0.08 | 0.11 | 97 | 201 | 40.8 | −0.08 (−0.20, 0.04) | 0.07 | 0.09 |
| Kenya | WASH−B (42) | 304 | 273 | 48.7 | −0.17 (−0.25, −0.10) | 0.09 | 0.11 | 45 | 27 | 33.3 | 0.04 (−0.15, 0.24) | 0.03 | 0.04 |
| Madagascar | MAHAY (43) | 359 | 340 | 69.4 | −0.12 (−0.21, −0.03) | 0.06 | 0.09 | 137 | 140 | 63.6 | 0.01 (−0.12, 0.14) | 0.06 | 0.08 |
| Malawi | iLiNS−DYADM (44) | 66 | 113 | 58.4 | −0.13 (−0.28, 0.02) | 0.02 | 0.03 | 143 | 314 | 50.0 | −0.03 (−0.13, 0.07) | 0.10 | 0.12 |
| Malawi | iLiNS−DOSE (45) | 50 | 21 | 61.9 | 0.07 (−0.17, 0.32) | 0.01 | 0.01 | 144 | 46 | 69.6 | −0.09 (−0.25, 0.06) | 0.04 | 0.06 |
| Mali | PROMIS CS (46) | | | | | | | | | | | | |
| Zimbabwe | SHINE (HIV−) (47) | 1253 | 1162 | 34.5 | −0.07 (−0.11, −0.03) | 0.40 | 0.29 | 264 | 293 | 36.9 | −0.15 (−0.22, −0.07) | 0.18 | 0.18 |
| Zimbabwe | SHINE (HIV+) (48) | 217 | 204 | 35.3 | −0.13 (−0.21, −0.04) | 0.07 | 0.09 | 75 | 66 | 42.4 | −0.17 (−0.33, −0.01) | 0.04 | 0.05 |
| | | **3837** | **3056** | | **I² = 0.27, Tau² = 0.00** | | | **2046** | **1536** | | **I² = 0.22, Tau² = 0.00** | | |
| **Fixed** | | | | | **−0.10 (−0.12, −0.08)** | | | | | | **−0.10 (−0.14, −0.07)** | | |
| **Random** | | | | | **−0.10 (−0.13, −0.07)** | | | | | | **−0.10 (−0.14, −0.06)** | | |

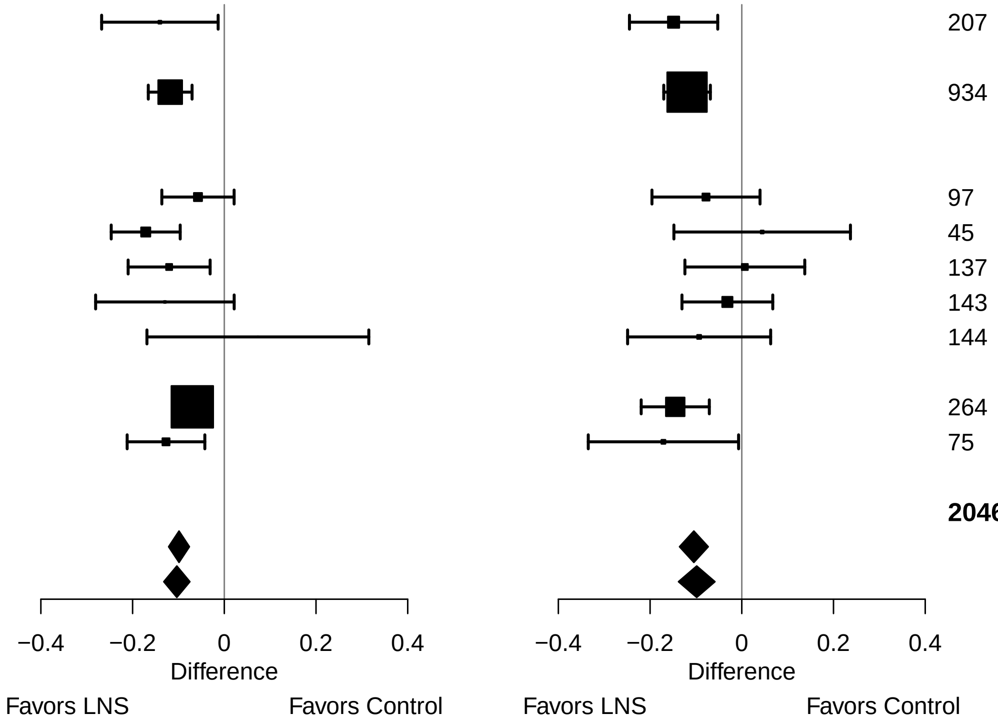

#### Supplemental figure 7C: Anemia prevalence difference

##### 7C3: Stratified by Household source water quality

**P−for−interaction = 0.005**
**Difference in PDs = 0.05 (0.02, 0.09)**

| Country | Trial | Improved LNS N | Improved Control N | Improved Control Prevalence | Improved PD (95% CI) | Improved Fixed W | Improved Random W | Unimproved LNS N | Unimproved Control N | Unimproved Control Prevalence | Unimproved PD (95% CI) | Unimproved Fixed W | Unimproved Random W |
|---|---|---|---|---|---|---|---|---|---|---|---|---|---|
| Bangladesh | JiVitA−4 (35) | | | | | | | | | | | | |
| Bangladesh | RDNS (36) | | | | | | | | | | | | |
| Bangladesh | WASH−B (37) | | | | | | | | | | | | |
| Burkina Faso | iLiNS−Zinc (38) | 555 | 141 | 96.5 | −0.19 (−0.24, −0.14) | 0.31 | 0.21 | 1394 | 519 | 89.6 | −0.10 (−0.14, −0.05) | 0.37 | 0.37 |
| Burkina Faso | PROMIS CS (39) | 373 | 363 | 71.1 | −0.05 (−0.13, 0.03) | 0.12 | 0.14 | 201 | 217 | 68.2 | −0.03 (−0.14, 0.08) | 0.08 | 0.08 |
| Ghana | GHANA (40) | | | | | | | | | | | | |
| Ghana | iLiNS−DYADG (41) | | | | | | | | | | | | |
| Kenya | WASH−B (42) | 128 | 121 | 52.9 | −0.15 (−0.26, −0.03) | 0.06 | 0.08 | 68 | 41 | 36.6 | −0.04 (−0.23, 0.15) | 0.03 | 0.03 |
| Madagascar | MAHAY (43) | 137 | 164 | 66.5 | −0.07 (−0.22, 0.08) | 0.03 | 0.06 | 433 | 408 | 64.2 | −0.08 (−0.17, 0.01) | 0.11 | 0.11 |
| Malawi | iLiNS−DYADM (44) | 191 | 397 | 51.1 | −0.05 (−0.13, 0.04) | 0.10 | 0.13 | 19 | 33 | 60.6 | −0.13 (−0.42, 0.15) | 0.01 | 0.01 |
| Malawi | iLiNS−DOSE (45) | | | | | | | | | | | | |
| Mali | PROMIS CS (46) | 548 | 567 | 86.8 | −0.17 (−0.24, −0.09) | 0.15 | 0.16 | 364 | 388 | 85.6 | −0.15 (−0.22, −0.08) | 0.18 | 0.18 |
| Zimbabwe | SHINE (HIV−) (47) | 469 | 436 | 33.9 | −0.10 (−0.17, −0.04) | 0.19 | 0.17 | 259 | 281 | 33.8 | −0.06 (−0.13, 0.01) | 0.20 | 0.20 |
| Zimbabwe | SHINE (HIV+) (48) | 77 | 77 | 36.4 | −0.12 (−0.27, 0.03) | 0.03 | 0.06 | 49 | 46 | 28.3 | 0.00 (−0.21, 0.21) | 0.02 | 0.02 |
| | | **2478** | **2266** | | **I² = 0.53, Tau² = 0.00** | | | **2787** | **1933** | | **I² = 0.00, Tau² = 0.00** | | |
| **Fixed** | | | | | **−0.13 (−0.16, −0.10)** | | | | | | **−0.09 (−0.12, −0.06)** | | |
| **Random** | | | | | **−0.12 (−0.16, −0.08)** | | | | | | **−0.09 (−0.12, −0.06)** | | |

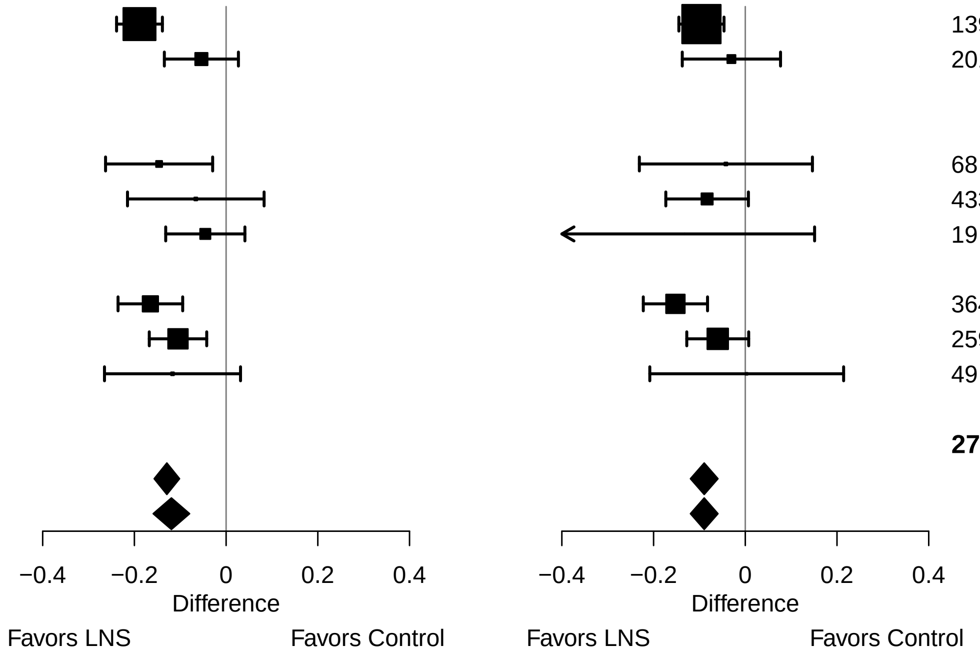

#### Supplemental figure 7C: Anemia prevalence difference

#### 7C4: Stratified by Household sanitation

**P−for−interaction = 0.036**
**Difference in PDs = −0.05 (−0.11, 0.00)**

| | | Improved | | | | | | Unimproved | | | | | |
|---|---|---|---|---|---|---|---|---|---|---|---|---|---|
| Country | Trial | LNS N | Control N | Control Prevalence | PD (95% CI) | Fixed W | Random W | LNS N | Control N | Control Prevalence | PD (95% CI) | Fixed W | Random W |
| Bangladesh | JiVitA−4 (35) | | | | | | | | | | | | |
| Bangladesh | RDNS (36) | 376 | 193 | 38.9 | −0.11 (−0.22, 0.00) | 0.11 | 0.13 | 173 | 79 | 49.4 | −0.23 (−0.33, −0.12) | 0.11 | 0.15 |
| Bangladesh | WASH−B (37) | | | | | | | | | | | | |
| Burkina Faso | iLiNS−Zinc (38) | | | | | | | | | | | | |
| Burkina Faso | PROMIS CS (39) | 244 | 229 | 69.9 | −0.06 (−0.14, 0.03) | 0.17 | 0.19 | 329 | 350 | 70.0 | −0.03 (−0.12, 0.05) | 0.19 | 0.18 |
| Ghana | GHANA (40) | | | | | | | | | | | | |
| Ghana | iLiNS−DYADG (41) | | | | | | | | | | | | |
| Kenya | WASH−B (42) | 24 | 25 | 44.0 | −0.23 (−0.47, 0.01) | 0.02 | 0.03 | 172 | 137 | 49.6 | −0.11 (−0.22, −0.01) | 0.12 | 0.16 |
| Madagascar | MAHAY (43) | | | | | | | | | | | | |
| Malawi | iLiNS−DYADM (44) | | | | | | | | | | | | |
| Malawi | iLiNS−DOSE (45) | | | | | | | | | | | | |
| Mali | PROMIS CS (46) | 703 | 696 | 86.1 | −0.14 (−0.20, −0.08) | 0.35 | 0.31 | 219 | 249 | 86.7 | −0.23 (−0.32, −0.15) | 0.18 | 0.18 |
| Zimbabwe | SHINE (HIV−) (47) | 262 | 218 | 31.7 | −0.04 (−0.11, 0.03) | 0.30 | 0.28 | 465 | 498 | 34.7 | −0.11 (−0.17, −0.05) | 0.34 | 0.21 |
| Zimbabwe | SHINE (HIV+) (48) | 46 | 30 | 30.0 | −0.08 (−0.27, 0.10) | 0.04 | 0.05 | 80 | 92 | 33.7 | −0.05 (−0.20, 0.10) | 0.06 | 0.12 |
| | | **1655** | **1391** | | **I² = 0.26, Tau² = 0.00** | | | **1438** | **1405** | | **I² = 0.68, Tau² = 0.00** | | |
| **Fixed** | | | | | **−0.09 (−0.13, −0.05)** | | | | | | **−0.13 (−0.17, −0.09)** | | |
| **Random** | | | | | **−0.09 (−0.13, −0.05)** | | | | | | **−0.13 (−0.20, −0.06)** | | |

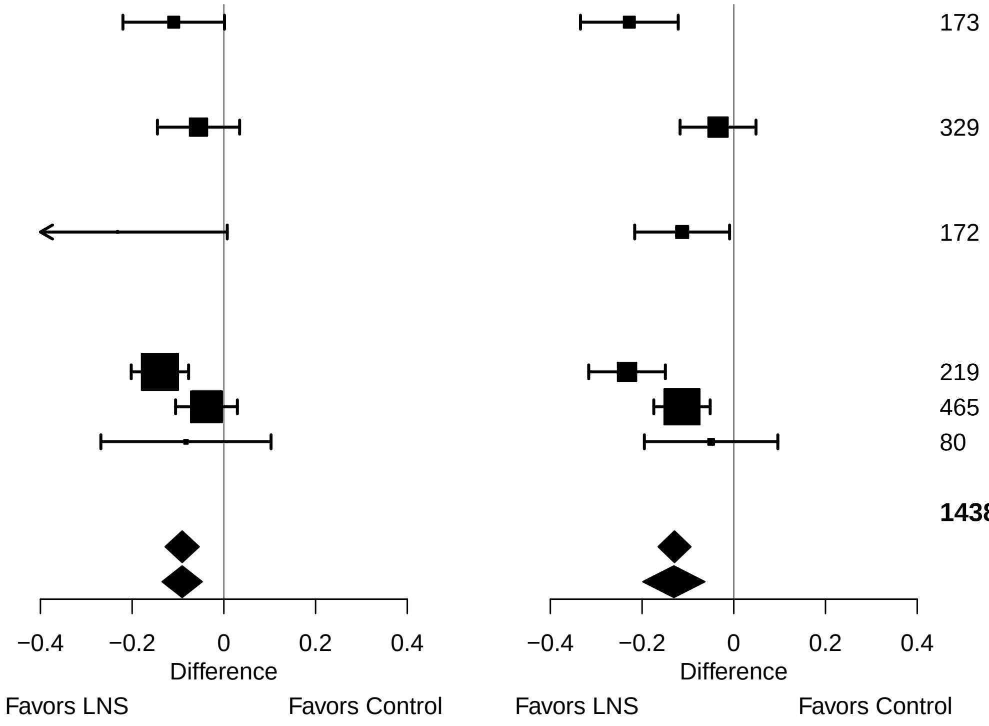

**Supplemental figure 7C: Anemia prevalence difference**

#### 7C5: Stratified by Season at the time of assessment

**P−for−interaction = 0.332**
**Difference in PDs = 0.02 (−0.02, 0.05)**

| | | Dry | | | | | | Rainy | | | | | |
|---|---|---|---|---|---|---|---|---|---|---|---|---|---|
| Country | Trial | LNS N | Control N | Control Prevalence | PD (95% CI) | Fixed W | Random W | LNS N | Control N | Control Prevalence | PD (95% CI) | Fixed W | Random W |
| Bangladesh | JiVitA−4 (35) | 264 | 72 | 13.9 | 0.01 (−0.08, 0.10) | 0.05 | 0.09 | 193 | 74 | 17.6 | −0.07 (−0.17, 0.02) | 0.09 | 0.10 |
| Bangladesh | RDNS (36) | 341 | 166 | 45.8 | −0.17 (−0.28, −0.07) | 0.04 | 0.07 | 208 | 106 | 35.8 | −0.10 (−0.24, 0.04) | 0.04 | 0.06 |
| Bangladesh | WASH−B (37) | 108 | 96 | 17.7 | −0.09 (−0.20, 0.01) | 0.04 | 0.07 | 126 | 90 | 14.4 | −0.09 (−0.17, −0.01) | 0.14 | 0.12 |
| Burkina Faso | iLiNS−Zinc (38) | 1398 | 475 | 92.6 | −0.13 (−0.17, −0.09) | 0.24 | 0.14 | 559 | 189 | 87.3 | −0.09 (−0.16, −0.01) | 0.16 | 0.12 |
| Burkina Faso | PROMIS CS (39) | 283 | 245 | 72.7 | −0.08 (−0.17, 0.02) | 0.04 | 0.08 | 291 | 336 | 68.2 | −0.02 (−0.10, 0.06) | 0.14 | 0.11 |
| Ghana | GHANA (40) | 57 | 55 | 60.0 | −0.30 (−0.48, −0.12) | 0.01 | 0.03 | 41 | 41 | 56.1 | −0.27 (−0.48, −0.06) | 0.02 | 0.04 |
| Ghana | iLiNS−DYADG (41) | 198 | 386 | 45.6 | −0.09 (−0.18, −0.01) | 0.06 | 0.09 | 130 | 274 | 43.8 | −0.01 (−0.12, 0.09) | 0.08 | 0.09 |
| Kenya | WASH−B (42) | 100 | 88 | 35.2 | −0.09 (−0.20, 0.02) | 0.04 | 0.07 | 250 | 212 | 52.4 | −0.18 (−0.26, −0.09) | 0.13 | 0.11 |
| Madagascar | MAHAY (43) | 517 | 537 | 66.7 | −0.04 (−0.12, 0.04) | 0.07 | 0.10 | 83 | 51 | 45.1 | −0.22 (−0.38, −0.07) | 0.04 | 0.06 |
| Malawi | iLiNS−DYADM (44) | 147 | 305 | 46.6 | −0.04 (−0.14, 0.05) | 0.04 | 0.08 | 63 | 125 | 64.8 | −0.08 (−0.22, 0.07) | 0.04 | 0.06 |
| Malawi | iLiNS−DOSE (45) | 123 | 32 | 68.8 | 0.00 (−0.17, 0.17) | 0.01 | 0.04 | 120 | 50 | 74.0 | −0.17 (−0.32, −0.01) | 0.03 | 0.05 |
| Mali | PROMIS CS (46) | | | | | | | | | | | | |
| Zimbabwe | SHINE (HIV−) (47) | 1558 | 1477 | 36.0 | −0.09 (−0.12, −0.05) | 0.36 | 0.15 | 124 | 117 | 25.6 | 0.03 (−0.08, 0.13) | 0.08 | 0.09 |
| Zimbabwe | SHINE (HIV+) (48) | | | | | | | | | | | | |
| | | **5094** | **3934** | | **I² = 0.45, Tau² = 0.00** | | | **2188** | **1665** | | **I² = 0.45, Tau² = 0.00** | | |
| **Fixed** | | | | | **−0.09 (−0.11, −0.07)** | | | | | | **−0.08 (−0.11, −0.05)** | | |
| **Random** | | | | | **−0.09 (−0.12, −0.05)** | | | | | | **−0.09 (−0.13, −0.05)** | | |

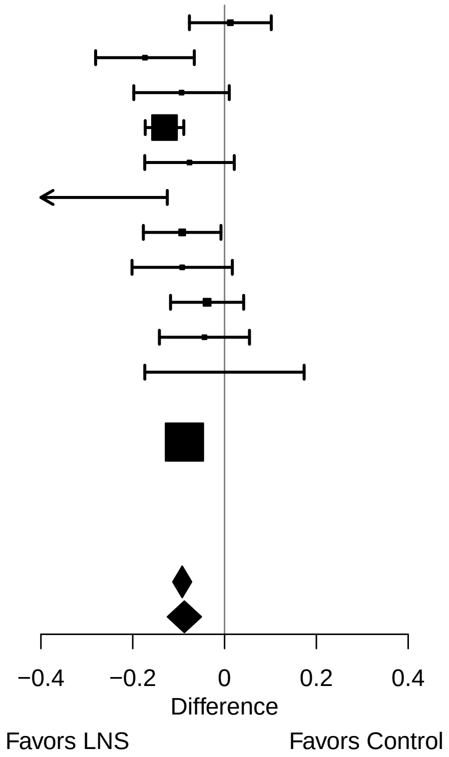

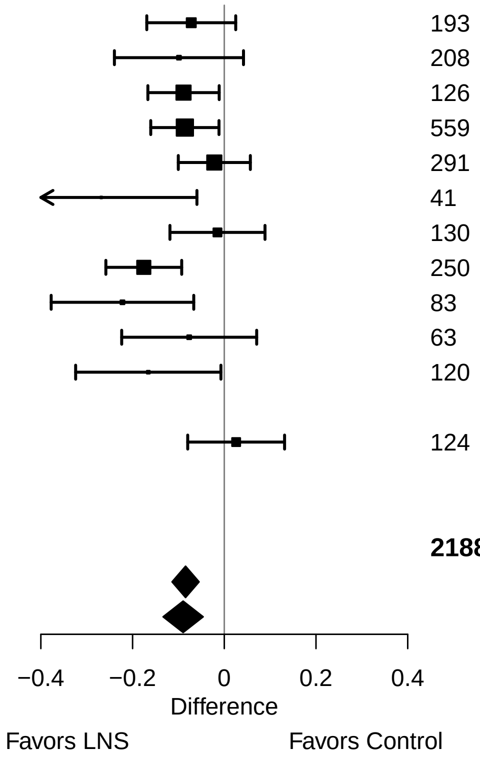

### Supplemental figure 7D: Moderate-to-severe anemia prevalence ratio

#### 7D1: Stratified by Household socio-economic status

**P−for−interaction = 0.244**
**Ratio of PRs = 0.95 (0.88, 1.03)**

**At least median**

| Country | Trial | LNS N | Control N | Control Prevalence | PR (95% CI) | Fixed W | Random W |
|---|---|---|---|---|---|---|---|
| Bangladesh | JiVitA−4 (35) | | | | | | |
| Bangladesh | RDNS (36) | 272 | 136 | 16.2 | 0.43 (0.24, 0.77) | 0.02 | 0.06 |
| Bangladesh | WASH−B (37) | | | | | | |
| Burkina Faso | iLiNS−Zinc (38) | 1232 | 299 | 73.6 | 0.73 (0.66, 0.82) | 0.51 | 0.16 |
| Burkina Faso | PROMIS CS (39) | 289 | 293 | 34.1 | 1.02 (0.76, 1.38) | 0.06 | 0.11 |
| Ghana | GHANA (40) | | | | | | |
| Ghana | iLiNS−DYADG (41) | 149 | 344 | 3.2 | 1.26 (0.47, 3.34) | 0.01 | 0.03 |
| Kenya | WASH−B (42) | 190 | 175 | 20.0 | 0.63 (0.41, 0.97) | 0.03 | 0.08 |
| Madagascar | MAHAY (43) | 278 | 311 | 30.2 | 0.73 (0.53, 0.99) | 0.06 | 0.11 |
| Malawi | iLiNS−DYADM (44) | 117 | 225 | 20.9 | 1.06 (0.70, 1.63) | 0.03 | 0.08 |
| Malawi | iLiNS−DOSE (45) | 86 | 32 | 53.1 | 0.55 (0.35, 0.87) | 0.03 | 0.08 |
| Mali | PROMIS CS (46) | 505 | 463 | 56.8 | 0.67 (0.56, 0.80) | 0.18 | 0.14 |
| Zimbabwe | SHINE (HIV−) (47) | 821 | 722 | 12.2 | 0.70 (0.53, 0.92) | 0.07 | 0.12 |
| Zimbabwe | SHINE (HIV+) (48) | 155 | 138 | 13.0 | 0.35 (0.15, 0.80) | 0.01 | 0.03 |
| | | **4094** | **3138** | | **I² = 0.46, Tau² = 0.04** | | |
| **Fixed** | | | | | **0.72 (0.67, 0.78)** | | |
| **Random** | | | | | **0.71 (0.60, 0.85)** | | |

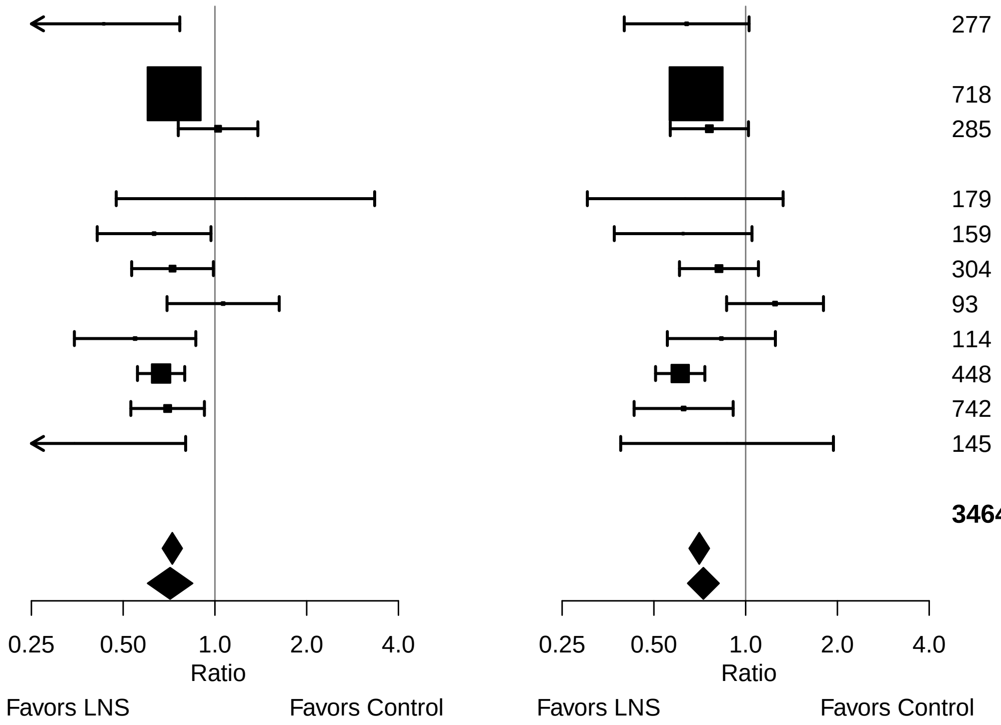

**Less than median**

| Country | Trial | LNS N | Control N | Control Prevalence | PR (95% CI) | Fixed W | Random W |
|---|---|---|---|---|---|---|---|
| Bangladesh | JiVitA−4 (35) | | | | | | |
| Bangladesh | RDNS (36) | 277 | 136 | 16.9 | 0.64 (0.40, 1.03) | 0.03 | 0.05 |
| Bangladesh | WASH−B (37) | | | | | | |
| Burkina Faso | iLiNS−Zinc (38) | 718 | 364 | 76.1 | 0.69 (0.62, 0.77) | 0.51 | 0.25 |
| Burkina Faso | PROMIS CS (39) | 285 | 288 | 39.2 | 0.76 (0.57, 1.02) | 0.07 | 0.11 |
| Ghana | GHANA (40) | | | | | | |
| Ghana | iLiNS−DYADG (41) | 179 | 315 | 7.9 | 0.63 (0.30, 1.33) | 0.01 | 0.02 |
| Kenya | WASH−B (42) | 159 | 125 | 23.2 | 0.62 (0.37, 1.05) | 0.02 | 0.04 |
| Madagascar | MAHAY (43) | 304 | 262 | 37.0 | 0.82 (0.61, 1.10) | 0.07 | 0.11 |
| Malawi | iLiNS−DYADM (44) | 93 | 206 | 26.7 | 1.25 (0.87, 1.80) | 0.04 | 0.08 |
| Malawi | iLiNS−DOSE (45) | 114 | 39 | 41.0 | 0.83 (0.55, 1.25) | 0.03 | 0.07 |
| Mali | PROMIS CS (46) | 448 | 507 | 63.3 | 0.61 (0.51, 0.73) | 0.17 | 0.18 |
| Zimbabwe | SHINE (HIV−) (47) | 742 | 760 | 9.5 | 0.63 (0.43, 0.91) | 0.04 | 0.08 |
| Zimbabwe | SHINE (HIV+) (48) | 145 | 140 | 7.1 | 0.87 (0.39, 1.94) | 0.01 | 0.02 |
| | | **3464** | **3142** | | **I² = 0.33, Tau² = 0.01** | | |
| **Fixed** | | | | | **0.70 (0.65, 0.76)** | | |
| **Random** | | | | | **0.73 (0.65, 0.82)** | | |

**Supplemental figure 7D: Moderate-to-severe anemia prevalence ratio**

#### 7D2: Stratified by Household food insecurity

**P−for−interaction = 0.938**
**Ratio of PRs = 1.00 (0.89, 1.11)**

| Country | Trial | LNS N | Control N | Control Prevalence | Mild to secure PR (95% CI) | Fixed W | Random W | LNS N | Control N | Control Prevalence | Moderate to severe PR (95% CI) | Fixed W | Random W |
|---|---|---|---|---|---|---|---|---|---|---|---|---|---|
| Bangladesh | JiVitA−4 (35) | | | | | | | | | | | | |
| Bangladesh | RDNS (36) | 342 | 166 | 13.3 | 0.71 (0.44, 1.14) | 0.02 | 0.02 | 207 | 106 | 21.7 | 0.38 (0.24, 0.61) | 0.05 | 0.13 |
| Bangladesh | WASH−B (37) | | | | | | | | | | | | |
| Burkina Faso | iLiNS−Zinc (38) | 1016 | 320 | 77.2 | 0.72 (0.66, 0.79) | 0.75 | 0.75 | 934 | 343 | 72.9 | 0.70 (0.62, 0.80) | 0.64 | 0.19 |
| Burkina Faso | PROMIS CS (39) | | | | | | | | | | | | |
| Ghana | GHANA (40) | | | | | | | | | | | | |
| Ghana | iLiNS−DYADG (41) | | | | | | | | | | | | |
| Kenya | WASH−B (42) | 304 | 273 | 21.2 | 0.63 (0.44, 0.92) | 0.04 | 0.04 | 45 | 27 | 22.2 | 0.60 (0.26, 1.36) | 0.02 | 0.08 |
| Madagascar | MAHAY (43) | 359 | 340 | 38.2 | 0.71 (0.54, 0.94) | 0.07 | 0.07 | 137 | 140 | 32.1 | 1.04 (0.74, 1.48) | 0.09 | 0.16 |
| Malawi | iLiNS−DYADM (44) | 66 | 113 | 28.3 | 0.80 (0.47, 1.37) | 0.02 | 0.02 | 143 | 314 | 22.0 | 1.34 (0.96, 1.86) | 0.10 | 0.16 |
| Malawi | iLiNS−DOSE (45) | 50 | 21 | 47.6 | 0.62 (0.34, 1.13) | 0.02 | 0.02 | 144 | 46 | 43.5 | 0.80 (0.55, 1.16) | 0.07 | 0.15 |
| Mali | PROMIS CS (46) | | | | | | | | | | | | |
| Zimbabwe | SHINE (HIV−) (47) | 1253 | 1162 | 10.2 | 0.76 (0.58, 1.00) | 0.08 | 0.08 | 264 | 293 | 12.6 | 0.51 (0.30, 0.87) | 0.04 | 0.12 |
| Zimbabwe | SHINE (HIV+) (48) | | | | | | | | | | | | |
| | | **3390** | **2395** | | **I² = 0.00, Tau² = 0.00** | | | **1874** | **1269** | | **I² = 0.77, Tau² = 0.14** | | |
| **Fixed** | | | | | **0.72 (0.67, 0.78)** | | | | | | **0.75 (0.68, 0.83)** | | |
| **Random** | | | | | **0.72 (0.67, 0.78)** | | | | | | **0.74 (0.54, 1.01)** | | |

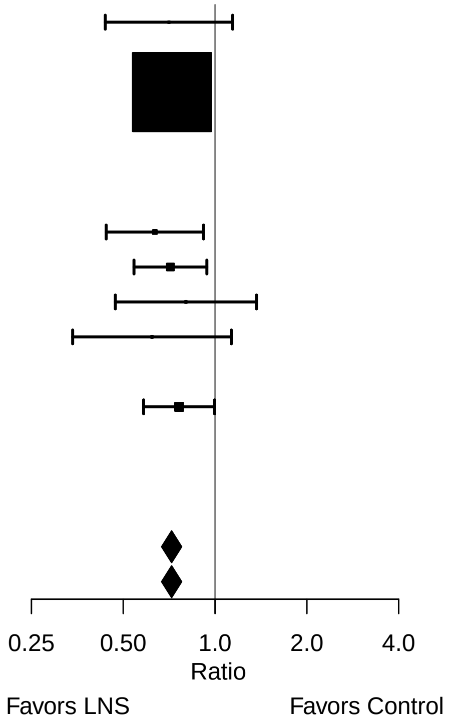

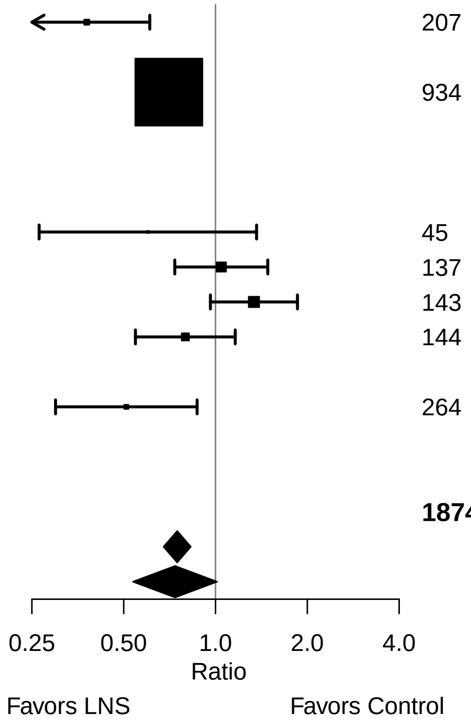

**Supplemental figure 7D: Moderate-to-severe anemia prevalence ratio**

#### 7D3: Stratified by Household source water quality

**P−for−interaction = 0.952**
**Ratio of PRs = 1.00 (0.89, 1.14)**

| Country | Trial | Improved LNS N | Improved Control N | Improved Control Prevalence | Improved PR (95% CI) | Improved Fixed W | Improved Random W | Unimproved LNS N | Unimproved Control N | Unimproved Control Prevalence | Unimproved PR (95% CI) | Unimproved Fixed W | Unimproved Random W |
|---|---|---|---|---|---|---|---|---|---|---|---|---|---|
| Bangladesh | JiVitA−4 (35) | | | | | | | | | | | | |
| Bangladesh | RDNS (36) | | | | | | | | | | | | |
| Bangladesh | WASH−B (37) | | | | | | | | | | | | |
| Burkina Faso | iLiNS−Zinc (38) | 555 | 141 | 75.9 | 0.66 (0.54, 0.80) | 0.24 | 0.19 | 1394 | 519 | 74.6 | 0.74 (0.68, 0.81) | 0.72 | 0.72 |
| Burkina Faso | PROMIS CS (39) | 373 | 363 | 35.8 | 0.94 (0.77, 1.16) | 0.24 | 0.19 | 201 | 217 | 37.8 | 0.79 (0.57, 1.09) | 0.05 | 0.05 |
| Ghana | GHANA (40) | | | | | | | | | | | | |
| Ghana | iLiNS−DYADG (41) | | | | | | | | | | | | |
| Kenya | WASH−B (42) | | | | | | | | | | | | |
| Madagascar | MAHAY (43) | 137 | 164 | 31.1 | 1.01 (0.72, 1.41) | 0.09 | 0.15 | 433 | 408 | 34.3 | 0.73 (0.55, 0.97) | 0.06 | 0.06 |
| Malawi | iLiNS−DYADM (44) | 191 | 397 | 23.2 | 1.17 (0.88, 1.57) | 0.12 | 0.16 | 19 | 33 | 30.3 | 0.87 (0.35, 2.16) | 0.01 | 0.01 |
| Malawi | iLiNS−DOSE (45) | | | | | | | | | | | | |
| Mali | PROMIS CS (46) | 548 | 567 | 59.8 | 0.62 (0.51, 0.76) | 0.25 | 0.19 | 364 | 388 | 60.8 | 0.67 (0.56, 0.81) | 0.15 | 0.15 |
| Zimbabwe | SHINE (HIV−) (47) | 469 | 436 | 9.4 | 0.70 (0.48, 1.04) | 0.07 | 0.13 | 259 | 281 | 10.3 | 0.41 (0.22, 0.76) | 0.01 | 0.01 |
| Zimbabwe | SHINE (HIV+) (48) | | | | | | | | | | | | |
| | | **2273** | **2068** | | **I² = 0.76, Tau² = 0.05** | | | **2670** | **1846** | | **I² = 0.00, Tau² = 0.00** | | |
| **Fixed** | | | | | **0.79 (0.71, 0.87)** | | | | | | **0.73 (0.68, 0.78)** | | |
| **Random** | | | | | **0.82 (0.66, 1.01)** | | | | | | **0.73 (0.68, 0.78)** | | |

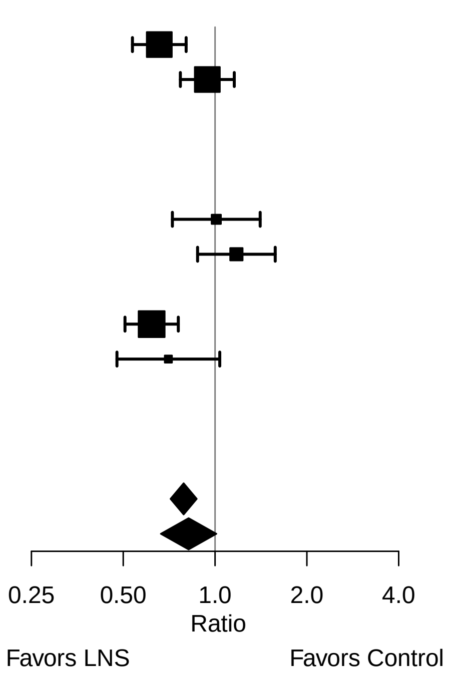

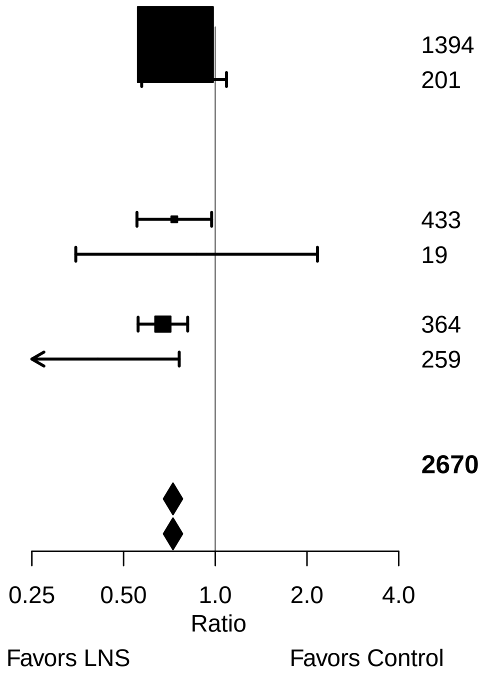

#### Supplemental figure 7D: Moderate-to-severe anemia prevalence ratio

#### 7D4: Stratified by Household sanitation

**P−for−interaction = 0.004**
**Ratio of PRs = 0.74 (0.60, 0.91)**

| Country | Trial | Improved: LNS N | Improved: Control N | Improved: Control Prevalence | Improved: PR (95% CI) | Improved: Fixed W | Improved: Random W | Unimproved: LNS N | Unimproved: Control N | Unimproved: Control Prevalence | Unimproved: PR (95% CI) | Unimproved: Fixed W | Unimproved: Random W |
|---|---|---|---|---|---|---|---|---|---|---|---|---|---|
| Bangladesh | JiVitA−4 (35) | | | | | | | | | | | | |
| Bangladesh | RDNS (36) | 376 | 193 | 13.5 | 0.71 (0.44, 1.15) | 0.08 | 0.12 | 173 | 79 | 24.1 | 0.31 (0.17, 0.56) | 0.08 | 0.19 |
| Bangladesh | WASH−B (37) | | | | | | | | | | | | |
| Burkina Faso | iLiNS−Zinc (38) | | | | | | | | | | | | |
| Burkina Faso | PROMIS CS (39) | 244 | 229 | 34.1 | 0.97 (0.73, 1.30) | 0.21 | 0.27 | 329 | 350 | 38.3 | 0.83 (0.65, 1.07) | 0.43 | 0.29 |
| Ghana | GHANA (40) | | | | | | | | | | | | |
| Ghana | iLiNS−DYADG (41) | | | | | | | | | | | | |
| Kenya | WASH−B (42) | | | | | | | | | | | | |
| Madagascar | MAHAY (43) | | | | | | | | | | | | |
| Malawi | iLiNS−DYADM (44) | | | | | | | | | | | | |
| Malawi | iLiNS−DOSE (45) | | | | | | | | | | | | |
| Mali | PROMIS CS (46) | 703 | 696 | 59.6 | 0.69 (0.58, 0.81) | 0.64 | 0.50 | 219 | 249 | 59.4 | 0.48 (0.37, 0.64) | 0.36 | 0.29 |
| Zimbabwe | SHINE (HIV−) (47) | 262 | 218 | 11.0 | 0.66 (0.40, 1.07) | 0.07 | 0.11 | 465 | 498 | 9.2 | 0.54 (0.34, 0.83) | 0.14 | 0.23 |
| Zimbabwe | SHINE (HIV+) (48) | | | | | | | | | | | | |
| | | **1585** | **1336** | | **I² = 0.35, Tau² = 0.01** | | | **1186** | **1176** | | **I² = 0.79, Tau² = 0.11** | | |
| **Fixed** | | | | | **0.74 (0.65, 0.84)** | | | | | | **0.60 (0.51, 0.71)** | | |
| **Random** | | | | | **0.75 (0.63, 0.90)** | | | | | | **0.54 (0.36, 0.79)** | | |

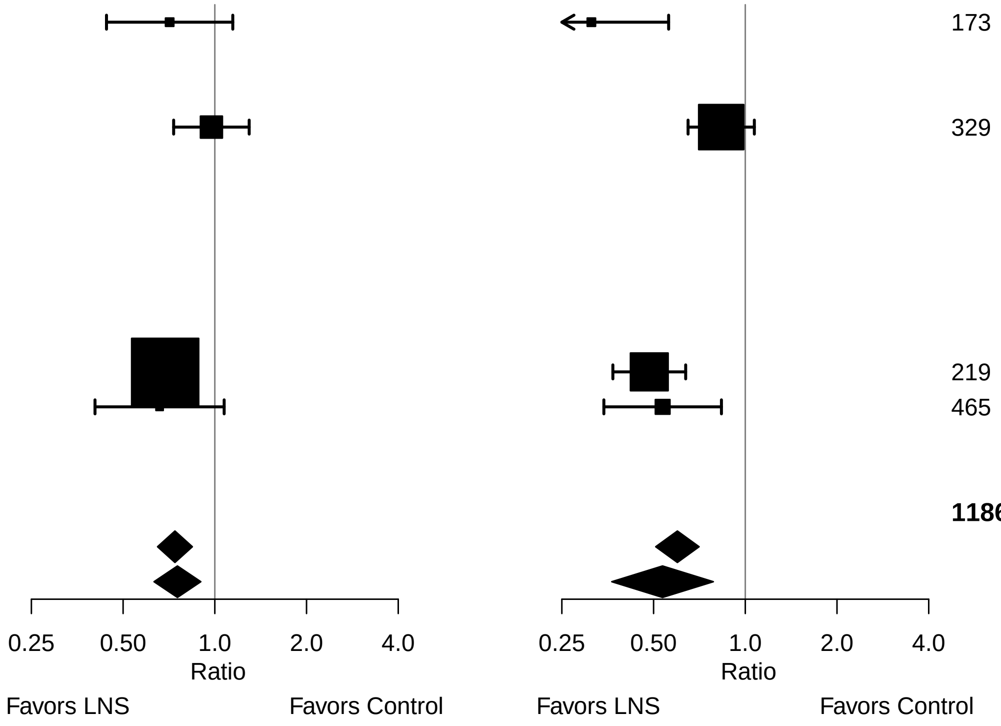

##### Supplemental figure 7D: Moderate-to-severe anemia prevalence ratio

#### 7D5: Stratified by Season at the time of assessment

**P−for−interaction = 0.189**
**Ratio of PRs = 1.08 (0.96, 1.20)**

| Country | Trial | Dry LNS N | Dry Control N | Dry Control Prevalence | Dry PR (95% CI) | Dry Fixed W | Dry Random W | Rainy LNS N | Rainy Control N | Rainy Control Prevalence | Rainy PR (95% CI) | Rainy Fixed W | Rainy Random W |
|---|---|---|---|---|---|---|---|---|---|---|---|---|---|
| Bangladesh | JiVitA−4 (35) | | | | | | | | | | | | |
| Bangladesh | RDNS (36) | 341 | 166 | 20.5 | 0.44 (0.29, 0.68) | 0.03 | 0.12 | 208 | 106 | 10.4 | 0.83 (0.38, 1.83) | 0.02 | 0.04 |
| Bangladesh | WASH−B (37) | | | | | | | | | | | | |
| Burkina Faso | iLiNS−Zinc (38) | 1398 | 475 | 75.8 | 0.70 (0.64, 0.77) | 0.68 | 0.16 | 559 | 189 | 73.0 | 0.75 (0.67, 0.86) | 0.59 | 0.30 |
| Burkina Faso | PROMIS CS (39) | 283 | 245 | 38.8 | 0.79 (0.58, 1.08) | 0.06 | 0.14 | 291 | 336 | 35.1 | 0.97 (0.77, 1.22) | 0.18 | 0.21 |
| Ghana | GHANA (40) | 57 | 55 | 34.5 | 0.25 (0.10, 0.63) | 0.01 | 0.06 | 41 | 41 | 29.3 | 0.42 (0.16, 1.08) | 0.01 | 0.03 |
| Ghana | iLiNS−DYADG (41) | 198 | 386 | 5.4 | 0.93 (0.45, 1.93) | 0.01 | 0.07 | 130 | 274 | 5.5 | 0.70 (0.26, 1.89) | 0.01 | 0.03 |
| Kenya | WASH−B (42) | 100 | 88 | 15.9 | 0.44 (0.20, 0.98) | 0.01 | 0.07 | 250 | 212 | 23.6 | 0.68 (0.48, 0.96) | 0.08 | 0.14 |
| Madagascar | MAHAY (43) | 517 | 537 | 34.6 | 0.85 (0.68, 1.07) | 0.12 | 0.15 | 83 | 51 | 13.7 | 0.61 (0.21, 1.79) | 0.01 | 0.02 |
| Malawi | iLiNS−DYADM (44) | 147 | 305 | 22.0 | 1.11 (0.78, 1.59) | 0.05 | 0.13 | 63 | 125 | 27.2 | 1.23 (0.78, 1.93) | 0.05 | 0.10 |
| Malawi | iLiNS−DOSE (45) | 123 | 32 | 40.6 | 0.96 (0.61, 1.51) | 0.03 | 0.11 | 120 | 50 | 50.0 | 0.58 (0.40, 0.84) | 0.07 | 0.13 |
| Mali | PROMIS CS (46) | | | | | | | | | | | | |
| Zimbabwe | SHINE (HIV−) (47) | | | | | | | | | | | | |
| Zimbabwe | SHINE (HIV+) (48) | | | | | | | | | | | | |
| | | **3164** | **2289** | | **I² = 0.64, Tau² = 0.11** | | | **1745** | **1384** | | **I² = 0.36, Tau² = 0.02** | | |
| **Fixed** | | | | | **0.73 (0.67, 0.79)** | | | | | | **0.78 (0.71, 0.86)** | | |
| **Random** | | | | | **0.71 (0.55, 0.93)** | | | | | | **0.78 (0.66, 0.92)** | | |

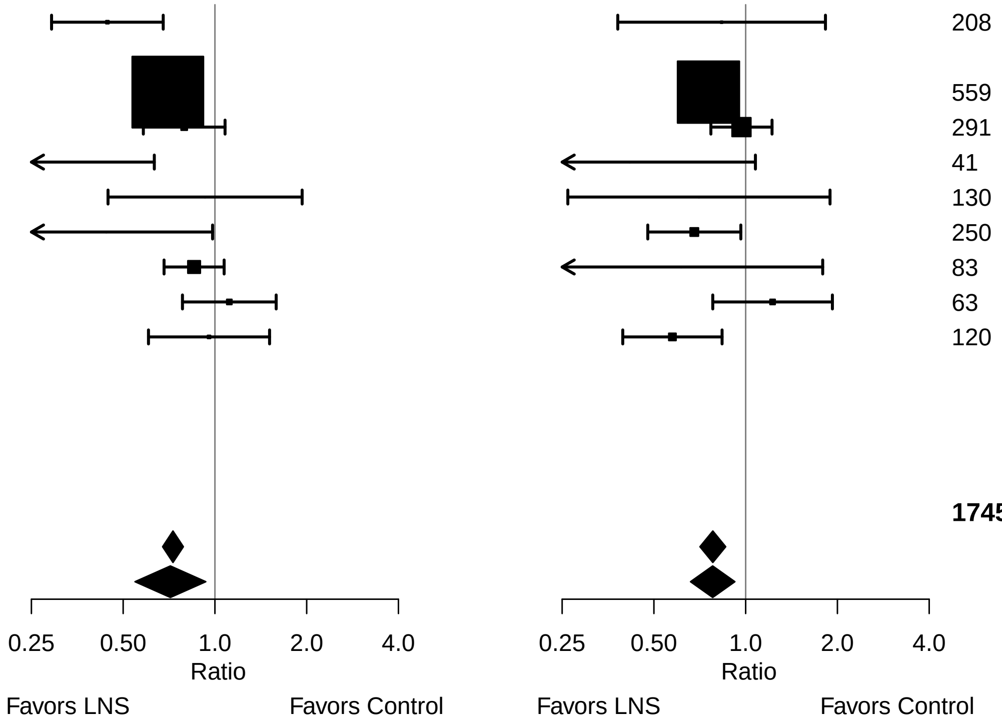

### Supplemental figure 7E: Moderate-to-severe anemia prevalence difference

#### 7E1: Stratified by Household socio-economic status

**P−for−interaction = 0.490**
**Difference in PDs = −0.01 (−0.03, 0.02)**

At least median:

| Country | Trial | LNS N | Control N | Control Prevalence | PD (95% CI) | Fixed W | Random W |
|---|---|---|---|---|---|---|---|
| Bangladesh | JiVitA−4 (35) | | | | | | |
| Bangladesh | RDNS (36) | 272 | 136 | 16.2 | −0.09 (−0.16, −0.02) | 0.06 | 0.10 |
| Bangladesh | WASH−B (37) | | | | | | |
| Burkina Faso | iLiNS−Zinc (38) | 1232 | 299 | 73.6 | −0.20 (−0.26, −0.13) | 0.06 | 0.10 |
| Burkina Faso | PROMIS CS (39) | 289 | 293 | 34.1 | 0.01 (−0.10, 0.11) | 0.03 | 0.08 |
| Ghana | GHANA (40) | | | | | | |
| Ghana | iLiNS−DYADG (41) | 149 | 344 | 3.2 | 0.01 (−0.03, 0.04) | 0.24 | 0.11 |
| Kenya | WASH−B (42) | 190 | 175 | 20.0 | −0.07 (−0.14, 0.00) | 0.06 | 0.10 |
| Madagascar | MAHAY (43) | 278 | 311 | 30.2 | −0.08 (−0.16, 0.00) | 0.04 | 0.09 |
| Malawi | iLiNS−DYADM (44) | 117 | 225 | 20.9 | 0.01 (−0.08, 0.11) | 0.04 | 0.08 |
| Malawi | iLiNS−DOSE (45) | 86 | 32 | 53.1 | −0.24 (−0.43, −0.05) | 0.01 | 0.04 |
| Mali | PROMIS CS (46) | 505 | 463 | 56.8 | −0.19 (−0.27, −0.11) | 0.05 | 0.09 |
| Zimbabwe | SHINE (HIV−) (47) | 821 | 722 | 12.2 | −0.04 (−0.07, −0.01) | 0.36 | 0.12 |
| Zimbabwe | SHINE (HIV+) (48) | 155 | 138 | 13.0 | −0.09 (−0.16, −0.01) | 0.05 | 0.09 |
| | | **4094** | **3138** | | **I² = 0.80, Tau² = 0.00** | | |
| **Fixed** | | | | | **−0.05 (−0.07, −0.04)** | | |
| **Random** | | | | | **−0.08 (−0.13, −0.03)** | | |

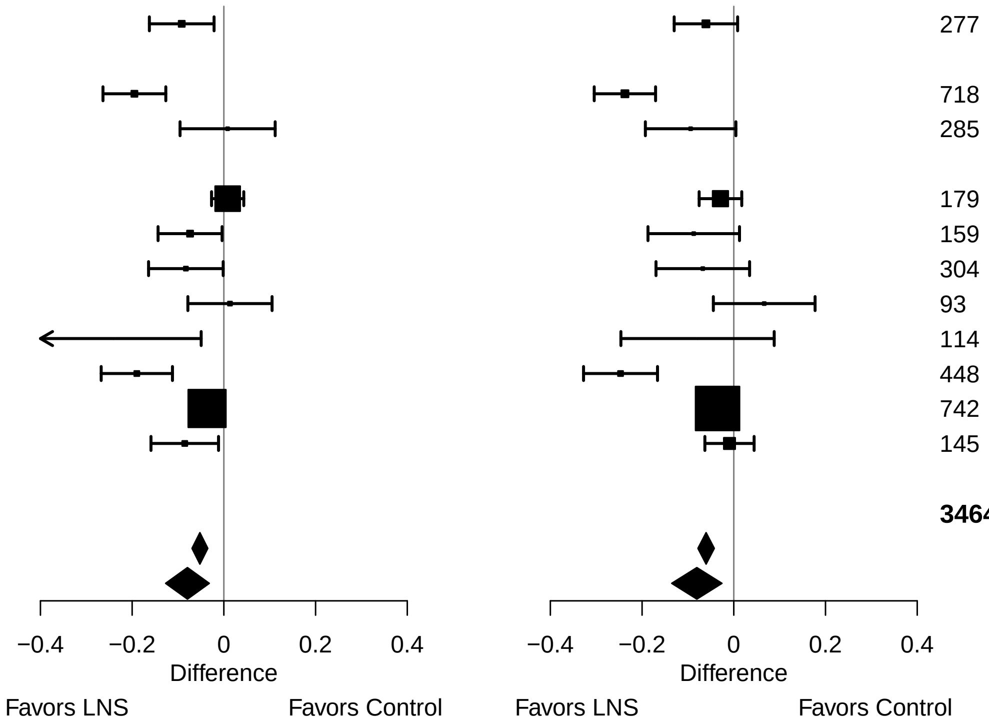

Less than median:

| Country | Trial | LNS N | Control N | Control Prevalence | PD (95% CI) | Fixed W | Random W |
|---|---|---|---|---|---|---|---|
| Bangladesh | JiVitA−4 (35) | | | | | | |
| Bangladesh | RDNS (36) | 277 | 136 | 16.9 | −0.06 (−0.13, 0.01) | 0.07 | 0.10 |
| Bangladesh | WASH−B (37) | | | | | | |
| Burkina Faso | iLiNS−Zinc (38) | 718 | 364 | 76.1 | −0.24 (−0.30, −0.17) | 0.07 | 0.10 |
| Burkina Faso | PROMIS CS (39) | 285 | 288 | 39.2 | −0.09 (−0.19, 0.00) | 0.03 | 0.08 |
| Ghana | GHANA (40) | | | | | | |
| Ghana | iLiNS−DYADG (41) | 179 | 315 | 7.9 | −0.03 (−0.08, 0.02) | 0.15 | 0.11 |
| Kenya | WASH−B (42) | 159 | 125 | 23.2 | −0.09 (−0.19, 0.01) | 0.03 | 0.08 |
| Madagascar | MAHAY (43) | 304 | 262 | 37.0 | −0.07 (−0.17, 0.03) | 0.03 | 0.08 |
| Malawi | iLiNS−DYADM (44) | 93 | 206 | 26.7 | 0.07 (−0.04, 0.18) | 0.03 | 0.08 |
| Malawi | iLiNS−DOSE (45) | 114 | 39 | 41.0 | −0.08 (−0.25, 0.09) | 0.01 | 0.06 |
| Mali | PROMIS CS (46) | 448 | 507 | 63.3 | −0.25 (−0.33, −0.17) | 0.05 | 0.09 |
| Zimbabwe | SHINE (HIV−) (47) | 742 | 760 | 9.5 | −0.04 (−0.06, −0.01) | 0.42 | 0.11 |
| Zimbabwe | SHINE (HIV+) (48) | 145 | 140 | 7.1 | −0.01 (−0.06, 0.04) | 0.11 | 0.10 |
| | | **3464** | **3142** | | **I² = 0.84, Tau² = 0.01** | | |
| **Fixed** | | | | | **−0.06 (−0.08, −0.04)** | | |
| **Random** | | | | | **−0.08 (−0.14, −0.03)** | | |

#### Supplemental figure 7E: Moderate-to-severe anemia prevalence difference

##### 7E2: Stratified by Household food insecurity

**P−for−interaction = 0.523**
**Difference in PDs = −0.01 (−0.05, 0.02)**

| Country | Trial | Mild to secure: LNS N | Control N | Control Prevalence | PD (95% CI) | Fixed W | Random W | Moderate to severe: LNS N | Control N | Control Prevalence | PD (95% CI) | Fixed W | Random W |
|---|---|---|---|---|---|---|---|---|---|---|---|---|---|
| Bangladesh | JiVitA−4 (35) | | | | | | | | | | | | |
| Bangladesh | RDNS (36) | 342 | 166 | 13.3 | −0.04 (−0.10, 0.02) | 0.10 | 0.18 | 207 | 106 | 21.7 | −0.13 (−0.22, −0.05) | 0.14 | 0.16 |
| Bangladesh | WASH−B (37) | | | | | | | | | | | | |
| Burkina Faso | iLiNS−Zinc (38) | 1016 | 320 | 77.2 | −0.21 (−0.27, −0.16) | 0.10 | 0.17 | 934 | 343 | 72.9 | −0.22 (−0.30, −0.14) | 0.15 | 0.16 |
| Burkina Faso | PROMIS CS (39) | | | | | | | | | | | | |
| Ghana | GHANA (40) | | | | | | | | | | | | |
| Ghana | iLiNS−DYADG (41) | | | | | | | | | | | | |
| Kenya | WASH−B (42) | 304 | 273 | 21.2 | −0.08 (−0.14, −0.02) | 0.09 | 0.17 | 45 | 27 | 22.2 | −0.09 (−0.23, 0.05) | 0.05 | 0.11 |
| Madagascar | MAHAY (43) | 359 | 340 | 38.2 | −0.11 (−0.20, −0.02) | 0.04 | 0.13 | 137 | 140 | 32.1 | 0.01 (−0.10, 0.13) | 0.07 | 0.13 |
| Malawi | iLiNS−DYADM (44) | 66 | 113 | 28.3 | −0.06 (−0.19, 0.08) | 0.02 | 0.09 | 143 | 314 | 22.0 | 0.07 (−0.01, 0.16) | 0.13 | 0.15 |
| Malawi | iLiNS−DOSE (45) | 50 | 21 | 47.6 | −0.18 (−0.43, 0.06) | 0.01 | 0.04 | 144 | 46 | 43.5 | −0.09 (−0.24, 0.06) | 0.04 | 0.11 |
| Mali | PROMIS CS (46) | | | | | | | | | | | | |
| Zimbabwe | SHINE (HIV−) (47) | 1253 | 1162 | 10.2 | −0.02 (−0.05, 0.00) | 0.64 | 0.21 | 264 | 293 | 12.6 | −0.06 (−0.11, −0.02) | 0.43 | 0.18 |
| Zimbabwe | SHINE (HIV+) (48) | | | | | | | | | | | | |
| | | **3390** | **2395** | | **I² = 0.84, Tau² = 0.00** | | | **1874** | **1269** | | **I² = 0.79, Tau² = 0.01** | | |
| **Fixed** | | | | | **−0.05 (−0.07, −0.04)** | | | | | | **−0.07 (−0.10, −0.04)** | | |
| **Random** | | | | | **−0.09 (−0.14, −0.04)** | | | | | | **−0.07 (−0.15, 0.00)** | | |

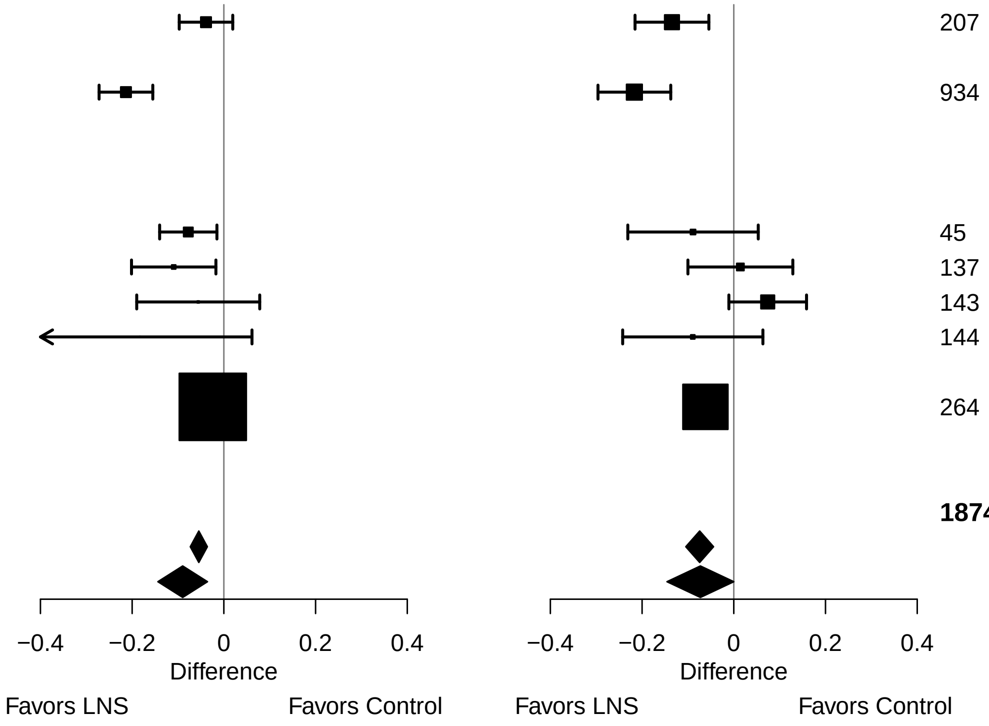

#### Supplemental figure 7E: Moderate-to-severe anemia prevalence difference

##### 7E3: Stratified by Household source water quality

**P−for−interaction = 0.407**
**Difference in PDs = −0.02 (−0.05, 0.02)**

| Country | Trial | Improved LNS N | Improved Control N | Improved Control Prevalence | Improved PD (95% CI) | Improved Fixed W | Improved Random W | Unimproved LNS N | Unimproved Control N | Unimproved Control Prevalence | Unimproved PD (95% CI) | Unimproved Fixed W | Unimproved Random W |
|---|---|---|---|---|---|---|---|---|---|---|---|---|---|
| Bangladesh | JiVitA−4 (35) | | | | | | | | | | | | |
| Bangladesh | RDNS (36) | | | | | | | | | | | | |
| Bangladesh | WASH−B (37) | | | | | | | | | | | | |
| Burkina Faso | iLiNS−Zinc (38) | 555 | 141 | 75.9 | −0.26 (−0.40, −0.12) | 0.03 | 0.14 | 1394 | 519 | 74.6 | −0.19 (−0.25, −0.14) | 0.26 | 0.23 |
| Burkina Faso | PROMIS CS (39) | 373 | 363 | 35.8 | −0.02 (−0.09, 0.05) | 0.12 | 0.17 | 201 | 217 | 37.8 | −0.08 (−0.19, 0.03) | 0.06 | 0.14 |
| Ghana | GHANA (40) | | | | | | | | | | | | |
| Ghana | iLiNS−DYADG (41) | | | | | | | | | | | | |
| Kenya | WASH−B (42) | | | | | | | | | | | | |
| Madagascar | MAHAY (43) | 137 | 164 | 31.1 | 0.00 (−0.10, 0.11) | 0.06 | 0.16 | 433 | 408 | 34.3 | −0.09 (−0.18, −0.01) | 0.10 | 0.17 |
| Malawi | iLiNS−DYADM (44) | 191 | 397 | 23.2 | 0.04 (−0.03, 0.11) | 0.11 | 0.17 | 19 | 33 | 30.3 | −0.04 (−0.30, 0.22) | 0.01 | 0.04 |
| Malawi | iLiNS−DOSE (45) | | | | | | | | | | | | |
| Mali | PROMIS CS (46) | 548 | 567 | 59.8 | −0.23 (−0.31, −0.14) | 0.08 | 0.17 | 364 | 388 | 60.8 | −0.20 (−0.29, −0.11) | 0.10 | 0.17 |
| Zimbabwe | SHINE (HIV−) (47) | 469 | 436 | 9.4 | −0.03 (−0.06, 0.00) | 0.61 | 0.19 | 259 | 281 | 10.3 | −0.06 (−0.10, −0.02) | 0.46 | 0.26 |
| Zimbabwe | SHINE (HIV+) (48) | | | | | | | | | | | | |
| | | **2273** | **2068** | | **I² = 0.85, Tau² = 0.01** | | | **2670** | **1846** | | **I² = 0.75, Tau² = 0.00** | | |
| **Fixed** | | | | | **−0.04 (−0.07, −0.02)** | | | | | | **−0.11 (−0.14, −0.09)** | | |
| **Random** | | | | | **−0.08 (−0.18, 0.02)** | | | | | | **−0.12 (−0.18, −0.07)** | | |

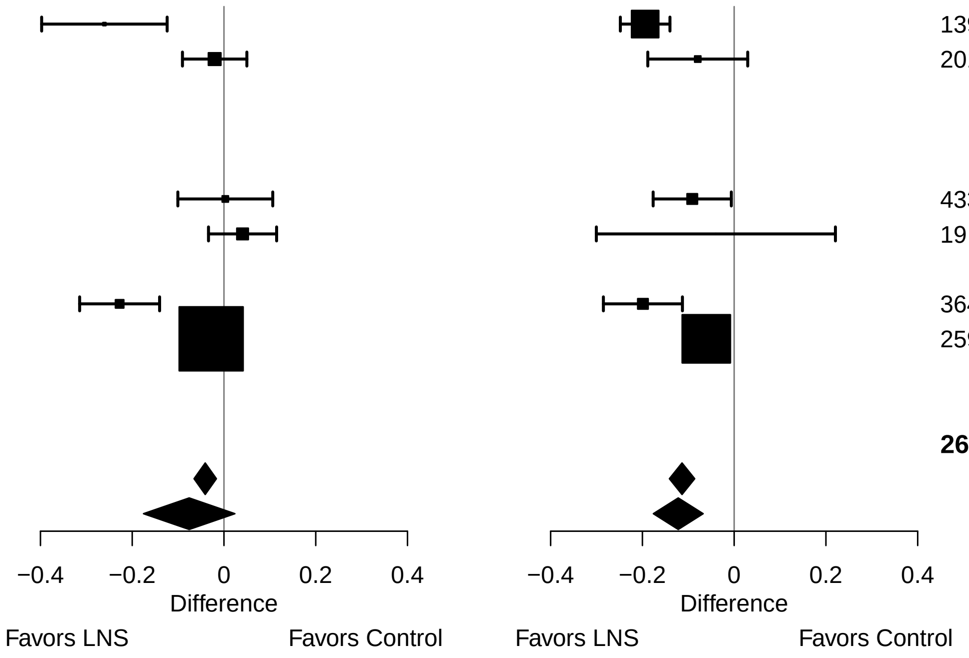

**Supplemental figure 7E: Moderate-to-severe anemia prevalence difference**

#### 7E4: Stratified by Household sanitation

**P−for−interaction = 0.020**
**Difference in PDs = −0.05 (−0.10, −0.01)**

| Country | Trial | LNS N | Control N | Control Prevalence | Improved PD (95% CI) | Fixed W | Random W | LNS N | Control N | Control Prevalence | Unimproved PD (95% CI) | Fixed W | Random W |
|---|---|---|---|---|---|---|---|---|---|---|---|---|---|
| Bangladesh | JiVitA−4 (35) | | | | | | | | | | | | |
| Bangladesh | RDNS (36) | 376 | 193 | 13.5 | −0.04 (−0.10, 0.02) | 0.27 | 0.27 | 173 | 79 | 24.1 | −0.17 (−0.25, −0.08) | 0.09 | 0.24 |
| Bangladesh | WASH−B (37) | | | | | | | | | | | | |
| Burkina Faso | iLiNS−Zinc (38) | | | | | | | | | | | | |
| Burkina Faso | PROMIS CS (39) | 244 | 229 | 34.1 | −0.01 (−0.10, 0.09) | 0.10 | 0.21 | 329 | 350 | 38.3 | −0.06 (−0.15, 0.02) | 0.09 | 0.24 |
| Ghana | GHANA (40) | | | | | | | | | | | | |
| Ghana | iLiNS−DYADG (41) | | | | | | | | | | | | |
| Kenya | WASH−B (42) | | | | | | | | | | | | |
| Madagascar | MAHAY (43) | | | | | | | | | | | | |
| Malawi | iLiNS−DYADM (44) | | | | | | | | | | | | |
| Malawi | iLiNS−DOSE (45) | | | | | | | | | | | | |
| Mali | PROMIS CS (46) | 703 | 696 | 59.6 | −0.19 (−0.26, −0.11) | 0.16 | 0.24 | 219 | 249 | 59.4 | −0.31 (−0.40, −0.21) | 0.08 | 0.24 |
| Zimbabwe | SHINE (HIV−) (47) | 262 | 218 | 11.0 | −0.04 (−0.08, 0.01) | 0.47 | 0.28 | 465 | 498 | 9.2 | −0.04 (−0.07, −0.01) | 0.74 | 0.27 |
| Zimbabwe | SHINE (HIV+) (48) | | | | | | | | | | | | |
| | | **1585** | **1336** | | **$I^2$ = 0.77, $Tau^2$ = 0.00** | | | **1186** | **1176** | | **$I^2$ = 0.91, $Tau^2$ = 0.01** | | |
| **Fixed** | | | | | **−0.06 (−0.09, −0.03)** | | | | | | **−0.08 (−0.10, −0.05)** | | |
| **Random** | | | | | **−0.07 (−0.14, 0.01)** | | | | | | **−0.14 (−0.26, −0.02)** | | |

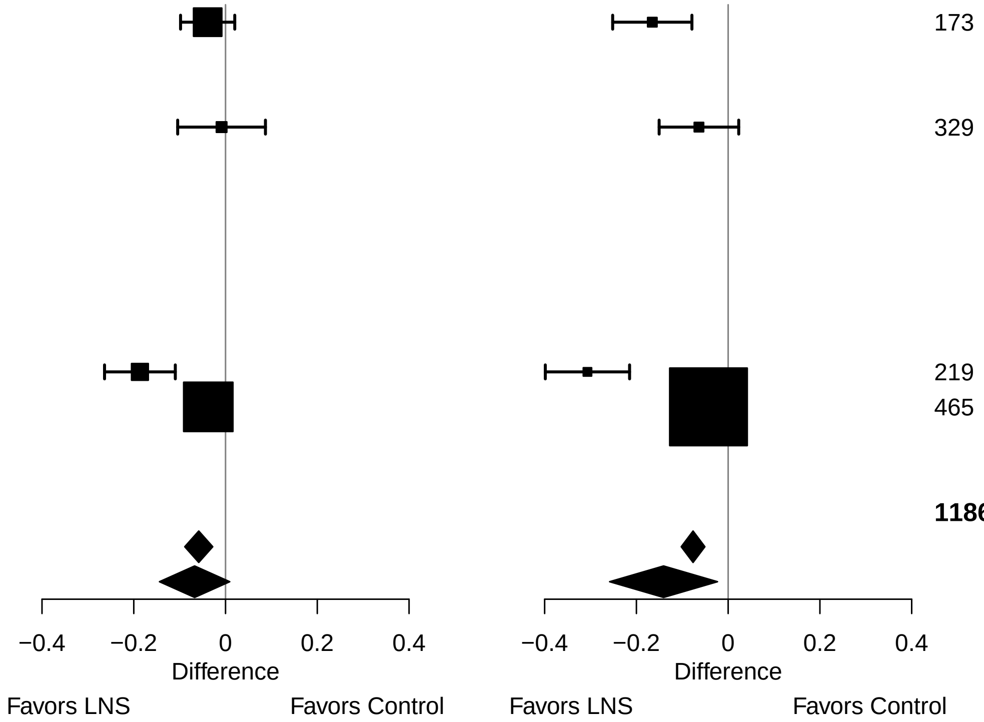

#### Supplemental figure 7E: Moderate-to-severe anemia prevalence difference

##### 7E5: Stratified by Season at the time of assessment

**P−for−interaction = 0.210**
**Difference in PDs = 0.02 (−0.01, 0.06)**

| | | Dry | | | | | | Rainy | | | | | |
|---|---|---|---|---|---|---|---|---|---|---|---|---|---|
| Country | Trial | LNS N | Control N | Control Prevalence | PD (95% CI) | Fixed W | Random W | LNS N | Control N | Control Prevalence | PD (95% CI) | Fixed W | Random W |
| Bangladesh | JiVitA−4 (35) | | | | | | | | | | | | |
| Bangladesh | RDNS (36) | 341 | 166 | 20.5 | −0.11 (−0.19, −0.04) | 0.11 | 0.12 | 208 | 106 | 10.4 | −0.02 (−0.10, 0.06) | 0.12 | 0.13 |
| Bangladesh | WASH−B (37) | | | | | | | | | | | | |
| Burkina Faso | iLiNS−Zinc (38) | 1398 | 475 | 75.8 | −0.23 (−0.29, −0.17) | 0.16 | 0.13 | 559 | 189 | 73.0 | −0.18 (−0.26, −0.09) | 0.10 | 0.13 |
| Burkina Faso | PROMIS CS (39) | 283 | 245 | 38.8 | −0.08 (−0.18, 0.02) | 0.06 | 0.11 | 291 | 336 | 35.1 | −0.01 (−0.09, 0.07) | 0.12 | 0.13 |
| Ghana | GHANA (40) | 57 | 55 | 34.5 | −0.26 (−0.40, −0.11) | 0.03 | 0.08 | 41 | 41 | 29.3 | −0.17 (−0.34, 0.00) | 0.03 | 0.07 |
| Ghana | iLiNS−DYADG (41) | 198 | 386 | 5.4 | 0.00 (−0.04, 0.03) | 0.38 | 0.14 | 130 | 274 | 5.5 | −0.02 (−0.06, 0.03) | 0.37 | 0.16 |
| Kenya | WASH−B (42) | 100 | 88 | 15.9 | −0.09 (−0.18, 0.00) | 0.07 | 0.11 | 250 | 212 | 23.6 | −0.08 (−0.15, 0.00) | 0.15 | 0.14 |
| Madagascar | MAHAY (43) | 517 | 537 | 34.6 | −0.05 (−0.12, 0.02) | 0.10 | 0.12 | 83 | 51 | 13.7 | −0.05 (−0.18, 0.08) | 0.04 | 0.09 |
| Malawi | iLiNS−DYADM (44) | 147 | 305 | 22.0 | 0.03 (−0.06, 0.11) | 0.08 | 0.12 | 63 | 125 | 27.2 | 0.06 (−0.08, 0.20) | 0.04 | 0.09 |
| Malawi | iLiNS−DOSE (45) | 123 | 32 | 40.6 | −0.02 (−0.21, 0.17) | 0.02 | 0.06 | 120 | 50 | 50.0 | −0.22 (−0.37, −0.06) | 0.03 | 0.08 |
| Mali | PROMIS CS (46) | | | | | | | | | | | | |
| Zimbabwe | SHINE (HIV−) (47) | | | | | | | | | | | | |
| Zimbabwe | SHINE (HIV+) (48) | | | | | | | | | | | | |
| | | **3164** | **2289** | | **I² = 0.85, Tau² = 0.01** | | | **1745** | **1384** | | **I² = 0.64, Tau² = 0.00** | | |
| **Fixed** | | | | | **−0.07 (−0.09, −0.05)** | | | | | | **−0.05 (−0.08, −0.02)** | | |
| **Random** | | | | | **−0.09 (−0.15, −0.03)** | | | | | | **−0.07 (−0.12, −0.01)** | | |

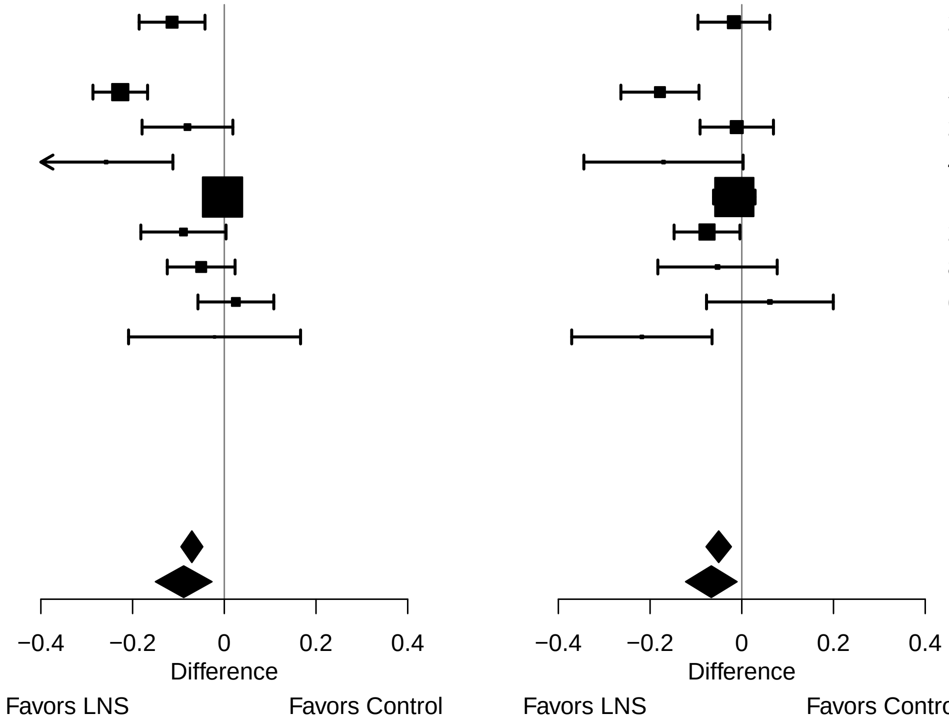

### Supplemental figure 7F: Geometric mean ratio of ferritin concentration

#### 7F1: Stratified by Household socio-economic status

**P−for−interaction = 0.204**
**Ratio of GMRs = 0.94 (0.86, 1.03)**

| | | At least median | | | | | | Less than median | | | | | |
|---|---|---|---|---|---|---|---|---|---|---|---|---|---|
| | | **LNS** | **Control** | **Control** | **GMR** | **Fixed** | **Random** | **LNS** | **Control** | **Control** | **GMR** | **Fixed** | **Random** |
| Country | Trial | N | N | Median | (95% CI) | W | W | N | N | Median | (95% CI) | W | W |
| Bangladesh | JiVitA−4 (35) | 243 | 78 | 21.4 | 1.45 (1.15, 1.83) | 0.08 | 0.13 | 211 | 66 | 27.2 | 1.30 (1.06, 1.61) | 0.12 | 0.15 |
| Bangladesh | RDNS (36) | 273 | 136 | 19.0 | 1.64 (1.47, 1.83) | 0.38 | 0.24 | 277 | 136 | 22.0 | 1.40 (1.24, 1.59) | 0.33 | 0.23 |
| Bangladesh | WASH−B (37) | 105 | 91 | 15.7 | 1.73 (1.42, 2.12) | 0.11 | 0.15 | 107 | 87 | 16.2 | 1.74 (1.46, 2.09) | 0.16 | 0.17 |
| Burkina Faso | iLiNS−Zinc (38) | 180 | 39 | 10.1 | 1.42 (1.13, 1.80) | 0.08 | 0.13 | 135 | 57 | 9.6 | 1.79 (1.47, 2.17) | 0.14 | 0.16 |
| Burkina Faso | PROMIS CS (39) | | | | | | | | | | | | |
| Ghana | GHANA (40) | 46 | 39 | 8.5 | 2.72 (1.26, 5.86) | 0.01 | 0.02 | 37 | 39 | 11.2 | 2.39 (1.15, 4.97) | 0.01 | 0.02 |
| Ghana | iLiNS−DYADG (41) | | | | | | | | | | | | |
| Kenya | WASH−B (42) | 163 | 155 | 9.5 | 1.79 (1.56, 2.04) | 0.26 | 0.22 | 134 | 104 | 11.6 | 1.57 (1.33, 1.87) | 0.18 | 0.18 |
| Madagascar | MAHAY (43) | 38 | 25 | 16.1 | 1.29 (1.02, 1.64) | 0.08 | 0.12 | 43 | 25 | 11.1 | 1.38 (1.01, 1.87) | 0.06 | 0.09 |
| Malawi | iLiNS−DYADM (44) | | | | | | | | | | | | |
| Malawi | iLiNS−DOSE (45) | | | | | | | | | | | | |
| Mali | PROMIS CS (46) | | | | | | | | | | | | |
| Zimbabwe | SHINE (HIV−) (47) | | | | | | | | | | | | |
| Zimbabwe | SHINE (HIV+) (48) | | | | | | | | | | | | |
| | | **1048** | **563** | | **I² = 0.39, Tau² = 0.01** | | | **944** | **514** | | **I² = 0.43, Tau² = 0.01** | | |
| **Fixed** | | | | | **1.63 (1.52, 1.74)** | | | | | | **1.53 (1.42, 1.64)** | | |
| **Random** | | | | | **1.60 (1.44, 1.77)** | | | | | | **1.54 (1.39, 1.71)** | | |

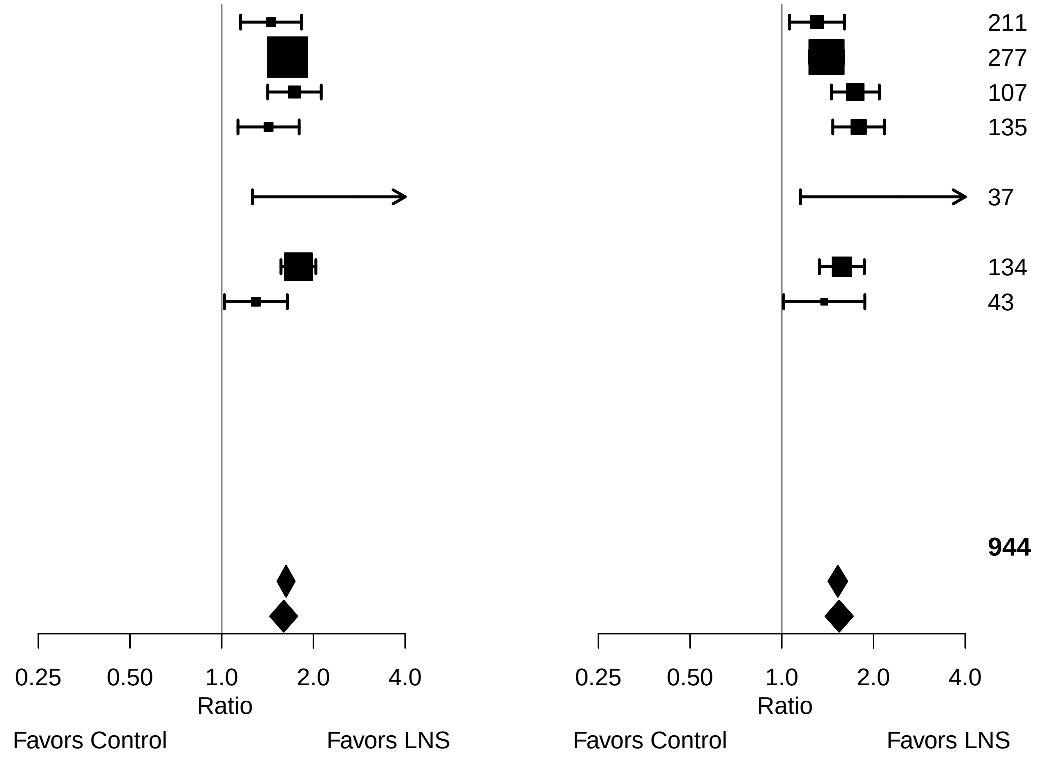

#### Supplemental figure 7F: Geometric mean ratio of ferritin concentration

#### 7F2: Stratified by Household food insecurity

**P−for−interaction = 0.245**
**Ratio of GMRs = 0.93 (0.82, 1.05)**

| | | Mild to secure | | | | | | Moderate to severe | | | | | |
|---|---|---|---|---|---|---|---|---|---|---|---|---|---|
| | | **LNS** | **Control** | **Control** | **GMR** | **Fixed** | **Random** | **LNS** | **Control** | **Control** | **GMR** | **Fixed** | **Random** |
| Country | Trial | N | N | Median | (95% CI) | W | W | N | N | Median | (95% CI) | W | W |
| Bangladesh | JiVitA−4 (35) | 337 | 111 | 22.9 | 1.43 (1.19, 1.73) | 0.13 | 0.15 | 117 | 33 | 33.0 | 1.23 (0.95, 1.58) | 0.13 | 0.13 |
| Bangladesh | RDNS (36) | 342 | 166 | 21.3 | 1.50 (1.33, 1.69) | 0.33 | 0.30 | 208 | 106 | 19.8 | 1.55 (1.36, 1.76) | 0.48 | 0.48 |
| Bangladesh | WASH−B (37) | 171 | 139 | 14.8 | 1.78 (1.49, 2.12) | 0.15 | 0.16 | 41 | 39 | 17.8 | 1.60 (1.20, 2.14) | 0.10 | 0.10 |
| Burkina Faso | iLiNS−Zinc (38) | 160 | 45 | 9.3 | 1.72 (1.31, 2.26) | 0.06 | 0.07 | 155 | 51 | 10.1 | 1.41 (1.13, 1.75) | 0.17 | 0.17 |
| Burkina Faso | PROMIS CS (39) | | | | | | | | | | | | |
| Ghana | GHANA (40) | | | | | | | | | | | | |
| Ghana | iLiNS−DYADG (41) | | | | | | | | | | | | |
| Kenya | WASH−B (42) | 259 | 235 | 10.1 | 1.71 (1.51, 1.94) | 0.29 | 0.28 | 38 | 24 | 10.3 | 1.55 (1.10, 2.18) | 0.07 | 0.07 |
| Madagascar | MAHAY (43) | 45 | 30 | 12.8 | 1.34 (0.93, 1.92) | 0.04 | 0.04 | 19 | 7 | 14.0 | 1.40 (0.98, 1.99) | 0.06 | 0.06 |
| Malawi | iLiNS−DYADM (44) | | | | | | | | | | | | |
| Malawi | iLiNS−DOSE (45) | | | | | | | | | | | | |
| Mali | PROMIS CS (46) | | | | | | | | | | | | |
| Zimbabwe | SHINE (HIV−) (47) | | | | | | | | | | | | |
| Zimbabwe | SHINE (HIV+) (48) | | | | | | | | | | | | |
| | | **1314** | **726** | | **I² = 0.18, Tau² = 0.00** | | | **578** | **260** | | **I² = 0.00, Tau² = 0.00** | | |
| **Fixed** | | | | | **1.60 (1.49, 1.71)** | | | | | | **1.47 (1.35, 1.61)** | | |
| **Random** | | | | | **1.60 (1.48, 1.73)** | | | | | | **1.47 (1.35, 1.61)** | | |

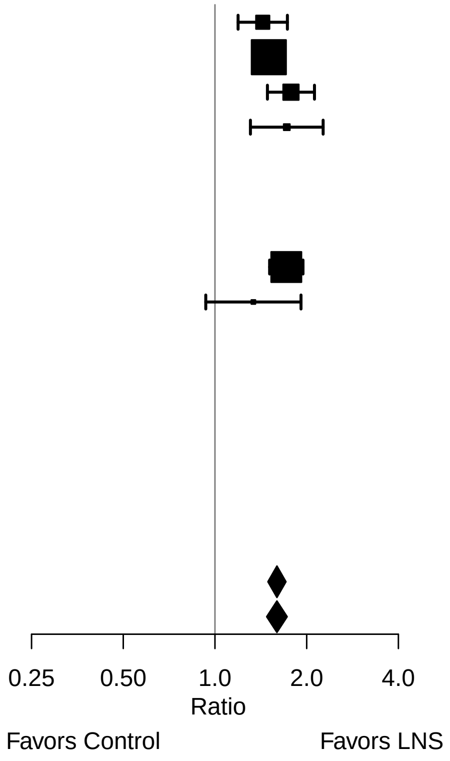

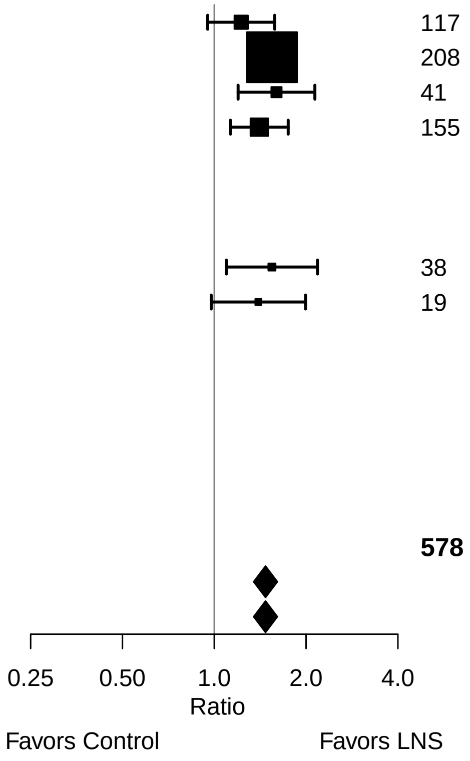

#### Supplemental figure 7F: Geometric mean ratio of ferritin concentration

#### 7F3: Stratified by Household source water quality

**P−for−interaction = 0.408**
**Ratio of GMRs = 0.93 (0.78, 1.11)**

| | | Improved | | | | | | Unimproved | | | | | |
|---|---|---|---|---|---|---|---|---|---|---|---|---|---|
| Country | Trial | LNS N | Control N | Control Median | GMR (95% CI) | Fixed W | Random W | LNS N | Control N | Control Median | GMR (95% CI) | Fixed W | Random W |
| Bangladesh | JiVitA−4 (35) | | | | | | | | | | | | |
| Bangladesh | RDNS (36) | | | | | | | | | | | | |
| Bangladesh | WASH−B (37) | 75 | 44 | 15.1 | 1.78 (1.33, 2.39) | 0.17 | 0.17 | 19 | 14 | 19.3 | 1.69 (1.06, 2.68) | 0.08 | 0.08 |
| Burkina Faso | iLiNS−Zinc (38) | 97 | 22 | 6.5 | 1.90 (1.50, 2.41) | 0.26 | 0.26 | 218 | 74 | 10.6 | 1.47 (1.20, 1.81) | 0.40 | 0.40 |
| Burkina Faso | PROMIS CS (39) | | | | | | | | | | | | |
| Ghana | GHANA (40) | | | | | | | | | | | | |
| Ghana | iLiNS−DYADG (41) | | | | | | | | | | | | |
| Kenya | WASH−B (42) | 111 | 107 | 10.9 | 1.63 (1.37, 1.94) | 0.49 | 0.49 | 55 | 37 | 9.9 | 1.84 (1.37, 2.48) | 0.19 | 0.19 |
| Madagascar | MAHAY (43) | 23 | 14 | 14.0 | 1.28 (0.83, 1.99) | 0.08 | 0.08 | 55 | 36 | 11.2 | 1.40 (1.12, 1.75) | 0.33 | 0.33 |
| Malawi | iLiNS−DYADM (44) | | | | | | | | | | | | |
| Malawi | iLiNS−DOSE (45) | | | | | | | | | | | | |
| Mali | PROMIS CS (46) | | | | | | | | | | | | |
| Zimbabwe | SHINE (HIV−) (47) | | | | | | | | | | | | |
| Zimbabwe | SHINE (HIV+) (48) | | | | | | | | | | | | |
| | | **306** | **187** | | **I² = 0.00, Tau² = 0.00** | | | **347** | **161** | | **I² = 0.00, Tau² = 0.00** | | |
| **Fixed** | | | | | **1.69 (1.50, 1.91)** | | | | | | **1.53 (1.34, 1.74)** | | |
| **Random** | | | | | **1.69 (1.50, 1.91)** | | | | | | **1.53 (1.34, 1.74)** | | |

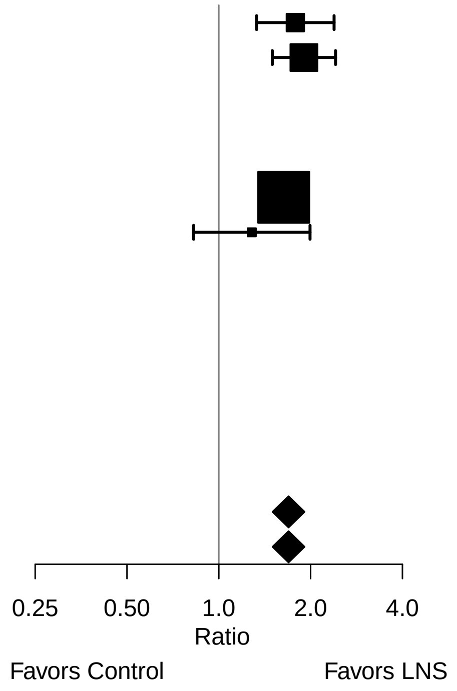

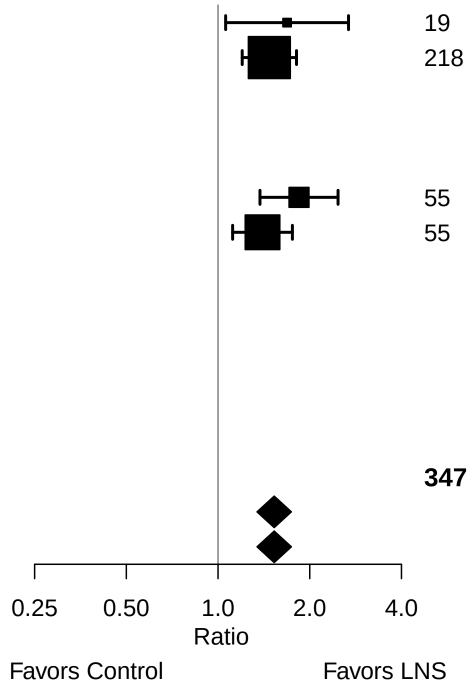

#### Supplemental figure 7F: Geometric mean ratio of ferritin concentration

##### 7F4: Stratified by Household sanitation

**P−for−interaction = 0.512**
**Ratio of GMRs = 1.05 (0.90, 1.23)**

| Country | Trial | **LNS** N | **Control** N | **Control** Median | **Improved GMR** (95% CI) | **Fixed** W | **Random** W | **LNS** N | **Control** N | **Control** Median | **Unimproved GMR** (95% CI) | **Fixed** W | **Random** W |
|---|---|---|---|---|---|---|---|---|---|---|---|---|---|
| Bangladesh | JiVitA−4 (35) | 362 | 113 | 23.2 | 1.44 (1.21, 1.72) | 0.20 | 0.20 | 92 | 31 | 30.4 | 1.19 (0.89, 1.60) | 0.15 | 0.26 |
| Bangladesh | RDNS (36) | 377 | 193 | 21.2 | 1.46 (1.33, 1.59) | 0.77 | 0.77 | 173 | 79 | 19.8 | 1.67 (1.41, 1.98) | 0.44 | 0.38 |
| Bangladesh | WASH−B (37) | | | | | | | | | | | | |
| Burkina Faso | iLiNS−Zinc (38) | | | | | | | | | | | | |
| Burkina Faso | PROMIS CS (39) | | | | | | | | | | | | |
| Ghana | GHANA (40) | | | | | | | | | | | | |
| Ghana | iLiNS−DYADG (41) | | | | | | | | | | | | |
| Kenya | WASH−B (42) | 21 | 22 | 10.5 | 1.43 (0.91, 2.22) | 0.03 | 0.03 | 145 | 122 | 10.6 | 1.71 (1.43, 2.05) | 0.40 | 0.37 |
| Madagascar | MAHAY (43) | | | | | | | | | | | | |
| Malawi | iLiNS−DYADM (44) | | | | | | | | | | | | |
| Malawi | iLiNS−DOSE (45) | | | | | | | | | | | | |
| Mali | PROMIS CS (46) | | | | | | | | | | | | |
| Zimbabwe | SHINE (HIV−) (47) | | | | | | | | | | | | |
| Zimbabwe | SHINE (HIV+) (48) | | | | | | | | | | | | |
| | | **760** | **328** | | **I² = 0.00, Tau² = 0.00** | | | **410** | **232** | | **I² = 0.57, Tau² = 0.02** | | |
| **Fixed** | | | | | **1.45 (1.34, 1.57)** | | | | | | **1.61 (1.43, 1.80)** | | |
| **Random** | | | | | **1.45 (1.34, 1.57)** | | | | | | **1.55 (1.25, 1.91)** | | |

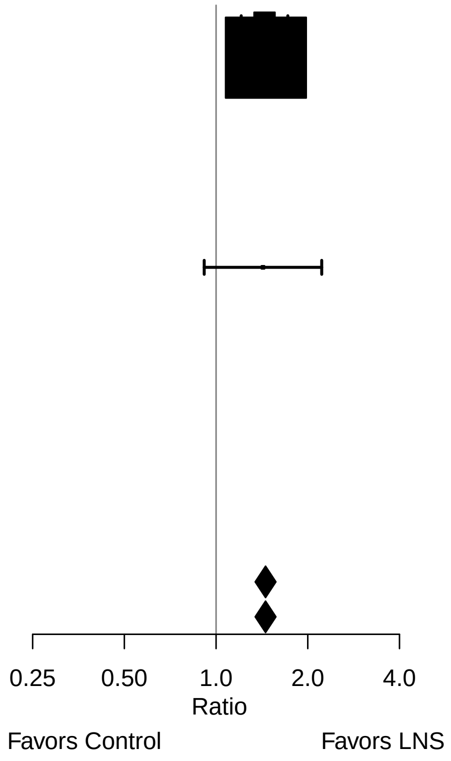

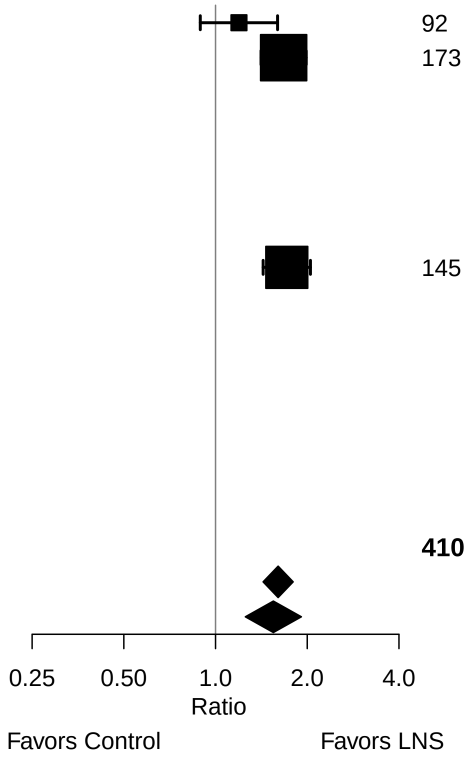

**Supplemental figure 7F: Geometric mean ratio of ferritin concentration**

#### 7F5: Stratified by Season at the time of assessment

**P−for−interaction = 0.012**
**Ratio of GMRs = 1.13 (1.03, 1.25)**

| Country | Trial | Dry LNS N | Dry Control N | Dry Control Median | Dry GMR (95% CI) | Dry Fixed W | Dry Random W | Rainy LNS N | Rainy Control N | Rainy Control Median | Rainy GMR (95% CI) | Rainy Fixed W | Rainy Random W |
|---|---|---|---|---|---|---|---|---|---|---|---|---|---|
| Bangladesh | JiVitA−4 (35) | 263 | 71 | 21.3 | 1.41 (1.17, 1.69) | 0.17 | 0.20 | 192 | 73 | 28.2 | 1.39 (1.15, 1.69) | 0.13 | 0.17 |
| Bangladesh | RDNS (36) | 341 | 166 | 21.2 | 1.39 (1.25, 1.54) | 0.52 | 0.37 | 208 | 106 | 20.5 | 1.77 (1.56, 2.00) | 0.32 | 0.25 |
| Bangladesh | WASH−B (37) | 96 | 93 | 17.7 | 1.50 (1.17, 1.91) | 0.09 | 0.13 | 116 | 85 | 13.2 | 2.03 (1.73, 2.38) | 0.20 | 0.21 |
| Burkina Faso | iLiNS−Zinc (38) | 174 | 61 | 9.2 | 1.54 (1.24, 1.92) | 0.12 | 0.16 | 126 | 30 | 10.4 | 1.51 (1.19, 1.92) | 0.09 | 0.13 |
| Burkina Faso | PROMIS CS (39) | | | | | | | | | | | | |
| Ghana | GHANA (40) | 48 | 47 | 9.1 | 3.14 (1.53, 6.42) | 0.01 | 0.02 | 35 | 35 | 11.2 | 1.88 (0.88, 4.01) | 0.01 | 0.02 |
| Ghana | iLiNS−DYADG (41) | | | | | | | | | | | | |
| Kenya | WASH−B (42) | 88 | 75 | 9.5 | 1.50 (1.17, 1.91) | 0.09 | 0.13 | 210 | 184 | 10.6 | 1.78 (1.55, 2.06) | 0.25 | 0.22 |
| Madagascar | MAHAY (43) | | | | | | | | | | | | |
| Malawi | iLiNS−DYADM (44) | | | | | | | | | | | | |
| Malawi | iLiNS−DOSE (45) | | | | | | | | | | | | |
| Mali | PROMIS CS (46) | | | | | | | | | | | | |
| Zimbabwe | SHINE (HIV−) (47) | | | | | | | | | | | | |
| Zimbabwe | SHINE (HIV+) (48) | | | | | | | | | | | | |
| | | **1010** | **513** | | **I² = 0.13, Tau² = 0.00** | | | **887** | **513** | | **I² = 0.51, Tau² = 0.01** | | |
| **Fixed** | | | | | **1.44 (1.34, 1.55)** | | | | | | **1.74 (1.62, 1.87)** | | |
| **Random** | | | | | **1.46 (1.33, 1.62)** | | | | | | **1.72 (1.54, 1.92)** | | |

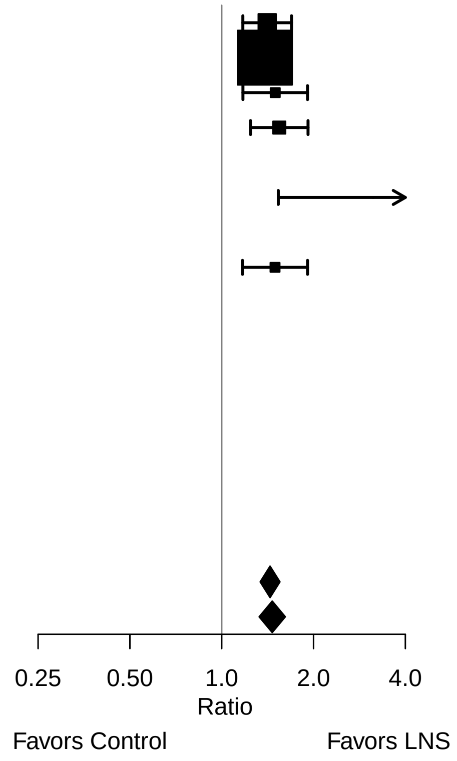

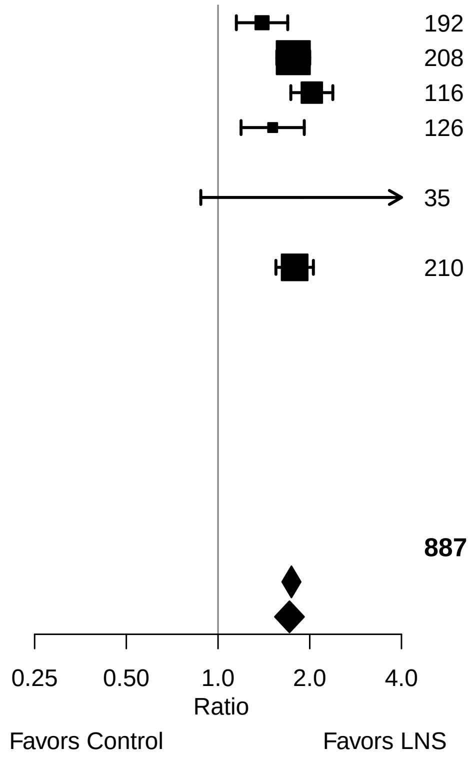

### Supplemental figure 7G: Iron deficiency (ferritin < 12 µg/L) prevalence ratio

#### 7G1: Stratified by Household socio-economic status

**P−for−interaction = 0.847**
**Ratio of PRs = 0.98 (0.78, 1.23)**

| | | **At least median** | | | | | | | **Less than median** | | | | |
|---|---|---|---|---|---|---|---|---|---|---|---|---|---|
| Country | Trial | **LNS** N | **Control** N | **Control** Prevalence | **PR** (95% CI) | **Fixed** W | **Random** W | **LNS** N | **Control** N | **Control** Prevalence | **PR** (95% CI) | **Fixed** W | **Random** W |
| Bangladesh | JiVitA−4 (35) | 243 | 78 | 23.1 | 0.43 (0.23, 0.81) | 0.06 | 0.11 | 211 | 66 | 21.2 | 0.27 (0.11, 0.64) | 0.05 | 0.05 |
| Bangladesh | RDNS (36) | 273 | 136 | 27.2 | 0.26 (0.15, 0.43) | 0.08 | 0.14 | 277 | 136 | 16.9 | 0.47 (0.29, 0.76) | 0.16 | 0.16 |
| Bangladesh | WASH−B (37) | 105 | 91 | 37.4 | 0.23 (0.10, 0.51) | 0.04 | 0.09 | 107 | 87 | 33.3 | 0.25 (0.13, 0.49) | 0.08 | 0.08 |
| Burkina Faso | iLiNS−Zinc (38) | 180 | 39 | 60.5 | 0.70 (0.55, 0.88) | 0.41 | 0.22 | 135 | 57 | 57.1 | 0.46 (0.32, 0.65) | 0.30 | 0.30 |
| Burkina Faso | PROMIS CS (39) | | | | | | | | | | | | |
| Ghana | GHANA (40) | 46 | 39 | 59.0 | 0.44 (0.25, 0.77) | 0.08 | 0.13 | 37 | 39 | 51.3 | 0.58 (0.32, 1.04) | 0.11 | 0.11 |
| Ghana | iLiNS−DYADG (41) | | | | | | | | | | | | |
| Kenya | WASH−B (42) | 163 | 155 | 62.6 | 0.37 (0.28, 0.49) | 0.30 | 0.21 | 134 | 104 | 53.8 | 0.47 (0.32, 0.69) | 0.25 | 0.25 |
| Madagascar | MAHAY (43) | 38 | 25 | 40.0 | 0.39 (0.18, 0.85) | 0.04 | 0.09 | 43 | 25 | 64.0 | 0.33 (0.15, 0.71) | 0.06 | 0.06 |
| Malawi | iLiNS−DYADM (44) | | | | | | | | | | | | |
| Malawi | iLiNS−DOSE (45) | | | | | | | | | | | | |
| Mali | PROMIS CS (46) | | | | | | | | | | | | |
| Zimbabwe | SHINE (HIV−) (47) | | | | | | | | | | | | |
| Zimbabwe | SHINE (HIV+) (48) | | | | | | | | | | | | |
| | | **1048** | **563** | | **I² = 0.73, Tau² = 0.08** | | | **944** | **514** | | **I² = 0.00, Tau² = 0.00** | | |
| **Fixed** | | | | | **0.47 (0.40, 0.54)** | | | | | | **0.43 (0.36, 0.52)** | | |
| **Random** | | | | | **0.41 (0.31, 0.54)** | | | | | | **0.43 (0.36, 0.52)** | | |

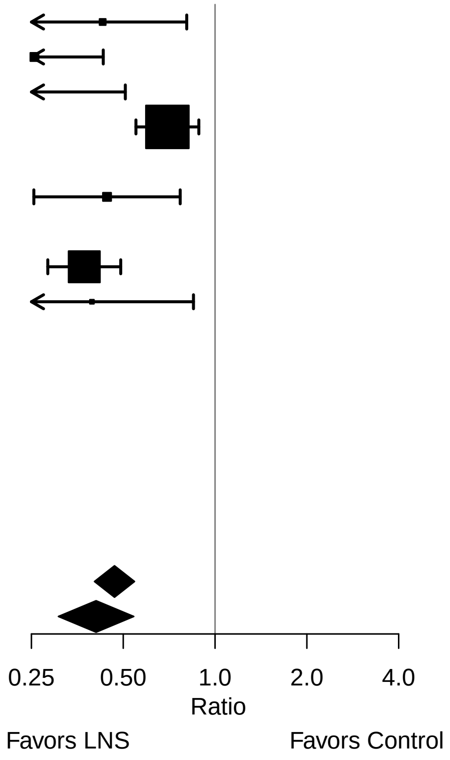

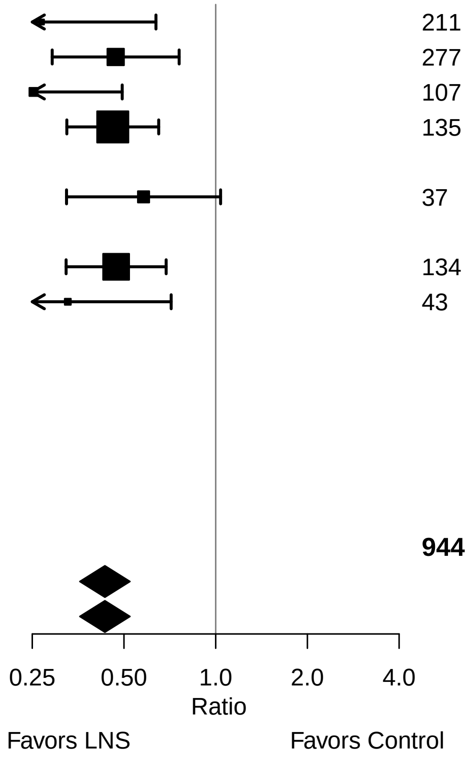

##### Supplemental figure 7G: Iron deficiency (ferritin < 12 µg/L) prevalence ratio

#### 7G2: Stratified by Household food insecurity

**P−for−interaction = 0.859**
**Ratio of PRs = 1.03 (0.77, 1.38)**

| Country | Trial | Mild to secure: LNS N | Control N | Control Prevalence | PR (95% CI) | Fixed W | Random W | Moderate to severe: LNS N | Control N | Control Prevalence | PR (95% CI) | Fixed W | Random W |
|---|---|---|---|---|---|---|---|---|---|---|---|---|---|
| Bangladesh | JiVitA−4 (35) | 337 | 111 | 22.5 | 0.38 (0.22, 0.66) | 0.09 | 0.16 | 117 | 33 | 21.2 | 0.28 (0.11, 0.69) | 0.07 | 0.14 |
| Bangladesh | RDNS (36) | 342 | 166 | 21.1 | 0.42 (0.26, 0.65) | 0.13 | 0.19 | 208 | 106 | 23.6 | 0.22 (0.13, 0.39) | 0.19 | 0.22 |
| Bangladesh | WASH−B (37) | 171 | 139 | 36.7 | 0.19 (0.10, 0.35) | 0.07 | 0.14 | 41 | 39 | 30.8 | 0.48 (0.19, 1.19) | 0.07 | 0.13 |
| Burkina Faso | iLiNS−Zinc (38) | 160 | 45 | 62.7 | 0.55 (0.41, 0.73) | 0.32 | 0.25 | 155 | 51 | 54.5 | 0.68 (0.48, 0.97) | 0.47 | 0.29 |
| Burkina Faso | PROMIS CS (39) | | | | | | | | | | | | |
| Ghana | GHANA (40) | | | | | | | | | | | | |
| Ghana | iLiNS−DYADG (41) | | | | | | | | | | | | |
| Kenya | WASH−B (42) | 259 | 235 | 58.3 | 0.41 (0.31, 0.54) | 0.38 | 0.26 | 38 | 24 | 66.7 | 0.39 (0.22, 0.69) | 0.19 | 0.22 |
| Madagascar | MAHAY (43) | | | | | | | | | | | | |
| Malawi | iLiNS−DYADM (44) | | | | | | | | | | | | |
| Malawi | iLiNS−DOSE (45) | | | | | | | | | | | | |
| Mali | PROMIS CS (46) | | | | | | | | | | | | |
| Zimbabwe | SHINE (HIV−) (47) | | | | | | | | | | | | |
| Zimbabwe | SHINE (HIV+) (48) | | | | | | | | | | | | |
| | | **1269** | **696** | | **I² = 0.59, Tau² = 0.08** | | | **559** | **253** | | **I² = 0.69, Tau² = 0.12** | | |
| **Fixed** | | | | | **0.42 (0.36, 0.50)** | | | | | | **0.45 (0.36, 0.58)** | | |
| **Random** | | | | | **0.39 (0.29, 0.54)** | | | | | | **0.40 (0.26, 0.61)** | | |

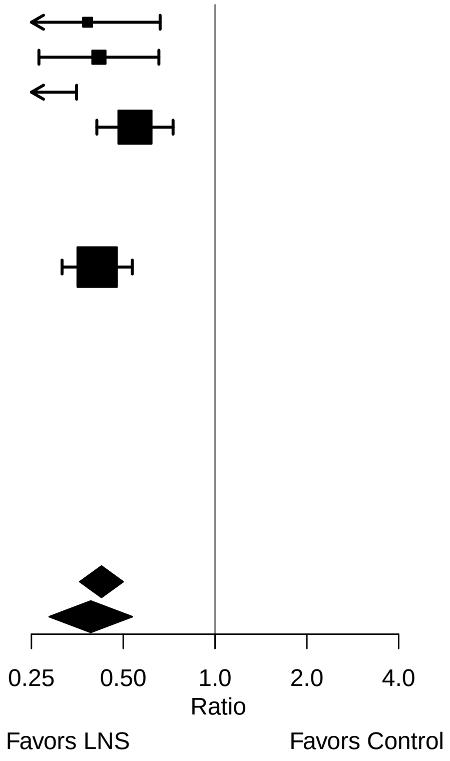

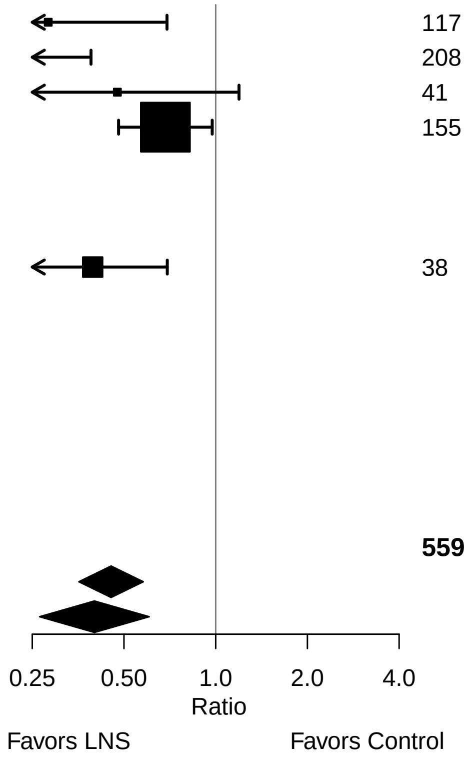

##### Supplemental figure 7G: Iron deficiency (ferritin < 12 µg/L) prevalence ratio

#### 7G3: Stratified by Household source water quality

**P−for−interaction = 0.976**
**Ratio of PRs = 1.00 (0.78, 1.29)**

| Country | Trial | Improved LNS N | Improved Control N | Improved Control Prevalence | Improved PR (95% CI) | Improved Fixed W | Improved Random W | Unimproved LNS N | Unimproved Control N | Unimproved Control Prevalence | Unimproved PR (95% CI) | Unimproved Fixed W | Unimproved Random W |
|---|---|---|---|---|---|---|---|---|---|---|---|---|---|
| Bangladesh | JiVitA−4 (35) | | | | | | | | | | | | |
| Bangladesh | RDNS (36) | | | | | | | | | | | | |
| Bangladesh | WASH−B (37) | | | | | | | | | | | | |
| Burkina Faso | iLiNS−Zinc (38) | 97 | 22 | 69.6 | 0.61 (0.47, 0.78) | 0.66 | 0.62 | 218 | 74 | 55.4 | 0.60 (0.45, 0.81) | 0.65 | 0.46 |
| Burkina Faso | PROMIS CS (39) | | | | | | | | | | | | |
| Ghana | GHANA (40) | | | | | | | | | | | | |
| Ghana | iLiNS−DYADG (41) | | | | | | | | | | | | |
| Kenya | WASH−B (42) | 111 | 107 | 57.0 | 0.43 (0.29, 0.62) | 0.30 | 0.33 | 55 | 37 | 64.9 | 0.48 (0.29, 0.79) | 0.22 | 0.31 |
| Madagascar | MAHAY (43) | 23 | 14 | 42.9 | 0.51 (0.19, 1.38) | 0.04 | 0.05 | 55 | 36 | 55.6 | 0.29 (0.15, 0.56) | 0.13 | 0.23 |
| Malawi | iLiNS−DYADM (44) | | | | | | | | | | | | |
| Malawi | iLiNS−DOSE (45) | | | | | | | | | | | | |
| Mali | PROMIS CS (46) | | | | | | | | | | | | |
| Zimbabwe | SHINE (HIV−) (47) | | | | | | | | | | | | |
| Zimbabwe | SHINE (HIV+) (48) | | | | | | | | | | | | |
| | | **231** | **143** | | **I² = 0.16, Tau² = 0.00** | | | **328** | **147** | | **I² = 0.51, Tau² = 0.06** | | |
| **Fixed** | | | | | **0.54 (0.44, 0.66)** | | | | | | **0.52 (0.41, 0.66)** | | |
| **Random** | | | | | **0.54 (0.43, 0.67)** | | | | | | **0.48 (0.32, 0.70)** | | |

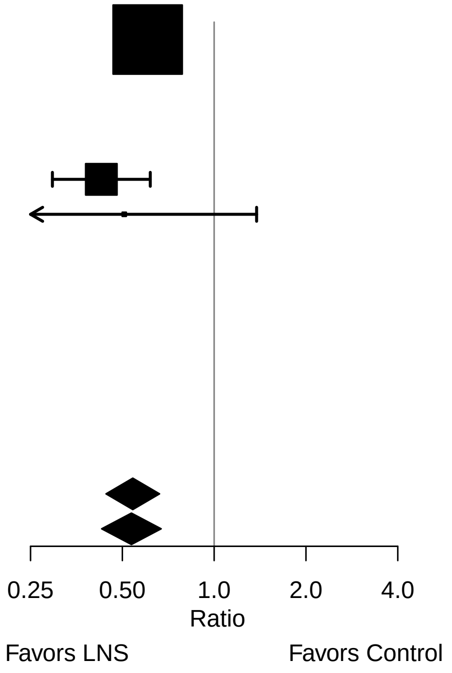

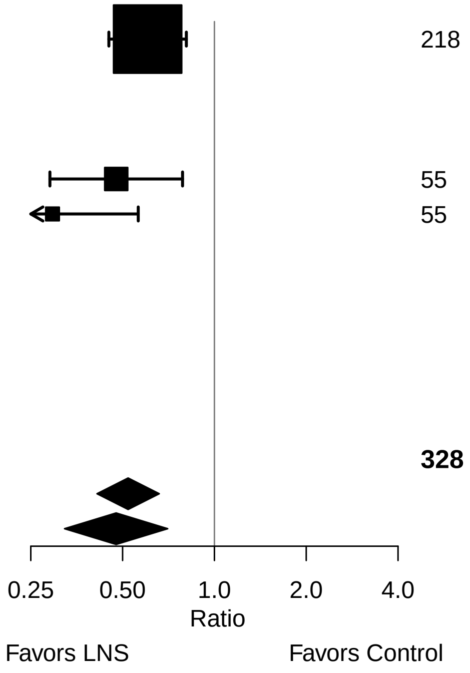

**Supplemental figure 7G: Iron deficiency (ferritin < 12 μg/L) prevalence ratio**

#### 7G4: Stratified by Household sanitation (insufficient comparisons)

**Supplemental figure 7G: Iron deficiency (ferritin < 12 µg/L) prevalence ratio**

#### 7G5: Stratified by Season at the time of assessment

**P−for−interaction = 0.002**
**Ratio of PRs = 0.68 (0.53, 0.87)**

| Country | Trial | Dry LNS N | Dry Control N | Dry Control Prevalence | Dry PR (95% CI) | Dry Fixed W | Dry Random W | Rainy LNS N | Rainy Control N | Rainy Control Prevalence | Rainy PR (95% CI) | Rainy Fixed W | Rainy Random W |
|---|---|---|---|---|---|---|---|---|---|---|---|---|---|
| Bangladesh | JiVitA−4 (35) | 263 | 71 | 21.1 | 0.43 (0.23, 0.81) | 0.08 | 0.08 | 192 | 73 | 23.3 | 0.27 (0.14, 0.53) | 0.08 | 0.16 |
| Bangladesh | RDNS (36) | 341 | 166 | 20.5 | 0.50 (0.35, 0.73) | 0.21 | 0.21 | 208 | 106 | 24.5 | 0.12 (0.05, 0.26) | 0.06 | 0.14 |
| Bangladesh | WASH−B (37) | 96 | 93 | 29.0 | 0.43 (0.22, 0.83) | 0.07 | 0.07 | 116 | 85 | 42.4 | 0.12 (0.05, 0.28) | 0.05 | 0.14 |
| Burkina Faso | iLiNS−Zinc (38) | 174 | 61 | 62.0 | 0.65 (0.49, 0.86) | 0.36 | 0.36 | 126 | 30 | 53.3 | 0.53 (0.38, 0.74) | 0.33 | 0.20 |
| Burkina Faso | PROMIS CS (39) | | | | | | | | | | | | |
| Ghana | GHANA (40) | 48 | 47 | 57.4 | 0.51 (0.31, 0.84) | 0.12 | 0.12 | 35 | 35 | 54.3 | 0.47 (0.25, 0.90) | 0.09 | 0.16 |
| Ghana | iLiNS−DYADG (41) | | | | | | | | | | | | |
| Kenya | WASH−B (42) | 88 | 75 | 64.0 | 0.48 (0.31, 0.74) | 0.16 | 0.16 | 210 | 184 | 57.1 | 0.38 (0.28, 0.52) | 0.39 | 0.20 |
| Madagascar | MAHAY (43) | | | | | | | | | | | | |
| Malawi | iLiNS−DYADM (44) | | | | | | | | | | | | |
| Malawi | iLiNS−DOSE (45) | | | | | | | | | | | | |
| Mali | PROMIS CS (46) | | | | | | | | | | | | |
| Zimbabwe | SHINE (HIV−) (47) | | | | | | | | | | | | |
| Zimbabwe | SHINE (HIV+) (48) | | | | | | | | | | | | |
| | | **1010** | **513** | | **I² = 0.00, Tau² = 0.00** | | | **887** | **513** | | **I² = 0.76, Tau² = 0.32** | | |
| **Fixed** | | | | | **0.53 (0.45, 0.64)** | | | | | | **0.37 (0.31, 0.45)** | | |
| **Random** | | | | | **0.53 (0.45, 0.64)** | | | | | | **0.29 (0.17, 0.48)** | | |

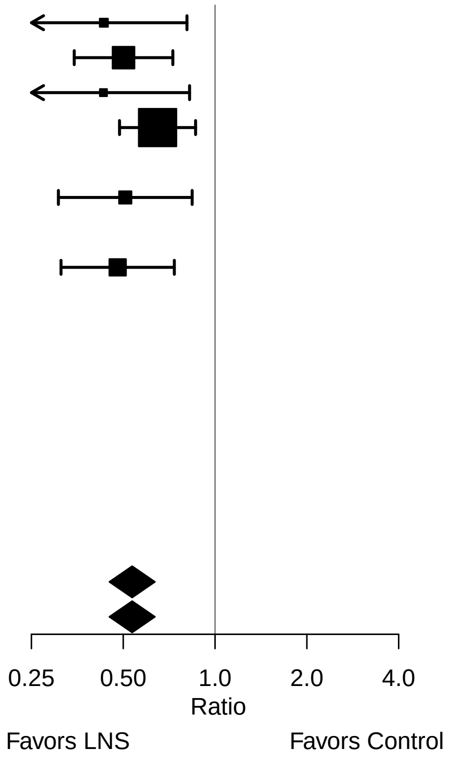

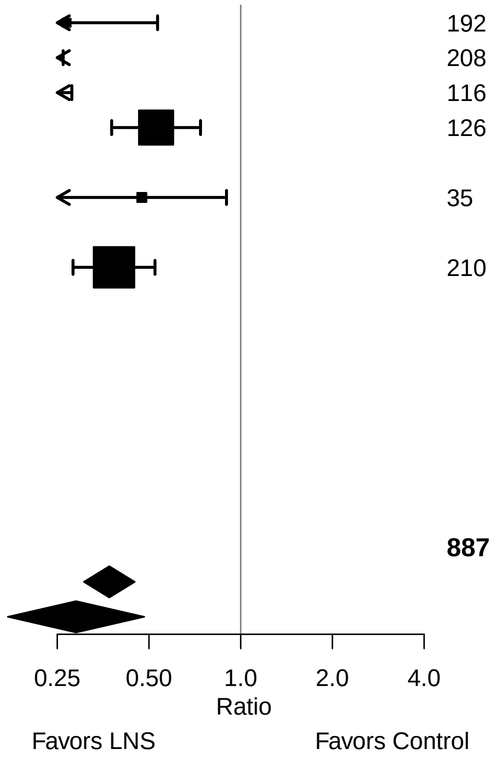

### Supplemental figure 7H: Iron deficiency (ferritin < 12 µg/L) prevalence difference

#### 7H1: Stratified by Household socio-economic status

**P−for−interaction = 0.158**
**Difference in PDs = 0.05 (−0.02, 0.11)**

| | | | | | **At least median** | | | | | | **Less than median** | | |
|---|---|---|---|---|---|---|---|---|---|---|---|---|---|
| | | **LNS** | **Control** | **Control** | **PD** | **Fixed** | **Random** | **LNS** | **Control** | **Control** | **PD** | **Fixed** | **Random** |
| Country | Trial | N | N | Prevalence | (95% CI) | W | W | N | N | Prevalence | (95% CI) | W | W |
| Bangladesh | JiVitA−4 (35) | 243 | 78 | 23.1 | −0.13 (−0.25, −0.01) | 0.12 | 0.15 | 211 | 66 | 21.2 | −0.16 (−0.28, −0.03) | 0.11 | 0.16 |
| Bangladesh | RDNS (36) | 273 | 136 | 27.2 | −0.20 (−0.27, −0.13) | 0.35 | 0.21 | 277 | 136 | 16.9 | −0.09 (−0.15, −0.03) | 0.48 | 0.22 |
| Bangladesh | WASH−B (37) | 105 | 91 | 37.4 | −0.29 (−0.42, −0.15) | 0.09 | 0.13 | 107 | 87 | 33.3 | −0.25 (−0.35, −0.14) | 0.16 | 0.17 |
| Burkina Faso | iLiNS−Zinc (38) | 180 | 39 | 60.5 | −0.18 (−0.31, −0.05) | 0.10 | 0.14 | 135 | 57 | 57.1 | −0.31 (−0.44, −0.17) | 0.09 | 0.14 |
| Burkina Faso | PROMIS CS (39) | | | | | | | | | | | | |
| Ghana | GHANA (40) | 46 | 39 | 59.0 | −0.33 (−0.53, −0.13) | 0.04 | 0.08 | 37 | 39 | 51.3 | −0.22 (−0.43, 0.00) | 0.04 | 0.08 |
| Ghana | iLiNS−DYADG (41) | | | | | | | | | | | | |
| Kenya | WASH−B (42) | 163 | 155 | 62.6 | −0.39 (−0.48, −0.30) | 0.21 | 0.18 | 134 | 104 | 53.8 | −0.28 (−0.42, −0.15) | 0.10 | 0.15 |
| Madagascar | MAHAY (43) | 38 | 25 | 40.0 | −0.24 (−0.40, −0.09) | 0.07 | 0.11 | 43 | 25 | 64.0 | −0.43 (−0.67, −0.19) | 0.03 | 0.07 |
| Malawi | iLiNS−DYADM (44) | | | | | | | | | | | | |
| Malawi | iLiNS−DOSE (45) | | | | | | | | | | | | |
| Mali | PROMIS CS (46) | | | | | | | | | | | | |
| Zimbabwe | SHINE (HIV−) (47) | | | | | | | | | | | | |
| Zimbabwe | SHINE (HIV+) (48) | | | | | | | | | | | | |
| | | **1048** | **563** | | **I² = 0.64, Tau² = 0.00** | | | **944** | **514** | | **I² = 0.71, Tau² = 0.01** | | |
| **Fixed** | | | | | **−0.25 (−0.29, −0.21)** | | | | | | **−0.18 (−0.22, −0.13)** | | |
| **Random** | | | | | **−0.25 (−0.32, −0.18)** | | | | | | **−0.22 (−0.30, −0.15)** | | |

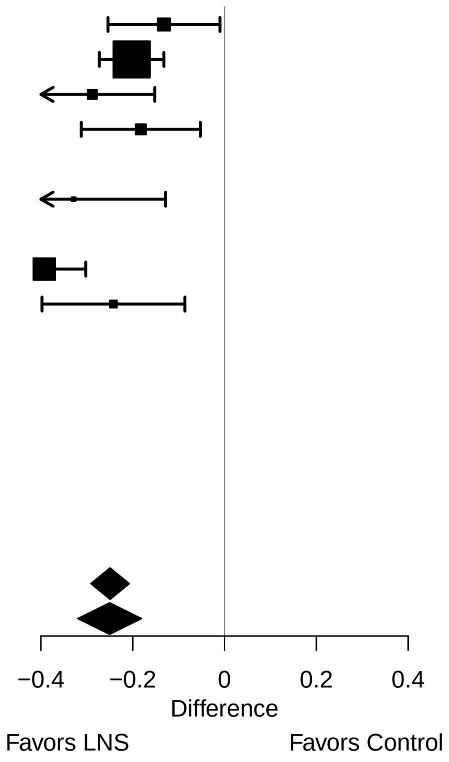

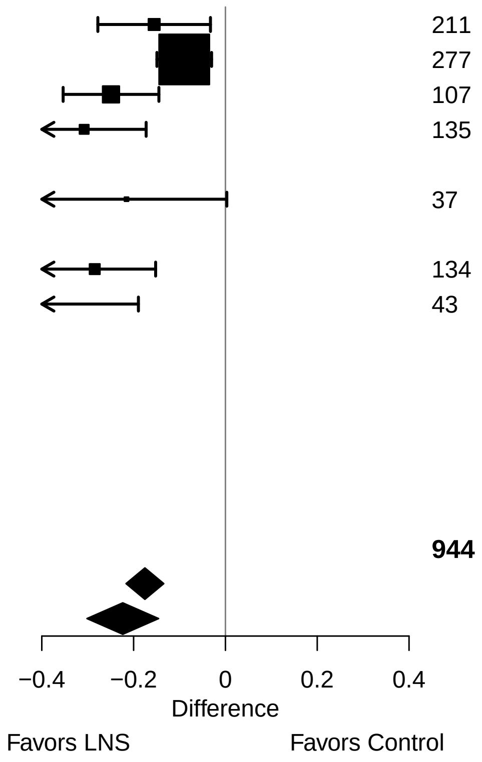

**Supplemental figure 7H: Iron deficiency (ferritin < 12 µg/L) prevalence difference**

#### 7H2: Stratified by Household food insecurity

**P−for−interaction = 0.908**
**Difference in PDs = 0.00 (−0.08, 0.07)**

| | | Mild to secure | | | | | | | Moderate to severe | | | | |
|---|---|---|---|---|---|---|---|---|---|---|---|---|---|
| Country | Trial | LNS N | Control N | Control Prevalence | PD (95% CI) | Fixed W | Random W | LNS N | Control N | Control Prevalence | PD (95% CI) | Fixed W | Random W |
| Bangladesh | JiVitA−4 (35) | 337 | 111 | 22.5 | −0.14 (−0.24, −0.04) | 0.16 | 0.20 | 117 | 33 | 21.2 | −0.15 (−0.28, −0.02) | 0.16 | 0.20 |
| Bangladesh | RDNS (36) | 342 | 166 | 21.1 | −0.12 (−0.19, −0.06) | 0.42 | 0.24 | 208 | 106 | 23.6 | −0.18 (−0.25, −0.11) | 0.58 | 0.44 |
| Bangladesh | WASH−B (37) | 171 | 139 | 36.7 | −0.30 (−0.40, −0.19) | 0.14 | 0.19 | 41 | 39 | 30.8 | −0.16 (−0.34, 0.01) | 0.09 | 0.13 |
| Burkina Faso | iLiNS−Zinc (38) | 160 | 45 | 62.7 | −0.28 (−0.41, −0.14) | 0.09 | 0.17 | 155 | 51 | 54.5 | −0.18 (−0.34, −0.01) | 0.10 | 0.14 |
| Burkina Faso | PROMIS CS (39) | | | | | | | | | | | | |
| Ghana | GHANA (40) | | | | | | | | | | | | |
| Ghana | iLiNS−DYADG (41) | | | | | | | | | | | | |
| Kenya | WASH−B (42) | 259 | 235 | 58.3 | −0.34 (−0.43, −0.25) | 0.20 | 0.21 | 38 | 24 | 66.7 | −0.40 (−0.61, −0.20) | 0.06 | 0.10 |
| Madagascar | MAHAY (43) | | | | | | | | | | | | |
| Malawi | iLiNS−DYADM (44) | | | | | | | | | | | | |
| Malawi | iLiNS−DOSE (45) | | | | | | | | | | | | |
| Mali | PROMIS CS (46) | | | | | | | | | | | | |
| Zimbabwe | SHINE (HIV−) (47) | | | | | | | | | | | | |
| Zimbabwe | SHINE (HIV+) (48) | | | | | | | | | | | | |
| | | **1269** | **696** | | **I² = 0.81, Tau² = 0.01** | | | **559** | **253** | | **I² = 0.14, Tau² = 0.00** | | |
| **Fixed** | | | | | **−0.21 (−0.25, −0.17)** | | | | | | **−0.19 (−0.24, −0.14)** | | |
| **Random** | | | | | **−0.23 (−0.32, −0.14)** | | | | | | **−0.19 (−0.26, −0.13)** | | |

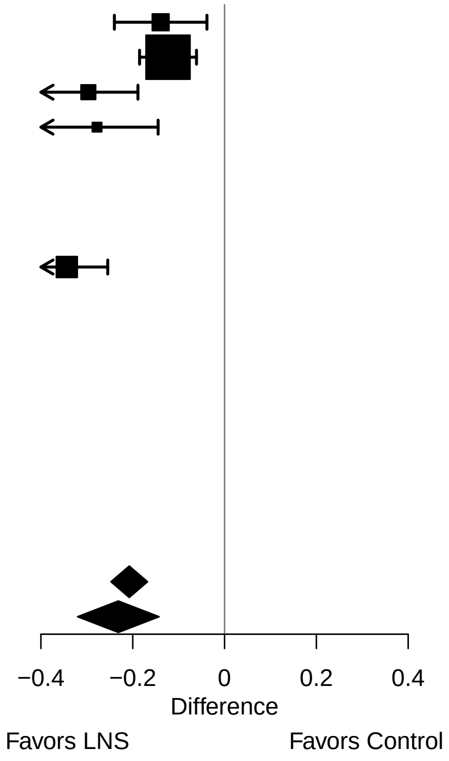

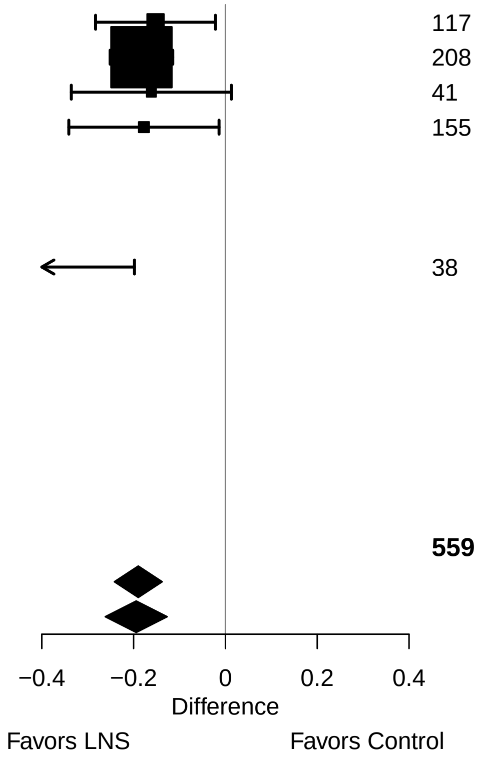

#### Supplemental figure 7H: Iron deficiency (ferritin < 12 µg/L) prevalence difference

##### 7H3: Stratified by Household source water quality

**P−for−interaction = 0.948**
**Difference in PDs = −0.01 (−0.16, 0.15)**

| Country | Trial | LNS N | Control N | Control Prevalence | Improved PD (95% CI) | Fixed W | Random W | LNS N | Control N | Control Prevalence | Unimproved PD (95% CI) | Fixed W | Random W |
|---|---|---|---|---|---|---|---|---|---|---|---|---|---|
| Bangladesh | JiVitA−4 (35) | | | | | | | | | | | | |
| Bangladesh | RDNS (36) | | | | | | | | | | | | |
| Bangladesh | WASH−B (37) | | | | | | | | | | | | |
| Burkina Faso | iLiNS−Zinc (38) | 97 | 22 | 69.6 | −0.28 (−0.42, −0.14) | 0.40 | 0.40 | 218 | 74 | 55.4 | −0.22 (−0.34, −0.10) | 0.52 | 0.45 |
| Burkina Faso | PROMIS CS (39) | | | | | | | | | | | | |
| Ghana | GHANA (40) | | | | | | | | | | | | |
| Ghana | iLiNS−DYADG (41) | | | | | | | | | | | | |
| Kenya | WASH−B (42) | 111 | 107 | 57.0 | −0.33 (−0.45, −0.20) | 0.51 | 0.51 | 55 | 37 | 64.9 | −0.34 (−0.55, −0.12) | 0.17 | 0.21 |
| Madagascar | MAHAY (43) | 23 | 14 | 42.9 | −0.21 (−0.50, 0.08) | 0.09 | 0.09 | 55 | 36 | 55.6 | −0.39 (−0.55, −0.24) | 0.32 | 0.34 |
| Malawi | iLiNS−DYADM (44) | | | | | | | | | | | | |
| Malawi | iLiNS−DOSE (45) | | | | | | | | | | | | |
| Mali | PROMIS CS (46) | | | | | | | | | | | | |
| Zimbabwe | SHINE (HIV−) (47) | | | | | | | | | | | | |
| Zimbabwe | SHINE (HIV+) (48) | | | | | | | | | | | | |
| | | **231** | **143** | | **I² = 0.00, Tau² = 0.00** | | | **328** | **147** | | **I² = 0.39, Tau² = 0.00** | | |
| **Fixed** | | | | | **−0.30 (−0.39, −0.21)** | | | | | | **−0.29 (−0.38, −0.21)** | | |
| **Random** | | | | | **−0.30 (−0.39, −0.21)** | | | | | | **−0.30 (−0.41, −0.19)** | | |

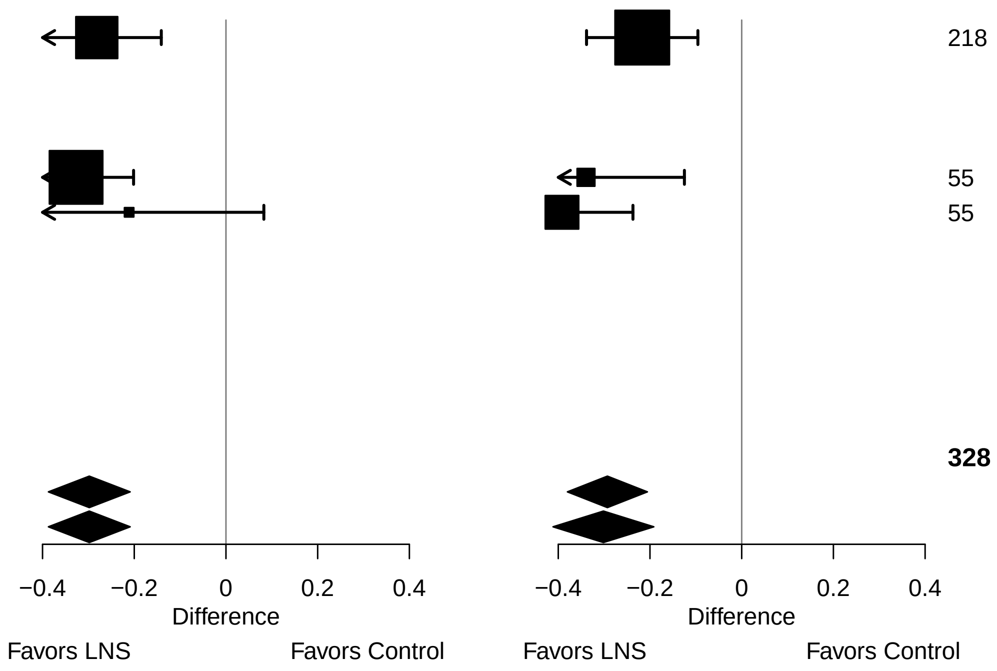

**Supplemental figure 7H: Iron deficiency (ferritin < 12 μg/L) prevalence difference**

#### 7H4: Stratified by Household sanitation (insufficient comparisons)

#### Supplemental figure 7H: Iron deficiency (ferritin < 12 µg/L) prevalence difference

#### 7H5: Stratified by Season at the time of assessment

**P−for−interaction = 0.007**
**Difference in PDs = −0.09 (−0.16, −0.02)**

| | | Dry | | | | | | Rainy | | | | | |
|---|---|---|---|---|---|---|---|---|---|---|---|---|---|
| Country | Trial | LNS N | Control N | Control Prevalence | PD (95% CI) | Fixed W | Random W | LNS N | Control N | Control Prevalence | PD (95% CI) | Fixed W | Random W |
| Bangladesh | JiVitA−4 (35) | 263 | 71 | 21.1 | −0.12 (−0.23, −0.01) | 0.13 | 0.19 | 192 | 73 | 23.3 | −0.17 (−0.28, −0.06) | 0.13 | 0.17 |
| Bangladesh | RDNS (36) | 341 | 166 | 20.5 | −0.10 (−0.16, −0.05) | 0.59 | 0.30 | 208 | 106 | 24.5 | −0.22 (−0.28, −0.16) | 0.43 | 0.25 |
| Bangladesh | WASH−B (37) | 96 | 93 | 29.0 | −0.17 (−0.30, −0.03) | 0.10 | 0.16 | 116 | 85 | 42.4 | −0.37 (−0.47, −0.27) | 0.15 | 0.18 |
| Burkina Faso | iLiNS−Zinc (38) | 174 | 61 | 62.0 | −0.22 (−0.37, −0.07) | 0.08 | 0.14 | 126 | 30 | 53.3 | −0.25 (−0.39, −0.12) | 0.09 | 0.14 |
| Burkina Faso | PROMIS CS (39) | | | | | | | | | | | | |
| Ghana | GHANA (40) | 48 | 47 | 57.4 | −0.28 (−0.48, −0.09) | 0.05 | 0.10 | 35 | 35 | 54.3 | −0.29 (−0.51, −0.06) | 0.03 | 0.07 |
| Ghana | iLiNS−DYADG (41) | | | | | | | | | | | | |
| Kenya | WASH−B (42) | 88 | 75 | 64.0 | −0.33 (−0.50, −0.16) | 0.06 | 0.12 | 210 | 184 | 57.1 | −0.35 (−0.45, −0.25) | 0.16 | 0.19 |
| Madagascar | MAHAY (43) | | | | | | | | | | | | |
| Malawi | iLiNS−DYADM (44) | | | | | | | | | | | | |
| Malawi | iLiNS−DOSE (45) | | | | | | | | | | | | |
| Mali | PROMIS CS (46) | | | | | | | | | | | | |
| Zimbabwe | SHINE (HIV−) (47) | | | | | | | | | | | | |
| Zimbabwe | SHINE (HIV+) (48) | | | | | | | | | | | | |
| | | **1010** | **513** | | **I² = 0.52, Tau² = 0.00** | | | **887** | **513** | | **I² = 0.61, Tau² = 0.00** | | |
| **Fixed** | | | | | **−0.14 (−0.18, −0.10)** | | | | | | **−0.26 (−0.30, −0.22)** | | |
| **Random** | | | | | **−0.18 (−0.25, −0.11)** | | | | | | **−0.27 (−0.34, −0.21)** | | |

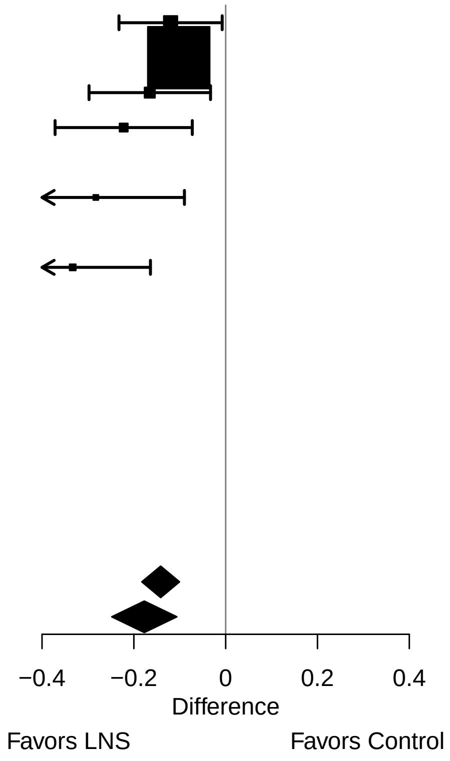

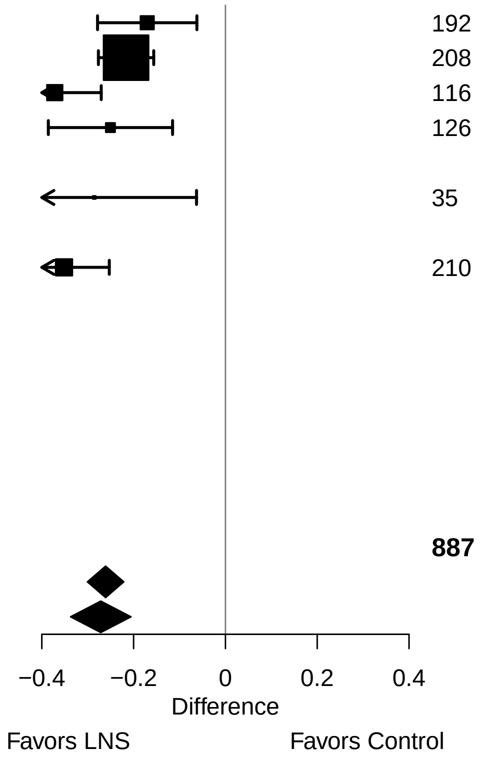

### Supplemental figure 7I: Iron deficiency anemia prevalence ratio

#### 7I1: Stratified by Household socio-economic status

**P−for−interaction = 0.751**
**Ratio of PRs = 1.06 (0.74, 1.52)**

| Country | Trial | At least median: LNS N | Control N | Control Prevalence | PR (95% CI) | Fixed W | Random W | Less than median: LNS N | Control N | Control Prevalence | PR (95% CI) | Fixed W | Random W |
|---|---|---|---|---|---|---|---|---|---|---|---|---|---|
| Bangladesh | JiVitA−4 (35) | | | | | | | | | | | | |
| Bangladesh | RDNS (36) | 272 | 136 | 17.6 | 0.17 (0.08, 0.33) | 0.13 | 0.29 | 277 | 136 | 11.8 | 0.37 (0.18, 0.76) | 0.18 | 0.18 |
| Bangladesh | WASH−B (37) | | | | | | | | | | | | |
| Burkina Faso | iLiNS−Zinc (38) | 188 | 41 | 58.5 | 0.56 (0.40, 0.77) | 0.61 | 0.37 | 139 | 60 | 51.7 | 0.40 (0.27, 0.59) | 0.62 | 0.62 |
| Burkina Faso | PROMIS CS (39) | | | | | | | | | | | | |
| Ghana | GHANA (40) | | | | | | | | | | | | |
| Ghana | iLiNS−DYADG (41) | | | | | | | | | | | | |
| Kenya | WASH−B (42) | 163 | 155 | 38.1 | 0.24 (0.15, 0.40) | 0.26 | 0.34 | 134 | 103 | 26.2 | 0.37 (0.19, 0.73) | 0.20 | 0.20 |
| Madagascar | MAHAY (43) | | | | | | | | | | | | |
| Malawi | iLiNS−DYADM (44) | | | | | | | | | | | | |
| Malawi | iLiNS−DOSE (45) | | | | | | | | | | | | |
| Mali | PROMIS CS (46) | | | | | | | | | | | | |
| Zimbabwe | SHINE (HIV−) (47) | | | | | | | | | | | | |
| Zimbabwe | SHINE (HIV+) (48) | | | | | | | | | | | | |
| | | **623** | **332** | | **I² = 0.86, Tau² = 0.32** | | | **550** | **299** | | **I² = 0.00, Tau² = 0.00** | | |
| **Fixed** | | | | | **0.38 (0.30, 0.49)** | | | | | | **0.39 (0.29, 0.52)** | | |
| **Random** | | | | | **0.30 (0.15, 0.60)** | | | | | | **0.39 (0.29, 0.52)** | | |

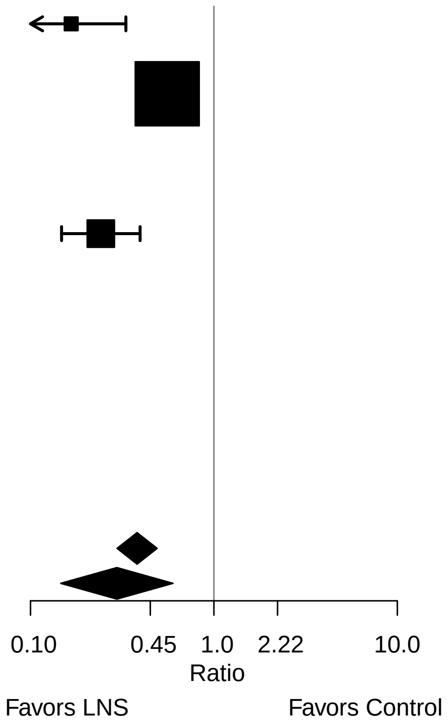

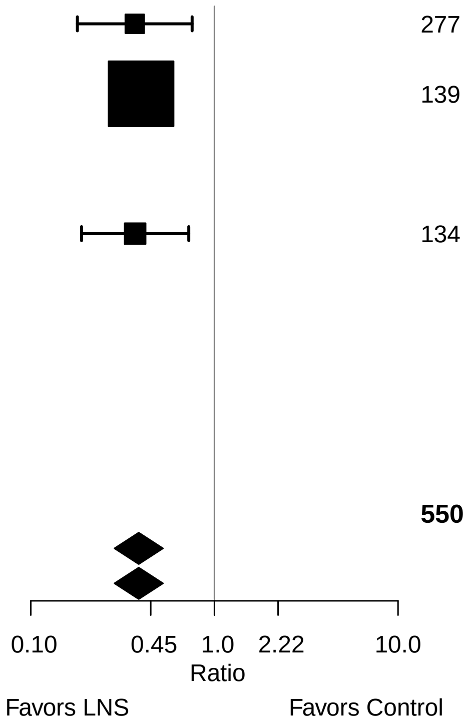

**Supplemental figure 7I: Iron deficiency anemia prevalence ratio**

#### 7I2: Stratified by Household food insecurity (insufficient comparisons)

**Supplemental figure 7I: Iron deficiency anemia prevalence ratio**

#### 7I3: Stratified by Household source water quality (insufficient comparisons)

**Supplemental figure 7I: Iron deficiency anemia prevalence ratio**

#### 7I4: Stratified by Household sanitation (insufficient comparisons)

**Supplemental figure 7I: Iron deficiency anemia prevalence ratio**

#### 7I5: Stratified by Season at the time of assessment (insufficient comparisons)

### Supplemental figure 7J: Iron deficiency anemia prevalence difference

#### 7J1: Stratified by Household socio-economic status

**P−for−interaction = 0.039**
**Difference in PDs = 0.07 (0.00, 0.14)**

| Country | Trial | At least median: LNS N | At least median: Control N | At least median: Control Prevalence | At least median: PD (95% CI) | At least median: Fixed W | At least median: Random W | Less than median: LNS N | Less than median: Control N | Less than median: Control Prevalence | Less than median: PD (95% CI) | Less than median: Fixed W | Less than median: Random W |
|---|---|---|---|---|---|---|---|---|---|---|---|---|---|
| Bangladesh | JiVitA−4 (35) | | | | | | | | | | | | |
| Bangladesh | RDNS (36) | 272 | 136 | 17.6 | −0.15 (−0.21, −0.09) | 0.60 | 0.43 | 277 | 136 | 11.8 | −0.07 (−0.14, −0.01) | 0.67 | 0.39 |
| Bangladesh | WASH−B (37) | | | | | | | | | | | | |
| Burkina Faso | iLiNS−Zinc (38) | 188 | 41 | 58.5 | −0.26 (−0.42, −0.10) | 0.09 | 0.20 | 139 | 60 | 51.7 | −0.31 (−0.45, −0.17) | 0.13 | 0.29 |
| Burkina Faso | PROMIS CS (39) | | | | | | | | | | | | |
| Ghana | GHANA (40) | | | | | | | | | | | | |
| Ghana | iLiNS−DYADG (41) | | | | | | | | | | | | |
| Kenya | WASH−B (42) | 163 | 155 | 38.1 | −0.29 (−0.37, −0.21) | 0.32 | 0.37 | 134 | 103 | 26.2 | −0.17 (−0.28, −0.05) | 0.20 | 0.32 |
| Madagascar | MAHAY (43) | | | | | | | | | | | | |
| Malawi | iLiNS−DYADM (44) | | | | | | | | | | | | |
| Malawi | iLiNS−DOSE (45) | | | | | | | | | | | | |
| Mali | PROMIS CS (46) | | | | | | | | | | | | |
| Zimbabwe | SHINE (HIV−) (47) | | | | | | | | | | | | |
| Zimbabwe | SHINE (HIV+) (48) | | | | | | | | | | | | |
| | | **623** | **332** | | **I² = 0.75, Tau² = 0.00** | | | **550** | **299** | | **I² = 0.80, Tau² = 0.01** | | |
| **Fixed** | | | | | **−0.20 (−0.25, −0.16)** | | | | | | **−0.12 (−0.17, −0.07)** | | |
| **Random** | | | | | **−0.22 (−0.31, −0.13)** | | | | | | **−0.17 (−0.30, −0.04)** | | |

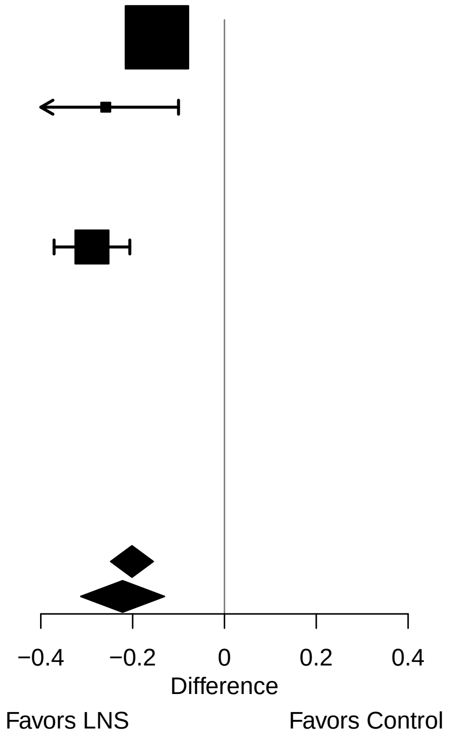

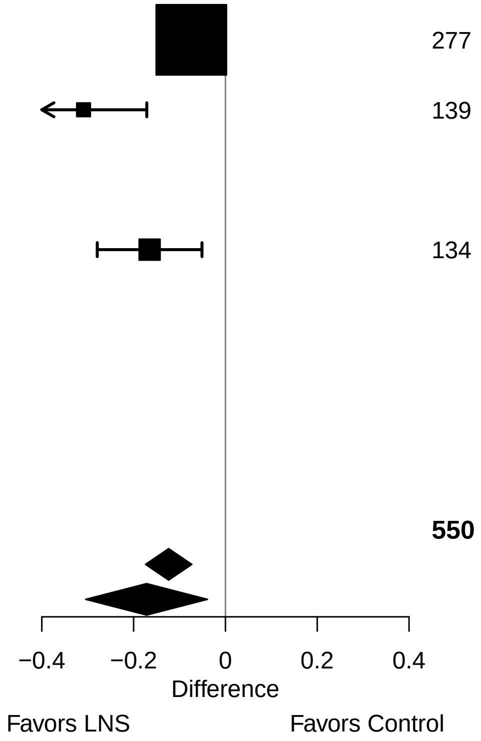

**Supplemental figure 7J: Iron deficiency anemia prevalence difference**

#### 7J2: Stratified by Household food insecurity (insufficient comparisons)

**Supplemental figure 7J: Iron deficiency anemia prevalence difference**

#### 7J3: Stratified by Household source water quality (insufficient comparisons)

**Supplemental figure 7J: Iron deficiency anemia prevalence difference**

#### 7J4: Stratified by Household sanitation (insufficient comparisons)

**Supplemental figure 7J: Iron deficiency anemia prevalence difference**

#### 7J5: Stratified by Season at the time of assessment (insufficient comparisons)

### Supplemental figure 7K: Geometric mean ratio of soluble transferrin receptor concentration

#### 7K1: Stratified by Household socio-economic status

**P−for−interaction = 0.373**
**Ratio of GMRs = 1.03 (0.97, 1.09)**

| | | **At least median** | | | | | | **Less than median** | | | | | |
|---|---|---|---|---|---|---|---|---|---|---|---|---|---|
| Country | Trial | **LNS** N | **Control** N | **Control** Median | **GMR** (95% CI) | **Fixed** W | **Random** W | **LNS** N | **Control** N | **Control** Median | **GMR** (95% CI) | **Fixed** W | **Random** W |
| Bangladesh | JiVitA−4 (35) | | | | | | | | | | | | |
| Bangladesh | RDNS (36) | 273 | 136 | 8.3 | 0.84 (0.78, 0.90) | 0.34 | 0.34 | 277 | 136 | 8.2 | 0.90 (0.84, 0.97) | 0.35 | 0.27 |
| Bangladesh | WASH−B (37) | 105 | 91 | 7.4 | 0.81 (0.73, 0.90) | 0.16 | 0.16 | 107 | 87 | 7.6 | 0.82 (0.76, 0.89) | 0.30 | 0.26 |
| Burkina Faso | iLiNS−Zinc (38) | 180 | 39 | 8.6 | 0.80 (0.71, 0.90) | 0.11 | 0.11 | 135 | 57 | 10.8 | 0.73 (0.64, 0.84) | 0.11 | 0.14 |
| Burkina Faso | PROMIS CS (39) | | | | | | | | | | | | |
| Ghana | GHANA (40) | 47 | 39 | 9.3 | 0.81 (0.69, 0.94) | 0.07 | 0.07 | 37 | 39 | 11.3 | 0.81 (0.69, 0.94) | 0.08 | 0.11 |
| Ghana | iLiNS−DYADG (41) | | | | | | | | | | | | |
| Kenya | WASH−B (42) | 163 | 155 | 11.0 | 0.78 (0.72, 0.84) | 0.26 | 0.26 | 134 | 104 | 10.9 | 0.81 (0.72, 0.91) | 0.13 | 0.16 |
| Madagascar | MAHAY (43) | 38 | 25 | 8.5 | 0.92 (0.77, 1.08) | 0.06 | 0.06 | 43 | 25 | 9.2 | 0.89 (0.70, 1.13) | 0.03 | 0.06 |
| Malawi | iLiNS−DYADM (44) | | | | | | | | | | | | |
| Malawi | iLiNS−DOSE (45) | | | | | | | | | | | | |
| Mali | PROMIS CS (46) | | | | | | | | | | | | |
| Zimbabwe | SHINE (HIV−) (47) | | | | | | | | | | | | |
| Zimbabwe | SHINE (HIV+) (48) | | | | | | | | | | | | |
| | | **806** | **485** | | **I² = 0.00, Tau² = 0.00** | | | **733** | **448** | | **I² = 0.45, Tau² = 0.00** | | |
| **Fixed** | | | | | **0.82 (0.78, 0.85)** | | | | | | **0.84 (0.80, 0.87)** | | |
| **Random** | | | | | **0.82 (0.78, 0.85)** | | | | | | **0.83 (0.78, 0.88)** | | |

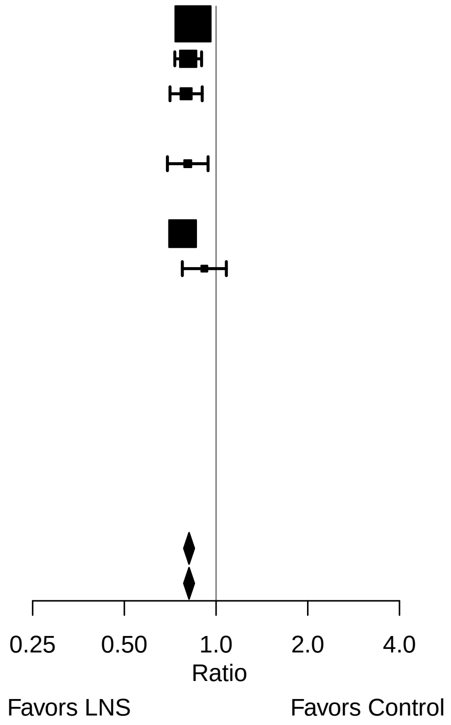

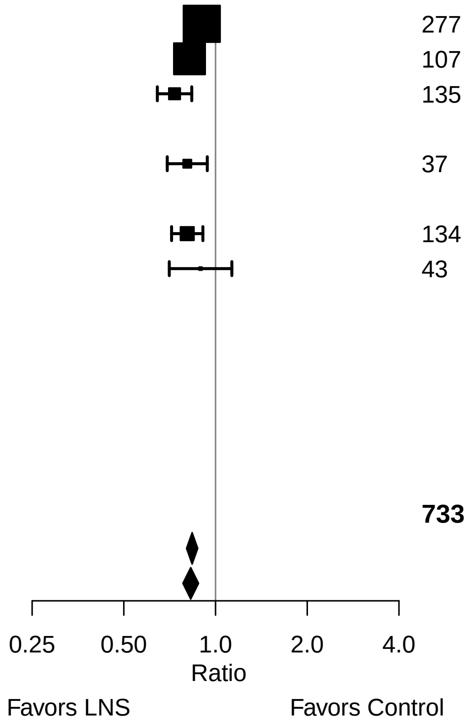

#### Supplemental figure 7K: Geometric mean ratio of soluble transferrin receptor concentration

##### 7K2: Stratified by Household food insecurity

**P−for−interaction = 0.189**
**Ratio of GMRs = 1.05 (0.98, 1.12)**

| | | Mild to secure | | | | | | Moderate to severe | | | | | |
|---|---|---|---|---|---|---|---|---|---|---|---|---|---|
| Country | Trial | LNS N | Control N | Control Median | GMR (95% CI) | Fixed W | Random W | LNS N | Control N | Control Median | GMR (95% CI) | Fixed W | Random W |
| Bangladesh | JiVitA−4 (35) | | | | | | | | | | | | |
| Bangladesh | RDNS (36) | 342 | 166 | 8.4 | 0.88 (0.82, 0.94) | 0.33 | 0.26 | 208 | 106 | 8.0 | 0.86 (0.80, 0.92) | 0.53 | 0.39 |
| Bangladesh | WASH−B (37) | 171 | 139 | 7.5 | 0.81 (0.74, 0.88) | 0.22 | 0.23 | 41 | 39 | 7.7 | 0.84 (0.76, 0.94) | 0.23 | 0.25 |
| Burkina Faso | iLiNS−Zinc (38) | 160 | 45 | 10.3 | 0.73 (0.66, 0.81) | 0.14 | 0.19 | 155 | 51 | 9.5 | 0.78 (0.67, 0.91) | 0.11 | 0.16 |
| Burkina Faso | PROMIS CS (39) | | | | | | | | | | | | |
| Ghana | GHANA (40) | | | | | | | | | | | | |
| Ghana | iLiNS−DYADG (41) | | | | | | | | | | | | |
| Kenya | WASH−B (42) | 259 | 235 | 11.0 | 0.77 (0.71, 0.83) | 0.27 | 0.24 | 38 | 24 | 9.4 | 0.97 (0.82, 1.14) | 0.10 | 0.13 |
| Madagascar | MAHAY (43) | 45 | 30 | 9.1 | 0.87 (0.71, 1.08) | 0.04 | 0.08 | 19 | 7 | 7.7 | 1.00 (0.77, 1.29) | 0.04 | 0.06 |
| Malawi | iLiNS−DYADM (44) | | | | | | | | | | | | |
| Malawi | iLiNS−DOSE (45) | | | | | | | | | | | | |
| Mali | PROMIS CS (46) | | | | | | | | | | | | |
| Zimbabwe | SHINE (HIV−) (47) | | | | | | | | | | | | |
| Zimbabwe | SHINE (HIV+) (48) | | | | | | | | | | | | |
| | | **977** | **615** | | **I² = 0.63, Tau² = 0.00** | | | **461** | **227** | | **I² = 0.19, Tau² = 0.00** | | |
| **Fixed** | | | | | **0.81 (0.78, 0.84)** | | | | | | **0.86 (0.82, 0.91)** | | |
| **Random** | | | | | **0.80 (0.75, 0.86)** | | | | | | **0.86 (0.81, 0.92)** | | |

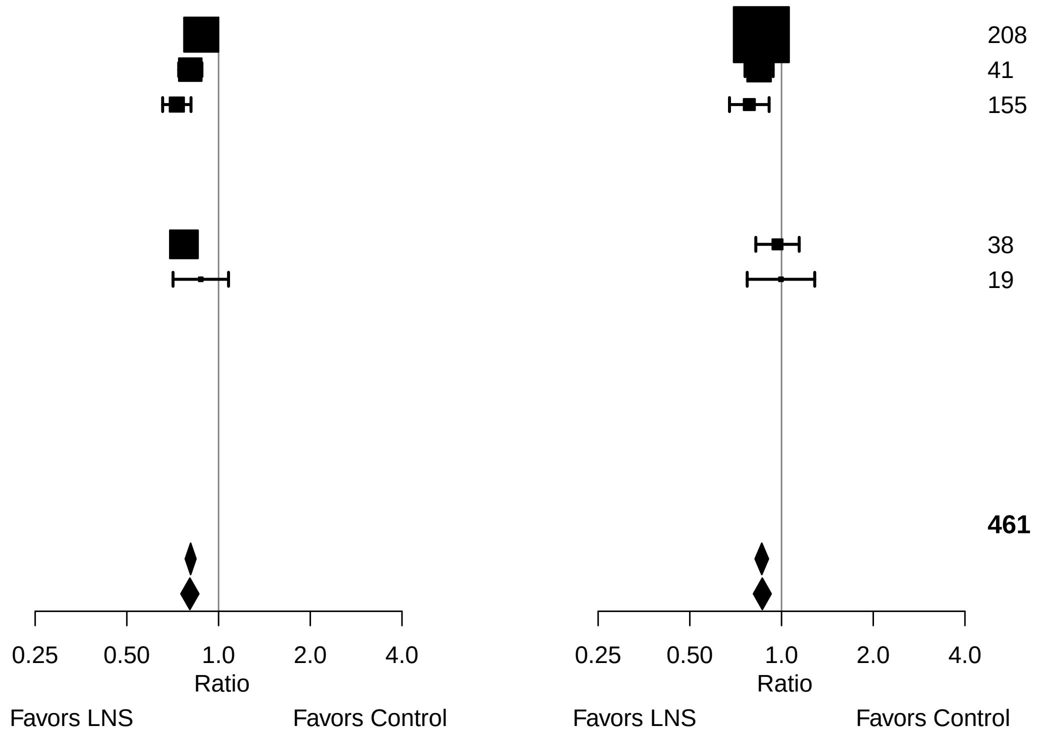

#### Supplemental figure 7K: Geometric mean ratio of soluble transferrin receptor concentration

##### 7K3: Stratified by Household source water quality

**P−for−interaction = 0.039**
**Ratio of GMRs = 1.11 (1.01, 1.22)**

| Country | Trial | Improved LNS N | Improved Control N | Improved Control Median | Improved GMR (95% CI) | Improved Fixed W | Improved Random W | Unimproved LNS N | Unimproved Control N | Unimproved Control Median | Unimproved GMR (95% CI) | Unimproved Fixed W | Unimproved Random W |
|---|---|---|---|---|---|---|---|---|---|---|---|---|---|
| Bangladesh | JiVitA−4 (35) | | | | | | | | | | | | |
| Bangladesh | RDNS (36) | | | | | | | | | | | | |
| Bangladesh | WASH−B (37) | 75 | 44 | 8.1 | 0.70 (0.60, 0.82) | 0.22 | 0.22 | 19 | 14 | 7.8 | 0.83 (0.70, 0.98) | 0.14 | 0.16 |
| Burkina Faso | iLiNS−Zinc (38) | 97 | 22 | 9.9 | 0.74 (0.58, 0.93) | 0.09 | 0.09 | 218 | 74 | 10.0 | 0.77 (0.70, 0.85) | 0.47 | 0.41 |
| Burkina Faso | PROMIS CS (39) | | | | | | | | | | | | |
| Ghana | GHANA (40) | | | | | | | | | | | | |
| Ghana | iLiNS−DYADG (41) | | | | | | | | | | | | |
| Kenya | WASH−B (42) | 111 | 107 | 10.8 | 0.76 (0.70, 0.84) | 0.64 | 0.64 | 55 | 37 | 10.5 | 0.84 (0.73, 0.97) | 0.19 | 0.21 |
| Madagascar | MAHAY (43) | 23 | 14 | 8.9 | 0.88 (0.64, 1.23) | 0.05 | 0.05 | 55 | 36 | 9.0 | 0.91 (0.80, 1.05) | 0.20 | 0.22 |
| Malawi | iLiNS−DYADM (44) | | | | | | | | | | | | |
| Malawi | iLiNS−DOSE (45) | | | | | | | | | | | | |
| Mali | PROMIS CS (46) | | | | | | | | | | | | |
| Zimbabwe | SHINE (HIV−) (47) | | | | | | | | | | | | |
| Zimbabwe | SHINE (HIV+) (48) | | | | | | | | | | | | |
| | | **306** | **187** | | **I² = 0.00, Tau² = 0.00** | | | **347** | **161** | | **I² = 0.28, Tau² = 0.00** | | |
| **Fixed** | | | | | **0.75 (0.70, 0.81)** | | | | | | **0.82 (0.77, 0.87)** | | |
| **Random** | | | | | **0.75 (0.70, 0.81)** | | | | | | **0.83 (0.77, 0.89)** | | |

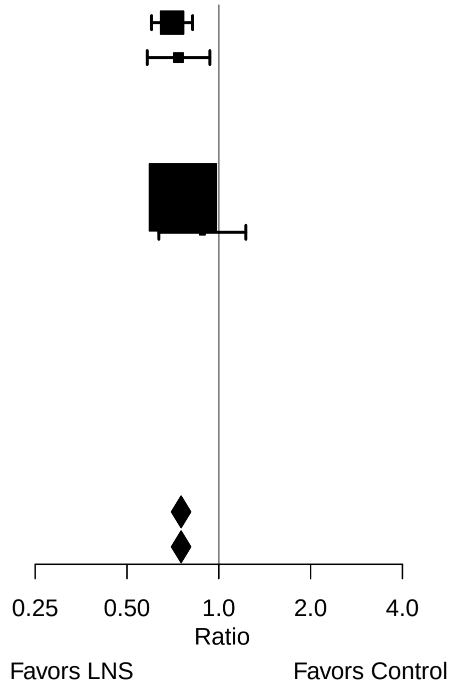

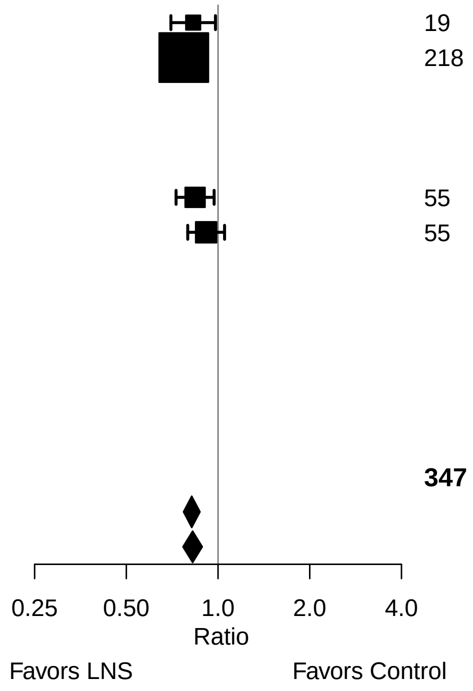

**Supplemental figure 7K: Geometric mean ratio of soluble transferrin receptor concentration**

#### 7K4: Stratified by Household sanitation (insufficient comparisons)

#### Supplemental figure 7K: Geometric mean ratio of soluble transferrin receptor concentration

#### 7K5: Stratified by Season at the time of assessment

**P−for−interaction = 0.865**
**Ratio of GMRs = 1.01 (0.95, 1.07)**

| Country | Trial | Dry LNS N | Dry Control N | Dry Control Median | Dry GMR (95% CI) | Dry Fixed W | Dry Random W | Rainy LNS N | Rainy Control N | Rainy Control Median | Rainy GMR (95% CI) | Rainy Fixed W | Rainy Random W |
|---|---|---|---|---|---|---|---|---|---|---|---|---|---|
| Bangladesh | JiVitA−4 (35) | | | | | | | | | | | | |
| Bangladesh | RDNS (36) | 341 | 166 | 8.2 | 0.86 (0.82, 0.91) | 0.58 | 0.55 | 208 | 106 | 8.4 | 0.88 (0.81, 0.97) | 0.26 | 0.25 |
| Bangladesh | WASH−B (37) | 96 | 93 | 7.6 | 0.82 (0.75, 0.91) | 0.17 | 0.18 | 116 | 85 | 7.1 | 0.82 (0.75, 0.89) | 0.29 | 0.27 |
| Burkina Faso | iLiNS−Zinc (38) | 174 | 61 | 9.6 | 0.77 (0.66, 0.91) | 0.06 | 0.06 | 126 | 30 | 10.2 | 0.72 (0.61, 0.84) | 0.08 | 0.11 |
| Burkina Faso | PROMIS CS (39) | | | | | | | | | | | | |
| Ghana | GHANA (40) | 48 | 47 | 9.7 | 0.80 (0.69, 0.92) | 0.08 | 0.08 | 36 | 35 | 9.6 | 0.80 (0.68, 0.94) | 0.08 | 0.11 |
| Ghana | iLiNS−DYADG (41) | | | | | | | | | | | | |
| Kenya | WASH−B (42) | 88 | 75 | 11.5 | 0.78 (0.70, 0.88) | 0.12 | 0.13 | 210 | 184 | 10.6 | 0.79 (0.73, 0.87) | 0.28 | 0.26 |
| Madagascar | MAHAY (43) | | | | | | | | | | | | |
| Malawi | iLiNS−DYADM (44) | | | | | | | | | | | | |
| Malawi | iLiNS−DOSE (45) | | | | | | | | | | | | |
| Mali | PROMIS CS (46) | | | | | | | | | | | | |
| Zimbabwe | SHINE (HIV−) (47) | | | | | | | | | | | | |
| Zimbabwe | SHINE (HIV+) (48) | | | | | | | | | | | | |
| | | **747** | **442** | | **$I^2$ = 0.04, $Tau^2$ = 0.00** | | | **696** | **440** | | **$I^2$ = 0.31, $Tau^2$ = 0.00** | | |
| **Fixed** | | | | | **0.84 (0.80, 0.87)** | | | | | | **0.82 (0.78, 0.86)** | | |
| **Random** | | | | | **0.83 (0.80, 0.87)** | | | | | | **0.81 (0.77, 0.86)** | | |

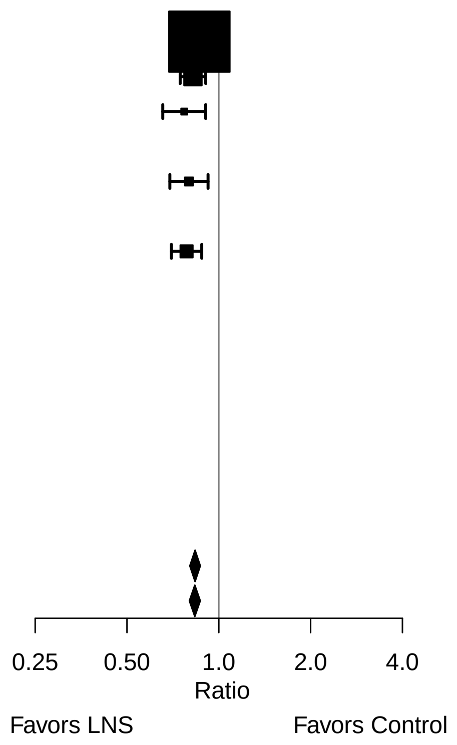

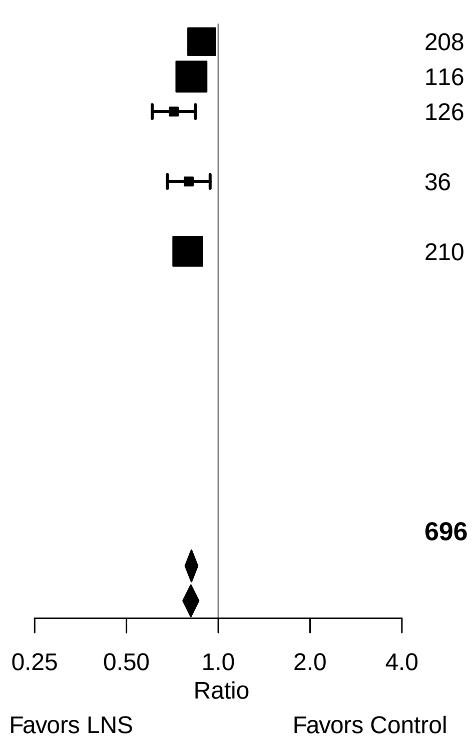

### Supplemental figure 7L: Elevated soluble transferrin receptor prevalence ratio

#### 7L1: Stratified by Household socio-economic status

**P−for−interaction = 0.066**
**Ratio of PRs = 1.18 (0.99, 1.41)**

| | | At least median | | | | | | Less than median | | | | | |
|---|---|---|---|---|---|---|---|---|---|---|---|---|---|
| Country | Trial | LNS N | Control N | Control Prevalence | PR (95% CI) | Fixed W | Random W | LNS N | Control N | Control Prevalence | PR (95% CI) | Fixed W | Random W |
| Bangladesh | JiVitA−4 (35) | | | | | | | | | | | | |
| Bangladesh | RDNS (36) | 273 | 136 | 51.5 | 0.60 (0.46, 0.80) | 0.18 | 0.19 | 277 | 136 | 48.5 | 0.80 (0.65, 0.99) | 0.35 | 0.29 |
| Bangladesh | WASH−B (37) | 105 | 91 | 36.3 | 0.16 (0.08, 0.33) | 0.03 | 0.12 | 107 | 87 | 26.4 | 0.53 (0.32, 0.89) | 0.06 | 0.09 |
| Burkina Faso | iLiNS−Zinc (38) | 180 | 39 | 53.5 | 0.53 (0.36, 0.79) | 0.09 | 0.17 | 135 | 57 | 66.7 | 0.48 (0.30, 0.76) | 0.08 | 0.11 |
| Burkina Faso | PROMIS CS (39) | | | | | | | | | | | | |
| Ghana | GHANA (40) | 47 | 39 | 59.0 | 0.61 (0.39, 0.97) | 0.07 | 0.16 | 37 | 39 | 64.1 | 0.55 (0.33, 0.90) | 0.07 | 0.09 |
| Ghana | iLiNS−DYADG (41) | | | | | | | | | | | | |
| Kenya | WASH−B (42) | 163 | 155 | 73.5 | 0.63 (0.53, 0.73) | 0.58 | 0.20 | 134 | 104 | 72.1 | 0.72 (0.58, 0.90) | 0.33 | 0.28 |
| Madagascar | MAHAY (43) | 38 | 25 | 52.0 | 0.81 (0.49, 1.33) | 0.06 | 0.16 | 43 | 25 | 64.0 | 0.80 (0.54, 1.18) | 0.11 | 0.14 |
| Malawi | iLiNS−DYADM (44) | | | | | | | | | | | | |
| Malawi | iLiNS−DOSE (45) | | | | | | | | | | | | |
| Mali | PROMIS CS (46) | | | | | | | | | | | | |
| Zimbabwe | SHINE (HIV−) (47) | | | | | | | | | | | | |
| Zimbabwe | SHINE (HIV+) (48) | | | | | | | | | | | | |
| | | **806** | **485** | | **I² = 0.66, Tau² = 0.22** | | | **733** | **448** | | **I² = 0.27, Tau² = 0.01** | | |
| **Fixed** | | | | | **0.60 (0.53, 0.68)** | | | | | | **0.71 (0.62, 0.80)** | | |
| **Random** | | | | | **0.53 (0.35, 0.80)** | | | | | | **0.69 (0.58, 0.81)** | | |

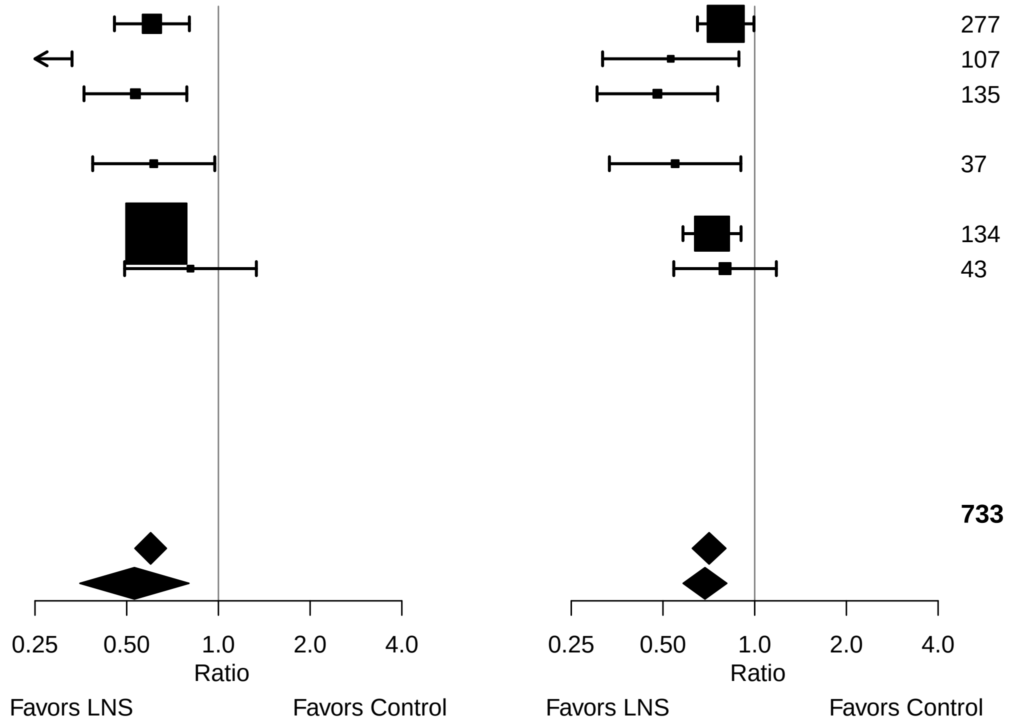

**Supplemental figure 7L: Elevated soluble transferrin receptor prevalence ratio**

#### 7L2: Stratified by Household food insecurity

**P−for−interaction = 0.366**
**Ratio of PRs = 1.10 (0.89, 1.36)**

| | | Mild to secure | | | | | | Moderate to severe | | | | | |
|---|---|---|---|---|---|---|---|---|---|---|---|---|---|
| Country | Trial | LNS N | Control N | Control Prevalence | PR (95% CI) | Fixed W | Random W | LNS N | Control N | Control Prevalence | PR (95% CI) | Fixed W | Random W |
| Bangladesh | JiVitA−4 (35) | | | | | | | | | | | | |
| Bangladesh | RDNS (36) | 342 | 166 | 51.8 | 0.73 (0.59, 0.91) | 0.27 | 0.33 | 208 | 106 | 47.2 | 0.64 (0.49, 0.84) | 0.49 | 0.39 |
| Bangladesh | WASH−B (37) | | | | | | | | | | | | |
| Burkina Faso | iLiNS−Zinc (38) | 160 | 45 | 64.7 | 0.48 (0.36, 0.64) | 0.16 | 0.27 | 155 | 51 | 58.2 | 0.54 (0.36, 0.82) | 0.21 | 0.28 |
| Burkina Faso | PROMIS CS (39) | | | | | | | | | | | | |
| Ghana | GHANA (40) | | | | | | | | | | | | |
| Ghana | iLiNS−DYADG (41) | | | | | | | | | | | | |
| Kenya | WASH−B (42) | 259 | 235 | 73.6 | 0.63 (0.55, 0.74) | 0.57 | 0.39 | 38 | 24 | 66.7 | 0.95 (0.67, 1.34) | 0.29 | 0.33 |
| Madagascar | MAHAY (43) | | | | | | | | | | | | |
| Malawi | iLiNS−DYADM (44) | | | | | | | | | | | | |
| Malawi | iLiNS−DOSE (45) | | | | | | | | | | | | |
| Mali | PROMIS CS (46) | | | | | | | | | | | | |
| Zimbabwe | SHINE (HIV−) (47) | | | | | | | | | | | | |
| Zimbabwe | SHINE (HIV+) (48) | | | | | | | | | | | | |
| | | **761** | **446** | | **I² = 0.63, Tau² = 0.03** | | | **401** | **181** | | **I² = 0.58, Tau² = 0.05** | | |
| **Fixed** | | | | | **0.63 (0.56, 0.71)** | | | | | | **0.69 (0.57, 0.84)** | | |
| **Random** | | | | | **0.62 (0.49, 0.78)** | | | | | | **0.70 (0.51, 0.95)** | | |

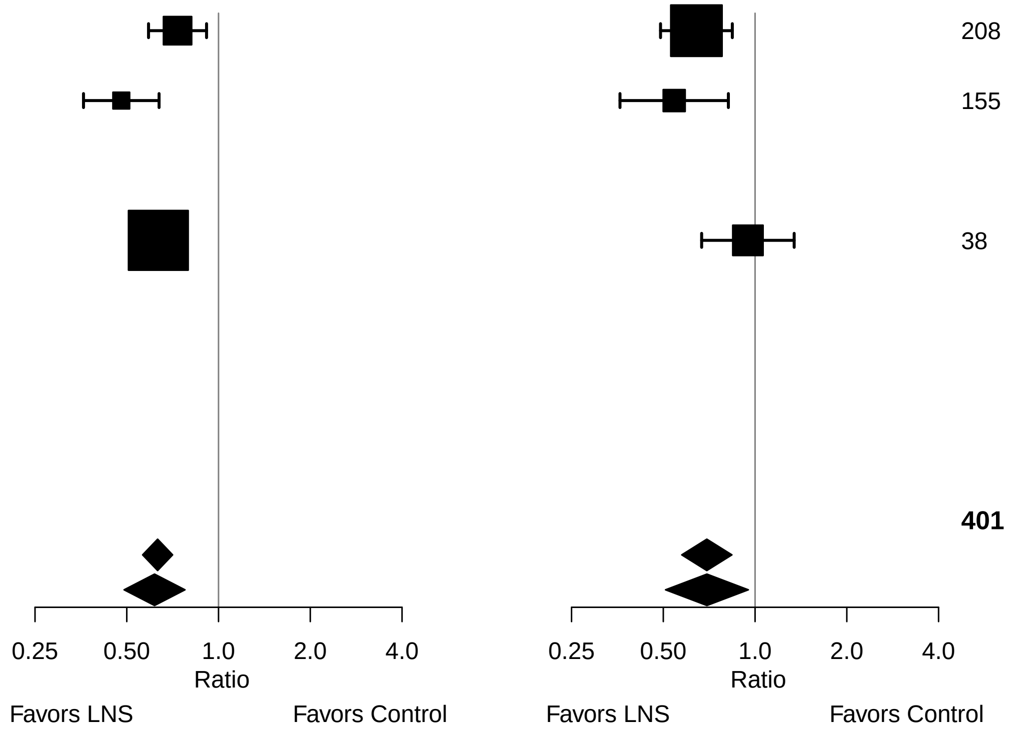

**Supplemental figure 7L: Elevated soluble transferrin receptor prevalence ratio**

#### 7L3: Stratified by Household source water quality

**P−for−interaction = 0.114**
**Ratio of PRs = 1.24 (0.95, 1.62)**

| | | | | | Improved | | | | | | Unimproved | | |
|---|---|---|---|---|---|---|---|---|---|---|---|---|---|
| | | **LNS** | **Control** | **Control** | **PR** | **Fixed** | **Random** | **LNS** | **Control** | **Control** | **PR** | **Fixed** | **Random** |
| Country | Trial | N | N | Prevalence | (95% CI) | W | W | N | N | Prevalence | (95% CI) | W | W |
| Bangladesh | JiVitA−4 (35) | | | | | | | | | | | | |
| Bangladesh | RDNS (36) | | | | | | | | | | | | |
| Bangladesh | WASH−B (37) | | | | | | | | | | | | |
| Burkina Faso | iLiNS−Zinc (38) | 97 | 22 | 65.2 | 0.42 (0.26, 0.67) | 0.12 | 0.31 | 218 | 74 | 60.2 | 0.56 (0.44, 0.73) | 0.48 | 0.45 |
| Burkina Faso | PROMIS CS (39) | | | | | | | | | | | | |
| Ghana | GHANA (40) | | | | | | | | | | | | |
| Ghana | iLiNS−DYADG (41) | | | | | | | | | | | | |
| Kenya | WASH−B (42) | 111 | 107 | 76.6 | 0.59 (0.49, 0.71) | 0.81 | 0.44 | 55 | 37 | 62.2 | 0.79 (0.54, 1.15) | 0.22 | 0.24 |
| Madagascar | MAHAY (43) | 23 | 14 | 57.1 | 0.99 (0.53, 1.85) | 0.07 | 0.25 | 55 | 36 | 58.3 | 0.72 (0.52, 0.99) | 0.29 | 0.31 |
| Malawi | iLiNS−DYADM (44) | | | | | | | | | | | | |
| Malawi | iLiNS−DOSE (45) | | | | | | | | | | | | |
| Mali | PROMIS CS (46) | | | | | | | | | | | | |
| Zimbabwe | SHINE (HIV−) (47) | | | | | | | | | | | | |
| Zimbabwe | SHINE (HIV+) (48) | | | | | | | | | | | | |
| | | **231** | **143** | | **I² = 0.57, Tau² = 0.11** | | | **328** | **147** | | **I² = 0.23, Tau² = 0.01** | | |
| **Fixed** | | | | | **0.59 (0.49, 0.69)** | | | | | | **0.65 (0.55, 0.78)** | | |
| **Random** | | | | | **0.60 (0.39, 0.94)** | | | | | | **0.66 (0.54, 0.81)** | | |

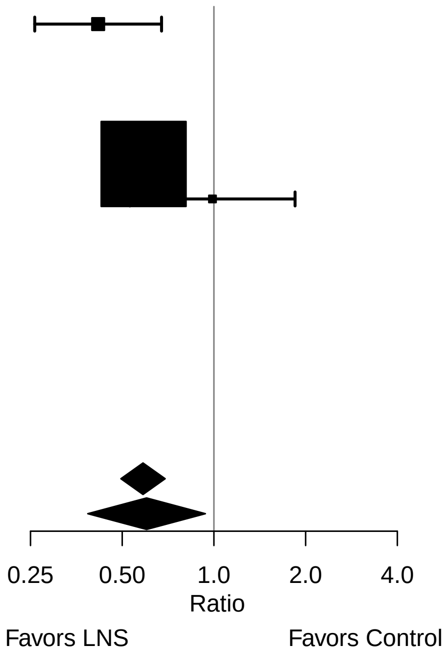

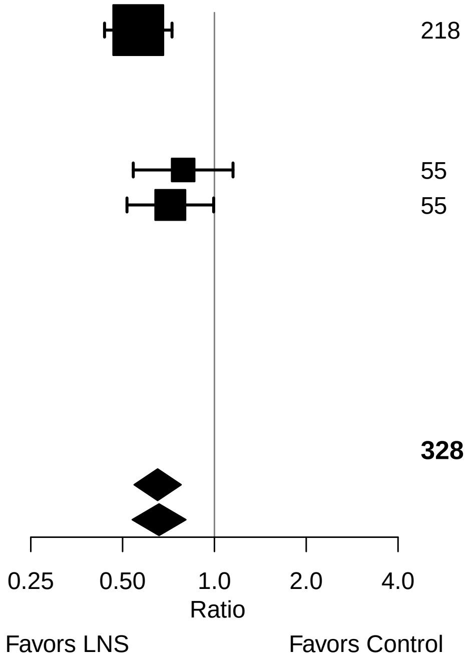

**Supplemental figure 7L: Elevated soluble transferrin receptor prevalence ratio**

#### 7L4: Stratified by Household sanitation (insufficient comparisons)

#### Supplemental figure 7L: Elevated soluble transferrin receptor prevalence ratio

##### 7L5: Stratified by Season at the time of assessment

**P−for−interaction = 0.154**
**Ratio of PRs = 1.14 (0.95, 1.35)**

| Country | Trial | Dry: LNS N | Dry: Control N | Dry: Control Prevalence | Dry: PR (95% CI) | Dry: Fixed W | Dry: Random W | Rainy: LNS N | Rainy: Control N | Rainy: Control Prevalence | Rainy: PR (95% CI) | Rainy: Fixed W | Rainy: Random W |
|---|---|---|---|---|---|---|---|---|---|---|---|---|---|
| Bangladesh | JiVitA−4 (35) | | | | | | | | | | | | |
| Bangladesh | RDNS (36) | 341 | 166 | 49.4 | 0.62 (0.50, 0.77) | 0.39 | 0.38 | 208 | 106 | 50.9 | 0.82 (0.65, 1.04) | 0.26 | 0.24 |
| Bangladesh | WASH−B (37) | 96 | 93 | 31.2 | 0.40 (0.26, 0.62) | 0.09 | 0.09 | 116 | 85 | 31.8 | 0.24 (0.12, 0.50) | 0.03 | 0.14 |
| Burkina Faso | iLiNS−Zinc (38) | 174 | 61 | 59.2 | 0.50 (0.34, 0.73) | 0.12 | 0.12 | 126 | 30 | 70.0 | 0.47 (0.29, 0.75) | 0.07 | 0.19 |
| Burkina Faso | PROMIS CS (39) | | | | | | | | | | | | |
| Ghana | GHANA (40) | 48 | 47 | 66.0 | 0.57 (0.37, 0.86) | 0.10 | 0.11 | 36 | 35 | 57.1 | 0.58 (0.34, 1.00) | 0.05 | 0.17 |
| Ghana | iLiNS−DYADG (41) | | | | | | | | | | | | |
| Kenya | WASH−B (42) | 88 | 75 | 72.0 | 0.63 (0.50, 0.80) | 0.31 | 0.30 | 210 | 184 | 73.4 | 0.69 (0.59, 0.81) | 0.60 | 0.26 |
| Madagascar | MAHAY (43) | | | | | | | | | | | | |
| Malawi | iLiNS−DYADM (44) | | | | | | | | | | | | |
| Malawi | iLiNS−DOSE (45) | | | | | | | | | | | | |
| Mali | PROMIS CS (46) | | | | | | | | | | | | |
| Zimbabwe | SHINE (HIV−) (47) | | | | | | | | | | | | |
| Zimbabwe | SHINE (HIV+) (48) | | | | | | | | | | | | |
| | | **747** | **442** | | **I² = 0.05, Tau² = 0.00** | | | **696** | **440** | | **I² = 0.69, Tau² = 0.14** | | |
| **Fixed** | | | | | **0.58 (0.51, 0.66)** | | | | | | **0.68 (0.60, 0.76)** | | |
| **Random** | | | | | **0.58 (0.50, 0.66)** | | | | | | **0.56 (0.39, 0.82)** | | |

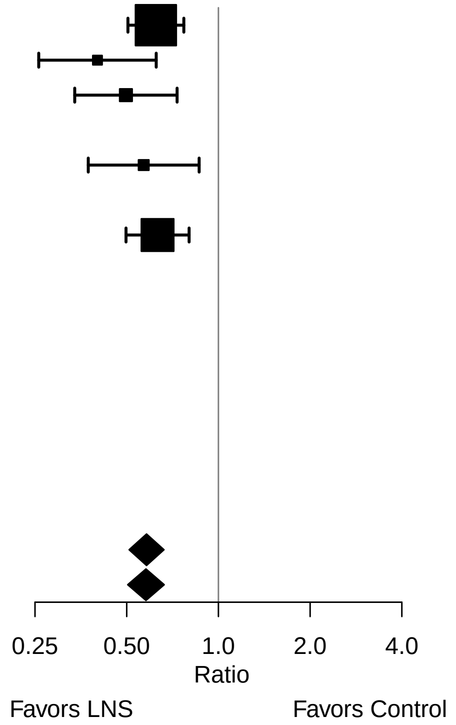

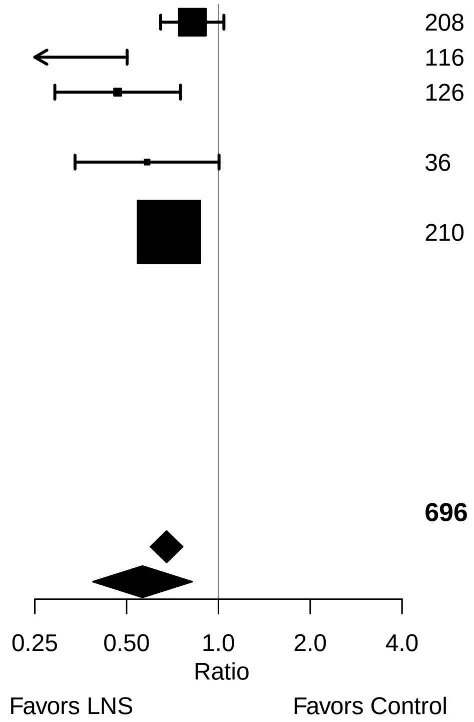

### Supplemental figure 7M: Elevated soluble transferrin receptor prevalence difference

#### 7M1: Stratified by Household socio-economic status

**P−for−interaction = 0.058**
**Difference in PDs = 0.07 (0.00, 0.15)**

| Country | Trial | At least median: LNS N | Control N | Control Prevalence | PD (95% CI) | Fixed W | Random W | Less than median: LNS N | Control N | Control Prevalence | PD (95% CI) | Fixed W | Random W |
|---|---|---|---|---|---|---|---|---|---|---|---|---|---|
| Bangladesh | JiVitA−4 (35) | | | | | | | | | | | | |
| Bangladesh | RDNS (36) | 273 | 136 | 51.5 | −0.20 (−0.32, −0.09) | 0.22 | 0.22 | 277 | 136 | 48.5 | −0.10 (−0.19, 0.00) | 0.32 | 0.25 |
| Bangladesh | WASH−B (37) | 105 | 91 | 36.3 | −0.31 (−0.42, −0.19) | 0.20 | 0.20 | 107 | 87 | 26.4 | −0.12 (−0.22, −0.03) | 0.30 | 0.24 |
| Burkina Faso | iLiNS−Zinc (38) | 180 | 39 | 53.5 | −0.25 (−0.45, −0.05) | 0.07 | 0.07 | 135 | 57 | 66.7 | −0.34 (−0.50, −0.17) | 0.11 | 0.14 |
| Burkina Faso | PROMIS CS (39) | | | | | | | | | | | | |
| Ghana | GHANA (40) | 47 | 39 | 59.0 | −0.23 (−0.44, −0.02) | 0.06 | 0.06 | 37 | 39 | 64.1 | −0.29 (−0.51, −0.07) | 0.06 | 0.09 |
| Ghana | iLiNS−DYADG (41) | | | | | | | | | | | | |
| Kenya | WASH−B (42) | 163 | 155 | 73.5 | −0.28 (−0.36, −0.19) | 0.39 | 0.39 | 134 | 104 | 72.1 | −0.20 (−0.34, −0.06) | 0.15 | 0.18 |
| Madagascar | MAHAY (43) | 38 | 25 | 52.0 | −0.10 (−0.33, 0.13) | 0.05 | 0.05 | 43 | 25 | 64.0 | −0.13 (−0.35, 0.09) | 0.06 | 0.09 |
| Malawi | iLiNS−DYADM (44) | | | | | | | | | | | | |
| Malawi | iLiNS−DOSE (45) | | | | | | | | | | | | |
| Mali | PROMIS CS (46) | | | | | | | | | | | | |
| Zimbabwe | SHINE (HIV−) (47) | | | | | | | | | | | | |
| Zimbabwe | SHINE (HIV+) (48) | | | | | | | | | | | | |
| | | **806** | **485** | | **I² = 0.00, Tau² = 0.00** | | | **733** | **448** | | **I² = 0.41, Tau² = 0.00** | | |
| **Fixed** | | | | | **−0.25 (−0.30, −0.20)** | | | | | | **−0.16 (−0.21, −0.11)** | | |
| **Random** | | | | | **−0.25 (−0.30, −0.20)** | | | | | | **−0.18 (−0.25, −0.10)** | | |

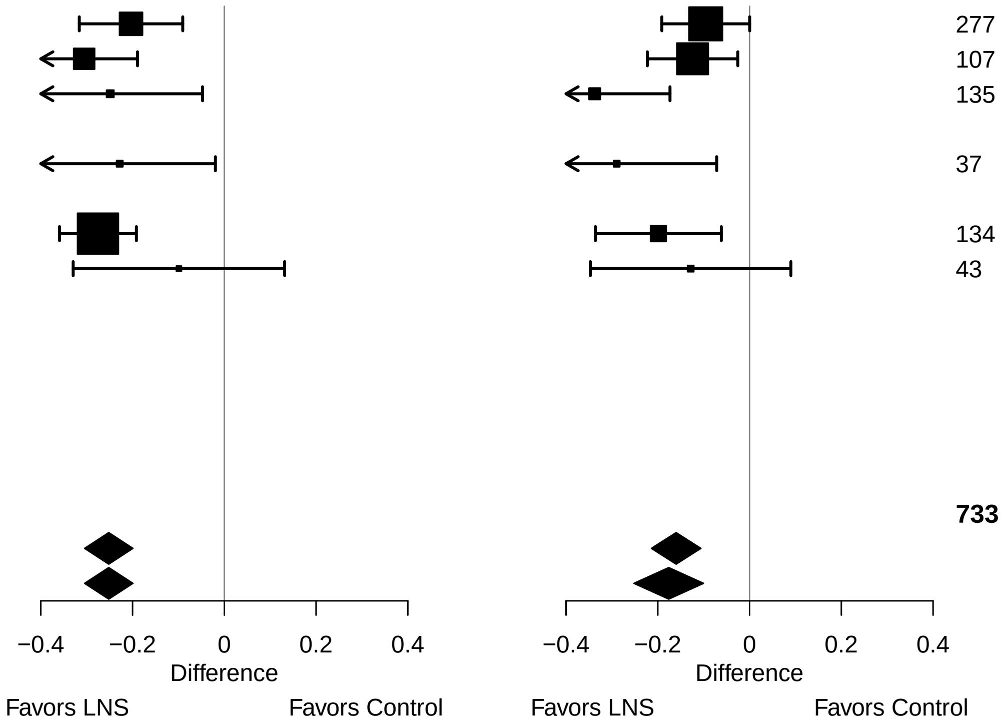

#### Supplemental figure 7M: Elevated soluble transferrin receptor prevalence difference

##### 7M2: Stratified by Household food insecurity

**P−for−interaction = 0.433**
**Difference in PDs = 0.04 (−0.07, 0.15)**

| Country | Trial | Mild to secure: LNS N | Control N | Control Prevalence | PD (95% CI) | Fixed W | Random W | Moderate to severe: LNS N | Control N | Control Prevalence | PD (95% CI) | Fixed W | Random W |
|---|---|---|---|---|---|---|---|---|---|---|---|---|---|
| Bangladesh | JiVitA−4 (35) | | | | | | | | | | | | |
| Bangladesh | RDNS (36) | 342 | 166 | 51.8 | −0.14 (−0.24, −0.04) | 0.33 | 0.35 | 208 | 106 | 47.2 | −0.17 (−0.27, −0.06) | 0.65 | 0.56 |
| Bangladesh | WASH−B (37) | | | | | | | | | | | | |
| Burkina Faso | iLiNS−Zinc (38) | 160 | 45 | 64.7 | −0.34 (−0.48, −0.19) | 0.16 | 0.27 | 155 | 51 | 58.2 | −0.27 (−0.45, −0.08) | 0.21 | 0.26 |
| Burkina Faso | PROMIS CS (39) | | | | | | | | | | | | |
| Ghana | GHANA (40) | | | | | | | | | | | | |
| Ghana | iLiNS−DYADG (41) | | | | | | | | | | | | |
| Kenya | WASH−B (42) | 259 | 235 | 73.6 | −0.27 (−0.35, −0.19) | 0.51 | 0.38 | 38 | 24 | 66.7 | −0.04 (−0.26, 0.19) | 0.14 | 0.18 |
| Madagascar | MAHAY (43) | | | | | | | | | | | | |
| Malawi | iLiNS−DYADM (44) | | | | | | | | | | | | |
| Malawi | iLiNS−DOSE (45) | | | | | | | | | | | | |
| Mali | PROMIS CS (46) | | | | | | | | | | | | |
| Zimbabwe | SHINE (HIV−) (47) | | | | | | | | | | | | |
| Zimbabwe | SHINE (HIV+) (48) | | | | | | | | | | | | |
| | | **761** | **446** | | **I² = 0.67, Tau² = 0.01** | | | **401** | **181** | | **I² = 0.16, Tau² = 0.00** | | |
| **Fixed** | | | | | **−0.24 (−0.29, −0.18)** | | | | | | **−0.17 (−0.26, −0.09)** | | |
| **Random** | | | | | **−0.24 (−0.35, −0.13)** | | | | | | **−0.17 (−0.27, −0.07)** | | |

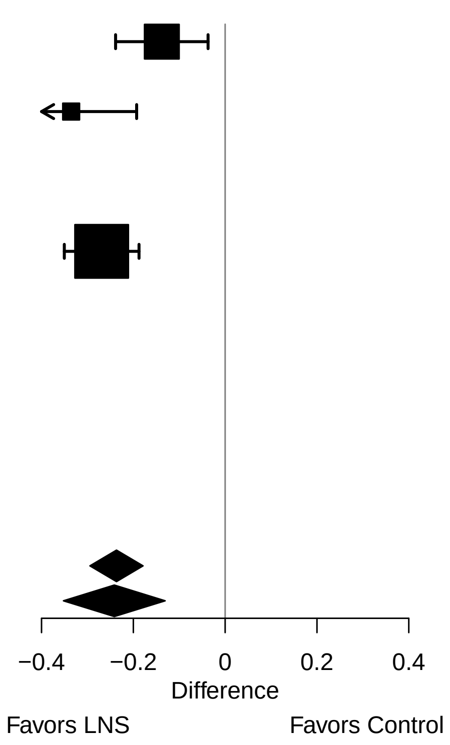

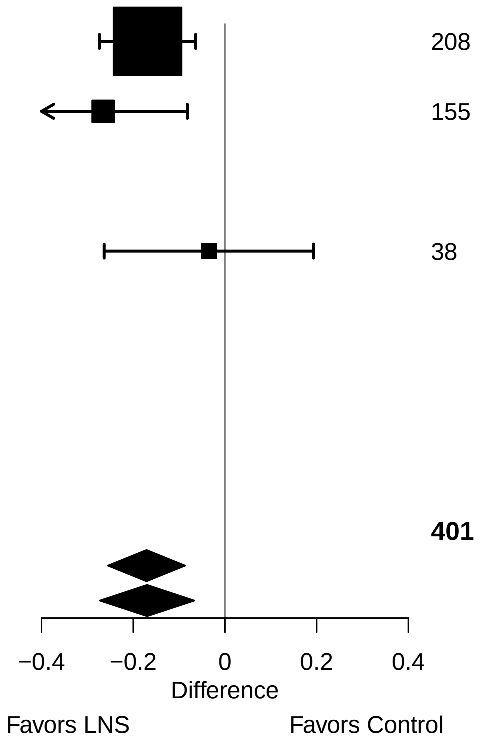

**Supplemental figure 7M: Elevated soluble transferrin receptor prevalence difference**

#### 7M3: Stratified by Household source water quality

**P−for−interaction = 0.231**
**Difference in PDs = 0.10 (−0.06, 0.25)**

| Country | Trial | Improved LNS N | Improved Control N | Improved Control Prevalence | Improved PD (95% CI) | Improved Fixed W | Improved Random W | Unimproved LNS N | Unimproved Control N | Unimproved Control Prevalence | Unimproved PD (95% CI) | Unimproved Fixed W | Unimproved Random W |
|---|---|---|---|---|---|---|---|---|---|---|---|---|---|
| Bangladesh | JiVitA−4 (35) | | | | | | | | | | | | |
| Bangladesh | RDNS (36) | | | | | | | | | | | | |
| Bangladesh | WASH−B (37) | | | | | | | | | | | | |
| Burkina Faso | iLiNS−Zinc (38) | 97 | 22 | 65.2 | −0.35 (−0.63, −0.08) | 0.12 | 0.25 | 218 | 74 | 60.2 | −0.26 (−0.38, −0.15) | 0.56 | 0.56 |
| Burkina Faso | PROMIS CS (39) | | | | | | | | | | | | |
| Ghana | GHANA (40) | | | | | | | | | | | | |
| Ghana | iLiNS−DYADG (41) | | | | | | | | | | | | |
| Kenya | WASH−B (42) | 111 | 107 | 76.6 | −0.32 (−0.42, −0.21) | 0.81 | 0.57 | 55 | 37 | 62.2 | −0.13 (−0.34, 0.08) | 0.16 | 0.16 |
| Madagascar | MAHAY (43) | 23 | 14 | 57.1 | −0.01 (−0.36, 0.35) | 0.07 | 0.18 | 55 | 36 | 58.3 | −0.17 (−0.33, −0.01) | 0.28 | 0.28 |
| Malawi | iLiNS−DYADM (44) | | | | | | | | | | | | |
| Malawi | iLiNS−DOSE (45) | | | | | | | | | | | | |
| Mali | PROMIS CS (46) | | | | | | | | | | | | |
| Zimbabwe | SHINE (HIV−) (47) | | | | | | | | | | | | |
| Zimbabwe | SHINE (HIV+) (48) | | | | | | | | | | | | |
| | | **231** | **143** | | **I² = 0.30, Tau² = 0.01** | | | **328** | **147** | | **I² = 0.00, Tau² = 0.00** | | |
| **Fixed** | | | | | **−0.30 (−0.39, −0.20)** | | | | | | **−0.22 (−0.30, −0.13)** | | |
| **Random** | | | | | **−0.27 (−0.44, −0.10)** | | | | | | **−0.22 (−0.30, −0.13)** | | |

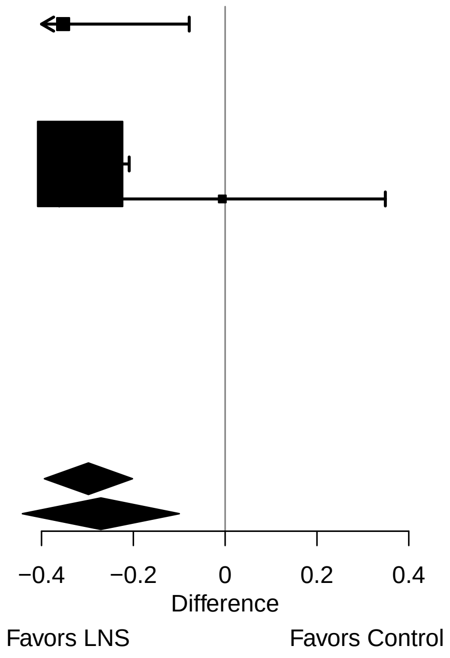

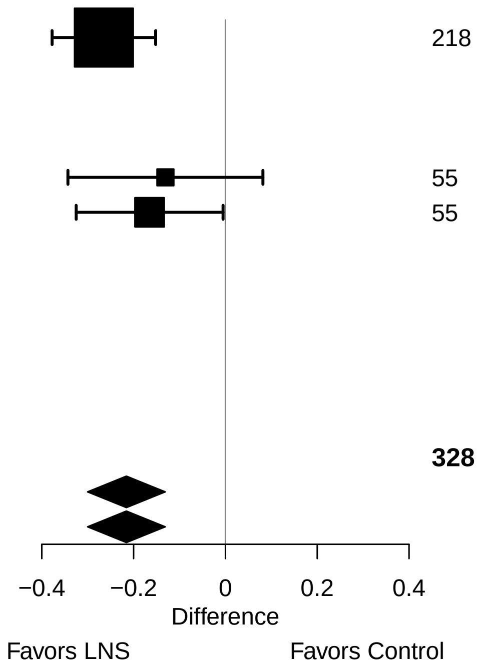

**Supplemental figure 7M: Elevated soluble transferrin receptor prevalence difference**

#### 7M4: Stratified by Household sanitation (insufficient comparisons)

**Supplemental figure 7M: Elevated soluble transferrin receptor prevalence difference**

#### 7M5: Stratified by Season at the time of assessment

**P−for−interaction = 0.684**
**Difference in PDs = 0.02 (−0.06, 0.10)**

| Country | Trial | Dry LNS N | Dry Control N | Dry Control Prevalence | Dry PD (95% CI) | Dry Fixed W | Dry Random W | Rainy LNS N | Rainy Control N | Rainy Control Prevalence | Rainy PD (95% CI) | Rainy Fixed W | Rainy Random W |
|---|---|---|---|---|---|---|---|---|---|---|---|---|---|
| Bangladesh | JiVitA−4 (35) | | | | | | | | | | | | |
| Bangladesh | RDNS (36) | 341 | 166 | 49.4 | −0.19 (−0.27, −0.10) | 0.36 | 0.36 | 208 | 106 | 50.9 | −0.09 (−0.20, 0.02) | 0.25 | 0.25 |
| Bangladesh | WASH−B (37) | 96 | 93 | 31.2 | −0.19 (−0.28, −0.09) | 0.29 | 0.29 | 116 | 85 | 31.8 | −0.24 (−0.35, −0.13) | 0.25 | 0.25 |
| Burkina Faso | iLiNS−Zinc (38) | 174 | 61 | 59.2 | −0.29 (−0.47, −0.11) | 0.08 | 0.08 | 126 | 30 | 70.0 | −0.37 (−0.59, −0.15) | 0.07 | 0.10 |
| Burkina Faso | PROMIS CS (39) | | | | | | | | | | | | |
| Ghana | GHANA (40) | 48 | 47 | 66.0 | −0.28 (−0.48, −0.09) | 0.07 | 0.07 | 36 | 35 | 57.1 | −0.24 (−0.47, −0.01) | 0.06 | 0.10 |
| Ghana | iLiNS−DYADG (41) | | | | | | | | | | | | |
| Kenya | WASH−B (42) | 88 | 75 | 72.0 | −0.27 (−0.38, −0.15) | 0.19 | 0.19 | 210 | 184 | 73.4 | −0.23 (−0.32, −0.14) | 0.37 | 0.30 |
| Madagascar | MAHAY (43) | | | | | | | | | | | | |
| Malawi | iLiNS−DYADM (44) | | | | | | | | | | | | |
| Malawi | iLiNS−DOSE (45) | | | | | | | | | | | | |
| Mali | PROMIS CS (46) | | | | | | | | | | | | |
| Zimbabwe | SHINE (HIV−) (47) | | | | | | | | | | | | |
| Zimbabwe | SHINE (HIV+) (48) | | | | | | | | | | | | |
| | | **747** | **442** | | **I² = 0.00, Tau² = 0.00** | | | **696** | **440** | | **I² = 0.40, Tau² = 0.00** | | |
| **Fixed** | | | | | **−0.22 (−0.27, −0.16)** | | | | | | **−0.21 (−0.26, −0.15)** | | |
| **Random** | | | | | **−0.22 (−0.27, −0.16)** | | | | | | **−0.21 (−0.29, −0.13)** | | |

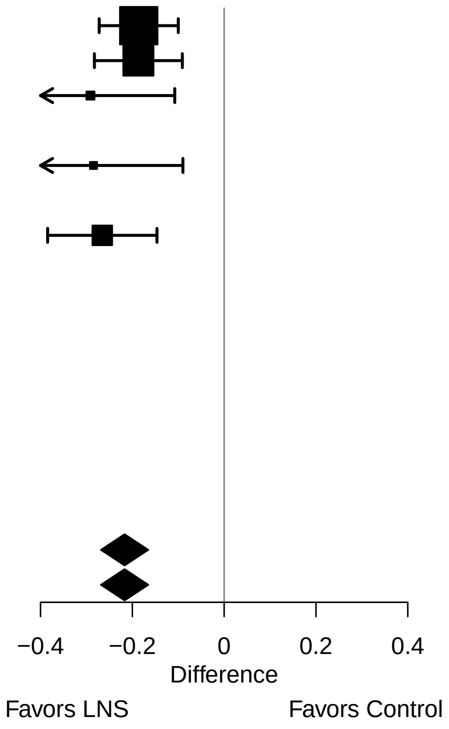

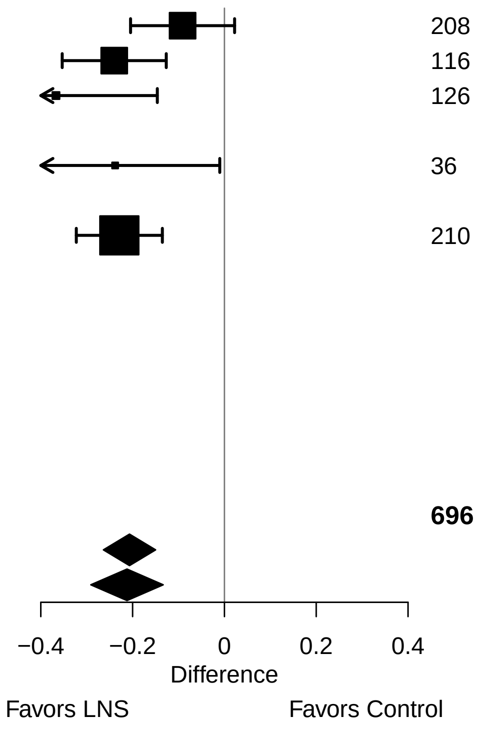

### Supplemental figure 7N: Geometric mean ratio of zinc protoporphyrin concentration

#### 7N1: Stratified by Household socio-economic status

**P−for−interaction = 0.157**
**Ratio of GMRs = 1.09 (0.97, 1.23)**

| Country | Trial | At least median: LNS N | Control N | Control Median | GMR (95% CI) | Fixed W | Random W | Less than median: LNS N | Control N | Control Median | GMR (95% CI) | Fixed W | Random W |
|---|---|---|---|---|---|---|---|---|---|---|---|---|---|
| Bangladesh | JiVitA−4 (35) | | | | | | | | | | | | |
| Bangladesh | RDNS (36) | | | | | | | | | | | | |
| Bangladesh | WASH−B (37) | | | | | | | | | | | | |
| Burkina Faso | iLiNS−Zinc (38) | 178 | 39 | 104.0 | 0.70 (0.57, 0.85) | 0.19 | 0.22 | 134 | 56 | 127.1 | 0.58 (0.49, 0.69) | 0.28 | 0.26 |
| Burkina Faso | PROMIS CS (39) | | | | | | | | | | | | |
| Ghana | GHANA (40) | | | | | | | | | | | | |
| Ghana | iLiNS−DYADG (41) | 58 | 109 | 47.1 | 0.81 (0.68, 0.96) | 0.26 | 0.26 | 42 | 93 | 49.3 | 0.95 (0.78, 1.15) | 0.20 | 0.25 |
| Kenya | WASH−B (42) | | | | | | | | | | | | |
| Madagascar | MAHAY (43) | | | | | | | | | | | | |
| Malawi | iLiNS−DYADM (44) | 106 | 186 | 56.0 | 0.83 (0.72, 0.96) | 0.37 | 0.31 | 84 | 189 | 51.7 | 0.90 (0.78, 1.04) | 0.38 | 0.27 |
| Malawi | iLiNS−DOSE (45) | 67 | 42 | 70.4 | 0.65 (0.53, 0.79) | 0.18 | 0.21 | 85 | 34 | 43.5 | 1.01 (0.80, 1.28) | 0.14 | 0.23 |
| Mali | PROMIS CS (46) | | | | | | | | | | | | |
| Zimbabwe | SHINE (HIV−) (47) | | | | | | | | | | | | |
| Zimbabwe | SHINE (HIV+) (48) | | | | | | | | | | | | |
| | | **409** | **376** | | **I² = 0.41, Tau² = 0.01** | | | **345** | **372** | | **I² = 0.87, Tau² = 0.05** | | |
| **Fixed** | | | | | **0.76 (0.70, 0.83)** | | | | | | **0.82 (0.75, 0.90)** | | |
| **Random** | | | | | **0.75 (0.67, 0.85)** | | | | | | **0.84 (0.65, 1.07)** | | |

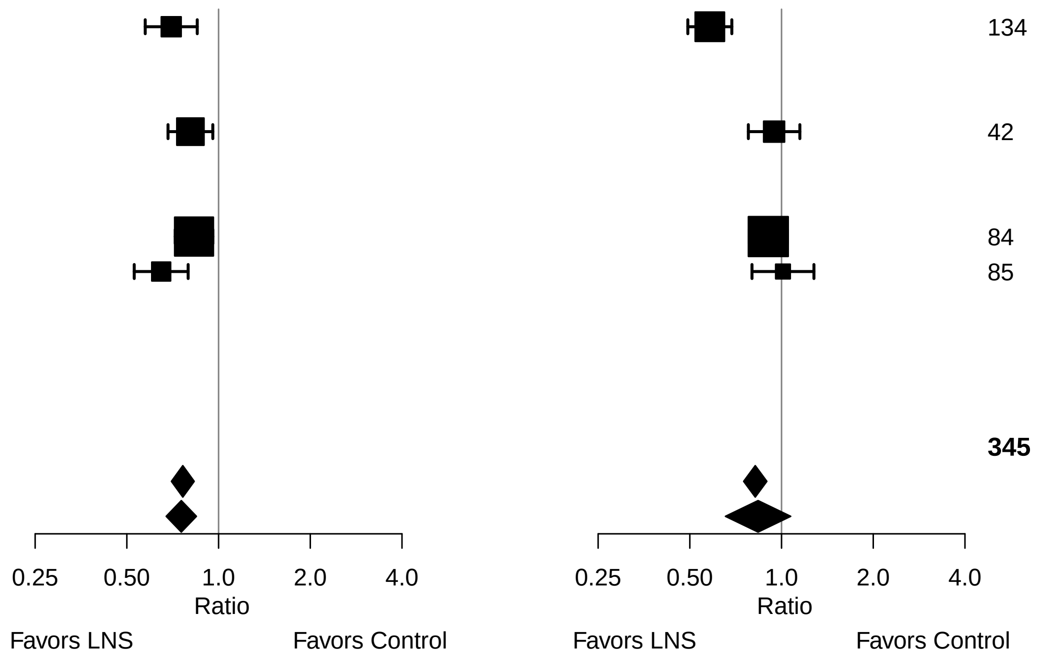

#### Supplemental figure 7N: Geometric mean ratio of zinc protoporphyrin concentration

#### 7N2: Stratified by Household food insecurity

**P−for−interaction = 0.193**
**Ratio of GMRs = 1.09 (0.96, 1.24)**

| | | Mild to secure | | | | | | Moderate to severe | | | | | |
|---|---|---|---|---|---|---|---|---|---|---|---|---|---|
| Country | Trial | LNS N | Control N | Control Median | GMR (95% CI) | Fixed W | Random W | LNS N | Control N | Control Median | GMR (95% CI) | Fixed W | Random W |
| Bangladesh | JiVitA−4 (35) | | | | | | | | | | | | |
| Bangladesh | RDNS (36) | | | | | | | | | | | | |
| Bangladesh | WASH−B (37) | | | | | | | | | | | | |
| Burkina Faso | iLiNS−Zinc (38) | 160 | 44 | 124.4 | 0.63 (0.54, 0.73) | 0.33 | 0.30 | 152 | 51 | 116.5 | 0.66 (0.54, 0.80) | 0.18 | 0.25 |
| Burkina Faso | PROMIS CS (39) | | | | | | | | | | | | |
| Ghana | GHANA (40) | | | | | | | | | | | | |
| Ghana | iLiNS−DYADG (41) | 79 | 146 | 47.8 | 0.83 (0.72, 0.95) | 0.39 | 0.32 | 21 | 56 | 48.6 | 1.01 (0.75, 1.37) | 0.08 | 0.17 |
| Kenya | WASH−B (42) | | | | | | | | | | | | |
| Madagascar | MAHAY (43) | | | | | | | | | | | | |
| Malawi | iLiNS−DYADM (44) | 57 | 100 | 56.2 | 0.81 (0.67, 0.98) | 0.21 | 0.25 | 132 | 270 | 52.7 | 0.88 (0.78, 0.99) | 0.50 | 0.32 |
| Malawi | iLiNS−DOSE (45) | 39 | 24 | 63.7 | 0.78 (0.56, 1.08) | 0.07 | 0.13 | 109 | 47 | 65.2 | 0.83 (0.70, 0.99) | 0.24 | 0.27 |
| Mali | PROMIS CS (46) | | | | | | | | | | | | |
| Zimbabwe | SHINE (HIV−) (47) | | | | | | | | | | | | |
| Zimbabwe | SHINE (HIV+) (48) | | | | | | | | | | | | |
| | | **335** | **314** | | **I² = 0.63, Tau² = 0.01** | | | **414** | **424** | | **I² = 0.62, Tau² = 0.02** | | |
| **Fixed** | | | | | **0.75 (0.69, 0.82)** | | | | | | **0.83 (0.77, 0.91)** | | |
| **Random** | | | | | **0.75 (0.66, 0.86)** | | | | | | **0.83 (0.70, 0.97)** | | |

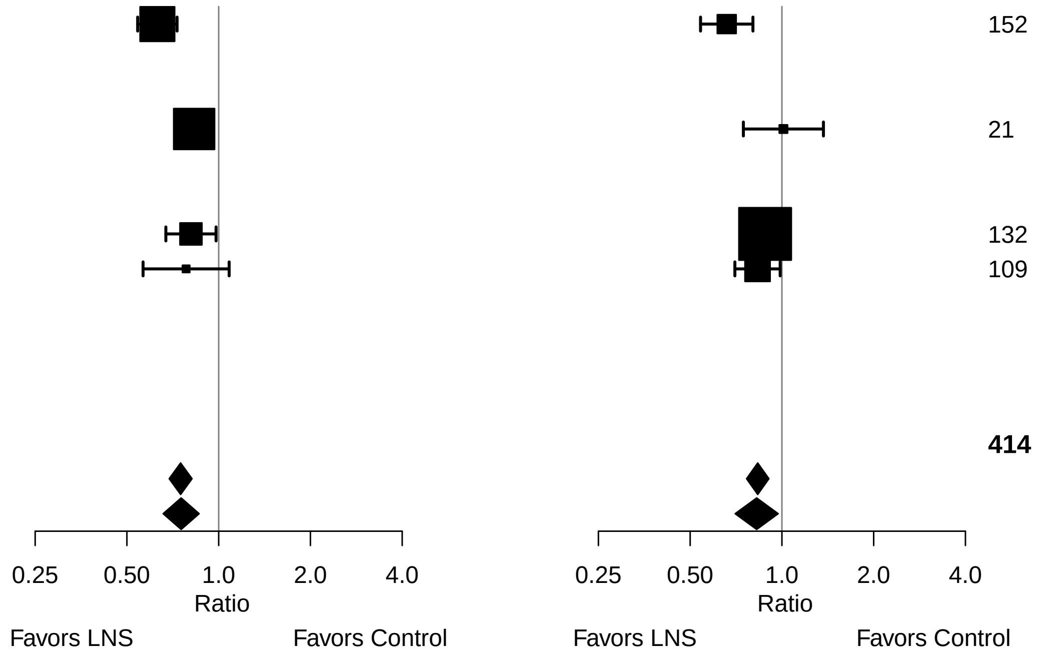

**Supplemental figure 7N: Geometric mean ratio of zinc protoporphyrin concentration**

#### 7N3: Stratified by Household source water quality (insufficient comparisons)

**Supplemental figure 7N: Geometric mean ratio of zinc protoporphyrin concentration**

#### 7N4: Stratified by Household sanitation (insufficient comparisons)

#### Supplemental figure 7N: Geometric mean ratio of zinc protoporphyrin concentration

##### 7N5: Stratified by Season at the time of assessment

**P−for−interaction = 0.079**
**Ratio of GMRs = 0.89 (0.77, 1.01)**

| Country | Trial | Dry LNS N | Dry Control N | Dry Control Median | Dry GMR (95% CI) | Dry Fixed W | Dry Random W | Rainy LNS N | Rainy Control N | Rainy Control Median | Rainy GMR (95% CI) | Rainy Fixed W | Rainy Random W |
|---|---|---|---|---|---|---|---|---|---|---|---|---|---|
| Bangladesh | JiVitA−4 (35) | | | | | | | | | | | | |
| Bangladesh | RDNS (36) | | | | | | | | | | | | |
| Bangladesh | WASH−B (37) | | | | | | | | | | | | |
| Burkina Faso | iLiNS−Zinc (38) | 173 | 60 | 127.5 | 0.63 (0.52, 0.76) | 0.18 | 0.23 | 124 | 30 | 77.5 | 0.67 (0.49, 0.90) | 0.12 | 0.16 |
| Burkina Faso | PROMIS CS (39) | | | | | | | | | | | | |
| Ghana | GHANA (40) | | | | | | | | | | | | |
| Ghana | iLiNS−DYADG (41) | 59 | 124 | 49.4 | 0.83 (0.70, 0.99) | 0.22 | 0.25 | 41 | 77 | 46.6 | 0.91 (0.75, 1.11) | 0.29 | 0.28 |
| Kenya | WASH−B (42) | | | | | | | | | | | | |
| Madagascar | MAHAY (43) | | | | | | | | | | | | |
| Malawi | iLiNS−DYADM (44) | 132 | 265 | 51.6 | 0.93 (0.83, 1.05) | 0.44 | 0.29 | 58 | 109 | 57.9 | 0.73 (0.61, 0.88) | 0.32 | 0.29 |
| Malawi | iLiNS−DOSE (45) | 103 | 46 | 65.6 | 0.87 (0.72, 1.06) | 0.17 | 0.23 | 78 | 41 | 74.0 | 0.68 (0.56, 0.83) | 0.27 | 0.27 |
| Mali | PROMIS CS (46) | | | | | | | | | | | | |
| Zimbabwe | SHINE (HIV−) (47) | | | | | | | | | | | | |
| Zimbabwe | SHINE (HIV+) (48) | | | | | | | | | | | | |
| | | **467** | **495** | | **I² = 0.75, Tau² = 0.02** | | | **301** | **257** | | **I² = 0.46, Tau² = 0.01** | | |
| **Fixed** | | | | | **0.84 (0.78, 0.91)** | | | | | | **0.76 (0.68, 0.84)** | | |
| **Random** | | | | | **0.82 (0.69, 0.96)** | | | | | | **0.75 (0.65, 0.87)** | | |

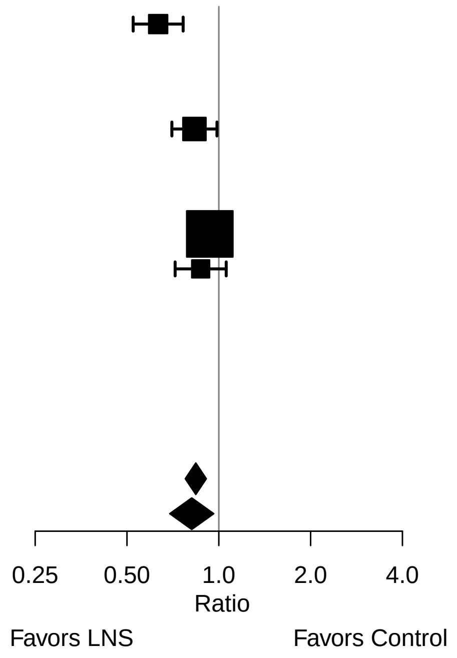

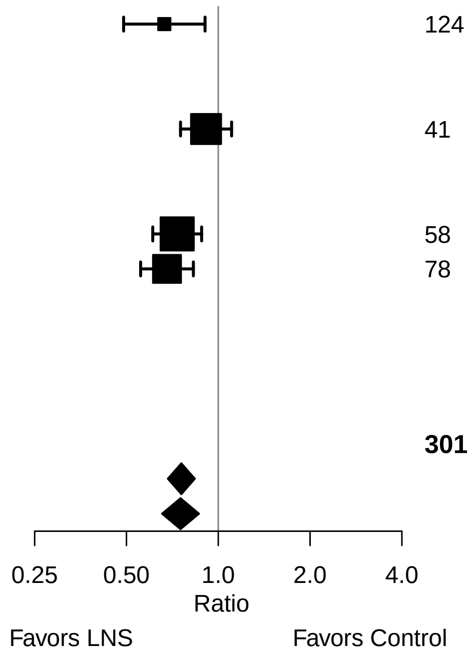

### Supplemental figure 7O: Elevated zinc protoporphyrin prevalence ratio

#### 7O1: Stratified by Household socio-economic status

**P−for−interaction = 0.894**
**Ratio of PRs = 0.98 (0.78, 1.24)**

| | | At least median | | | | | | Less than median | | | | | |
|---|---|---|---|---|---|---|---|---|---|---|---|---|---|
| Country | Trial | LNS N | Control N | Control Prevalence | PR (95% CI) | Fixed W | Random W | LNS N | Control N | Control Prevalence | PR (95% CI) | Fixed W | Random W |
| Bangladesh | JiVitA−4 (35) | | | | | | | | | | | | |
| Bangladesh | RDNS (36) | | | | | | | | | | | | |
| Bangladesh | WASH−B (37) | | | | | | | | | | | | |
| Burkina Faso | iLiNS−Zinc (38) | 178 | 39 | 72.1 | 0.77 (0.61, 0.96) | 0.62 | 0.60 | 134 | 56 | 77.4 | 0.63 (0.48, 0.82) | 0.54 | 0.54 |
| Burkina Faso | PROMIS CS (39) | | | | | | | | | | | | |
| Ghana | GHANA (40) | | | | | | | | | | | | |
| Ghana | iLiNS−DYADG (41) | 58 | 109 | 24.8 | 0.49 (0.23, 1.05) | 0.05 | 0.06 | 42 | 93 | 23.7 | 1.11 (0.59, 2.07) | 0.10 | 0.10 |
| Kenya | WASH−B (42) | | | | | | | | | | | | |
| Madagascar | MAHAY (43) | | | | | | | | | | | | |
| Malawi | iLiNS−DYADM (44) | 106 | 186 | 35.5 | 0.64 (0.43, 0.95) | 0.20 | 0.21 | 84 | 189 | 31.7 | 0.79 (0.51, 1.20) | 0.21 | 0.22 |
| Malawi | iLiNS−DOSE (45) | 67 | 42 | 50.0 | 0.51 (0.31, 0.84) | 0.13 | 0.14 | 85 | 34 | 35.3 | 0.77 (0.46, 1.30) | 0.14 | 0.15 |
| Mali | PROMIS CS (46) | | | | | | | | | | | | |
| Zimbabwe | SHINE (HIV−) (47) | | | | | | | | | | | | |
| Zimbabwe | SHINE (HIV+) (48) | | | | | | | | | | | | |
| | | **409** | **376** | | **I² = 0.06, Tau² = 0.00** | | | **345** | **372** | | **I² = 0.02, Tau² = 0.00** | | |
| **Fixed** | | | | | **0.69 (0.58, 0.82)** | | | | | | **0.72 (0.59, 0.87)** | | |
| **Random** | | | | | **0.68 (0.56, 0.82)** | | | | | | **0.72 (0.59, 0.88)** | | |

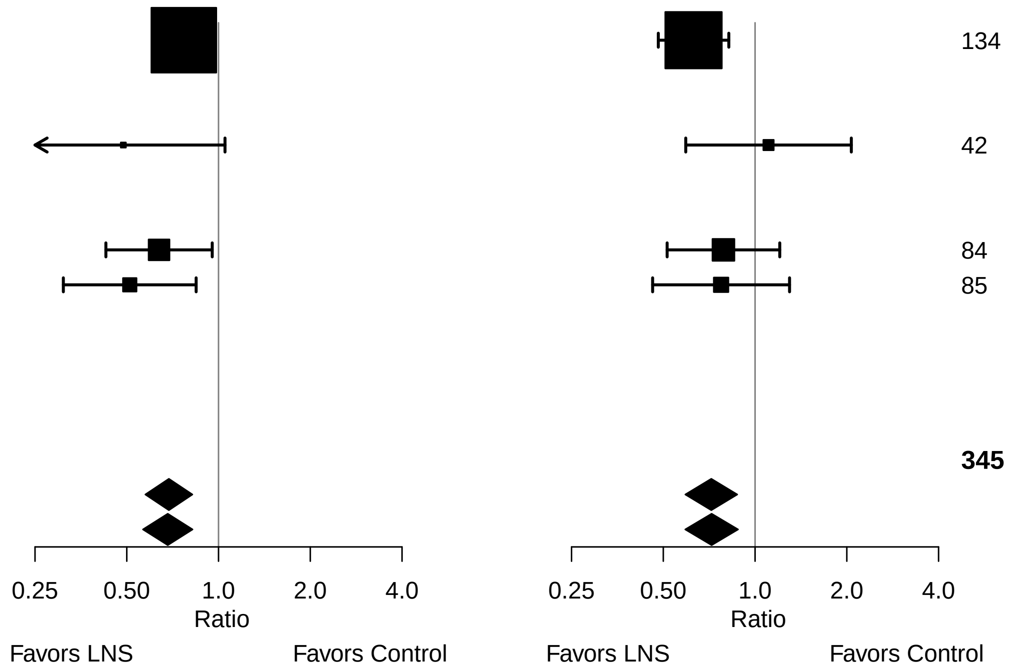

#### Supplemental figure 7O: Elevated zinc protoporphyrin prevalence ratio

### 7O2: Stratified by Household food insecurity

**P−for−interaction = 0.790**
**Ratio of PRs = 0.97 (0.76, 1.23)**

| Country | Trial | LNS N | Control N | Control Prevalence | Mild to secure PR (95% CI) | Fixed W | Random W | LNS N | Control N | Control Prevalence | Moderate to severe PR (95% CI) | Fixed W | Random W |
|---|---|---|---|---|---|---|---|---|---|---|---|---|---|
| Bangladesh | JiVitA−4 (35) | | | | | | | | | | | | |
| Bangladesh | RDNS (36) | | | | | | | | | | | | |
| Bangladesh | WASH−B (37) | | | | | | | | | | | | |
| Burkina Faso | iLiNS−Zinc (38) | 160 | 44 | 76.0 | 0.72 (0.58, 0.90) | 0.67 | 0.67 | 152 | 51 | 74.5 | 0.70 (0.55, 0.88) | 0.55 | 0.55 |
| Burkina Faso | PROMIS CS (39) | | | | | | | | | | | | |
| Ghana | GHANA (40) | | | | | | | | | | | | |
| Ghana | iLiNS−DYADG (41) | 79 | 146 | 20.5 | 0.74 (0.40, 1.36) | 0.09 | 0.09 | 21 | 56 | 33.9 | 0.84 (0.39, 1.82) | 0.05 | 0.05 |
| Kenya | WASH−B (42) | | | | | | | | | | | | |
| Madagascar | MAHAY (43) | | | | | | | | | | | | |
| Malawi | iLiNS−DYADM (44) | 57 | 100 | 38.0 | 0.69 (0.42, 1.14) | 0.13 | 0.13 | 132 | 270 | 32.2 | 0.68 (0.47, 0.98) | 0.23 | 0.23 |
| Malawi | iLiNS−DOSE (45) | 39 | 24 | 45.8 | 0.67 (0.39, 1.16) | 0.11 | 0.11 | 109 | 47 | 42.6 | 0.59 (0.39, 0.91) | 0.17 | 0.17 |
| Mali | PROMIS CS (46) | | | | | | | | | | | | |
| Zimbabwe | SHINE (HIV−) (47) | | | | | | | | | | | | |
| Zimbabwe | SHINE (HIV+) (48) | | | | | | | | | | | | |
| | | **335** | **314** | | **I² = 0.00, Tau² = 0.00** | | | **414** | **424** | | **I² = 0.00, Tau² = 0.00** | | |
| **Fixed** | | | | | **0.71 (0.59, 0.85)** | | | | | | **0.68 (0.57, 0.81)** | | |
| **Random** | | | | | **0.71 (0.59, 0.85)** | | | | | | **0.68 (0.57, 0.81)** | | |

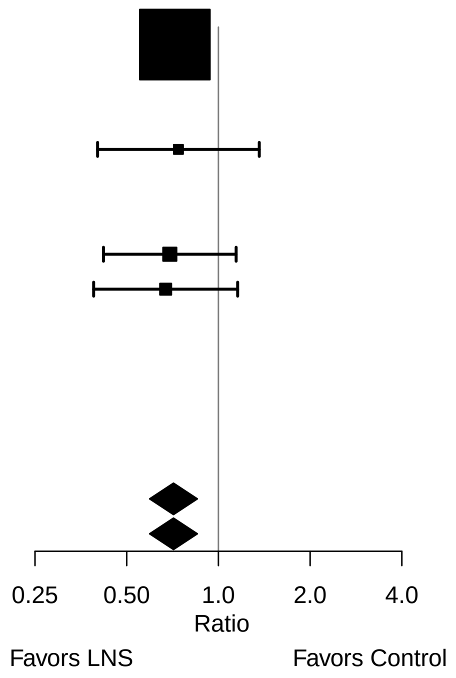

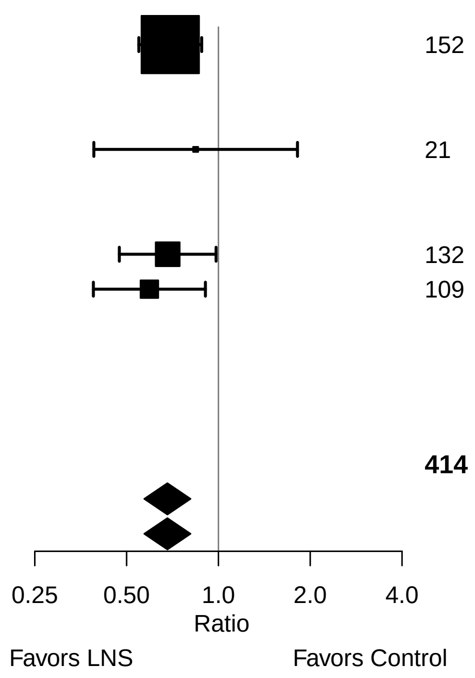

**Supplemental figure 7O: Elevated zinc protoporphyrin prevalence ratio**

#### 7O3: Stratified by Household source water quality (insufficient comparisons)

**Supplemental figure 7O: Elevated zinc protoporphyrin prevalence ratio**

#### 7O4: Stratified by Household sanitation (insufficient comparisons)

**Supplemental figure 7O: Elevated zinc protoporphyrin prevalence ratio**

#### 7O5: Stratified by Season at the time of assessment

**P−for−interaction = 0.680**
**Ratio of PRs = 0.94 (0.69, 1.28)**

| Country | Trial | Dry LNS N | Dry Control N | Dry Control Prevalence | Dry PR (95% CI) | Dry Fixed W | Dry Random W | Rainy LNS N | Rainy Control N | Rainy Control Prevalence | Rainy PR (95% CI) | Rainy Fixed W | Rainy Random W |
|---|---|---|---|---|---|---|---|---|---|---|---|---|---|
| Bangladesh | JiVitA−4 (35) | | | | | | | | | | | | |
| Bangladesh | RDNS (36) | | | | | | | | | | | | |
| Bangladesh | WASH−B (37) | | | | | | | | | | | | |
| Burkina Faso | iLiNS−Zinc (38) | 173 | 60 | 82.9 | 0.72 (0.58, 0.88) | 0.60 | 0.60 | 124 | 30 | 63.3 | 0.75 (0.48, 1.15) | 0.33 | 0.29 |
| Burkina Faso | PROMIS CS (39) | | | | | | | | | | | | |
| Ghana | GHANA (40) | | | | | | | | | | | | |
| Ghana | iLiNS−DYADG (41) | 59 | 124 | 25.0 | 0.41 (0.18, 0.92) | 0.04 | 0.04 | 41 | 77 | 23.4 | 1.25 (0.67, 2.34) | 0.16 | 0.20 |
| Kenya | WASH−B (42) | | | | | | | | | | | | |
| Madagascar | MAHAY (43) | | | | | | | | | | | | |
| Malawi | iLiNS−DYADM (44) | 132 | 265 | 33.2 | 0.75 (0.54, 1.06) | 0.22 | 0.22 | 58 | 109 | 33.9 | 0.61 (0.35, 1.08) | 0.19 | 0.23 |
| Malawi | iLiNS−DOSE (45) | 103 | 46 | 39.1 | 0.78 (0.51, 1.19) | 0.14 | 0.14 | 78 | 41 | 51.2 | 0.49 (0.31, 0.76) | 0.31 | 0.28 |
| Mali | PROMIS CS (46) | | | | | | | | | | | | |
| Zimbabwe | SHINE (HIV−) (47) | | | | | | | | | | | | |
| Zimbabwe | SHINE (HIV+) (48) | | | | | | | | | | | | |
| | | **467** | **495** | | **$I^2$ = 0.00, $Tau^2$ = 0.00** | | | **301** | **257** | | **$I^2$ = 0.51, $Tau^2$ = 0.08** | | |
| **Fixed** | | | | | **0.72 (0.61, 0.84)** | | | | | | **0.68 (0.53, 0.87)** | | |
| **Random** | | | | | **0.72 (0.61, 0.84)** | | | | | | **0.70 (0.48, 1.03)** | | |

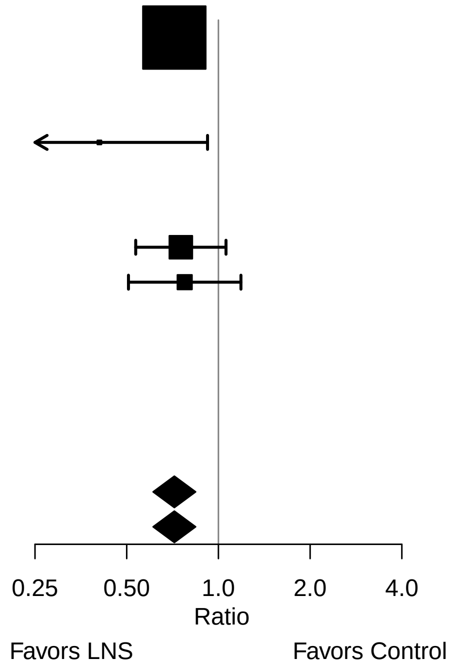

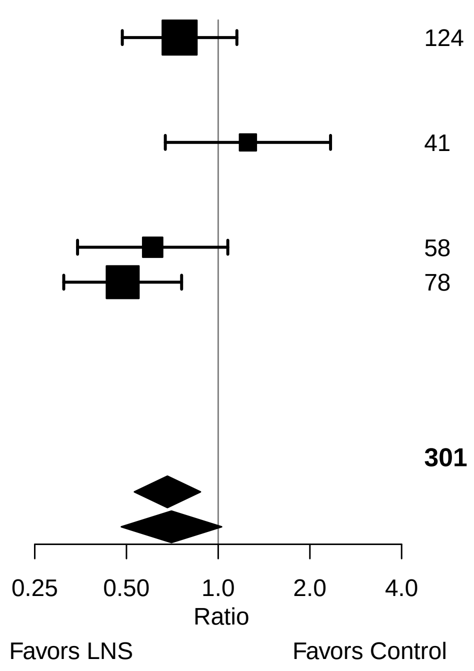

### Supplemental figure 7P: Elevated zinc protoporphyrin prevalence difference

#### 7P1: Stratified by Household socio-economic status

**P−for−interaction = 0.191**
**Difference in PDs = 0.06 (−0.03, 0.16)**

| | | At least median | | | | | | Less than median | | | | | |
|---|---|---|---|---|---|---|---|---|---|---|---|---|---|
| Country | Trial | LNS N | Control N | Control Prevalence | PD (95% CI) | Fixed W | Random W | LNS N | Control N | Control Prevalence | PD (95% CI) | Fixed W | Random W |
| Bangladesh | JiVitA−4 (35) | | | | | | | | | | | | |
| Bangladesh | RDNS (36) | | | | | | | | | | | | |
| Bangladesh | WASH−B (37) | | | | | | | | | | | | |
| Burkina Faso | iLiNS−Zinc (38) | 178 | 39 | 72.1 | −0.17 (−0.33, −0.02) | 0.18 | 0.18 | 134 | 56 | 77.4 | −0.28 (−0.42, −0.14) | 0.26 | 0.26 |
| Burkina Faso | PROMIS CS (39) | | | | | | | | | | | | |
| Ghana | GHANA (40) | | | | | | | | | | | | |
| Ghana | iLiNS−DYADG (41) | 58 | 109 | 24.8 | −0.13 (−0.25, 0.00) | 0.28 | 0.28 | 42 | 93 | 23.7 | 0.03 (−0.13, 0.18) | 0.20 | 0.24 |
| Kenya | WASH−B (42) | | | | | | | | | | | | |
| Madagascar | MAHAY (43) | | | | | | | | | | | | |
| Malawi | iLiNS−DYADM (44) | 106 | 186 | 35.5 | −0.13 (−0.24, −0.02) | 0.38 | 0.38 | 84 | 189 | 31.7 | −0.07 (−0.19, 0.05) | 0.37 | 0.28 |
| Malawi | iLiNS−DOSE (45) | 67 | 42 | 50.0 | −0.25 (−0.42, −0.09) | 0.17 | 0.17 | 85 | 34 | 35.3 | −0.08 (−0.25, 0.09) | 0.17 | 0.22 |
| Mali | PROMIS CS (46) | | | | | | | | | | | | |
| Zimbabwe | SHINE (HIV−) (47) | | | | | | | | | | | | |
| Zimbabwe | SHINE (HIV+) (48) | | | | | | | | | | | | |
| | | **409** | **376** | | **I² = 0.00, Tau² = 0.00** | | | **345** | **372** | | **I² = 0.66, Tau² = 0.01** | | |
| **Fixed** | | | | | **−0.16 (−0.22, −0.09)** | | | | | | **−0.10 (−0.18, −0.03)** | | |
| **Random** | | | | | **−0.16 (−0.22, −0.09)** | | | | | | **−0.10 (−0.23, 0.02)** | | |

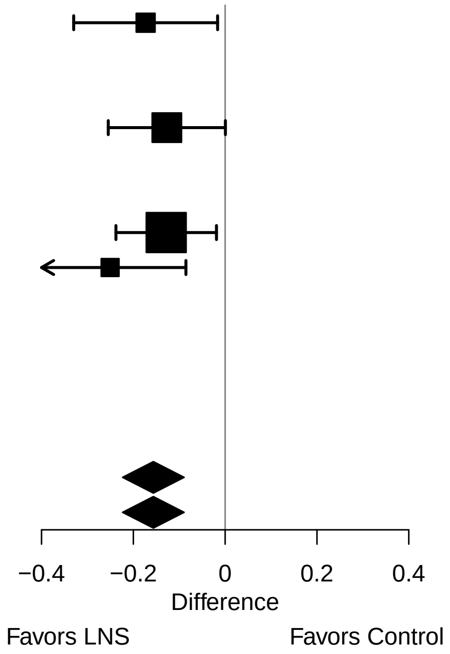

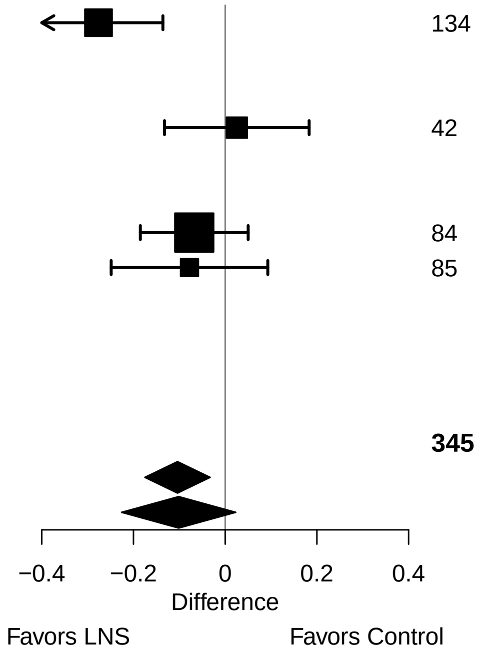

**Supplemental figure 7P: Elevated zinc protoporphyrin prevalence difference**

#### 7P2: Stratified by Household food insecurity

**P−for−interaction = 0.945**
**Difference in PDs = 0.00 (−0.11, 0.10)**

| | | Mild to secure | | | | | | Moderate to severe | | | | | |
|---|---|---|---|---|---|---|---|---|---|---|---|---|---|
| Country | Trial | LNS N | Control N | Control Prevalence | PD (95% CI) | Fixed W | Random W | LNS N | Control N | Control Prevalence | PD (95% CI) | Fixed W | Random W |
| Bangladesh | JiVitA−4 (35) | | | | | | | | | | | | |
| Bangladesh | RDNS (36) | | | | | | | | | | | | |
| Bangladesh | WASH−B (37) | | | | | | | | | | | | |
| Burkina Faso | iLiNS−Zinc (38) | 160 | 44 | 76.0 | −0.21 (−0.35, −0.07) | 0.26 | 0.26 | 152 | 51 | 74.5 | −0.23 (−0.38, −0.08) | 0.19 | 0.19 |
| Burkina Faso | PROMIS CS (39) | | | | | | | | | | | | |
| Ghana | GHANA (40) | | | | | | | | | | | | |
| Ghana | iLiNS−DYADG (41) | 79 | 146 | 20.5 | −0.05 (−0.16, 0.05) | 0.44 | 0.43 | 21 | 56 | 33.9 | −0.05 (−0.29, 0.18) | 0.08 | 0.08 |
| Kenya | WASH−B (42) | | | | | | | | | | | | |
| Madagascar | MAHAY (43) | | | | | | | | | | | | |
| Malawi | iLiNS−DYADM (44) | 57 | 100 | 38.0 | −0.12 (−0.27, 0.04) | 0.21 | 0.22 | 132 | 270 | 32.2 | −0.10 (−0.20, −0.01) | 0.50 | 0.50 |
| Malawi | iLiNS−DOSE (45) | 39 | 24 | 45.8 | −0.15 (−0.38, 0.08) | 0.09 | 0.10 | 109 | 47 | 42.6 | −0.18 (−0.32, −0.04) | 0.23 | 0.23 |
| Mali | PROMIS CS (46) | | | | | | | | | | | | |
| Zimbabwe | SHINE (HIV−) (47) | | | | | | | | | | | | |
| Zimbabwe | SHINE (HIV+) (48) | | | | | | | | | | | | |
| | | **335** | **314** | | **I² = 0.06, Tau² = 0.00** | | | **414** | **424** | | **I² = 0.00, Tau² = 0.00** | | |
| **Fixed** | | | | | **−0.12 (−0.19, −0.05)** | | | | | | **−0.14 (−0.21, −0.07)** | | |
| **Random** | | | | | **−0.12 (−0.19, −0.05)** | | | | | | **−0.14 (−0.21, −0.07)** | | |

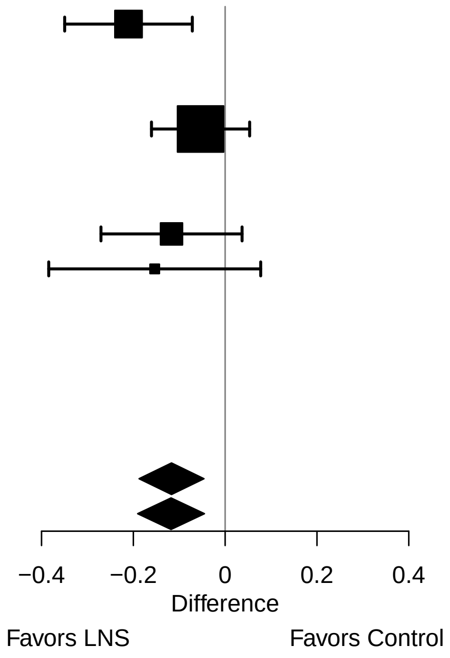

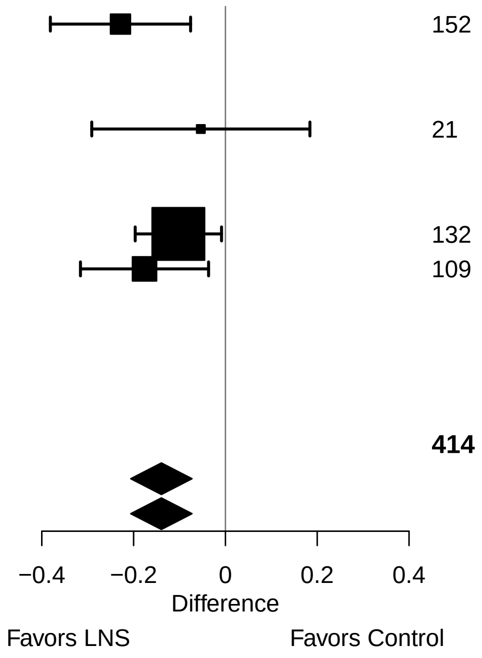

**Supplemental figure 7P: Elevated zinc protoporphyrin prevalence difference**

#### 7P3: Stratified by Household source water quality (insufficient comparisons)

**Supplemental figure 7P: Elevated zinc protoporphyrin prevalence difference**

#### 7P4: Stratified by Household sanitation (insufficient comparisons)

#### Supplemental figure 7P: Elevated zinc protoporphyrin prevalence difference

#### 7P5: Stratified by Season at the time of assessment

**P−for−interaction = 0.874**
**Difference in PDs = 0.01 (−0.09, 0.11)**

| | | Dry | | | | | | Rainy | | | | | |
|---|---|---|---|---|---|---|---|---|---|---|---|---|---|
| Country | Trial | LNS N | Control N | Control Prevalence | PD (95% CI) | Fixed W | Random W | LNS N | Control N | Control Prevalence | PD (95% CI) | Fixed W | Random W |
| Bangladesh | JiVitA−4 (35) | | | | | | | | | | | | |
| Bangladesh | RDNS (36) | | | | | | | | | | | | |
| Bangladesh | WASH−B (37) | | | | | | | | | | | | |
| Burkina Faso | iLiNS−Zinc (38) | 173 | 60 | 82.9 | −0.23 (−0.38, −0.09) | 0.18 | 0.20 | 124 | 30 | 63.3 | −0.17 (−0.41, 0.07) | 0.12 | 0.18 |
| Burkina Faso | PROMIS CS (39) | | | | | | | | | | | | |
| Ghana | GHANA (40) | | | | | | | | | | | | |
| Ghana | iLiNS−DYADG (41) | 59 | 124 | 25.0 | −0.15 (−0.27, −0.02) | 0.25 | 0.25 | 41 | 77 | 23.4 | 0.06 (−0.11, 0.22) | 0.25 | 0.26 |
| Kenya | WASH−B (42) | | | | | | | | | | | | |
| Madagascar | MAHAY (43) | | | | | | | | | | | | |
| Malawi | iLiNS−DYADM (44) | 132 | 265 | 33.2 | −0.08 (−0.18, 0.01) | 0.40 | 0.38 | 58 | 109 | 33.9 | −0.13 (−0.28, 0.01) | 0.34 | 0.29 |
| Malawi | iLiNS−DOSE (45) | 103 | 46 | 39.1 | −0.08 (−0.23, 0.07) | 0.17 | 0.18 | 78 | 41 | 51.2 | −0.26 (−0.42, −0.11) | 0.29 | 0.27 |
| Mali | PROMIS CS (46) | | | | | | | | | | | | |
| Zimbabwe | SHINE (HIV−) (47) | | | | | | | | | | | | |
| Zimbabwe | SHINE (HIV+) (48) | | | | | | | | | | | | |
| | | **467** | **495** | | **I² = 0.14, Tau² = 0.00** | | | **301** | **257** | | **I² = 0.62, Tau² = 0.01** | | |
| **Fixed** | | | | | **−0.13 (−0.19, −0.07)** | | | | | | **−0.13 (−0.21, −0.04)** | | |
| **Random** | | | | | **−0.13 (−0.20, −0.06)** | | | | | | **−0.13 (−0.26, 0.01)** | | |

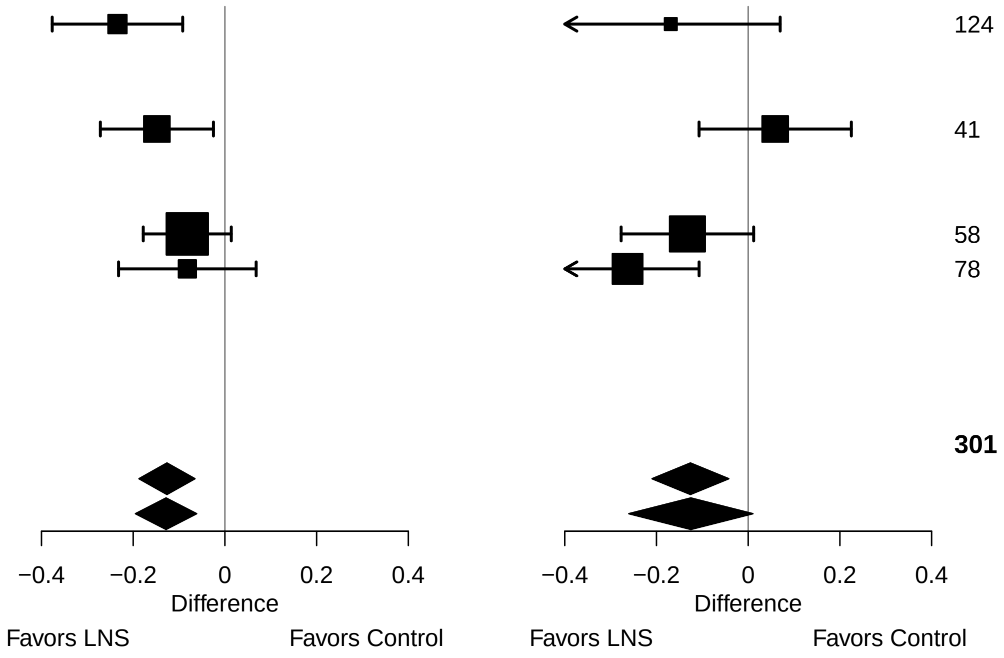

### Supplemental figure 7Q: Geometric mean ratio of plasma zinc concentration

#### 7Q1: Stratified by Household socio-economic status

**P−for−interaction = 0.696**
**Ratio of GMRs = 0.99 (0.94, 1.04)**

| Country | Trial | LNS N | Control N | Control Median | At least median GMR (95% CI) | Fixed W | Random W | LNS N | Control N | Control Median | Less than median GMR (95% CI) | Fixed W | Random W |
|---|---|---|---|---|---|---|---|---|---|---|---|---|---|
| Bangladesh | JiVitA−4 (35) | 242 | 78 | 88.3 | 0.98 (0.92, 1.05) | 0.27 | 0.27 | 209 | 66 | 88.7 | 0.94 (0.87, 1.03) | 0.20 | 0.20 |
| Bangladesh | RDNS (36) | | | | | | | | | | | | |
| Bangladesh | WASH−B (37) | | | | | | | | | | | | |
| Burkina Faso | iLiNS−Zinc (38) | 180 | 39 | 67.8 | 1.01 (0.96, 1.06) | 0.55 | 0.55 | 134 | 57 | 68.8 | 1.00 (0.96, 1.05) | 0.69 | 0.69 |
| Burkina Faso | PROMIS CS (39) | | | | | | | | | | | | |
| Ghana | GHANA (40) | 34 | 34 | 61.0 | 0.99 (0.91, 1.08) | 0.18 | 0.18 | 24 | 31 | 58.9 | 1.00 (0.89, 1.12) | 0.11 | 0.11 |
| Ghana | iLiNS−DYADG (41) | | | | | | | | | | | | |
| Kenya | WASH−B (42) | | | | | | | | | | | | |
| Madagascar | MAHAY (43) | | | | | | | | | | | | |
| Malawi | iLiNS−DYADM (44) | | | | | | | | | | | | |
| Malawi | iLiNS−DOSE (45) | | | | | | | | | | | | |
| Mali | PROMIS CS (46) | | | | | | | | | | | | |
| Zimbabwe | SHINE (HIV−) (47) | | | | | | | | | | | | |
| Zimbabwe | SHINE (HIV+) (48) | | | | | | | | | | | | |
| | | **456** | **151** | | **I² = 0.00, Tau² = 0.00** | | | **367** | **154** | | **I² = 0.00, Tau² = 0.00** | | |
| **Fixed** | | | | | **1.00 (0.96, 1.03)** | | | | | | **0.99 (0.95, 1.03)** | | |
| **Random** | | | | | **1.00 (0.96, 1.03)** | | | | | | **0.99 (0.95, 1.03)** | | |

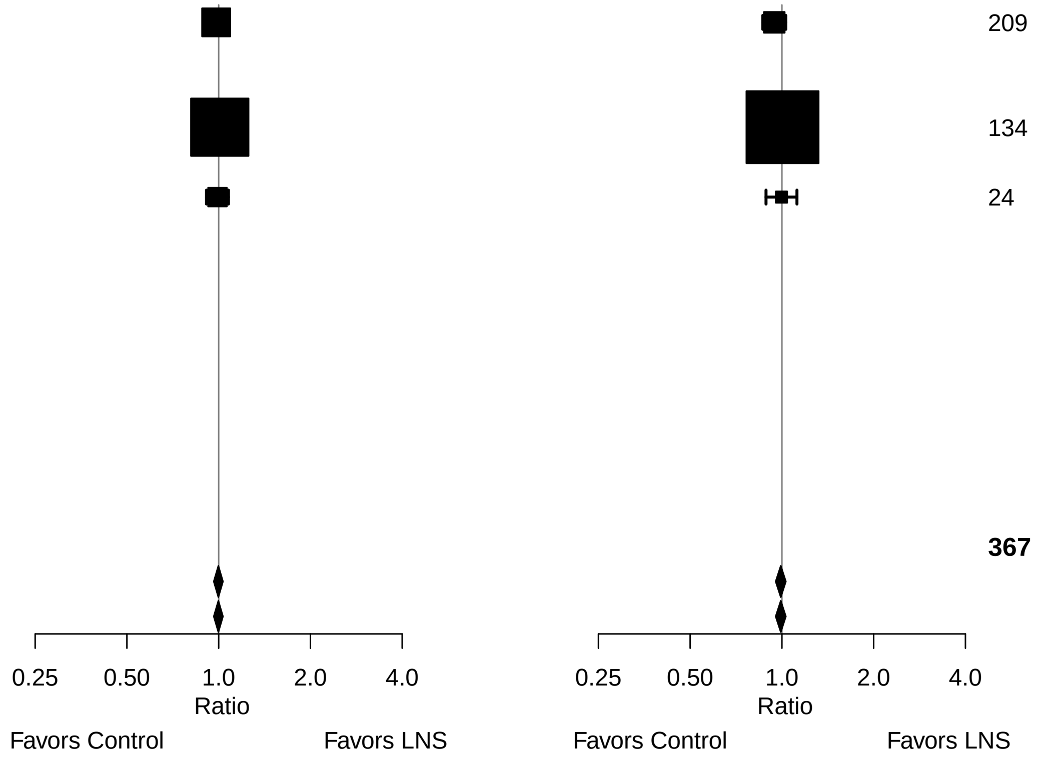

**Supplemental figure 7Q: Geometric mean ratio of plasma zinc concentration**

#### 7Q2: Stratified by Household food insecurity (insufficient comparisons)

**Supplemental figure 7Q: Geometric mean ratio of plasma zinc concentration**

#### 7Q3: Stratified by Household source water quality (insufficient comparisons)

**Supplemental figure 7Q: Geometric mean ratio of plasma zinc concentration**

#### 7Q4: Stratified by Household sanitation (insufficient comparisons)

**Supplemental figure 7Q: Geometric mean ratio of plasma zinc concentration**

#### 7Q5: Stratified by Season at the time of assessment

**P−for−interaction = 0.044**
**Ratio of GMRs = 1.04 (1.00, 1.08)**

| Country | Trial | Dry: LNS N | Dry: Control N | Dry: Control Median | Dry: GMR (95% CI) | Dry: Fixed W | Dry: Random W | Rainy: LNS N | Rainy: Control N | Rainy: Control Median | Rainy: GMR (95% CI) | Rainy: Fixed W | Rainy: Random W |
|---|---|---|---|---|---|---|---|---|---|---|---|---|---|
| Bangladesh | JiVitA−4 (35) | 261 | 71 | 88.0 | 0.98 (0.93, 1.04) | 0.30 | 0.30 | 191 | 73 | 88.9 | 0.95 (0.87, 1.03) | 0.19 | 0.29 |
| Bangladesh | RDNS (36) | | | | | | | | | | | | |
| Bangladesh | WASH−B (37) | | | | | | | | | | | | |
| Burkina Faso | iLiNS−Zinc (38) | 174 | 61 | 68.7 | 0.99 (0.95, 1.03) | 0.61 | 0.61 | 125 | 30 | 67.3 | 1.05 (1.00, 1.10) | 0.64 | 0.45 |
| Burkina Faso | PROMIS CS (39) | | | | | | | | | | | | |
| Ghana | GHANA (40) | 30 | 36 | 59.7 | 1.03 (0.93, 1.14) | 0.08 | 0.08 | 28 | 33 | 60.0 | 0.98 (0.90, 1.07) | 0.17 | 0.27 |
| Ghana | iLiNS−DYADG (41) | | | | | | | | | | | | |
| Kenya | WASH−B (42) | | | | | | | | | | | | |
| Madagascar | MAHAY (43) | | | | | | | | | | | | |
| Malawi | iLiNS−DYADM (44) | | | | | | | | | | | | |
| Malawi | iLiNS−DOSE (45) | | | | | | | | | | | | |
| Mali | PROMIS CS (46) | | | | | | | | | | | | |
| Zimbabwe | SHINE (HIV−) (47) | | | | | | | | | | | | |
| Zimbabwe | SHINE (HIV+) (48) | | | | | | | | | | | | |
| | | **465** | **168** | | **$I^2$ = 0.00, $Tau^2$ = 0.00** | | | **344** | **136** | | **$I^2$ = 0.62, $Tau^2$ = 0.00** | | |
| **Fixed** | | | | | **0.99 (0.96, 1.02)** | | | | | | **1.02 (0.98, 1.06)** | | |
| **Random** | | | | | **0.99 (0.96, 1.02)** | | | | | | **1.00 (0.94, 1.07)** | | |

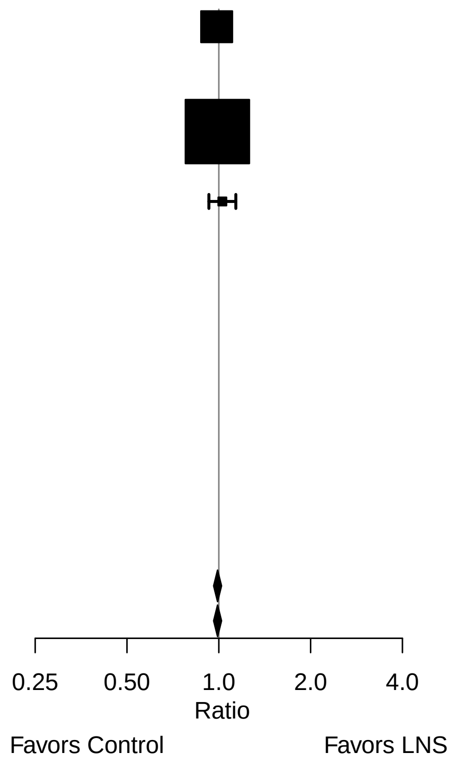

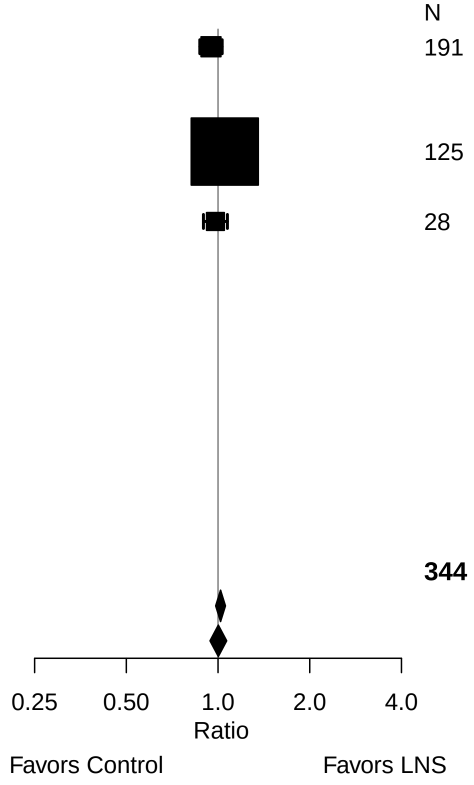

### Supplemental figure 7R: Geometric mean ratio of retinol concentration

#### 7R1: Stratified by Household socio-economic status

**P−for−interaction = 0.882**
**Ratio of GMRs = 1.01 (0.94, 1.08)**

| | | **At least median** | | | | | | **Less than median** | | | | | |
|---|---|---|---|---|---|---|---|---|---|---|---|---|---|
| | | **LNS** | **Control** | **Control** | **GMR** | **Fixed** | **Random** | **LNS** | **Control** | **Control** | **GMR** | **Fixed** | **Random** |
| Country | Trial | N | N | Median | (95% CI) | W | W | N | N | Median | (95% CI) | W | W |
| Bangladesh | JiVitA−4 (35) | 232 | 78 | 1.4 | 1.07 (1.00, 1.14) | 0.50 | 0.50 | 201 | 61 | 1.3 | 1.10 (1.00, 1.21) | 0.36 | 0.34 |
| Bangladesh | RDNS (36) | | | | | | | | | | | | |
| Bangladesh | WASH−B (37) | | | | | | | | | | | | |
| Burkina Faso | iLiNS−Zinc (38) | | | | | | | | | | | | |
| Burkina Faso | PROMIS CS (39) | | | | | | | | | | | | |
| Ghana | GHANA (40) | 38 | 37 | 1.1 | 1.08 (0.90, 1.29) | 0.07 | 0.07 | 25 | 33 | 1.0 | 1.04 (0.83, 1.29) | 0.07 | 0.08 |
| Ghana | iLiNS−DYADG (41) | 56 | 106 | 1.0 | 1.04 (0.96, 1.13) | 0.31 | 0.31 | 40 | 90 | 1.0 | 0.96 (0.87, 1.06) | 0.32 | 0.31 |
| Kenya | WASH−B (42) | | | | | | | | | | | | |
| Madagascar | MAHAY (43) | | | | | | | | | | | | |
| Malawi | iLiNS−DYADM (44) | 36 | 73 | 1.0 | 0.97 (0.85, 1.11) | 0.13 | 0.13 | 35 | 90 | 1.1 | 1.05 (0.94, 1.17) | 0.26 | 0.26 |
| Malawi | iLiNS−DOSE (45) | | | | | | | | | | | | |
| Mali | PROMIS CS (46) | | | | | | | | | | | | |
| Zimbabwe | SHINE (HIV−) (47) | | | | | | | | | | | | |
| Zimbabwe | SHINE (HIV+) (48) | | | | | | | | | | | | |
| | | **362** | **294** | | **I² = 0.00, Tau² = 0.00** | | | **301** | **274** | | **I² = 0.22, Tau² = 0.00** | | |
| **Fixed** | | | | | **1.05 (1.00, 1.10)** | | | | | | **1.04 (0.98, 1.10)** | | |
| **Random** | | | | | **1.05 (1.00, 1.10)** | | | | | | **1.04 (0.97, 1.10)** | | |

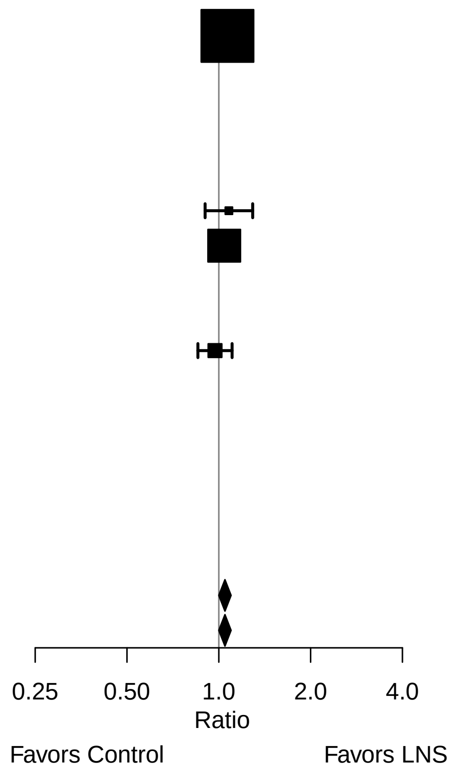

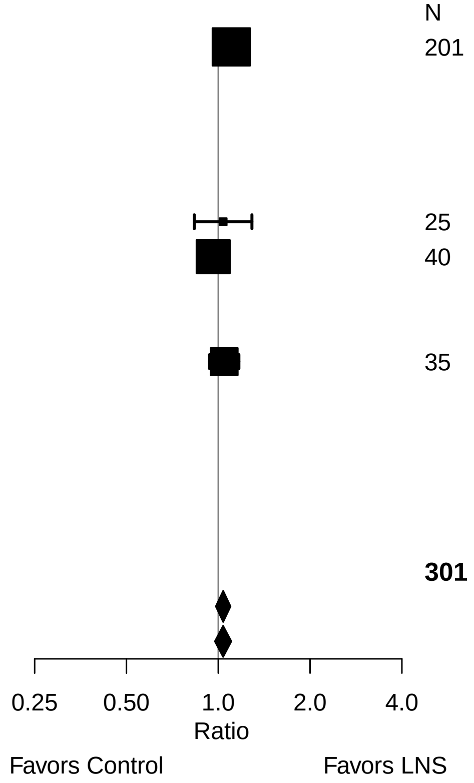

#### Supplemental figure 7R: Geometric mean ratio of retinol concentration

#### 7R2: Stratified by Household food insecurity

**P−for−interaction = 0.084**
**Ratio of GMRs = 0.93 (0.85, 1.01)**

| | | Mild to secure | | | | | | Moderate to severe | | | | | |
|---|---|---|---|---|---|---|---|---|---|---|---|---|---|
| | | **LNS** | **Control** | **Control** | **GMR** | **Fixed** | **Random** | **LNS** | **Control** | **Control** | **GMR** | **Fixed** | **Random** |
| Country | Trial | N | N | Median | (95% CI) | W | W | N | N | Median | (95% CI) | W | W |
| Bangladesh | JiVitA−4 (35) | 320 | 106 | 1.3 | 1.12 (1.04, 1.20) | 0.46 | 0.44 | 113 | 33 | 1.4 | 0.98 (0.88, 1.10) | 0.32 | 0.32 |
| Bangladesh | RDNS (36) | | | | | | | | | | | | |
| Bangladesh | WASH−B (37) | | | | | | | | | | | | |
| Burkina Faso | iLiNS−Zinc (38) | | | | | | | | | | | | |
| Burkina Faso | PROMIS CS (39) | | | | | | | | | | | | |
| Ghana | GHANA (40) | | | | | | | | | | | | |
| Ghana | iLiNS−DYADG (41) | 75 | 144 | 1.0 | 1.01 (0.94, 1.09) | 0.46 | 0.44 | 21 | 52 | 1.0 | 1.00 (0.88, 1.13) | 0.26 | 0.26 |
| Kenya | WASH−B (42) | | | | | | | | | | | | |
| Madagascar | MAHAY (43) | | | | | | | | | | | | |
| Malawi | iLiNS−DYADM (44) | 19 | 34 | 1.1 | 1.06 (0.88, 1.28) | 0.08 | 0.11 | 51 | 127 | 1.0 | 0.98 (0.89, 1.08) | 0.42 | 0.42 |
| Malawi | iLiNS−DOSE (45) | | | | | | | | | | | | |
| Mali | PROMIS CS (46) | | | | | | | | | | | | |
| Zimbabwe | SHINE (HIV−) (47) | | | | | | | | | | | | |
| Zimbabwe | SHINE (HIV+) (48) | | | | | | | | | | | | |
| | | **414** | **284** | | **I² = 0.43, Tau² = 0.00** | | | **185** | **212** | | **I² = 0.00, Tau² = 0.00** | | |
| **Fixed** | | | | | **1.06 (1.01, 1.12)** | | | | | | **0.99 (0.93, 1.05)** | | |
| **Random** | | | | | **1.06 (0.99, 1.14)** | | | | | | **0.99 (0.93, 1.05)** | | |

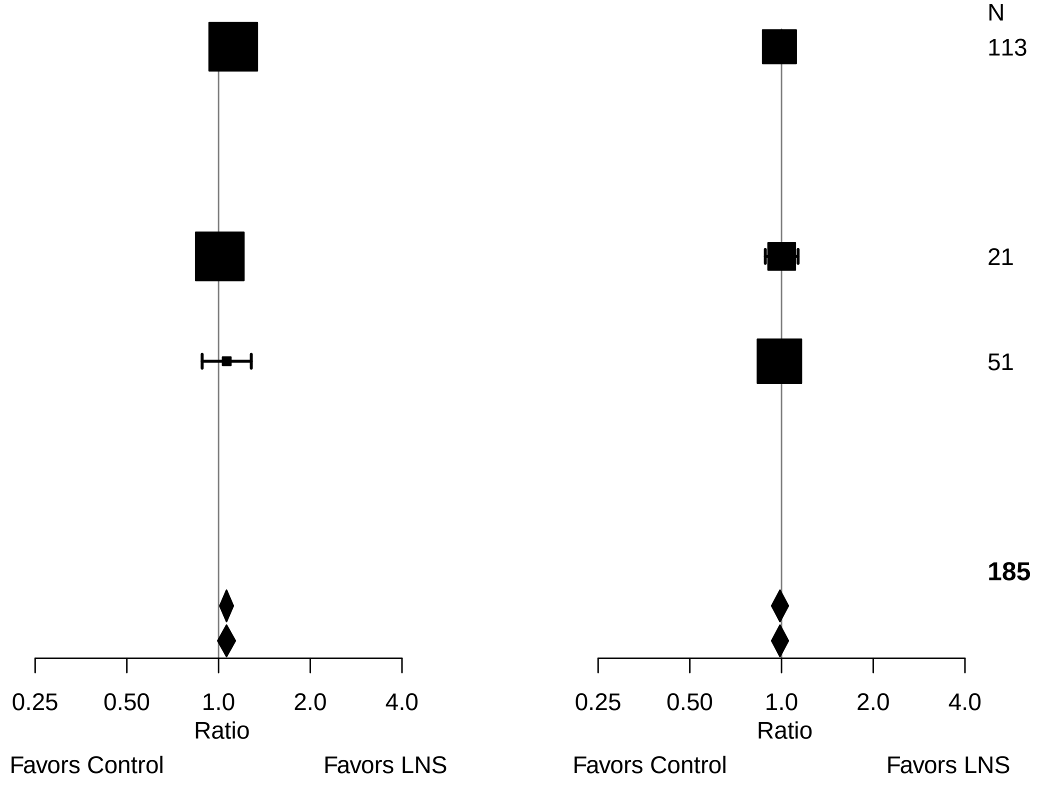

**Supplemental figure 7R: Geometric mean ratio of retinol concentration**

#### 7R3: Stratified by Household source water quality (insufficient comparisons)

**Supplemental figure 7R: Geometric mean ratio of retinol concentration**

#### 7R4: Stratified by Household sanitation (insufficient comparisons)

#### Supplemental figure 7R: Geometric mean ratio of retinol concentration

##### 7R5: Stratified by Season at the time of assessment

**P−for−interaction = 0.952**
**Ratio of GMRs = 1.00 (0.93, 1.08)**

| Country | Trial | **Dry** LNS N | Control N | Control Median | GMR (95% CI) | Fixed W | Random W | **Rainy** LNS N | Control N | Control Median | GMR (95% CI) | Fixed W | Random W |
|---|---|---|---|---|---|---|---|---|---|---|---|---|---|
| Bangladesh | JiVitA−4 (35) | 257 | 69 | 1.4 | 1.10 (1.00, 1.21) | 0.30 | 0.28 | 177 | 70 | 1.3 | 1.04 (0.96, 1.13) | 0.48 | 0.30 |
| Bangladesh | RDNS (36) | | | | | | | | | | | | |
| Bangladesh | WASH−B (37) | | | | | | | | | | | | |
| Burkina Faso | iLiNS−Zinc (38) | | | | | | | | | | | | |
| Burkina Faso | PROMIS CS (39) | | | | | | | | | | | | |
| Ghana | GHANA (40) | 35 | 41 | 0.9 | 1.15 (0.97, 1.36) | 0.09 | 0.16 | 28 | 33 | 1.4 | 0.99 (0.84, 1.16) | 0.11 | 0.21 |
| Ghana | iLiNS−DYADG (41) | 59 | 122 | 1.0 | 1.02 (0.94, 1.11) | 0.35 | 0.29 | 37 | 73 | 1.0 | 0.99 (0.90, 1.09) | 0.30 | 0.28 |
| Kenya | WASH−B (42) | | | | | | | | | | | | |
| Madagascar | MAHAY (43) | | | | | | | | | | | | |
| Malawi | iLiNS−DYADM (44) | 54 | 120 | 1.1 | 0.92 (0.84, 1.02) | 0.27 | 0.27 | 17 | 43 | 1.0 | 1.30 (1.10, 1.52) | 0.11 | 0.21 |
| Malawi | iLiNS−DOSE (45) | | | | | | | | | | | | |
| Mali | PROMIS CS (46) | | | | | | | | | | | | |
| Zimbabwe | SHINE (HIV−) (47) | | | | | | | | | | | | |
| Zimbabwe | SHINE (HIV+) (48) | | | | | | | | | | | | |
| | | **405** | **352** | | **I² = 0.64, Tau² = 0.01** | | | **259** | **219** | | **I² = 0.65, Tau² = 0.01** | | |
| **Fixed** | | | | | **1.03 (0.98, 1.08)** | | | | | | **1.04 (0.99, 1.10)** | | |
| **Random** | | | | | **1.03 (0.95, 1.13)** | | | | | | **1.06 (0.94, 1.20)** | | |

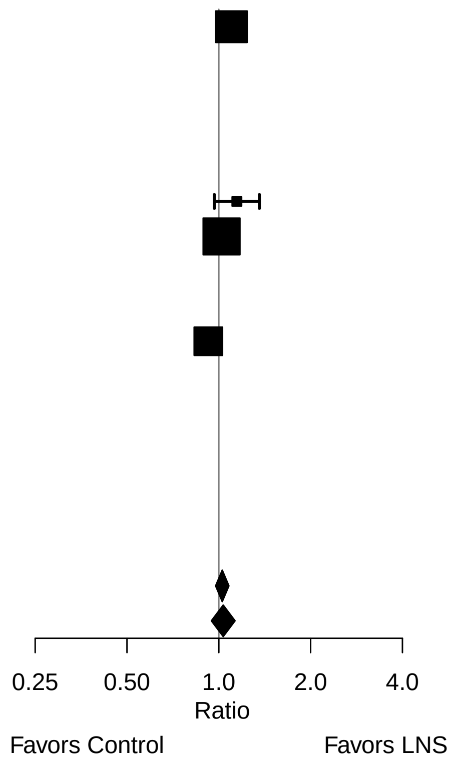

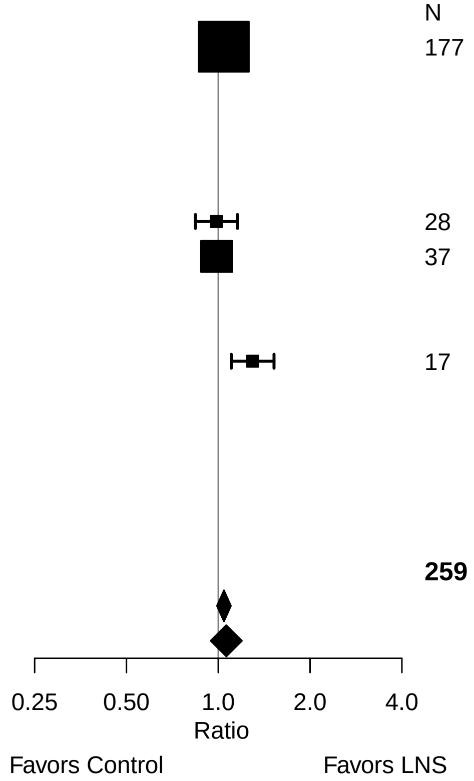

### Supplemental figure 7S: Low vitamin A (retinol < 0.70 µmol/L) prevalence ratio

#### 7S1: Stratified by Household socio-economic status (insufficient comparisons)

**Supplemental figure 7S: Low vitamin A (retinol < 0.70 µmol/L) prevalence ratio**

#### 7S2: Stratified by Household food insecurity (insufficient comparisons)

**Supplemental figure 7S: Low vitamin A (retinol < 0.70 µmol/L) prevalence ratio**

#### 7S3: Stratified by Household source water quality (insufficient comparisons)

**Supplemental figure 7S: Low vitamin A (retinol < 0.70 µmol/L) prevalence ratio**

#### 7S4: Stratified by Household sanitation (insufficient comparisons)

**Supplemental figure 7S: Low vitamin A (retinol < 0.70 µmol/L) prevalence ratio**

#### 7S5: Stratified by Season at the time of assessment (insufficient comparisons)

#### Supplemental figure 7T: Low vitamin A (retinol < 0.70 µmol/L) prevalence difference

##### 7T1: Stratified by Household socio-economic status (insufficient comparisons)

**Supplemental figure 7T: Low vitamin A (retinol < 0.70 µmol/L) prevalence difference**

#### 7T2: Stratified by Household food insecurity (insufficient comparisons)

**Supplemental figure 7T: Low vitamin A (retinol < 0.70 µmol/L) prevalence difference**

#### 7T3: Stratified by Household source water quality (insufficient comparisons)

**Supplemental figure 7T: Low vitamin A (retinol < 0.70 µmol/L) prevalence difference**

#### 7T4: Stratified by Household sanitation (insufficient comparisons)

**Supplemental figure 7T: Low vitamin A (retinol < 0.70 µmol/L) prevalence difference**

#### 7T5: Stratified by Season at the time of assessment (insufficient comparisons)

### Supplemental figure 7U: Marginal vitamin A (retinol < 1.05 µmol/L) prevalence ratio

#### 7U1: Stratified by Household socio-economic status

**P−for−interaction = 0.579**
**Ratio of PRs = 1.08 (0.82, 1.44)**

| | | At least median | | | | | | Less than median | | | | | |
|---|---|---|---|---|---|---|---|---|---|---|---|---|---|
| Country | Trial | LNS N | Control N | Control Prevalence | PR (95% CI) | Fixed W | Random W | LNS N | Control N | Control Prevalence | PR (95% CI) | Fixed W | Random W |
| Bangladesh | JiVitA−4 (35) | 232 | 78 | 17.9 | 0.67 (0.42, 1.09) | 0.16 | 0.19 | 201 | 61 | 21.3 | 0.79 (0.43, 1.48) | 0.11 | 0.11 |
| Bangladesh | RDNS (36) | | | | | | | | | | | | |
| Bangladesh | WASH−B (37) | | | | | | | | | | | | |
| Burkina Faso | iLiNS−Zinc (38) | | | | | | | | | | | | |
| Burkina Faso | PROMIS CS (39) | | | | | | | | | | | | |
| Ghana | GHANA (40) | 38 | 37 | 45.9 | 1.03 (0.64, 1.67) | 0.16 | 0.19 | 25 | 33 | 51.5 | 1.01 (0.61, 1.67) | 0.17 | 0.17 |
| Ghana | iLiNS−DYADG (41) | 56 | 106 | 55.7 | 0.83 (0.60, 1.16) | 0.34 | 0.31 | 40 | 90 | 57.8 | 1.04 (0.76, 1.41) | 0.45 | 0.45 |
| Kenya | WASH−B (42) | | | | | | | | | | | | |
| Madagascar | MAHAY (43) | | | | | | | | | | | | |
| Malawi | iLiNS−DYADM (44) | 36 | 73 | 52.1 | 1.23 (0.88, 1.71) | 0.34 | 0.31 | 35 | 90 | 45.6 | 1.13 (0.76, 1.67) | 0.27 | 0.27 |
| Malawi | iLiNS−DOSE (45) | | | | | | | | | | | | |
| Mali | PROMIS CS (46) | | | | | | | | | | | | |
| Zimbabwe | SHINE (HIV−) (47) | | | | | | | | | | | | |
| Zimbabwe | SHINE (HIV+) (48) | | | | | | | | | | | | |
| | | **362** | **294** | | **I² = 0.40, Tau² = 0.03** | | | **301** | **274** | | **I² = 0.00, Tau² = 0.00** | | |
| **Fixed** | | | | | **0.95 (0.78, 1.15)** | | | | | | **1.03 (0.84, 1.26)** | | |
| **Random** | | | | | **0.94 (0.73, 1.21)** | | | | | | **1.03 (0.84, 1.26)** | | |

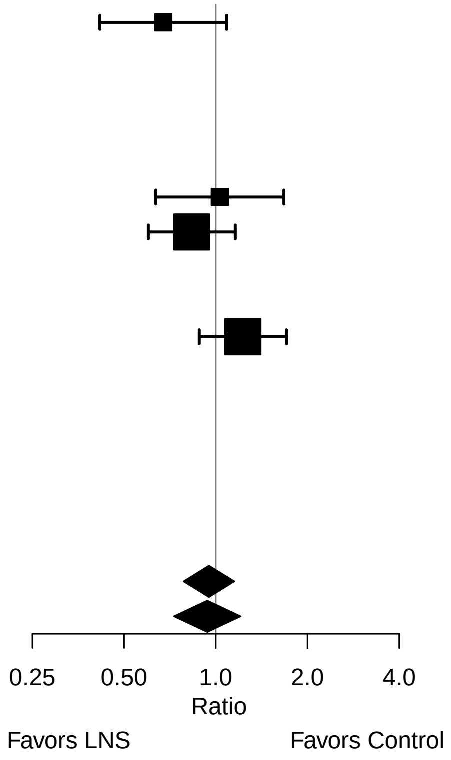

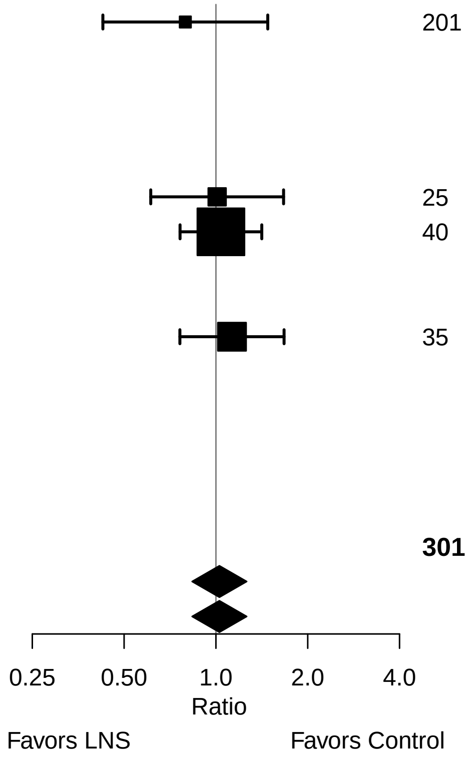

**Supplemental figure 7U: Marginal vitamin A (retinol < 1.05 µmol/L) prevalence ratio**

#### 7U2: Stratified by Household food insecurity (insufficient comparisons)

**Supplemental figure 7U: Marginal vitamin A (retinol < 1.05 µmol/L) prevalence ratio**

#### 7U3: Stratified by Household source water quality (insufficient comparisons)

**Supplemental figure 7U: Marginal vitamin A (retinol < 1.05 µmol/L) prevalence ratio**

#### 7U4: Stratified by Household sanitation (insufficient comparisons)

**Supplemental figure 7U: Marginal vitamin A (retinol < 1.05 µmol/L) prevalence ratio**

#### 7U5: Stratified by Season at the time of assessment

**P−for−interaction = 0.615**
**Ratio of PRs = 1.09 (0.77, 1.54)**

| | | Dry | | | | | | Rainy | | | | | |
|---|---|---|---|---|---|---|---|---|---|---|---|---|---|
| | | **LNS** | **Control** | **Control** | **PR** | **Fixed** | **Random** | **LNS** | **Control** | **Control** | **PR** | **Fixed** | **Random** |
| Country | Trial | N | N | Prevalence | (95% CI) | W | W | N | N | Prevalence | (95% CI) | W | W |
| Bangladesh | JiVitA−4 (35) | 257 | 69 | 15.9 | 0.61 (0.33, 1.13) | 0.07 | 0.16 | 177 | 70 | 22.9 | 0.91 (0.51, 1.64) | 0.21 | 0.23 |
| Bangladesh | RDNS (36) | | | | | | | | | | | | |
| Bangladesh | WASH−B (37) | | | | | | | | | | | | |
| Burkina Faso | iLiNS−Zinc (38) | | | | | | | | | | | | |
| Burkina Faso | PROMIS CS (39) | | | | | | | | | | | | |
| Ghana | GHANA (40) | 35 | 41 | 70.7 | 0.93 (0.68, 1.27) | 0.27 | 0.27 | 28 | 33 | 21.2 | 1.35 (0.56, 3.25) | 0.09 | 0.11 |
| Ghana | iLiNS−DYADG (41) | 59 | 122 | 59.8 | 0.82 (0.61, 1.11) | 0.30 | 0.28 | 37 | 73 | 52.1 | 1.09 (0.76, 1.56) | 0.57 | 0.52 |
| Kenya | WASH−B (42) | | | | | | | | | | | | |
| Madagascar | MAHAY (43) | | | | | | | | | | | | |
| Malawi | iLiNS−DYADM (44) | 54 | 120 | 45.8 | 1.45 (1.11, 1.91) | 0.36 | 0.29 | 17 | 43 | 55.8 | 0.53 (0.24, 1.15) | 0.12 | 0.14 |
| Malawi | iLiNS−DOSE (45) | | | | | | | | | | | | |
| Mali | PROMIS CS (46) | | | | | | | | | | | | |
| Zimbabwe | SHINE (HIV−) (47) | | | | | | | | | | | | |
| Zimbabwe | SHINE (HIV+) (48) | | | | | | | | | | | | |
| | | **405** | **352** | | **I² = 0.74, Tau² = 0.09** | | | **259** | **219** | | **I² = 0.09, Tau² = 0.01** | | |
| **Fixed** | | | | | **1.02 (0.87, 1.20)** | | | | | | **0.98 (0.75, 1.29)** | | |
| **Random** | | | | | **0.95 (0.68, 1.34)** | | | | | | **0.97 (0.72, 1.31)** | | |

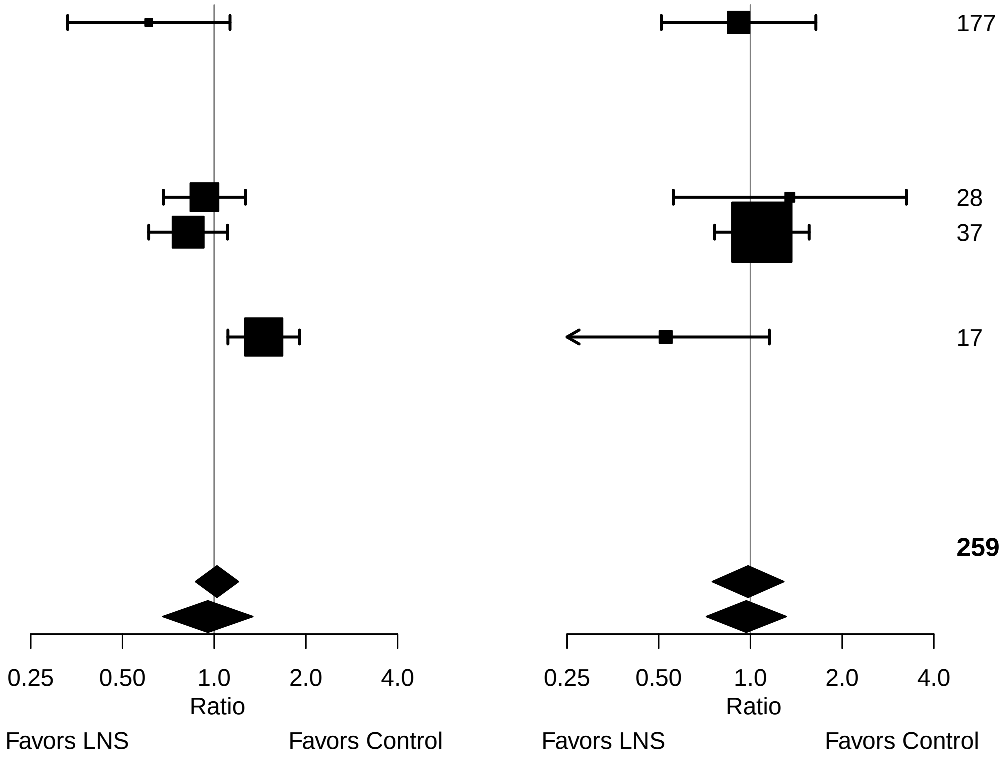

### Supplemental figure 7V: Marginal vitamin A (retinol < 1.05 µmol/L) prevalence difference

#### 7V1: Stratified by Household socio-economic status

**P−for−interaction = 0.712**
**Difference in PDs = 0.02 (−0.09, 0.13)**

| Country | Trial | At least median: LNS N | Control N | Control Prevalence | PD (95% CI) | Fixed W | Random W | Less than median: LNS N | Control N | Control Prevalence | PD (95% CI) | Fixed W | Random W |
|---|---|---|---|---|---|---|---|---|---|---|---|---|---|
| Bangladesh | JiVitA−4 (35) | 232 | 78 | 17.9 | −0.06 (−0.13, 0.01) | 0.70 | 0.62 | 201 | 61 | 21.3 | −0.04 (−0.17, 0.08) | 0.47 | 0.47 |
| Bangladesh | RDNS (36) | | | | | | | | | | | | |
| Bangladesh | WASH−B (37) | | | | | | | | | | | | |
| Burkina Faso | iLiNS−Zinc (38) | | | | | | | | | | | | |
| Burkina Faso | PROMIS CS (39) | | | | | | | | | | | | |
| Ghana | GHANA (40) | 38 | 37 | 45.9 | 0.01 (−0.21, 0.24) | 0.07 | 0.09 | 25 | 33 | 51.5 | 0.00 (−0.26, 0.27) | 0.11 | 0.11 |
| Ghana | iLiNS−DYADG (41) | 56 | 106 | 55.7 | −0.09 (−0.25, 0.07) | 0.14 | 0.17 | 40 | 90 | 57.8 | 0.02 (−0.16, 0.21) | 0.22 | 0.22 |
| Kenya | WASH−B (42) | | | | | | | | | | | | |
| Madagascar | MAHAY (43) | | | | | | | | | | | | |
| Malawi | iLiNS−DYADM (44) | 36 | 73 | 52.1 | 0.12 (−0.08, 0.32) | 0.09 | 0.12 | 35 | 90 | 45.6 | 0.06 (−0.14, 0.25) | 0.20 | 0.20 |
| Malawi | iLiNS−DOSE (45) | | | | | | | | | | | | |
| Mali | PROMIS CS (46) | | | | | | | | | | | | |
| Zimbabwe | SHINE (HIV−) (47) | | | | | | | | | | | | |
| Zimbabwe | SHINE (HIV+) (48) | | | | | | | | | | | | |
| | | **362** | **294** | | **I² = 0.09, Tau² = 0.00** | | | **301** | **274** | | **I² = 0.00, Tau² = 0.00** | | |
| **Fixed** | | | | | **−0.04 (−0.10, 0.02)** | | | | | | **0.00 (−0.09, 0.08)** | | |
| **Random** | | | | | **−0.04 (−0.11, 0.03)** | | | | | | **0.00 (−0.09, 0.08)** | | |

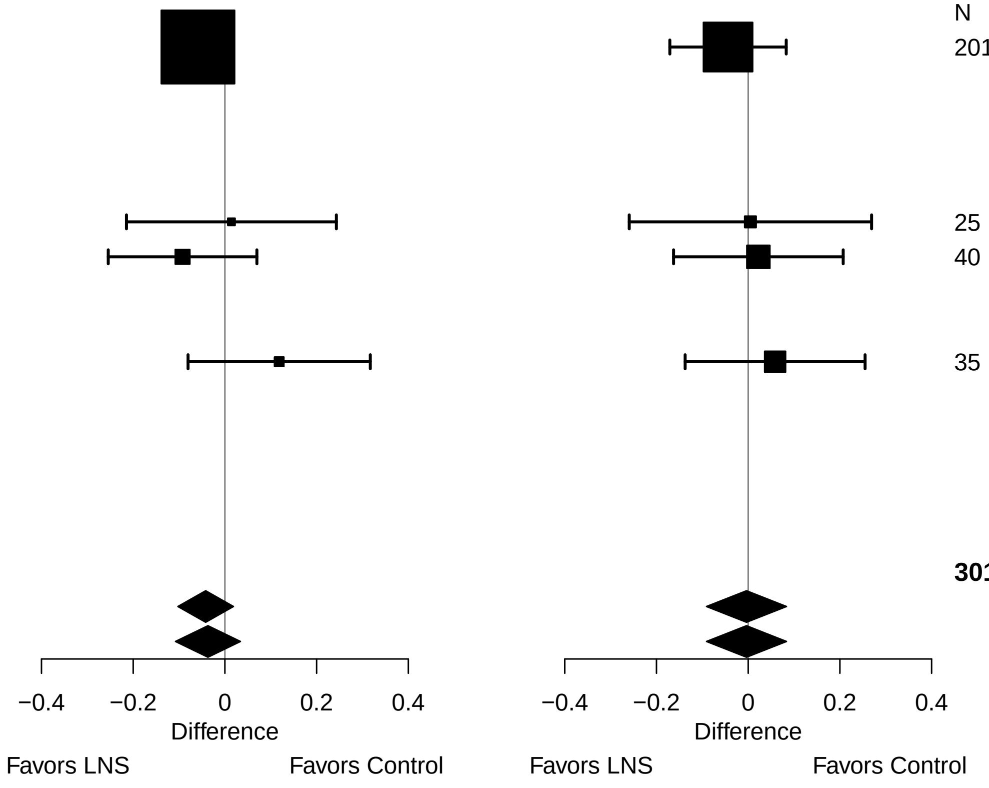

**Supplemental figure 7V: Marginal vitamin A (retinol < 1.05 µmol/L) prevalence difference**

#### 7V2: Stratified by Household food insecurity (insufficient comparisons)

**Supplemental figure 7V: Marginal vitamin A (retinol < 1.05 µmol/L) prevalence difference**

#### 7V3: Stratified by Household source water quality (insufficient comparisons)

**Supplemental figure 7V: Marginal vitamin A (retinol < 1.05 µmol/L) prevalence difference**

#### 7V4: Stratified by Household sanitation (insufficient comparisons)

#### Supplemental figure 7V: Marginal vitamin A (retinol < 1.05 μmol/L) prevalence difference

#### 7V5: Stratified by Season at the time of assessment

**P−for−interaction = 0.888**
**Difference in PDs = 0.01 (−0.10, 0.12)**

| Country | Trial | Dry LNS N | Dry Control N | Dry Control Prevalence | Dry PD (95% CI) | Dry Fixed W | Dry Random W | Rainy LNS N | Rainy Control N | Rainy Control Prevalence | Rainy PD (95% CI) | Rainy Fixed W | Rainy Random W |
|---|---|---|---|---|---|---|---|---|---|---|---|---|---|
| Bangladesh | JiVitA−4 (35) | 257 | 69 | 15.9 | −0.06 (−0.15, 0.03) | 0.55 | 0.31 | 177 | 70 | 22.9 | −0.02 (−0.15, 0.11) | 0.49 | 0.38 |
| Bangladesh | RDNS (36) | | | | | | | | | | | | |
| Bangladesh | WASH−B (37) | | | | | | | | | | | | |
| Burkina Faso | iLiNS−Zinc (38) | | | | | | | | | | | | |
| Burkina Faso | PROMIS CS (39) | | | | | | | | | | | | |
| Ghana | GHANA (40) | 35 | 41 | 70.7 | −0.05 (−0.26, 0.16) | 0.10 | 0.19 | 28 | 33 | 21.2 | 0.07 (−0.15, 0.29) | 0.18 | 0.22 |
| Ghana | iLiNS−DYADG (41) | 59 | 122 | 59.8 | −0.11 (−0.26, 0.05) | 0.18 | 0.25 | 37 | 73 | 52.1 | 0.05 (−0.15, 0.25) | 0.22 | 0.25 |
| Kenya | WASH−B (42) | | | | | | | | | | | | |
| Madagascar | MAHAY (43) | | | | | | | | | | | | |
| Malawi | iLiNS−DYADM (44) | 54 | 120 | 45.8 | 0.21 (0.05, 0.37) | 0.17 | 0.24 | 17 | 43 | 55.8 | −0.26 (−0.54, 0.01) | 0.11 | 0.16 |
| Malawi | iLiNS−DOSE (45) | | | | | | | | | | | | |
| Mali | PROMIS CS (46) | | | | | | | | | | | | |
| Zimbabwe | SHINE (HIV−) (47) | | | | | | | | | | | | |
| Zimbabwe | SHINE (HIV+) (48) | | | | | | | | | | | | |
| | | **405** | **352** | | **I² = 0.70, Tau² = 0.01** | | | **259** | **219** | | **I² = 0.27, Tau² = 0.01** | | |
| **Fixed** | | | | | **−0.02 (−0.09, 0.04)** | | | | | | **−0.02 (−0.11, 0.08)** | | |
| **Random** | | | | | **0.00 (−0.14, 0.13)** | | | | | | **−0.02 (−0.15, 0.10)** | | |

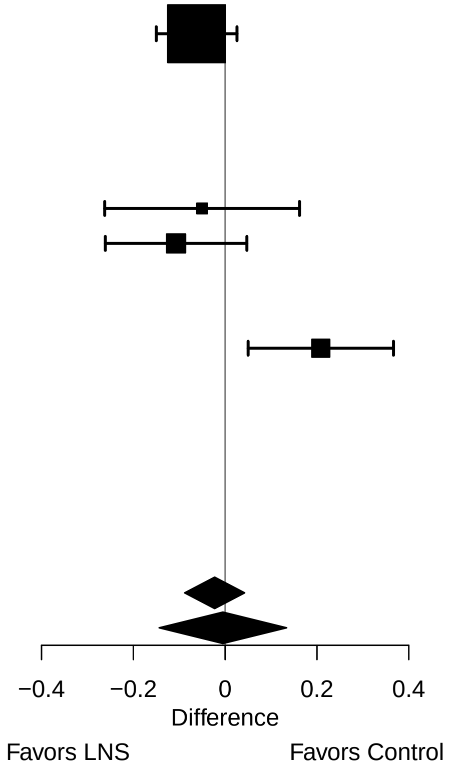

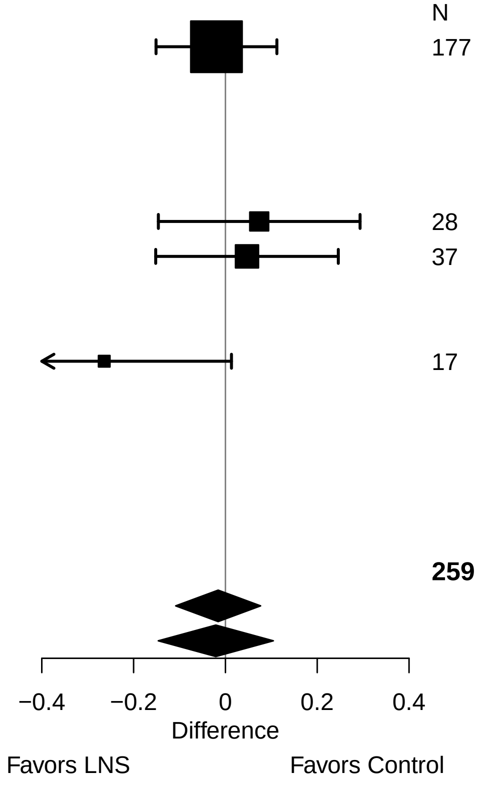

### Supplemental figure 7W: Geometric mean ratio of retinol binding protein concentration

#### 7W1: Stratified by Household socio-economic status

**P−for−interaction = 0.108**
**Ratio of GMRs = 0.96 (0.92, 1.01)**

| | | **At least median** | | | | | | | **Less than median** | | | | |
|---|---|---|---|---|---|---|---|---|---|---|---|---|---|
| | | **LNS** | **Control** | **Control** | **GMR** | **Fixed** | **Random** | **LNS** | **Control** | **Control** | **GMR** | **Fixed** | **Random** |
| Country | Trial | N | N | Median | (95% CI) | W | W | N | N | Median | (95% CI) | W | W |
| Bangladesh | JiVitA−4 (35) | | | | | | | | | | | | |
| Bangladesh | RDNS (36) | 273 | 136 | 1.2 | 1.03 (0.96, 1.11) | 0.23 | 0.23 | 277 | 136 | 1.2 | 1.01 (0.95, 1.07) | 0.28 | 0.25 |
| Bangladesh | WASH−B (37) | 105 | 91 | 1.1 | 1.08 (1.01, 1.16) | 0.24 | 0.23 | 107 | 87 | 1.1 | 1.00 (0.94, 1.07) | 0.20 | 0.21 |
| Burkina Faso | iLiNS−Zinc (38) | 180 | 39 | 1.1 | 1.08 (0.98, 1.20) | 0.11 | 0.13 | 135 | 57 | 1.0 | 1.09 (1.03, 1.17) | 0.22 | 0.22 |
| Burkina Faso | PROMIS CS (39) | | | | | | | | | | | | |
| Ghana | GHANA (40) | | | | | | | | | | | | |
| Ghana | iLiNS−DYADG (41) | | | | | | | | | | | | |
| Kenya | WASH−B (42) | 163 | 155 | 1.0 | 1.16 (1.09, 1.23) | 0.30 | 0.27 | 134 | 104 | 1.0 | 1.11 (1.04, 1.18) | 0.25 | 0.24 |
| Madagascar | MAHAY (43) | 38 | 25 | 1.1 | 1.05 (0.95, 1.15) | 0.12 | 0.14 | 43 | 25 | 1.1 | 1.04 (0.91, 1.18) | 0.06 | 0.09 |
| Malawi | iLiNS−DYADM (44) | | | | | | | | | | | | |
| Malawi | iLiNS−DOSE (45) | | | | | | | | | | | | |
| Mali | PROMIS CS (46) | | | | | | | | | | | | |
| Zimbabwe | SHINE (HIV−) (47) | | | | | | | | | | | | |
| Zimbabwe | SHINE (HIV+) (48) | | | | | | | | | | | | |
| | | **759** | **446** | | **I² = 0.41, Tau² = 0.00** | | | **696** | **409** | | **I² = 0.50, Tau² = 0.00** | | |
| **Fixed** | | | | | **1.09 (1.05, 1.13)** | | | | | | **1.05 (1.02, 1.08)** | | |
| **Random** | | | | | **1.09 (1.04, 1.13)** | | | | | | **1.05 (1.01, 1.10)** | | |

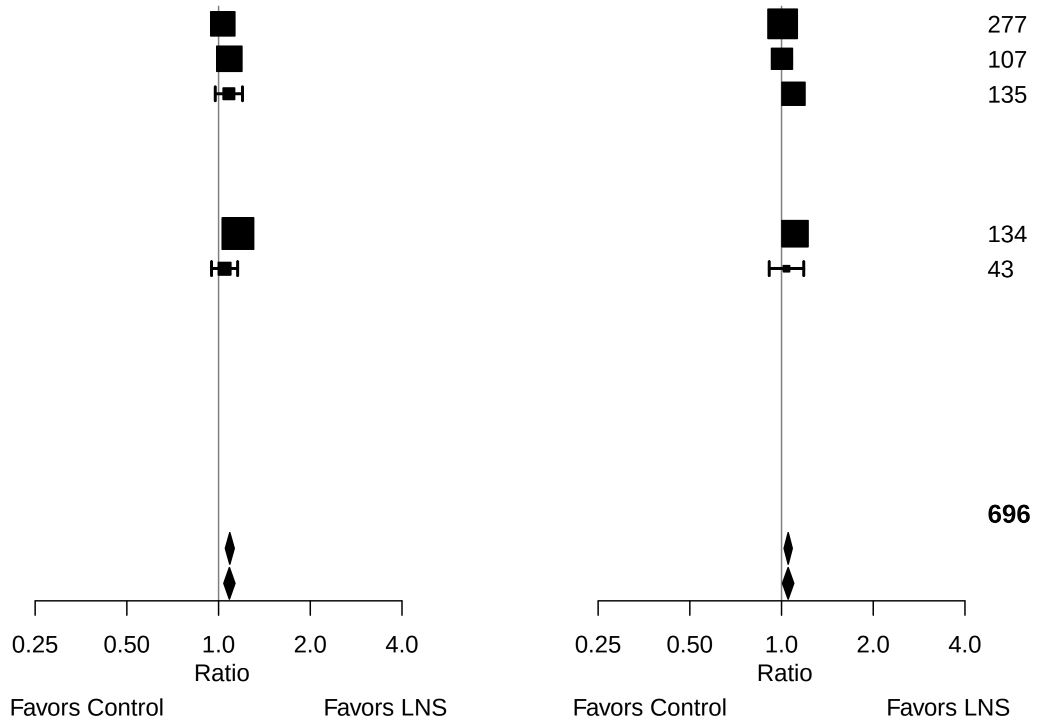

#### Supplemental figure 7W: Geometric mean ratio of retinol binding protein concentration

#### 7W2: Stratified by Household food insecurity

**P−for−interaction = 0.183**
**Ratio of GMRs = 1.04 (0.98, 1.09)**

| Country | Trial | Mild to secure: LNS N | Control N | Control Median | GMR (95% CI) | Fixed W | Random W | Moderate to severe: LNS N | Control N | Control Median | GMR (95% CI) | Fixed W | Random W |
|---|---|---|---|---|---|---|---|---|---|---|---|---|---|
| Bangladesh | JiVitA−4 (35) | | | | | | | | | | | | |
| Bangladesh | RDNS (36) | 342 | 166 | 1.2 | 1.03 (0.96, 1.10) | 0.19 | 0.21 | 208 | 106 | 1.2 | 1.01 (0.95, 1.07) | 0.29 | 0.25 |
| Bangladesh | WASH−B (37) | 171 | 139 | 1.2 | 1.02 (0.97, 1.08) | 0.29 | 0.27 | 41 | 39 | 1.0 | 1.11 (1.00, 1.23) | 0.09 | 0.18 |
| Burkina Faso | iLiNS−Zinc (38) | 160 | 45 | 1.1 | 1.10 (1.00, 1.21) | 0.10 | 0.14 | 155 | 51 | 1.1 | 1.08 (1.03, 1.13) | 0.49 | 0.26 |
| Burkina Faso | PROMIS CS (39) | | | | | | | | | | | | |
| Ghana | GHANA (40) | | | | | | | | | | | | |
| Ghana | iLiNS−DYADG (41) | | | | | | | | | | | | |
| Kenya | WASH−B (42) | 259 | 235 | 1.0 | 1.13 (1.07, 1.18) | 0.38 | 0.30 | 38 | 24 | 0.8 | 1.27 (1.13, 1.42) | 0.07 | 0.16 |
| Madagascar | MAHAY (43) | 45 | 30 | 1.1 | 1.07 (0.93, 1.23) | 0.05 | 0.08 | 19 | 7 | 1.0 | 1.09 (0.96, 1.24) | 0.06 | 0.15 |
| Malawi | iLiNS−DYADM (44) | | | | | | | | | | | | |
| Malawi | iLiNS−DOSE (45) | | | | | | | | | | | | |
| Mali | PROMIS CS (46) | | | | | | | | | | | | |
| Zimbabwe | SHINE (HIV−) (47) | | | | | | | | | | | | |
| Zimbabwe | SHINE (HIV+) (48) | | | | | | | | | | | | |
| | | **977** | **615** | | **I² = 0.53, Tau² = 0.00** | | | **461** | **227** | | **I² = 0.68, Tau² = 0.00** | | |
| **Fixed** | | | | | **1.07 (1.04, 1.10)** | | | | | | **1.07 (1.04, 1.11)** | | |
| **Random** | | | | | **1.07 (1.02, 1.11)** | | | | | | **1.10 (1.02, 1.18)** | | |

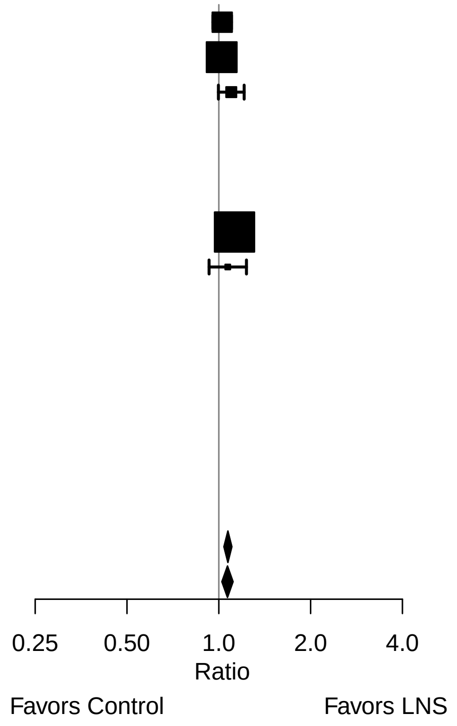

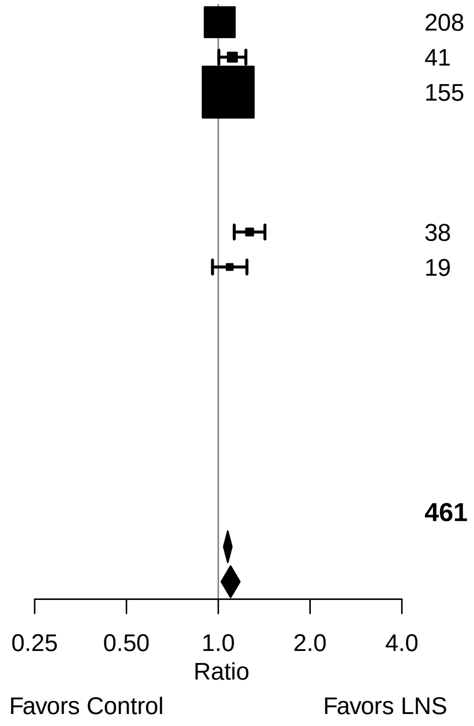

#### Supplemental figure 7W: Geometric mean ratio of retinol binding protein concentration

#### 7W3: Stratified by Household source water quality

**P−for−interaction = 0.959**
**Ratio of GMRs = 1.00 (0.92, 1.09)**

| Country | Trial | Improved LNS N | Improved Control N | Improved Control Median | Improved GMR (95% CI) | Improved Fixed W | Improved Random W | Unimproved LNS N | Unimproved Control N | Unimproved Control Median | Unimproved GMR (95% CI) | Unimproved Fixed W | Unimproved Random W |
|---|---|---|---|---|---|---|---|---|---|---|---|---|---|
| Bangladesh | JiVitA−4 (35) | | | | | | | | | | | | |
| Bangladesh | RDNS (36) | | | | | | | | | | | | |
| Bangladesh | WASH−B (37) | 75 | 44 | 1.1 | 1.11 (1.00, 1.22) | 0.29 | 0.29 | 19 | 14 | 1.0 | 1.05 (0.84, 1.30) | 0.04 | 0.04 |
| Burkina Faso | iLiNS−Zinc (38) | 97 | 22 | 1.0 | 1.12 (0.97, 1.30) | 0.13 | 0.18 | 218 | 74 | 1.1 | 1.09 (1.03, 1.15) | 0.64 | 0.64 |
| Burkina Faso | PROMIS CS (39) | | | | | | | | | | | | |
| Ghana | GHANA (40) | | | | | | | | | | | | |
| Ghana | iLiNS−DYADG (41) | | | | | | | | | | | | |
| Kenya | WASH−B (42) | 111 | 107 | 1.0 | 1.17 (1.08, 1.27) | 0.45 | 0.36 | 55 | 37 | 0.9 | 1.17 (1.05, 1.30) | 0.16 | 0.16 |
| Madagascar | MAHAY (43) | 23 | 14 | 1.2 | 0.97 (0.83, 1.13) | 0.12 | 0.17 | 55 | 36 | 1.1 | 1.07 (0.96, 1.19) | 0.16 | 0.16 |
| Malawi | iLiNS−DYADM (44) | | | | | | | | | | | | |
| Malawi | iLiNS−DOSE (45) | | | | | | | | | | | | |
| Mali | PROMIS CS (46) | | | | | | | | | | | | |
| Zimbabwe | SHINE (HIV−) (47) | | | | | | | | | | | | |
| Zimbabwe | SHINE (HIV+) (48) | | | | | | | | | | | | |
| | | **306** | **187** | | **I² = 0.37, Tau² = 0.00** | | | **347** | **161** | | **I² = 0.00, Tau² = 0.00** | | |
| **Fixed** | | | | | **1.12 (1.06, 1.18)** | | | | | | **1.09 (1.05, 1.14)** | | |
| **Random** | | | | | **1.11 (1.03, 1.19)** | | | | | | **1.09 (1.05, 1.14)** | | |

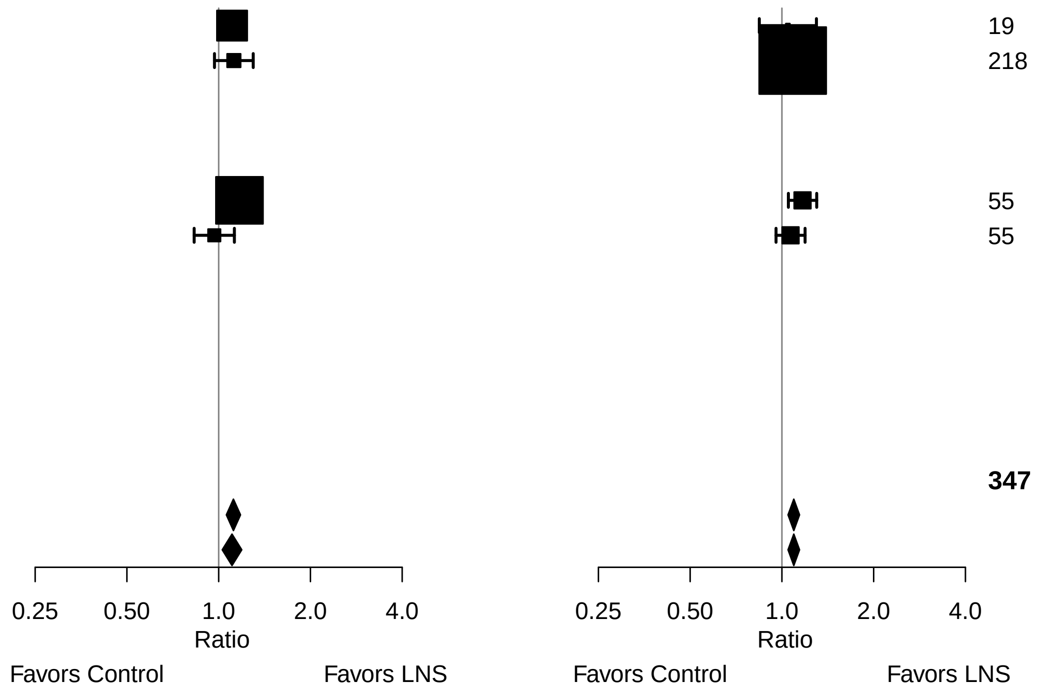

**Supplemental figure 7W: Geometric mean ratio of retinol binding protein concentration**

#### 7W4: Stratified by Household sanitation (insufficient comparisons)

#### Supplemental figure 7W: Geometric mean ratio of retinol binding protein concentration

#### 7W5: Stratified by Season at the time of assessment

**P−for−interaction = 0.101**
**Ratio of GMRs = 0.96 (0.92, 1.01)**

| Country | Trial | Dry LNS N | Dry Control N | Dry Control Median | Dry GMR (95% CI) | Dry Fixed W | Dry Random W | Rainy LNS N | Rainy Control N | Rainy Control Median | Rainy GMR (95% CI) | Rainy Fixed W | Rainy Random W |
|---|---|---|---|---|---|---|---|---|---|---|---|---|---|
| Bangladesh | JiVitA−4 (35) | | | | | | | | | | | | |
| Bangladesh | RDNS (36) | 341 | 166 | 1.2 | 1.03 (0.98, 1.08) | 0.30 | 0.28 | 208 | 106 | 1.3 | 1.01 (0.94, 1.08) | 0.28 | 0.26 |
| Bangladesh | WASH−B (37) | 96 | 93 | 1.1 | 1.10 (1.05, 1.15) | 0.35 | 0.29 | 116 | 85 | 1.2 | 0.98 (0.90, 1.06) | 0.20 | 0.24 |
| Burkina Faso | iLiNS−Zinc (38) | 174 | 61 | 1.0 | 1.09 (1.02, 1.15) | 0.23 | 0.25 | 126 | 30 | 1.1 | 1.09 (1.00, 1.19) | 0.17 | 0.22 |
| Burkina Faso | PROMIS CS (39) | | | | | | | | | | | | |
| Ghana | GHANA (40) | | | | | | | | | | | | |
| Ghana | iLiNS−DYADG (41) | | | | | | | | | | | | |
| Kenya | WASH−B (42) | 88 | 75 | 1.0 | 1.16 (1.06, 1.26) | 0.12 | 0.18 | 210 | 184 | 1.0 | 1.13 (1.06, 1.20) | 0.35 | 0.28 |
| Madagascar | MAHAY (43) | | | | | | | | | | | | |
| Malawi | iLiNS−DYADM (44) | | | | | | | | | | | | |
| Malawi | iLiNS−DOSE (45) | | | | | | | | | | | | |
| Mali | PROMIS CS (46) | | | | | | | | | | | | |
| Zimbabwe | SHINE (HIV−) (47) | | | | | | | | | | | | |
| Zimbabwe | SHINE (HIV+) (48) | | | | | | | | | | | | |
| | | **699** | **395** | | **$I^2$ = 0.53, $Tau^2$ = 0.00** | | | **660** | **405** | | **$I^2$ = 0.71, $Tau^2$ = 0.00** | | |
| **Fixed** | | | | | **1.08 (1.05, 1.11)** | | | | | | **1.06 (1.02, 1.09)** | | |
| **Random** | | | | | **1.09 (1.04, 1.14)** | | | | | | **1.05 (0.98, 1.12)** | | |

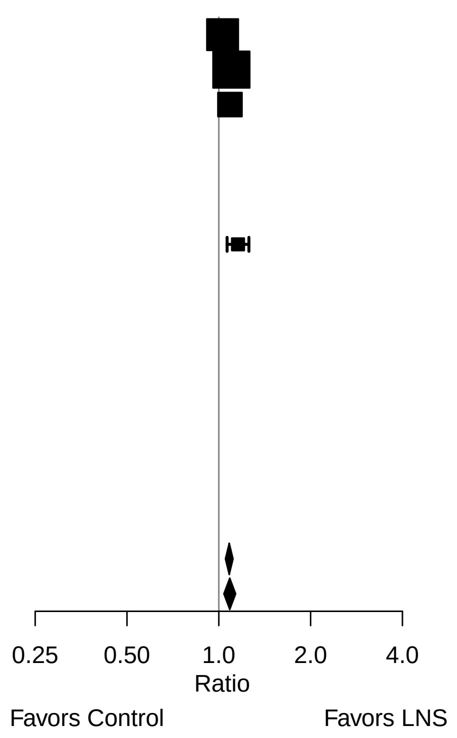

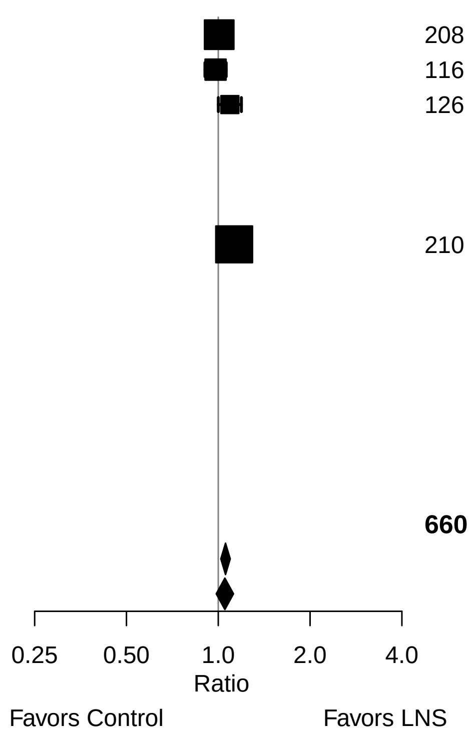

#### Supplemental figure 7X: Low vitamin A status (RBP < 0.70 µmol/L) prevalence ratio

##### 7X1: Stratified by Household socio-economic status (insufficient comparisons)

**Supplemental figure 7X: Low vitamin A status (RBP < 0.70 µmol/L) prevalence ratio**

#### 7X2: Stratified by Household food insecurity (insufficient comparisons)

**Supplemental figure 7X: Low vitamin A status (RBP < 0.70 µmol/L) prevalence ratio**

#### 7X3: Stratified by Household source water quality (insufficient comparisons)

**Supplemental figure 7X: Low vitamin A status (RBP < 0.70 µmol/L) prevalence ratio**

#### 7X4: Stratified by Household sanitation (insufficient comparisons)

**Supplemental figure 7X: Low vitamin A status (RBP < 0.70 μmol/L) prevalence ratio**

#### 7X5: Stratified by Season at the time of assessment (insufficient comparisons)

#### Supplemental figure 7Y: Low vitamin A status (RBP < 0.70 µmol/L) prevalence difference

##### 7Y1: Stratified by Household socio-economic status (insufficient comparisons)

**Supplemental figure 7Y: Low vitamin A status (RBP < 0.70 µmol/L) prevalence difference**

#### 7Y2: Stratified by Household food insecurity (insufficient comparisons)

**Supplemental figure 7Y: Low vitamin A status (RBP < 0.70 µmol/L) prevalence difference**

#### 7Y3: Stratified by Household source water quality (insufficient comparisons)

**Supplemental figure 7Y: Low vitamin A status (RBP < 0.70 µmol/L) prevalence difference**

#### 7Y4: Stratified by Household sanitation (insufficient comparisons)

**Supplemental figure 7Y: Low vitamin A status (RBP < 0.70 µmol/L) prevalence difference**

#### 7Y5: Stratified by Season at the time of assessment (insufficient comparisons)

### Supplemental figure 7Z: Marginal vitamin A status (RBP < 1.05 µmol/L) prevalence ratio

#### 7Z1: Stratified by Household socio-economic status

**P−for−interaction = 0.026**
**Ratio of PRs = 1.26 (1.03, 1.56)**

| | | At least median | | | | | | Less than median | | | | | |
|---|---|---|---|---|---|---|---|---|---|---|---|---|---|
| Country | Trial | LNS N | Control N | Control Prevalence | PR (95% CI) | Fixed W | Random W | LNS N | Control N | Control Prevalence | PR (95% CI) | Fixed W | Random W |
| Bangladesh | JiVitA−4 (35) | | | | | | | | | | | | |
| Bangladesh | RDNS (36) | 273 | 136 | 30.9 | 0.75 (0.53, 1.05) | 0.21 | 0.21 | 277 | 136 | 27.2 | 1.01 (0.71, 1.43) | 0.17 | 0.17 |
| Bangladesh | WASH−B (37) | 105 | 91 | 34.1 | 0.73 (0.47, 1.12) | 0.13 | 0.13 | 107 | 87 | 36.8 | 0.97 (0.72, 1.30) | 0.24 | 0.24 |
| Burkina Faso | iLiNS−Zinc (38) | 180 | 39 | 44.2 | 0.86 (0.55, 1.35) | 0.12 | 0.12 | 135 | 57 | 52.4 | 0.69 (0.49, 0.96) | 0.19 | 0.19 |
| Burkina Faso | PROMIS CS (39) | | | | | | | | | | | | |
| Ghana | GHANA (40) | | | | | | | | | | | | |
| Ghana | iLiNS−DYADG (41) | | | | | | | | | | | | |
| Kenya | WASH−B (42) | 163 | 155 | 63.2 | 0.63 (0.50, 0.79) | 0.48 | 0.48 | 134 | 104 | 53.8 | 0.86 (0.67, 1.10) | 0.35 | 0.35 |
| Madagascar | MAHAY (43) | 38 | 25 | 36.0 | 0.80 (0.41, 1.58) | 0.05 | 0.05 | 43 | 25 | 40.0 | 0.87 (0.43, 1.78) | 0.04 | 0.04 |
| Malawi | iLiNS−DYADM (44) | | | | | | | | | | | | |
| Malawi | iLiNS−DOSE (45) | | | | | | | | | | | | |
| Mali | PROMIS CS (46) | | | | | | | | | | | | |
| Zimbabwe | SHINE (HIV−) (47) | | | | | | | | | | | | |
| Zimbabwe | SHINE (HIV+) (48) | | | | | | | | | | | | |
| | | **759** | **446** | | **I² = 0.00, Tau² = 0.00** | | | **696** | **409** | | **I² = 0.00, Tau² = 0.00** | | |
| **Fixed** | | | | | **0.70 (0.60, 0.82)** | | | | | | **0.87 (0.75, 1.01)** | | |
| **Random** | | | | | **0.70 (0.60, 0.82)** | | | | | | **0.87 (0.75, 1.01)** | | |

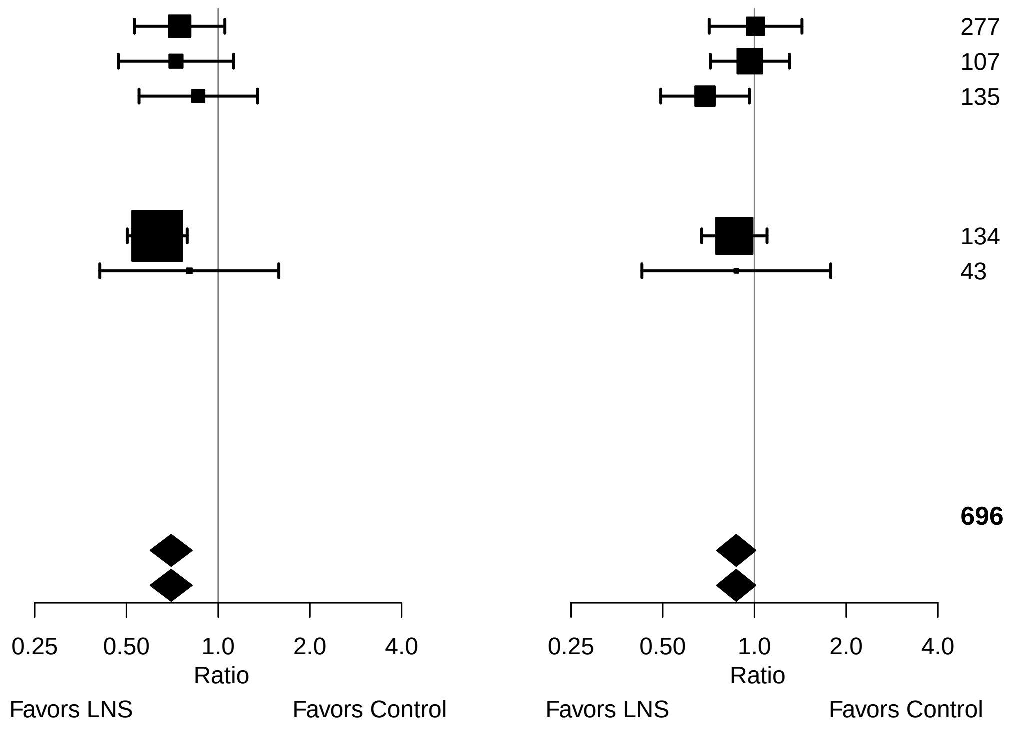

**Supplemental figure 7Z: Marginal vitamin A status (RBP < 1.05 µmol/L) prevalence ratio**

#### 7Z2: Stratified by Household food insecurity

**P−for−interaction = 0.208**
**Ratio of PRs = 0.85 (0.67, 1.09)**

| | | Mild to secure | | | | | | Moderate to severe | | | | | |
|---|---|---|---|---|---|---|---|---|---|---|---|---|---|
| Country | Trial | LNS N | Control N | Control Prevalence | PR (95% CI) | Fixed W | Random W | LNS N | Control N | Control Prevalence | PR (95% CI) | Fixed W | Random W |
| Bangladesh | JiVitA−4 (35) | | | | | | | | | | | | |
| Bangladesh | RDNS (36) | 342 | 166 | 28.9 | 0.89 (0.63, 1.26) | 0.16 | 0.16 | 208 | 106 | 29.2 | 0.84 (0.57, 1.22) | 0.21 | 0.21 |
| Bangladesh | WASH−B (37) | 171 | 139 | 29.5 | 0.95 (0.67, 1.35) | 0.16 | 0.16 | 41 | 39 | 56.4 | 0.69 (0.46, 1.03) | 0.19 | 0.19 |
| Burkina Faso | iLiNS−Zinc (38) | 160 | 45 | 49.0 | 0.73 (0.49, 1.08) | 0.13 | 0.13 | 155 | 51 | 49.1 | 0.80 (0.61, 1.04) | 0.42 | 0.42 |
| Burkina Faso | PROMIS CS (39) | | | | | | | | | | | | |
| Ghana | GHANA (40) | | | | | | | | | | | | |
| Ghana | iLiNS−DYADG (41) | | | | | | | | | | | | |
| Kenya | WASH−B (42) | 259 | 235 | 57.4 | 0.74 (0.61, 0.89) | 0.56 | 0.56 | 38 | 24 | 79.2 | 0.57 (0.37, 0.85) | 0.18 | 0.18 |
| Madagascar | MAHAY (43) | | | | | | | | | | | | |
| Malawi | iLiNS−DYADM (44) | | | | | | | | | | | | |
| Malawi | iLiNS−DOSE (45) | | | | | | | | | | | | |
| Mali | PROMIS CS (46) | | | | | | | | | | | | |
| Zimbabwe | SHINE (HIV−) (47) | | | | | | | | | | | | |
| Zimbabwe | SHINE (HIV+) (48) | | | | | | | | | | | | |
| | | **932** | **585** | | **I² = 0.00, Tau² = 0.00** | | | **442** | **220** | | **I² = 0.00, Tau² = 0.00** | | |
| **Fixed** | | | | | **0.79 (0.69, 0.91)** | | | | | | **0.74 (0.62, 0.88)** | | |
| **Random** | | | | | **0.79 (0.69, 0.91)** | | | | | | **0.74 (0.62, 0.88)** | | |

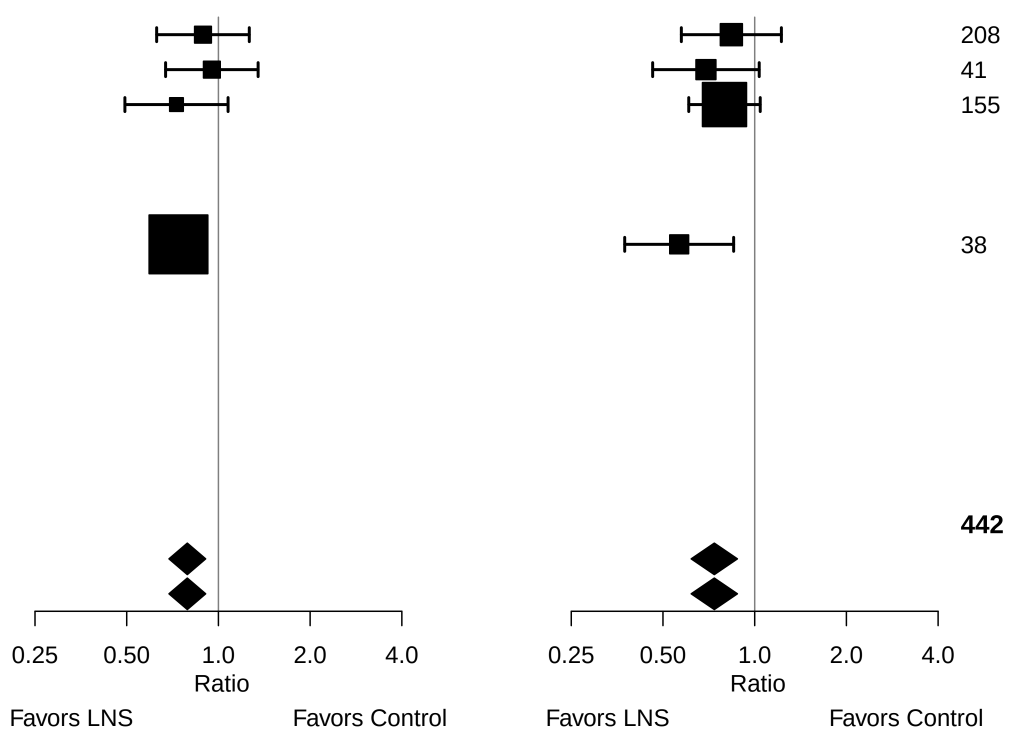

#### Supplemental figure 7Z: Marginal vitamin A status (RBP < 1.05 μmol/L) prevalence ratio

##### 7Z3: Stratified by Household source water quality

**P−for−interaction = 0.520**
**Ratio of PRs = 1.10 (0.82, 1.48)**

| Country | Trial | Improved LNS N | Improved Control N | Improved Control Prevalence | Improved PR (95% CI) | Improved Fixed W | Improved Random W | Unimproved LNS N | Unimproved Control N | Unimproved Control Prevalence | Unimproved PR (95% CI) | Unimproved Fixed W | Unimproved Random W |
|---|---|---|---|---|---|---|---|---|---|---|---|---|---|
| Bangladesh | JiVitA−4 (35) | | | | | | | | | | | | |
| Bangladesh | RDNS (36) | | | | | | | | | | | | |
| Bangladesh | WASH−B (37) | 75 | 44 | 45.5 | 0.67 (0.40, 1.13) | 0.16 | 0.16 | 19 | 14 | 50.0 | 0.84 (0.32, 2.22) | 0.04 | 0.04 |
| Burkina Faso | iLiNS−Zinc (38) | 97 | 22 | 56.5 | 0.74 (0.51, 1.08) | 0.29 | 0.29 | 218 | 74 | 47.0 | 0.76 (0.60, 0.96) | 0.69 | 0.69 |
| Burkina Faso | PROMIS CS (39) | | | | | | | | | | | | |
| Ghana | GHANA (40) | | | | | | | | | | | | |
| Ghana | iLiNS−DYADG (41) | | | | | | | | | | | | |
| Kenya | WASH−B (42) | 111 | 107 | 61.7 | 0.67 (0.51, 0.88) | 0.55 | 0.55 | 55 | 37 | 67.6 | 0.78 (0.53, 1.14) | 0.26 | 0.26 |
| Madagascar | MAHAY (43) | | | | | | | | | | | | |
| Malawi | iLiNS−DYADM (44) | | | | | | | | | | | | |
| Malawi | iLiNS−DOSE (45) | | | | | | | | | | | | |
| Mali | PROMIS CS (46) | | | | | | | | | | | | |
| Zimbabwe | SHINE (HIV−) (47) | | | | | | | | | | | | |
| Zimbabwe | SHINE (HIV+) (48) | | | | | | | | | | | | |
| | | **283** | **173** | | **I² = 0.00, Tau² = 0.00** | | | **292** | **125** | | **I² = 0.00, Tau² = 0.00** | | |
| **Fixed** | | | | | **0.69 (0.56, 0.85)** | | | | | | **0.77 (0.63, 0.93)** | | |
| **Random** | | | | | **0.69 (0.56, 0.85)** | | | | | | **0.77 (0.63, 0.93)** | | |

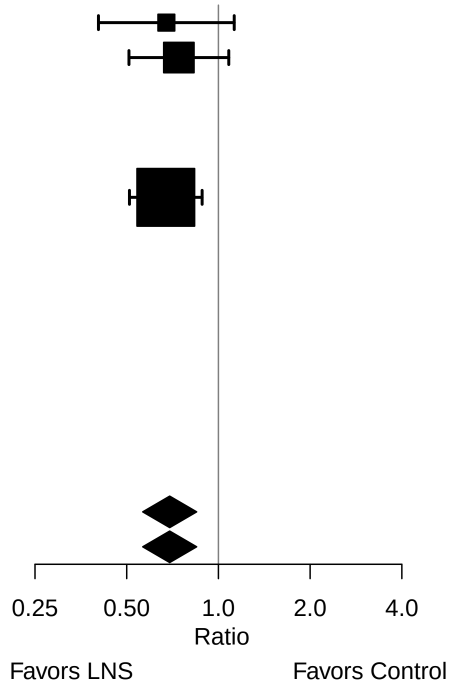

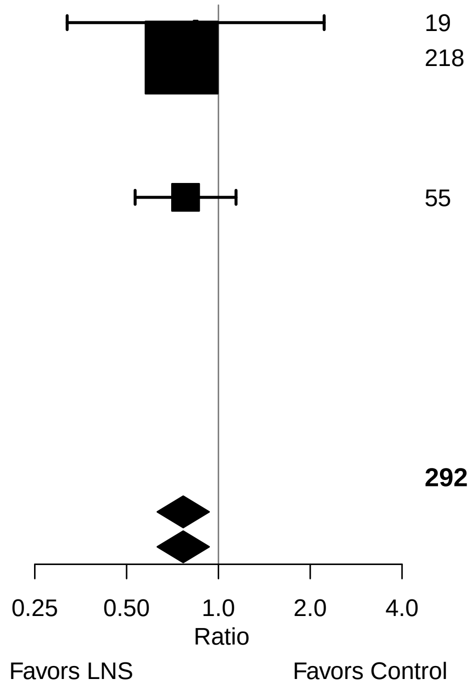

**Supplemental figure 7Z: Marginal vitamin A status (RBP < 1.05 µmol/L) prevalence ratio**

#### 7Z4: Stratified by Household sanitation (insufficient comparisons)

**Supplemental figure 7Z: Marginal vitamin A status (RBP < 1.05 µmol/L) prevalence ratio**

#### 7Z5: Stratified by Season at the time of assessment

**P−for−interaction = 0.552**
**Ratio of PRs = 1.08 (0.84, 1.37)**

| Country | Trial | Dry LNS N | Dry Control N | Dry Control Prevalence | Dry PR (95% CI) | Dry Fixed W | Dry Random W | Rainy LNS N | Rainy Control N | Rainy Control Prevalence | Rainy PR (95% CI) | Rainy Fixed W | Rainy Random W |
|---|---|---|---|---|---|---|---|---|---|---|---|---|---|
| Bangladesh | JiVitA−4 (35) | | | | | | | | | | | | |
| Bangladesh | RDNS (36) | 341 | 166 | 28.9 | 0.85 (0.63, 1.15) | 0.26 | 0.26 | 208 | 106 | 29.2 | 0.90 (0.59, 1.38) | 0.15 | 0.17 |
| Bangladesh | WASH−B (37) | 96 | 93 | 44.1 | 0.71 (0.49, 1.02) | 0.17 | 0.17 | 116 | 85 | 25.9 | 1.13 (0.68, 1.89) | 0.11 | 0.12 |
| Burkina Faso | iLiNS−Zinc (38) | 174 | 61 | 50.7 | 0.75 (0.58, 0.97) | 0.35 | 0.35 | 126 | 30 | 46.7 | 0.82 (0.52, 1.28) | 0.14 | 0.15 |
| Burkina Faso | PROMIS CS (39) | | | | | | | | | | | | |
| Ghana | GHANA (40) | | | | | | | | | | | | |
| Ghana | iLiNS−DYADG (41) | | | | | | | | | | | | |
| Kenya | WASH−B (42) | 88 | 75 | 56.0 | 0.75 (0.55, 1.03) | 0.23 | 0.23 | 210 | 184 | 60.9 | 0.71 (0.58, 0.88) | 0.60 | 0.57 |
| Madagascar | MAHAY (43) | | | | | | | | | | | | |
| Malawi | iLiNS−DYADM (44) | | | | | | | | | | | | |
| Malawi | iLiNS−DOSE (45) | | | | | | | | | | | | |
| Mali | PROMIS CS (46) | | | | | | | | | | | | |
| Zimbabwe | SHINE (HIV−) (47) | | | | | | | | | | | | |
| Zimbabwe | SHINE (HIV+) (48) | | | | | | | | | | | | |
| | | **699** | **395** | | **I² = 0.00, Tau² = 0.00** | | | **660** | **405** | | **I² = 0.08, Tau² = 0.00** | | |
| **Fixed** | | | | | **0.77 (0.66, 0.89)** | | | | | | **0.79 (0.67, 0.93)** | | |
| **Random** | | | | | **0.77 (0.66, 0.89)** | | | | | | **0.80 (0.67, 0.95)** | | |

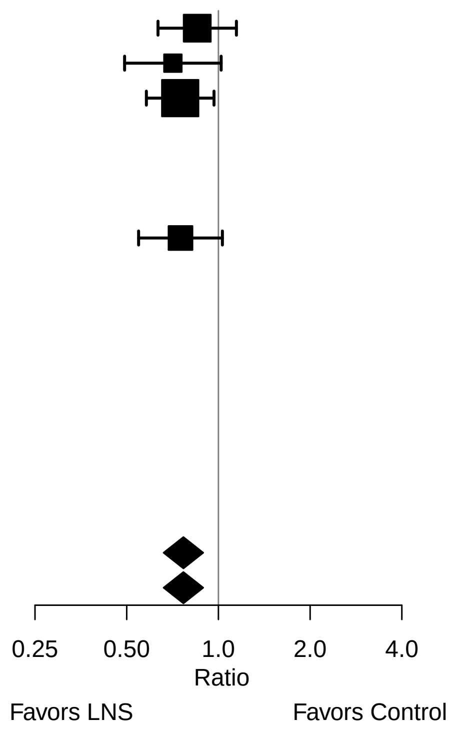

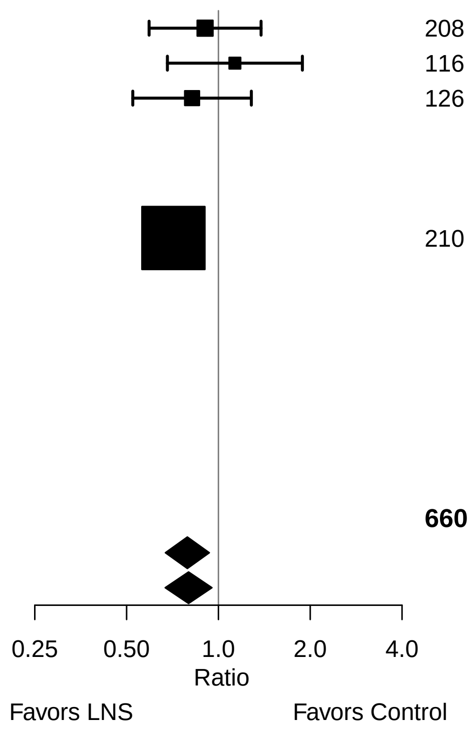

### Supplemental figure 7AA: Marginal vitamin A status (RBP < 1.05 µmol/L) prevalence difference

#### 7AA1: Stratified by Household socio-economic status

**P−for−interaction = 0.076**
**Difference in PDs = 0.07 (−0.01, 0.15)**

| Country | Trial | LNS N | Control N | Control Prevalence | At least median PD (95% CI) | Fixed W | Random W | LNS N | Control N | Control Prevalence | Less than median PD (95% CI) | Fixed W | Random W |
|---|---|---|---|---|---|---|---|---|---|---|---|---|---|
| Bangladesh | JiVitA−4 (35) | | | | | | | | | | | | |
| Bangladesh | RDNS (36) | 273 | 136 | 30.9 | −0.08 (−0.17, 0.02) | 0.36 | 0.33 | 277 | 136 | 27.2 | 0.00 (−0.09, 0.10) | 0.34 | 0.34 |
| Bangladesh | WASH−B (37) | 105 | 91 | 34.1 | −0.09 (−0.22, 0.03) | 0.21 | 0.22 | 107 | 87 | 36.8 | −0.01 (−0.12, 0.10) | 0.26 | 0.26 |
| Burkina Faso | iLiNS−Zinc (38) | 180 | 39 | 44.2 | −0.07 (−0.25, 0.11) | 0.10 | 0.12 | 135 | 57 | 52.4 | −0.15 (−0.29, −0.01) | 0.16 | 0.16 |
| Burkina Faso | PROMIS CS (39) | | | | | | | | | | | | |
| Ghana | GHANA (40) | | | | | | | | | | | | |
| Ghana | iLiNS−DYADG (41) | | | | | | | | | | | | |
| Kenya | WASH−B (42) | 163 | 155 | 63.2 | −0.23 (−0.34, −0.12) | 0.27 | 0.26 | 134 | 104 | 53.8 | −0.08 (−0.20, 0.05) | 0.20 | 0.20 |
| Madagascar | MAHAY (43) | 38 | 25 | 36.0 | −0.07 (−0.30, 0.16) | 0.06 | 0.08 | 43 | 25 | 40.0 | −0.05 (−0.32, 0.22) | 0.04 | 0.04 |
| Malawi | iLiNS−DYADM (44) | | | | | | | | | | | | |
| Malawi | iLiNS−DOSE (45) | | | | | | | | | | | | |
| Mali | PROMIS CS (46) | | | | | | | | | | | | |
| Zimbabwe | SHINE (HIV−) (47) | | | | | | | | | | | | |
| Zimbabwe | SHINE (HIV+) (48) | | | | | | | | | | | | |
| | | **759** | **446** | | **I² = 0.26, Tau² = 0.00** | | | **696** | **409** | | **I² = 0.00, Tau² = 0.00** | | |
| **Fixed** | | | | | **−0.12 (−0.18, −0.06)** | | | | | | **−0.04 (−0.10, 0.01)** | | |
| **Random** | | | | | **−0.12 (−0.19, −0.05)** | | | | | | **−0.04 (−0.10, 0.01)** | | |

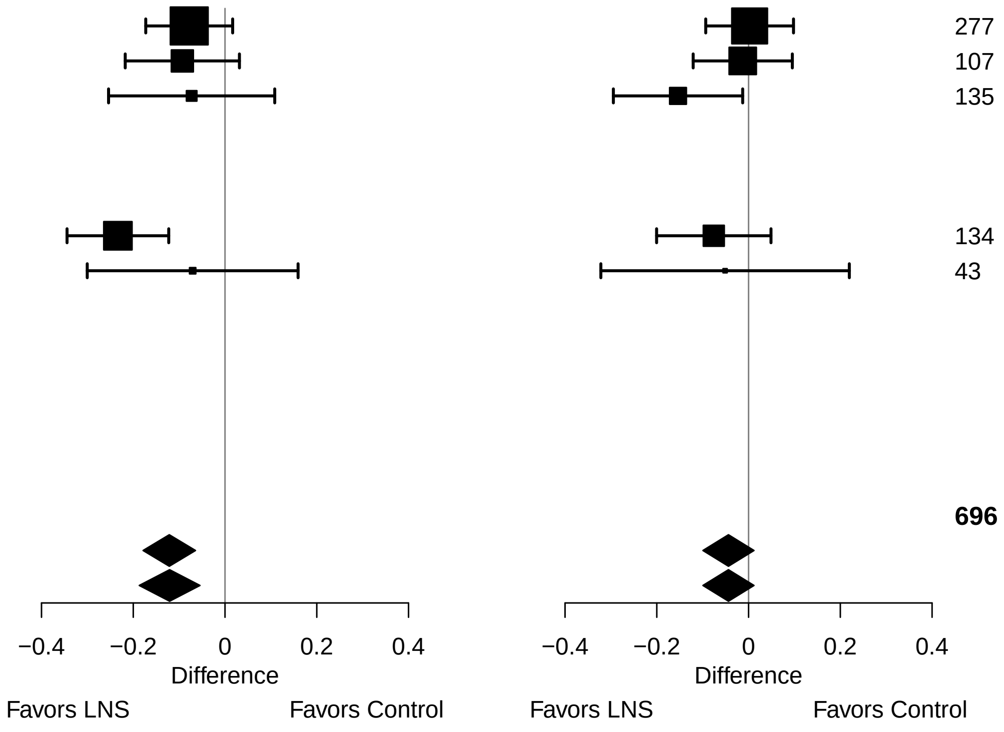

#### Supplemental figure 7AA: Marginal vitamin A status (RBP < 1.05 µmol/L) prevalence difference

#### 7AA2: Stratified by Household food insecurity

**P−for−interaction = 0.231**
**Difference in PDs = −0.06 (−0.15, 0.04)**

| | | Mild to secure | | | | | | Moderate to severe | | | | | |
|---|---|---|---|---|---|---|---|---|---|---|---|---|---|
| Country | Trial | LNS N | Control N | Control Prevalence | PD (95% CI) | Fixed W | Random W | LNS N | Control N | Control Prevalence | PD (95% CI) | Fixed W | Random W |
| Bangladesh | JiVitA−4 (35) | | | | | | | | | | | | |
| Bangladesh | RDNS (36) | 342 | 166 | 28.9 | −0.03 (−0.13, 0.07) | 0.29 | 0.28 | 208 | 106 | 29.2 | −0.05 (−0.15, 0.06) | 0.44 | 0.34 |
| Bangladesh | WASH−B (37) | 171 | 139 | 29.5 | −0.01 (−0.11, 0.09) | 0.28 | 0.27 | 41 | 39 | 56.4 | −0.17 (−0.36, 0.02) | 0.13 | 0.20 |
| Burkina Faso | iLiNS−Zinc (38) | 160 | 45 | 49.0 | −0.12 (−0.29, 0.04) | 0.10 | 0.14 | 155 | 51 | 49.1 | −0.10 (−0.22, 0.02) | 0.34 | 0.31 |
| Burkina Faso | PROMIS CS (39) | | | | | | | | | | | | |
| Ghana | GHANA (40) | | | | | | | | | | | | |
| Ghana | iLiNS−DYADG (41) | | | | | | | | | | | | |
| Kenya | WASH−B (42) | 259 | 235 | 57.4 | −0.15 (−0.24, −0.06) | 0.33 | 0.30 | 38 | 24 | 79.2 | −0.34 (−0.59, −0.10) | 0.08 | 0.15 |
| Madagascar | MAHAY (43) | | | | | | | | | | | | |
| Malawi | iLiNS−DYADM (44) | | | | | | | | | | | | |
| Malawi | iLiNS−DOSE (45) | | | | | | | | | | | | |
| Mali | PROMIS CS (46) | | | | | | | | | | | | |
| Zimbabwe | SHINE (HIV−) (47) | | | | | | | | | | | | |
| Zimbabwe | SHINE (HIV+) (48) | | | | | | | | | | | | |
| | | **932** | **585** | | **I² = 0.40, Tau² = 0.00** | | | **442** | **220** | | **I² = 0.45, Tau² = 0.01** | | |
| **Fixed** | | | | | **−0.07 (−0.13, −0.02)** | | | | | | **−0.11 (−0.18, −0.04)** | | |
| **Random** | | | | | **−0.08 (−0.14, −0.01)** | | | | | | **−0.13 (−0.25, −0.02)** | | |

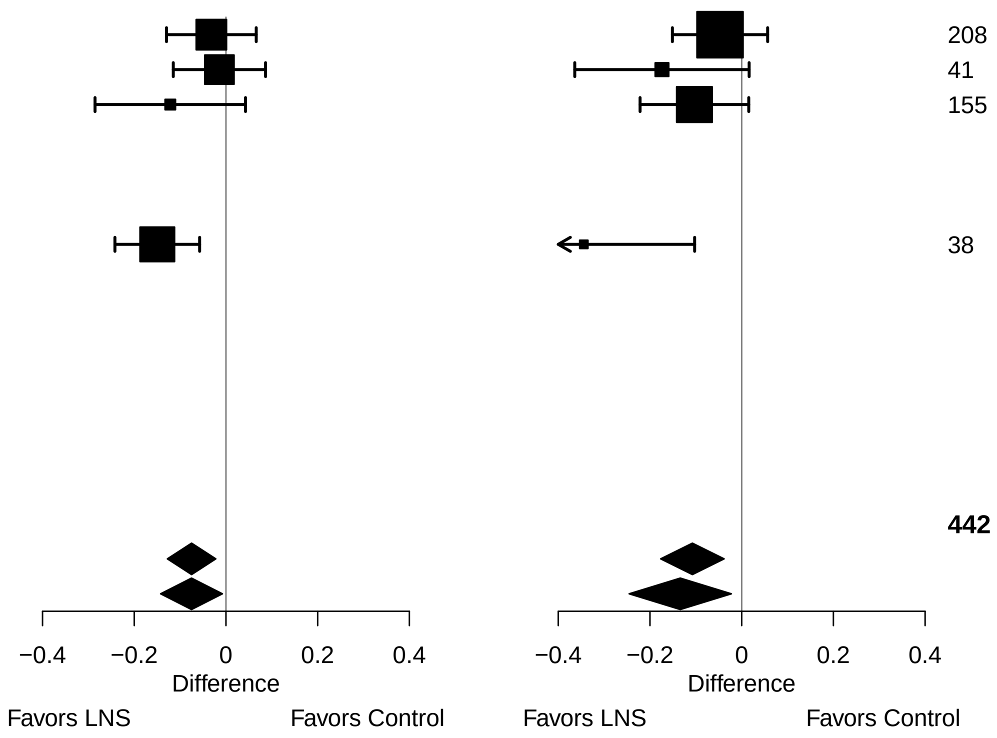

**Supplemental figure 7AA: Marginal vitamin A status (RBP < 1.05 µmol/L) prevalence difference**

#### 7AA3: Stratified by Household source water quality

**P−for−interaction = 0.499**
**Difference in PDs = 0.05 (−0.10, 0.21)**

| Country | Trial | Improved LNS N | Improved Control N | Improved Control Prevalence | Improved PD (95% CI) | Improved Fixed W | Improved Random W | Unimproved LNS N | Unimproved Control N | Unimproved Control Prevalence | Unimproved PD (95% CI) | Unimproved Fixed W | Unimproved Random W |
|---|---|---|---|---|---|---|---|---|---|---|---|---|---|
| Bangladesh | JiVitA−4 (35) | | | | | | | | | | | | |
| Bangladesh | RDNS (36) | | | | | | | | | | | | |
| Bangladesh | WASH−B (37) | 75 | 44 | 45.5 | −0.15 (−0.35, 0.05) | 0.22 | 0.22 | 19 | 14 | 50.0 | −0.08 (−0.52, 0.36) | 0.03 | 0.03 |
| Burkina Faso | iLiNS−Zinc (38) | 97 | 22 | 56.5 | −0.15 (−0.34, 0.03) | 0.27 | 0.27 | 218 | 74 | 47.0 | −0.11 (−0.20, −0.02) | 0.84 | 0.84 |
| Burkina Faso | PROMIS CS (39) | | | | | | | | | | | | |
| Ghana | GHANA (40) | | | | | | | | | | | | |
| Ghana | iLiNS−DYADG (41) | | | | | | | | | | | | |
| Kenya | WASH−B (42) | 111 | 107 | 61.7 | −0.20 (−0.33, −0.07) | 0.51 | 0.51 | 55 | 37 | 67.6 | −0.15 (−0.38, 0.08) | 0.13 | 0.13 |
| Madagascar | MAHAY (43) | | | | | | | | | | | | |
| Malawi | iLiNS−DYADM (44) | | | | | | | | | | | | |
| Malawi | iLiNS−DOSE (45) | | | | | | | | | | | | |
| Mali | PROMIS CS (46) | | | | | | | | | | | | |
| Zimbabwe | SHINE (HIV−) (47) | | | | | | | | | | | | |
| Zimbabwe | SHINE (HIV+) (48) | | | | | | | | | | | | |
| | | **283** | **173** | | **I² = 0.00, Tau² = 0.00** | | | **292** | **125** | | **I² = 0.00, Tau² = 0.00** | | |
| **Fixed** | | | | | **−0.18 (−0.27, −0.08)** | | | | | | **−0.11 (−0.20, −0.03)** | | |
| **Random** | | | | | **−0.18 (−0.27, −0.08)** | | | | | | **−0.11 (−0.20, −0.03)** | | |

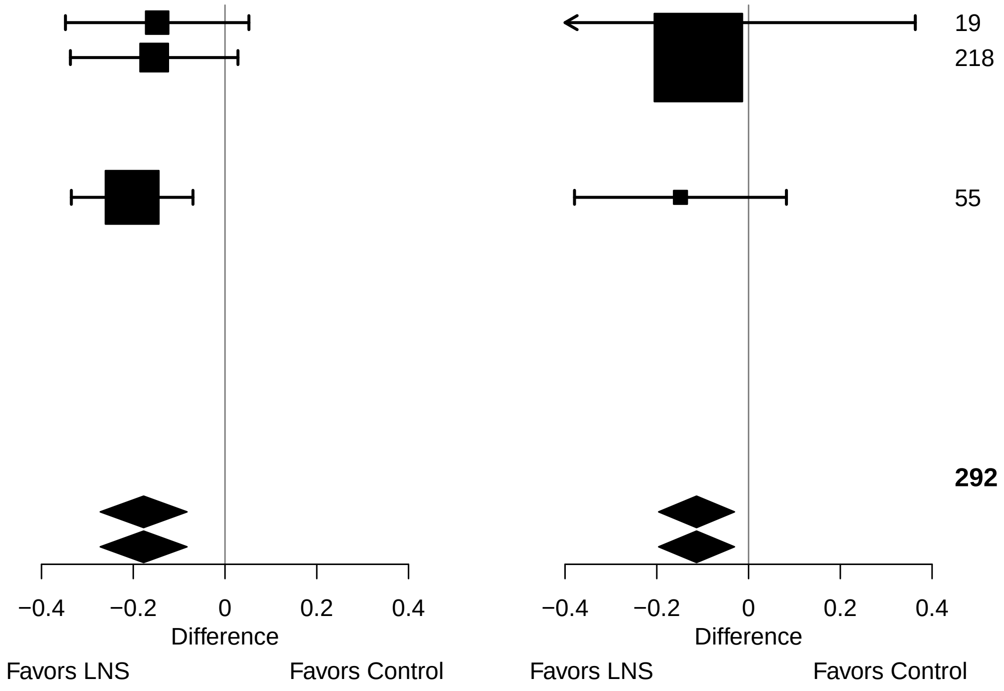

**Supplemental figure 7AA: Marginal vitamin A status (RBP < 1.05 μmol/L) prevalence difference**

#### 7AA4: Stratified by Household sanitation (insufficient comparisons)

#### Supplemental figure 7AA: Marginal vitamin A status (RBP < 1.05 µmol/L) prevalence difference

##### 7AA5: Stratified by Season at the time of assessment

**P−for−interaction = 0.397**
**Difference in PDs = 0.04 (−0.05, 0.12)**

| Country | Trial | Dry: LNS N | Dry: Control N | Dry: Control Prevalence | Dry: PD (95% CI) | Dry: Fixed W | Dry: Random W | Rainy: LNS N | Rainy: Control N | Rainy: Control Prevalence | Rainy: PD (95% CI) | Rainy: Fixed W | Rainy: Random W |
|---|---|---|---|---|---|---|---|---|---|---|---|---|---|
| Bangladesh | JiVitA−4 (35) | | | | | | | | | | | | |
| Bangladesh | RDNS (36) | 341 | 166 | 28.9 | −0.04 (−0.12, 0.04) | 0.46 | 0.46 | 208 | 106 | 29.2 | −0.03 (−0.15, 0.09) | 0.29 | 0.28 |
| Bangladesh | WASH−B (37) | 96 | 93 | 44.1 | −0.13 (−0.26, 0.00) | 0.17 | 0.17 | 116 | 85 | 25.9 | 0.03 (−0.10, 0.17) | 0.22 | 0.24 |
| Burkina Faso | iLiNS−Zinc (38) | 174 | 61 | 50.7 | −0.13 (−0.24, −0.02) | 0.24 | 0.24 | 126 | 30 | 46.7 | −0.09 (−0.26, 0.08) | 0.14 | 0.18 |
| Burkina Faso | PROMIS CS (39) | | | | | | | | | | | | |
| Ghana | GHANA (40) | | | | | | | | | | | | |
| Ghana | iLiNS−DYADG (41) | | | | | | | | | | | | |
| Kenya | WASH−B (42) | 88 | 75 | 56.0 | −0.14 (−0.30, 0.02) | 0.12 | 0.12 | 210 | 184 | 60.9 | −0.18 (−0.29, −0.06) | 0.34 | 0.30 |
| Madagascar | MAHAY (43) | | | | | | | | | | | | |
| Malawi | iLiNS−DYADM (44) | | | | | | | | | | | | |
| Malawi | iLiNS−DOSE (45) | | | | | | | | | | | | |
| Mali | PROMIS CS (46) | | | | | | | | | | | | |
| Zimbabwe | SHINE (HIV−) (47) | | | | | | | | | | | | |
| Zimbabwe | SHINE (HIV+) (48) | | | | | | | | | | | | |
| | | **699** | **395** | | **I² = 0.00, Tau² = 0.00** | | | **660** | **405** | | **I² = 0.52, Tau² = 0.00** | | |
| **Fixed** | | | | | **−0.09 (−0.14, −0.04)** | | | | | | **−0.07 (−0.14, −0.01)** | | |
| **Random** | | | | | **−0.09 (−0.14, −0.04)** | | | | | | **−0.07 (−0.16, 0.02)** | | |

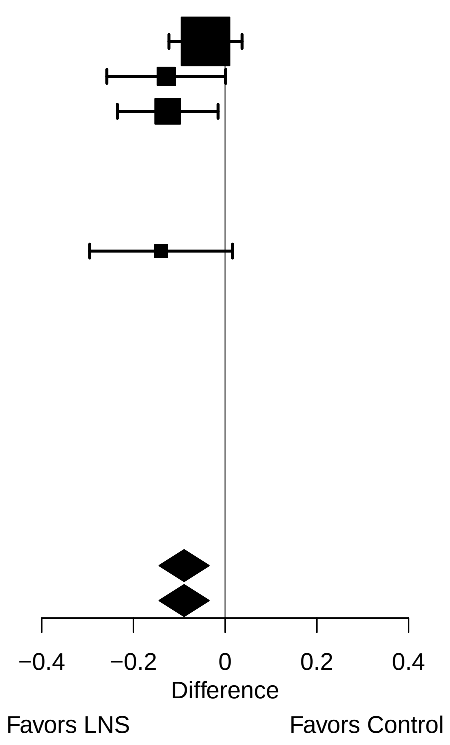

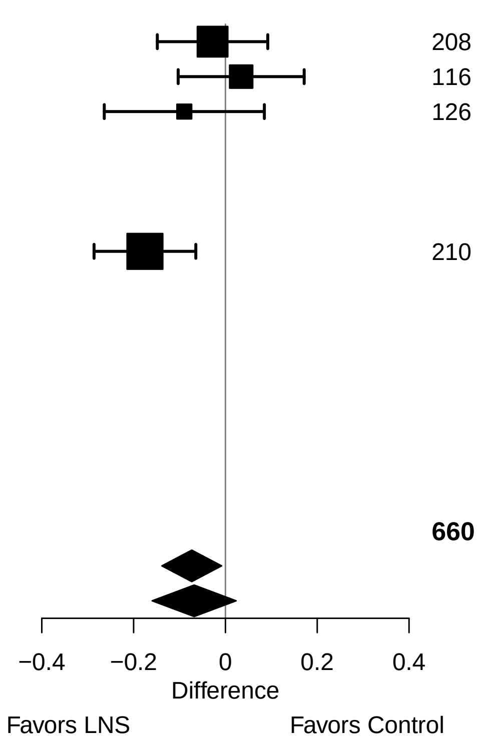

### Supplemental figure 8: Sensitivity analyses of effect modification of SQ-LNS on biochemical outcomes by individual-level effect modifiers

These figures show the pooled estimates of effect modification by different sensitivity analyses. For continuous outcomes, the intervention effect is measured by the difference in mean of the LNS group minus control. For log transformed continuous outcomes, the intervention effect is measured by the ratio of geometric means, the effect estimate is the geometric mean in the LNS group divided by the geometric mean in the control group. For dichotomous outcomes analyzed via prevalence ratios, the effect estimate is the prevalence in the LNS group divided by the prevalence in the control group. For dichotomous outcomes analyzed via prevalence differences, the effect estimate is the prevalence in the LNS group minus the prevalence in the control group. The labels on the left y-axis indicate which outcome is assessed. The different columns correspond to sensitivity analyses in which intervention group categorization differs. All-trial analysis includes all trials; Child-LNS-only excludes trial arms that provided both maternal and child LNS; Multi-component analysis separates comparisons within trials that included multi-component interventions, so that the SQ-LNS vs. no SQ-LNS comparisons were conducted separately between pairs of arms that included the same non-nutrition components (e.g. SQ-LNS+WASH vs. WASH; SQ-LNS vs. Control); Passive arms excluded analysis excludes passive control arms. Depending on the sensitivity analysis, there may not have been enough comparisons available to generate a pooled estimate.
sTfR, soluble transferrin receptor; ZPP, zinc protoporphyrin; RBP, retinol binding protein.

#### Supplemental figure 8A: Difference in mean differences in hemoglobin concentration

##### 8A1: By maternal effect modifiers

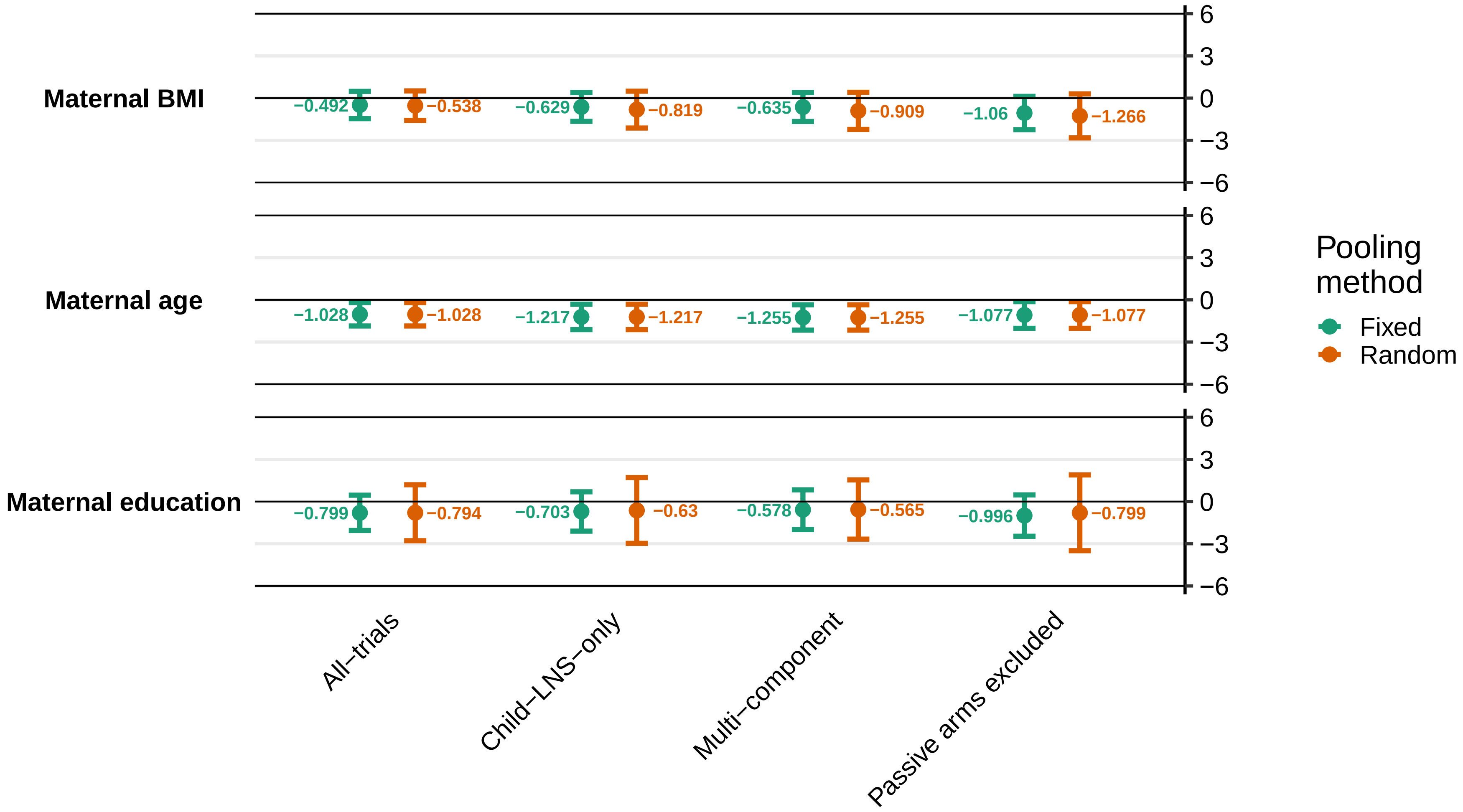

**Supplemental figure 8A: Difference in mean differences in hemoglobin concentration**

**8A2: By child effect modifiers**

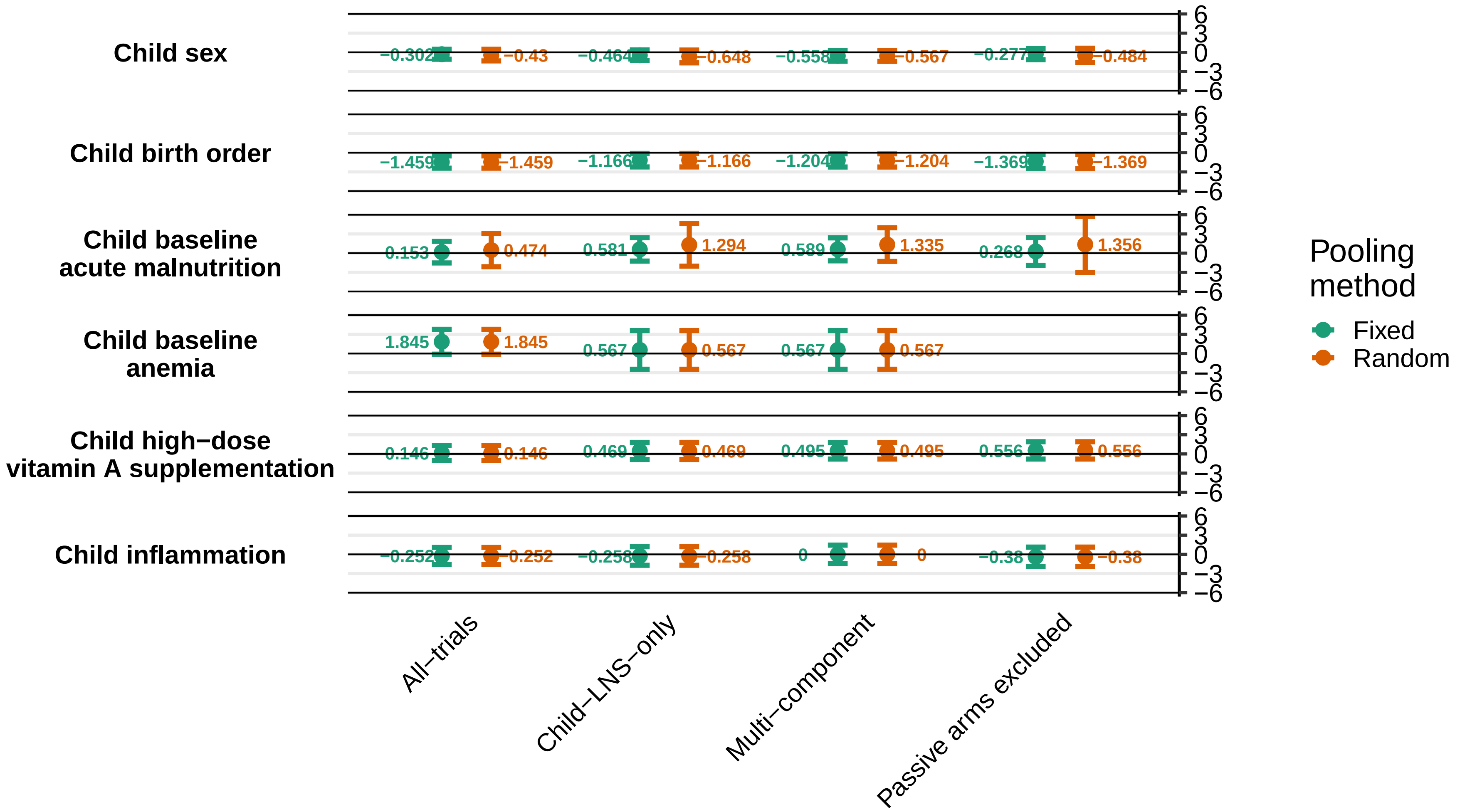

Supplemental figure 8A: Difference in mean differences in hemoglobin concentration

#### 8A3: By household effect modifiers

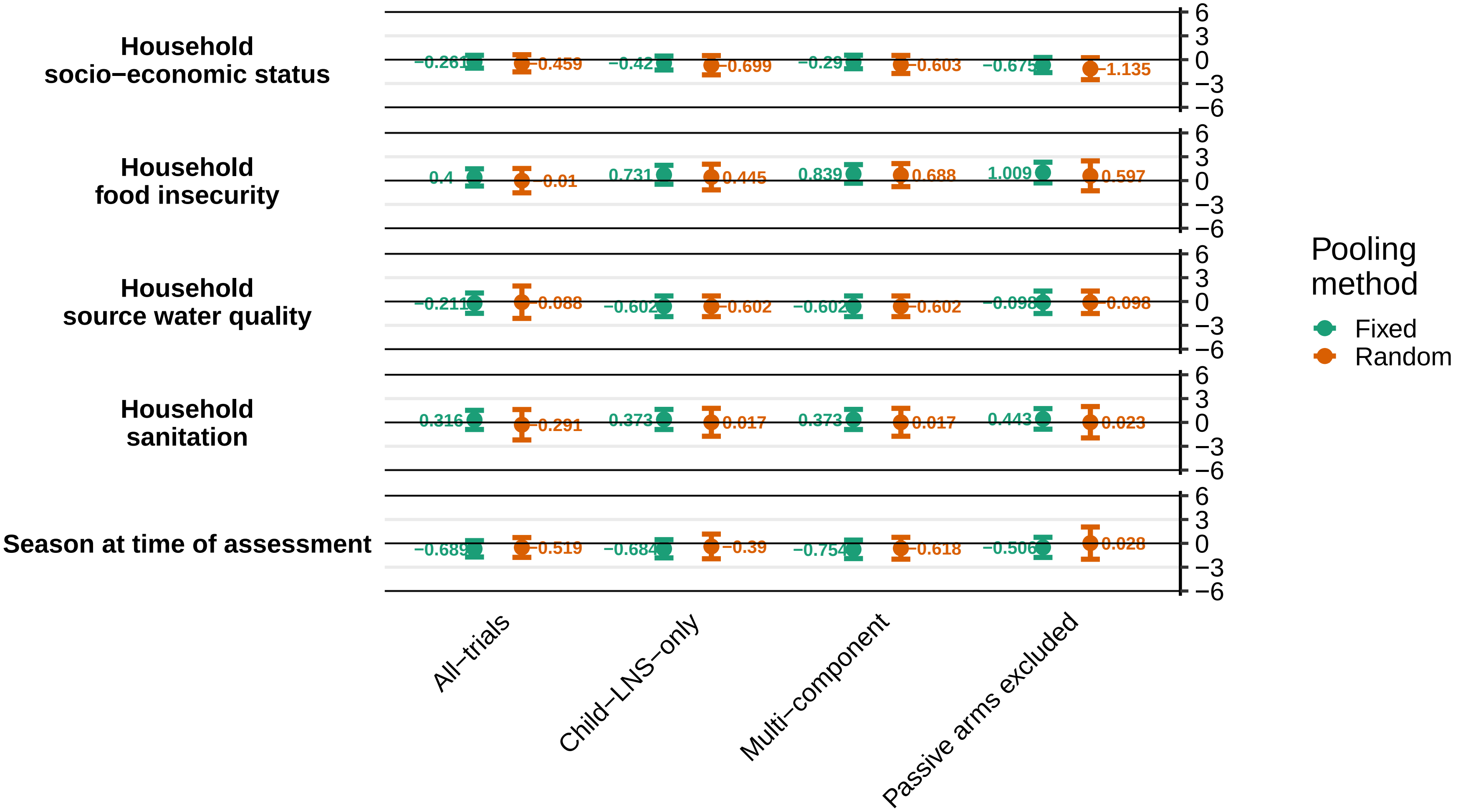

#### Supplemental figure 8B: Ratio of anemia prevalence ratios

##### 8B1: By maternal effect modifiers

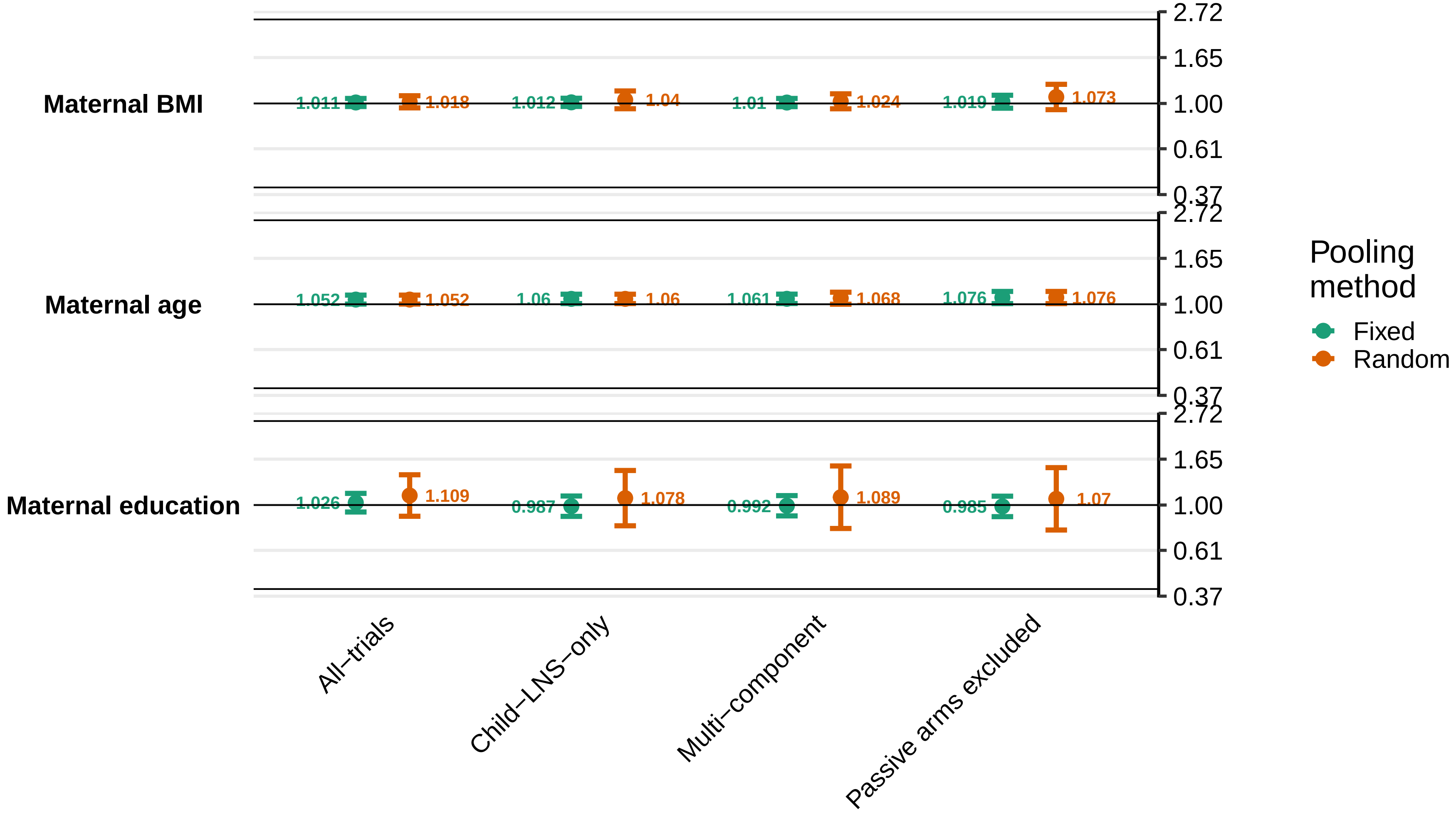

Supplemental figure 8B: Ratio of anemia prevalence ratios

#### 8B2: By child effect modifiers

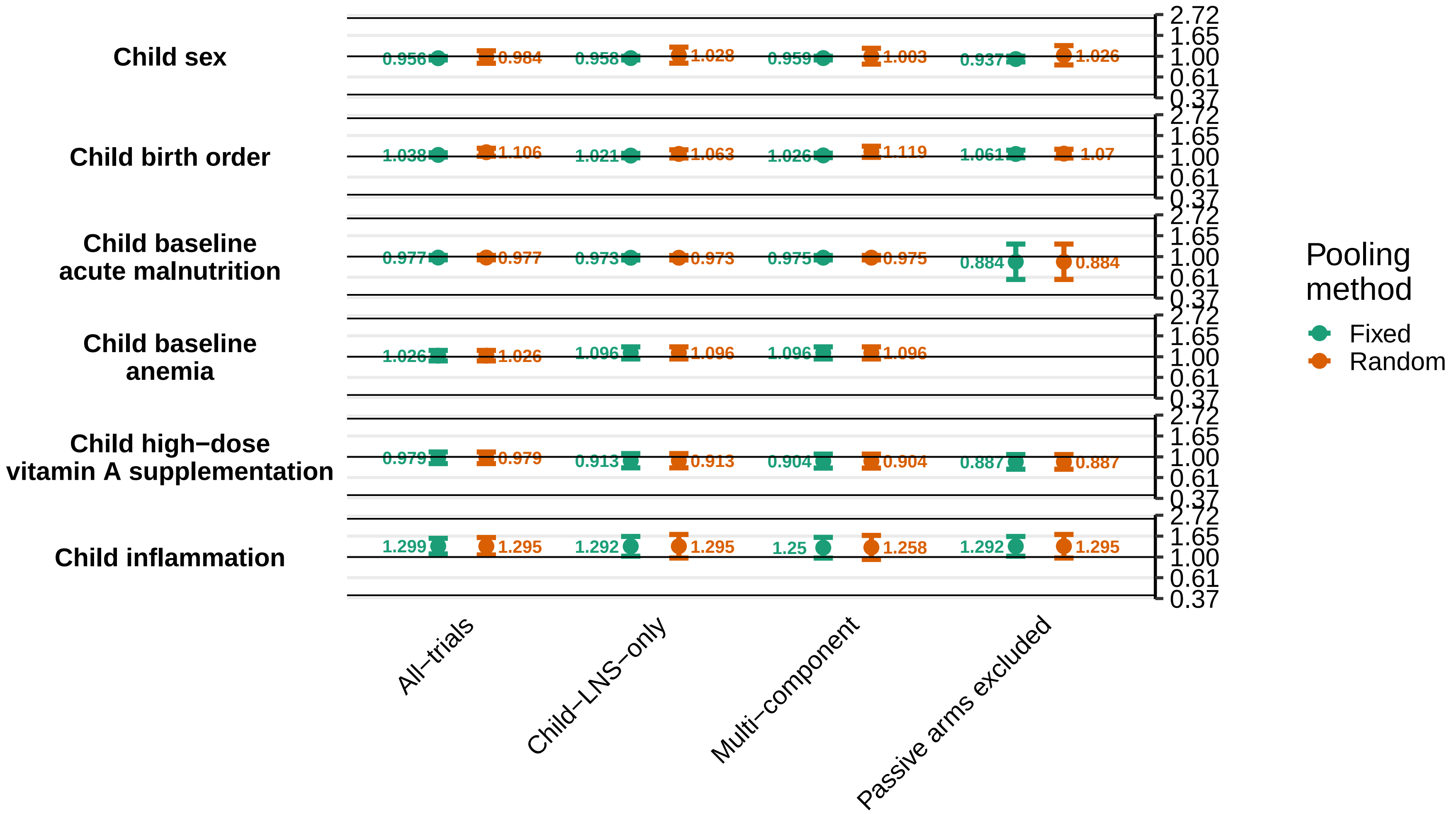

Supplemental figure 8B: Ratio of anemia prevalence ratios

#### 8B3: By household effect modifiers

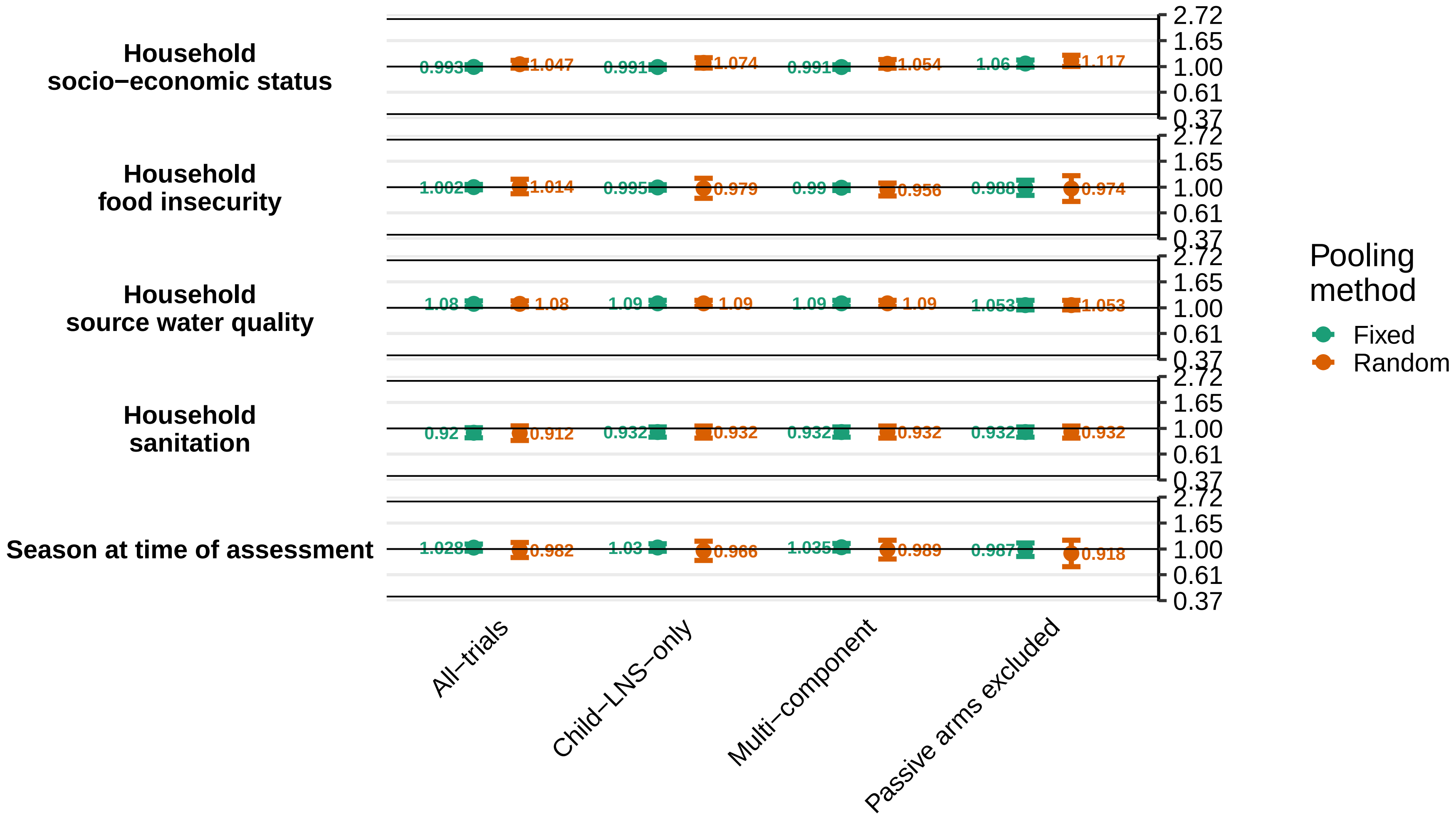

#### Supplemental figure 8C: Difference in anemia prevalence differences

##### 8C1: By maternal effect modifiers

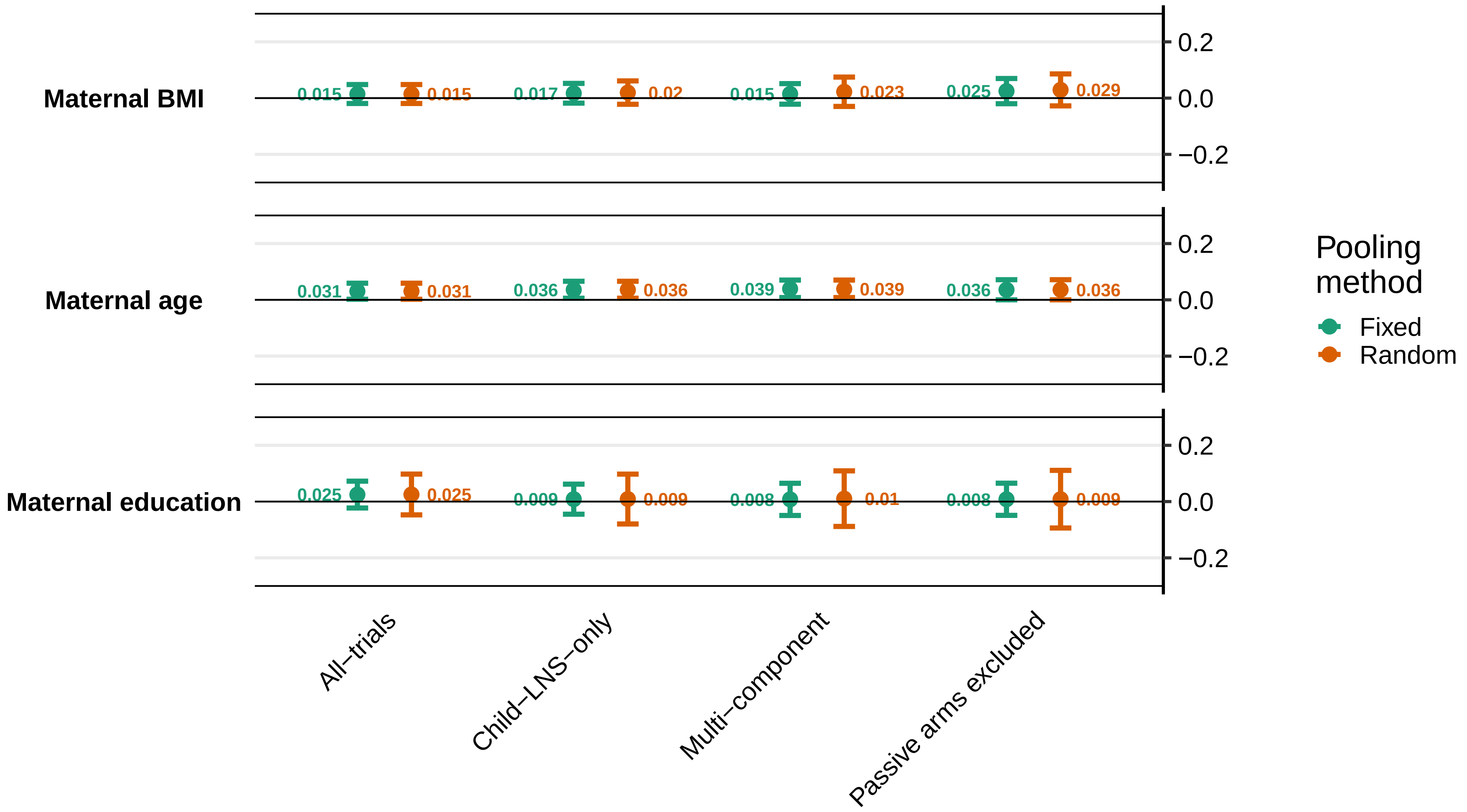

Supplemental figure 8C: Difference in anemia prevalence differences

8C2: By child effect modifiers

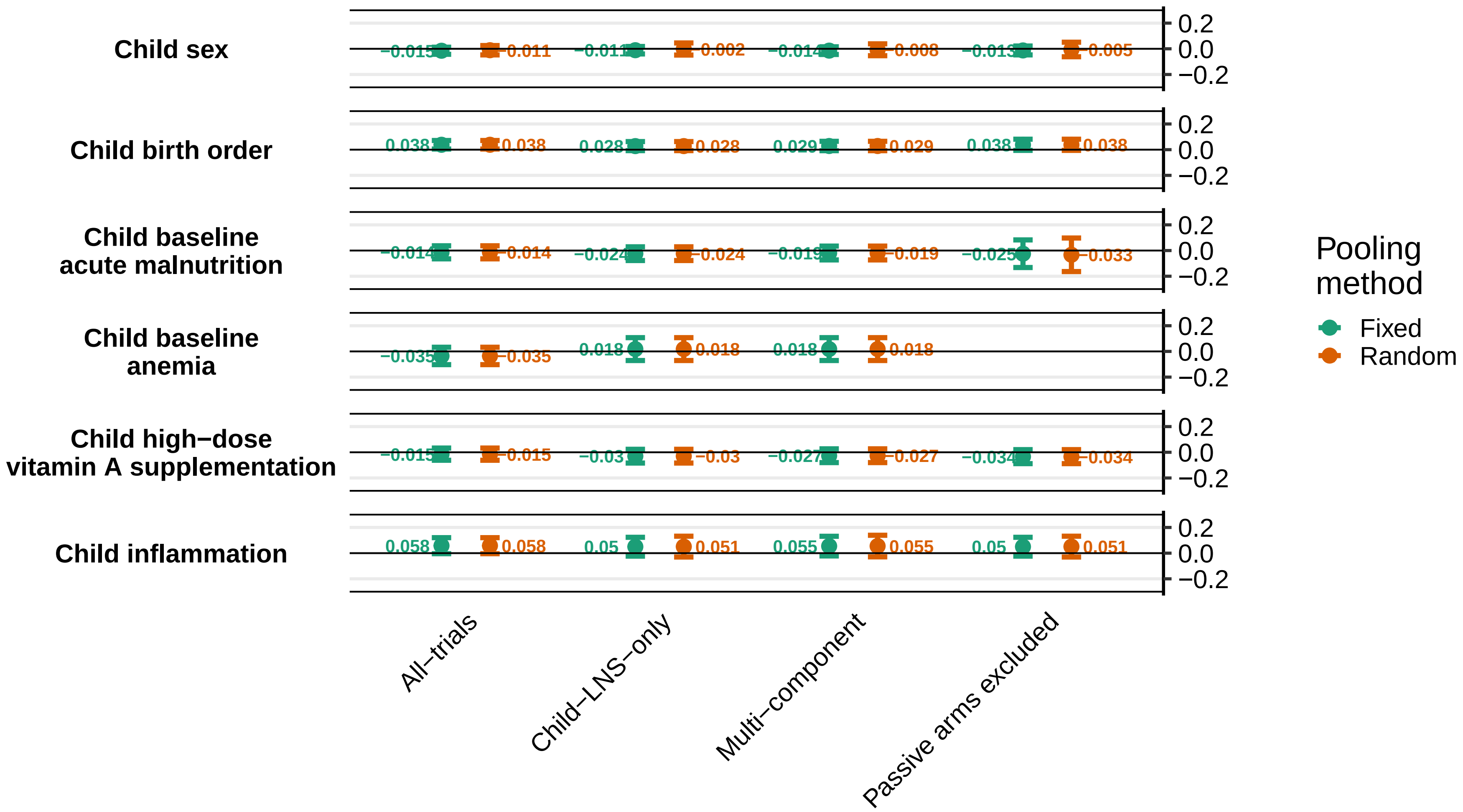

Supplemental figure 8C: Difference in anemia prevalence differences

#### 8C3: By household effect modifiers

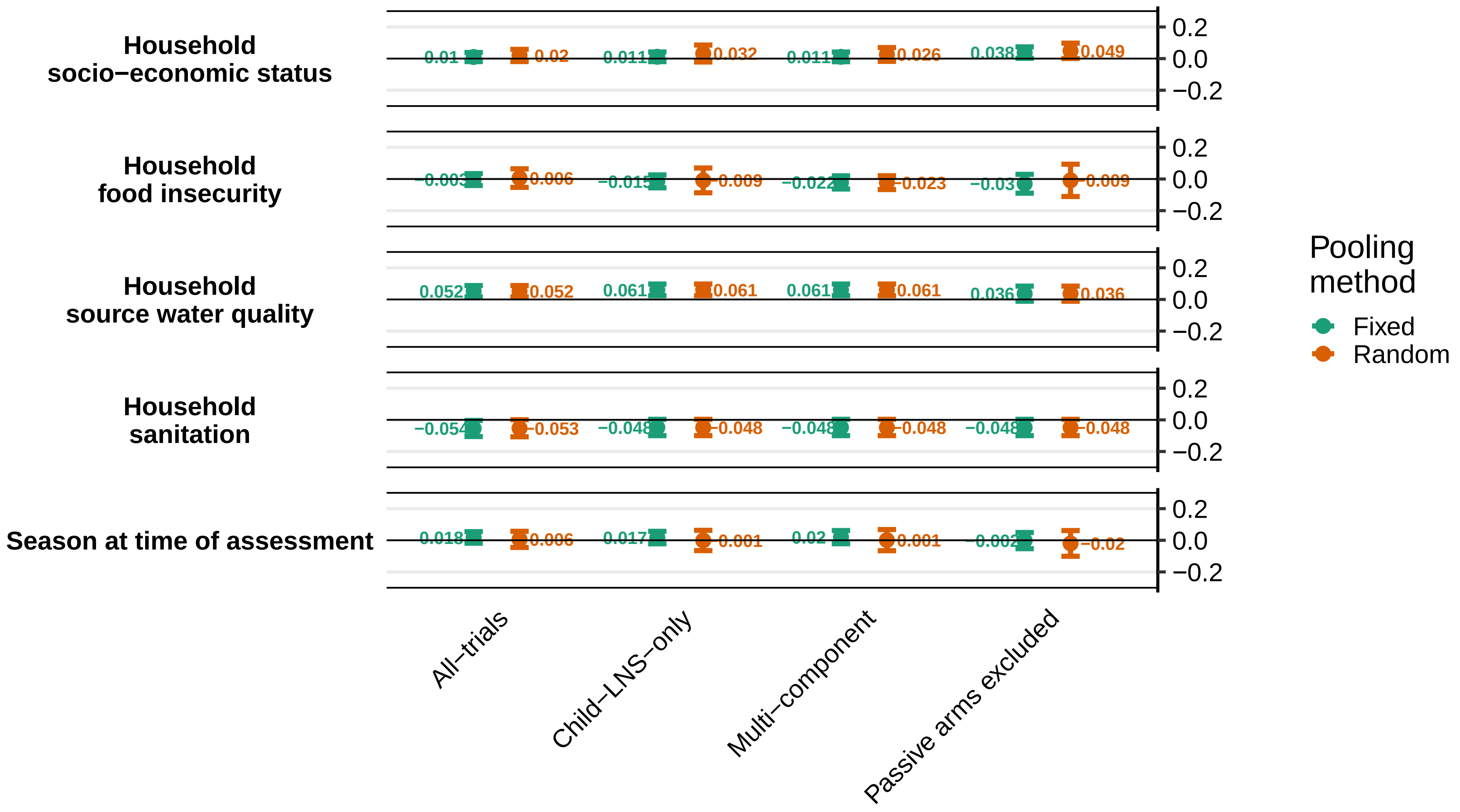

#### Supplemental figure 8D: Ratio of moderate-to-severe anemia prevalence ratios

##### 8D1: By maternal effect modifiers

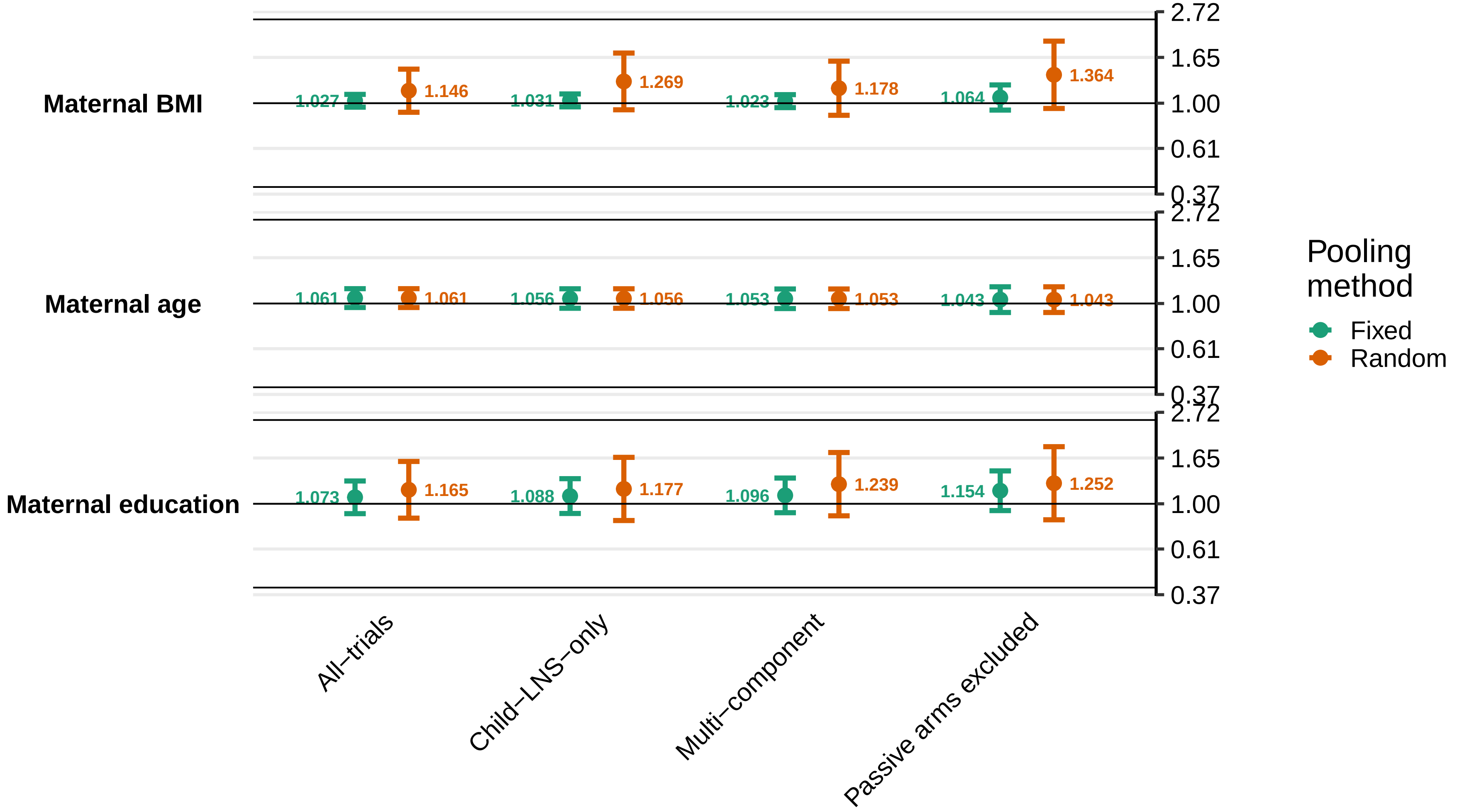

Supplemental figure 8D: Ratio of moderate-to-severe anemia prevalence ratios

8D2: By child effect modifiers

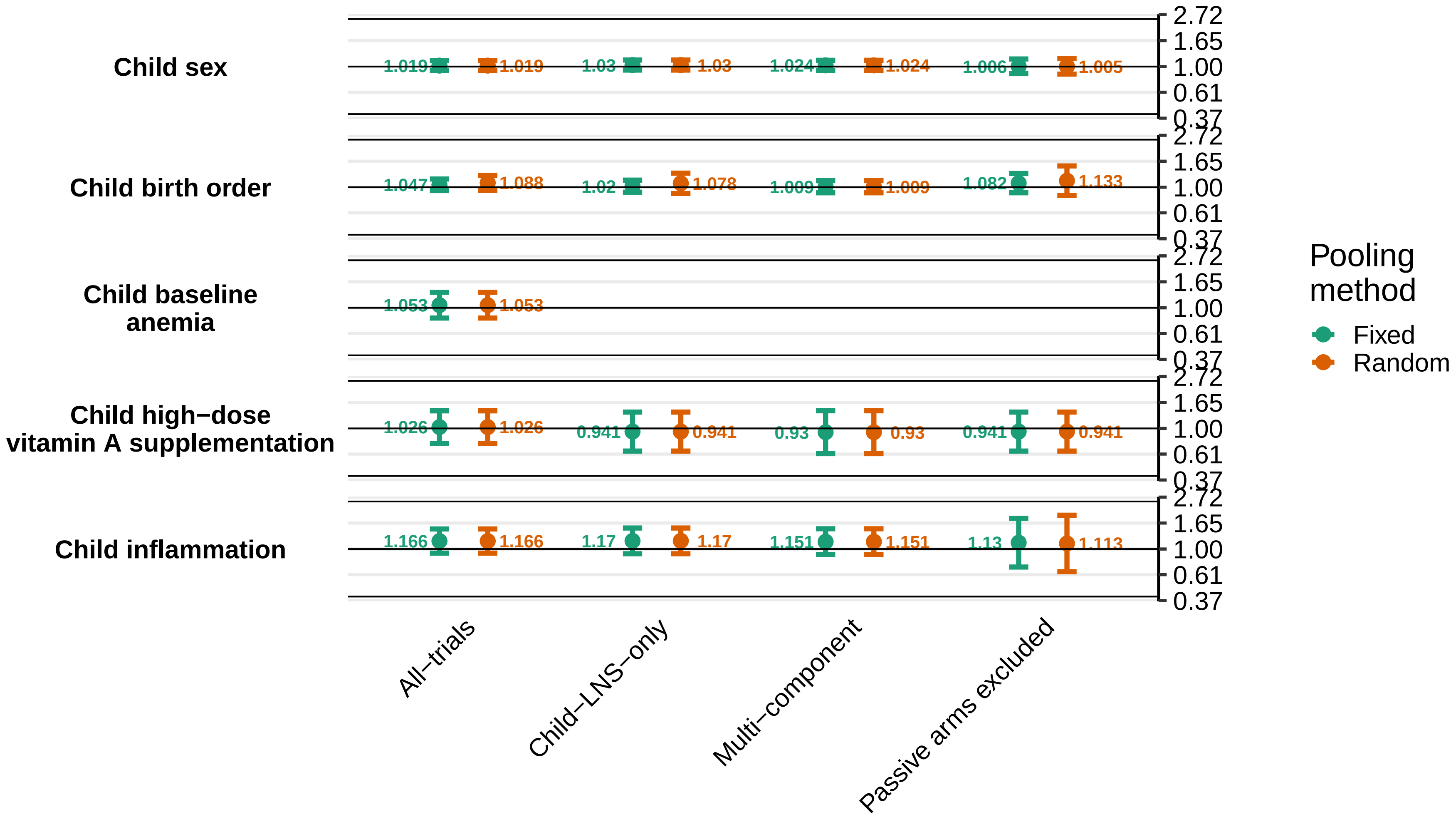

Supplemental figure 8D: Ratio of moderate-to-severe anemia prevalence ratios

#### 8D3: By household effect modifiers

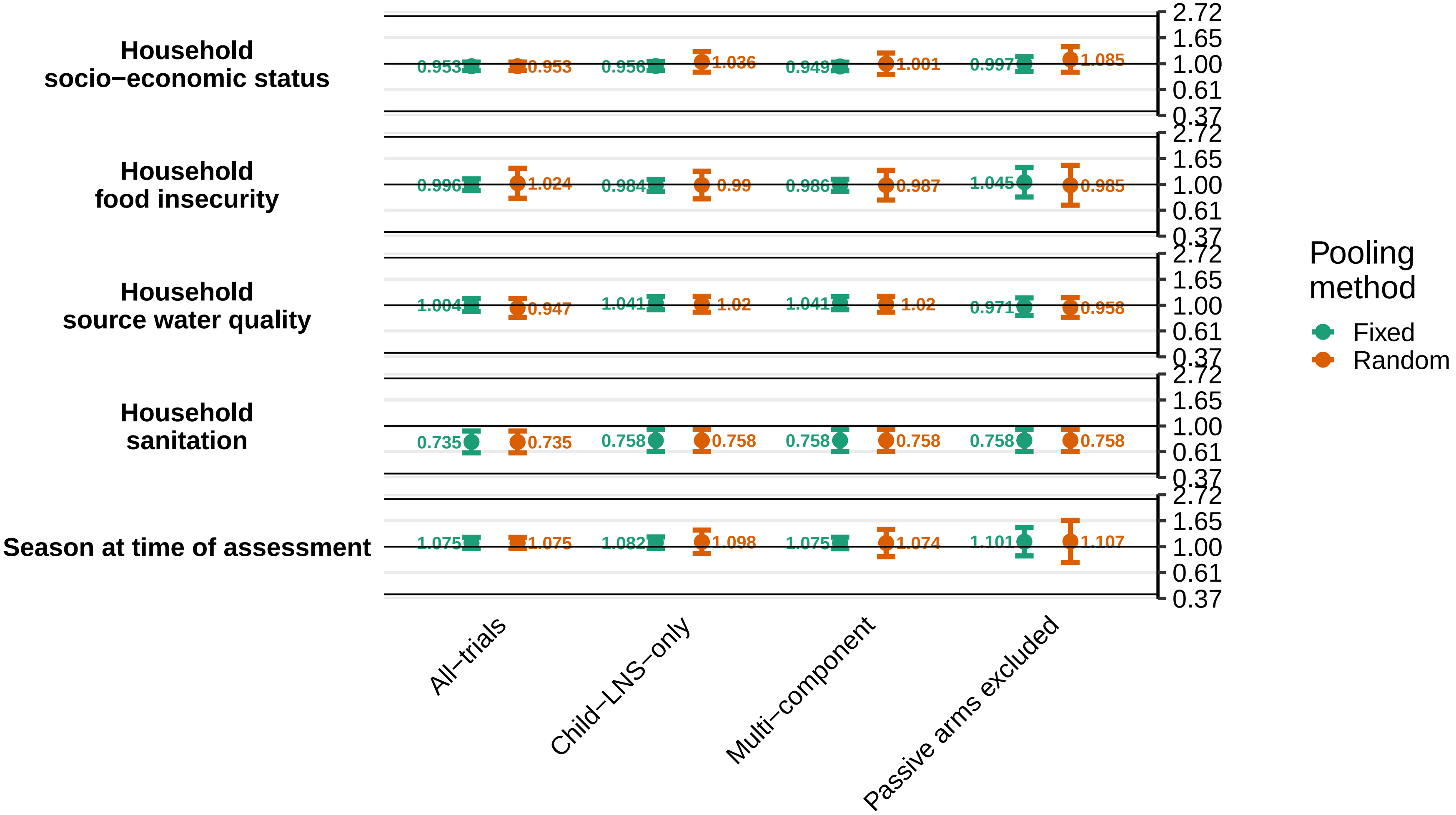

#### Supplemental figure 8E: Difference in moderate-to-severe anemia prevalence differences

##### 8E1: By maternal effect modifiers

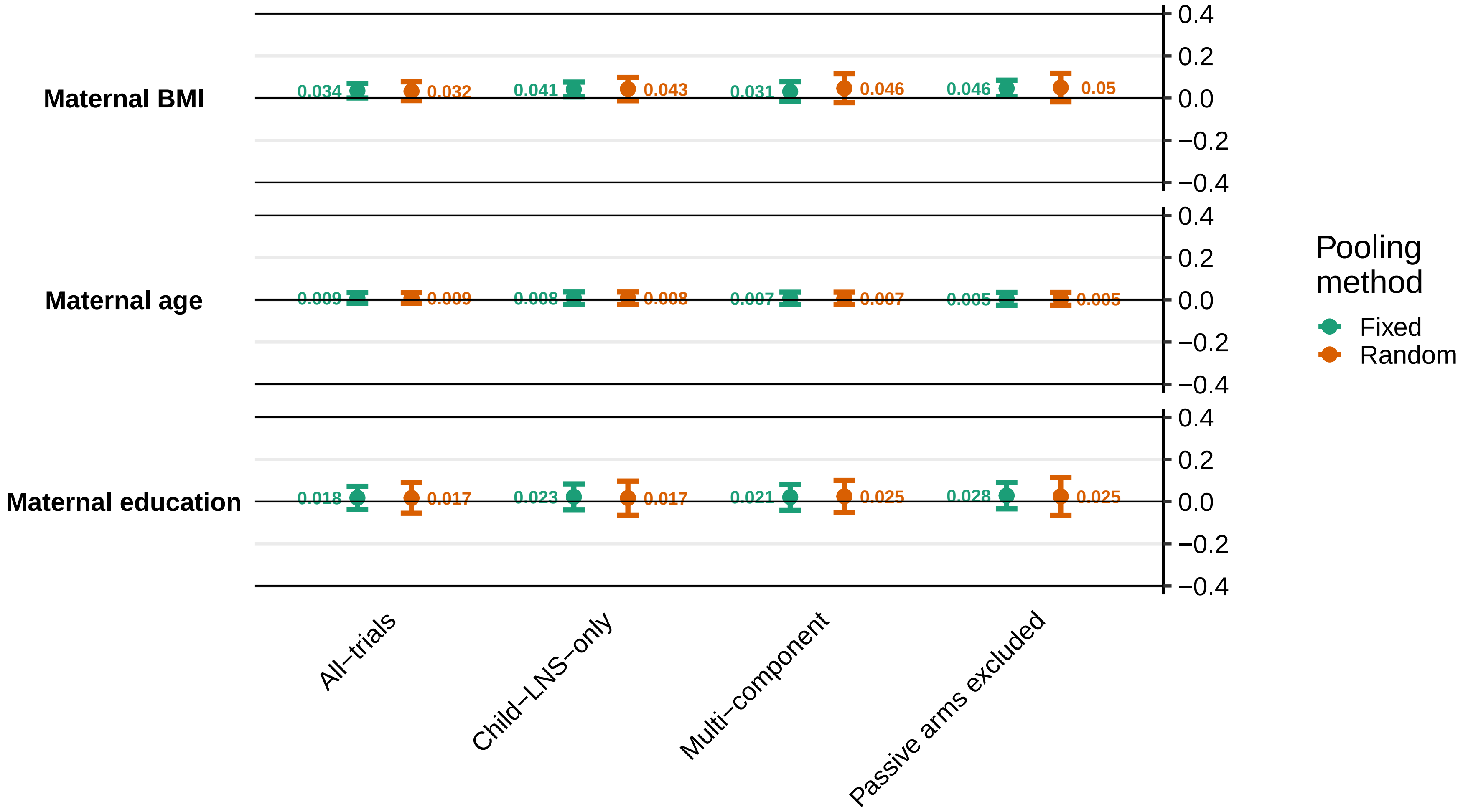

Supplemental figure 8E: Difference in moderate-to-severe anemia prevalence differences

#### 8E2: By child effect modifiers

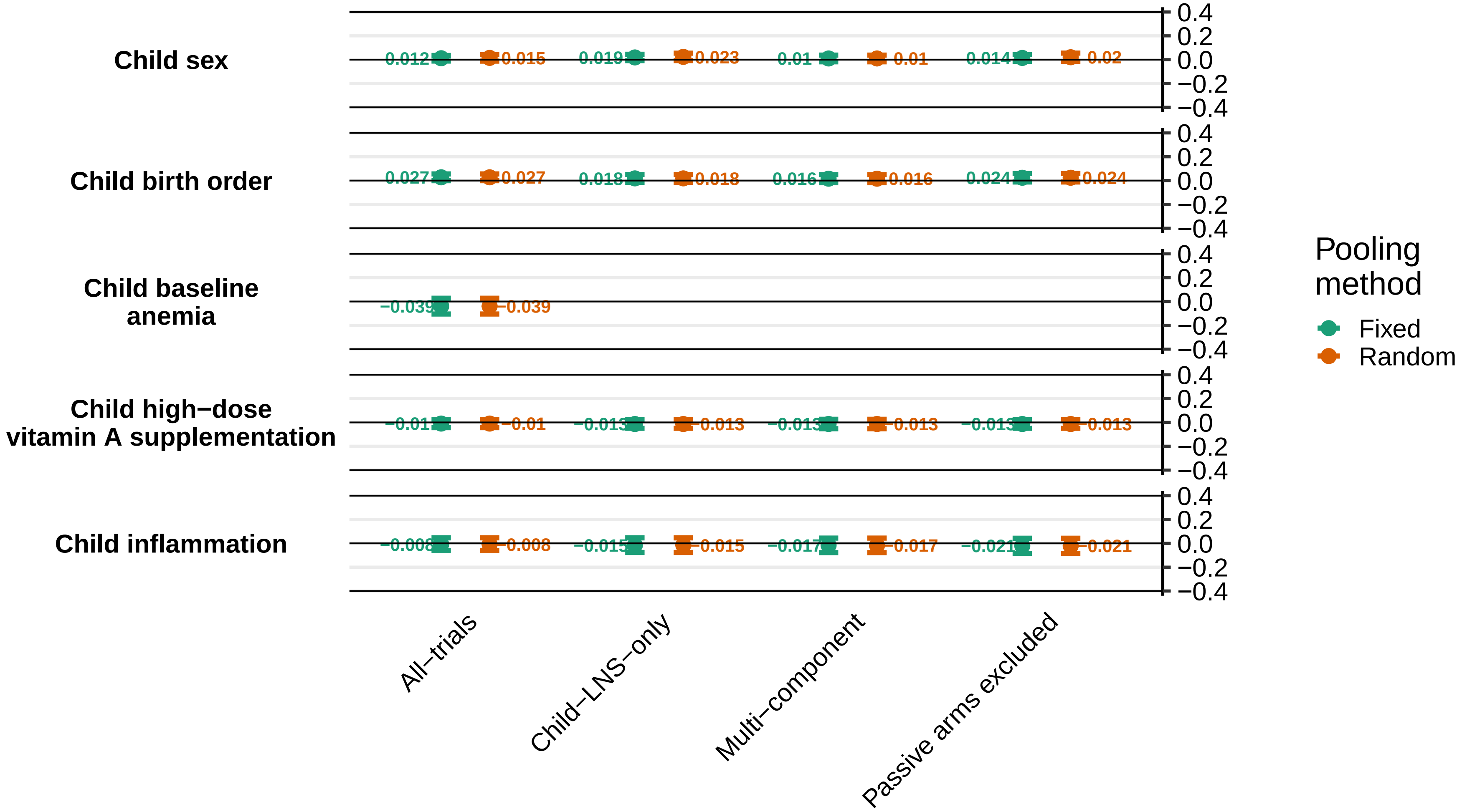

Supplemental figure 8E: Difference in moderate-to-severe anemia prevalence differences

#### 8E3: By household effect modifiers

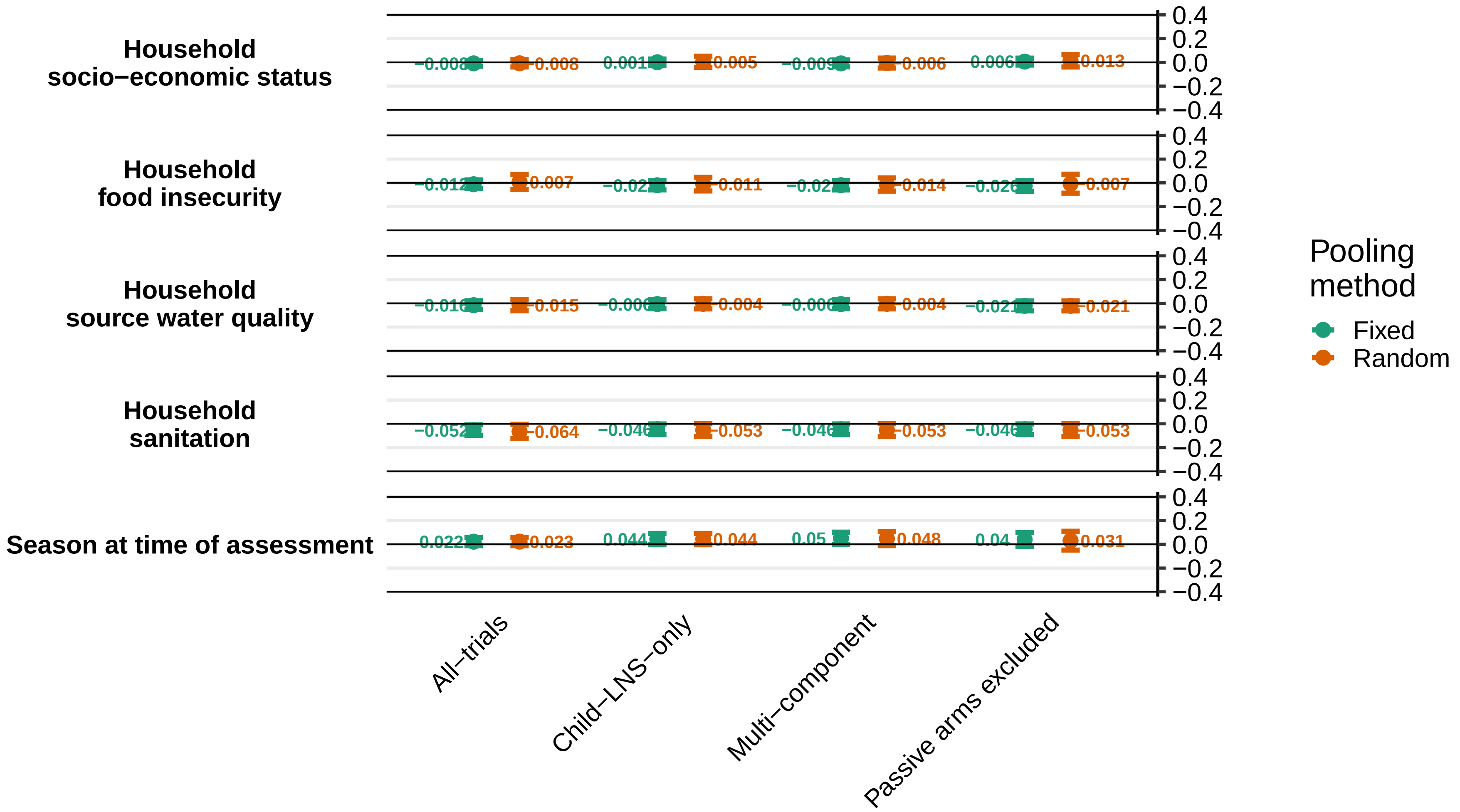

#### Supplemental figure 8F: Ratio of geometric mean ratios of ferritin concentration

##### 8F1: By maternal effect modifiers

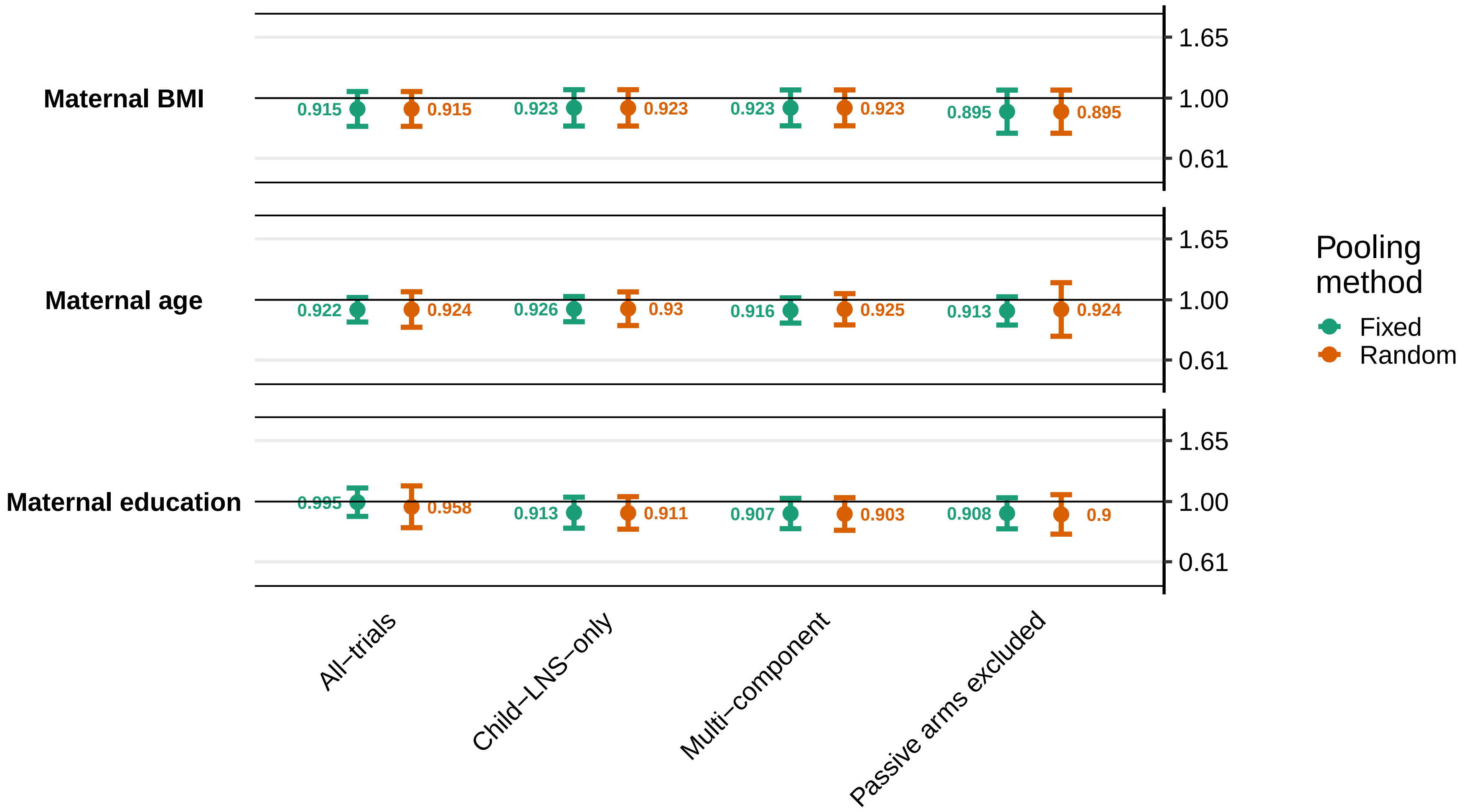

Supplemental figure 8F: Ratio of geometric mean ratios of ferritin concentration

#### 8F2: By child effect modifiers

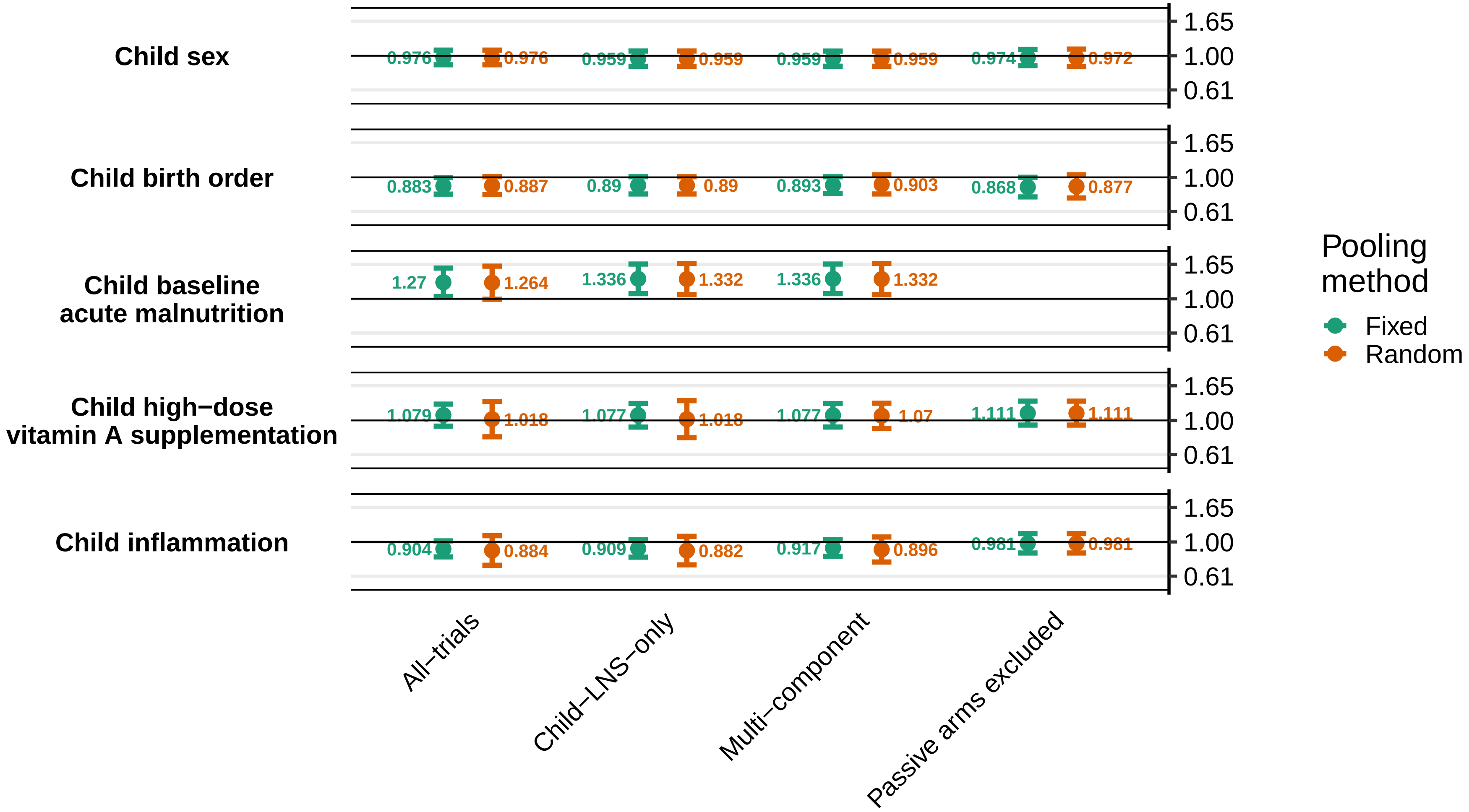

Supplemental figure 8F: Ratio of geometric mean ratios of ferritin concentration

#### 8F3: By household effect modifiers

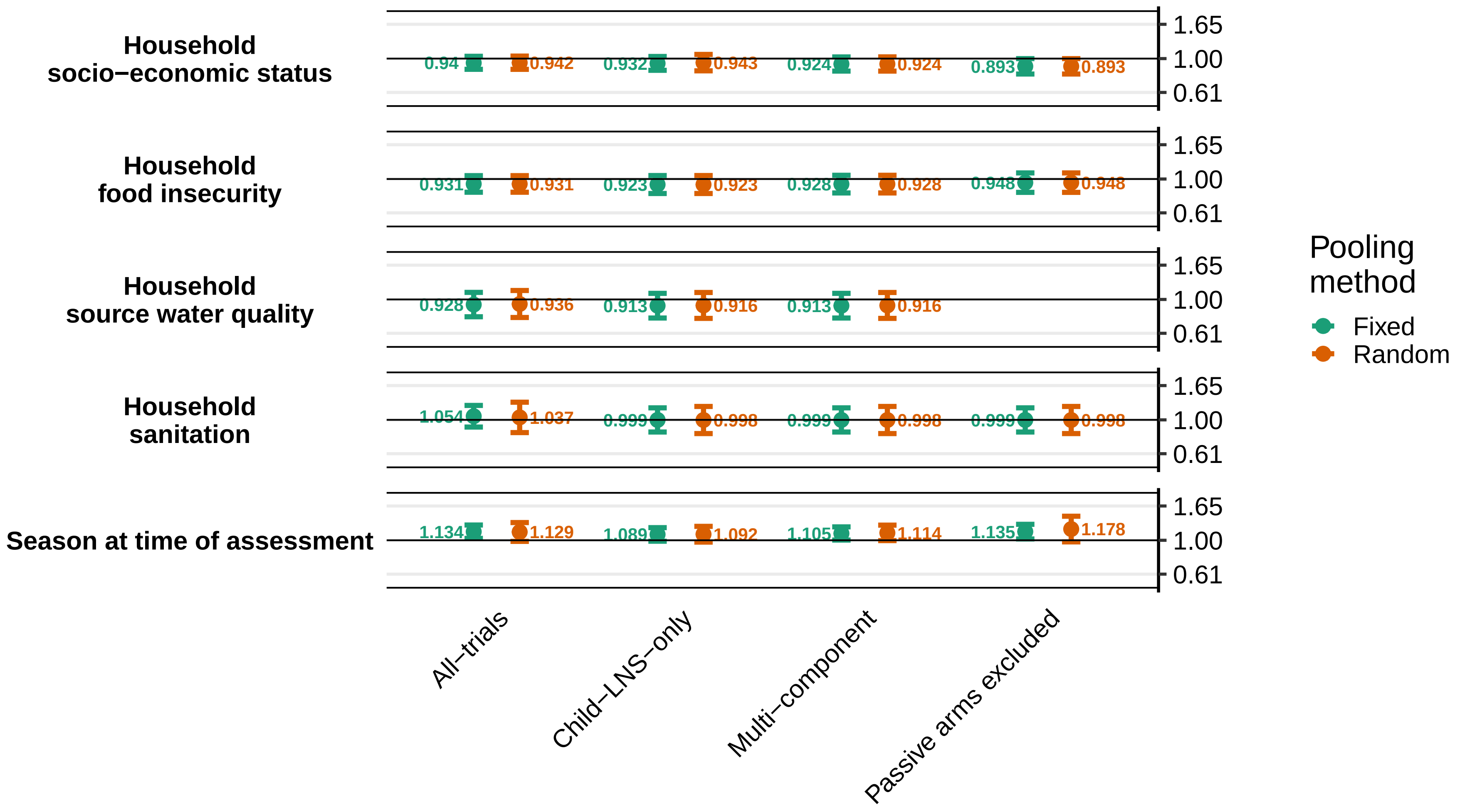

#### Supplemental figure 8G: Ratio of iron deficiency (ferritin < 12 µg/L) prevalence ratios

##### 8G1: By maternal effect modifiers

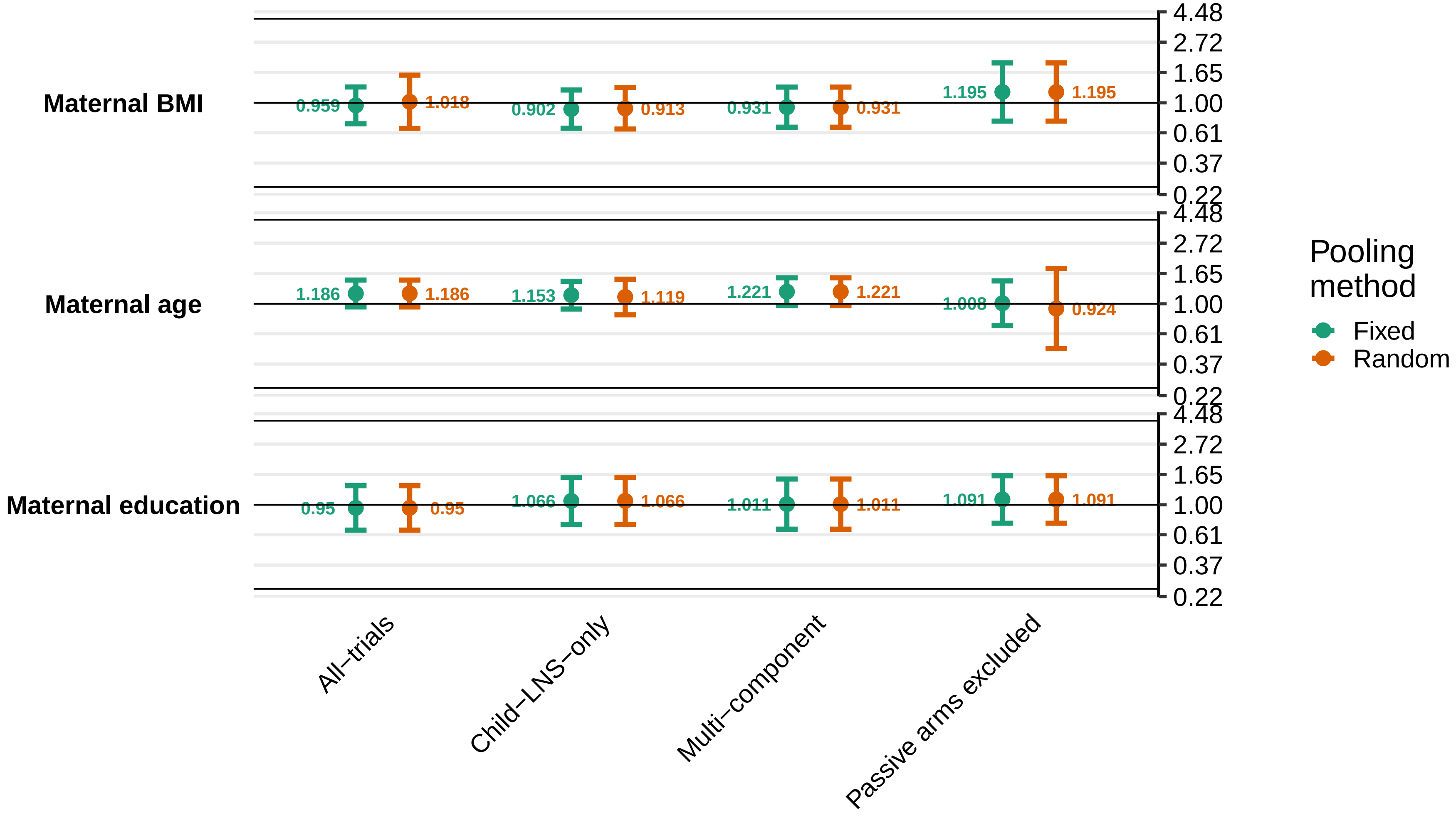

Supplemental figure 8G: Ratio of iron deficiency (ferritin < 12 µg/L) prevalence ratios

#### 8G2: By child effect modifiers

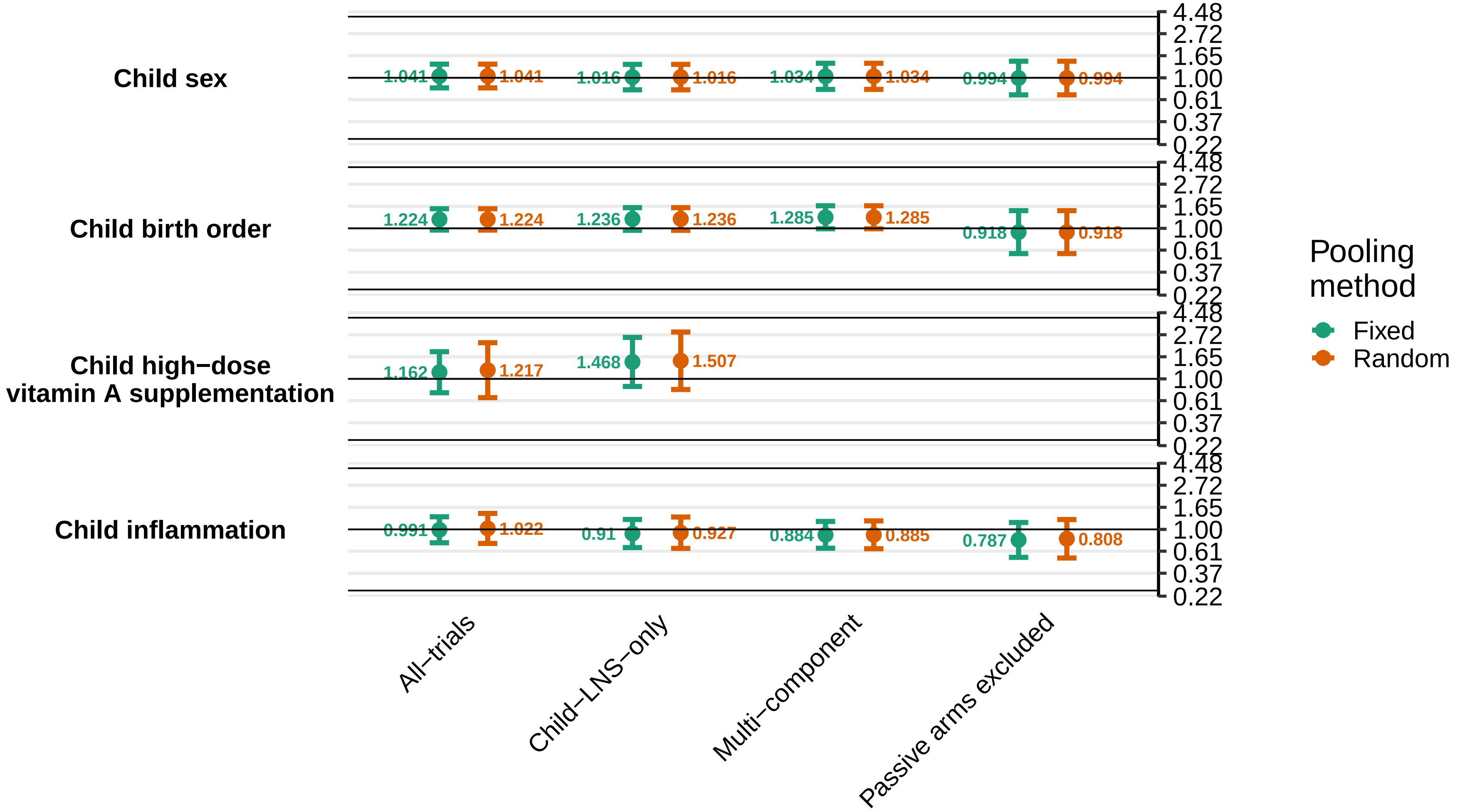

Supplemental figure 8G: Ratio of iron deficiency (ferritin < 12 µg/L) prevalence ratios

#### 8G3: By household effect modifiers

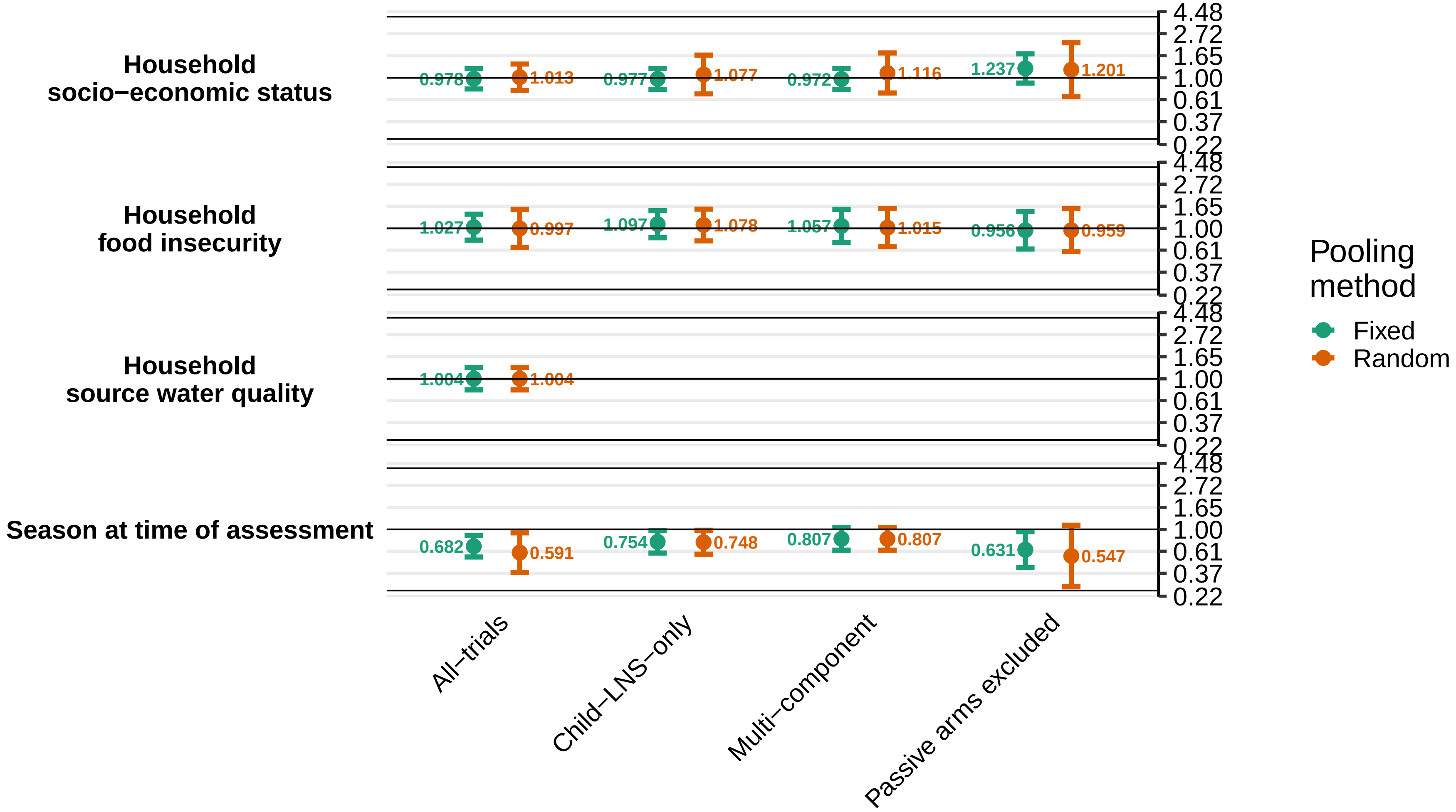

#### Supplemental figure 8H: Difference in iron deficiency (ferritin < 12 µg/L) prevalence differences

##### 8H1: By maternal effect modifiers

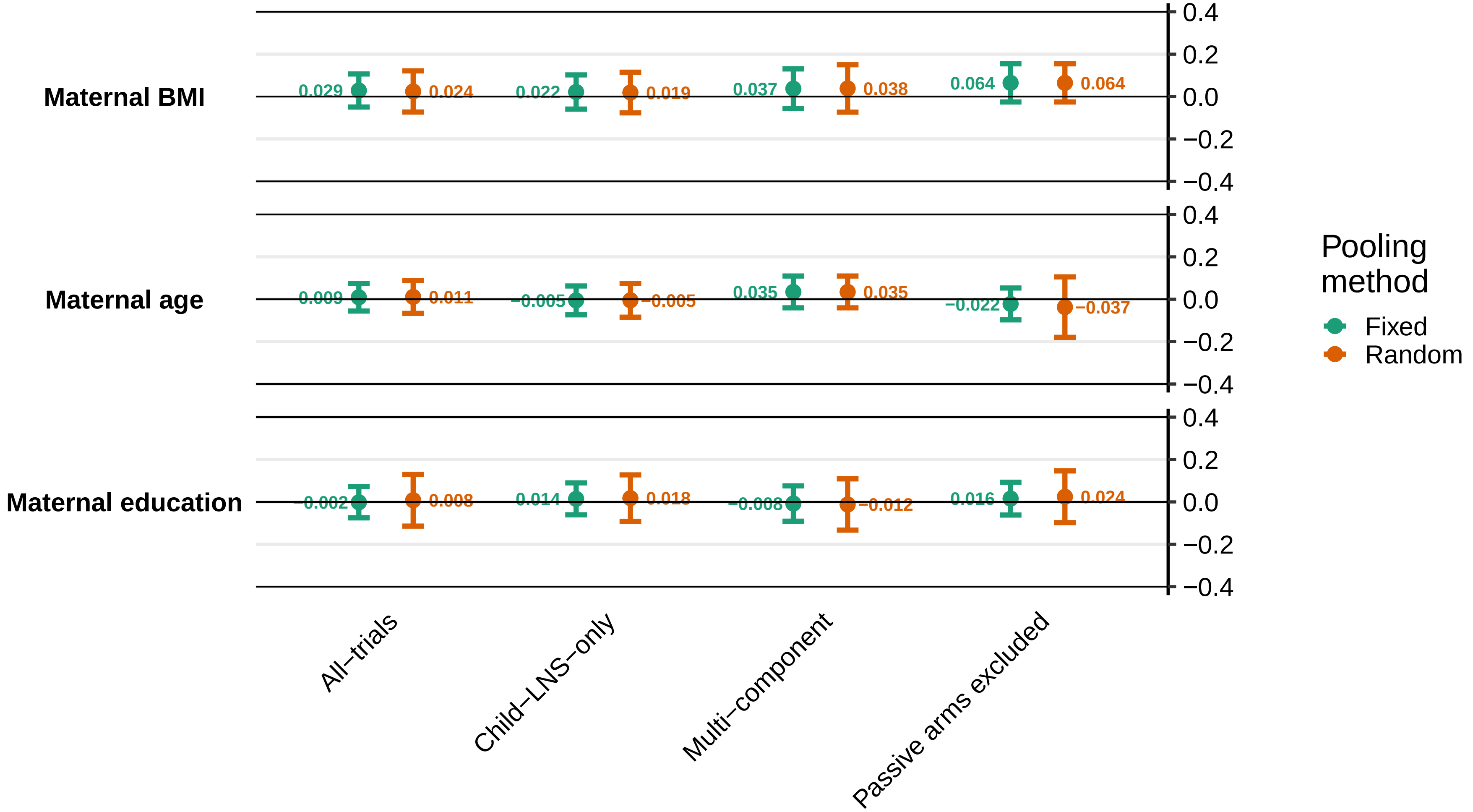

**Supplemental figure 8H: Difference in iron deficiency (ferritin < 12 µg/L) prevalence differences**

**8H2: By child effect modifiers**

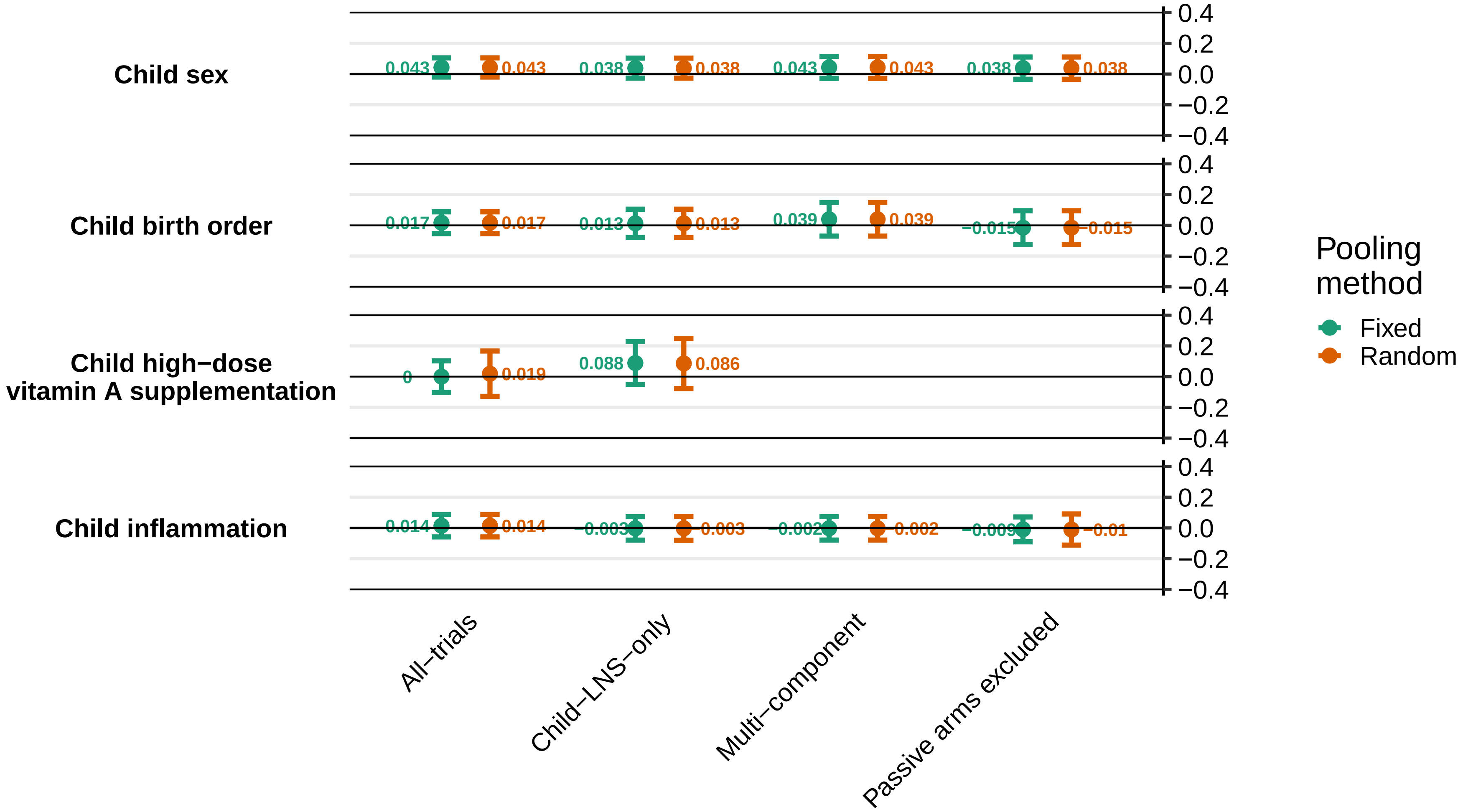

**Supplemental figure 8H: Difference in iron deficiency (ferritin < 12 µg/L) prevalence differences**

#### 8H3: By household effect modifiers

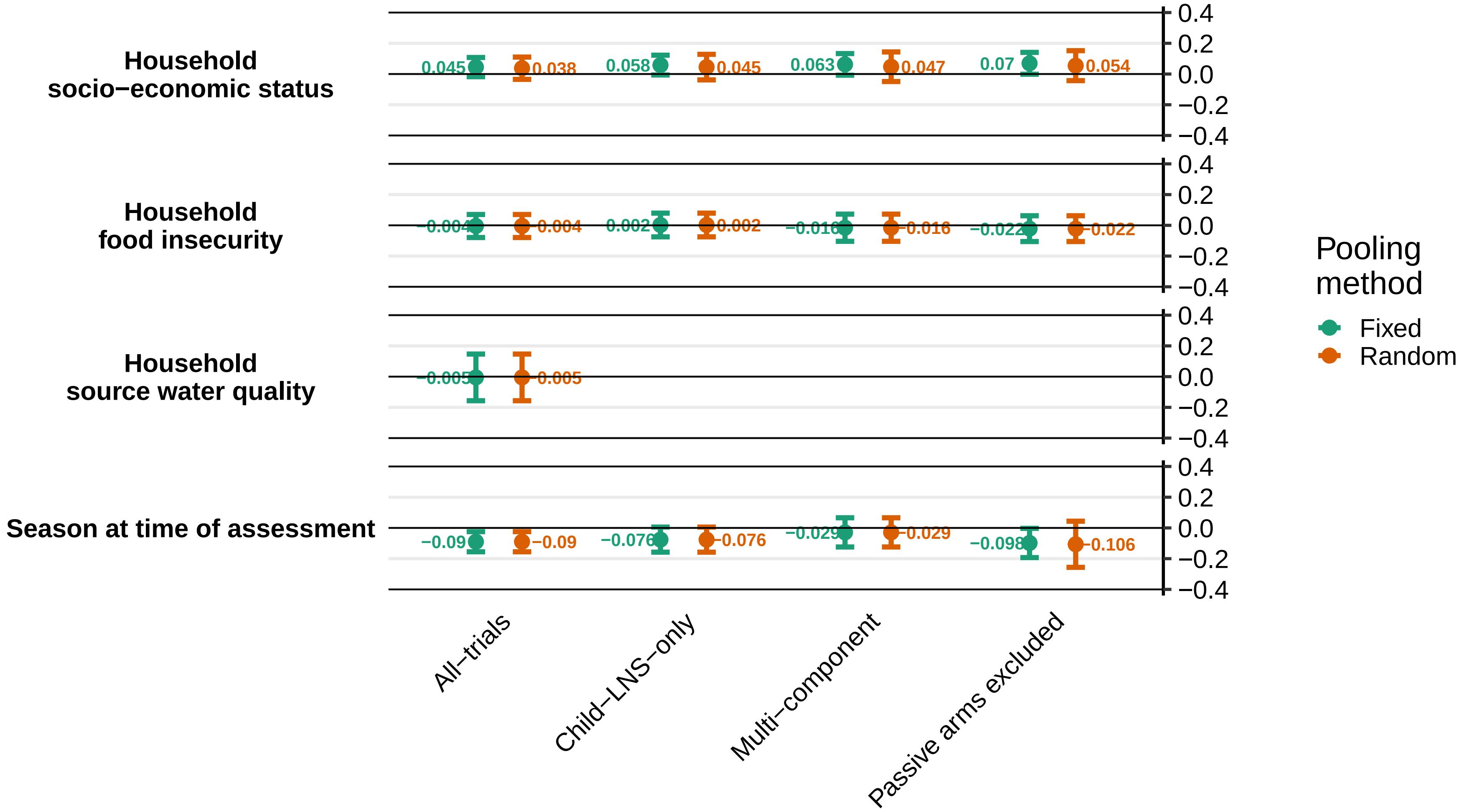

#### Supplemental figure 8I: Ratio of iron deficiency anemia prevalence ratios

##### 8I1: By maternal effect modifiers

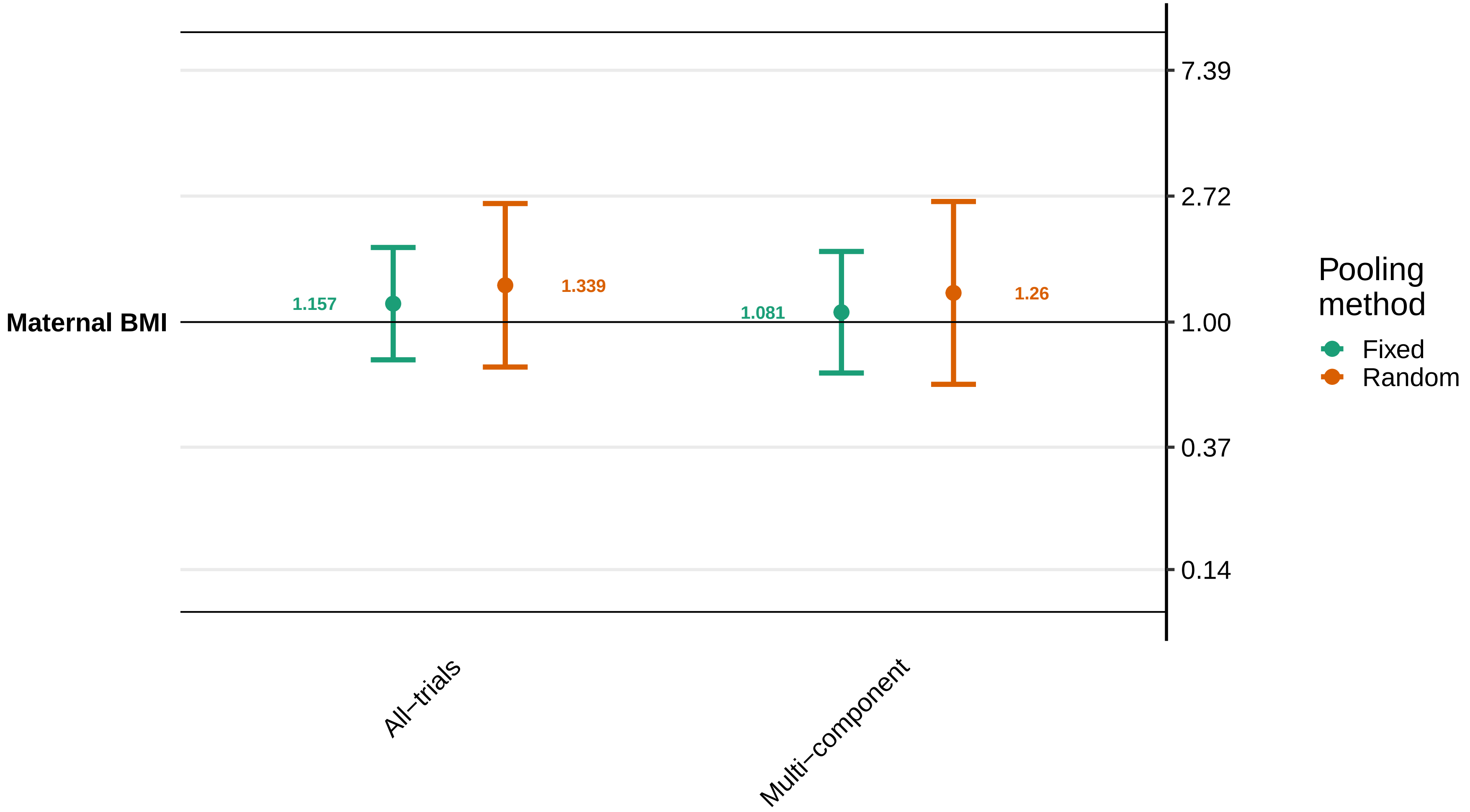

**Supplemental figure 8I: Ratio of iron deficiency anemia prevalence ratios**

#### 8I2: By child effect modifiers

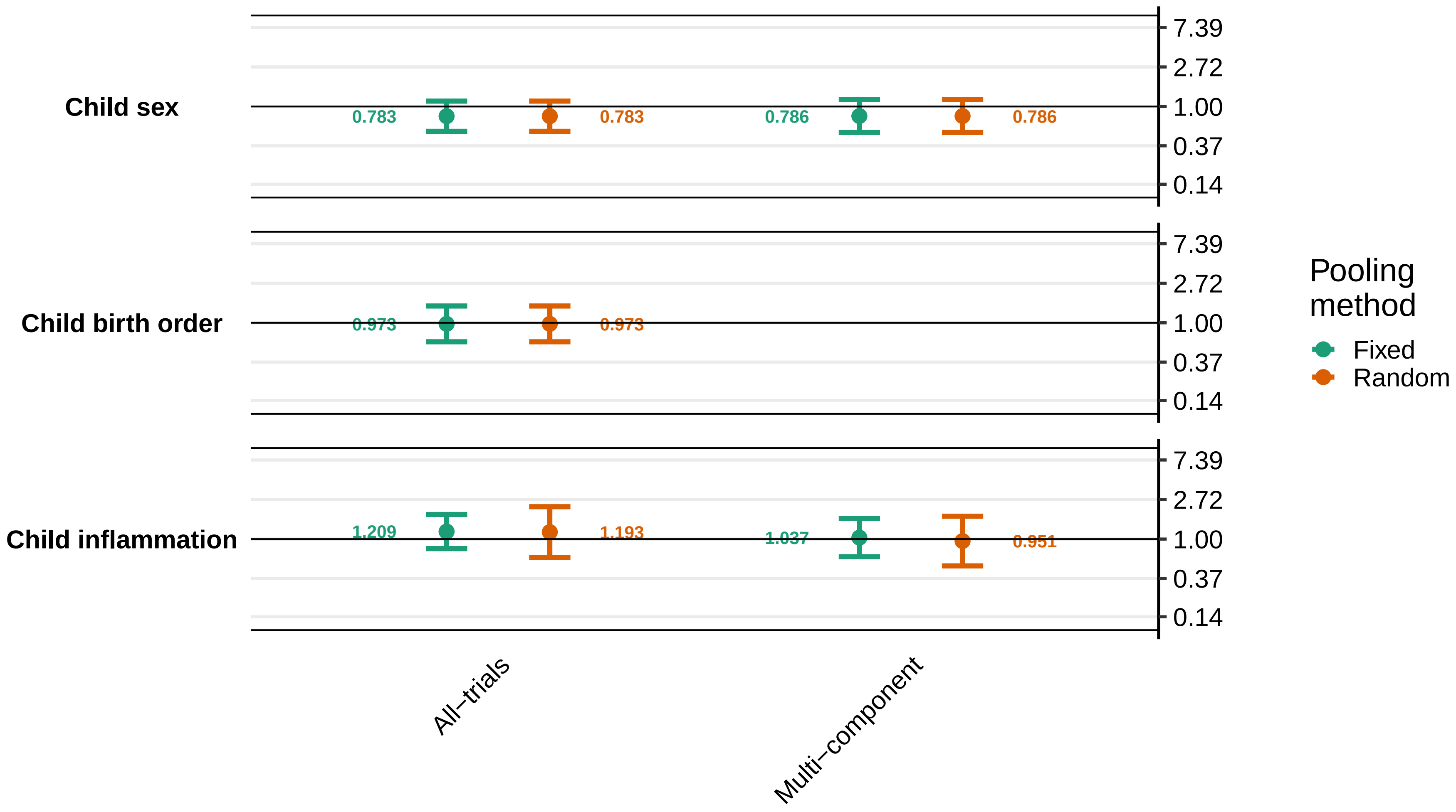

Supplemental figure 8I: Ratio of iron deficiency anemia prevalence ratios

##### 8I3: By household effect modifiers

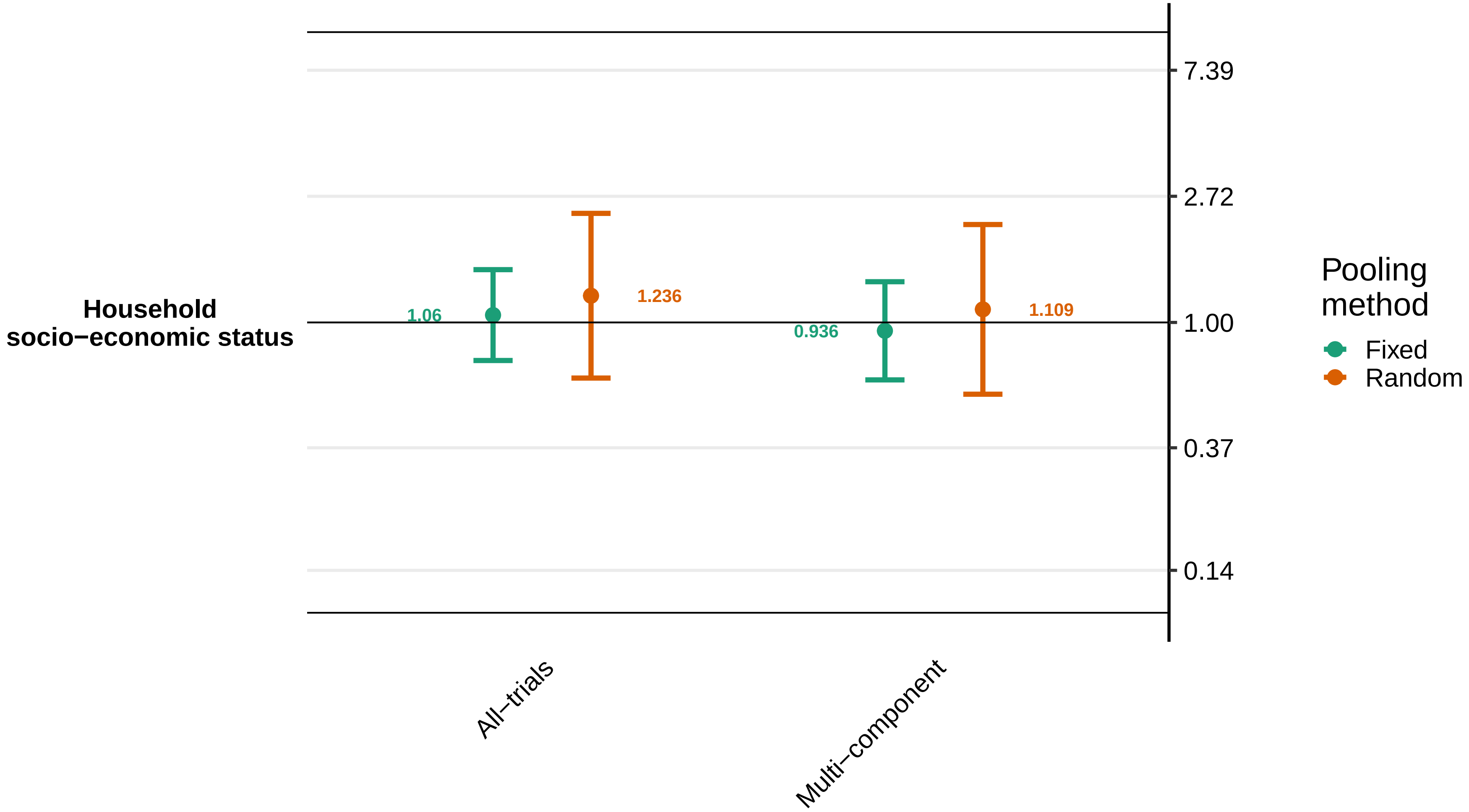

#### Supplemental figure 8J: Difference in iron deficiency anemia prevalence differences

##### 8J1: By maternal effect modifiers

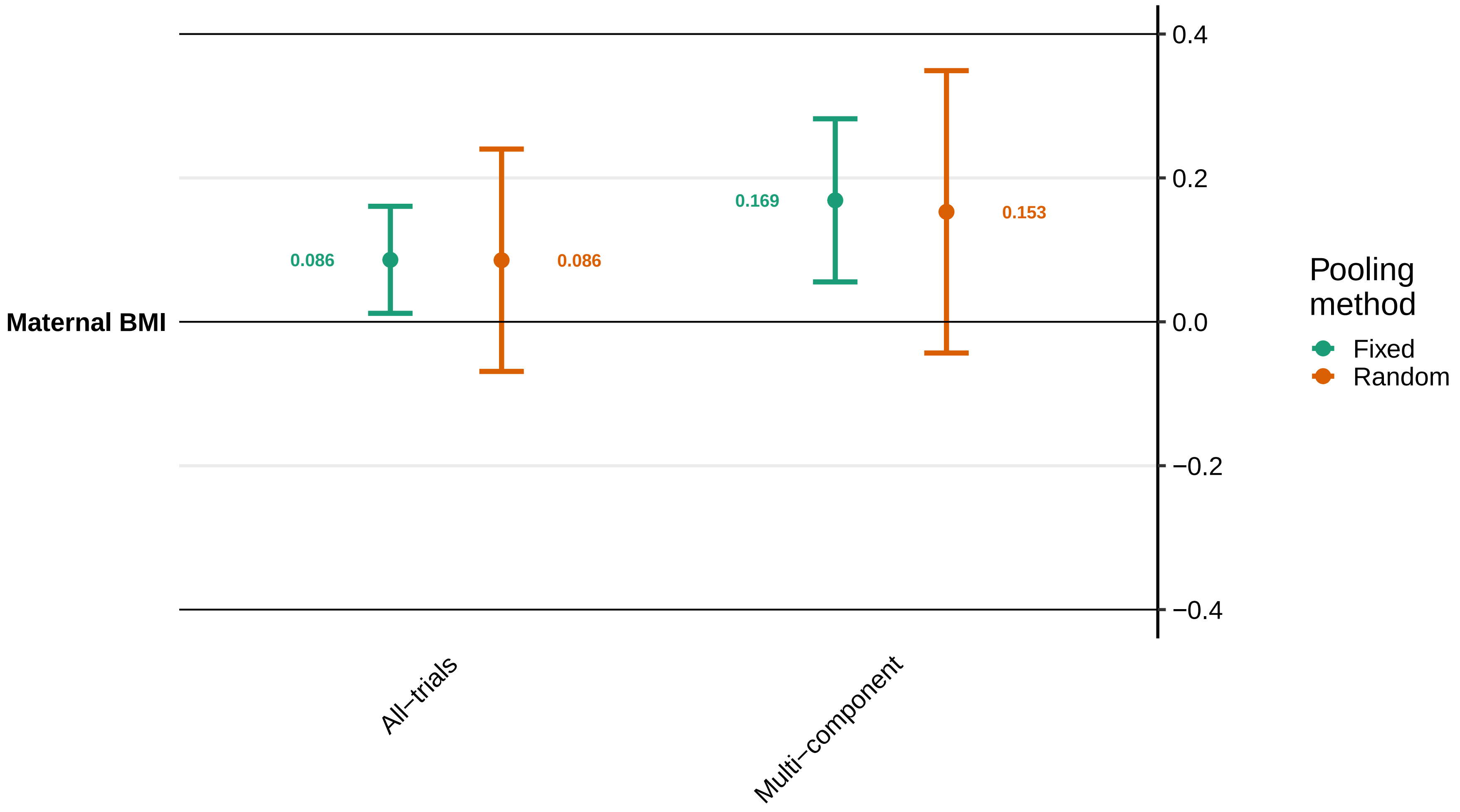

Supplemental figure 8J: Difference in iron deficiency anemia prevalence differences

#### 8J2: By child effect modifiers

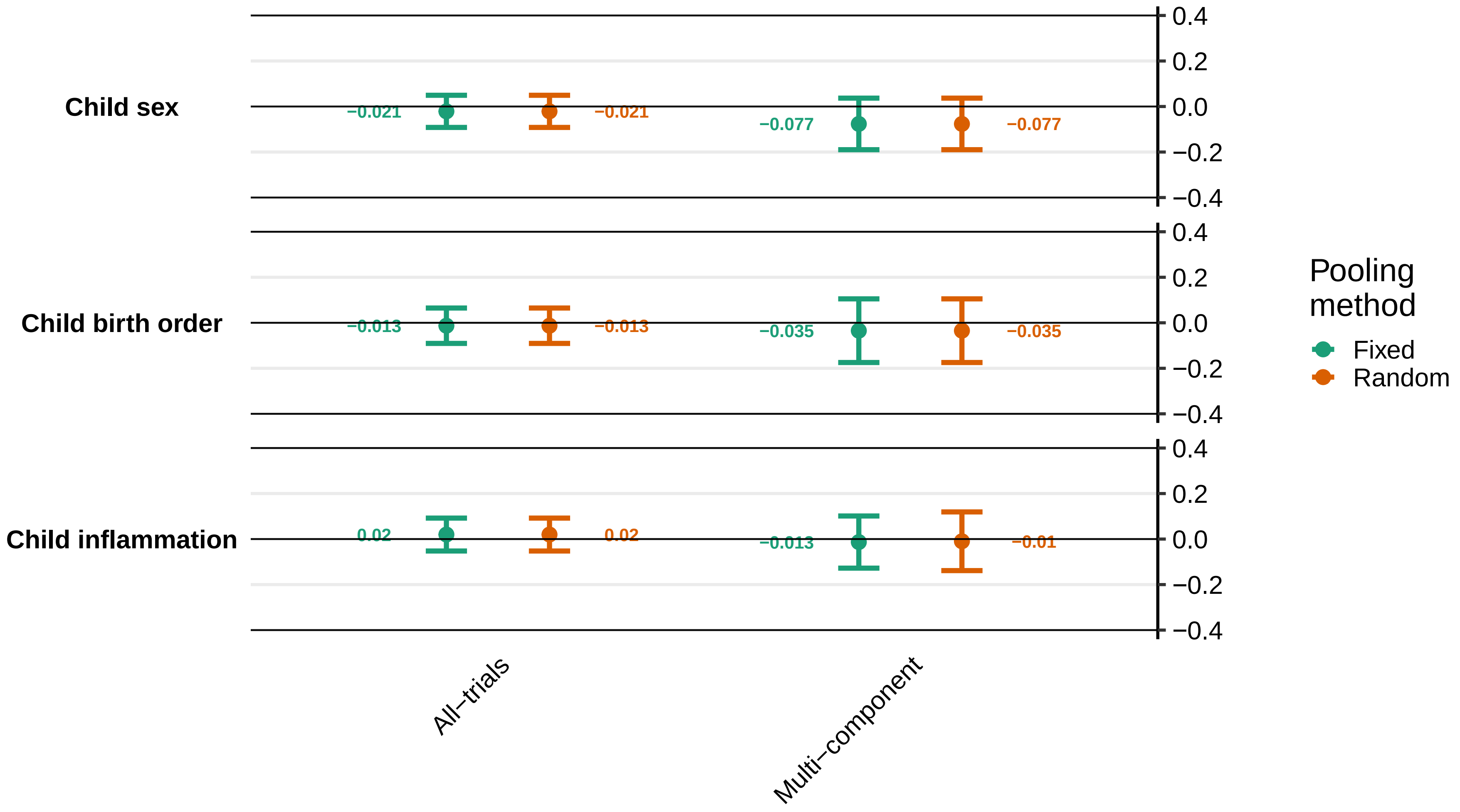

Supplemental figure 8J: Difference in iron deficiency anemia prevalence differences

8J3: By household effect modifiers

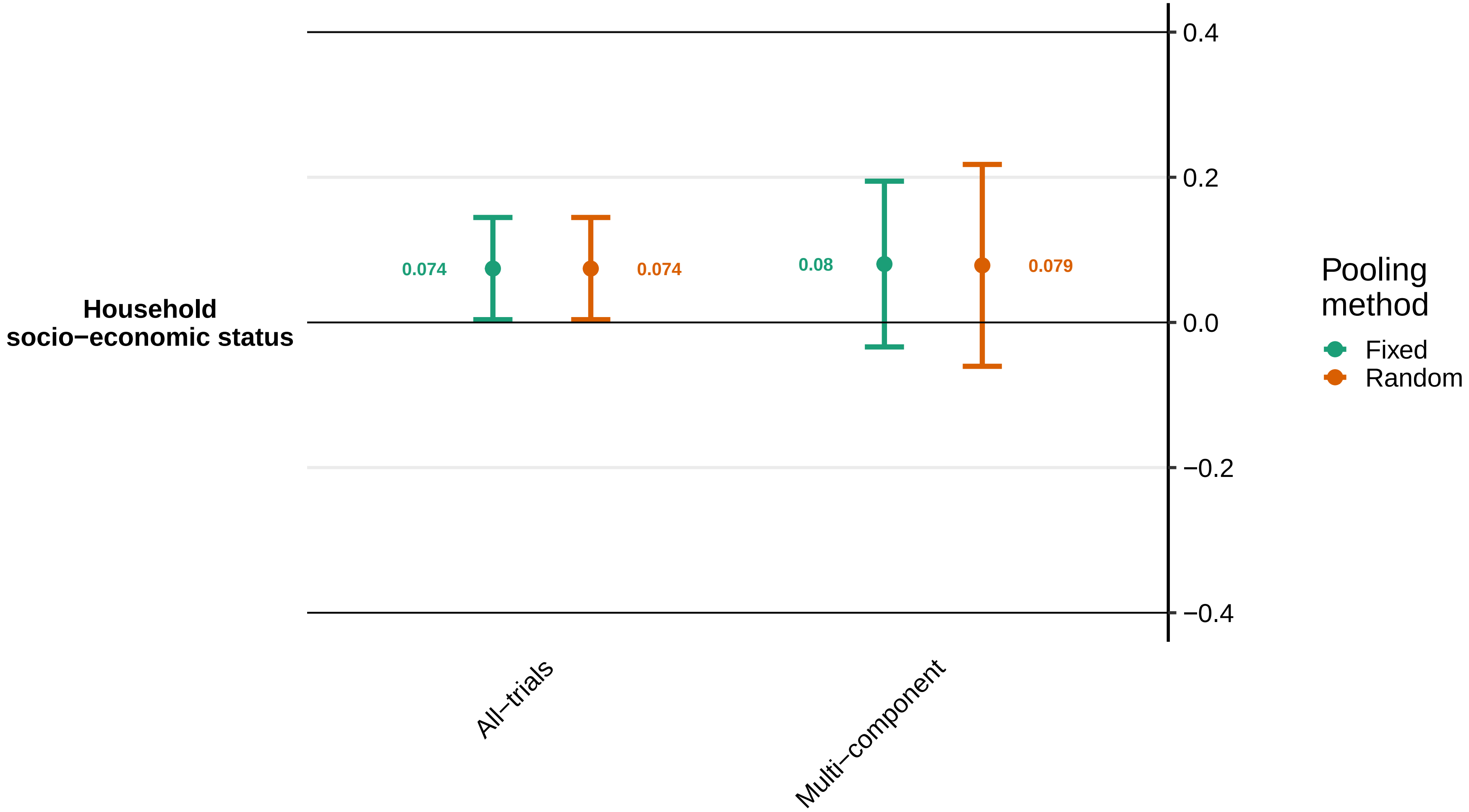

#### Supplemental figure 8K: Ratio of geometric mean ratios of soluble transferrin receptor concentration

##### 8K1: By maternal effect modifiers

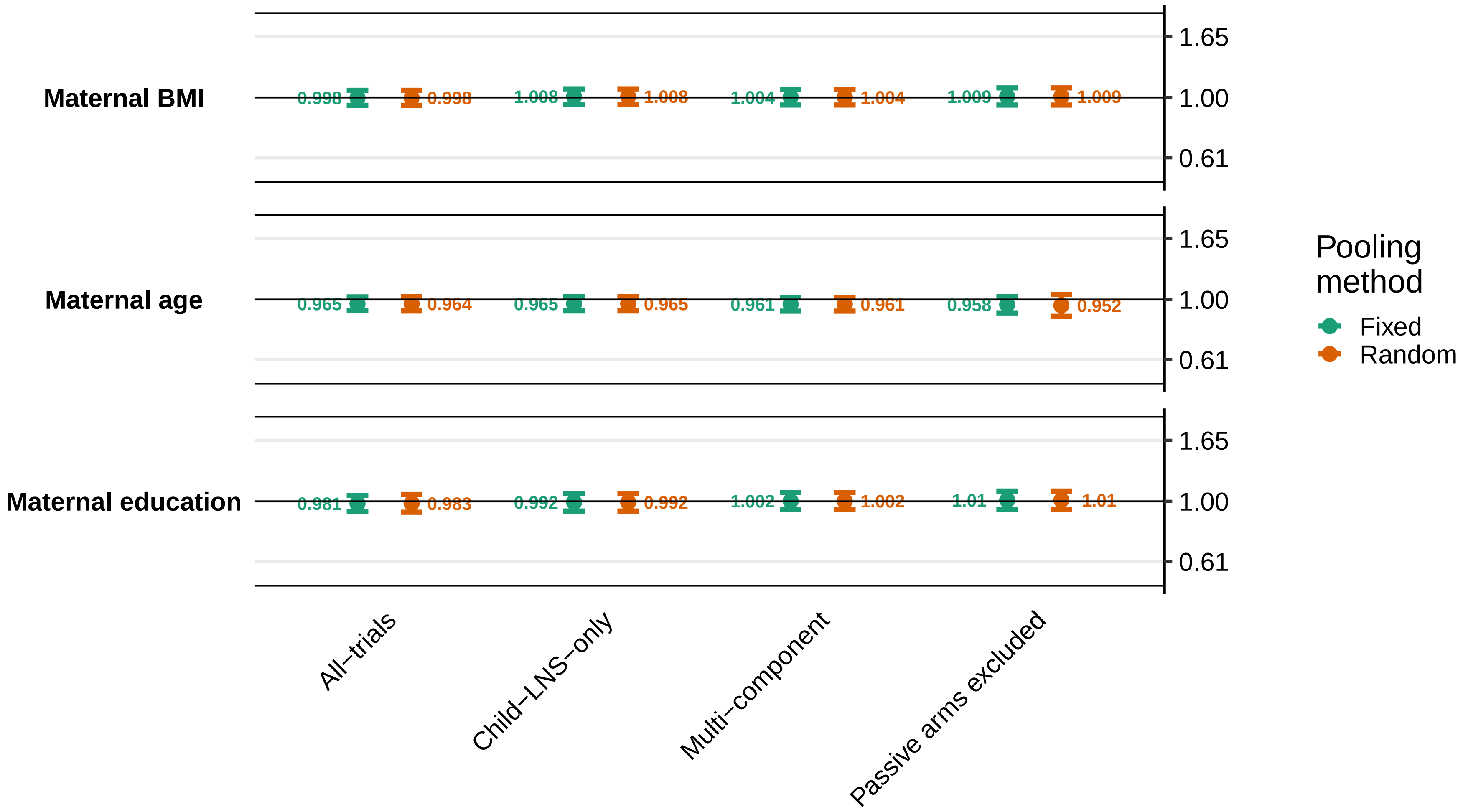

Supplemental figure 8K: Ratio of geometric mean ratios of soluble transferrin receptor concentration

#### 8K2: By child effect modifiers

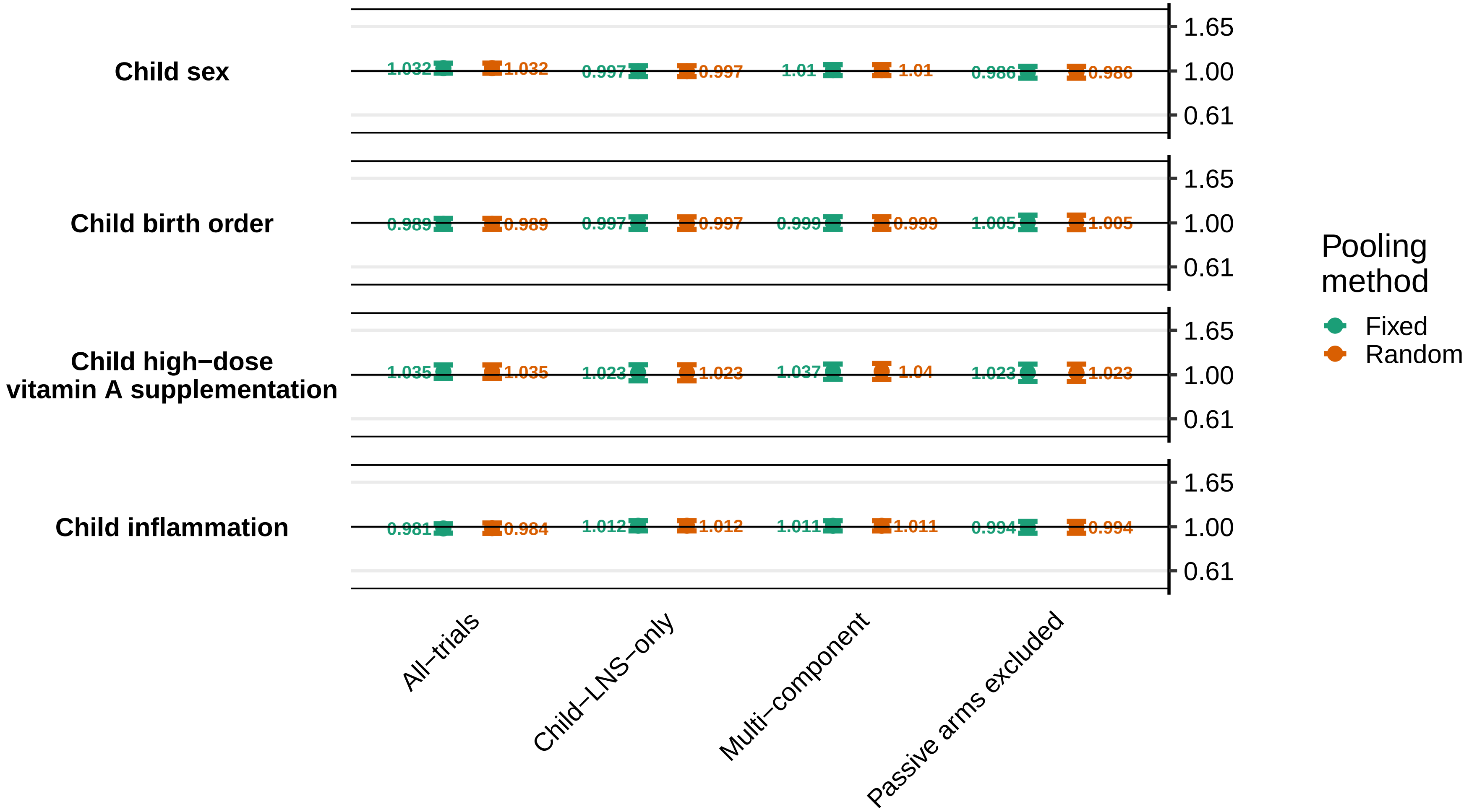

Supplemental figure 8K: Ratio of geometric mean ratios of soluble transferrin receptor concentration

#### 8K3: By household effect modifiers

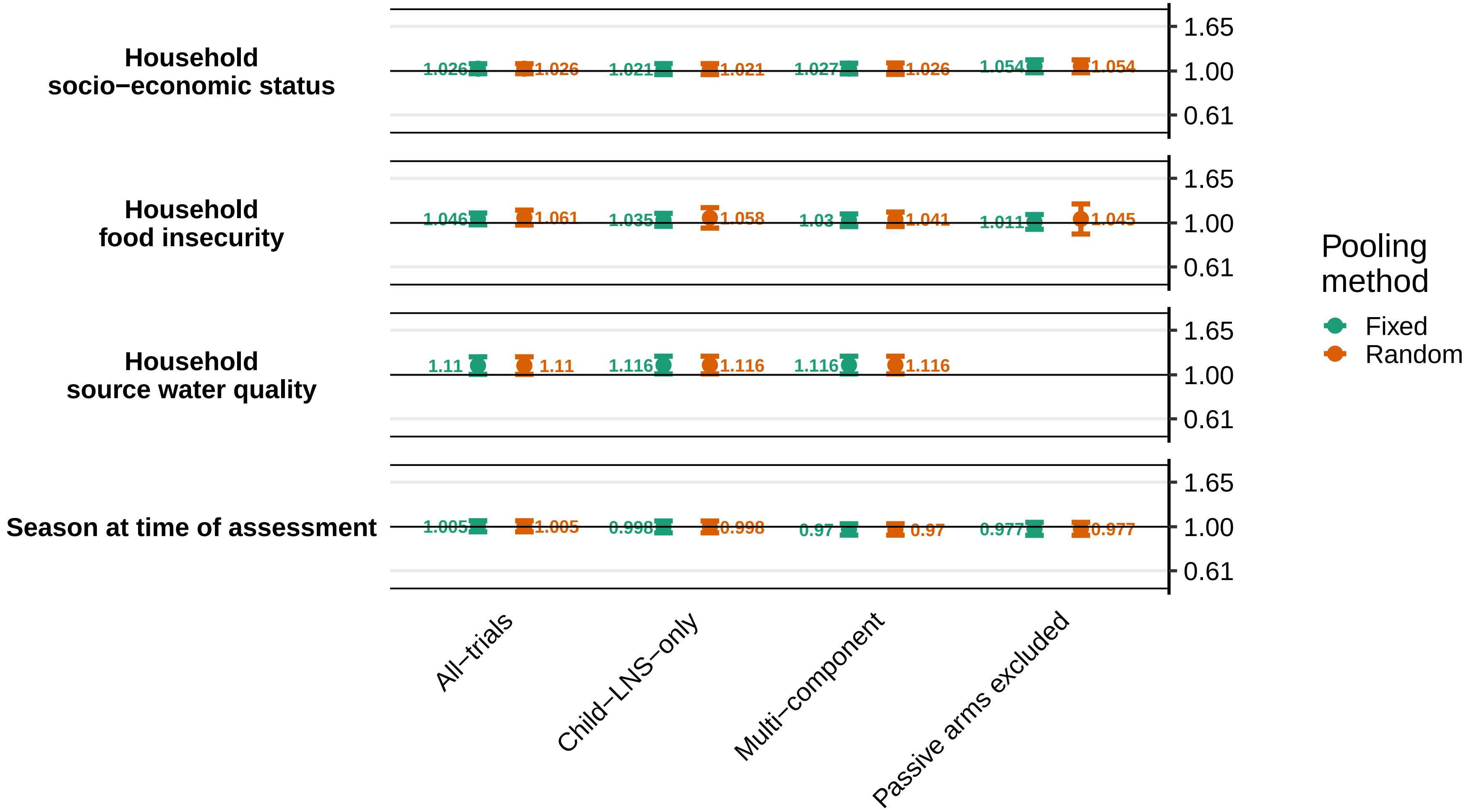

Supplemental figure 8L: Ratio of elevated soluble transferrin receptor prevalence ratios

8L1: By maternal effect modifiers

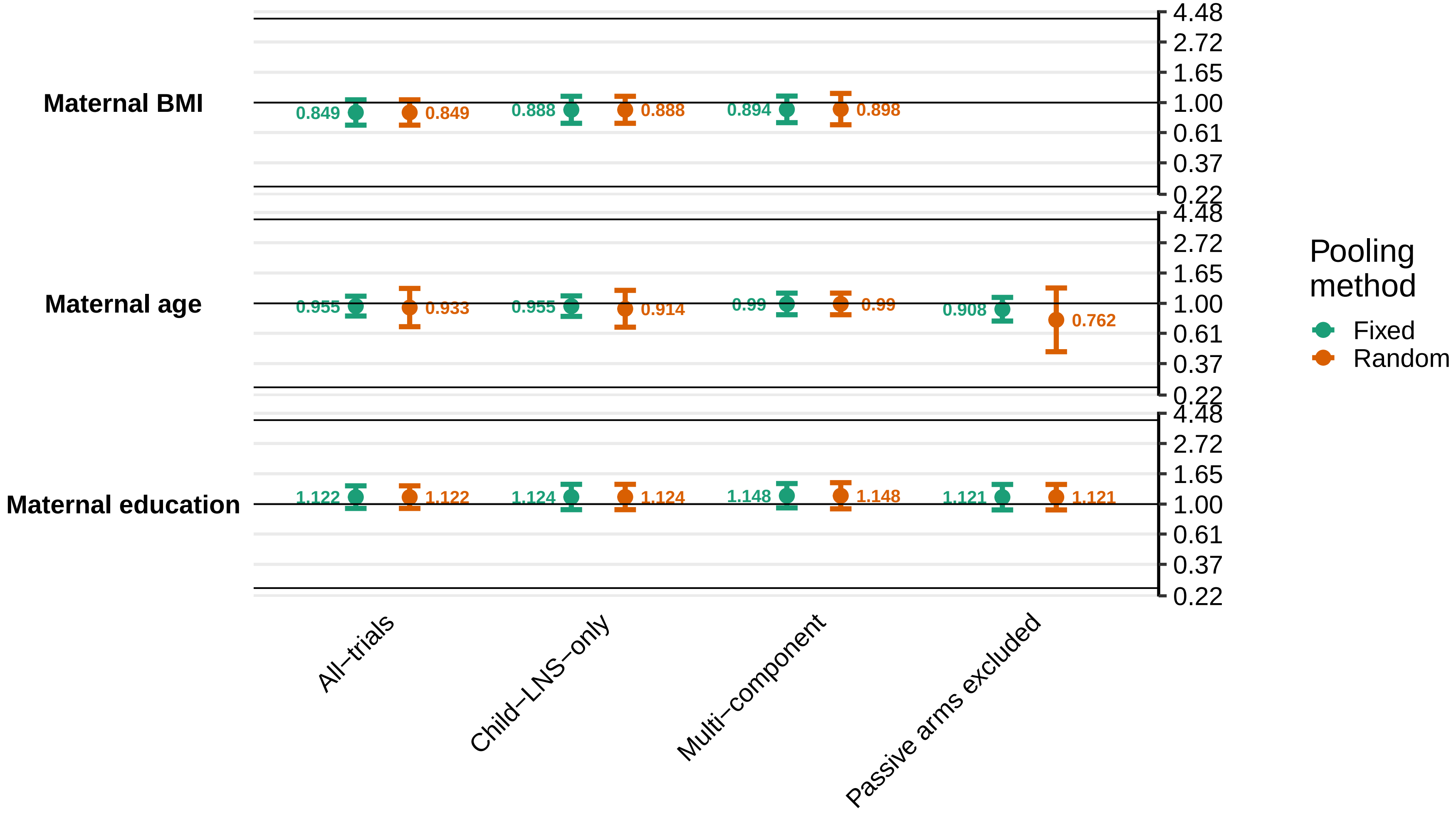

**Supplemental figure 8L: Ratio of elevated soluble transferrin receptor prevalence ratios**

#### 8L2: By child effect modifiers

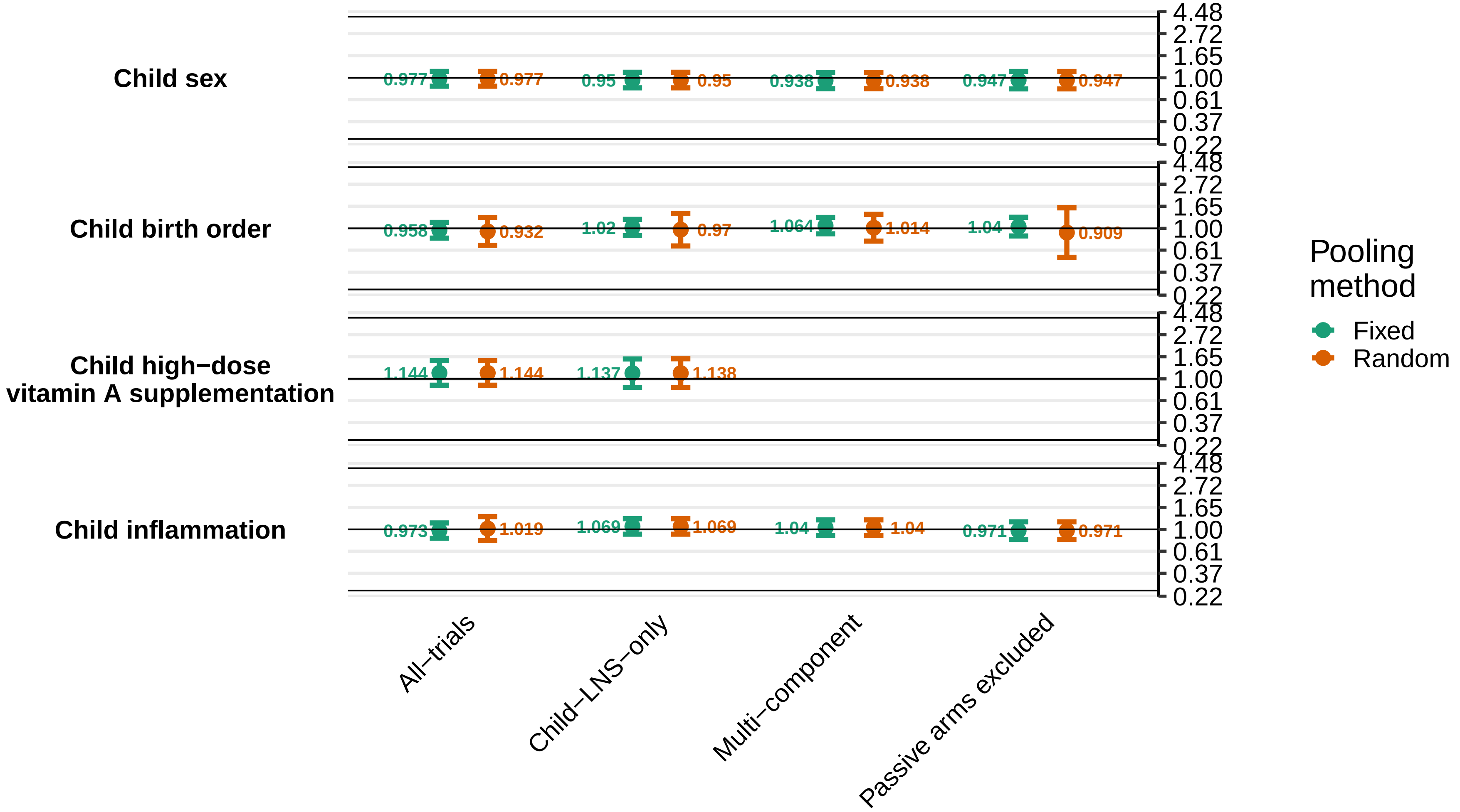

Supplemental figure 8L: Ratio of elevated soluble transferrin receptor prevalence ratios

#### 8L3: By household effect modifiers

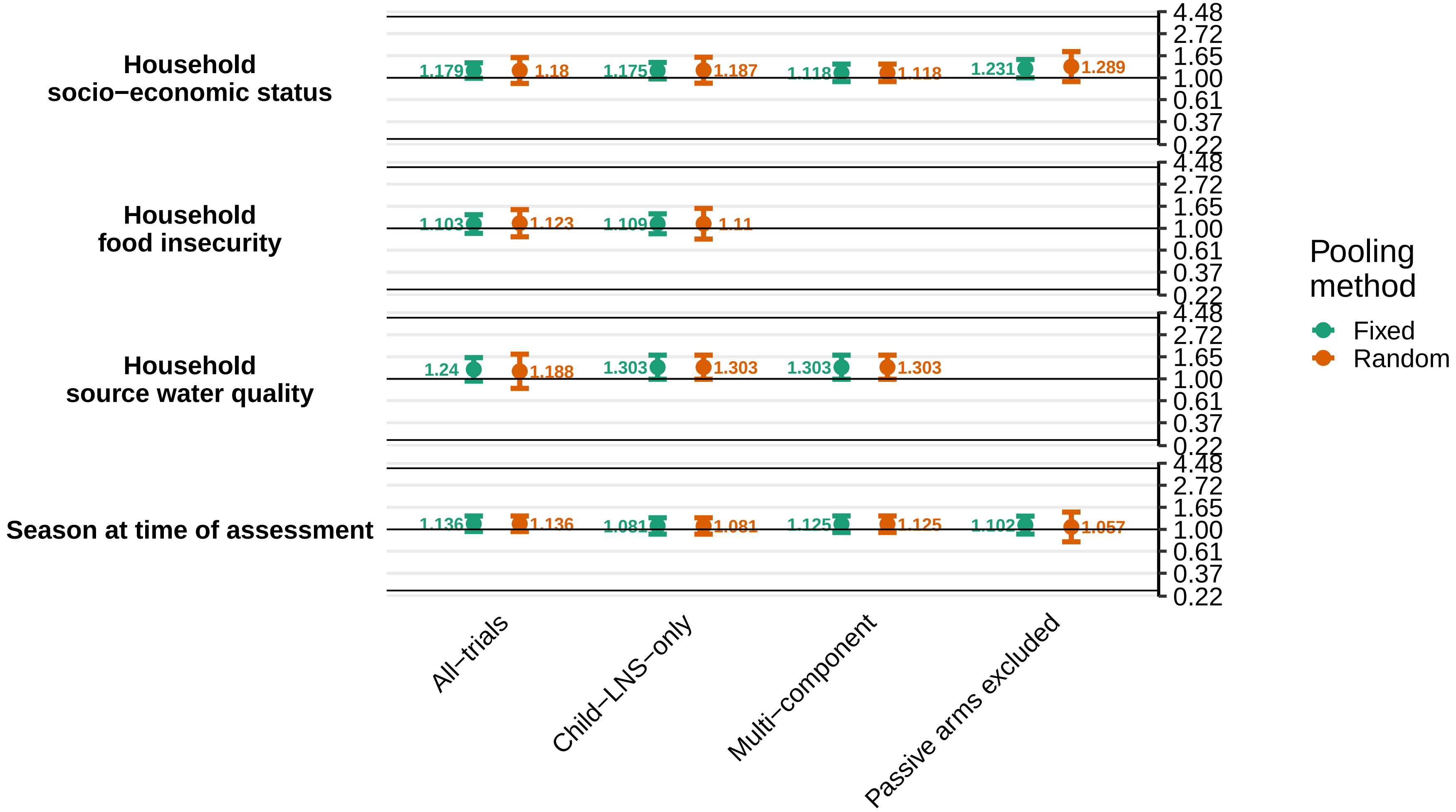

**Supplemental figure 8M: Difference in elevated soluble transferrin receptor prevalence differences**

**8M1: By maternal effect modifiers**

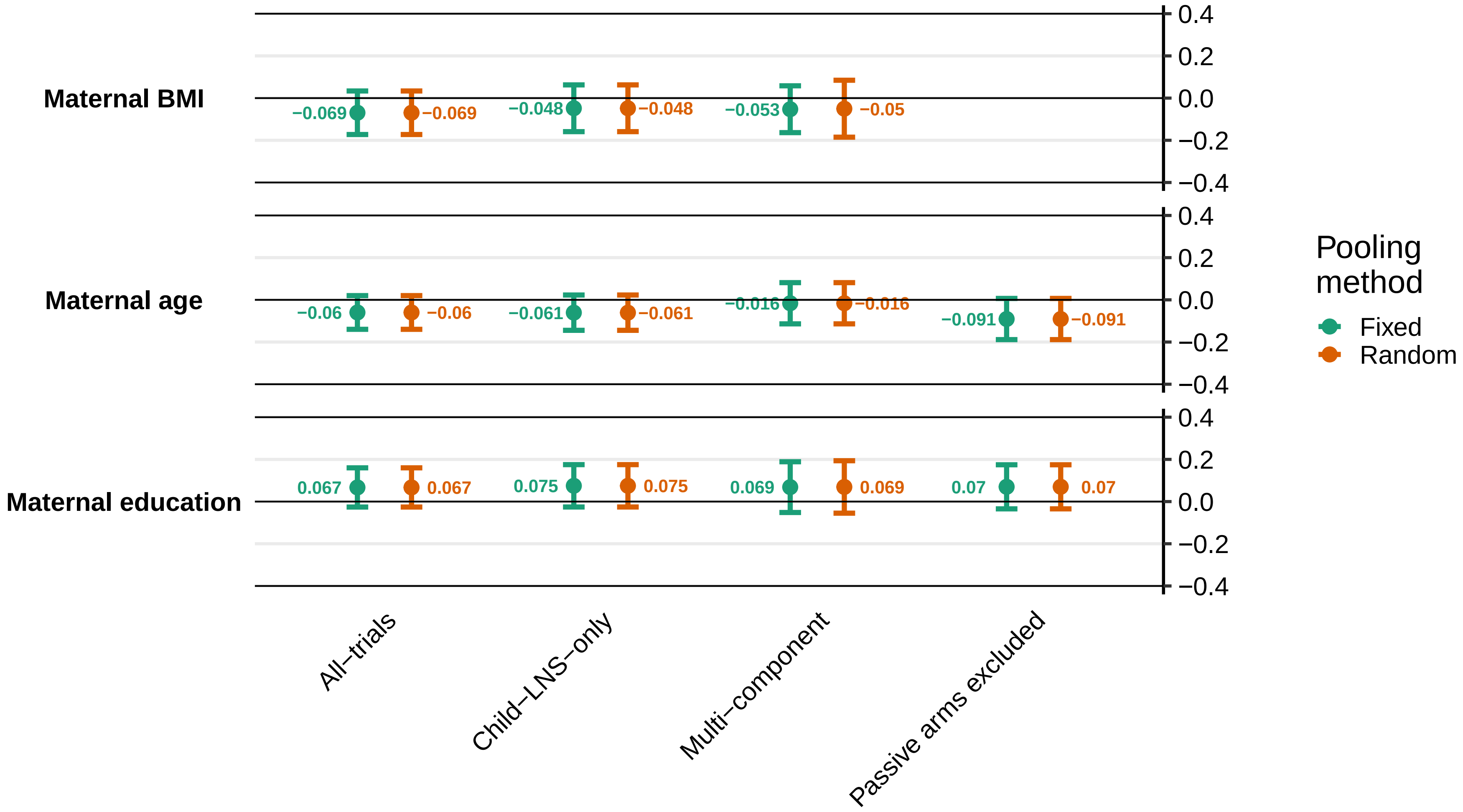

Supplemental figure 8M: Difference in elevated soluble transferrin receptor prevalence differences

#### 8M2: By child effect modifiers

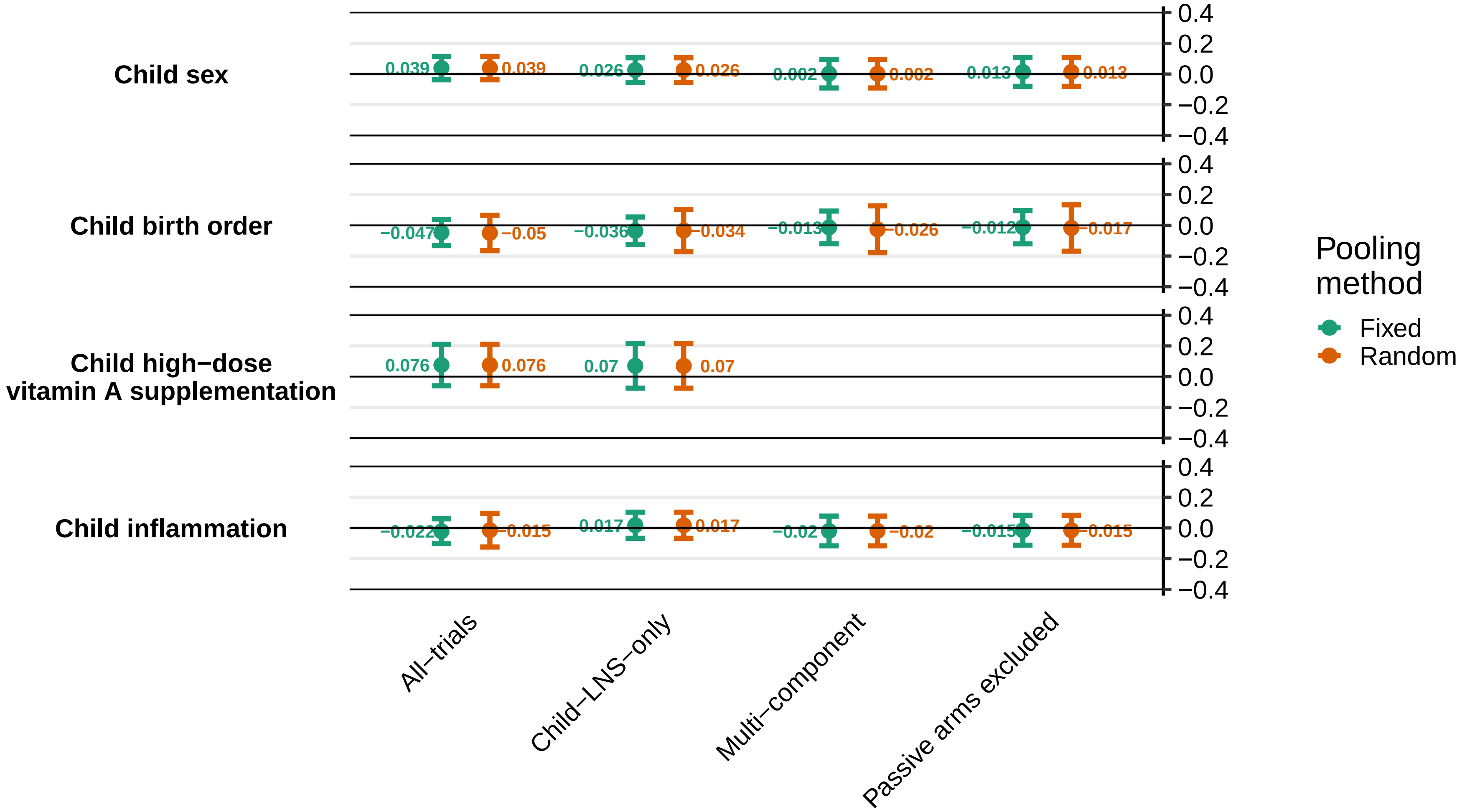

Supplemental figure 8M: Difference in elevated soluble transferrin receptor prevalence differences

8M3: By household effect modifiers

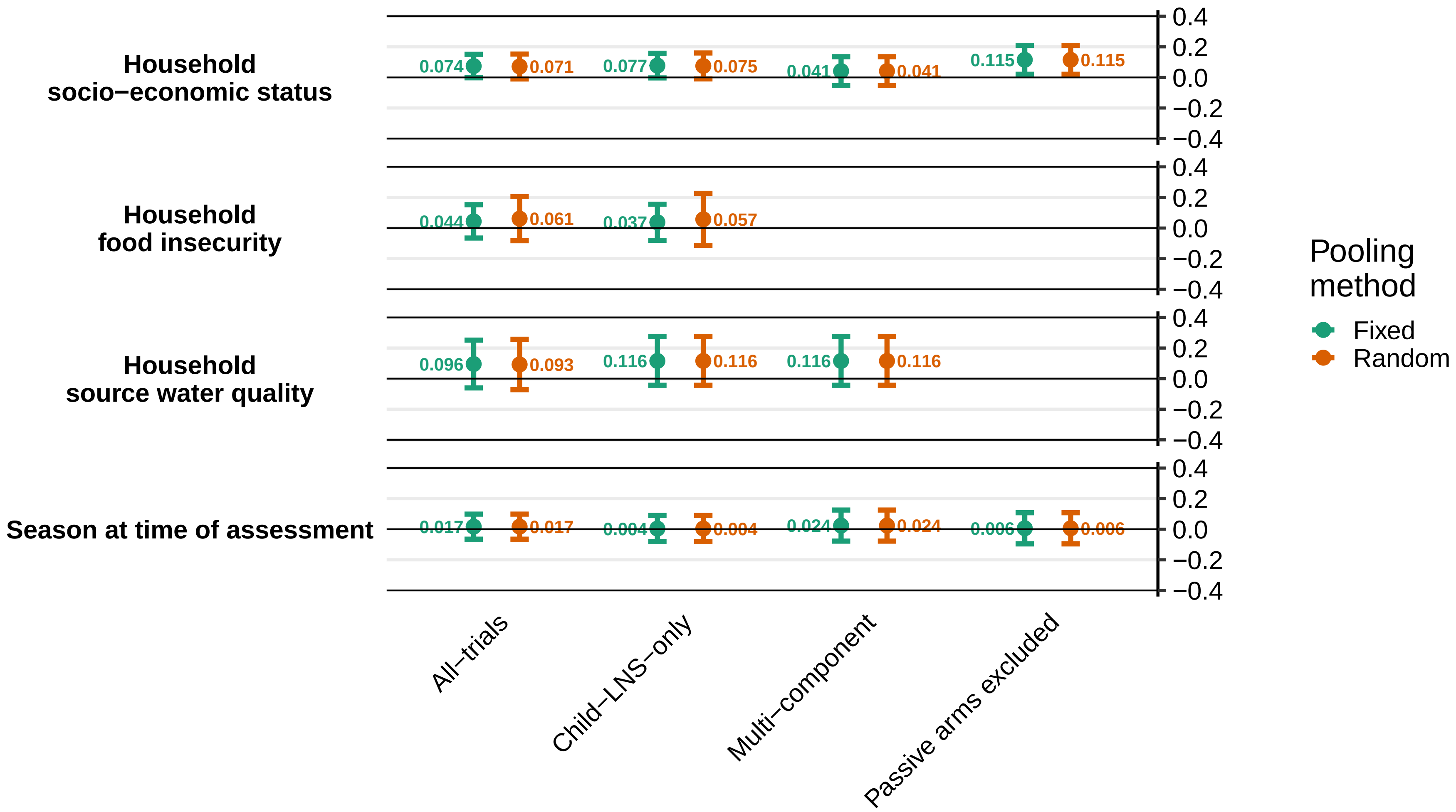

#### Supplemental figure 8N: Ratio of geometric mean ratios of zinc protoporphyrin concentration

##### 8N1: By maternal effect modifiers

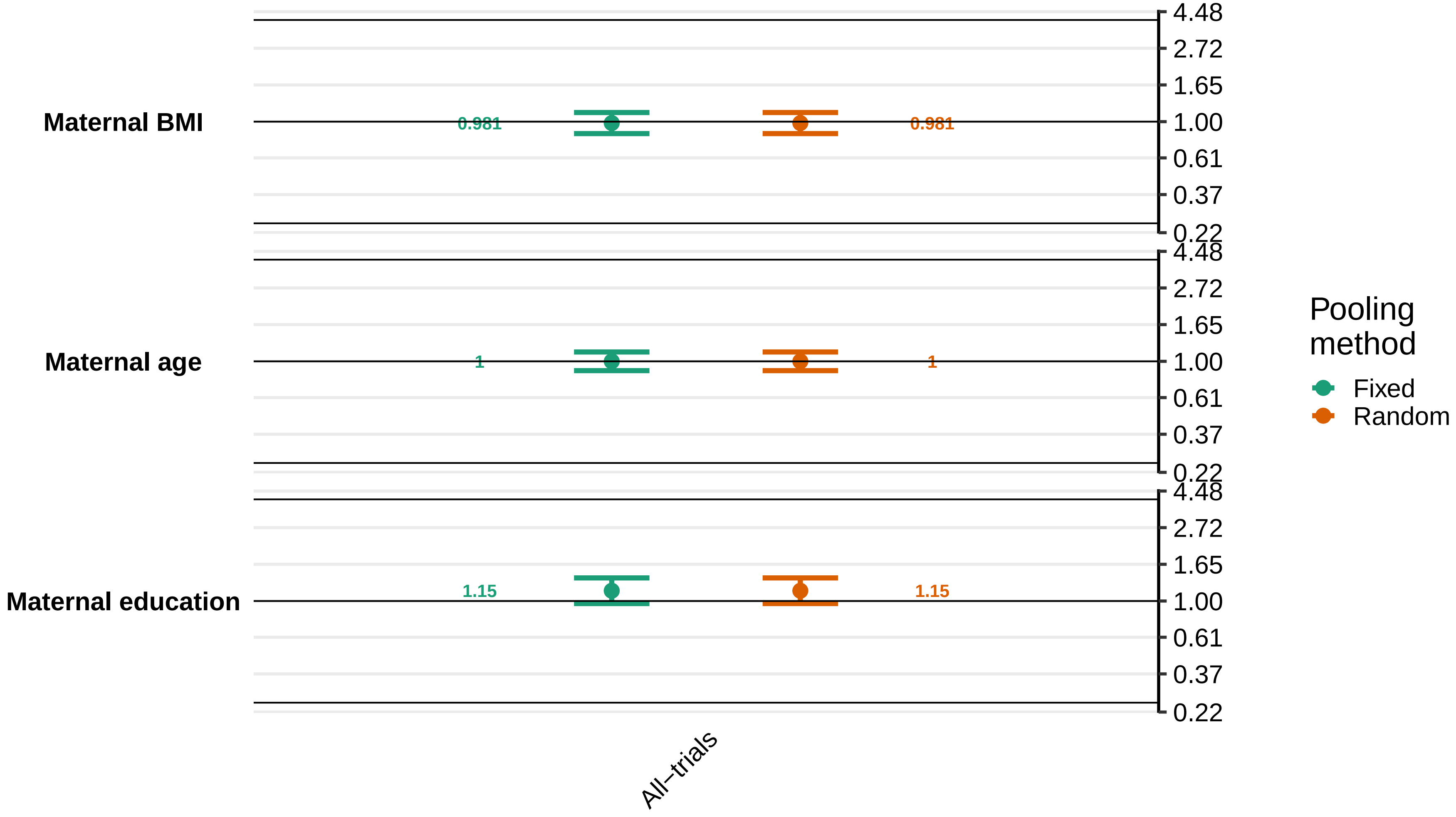

**Supplemental figure 8N: Ratio of geometric mean ratios of zinc protoporphyrin concentration**

#### 8N2: By child effect modifiers

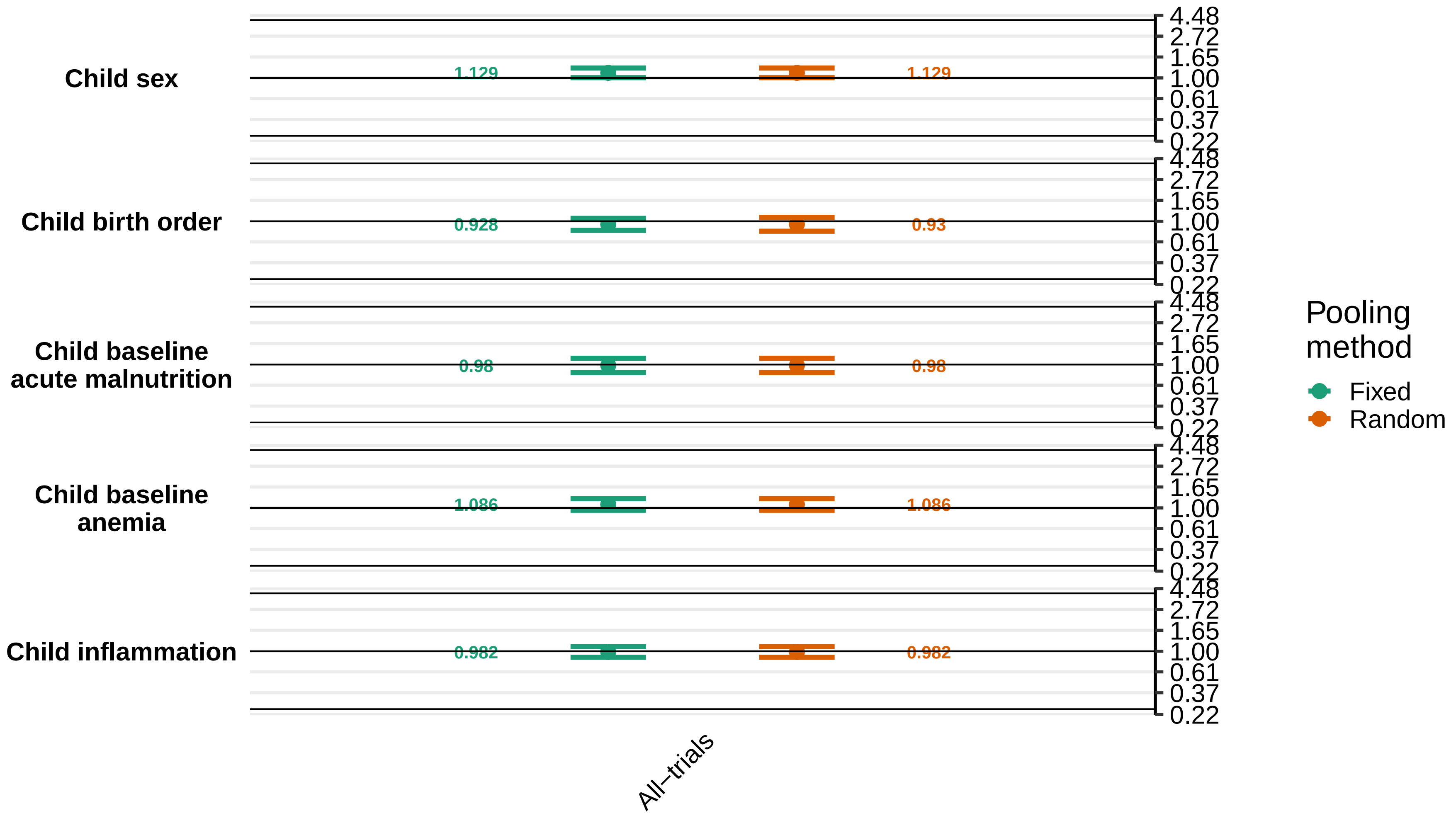

**Supplemental figure 8N: Ratio of geometric mean ratios of zinc protoporphyrin concentration**

#### 8N3: By household effect modifiers

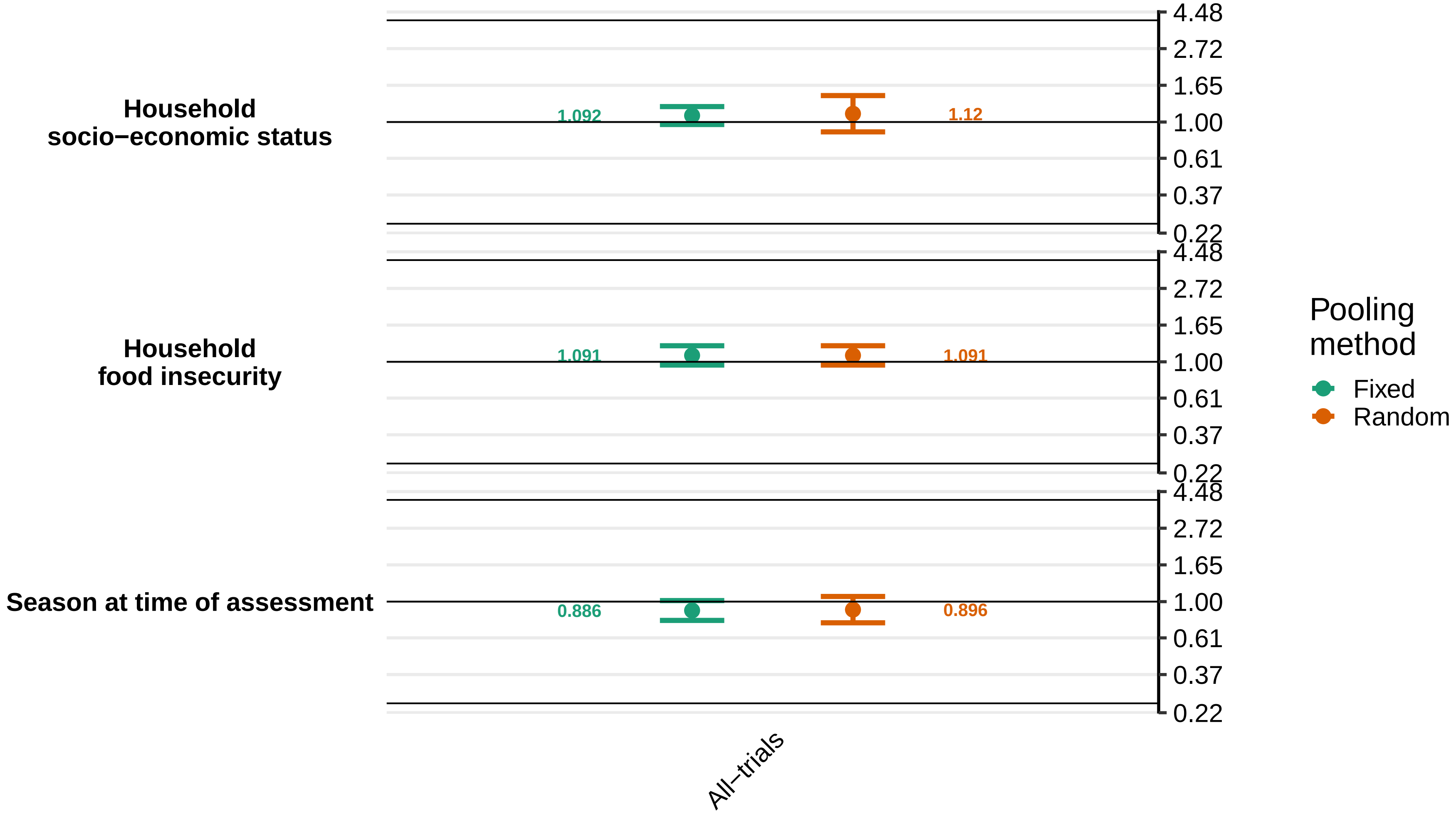

#### Supplemental figure 8O: Ratio of elevated zinc protoporphyrin prevalence ratios

##### 8O1: By maternal effect modifiers

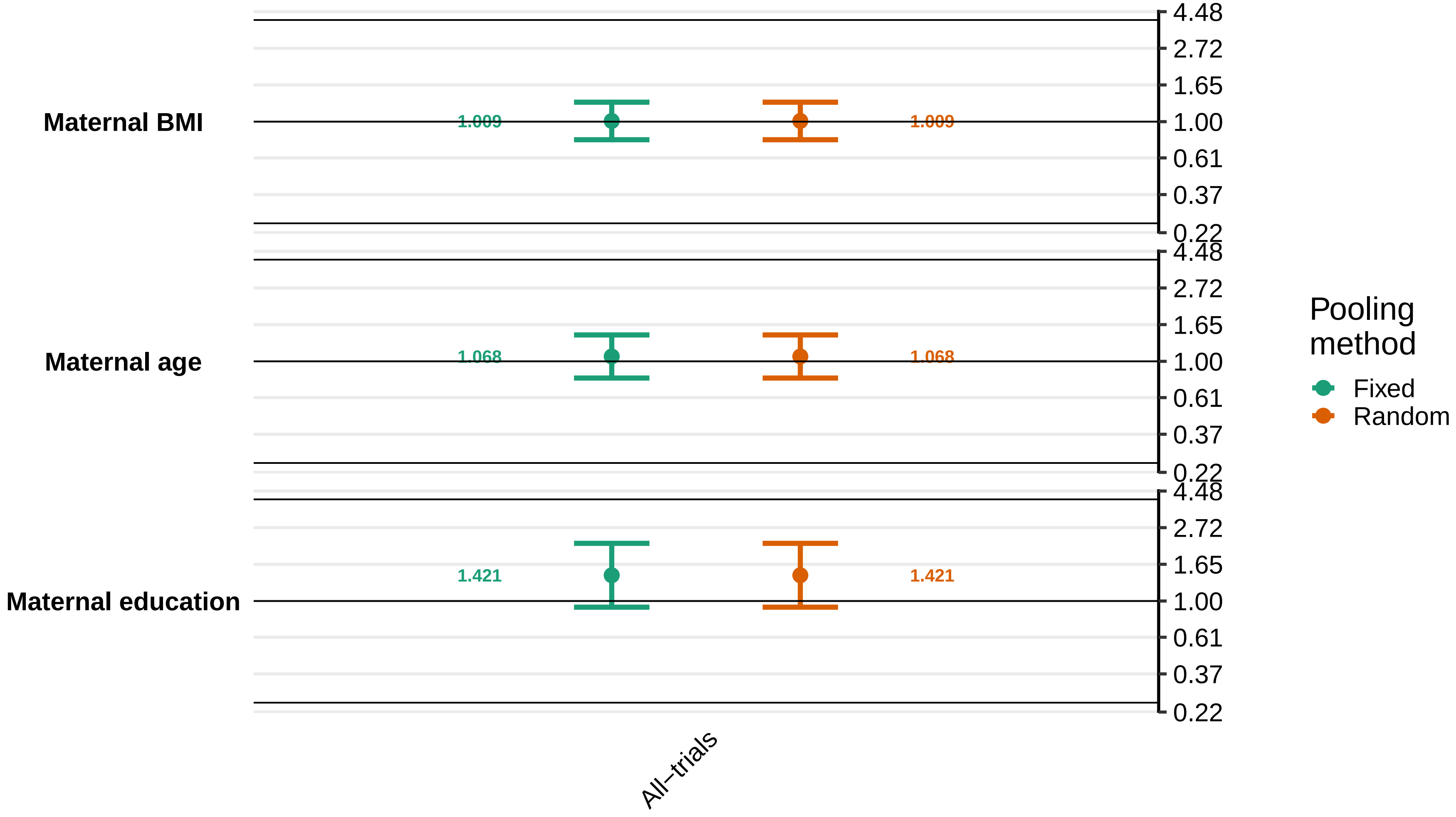

Supplemental figure 8O: Ratio of elevated zinc protoporphyrin prevalence ratios

#### 8O2: By child effect modifiers

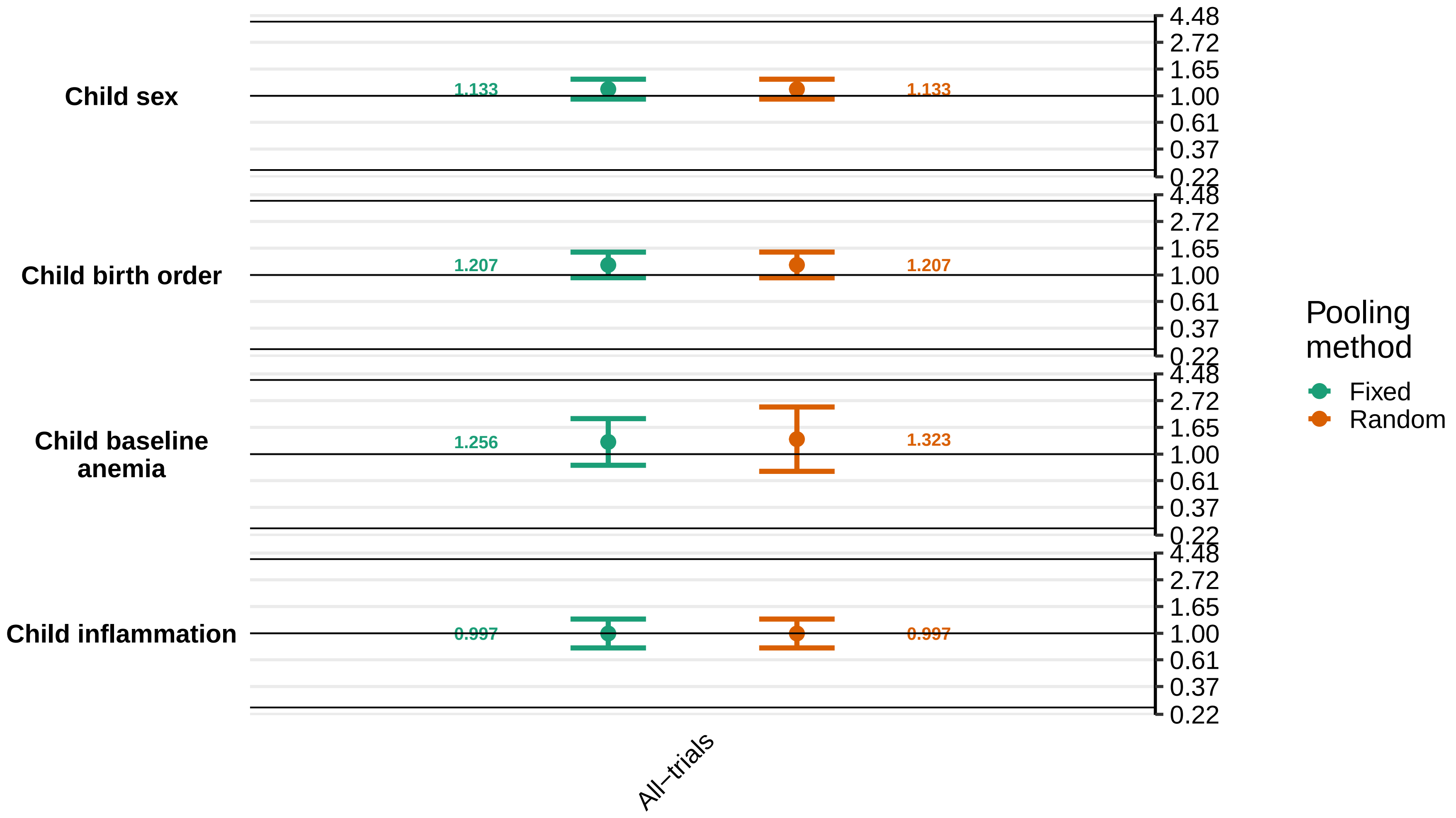

Supplemental figure 8O: Ratio of elevated zinc protoporphyrin prevalence ratios

#### 8O3: By household effect modifiers

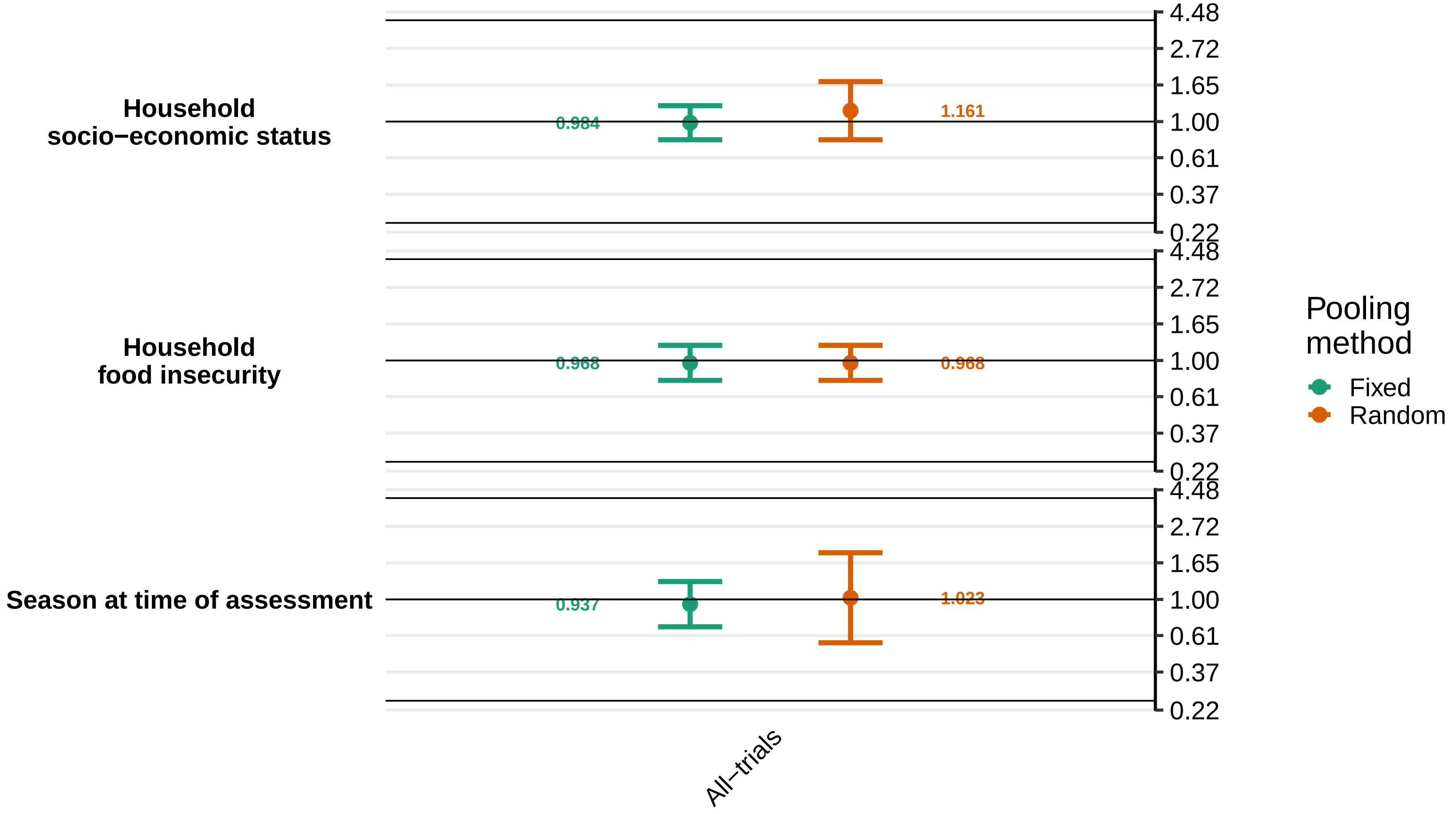

#### Supplemental figure 8P: Difference in elevated zinc protoporphyrin prevalence differences

##### 8P1: By maternal effect modifiers

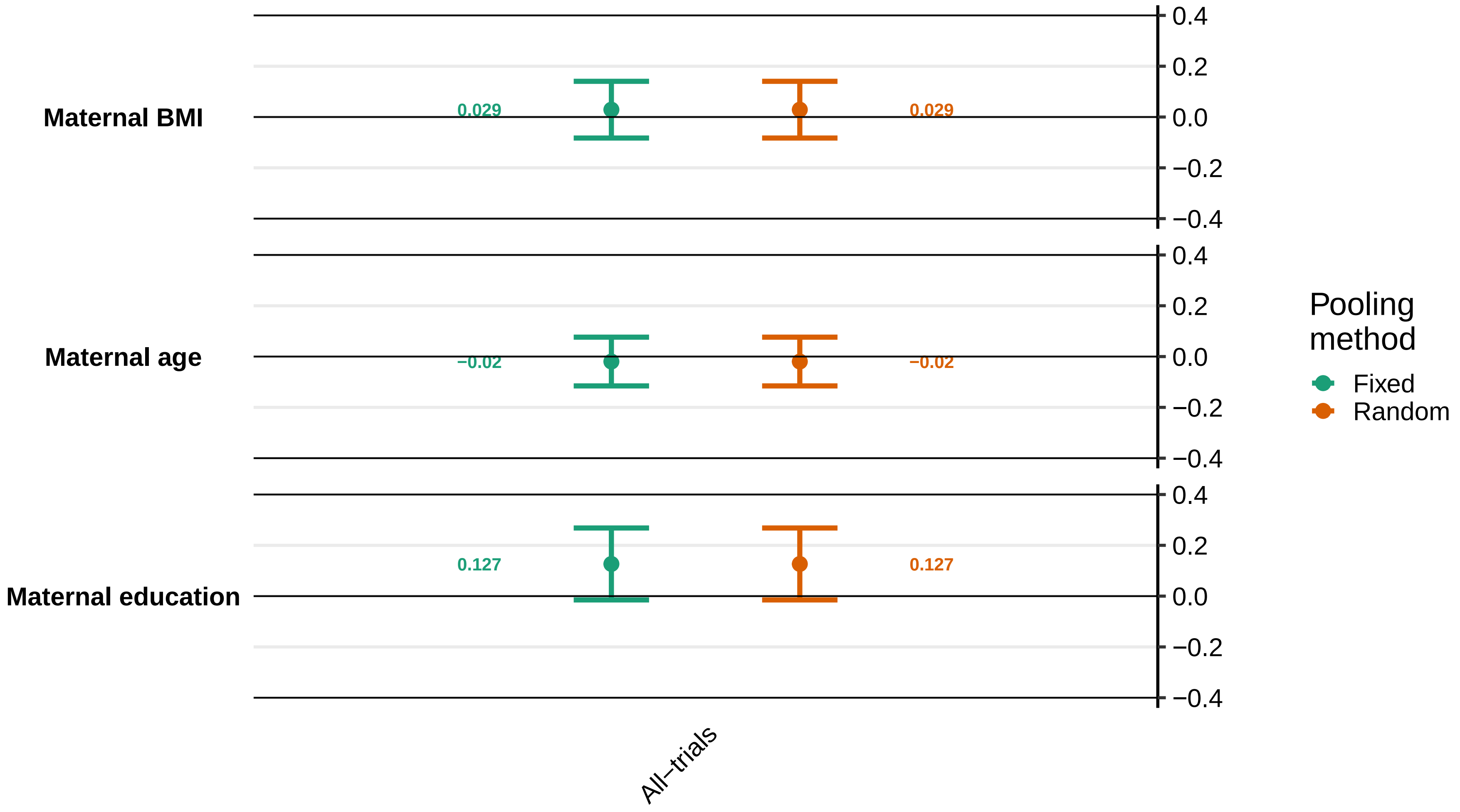

Supplemental figure 8P: Difference in elevated zinc protoporphyrin prevalence differences

8P2: By child effect modifiers

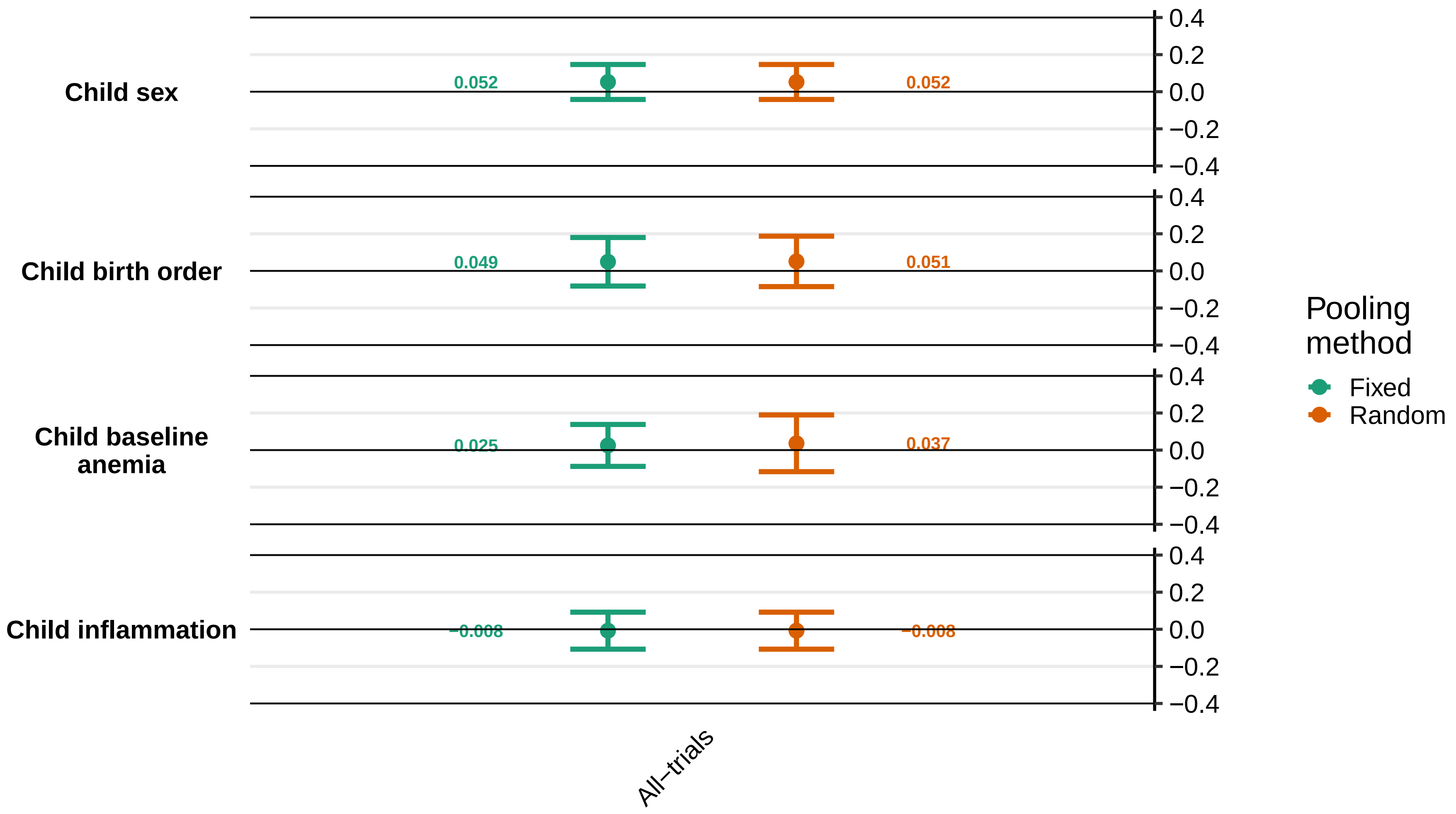

Supplemental figure 8P: Difference in elevated zinc protoporphyrin prevalence differences

#### 8P3: By household effect modifiers

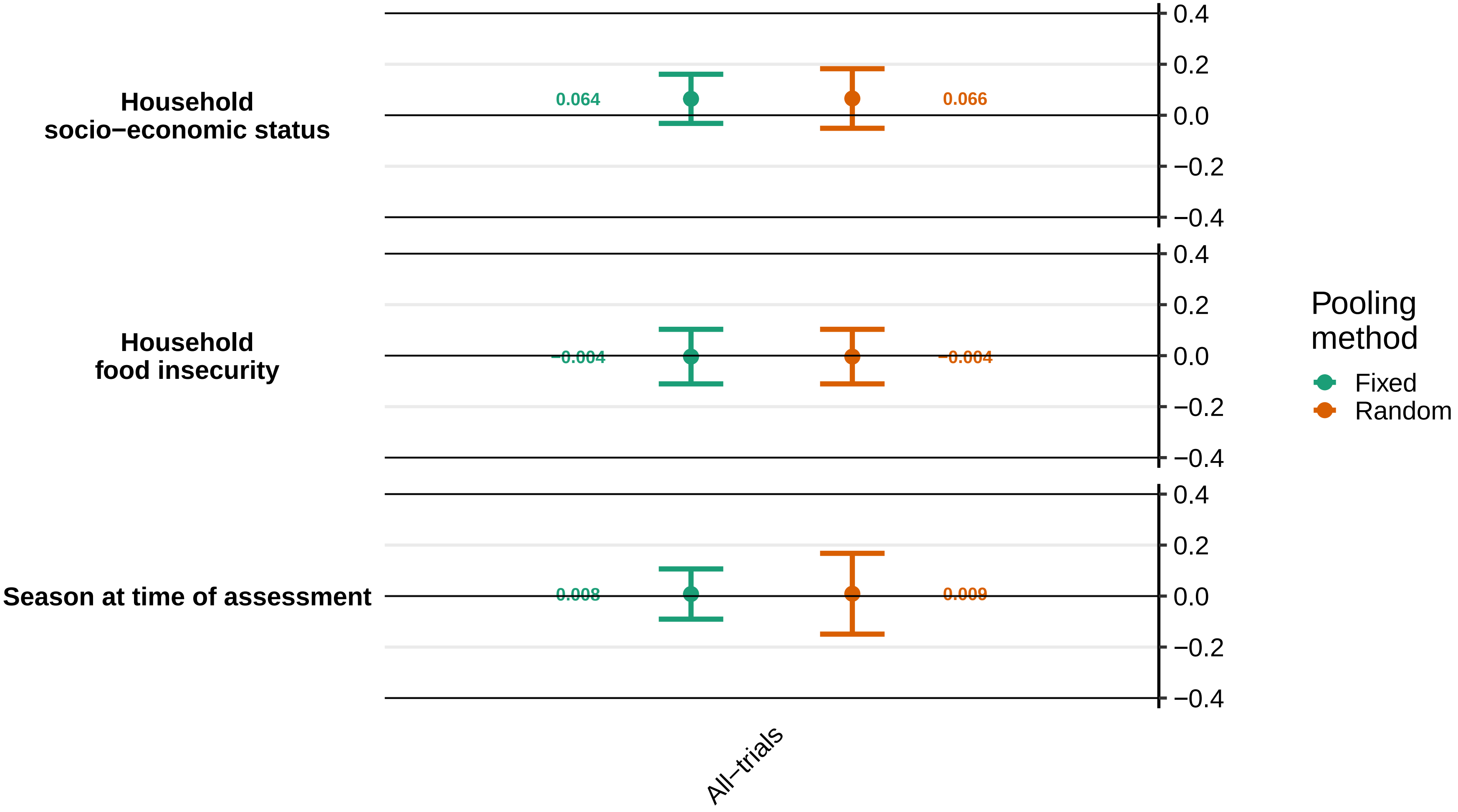

#### Supplemental figure 8Q: Ratio of geometric mean ratios of plasma zinc concentration

##### 8Q1: By maternal effect modifiers

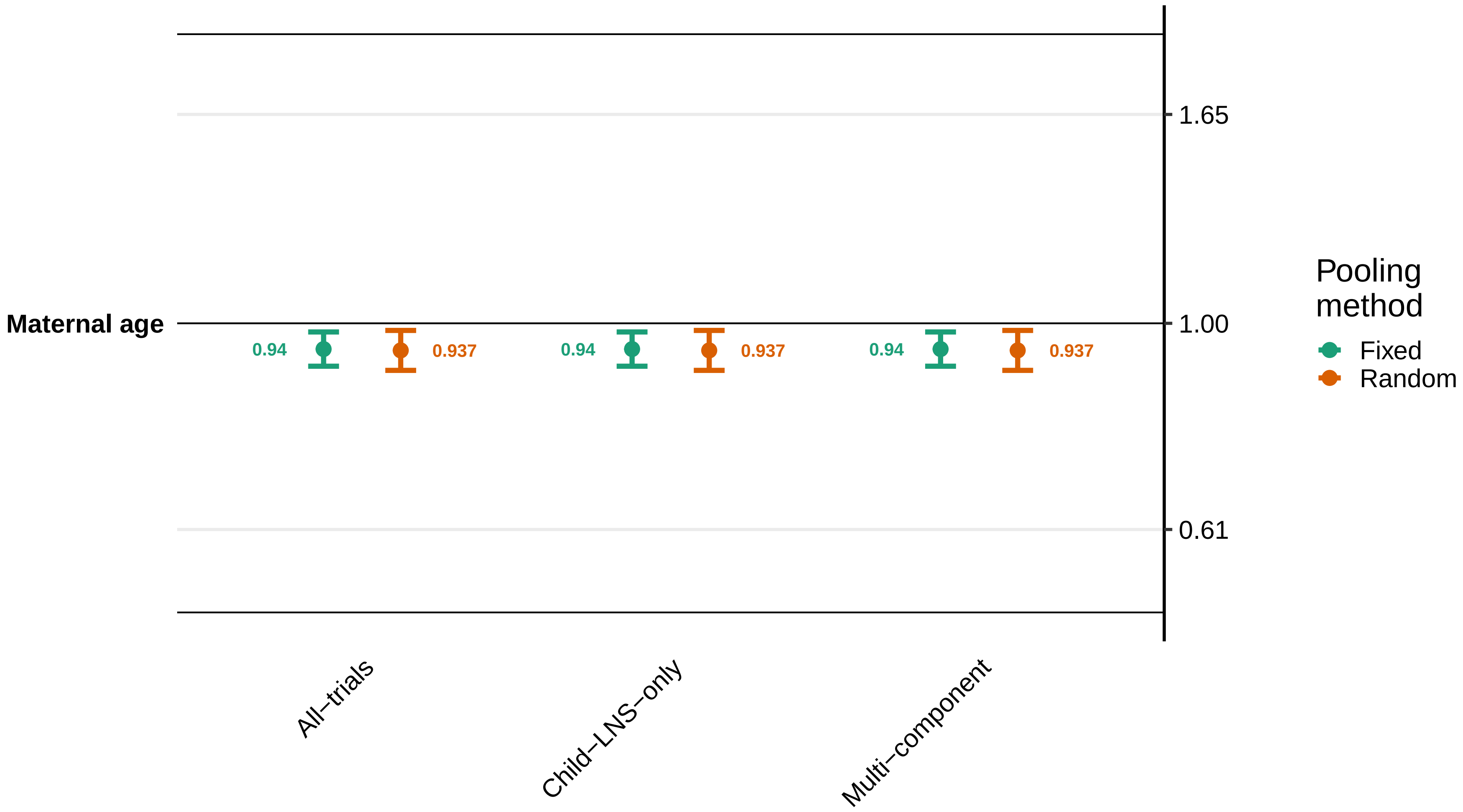

Supplemental figure 8Q: Ratio of geometric mean ratios of plasma zinc concentration

#### 8Q2: By child effect modifiers

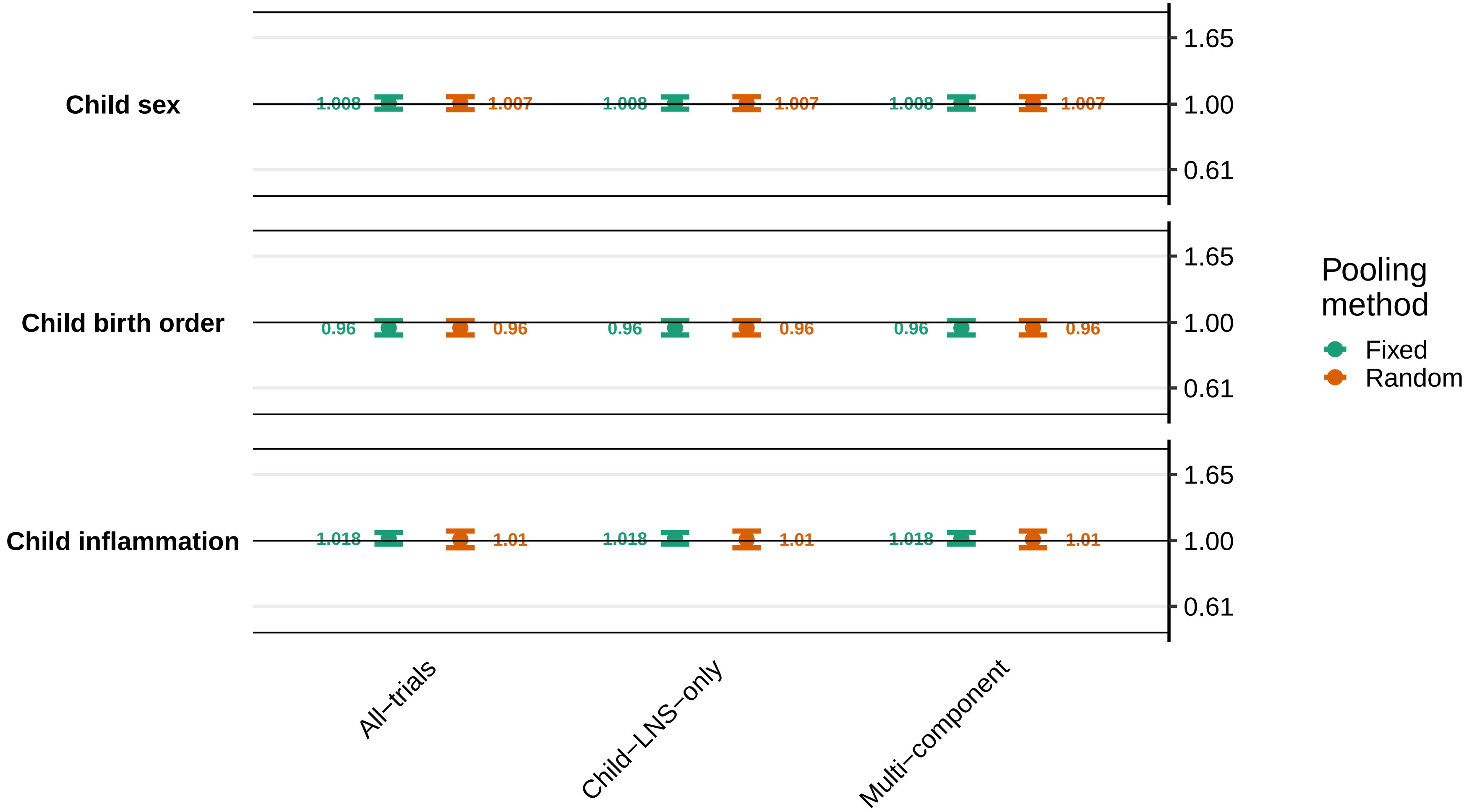

Supplemental figure 8Q: Ratio of geometric mean ratios of plasma zinc concentration

#### 8Q3: By household effect modifiers

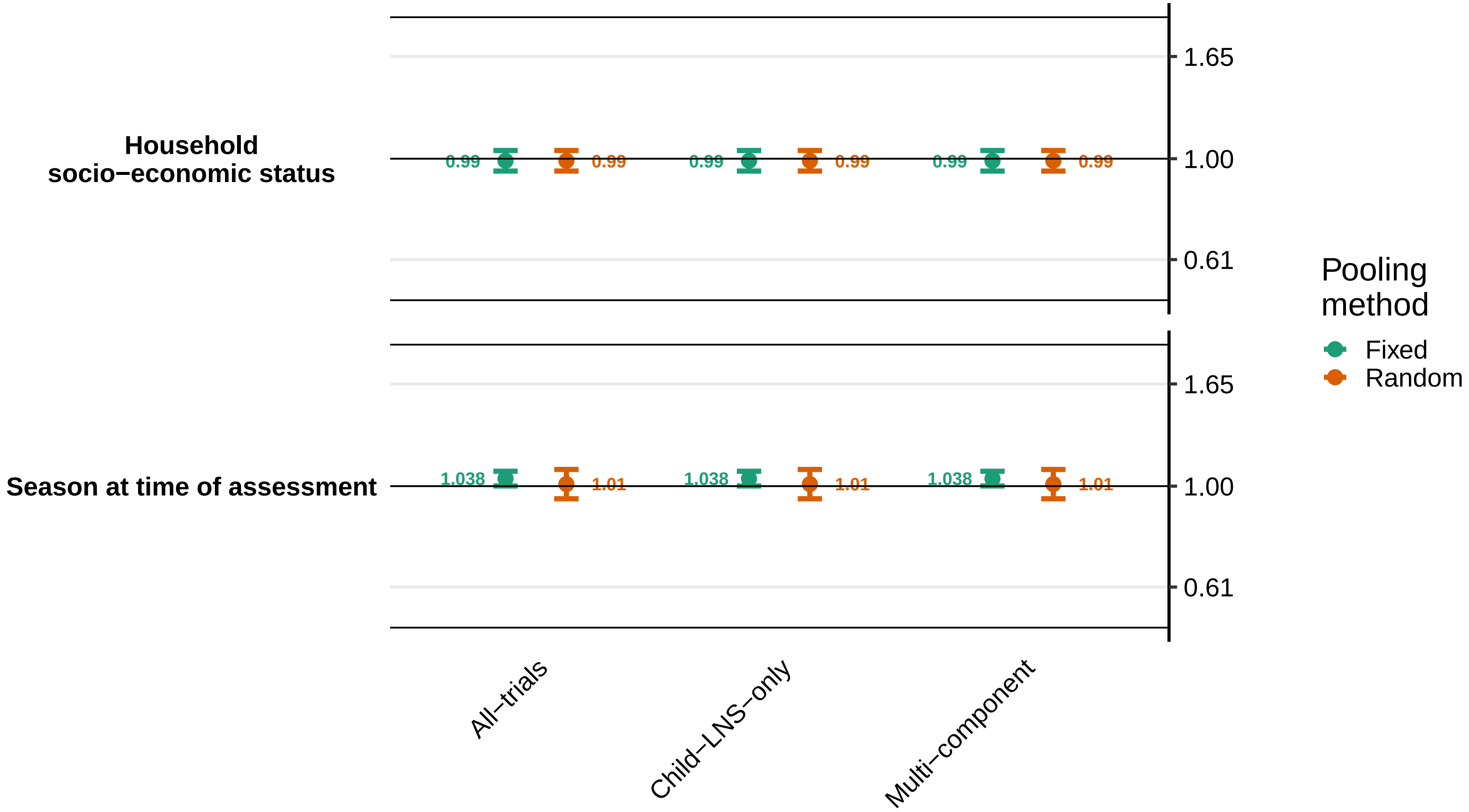

#### Supplemental figure 8R: Ratio of geometric mean ratios of retinol concentration

##### 8R1: By maternal effect modifiers

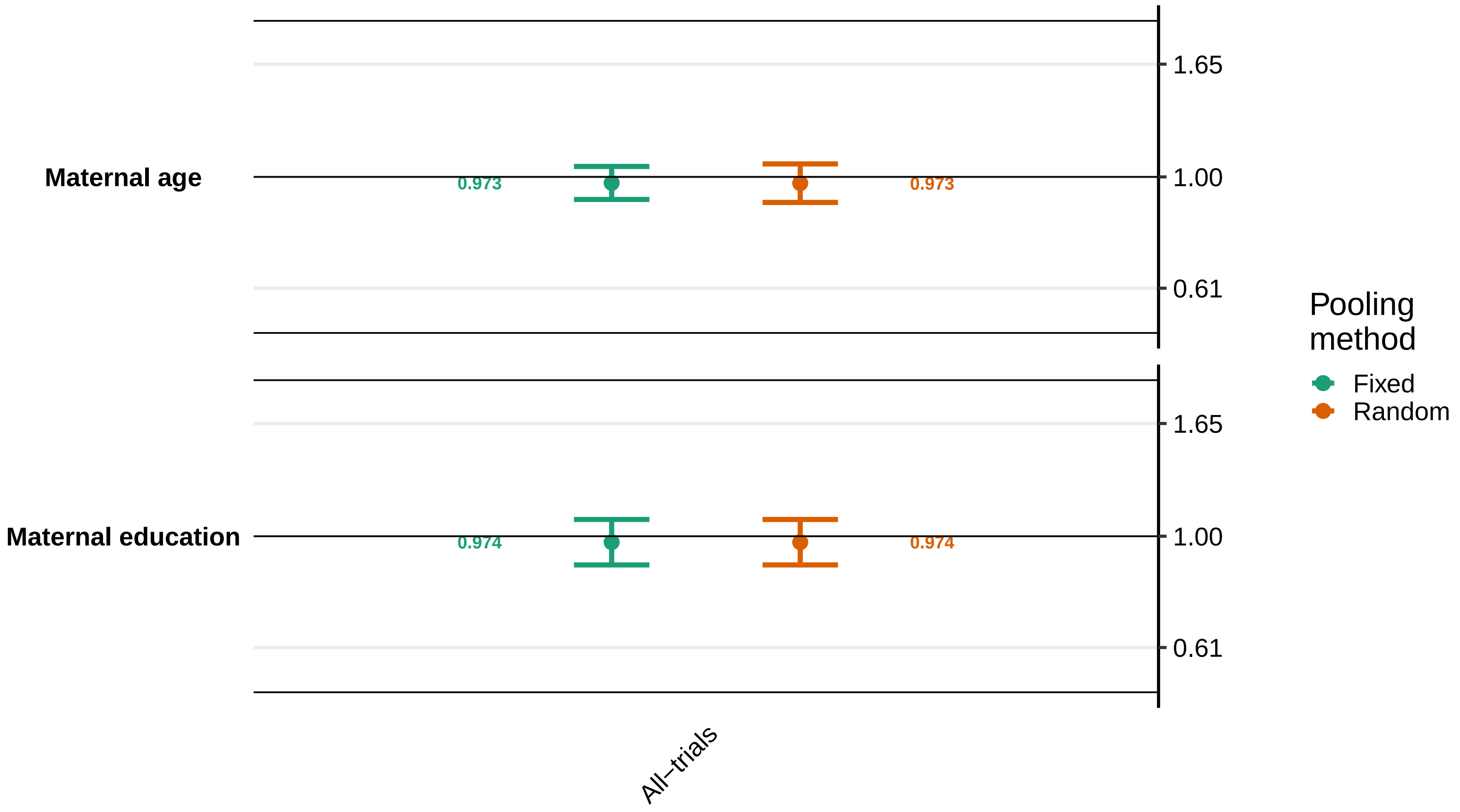

Supplemental figure 8R: Ratio of geometric mean ratios of retinol concentration

#### 8R2: By child effect modifiers

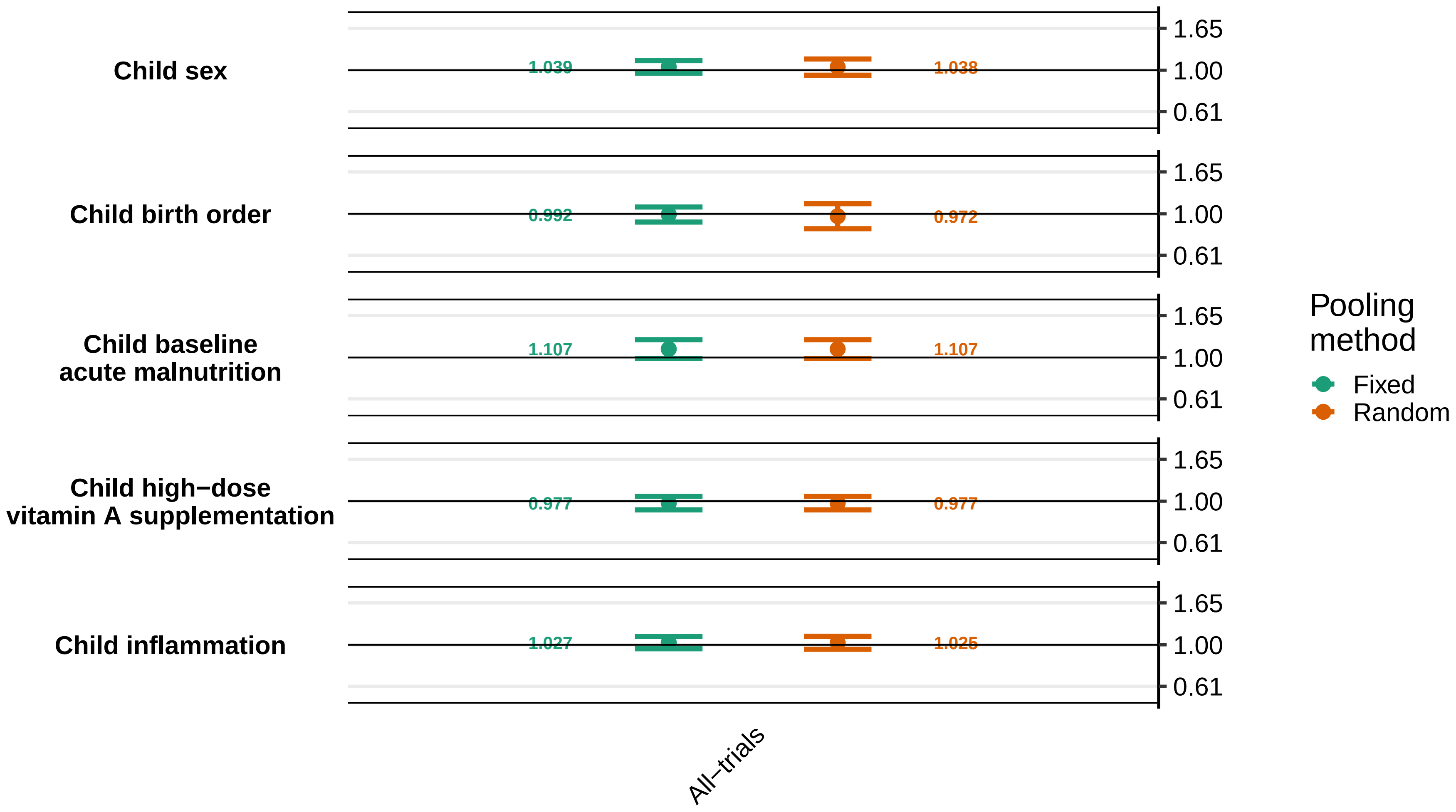

**Supplemental figure 8R: Ratio of geometric mean ratios of retinol concentration**

#### 8R3: By household effect modifiers

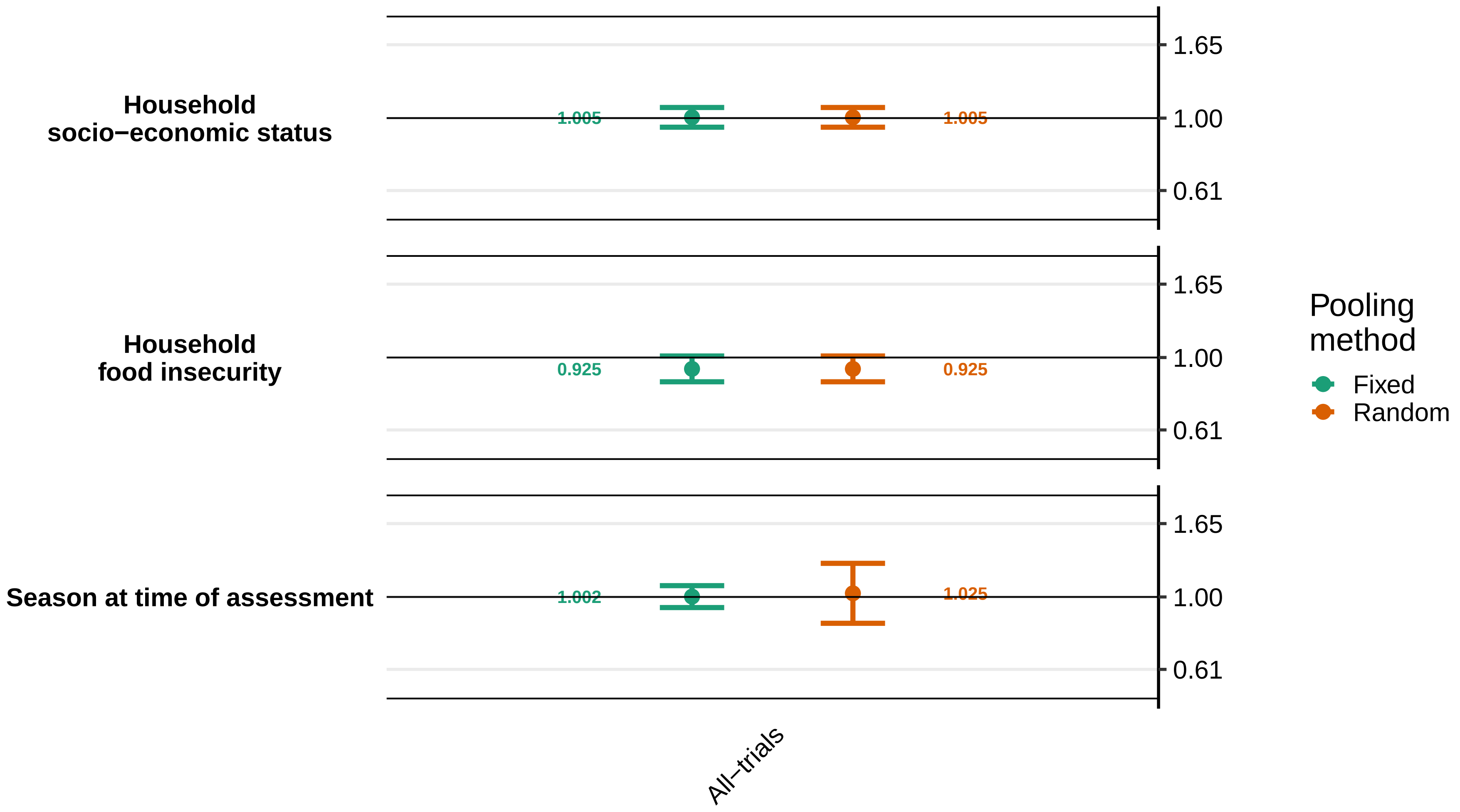

#### Supplemental figure 8S: Ratio of low vitamin A (retinol < 0.70 µmol/L) prevalence ratios

##### 8S1: By maternal effect modifiers (insufficient comparisons)

**Supplemental figure 8S: Ratio of low vitamin A (retinol < 0.70 μmol/L) prevalence ratios**

#### 8S2: By child effect modifiers (insufficient comparisons)

**Supplemental figure 8S: Ratio of low vitamin A (retinol < 0.70 µmol/L) prevalence ratios**

#### 8S3: By household effect modifiers (insufficient comparisons)

#### Supplemental figure 8T: Difference in low vitamin A (retinol $< 0.70$ µmol/L) prevalence differences

##### 8T1: By maternal effect modifiers (insufficient comparisons)

**Supplemental figure 8T: Difference in low vitamin A (retinol $< 0.70$ µmol/L) prevalence differences**

#### 8T2: By child effect modifiers (insufficient comparisons)

**Supplemental figure 8T: Difference in low vitamin A (retinol < 0.70 µmol/L) prevalence differences**

#### 8T3: By household effect modifiers (insufficient comparisons)

#### Supplemental figure 8U: Ratio of marginal vitamin A (retinol < 1.05 µmol/L) prevalence ratios

##### 8U1: By maternal effect modifiers

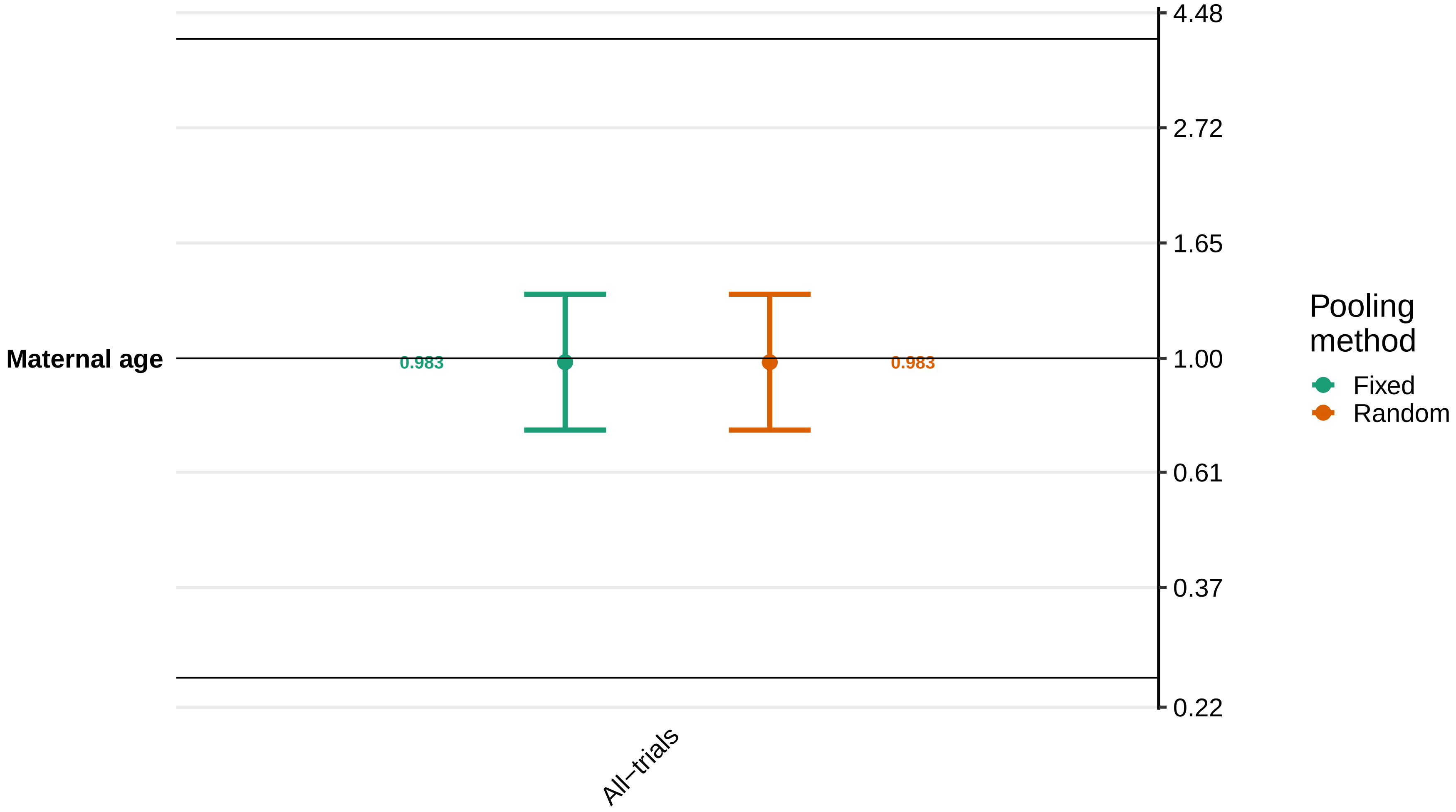

Supplemental figure 8U: Ratio of marginal vitamin A (retinol < 1.05 µmol/L) prevalence ratios

#### 8U2: By child effect modifiers

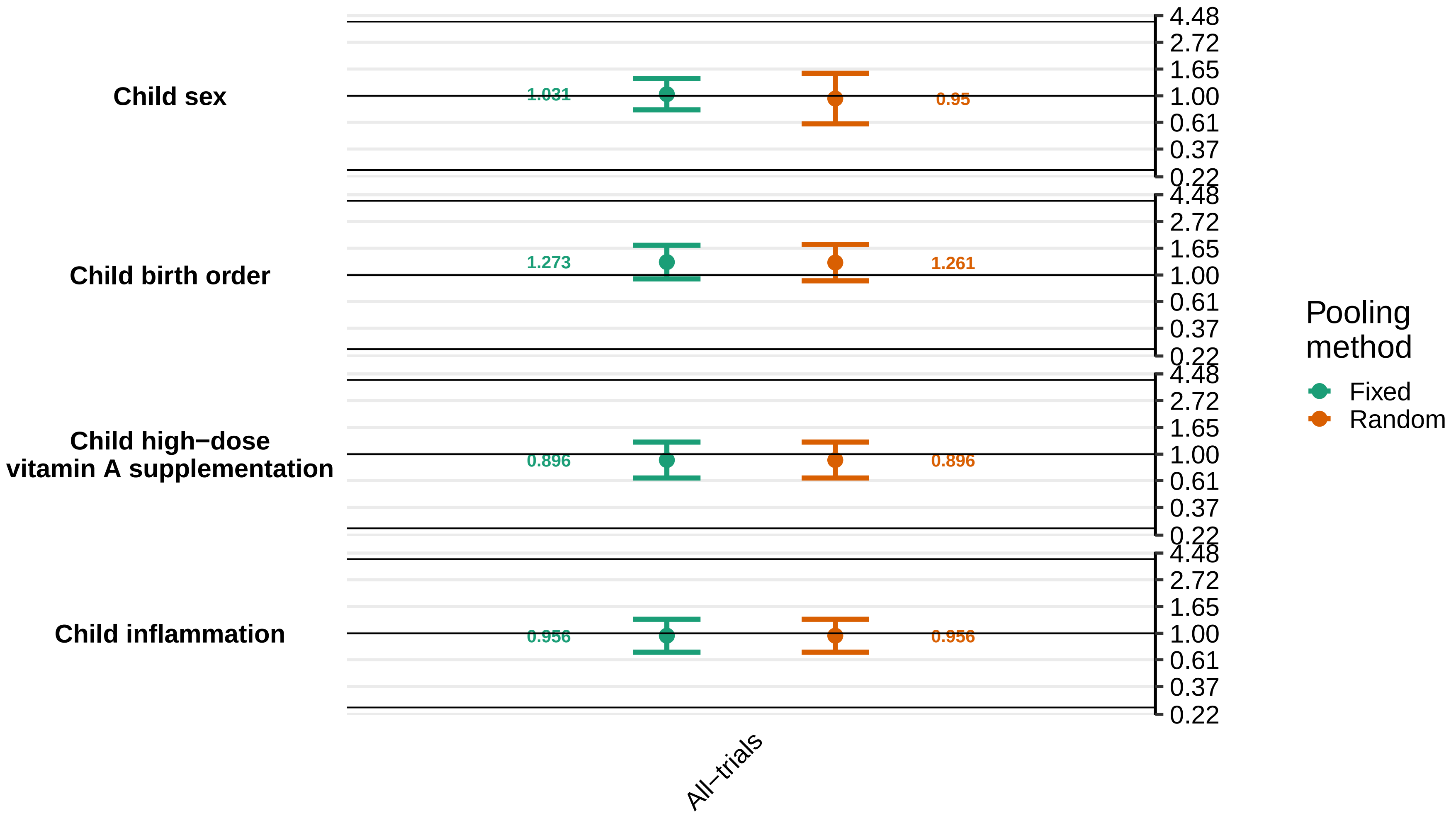

Supplemental figure 8U: Ratio of marginal vitamin A (retinol < 1.05 µmol/L) prevalence ratios

#### 8U3: By household effect modifiers

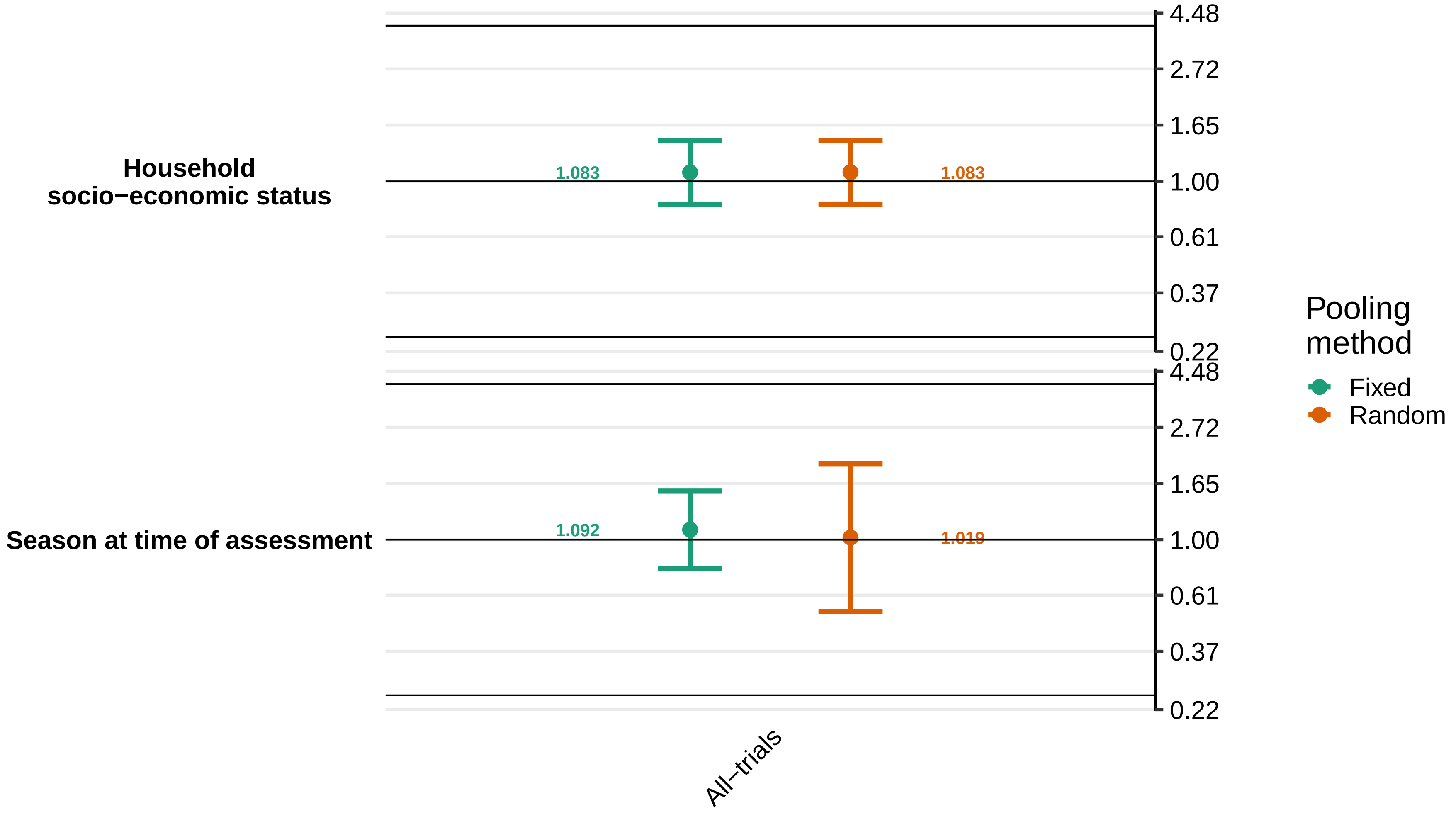

#### Supplemental figure 8V: Difference in marginal vitamin A (retinol < 1.05 µmol/L) prevalence differences

##### 8V1: By maternal effect modifiers

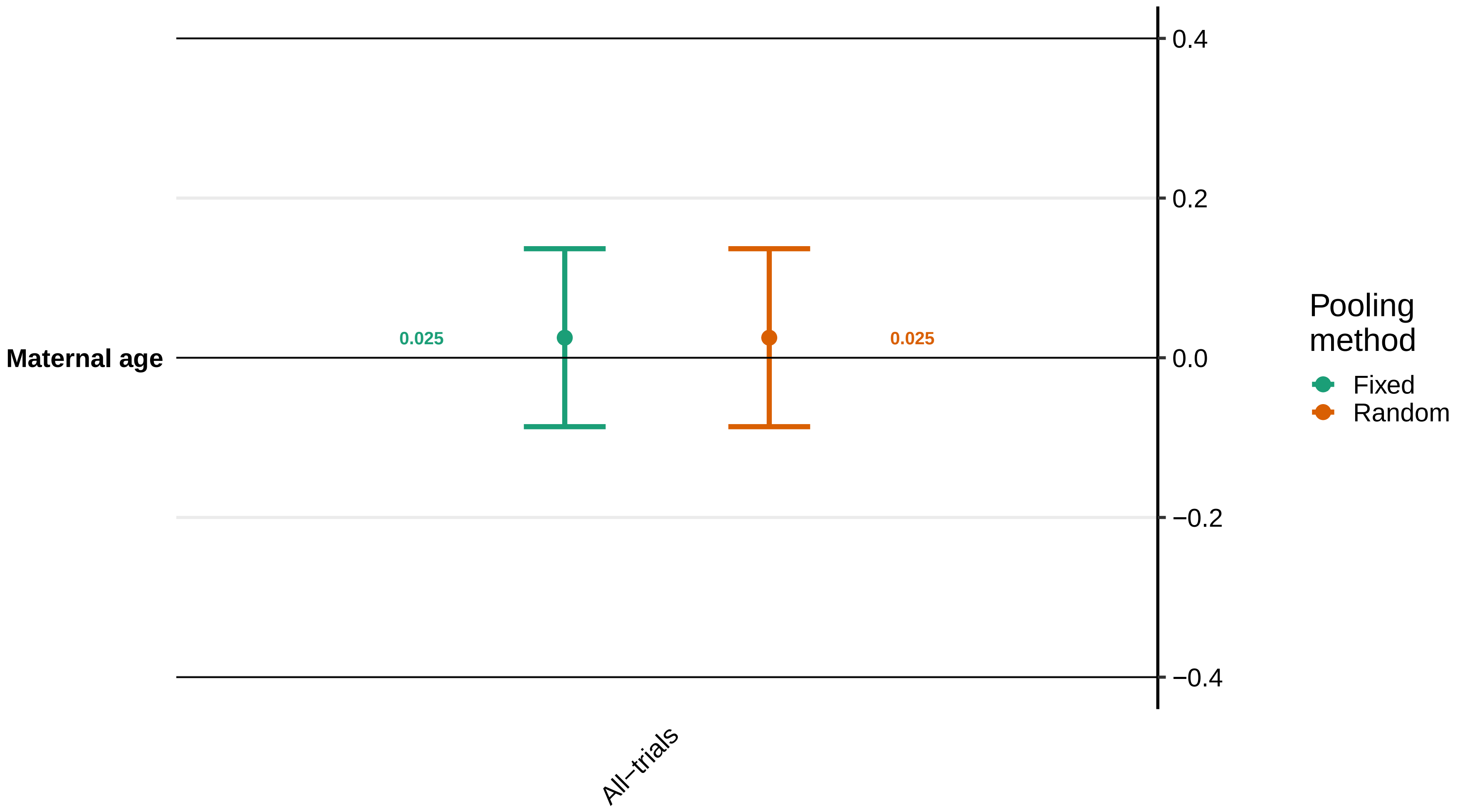

Supplemental figure 8V: Difference in marginal vitamin A (retinol < 1.05 µmol/L) prevalence differences

8V2: By child effect modifiers

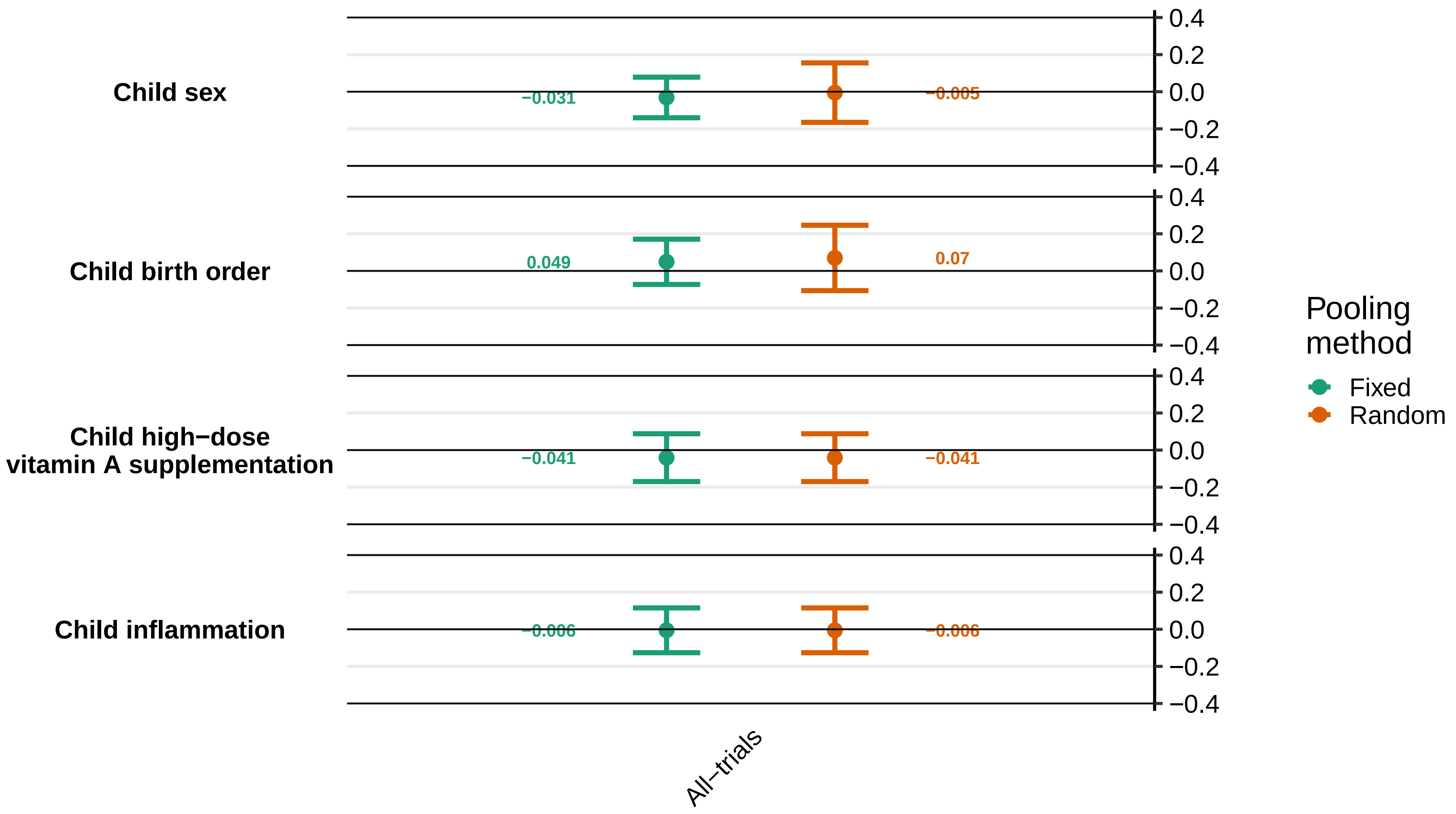

Supplemental figure 8V: Difference in marginal vitamin A (retinol < 1.05 µmol/L) prevalence differences

#### 8V3: By household effect modifiers

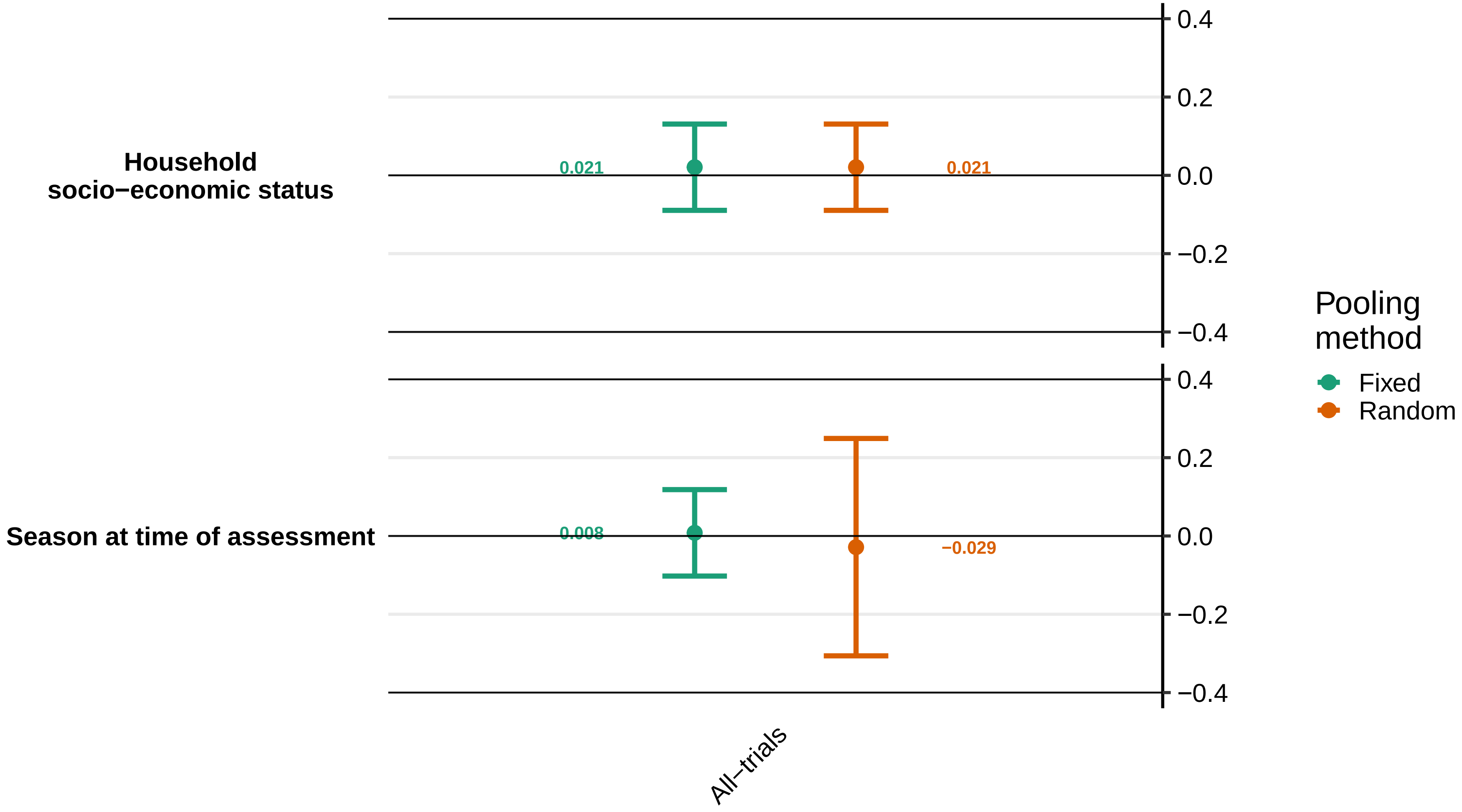

#### Supplemental figure 8W: Ratio of geometric mean ratio of retinol binding protein concentrations

##### 8W1: By maternal effect modifiers

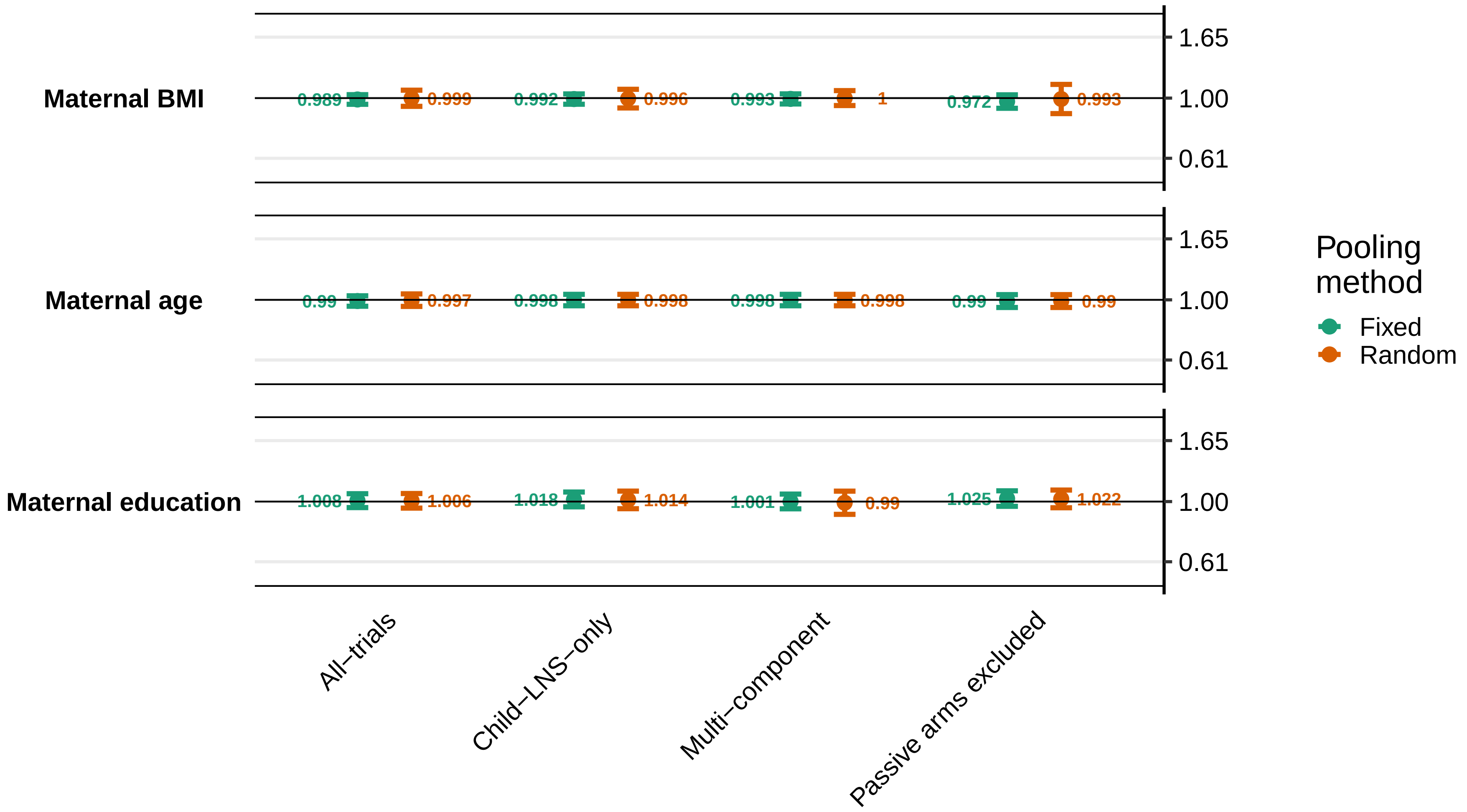

Supplemental figure 8W: Ratio of geometric mean ratio of retinol binding protein concentrations

#### 8W2: By child effect modifiers

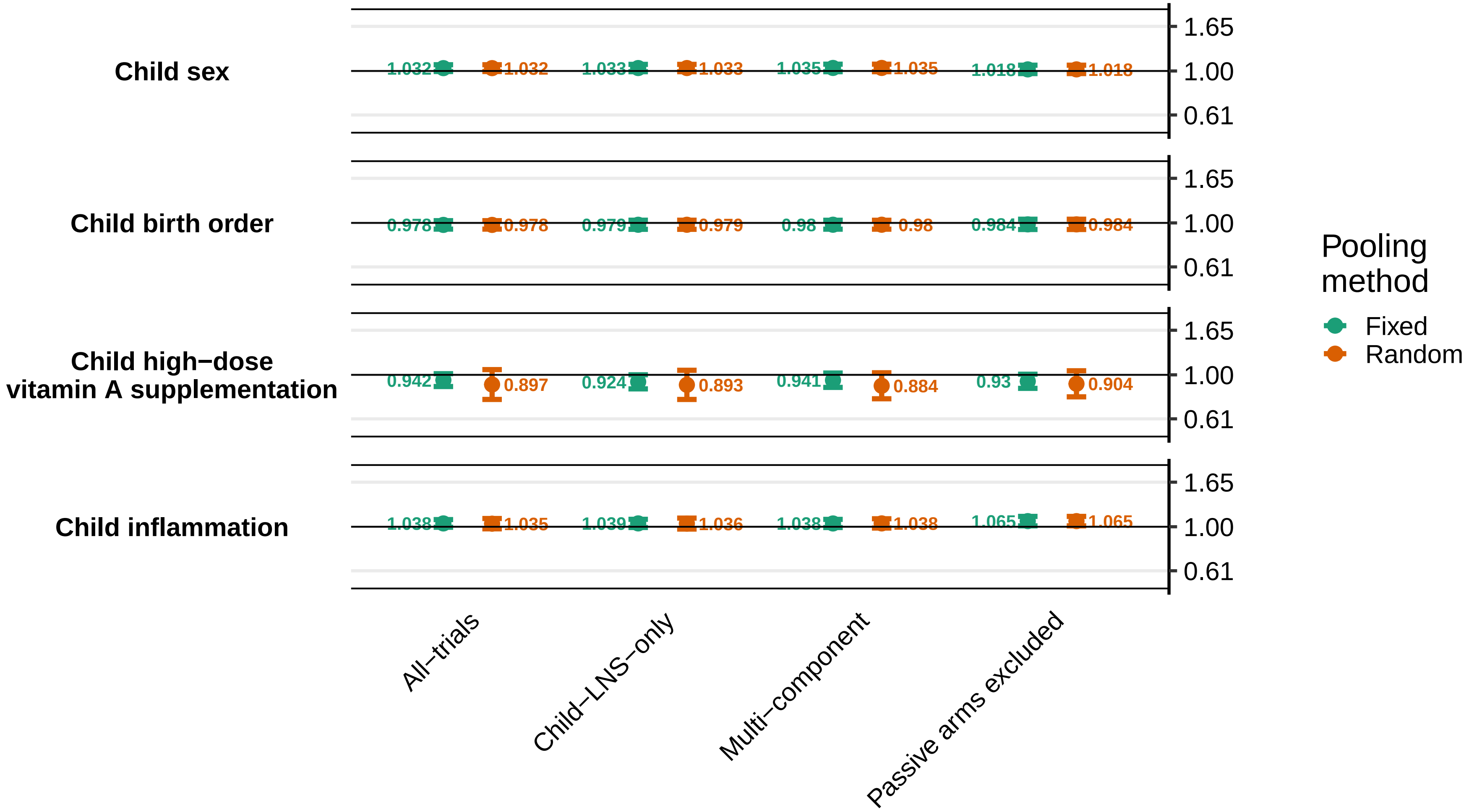

Supplemental figure 8W: Ratio of geometric mean ratio of retinol binding protein concentrations

#### 8W3: By household effect modifiers

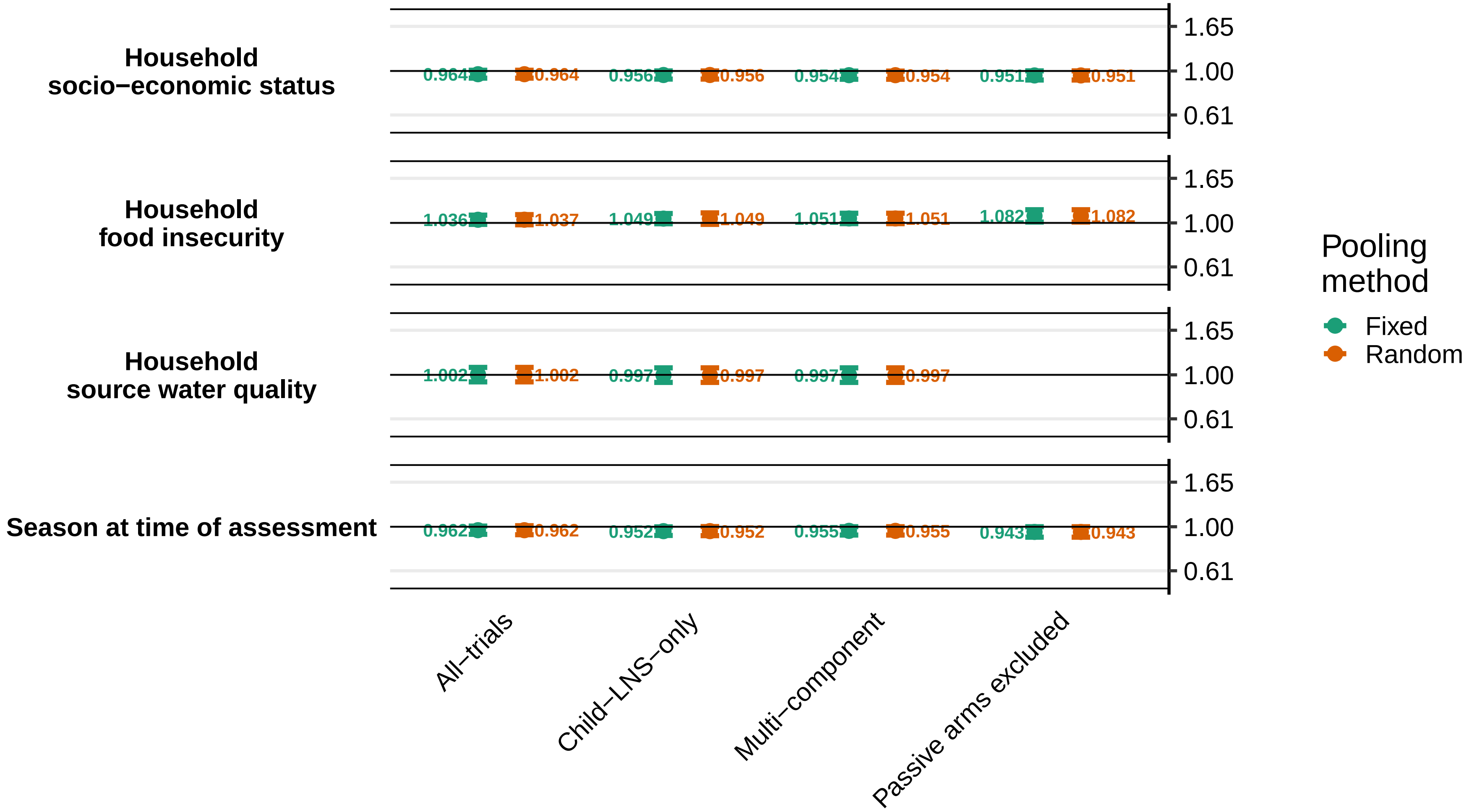

#### Supplemental figure 8X: Ratio of Low vitamin A status (RBP $< 0.70$ µmol/L) prevalence ratios

##### 8X1: By maternal effect modifiers (insufficient comparisons)

**Supplemental figure 8X: Ratio of Low vitamin A status (RBP $<$ 0.70 µmol/L) prevalence ratios**

#### 8X2: By child effect modifiers (insufficient comparisons)

**Supplemental figure 8X: Ratio of Low vitamin A status (RBP $< 0.70$ µmol/L) prevalence ratios**

#### 8X3: By household effect modifiers (insufficient comparisons)

#### Supplemental figure 8Y: Difference in low vitamin A status (RBP < 0.70 µmol/L) prevalence differences

##### 8Y1: By maternal effect modifiers (insufficient comparisons)

**Supplemental figure 8Y: Difference in low vitamin A status (RBP $< 0.70$ µmol/L) prevalence differences**

#### 8Y2: By child effect modifiers (insufficient comparisons)

**Supplemental figure 8Y: Difference in low vitamin A status (RBP < 0.70 µmol/L) prevalence differences**

#### 8Y3: By household effect modifiers (insufficient comparisons)

**Supplemental figure 8Z: Ratio of marginal vitamin A status (RBP < 1.05 µmol/L) prevalence ratios**

**8Z1: By maternal effect modifiers**

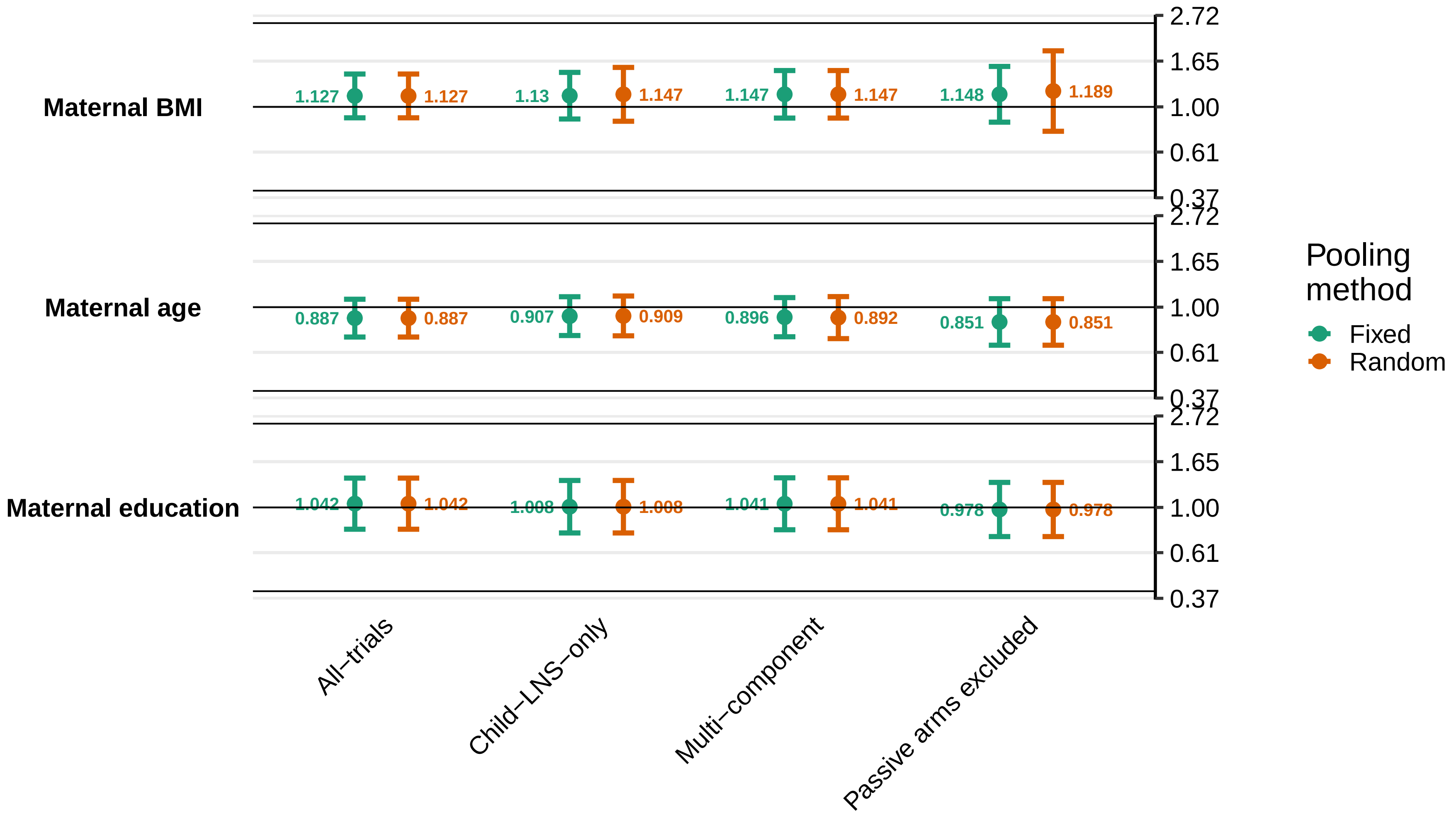

**Supplemental figure 8Z: Ratio of marginal vitamin A status (RBP < 1.05 μmol/L) prevalence ratios**

#### 8Z2: By child effect modifiers

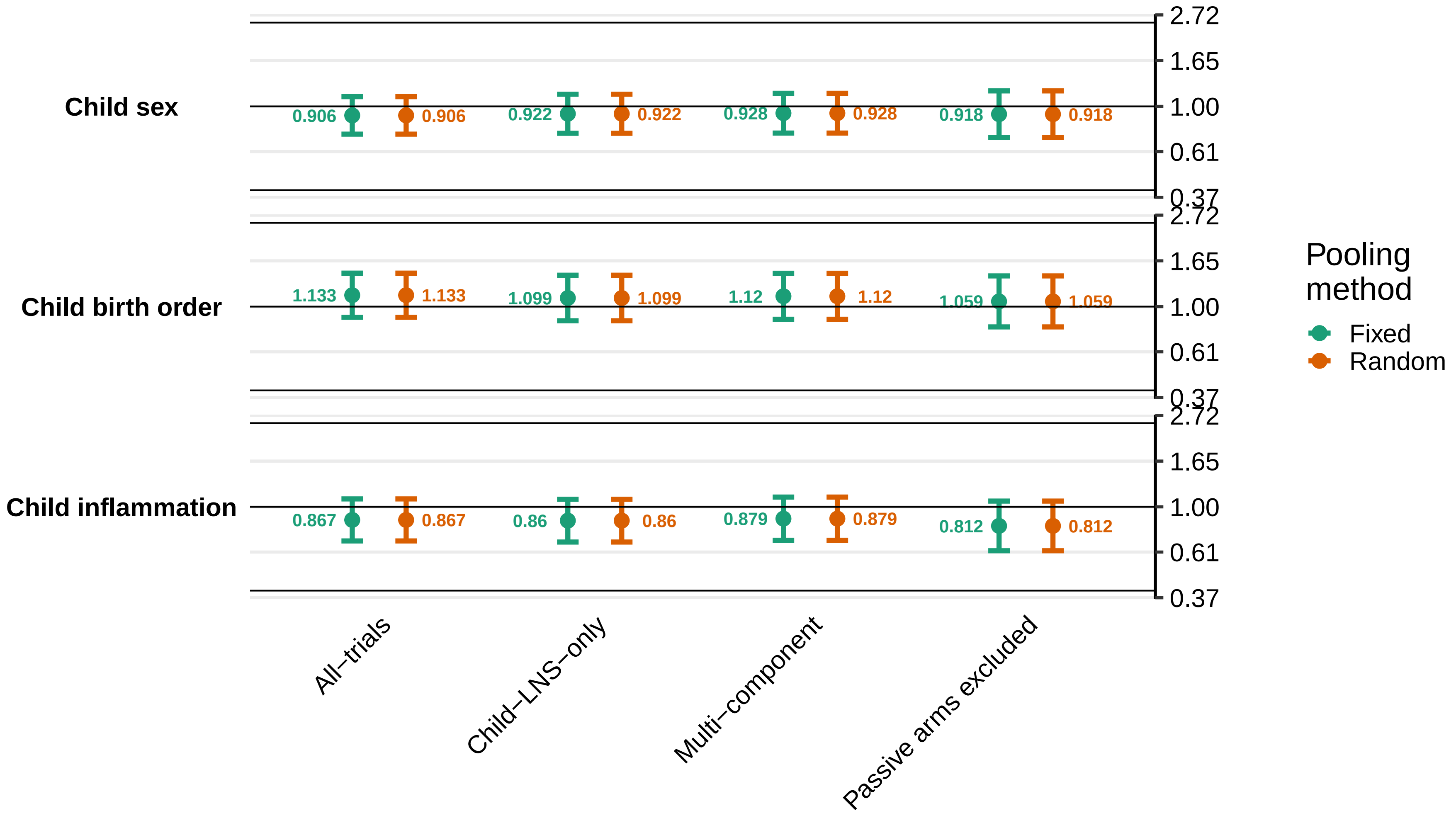

Supplemental figure 8Z: Ratio of marginal vitamin A status (RBP < 1.05 µmol/L) prevalence ratios

#### 8Z3: By household effect modifiers

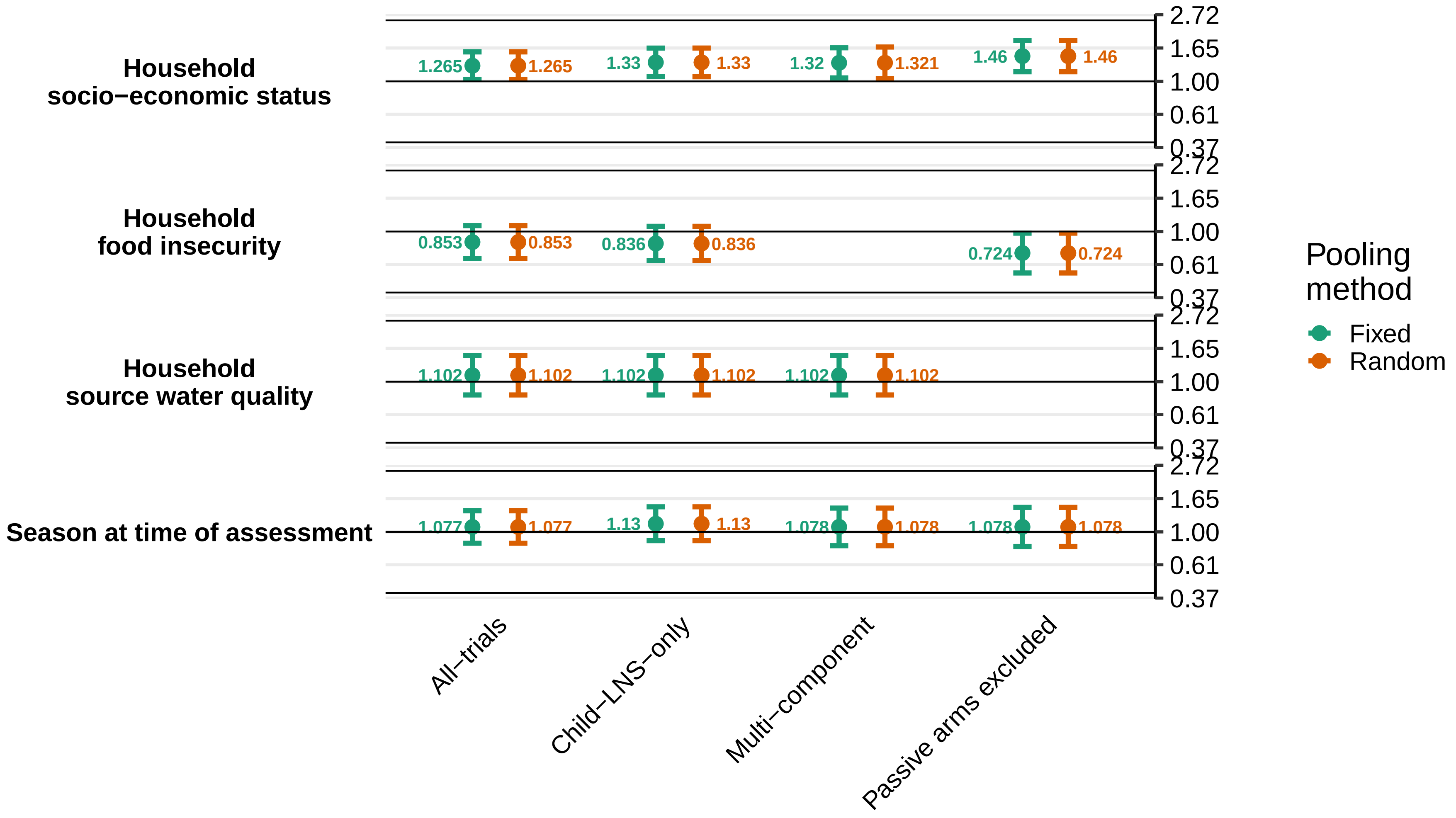

**Supplemental figure 8AA: Difference in marginal vitamin A status (RBP < 1.05 µmol/L) prevalence differences**

**8AA1: By maternal effect modifiers**

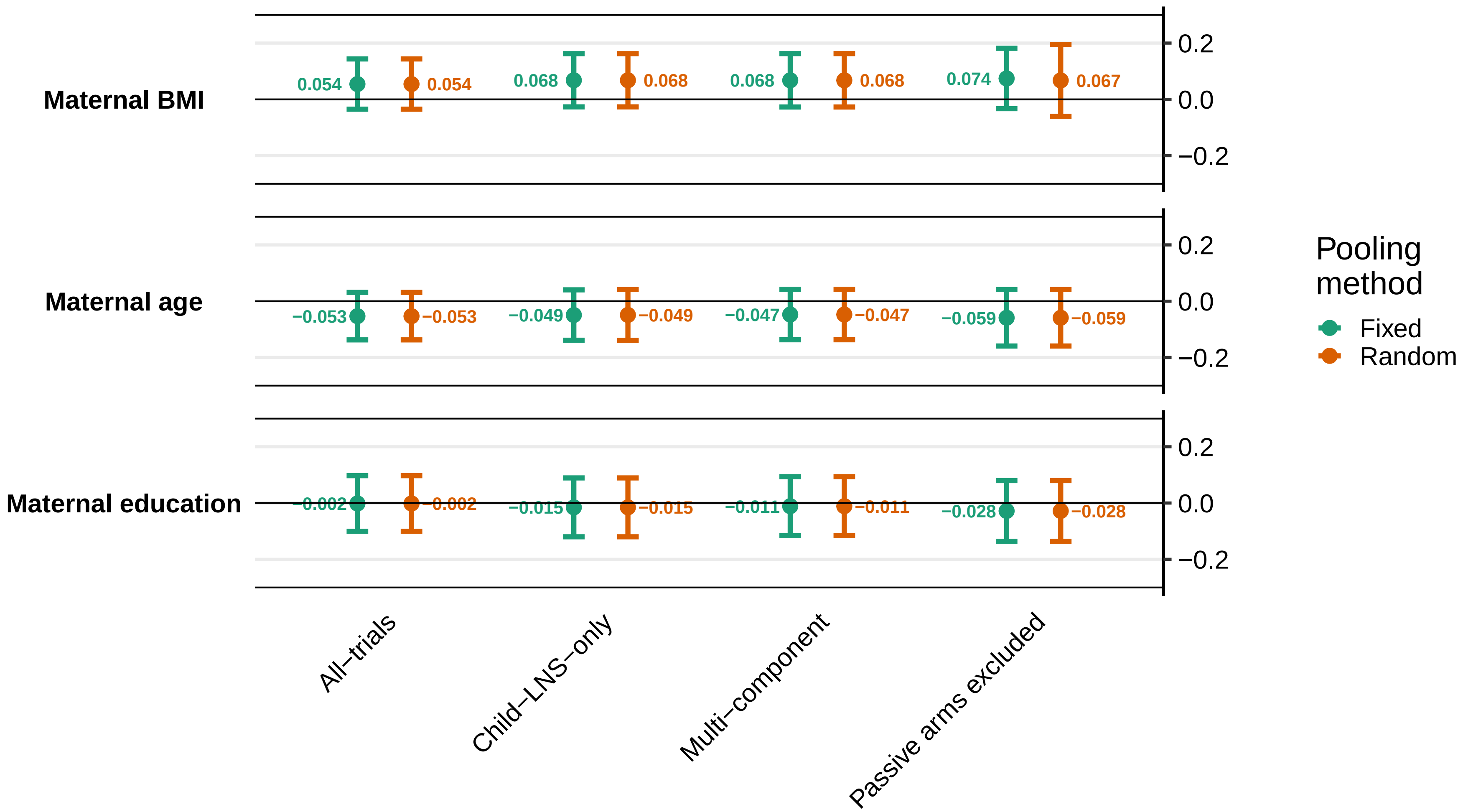

**Supplemental figure 8AA: Difference in marginal vitamin A status (RBP < 1.05 µmol/L) prevalence differences**

**8AA2: By child effect modifiers**

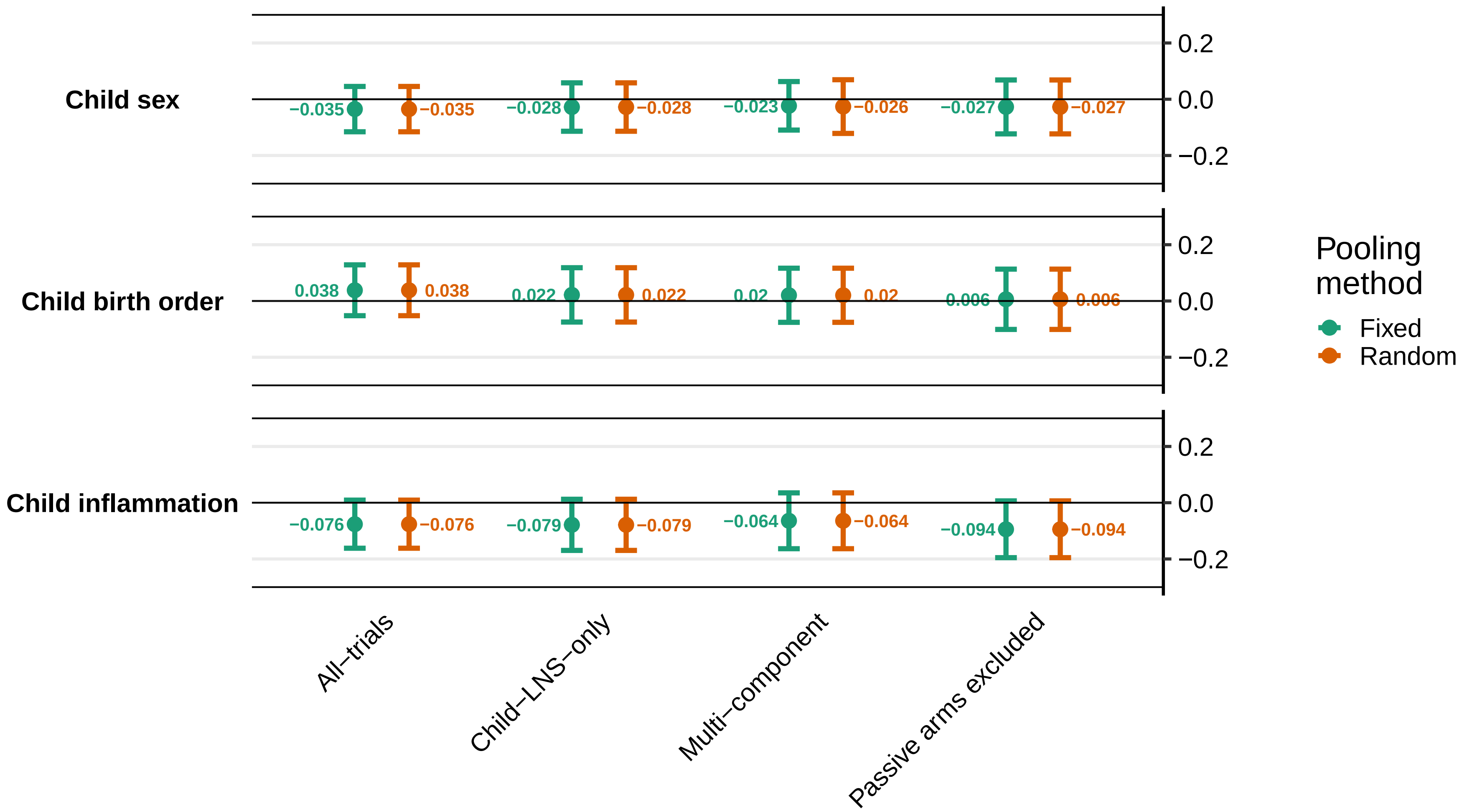

**Supplemental figure 8AA: Difference in marginal vitamin A status (RBP < 1.05 µmol/L) prevalence differences**

#### 8AA3: By household effect modifiers

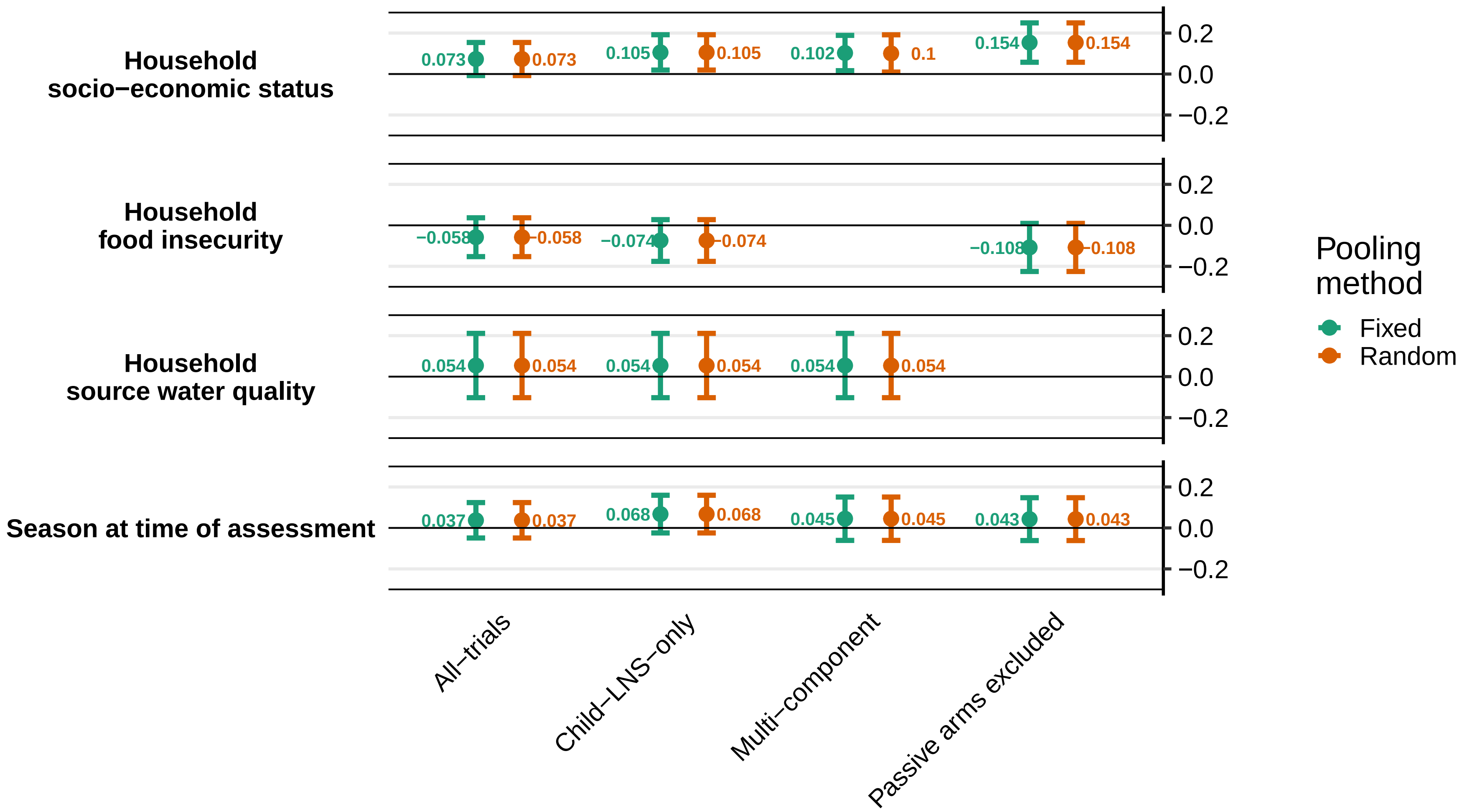
